# Novel antibiotic resistance genes from the hospital effluent are disseminated into the marine environment in Norway

**DOI:** 10.1101/2024.09.23.24313887

**Authors:** Vera Radisic, Manish P. Victor, Didrik H. Grevskott, Nachiket P. Marathe

## Abstract

Hospital effluent comprises feces of many individuals, including patients undergoing antibiotic treatment. Although, hospital effluent is an important source for contamination of the environment with antibiotic resistance genes (ARGs) and pathogens, how hospital effluent in a low resistance setting contributes to antimicrobial resistance (AMR) in the environment is largely understudied. The aim of our study was to understand the microbiota and resistome of hospital effluent, and its role in the spread of AMR in the marine environment in Norway. We further aimed at describing/characterizing novel resistance factors from hospital effluent and the receiving sewage treatment plant (STP). 24-hour composite samples of the hospital effluent and the influent and effluent of the receiving STP were collected at two sampling time-points (February and April 2023) in Bergen city, Norway. Isolation of *Escherichia coli* and *Klebsiella* spp. was performed, using ECC and SCAI plates with cefotaxime, tigecycline or meropenem, followed by antibiotic susceptibility testing, using EUVSEC3 plates. Whole-genome sequencing of selected strains (n=36) and shotgun metagenomics of sewage samples (n=6) were performed, using Illumina NovaSeq. ARGs were identified with USEARCH, and known and novel ARGs were assembled with fARGene. All *E. coli* strains (n=66) were multidrug-resistant (MDR), while 92.3% of the *Klebsiella* spp. strains (n=55) showed MDR phenotype. The sequenced strains carried multiple clinically important ARGs, including carbapenemases such as NDM-5 (n=3) and KPC-3 (n=3). We obtained 238 Gigabases of sequence data from which we identified 676 unique ARGs with >200 ARGs shared across samples. We assembled 1,205 ARGs using fARGene, 365 gene sequences represented novel ARGs (< 90% amino acid (aa) identity). Both known and novel ARGs (n=54) were shared between the hospital effluent and the treated effluent of the receiving STP. We show that hospital effluent in Norway has a high diversity of both known and novel ARGs. Our study demonstrates that hospital effluent is a source of clinically relevant pathogens, as well as known and novel ARGs, reaching the marine environment in Norway through treated sewage.

## 1. Introduction

Antimicrobial resistance (AMR) is an emerging global health problem (WHO, 2017). Pathogens of particular concern to the public health include extended spectrum β-lactamases (ESBL)- and carbapenemase-producing *Enterobacterales* (WHO, 2017). Carbapenems are last resort antibiotics used for treatment of severe infections cause by multidrug-resistant (MDR) pathogens (Sheu et al., 2019; Zhu et al., 2020). Resistance to carbapenems is largely due to the presence of carbapenem hydrolyzing β-lactamases or carbapenemases (Sheu et al., 2019; Tamma & Simner, 2018). The five clinically most relevant carbapenemases are KPC, IMP, VIM, NDM and OXA-48 variants (Bonnin et al., 2021). KPC belongs to Ambler class A, while IMP, VIM and NDM belongs to class B (metallo-β-lactamases (MBLs)) and OXA-48 variants belong to class D of β-lactamases (van Duin & Doi, 2017). MBLs are of particular concern as they hydrolyze most β-lactam antibiotics, and have successfully being disseminated globally (Boyd et al., 2020; Logan & Bonomo, 2016). Although, the use of carbapenems in clinics is low in Norway, the prevalence of carbapenemase-producing clinical strains has increased over the past few years (NORM/NORM-VET, 2023).

Pathogens, antibiotic resistance genes (ARGs) and residues of antibiotics may be introduced into the environment through different routes, such as pollution with untreated sewage from the population, industries and/or hospitals (Stalder et al., 2012). The hospital effluent, in particular, is an important source of clinically relevant pathogens, ARGs and residues of antibiotics reaching the receiving environment (Karkman et al., 2019; Khan et al., 2021; Marathe et al., 2019). Hospital effluent contains fecal material from many individuals undergoing antibiotic treatment, and invariably contains antibiotic resistant bacteria, resistant genes, as well as residues of different antimicrobial compounds (Deguenon et al., 2022; Orias & Perrodin, 2013). Hospital effluents are treated on-site in some countries, but not in Norway (VKM, 2020). How untreated hospital effluent contributes to the spread of AMR in the receiving STP and eventually to the marine environment in Norway is largely unknown.

The aim our study was to understand the microbiota and resistome of hospital effluent and its role in the spread of AMR in the marine environment in Norway. We further aimed at describing/characterizing novel resistance genes from hospital effluent and the receiving sewage treatment plant (STP).

## 2. Materials and Methods

### 2.1 Collection of sewage samples

Composite samples, representing a 24-hour period, from both raw sewage (influent) and treated sewage (effluent) were collected on two occasions (February 7^th^ and April 12^th^, 2023) from the second largest sewage treatment plant (STP), Holen, in Bergen, Norway. This treatment plant, serving 132,000 inhabitants, receives sewage from two hospitals: a 933-bed and a 170-bed hospital. Moreover, composite sewage samples were also collected directly from the sewer line from the Haukeland University hospital in Bergen, on the same two occasions. The sewage samples were transported to the laboratory in sterile containers at 4°C, and analyses were initiated within 6 h after collection.

### 2.2 Isolation and identification of pathogens

The samples were serially diluted ten-fold with sterile saline (0.85% NaCl) before plating on ECC (CHROMagar™, France) chromogenic media and on Simmons Citrate Agar with myo-Inositol (SCAI) (Sigma-Aldrich, Germany) media supplemented with three different antibiotics: 2 mg/L cefotaxime (Sigma-Aldrich, Germany), 0.5 mg/L tigecycline (Glentham Life Sciences, UK), or 0.125 mg/L meropenem (Sigma-Aldrich, Germany). The ECC plates were incubated at 37°C for 20-24 h, while the SCAI plates were incubated 37°C for 24-48 h. *E. coli* CCUG 17620 and *K. pneumoniae* CCUG 225T were included as negative controls for verifying the ECC and SCAI plates, respectively. From each sample, 10-15 isolated colonies were picked depending on growth, restreaked and incubated at 37°C overnight. Identification of strains was performed, using matrix-assisted laser desorption ionization-time of flight mass spectrometry (MALDI-TOF MS) (Bruker Daltonics, Germany) at the Institute of Marine Research (IMR, Norway). Subsequently, confirmed strains were stored at –80°C in Mueller-Hinton (MH) broth (Oxoid, UK) with respective antibiotics and 20% glycerol until further use.

### 2.3 Antibiotic sensitivity testing

The resistance profile of 66 *E. coli* and 55 *Klebsiella* spp. strains were determined against 15 antibiotics, using a broth microdilution assay with Sensititre® EUVSEC3 plates (Thermo Scientific, USA), following manufacturer’s protocol. The plates were incubated at 37°C for 20-22 h. The strains were defined as ‘susceptible’ or ‘resistant’, according to the EUCAST clinical breakpoints tables v.13.1 (EUCAST, 2023). For certain antibiotics, like azithromycin, nalidixic acid, sulfamethoxazole and tetracyclines, not having defined breakpoints in EUCAST, resistance was determined if growth was observed in the highest antibiotic concentration tested. *E. coli* CCUG 17620 was included as negative controls, while *E. coli* CCUG 73937 was included as positive controls (Grevskott et al., 2020).

### 2.4 DNA extraction and sequencing

Based on phenotypic resistance profiles, 19 presumptive *E. coli* and 17 presumptive *K. pneumoniae* strains were selected for whole genome sequencing (WGS). These isolates were grown overnight on MH agar (Oxoid, UK) containing 2 mg/L cefotaxime (Sigma-Aldrich, Germany), 0.5 mg/L tigecycline (Glentham Life Sciences, UK), or 0.125 mg/L meropenem (Sigma-Aldrich, Germany), based on the resistance profiles, at 37°C. Genomic DNA was extracted from the strains, using the DNeasy Blood and Tissue kit (Qiagen, Germany) following the manufacturer’s protocol. The extracted DNA was quantified, using Qubit™ 2.0 Fluorometer with the dsDNA BR (Broad-Range) kit and NanoDrop™ 2000 Spectrophotometer (Thermo Scientific, USA) assay. Sequencing libraries was prepared, using Nextera DNA Flex Library Prep kit (Illumina, USA). Sequencing was performed, using Illumina NovaSeq platform (Illumina, USA), with 2 × 150 bp on a S4 flow cell, at Eurofins Genomics A/S Germany.

### 2.5 Genome assembly and sequencing analysis

Obtained raw reads from the Illumina NovaSeq sequencing were quality trimmed and assembled as previously described (Radisic et al., 2020), follow by annotation using National Center for Biotechnology Information (NCBI) Prokaryotic Genomes Automatic Annotation Pipeline (PGAAP) (Tatusova et al., 2016). Antibiotic resistance genes (ARGs) and sequence types (STs) of *Klebsiella* spp. were screened, using Kleborate v.2.1.0 (Lam et al., 2021), while the PubMLST database (https://pubmlst.org/bigsdb?db=pubmlst_escherichia_seqdef) was used for identification of STs among *E. coli* strains. ResFinder v.4.1 (Bortolaia et al., 2020) and the comprehensive antibiotic resistance database (CARD) v.3.1.4 (Alcock et al., 2020) were used for further identification of ARGs among strains of *Klebsiella* spp. and *E. coli*. PlasmidFinder v.2.1 was used for identification of plasmid replicons (Carattoli et al., 2014).

### 2.6 Sampling, DNA extraction and shotgun sequencing

From each sample, 30-50 mL was vacuum-filtered through S-Pak filters (pore size 0.22 μm) (Millipore, USA), after which the filters were transferred to 50 mL Falcon tubes and stored at –20°C until further use. Genomic DNA was extracted from the filters, using DNeasy PowerWater kit (Qiagen, Germany) following the manufacturer’s protocol. The extracted DNA was quantified, using dsDNA BR (Broad-Range) assay kit on the Qubit™ 2.0 Fluorometer and NanoDrop™ 2000 Spectrophotometer (Thermo Scientific, USA) assay. Sequencing libraries for the shotgun metagenomics were prepared, using Nextera DNA Flex Library Prep kit (Illumina, USA) and sequencing was performed, using Illumina NovaSeq platform (Illumina, USA), with 2 × 150 bp chemistry on a S4 flow cell, at the Norwegian Sequencing Centre (Oslo University Hospital, Ullevål, Oslo, Norway).

### 2.7 Sequence analysis of shotgun metagenomic data

The sequences were quality-trimmed with Trim Galore v.0.6.10 (http://www.bioinformatics.babraham.ac.uk/projects/trim_galore/), using the default settings; trim_galore --quality 30 --phred33 --trim_n --cores 50 --paired seqForward.fq seqReverse.fq with a phred quality score of 33 and a maximum error rate of 0.5. The quality-processed reads from the metagenomes were mapped against protein sequences from a high-quality and manually curated database of ARGs, mobile genetic elements (MGEs), biocide/metal resistance genes (BMRGs) and virulence genes, using USEARCH v.usearch11.0.667_i86linux64 with the default settings; usearch -usearch_global reads.fastq - db DB_seq.fasta -id 0.9 -query_cov 1.0 -blast6out hits.b6 -strand plus -maxaccepts 1. The database for BMRGs were obtained from Pal et al. (2014), virulence factors (experimentally verified) from Chen et al. (2016), MGEs from Pärnänen et al. (2018), while for the ARGs database we used a combination of the NCBI AMR database https://www.ncbi.nlm.nih.gov/pathogens/antimicrobial-resistance/, CARD (Alcock et al., 2020), and Antibiotic Resistance Gene-ANNOTation (ARG-ANNOT) (Gupta et al., 2014). For quantitative taxonomic distribution, quality-filtered shotgun reads were used as input to extract the reads corresponding to small subunit (SSU) 16S bacterial ribosomal RNA (rRNA) genes from the metagenomes and assigned them to different taxonomic groups, using Metaxa2 v.2.2.3 (Bengtsson-Palme et al., 2015) with the default settings; metaxa2 -o metaxa_out -1 (sequence1) -2 (sequence2) -f fastq -g ssu --mode metagenome –usearch v11.0.667_i86linux64 --usearch_bin usearch11 --cpu 25 --table T --reltax T and metaxa2_ttt -i (taxonomy_text_file) -o metaxa_ttt_out. fARGene was used to predict probable ARGs for β-lactamases (class A, subclass B1-B3, class C and D1-2), quinolones, tetracyclines, macrolides, and aminoglycosides. Known and novel ARGs were assembled from shotgun sequence data, using fARGene as previously described by Berglund et al. (2019) with the default settings; Fargene --infiles (sequence1) (sequence2) --hmm-model (model_name) -- meta --output (folder_out) --force --processes (cpu) --rerun --amino-dir class_a_out tmpdir -- orf-finder’. ARGs in different samples were clustered at 100/95/90% amino acid (aa) sequence similarity together with previously characterized ARGs with USEARCH. The query (obtained fARGenes) was blasted against nucleotide non-redundant (nr) database (https://ftp.ncbi.nlm.nih.gov/blast/db/FASTA/) with query coverage of > 90% and E-value = 10^−5^, using diamond v.2.1.9 (https://github.com/bbuchfink/diamond). Novel ARGs were identified, using cutoff < 90% aa identity (Inda-Díaz et al., 2023). The number of shared ARGs, both known and novel, across samples were clustered at 100% identity for each sample and then presented in a Venn diagram, using pyvenn (https://github.com/tctianchi/pyvenn).

## 3. Results

### 3.1 Strains obtained from media with antibiotics

Most *Klebsiella* spp. isolates and *E. coli* isolates were obtained from the hospital effluent samples (n=31; n=31), followed by STP influent samples (n=19; n=21) and STP treated effluent samples (n=10; n=14). In total, 130 isolates were obtained on both ECC plates with antibiotics (n=66) and SCAI plates with antibiotics (n=64) and identified with MALDI-TOF MS. Most of the obtained isolates on the SCAI plates (n=64) were identified as *K. pneumoniae* (n=34, 53.1%), followed by *K. oxytoca* (n=21, 32.8%), *Raoultella ornithinolytica* (n=5, 7.8%), *Acinetobacter bereziniae* (n=2, 3.1%), *A. baumannii* (n=1, 1.6%) and *Citrobacter freundii* (n=1, 1.6%). Isolates identified as other genera than *K. pneumoniae, K. oxytoca* and *E. coli* were not included in further analysis.

### 3.2 Resistance rates in *E. coli* and *Klebsiella* spp. strains

We obtained a total of 64 *E. coli* isolates on cefotaxime-containing plates and two *E. coli* isolates on meropenem-containing plates. All *E. coli* isolates (n=64) on cefotaxime-containing plates were multidrug-resistant (MDR), showing resistance against at least three different classes of antibiotics (Supplementary Table S1). *E. coli* isolates obtained on cefotaxime-containing plates from the hospital effluent samples (n=31) showed highest resistance against ampicillin (100%), ceftazidime (100%), nalidixic acid (71%) and ciprofloxacin (71%). Colistin resistance was detected in one *E. coli* isolate (3.2%). Further, *E. coli* isolates (n=20) obtained on cefotaxime-containing plates from STP influent samples showed highest resistance against ampicillin (100%), trimethoprim (81%) and sulfamethoxazole (76%). Highest resistance against ampicillin (100%), nalidixic acid (79%), ceftazidime (45%) and tetracycline (45%) was observed in *E. coli* isolates (n=13) on cefotaxime-containing plates from STP effluent samples. *E. coli* isolates (n=2), obtained from both STP influent (n=1) and STP effluent samples (n=1) on meropenem-containing plates, were MDR (Supplementary Table S1). Several *E. coli* isolates from hospital effluent (February: n=3; April: n=5) and STP influent (February: n=4) samples on cefotaxime-containing plates displaying reduced susceptibility against meropenem. No resistance was observed against tigecycline in these isolates.

On the cefotaxime-containing plates we obtained a total of 31 *Klebsiella* spp. isolates (n=21 hospital effluent; n=10 STP influent; n=10 STP effluent). All *Klebsiella* spp. isolates on cefotaxime-resistant were MDR (Supplementary Table S1). *Klebsiella* spp. isolates (n=21) obtained on cefotaxime-containing plates from the hospital effluent samples showed highest resistance against ampicillin (100%), cefotaxime (100%), nalidixic acid (100%) and ciprofloxacin (100%). Resistance against colistin and tigecycline was observed in 9.5% and 57.1% of the isolates, respectively. Further, *Klebsiella* spp. isolates (n=10) obtained on cefotaxime-containing plates from STP influent samples showed highest resistance against ampicillin (100%), cefotaxime (100%), ceftazidime (100%) and trimethoprim (100%). Highest resistance against ampicillin (100%), cefotaxime (100%), ceftazidime (100%), and sulfamethoxazole (100%) was observed in *Klebsiella* spp. isolates (n=10) on the cefotaxime-containing plates from STP effluent samples. During the two-sampling time-points no resistance against meropenem or amikacin was detected among these *Klebsiella* spp. isolates.

We further detected 11 *K. pneumoniae* isolates (n=7 hospital effluent; n=4 STP influent) on tigecycline-containing plates and three *K. oxytoca* isolates (n=3 hospital effluent) on meropenem-containing plates. Most tigecycline-resistant *K. pneumoniae* isolates (91.7%) were MDR and showed resistance against third generation cephalosporins, such as cefotaxime (63.6%) and ceftazidime (63.6%) (Supplementary Table S1). None of the tigecycline-resistant *K. pneumoniae* isolates were resistant to meropenem, amikacin or colistin. Three presumptive *K. oxytoca* isolates obtained on meropenem-containing plates showed intermediate resistance for meropenem (Supplementary Table S1).

### 3.3 Genome sequence and STs of sequenced strains of *E. coli* and *Klebsiella* spp

A complete overview of the genome sequence statistics is presented in Supplementary Table S2. The total length ranged from 5,857,362 bp - 6,384,740 bp and 4,590,194 bp - 5,361,939 bp for *Klebsiella* spp. strains and *E. coli* strains, respectively. Six *Klebsiella* spp. strains which were initially identified as *K. oxytoca* by MALDI TOF MS were after sequencing re-identified as either *K. michiganensis* (n=3) (K11-303, K11-314 and K11-325) or *K. pneumoniae* (n=3) (K10-M302, K10-M304 and K10-M306) with Kleborate v.2.1.0. Additionally, four *K. pneumoniae* strains (K10-T302, K10-T303, K10-T306 and K10-321) were re-identified as *K. michiganensis*. A high ST diversity was observed among the sequenced *E. coli* strains, including pathogenic ST131 and ST38. The most common ST among the sequenced *K. pneumoniae* strains was ST307 (n=6), followed by ST2947 (n=4) (Supplementary Table S2).

### 3.4 Clinically important ARGs detected in *E. coli* and *Klebsiella* spp. strains

Acquired ARGs conferring resistance against tetracyclines, aminoglycosides, sulfonamides and quinolones were detected in sequenced *Klebsiella* spp. and *E. coli* strains (Supplementary Table S3). In addition, different β-lactamase genes with *bla*_OXA-1_ (52.65%) as most dominant, as well as carbapenemases, such as NDM-5 (n=3) and KPC-3 (n=3), were detected among the sequenced strains. Four *K. pneumoniae* strains (K11-304, K11-305, K11-327 and K11-329) with MIC of ≥ 2μg/mL for colistin carried mutations in the *mgrB* gene, contributing to a truncated protein of only 24 aa instead of 47 aa long protein.

### 3.5 Shotgun sequencing

In total, 238 Gigabases of sequence data was generated from the shotgun metagenomic sequencing of the hospital effluent samples and STP influent and STP effluent samples, representing 2.1×10^11^ quality filtered reads from six samples. In the hospital effluent samples, 109,987 reads (0.000053% of the total reads from hospital effluent samples) matched to ARGs, while 39,737 reads (0.000019% of the total reads from STP influent samples) and 45,069 reads (0.000022% of the total reads from STP effluent samples) matched to ARGs in the STP influent and STP effluent samples, respectively. All together, these accounted for between 0.064 (STP effluent from February) to 0.376 (hospital effluent from April) copies of ARGs per 16S rRNA gene. We detected 676 unique ARGs in hospital effluent samples, as well as 517 unique ARGs in STP influent and 567 unique ARGs in STP effluent samples, respectively (Supplementary Table S4). These ARGs conferred resistance to multiple classes of antibiotics, including glycylcyclines, polymyxins, glycopeptides and oxazolidinones.

### 3.6 Bacterial diversity

A total of 28 bacterial phyla were detected across all samples, with *Pseudomonadota* as most dominant followed by *Bacillota* and *Bacteroidota* for most of the samples (Figure 1). Within *Pseudomonadota, β-proteobacteria* was most dominant class followed by *γ-proteobacteria* for most samples. At genus level, *Acinetobacter* was found to be the dominant in the majority of the samples. For STP influent samples collected in February, *Bacillota* dominated followed by *Pseudomonadota* and *Bacteroidota*. Within *Pseudomonadota, β-proteobacteria* was most dominant class followed by *γ-protebacteria*. At genus level, *Lachnospiraceae* was found to be most dominant. For STP effluent samples collected in February, *Pseudomonadota* dominated followed by *Bacteroidota* and *Bacillota*. Within *Pseudomonadota, β-proteobacteria* was most dominant class followed by *γ-protebacteria*.

**Figure 1:**
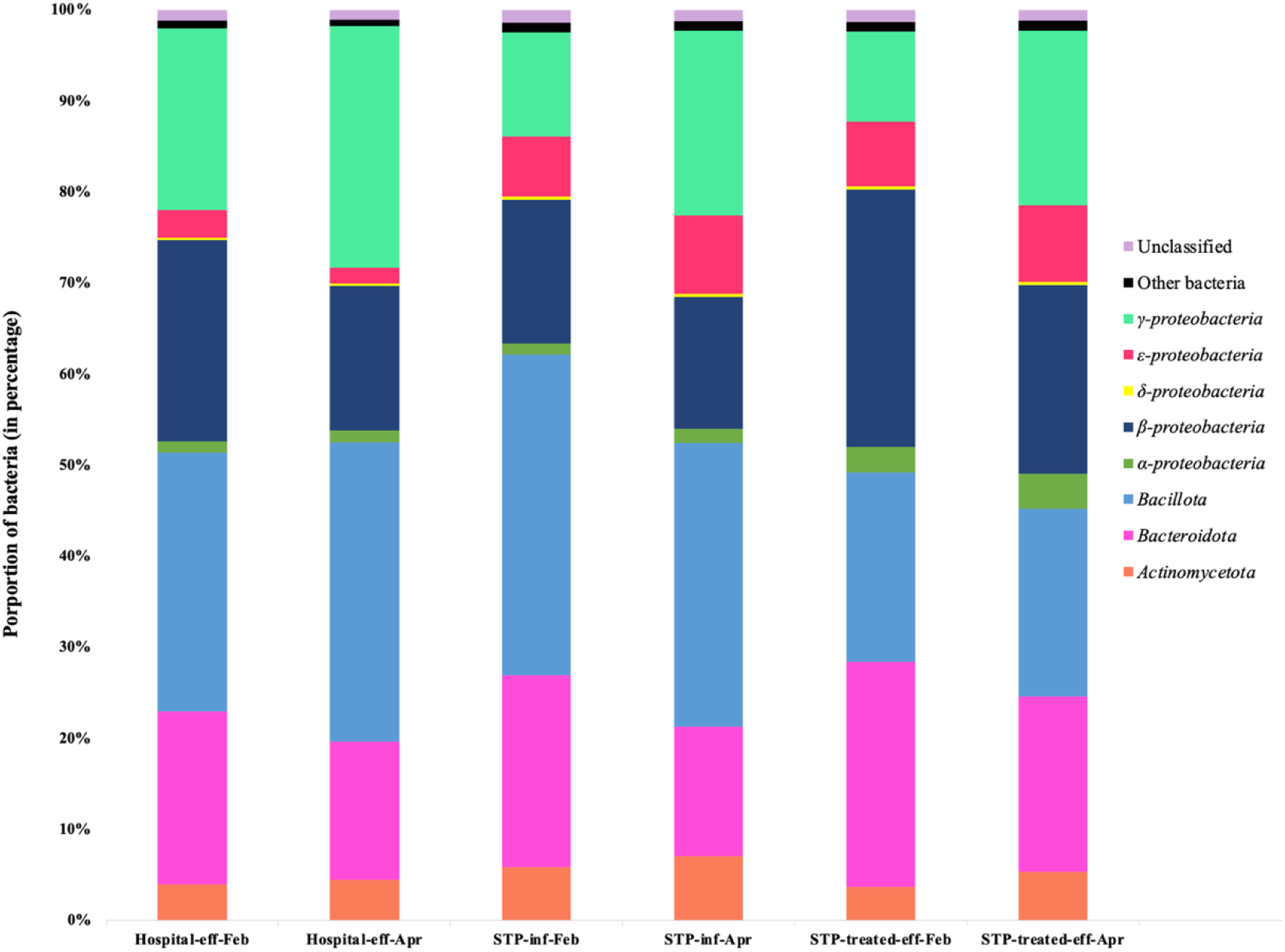
Relative abundance of bacterial diversity detected in sewage from two sampling timepoints February (Feb) and April (Apr), including hospital effluent (eff), STP influent (inf) and STP treated effluent (eff) samples. Only *Pseudomonadota* are represented at class level.

### 3.7 Abundance of ARGs, MGEs, BMRGs and virulence genes

In total, 676 unique ARGs were detected across samples. The macrolide resistance genes *msr*(E) and *mph*(E) were the most abundant ARGs in hospital effluent collected in February and April, and in STP influent and STP effluent samples collected in April. For STP influent and STP effluent collected in February, the tetracycline resistance genes *tet*(Q) and *tet*(W) were the most abundant ARGs (Figure 2). For majority of the samples, we detected genes conferring resistance to macrolides as most abundant, followed by genes conferring resistance to β-lactams and tetracyclines. Genes conferring resistance against last resort antibiotics, such as tigecycline (*tet*(X)), colistin (*mcr*), vancomycin (*van* type genes), and linezolid (*cfr*(C)) were detected across all samples (Supplementary Table S4).

**Figure 2:**
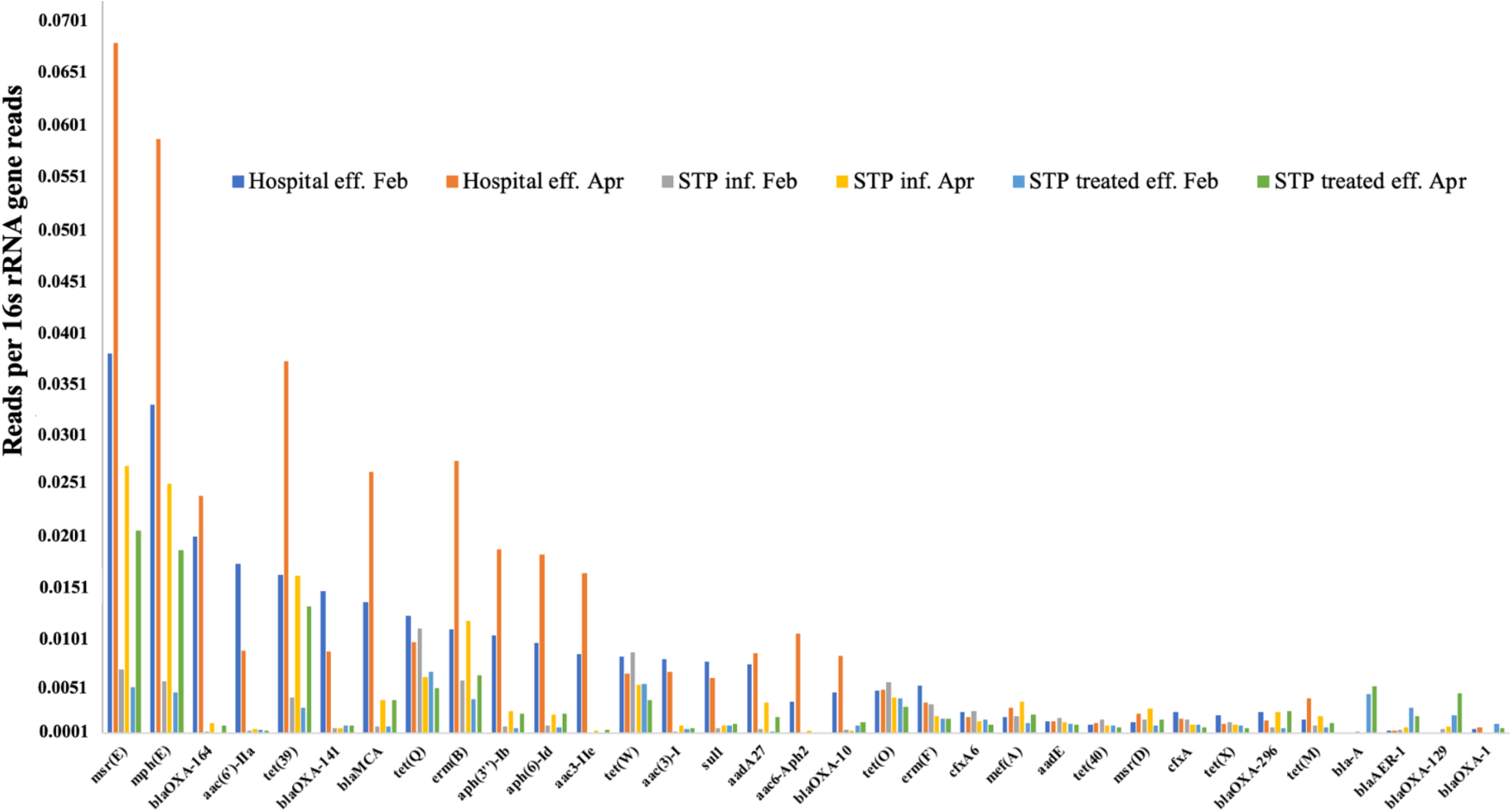
Relative abundance of antibiotic resistance genes (ARGs) representing the 15 ARGs across samples. Hospital effluent (eff.) February (Feb) is highlighted in blue, hospital eff. April (Apr) in orange, STP influent (inf) Feb in grey, STP inf. Apr in orange, STP treated eff. Feb in blue and STP treated eff. Apr in green.

Overall, there was a higher abundance of integron-associated integrases (*intI*) and IS*CR* transposases in hospital effluent samples compared to STP influent and STP effluent samples. Among the studied MGEs, class 1 integrase was the most abundant followed by class 2 integrase and IS*CR2* for most of the samples (Supplementary Table S5).

Several BMRGs conferring resistance to a range of compounds, such as arsenic, cobalt, copper, lead, mercury, nickel, silver, and zinc compounds were detected across samples and found in higher abundance in hospital effluent compared to STP influent and STP effluent. For all samples, the relative abundance of genes conferring resistance to mercury (*mer* genes) was higher compared to other BMRGs. A complete overview of the BMRGs is presented in Supplementary Table S6.

Virulence genes involved in adhesion factors, capsule formation, pilus formation, siderophore production, and toxin productions were also detected in all samples. Important virulence genes, such as aerobactin, enterobactin, yersiniabactin, salmochelin, cytotoxic necrotizing factor 1, colibactin, and hemolysin, were detected across all samples (Supplementary Table S7). The important virulence factor, yersiniabactin, was also detected in five (K11-302, K11-304, K11-305, K11-327 and K11-329) out of 17 sequenced *K. pneumoniae* strains.

### 3.8 Shared ARGs across samples

More than 200 ARGs were shared among the samples, including genes conferring resistance against last resort antibiotics, such as colistin, carbapenems, tigecycline, linezolid and vancomycin. A total of 1,205 predicted gene sequences were assembled, using fARGene. Several ARGs conferring resistance to aminoglycosides, macrolides, tetracyclines and quinolones were shared across samples (Supplementary Table S8). Among all assembled ARGs, 365 represented novel ARGs using cutoff < 90% aa identity (Table 1). The majority of the predicted novel ARGs (n=244) were β-lactamases, followed by aminoglycoside resistance genes (n=98) (Table 1). Among novel β-lactamases, we detected 32 novel MBLs across all samples, where most of the novel MBLs were detected in the STP treated effluent samples.

**Table 1:** Number of antibiotic resistance genes (ARGs) assembled using fARGene and the antibiotic classes they confer resistant against. UA (unique ARGs); KA (known ARGs); NA (novel ARGs); Hosp (hospital); eff (effluent); F (February); A (April); STP (sewage treatment plant); inf (influent); T (treated). Novel genes <90% aa identity.

| Antibiotic class | Aminoglycosides | | | β-lactams | | | Macrolides | | | Quinolones | | | Tetracyclines | | |
| --- | --- | --- | --- | --- | --- | --- | --- | --- | --- | --- | --- | --- | --- | --- | --- |
| Sample | UA | KA | NA | UA | KA | NA | UA | KA | NA | UA | KA | NA | UA | KA | NA |
| Hosp-eff-F | 55 | 35 | 20 | 136 | 105 | 31 | 23 | 20 | 3 | 6 | 5 | 1 | 17 | 17 | 0 |
| Hosp-eff-A | 39 | 12 | 27 | 92 | 78 | 14 | 16 | 15 | 1 | 5 | 4 | 1 | 12 | 11 | 1 |
| STP-inf-F | 34 | 24 | 10 | 111 | 79 | 32 | 23 | 21 | 2 | 5 | 4 | 1 | 17 | 17 | 0 |
| STP-inf-A | 38 | 24 | 14 | 94 | 65 | 29 | 14 | 13 | 1 | 2 | 1 | 1 | 12 | 12 | 0 |
| STP-T-eff-F | 25 | 19 | 6 | 120 | 65 | 56 | 11 | 11 | 0 | 2 | 1 | 1 | 11 | 11 | 0 |
| STP-T-eff-A | 49 | 28 | 21 | 187 | 104 | 83 | 23 | 18 | 5 | 3 | 2 | 1 | 22 | 19 | 3 |

In total, 40 unique assembled ARGs were shared across all samples (Figure 3), representing both novel (n=1) (belonging to class A β-lactamase) and known ARGs (n=39). A total of 54 unique ARGs were shared between hospital effluent and STP effluent, including three novel ARGs (two β-lactamases and one ribosomal RNA methyltransferase Erm).

**Figure 3:**
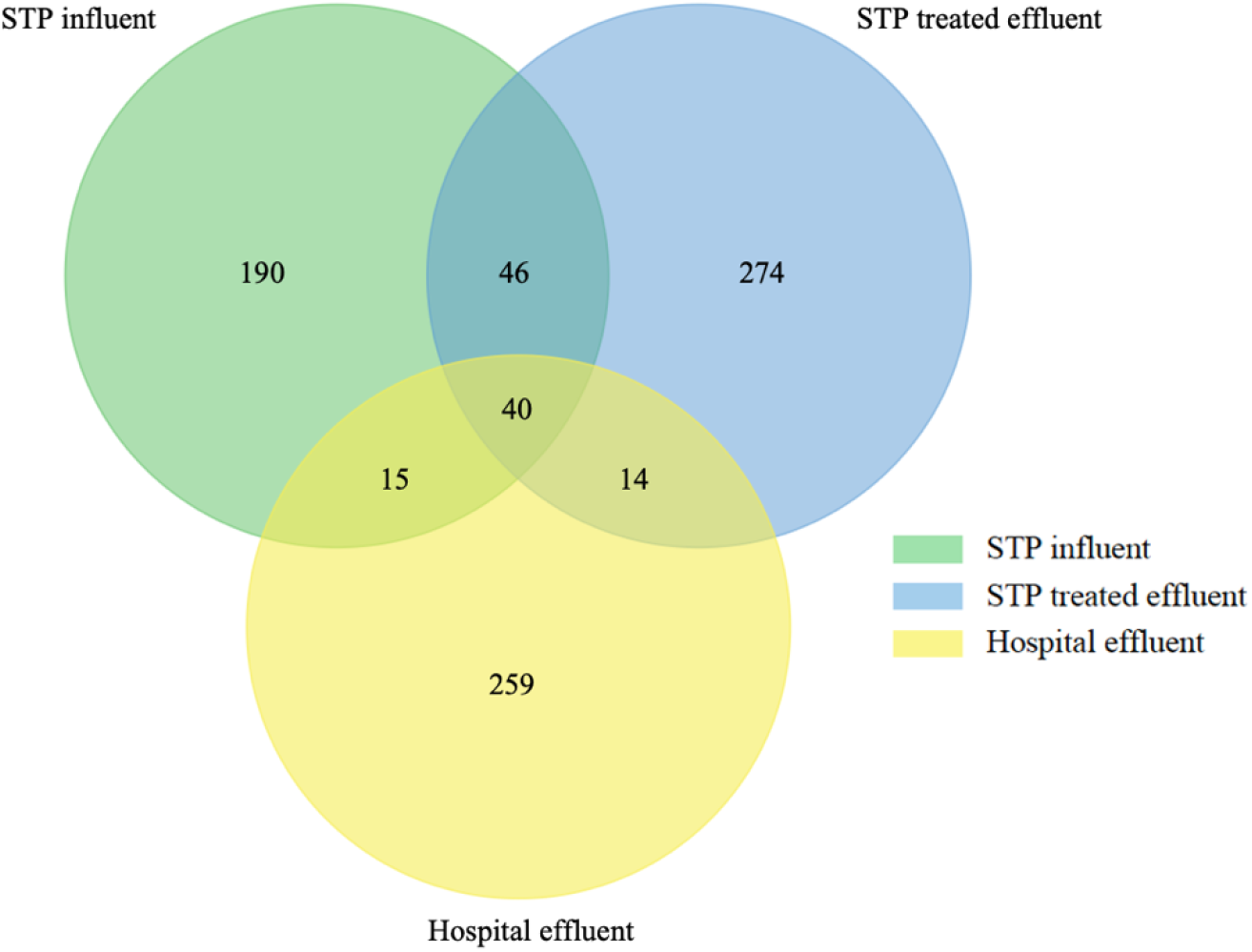
Venn diagram showing the shared (46, 40, 14 and 15) and exclusive (190, 274 and 259) antibiotic resistance genes (ARGs), including both known and novel, across the three samples (hospital effluent, sewage treatment plant (STP) influent and STP treated effluent).

## 4. Discussion

To the best of our knowledge this is the first comprehensive study, investigating the microbiota and resistome of hospital effluent and receiving STP in Norway. Although resistance is low in Norway, we show high diversity of ARGs in hospital effluent and the receiving STP, including genes for resistance against last resort antibiotics. We further show that both novel genes and pathogenic strains are shared between the hospital effluent and STP effluent (Grevskott et al., 2024). Thus, suggesting that hospital effluent is contributing to the spread of AMR even in a low prevalence setting like Norway.

We detected resistance against meropenem (with a MIC of > 16μg/mL) in three out of 66 MDR *E. coli* isolates (4.5%) from STP influent samples (n=2) and STP effluent (n=1) samples. All sequenced meropenem-resistant *E. coli* strains (11-330, 11-M301 and 11-M302) carried NDM-5 and belonged to ST11681 (Supplementary Table S3). The *bla*_NDM-5_ gene was located on a highly conserved region (IS*30*Δ-*bla*_NDM_-*ble*-*trpT*-IS*26*), which has previously been observed in NDM-carrying *E. coli* strains in Norway and in Europe (Chakraborty et al., 2021; Grevskott et al., 2023; Turton et al., 2022). In the clinics in Norway, the occurrence of NDM-5-producing *E. coli* strains has increased during the last decade (NORM/NORM-VET, 2023). The spread of NDM-5 carrying *E. coli* strains in the environment through sewage may have contributed to this increased detection in clinics in Norway.

The *bla*_KPC_ genes, initially described in *K. pneumoniae*, represent globally the most disseminated carbapenemase genes in pathogens like *Enterobacterales* (Hansen, 2021; Munoz-Price et al., 2013), *Acinetobacter* spp. and *Pseudomonas* spp. (Robledo et al., 2011).

Among the KPC variants, KPC-2 and KPC-3 are responsible for most epidemic outbreaks worldwide (Chen et al., 2011). We detected three *bla*_KPC-3_ carrying *K. oxytoca* isolates, (K10-M302, K10-M304 and K10-M306) which were later reidentified as *K. pneumoniae*, using Kleborate. These stains belonged to ST307 which represents a globally disseminated high risk clone (Fostervold et al., 2022; NORM/NORM-VET, 2023; Peirano et al., 2020). This ST is also the most common among *K. pneumoniae* strains in clinics in Norway and is often associated with carbapenemases and ESBLs (NORM/NORM-VET, 2023; Peirano et al., 2020). KPC-3-producing *K. pneumoniae* belonging to ST307 have also been reported from clinics in Norway (NORM/NORM-VET, 2023).

We assembled 365 novel ARGs across samples, representing 32 novel MBLs. The MBLs are enzymes that confer resistance against nearly all β-lactams, including carbapenems, thus limiting available treatment options (Yang et al., 2023). MBLs, such as IMP, VIM, and NDM have been successfully spread among pathogens (Bakthavatchalam et al., 2016; Cornaglia et al., 2011). MBLs can be divided into subclass B1 and B3 (two zinc ions) or subclass B2 (one zinc ion) (Berglund et al., 2017; Palzkill, 2013). Most of the novel MBLs in our study belonged to subclass B1, which also include NDM, VIM and IMP (Palzkill, 2013). These were detected in the STP effluent, suggesting that these ARGs are reaching the receiving environment. Presence of novel MBLs along with clinical strains may provide an opportunity for pathogens to acquire these genes and this may eventually become a problem in clinics (Berglund et al., 2017).

Hospital effluent has been shown to contribute to dissemination of carbapenem-resistant pathogens and genes in the environment (Lamba et al., 2017). In a previous longitudinal temporal study, we showed the presence of carbapenem-resistant *E. coli* strains only in the STP (Holen) receiving hospital sewage on different time-points (Grevskott et al., 2021; Grevskott et al., 2024; Marathe et al., 2023). Further, we showed that OXA-244 carbapenemase-producing *E. coli* ST38 from raw and treated sewage from STP receiving hospital effluent were identical to OXA-244 carbapenemase-producing *E. coli* ST38 from clinics in Norway (Grevskott et al., 2024). Although, the prevalence of resistance in clinics in Norway is low, we detected a high diversity of ARGs in our study. A total of 54 unique ARGs, including both known (n=51) and novel (n=3), were present in the hospital effluent and STP effluent, suggesting that the genes from the hospital effluent are introduced in the environment through treated sewage. Put together, our studies strongly suggest that the hospital effluent contributes to the spread of epidemic-causing pathogens, carbapenemaseses as well as novel ARGs into the receiving marine environment in Norway (Grevskott et al., 2024; Radisic et al., 2023).

## 5. Conclusions

Our study shows that hospital effluent is a rich source of both known and novel ARGs, although Norway has low prevalence of AMR and low usage of antibiotics. We detected *E. coli* and *Klebsiella* spp. strains resistant to last resort antibiotics in hospital effluent as well as in treated effluent of the receiving plant. We further show that clinical strains carrying carbapenemases, such as NDM-5 and KPC-3, are reaching the marine environment through treated sewage, in Norway. We further show that the hospital effluent contributes to the spread of both known and novel ARGs into the receiving environment in Norway. Our study suggests that mitigation strategies, such as on-site treatment of hospital effluent or bacterial removal from sewage, are needed in order to limit the ongoing dissemination of AMR into the receiving environment in Norway.

## Data Availability

All data produced in the present study are available upon reasonable request to the authors.

## Author contributions

D.H.G.: Methodology, Investigation, Formal analysis, Visualization, Writing – original draft, Writing - review & editing. V.R.: Methodology, Investigation, Formal analysis, Visualization, Writing - original draft, Writing - review & editing. M.P.V.: Methodology, Investigation, Formal analysis, Data curation, Writing - original draft. N.P.M.: Conceptualization, Methodology, Investigation, Validation, Writing - review & editing, Supervision, Project administration, Funding acquisition.

## Acknowledgements

The authors would like to thank Kristine S. Akervold (Bergen municipality), Ingvild Fenne (Bergen municipality) for permission and support as well as Ole Gunnar (Bergen Vann) and the people working at the STP for sewage sample collection.

## Supplementary Materials

**Supplementary Table S1:** Minimum inhibitory concentrations (MICs) of *Escherichia coli* (n= 66) and *Klebsiella* spp. (n=55) strains in this study.

**Supplementary Table S2:** Genome assembly statistics of sequenced strains (n=36) in this study.

**Supplementary Table S3:** Detected antibiotic resistance genes (ARGs) and sequence types (STs) in sequenced strains (n=36).

**Supplementary Table S4:** Detected antibiotic resistance genes (ARGs) across six samples.

**Supplementary Table S5:** Relative abundance of most detected mobile genetic elements (MGEs) across samples.

**Supplementary Table S6:** Detected biocide/metal resistance genes (BMRGs) across all samples.

**Supplementary Table S7:** Detected virulence genes across all samples.

**Supplementary Table S8:** Assembled known and novel antibiotic resistance genes (ARGs) across all samples.

