## Supplementary material for "Novel antibiotic resistance genes from the hospital effluent are disseminated into the marine environment in Norway": supplemantary table S4

**Supplementary Table S4:** Detected antibiotic resistance genes (ARG

| Sample | ARGs |
|---|---|
| Hospital-eff-F | *msr*(E) |
| Hospital-eff-F | *mph*(E) |
| Hospital-eff-F | *blaOXA-164* |
| Hospital-eff-F | *AAC(6')-IIa* |
| Hospital-eff-F | *tet*(39) |
| Hospital-eff-F | *blaOXA-141* |
| Hospital-eff-F | *blaMCA* |
| Hospital-eff-F | *tet*(Q) |
| Hospital-eff-F | *erm*(B) |
| Hospital-eff-F | *aph(3'')-Ib* |
| Hospital-eff-F | *aph(6)-Id* |
| Hospital-eff-F | *aac3-IIe* |
| Hospital-eff-F | *tet*(W) |
| Hospital-eff-F | *aac(3)-I* |
| Hospital-eff-F | *sul1* |
| Hospital-eff-F | *aadA27* |
| Hospital-eff-F | *blaOXA-139* |
| Hospital-eff-F | *erm*(F) |
| Hospital-eff-F | *tet*(C) |
| Hospital-eff-F | *aacA38* |
| Hospital-eff-F | *tet*(O) |
| Hospital-eff-F | *blaOXA-10* |
| Hospital-eff-F | *aac3-Ib* |
| Hospital-eff-F | *qacH* |
| Hospital-eff-F | *sul2* |
| Hospital-eff-F | *aacA-ACI1* |
| Hospital-eff-F | *ere(B)* |
| Hospital-eff-F | *aac6-Aph2* |
| Hospital-eff-F | *blaOXA-280* |
| Hospital-eff-F | *blaOXA-211* |
| Hospital-eff-F | *blaOXA-333* |
| Hospital-eff-F | *blaPSE-4* |
| Hospital-eff-F | *AAC(6')-Ib7* |
| Hospital-eff-F | *ant(3'')-IIa* |
| Hospital-eff-F | *aac3-IId* |
| Hospital-eff-F | *AAC(6')-30/AAC(6')-Ib'_fusion_* |
| Hospital-eff-F | *blaOXA-2* |
| Hospital-eff-F | *cfxA6* |
| Hospital-eff-F | *cfxA* |
| Hospital-eff-F | *blaOXA-296* |
| Hospital-eff-F | *blaOXA-160* |

| | |
|---|---|
| Hospital-eff-F | *aadS* |
| Hospital-eff-F | *tet*(X) |
| Hospital-eff-F | *blaOXA-281* |
| Hospital-eff-F | *ant(2'')-Ia* |
| Hospital-eff-F | *mef*(A) |
| Hospital-eff-F | *tet*(A) |
| Hospital-eff-F | *aadA1* |
| Hospital-eff-F | *tet*(M) |
| Hospital-eff-F | *aph(2'')-IIa* |
| Hospital-eff-F | *tet*(R) |
| Hospital-eff-F | *aac(6')-Ib-G* |
| Hospital-eff-F | *Mef(En2)* |
| Hospital-eff-F | *lnu(AN2)* |
| Hospital-eff-F | *aac3-Ib-Aac6-Ib* |
| Hospital-eff-F | *blaOXA-18* |
| Hospital-eff-F | *aadE* |
| Hospital-eff-F | *aac(3)-IIe* |
| Hospital-eff-F | *mef*(C) |
| Hospital-eff-F | *Bifidobacterium_adolescentis_* |
| Hospital-eff-F | *aadA5* |
| Hospital-eff-F | *msr*(D) |
| Hospital-eff-F | *aac3-I* |
| Hospital-eff-F | *blaOXA-347* |
| Hospital-eff-F | *vanZ-A* |
| Hospital-eff-F | *aac(6')-Im* |
| Hospital-eff-F | *mph*(G) |
| Hospital-eff-F | *blaOXA-15* |
| Hospital-eff-F | *strB* |
| Hospital-eff-F | *dfrF* |
| Hospital-eff-F | *tet*(40) |
| Hospital-eff-F | *vanA-G* |
| Hospital-eff-F | *vanY-A* |
| Hospital-eff-F | *ere(D)* |
| Hospital-eff-F | *erm*(G) |
| Hospital-eff-F | *TEM-1* |
| Hospital-eff-F | *tet*(32) |
| Hospital-eff-F | *ant(3'')-IIc* |
| Hospital-eff-F | *CblA-1* |
| Hospital-eff-F | *aadA2* |
| Hospital-eff-F | *blaOXA-9* |
| Hospital-eff-F | *aac(3)-Ia* |
| Hospital-eff-F | *aph(3')-VIa* |
| Hospital-eff-F | *aadA11* |
| Hospital-eff-F | *catB3* |

| | |
|---|---|
| Hospital-eff-F | *catA13* |
| Hospital-eff-F | *aadA6* |
| Hospital-eff-F | *aacA-ACI6* |
| Hospital-eff-F | *blaOXA-212* |
| Hospital-eff-F | *blaOXA-1* |
| Hospital-eff-F | *tet*(G) |
| Hospital-eff-F | *catS* |
| Hospital-eff-F | *aac3-IIa* |
| Hospital-eff-F | *aac6-Ig* |
| Hospital-eff-F | *blaOXA-652* |
| Hospital-eff-F | *blaOXA-650* |
| Hospital-eff-F | *vanY-B* |
| Hospital-eff-F | *vanH-B* |
| Hospital-eff-F | *blaOXA-643* |
| Hospital-eff-F | *blaOXA-662* |
| Hospital-eff-F | *CARB-3* |
| Hospital-eff-F | *blaOXA-209* |
| Hospital-eff-F | *aac3-IIc* |
| Hospital-eff-F | *tetR*(G) |
| Hospital-eff-F | *mph*(A) |
| Hospital-eff-F | *blaOXA-373* |
| Hospital-eff-F | *aph(3'')-III* |
| Hospital-eff-F | *Klebsiella_pneumoniae_KpnG* |
| Hospital-eff-F | *floR* |
| Hospital-eff-F | *aph3-VIb* |
| Hospital-eff-F | *aph(3'')-Ia* |
| Hospital-eff-F | *vanS-B* |
| Hospital-eff-F | *vanX-B* |
| Hospital-eff-F | *Bifidobacteria_intrinsic_ileS_* |
| Hospital-eff-F | *aadA24* |
| Hospital-eff-F | *aac(6')-30* |
| Hospital-eff-F | *vanH-A* |
| Hospital-eff-F | *bacA* |
| Hospital-eff-F | *CRP* |
| Hospital-eff-F | *ampH* |
| Hospital-eff-F | *tolC* |
| Hospital-eff-F | *cpxA* |
| Hospital-eff-F | *blaOXA-427* |
| Hospital-eff-F | *vanR-A* |
| Hospital-eff-F | *mdtP* |
| Hospital-eff-F | *blaAER-1* |
| Hospital-eff-F | *mdtE* |
| Hospital-eff-F | *vanX-A* |
| Hospital-eff-F | *lnu(C)* |

| | |
|---|---|
| Hospital-eff-F | *Klebsiella_pneumoniae_acrA* |
| Hospital-eff-F | *mdtF* |
| Hospital-eff-F | *vanA-A* |
| Hospital-eff-F | *acrB* |
| Hospital-eff-F | *cfxA_gen* |
| Hospital-eff-F | *aadB* |
| Hospital-eff-F | *H-NS* |
| Hospital-eff-F | *blaOXA-334* |
| Hospital-eff-F | *gadX* |
| Hospital-eff-F | *cmxA* |
| Hospital-eff-F | *blaMOX-9* |
| Hospital-eff-F | *vanW-B* |
| Hospital-eff-F | *baeR* |
| Hospital-eff-F | *blaOXA-101* |
| Hospital-eff-F | *gadW* |
| Hospital-eff-F | *kdpE* |
| Hospital-eff-F | *vanB* |
| Hospital-eff-F | *blaPER-1* |
| Hospital-eff-F | *dfrA14* |
| Hospital-eff-F | *vanS-A* |
| Hospital-eff-F | *CARB-2* |
| Hospital-eff-F | *qnrS2* |
| Hospital-eff-F | *blaOXA-24* |
| Hospital-eff-F | *floR2* |
| Hospital-eff-F | *Penicillin_Binding_Protein_Eco* |
| Hospital-eff-F | *mdtN* |
| Hospital-eff-F | *aph7* |
| Hospital-eff-F | *vanR-B* |
| Hospital-eff-F | *cfr(C)* |
| Hospital-eff-F | *acrF* |
| Hospital-eff-F | *aadA17* |
| Hospital-eff-F | *AAC(6')-Ib8* |
| Hospital-eff-F | *mdtO* |
| Hospital-eff-F | *aph(3')-VI* |
| Hospital-eff-F | *mdtB* |
| Hospital-eff-F | *blaBES-1* |
| Hospital-eff-F | *ampH_Ecoli* |
| Hospital-eff-F | *blaOXA-205* |
| Hospital-eff-F | *evgS* |
| Hospital-eff-F | *aad9* |
| Hospital-eff-F | *marA* |
| Hospital-eff-F | *Klebsiella_pneumoniae_OmpK* |
| Hospital-eff-F | *eptA* |
| Hospital-eff-F | *tet*(44) |

| | |
|---|---|
| Hospital-eff-F | *mdtC* |
| Hospital-eff-F | *Escherichia_coli_emrE* |
| Hospital-eff-F | *OqxA* |
| Hospital-eff-F | *blaOXA-836* |
| Hospital-eff-F | *emrK* |
| Hospital-eff-F | *Escherichia_coli_acrA* |
| Hospital-eff-F | *evgA* |
| Hospital-eff-F | *blaOXA-645* |
| Hospital-eff-F | *acrS* |
| Hospital-eff-F | *emrY* |
| Hospital-eff-F | *acrD* |
| Hospital-eff-F | *fosA_gen* |
| Hospital-eff-F | *aac(3)-IVa* |
| Hospital-eff-F | *Escherichia_coli_mdfA* |
| Hospital-eff-F | *dfrB1* |
| Hospital-eff-F | *mdtG* |
| Hospital-eff-F | *pmrF* |
| Hospital-eff-F | *acrE* |
| Hospital-eff-F | *yojI* |
| Hospital-eff-F | *nimj_Nitroimidazole_Gene* |
| Hospital-eff-F | *cblA* |
| Hospital-eff-F | *ugd* |
| Hospital-eff-F | *mdtA* |
| Hospital-eff-F | *lsa(E)* |
| Hospital-eff-F | *emrA* |
| Hospital-eff-F | *oqxA10* |
| Hospital-eff-F | *blaOXA-161* |
| Hospital-eff-F | *bla-A* |
| Hospital-eff-F | *emrR* |
| Hospital-eff-F | *ant3''Ih-Aac6-IId* |
| Hospital-eff-F | *blaSHV-1* |
| Hospital-eff-F | *qnr-S1* |
| Hospital-eff-F | *aac6-Im* |
| Hospital-eff-F | *AmpC1_Ecoli* |
| Hospital-eff-F | *AAC(6')-Ib-cr* |
| Hospital-eff-F | *blaOXA-13* |
| Hospital-eff-F | *blaOXA-256* |
| Hospital-eff-F | *Klebsiella_pneumoniae_KpnH* |
| Hospital-eff-F | *msbA* |
| Hospital-eff-F | *mdtH* |
| Hospital-eff-F | *blaOXA-504* |
| Hospital-eff-F | *baeS* |
| Hospital-eff-F | *blaTEM-112* |
| Hospital-eff-F | *catB4* |

| | |
|---|---|
| Hospital-eff-F | *OqxBgb* |
| Hospital-eff-F | *blaIMP-22* |
| Hospital-eff-F | *blaVIM-10* |
| Hospital-eff-F | *catA2* |
| Hospital-eff-F | *aph4-Ia* |
| Hospital-eff-F | *cepS* |
| Hospital-eff-F | *blaOXA-275* |
| Hospital-eff-F | *lin(B)* |
| Hospital-eff-F | *mdtM* |
| Hospital-eff-F | *blaOXA-667* |
| Hospital-eff-F | *Klebsiella_pneumoniae_KpnE* |
| Hospital-eff-F | *tet*(E) |
| Hospital-eff-F | *erm*(T) |
| Hospital-eff-F | *tet*(36) |
| Hospital-eff-F | *aph(3')-Ia* |
| Hospital-eff-F | *blaCTX-M-101* |
| Hospital-eff-F | *msr*(C) |
| Hospital-eff-F | *efmA* |
| Hospital-eff-F | *vanT-G* |
| Hospital-eff-F | *aadA10* |
| Hospital-eff-F | *mcr-5.1* |
| Hospital-eff-F | *arr* |
| Hospital-eff-F | *arr2* |
| Hospital-eff-F | *blaOXA-299* |
| Hospital-eff-F | *aadA16* |
| Hospital-eff-F | *aadA25* |
| Hospital-eff-F | *qnrB19* |
| Hospital-eff-F | *blaSHV-103* |
| Hospital-eff-F | *blaEC* |
| Hospital-eff-F | *cfxA3* |
| Hospital-eff-F | *blaRCP* |
| Hospital-eff-F | *blaMOX-2* |
| Hospital-eff-F | *aph(2'')-If* |
| Hospital-eff-F | *blaVEB-1* |
| Hospital-eff-F | *cepA* |
| Hospital-eff-F | *arr3* |
| Hospital-eff-F | *blaAIM-1* |
| Hospital-eff-F | *aadA1-pm* |
| Hospital-eff-F | *blaSHV-12* |
| Hospital-eff-F | *eat(A)* |
| Hospital-eff-F | *blaOXA-726* |
| Hospital-eff-F | *blaOXA-46* |
| Hospital-eff-F | *mcr-3.6* |
| Hospital-eff-F | *blaOXA-230* |

| | |
|---|---|
| Hospital-eff-F | *sat-2A* |
| Hospital-eff-F | *Nocardia_rifampin_resistant_* |
| Hospital-eff-F | *blaOXA-668* |
| Hospital-eff-F | *fosA* |
| Hospital-eff-F | *ant6-Ib* |
| Hospital-eff-F | *cfxA5* |
| Hospital-eff-F | *sul4* |
| Hospital-eff-F | *sat4* |
| Hospital-eff-F | *blaOXY2-1* |
| Hospital-eff-F | *vanXY-G2* |
| Hospital-eff-F | *qnrVC1* |
| Hospital-eff-F | *catA1* |
| Hospital-eff-F | *aac(6')-Ib-cr3* |
| Hospital-eff-F | *ere(A)* |
| Hospital-eff-F | *blaNPS-1* |
| Hospital-eff-F | *AmpC2_Ecoli* |
| Hospital-eff-F | *blaOXA-780* |
| Hospital-eff-F | *spw* |
| Hospital-eff-F | *blaOXA-309* |
| Hospital-eff-F | *dfrB3* |
| Hospital-eff-F | *Enterobacter_cloacae_acrA* |
| Hospital-eff-F | *vanW-G* |
| Hospital-eff-F | *tet*(33) |
| Hospital-eff-F | *blaSHV-67* |
| Hospital-eff-F | *blaOXY1-1* |
| Hospital-eff-F | *oqxB11* |
| Hospital-eff-F | *qnrD* |
| Hospital-eff-F | *oqxB15* |
| Hospital-eff-F | *aadA12* |
| Hospital-eff-F | *MexF* |
| Hospital-eff-F | *aadA13* |
| Hospital-eff-F | *smeE* |
| Hospital-eff-F | *ere(A2)* |
| Hospital-eff-F | *mef*(B) |
| Hospital-eff-F | *aac6-Ii* |
| Hospital-eff-F | *blaOXA-651* |
| Hospital-eff-F | *adeJ* |
| Hospital-eff-F | *aad(6)* |
| Hospital-eff-F | *aac6-IIa* |
| Hospital-eff-F | *FosA6* |
| Hospital-eff-F | *dfr23* |
| Hospital-eff-F | *MexB* |
| Hospital-eff-F | *blaSHV-149* |
| Hospital-eff-F | *blaCMY-103* |

| | |
|---|---|
| Hospital-eff-F | *blaTEM-156* |
| Hospital-eff-F | *blaACC-1* |
| Hospital-eff-F | *AAC(6')-Ib10* |
| Hospital-eff-F | *blaBKC-1* |
| Hospital-eff-F | *dfrA1* |
| Hospital-eff-F | *dfrA27* |
| Hospital-eff-F | *blaOXA-724* |
| Hospital-eff-F | *blaOXA-779* |
| Hospital-eff-F | *aph(3')-IIc* |
| Hospital-eff-F | *qnrB1* |
| Hospital-eff-F | *aac3-Vb* |
| Hospital-eff-F | *Klebsiella_pneumoniae_KpnF* |
| Hospital-eff-F | *dfrA12* |
| Hospital-eff-F | *dfrG* |
| Hospital-eff-F | *aci1* |
| Hospital-eff-F | *nimE_Nitroimidazole_Gene* |
| Hospital-eff-F | *blaSHV-102* |
| Hospital-eff-F | *tet*(B) |
| Hospital-eff-F | *cfr-Cb* |
| Hospital-eff-F | *blaCPS-1* |
| Hospital-eff-F | *blaCTX-M-103* |
| Hospital-eff-F | *tetA(P)* |
| Hospital-eff-F | *MexD* |
| Hospital-eff-F | *aacA47* |
| Hospital-eff-F | *blaEC-13* |
| Hospital-eff-F | *blaEC-19* |
| Hospital-eff-F | *blaEC-5* |
| Hospital-eff-F | *Salmonella_enterica_cmlA* |
| Hospital-eff-F | *oqxB10* |
| Hospital-eff-F | *blaOXA-725* |
| Hospital-eff-F | *blaOXA-118* |
| Hospital-eff-F | *lnu(G)* |
| Hospital-eff-F | *mcr-3.1* |
| Hospital-eff-F | *blaOXA-644* |
| Hospital-eff-F | *vanD* |
| Hospital-eff-F | *blaOXA-228* |
| Hospital-eff-F | *blaOXA-257* |
| Hospital-eff-F | *blaOXA-392* |
| Hospital-eff-F | *cmlA1* |
| Hospital-eff-F | *lsa(C)* |
| Hospital-eff-F | *FosA2* |
| Hospital-eff-F | *aac(3)-IIa* |
| Hospital-eff-F | *aac(3)-IId* |
| Hospital-eff-F | *blaSHV-110* |

| | |
|---|---|
| Hospital-eff-F | *blaSHV-35* |
| Hospital-eff-F | *blaCTX-M-1* |
| Hospital-eff-F | *blaCTX-M-182* |
| Hospital-eff-F | *blaPAU-1* |
| Hospital-eff-F | *blaCMY-113* |
| Hospital-eff-F | *smeR* |
| Hospital-eff-F | *TriA* |
| Hospital-eff-F | *oqxA11* |
| Hospital-eff-F | *dfrA17* |
| Hospital-eff-F | *dfrA5* |
| Hospital-eff-F | *dhfr7* |
| Hospital-eff-F | *mphB* |
| Hospital-eff-F | *oqxB14* |
| Hospital-eff-F | *oqxB9* |
| Hospital-eff-F | *cmlA* |
| Hospital-eff-F | *blaOXA-464* |
| Hospital-eff-F | *cphA2* |
| Hospital-eff-F | *cphA8* |
| Hospital-eff-F | *imiH* |
| Hospital-eff-F | *vanXY-G* |
| Hospital-eff-F | *AAC(6')-Ib-Suzhou* |
| Hospital-eff-F | *vanG2* |
| Hospital-eff-F | *aph(3')-IIa* |
| Hospital-eff-F | *nimA_Nitroimidazole_Gene* |
| Hospital-eff-F | *aadA7* |
| Hospital-eff-F | *blaOXA-20* |
| Hospital-eff-F | *blaVIM-11* |
| Hospital-eff-F | *erm*(35) |
| Hospital-eff-F | *lnu((B)* |
| Hospital-eff-F | *aph3-Ib* |
| Hospital-eff-F | *car(A)* |
| Hospital-eff-F | *aac(6')-Ib-AKT* |
| Hospital-eff-F | *ICR-Mo* |
| Hospital-eff-F | *mexK* |
| Hospital-eff-F | *dfrA10* |
| Hospital-eff-F | *Clostridium_perfringens_mprF* |
| Hospital-eff-F | *lmrD* |
| Hospital-eff-F | *AxyY* |
| Hospital-eff-F | *blaCMY-105* |
| Hospital-eff-F | *blaSHV-107* |
| Hospital-eff-F | *blaSHV-179* |
| Hospital-eff-F | *blaTEM-102* |
| Hospital-eff-F | *blaTEM-93* |
| Hospital-eff-F | *blaGES-1* |

| | |
|---|---|
| Hospital-eff-F | *efrA* |
| Hospital-eff-F | *blaLHK-6* |
| Hospital-eff-F | *blaKPC-15* |
| Hospital-eff-F | *blaRSC1-1* |
| Hospital-eff-F | *catP* |
| Hospital-eff-F | *tet*(U) |
| Hospital-eff-F | *cfxA4* |
| Hospital-eff-F | *qnrB21* |
| Hospital-eff-F | *mcr-3.12* |
| Hospital-eff-F | *qnrVC6* |
| Hospital-eff-F | *mtrA* |
| Hospital-eff-F | *oqxB12* |
| Hospital-eff-F | *smeC* |
| Hospital-eff-F | *ble-MBL* |
| Hospital-eff-F | *OpmB* |
| Hospital-eff-F | *blaCCR-A* |
| Hospital-eff-F | *ramA* |
| Hospital-eff-F | *blaEC-18* |
| Hospital-eff-F | *blaEC-8* |
| Hospital-eff-F | *blaACT-58* |
| Hospital-eff-F | *vanS-D* |
| Hospital-eff-F | *mexW* |
| Hospital-eff-F | *cepH-A3* |
| Hospital-eff-F | *cphA1* |
| Hospital-eff-F | *tet*(T) |
| Hospital-eff-F | *blaLCR-1* |
| Hospital-eff-F | *tetB-P* |
| Hospital-eff-F | *oqxA3* |
| Hospital-eff-F | *aadA15* |
| Hospital-eff-F | *aadA3* |
| Hospital-eff-F | *beta-lactamase_class-C* |
| Hospital-eff-F | *blaOXA-145* |
| Hospital-eff-F | *smeA* |
| Hospital-eff-F | *blaOXA-119* |
| Hospital-eff-F | *blaOXA-240* |
| Hospital-eff-F | *aac6* |
| Hospital-eff-F | *mcr-9.1* |
| Hospital-eff-F | *nshR* |
| Hospital-eff-F | *blaOXA-226* |
| Hospital-eff-F | *blaOXA-31* |
| Hospital-eff-F | *blaOXA-320* |
| Hospital-eff-F | *blaOXA-539* |
| Hospital-eff-F | *blaOXA-675* |
| Hospital-eff-F | *mepR* |

| | |
|---|---|
| Hospital-eff-F | *Streptomyces_rishiriensis_pa* |
| Hospital-eff-F | *erm*(36) |
| Hospital-eff-F | *fosA8* |
| Hospital-eff-F | *blaOXA-666* |
| Hospital-eff-F | *blaOKP-B-10* |
| Hospital-eff-F | *blaSHV-115* |
| Hospital-eff-F | *blaTEM-10* |
| Hospital-eff-F | *blaTEM-106* |
| Hospital-eff-F | *blaTEM-108* |
| Hospital-eff-F | *blaTEM-125* |
| Hospital-eff-F | *blaCTX-M-115* |
| Hospital-eff-F | *blaORN1a* |
| Hospital-eff-F | *aacA34* |
| Hospital-eff-F | *mexI* |
| Hospital-eff-F | *blaGPC-1* |
| Hospital-eff-F | *mexG* |
| Hospital-eff-F | *smeB* |
| Hospital-eff-F | *aph(2'')-Id* |
| Hospital-eff-F | *erm*(49) |
| Hospital-eff-F | *aac(6')-Iz* |
| Hospital-eff-F | *aac(6')_Steno* |
| Hospital-eff-F | *aac(6')_Strep* |
| Hospital-eff-F | *PmpM* |
| Hospital-eff-F | *vanH-D* |
| Hospital-eff-F | *nimD_Nitroimidazole_Gene* |
| Hospital-eff-F | *BlaA1* |
| Hospital-eff-F | *qepA2* |
| Hospital-eff-F | *MuxC* |
| Hospital-eff-F | *mcr-3.17* |
| Hospital-eff-F | *sat4A* |
| Hospital-eff-F | *mfpA* |
| Hospital-eff-F | *ACT-16* |
| Hospital-eff-F | *blaACT-59* |
| Hospital-eff-F | *blaCMY-115* |
| Hospital-eff-F | *blaFOX-15* |
| Hospital-eff-F | *blaMOX-4* |
| Hospital-eff-F | *cepH* |
| Hospital-eff-F | *tetA(46)* |
| Hospital-eff-F | *tet*(Y) |
| Hospital-eff-F | *patB* |
| Hospital-eff-F | *smeD* |
| Hospital-eff-F | *tet*(D) |
| Hospital-eff-F | *tet*(H) |
| Hospital-eff-F | *bcr1* |

| | |
|---|---|
| Hospital-eff-F | *aac6-Ib* |
| Hospital-eff-F | *vanX-D* |
| Hospital-eff-F | *MuxB* |
| Hospital-eff-F | *cmlA5* |
| Hospital-eff-F | *catB2* |
| Hospital-eff-F | *catB8* |
| Hospital-eff-F | *MuxA* |
| Hospital-eff-F | *qnrE1* |
| Hospital-eff-F | *catA14* |
| Hospital-eff-F | *catA4* |
| Hospital-eff-F | *qnrA9* |
| Hospital-eff-F | *arlR* |
| Hospital-eff-F | *catQ* |
| Hospital-eff-F | *arnA* |
| Hospital-eff-F | *Pseudomonas_aeruginosa_Cp* |
| Hospital-eff-F | *qnrB5* |
| Hospital-eff-F | *vanR-C* |
| Hospital-eff-F | *vanT* |
| Hospital-eff-F | *iri* |
| Hospital-eff-F | *OpmH* |
| Hospital-eff-F | *adeK* |
| Hospital-eff-F | *erm*(A) |
| Hospital-eff-F | *erm*(C) |
| Hospital-eff-F | *rmtD2* |
| Hospital-eff-F | *cfiA11* |
| Hospital-eff-F | *cfiA19* |
| Hospital-eff-F | *cfiA23* |
| Hospital-eff-F | *cphA4* |
| Hospital-eff-F | *cphA5* |
| Hospital-eff-F | *blaOXA-33* |
| Hospital-eff-F | *imiS* |
| Hospital-eff-F | *TetAB* |
| Hospital-eff-F | *blaOXA-641* |
| Hospital-eff-F | *erm*(Q) |
| Hospital-eff-F | *erm*(V) |
| Hospital-eff-F | *erm*(O) |
| Hospital-eff-F | *oqxB18* |
| Hospital-eff-F | *oqxB21* |
| Hospital-eff-F | *blaOXA-198* |
| Hospital-eff-F | *aph(3')-IIIa* |
| Hospital-eff-F | *blaOXA-520* |
| Hospital-eff-F | *blaOXA-732* |
| Hospital-eff-F | *blaOXA-736* |
| Hospital-eff-F | *blaVIM-12* |

| | |
|---|---|
| Hospital-eff-F | *blaVIM-8* |
| Hospital-eff-F | *blaOXA-129* |
| Hospital-eff-F | *blaOXA-5* |
| Hospital-eff-F | *aph(3')-IIb* |
| Hospital-eff-F | *mcr-10.1* |
| Hospital-eff-F | *mcr-3.9* |
| Hospital-eff-F | *blaOXA-22* |
| Hospital-eff-F | *blaOXA-258* |
| Hospital-eff-F | *sgm* |
| Hospital-eff-F | *blaOXA-114n* |
| Hospital-eff-F | *blaOXA-114p* |
| Hospital-eff-F | *blaOXA-210* |
| Hospital-eff-F | *blaOXA-3* |
| Hospital-eff-F | *blaOXA-737* |
| Hospital-eff-F | *blaOXA-838* |
| Hospital-eff-F | *blaOXA-355* |
| Hospital-eff-F | *blaOXA-356* |
| Hospital-eff-F | *mph*(D) |
| Hospital-eff-F | *blaLEN-1* |
| Hospital-eff-F | *blaOXA-420* |
| Hospital-eff-F | *blaOXA-512* |
| Hospital-eff-F | *str* |
| Hospital-eff-F | *blaCARB-42* |
| Hospital-eff-F | *blaTEM-199* |
| Hospital-eff-F | *aac3-Xa* |
| Hospital-eff-F | *blaTER-1* |
| Hospital-eff-F | *erm*(X) |
| Hospital-eff-F | *ole(B)* |
| Hospital-eff-F | *blaTEM-178* |
| Hospital-eff-F | *blaGES-23* |
| Hospital-eff-F | *blaOKP-B-11* |
| Hospital-eff-F | *blaOKP-B-17* |
| Hospital-eff-F | *blaOKP-B-4* |
| Hospital-eff-F | *blaSHV-105* |
| Hospital-eff-F | *blaSHV-134* |
| Hospital-eff-F | *blaSHV-142* |
| Hospital-eff-F | *blaSHV-143* |
| Hospital-eff-F | *blaSHV-164* |
| Hospital-eff-F | *blaTEM-107* |
| Hospital-eff-F | *blaTEM-11* |
| Hospital-eff-F | *blaTEM-110* |
| Hospital-eff-F | *blaGES-10* |
| Hospital-eff-F | *blaGES-11* |
| Hospital-eff-F | *blaGES-14* |

| | |
|---|---|
| Hospital-eff-F | *blaGES-9* |
| Hospital-eff-F | *rph* |
| Hospital-eff-F | *tetB*(46) |
| Hospital-eff-F | *blaMBL1b* |
| Hospital-eff-F | *blaCRH-2* |
| Hospital-eff-F | *blaESP-1* |
| Hospital-eff-F | *blaL1* |
| Hospital-eff-F | *blaOXY6-1* |
| Hospital-eff-F | *blaERP-1* |
| Hospital-eff-F | *blaKPC-10* |
| Hospital-eff-F | *blaKPC-21* |
| Hospital-eff-F | *penI_Bp* |
| Hospital-eff-F | *blaSPR-1* |
| Hospital-eff-F | *aph(2'')-Ib* |
| Hospital-eff-F | *blaVEB-3* |
| Hospital-eff-F | *mph*(C) |
| Hospital-eff-F | *mph*(F) |
| Hospital-eff-F | *blaEFM-1* |
| Hospital-eff-F | *blaLRA-7* |
| Hospital-eff-F | *blaRm3* |
| Hospital-eff-F | *aph(2'')-Ic* |
| Hospital-eff-F | *aph(6)-Ia* |
| Hospital-eff-F | *aph9-Ib* |
| Hospital-eff-F | *blaPER-2* |
| Hospital-eff-F | *erm*(S) |
| Hospital-eff-F | *blaDES-1* |
| Hospital-eff-F | *blaL* |
| Hospital-eff-F | *ole(C)* |
| Hospital-eff-F | *blaSGM-6* |
| Hospital-eff-F | *erm*(30) |
| Hospital-eff-F | *vanA-D* |
| Hospital-eff-F | *vanN* |
| Hospital-eff-F | *vanY-D* |
| Hospital-eff-F | *cfr(B)* |
| Hospital-eff-F | *oqxB13* |
| Hospital-eff-F | *oqxB16* |
| Hospital-eff-F | *oqxB17* |
| Hospital-eff-F | *oqxB8* |
| Hospital-eff-F | *vanSc4* |
| Hospital-eff-F | *vanS-E* |
| Hospital-eff-F | *vanG-Cd* |
| Hospital-eff-F | *mexJ* |
| Hospital-eff-F | *mexV* |
| Hospital-eff-F | *blaSRT-1* |

| | |
|---|---|
| Hospital-eff-F | *blaDHA-13* |
| Hospital-eff-F | *vanS-Cd* |
| Hospital-eff-F | *blaACT-2* |
| Hospital-eff-F | *blaACT-57* |
| Hospital-eff-F | *blaACT-8* |
| Hospital-eff-F | *CMY2-MIR-ACT-EC* |
| Hospital-eff-F | *blaFOX-2* |
| Hospital-eff-F | *blaFOX-4* |
| Hospital-eff-F | *blaTRU-1* |
| Hospital-eff-F | *blaMOX-12* |
| Hospital-eff-F | *blaMOX-6* |
| Hospital-eff-F | *MexA* |
| Hospital-eff-F | *vanS-Pt* |
| Hospital-eff-F | *MexC* |
| Hospital-eff-F | *Tet*(53) |
| Hospital-eff-F | *blaLHK-2* |
| Hospital-eff-F | *blaACC-1c* |
| Hospital-eff-F | *AxyX* |
| Hospital-eff-F | *emeA* |
| Hospital-eff-F | *tet*(30) |
| Hospital-eff-F | *blaPDC-1* |
| Hospital-eff-F | *blaPDC-133* |
| Hospital-eff-F | *tet*(J) |
| Hospital-eff-F | *ceoA* |
| Hospital-eff-F | *sulX* |
| Hospital-eff-F | *tet*(42) |
| Hospital-eff-F | *cmlV* |
| Hospital-eff-F | *abeM* |
| Hospital-eff-F | *tet*(38) |
| Hospital-eff-F | *mtrE* |
| Hospital-eff-F | *rox* |
| Hospital-eff-F | *opmD* |
| Hospital-eff-F | *opmE* |
| Hospital-eff-F | *lsa(A)* |
| Hospital-eff-F | *TriC* |
| Hospital-eff-F | *emrB* |
| Hospital-eff-F | *tcr-3* |
| Hospital-eff-F | *mexN* |
| Hospital-eff-F | *oqxB24* |
| Hospital-eff-F | *oqxB31* |
| Hospital-eff-F | *tcmA* |
| Hospital-eff-F | *mcr-3.15* |
| Hospital-eff-F | *TlrC* |
| Hospital-eff-F | *mcr-3.10* |

| | |
|---|---|
| Hospital-eff-F | *srm(B)* |
| Hospital-eff-F | *facT* |
| Hospital-eff-F | *tet*(S) |
| Hospital-eff-F | *taeA* |
| Hospital-eff-F | *otrA* |
| Hospital-eff-F | *vanT-C* |
| Hospital-eff-F | *PBP1a* |
| Hospital-eff-F | *Listeria_monocytogenes_mprF* |
| Hospital-eff-F | *ceoB* |
| Hospital-eff-F | *adeB* |
| Hospital-eff-F | *cmeB* |
| Hospital-eff-F | *mexY* |
| Hospital-eff-F | *oqxB20* |
| Hospital-eff-F | *oqxB22* |
| Hospital-eff-F | *oqxB23* |
| Hospital-eff-F | *oqxB26* |
| Hospital-eff-F | *oqxB27* |
| Hospital-eff-F | *oqxB7* |
| Hospital-eff-F | *mdsB* |
| Hospital-eff-A | *msr*(E) |
| Hospital-eff-A | *mph*(E) |
| Hospital-eff-A | *tet*(39) |
| Hospital-eff-A | *erm*(B) |
| Hospital-eff-A | *blaMCA* |
| Hospital-eff-A | *blaOXA-164* |
| Hospital-eff-A | *aph(3'')-Ib* |
| Hospital-eff-A | *aph(6)-Id* |
| Hospital-eff-A | *aac3-IIe* |
| Hospital-eff-A | *aac6-Aph2* |
| Hospital-eff-A | *tet*(Q) |
| Hospital-eff-A | *AAC(6')-IIa* |
| Hospital-eff-A | *blaOXA-141* |
| Hospital-eff-A | *aadA27* |
| Hospital-eff-A | *blaOXA-10* |
| Hospital-eff-A | *blaOXA-139* |
| Hospital-eff-A | *blaOXA-211* |
| Hospital-eff-A | *aac(3)-I* |
| Hospital-eff-A | *tet*(W) |
| Hospital-eff-A | *blaOXA-280* |
| Hospital-eff-A | *sul1* |
| Hospital-eff-A | *sul2* |
| Hospital-eff-A | *te*t(O) |
| Hospital-eff-A | *ant(3'')-IIa* |
| Hospital-eff-A | *vanZ-A* |

| | |
|---|---|
| Hospital-eff-A | *vanY-A* |
| Hospital-eff-A | *blaOXA-281* |
| Hospital-eff-A | *tet*(M) |
| Hospital-eff-A | *AAC(6')-30/AAC(6')-Ib'_fusion_prote* |
| Hospital-eff-A | *aadA1* |
| Hospital-eff-A | *blaOXA-160* |
| Hospital-eff-A | *erm*(F) |
| Hospital-eff-A | *aacA-ACI1* |
| Hospital-eff-A | *blaOXA-333* |
| Hospital-eff-A | *mef*(A) |
| Hospital-eff-A | *aac3-IId* |
| Hospital-eff-A | *ant(2'')-Ia* |
| Hospital-eff-A | *aac(3)-IIe* |
| Hospital-eff-A | *msr*(D) |
| Hospital-eff-A | *blaPER-1* |
| Hospital-eff-A | *aph3-VIb* |
| Hospital-eff-A | *aac3-Ib* |
| Hospital-eff-A | *aacA38* |
| Hospital-eff-A | *tet*(C) |
| Hospital-eff-A | *aac3-Ib-Aac6-Ib* |
| Hospital-eff-A | *qacH* |
| Hospital-eff-A | *cfxA6* |
| Hospital-eff-A | *cfxA* |
| Hospital-eff-A | *ant(3'')-IIc* |
| Hospital-eff-A | *aph(3')-VIa* |
| Hospital-eff-A | *AAC(6')-Ib7* |
| Hospital-eff-A | *lnu(AN2)* |
| Hospital-eff-A | *blaOXA-2* |
| Hospital-eff-A | *aac3-IIa* |
| Hospital-eff-A | *blaOXA-296* |
| Hospital-eff-A | *aadE* |
| Hospital-eff-A | *Bifidobacterium_adolescentis_rpoB_* |
| Hospital-eff-A | *tet*(A) |
| Hospital-eff-A | *blaOXA-652* |
| Hospital-eff-A | *aadS* |
| Hospital-eff-A | *strB* |
| Hospital-eff-A | *Mef(En2)* |
| Hospital-eff-A | *ere(B)* |
| Hospital-eff-A | *TEM-1* |
| Hospital-eff-A | *blaOXA-643* |
| Hospital-eff-A | *tet*(40) |
| Hospital-eff-A | *tet*(X) |
| Hospital-eff-A | *mef*(C) |
| Hospital-eff-A | *cmxA* |

| | |
|---|---|
| Hospital-eff-A | *dfrF* |
| Hospital-eff-A | *aac3-I* |
| Hospital-eff-A | *blaOXA-212* |
| Hospital-eff-A | *blaOXA-662* |
| Hospital-eff-A | *blaOXA-18* |
| Hospital-eff-A | *blaOXA-373* |
| Hospital-eff-A | *erm*(G) |
| Hospital-eff-A | *aph(2")-IIa* |
| Hospital-eff-A | *blaPSE-4* |
| Hospital-eff-A | *blaOXA-347* |
| Hospital-eff-A | *tet*(32) |
| Hospital-eff-A | *aac(6')-Ib-G* |
| Hospital-eff-A | *vanA-G* |
| Hospital-eff-A | *catB3* |
| Hospital-eff-A | *tet*(R) |
| Hospital-eff-A | *aac6-Ig* |
| Hospital-eff-A | *gadX* |
| Hospital-eff-A | *aac3-IIc* |
| Hospital-eff-A | *blaOXA-334* |
| Hospital-eff-A | *CblA-1* |
| Hospital-eff-A | *blaOXA-15* |
| Hospital-eff-A | *blaOXA-1* |
| Hospital-eff-A | *aadA24* |
| Hospital-eff-A | *blaOXA-650* |
| Hospital-eff-A | *blaOXA-9* |
| Hospital-eff-A | *mph*(A) |
| Hospital-eff-A | *ere(D)* |
| Hospital-eff-A | *aph(3')-VI* |
| Hospital-eff-A | *aadA2* |
| Hospital-eff-A | *aadA5* |
| Hospital-eff-A | *aac(6')-Im* |
| Hospital-eff-A | *catA13* |
| Hospital-eff-A | *tet*(G) |
| Hospital-eff-A | *blaOXA-275* |
| Hospital-eff-A | *mdtF* |
| Hospital-eff-A | *vanA-A* |
| Hospital-eff-A | *mph*(G) |
| Hospital-eff-A | *vanH-A* |
| Hospital-eff-A | *vanS-A* |
| Hospital-eff-A | *vanX-A* |
| Hospital-eff-A | *blaOXA-24* |
| Hospital-eff-A | *floR* |
| Hospital-eff-A | *aph(3")-Ia* |
| Hospital-eff-A | *vanY-B* |

| | |
|---|---|
| Hospital-eff-A | *blaCTX-M-101* |
| Hospital-eff-A | *cpxA* |
| Hospital-eff-A | *aph4-Ia* |
| Hospital-eff-A | *erm*(T) |
| Hospital-eff-A | *CRP* |
| Hospital-eff-A | *vanX-B* |
| Hospital-eff-A | *vanR-A* |
| Hospital-eff-A | *aadA6* |
| Hospital-eff-A | *AAC(6')-Ib-cr* |
| Hospital-eff-A | *aph(3'')-III* |
| Hospital-eff-A | *aadB* |
| Hospital-eff-A | *mdtP* |
| Hospital-eff-A | *aac(3)-IVa* |
| Hospital-eff-A | *gadW* |
| Hospital-eff-A | *vanH-B* |
| Hospital-eff-A | *mdtE* |
| Hospital-eff-A | *emrA* |
| Hospital-eff-A | *tetR*(G) |
| Hospital-eff-A | *acrE* |
| Hospital-eff-A | *dfrA14* |
| Hospital-eff-A | *arr-8* |
| Hospital-eff-A | *Escherichia_coli_acrA* |
| Hospital-eff-A | *aac(3)-Ia* |
| Hospital-eff-A | *Penicillin_Binding_Protein_Ecoli* |
| Hospital-eff-A | *acrF* |
| Hospital-eff-A | *bacA* |
| Hospital-eff-A | *mdtN* |
| Hospital-eff-A | *vanW-B* |
| Hospital-eff-A | *aadA17* |
| Hospital-eff-A | *blaOXA-299* |
| Hospital-eff-A | *blaOXA-836* |
| Hospital-eff-A | *blaAER-1* |
| Hospital-eff-A | *eptA* |
| Hospital-eff-A | *blaOXA-645* |
| Hospital-eff-A | *Klebsiella_pneumoniae_KpnH* |
| Hospital-eff-A | *mdtO* |
| Hospital-eff-A | *mdtG* |
| Hospital-eff-A | *Bifidobacteria_intrinsic_ileS_confer* |
| Hospital-eff-A | *vanS-B* |
| Hospital-eff-A | *catS* |
| Hospital-eff-A | *baeS* |
| Hospital-eff-A | *yojI* |
| Hospital-eff-A | *acrB* |
| Hospital-eff-A | *acrS* |

| | |
|---|---|
| Hospital-eff-A | *msbA* |
| Hospital-eff-A | *dfr23* |
| Hospital-eff-A | *qnr-S1* |
| Hospital-eff-A | *lnu(C)* |
| Hospital-eff-A | *aac6-Im* |
| Hospital-eff-A | *vanR-B* |
| Hospital-eff-A | *mdtC* |
| Hospital-eff-A | *emrK* |
| Hospital-eff-A | *catA2* |
| Hospital-eff-A | *blaOXA-209* |
| Hospital-eff-A | *ant3''Ih-Aac6-IId* |
| Hospital-eff-A | *aacA-ACI6* |
| Hospital-eff-A | *aac(6')-30* |
| Hospital-eff-A | *mdtH* |
| Hospital-eff-A | *vanB* |
| Hospital-eff-A | *tolC* |
| Hospital-eff-A | *acrD* |
| Hospital-eff-A | *emrY* |
| Hospital-eff-A | *aad9* |
| Hospital-eff-A | *lin(B)* |
| Hospital-eff-A | *mdtB* |
| Hospital-eff-A | *baeR* |
| Hospital-eff-A | *emrR* |
| Hospital-eff-A | *mdtM* |
| Hospital-eff-A | *sat4* |
| Hospital-eff-A | *evgS* |
| Hospital-eff-A | *floR2* |
| Hospital-eff-A | *pmrF* |
| Hospital-eff-A | *aadA1-pm* |
| Hospital-eff-A | *ugd* |
| Hospital-eff-A | *Escherichia_coli_mdfA* |
| Hospital-eff-A | *qnrS2* |
| Hospital-eff-A | *kdpE* |
| Hospital-eff-A | *lsa(E)* |
| Hospital-eff-A | *ampH_Ecoli* |
| Hospital-eff-A | *dfrB1* |
| Hospital-eff-A | *cfxA_gen* |
| Hospital-eff-A | *aadA11* |
| Hospital-eff-A | *blaOXA-427* |
| Hospital-eff-A | *bla-A* |
| Hospital-eff-A | *cfr(C)* |
| Hospital-eff-A | *blaMOX-9* |
| Hospital-eff-A | *arr3* |
| Hospital-eff-A | *blaOXA-667* |

| | |
|---|---|
| Hospital-eff-A | *AmpC1_Ecoli* |
| Hospital-eff-A | *blaTEM-112* |
| Hospital-eff-A | *blaOXA-205* |
| Hospital-eff-A | *blaVIM-10* |
| Hospital-eff-A | *blaIMP-22* |
| Hospital-eff-A | *AAC(6')-Ib8* |
| Hospital-eff-A | *blaOXA-161* |
| Hospital-eff-A | *aac(6')-Ib-AKT* |
| Hospital-eff-A | *nimD_Nitroimidazole_Gene* |
| Hospital-eff-A | *aadA13* |
| Hospital-eff-A | *blaOXA-13* |
| Hospital-eff-A | *arr* |
| Hospital-eff-A | *arr2* |
| Hospital-eff-A | *blaOXA-668* |
| Hospital-eff-A | *ampH* |
| Hospital-eff-A | *mdtA* |
| Hospital-eff-A | *blaOXA-101* |
| Hospital-eff-A | *AmpC2_Ecoli* |
| Hospital-eff-A | *aph(3')-Ia* |
| Hospital-eff-A | *H-NS* |
| Hospital-eff-A | *Klebsiella_pneumoniae_KpnG* |
| Hospital-eff-A | *aadA16* |
| Hospital-eff-A | *blaSHV-107* |
| Hospital-eff-A | *AAC(6')-Ib-Suzhou* |
| Hospital-eff-A | *sat-2A* |
| Hospital-eff-A | *cblA* |
| Hospital-eff-A | *Salmonella_enterica_cmlA* |
| Hospital-eff-A | *aph7* |
| Hospital-eff-A | *Klebsiella_pneumoniae_acrA* |
| Hospital-eff-A | *blaOXA-230* |
| Hospital-eff-A | *msr*(C) |
| Hospital-eff-A | *aadA10* |
| Hospital-eff-A | *SHV-1* |
| Hospital-eff-A | *marA* |
| Hospital-eff-A | *aadA12* |
| Hospital-eff-A | *aadA25* |
| Hospital-eff-A | *blaOXA-504* |
| Hospital-eff-A | *tet*(B) |
| Hospital-eff-A | *tet*(E) |
| Hospital-eff-A | *evgA* |
| Hospital-eff-A | *fosA_gen* |
| Hospital-eff-A | *str* |
| Hospital-eff-A | *efmA* |
| Hospital-eff-A | *blaTEM-156* |

| | |
|---|---|
| Hospital-eff-A | *Escherichia_coli_emrE* |
| Hospital-eff-A | *tet*(44) |
| Hospital-eff-A | *eat(A)* |
| Hospital-eff-A | *spw* |
| Hospital-eff-A | *dfrA17* |
| Hospital-eff-A | *mphB* |
| Hospital-eff-A | *ICR-Mo* |
| Hospital-eff-A | *aac(3)-IIa* |
| Hospital-eff-A | *blaTEM-102* |
| Hospital-eff-A | *dfrG* |
| Hospital-eff-A | *blaCTX-M-103* |
| Hospital-eff-A | *blaRCP* |
| Hospital-eff-A | *tet*(36) |
| Hospital-eff-A | *CARB-3* |
| Hospital-eff-A | *catA1* |
| Hospital-eff-A | *MexF* |
| Hospital-eff-A | *catB4* |
| Hospital-eff-A | *blaOXA-309* |
| Hospital-eff-A | *aad(6)* |
| Hospital-eff-A | *aac(6')-Ib-cr3* |
| Hospital-eff-A | *blaOXA-420* |
| Hospital-eff-A | *blaTEM-10* |
| Hospital-eff-A | *CARB-2* |
| Hospital-eff-A | *blaBES-1* |
| Hospital-eff-A | *cphA2* |
| Hospital-eff-A | *cmlA1* |
| Hospital-eff-A | *tetA(P)* |
| Hospital-eff-A | *blaOXA-724* |
| Hospital-eff-A | *blaOXA-256* |
| Hospital-eff-A | *blaOXA-46* |
| Hospital-eff-A | *blaEC* |
| Hospital-eff-A | *vanT-G* |
| Hospital-eff-A | *nimj_Nitroimidazole_Gene* |
| Hospital-eff-A | *blaOXA-356* |
| Hospital-eff-A | *SHV-12* |
| Hospital-eff-A | *erm*(X) |
| Hospital-eff-A | *ant6-Ib* |
| Hospital-eff-A | *cfr-Cb* |
| Hospital-eff-A | *nimA_Nitroimidazole_Gene* |
| Hospital-eff-A | *tetB-P* |
| Hospital-eff-A | *aph(2'')-If* |
| Hospital-eff-A | *cepA_beta-lactamase* |
| Hospital-eff-A | *Klebsiella_pneumoniae_OmpK37* |
| Hospital-eff-A | *ramA* |

| | |
|---|---|
| Hospital-eff-A | *cepS* |
| Hospital-eff-A | *cfxA3* |
| Hospital-eff-A | *OqxA* |
| Hospital-eff-A | *oqxB11* |
| Hospital-eff-A | *blaOXA-726* |
| Hospital-eff-A | *mcr-10.1* |
| Hospital-eff-A | *ere(A)* |
| Hospital-eff-A | *blaOXA-228* |
| Hospital-eff-A | *mph*(D) |
| Hospital-eff-A | *vanYD* |
| Hospital-eff-A | *OqxBgb* |
| Hospital-eff-A | *qnrVC1* |
| Hospital-eff-A | *blaEC-19* |
| Hospital-eff-A | *qnrB17* |
| Hospital-eff-A | *blaMOX-2* |
| Hospital-eff-A | *mcr-5.1* |
| Hospital-eff-A | *dfrA27* |
| Hospital-eff-A | *Enterobacter_cloacae_acrA* |
| Hospital-eff-A | *lsa(C)* |
| Hospital-eff-A | *tetB*(46) |
| Hospital-eff-A | *erm*(Q) |
| Hospital-eff-A | *blaNPS-1* |
| Hospital-eff-A | *cfr(B)* |
| Hospital-eff-A | *aadA15* |
| Hospital-eff-A | *aac6-Ii* |
| Hospital-eff-A | *blaOXA-229* |
| Hospital-eff-A | *aac(6')-set_A* |
| Hospital-eff-A | *blaOXA-512* |
| Hospital-eff-A | *blaOXA-96* |
| Hospital-eff-A | *aci1* |
| Hospital-eff-A | *blaDHA-1* |
| Hospital-eff-A | *RSA-1* |
| Hospital-eff-A | *blaSHV-35* |
| Hospital-eff-A | *sul4* |
| Hospital-eff-A | *Nocardia_rifampin_resistant_beta-s* |
| Hospital-eff-A | *blaCTX-M-1* |
| Hospital-eff-A | *blaCTX-M-182* |
| Hospital-eff-A | *oqxA10* |
| Hospital-eff-A | *tet*(H) |
| Hospital-eff-A | *aph(2'')-Id* |
| Hospital-eff-A | *AAC(6')-Ib10* |
| Hospital-eff-A | *ere(A2)* |
| Hospital-eff-A | *catP* |
| Hospital-eff-A | *blaBKC-1* |

| | |
|---|---|
| Hospital-eff-A | *cfxA5* |
| Hospital-eff-A | *qnrB1* |
| Hospital-eff-A | *vanH-D* |
| Hospital-eff-A | *qnrB10* |
| Hospital-eff-A | *qnrB26* |
| Hospital-eff-A | *tet*(L) |
| Hospital-eff-A | *tetA(46)* |
| Hospital-eff-A | *vanG2* |
| Hospital-eff-A | *oqxB15* |
| Hospital-eff-A | *oqxB9* |
| Hospital-eff-A | *efrB* |
| Hospital-eff-A | *Klebsiella_pneumoniae_KpnE* |
| Hospital-eff-A | *erm*(47) |
| Hospital-eff-A | *MecI* |
| Hospital-eff-A | *blaCMY-103* |
| Hospital-eff-A | *cphA1* |
| Hospital-eff-A | *vanXY-G* |
| Hospital-eff-A | *blaLCR-1* |
| Hospital-eff-A | *erm*(O) |
| Hospital-eff-A | *oqxA3* |
| Hospital-eff-A | *blaOXA-142* |
| Hospital-eff-A | *blaOXA-240* |
| Hospital-eff-A | *blaOXA-824* |
| Hospital-eff-A | *blaOXA-827* |
| Hospital-eff-A | *blaVIM-11* |
| Hospital-eff-A | *vanZ-Pa* |
| Hospital-eff-A | *mef*(B) |
| Hospital-eff-A | *blaOXA-644* |
| Hospital-eff-A | *nshR* |
| Hospital-eff-A | *blaOXA-392* |
| Hospital-eff-A | *blaOXA-257* |
| Hospital-eff-A | *cmlA5* |
| Hospital-eff-A | *FosA5* |
| Hospital-eff-A | *FosA6* |
| Hospital-eff-A | *blaOXA-397* |
| Hospital-eff-A | *vanW-G* |
| Hospital-eff-A | *fosA* |
| Hospital-eff-A | *blaLAP-1* |
| Hospital-eff-A | *fosB-251804940* |
| Hospital-eff-A | *blaSHV-67* |
| Hospital-eff-A | *blaGES-1* |
| Hospital-eff-A | *TriC* |
| Hospital-eff-A | *blaCPS-1* |
| Hospital-eff-A | *blaCTX-M-105* |

| | |
|---|---|
| Hospital-eff-A | *cepA* |
| Hospital-eff-A | *blaL2* |
| Hospital-eff-A | *erm*(49) |
| Hospital-eff-A | *aac3-Ih* |
| Hospital-eff-A | *aph(2")-Ig* |
| Hospital-eff-A | *blaPER-12* |
| Hospital-eff-A | *dfrA1* |
| Hospital-eff-A | *dhfr7* |
| Hospital-eff-A | *PmpM* |
| Hospital-eff-A | *dfrA3* |
| Hospital-eff-A | *dfrE* |
| Hospital-eff-A | *dfrA12* |
| Hospital-eff-A | *BlaA1* |
| Hospital-eff-A | *emrB* |
| Hospital-eff-A | *mcr-9.1* |
| Hospital-eff-A | *mcr-3.17* |
| Hospital-eff-A | *aac(6')-Ib-cr10* |
| Hospital-eff-A | *satA_Ba* |
| Hospital-eff-A | *dfr1_rpt* |
| Hospital-eff-A | *dfrA10* |
| Hospital-eff-A | *mexV* |
| Hospital-eff-A | *blaEC-13* |
| Hospital-eff-A | *blaEC-5* |
| Hospital-eff-A | *dfrA19* |
| Hospital-eff-A | *blaACT-13* |
| Hospital-eff-A | *efrA* |
| Hospital-eff-A | *mexM* |
| Hospital-eff-A | *blaACC-1* |
| Hospital-eff-A | *vanS-Pt* |
| Hospital-eff-A | *blaOCH-1* |
| Hospital-eff-A | *oqxA11* |
| Hospital-eff-A | *tet*(D) |
| Hospital-eff-A | *beta-lactamase_class-C* |
| Hospital-eff-A | *vanX-D* |
| Hospital-eff-A | *tet*(33) |
| Hospital-eff-A | *MexB* |
| Hospital-eff-A | *cmlA* |
| Hospital-eff-A | *catB7* |
| Hospital-eff-A | *tet*(S) |
| Hospital-eff-A | *qnrD* |
| Hospital-eff-A | *qnrB12* |
| Hospital-eff-A | *qnr-A1* |
| Hospital-eff-A | *catQ* |
| Hospital-eff-A | *Pseudomonas_aeruginosa_CpxR* |

| | |
|---|---|
| Hospital-eff-A | *smeR* |
| Hospital-eff-A | *vanRc3* |
| Hospital-eff-A | *vanR-D* |
| Hospital-eff-A | *vanR-O* |
| Hospital-eff-A | *Streptomyces_rishiriensis_parY_mut* |
| Hospital-eff-A | *blaCAM-1* |
| Hospital-eff-A | *erm*(A) |
| Hospital-eff-A | *blaIMP-13* |
| Hospital-eff-A | *lsa(A)* |
| Hospital-eff-A | *rmtH* |
| Hospital-eff-A | *blaOXA-460* |
| Hospital-eff-A | *mexW* |
| Hospital-eff-A | *cepH-A3* |
| Hospital-eff-A | *imiH* |
| Hospital-eff-A | *blaOXA-33* |
| Hospital-eff-A | *qepA2* |
| Hospital-eff-A | *blaOXA-589* |
| Hospital-eff-A | *aphA6* |
| Hospital-eff-A | *oqxB19* |
| Hospital-eff-A | *aadA4* |
| Hospital-eff-A | *blaLEN-11* |
| Hospital-eff-A | *blaOXA-485* |
| Hospital-eff-A | *aadA3* |
| Hospital-eff-A | *aph3-Vb* |
| Hospital-eff-A | *adeJ* |
| Hospital-eff-A | *blaOXA-725* |
| Hospital-eff-A | *blaOXA-780* |
| Hospital-eff-A | *aadA8* |
| Hospital-eff-A | *aph(3')-IX* |
| Hospital-eff-A | *blaOXA-118* |
| Hospital-eff-A | *blaOXA-119* |
| Hospital-eff-A | *blaOXA-20* |
| Hospital-eff-A | *blaOXA-233* |
| Hospital-eff-A | *blaOXA-251* |
| Hospital-eff-A | *blaOXA-655* |
| Hospital-eff-A | *blaOXA-779* |
| Hospital-eff-A | *aph(3')-IIc* |
| Hospital-eff-A | *blaOXA-129* |
| Hospital-eff-A | *lnu((B)* |
| Hospital-eff-A | *blaOXA-136* |
| Hospital-eff-A | *sph-I* |
| Hospital-eff-A | *tsnR* |
| Hospital-eff-A | *aph(3')-Ib* |
| Hospital-eff-A | *lnu(F)* |

| | |
|---|---|
| Hospital-eff-A | *tlr(C)* |
| Hospital-eff-A | *blaOXA-22* |
| Hospital-eff-A | *blaOXA-441* |
| Hospital-eff-A | *blaOXA-651* |
| Hospital-eff-A | *blaOXA-114* |
| Hospital-eff-A | *blaOXA-114u* |
| Hospital-eff-A | *blaOXA-25* |
| Hospital-eff-A | *blaOXA-26* |
| Hospital-eff-A | *blaOXA-32* |
| Hospital-eff-A | *blaOXA-364* |
| Hospital-eff-A | *blaOXA-415* |
| Hospital-eff-A | *blaOXA-224* |
| Hospital-eff-A | *blaOXA-355* |
| Hospital-eff-A | *blaOXA-568* |
| Hospital-eff-A | *blaSPN79-1* |
| Hospital-eff-A | *blaLEN-1* |
| Hospital-eff-A | *erm*(32) |
| Hospital-eff-A | *otr(B)* |
| Hospital-eff-A | *aac3-Xa* |
| Hospital-eff-A | *blaTER-2* |
| Hospital-eff-A | *ant(6)-Ic* |
| Hospital-eff-A | *blaOKP-B-1* |
| Hospital-eff-A | *blaSHV-105* |
| Hospital-eff-A | *blaSHV-11* |
| Hospital-eff-A | *blaSHV-115* |
| Hospital-eff-A | *blaSHV-119* |
| Hospital-eff-A | *blaSHV-128* |
| Hospital-eff-A | *blaSHV-164* |
| Hospital-eff-A | *blaSHV-179* |
| Hospital-eff-A | *blaSHV-40* |
| Hospital-eff-A | *blaTEM-145* |
| Hospital-eff-A | *blaTEM-21* |
| Hospital-eff-A | *blaTEM-213* |
| Hospital-eff-A | *blaTEM-93* |
| Hospital-eff-A | *CARB-1* |
| Hospital-eff-A | *CARB-6* |
| Hospital-eff-A | *blaL1* |
| Hospital-eff-A | *blaESP-1* |
| Hospital-eff-A | *blaOXY5-1* |
| Hospital-eff-A | *blaOXY1-1* |
| Hospital-eff-A | *blaIMI-16* |
| Hospital-eff-A | *blaOXA-154* |
| Hospital-eff-A | *blaPAU-1* |
| Hospital-eff-A | *blaA* |

| | |
|---|---|
| Hospital-eff-A | *blaSED* |
| Hospital-eff-A | *aph(2")-Ib* |
| Hospital-eff-A | *blaVEB-1* |
| Hospital-eff-A | *mph*(F) |
| Hospital-eff-A | *blaAIM-1* |
| Hospital-eff-A | *blaR39* |
| Hospital-eff-A | *blaCBP-1* |
| Hospital-eff-A | *blaPER-11* |
| Hospital-eff-A | *blaPER-7* |
| Hospital-eff-A | *blaAST-1* |
| Hospital-eff-A | *erm*(W) |
| Hospital-eff-A | *blaTHIN-B* |
| Hospital-eff-A | *erm*(S) |
| Hospital-eff-A | *cfxA4* |
| Hospital-eff-A | *tet*(T) |
| Hospital-eff-A | *otrA* |
| Hospital-eff-A | *lmrD* |
| Hospital-eff-A | *aph4-Ib* |
| Hospital-eff-A | *MSI-1* |
| Hospital-eff-A | *ceoB* |
| Hospital-eff-A | *vanD* |
| Hospital-eff-A | *MuxC* |
| Hospital-eff-A | *oqxB10* |
| Hospital-eff-A | *oqxB14* |
| Hospital-eff-A | *oqxB16* |
| Hospital-eff-A | *vanE* |
| Hospital-eff-A | *vanY-D* |
| Hospital-eff-A | *blaCSA-2* |
| Hospital-eff-A | *vanS-C* |
| Hospital-eff-A | *blaEC-14* |
| Hospital-eff-A | *blaEC-18* |
| Hospital-eff-A | *blaDHA-13* |
| Hospital-eff-A | *blaDHA-29* |
| Hospital-eff-A | *blaDHA-4* |
| Hospital-eff-A | *blaACT-3* |
| Hospital-eff-A | *blaACT-1* |
| Hospital-eff-A | *blaACT-28* |
| Hospital-eff-A | *CMY2-MIR-ACT-EC* |
| Hospital-eff-A | *erm*(E) |
| Hospital-eff-A | *blaCMY-FOX* |
| Hospital-eff-A | *blaFOX-15* |
| Hospital-eff-A | *cepH* |
| Hospital-eff-A | *blaMOX-8* |
| Hospital-eff-A | *blaLHK-1* |

| | |
|---|---|
| Hospital-eff-A | *Tet*(54) |
| Hospital-eff-A | *bmr* |
| Hospital-eff-A | *blaLHK-6* |
| Hospital-eff-A | *oqxA5* |
| Hospital-eff-A | *oqxA6* |
| Hospital-eff-A | *emeA* |
| Hospital-eff-A | *smeD* |
| Hospital-eff-A | *dha1* |
| Hospital-eff-A | *blaPDC-109* |
| Hospital-eff-A | *amrA* |
| Hospital-eff-A | *ceoA* |
| Hospital-eff-A | *tap* |
| Hospital-eff-A | *mgt* |
| Hospital-eff-A | *cmlA4* |
| Hospital-eff-A | *tetA-P* |
| Hospital-eff-A | *BlaA2* |
| Hospital-eff-A | *oleI* |
| Hospital-eff-A | *Acinetobacter_baumannii_AbaQ* |
| Hospital-eff-A | *cdeA* |
| Hospital-eff-A | *tet*(K) |
| Hospital-eff-A | *OprZ* |
| Hospital-eff-A | *smeS* |
| Hospital-eff-A | *basS* |
| Hospital-eff-A | *lmr(A)* |
| Hospital-eff-A | *adeK* |
| Hospital-eff-A | *Acinetobacter_baumannii_AmvA* |
| Hospital-eff-A | *lrfA* |
| Hospital-eff-A | *opcM* |
| Hospital-eff-A | *mexK* |
| Hospital-eff-A | *tcr-3* |
| Hospital-eff-A | *smeE* |
| Hospital-eff-A | *AxyY* |
| Hospital-eff-A | *vga(A)* |
| Hospital-eff-A | *vga(A)-LC* |
| Hospital-eff-A | *oqxB17* |
| Hospital-eff-A | *oqxB18* |
| Hospital-eff-A | *oqxB21* |
| Hospital-eff-A | *oqxB22* |
| Hospital-eff-A | *mcr-2.3* |
| Hospital-eff-A | *mcr-3.15* |
| Hospital-eff-A | *mcr-3.3* |
| Hospital-eff-A | *mcr-3.6* |
| Hospital-eff-A | *mcr-3.1* |
| Hospital-eff-A | *poxtA* |

| | |
|---|---|
| Hospital-eff-A | *lmrC* |
| Hospital-eff-A | *facT* |
| Hospital-eff-A | *novA* |
| Hospital-eff-A | *arnA* |
| Hospital-eff-A | *mecB* |
| Hospital-eff-A | *vanTc2* |
| Hospital-eff-A | *PBP1a* |
| Hospital-eff-A | *Streptococcus_agalactiae_mprF* |
| Hospital-eff-A | *rph* |
| Hospital-eff-A | *mupA* |
| Hospital-eff-A | *mexI* |
| Hospital-eff-A | *cmeB* |
| Hospital-eff-A | *MexD* |
| Hospital-eff-A | *smeB* |
| Hospital-eff-A | *oqxB13* |
| Hospital-eff-A | *oqxB20* |
| Hospital-eff-A | *oqxB24* |
| Hospital-eff-A | *oqxB8* |
| Hospital-eff-A | *mexQ* |
| Hospital-eff-A | *adeF* |
| Hospital-eff-A | *abcA* |
| STP-inf-F | *tet*(Q) |
| STP-inf-F | *tet*(W) |
| STP-inf-F | *msr*(E) |
| STP-inf-F | *erm*(B) |
| STP-inf-F | *mph*(E) |
| STP-inf-F | *tet*(O) |
| STP-inf-F | *tet*(39) |
| STP-inf-F | *erm*(F) |
| STP-inf-F | *cfxA6* |
| STP-inf-F | *mef*(A) |
| STP-inf-F | *aadE* |
| STP-inf-F | *qacH* |
| STP-inf-F | *tet*(40) |
| STP-inf-F | *msr*(D) |
| STP-inf-F | *cfxA* |
| STP-inf-F | *tet*(X) |
| STP-inf-F | *Mef(En2)* |
| STP-inf-F | *vanA-G* |
| STP-inf-F | *lnu(AN2)* |
| STP-inf-F | *dfrF* |
| STP-inf-F | *aadS* |
| STP-inf-F | *mef*(C) |
| STP-inf-F | *tet*(32) |

| | |
|---|---|
| STP-inf-F | *tet*(M) |
| STP-inf-F | *aph(6)-Id* |
| STP-inf-F | *aph(3")-Ib* |
| STP-inf-F | *CblA-1* |
| STP-inf-F | *blaMCA* |
| STP-inf-F | *blaOXA-296* |
| STP-inf-F | *catA13* |
| STP-inf-F | *sul1* |
| STP-inf-F | *blaOXA-141* |
| STP-inf-F | *Bifidobacteria_intrinsic_ileS_confer* |
| STP-inf-F | *mph*(G) |
| STP-inf-F | *blaOXA-333* |
| STP-inf-F | *tet*(C) |
| STP-inf-F | *blaOXA-129* |
| STP-inf-F | *aadA27* |
| STP-inf-F | *ere(D)* |
| STP-inf-F | *sul2* |
| STP-inf-F | *aph(3")-III* |
| STP-inf-F | *blaOXA-10* |
| STP-inf-F | *blaAER-1* |
| STP-inf-F | *tet*(A) |
| STP-inf-F | *tet*(G) |
| STP-inf-F | *erm*(G) |
| STP-inf-F | *lnu(C)* |
| STP-inf-F | *AAC(6')-IIa* |
| STP-inf-F | *cmxA* |
| STP-inf-F | *tet*(R) |
| STP-inf-F | *aac(3)-I* |
| STP-inf-F | *aadA5* |
| STP-inf-F | *cfr(C)* |
| STP-inf-F | *blaOXA-164* |
| STP-inf-F | *ant(3")-IIa* |
| STP-inf-F | *aad9* |
| STP-inf-F | *catS* |
| STP-inf-F | *blaOXA-211* |
| STP-inf-F | *lsa(E)* |
| STP-inf-F | *lin(B)* |
| STP-inf-F | *blaOXA-118* |
| STP-inf-F | *blaOXA-20* |
| STP-inf-F | *bla-A* |
| STP-inf-F | *cat-TC* |
| STP-inf-F | *cblA* |
| STP-inf-F | *blaOXA-427* |
| STP-inf-F | *tetR*(G) |

| | |
|---|---|
| STP-inf-F | *blaOXA-392* |
| STP-inf-F | *sat4* |
| STP-inf-F | *tet*(44) |
| STP-inf-F | *aadA6* |
| STP-inf-F | *cfxA_gen* |
| STP-inf-F | *mdtO* |
| STP-inf-F | *cfxA3* |
| STP-inf-F | *blaOXA-280* |
| STP-inf-F | *blaOXA-1* |
| STP-inf-F | *tet*(S) |
| STP-inf-F | *aac6-Aph2* |
| STP-inf-F | *aadA2* |
| STP-inf-F | *spw* |
| STP-inf-F | *Klebsiella_pneumoniae_KpnG* |
| STP-inf-F | *blaOXA-347* |
| STP-inf-F | *gadX* |
| STP-inf-F | *mdtE* |
| STP-inf-F | *CRP* |
| STP-inf-F | *aadA1* |
| STP-inf-F | *lnu(G)* |
| STP-inf-F | *tolC* |
| STP-inf-F | *ampH* |
| STP-inf-F | *aadA11* |
| STP-inf-F | *AAC(6')-Ib7* |
| STP-inf-F | *msbA* |
| STP-inf-F | *mdtP* |
| STP-inf-F | *blaOXA-205* |
| STP-inf-F | *Escherichia_coli_mdfA* |
| STP-inf-F | *acrF* |
| STP-inf-F | *cfr-Cb* |
| STP-inf-F | *acrE* |
| STP-inf-F | *mdtB* |
| STP-inf-F | *ICR-Mo* |
| STP-inf-F | *mdtF* |
| STP-inf-F | *Escherichia_coli_acrA* |
| STP-inf-F | *Klebsiella_pneumoniae_acrA* |
| STP-inf-F | *acrB* |
| STP-inf-F | *blaOXA-666* |
| STP-inf-F | *blaOXA-119* |
| STP-inf-F | *baeS* |
| STP-inf-F | *Klebsiella_pneumoniae_KpnH* |
| STP-inf-F | *mdtM* |
| STP-inf-F | *H-NS* |
| STP-inf-F | *evgS* |

| | |
|---|---|
| STP-inf-F | *floR2* |
| STP-inf-F | *ampH_Ecoli* |
| STP-inf-F | *emrA* |
| STP-inf-F | *aac(6')-30* |
| STP-inf-F | *yojI* |
| STP-inf-F | *Klebsiella_pneumoniae_OmpK37* |
| STP-inf-F | *emrR* |
| STP-inf-F | *strB* |
| STP-inf-F | *cpxA* |
| STP-inf-F | *mdtC* |
| STP-inf-F | *aadA17* |
| STP-inf-F | *Nocardia_rifampin_resistant_beta-s* |
| STP-inf-F | *pmrF* |
| STP-inf-F | *emrY* |
| STP-inf-F | *catA9* |
| STP-inf-F | *mef*(B) |
| STP-inf-F | *blaOXA-464* |
| STP-inf-F | *mdtN* |
| STP-inf-F | *acrD* |
| STP-inf-F | *catP* |
| STP-inf-F | *blaOXA-5* |
| STP-inf-F | *aph7* |
| STP-inf-F | *aadA10* |
| STP-inf-F | *qnrS2* |
| STP-inf-F | *mdtG* |
| STP-inf-F | *Penicillin_Binding_Protein_Ecoli* |
| STP-inf-F | *mphB* |
| STP-inf-F | *eptA* |
| STP-inf-F | *ugd* |
| STP-inf-F | *aadA24* |
| STP-inf-F | *nimD_Nitroimidazole_Gene* |
| STP-inf-F | *blaAIM-1* |
| STP-inf-F | *bacA* |
| STP-inf-F | *MexF* |
| STP-inf-F | *baeR* |
| STP-inf-F | *blaOXA-2* |
| STP-inf-F | *mdtA* |
| STP-inf-F | *blaMOX-9* |
| STP-inf-F | *emrK* |
| STP-inf-F | *nimA_Nitroimidazole_Gene* |
| STP-inf-F | *erm*(X) |
| STP-inf-F | *OqxA* |
| STP-inf-F | *ant3''Ih-Aac6-IId* |
| STP-inf-F | *ant6-Ib* |

| | |
|---|---|
| STP-inf-F | *aac3-IIe* |
| STP-inf-F | *mdtH* |
| STP-inf-F | *catQ* |
| STP-inf-F | *blaRCP* |
| STP-inf-F | *aadA13* |
| STP-inf-F | *RbpA* |
| STP-inf-F | *aci1* |
| STP-inf-F | *evgA* |
| STP-inf-F | *aac3-Ib-Aac6-Ib* |
| STP-inf-F | *OqxBgb* |
| STP-inf-F | *cepA* |
| STP-inf-F | *erm*(Q) |
| STP-inf-F | *mph*(A) |
| STP-inf-F | *ant(3")-IIc* |
| STP-inf-F | *acrS* |
| STP-inf-F | *tet*(E) |
| STP-inf-F | *blaOXA-209* |
| STP-inf-F | *blaOXA-281* |
| STP-inf-F | *fosA_gen* |
| STP-inf-F | *FosA5* |
| STP-inf-F | *vanW-G* |
| STP-inf-F | *AmpC1_Ecoli* |
| STP-inf-F | *gadW* |
| STP-inf-F | *floR* |
| STP-inf-F | *aac(3)-Ia* |
| STP-inf-F | *catB3* |
| STP-inf-F | *aadA12* |
| STP-inf-F | *iri* |
| STP-inf-F | *blaEC* |
| STP-inf-F | *blaOXA-643* |
| STP-inf-F | *blaOXA-650* |
| STP-inf-F | *aad(6)* |
| STP-inf-F | *Escherichia_coli_emrE* |
| STP-inf-F | *nimE_Nitroimidazole_Gene* |
| STP-inf-F | *aac3-Ib* |
| STP-inf-F | *ant(2")-Ia* |
| STP-inf-F | *aph(2")-IIa* |
| STP-inf-F | *mcr-3.6* |
| STP-inf-F | *Klebsiella_pneumoniae_KpnE* |
| STP-inf-F | *erm*(T) |
| STP-inf-F | *aac(6')-Ib-G* |
| STP-inf-F | *erm*(47) |
| STP-inf-F | *marA* |
| STP-inf-F | *AAC(6')-30/AAC(6')-Ib'_fusion_prote* |

| | |
|---|---|
| STP-inf-F | *blaRSC1-1* |
| STP-inf-F | *blaOXA-725* |
| STP-inf-F | *lmrD* |
| STP-inf-F | *blaOXA-101* |
| STP-inf-F | *blaOXA-37* |
| STP-inf-F | *lnu((B)* |
| STP-inf-F | *vanX-A* |
| STP-inf-F | *blaOXA-212* |
| STP-inf-F | *blaOXA-662* |
| STP-inf-F | *ere(B)* |
| STP-inf-F | *FosA6* |
| STP-inf-F | *str* |
| STP-inf-F | *blaBEL-1* |
| STP-inf-F | *tet*(36) |
| STP-inf-F | *qnrB19* |
| STP-inf-F | *aacA-ACI1* |
| STP-inf-F | *blaPAU-1* |
| STP-inf-F | *CARB-10* |
| STP-inf-F | *Pseudomonas_aeruginosa_CpxR* |
| STP-inf-F | *blaEC-19* |
| STP-inf-F | *mcr-3.1* |
| STP-inf-F | *oqxA10* |
| STP-inf-F | *vanT-G* |
| STP-inf-F | *vanZ-A* |
| STP-inf-F | *erm*(A) |
| STP-inf-F | *tetB*(46) |
| STP-inf-F | *nimj_Nitroimidazole_Gene* |
| STP-inf-F | *aph(3')-VI* |
| STP-inf-F | *cfr(B)* |
| STP-inf-F | *sat-2A* |
| STP-inf-F | *vanG2* |
| STP-inf-F | *oqxB11* |
| STP-inf-F | *blaOXA-780* |
| STP-inf-F | *aadB* |
| STP-inf-F | *lnu(F)* |
| STP-inf-F | *blaOXA-224* |
| STP-inf-F | *aacA38* |
| STP-inf-F | *tet*(T) |
| STP-inf-F | *mexK* |
| STP-inf-F | *aadA16* |
| STP-inf-F | *blaEC-5* |
| STP-inf-F | *aac(3)-IIe* |
| STP-inf-F | *aac3-IId* |
| STP-inf-F | *blaSHV-1* |

| | |
|---|---|
| STP-inf-F | *blaGES-10* |
| STP-inf-F | *cepA_beta-lactamase* |
| STP-inf-F | *vanY-A* |
| STP-inf-F | *tet*(33) |
| STP-inf-F | *ere(A2)* |
| STP-inf-F | *tetA(P)* |
| STP-inf-F | *mcr-3.17* |
| STP-inf-F | *mcr-5.1* |
| STP-inf-F | *Klebsiella_pneumoniae_KpnF* |
| STP-inf-F | *kdpE* |
| STP-inf-F | *mtrA* |
| STP-inf-F | *vanR-D* |
| STP-inf-F | *vanS-D* |
| STP-inf-F | *mexW* |
| STP-inf-F | *cepH-A3* |
| STP-inf-F | *cphA2* |
| STP-inf-F | *vanXY-G* |
| STP-inf-F | *blaOXA-33* |
| STP-inf-F | *tet*(Z) |
| STP-inf-F | *smeE* |
| STP-inf-F | *tetB-P* |
| STP-inf-F | *oqxB10* |
| STP-inf-F | *aadA25* |
| STP-inf-F | *blaOXA-504* |
| STP-inf-F | *blaOXA-726* |
| STP-inf-F | *blaOXA-142* |
| STP-inf-F | *erm*(35) |
| STP-inf-F | *blaOXA-309* |
| STP-inf-F | *blaOXA-373* |
| STP-inf-F | *blaOXA-652* |
| STP-inf-F | *blaOXA-9* |
| STP-inf-F | *blaOXA-15* |
| STP-inf-F | *blaOXA-18* |
| STP-inf-F | *vanW-B* |
| STP-inf-F | *aadA1-pm* |
| STP-inf-F | *blaOXA-675* |
| STP-inf-F | *FosB6* |
| STP-inf-F | *tetA-P* |
| STP-inf-F | *blaSHV-179* |
| STP-inf-F | *blaTEM-93* |
| STP-inf-F | *blaTEM-1* |
| STP-inf-F | *tetA(46)* |
| STP-inf-F | *blaL1* |
| STP-inf-F | *blaOXY2-1* |

| | |
|---|---|
| STP-inf-F | *aac6-Ik* |
| STP-inf-F | *aacA34* |
| STP-inf-F | *blaLRA-1* |
| STP-inf-F | *aph(2")-If* |
| STP-inf-F | *CARB-14/blaRTG* |
| STP-inf-F | *blaVEB-1* |
| STP-inf-F | *blaVEB-1a* |
| STP-inf-F | *oqxB15* |
| STP-inf-F | *blaPER-7* |
| STP-inf-F | *dfrA14* |
| STP-inf-F | *eat(A)* |
| STP-inf-F | *MuxB* |
| STP-inf-F | *oqxB12* |
| STP-inf-F | *oqxB19* |
| STP-inf-F | *adeJ* |
| STP-inf-F | *aac3-I* |
| STP-inf-F | *vanY-D* |
| STP-inf-F | *aac(6')-Im* |
| STP-inf-F | *aac6-Im* |
| STP-inf-F | *mcr-4.1* |
| STP-inf-F | *sat3* |
| STP-inf-F | *aac(6')-Iid* |
| STP-inf-F | *catB4* |
| STP-inf-F | *aac(6')-Iaj* |
| STP-inf-F | *blaACT-1* |
| STP-inf-F | *vanXY-c4* |
| STP-inf-F | *blaMOX-2* |
| STP-inf-F | *blaMOX-12* |
| STP-inf-F | *blaMOX-6* |
| STP-inf-F | *mexP* |
| STP-inf-F | *aac6-IIc* |
| STP-inf-F | *blaLHK-1* |
| STP-inf-F | *Salmonella_enterica_cmlA* |
| STP-inf-F | *cmr* |
| STP-inf-F | *dfrA36* |
| STP-inf-F | *oqxA11* |
| STP-inf-F | *tet*(Y) |
| STP-inf-F | *smeD* |
| STP-inf-F | *Enterobacter_cloacae_acrA* |
| STP-inf-F | *vanX-B* |
| STP-inf-F | *vanX-D* |
| STP-inf-F | *ere(A)* |
| STP-inf-F | *cmlA5* |
| STP-inf-F | *smeB* |

| | |
|---|---|
| STP-inf-F | *oqxB14* |
| STP-inf-F | *catB6* |
| STP-inf-F | *catB8* |
| STP-inf-F | *fusF* |
| STP-inf-F | *catA16* |
| STP-inf-F | *vanS-B* |
| STP-inf-F | *tet*(L) |
| STP-inf-F | *Mbl* |
| STP-inf-F | *vanR-A* |
| STP-inf-F | *blaSHN-1* |
| STP-inf-F | *basS* |
| STP-inf-F | *adeK* |
| STP-inf-F | *msr*(A) |
| STP-inf-F | *lsa(C)* |
| STP-inf-F | *nd* |
| STP-inf-F | *blaSIM-1* |
| STP-inf-F | *blaOXA-448* |
| STP-inf-F | *aph3-VIIa* |
| STP-inf-F | *cphA1* |
| STP-inf-F | *imiH* |
| STP-inf-F | *opcM* |
| STP-inf-F | *mexI* |
| STP-inf-F | *armA* |
| STP-inf-F | *blaRSD2-1* |
| STP-inf-F | *blaNPS-1* |
| STP-inf-F | *MexB* |
| STP-inf-F | *oqxB17* |
| STP-inf-F | *oqxB7* |
| STP-inf-F | *blaOXA-198* |
| STP-inf-F | *ant(3")-IIb* |
| STP-inf-F | *adeF* |
| STP-inf-F | *aphA15* |
| STP-inf-F | *blaOXA-12* |
| STP-inf-F | *blaOXA-724* |
| STP-inf-F | *aadA7* |
| STP-inf-F | *blaOXA-13* |
| STP-inf-F | *blaOXA-46* |
| STP-inf-F | *blaOXA-732* |
| STP-inf-F | *blaOXA-836* |
| STP-inf-F | *vanY-B* |
| STP-inf-F | *mcr-10.1* |
| STP-inf-F | *mcr-3.10* |
| STP-inf-F | *aph(3")-Ia* |
| STP-inf-F | *blaOXA-444* |

| | |
|---|---|
| STP-inf-F | *aphEII* |
| STP-inf-F | *blaOXA-334* |
| STP-inf-F | *srm(B)* |
| STP-inf-F | *blaOXA-160* |
| STP-inf-F | *blaOXA-3* |
| STP-inf-F | *blaOXA-737* |
| STP-inf-F | *blaOXA-228* |
| STP-inf-F | *blaOXA-320* |
| STP-inf-F | *blaOXA-299* |
| STP-inf-F | *blaLEN-1* |
| STP-inf-F | *aac3-IXa* |
| STP-inf-F | *blaTER-1* |
| STP-inf-F | *aac3-IIa* |
| STP-inf-F | *blaGIL-1* |
| STP-inf-F | *blaOKP-A-12* |
| STP-inf-F | *blaOKP-A-16* |
| STP-inf-F | *blaOKP-D-1* |
| STP-inf-F | *blaSHV-102* |
| STP-inf-F | *blaSHV-107* |
| STP-inf-F | *blaSHV-35* |
| STP-inf-F | *blaSHV-63* |
| STP-inf-F | *blaTEM-102* |
| STP-inf-F | *blaTEM-156* |
| STP-inf-F | *blaGES-1* |
| STP-inf-F | *blaGES-13* |
| STP-inf-F | *blaCRH-1* |
| STP-inf-F | *blaCTX-M-74* |
| STP-inf-F | *blaGOB-27* |
| STP-inf-F | *blaCTX-M-12* |
| STP-inf-F | *blaORN1a* |
| STP-inf-F | *blaOXY1-1* |
| STP-inf-F | *blaOXY1-2* |
| STP-inf-F | *blaPLA1a* |
| STP-inf-F | *blaSFO-1* |
| STP-inf-F | *blaBES-1* |
| STP-inf-F | *blaERP-1* |
| STP-inf-F | *vanYF-Pp* |
| STP-inf-F | *patB* |
| STP-inf-F | *aadA9* |
| STP-inf-F | *aph(2'')-Id* |
| STP-inf-F | *blaRm3* |
| STP-inf-F | *blaL2* |
| STP-inf-F | *CARB-3* |
| STP-inf-F | *blaM-1* |

| | |
|---|---|
| STP-inf-F | *blaPER-1* |
| STP-inf-F | *penA-A_Burk* |
| STP-inf-F | *blaBKC-1* |
| STP-inf-F | *erm*(S) |
| STP-inf-F | *otr(A)* |
| STP-inf-F | *cfxA4* |
| STP-inf-F | *cfxA5* |
| STP-inf-F | *erm*(31) |
| STP-inf-F | *vanH-A* |
| STP-inf-F | *vanH-B* |
| STP-inf-F | *blaDES-1* |
| STP-inf-F | *blaSGM-6* |
| STP-inf-F | *vanJ* |
| STP-inf-F | *MSI-1* |
| STP-inf-F | *vanB* |
| STP-inf-F | *vanD* |
| STP-inf-F | *vanSO* |
| STP-inf-F | *AxyY* |
| STP-inf-F | *oqxB8* |
| STP-inf-F | *Streptomyces_rishiriensis_parY_mut* |
| STP-inf-F | *vanE* |
| STP-inf-F | *TriB* |
| STP-inf-F | *efrB* |
| STP-inf-F | *mexH* |
| STP-inf-F | *pp-flo* |
| STP-inf-F | *mexV* |
| STP-inf-F | *blaEC-13* |
| STP-inf-F | *blaEC-18* |
| STP-inf-F | *blaDHA-16* |
| STP-inf-F | *blaACT-15* |
| STP-inf-F | *blaCMY-103* |
| STP-inf-F | *blaCMY-FOX* |
| STP-inf-F | *blaFOX-1* |
| STP-inf-F | *blaFOX-13* |
| STP-inf-F | *blaMOX-3* |
| STP-inf-F | *blaADC-120* |
| STP-inf-F | *blaADC-208* |
| STP-inf-F | *blaMOX-13* |
| STP-inf-F | *blaMOX-8* |
| STP-inf-F | *MexA* |
| STP-inf-F | *oqxA3* |
| STP-inf-F | *oqxA7* |
| STP-inf-F | *beta-lactamase_class-C* |
| STP-inf-F | *blaPDC-122* |

| | |
|---|---|
| STP-inf-F | *blaPDC-125* |
| STP-inf-F | *blaPDC-148* |
| STP-inf-F | *blaPDC-275* |
| STP-inf-F | *vanK-Sc* |
| STP-inf-F | *smeA* |
| STP-inf-F | *tet*(B) |
| STP-inf-F | *MexE* |
| STP-inf-F | *mgt* |
| STP-inf-F | *cmlA* |
| STP-inf-F | *cmlA1* |
| STP-inf-F | *efmA* |
| STP-inf-F | *tet*(42) |
| STP-inf-F | *abeM* |
| STP-inf-F | *mepA* |
| STP-inf-F | *smeF* |
| STP-inf-F | *smeS* |
| STP-inf-F | *cmeC* |
| STP-inf-F | *lsa(A)* |
| STP-inf-F | *lrfA* |
| STP-inf-F | *tva(A)* |
| STP-inf-F | *TriC* |
| STP-inf-F | *qepA2* |
| STP-inf-F | *tcr-3* |
| STP-inf-F | *mexN* |
| STP-inf-F | *vga(A)* |
| STP-inf-F | *vga(C)* |
| STP-inf-F | *mexY* |
| STP-inf-F | *vga(E)* |
| STP-inf-F | *oqxB16* |
| STP-inf-F | *oqxB18* |
| STP-inf-F | *oqxB22* |
| STP-inf-F | *oqxB24* |
| STP-inf-F | *efpA* |
| STP-inf-F | *tcmA* |
| STP-inf-F | *mcr-9.1* |
| STP-inf-F | *mcr-3.3* |
| STP-inf-F | *mcr-3.12* |
| STP-inf-F | *mcr-3.23* |
| STP-inf-F | *tlr(C)* |
| STP-inf-F | *car(A)* |
| STP-inf-F | *patA* |
| STP-inf-F | *mcr-8.1* |
| STP-inf-F | *efrA* |
| STP-inf-F | *macB* |

| | |
|---|---|
| STP-inf-F | *arnA* |
| STP-inf-F | *mecC* |
| STP-inf-F | *mecA2* |
| STP-inf-F | *mecB* |
| STP-inf-F | *PBP1b* |
| STP-inf-F | *MexD* |
| STP-inf-F | *oqxB20* |
| STP-inf-F | *oqxB21* |
| STP-inf-F | *oqxB26* |
| STP-inf-F | *mtrD* |
| STP-inf-A | *msr*(E) |
| STP-inf-A | *mph*(E) |
| STP-inf-A | *tet*(39) |
| STP-inf-A | *erm*(B) |
| STP-inf-A | *tet*(Q) |
| STP-inf-A | *tet*(W) |
| STP-inf-A | *tet*(O) |
| STP-inf-A | *blaMCA* |
| STP-inf-A | *mef*(A) |
| STP-inf-A | *aadA27* |
| STP-inf-A | *msr*(D) |
| STP-inf-A | *aph(3'')-Ib* |
| STP-inf-A | *blaOXA-296* |
| STP-inf-A | *aph(6)-Id* |
| STP-inf-A | *qacH* |
| STP-inf-A | *tet*(M) |
| STP-inf-A | *erm*(F) |
| STP-inf-A | *cfxA6* |
| STP-inf-A | *blaOXA-333* |
| STP-inf-A | *aadE* |
| STP-inf-A | *blaOXA-164* |
| STP-inf-A | *mef*(C) |
| STP-inf-A | *cfxA* |
| STP-inf-A | *blaOXA-211* |
| STP-inf-A | *sul1* |
| STP-inf-A | *dfrF* |
| STP-inf-A | *tet*(40) |
| STP-inf-A | *aac(3)-I* |
| STP-inf-A | *blaOXA-129* |
| STP-inf-A | *lsa(E)* |
| STP-inf-A | *Mef(En2)* |
| STP-inf-A | *cmxA* |
| STP-inf-A | *tet*(32) |
| STP-inf-A | *cfr-Cb* |

| | |
|---|---|
| STP-inf-A | *blaAER-1* |
| STP-inf-A | *tet*(X) |
| STP-inf-A | *aadS* |
| STP-inf-A | *vanA-G* |
| STP-inf-A | *mph*(G) |
| STP-inf-A | *tet*(A) |
| STP-inf-A | *blaOXA-141* |
| STP-inf-A | *blaOXA-280* |
| STP-inf-A | *aadA5* |
| STP-inf-A | *sul2* |
| STP-inf-A | *lnu(AN2)* |
| STP-inf-A | *tet*(C) |
| STP-inf-A | *lin(B)* |
| STP-inf-A | *AAC(6')-IIa* |
| STP-inf-A | *erm*(G) |
| STP-inf-A | *tet*(R) |
| STP-inf-A | *ere(D)* |
| STP-inf-A | *tetR*(G) |
| STP-inf-A | *tet*(S) |
| STP-inf-A | *Bifidobacteria_intrinsic_ileS_confer* |
| STP-inf-A | *lnu(C)* |
| STP-inf-A | *blaOXA-666* |
| STP-inf-A | *blaOXA-10* |
| STP-inf-A | *catA9* |
| STP-inf-A | *CblA-1* |
| STP-inf-A | *aph(3'')-III* |
| STP-inf-A | *aac3-IIe* |
| STP-inf-A | *tet*(G) |
| STP-inf-A | *blaOXA-392* |
| STP-inf-A | *aac6-Aph2* |
| STP-inf-A | *aadA1* |
| STP-inf-A | *catA13* |
| STP-inf-A | *blaOXA-427* |
| STP-inf-A | *lnu(G)* |
| STP-inf-A | *blaOXA-643* |
| STP-inf-A | *cat-TC* |
| STP-inf-A | *aadA6* |
| STP-inf-A | *blaOXA-118* |
| STP-inf-A | *catQ* |
| STP-inf-A | *blaOXA-650* |
| STP-inf-A | *ICR-Mo* |
| STP-inf-A | *ant(3'')-IIc* |
| STP-inf-A | *blaMOX-9* |
| STP-inf-A | *blaRCP* |

| | |
|---|---|
| STP-inf-A | *blaOXA-212* |
| STP-inf-A | *aadA11* |
| STP-inf-A | *blaPER-1* |
| STP-inf-A | *ant(3'')-IIa* |
| STP-inf-A | *spw* |
| STP-inf-A | *bla-A* |
| STP-inf-A | *blaOXA-1* |
| STP-inf-A | *aad9* |
| STP-inf-A | *aadA13* |
| STP-inf-A | *aph3-VIb* |
| STP-inf-A | *AAC(6')-Ib7* |
| STP-inf-A | *blaOXA-20* |
| STP-inf-A | *lnu((B)* |
| STP-inf-A | *blaOXA-347* |
| STP-inf-A | *aac3-I* |
| STP-inf-A | *sat4* |
| STP-inf-A | *aadA2* |
| STP-inf-A | *iri* |
| STP-inf-A | *erm*(T) |
| STP-inf-A | *qnrS2* |
| STP-inf-A | *blaOXA-662* |
| STP-inf-A | *blaOXA-139* |
| STP-inf-A | *tet*(44) |
| STP-inf-A | *strB* |
| STP-inf-A | *aac3-IId* |
| STP-inf-A | *CARB-14/blaRTG* |
| STP-inf-A | *blaOXA-645* |
| STP-inf-A | *kdpE* |
| STP-inf-A | *Nocardia_rifampin_resistant_beta-s* |
| STP-inf-A | *mdtP* |
| STP-inf-A | *catS* |
| STP-inf-A | *ant3''Ih-Aac6-IId* |
| STP-inf-A | *blaOXA-119* |
| STP-inf-A | *floR2* |
| STP-inf-A | *vanY-A* |
| STP-inf-A | *evgA* |
| STP-inf-A | *blaOXA-334* |
| STP-inf-A | *blaOXA-373* |
| STP-inf-A | *cfr(C)* |
| STP-inf-A | *tetA(P)* |
| STP-inf-A | *CRP* |
| STP-inf-A | *Escherichia_coli_acrA* |
| STP-inf-A | *Klebsiella_pneumoniae_acrA* |
| STP-inf-A | *AAC(6')-30/AAC(6')-Ib'_fusion_prot* |

| | |
|---|---|
| STP-inf-A | *tet*(E) |
| STP-inf-A | *aadA17* |
| STP-inf-A | *mph*(A) |
| STP-inf-A | *mdtF* |
| STP-inf-A | *mdtE* |
| STP-inf-A | *aac(3)-Ia* |
| STP-inf-A | *blaOXA-281* |
| STP-inf-A | *Klebsiella_pneumoniae_KpnH* |
| STP-inf-A | *dfrB3* |
| STP-inf-A | *aad(6)* |
| STP-inf-A | *acrF* |
| STP-inf-A | *cfxA_gen* |
| STP-inf-A | *acrB* |
| STP-inf-A | *gadW* |
| STP-inf-A | *mef*(B) |
| STP-inf-A | *cpxA* |
| STP-inf-A | *Penicillin_Binding_Protein_Ecoli* |
| STP-inf-A | *CARB-10* |
| STP-inf-A | *emrA* |
| STP-inf-A | *acrS* |
| STP-inf-A | *ant(2")-Ia* |
| STP-inf-A | *mdtC* |
| STP-inf-A | *H-NS* |
| STP-inf-A | *aadA10* |
| STP-inf-A | *fosA_gen* |
| STP-inf-A | *blaTEM-1* |
| STP-inf-A | *aacA-ACI1* |
| STP-inf-A | *mdtB* |
| STP-inf-A | *evgS* |
| STP-inf-A | *mdtH* |
| STP-inf-A | *MexF* |
| STP-inf-A | *floR* |
| STP-inf-A | *mdtG* |
| STP-inf-A | *erm*(Q) |
| STP-inf-A | *aadA24* |
| STP-inf-A | *mphB* |
| STP-inf-A | *blaOXA-205* |
| STP-inf-A | *lnu(A)* |
| STP-inf-A | *vanZ-A* |
| STP-inf-A | *eptA* |
| STP-inf-A | *blaOXA-209* |
| STP-inf-A | *blaOXA-309* |
| STP-inf-A | *blaOXA-652* |
| STP-inf-A | *gadX* |

| | |
|---|---|
| STP-inf-A | *blaOXA-18* |
| STP-inf-A | *ampH* |
| STP-inf-A | *Escherichia_coli_emrE* |
| STP-inf-A | *ugd* |
| STP-inf-A | *erm*(X) |
| STP-inf-A | *ant6-Ib* |
| STP-inf-A | *aac(3)-IIe* |
| STP-inf-A | *ere(A2)* |
| STP-inf-A | *emrK* |
| STP-inf-A | *tet*(T) |
| STP-inf-A | *tetB-P* |
| STP-inf-A | *cblA* |
| STP-inf-A | *aac(6')-Im* |
| STP-inf-A | *tetA-P* |
| STP-inf-A | *baeR* |
| STP-inf-A | *Salmonella_enterica_cmlA* |
| STP-inf-A | *blaOXA-5* |
| STP-inf-A | *blaOXA-836* |
| STP-inf-A | *mcr-3.1* |
| STP-inf-A | *Escherichia_coli_mdfA* |
| STP-inf-A | *mdtM* |
| STP-inf-A | *mdtN* |
| STP-inf-A | *catP* |
| STP-inf-A | *mdtA* |
| STP-inf-A | *catB3* |
| STP-inf-A | *tolC* |
| STP-inf-A | *emrY* |
| STP-inf-A | *aac(6')-30* |
| STP-inf-A | *lmrD* |
| STP-inf-A | *Pseudomonas_aeruginosa_CpxR* |
| STP-inf-A | *ampH_Ecoli* |
| STP-inf-A | *mcr-3.6* |
| STP-inf-A | *baeS* |
| STP-inf-A | *Klebsiella_pneumoniae_KpnG* |
| STP-inf-A | *dfrB1* |
| STP-inf-A | *dhfr7* |
| STP-inf-A | *tet*(36) |
| STP-inf-A | *pmrF* |
| STP-inf-A | *erm*(A) |
| STP-inf-A | *tetA(46)* |
| STP-inf-A | *erm*(47) |
| STP-inf-A | *msbA* |
| STP-inf-A | *cphA5* |
| STP-inf-A | *nimE_Nitroimidazole_Gene* |

| | |
|---|---|
| STP-inf-A | *blaNPS-1* |
| STP-inf-A | *aac3-Ib* |
| STP-inf-A | *yojI* |
| STP-inf-A | *blaOXA-9* |
| STP-inf-A | *vanW-G* |
| STP-inf-A | *str* |
| STP-inf-A | *acrD* |
| STP-inf-A | *blaOXY1-1* |
| STP-inf-A | *blaPAU-1* |
| STP-inf-A | *cmr* |
| STP-inf-A | *oqxA10* |
| STP-inf-A | *oqxA11* |
| STP-inf-A | *cepA* |
| STP-inf-A | *vanT-G* |
| STP-inf-A | *aph7* |
| STP-inf-A | *vanH-B* |
| STP-inf-A | *mcr-3.17* |
| STP-inf-A | *qnrVC4* |
| STP-inf-A | *Klebsiella_pneumoniae_KpnF* |
| STP-inf-A | *aac3-Ib-Aac6-Ib* |
| STP-inf-A | *vanS-B* |
| STP-inf-A | *vanB* |
| STP-inf-A | *tet*(L) |
| STP-inf-A | *Klebsiella_pneumoniae_OmpK37* |
| STP-inf-A | *blaEC* |
| STP-inf-A | *blaOXA-464* |
| STP-inf-A | *marA* |
| STP-inf-A | *vanS-A* |
| STP-inf-A | *acrE* |
| STP-inf-A | *aph(3')-VIa* |
| STP-inf-A | *smeE* |
| STP-inf-A | *aadA25* |
| STP-inf-A | *beta-lactamase_class-C* |
| STP-inf-A | *blaOXA-12* |
| STP-inf-A | *blaOXA-504* |
| STP-inf-A | *blaOXA-725* |
| STP-inf-A | *blaOXA-726* |
| STP-inf-A | *blaOXA-13* |
| STP-inf-A | *blaOXA-46* |
| STP-inf-A | *aph(3')-IIc* |
| STP-inf-A | *bacA* |
| STP-inf-A | *blaOXA-644* |
| STP-inf-A | *arr-269927220* |
| STP-inf-A | *blaOXA-2* |

| | |
|---|---|
| STP-inf-A | *MexE* |
| STP-inf-A | *FosA* |
| STP-inf-A | *blaOXA-512* |
| STP-inf-A | *FosA2* |
| STP-inf-A | *blaGES-1* |
| STP-inf-A | *blaOXY1-4* |
| STP-inf-A | *aph(2'')-IIa* |
| STP-inf-A | *blaVEB-3* |
| STP-inf-A | *arr* |
| STP-inf-A | *arr-8* |
| STP-inf-A | *arr2* |
| STP-inf-A | *dfrA1* |
| STP-inf-A | *cfxA3* |
| STP-inf-A | *dfrA14* |
| STP-inf-A | *vanH-A* |
| STP-inf-A | *vanH-D* |
| STP-inf-A | *nimD_Nitroimidazole_Gene* |
| STP-inf-A | *dfrG* |
| STP-inf-A | *aph4-Ia* |
| STP-inf-A | *mdtO* |
| STP-inf-A | *AxyY* |
| STP-inf-A | *MexB* |
| STP-inf-A | *cfr(B)* |
| STP-inf-A | *vanG* |
| STP-inf-A | *oqxB11* |
| STP-inf-A | *nimA_Nitroimidazole_Gene* |
| STP-inf-A | *aadB* |
| STP-inf-A | *mcr-9.1* |
| STP-inf-A | *mcr-3.3* |
| STP-inf-A | *mcr-5.1* |
| STP-inf-A | *aacA-ENT1* |
| STP-inf-A | *catB4* |
| STP-inf-A | *aac(6')-Ib-AKT* |
| STP-inf-A | *satA_Ba* |
| STP-inf-A | *dfr24* |
| STP-inf-A | *blaEC-18* |
| STP-inf-A | *tet*(Z) |
| STP-inf-A | *tet*(Y) |
| STP-inf-A | *patB* |
| STP-inf-A | *blaRSC1-1* |
| STP-inf-A | *tet*(B) |
| STP-inf-A | *mexW* |
| STP-inf-A | *vanX-D* |
| STP-inf-A | *cmlA* |

| | |
|---|---|
| STP-inf-A | *cmlA1* |
| STP-inf-A | *OqxBgb* |
| STP-inf-A | *cmlB1* |
| STP-inf-A | *MuxA* |
| STP-inf-A | *tet*(42) |
| STP-inf-A | *qnrB19* |
| STP-inf-A | *catA16* |
| STP-inf-A | *catB* |
| STP-inf-A | *mtrA* |
| STP-inf-A | *Mbl* |
| STP-inf-A | *vanR-A* |
| STP-inf-A | *oprA* |
| STP-inf-A | *smeS* |
| STP-inf-A | *blaEBR-2* |
| STP-inf-A | *blaEBR-3* |
| STP-inf-A | *blaIND-11* |
| STP-inf-A | *erm*(48) |
| STP-inf-A | *blaZOG-1* |
| STP-inf-A | *TriC* |
| STP-inf-A | *cphA2* |
| STP-inf-A | *cphA6* |
| STP-inf-A | *imiH* |
| STP-inf-A | *vanXY-G* |
| STP-inf-A | *imiS* |
| STP-inf-A | *aph(3')-VI* |
| STP-inf-A | *blaLCR-1* |
| STP-inf-A | *vga(A)* |
| STP-inf-A | *aadA4* |
| STP-inf-A | *aadA7* |
| STP-inf-A | *blaOXA-101* |
| STP-inf-A | *blaOXA-732* |
| STP-inf-A | *aac(3)-II* |
| STP-inf-A | *mcr-3.12* |
| STP-inf-A | *MSI-OXA* |
| STP-inf-A | *lnu(F)* |
| STP-inf-A | *blaOXA-15* |
| STP-inf-A | *blaOXA-21* |
| STP-inf-A | *vanW-B* |
| STP-inf-A | *blaOXA-224* |
| STP-inf-A | *blaOXA-320* |
| STP-inf-A | *blaOXA-299* |
| STP-inf-A | *blaLEN-1* |
| STP-inf-A | *blaOXA-279* |
| STP-inf-A | *blaOXA-669* |

| | |
|---|---|
| STP-inf-A | *blaOXA-420* |
| STP-inf-A | *blaOXA-96* |
| STP-inf-A | *rosB* |
| STP-inf-A | *patA* |
| STP-inf-A | *blaBEL-1* |
| STP-inf-A | *aac3-Xa* |
| STP-inf-A | *aci1* |
| STP-inf-A | *ole(B)* |
| STP-inf-A | *blaOXA-62* |
| STP-inf-A | *blaSHV-149* |
| STP-inf-A | *RSA-1* |
| STP-inf-A | *aac3-IIa* |
| STP-inf-A | *aac3-IIc* |
| STP-inf-A | *blaOKP-A-1* |
| STP-inf-A | *blaOKP-A-10* |
| STP-inf-A | *blaOKP-B-10* |
| STP-inf-A | *blaSHV-108* |
| STP-inf-A | *blaTEM-93* |
| STP-inf-A | *blaPAM-1* |
| STP-inf-A | *sul4* |
| STP-inf-A | *tetB*(46) |
| STP-inf-A | *blaCPS-1* |
| STP-inf-A | *blaGOB-33* |
| STP-inf-A | *blaCTX-M-115* |
| STP-inf-A | *blaCTX-M-77* |
| STP-inf-A | *blaORN1a* |
| STP-inf-A | *blaR1* |
| STP-inf-A | *blaKPC-2* |
| STP-inf-A | *aph(2")-If* |
| STP-inf-A | *blaCARB-16* |
| STP-inf-A | *CARB-8_OU* |
| STP-inf-A | *blaSHV-100* |
| STP-inf-A | *mph*(F) |
| STP-inf-A | *vgb(A)* |
| STP-inf-A | *hugA* |
| STP-inf-A | *erm*(42) |
| STP-inf-A | *blaAIM-1* |
| STP-inf-A | *CARB-3* |
| STP-inf-A | *aph(2")-Ic* |
| STP-inf-A | *blaSFC-1* |
| STP-inf-A | *blaAST-1* |
| STP-inf-A | *erm*(W) |
| STP-inf-A | *blaBKC-1* |
| STP-inf-A | *blaBRO-1* |

| | |
|---|---|
| STP-inf-A | *mphO* |
| STP-inf-A | *blaDES-1* |
| STP-inf-A | *adeL* |
| STP-inf-A | *mexK* |
| STP-inf-A | *ceoB* |
| STP-inf-A | *vanA-A* |
| STP-inf-A | *MuxB* |
| STP-inf-A | *smeB* |
| STP-inf-A | *oqxB12* |
| STP-inf-A | *oqxB15* |
| STP-inf-A | *adeJ* |
| STP-inf-A | *adeF* |
| STP-inf-A | *efrB* |
| STP-inf-A | *Burkholderia_pseudomallei_Omp38* |
| STP-inf-A | *AmpC2_Ecoli* |
| STP-inf-A | *blaEC-13* |
| STP-inf-A | *blaACT-1* |
| STP-inf-A | *blaACT-50* |
| STP-inf-A | *blaACT-8* |
| STP-inf-A | *blaCMY-115* |
| STP-inf-A | *blaCMY-40* |
| STP-inf-A | *vanS-D* |
| STP-inf-A | *blaCMY-FOX* |
| STP-inf-A | *blaFOX-13* |
| STP-inf-A | *blaFOX-4* |
| STP-inf-A | *blaFOX-9* |
| STP-inf-A | *blaTRU-1* |
| STP-inf-A | *blaADC-120* |
| STP-inf-A | *blaADC-18* |
| STP-inf-A | *TriA* |
| STP-inf-A | *mexM* |
| STP-inf-A | *mexP* |
| STP-inf-A | *tet*(55) |
| STP-inf-A | *blaACC-1* |
| STP-inf-A | *erm*(38) |
| STP-inf-A | *MexC* |
| STP-inf-A | *blaADC-10* |
| STP-inf-A | *AxyX* |
| STP-inf-A | *emeA* |
| STP-inf-A | *adeA* |
| STP-inf-A | *blaPDC-131* |
| STP-inf-A | *Enterobacter_cloacae_acrA* |
| STP-inf-A | *tet*(H) |
| STP-inf-A | *ceoA* |

| | |
|---|---|
| STP-inf-A | *oleD* |
| STP-inf-A | *adeI* |
| STP-inf-A | *ere(B)* |
| STP-inf-A | *tet*(V) |
| STP-inf-A | *AmpC1_Ecoli* |
| STP-inf-A | *OprZ* |
| STP-inf-A | *smeF* |
| STP-inf-A | *smeC* |
| STP-inf-A | *OpmH* |
| STP-inf-A | *adeH* |
| STP-inf-A | *adeK* |
| STP-inf-A | *opmE* |
| STP-inf-A | *msr*(C) |
| STP-inf-A | *lsa(A)* |
| STP-inf-A | *lrfA* |
| STP-inf-A | *tva(A)* |
| STP-inf-A | *qepA2* |
| STP-inf-A | *emrB* |
| STP-inf-A | *TetAB* |
| STP-inf-A | *adeB* |
| STP-inf-A | *vga(C)* |
| STP-inf-A | *oqxB10* |
| STP-inf-A | *oqxB14* |
| STP-inf-A | *oqxB22* |
| STP-inf-A | *oqxB9* |
| STP-inf-A | *mcr-10.1* |
| STP-inf-A | *mcr-3.8* |
| STP-inf-A | *TlrC* |
| STP-inf-A | *mcr-3.7* |
| STP-inf-A | *mcr-3.9* |
| STP-inf-A | *sal(A)* |
| STP-inf-A | *vmlR* |
| STP-inf-A | *tlr(C)* |
| STP-inf-A | *srm(B)* |
| STP-inf-A | *otr(B)* |
| STP-inf-A | *Clostridium_perfringens_mprF* |
| STP-inf-A | *efrA* |
| STP-inf-A | *otr(A)* |
| STP-inf-A | *mecB* |
| STP-inf-A | *vanT* |
| STP-inf-A | *vanTc3* |
| STP-inf-A | *Streptomyces_rishiriensis_parY_mut* |
| STP-inf-A | *mupA* |
| STP-inf-A | *mexI* |

| | |
|---|---|
| STP-inf-A | *mupB* |
| STP-inf-A | *mexN* |
| STP-inf-A | *MexD* |
| STP-inf-A | *oqxB16* |
| STP-inf-A | *mexQ* |
| STP-T-eff-F | *tet*(Q) |
| STP-T-eff-F | *tet*(W) |
| STP-T-eff-F | *msr*(E) |
| STP-T-eff-F | *mph*(E) |
| STP-T-eff-F | *bla-A* |
| STP-T-eff-F | *tet*(O) |
| STP-T-eff-F | *erm*(B) |
| STP-T-eff-F | *blaAER-1* |
| STP-T-eff-F | *tet*(39) |
| STP-T-eff-F | *qacH* |
| STP-T-eff-F | *blaOXA-129* |
| STP-T-eff-F | *erm*(F) |
| STP-T-eff-F | *cfxA6* |
| STP-T-eff-F | *mef*(A) |
| STP-T-eff-F | *blaOXA-1* |
| STP-T-eff-F | *aadE* |
| STP-T-eff-F | *cfxA* |
| STP-T-eff-F | *blaOXA-10* |
| STP-T-eff-F | *blaOXA-141* |
| STP-T-eff-F | *sul1* |
| STP-T-eff-F | *mef*(C) |
| STP-T-eff-F | *tet*(X) |
| STP-T-eff-F | *msr*(D) |
| STP-T-eff-F | *tet*(40) |
| STP-T-eff-F | *blaMCA* |
| STP-T-eff-F | *vanA-G* |
| STP-T-eff-F | *aph(6)-Id* |
| STP-T-eff-F | *tet*(M) |
| STP-T-eff-F | *Mef(En2)* |
| STP-T-eff-F | *tet*(32) |
| STP-T-eff-F | *lnu(AN2)* |
| STP-T-eff-F | *blaOXA-296* |
| STP-T-eff-F | *aph(3")-Ib* |
| STP-T-eff-F | *mph*(G) |
| STP-T-eff-F | *aadS* |
| STP-T-eff-F | *blaOXA-20* |
| STP-T-eff-F | *dfrF* |
| STP-T-eff-F | *blaOXA-392* |
| STP-T-eff-F | *aac(3)-I* |

| | |
|---|---|
| STP-T-eff-F | *CblA-1* |
| STP-T-eff-F | *catA13* |
| STP-T-eff-F | *blaOXA-224* |
| STP-T-eff-F | *aadA1* |
| STP-T-eff-F | *AAC(6')-IIa* |
| STP-T-eff-F | *tet*(C) |
| STP-T-eff-F | *blaPAU-1* |
| STP-T-eff-F | *Bifidobacteria_intrinsic_ileS_confer* |
| STP-T-eff-F | *aadA5* |
| STP-T-eff-F | *erm*(G) |
| STP-T-eff-F | *ere(D)* |
| STP-T-eff-F | *lnu(C)* |
| STP-T-eff-F | *sul2* |
| STP-T-eff-F | *blaBEL-1* |
| STP-T-eff-F | *tet*(A) |
| STP-T-eff-F | *aadA27* |
| STP-T-eff-F | *blaOXA-118* |
| STP-T-eff-F | *aadA11* |
| STP-T-eff-F | *aph(3'')-III* |
| STP-T-eff-F | *blaOXA-33* |
| STP-T-eff-F | *AAC(6')-Ib7* |
| STP-T-eff-F | *cmxA* |
| STP-T-eff-F | *ant(3'')-IIa* |
| STP-T-eff-F | *aad9* |
| STP-T-eff-F | *tet*(G) |
| STP-T-eff-F | *tet*(R) |
| STP-T-eff-F | *lsa(E)* |
| STP-T-eff-F | *aadA6* |
| STP-T-eff-F | *lnu(G)* |
| STP-T-eff-F | *blaOXA-205* |
| STP-T-eff-F | *lin(B)* |
| STP-T-eff-F | *tetR*(G) |
| STP-T-eff-F | *blaOXA-211* |
| STP-T-eff-F | *blaOXA-164* |
| STP-T-eff-F | *blaOXA-5* |
| STP-T-eff-F | *blaOXA-2* |
| STP-T-eff-F | *cfr(C)* |
| STP-T-eff-F | *blaOXA-101* |
| STP-T-eff-F | *cfxA_gen* |
| STP-T-eff-F | *gadW* |
| STP-T-eff-F | *blaOXA-119* |
| STP-T-eff-F | *aadA24* |
| STP-T-eff-F | *tet*(44) |
| STP-T-eff-F | *aac6-Aph2* |

| | |
|---|---|
| STP-T-eff-F | *blaOXA-31* |
| STP-T-eff-F | *cblA* |
| STP-T-eff-F | *blaOXA-464* |
| STP-T-eff-F | *floR2* |
| STP-T-eff-F | *aac(6')-30* |
| STP-T-eff-F | *CRP* |
| STP-T-eff-F | *mdtO* |
| STP-T-eff-F | *blaRCP* |
| STP-T-eff-F | *catS* |
| STP-T-eff-F | *blaOXA-333* |
| STP-T-eff-F | *mdtP* |
| STP-T-eff-F | *fosA_gen* |
| STP-T-eff-F | *ampH* |
| STP-T-eff-F | *Klebsiella_pneumoniae_KpnG* |
| STP-T-eff-F | *tet*(S) |
| STP-T-eff-F | *aadA2* |
| STP-T-eff-F | *aadA13* |
| STP-T-eff-F | *mdtF* |
| STP-T-eff-F | *spw* |
| STP-T-eff-F | *cat-TC* |
| STP-T-eff-F | *aac3-IIe* |
| STP-T-eff-F | *mdtA* |
| STP-T-eff-F | *acrB* |
| STP-T-eff-F | *Klebsiella_pneumoniae_KpnH* |
| STP-T-eff-F | *cfr-Cb* |
| STP-T-eff-F | *catA9* |
| STP-T-eff-F | *aac3-Ib* |
| STP-T-eff-F | *emrR* |
| STP-T-eff-F | *aac3-I* |
| STP-T-eff-F | *tolC* |
| STP-T-eff-F | *sat4* |
| STP-T-eff-F | *cpxA* |
| STP-T-eff-F | *cfxA4* |
| STP-T-eff-F | *blaOXA-320* |
| STP-T-eff-F | *aac(3)-Ia* |
| STP-T-eff-F | *catP* |
| STP-T-eff-F | *Penicillin_Binding_Protein_Ecoli* |
| STP-T-eff-F | *mdtC* |
| STP-T-eff-F | *blaOXA-209* |
| STP-T-eff-F | *blaOXA-347* |
| STP-T-eff-F | *blaOXA-280* |
| STP-T-eff-F | *acrS* |
| STP-T-eff-F | *oqxA10* |
| STP-T-eff-F | *Escherichia_coli_acrA* |

| | |
|---|---|
| STP-T-eff-F | *nimE_Nitroimidazole_Gene* |
| STP-T-eff-F | *mdtN* |
| STP-T-eff-F | *Escherichia_coli_mdfA* |
| STP-T-eff-F | *mdtM* |
| STP-T-eff-F | *baeR* |
| STP-T-eff-F | *acrF* |
| STP-T-eff-F | *eptA* |
| STP-T-eff-F | *blaMOX-9* |
| STP-T-eff-F | *pmrF* |
| STP-T-eff-F | *Nocardia_rifampin_resistant_beta-s* |
| STP-T-eff-F | *mdtB* |
| STP-T-eff-F | *aadA12* |
| STP-T-eff-F | *aadA17* |
| STP-T-eff-F | *blaOXA-427* |
| STP-T-eff-F | *blaOXA-504* |
| STP-T-eff-F | *ant3''Ih-Aac6-IId* |
| STP-T-eff-F | *fosE* |
| STP-T-eff-F | *tet*(E) |
| STP-T-eff-F | *bacA* |
| STP-T-eff-F | *yojI* |
| STP-T-eff-F | *gadX* |
| STP-T-eff-F | *blaOXA-15* |
| STP-T-eff-F | *blaOXA-18* |
| STP-T-eff-F | *H-NS* |
| STP-T-eff-F | *aad(6)* |
| STP-T-eff-F | *evgS* |
| STP-T-eff-F | *aci1* |
| STP-T-eff-F | *aac3-IId* |
| STP-T-eff-F | *AmpC1_Ecoli* |
| STP-T-eff-F | *acrD* |
| STP-T-eff-F | *CARB-10* |
| STP-T-eff-F | *cepA* |
| STP-T-eff-F | *blaEC* |
| STP-T-eff-F | *ampH_Ecoli* |
| STP-T-eff-F | *dfrA15* |
| STP-T-eff-F | *dhfr7* |
| STP-T-eff-F | *ICR-Mo* |
| STP-T-eff-F | *tet*(36) |
| STP-T-eff-F | *cfxA3* |
| STP-T-eff-F | *mdtH* |
| STP-T-eff-F | *mdtG* |
| STP-T-eff-F | *AAC(6')-30/AAC(6')-Ib'_fusion_prote* |
| STP-T-eff-F | *msbA* |
| STP-T-eff-F | *emrY* |

| | |
|---|---|
| STP-T-eff-F | *tet*(42) |
| STP-T-eff-F | *ant(3")-IIc* |
| STP-T-eff-F | *blaLCR-1* |
| STP-T-eff-F | *blaOXA-37* |
| STP-T-eff-F | *mcr-3.17* |
| STP-T-eff-F | *blaOXA-281* |
| STP-T-eff-F | *aac(6')-Ib-G* |
| STP-T-eff-F | *aacA38* |
| STP-T-eff-F | *strB* |
| STP-T-eff-F | *Klebsiella_pneumoniae_OmpK37* |
| STP-T-eff-F | *acrE* |
| STP-T-eff-F | *ugd* |
| STP-T-eff-F | *emrA* |
| STP-T-eff-F | *blaGPC-1* |
| STP-T-eff-F | *Klebsiella_pneumoniae_acrA* |
| STP-T-eff-F | *aph(2")-IIa* |
| STP-T-eff-F | *blaVEB-1* |
| STP-T-eff-F | *blaAIM-1* |
| STP-T-eff-F | *evgA* |
| STP-T-eff-F | *ere(B)* |
| STP-T-eff-F | *vanH-B* |
| STP-T-eff-F | *qnrS2* |
| STP-T-eff-F | *catQ* |
| STP-T-eff-F | *aac3-Ib-Aac6-Ib* |
| STP-T-eff-F | *kdpE* |
| STP-T-eff-F | *Pseudomonas_aeruginosa_CpxR* |
| STP-T-eff-F | *RbpA* |
| STP-T-eff-F | *tetA(46)* |
| STP-T-eff-F | *emrK* |
| STP-T-eff-F | *erm*(A) |
| STP-T-eff-F | *erm*(T) |
| STP-T-eff-F | *MecI* |
| STP-T-eff-F | *blaEC-18* |
| STP-T-eff-F | *mexW* |
| STP-T-eff-F | *erm*(Q) |
| STP-T-eff-F | *blaNPS-1* |
| STP-T-eff-F | *MexF* |
| STP-T-eff-F | *lnu((B)* |
| STP-T-eff-F | *mef*(B) |
| STP-T-eff-F | *ere(A)* |
| STP-T-eff-F | *blaOXA-373* |
| STP-T-eff-F | *blaOXA-650* |
| STP-T-eff-F | *blaOXA-139* |
| STP-T-eff-F | *aadA10* |

| | |
|---|---|
| STP-T-eff-F | *FosA5* |
| STP-T-eff-F | *vanW-G* |
| STP-T-eff-F | *blaOXA-666* |
| STP-T-eff-F | *erm*(X) |
| STP-T-eff-F | *ant6-Ib* |
| STP-T-eff-F | *aac(3)-IIe* |
| STP-T-eff-F | *blaPSE-4* |
| STP-T-eff-F | *blaL1* |
| STP-T-eff-F | *blaCPS-1* |
| STP-T-eff-F | *aac6-Ig* |
| STP-T-eff-F | *aacA-ACI1* |
| STP-T-eff-F | *mexK* |
| STP-T-eff-F | *aacA-ACI5* |
| STP-T-eff-F | *aph(2")-If* |
| STP-T-eff-F | *CARB-14/blaRTG* |
| STP-T-eff-F | *aph(2")-Ib* |
| STP-T-eff-F | *OqxBgb* |
| STP-T-eff-F | *cepA_beta-lactamase* |
| STP-T-eff-F | *mph*(A) |
| STP-T-eff-F | *erm*(49) |
| STP-T-eff-F | *aph7* |
| STP-T-eff-F | *baeS* |
| STP-T-eff-F | *mphB* |
| STP-T-eff-F | *iri* |
| STP-T-eff-F | *vanZ-Pa* |
| STP-T-eff-F | *aac(6')* |
| STP-T-eff-F | *aacA-STR-10* |
| STP-T-eff-F | *cfxA5* |
| STP-T-eff-F | *dfrA14* |
| STP-T-eff-F | *lin(A)* |
| STP-T-eff-F | *vanH-D* |
| STP-T-eff-F | *nimj_Nitroimidazole_Gene* |
| STP-T-eff-F | *aac(6')-Iag* |
| STP-T-eff-F | *aph4-Ia* |
| STP-T-eff-F | *sat-2A* |
| STP-T-eff-F | *vanG2* |
| STP-T-eff-F | *nimA_Nitroimidazole_Gene* |
| STP-T-eff-F | *vanT-G* |
| STP-T-eff-F | *mcr-3.12* |
| STP-T-eff-F | *blaDHA-12* |
| STP-T-eff-F | *vanS-D* |
| STP-T-eff-F | *blaFOX-2* |
| STP-T-eff-F | *mdtE* |
| STP-T-eff-F | *Salmonella_enterica_cmlA* |

| | |
|---|---|
| STP-T-eff-F | *bcr1* |
| STP-T-eff-F | *floR* |
| STP-T-eff-F | *vanX-D* |
| STP-T-eff-F | *cmlA1* |
| STP-T-eff-F | *cmlA4* |
| STP-T-eff-F | *tetA(P)* |
| STP-T-eff-F | *catB3* |
| STP-T-eff-F | *vat(E)* |
| STP-T-eff-F | *cat-pC223* |
| STP-T-eff-F | *kamB* |
| STP-T-eff-F | *Acinetobacter_baumannii_AbaQ* |
| STP-T-eff-F | *qnrA6* |
| STP-T-eff-F | *catB* |
| STP-T-eff-F | *npmA* |
| STP-T-eff-F | *tet*(L) |
| STP-T-eff-F | *blaOXA-664* |
| STP-T-eff-F | *vanR-O* |
| STP-T-eff-F | *blaIMP-22* |
| STP-T-eff-F | *blaOXA-184* |
| STP-T-eff-F | *cfiA13* |
| STP-T-eff-F | *cfiA18* |
| STP-T-eff-F | *blaOXA-85* |
| STP-T-eff-F | *cphA1* |
| STP-T-eff-F | *cphA5* |
| STP-T-eff-F | *vanXY-G* |
| STP-T-eff-F | *aph(3')-VI* |
| STP-T-eff-F | *aph3-VIb* |
| STP-T-eff-F | *erm*(O) |
| STP-T-eff-F | *AxyY* |
| STP-T-eff-F | *vga(A)* |
| STP-T-eff-F | *mexY* |
| STP-T-eff-F | *oqxB12* |
| STP-T-eff-F | *aadA25* |
| STP-T-eff-F | *blaOXA-12* |
| STP-T-eff-F | *APH(6)-Ic* |
| STP-T-eff-F | *blaOXA-732* |
| STP-T-eff-F | *blaOXA-836* |
| STP-T-eff-F | *aac3-Vb* |
| STP-T-eff-F | *mcr-3.3* |
| STP-T-eff-F | *mcr-3.6* |
| STP-T-eff-F | *TlrC* |
| STP-T-eff-F | *mcr-4.1* |
| STP-T-eff-F | *aph(3'')-Ia* |
| STP-T-eff-F | *blaOXA-60* |

| | |
|---|---|
| STP-T-eff-F | *blaOXA-527* |
| STP-T-eff-F | *blaOXA-643* |
| STP-T-eff-F | *blaOXA-645* |
| STP-T-eff-F | *blaOXA-652* |
| STP-T-eff-F | *blaOXA-662* |
| STP-T-eff-F | *blaOXA-3* |
| STP-T-eff-F | *blaOXA-675* |
| STP-T-eff-F | *blaOXA-299* |
| STP-T-eff-F | *aadA9* |
| STP-T-eff-F | *facT* |
| STP-T-eff-F | *aadA16* |
| STP-T-eff-F | *blaOXA-275* |
| STP-T-eff-F | *str* |
| STP-T-eff-F | *aac3-IIa* |
| STP-T-eff-F | *blaLEN-12* |
| STP-T-eff-F | *blaTEM-107* |
| STP-T-eff-F | *blaTEM-156* |
| STP-T-eff-F | *SHV-1* |
| STP-T-eff-F | *blaTEM-1* |
| STP-T-eff-F | *sul4* |
| STP-T-eff-F | *blaOXA-154* |
| STP-T-eff-F | *blaCRP-1* |
| STP-T-eff-F | *blaLRA-1* |
| STP-T-eff-F | *blaLUT-1* |
| STP-T-eff-F | *vgb(C)* |
| STP-T-eff-F | *mphH* |
| STP-T-eff-F | *blaRm3* |
| STP-T-eff-F | *mph*(B) |
| STP-T-eff-F | *blaECM-1* |
| STP-T-eff-F | *vanY-A* |
| STP-T-eff-F | *blaCSP-1* |
| STP-T-eff-F | *aph(2")-Ic* |
| STP-T-eff-F | *aph(2")-Ig* |
| STP-T-eff-F | *blaSGM-2* |
| STP-T-eff-F | *blaCDD-1* |
| STP-T-eff-F | *tet*(T) |
| STP-T-eff-F | *erm*(R) |
| STP-T-eff-F | *vanB* |
| STP-T-eff-F | *smeE* |
| STP-T-eff-F | *oqxB15* |
| STP-T-eff-F | *oqxB17* |
| STP-T-eff-F | *oqxB9* |
| STP-T-eff-F | *otrC* |
| STP-T-eff-F | *Streptomyces_rishiriensis_parY_mut* |

| | |
|---|---|
| STP-T-eff-F | *adeS* |
| STP-T-eff-F | *AmpC2_Ecoli* |
| STP-T-eff-F | *blaEC-14* |
| STP-T-eff-F | *blaEC-5* |
| STP-T-eff-F | *blaEC-8* |
| STP-T-eff-F | *blaSRT-2* |
| STP-T-eff-F | *blaCMY-13* |
| STP-T-eff-F | *blaMIR-11* |
| STP-T-eff-F | *blaMOX-4* |
| STP-T-eff-F | *cepS* |
| STP-T-eff-F | *blaADC-130* |
| STP-T-eff-F | *blaADC-135* |
| STP-T-eff-F | *TriA* |
| STP-T-eff-F | *norA* |
| STP-T-eff-F | *cmr* |
| STP-T-eff-F | *cmrA_variant1* |
| STP-T-eff-F | *oqxA11* |
| STP-T-eff-F | *tet*(Y) |
| STP-T-eff-F | *emeA* |
| STP-T-eff-F | *tet*(41) |
| STP-T-eff-F | *smeD* |
| STP-T-eff-F | *blaRSC1-1* |
| STP-T-eff-F | *Enterobacter_cloacae_acrA* |
| STP-T-eff-F | *Tet*(57) |
| STP-T-eff-F | *tet*(B) |
| STP-T-eff-F | *ere(A2)* |
| STP-T-eff-F | *tet*(V) |
| STP-T-eff-F | *tetA-P* |
| STP-T-eff-F | *MuxA* |
| STP-T-eff-F | *efmA* |
| STP-T-eff-F | *cdeA* |
| STP-T-eff-F | *hp1181* |
| STP-T-eff-F | *tet*(K) |
| STP-T-eff-F | *smeC* |
| STP-T-eff-F | *PmpM* |
| STP-T-eff-F | *OprM* |
| STP-T-eff-F | *lsa(C)* |
| STP-T-eff-F | *msr*(C) |
| STP-T-eff-F | *qepA2* |
| STP-T-eff-F | *emrB* |
| STP-T-eff-F | *tcr-3* |
| STP-T-eff-F | *TetAB* |
| STP-T-eff-F | *MexB* |
| STP-T-eff-F | *smeB* |

| | |
|---|---|
| STP-T-eff-F | *oqxB11* |
| STP-T-eff-F | *oqxB21* |
| STP-T-eff-F | *efpA* |
| STP-T-eff-F | *mcr-2.2* |
| STP-T-eff-F | *mcr-9.1* |
| STP-T-eff-F | *mcr-3.10* |
| STP-T-eff-F | *sal(A)* |
| STP-T-eff-F | *mcr-5.1* |
| STP-T-eff-F | *patB* |
| STP-T-eff-F | *taeA* |
| STP-T-eff-F | *tetB-P* |
| STP-T-eff-F | *mecC3* |
| STP-T-eff-F | *mecA* |
| STP-T-eff-F | *vanT-C* |
| STP-T-eff-F | *vanT-Cd* |
| STP-T-eff-F | *Listeria_monocytogenes_mprF* |
| STP-T-eff-F | *rph* |
| STP-T-eff-F | *Brucella_suis_mprF* |
| STP-T-eff-F | *rphB* |
| STP-T-eff-F | *mupA* |
| STP-T-eff-F | *mexI* |
| STP-T-eff-F | *MuxB* |
| STP-T-eff-F | *oqxB10* |
| STP-T-eff-F | *oqxB14* |
| STP-T-eff-F | *oqxB20* |
| STP-T-eff-F | *oqxB26* |
| STP-T-eff-F | *oqxB3* |
| STP-T-eff-F | *adeJ* |
| STP-T-eff-F | *mtrD* |
| STP-T-eff-A | *msr*(E) |
| STP-T-eff-A | *mph*(E) |
| STP-T-eff-A | *tet*(39) |
| STP-T-eff-A | *erm*(B) |
| STP-T-eff-A | *bla-A* |
| STP-T-eff-A | *tet*(Q) |
| STP-T-eff-A | *blaOXA-129* |
| STP-T-eff-A | *blaMCA* |
| STP-T-eff-A | *te*t(W) |
| STP-T-eff-A | *tet*(O) |
| STP-T-eff-A | *blaOXA-296* |
| STP-T-eff-A | *aph(6)-Id* |
| STP-T-eff-A | *aph(3'')-Ib* |
| STP-T-eff-A | *mef*(A) |
| STP-T-eff-A | *blaAER-1* |

| | |
|---|---|
| STP-T-eff-A | *aadA27* |
| STP-T-eff-A | *qacH* |
| STP-T-eff-A | *erm*(F) |
| STP-T-eff-A | *msr*(D) |
| STP-T-eff-A | *blaOXA-10* |
| STP-T-eff-A | *Bifidobacterium_adolescentis_rpoB_* |
| STP-T-eff-A | *tet*(M) |
| STP-T-eff-A | *sul1* |
| STP-T-eff-A | *cfxA6* |
| STP-T-eff-A | *aadE* |
| STP-T-eff-A | *mef*(C) |
| STP-T-eff-A | *blaOXA-164* |
| STP-T-eff-A | *blaOXA-141* |
| STP-T-eff-A | *blaOXA-333* |
| STP-T-eff-A | *tet*(40) |
| STP-T-eff-A | *cfxA* |
| STP-T-eff-A | *aac(3)-I* |
| STP-T-eff-A | *blaOXA-1* |
| STP-T-eff-A | *lnu(AN2)* |
| STP-T-eff-A | *mph*(G) |
| STP-T-eff-A | *blaOXA-211* |
| STP-T-eff-A | *tet*(X) |
| STP-T-eff-A | *dfrF* |
| STP-T-eff-A | *Mef(En2)* |
| STP-T-eff-A | *tet*(32) |
| STP-T-eff-A | *cmxA* |
| STP-T-eff-A | *blaOXA-118* |
| STP-T-eff-A | *vanA-G* |
| STP-T-eff-A | *lsa(E)* |
| STP-T-eff-A | *ere(D)* |
| STP-T-eff-A | *aadA5* |
| STP-T-eff-A | *blaOXA-20* |
| STP-T-eff-A | *blaOXA-392* |
| STP-T-eff-A | *tet*(C) |
| STP-T-eff-A | *tet*(G) |
| STP-T-eff-A | *tet*(R) |
| STP-T-eff-A | *tet*(A) |
| STP-T-eff-A | *aac3-IIe* |
| STP-T-eff-A | *aadS* |
| STP-T-eff-A | *blaOXA-280* |
| STP-T-eff-A | *sul2* |
| STP-T-eff-A | *lnu(C)* |
| STP-T-eff-A | *blaOXA-666* |
| STP-T-eff-A | *cfr-Cb* |

| | |
|---|---|
| STP-T-eff-A | *aadA11* |
| STP-T-eff-A | *blaPAU-1* |
| STP-T-eff-A | *CblA-1* |
| STP-T-eff-A | *lin(B)* |
| STP-T-eff-A | *erm*(G) |
| STP-T-eff-A | *AAC(6')-IIa* |
| STP-T-eff-A | *blaRCP* |
| STP-T-eff-A | *tetR*(G) |
| STP-T-eff-A | *Bifidobacteria_intrinsic_ileS_confer* |
| STP-T-eff-A | *strB* |
| STP-T-eff-A | *aadA1* |
| STP-T-eff-A | *AAC(6')-Ib7* |
| STP-T-eff-A | *blaOXA-224* |
| STP-T-eff-A | *blaOXA-101* |
| STP-T-eff-A | *ant(3'')-IIc* |
| STP-T-eff-A | *catA13* |
| STP-T-eff-A | *spw* |
| STP-T-eff-A | *tet*(S) |
| STP-T-eff-A | *ant(3'')-IIa* |
| STP-T-eff-A | *blaOXA-119* |
| STP-T-eff-A | *aac6-Aph2* |
| STP-T-eff-A | *aph(3'')-III* |
| STP-T-eff-A | *aadA6* |
| STP-T-eff-A | *blaOXA-427* |
| STP-T-eff-A | *blaOXA-347* |
| STP-T-eff-A | *blaOXA-5* |
| STP-T-eff-A | *lnu(G)* |
| STP-T-eff-A | *catQ* |
| STP-T-eff-A | *floR2* |
| STP-T-eff-A | *aad9* |
| STP-T-eff-A | *blaMOX-9* |
| STP-T-eff-A | *blaOXA-209* |
| STP-T-eff-A | *blaOXA-2* |
| STP-T-eff-A | *blaOXA-281* |
| STP-T-eff-A | *blaOXA-464* |
| STP-T-eff-A | *blaOXA-33* |
| STP-T-eff-A | *cfxA_gen* |
| STP-T-eff-A | *blaBEL-1* |
| STP-T-eff-A | *blaOXA-643* |
| STP-T-eff-A | *aadA13* |
| STP-T-eff-A | *cat-TC* |
| STP-T-eff-A | *tet*(44) |
| STP-T-eff-A | *catA9* |
| STP-T-eff-A | *blaOXA-205* |

| | |
|---|---|
| STP-T-eff-A | *lnu((B)* |
| STP-T-eff-A | *aadA2* |
| STP-T-eff-A | *tet*(E) |
| STP-T-eff-A | *blaOXA-650* |
| STP-T-eff-A | *blaPER-1* |
| STP-T-eff-A | *catS* |
| STP-T-eff-A | *aph3-VIb* |
| STP-T-eff-A | *ICR-Mo* |
| STP-T-eff-A | *ant3''Ih-Aac6-IId* |
| STP-T-eff-A | *bacA* |
| STP-T-eff-A | *gadX* |
| STP-T-eff-A | *aac3-Ib-Aac6-Ib* |
| STP-T-eff-A | *blaOXA-18* |
| STP-T-eff-A | *tolC* |
| STP-T-eff-A | *Nocardia_rifampin_resistant_beta-s* |
| STP-T-eff-A | *aac(6')-30* |
| STP-T-eff-A | *blaOXA-139* |
| STP-T-eff-A | *mdtG* |
| STP-T-eff-A | *mph*(A) |
| STP-T-eff-A | *mef*(B) |
| STP-T-eff-A | *aadA24* |
| STP-T-eff-A | *aph7* |
| STP-T-eff-A | *acrB* |
| STP-T-eff-A | *qnrS2* |
| STP-T-eff-A | *aac(3)-IIe* |
| STP-T-eff-A | *erm*(A) |
| STP-T-eff-A | *emrY* |
| STP-T-eff-A | *aph(3'')-Ia* |
| STP-T-eff-A | *blaVEB-1* |
| STP-T-eff-A | *catB3* |
| STP-T-eff-A | *aadA17* |
| STP-T-eff-A | *mdtB* |
| STP-T-eff-A | *aacA-ACI1* |
| STP-T-eff-A | *blaOXA-836* |
| STP-T-eff-A | *blaOXA-212* |
| STP-T-eff-A | *blaOXA-15* |
| STP-T-eff-A | *kdpE* |
| STP-T-eff-A | *cfr(C)* |
| STP-T-eff-A | *mdtO* |
| STP-T-eff-A | *aac3-I* |
| STP-T-eff-A | *tetA(P)* |
| STP-T-eff-A | *Klebsiella_pneumoniae_acrA* |
| STP-T-eff-A | *mdtF* |
| STP-T-eff-A | *cfxA3* |

| | |
|---|---|
| STP-T-eff-A | *fosE* |
| STP-T-eff-A | *mdtP* |
| STP-T-eff-A | *CARB-10* |
| STP-T-eff-A | *AAC(6')-30/AAC(6')-Ib'_fusion_prote* |
| STP-T-eff-A | *erm*(47) |
| STP-T-eff-A | *blaOXA-645* |
| STP-T-eff-A | *emrA* |
| STP-T-eff-A | *acrF* |
| STP-T-eff-A | *mcr-3.17* |
| STP-T-eff-A | *ant6-Ib* |
| STP-T-eff-A | *Escherichia_coli_mdfA* |
| STP-T-eff-A | *cblA* |
| STP-T-eff-A | *ampH* |
| STP-T-eff-A | *CRP* |
| STP-T-eff-A | *blaOXA-373* |
| STP-T-eff-A | *blaOXA-662* |
| STP-T-eff-A | *msbA* |
| STP-T-eff-A | *aac(3)-Ia* |
| STP-T-eff-A | *mdtH* |
| STP-T-eff-A | *floR* |
| STP-T-eff-A | *MexF* |
| STP-T-eff-A | *acrE* |
| STP-T-eff-A | *evgS* |
| STP-T-eff-A | *vanZ-A* |
| STP-T-eff-A | *Klebsiella_pneumoniae_KpnG* |
| STP-T-eff-A | *tet*(42) |
| STP-T-eff-A | *Penicillin_Binding_Protein_Ecoli* |
| STP-T-eff-A | *baeS* |
| STP-T-eff-A | *evgA* |
| STP-T-eff-A | *Klebsiella_pneumoniae_KpnH* |
| STP-T-eff-A | *eptA* |
| STP-T-eff-A | *tet*(T) |
| STP-T-eff-A | *tetB-P* |
| STP-T-eff-A | *mdtN* |
| STP-T-eff-A | *baeR* |
| STP-T-eff-A | *H-NS* |
| STP-T-eff-A | *aadA10* |
| STP-T-eff-A | *tetA-P* |
| STP-T-eff-A | *erm*(T) |
| STP-T-eff-A | *cpxA* |
| STP-T-eff-A | *emrR* |
| STP-T-eff-A | *ant(2")-Ia* |
| STP-T-eff-A | *blaRSC1-1* |
| STP-T-eff-A | *erm*(Q) |

| | |
|---|---|
| STP-T-eff-A | *Escherichia_coli_emrE* |
| STP-T-eff-A | *CARB-14/blaRTG* |
| STP-T-eff-A | *acrD* |
| STP-T-eff-A | *ampH_Ecoli* |
| STP-T-eff-A | *blaOXA-334* |
| STP-T-eff-A | *blaOXA-652* |
| STP-T-eff-A | *blaOXA-31* |
| STP-T-eff-A | *mphB* |
| STP-T-eff-A | *iri* |
| STP-T-eff-A | *gadW* |
| STP-T-eff-A | *ere(A)* |
| STP-T-eff-A | *mexW* |
| STP-T-eff-A | *aac3-IId* |
| STP-T-eff-A | *catP* |
| STP-T-eff-A | *tet*(36) |
| STP-T-eff-A | *nimE_Nitroimidazole_Gene* |
| STP-T-eff-A | *mdtE* |
| STP-T-eff-A | *vanY-A* |
| STP-T-eff-A | *emrK* |
| STP-T-eff-A | *mcr-3.6* |
| STP-T-eff-A | *sat4* |
| STP-T-eff-A | *mdtM* |
| STP-T-eff-A | *mdtC* |
| STP-T-eff-A | *str* |
| STP-T-eff-A | *aac3-IIa* |
| STP-T-eff-A | *blaTEM-1* |
| STP-T-eff-A | *Salmonella_enterica_cmlA* |
| STP-T-eff-A | *dfrB1* |
| STP-T-eff-A | *aph(2'')-IIa* |
| STP-T-eff-A | *cphA1* |
| STP-T-eff-A | *cphA5* |
| STP-T-eff-A | *ere(A2)* |
| STP-T-eff-A | *dfrA14* |
| STP-T-eff-A | *lnu(A)* |
| STP-T-eff-A | *nimD_Nitroimidazole_Gene* |
| STP-T-eff-A | *acrS* |
| STP-T-eff-A | *nimj_Nitroimidazole_Gene* |
| STP-T-eff-A | *aad(6)* |
| STP-T-eff-A | *cmr* |
| STP-T-eff-A | *aadA16* |
| STP-T-eff-A | *aph4-Ia* |
| STP-T-eff-A | *vanY-D* |
| STP-T-eff-A | *aac6-Im* |
| STP-T-eff-A | *ere(B)* |

| | |
|---|---|
| STP-T-eff-A | *mexK* |
| STP-T-eff-A | *blaAIM-1* |
| STP-T-eff-A | *Klebsiella_pneumoniae_OmpK37* |
| STP-T-eff-A | *cfxA4* |
| STP-T-eff-A | *pmrF* |
| STP-T-eff-A | *vanH-A* |
| STP-T-eff-A | *aac6-IIc* |
| STP-T-eff-A | *blaNPS-1* |
| STP-T-eff-A | *Enterobacter_cloacae_acrA* |
| STP-T-eff-A | *Escherichia_coli_acrA* |
| STP-T-eff-A | *blaOXA-37* |
| STP-T-eff-A | *mcr-3.3* |
| STP-T-eff-A | *vanX-A* |
| STP-T-eff-A | *tet*(33) |
| STP-T-eff-A | *yojI* |
| STP-T-eff-A | *blaOXA-644* |
| STP-T-eff-A | *blaOXA-9* |
| STP-T-eff-A | *blaOXA-320* |
| STP-T-eff-A | *MexB* |
| STP-T-eff-A | *aci1* |
| STP-T-eff-A | *SHV-1* |
| STP-T-eff-A | *blaGES-1* |
| STP-T-eff-A | *AmpC1_Ecoli* |
| STP-T-eff-A | *OqxBgb* |
| STP-T-eff-A | *cepA* |
| STP-T-eff-A | *Pseudomonas_aeruginosa_CpxR* |
| STP-T-eff-A | *blaRm3* |
| STP-T-eff-A | *mcr-3.12* |
| STP-T-eff-A | *oqxA10* |
| STP-T-eff-A | *blaBKC-1* |
| STP-T-eff-A | *tetB*(46) |
| STP-T-eff-A | *mdtA* |
| STP-T-eff-A | *lnu(P)* |
| STP-T-eff-A | *bla2* |
| STP-T-eff-A | *aph(3')-VIa* |
| STP-T-eff-A | *MuxB* |
| STP-T-eff-A | *blaOXA-725* |
| STP-T-eff-A | *blaOXA-726* |
| STP-T-eff-A | *nimA_Nitroimidazole_Gene* |
| STP-T-eff-A | *blaOXA-46* |
| STP-T-eff-A | *aac(6')-Im* |
| STP-T-eff-A | *mcr-3.1* |
| STP-T-eff-A | *lnu(F)* |
| STP-T-eff-A | *blaOXA-309* |

| | |
|---|---|
| STP-T-eff-A | *blaOXA-160* |
| STP-T-eff-A | *blaOXA-161* |
| STP-T-eff-A | *blaOXA-21* |
| STP-T-eff-A | *smeE* |
| STP-T-eff-A | *lmrD* |
| STP-T-eff-A | *RSA-1* |
| STP-T-eff-A | *tetA(46)* |
| STP-T-eff-A | *blaPSE-4* |
| STP-T-eff-A | *cmrA_variant1* |
| STP-T-eff-A | *OqxA* |
| STP-T-eff-A | *lsa(C)* |
| STP-T-eff-A | *aph(2")-If* |
| STP-T-eff-A | *eat(A)* |
| STP-T-eff-A | *cepA_beta-lactamase* |
| STP-T-eff-A | *vanT-G* |
| STP-T-eff-A | *cmlA1* |
| STP-T-eff-A | *catA8* |
| STP-T-eff-A | *mcr-5.1* |
| STP-T-eff-A | *vanR-A* |
| STP-T-eff-A | *vanR-D* |
| STP-T-eff-A | *ble-MBL* |
| STP-T-eff-A | *blaIMP-16* |
| STP-T-eff-A | *vanS-D* |
| STP-T-eff-A | *cphA7* |
| STP-T-eff-A | *imiH* |
| STP-T-eff-A | *vanXY-G* |
| STP-T-eff-A | *blaFOX-10* |
| STP-T-eff-A | *blaFOX-4* |
| STP-T-eff-A | *marA* |
| STP-T-eff-A | *aac(3)-IVa* |
| STP-T-eff-A | *ugd* |
| STP-T-eff-A | *aph(3')-VI* |
| STP-T-eff-A | *AxyY* |
| STP-T-eff-A | *aadA25* |
| STP-T-eff-A | *adeJ* |
| STP-T-eff-A | *aphA15* |
| STP-T-eff-A | *blaOXA-504* |
| STP-T-eff-A | *aadA7* |
| STP-T-eff-A | *blaOXA-13* |
| STP-T-eff-A | *blaOXA-256* |
| STP-T-eff-A | *blaOXA-732* |
| STP-T-eff-A | *tet*(H) |
| STP-T-eff-A | *vanY-B* |
| STP-T-eff-A | *mcr-4.1* |

| | |
|---|---|
| STP-T-eff-A | *srm(B)* |
| STP-T-eff-A | *blaOXA-299* |
| STP-T-eff-A | *vanYD* |
| STP-T-eff-A | *fosA_gen* |
| STP-T-eff-A | *FosA6* |
| STP-T-eff-A | *Streptomyces_rishiriensis_parY_mut* |
| STP-T-eff-A | *efmA* |
| STP-T-eff-A | *blaCPS-1* |
| STP-T-eff-A | *blaCTX-M-101* |
| STP-T-eff-A | *penI_Bp* |
| STP-T-eff-A | *blaGPC-1* |
| STP-T-eff-A | *aadA9* |
| STP-T-eff-A | *oqxB11* |
| STP-T-eff-A | *arr-8* |
| STP-T-eff-A | *aac(6')_Strep* |
| STP-T-eff-A | *dfr32* |
| STP-T-eff-A | *OpmH* |
| STP-T-eff-A | *dfrG* |
| STP-T-eff-A | *vanZ1* |
| STP-T-eff-A | *emrB* |
| STP-T-eff-A | *sat-2A* |
| STP-T-eff-A | *oqxB15* |
| STP-T-eff-A | *aac3-Ib* |
| STP-T-eff-A | *LpeA* |
| STP-T-eff-A | *catB4* |
| STP-T-eff-A | *aac(6')-Ib-cr3* |
| STP-T-eff-A | *aac(6')-Ib-G* |
| STP-T-eff-A | *aac(6')-Ib-generic* |
| STP-T-eff-A | *aacA38* |
| STP-T-eff-A | *otr(B)* |
| STP-T-eff-A | *mexV* |
| STP-T-eff-A | *sta* |
| STP-T-eff-A | *blaFOX-3* |
| STP-T-eff-A | *blaFOX-9* |
| STP-T-eff-A | *blaADC-18* |
| STP-T-eff-A | *MexA* |
| STP-T-eff-A | *blaACC-1* |
| STP-T-eff-A | *Tet*(53) |
| STP-T-eff-A | *dfrA36* |
| STP-T-eff-A | *smeD* |
| STP-T-eff-A | *AAC(6')-Ib-cr* |
| STP-T-eff-A | *Tet*(48) |
| STP-T-eff-A | *tetA(D)* |
| STP-T-eff-A | *vanX-B* |

| | |
|---|---|
| STP-T-eff-A | *AAC(6')-Ib10* |
| STP-T-eff-A | *vanX-D* |
| STP-T-eff-A | *cmlA* |
| STP-T-eff-A | *cmlA5* |
| STP-T-eff-A | *smeB* |
| STP-T-eff-A | *aac(2')-Id* |
| STP-T-eff-A | *catB8* |
| STP-T-eff-A | *fusC* |
| STP-T-eff-A | *Acinetobacter_baumannii_AbaQ* |
| STP-T-eff-A | *catA1* |
| STP-T-eff-A | *catA16* |
| STP-T-eff-A | *catB* |
| STP-T-eff-A | *npmA* |
| STP-T-eff-A | *vanS-B* |
| STP-T-eff-A | *AAC(6')-Ib8* |
| STP-T-eff-A | *mtrA* |
| STP-T-eff-A | *tet*(L) |
| STP-T-eff-A | *Mbl* |
| STP-T-eff-A | *smeC* |
| STP-T-eff-A | *PmpM* |
| STP-T-eff-A | *erm*(44) |
| STP-T-eff-A | *erm*(C) |
| STP-T-eff-A | *msr*(C) |
| STP-T-eff-A | *blaIMP-39* |
| STP-T-eff-A | *OpmB* |
| STP-T-eff-A | *cfiA10* |
| STP-T-eff-A | *blaOXA-85* |
| STP-T-eff-A | *cepH-A3* |
| STP-T-eff-A | *vanXY-G2* |
| STP-T-eff-A | *qepA2* |
| STP-T-eff-A | *BcII* |
| STP-T-eff-A | *blaOXA-34* |
| STP-T-eff-A | *aph3-Va* |
| STP-T-eff-A | *blaLCR-1* |
| STP-T-eff-A | *vga(E)* |
| STP-T-eff-A | *aadA4* |
| STP-T-eff-A | *blaFIM-1* |
| STP-T-eff-A | *blaOXA-198* |
| STP-T-eff-A | *aadA12* |
| STP-T-eff-A | *aadA15* |
| STP-T-eff-A | *aadA3* |
| STP-T-eff-A | *blaOXA-780* |
| STP-T-eff-A | *blaOXA-663* |
| STP-T-eff-A | *blaOXA-779* |

| | |
|---|---|
| STP-T-eff-A | *blaOXA-827* |
| STP-T-eff-A | *blaOXA-830* |
| STP-T-eff-A | *aph3-IIc* |
| STP-T-eff-A | *blaAFM-1* |
| STP-T-eff-A | *mcr-10.1* |
| STP-T-eff-A | *aph(3')-Ia* |
| STP-T-eff-A | *aph(3')-Ib* |
| STP-T-eff-A | *blaOXA-304* |
| STP-T-eff-A | *blaOXA-651* |
| STP-T-eff-A | *blaOXA-24* |
| STP-T-eff-A | *blaOXA-458* |
| STP-T-eff-A | *blaOXA-675* |
| STP-T-eff-A | *blaOXA-669* |
| STP-T-eff-A | *blaOXA-274* |
| STP-T-eff-A | *blaZ* |
| STP-T-eff-A | *vanW-G* |
| STP-T-eff-A | *blaOXA-727* |
| STP-T-eff-A | *erm*(X) |
| STP-T-eff-A | *blaPEDO-2* |
| STP-T-eff-A | *blaSHV-149* |
| STP-T-eff-A | *aac3-IIc* |
| STP-T-eff-A | *blaPOM-1* |
| STP-T-eff-A | *blaSHV-128* |
| STP-T-eff-A | *blaSHV-42* |
| STP-T-eff-A | *blaSHV-67* |
| STP-T-eff-A | *blaTEM-106* |
| STP-T-eff-A | *aac(3)-VIII* |
| STP-T-eff-A | *blaGES-10* |
| STP-T-eff-A | *sul4* |
| STP-T-eff-A | *CARB-1* |
| STP-T-eff-A | *CARB-2* |
| STP-T-eff-A | *CARB-4* |
| STP-T-eff-A | *blaCAU-1* |
| STP-T-eff-A | *blaESP-1* |
| STP-T-eff-A | *blaCTX-M-115* |
| STP-T-eff-A | *blaSFO-1* |
| STP-T-eff-A | *blaA_Yent* |
| STP-T-eff-A | *blaFRI-1* |
| STP-T-eff-A | *blaCME-1* |
| STP-T-eff-A | *blaLRA-1* |
| STP-T-eff-A | *blaCARB-16* |
| STP-T-eff-A | *mphH* |
| STP-T-eff-A | *blaVEB-24* |
| STP-T-eff-A | *vanY-Pt2* |

| | |
|---|---|
| STP-T-eff-A | *cepA-44* |
| STP-T-eff-A | *cepA-49* |
| STP-T-eff-A | *hugA* |
| STP-T-eff-A | *aph(2")-Id* |
| STP-T-eff-A | *erm*(42) |
| STP-T-eff-A | *blaEFM-1* |
| STP-T-eff-A | *blaL2* |
| STP-T-eff-A | *blaR39* |
| STP-T-eff-A | *blaEVM-1* |
| STP-T-eff-A | *aph(2")-Ic* |
| STP-T-eff-A | *aph(2")-Ig* |
| STP-T-eff-A | *aph(6)-Ia* |
| STP-T-eff-A | *aph6-Ib* |
| STP-T-eff-A | *blaM-1* |
| STP-T-eff-A | *vanH-B* |
| STP-T-eff-A | *vanH-M* |
| STP-T-eff-A | *vanJ* |
| STP-T-eff-A | *erm*(30) |
| STP-T-eff-A | *TriC* |
| STP-T-eff-A | *vanB* |
| STP-T-eff-A | *mexI* |
| STP-T-eff-A | *vanA-D* |
| STP-T-eff-A | *vanN* |
| STP-T-eff-A | *MuxC* |
| STP-T-eff-A | *mexY* |
| STP-T-eff-A | *vanH-Ao1* |
| STP-T-eff-A | *cfr(B)* |
| STP-T-eff-A | *vanG* |
| STP-T-eff-A | *vanG2* |
| STP-T-eff-A | *oqxB17* |
| STP-T-eff-A | *otrC* |
| STP-T-eff-A | *vanS-G* |
| STP-T-eff-A | *mexH* |
| STP-T-eff-A | *AmpC2_Ecoli* |
| STP-T-eff-A | *blaEC* |
| STP-T-eff-A | *blaEC-13* |
| STP-T-eff-A | *blaEC-19* |
| STP-T-eff-A | *blaEC-5* |
| STP-T-eff-A | *blaACT-28* |
| STP-T-eff-A | *blaACT-3* |
| STP-T-eff-A | *blaACT-38* |
| STP-T-eff-A | *blaCFE-1* |
| STP-T-eff-A | *blaCMY-101* |
| STP-T-eff-A | *blaCMY-105* |

| | |
|---|---|
| STP-T-eff-A | *blaCMY-FOX* |
| STP-T-eff-A | *blaFOX-13* |
| STP-T-eff-A | *blaFOX-16* |
| STP-T-eff-A | *blaFOX-2* |
| STP-T-eff-A | *blaMOX-3* |
| STP-T-eff-A | *blaADC-133* |
| STP-T-eff-A | *blaADC-2* |
| STP-T-eff-A | *blaMOX-6* |
| STP-T-eff-A | *vanS-A* |
| STP-T-eff-A | *erm*(38) |
| STP-T-eff-A | *mexX* |
| STP-T-eff-A | *oqxA11* |
| STP-T-eff-A | *macA* |
| STP-T-eff-A | *blaADC-8* |
| STP-T-eff-A | *blaPDC-125* |
| STP-T-eff-A | *blaPDC-160* |
| STP-T-eff-A | *amrA* |
| STP-T-eff-A | *bcr1* |
| STP-T-eff-A | *adeG* |
| STP-T-eff-A | *mdsA* |
| STP-T-eff-A | *adeH* |
| STP-T-eff-A | *opmD* |
| STP-T-eff-A | *Acinetobacter_baumannii_AmvA* |
| STP-T-eff-A | *cmeC* |
| STP-T-eff-A | *lsa(B)* |
| STP-T-eff-A | *lsa(A)* |
| STP-T-eff-A | *farB* |
| STP-T-eff-A | *opcM* |
| STP-T-eff-A | *mupA* |
| STP-T-eff-A | *MexD* |
| STP-T-eff-A | *lin* |
| STP-T-eff-A | *oqxB10* |
| STP-T-eff-A | *oqxB20* |
| STP-T-eff-A | *oqxB7* |
| STP-T-eff-A | *tetAB_B* |
| STP-T-eff-A | *tcmA* |
| STP-T-eff-A | *mcr-7.1* |
| STP-T-eff-A | *mcr-9.1* |
| STP-T-eff-A | *mcr-3.13* |
| STP-T-eff-A | *mcr-4.3* |
| STP-T-eff-A | *rif* |
| STP-T-eff-A | *tlr(C)* |
| STP-T-eff-A | *vga(B)* |
| STP-T-eff-A | *rosB* |

| | |
|---|---|
| STP-T-eff-A | *Clostridium_perfringens_mprF* |
| STP-T-eff-A | *ole(B)* |
| STP-T-eff-A | *Tet*(60) |
| STP-T-eff-A | *blaR1-2* |
| STP-T-eff-A | *novA* |
| STP-T-eff-A | *mecA1* |
| STP-T-eff-A | *mecC3* |
| STP-T-eff-A | *mexN* |
| STP-T-eff-A | *cmeB* |
| STP-T-eff-A | *amrB* |
| STP-T-eff-A | *oqxB12* |
| STP-T-eff-A | *oqxB13* |
| STP-T-eff-A | *oqxB14* |
| STP-T-eff-A | *oqxB16* |
| STP-T-eff-A | *oqxB18* |
| STP-T-eff-A | *oqxB2* |
| STP-T-eff-A | *oqxB21* |
| STP-T-eff-A | *oqxB24* |
| STP-T-eff-A | *oqxB26* |
| STP-T-eff-A | *oqxB28* |
| STP-T-eff-A | *oqxB31* |
| STP-T-eff-A | *oqxB9* |
| STP-T-eff-A | *mexQ* |
| STP-T-eff-A | *mtrD* |

**Legend:** eff (effluent); F (February); A (April); STP (sewage treatme

s) across six samples.

| Relativ abundance |
|---|
| 0.0372923062621251 |
| 0.032236423540148 |
| 0.0193719187723776 |
| 0.0166497703688608 |
| 0.015556603627471 |
| 0.0139625061479274 |
| 0.0129271753275136 |
| 0.011574584145 |
| 0.0102528809466608 |
| 0.00965529656184482 |
| 0.00891643711299312 |
| 0.0078168655903987 |
| 0.007600594376 |
| 0.0073244477957278 |
| 0.007089543107832 |
| 0.0068017960249891 |
| 0.0047688841420738 |
| 0.004717879048929 |
| 0.004665479269 |
| 0.0043274733557832 |
| 0.004215638272 |
| 0.0040741468986264 |
| 0.0038586602889911 |
| 0.0037488469748781 |
| 0.00347193046150099 |
| 0.00342327892870512 |
| 0.00316614576030465 |
| 0.0031520111810176 |
| 0.0029989889965618 |
| 0.0025164690978005 |
| 0.0024589028112496 |
| 0.0023710879025514 |
| 0.0023564825315244 |
| 0.002343886532 |
| 0.002285172400419 |
| 0.0022684066454636 |
| 0.0022369508089109 |
| 0.0021934142315363 |
| 0.0021702285712824 |
| 0.0021583750546437 |
| 0.0019993259977079 |

| |
|---|
| 0.0018767580275615 |
| 0.001851820095 |
| 0.0017105410860866 |
| 0.00170250213435309 |
| 0.00166732799939058 |
| 0.00166289168083224 |
| 0.00150778617301045 |
| 0.00138518877 |
| 0.0013418427269861 |
| 0.0013339916304114 |
| 0.00133247060954897 |
| 0.0013276659549252 |
| 0.0013093083971184 |
| 0.0012826603293345 |
| 0.0012700636460849 |
| 0.001236346191 |
| 0.0012292651533288 |
| 0.001219453899277 |
| 0.0012193605046305 |
| 0.00114366481836079 |
| 0.001126132054675 |
| 0.0010291244244971 |
| 0.0010115218922531 |
| 0.0009911655598837 |
| 0.0009854723435904 |
| 0.0009812751993196 |
| 0.0009750811218329 |
| 0.0009645963785874 |
| 0.0009183193330753 |
| 0.000911280984011791 |
| 0.0009046130743727 |
| 0.0008927102707625 |
| 0.0008736483236607 |
| 0.000812305617804049 |
| 0.0008116301973901 |
| 0.000798603729719586 |
| 0.0007874684593744 |
| 0.0007690734049802 |
| 0.000738424687603452 |
| 0.000723396753954513 |
| 0.000714936139423551 |
| 0.0006784598057795 |
| 0.0006656284146537 |
| 0.00056806271258 |

| |
|---|
| 0.0005653831714828 |
| 0.000565030444597184 |
| 0.000523075587222249 |
| 0.0005180965789589 |
| 0.0005143558094357 |
| 0.000513460227 |
| 0.0004826441707781 |
| 0.0004806741945708 |
| 0.0004801884470129 |
| 0.0004687540476295 |
| 0.0004440827819648 |
| 0.0004371736047154 |
| 0.00043276242755484 |
| 0.0004276352715216 |
| 0.0004276352715216 |
| 0.0004226470921249 |
| 0.0004194115163 |
| 0.0004097550511096 |
| 0.0004072902467555 |
| 0.0004043800166898 |
| 0.0004029640058569 |
| 0.0004011020235426 |
| 0.0003990940033997 |
| 0.000396466223953484 |
| 0.0003914191187189 |
| 0.0003907795449955 |
| 0.000378294152238062 |
| 0.000356497763299512 |
| 0.0003490271564028 |
| 0.000339229902889698 |
| 0.0003230760979902 |
| 0.0003220758623927 |
| 0.0003218969881435 |
| 0.000321544931649 |
| 0.000321406454073 |
| 0.0003191679193855 |
| 0.0003160220347153 |
| 0.0003157611674697 |
| 0.0003119355428871 |
| 0.0003098623516512 |
| 0.0003040093118793 |
| 0.0002929446484367 |
| 0.0002896544326809 |
| 0.0002878314327549 |

| |
|---|
| 0.0002834000859563 |
| 0.0002832362708777 |
| 0.0002826915857414 |
| 0.0002821531255781 |
| 0.000280935737382813 |
| 0.0002795152757893 |
| 0.0002785946062379 |
| 0.0002713839223118 |
| 0.0002713839223118 |
| 0.0002711531536704 |
| 0.0002709127696689 |
| 0.00026317068340119 |
| 0.0002627506855024 |
| 0.0002625749560445 |
| 0.0002605881284201 |
| 0.0002601763267001 |
| 0.000256969698965086 |
| 0.0002561606602188 |
| 0.000253108874787627 |
| 0.0002525867675196 |
| 0.0002504119237018 |
| 0.0002478391984582 |
| 0.0002458187702099 |
| 0.0002456973782247 |
| 0.0002389947791126 |
| 0.0002366720252719 |
| 0.0002365001501628 |
| 0.0002353631302101 |
| 0.0002321046703982 |
| 0.0002316159079311 |
| 0.0002312931156066 |
| 0.0002301559813116 |
| 0.0002281370691948 |
| 0.0002261532685931 |
| 0.0002259360224177 |
| 0.0002238377061161 |
| 0.0002226379328119 |
| 0.0002202241566825 |
| 0.0002189797926277 |
| 0.000218396986426421 |
| 0.0002120186893061 |
| 0.0002110763840203 |
| 0.0002104711076323 |
| 0.000208159794804906 |

0.0002071969517325
0.0002037416834172
0.0002019225612439
0.0002017724329455
0.0001991690717724
0.0001988785025317
0.0001985736016915
0.0001973701253176
0.0001944304119126
0.0001939716143879
0.000193907908524
0.00019384565879416З
0.0001920993015077
0.0001870854290065
0.0001846149131372
0.0001824708524101
0.0001820428787437
0.0001816256820308
0.0001815829163886
0.0001771079814284
0.0001751355278667
0.0001744112611258
0.0001739640527639
0.0001736126102331
0.0001735191319129
0.0001730764810662
0.0001720731391469
0.0001692303370425
0.0001661012707181
0.0001657157571588
0.0001654780014096
0.0001652261323055
0.0001642453905984
0.0001611666971583
0.0001583072880152
0.0001524628777032
0.0001524628777032
0.0001498871565724
0.0001474069332168
0.0001459053345762
0.0001450794553239
0.0001449700439699
0.0001418382869225
0.000135939123198

| |
|---|
| 0.0001291074796916 |
| 0.0001281840388787 |
| 0.000127052398086 |
| 0.000126814916968045 |
| 0.0001256407047739 |
| 0.000124000486696 |
| 0.0001202942918048 |
| 0.0001181397671755 |
| 0.0001155527649746 |
| 0.0001122746723512 |
| 0.0001121421166577 |
| 0.000111405551 |
| 0.0001103186676064 |
| 0.0001093720955754 |
| 0.000108703845061814 |
| 0.0001084296493255 |
| 0.000105507610094186 |
| 0.0001054327592509 |
| 0.000104675673135551 |
| 0.0000966969942757763 |
| 0.0000949183426577261 |
| 0.0000898622259310692 |
| 0.0000898622259310692 |
| 0.0000894851062778813 |
| 0.0000882158139902518 |
| 0.0000856641168913602 |
| 0.0000841500534300245 |
| 0.0000787990482903104 |
| 0.0000777775791458064 |
| 0.0000772573277802826 |
| 0.0000769228804738745 |
| 0.0000767622060499081 |
| 0.0000759670155161861 |
| 0.000075384422864397 |
| 0.000075133976 |
| 0.000074885188275891 |
| 0.0000743925225635496 |
| 0.0000734794013479682 |
| 0.0000729526672881261 |
| 0.0000722245967563085 |
| 0.0000682726848583218 |
| 0.0000677612789792332 |
| 0.000066884515665269 |
| 0.0000653150234204161 |

0.0000646152195980545
0.0000641707468923069
0.000064156955629274
0.000063705146082589
0.0000632596555505429
0.0000632105409111403
0.0000628203523869975
0.0000624732786169035
0.0000621727198881624
0.0000620812894177387
0.0000619597996145728
0.0000616781641617793
0.0000611225050251867
0.0000611225050251867
0.00006065413333917
0.0000598289070352357
0.0000597385992510315
0.000058632328894531
0.000057566286550994
0.0000572539920489091
0.0000568224292947716
0.0000561373361756147
0.0000554297226944095
0.0000551593338032173
0.0000542148246627512
0.0000537947832048503
0.0000525937833937653
0.0000516429918766563
0.0000513984701348161
0.0000510600041978982
0.0000510120154721483
0.0000499666203423957
0.000049764777930042
0.0000496434004228955
0.0000494324084356701
0.0000493425313294234
0.0000491173293450745
0.0000489862675653121
0.0000488980040201494
0.0000484614146985409
0.000048375030715121
0.000047520266562084
0.0000474447416629072
0.0000473619410666368

0.0000472794289741862
0.0000467500296833469
0.0000443437781555276
0.0000432140003681256
0.0000429404940366818
0.0000429404940366818
0.0000426704280364511
0.0000423507993620207
0.0000421927739912669
0.0000420750267150122
0.000041880234924665
0.0000411187761078529
0.0000408710726373236
0.0000408710726373236
0.0000396760120338931
0.0000396760120338931
0.0000393995241451552
0.0000393799223918491
0.0000393310032335984
0.0000388579499301015
0.000038724874759108
0.0000376026812387728
0.0000368257238130675
0.000036673503015112
0.0000358973442211414
0.0000358973442211414
0.0000358973442211414
0.0000347038263825868
0.0000344286612511042
0.0000341363424291609
0.0000338806394896166
0.0000337542191930135
0.0000333805562499174
0.0000328950208862823
0.0000328711146211033
0.000032657511710208
0.000032657511710208
0.000032657511710208
0.0000323076097990272
0.0000321110117677958
0.0000318525730412945
0.0000315196193161241
0.0000315196193161241
0.0000315196193161241

0.0000315196193161241
0.0000309798998072864
0.0000309798998072864
0.0000307691521895498
0.000029601213166648
0.0000294982524251988
0.000029447040181405
0.0000288460801777029
0.0000286269960244545
0.0000286269960244545
0.0000286269960244545
0.0000284469520243007
0.0000279732872665222
0.0000279732872665222
0.000026923008165856
0.000026711016
0.0000266062668933165
0.0000266062668933165
0.0000266062668933165
0.0000266062668933165
0.0000261448865425654
0.0000258460878392218
0.0000256022568218706
0.0000255540416489481
0.0000255060077360741
0.00002541048
0.0000254104796172124
0.0000254104796172124
0.0000253156643947601
0.0000249433752124843
0.000024581877020999
0.0000244490020100747
0.0000242740538740455
0.0000242464517984902
0.0000240588583609777
0.0000238056072203358
0.0000238056072203358
0.0000237828485136242
0.0000236809705333184
0.0000236397144870931
0.0000236397144870931
0.0000236397144870931
0.0000236397144870931
0.000023557632145124

0.000023557632145124
0.0000231358842550579
0.0000230768641421623
0.0000227862235358378
0.0000217455065954991
0.0000213352140182255
0.0000210701803037134
0.0000210375133575061
0.0000208628476561984
0.0000206532665381909
0.000019751377169711
0.0000193661219537461
0.0000191655312367111
0.0000185371531633763
0.0000181285185245042
0.0000180922614874552
0.0000180922614874552
0.0000179486721105707
0.0000179486721105707
0.0000177607278999888
0.0000177607278999888
0.0000177549180446077
0.000017737511262211
0.000017737511262211
0.000017343042
0.00001732975238262
0.0000173164830469518
0.0000173076481066217
0.000017132823378272
0.000017132823378272
0.0000170896676518784
0.0000170040051573827
0.0000170040051573827
0.0000169403197448083
0.0000169403197448083
0.0000168771095965067
0.000016752093969866
0.0000164475104431411
0.0000163879180139993
0.000016328755855104
0.000016328755855104
0.000016328755855104
0.000016328755855104
0.0000161538048995136

| |
|---|
| 0.00001606202191713 |
| 0.000016039238907318 |
| 0.000015926286520647 |
| 0.000015870404813557 |
| 0.00001575980965806 |
| 0.00001575980965806 |
| 0.00001575980965806 |
| 0.00001575980965806 |
| 0.00001575980965806 |
| 0.00001575980965806 |
| 0.000015489949903643 |
| 0.000015489949903643 |
| 0.000015384576094775 |
| 0.000015369639613129 |
| 0.000015178071717664 |
| 0.000015178071717664 |
| 0.000015076884572879 |
| 0.000014977037655178 |
| 0.00001482972253070 |
| 0.00001468527718138 |
| 0.000014497004396999 |
| 0.000014313498012227 |
| 0.000014193719786184 |
| 0.000013960078308222 |
| 0.000013706258702618 |
| 0.000013303133446658 |
| 0.000013251168081632 |
| 0.00001308504929180 |
| 0.000012540846687238 |
| 0.000012494655723381 |
| 0.000012290938510500 |
| 0.000011902803610168 |
| 0.000011840485266659 |
| 0.000011840485266659 |
| 0.000011809570161524 |
| 0.000011809570161524 |
| 0.000011809570161524 |
| 0.000011799300970080 |
| 0.000011538432071081 |
| 0.000011518842203388 |
| 0.000011450798409782 |
| 0.000011450798409782 |
| 0.000011279464767740 |
| 0.000011223487275096 |

0.0000111957063659995
0.0000111405551031128
0.0000108310952391375
0.0000107692032663424
0.0000107181643883028
0.0000107181643883028
0.0000105926589505007
0.000010518756678753
0.0000104218096125894
0.0000103740031464766
0.0000103266332690954
0.0000102796940269632
0.0000102796940269632
0.0000102331795743525
0.0000100067817961588
9.96269905696876E-06
9.74798571522375E-06
9.70614886666055E-06
9.42305285804962E-06
0.0000093645245794282
9.32590798322437E-06
9.26857658168815E-06
9.23074565686493E-06
9.11908341101576E-06
9.04613074372764E-06
9.04613074372764E-06
9.04613074372764E-06
8.86875563110552E-06
8.86875563110552E-06
8.83411205442152E-06
8.83411205442152E-06
8.79973807755607E-06
8.76563056562755E-06
8.76563056562755E-06
8.69820263819965E-06
0.00000866487619131
8.60716531277606E-06
8.60716531277606E-06
8.59898359669927E-06
8.53408560729022E-06
8.47015987240415E-06
8.47015987240415E-06
8.47015987240415E-06
8.47015987240415E-06

8.47015987240415E-06
8.43855479825339E-06
8.43855479825339E-06
8.40718470606658E-06
0.000008376046984933
8.34513906247937E-06
8.22375522157058E-06
8.22375522157058E-06
8.22375522157058E-06
8.19395900699967E-06
8.19395900699967E-06
8.19395900699967E-06
8.19395900699967E-06
8.19395900699967E-06
8.19395900699967E-06
8.16437792755202E-06
8.16437792755202E-06
8.13500966162557E-06
8.07690244975682E-06
8.04815902466872E-06
8.04815902466872E-06
7.99128157573113E-06
7.96314326032362E-06
7.95380780515325E-06
7.93520240677863E-06
7.93520240677863E-06
7.93520240677863E-06
7.93520240677863E-06
7.90745694381786E-06
7.87990482903104E-06
7.87990482903104E-06
7.87990482903104E-06
7.87990482903104E-06
7.87990482903104E-06
7.87990482903104E-06
7.87990482903104E-06
7.87990482903104E-06
7.87990482903104E-06
7.87990482903104E-06
7.87990482903104E-06
7.87990482903104E-06
7.85254404837468E-06
7.85254404837468E-06
7.85254404837468E-06

7.85254404837468E-06
7.83440884272024E-06
7.81185729164735E-06
0.000007798388572179
0.0000077715899860203
0.0000077715899860203
0.0000077715899860203
0.0000077449749518216
7.69228804738745E-06
7.69228804738745E-06
7.69228804738745E-06
7.66621249468444E-06
7.56365446799969E-06
0.0000075384422864397
0.0000075384422864397
0.0000075384422864397
0.0000075384422864397
0.0000074638042439997
0.0000074638042439997
0.0000074638042439997
7.36655598023423E-06
7.34263859068801E-06
7.34263859068801E-06
7.31887600625213E-06
7.06728964353721E-06
6.95856211055972E-06
6.93721682801199E-06
6.93721682801199E-06
0.0000069160020976511
6.71077948347747E-06
6.57422292422066E-06
6.57422292422066E-06
6.51738526205161E-06
6.46152195980545E-06
6.45537398458204E-06
6.45537398458204E-06
6.45537398458204E-06
6.45537398458204E-06
0.0000063705146082589
6.31713040763103E-06
6.16221440308422E-06
6.14546925524975E-06
5.99876044013769E-06
5.96710471222139E-06

| |
|---|
| 5.95140180508397E-06 |
| 5.93578132790527E-06 |
| 0.0000059202426333296 |
| 0.0000059202426333296 |
| 0.0000059202426333296 |
| 0.0000059202426333296 |
| 5.90478508076216E-06 |
| 5.90478508076216E-06 |
| 5.90478508076216E-06 |
| 5.88940803628101E-06 |
| 5.88940803628101E-06 |
| 5.88940803628101E-06 |
| 5.84375371041837E-06 |
| 5.82869248951523E-06 |
| 5.82869248951523E-06 |
| 5.79880175879976E-06 |
| 5.78397106376447E-06 |
| 5.76921603554058E-06 |
| 5.73993067495408E-06 |
| 5.72539920489091E-06 |
| 5.68224292947716E-06 |
| 5.68224292947716E-06 |
| 5.66800171912759E-06 |
| 5.57027755155642E-06 |
| 5.33380350455639E-06 |
| 5.27163796254524E-06 |
| 5.17513200442084E-06 |
| 5.03682112679712E-06 |
| 5.01448489120157E-06 |
| 0.0000048323347989998 |
| 4.77116600407576E-06 |
| 4.63428829084407E-06 |
| 4.59661115026811E-06 |
| 4.53212963112607E-06 |
| 4.45183599592895E-06 |
| 4.40844578154368E-06 |
| 4.40844578154368E-06 |
| 4.36168309726501E-06 |
| 4.30358265638803E-06 |
| 4.30358265638803E-06 |
| 4.19579348039315E-06 |
| 4.18028222907931E-06 |
| 4.18028222907931E-06 |
| 4.17256953123968E-06 |

| |
|---|
| 4.10441503798894E-06 |
| 4.03845122487841E-06 |
| 3.52263658244845E-06 |
| 3.48464204303838E-06 |
| 3.40592271977697E-06 |
| 3.22615219105835E-06 |
| 3.14101761934987E-06 |
| 2.61146961424008E-06 |
| 2.19993451938901E-06 |
| 2.18294660804238E-06 |
| 0.0000021724617540172 |
| 2.16001211645836E-06 |
| 2.15179132819401E-06 |
| 2.15179132819401E-06 |
| 2.15179132819401E-06 |
| 2.15179132819401E-06 |
| 2.15179132819401E-06 |
| 2.15179132819401E-06 |
| 0.000002141602922284 |
| 0.0677224472176267 |
| 0.0583401140368887 |
| 0.0365458507813935 |
| 0.0267970646865953 |
| 0.0257189331463022 |
| 0.0233680272851706 |
| 0.018102599311392 |
| 0.0176016503307348 |
| 0.0157499087671951 |
| 0.0097891678323759 |
| 0.008976934477 |
| 0.0081847674037917 |
| 0.0080472762517517 |
| 0.0078933636559208 |
| 0.0076636398911908 |
| 0.0066858442560617 |
| 0.0064936997583802 |
| 0.0060965716145545 |
| 0.00587667524 |
| 0.0057692365757952 |
| 0.0054719105743607 |
| 0.0050276337770239 |
| 0.004290027233 |
| 0.004256710896 |
| 0.0041107557948336 |

| |
|---|
| 0.0039773466002277 |
| 0.0036932917375787 |
| 0.003482012974 |
| 0.0032793348260563 |
| 0.00322728150453527 |
| 0.0030733316338348 |
| 0.00305152206576504 |
| 0.00277752069431282 |
| 0.0026651226091685 |
| 0.00260559392261993 |
| 0.0025796146869727 |
| 0.00240359614769158 |
| 0.0021259136113744 |
| 0.0019669057285564 |
| 0.0019625141024096 |
| 0.0019603376474116 |
| 0.0019547595494417 |
| 0.0019506693812476 |
| 0.001864859988 |
| 0.0017768830184622 |
| 0.0017763827698647 |
| 0.0016136452712842 |
| 0.0014904503036266 |
| 0.001427802194805 |
| 0.0014022853244258 |
| 0.0013773810516443 |
| 0.0013488984025387 |
| 0.0013344729462703 |
| 0.0013092516752977 |
| 0.0012792778398271 |
| 0.00126192916 |
| 0.0012004158365827 |
| 0.00119926538429875 |
| 0.0011905116223697 |
| 0.0011755268840673 |
| 0.0011601087717984 |
| 0.0011383156837023 |
| 0.00111610464597152 |
| 0.0011148083571841 |
| 0.0010552262107367 |
| 0.00103292239962957 |
| 0.001015197155 |
| 0.0009847982170337 |
| 0.0009775406338016 |

0.0009695454500359
0.0009405376230097
0.0009334693402671
0.0009334693402671
0.0009031281555567
0.0008928837167772
0.000880735638997917
0.000830877903112099
0.0008238834756883
0.0008117124697975
0.000808013259323099
0.00076417975760213
0.000744069763981
0.0007405433669953
0.0007372119781379
0.0007134915545023
0.0007034841404911
0.0006999959452086
0.0006764270581646
0.0006764270581646
0.0006470171860704
0.0006446813839547
0.000643906526522
0.0006358414346747
0.000635116691398071
0.0006036327555475
0.0006003496873196
0.0005866703908312
0.000570843732098505
0.000565832520137542
0.0005611699337287
0.000554475064505055
0.000550459774
0.0005409017787805
0.0005340385107571
0.0005299334074865
0.000529676781139
0.0004952786354672
0.0004928254280913
0.0004764978784115
0.000471783364843
0.000468488369913916
0.0004650436024881
0.0004425693763456

0.0004331913009478
0.0004305207149672
0.0004242503040242
0.000423701249374913
0.000423167638283
0.000421517353979366
0.00041693564361
0.000413331825530776
0.0004092383701895
0.000407132512367
0.0003971158852707
0.0003956199154091
0.0003878355912642
0.0003827519362042
0.000378924416842138
0.0003758901657417
0.0003710833861286
0.0003702716834976
0.0003566137469857
0.000347055104553588
0.00034493300317
0.0003365139636095
0.000336031506314006
0.0003344793103069
0.0003306976728804
0.0003258699696267
0.0003244490249917
0.000324301506253435
0.0003241212987038
0.0003211811930853
0.0003180967392484
0.0003171444895657
0.0003122920542256
0.0003111564467557
0.0003045899618635
0.0003045899618635
0.0003001748436598
0.0002988366831873
0.000298701083136589
0.0002949056991388
0.0002941301417446
0.0002919251811239
0.0002905415268878
0.0002861806784542

0.0002807810430117
0.000278528788656
0.0002718063064771
0.0002705708232658
0.0002701929310545
0.0002693465208981
0.0002683292521179
0.0002642293195955
0.0002607721135447
0.0002570422821025
0.0002565757806831
0.000253084953735005
0.000253084953735
0.0002492541392988
0.000249248740773474
0.0002475232077759
0.0002473064244445
0.0002465728262705
0.0002441927796849
0.0002359922758895
0.000232298437362
0.000231556980492
0.0002312081187511
0.0002262985900185
0.000226098547066
0.0002204881187089
0.0002204651152536
0.0002188440482297
0.0002148937946515
0.000210405331717
0.000208194702817
0.0002038547298578
0.0001975406453047
0.0001954122612475
0.0001927641875598
0.0001898137153012
0.000184862053162962
0.000184698168364
0.0001825076779576
0.0001771594676145
0.0001762270493639
0.000174391350933
0.000172466501585
0.0001715054420523

0.0001710505204554
0.000168517542365
0.000167206688535
0.000167206688535
0.000165683550684
0.0001646172044205
0.0001617542965176
0.00016087994896881
0.0001578329802384
0.0001538490113494
0.0001532727978238
0.00014782842993
0.00014782842993
0.0001451199894289
0.0001441995666629
0.0001430903392271
0.0001393389071125
0.0001377906970335
0.000137588063655515
0.000134795247098
0.0001332094206615
0.000131927263117211
0.0001296288787423
0.0001290294388406
0.0001275548166824
0.0001252642700304
0.0001236944620428
0.0001215800267942
0.0001212143725783
0.0001208777594915
0.00012074154385086
0.000119873477206501
0.0001166659908681
0.000116260900622
0.0001127378430274
0.0001127378430274
0.0001123124172046
0.000111055188653891
0.000109961049
0.0001088882581435
0.0001062956805687
0.000105168871234073
0.000104065701256
0.0001037031029938

0.0001005499681055
0.0000986675974077863
0.0000965359973228934
0.0000964534879234721
0.0000941860460735532
0.0000935936810039082
0.0000931751046130142
0.0000907402151196428
0.0000907402151196428
0.0000896469595157916
0.0000891864443127995
0.0000885797338072703
0.0000870596447715761
0.0000853850548830737
0.0000845533822705762
0.0000839965867147022
0.0000813190998886416
0.0000811712469797531
0.0000805851729943398
0.00008043997448444
0.0000794380530940645
0.0000777773272454081
0.0000772390758457859
0.0000761846174724646
0.0000729480160765755
0.0000708637870458162
0.0000706954645112656
0.0000701952607529312
0.00006966945355628
0.00006966945355628
0.0000688953485167658
0.0000678324469267456
0.0000672352196368437
0.0000671543108286165
0.0000666729179194507
0.0000652692775421992
0.0000650410632850586
0.0000647017186070496
0.0000630567596594127
0.0000626705007947303
0.000062421960065526
0.0000617998142841421
0.0000595255811184856
0.0000595255811184856

0.000058282226943687
0.0000577693916134371
0.000056944114590388
0.000056637089551366
0.0000561562086023449
0.0000551162788134126
0.0000537982688974368
0.0000537234486628932
0.000053530198847559
0.000053530198847559
0.0000530972714544056
0.0000509636824644569
0.0000492109632262613
0.0000491676055494099
0.0000485685224530725
0.0000475227039039005
0.0000470930230367766
0.0000467380505013235
0.0000452780789440813
0.0000449783104306346
0.0000432598699988994
0.0000427626301138546
0.0000425182722274897
0.0000422766911352881
0.0000406595499443208
0.0000402925864971699
0.00004021998724222
0.0000397190265470322
0.0000397190265470322
0.0000391615665253195
0.0000391615665253195
0.0000390246379710351
0.000038888663622704
0.0000387536335406807
0.0000383870901451971
0.0000382227618483426
0.0000382227618483426
0.0000379627430602587
0.0000371107114205022
0.0000369571074825035
0.0000364740080382877
0.0000363848295345266
0.0000357725848067822
0.0000355447339481403

0.0000346616349680623
0.0000346078959991195
0.0000344832239738198
0.0000327784036996066
0.0000327784036996066
0.00003242134
0.0000323508593035248
0.0000318887041706173
0.0000318583628726433
0.0000318583628726433
0.0000308316200544642
0.0000307466844620277
0.0000301242819425534
0.0000297627905592428
0.0000292173991092043
0.0000291792064306302
0.0000291792064306302
0.000028508420075903
0.000028508420075903
0.000028472057295194
0.000027867781422512
0.000027867781422512
0.000027867781422512
0.000027867781422512
0.000027867781422512
0.0000273555060287158
0.0000272220645358928
0.0000270570823265844
0.0000270570823265844
0.0000268617243314466
0.0000268617243314466
0.0000265739201421811
0.0000265739201421811
0.0000265739201421811
0.0000264793510313548
0.0000263854526234422
0.0000261996395767982
0.0000260164253140234
0.0000260164253140234
0.0000259257757484693
0.0000258357556937871
0.000025632324546592
0.000025569407696944
0.0000254818412322284

| |
|---|
| 0.0000247199257136568 |
| 0.0000243957299665924 |
| 0.0000243957299665924 |
| 0.0000243160053588585 |
| 0.0000242368001296765 |
| 0.0000240799276369278 |
| 0.0000235465115183883 |
| 0.0000235465115183883 |
| 0.0000233494695809959 |
| 0.0000228242258889899 |
| 0.000022547568605487 |
| 0.0000224117398789479 |
| 0.0000218844048229726 |
| 0.0000217564258473997 |
| 0.0000206686045550297 |
| 0.0000206304001103808 |
| 0.00002010999362111 |
| 0.00002010999362111 |
| 0.0000197890894675816 |
| 0.0000197890894675816 |
| 0.0000197365985140867 |
| 0.0000196843852905045 |
| 0.0000196843852905045 |
| 0.0000195807832626597 |
| 0.0000194782660728029 |
| 0.0000193768167703403 |
| 0.0000192764187559862 |
| 0.0000192266088883997 |
| 0.0000192266088883997 |
| 0.0000190299172373675 |
| 0.0000189813715301293 |
| 0.0000188372092147106 |
| 0.0000187423114352914 |
| 0.0000183268414773662 |
| 0.0000182370040191438 |
| 0.0000177667087865585 |
| 0.000017715946761454 |
| 0.0000174664263845321 |
| 0.0000173848075696511 |
| 0.0000173039479995597 |
| 0.0000172238371291914 |
| 0.0000169878941548189 |
| 0.0000169106764541152 |
| 0.000016461720442059 |

0.0000161754296517624
0.0000160359862926954
0.0000159671623171903
0.0000158989265807921
0.000015853759175733
0.0000153733422310138
0.0000152473312291203
0.0000150621409712767
0.0000149112177150515
0.0000147049360470567
0.0000146470425980525
0.0000146039207847118
0.0000145896032153151
0.0000145896032153151
0.0000145326125777552
0.0000145326125777552
0.0000144199566662998
0.0000143090339227129
0.0000141592723878415
0.0000141458130034424
0.0000141458130034424
0.0000141458130034424
0.0000140922303784293
0.0000140922303784293
0.0000140523090459125
0.0000140390521505862
0.0000140390521505862
0.0000139862737590426
0.000013933890711256
0.000013933890711256
0.000013933890711256
0.000013933890711256
0.000013933890711256
0.000013933890711256
0.000013933890711256
0.000013933890711256
0.0000138818985817363
0.0000138818985817363
0.0000138818985817363
0.0000137790697033531
0.000013728224427695
0.000013728224427695
0.0000136777530143579
0.0000135779154011144

0.0000135531833147736
0.0000135285411632922
0.0000135285411632922
0.0000135285411632922
0.000013479524709802
0.000013479524709802
0.000013479524709802
0.000013479524709802
0.000013479524709802
0.000013479524709802
0.000013479524709802
0.0000134308621657233
0.0000134308621657233
0.0000134308621657233
0.0000133345835838901
0.0000132869600710905
0.0000132396755156774
0.0000131927263117211
0.0000130538555084398
0.0000130538555084398
0.0000129628878742346
0.0000129628878742346
0.0000129628878742346
0.0000129628878742346
0.0000129628878742346
0.0000129628878742346
0.0000129628878742346
0.0000129628878742346
0.0000129628878742346
0.0000129628878742346
0.0000129628878742346
0.0000129628878742346
0.0000129628878742346
0.0000129628878742346
0.0000128731793076309
0.0000128731793076309
0.0000128287890341563
0.000012784703848472
0.000012784703848472
0.0000127409206161142
0.0000126974362454107
0.0000126974362454107
0.0000126542476867529
0.0000126113519318825

0.0000126113519318825
0.0000124011627330178
0.0000124011627330178
0.0000124011627330178
0.0000122379895391623
0.0000121978649832962
0.0000120399638184639
0.0000120399638184639
0.0000120399638184639
0.0000119625363984095
0.000011924194935594
0.0000117361161511209
0.0000116260900622042
0.0000115538783226874
0.0000114121129444949
0.0000112058699394739
0.0000111890190072341
0.0000111722186783944
0.0000108782129236998
0.0000108570490853269
0.0000108149674997248
0.0000107628220440849
0.0000106194542908811
0.0000106194542908811
0.0000106194542908811
0.0000105392317844344
0.000010450418033442
9.89454473379083E-06
9.86829925704338E-06
9.84219264525226E-06
9.84219264525226E-06
9.79039163132988E-06
9.79039163132988E-06
9.79039163132988E-06
9.76469506536838E-06
9.73913303640145E-06
9.73913303640145E-06
9.73913303640145E-06
9.73913303640145E-06
0.0000097137044906145
0.0000097137044906145
0.0000097137044906145
9.68840838517019E-06
9.56387871440965E-06

9.56387871440965E-06
9.53935594847526E-06
9.51495861868377E-06
9.49068576506468E-06
9.49068576506468E-06
9.44250969519125E-06
9.41860460735532E-06
9.39482025228625E-06
9.34761010026471E-06
9.30087204976338E-06
9.16342073868314E-06
9.07402151196427E-06
0.0000089003560284817
8.85797338072703E-06
0.0000088369330639082
8.75376192918906E-06
8.75376192918906E-06
8.55252602277093E-06
8.41707877806641E-06
0.0000080877148258812
8.01799314634774E-06
7.94946329039605E-06
7.78315652699865E-06
7.71856601640115E-06
0.0000076708223090832
7.54634649068023E-06
7.36702736614921E-06
7.26630628887764E-06
7.25214194913324E-06
7.25214194913324E-06
7.14764422652325E-06
7.11347766712304E-06
7.11347766712304E-06
7.11347766712304E-06
7.07963619392075E-06
7.07963619392075E-06
7.07963619392075E-06
7.07963619392075E-06
6.90231692004704E-06
6.87680003679363E-06
6.87680003679363E-06
6.87680003679363E-06
6.86411221384751E-06
6.85147112321428E-06

| |
|---|
| 6.75199422850336E-06 |
| 6.64348003554527E-06 |
| 6.03952730504115E-06 |
| 5.61138585204427E-06 |
| 5.51162788134126E-06 |
| 5.26216240439229E-06 |
| 5.16715113875743E-06 |
| 0.0000043872037970582 |
| 4.29601480358585E-06 |
| 3.62960860478571E-06 |
| 3.61198914553917E-06 |
| 3.57382211326162E-06 |
| 3.56355250948788E-06 |
| 3.54318935229081E-06 |
| 3.53981809696037E-06 |
| 3.53981809696037E-06 |
| 3.53981809696037E-06 |
| 3.53981809696037E-06 |
| 3.52974271338268E-06 |
| 3.50976303764656E-06 |
| 2.56046030275661E-06 |
| 0.010308547969 |
| 0.008031538621 |
| 0.0063003712973163 |
| 0.0052812693864861 |
| 0.0051824848572932 |
| 0.00507702602 |
| 0.0035575774932906 |
| 0.00294185326849832 |
| 0.0022453736169767 |
| 0.00170173536307392 |
| 0.001573664772 |
| 0.0015177157649234 |
| 0.0014073364365652 |
| 0.001402448673658 |
| 0.0013733677981508 |
| 0.001124499697 |
| 0.001084345619734 |
| 0.001010798699439 |
| 0.0009605543196423 |
| 0.0009444331282637 |
| 0.0009213008979261 |
| 0.0009032361744374 |
| 0.000878526213379508 |

0.000806138286
0.00080006468145556
0.000725516272140771
0.0007090338800778
0.0007055340453245
0.0006649991443677
0.000597326955798278
0.000586518318116286
0.0005722365825628
0.0005682701773696
0.0005496574848644
0.0004747690861001
0.000472087974
0.0004714546172756
0.0004563967511725
0.0004376420611874
0.000417415059446197
0.0004132195941102
0.0004101243162517
0.000407357399364
0.000401965904372456
0.000354553115
0.000352404308477721
0.0003445904657178
0.0003301031788708
0.0002900889124665
0.0002814243690834
0.0002717200804943
0.000256222737500295
0.0002438330196015
0.0002398098931053
0.00023850363
0.0002357879849077
0.0002311277513961
0.0002220694112403
0.0002212185705842
0.0002200121547286
0.0002050621581258
0.000205062158
0.0002005552975077
0.0001850311426797
0.0001772584700194
0.0001748236744312
0.0001696640786973

0.0001672500856472
0.000162881926705084
0.000160976131930932
0.000157032065754802
0.000156956319788644
0.0001539349871221
0.0001504164731308
0.0001449666009159
0.0001368409791659
0.0001344845102446
0.0001316144139894
0.000129466859448776
0.0001247899628937
0.0001238723896371
0.0001225210544774
0.0001225210544774
0.0001200214345188
0.0001197628790804
0.000119649467263139
0.0001178636543189
0.0001146319089585
0.0001142698788125
0.000114048853901886
0.00011252530051
0.0001119738410613
0.0001119664544368
0.0001104180851447
0.0001075971851592
0.0001037655669713
0.0001037655669713
0.0001036548752663
0.0001031674945392
0.0001017127492727
0.0001014369279302
0.0001005296026452
0.0001002776487538
0.0000982720957787923
0.000096055431964233
0.0000946440729811804
0.0000944923997873003
0.0000944134587682632
0.0000922261587079385
0.0000915578532100549
0.0000914050020360815

0.0000883928903830407
0.0000872883160137311
0.0000861720971388752
0.0000859522703604597
0.000084540645482277
0.0000842332249532505
0.0000832814371006714
0.0000830255801510534
0.0000827619022466435
0.0000800461933035275
0.0000797663115087599
0.0000796702901535473
0.0000782351934550314
0.0000779937268085653
0.0000776343087126733
0.0000770425837987047
0.000074616046
0.0000734592078080673
0.0000730345881097548
0.0000708692998404752
0.0000707181925913484
0.0000688179942428517
0.0000674222519748006
0.0000673096546430084
0.0000669334917110181
0.0000664299881334783
0.0000662210887997252
0.0000653268624181231
0.0000649613560050775
0.0000647947884255773
0.000063813049207008
0.0000623436697844782
0.0000614841058052923
0.0000614118055868007
0.0000611652048415719
0.0000610385688067033
0.0000607451141489787
0.0000603232730784997
0.0000598247336315699
0.0000594867407861938
0.0000591110350549126
0.000059092185872816
0.0000589995648918242
0.0000589043531141612

0.0000586991114656798
0.0000574792477968831
0.0000574317442863071
0.0000573015135736398
0.0000554165953639806
0.0000549347119260329
0.0000517221556730485
0.0000513617225324698
0.0000502884925094033
0.0000500911185497446
0.0000489728052053782
0.0000489728052053782
0.0000488106435987379
0.0000484113153065625
0.0000476432267835127
0.0000466809744199048
0.0000459453954290457
0.0000459453954290457
0.0000451249419392413
0.0000451249419392413
0.0000448049068900268
0.0000435689094585778
0.0000433298482269807
0.0000415966542979015
0.0000407580120741534
0.0000399209596934836
0.0000398831557543799
0.0000394843241968362
0.0000389968634042826
0.0000382878295242047
0.0000382878295242047
0.0000380113831016473
0.0000379428941230858
0.0000369443969093204
0.0000356920444717163
0.0000354915273679426
0.0000350971770638543
0.0000350323024297252
0.000034807117749277
0.0000342411483549799
0.0000341486047107772
0.000034102520223988
0.0000329036034973634
0.0000320035049214477

| |
|---|
| 0.0000318261555238981 |
| 0.0000317861226238681 |
| 0.0000316666259222746 |
| 0.0000315480243270601 |
| 0.0000315480243270601 |
| 0.000031430307818377 |
| 0.0000311206496132699 |
| 0.0000306302636193638 |
| 0.0000306302636193638 |
| 0.0000300832946261609 |
| 0.0000300832946261609 |
| 0.0000297643904428447 |
| 0.0000296595862511445 |
| 0.0000295670446403765 |
| 0.0000293836831232269 |
| 0.0000288469948470036 |
| 0.0000286507567868199 |
| 0.0000281716471415553 |
| 0.0000279535038561672 |
| 0.0000278549024316304 |
| 0.0000271970744775255 |
| 0.0000268600844876436 |
| 0.000026581312222274 |
| 0.0000259979089361884 |
| 0.0000258913601290729 |
| 0.0000254590921706715 |
| 0.0000253714532991718 |
| 0.0000242980456595915 |
| 0.0000240666357009287 |
| 0.0000240666357009287 |
| 0.0000240666357009287 |
| 0.0000240437369038774 |
| 0.0000238395919679011 |
| 0.0000236610182452951 |
| 0.0000230565396769846 |
| 0.0000228068298609884 |
| 0.0000227657364738515 |
| 0.000022608611 |
| 0.0000225771314445846 |
| 0.0000224024534450134 |
| 0.0000222839219453043 |
| 0.0000220121667996299 |
| 0.0000220121667996299 |
| 0.0000220121667996299 |

0.000021935735664909
0.0000209883450880192
0.0000207812232614927
0.0000206453982728555
0.0000205949205264671
0.0000200078919128861
0.0000194623902387362
0.0000192137830641538
0.0000191439147621024
0.0000186356692374448
0.0000183915338325874
0.0000180757993461911
0.0000165379368363711
0.0000165325269780668
0.0000165163186182844
0.0000165163186182844
0.0000165163186182844
0.0000164518017486817
0.0000164090697960877
0.000016183136398319
0.000016124277364711
0.0000160291579359182
0.000015953262301752
0.000015893061311934
0.000015893061311934
0.00001577401216353
0.00001577401216353
0.0000153151318096819
0.0000153151318096819
0.0000153151318096819
0.0000153151318096819
0.0000152596422016758
0.0000152596422016758
0.0000152596422016758
0.0000152045532406589
0.0000152045532406589
0.0000150416473130804
0.0000150059189346645
0.0000146747778664199
0.0000146747778664199
0.0000146747778664199
0.0000146492565136087
0.000014498016263135
0.000014473062706744

0.0000144234974235018
0.0000143253783934099
0.0000142285852961571
0.0000141330914351091
0.0000140858235707776
0.0000140388708255417
0.0000140388708255417
0.0000140255131939284
0.0000136299716752832
0.0000130796933157221
0.0000126097642145584
0.0000121024748496049
0.0000120218684519387
0.0000120218684519387
0.0000119310516930949
0.0000118305091226475
0.0000118305091226475
0.000011764416892912
0.000011764416892912
0.0000116558890617966
0.0000116344233360843
0.0000115072711684768
0.0000115072711684768
0.0000113828682369257
0.0000110252912242474
0.0000110252912242474
0.0000109965045630875
0.0000109678678324545
0.0000109678678324545
0.0000109110395017163
0.000010854797030058
0.0000108268926675129
0.0000107715121423594
0.0000107440337950574
0.0000107440337950574
0.0000107440337950574
0.0000107440337950574
0.0000106624335383861
0.000010582063436338
0.0000103735498710899
0.0000103735498710899
0.0000103480620335688
0.0000100277648753869
0.0000100277648753869

0.0000100182237099489
9.98023992337092E-06
9.98023992337092E-06
9.79456104107564E-06
9.57195738105119E-06
9.40102957067528E-06
9.15582859051679E-06
9.07685613720372E-06
9.07685613720372E-06
8.84802783122379E-06
8.81100679427307E-06
8.68383762404645E-06
8.61280418744893E-06
8.54292342325056E-06
8.52563005599701E-06
8.52563005599701E-06
8.45715109972395E-06
8.38976344155881E-06
8.25815930914221E-06
8.25815930914221E-06
8.22590087434087E-06
8.17798300516995E-06
8.16213420089637E-06
8.16213420089637E-06
8.06831656640331E-06
8.04519818082622E-06
8.01457896795914E-06
8.01457896795914E-06
8.00696054688693E-06
0.000007976631150876
7.94653065596703E-06
7.94653065596703E-06
7.94653065596703E-06
7.94653065596703E-06
7.91665648056866E-06
7.88700608176503E-06
7.88700608176503E-06
7.88700608176503E-06
7.82836663134298E-06
7.82836663134298E-06
7.79937268085653E-06
7.77059270786444E-06
7.74202435232082E-06
7.74202435232082E-06

0.0000077136652887592
7.65756590484095E-06
7.64366832606629E-06
7.62982110083791E-06
7.62982110083791E-06
7.62982110083791E-06
7.60227662032947E-06
7.60227662032947E-06
0.0000075749303015513
7.52082365654022E-06
7.46748448167114E-06
7.38887938186408E-06
7.33738893320997E-06
7.33738893320997E-06
7.33738893320997E-06
7.33738893320997E-06
7.33738893320997E-06
7.33738893320997E-06
7.33738893320997E-06
7.33738893320997E-06
7.33738893320997E-06
7.33738893320997E-06
7.33738893320997E-06
7.31191188830299E-06
7.31191188830299E-06
7.23653135337204E-06
7.23653135337204E-06
7.23653135337204E-06
0.0000072117487117509
0.0000072117487117509
0.0000072117487117509
0.0000072117487117509
0.0000072117487117509
0.0000072117487117509
7.18713523491898E-06
7.16268919670497E-06
7.16268919670497E-06
7.15052843406201E-06
7.01943541277087E-06
6.97294908553398E-06
6.94993605224839E-06
6.90436270108611E-06
6.90436270108611E-06
6.81498583764162E-06

6.81498583764162E-06
6.72789336687304E-06
6.70646695487663E-06
6.58072069947269E-06
6.58072069947269E-06
6.53984665786106E-06
6.53984665786106E-06
6.51959945458595E-06
6.51959945458595E-06
6.49947723404711E-06
6.47947884255773E-06
6.43984900254209E-06
6.36202605387088E-06
6.15739948488673E-06
6.13944788288998E-06
6.12160065067227E-06
0.0000060862156758129
6.03966718116041E-06
6.01093422596935E-06
0.000005982473363157
5.96552584654749E-06
5.89868522081586E-06
5.81721166804216E-06
5.67609332569073E-06
0.0000056155483302167
5.58575762289459E-06
5.57098048632609E-06
5.57098048632609E-06
5.54165953639806E-06
5.51264561212372E-06
5.51264561212372E-06
5.49825228154377E-06
5.49825228154377E-06
5.49825228154377E-06
5.49825228154377E-06
5.48393391622724E-06
5.48393391622724E-06
5.48393391622724E-06
5.48393391622724E-06
5.48393391622724E-06
5.37201689752873E-06
5.37201689752873E-06
5.30435925398303E-06
0.000005291031718169

0.000005291031718169
0.000005291031718169
0.000005291031718169
0.000005291031718169
5.27777098704577E-06
5.23838463639617E-06
5.07429065983437E-06
5.03787230581642E-06
5.01388243769348E-06
5.01388243769348E-06
4.90869609284676E-06
4.90869609284676E-06
4.69004593280905E-06
0.0000046589173093612
4.50927328443525E-06
4.49963808510953E-06
4.27146171162528E-06
4.22010145056365E-06
4.16996163134903E-06
4.16172060045704E-06
4.14533587368358E-06
4.11295043717043E-06
4.10493298992449E-06
0.0000040613898241683
4.02644478744027E-06
4.02644478744027E-06
4.02259909041311E-06
4.01110595015478E-06
4.00728948397957E-06
4.00728948397957E-06
4.00728948397957E-06
4.00728948397957E-06
3.96578271907959E-06
3.90692138002089E-06
3.89968634042826E-06
3.89247804774725E-06
3.88529635393222E-06
3.88529635393222E-06
3.83575705615895E-06
3.81491055041895E-06
3.72713384748896E-06
3.72054880535558E-06
0.0000036559559441515
3.26485368035854E-06

3.17621511890085E-06
3.16190784359048E-06
3.14772888465061E-06
3.11974907234261E-06
2.56183774188718E-06
2.01707914160082E-06
2.00364474198978E-06
2.00364474198978E-06
2.00364474198978E-06
1.97175152044125E-06
0.0262490517530054
0.024502323086024
0.0154992371679793
0.0110797265269075
0.005583639986
0.004828327708
0.003536465437
0.0033029497346492
0.00317092549109561
0.0030690431232126
0.002464141808404
0.00221737203890158
0.0021385505644097
0.00193656598625418
0.0018105040936091
0.001755363485
0.00170278760397269
0.001270177866976
0.0011740484965889
0.001116793635
0.001055185572321
0.0010335761074412
0.000910598204906
0.0009104865891914
0.000846831832747263
0.0008186392578256
0.000809465317559795
0.0007864347236861
0.0007744589629684
0.0007188052019932
0.0007129939659074
0.0007059693948147
0.00070008631652452
0.0006780034574356

| |
|---|
| 0.0006373013334612 |
| 0.000636769043 |
| 0.000594844582668 |
| 0.0005710507993613 |
| 0.000569561749037 |
| 0.000566851896424745 |
| 0.0005490873070781 |
| 0.0005252139459008 |
| 0.00050106826501423 |
| 0.000483977016418986 |
| 0.0004816555325247 |
| 0.000481315826 |
| 0.0004671339776635 |
| 0.0004630141616442 |
| 0.0004437521910263 |
| 0.000440281988164 |
| 0.0004108085964936 |
| 0.0004059602839061 |
| 0.000374527003 |
| 0.0003657278272777 |
| 0.0003594026009966 |
| 0.0003583517161983 |
| 0.0003578321776464 |
| 0.000334007025503698 |
| 0.0003327801861079 |
| 0.0003232363015252 |
| 0.0003214169602408 |
| 0.000319324445 |
| 0.0003092332848526 |
| 0.0002951344275545 |
| 0.000287117809810658 |
| 0.000285103024828994 |
| 0.0002735076397521 |
| 0.0002704459870683 |
| 0.0002635619073975 |
| 0.0002343385159915 |
| 0.000234158462807212 |
| 0.0002097636903444 |
| 0.0001946764088731 |
| 0.0001916813871981 |
| 0.0001827016799938 |
| 0.000176096587367989 |
| 0.0001715897834619 |
| 0.0001680879511463 |

0.0001677212137984
0.0001635579212573
0.0001599283418674
0.000159599521
0.0001586252220448
0.0001568820877366
0.0001546166424263
0.000152791095794944
0.0001486251357504
0.0001393837010275
0.0001383197033097
0.000135729422
0.0001352229935341
0.0001317809536987
0.0001295599263892
0.000127412524294311
0.0001267124554795
0.0001235446440925
0.000120640685587011
0.0001203479029212
0.0001198008669988
0.0001193668058865
0.000118212243957547
0.0001180832918447
0.0001147917715146
0.0001101847438952
0.0001078207802989
0.0001020427738817
0.0001019801017445
0.0001010590135726
0.0001004428000752
0.0000994037366262212
0.0000987123248679864
0.0000976155212583421
0.0000975352453362547
0.0000964250880722648
0.0000958406935990996
0.0000958406935990996
0.0000953677954398935
0.0000939056677188327
0.0000936831424398781
0.0000910546790632149
0.0000908264718475176
0.0000901237525295484

0.000089260498
0.0000873547988533459
0.000087272154767392
0.000085695706885033
0.0000853503586131877
0.0000850199701282334
0.0000838606068992121
0.0000834869589709505
0.0000834056668979505
0.0000832551151526474
0.0000827609854146814
0.0000818515240364981
0.0000815786856230431
0.0000813462677152851
0.0000803542400602207
0.0000791261184872478
0.0000779461476924853
0.0000771293207267001
0.0000758330296220497
0.0000745367385173992
0.0000740342436509898
0.0000738538483204562
0.0000716200835319358
0.0000711048311324255
0.0000705969394814796
0.0000688750629087605
0.0000676956953931996
0.0000664601351506724
0.0000660004776454567
0.000065399977021718
0.0000650846666903574
0.0000650770141722281
0.0000644405641558738
0.0000638473612881598
0.0000633562277397894
0.0000621608272163971
0.0000616952030424915
0.0000610097007864638
0.0000610097007864638
0.0000601190482202381
0.0000599004334994372
0.0000599004334994372
0.0000599004334994372
0.0000599004334994372

| |
|---|
| 0.0000596834029432798 |
| 0.0000595908705356158 |
| 0.0000593607899543972 |
| 0.0000592844907385175 |
| 0.0000577986639029657 |
| 0.0000575965706725358 |
| 0.0000573958857573004 |
| 0.0000563854936363895 |
| 0.0000561566564057224 |
| 0.000055582457 |
| 0.0000554973388471049 |
| 0.0000554633643513307 |
| 0.000055215483393336 |
| 0.000054778306169319 |
| 0.0000546808936509385 |
| 0.0000505553530813664 |
| 0.0000491719976487917 |
| 0.0000489892021184988 |
| 0.000048627658191425 |
| 0.0000480952385761904 |
| 0.0000480952385761904 |
| 0.0000478855209661198 |
| 0.000047517170804842 |
| 0.000047517170804842 |
| 0.000046841571219939 |
| 0.0000464952961638777 |
| 0.0000449545163689733 |
| 0.0000448234536390346 |
| 0.000044587540725145 |
| 0.0000437326173779077 |
| 0.0000426751793065938 |
| 0.0000426278500873998 |
| 0.0000422374851598596 |
| 0.0000421294609011387 |
| 0.0000417028334489752 |
| 0.0000417028334489752 |
| 0.0000411173022461035 |
| 0.0000407990568726817 |
| 0.0000405064406860948 |
| 0.000040107246777884 |
| 0.0000400144596251301 |
| 0.0000395568900467981 |
| 0.0000387591040290476 |
| 0.0000385324426019771 |

0.0000378680901433224
0.0000372262581069948
0.0000360714289321428
0.0000359402600996623
0.0000350481259837132
0.0000349242810155729
0.0000349130657679764
0.0000338478476965998
0.000033617590229276
0.000033617590229276
0.000033617590229276
0.000033617590229276
0.0000328357858053393
0.0000323445538531323
0.0000322992533575396
0.0000305048503932319
0.0000304484643481427
0.0000300869757303109
0.0000299502167497186
0.000029503198589275
0.0000294153914506164
0.0000288150773393794
0.0000286481688010957
0.0000263561907397523
0.0000261470146227702
0.0000259411326178665
0.0000257384675192894
0.0000256716143569016
0.0000256051075839563
0.0000253424910959157
0.0000253181467240656
0.0000249585139580988
0.0000248956461647535
0.0000248643308865588
0.0000248643308865588
0.0000248643308865588
0.0000248643308865588
0.0000246780812169966
0.0000246780812169966
0.0000245859988243958
0.0000240476192880952
0.0000239601733997748
0.0000238733611773119
0.0000238733611773119

0.0000238158350058003
0.0000237016103774751
0.000023448568273801
0.0000232008721300637
0.0000228786377949239
0.0000226427755496154
0.0000219634922831269
0.0000219634922831269
0.000021818038691848
0.000021818038691848
0.000021818038691848
0.0000208514167244876
0.0000204628810091245
0.0000204628810091245
0.0000203995284363408
0.0000203365669288212
0.000019966811166479
0.0000198465291715002
0.0000192662213009885
0.0000192662213009885
0.0000188978423086178
0.0000188797927935189
0.0000188258505283945
0.0000188258505283945
0.0000188079382062933
0.0000186131290534974
0.0000185085609127474
0.0000183029102359391
0.0000182690786088856
0.0000180357144660714
0.0000180028625271532
0.0000180028625271532
0.0000178082369863191
0.0000178082369863191
0.0000177124937767153
0.0000174313430818468
0.0000171144095712677
0.000016808795114638
0.0000167802572621513
0.000016597097443169
0.0000163906658829305
0.000016165475183852
0.0000161496266787698
0.0000156882087736621

0.0000156882087736621
0.0000156732818385777
0.000015613857073313
0.0000154310250232742
0.0000153590855126762
0.0000153233667091583
0.0000149751083748593
0.0000149751083748593
0.0000143865669976814
0.0000142005338037458
0.0000142005338037458
0.0000140791617199532
0.0000140791617199532
0.000013959846790123
0.000013959846790123
0.0000135021468953649
0.0000135021468953649
0.0000132843703325364
0.0000129705663089332
0.0000129197013430158
0.0000129197013430158
0.0000129197013430158
0.0000129197013430158
0.0000128692337596447
0.0000126712455479578
0.0000126226967144407
0.0000125745642790041
0.0000125267066253575
0.0000123854279792069
0.0000123390406084983
0.0000123390406084983
0.0000122019401572927
0.0000121569145478562
0.000012067852902817
0.0000120238096440476
0.0000119366805886559
0.0000119366805886559
0.0000119366805886559
0.0000118935878789496
0.0000118935878789496
0.0000118508051887375
0.0000117661565802466
0.0000117661565802466
0.0000117661565802466

0.0000117242841369005
0.0000117242841369005
0.0000116827086612377
0.0000116620313007753
0.0000116004360650318
0.0000115597327805931
0.0000115597327805931
0.0000115597327805931
0.0000115193141345071
0.0000115193141345071
0.0000115193141345071
0.00001147917715146
0.00001147917715146
0.00001147917715146
0.00001147917715146
0.00001147917715146
0.00001147917715146
0.00001147917715146
0.0000114393188974619
0.0000114393188974619
0.0000113800478150917
0.0000113213877748077
0.0000113213877748077
0.0000112826158988666
0.0000112826158988666
0.0000112826158988666
0.0000112154734260681
0.0000112058634097586
0.0000110926728702661
0.0000110184743895285
0.0000110184743895285
0.0000109817461415634
0.0000109817461415634
0.0000109817461415634
0.0000109452619351131
0.000010909019345924
0.000010837249481806
0.0000108017175162919
0.0000107313480210718
0.0000106274962660291
0.000010593324252312
0.0000105593712899649
0.0000104921141479906
0.0000104921141479906

0.0000103601378693995
0.0000101369964383663
0.0000097471119599676
9.63311065049429E-06
9.61436919008477E-06
9.57710419322397E-06
0.0000094670225358306
9.41292526419728E-06
9.40396910314666E-06
9.40396910314666E-06
9.33292873220693E-06
9.32412408245957E-06
9.10089459245593E-06
0.000008808887279329
0.0000087156715409234
0.0000087156715409234
8.62440796457866E-06
8.62440796457866E-06
8.62440796457866E-06
8.62440796457866E-06
8.62440796457866E-06
8.62440796457866E-06
8.60188992811762E-06
8.60188992811762E-06
8.60188992811762E-06
8.60188992811762E-06
8.60188992811762E-06
8.57948917309648E-06
8.57948917309648E-06
8.57948917309648E-06
8.53503586131877E-06
8.53503586131877E-06
8.53503586131877E-06
8.51298150508798E-06
8.51298150508798E-06
8.49104083110579E-06
8.46921296264536E-06
0.000008404397557319
8.36173564078438E-06
0.0000082985487215845
0.0000082776980966559
0.0000082776980966559
8.21577018072081E-06
8.11459074499765E-06

7.91952846747367E-06
7.90053679249172E-06
7.84410438683106E-06
7.84410438683106E-06
7.57361802866447E-06
7.10026690187294E-06
7.05465490892729E-06
6.97992339506154E-06
6.82096033637484E-06
6.80686744311786E-06
6.79283266488463E-06
6.69618667168505E-06
6.68260414293924E-06
6.60225218931673E-06
6.52380958904762E-06
6.51091668472144E-06
6.43461687982235E-06
6.42207376699619E-06
6.40957946005651E-06
6.36008463797113E-06
6.29928076953928E-06
6.26931273543111E-06
6.26931273543111E-06
6.26931273543111E-06
6.26931273543111E-06
6.10097007864638E-06
6.08969286962855E-06
6.08969286962855E-06
6.07845727392813E-06
6.07845727392813E-06
6.07845727392813E-06
6.01190482202381E-06
6.00095417571775E-06
5.97917212789301E-06
5.84135433061888E-06
5.77986639029657E-06
5.71965944873098E-06
5.14769350385788E-06
0.0000048807760629171
4.71319576891137E-06
4.69974870537667E-06
4.67972136714353E-06
3.21416960240882E-06
3.19856683734859E-06

3.18619327124666E-06
3.17697574008587E-06
3.15567417861019E-06
3.13465636771555E-06
3.12573419589093E-06
0.006062095022
0.004896225154
0.0045761687161567
0.0040754947669515
0.0039000349109921
0.00346139931
0.00338546835092394
0.0025731664364199
0.00257219866481 8
0.0019556459498572
0.0018484334422732
0.00146969244027729
0.0013663694072739
0.00105181408066136
0.0010147526957257
0.000963255255
0.0009247972187631
0.0008442914018614
0.0008369270780395
0.00083463040292286
0.0008253607576839
0.00081021726
0.0008098174511782
0.000786361133683782
0.000768715445173
0.0006864818731678
0.000678302344891182
0.000634879773
0.0006161444807577
0.000608788823332848
0.0006021770817261
0.000595667059221
0.0005559447268976
0.0005377388928616
0.0005314823060843
0.000510744181
0.0005060062947183
0.0004923057632728
0.000447686758496407

0.0004216719122652
0.000414779198314671
0.0004018822557329
0.000368962923232053
0.0003610423292044
0.000315457325
0.0003123814370454
0.0002988999277013
0.000296292659264705
0.000295342449570116
0.0002925928819092
0.0002867369003403
0.000275824785858501
0.0002645842773439
0.000255793623249076
0.0002539651365126
0.0002501604153663
0.000246274340248529
0.0002415464010636
0.0002391670377379
0.0002230676604578
0.0002200869215547
0.000216732593
0.0002149061483359
0.000205887765
0.0002052007093788
0.0001967802257237
0.000188503416465702
0.0001869202357354
0.0001771969608845
0.0001765357781946
0.0001582768504806
0.0001518018884154
0.0001485605669546
0.000145382405572
0.0001411684228018
0.0001391517310475
0.0001355035583234
0.00012964446992
0.0001259809910718
0.0001250802076831
0.000117743772424836
0.000117226107278734
0.0001159597758729

0.0001105176203265
0.0001030753563315
0.000098611463
0.0000962036659094064
0.0000946610415289348
0.0000923281627803544
0.0000854440453800648
0.0000851949373760413
0.0000848486164923989
0.000080961007154929
0.000079677882808404
0.0000795152748843053
0.0000791043432311564
0.0000782950916379988
0.0000780290080640378
0.0000749278551794415
0.0000732377531829127
0.0000723910739553646
0.0000721527494320548
0.0000698669779318163
0.0000678788931939191
0.0000668998706959299
0.0000662627290702544
0.000065100225051478
0.0000645341361379869
0.0000641252216808913
0.0000628934377615974
0.0000628934377615974
0.0000625401038415884
0.0000617205258678579
0.0000615035275348217
0.0000607649480556918
0.0000605007526293627
0.0000602823383599426
0.0000538651862119487
0.0000535198965567439
0.0000526757341504861
0.0000515376781657534
0.0000506006294718306
0.0000506006294718306
0.0000504172938578022
0.0000503716673475237
0.0000496970468026908
0.0000489478450920974

0.0000488251687886085
0.0000485413015282096
0.0000473996164152184
0.0000473996164152184
0.0000461914459908827
0.0000457116797644073
0.0000457067729718178
0.0000434849159523544
0.0000430810312840663
0.0000430736226802341
0.0000427747876418933
0.0000421671912265255
0.0000421671912265255
0.000042008069750199
0.000042008069750199
0.0000419854360919284
0.0000412301425326027
0.00004112859
0.0000406282426416158
0.0000406282426416158
0.0000404805035774645
0.0000403338350862418
0.0000403338350862418
0.0000403338350862418
0.000040188225573295
0.0000394921440368628
0.0000390601350308868
0.0000387879389679538
0.000038386684426906
0.0000375361124213001
0.0000372312323873001
0.0000369838487833978
0.0000368126272612524
0.0000360496712558378
0.0000352282863411479
0.0000352282863411479
0.0000348501651997402
0.0000347336614159211
0.0000345718586453501
0.0000345289655204799
0.0000340224281289814
0.0000338362871848107
0.0000334155100285674
0.000032550112525739

0.0000324363009434811
0.0000319898393107229
0.0000319889036890883
0.0000312700519207942
0.0000308654486611905
0.0000303603776830983
0.0000300868607670344
0.0000300868607670344
0.0000299251034510826
0.0000296857026234739
0.0000288397370046703
0.0000286173225804697
0.0000284709424138177
0.0000280171270565505
0.0000279000964506334
0.0000278303462095068
0.0000278303462095068
0.0000274641574435922
0.0000271515572775676
0.0000265050916281017
0.0000257688390828767
0.0000254158412872208
0.0000253003147359153
0.0000249226980980658
0.0000246286249641653
0.0000246286249641653
0.000024200301051745
0.000024200301051745
0.0000237190450649206
0.0000228117591881203
0.0000226724738491253
0.0000222642769676054
0.0000220875763567514
0.0000218491432459327
0.0000215739117903153
0.0000213259357927255
0.0000209447572602121
0.0000207689150817215
0.0000203636679581757
0.0000203179332040344
0.0000202402517887322
0.0000202402517887322
0.0000201669175431209
0.0000198798329982701

0.0000198788187210763
0.0000197378341911396
0.0000195300675154434
0.0000195300675154434
0.0000194617805660887
0.0000193939694839769
0.0000192597551622884
0.0000191603639622271
0.0000191273857110012
0.0000190618809654156
0.0000190618809654156
0.0000189875656400144
0.0000189322083057869
0.000018678084704367
0.00001861561619365
0.0000185535641396712
0.0000185359108912034
0.0000184919243916989
0.000018430692853978
0.0000182494073504962
0.0000181897687643835
0.0000178399655189146
0.0000175033623959162
0.0000173939663809417
0.0000173939663809417
0.000017285929322675
0.000017285929322675
0.000017285929322675
0.000017285929322675
0.0000171792260552511
0.0000171792260552511
0.0000167652688009077
0.0000165656822675636
0.0000162750562628695
0.000015903054976861
0.000015903054976861
0.0000157233594403993
0.0000156130974527387
0.0000154042506694687
0.0000146475506365825
0.0000145708618897941
0.0000145328178639722
0.0000144198685023351
0.0000142354712069088

0.0000138115862081919
0.0000137433808442009
0.0000137095301524664
0.0000132525458140508
0.0000132525458140508
0.0000132210670829011
0.0000131897375400506
0.0000129443470741892
0.0000128844195414383
0.0000128844195414383
0.0000127955614756353
0.0000127079206436104
0.0000126501573679576
0.0000126501573679576
0.000012126512509589
0.0000119958388834081
0.0000118933103459431
0.000011267346643525
0.0000111768458672718
0.0000111321384838027
0.0000111321384838027
0.0000109138612586301
0.0000109138612586301
0.0000109138612586301
0.0000109138612586301
0.0000107039793113487
0.0000107039793113487
0.0000106629678963627
0.0000106425798124309
0.0000106425798124309
0.0000106324149797542
0.0000105919490806876
0.0000105417978066313
0.0000105020174375497
0.0000104233506402647
0.0000104233506402647
0.0000103458536094821
0.0000103075356331506
0.0000102884828870635
0.0000102884828870635
0.0000102884828870635
0.0000102695004463124
0.0000102317449299657
0.0000102317449299657

0.0000101201258943661
0.0000101201258943661
0.0000101201258943661
0.0000101201258943661
0.0000101201258943661
0.0000100834587715604
0.0000100470563933237
0.0000100109159027003
9.97503448369421E-06
9.93940936053816E-06
0.0000098689170955698
0.0000098689170955698
9.83404459699888E-06
9.69698474198845E-06
9.69698474198845E-06
9.69698474198845E-06
9.69698474198845E-06
9.69698474198845E-06
9.69698474198845E-06
9.66331465607876E-06
0.0000094984116755996
9.46610415289348E-06
9.40214398969826E-06
9.37048693922789E-06
9.37048693922789E-06
9.30780809682503E-06
0.0000091849327424115
0.0000091849327424115
9.15471914786409E-06
9.15471914786409E-06
9.09488438219178E-06
9.06525935163089E-06
9.06525935163089E-06
8.97753103532479E-06
8.89148441198302E-06
0.000008536916
8.16139185029526E-06
8.09021692136827E-06
0.000008020272682855
7.94396181051575E-06
7.94396181051575E-06
7.94396181051575E-06
7.90634835497353E-06
7.90634835497353E-06

| |
|---|
| 7.68794094185272E-06 |
| 7.36252545225048E-06 |
| 7.36252545225048E-06 |
| 7.36252545225048E-06 |
| 7.36252545225048E-06 |
| 7.34309926372212E-06 |
| 7.28543094489708E-06 |
| 7.28543094489708E-06 |
| 7.26640893198612E-06 |
| 7.26640893198612E-06 |
| 7.24748599205907E-06 |
| 7.24748599205907E-06 |
| 7.24748599205907E-06 |
| 7.15433064511744E-06 |
| 7.09957811467011E-06 |
| 7.09957811467011E-06 |
| 7.09957811467011E-06 |
| 7.09957811467011E-06 |
| 7.06353964708295E-06 |
| 7.06353964708295E-06 |
| 7.04565726822958E-06 |
| 7.01016277317552E-06 |
| 6.99254929887106E-06 |
| 6.97502411265835E-06 |
| 6.92297169390717E-06 |
| 6.80448562579629E-06 |
| 6.62627290702544E-06 |
| 6.61053354145055E-06 |
| 0.0000065176454823201 |
| 6.48726018869623E-06 |
| 6.29645841844046E-06 |
| 6.26809599313217E-06 |
| 6.05007526293627E-06 |
| 5.89625979014975E-06 |
| 5.82224816098469E-06 |
| 5.72640868508371E-06 |
| 5.64510065101558E-06 |
| 5.64510065101558E-06 |
| 0.0000054356144940443 |
| 5.42501875428983E-06 |
| 5.42501875428983E-06 |
| 5.41446424309471E-06 |
| 5.31620748987714E-06 |
| 5.30101832562035E-06 |

| |
|---|
| 5.29597454034383E-06 |
| 5.29597454034383E-06 |
| 5.24111981346645E-06 |
| 0.0000051633295379419 |
| 5.15376781657534E-06 |
| 5.13475022315624E-06 |
| 5.13475022315624E-06 |
| 5.07853033020198E-06 |
| 4.72501633438146E-06 |
| 4.28818893829073E-06 |
| 4.26192131845434E-06 |
| 4.17873066208811E-06 |
| 4.15999195956754E-06 |
| 3.97009218395247E-06 |
| 3.90327436318469E-06 |
| 0.0000032136658440539 |
| 0.0000032136658440539 |
| 3.15894962650475E-06 |
| 3.14467188807987E-06 |
| 2.71515572775676E-06 |
| 2.70197536014629E-06 |
| 2.66574197409069E-06 |
| 2.64798727017191E-06 |
| 2.64798727017191E-06 |
| 2.64798727017191E-06 |
| 2.64798727017191E-06 |
| 2.64798727017191E-06 |
| 2.62798358918856E-06 |
| 2.60583766006618E-06 |
| 0.0199272779964764 |
| 0.0179906772289908 |
| 0.0124927421098815 |
| 0.00573318274855665 |
| 0.0046401298987237 |
| 0.004492105105 |
| 0.0040027685721059 |
| 0.0033641120970544 |
| 0.003356949642 |
| 0.002656104558 |
| 0.0022380581793396 |
| 0.00199631289212846 |
| 0.00198574685961105 |
| 0.00194456888731373 |
| 0.0017310762966874 |

0.0016710009220095
0.00159468518921
0.00145736679196006
0.0014446482100245
0.0011830389252382
0.0011055736583243
0.001032399707
0.000953410942207028
0.0008732839281377
0.000865961733
0.0008340357185707
0.000830854662178
0.0008293199775221
0.0007712977361856
0.0006486219539185
0.0006160662690164
0.0005808450113535
0.0005784282298096
0.0005711153248713
0.0005638251345485
0.0005548904576875
0.000530752739
0.0005178977605083
0.0004706906618692
0.000467321807333684
0.0004593417181622
0.0004572131112032
0.0004476116358679
0.00044391236615
0.0004327874229885
0.000429354403571864
0.000428637292
0.0004131630212926
0.000399744763
0.000396959295
0.000386761206107
0.000385536026868357
0.0003774995186881
0.0003655918900909
0.0003483143905592
0.000336606343817786
0.0003236861003177
0.0003158981640694
0.0003096127916082

0.00030720664080289
0.0003062278121081
0.0003031346018848
0.0002903857712338
0.000286504664887496
0.0002804446367231
0.0002750860007073
0.0002531194623337
0.0002478978127755
0.0002406514171332
0.000231270290171332
0.0002213208123219
0.0002093359307882
0.0002000307361514
0.000198141255161898
0.000198079886938897
0.0001978081724163
0.0001972799796996
0.000196035313
0.0001943155722613
0.00019074359483
0.0001900238076797
0.000189780095881668
0.0001842655104773
0.00017756494646
0.0001480398049427
0.0001480398049427
0.0001456587451429
0.0001431754390823
0.0001414006571713
0.0001311362214456
0.0001276248052681
0.0001271623965533
0.0001220759006912
0.000120153446
0.0001192147467688
0.000113735311203
0.0001128342391952
0.0001109780915375
0.000108996339902906
0.0001085402882715
0.000107125887892091
0.0001054803289382
0.0001028729500207

0.0001024890957295
0.0000996846479275179
0.000097720856
0.0000943313778068787
0.0000938285644471618
0.0000930456560146759
0.0000880355053061933
0.0000873530595286259
0.0000822170667371058
0.0000779681847480642
0.000077684664076253
0.0000774362056623631
0.0000774031979020637
0.0000769127398508409
0.0000747885462102485
0.0000726642266019373
0.0000718743980519163
0.0000708875951446773
0.0000707393464270516
0.0000707146985711537
0.0000704284042449547
0.0000698146490881359
0.0000697576575378598
0.000069678025508707
0.0000691196301823306
0.0000687927719059325
0.0000684148566252159
0.0000673212687635596
0.000066124446207763
0.0000650878617429676
0.0000635811982766952
0.0000630315049198361
0.0000627102229578363
0.000062399391617277
0.0000610379503456274
0.0000608167983516215
0.0000607678709193193
0.000060234819420027
0.000060234819420027
0.0000600092208454202
0.0000579933019910949
0.00005736649468574
0.0000573333348622222
0.0000568676556015162

0.0000565166206903957
0.0000561698929560988
0.0000561385830938044
0.000055657705482335
0.0000556013717723326
0.0000554890457687521
0.0000546375514602802
0.0000545508251881211
0.0000535915460520758
0.0000533548517007232
0.0000519787898320428
0.0000513787460821779
0.0000512592606261728
0.0000506238924667525
0.0000499401411918769
0.0000499401411918769
0.0000497307485663345
0.0000492241535045382
0.0000492241535045382
0.0000489810712650096
0.000048807392091988
0.0000474388215121456
0.0000471286344488358
0.0000470971839086631
0.0000468321869659545
0.0000462408714739601
0.0000457303255744053
0.0000456480397883965
0.0000446619148870444
0.0000446183847555755
0.0000445532484274652
0.000044467832
0.000044399734171781
0.0000443589755418803
0.0000443221631140448
0.0000442303988011792
0.0000436786189906334
0.0000434949764933212
0.0000434466246860364
0.0000433129560312858
0.0000431058971367425
0.0000428637291753001
0.0000422806960832431
0.000041401710505755

0.0000412418583416401
0.0000408280604318577
0.0000396923087507692
0.0000395323512601213
0.0000388423320381265
0.0000388423320381265
0.0000385618819873097
0.0000383886480161178
0.0000381487189660171
0.0000376777471269305
0.0000374925984924001
0.0000374374082100266
0.0000372182624058703
0.0000366814605442472
0.0000357086292973639
0.0000356947078044604
0.0000355791161341092
0.0000351369779950157
0.0000346806536054701
0.0000338472922434163
0.0000337226245003466
0.0000334149363205989
0.0000327201488207554
0.0000323522705012159
0.0000319013677764603
0.0000319013677764603
0.000031221457977303
0.0000311418114008303
0.0000305189751728137
0.0000299205638949153
0.0000299205638949153
0.0000298474084819693
0.0000284338278007581
0.0000282583103451978
0.000027744522884376
0.0000276189820568449
0.0000275773872043497
0.0000275442014195069
0.0000272490849757265
0.0000270558290539128
0.0000267710308533453
0.0000258174047956945
0.0000255745602006818
0.000025432479310678

0.0000252837513614928
0.0000250978414250112
0.0000244151801382509
0.0000236948565006317
0.0000236214978117753
0.0000236214978117753
0.0000235971457521755
0.0000233861878718878
0.0000230042526428244
0.0000230042526428244
0.00002286065556016
0.0000225648614956108
0.0000225509668764633
0.0000224404229211865
0.0000222766242137326
0.0000221956183075008
0.0000221956183075008
0.0000220353611356055
0.0000218617300664854
0.0000214168246826762
0.0000212675785176402
0.0000211937327588984
0.0000210475690846991
0.0000203266247582964
0.0000202783888191453
0.0000202559569731064
0.0000201445380678638
0.0000197078252960974
0.0000194636321255189
0.0000194388376896902
0.0000184484305880566
0.0000183407302721236
0.0000182748354328225
0.0000177435902167521
0.0000176071010612386
0.0000175396409039159
0.000017274891607253
0.000017274891607253
0.000017242358854697
0.00001714549167012
0.0000170497068004545
0.0000168924216823692
0.0000167074681602994
0.0000166467137306256

0.0000165863995504422
0.0000165863995504422
0.0000165863995504422
0.000016124338467865
0.0000160626185120072
0.0000160064555102169
0.0000159229435684245
0.0000158402985326022
0.0000155709057004151
0.0000155709057004151
0.0000154761537387493
0.0000153963826346929
0.0000152290295273521
0.0000152087916143589
0.0000149812641100768
0.0000145328453203874
0.0000141291551725989
0.0000139228901335828
0.0000131547306779369
0.0000130982726063578
0.0000125077767101695
0.0000123558603938517
0.000011983890774665
0.0000119682255579661
0.0000119682255579661
0.0000119682255579661
0.0000119526012426163
0.0000119526012426163
0.0000119214746768803
0.0000117833880976114
0.0000117682423545554
0.0000117380673741591
0.0000116707362037528
0.00001156021786849
0.0000115274693759447
0.0000115165944048353
0.0000115165944048353
0.000011473298937148
0.00001143032778008
0.00001143032778008
0.00001143032778008
0.0000114160755010525
0.000011345343930414
0.0000112616144549128

0.0000110776679393153
0.0000109780486233142
0.0000109780486233142
0.0000108996339902906
0.0000108996339902906
0.0000108377042517094
0.0000106709703401446
0.0000104876203343002
0.000010451703826306
0.0000103279399491141
0.0000102412668365146
0.0000101729917242712
0.0000101633123791482
0.0000101056209181502
9.65790353570054E-06
9.65790353570054E-06
0.0000094779426002527
9.19246240144991E-06
8.97616916847461E-06
8.92367695111511E-06
8.71970719223248E-06
8.71141061069848E-06
0.0000086211794273485
8.57274583506003E-06
0.0000083385178067797
8.24837166832802E-06
8.24837166832802E-06
8.24837166832802E-06
8.24837166832802E-06
8.11674871617385E-06
8.09521887873042E-06
0.0000080313092560036
7.96840082841088E-06
7.96840082841088E-06
0.0000079476497845869
0.0000079476497845869
7.88604009633429E-06
7.86571525072518E-06
7.78545285020757E-06
7.72632282856043E-06
7.62974379320342E-06
7.62974379320342E-06
7.61071700070167E-06
0.0000075169889588211

| |
|---|
| 7.48014097372884E-06 |
| 7.48014097372884E-06 |
| 7.26642266019373E-06 |
| 7.26642266019373E-06 |
| 7.26642266019373E-06 |
| 7.23198463810751E-06 |
| 7.23198463810751E-06 |
| 7.16407867906424E-06 |
| 7.01585636156636E-06 |
| 6.93613072109402E-06 |
| 6.93613072109402E-06 |
| 6.93613072109402E-06 |
| 6.93613072109402E-06 |
| 6.75986915743984E-06 |
| 6.75198565770214E-06 |
| 6.66353169712089E-06 |
| 6.64901419887008E-06 |
| 6.57736533896847E-06 |
| 6.46588457051137E-06 |
| 6.38472283950077E-06 |
| 6.25388835508477E-06 |
| 6.22836228016606E-06 |
| 6.19046149549973E-06 |
| 6.17793019692585E-06 |
| 6.11602708874022E-06 |
| 6.10379503456274E-06 |
| 5.98411277898307E-06 |
| 5.98411277898307E-06 |
| 5.98411277898307E-06 |
| 5.96073733844017E-06 |
| 0.0000059375438079404 |
| 0.0000059375438079404 |
| 5.89169404880573E-06 |
| 5.84654696797197E-06 |
| 5.81313812815499E-06 |
| 5.80208653475545E-06 |
| 5.80208653475545E-06 |
| 5.80208653475545E-06 |
| 5.78010893424501E-06 |
| 5.78010893424501E-06 |
| 5.78010893424501E-06 |
| 5.75829720241767E-06 |
| 5.71516389004001E-06 |
| 5.71516389004001E-06 |

| |
|---|
| 5.71516389004001E-06 |
| 5.71516389004001E-06 |
| 5.69383865164434E-06 |
| 5.69383865164434E-06 |
| 5.65166206903957E-06 |
| 5.61010573029663E-06 |
| 5.61010573029663E-06 |
| 5.56915605343315E-06 |
| 5.54890457687521E-06 |
| 0.0000055287998501474 |
| 0.0000055287998501474 |
| 5.50884028390139E-06 |
| 0.0000054498169951453 |
| 5.41116581078257E-06 |
| 5.41116581078257E-06 |
| 5.41116581078257E-06 |
| 5.37305900929818E-06 |
| 5.35420617066907E-06 |
| 5.33548517007232E-06 |
| 5.33548517007232E-06 |
| 5.31689462941005E-06 |
| 5.31689462941005E-06 |
| 5.31689462941005E-06 |
| 5.31689462941005E-06 |
| 5.31689462941005E-06 |
| 5.31689462941005E-06 |
| 0.0000052984331897246 |
| 0.0000052984331897246 |
| 0.0000052984331897246 |
| 5.28009951086742E-06 |
| 5.28009951086742E-06 |
| 5.28009951086742E-06 |
| 5.26189227117477E-06 |
| 5.24381016715012E-06 |
| 5.22585191315303E-06 |
| 5.22585191315303E-06 |
| 0.0000051727076564091 |
| 0.0000051727076564091 |
| 5.15523229270501E-06 |
| 5.15523229270501E-06 |
| 5.10350755398222E-06 |
| 5.10350755398222E-06 |
| 5.08649586213561E-06 |
| 5.08649586213561E-06 |

5.06959720478632E-06
5.06959720478632E-06
5.06959720478632E-06
5.05281045907511E-06
5.05281045907511E-06
5.03613451696595E-06
5.00311068406782E-06
5.00311068406782E-06
4.98676064915256E-06
4.97051712912275E-06
4.97051712912275E-06
4.95437908649573E-06
4.95437908649573E-06
4.93834549721904E-06
4.70971839086631E-06
4.66650996526203E-06
4.61011709559119E-06
0.0000045280378594679
4.50575420858469E-06
4.44883020011861E-06
4.44451094749714E-06
4.43589755418803E-06
4.43589755418803E-06
4.41450942711866E-06
4.37234601329709E-06
4.37234601329709E-06
4.35985359611624E-06
4.35985359611624E-06
4.35985359611624E-06
4.35570530534924E-06
4.33508170068376E-06
4.13536248954115E-06
4.11306943029834E-06
4.03690147788541E-06
4.03690147788541E-06
4.03690147788541E-06
4.03690147788541E-06
4.03690147788541E-06
3.99463025822168E-06
3.99463025822168E-06
3.99463025822168E-06
3.99463025822168E-06
3.99463025822168E-06
3.99463025822168E-06

| |
|---|
| 3.98420041420544E-06 |

| |
| ---: |
| 2.67710308533453E-06 |
| 2.67710308533453E-06 |
| 2.63094613558738E-06 |
| 2.60400812054724E-06 |
| 2.47718954324786E-06 |
| 2.29121435231334E-06 |
| 2.29121435231334E-06 |
| 1.47150314237288E-06 |
| 1.46584895162409E-06 |
| 1.46163674199299E-06 |
| 1.45190176844974E-06 |
| 1.45190176844974E-06 |
| 1.45190176844974E-06 |
| 1.45190176844974E-06 |
| 1.45190176844974E-06 |
| 1.45190176844974E-06 |
| 1.45190176844974E-06 |
| 1.45190176844974E-06 |
| 1.45190176844974E-06 |
| 1.45190176844974E-06 |
| 1.45190176844974E-06 |
| 1.45190176844974E-06 |
| 1.44776922072171E-06 |
| 0.00000142879097251 |

nt plant); inf (influent); T (treated).
