## Supplementary material for "Novel antibiotic resistance genes from the hospital effluent are disseminated into the marine environment in Norway": supplemantary table S1

**Supplementary Table S1:** Minimum inhibitory concentrations (MICs) of *Escherichia coli* (n= 66) a

| Strain | Month Year | Sample | Type | MALDI-TOF MS | AMP |
|---|---|---|---|---|---|
| 11-301 | Feb 23 | Hospital | Eff. | *E. coli* | >32 |
| 11-302 | Feb 23 | Hospital | Eff. | *E. coli* | >32 |
| 11-303 | Feb 23 | Hospital | Eff. | *E. coli* | >32 |
| 11-304 | Feb 23 | Hospital | Eff. | *E. coli* | >32 |
| 11-305 | Feb 23 | Hospital | Eff. | *E. coli* | >32 |
| 11-306 | Feb 23 | Hospital | Eff. | *E. coli* | >32 |
| 11-307 | Feb 23 | Hospital | Eff. | *E. coli* | >32 |
| 11-308 | Feb 23 | Hospital | Eff. | *E. coli* | >32 |
| 11-310 | Feb 23 | Hospital | Eff. | *E. coli* | >32 |
| 11-311 | Feb 23 | Hospital | Eff. | *E. coli* | >32 |
| 11-312 | Feb 23 | Hospital | Eff. | *E. coli* | >32 |
| 11-313 | Feb 23 | Hospital | Eff. | *E. coli* | >32 |
| 11-314 | Feb 23 | Hospital | Eff. | *E. coli* | >32 |
| 11-315 | Feb 23 | Hospital | Eff. | *E. coli* | >32 |
| 11-316 | Feb 23 | Hospital | Eff. | *E. coli* | >32 |
| 11-317 | Feb 23 | Hospital | Eff. | *E. coli* | >32 |
| 11-318 | Feb 23 | Hospital | Eff. | *E. coli* | >32 |
| 11-319 | Feb 23 | Hospital | Eff. | *E. coli* | >32 |
| 11-320 | Feb 23 | Hospital | Eff. | *E. coli* | >32 |
| 11-321 | Feb 23 | R-STP | Inf. | *E. coli* | >32 |
| 11-323 | Feb 23 | R-STP | Inf. | *E. coli* | >32 |
| 11-324 | Feb 23 | R-STP | Inf. | *E. coli* | >32 |
| 11-325 | Feb 23 | R-STP | Inf. | *E. coli* | >32 |
| 11-326 | Feb 23 | R-STP | Inf. | *E. coli* | >32 |
| 11-327 | Feb 23 | R-STP | Inf. | *E. coli* | >32 |
| 11-328 | Feb 23 | R-STP | Inf. | *E. coli* | >32 |
| 11-329 | Feb 23 | R-STP | Inf. | *E. coli* | >32 |
| 11-330 | Feb 23 | R-STP | Inf. | *E. coli* | >32 |
| 11-331 | Feb 23 | R-STP | Inf. | *E. coli* | >32 |
| 11-332 | Feb 23 | R-STP | Inf. | *E. coli* | >32 |
| 11-333 | Feb 23 | R-STP | Inf. | *E. coli* | >32 |
| 11-334 | Feb 23 | R-STP | Inf. | *E. coli* | >32 |
| 11-335 | Feb 23 | R-STP | Inf. | *E. coli* | >32 |
| 11-336 | Feb 23 | R-STP | T-Eff. | *E. coli* | >32 |
| 11-337 | Feb 23 | R-STP | T-Eff. | *E. coli* | >32 |
| 11-338 | Feb 23 | R-STP | T-Eff. | *E. coli* | >32 |
| 11-339 | Feb 23 | R-STP | T-Eff. | *E. coli* | >32 |
| 11-340 | Feb 23 | R-STP | T-Eff. | *E. coli* | >32 |
| 11-M301 | Feb 23 | R-STP | Inf. | *E. coli* | >32 |
| 11-M302 | Feb 23 | R-STP | T-Eff. | *E. coli* | >32 |
| K10-301 | Feb 23 | Hospital | Eff. | *K. oxytoca* | >32 |
| K10-304 | Feb 23 | Hospital | Eff. | *K. oxytoca* | >32 |

| | | | | | |
|---|---|---|---|---|---|
| K10-309 | Feb 23 | Hospital | Eff. | *K. oxytoca* | >32 |
| K10-311 | Feb 23 | Hospital | Eff. | *K. pneumoniae* | >32 |
| K10-313 | Feb 23 | Hospital | Eff. | *K. pneumoniae* | >32 |
| K10-314 | Feb 23 | Hospital | Eff. | *K. oxytoca* | >32 |
| K10-315 | Feb 23 | Hospital | Eff. | *K. oxytoca* | >32 |
| K10-316 | Feb 23 | R-STP | Inf. | *K. oxytoca* | >32 |
| K10-317 | Feb 23 | R-STP | Inf. | *K. oxytoca* | >32 |
| K10-318 | Feb 23 | R-STP | T-Eff. | *K. oxytoca* | >32 |
| K10-319 | Feb 23 | R-STP | T-Eff. | *K. oxytoca* | >32 |
| K10-320 | Feb 23 | R-STP | T-Eff. | *K. oxytoca* | >32 |
| K10-321 | Feb 23 | R-STP | T-Eff. | *K. oxytoca* | >32 |
| K10-T301 | Feb 23 | Hospital | Eff. | *K. pneumoniae* | >32 |
| K10-T302 | Feb 23 | Hospital | Eff. | *K. pneumoniae* | >32 |
| K10-T303 | Feb 23 | Hospital | Eff. | *K. pneumoniae* | >32 |
| K10-T304 | Feb 23 | Hospital | Eff. | *K. pneumoniae* | >32 |
| K10-T305 | Feb 23 | Hospital | Eff. | *K. pneumoniae* | >32 |
| K10-T306 | Feb 23 | Hospital | Eff. | *K. pneumoniae* | >32 |
| K10-T307 | Feb 23 | Hospital | Eff. | *K. pneumoniae* | >32 |
| K10-T309 | Feb 23 | R-STP | Inf. | *K. pneumoniae* | 32 |
| K10-T310 | Feb 23 | R-STP | Inf. | *K. pneumoniae* | 32 |
| K10-M302 | Feb 23 | Hospital | Eff. | *K. oxytoca* | >32 |
| K10-M304 | Feb 23 | Hospital | Eff. | *K. oxytoca* | >32 |
| K10-M306 | Feb 23 | Hospital | Eff. | *K. oxytoca* | >32 |
| 12-301 | Apr 23 | R-STP | Inf. | *E. coli* | >32 |
| 12-305 | Apr 23 | R-STP | Inf. | *E. coli* | >32 |
| 12-306 | Apr 23 | R-STP | Inf. | *E. coli* | >32 |
| 12-307 | Apr 23 | R-STP | Inf. | *E. coli* | >32 |
| 12-310 | Apr 23 | R-STP | Inf. | *E. coli* | >32 |
| 12-313 | Apr 23 | R-STP | Inf. | *E. coli* | >32 |
| 12-316 | Apr 23 | R-STP | T-Eff. | *E. coli* | >32 |
| 12-317 | Apr 23 | R-STP | T-Eff. | *E. coli* | >32 |
| 12-318 | Apr 23 | R-STP | T-Eff. | *E. coli* | >32 |
| 12-319 | Apr 23 | R-STP | T-Eff. | *E. coli* | >32 |
| 12-327 | Apr 23 | R-STP | T-Eff. | *E. coli* | >32 |
| 12-328 | Apr 23 | R-STP | T-Eff. | *E. coli* | >32 |
| 12-329 | Apr 23 | R-STP | T-Eff. | *E. coli* | >32 |
| 12-330 | Apr 23 | R-STP | T-Eff. | *E. coli* | >32 |
| 12-331 | Apr 23 | Hospital | Eff. | *E. coli* | >32 |
| 12-332 | Apr 23 | Hospital | Eff. | *E. coli* | >32 |
| 12-333 | Apr 23 | Hospital | Eff. | *E. coli* | >32 |
| 12-336 | Apr 23 | Hospital | Eff. | *E. coli* | >32 |
| 12-337 | Apr 23 | Hospital | Eff. | *E. coli* | >32 |
| 12-338 | Apr 23 | Hospital | Eff. | *E. coli* | >32 |
| 12-340 | Apr 23 | Hospital | Eff. | *E. coli* | >32 |
| 12-341 | Apr 23 | Hospital | Eff. | *E. coli* | >32 |
| 12-342 | Apr 23 | Hospital | Eff. | *E. coli* | >32 |

| | | | | | |
|---|---|---|---|---|---|
| 12-343 | Apr 23 | Hospital | Eff. | *E. coli* | >32 |
| 12-344 | Apr 23 | Hospital | Eff. | *E. coli* | >32 |
| 12-345 | Apr 23 | Hospital | Eff. | *E. coli* | >32 |
| K11-301 | Apr 23 | R-STP | Inf. | *K. pneumoniae* | >32 |
| K11-302 | Apr 23 | R-STP | Inf. | *K. pneumoniae* | >32 |
| K11-303 | Apr 23 | R-STP | Inf. | *K. oxytoca* | >32 |
| K11-304 | Apr 23 | R-STP | Inf. | *K. pneumoniae* | >32 |
| K11-305 | Apr 23 | R-STP | Inf. | *K. pneumoniae* | >32 |
| K11-306 | Apr 23 | R-STP | Inf. | *K. pneumoniae* | >32 |
| K11-307 | Apr 23 | R-STP | Inf. | *K. oxytoca* | >32 |
| K11-308 | Apr 23 | R-STP | Inf. | *K. oxytoca* | >32 |
| K11-309 | Apr 23 | R-STP | T-Eff. | *K. oxytoca* | >32 |
| K11-310 | Apr 23 | R-STP | T-Eff. | *K. pneumoniae* | >32 |
| K11-311 | Apr 23 | R-STP | T-Eff. | *K. pneumoniae* | >32 |
| K11-312 | Apr 23 | R-STP | T-Eff. | *K. pneumoniae* | >32 |
| K11-313 | Apr 23 | R-STP | T-Eff. | *K. oxytoca* | >32 |
| K11-314 | Apr 23 | R-STP | T-Eff. | *K. oxytoca* | >32 |
| K11-315 | Apr 23 | Hospital | Eff. | *K. pneumoniae* | >32 |
| K11-316 | Apr 23 | Hospital | Eff. | *K. pneumoniae* | >32 |
| K11-317 | Apr 23 | Hospital | Eff. | *K. pneumoniae* | >32 |
| K11-318 | Apr 23 | Hospital | Eff. | *K. pneumoniae* | >32 |
| K11-319 | Apr 23 | Hospital | Eff. | *K. pneumoniae* | >32 |
| K11-320 | Apr 23 | Hospital | Eff. | *K. pneumoniae* | >32 |
| K11-321 | Apr 23 | Hospital | Eff. | *K. pneumoniae* | >32 |
| K11-322 | Apr 23 | Hospital | Eff. | *K. pneumoniae* | >32 |
| K11-323 | Apr 23 | Hospital | Eff. | *K. pneumoniae* | >32 |
| K11-324 | Apr 23 | Hospital | Eff. | *K. pneumoniae* | >32 |
| K11-325 | Apr 23 | Hospital | Eff. | *K. oxytoca* | >32 |
| K11-327 | Apr 23 | Hospital | Eff. | *K. pneumoniae* | >32 |
| K11-328 | Apr 23 | Hospital | Eff. | *K. oxytoca* | >32 |
| K11-329 | Apr 23 | Hospital | Eff. | *K. pneumoniae* | >32 |
| K11-T301 | Apr 23 | R-STP | T-Eff. | *K. pneumoniae* | 32 |
| K11-T302 | Apr 23 | R-STP | T-Eff. | *K. pneumoniae* | >32 |

**Legend:** R-STP (receiving sewage treatment plant); Inf (influent); Eff (effluent); T-EFF (treated efflu (sulfamethoxazole); TET (tetracycline); TGC (tigecycline); GEN (gentamicin); AMK (amikacin); CH nd *Klebsiella* spp. (n=55) strains in this study.

| | | | | | | MIC (μg/mL |
|---|---|---|---|---|---|---|
| AZI | FOT | TAZ | MERO | NAL | CIP | TMP |
| 4 | >4 | >8 | <0.03 | >64 | >8 | 0.5 |
| 4 | >4 | >8 | <0.03 | >64 | >8 | 0.5 |
| <2 | >4 | 8 | <0.03 | 64 | 0.25 | >16 |
| 8 | >4 | >8 | <0.03 | >64 | >8 | 1 |
| 32 | >4 | 8 | <0.03 | 8 | 0.5 | >16 |
| 4 | >4 | >8 | <0.03 | >64 | >8 | 0.5 |
| <2 | 4 | >8 | <0.03 | 16 | 0.5 | >16 |
| <2 | >4 | 8 | <0.03 | >64 | >8 | 0.5 |
| 4 | >4 | 8 | <0.03 | >64 | >8 | 0.5 |
| 8 | >4 | >8 | <0.03 | >64 | >8 | 0.5 |
| <2 | >4 | 8 | <0.03 | >64 | >8 | 0.5 |
| 8 | >4 | >8 | <0.03 | >64 | >8 | 0.5 |
| 8 | >4 | >8 | <0.03 | >64 | >8 | 0.5 |
| >64 | >4 | >8 | 0.5 | 64 | 1 | 0.5 |
| >64 | >4 | >8 | 0.06 | >64 | 0.5 | >16 |
| 8 | >4 | >8 | <0.03 | >64 | >8 | 0.5 |
| 4 | >4 | >8 | <0.03 | >64 | >8 | 0.5 |
| 4 | >4 | 8 | <0.03 | >64 | >8 | 0.5 |
| >64 | >4 | >8 | 0.06 | 64 | 1 | 1 |
| 32 | >4 | 2 | <0.03 | <4 | 0.25 | >16 |
| 32 | >4 | 4 | <0.03 | >64 | >8 | >16 |
| 8 | >4 | 4 | <0.03 | 32 | 0.06 | >16 |
| 64 | >4 | 4 | <0.03 | >64 | 0.25 | >16 |
| >64 | >4 | >8 | 0.06 | >64 | 8 | >16 |
| 4 | >4 | 8 | <0.03 | <4 | <0.015 | <0.25 |
| 4 | >4 | 8 | 0.06 | <4 | 1 | >16 |
| 64 | >4 | >8 | 0.06 | >64 | 8 | >16 |
| >64 | >4 | >8 | >16 | >64 | >8 | >16 |
| >64 | >4 | 8 | <0.03 | <4 | <0.015 | >16 |
| 4 | >4 | >8 | <0.03 | >64 | >8 | <0.25 |
| 4 | >4 | 4 | <0.03 | >64 | 0.25 | 0.5 |
| 8 | >4 | >8 | 0.06 | <4 | 1 | >16 |
| 64 | >4 | 8 | <0.03 | 64 | 0.25 | >16 |
| 16 | >4 | 2 | <0.03 | >64 | >8 | 1 |
| 4 | >4 | 8 | <0.03 | <4 | <0.015 | <0.25 |
| 64 | >4 | 2 | <0.03 | >64 | 0.25 | >16 |
| 4 | >4 | 4 | <0.03 | <4 | <0.015 | <0.25 |
| 4 | >4 | 4 | <0.03 | <4 | <0.015 | <0.25 |
| >64 | >4 | >8 | >16 | >64 | >8 | >16 |
| >64 | >4 | >8 | >16 | >64 | >8 | >16 |
| >64 | >4 | >8 | 0.06 | >64 | >8 | >16 |
| 64 | >4 | 4 | <0.03 | >64 | >8 | >16 |

| | | | | | | |
|---|---|---|---|---|---|---|
| 16 | >4 | >8 | <0.03 | >64 | 4 | <0.25 |
| 16 | >4 | >8 | <0.03 | >64 | >8 | >16 |
| 16 | >4 | >8 | <0.03 | >64 | >8 | >16 |
| 64 | >4 | >8 | 0.06 | >64 | >8 | >16 |
| 64 | >4 | >8 | <0.03 | >64 | >8 | >16 |
| 16 | >4 | 8 | 0.06 | <4 | 1 | >16 |
| 64 | >4 | >8 | 0.06 | >64 | >8 | >16 |
| >64 | >4 | >8 | <0.03 | <4 | 1 | >16 |
| 64 | >4 | >8 | <0.03 | <4 | 0.5 | <0.25 |
| 16 | >4 | >8 | <0.03 | 8 | 0.5 | >16 |
| 64 | >4 | >8 | <0.03 | <4 | 1 | >16 |
| 32 | >4 | >8 | <0.03 | >64 | >8 | >16 |
| 32 | >4 | >8 | <0.03 | >64 | 2 | >16 |
| 32 | >4 | >8 | <0.03 | >64 | >8 | >16 |
| 8 | >4 | >8 | <0.03 | >64 | 8 | >16 |
| 32 | >4 | >8 | <0.03 | >64 | >8 | >16 |
| 8 | >4 | >8 | <0.03 | >64 | >8 | >16 |
| 16 | >4 | >8 | <0.03 | >64 | >8 | >16 |
| 32 | <0.25 | <0.25 | <0.03 | <4 | 0.06 | 1 |
| 32 | <0.25 | 1 | <0.03 | 16 | 0.06 | 8 |
| 16 | 4 | >8 | 2 | >64 | 0.5 | <0.25 |
| 16 | 4 | >8 | 2 | >64 | 0.5 | <0.25 |
| 16 | 2 | >8 | 1 | 64 | 1 | <0.25 |
| 32 | >4 | >8 | <0.03 | >64 | 0.5 | >16 |
| 4 | 4 | >8 | <0.03 | >64 | 4 | >16 |
| 32 | >4 | 0.5 | <0.03 | >64 | 0.5 | >16 |
| 32 | >4 | 0.5 | <0.03 | >64 | 0.5 | >16 |
| 16 | >4 | 2 | <0.03 | <4 | 0.5 | >16 |
| 4 | >4 | 4 | <0.03 | >64 | 0.25 | 0.5 |
| 4 | >4 | 4 | <0.03 | >64 | 0.25 | 0.5 |
| 4 | >4 | 4 | <0.03 | >64 | 0.25 | 0.5 |
| 4 | >4 | 4 | <0.03 | >64 | 0.25 | 0.5 |
| 4 | >4 | 4 | <0.03 | >64 | 0.5 | 0.5 |
| 8 | >4 | 8 | <0.03 | 32 | 0.06 | >16 |
| >64 | >4 | 1 | <0.03 | >64 | 0.25 | <0.25 |
| 64 | >4 | 8 | <0.03 | >64 | >8 | <0.25 |
| 4 | >4 | >8 | <0.03 | >64 | 0.25 | >16 |
| 32 | >4 | 8 | <0.03 | >64 | 8 | 0.5 |
| >64 | >4 | 8 | <0.03 | >64 | 0.5 | >16 |
| <2 | >4 | >8 | <0.03 | <4 | 0.5 | >16 |
| 8 | >4 | >8 | 0.12 | <4 | 2 | >16 |
| 4 | >4 | >8 | <0.03 | 32 | 0.12 | 0.5 |
| >64 | >4 | >8 | 0.12 | 32 | 1 | 0.5 |
| 64 | >4 | 8 | <0.03 | >64 | 0.5 | >16 |
| 64 | >4 | >8 | 0.06 | 8 | 2 | >16 |
| 4 | >4 | >8 | <0.03 | <4 | 0.25 | >16 |

| | | | | | | |
|---|---|---|---|---|---|---|
| >64 | >4 | 8 | 0.06 | >64 | 1 | >16 |
| 64 | >4 | >8 | <0.03 | >64 | 8 | <0.25 |
| 8 | >4 | >8 | 0.06 | <4 | 1 | >16 |
| 16 | >4 | >8 | <0.03 | 16 | 1 | >16 |
| 16 | >4 | 8 | <0.03 | >64 | >8 | >16 |
| 32 | >4 | >8 | 0.06 | >64 | >8 | >16 |
| 16 | >4 | >8 | <0.03 | >64 | 4 | >16 |
| 8 | >4 | >8 | <0.03 | >64 | 8 | >16 |
| 16 | >4 | 8 | <0.03 | >64 | >8 | >16 |
| 32 | >4 | >8 | 0.06 | >64 | >8 | >16 |
| 32 | >4 | >8 | 0.06 | >64 | >8 | >16 |
| 16 | >4 | >8 | <0.03 | 16 | 2 | >16 |
| 8 | >4 | 8 | <0.03 | >64 | >8 | >16 |
| 16 | >4 | >8 | <0.03 | >64 | >8 | >16 |
| 16 | >4 | >8 | <0.03 | >64 | >8 | >16 |
| 16 | >4 | >8 | <0.03 | 16 | 2 | >16 |
| 64 | >4 | >8 | 0.06 | >64 | >8 | >16 |
| 16 | >4 | >8 | <0.03 | >64 | >8 | >16 |
| 16 | >4 | >8 | <0.03 | >64 | >8 | >16 |
| 16 | >4 | >8 | <0.03 | >64 | >8 | >16 |
| 16 | >4 | >8 | <0.03 | >64 | >8 | >16 |
| 16 | >4 | >8 | <0.03 | >64 | >8 | >16 |
| 16 | >4 | >8 | <0.03 | >64 | >8 | >16 |
| 16 | >4 | >8 | <0.03 | >64 | >8 | >16 |
| 16 | >4 | >8 | <0.03 | >64 | >8 | >16 |
| 16 | >4 | >8 | <0.03 | >64 | >8 | >16 |
| 16 | >4 | >8 | <0.03 | >64 | >8 | >16 |
| 64 | >4 | >8 | 0.06 | >64 | >8 | >16 |
| 16 | >4 | >8 | <0.03 | >64 | 8 | >16 |
| 64 | >4 | >8 | 0.06 | >64 | >8 | >16 |
| 16 | >4 | >8 | <0.03 | >64 | >8 | >16 |
| 16 | <0.25 | 0.5 | <0.03 | 16 | 2 | >16 |
| 16 | 0.5 | 1 | <0.03 | 16 | 0.25 | >16 |

ıent); AMP (ampicillin); AZM (azithromycin); CTX (cefotaxime); CAZ (ceftazidime); MEM (ı
[L (chloramphenicol); CST (colistin).

| ) | | | | | | |
|---|---|---|---|---|---|---|
| **SMX** | **TET** | **TGC** | **GEN** | **AMI** | **CHL** | **CST** |
| >512 | <2 | <0.25 | <0.5 | <4 | <8 | <1 |
| >512 | <2 | <0.25 | <0.5 | <4 | <8 | <1 |
| 64 | >32 | 0.5 | <0.5 | <4 | <8 | <1 |
| 32 | <2 | <0.25 | >16 | <4 | 16 | <1 |
| >512 | <2 | 0.5 | >16 | 8 | >64 | <1 |
| 64 | <2 | 0.5 | >16 | <4 | 16 | <1 |
| >512 | <2 | <0.25 | >16 | <4 | <8 | <1 |
| 64 | <2 | <0.25 | <0.5 | <4 | <8 | <1 |
| 16 | <2 | <0.25 | >16 | <4 | <8 | <1 |
| 64 | <2 | <0.25 | >16 | <4 | 16 | <1 |
| 32 | <2 | <0.25 | >16 | <4 | <8 | <1 |
| 64 | <2 | <0.25 | >16 | <4 | 16 | <1 |
| 64 | <2 | <0.25 | >16 | <4 | <8 | <1 |
| 32 | 4 | 0.5 | 1 | <4 | 16 | <1 |
| >512 | 32 | <0.25 | 1 | <4 | <8 | <1 |
| 64 | <2 | <0.25 | >16 | <4 | 16 | <1 |
| 64 | <2 | <0.25 | >16 | <4 | <8 | <1 |
| 32 | <2 | <0.25 | >16 | <4 | <8 | <1 |
| 16 | 4 | <0.25 | <0.5 | <4 | <8 | <1 |
| >512 | >32 | <0.25 | <0.5 | <4 | <8 | <1 |
| >512 | >32 | <0.25 | 1 | <4 | <8 | <1 |
| >512 | >32 | 0.5 | 1 | <4 | <8 | <1 |
| >512 | >32 | <0.25 | >16 | <4 | <8 | <1 |
| >512 | <2 | <0.25 | >16 | <4 | <8 | <1 |
| <8 | <2 | <0.25 | 1 | 8 | <8 | <1 |
| >512 | <2 | <0.25 | >16 | 8 | <8 | <1 |
| >512 | <2 | <0.25 | 16 | <4 | <8 | <1 |
| >512 | >32 | <0.25 | <0.5 | <4 | >64 | <1 |
| >512 | >32 | <0.25 | 1 | <4 | <8 | <1 |
| 32 | <2 | <0.25 | >16 | <4 | <8 | <1 |
| 32 | <2 | <0.25 | <0.5 | <4 | <8 | <1 |
| >512 | >32 | <0.25 | >16 | 8 | <8 | <1 |
| 32 | >32 | <0.25 | 1 | <4 | <8 | <1 |
| >512 | 32 | <0.25 | >16 | 8 | 16 | <1 |
| <8 | <2 | <0.25 | <0.5 | <4 | <8 | <1 |
| >512 | >32 | <0.25 | <0.5 | <4 | <8 | <1 |
| <8 | <2 | <0.25 | <0.5 | <4 | <8 | <1 |
| <8 | <2 | <0.25 | <0.5 | <4 | <8 | <1 |
| >512 | >32 | 0.5 | <0.5 | <4 | >64 | <1 |
| >512 | >32 | <0.25 | <0.5 | <4 | >64 | <1 |
| >512 | >32 | <0.25 | >16 | <4 | <8 | <1 |
| >512 | 32 | 0.5 | 16 | <4 | <8 | <1 |

| | | | | | | |
|---|---|---|---|---|---|---|
| <8 | <2 | <0.25 | <0.5 | <4 | <8 | <1 |
| >512 | >32 | 1 | 16 | <4 | <8 | <1 |
| >512 | >32 | 1 | 16 | <4 | <8 | <1 |
| >512 | 32 | <0.25 | 16 | <4 | <8 | <1 |
| >512 | >32 | <0.25 | 16 | <4 | >64 | <1 |
| >512 | 32 | 0.5 | >16 | <4 | 16 | <1 |
| >512 | >32 | <0.25 | 16 | <4 | >64 | <1 |
| >512 | <2 | <0.25 | >16 | <4 | >64 | <1 |
| >512 | <2 | <0.25 | <0.5 | <4 | >64 | <1 |
| >512 | <2 | <0.25 | <0.5 | <4 | >64 | <1 |
| >512 | <2 | <0.25 | >16 | <4 | >64 | <1 |
| >512 | >32 | 4 | 16 | <4 | 16 | <1 |
| >512 | >32 | 4 | 16 | <4 | 32 | <1 |
| >512 | >32 | 4 | 16 | <4 | 32 | <1 |
| >512 | >32 | 1 | >16 | <4 | <8 | <1 |
| >512 | >32 | 2 | 4 | <4 | <8 | <1 |
| >512 | >32 | 2 | >16 | <4 | <8 | <1 |
| >512 | >32 | 2 | 16 | <4 | 16 | <1 |
| <8 | 8 | 1 | <0.5 | <4 | 8 | <1 |
| 32 | 16 | 4 | <0.5 | <4 | 32 | <1 |
| <8 | <2 | <0.25 | 4 | <4 | <8 | <1 |
| <8 | <2 | <0.25 | 4 | <4 | <8 | <1 |
| <8 | <2 | <0.25 | 4 | <4 | <8 | <1 |
| <8 | >32 | <0.25 | <0.5 | <4 | <8 | <1 |
| >512 | <2 | <0.25 | >16 | <4 | <8 | <1 |
| >512 | >32 | <0.25 | >16 | <4 | <8 | <1 |
| >512 | >32 | <0.25 | >16 | <4 | <8 | <1 |
| >512 | >32 | <0.25 | 1 | <4 | <8 | <1 |
| 128 | <2 | <0.25 | <0.5 | <4 | <8 | <1 |
| 64 | <2 | <0.25 | <0.5 | <4 | <8 | <1 |
| 64 | <2 | <0.25 | 1 | <4 | <8 | <1 |
| 128 | <2 | <0.25 | 1 | <4 | <8 | <1 |
| 128 | <2 | <0.25 | <0.5 | <4 | <8 | <1 |
| >512 | >32 | 0.5 | <0.5 | <4 | <8 | <1 |
| <8 | >32 | <0.25 | <0.5 | <4 | <8 | <1 |
| 16 | <2 | <0.25 | >16 | <4 | <8 | <1 |
| >512 | >32 | <0.25 | 1 | <4 | <8 | <1 |
| >512 | 32 | <0.25 | >16 | 8 | >64 | <1 |
| >512 | >32 | <0.25 | >16 | <4 | 16 | <1 |
| >512 | >32 | <0.25 | <0.5 | <4 | <8 | <1 |
| >512 | >32 | <0.25 | >16 | 16 | <8 | <1 |
| >512 | <2 | <0.25 | <0.5 | <4 | <8 | 8 |
| 16 | <2 | <0.25 | 1 | <4 | <8 | <1 |
| >512 | >32 | <0.25 | >16 | <4 | 16 | <1 |
| >512 | >32 | <0.25 | >16 | 8 | 16 | <1 |
| >512 | >32 | <0.25 | <0.5 | 4 | <8 | <1 |

| | | | | | | |
|---|---|---|---|---|---|---|
| >512 | 32 | <0.25 | 8 | <4 | <8 | <1 |
| >512 | >32 | <0.25 | >16 | 8 | >64 | <1 |
| >512 | <2 | <0.25 | >16 | 16 | <8 | <1 |
| 64 | <2 | <0.25 | <0.5 | <4 | <8 | <1 |
| >512 | >32 | 0.5 | 16 | <4 | <8 | <1 |
| >512 | >32 | <0.25 | 16 | <4 | >64 | <1 |
| >512 | 4 | 0.5 | 16 | <4 | <8 | 2 |
| >512 | 4 | 0.5 | 16 | <4 | <8 | 2 |
| >512 | >32 | 0.5 | >16 | <4 | <8 | <1 |
| >512 | >32 | <0.25 | 16 | <4 | >64 | <1 |
| >512 | >32 | <0.25 | 16 | <4 | >64 | <1 |
| >512 | >32 | 0.5 | 16 | <4 | <8 | <1 |
| >512 | >32 | 0.5 | 16 | <4 | <8 | <1 |
| >512 | >32 | 0.5 | 16 | <4 | <8 | <1 |
| >512 | >32 | 0.5 | >16 | <4 | <8 | <1 |
| >512 | >32 | 0.5 | 16 | <4 | <8 | <1 |
| >512 | >32 | <0.25 | 16 | <4 | >64 | <1 |
| >512 | >32 | 1 | 16 | <4 | <8 | <1 |
| >512 | >32 | 1 | >16 | <4 | <8 | <1 |
| >512 | >32 | 1 | 16 | <4 | <8 | <1 |
| >512 | >32 | 1 | 16 | <4 | <8 | <1 |
| >512 | >32 | 1 | 16 | <4 | <8 | <1 |
| >512 | >32 | 1 | 16 | <4 | <8 | <1 |
| >512 | >32 | 1 | 16 | <4 | <8 | <1 |
| >512 | >32 | 1 | 16 | <4 | <8 | <1 |
| >512 | >32 | 1 | 16 | <4 | <8 | <1 |
| >512 | >32 | 1 | 16 | <4 | <8 | <1 |
| >512 | >32 | <0.25 | 16 | <4 | >64 | <1 |
| >512 | 4 | 0.5 | 16 | <4 | <8 | 4 |
| >512 | >32 | <0.25 | 16 | <4 | >64 | |
| >512 | 4 | 0.5 | 16 | <4 | <8 | 4 |
| >512 | >32 | 2 | <0.5 | <4 | 16 | <1 |
| >512 | >32 | 4 | <0.5 | <4 | >64 | <1 |

neropenem); NAL (nalidixic acid); CIP (ciprofloxacin); TMP (trimethoprim); SMX
