## Supplementary material for "Novel antibiotic resistance genes from the hospital effluent are disseminated into the marine environment in Norway": supplemantary table S2

**Supplementary Table S2:** Genome assembly statistics of sequenced strains (n=36) in this stud

| Strain | Species | Accession number | Genome size (bp) |
|---|---|---|---|
| 11-303 | *E. coli* | | 5,361,939 |
| 11-305 | *E. coli* | | 5,172,230 |
| 11-315 | *E. coli* | | 5,134,302 |
| 11-317 | *E. coli* | | 5,058,423 |
| 11-320 | *E. coli* | | 5,121,918 |
| 11-321 | *E. coli* | | 5,121,196 |
| 11-330 | *E. coli* | | 4,889,655 |
| 11-336 | *E. coli* | | 5,161,917 |
| 11-338 | *E. coli* | | 4,937,829 |
| 12-306 | *E. coli* | | 4,967,142 |
| 12-310 | *E. coli* | | 5,219,703 |
| 12-327 | *E. coli* | | 4,590,194 |
| 12-328 | *E. coli* | | 4,860,094 |
| 12-336 | *E. coli* | | 4,806,073 |
| 12-337 | *E. coli* | | 5,288,896 |
| 12-341 | *E. coli* | | 4,878,940 |
| 12-343 | *E. coli* | | 4,866,874 |
| 11-M301 | *E. coli* | | 4,893,152 |
| 11-M302 | *E. coli* | | 4,888,903 |
| K10-311 | *K. pneumoniae* | | 5,857,362 |
| K10-321 | *K. michiganensis* | | 6,384,740 |
| K10-M302 | *K. michiganensis* | | 6,109,672 |
| K10-M304 | *K. michiganensis* | | 6,168,049 |
| K10-M306 | *K. michiganensis* | | 6,180,804 |
| K10-T302 | *K. pneumoniae* | | 5,858,561 |
| K10-T303 | *K. pneumoniae* | | 5,869,222 |
| K10-T306 | *K. pneumoniae* | | 5,884,286 |
| K11-302 | *K. pneumoniae* | | 5,476,119 |
| K11-303 | *K. michiganensis* | | 6,279,332 |
| K11-304 | *K. pneumoniae* | | 5,808,781 |
| K11-305 | *K. pneumoniae* | | 5,804,183 |
| K11-314 | *K. michiganensis* | | 6,259,219 |
| K11-324 | *K. pneumoniae* | | 5,874,880 |
| K11-325 | *K. michiganensis* | | 6,346,676 |
| K11-327 | *K. pneumoniae* | | 5,803,532 |
| K11-329 | *K. pneumoniae* | | 5,814,628 |

y.

| No. of contigs (≥500 bp) | Largest contig | N50 | GC content (%) | tRNA genes |
|---|---|---|---|---|
| 129 | 391.648 | 137.123 | 50.71 | 78 |
| 154 | 271.157 | 100.393 | 50.78 | 77 |
| 206 | 300.645 | 81.982 | 50.61 | 72 |
| 75 | 602.478 | 201.086 | 50.52 | 73 |
| 168 | 313.888 | 113.319 | 50.61 | 75 |
| 109 | 531.044 | 190.924 | 50.81 | 80 |
| 95 | 527.397 | 147.301 | 50.64 | 75 |
| 135 | 375.261 | 112.495 | 50.69 | 74 |
| 125 | 418.355 | 107.152 | 50.61 | 79 |
| 66 | 599.233 | 262.718 | 50.61 | 76 |
| 67 | 462.721 | 225.822 | 50.77 | 81 |
| 97 | 356.940 | 134.206 | 50.61 | 73 |
| 135 | 523.725 | 125.474 | 50.63 | 75 |
| 172 | 150.488 | 64.731 | 50.91 | 69 |
| 191 | 343.918 | 118.963 | 50.73 | 74 |
| 157 | 162.241 | 71.476 | 51.03 | 72 |
| 177 | 184.106 | 75.268 | 50.88 | 74 |
| 91 | 543.390 | 147.301 | 50.64 | 75 |
| 96 | 527.397 | 147.301 | 50.63 | 75 |
| 104 | 699.965 | 307.14 | 56.64 | 77 |
| 113 | 548.666 | 157.103 | 55.46 | 71 |
| 173 | 342.450 | 198.305 | 55.62 | 70 |
| 165 | 342.450 | 194.646 | 55.63 | 71 |
| 178 | 425.784 | 219.167 | 55.51 | 69 |
| 109 | 559.528 | 354.606 | 56.62 | 73 |
| 97 | 559.528 | 307.140 | 56.63 | 73 |
| 88 | 559.528 | 354.606 | 56.62 | 77 |
| 46 | 593.057 | 377.679 | 57.28 | 76 |
| 96 | 478.315 | 280.717 | 55.94 | 71 |
| 131 | 595.088 | 264.543 | 56.93 | 73 |
| 105 | 595.088 | 302.134 | 56.94 | 75 |
| 103 | 687.219 | 324.010 | 55.94 | 71 |
| 84 | 946.133 | 354.606 | 56.63 | 77 |
| 94 | 950.607 | 346.128 | 55.91 | 75 |
| 107 | 595.088 | 302.134 | 56.94 | 75 |
| 116 | 595.088 | 302.134 | 56.81 | 75 |

| No. of CDSs (total) |
|---|
| 5.277 |
| 5.072 |
| 5.164 |
| 4.938 |
| 5.109 |
| 5.041 |
| 4.701 |
| 5.073 |
| 4.836 |
| 4.713 |
| 5.017 |
| 4.449 |
| 4.876 |
| 4.744 |
| 5.273 |
| 4.773 |
| 4.749 |
| 4.704 |
| 4.705 |
| 5.751 |
| 6.064 |
| 5.878 |
| 5.947 |
| 5.985 |
| 5.771 |
| 5.768 |
| 5.780 |
| 5.248 |
| 5.959 |
| 5.758 |
| 5.747 |
| 5.931 |
| 5.765 |
| 6.053 |
| 5.745 |
| 5.756 |
