## Supplementary material for "Novel antibiotic resistance genes from the hospital effluent are disseminated into the marine environment in Norway": supplemantary table S3

**Supplementary Table S3:** Detected antibiotic resistance genes (ARGs) and sequen

| Strain | Species | Sample type | ST |
|---|---|---|---|
| 11-303 | *E. coli* | Hospital-eff. | 69 |
| 11-305 | *E. coli* | Hospital-eff. | 399 |
| 11-315 | *E. coli* | Hospital-eff. | 401 |
| 11-317 | *E. coli* | Hospital-eff. | 131 |
| 11-320 | *E. coli* | Hospital-eff. | 401 |
| 11-321 | *E. coli* | STP-inf. | 5614 |
| 11-330 | *E. coli* | STP-inf. | 11681 |
| 11-336 | *E. coli* | STP-T-eff. | 448 |
| 11-338 | *E. coli* | STP-T-eff. | 2253 |
| 12-306 | *E. coli* | STP-inf. | 38 |
| 12-310 | *E. coli* | STP-inf. | 219 |
| 12-327 | *E. coli* | STP-T-eff. | 10 |
| 12-328 | *E. coli* | STP-T-eff. | 10 |
| 12-336 | *E. coli* | Hospital-eff. | 11873 |
| 12-337 | *E. coli* | Hospital-eff. | 10 |
| 12-341 | *E. coli* | Hospital-eff. | 11873 |
| 12-343 | *E. coli* | Hospital-eff. | 398 |
| 11-M301 | *E. coli* | STP-inf. | 11681 |
| 11-M302 | *E. coli* | STP-T-eff. | 11681 |
| K10-311 | *K. pneumoniae* | STP-T-eff. | ST307 |
| K10-321 | *K. michiganensis* | STP-T-eff. | NA |
| K10-T302 | *K. michiganensis* | Hospital-eff. | NA |
| K10-T303 | *K. michiganensis* | Hospital-eff. | NA |
| K10-T306 | *K. michiganensis* | Hospital-eff. | NA |
| K10-M302 | *K. pneumoniae* | Hospital-eff. | ST307 |
| K10-M304 | *K. pneumoniae* | Hospital-eff. | ST307 |

| | | | |
|---|---|---|---|
| K10-M306 | *K. pneumoniae* | Hospital-eff. | ST307 |
| K11-302 | *K. pneumoniae* | STP-inf. | ST307 |
| K11-303 | *K. michiganensis* | STP-inf. | NA |
| K11-304 | *K. pneumoniae* | STP-inf. | ST2947 |
| K11-305 | *K. pneumoniae* | STP-inf. | ST2947 |
| K11-314 | *K. michiganensis* | STP-T-eff. | NA |
| K11-324 | *K. pneumoniae* | Hospital-eff. | ST307 |
| K11-325 | *K. michiganensis* | Hospital-eff. | NA |
| K11-327 | *K. pneumoniae* | Hospital-eff. | ST2947 |
| K11-329 | *K. pneumoniae* | Hospital-eff. | ST2947 |

**Legend:** eff (effluent); STP (sewage treatment plant); inf (influent); T (treated); NA ce types (STs) in sequenced strains (n=36).

| ARGs |
|---|
| *mph(A), dfrA14, tet(B), blaCTX-M-15* |
| *aadA1, aac(3)-IIa, aac(6')-Ib-cr, aph(3')-Ia, dfrA14, sul1, sul2, qnrS1, ARR-2, blaCTX-M-15, blaOXA-1. blaDHA* <br> *blaOXA-10, blaTEM-1B, mph(A), catB3, catA2, floR, cmlA1* |
| *msr(E), mph(E), blaSHV-12, blaOXA-2* |
| *aac(3)-IId, blaTEM-1B, blaCTX-M-15* |
| *msr(E), mph(E), blaSHV-12, blaOXA-2* |
| *aph(3")-lb, aph(6)-Id, aadA5, qnrS1, dfrA17, sul1, sul2, tet(A), blaCTX-M-15, mph(A)* |
| *dfrA1, dfrA12, sul1, aadA2, aadA1, mph(A), tet(A), tet(B), blaCMY-145, blaNDM-5, blaOXA-1, catA1* |
| *sul1, sul2, ant(2``)-Ia, aadA1, aph(3")-Ib, aac(6')-Ib-cr, aac(3)-Iia, aph(6)-Id, mph(A), tet(A), blaOXA-1, blaTEM* <br> *blaCTX-M-15, catB3, cmlA1* |
| *dfrA17, sul1, aadA5, mph(A), tet(A), blaCTX-M-15* |
| *dfrA17, sul1, sul2, aph(3``)-Ib, aadA5, aac(3)-IId, aph(6)-Id, mph(A), tet(D), blaTEM-1B, blaCTX-M-14, catA1* |
| *aadA5, aph(3")-lb, aph(6)-ld, mph(A), qnrS1, dfrA17, sul1, sul2, tet(A), blaCTX-M-15* |
| *aph(6)-Id, aadA1, aph(3")-Ib, dfrA1, sul2, tet(A), blaCTX-M-15* |
| *tet(B), blaCTX-M-14b, mph(A)* |
| *aac(3)-lld, aph(6)-ld, aac(6')-lb-cr, aph(3")-lb, qnrS1, dfrA14, sul2, tet(A), blaLAP-2, blaOXA-1, blaCTX-M-105,* |
| *sul2, blaSHV-12, blaOXA-2* |
| *dfrA12, dfrA14, sul1, sul2, aac(3)ll, aac(3)-IV, aac(6')-lb-cr, aph(4)-la, aph(3")-lb, aadA2, aph(6)-ld, aph(3')-la,* <br> *qnrS1, tet(A), blaCTX-M-105, blaLAP-2, blaOXA-1, catB3* |
| *msr(E), aadA2, aph(6)-ld, aph(3")-lb, aac(3)-I, mph(E), dfrA23, sul1, sul2, tet(C), blaOXA-2, blaCTX-M-3, blaTE* |
| *blaNDM-5, blaOXA-1, blaCMY-145, aadA1, aadA2, mph(A), dfrA1, dfrA12, sul1, tet(A), tet(B), catA1* |
| *blaNDM-5, blaOXA-1, blaCMY-145, aadA1, aadA2, mph(A), dfrA1, dfrA12, sul1, tet(A), tet(B), catA1* |
| *aph(3")-Ib, aph(6)-Id, aac(3)-IIe, aac(6')-Ib-cr5, bla,class A beta-lactamase , blaCTX-M-15, blaOXA-1, sul2, dfr* <br> *oqxB19, fosA5, tet(A), qnrB1, catB* |
| *aph(3')-Ia, aac(3)-IIe, aac(6')-Ib-cr5,aadA1, blaTEM-1, blaOXY-1-4, blaCTX-M-15, blaOXA-1,blaOXA-10, sul1,* <br> *dfrA14, mph(A), qnrS1, arr-2, catA2* |
| *aph(3")-Ib , aph(6)-Id , aac(3)-IIe, aac(6')-Ib-cr5, blaCTX-M-15, blaOXA-1, blaTEM-1, class A beta-lactamase,* <br> *dfrA14, oqxA, oqxB19, fosA5, tet(A), qnrB1, catB* |
| *aph(3")-Ib, aph(6)-Id, aac(3)-IIe, aac(6')-Ib-cr5, blaCTX-M-15, blaOXA-1,class A beta-lactamase, sul2, dfrA14,* <br> *oqxA,oqxB19, fosA5, tet(A), qnrB1,catB* |
| *aph(3")-Ib, aph(6)-Id, aac(3)-IIe, aac(6')-Ib-cr5, blaCTX-M-15, blaOXA-1,, class A beta-lactamase, sul2, dfrA14* <br> *oqxB19, fosA5, tet(A), qnrB1, catB* |
| *aac(3)-I, aac(6')-4, blaOXA-2, blaOXA-9, blaOXY-5-6, blaTEM-1, blaKPC-3, oqxB* |
| *aac(3)-I,aac(6')-4,blaOXY-5-6, blaOXA-2, blaOXA-9, blaTEM-1, blaKPC-3, oqxB* |

*ant(3''), aac(6')-Ib4, blaOXA-9, blaOXY-5-6, blaTEM-1, blaKPC-3, qnrS1, oqxB*

*aph(3'')-Ib, aph(6)-Id, aac(3)-IIe, aac(6')-Ib-cr5, blaCTX-M-15, blaOXA-1, blaTEM-1,blaSHV-28, sul2, dfrA14, o[illegible] oqxB19, fosA5, tet(A), qnrB1, catB*

*ant(3'')-Ia, aph(3')-I, aph(3'')-Ib, aph(6)-Id, aac(6')-Ib-cr5, blaCTX-M-15, blaOXA-1, blaOXY-1-1, blaTEM-1, su[illegible] dfrA17,oqxB, mph(A), tet(A), tet(B), qnrB1, catA1*

*aph(3'')-Ib, aph(6)-Id, aac(3)-IId, class A beta-lactamase X2, sul2, dfrA14,oqxA, oqxB19, fosA5*

*aph(3'')-Ib , aph(6)-Id, aac(3)-Ild, class A beta-lactamase X2, sul2, dfrA14, oqxA, oqxB19, fosA5*

*ant(3'')-Ia, aph(3')-I, aph(3'')-Ib, aph(6)-Id, aac(3)-IId,aac(3)-IIe, aac(6')-Ib-cr5, blaCTX-M-15, blaOXA-1, blaO[illegible] blaTEM-1, sul1,dfrA14, dfrA17, oqxB, mph(A), tet(A), tet(B), qnrB1, catB*

*aph(3'')-Ib, aph(6)-Id, aac(3)-IIe, aac(6')-Ib-cr5, blaCTX-M-15, blaOXA-1, class A beta-lactamase X2, sul2, dfrA[illegible] oqxB19, fosA5, tet(A), qnrB1, catB*

*ant(3'')-Ia, aph(3')-I, aph(3'')-Ib,aph(6)-Id, aac(3)-IIe, aac(3)-IId, aac(6')-Ib-cr5, blaCTX-M-15, blaOXA-1, blaO[illegible] blaTEM-1, sul1, dfrA14, dfrA17, oqxB, mph(A), tet(A), tet(B), qnrB1,catB*

*aac(3)-IId, aph(3'')-Ib, aph(6)-Id, class A beta-lactamase X2, sul2, dfrA14, oqxA, oqxB19, fosA5*

*aph(3'')-Ib, aph(6)-Id, aac(3)-IId, blaTEM,class A beta-lactamase X2, sul2, dfrA14, oqxA, oqxB19, fosA5*

(not available).
