## Supplementary material for "Novel antibiotic resistance genes from the hospital effluent are disseminated into the marine environment in Norway": supplemantary table S5

**Supplementary Table S5**: Relative abundance of most detected mobile genetic elements

| MGEs | Hospital effluent February | Hospital effluent April | STP influent February |
|---|---|---|---|
| IS*CR1* | 0.0004981 | 0.0007864 | 0.0000293 |
| IS*CR2* | 0.0028998 | 0.0015079 | 0.0004916 |
| IS*CR3* | 0 | 0 | 0 |
| IS*CR4* | 0 | 0 | 0 |
| IS*CR5* | 0.0005498 | 0.0003501 | 0.0001527 |
| IS*CR6* | 0 | 0 | 0 |
| IS*CR7* | 0 | 0 | 0 |
| IS*CR8* | 0 | 0 | 0 |
| IS*CR14* | 0 | 0 | 0 |
| *intI1* | 0.0609183 | 0.0397664 | 0.0053216 |
| *intI2* | 0.0026767 | 0.0022447 | 0.0012776 |
| *intI3* | 0.0000519 | 0.0000783 | 0.0000847 |
| *intI6* | 0 | 0 | 0 |
| *intI7* | 0 | 0 | 0 |
| *intI8* | 0 | 0 | 0 |
| *intI9* | 0 | 0 | 0 |
| *intI10* | 0 | 0 | 0 |
| other | 0.1736948 | 0.1484144 | 0.0731509 |

**Legend:** STP (sewage treatment plant).

(MGEs) across samples.

| STP influent April | STP treated effluent February | STP treated effluent April |
|---|---|---|
| 0.0000750 | 0.0000281 | 0.0000714 |
| 0.0004902 | 0.0002956 | 0.0003580 |
| 0 | 0 | 0 |
| 0 | 0 | 0 |
| 0.0001033 | 0.0000763 | 0.0000867 |
| 0 | 0 | 0 |
| 0 | 0 | 0 |
| 0 | 0 | 0 |
| 0 | 0 | 0 |
| 0.0061597 | 0.0070679 | 0.0063007 |
| 0.0010418 | 0.0008162 | 0.0008017 |
| 0.0000526 | 0.0000934 | 0.0000563 |
| 0 | 0 | 0 |
| 0 | 0 | 0 |
| 0 | 0 | 0 |
| 0 | 0 | 0 |
| 0 | 0 | 0 |
| 0.1019743 | 0.0650622 | 0.0871719 |
