## Supplementary material for "Novel antibiotic resistance genes from the hospital effluent are disseminated into the marine environment in Norway": supplemantary table S6

**Supplementary Table S6:** D

| Sample |
|---|
| Hospital-eff-F |
| Hospital-eff-F |
| Hospital-eff-F |
| Hospital-eff-F |
| Hospital-eff-F |
| Hospital-eff-F |
| Hospital-eff-F |
| Hospital-eff-F |
| Hospital-eff-F |
| Hospital-eff-F |
| Hospital-eff-F |
| Hospital-eff-F |
| Hospital-eff-F |
| Hospital-eff-F |
| Hospital-eff-F |
| Hospital-eff-F |
| Hospital-eff-F |
| Hospital-eff-F |
| Hospital-eff-F |
| Hospital-eff-F |
| Hospital-eff-F |
| Hospital-eff-F |
| Hospital-eff-F |
| Hospital-eff-F |
| Hospital-eff-F |
| Hospital-eff-F |
| Hospital-eff-F |
| Hospital-eff-F |
| Hospital-eff-F |
| Hospital-eff-F |
| Hospital-eff-F |
| Hospital-eff-F |
| Hospital-eff-F |
| Hospital-eff-F |
| Hospital-eff-F |
| Hospital-eff-F |
| Hospital-eff-F |
| Hospital-eff-F |
| Hospital-eff-F |
| Hospital-eff-F |
| Hospital-eff-F |
| Hospital-eff-F |
| Hospital-eff-F |

Hospital-eff-F
Hospital-eff-F
Hospital-eff-F
Hospital-eff-F
Hospital-eff-F
Hospital-eff-F
Hospital-eff-F
Hospital-eff-F
Hospital-eff-F
Hospital-eff-F
Hospital-eff-F
Hospital-eff-F
Hospital-eff-F
Hospital-eff-F
Hospital-eff-F
Hospital-eff-F
Hospital-eff-F
Hospital-eff-F
Hospital-eff-F
Hospital-eff-F
Hospital-eff-F
Hospital-eff-F
Hospital-eff-F
Hospital-eff-F
Hospital-eff-F
Hospital-eff-F
Hospital-eff-F
Hospital-eff-F
Hospital-eff-F
Hospital-eff-F
Hospital-eff-F
Hospital-eff-F
Hospital-eff-F
Hospital-eff-F
Hospital-eff-F
Hospital-eff-F
Hospital-eff-F
Hospital-eff-F
Hospital-eff-F
Hospital-eff-F
Hospital-eff-F
Hospital-eff-F
Hospital-eff-F
Hospital-eff-F
Hospital-eff-F
Hospital-eff-F

Hospital-eff-F
Hospital-eff-F
Hospital-eff-F
Hospital-eff-F
Hospital-eff-F
Hospital-eff-F
Hospital-eff-F
Hospital-eff-F
Hospital-eff-F
Hospital-eff-F
Hospital-eff-F
Hospital-eff-F
Hospital-eff-F
Hospital-eff-F
Hospital-eff-F
Hospital-eff-F
Hospital-eff-F
Hospital-eff-F
Hospital-eff-F
Hospital-eff-F
Hospital-eff-F
Hospital-eff-F
Hospital-eff-F
Hospital-eff-F
Hospital-eff-F
Hospital-eff-F
Hospital-eff-F
Hospital-eff-F
Hospital-eff-F
Hospital-eff-F
Hospital-eff-F
Hospital-eff-F
Hospital-eff-F
Hospital-eff-F
Hospital-eff-F
Hospital-eff-F
Hospital-eff-F
Hospital-eff-F
Hospital-eff-F
Hospital-eff-F
Hospital-eff-F
Hospital-eff-F
Hospital-eff-F
Hospital-eff-F
Hospital-eff-F
Hospital-eff-F

Hospital-eff-F
Hospital-eff-F
Hospital-eff-F
Hospital-eff-F
Hospital-eff-F
Hospital-eff-F
Hospital-eff-F
Hospital-eff-F
Hospital-eff-F
Hospital-eff-F
Hospital-eff-F
Hospital-eff-F
Hospital-eff-F
Hospital-eff-F
Hospital-eff-F
Hospital-eff-F
Hospital-eff-F
Hospital-eff-F
Hospital-eff-F
Hospital-eff-F
Hospital-eff-F
Hospital-eff-F
Hospital-eff-F
Hospital-eff-F
Hospital-eff-F
Hospital-eff-F
Hospital-eff-F
Hospital-eff-F
Hospital-eff-F
Hospital-eff-F
Hospital-eff-F
Hospital-eff-F
Hospital-eff-F
Hospital-eff-F
Hospital-eff-F
Hospital-eff-F
Hospital-eff-F
Hospital-eff-F
Hospital-eff-F
Hospital-eff-F
Hospital-eff-F
Hospital-eff-F
Hospital-eff-F
Hospital-eff-F
Hospital-eff-F
Hospital-eff-F

Hospital-eff-F
Hospital-eff-F
Hospital-eff-F
Hospital-eff-F
Hospital-eff-F
Hospital-eff-F
Hospital-eff-F
Hospital-eff-F
Hospital-eff-F
Hospital-eff-F
Hospital-eff-F
Hospital-eff-F
Hospital-eff-F
Hospital-eff-F
Hospital-eff-F
Hospital-eff-F
Hospital-eff-F
Hospital-eff-F
Hospital-eff-F
Hospital-eff-F
Hospital-eff-F
Hospital-eff-F
Hospital-eff-F
Hospital-eff-F
Hospital-eff-F
Hospital-eff-F
Hospital-eff-F
Hospital-eff-F
Hospital-eff-F
Hospital-eff-F
Hospital-eff-F
Hospital-eff-F
Hospital-eff-F
Hospital-eff-F
Hospital-eff-F
Hospital-eff-A
Hospital-eff-A
Hospital-eff-A
Hospital-eff-A
Hospital-eff-A
Hospital-eff-A
Hospital-eff-A
Hospital-eff-A
Hospital-eff-A
Hospital-eff-A
Hospital-eff-A

Hospital-eff-A
Hospital-eff-A
Hospital-eff-A
Hospital-eff-A
Hospital-eff-A
Hospital-eff-A
Hospital-eff-A
Hospital-eff-A
Hospital-eff-A
Hospital-eff-A
Hospital-eff-A
Hospital-eff-A
Hospital-eff-A
Hospital-eff-A
Hospital-eff-A
Hospital-eff-A
Hospital-eff-A
Hospital-eff-A
Hospital-eff-A
Hospital-eff-A
Hospital-eff-A
Hospital-eff-A
Hospital-eff-A
Hospital-eff-A
Hospital-eff-A
Hospital-eff-A
Hospital-eff-A
Hospital-eff-A
Hospital-eff-A
Hospital-eff-A
Hospital-eff-A
Hospital-eff-A
Hospital-eff-A
Hospital-eff-A
Hospital-eff-A
Hospital-eff-A
Hospital-eff-A
Hospital-eff-A
Hospital-eff-A
Hospital-eff-A
Hospital-eff-A
Hospital-eff-A
Hospital-eff-A
Hospital-eff-A
Hospital-eff-A
Hospital-eff-A

Hospital-eff-A
Hospital-eff-A
Hospital-eff-A
Hospital-eff-A
Hospital-eff-A
Hospital-eff-A
Hospital-eff-A
Hospital-eff-A
Hospital-eff-A
Hospital-eff-A
Hospital-eff-A
Hospital-eff-A
Hospital-eff-A
Hospital-eff-A
Hospital-eff-A
Hospital-eff-A
Hospital-eff-A
Hospital-eff-A
Hospital-eff-A
Hospital-eff-A
Hospital-eff-A
Hospital-eff-A
Hospital-eff-A
Hospital-eff-A
Hospital-eff-A
Hospital-eff-A
Hospital-eff-A
Hospital-eff-A
Hospital-eff-A
Hospital-eff-A
Hospital-eff-A
Hospital-eff-A
Hospital-eff-A
Hospital-eff-A
Hospital-eff-A
Hospital-eff-A
Hospital-eff-A
Hospital-eff-A
Hospital-eff-A
Hospital-eff-A
Hospital-eff-A
Hospital-eff-A
Hospital-eff-A
Hospital-eff-A
Hospital-eff-A
Hospital-eff-A

Hospital-eff-A
Hospital-eff-A
Hospital-eff-A
Hospital-eff-A
Hospital-eff-A
Hospital-eff-A
Hospital-eff-A
Hospital-eff-A
Hospital-eff-A
Hospital-eff-A
Hospital-eff-A
Hospital-eff-A
Hospital-eff-A
Hospital-eff-A
Hospital-eff-A
Hospital-eff-A
Hospital-eff-A
Hospital-eff-A
Hospital-eff-A
Hospital-eff-A
Hospital-eff-A
Hospital-eff-A
Hospital-eff-A
Hospital-eff-A
Hospital-eff-A
Hospital-eff-A
Hospital-eff-A
Hospital-eff-A
Hospital-eff-A
Hospital-eff-A
Hospital-eff-A
Hospital-eff-A
Hospital-eff-A
Hospital-eff-A
Hospital-eff-A
Hospital-eff-A
Hospital-eff-A
Hospital-eff-A
Hospital-eff-A
Hospital-eff-A
Hospital-eff-A
Hospital-eff-A
Hospital-eff-A
Hospital-eff-A
Hospital-eff-A
Hospital-eff-A

Hospital-eff-A
Hospital-eff-A
Hospital-eff-A
Hospital-eff-A
Hospital-eff-A
Hospital-eff-A
Hospital-eff-A
Hospital-eff-A
Hospital-eff-A
Hospital-eff-A
Hospital-eff-A
Hospital-eff-A
Hospital-eff-A
Hospital-eff-A
Hospital-eff-A
Hospital-eff-A
Hospital-eff-A
Hospital-eff-A
Hospital-eff-A
Hospital-eff-A
Hospital-eff-A
Hospital-eff-A
Hospital-eff-A
Hospital-eff-A
Hospital-eff-A
Hospital-eff-A
Hospital-eff-A
Hospital-eff-A
Hospital-eff-A
Hospital-eff-A
Hospital-eff-A
Hospital-eff-A
Hospital-eff-A
Hospital-eff-A
Hospital-eff-A
Hospital-eff-A
Hospital-eff-A
Hospital-eff-A
Hospital-eff-A
Hospital-eff-A
Hospital-eff-A
Hospital-eff-A
Hospital-eff-A
Hospital-eff-A
Hospital-eff-A
Hospital-eff-A

Hospital-eff-A
Hospital-eff-A
Hospital-eff-A
STP-inf-F
STP-inf-F
STP-inf-F
STP-inf-F
STP-inf-F
STP-inf-F
STP-inf-F
STP-inf-F
STP-inf-F
STP-inf-F
STP-inf-F
STP-inf-F
STP-inf-F
STP-inf-F
STP-inf-F
STP-inf-F
STP-inf-F
STP-inf-F
STP-inf-F
STP-inf-F
STP-inf-F
STP-inf-F
STP-inf-F
STP-inf-F
STP-inf-F
STP-inf-F
STP-inf-F
STP-inf-F
STP-inf-F
STP-inf-F
STP-inf-F
STP-inf-F
STP-inf-F
STP-inf-F
STP-inf-F
STP-inf-F
STP-inf-F
STP-inf-F
STP-inf-F
STP-inf-F
STP-inf-F
STP-inf-F
STP-inf-F

STP-inf-F
STP-inf-F
STP-inf-F
STP-inf-F
STP-inf-F
STP-inf-F
STP-inf-F
STP-inf-F
STP-inf-F
STP-inf-F
STP-inf-F
STP-inf-F
STP-inf-F
STP-inf-F
STP-inf-F
STP-inf-F
STP-inf-F
STP-inf-F
STP-inf-F
STP-inf-F
STP-inf-F
STP-inf-F
STP-inf-F
STP-inf-F
STP-inf-F
STP-inf-F
STP-inf-F
STP-inf-F
STP-inf-F
STP-inf-F
STP-inf-F
STP-inf-F
STP-inf-F
STP-inf-F
STP-inf-F
STP-inf-F
STP-inf-F
STP-inf-F
STP-inf-F
STP-inf-F
STP-inf-F
STP-inf-F
STP-inf-F
STP-inf-F
STP-inf-F
STP-inf-F

STP-inf-F
STP-inf-F
STP-inf-F
STP-inf-F
STP-inf-F
STP-inf-F
STP-inf-F
STP-inf-F
STP-inf-F
STP-inf-F
STP-inf-F
STP-inf-F
STP-inf-F
STP-inf-F
STP-inf-F
STP-inf-F
STP-inf-F
STP-inf-F
STP-inf-F
STP-inf-F
STP-inf-F
STP-inf-F
STP-inf-F
STP-inf-F
STP-inf-F
STP-inf-F
STP-inf-F
STP-inf-F
STP-inf-F
STP-inf-F
STP-inf-F
STP-inf-F
STP-inf-F
STP-inf-F
STP-inf-F
STP-inf-F
STP-inf-F
STP-inf-F
STP-inf-F
STP-inf-F
STP-inf-F
STP-inf-F
STP-inf-F
STP-inf-F
STP-inf-F
STP-inf-F

STP-inf-F
STP-inf-F
STP-inf-F
STP-inf-F
STP-inf-F
STP-inf-F
STP-inf-F
STP-inf-F
STP-inf-F
STP-inf-F
STP-inf-F
STP-inf-F
STP-inf-F
STP-inf-F
STP-inf-F
STP-inf-F
STP-inf-F
STP-inf-F
STP-inf-F
STP-inf-F
STP-inf-F
STP-inf-F
STP-inf-F
STP-inf-F
STP-inf-F
STP-inf-F
STP-inf-F
STP-inf-F
STP-inf-F
STP-inf-F
STP-inf-F
STP-inf-F
STP-inf-F
STP-inf-F
STP-inf-F
STP-inf-F
STP-inf-F
STP-inf-F
STP-inf-F
STP-inf-F
STP-inf-F
STP-inf-F
STP-inf-F
STP-inf-F
STP-inf-F
STP-inf-F

STP-inf-F
STP-inf-F
STP-inf-F
STP-inf-F
STP-inf-F
STP-inf-F
STP-inf-F
STP-inf-F
STP-inf-F
STP-inf-F
STP-inf-F
STP-inf-F
STP-inf-F
STP-inf-F
STP-inf-F
STP-inf-F
STP-inf-F
STP-inf-F
STP-inf-A
STP-inf-A
STP-inf-A
STP-inf-A
STP-inf-A
STP-inf-A
STP-inf-A
STP-inf-A
STP-inf-A
STP-inf-A
STP-inf-A
STP-inf-A
STP-inf-A
STP-inf-A
STP-inf-A
STP-inf-A
STP-inf-A
STP-inf-A
STP-inf-A
STP-inf-A
STP-inf-A
STP-inf-A
STP-inf-A
STP-inf-A
STP-inf-A
STP-inf-A
STP-inf-A
STP-inf-A

STP-inf-A
STP-inf-A
STP-inf-A
STP-inf-A
STP-inf-A
STP-inf-A
STP-inf-A
STP-inf-A
STP-inf-A
STP-inf-A
STP-inf-A
STP-inf-A
STP-inf-A
STP-inf-A
STP-inf-A
STP-inf-A
STP-inf-A
STP-inf-A
STP-inf-A
STP-inf-A
STP-inf-A
STP-inf-A
STP-inf-A
STP-inf-A
STP-inf-A
STP-inf-A
STP-inf-A
STP-inf-A
STP-inf-A
STP-inf-A
STP-inf-A
STP-inf-A
STP-inf-A
STP-inf-A
STP-inf-A
STP-inf-A
STP-inf-A
STP-inf-A
STP-inf-A
STP-inf-A
STP-inf-A
STP-inf-A
STP-inf-A
STP-inf-A
STP-inf-A
STP-inf-A

STP-inf-A
STP-inf-A
STP-inf-A
STP-inf-A
STP-inf-A
STP-inf-A
STP-inf-A
STP-inf-A
STP-inf-A
STP-inf-A
STP-inf-A
STP-inf-A
STP-inf-A
STP-inf-A
STP-inf-A
STP-inf-A
STP-inf-A
STP-inf-A
STP-inf-A
STP-inf-A
STP-inf-A
STP-inf-A
STP-inf-A
STP-inf-A
STP-inf-A
STP-inf-A
STP-inf-A
STP-inf-A
STP-inf-A
STP-inf-A
STP-inf-A
STP-inf-A
STP-inf-A
STP-inf-A
STP-inf-A
STP-inf-A
STP-inf-A
STP-inf-A
STP-inf-A
STP-inf-A
STP-inf-A
STP-inf-A
STP-inf-A
STP-inf-A
STP-inf-A
STP-inf-A

STP-inf-A
STP-inf-A
STP-inf-A
STP-inf-A
STP-inf-A
STP-inf-A
STP-inf-A
STP-inf-A
STP-inf-A
STP-inf-A
STP-inf-A
STP-inf-A
STP-inf-A
STP-inf-A
STP-inf-A
STP-inf-A
STP-inf-A
STP-inf-A
STP-inf-A
STP-inf-A
STP-inf-A
STP-inf-A
STP-inf-A
STP-inf-A
STP-inf-A
STP-inf-A
STP-inf-A
STP-inf-A
STP-inf-A
STP-inf-A
STP-inf-A
STP-inf-A
STP-inf-A
STP-inf-A
STP-inf-A
STP-inf-A
STP-inf-A
STP-inf-A
STP-inf-A
STP-inf-A
STP-inf-A
STP-inf-A
STP-inf-A
STP-inf-A
STP-inf-A
STP-inf-A

STP-inf-A
STP-inf-A
STP-inf-A
STP-inf-A
STP-inf-A
STP-inf-A
STP-inf-A
STP-inf-A
STP-inf-A
STP-inf-A
STP-inf-A
STP-inf-A
STP-inf-A
STP-inf-A
STP-inf-A
STP-inf-A
STP-inf-A
STP-inf-A
STP-inf-A
STP-inf-A
STP-inf-A
STP-inf-A
STP-inf-A
STP-inf-A
STP-inf-A
STP-inf-A
STP-inf-A
STP-inf-A
STP-inf-A
STP-inf-A
STP-inf-A
STP-inf-A
STP-inf-A
STP-inf-A
STP-inf-A
STP-inf-A
STP-inf-A
STP-inf-A
STP-inf-A
STP-inf-A
STP-inf-A
STP-inf-A
STP-inf-A
STP-T-eff-F
STP-T-eff-F
STP-T-eff-F

STP-T-eff-F
STP-T-eff-F
STP-T-eff-F
STP-T-eff-F
STP-T-eff-F
STP-T-eff-F
STP-T-eff-F
STP-T-eff-F
STP-T-eff-F
STP-T-eff-F
STP-T-eff-F
STP-T-eff-F
STP-T-eff-F
STP-T-eff-F
STP-T-eff-F
STP-T-eff-F
STP-T-eff-F
STP-T-eff-F
STP-T-eff-F
STP-T-eff-F
STP-T-eff-F
STP-T-eff-F
STP-T-eff-F
STP-T-eff-F
STP-T-eff-F
STP-T-eff-F
STP-T-eff-F
STP-T-eff-F
STP-T-eff-F
STP-T-eff-F
STP-T-eff-F
STP-T-eff-F
STP-T-eff-F
STP-T-eff-F
STP-T-eff-F
STP-T-eff-F
STP-T-eff-F
STP-T-eff-F
STP-T-eff-F
STP-T-eff-F
STP-T-eff-F
STP-T-eff-F
STP-T-eff-F
STP-T-eff-F
STP-T-eff-F
STP-T-eff-F

STP-T-eff-F
STP-T-eff-F
STP-T-eff-F
STP-T-eff-F
STP-T-eff-F
STP-T-eff-F
STP-T-eff-F
STP-T-eff-F
STP-T-eff-F
STP-T-eff-F
STP-T-eff-F
STP-T-eff-F
STP-T-eff-F
STP-T-eff-F
STP-T-eff-F
STP-T-eff-F
STP-T-eff-F
STP-T-eff-F
STP-T-eff-F
STP-T-eff-F
STP-T-eff-F
STP-T-eff-F
STP-T-eff-F
STP-T-eff-F
STP-T-eff-F
STP-T-eff-F
STP-T-eff-F
STP-T-eff-F
STP-T-eff-F
STP-T-eff-F
STP-T-eff-F
STP-T-eff-F
STP-T-eff-F
STP-T-eff-F
STP-T-eff-F
STP-T-eff-F
STP-T-eff-F
STP-T-eff-F
STP-T-eff-F
STP-T-eff-F
STP-T-eff-F
STP-T-eff-F
STP-T-eff-F
STP-T-eff-F
STP-T-eff-F
STP-T-eff-F

STP-T-eff-F
STP-T-eff-F
STP-T-eff-F
STP-T-eff-F
STP-T-eff-F
STP-T-eff-F
STP-T-eff-F
STP-T-eff-F
STP-T-eff-F
STP-T-eff-F
STP-T-eff-F
STP-T-eff-F
STP-T-eff-F
STP-T-eff-F
STP-T-eff-F
STP-T-eff-F
STP-T-eff-F
STP-T-eff-F
STP-T-eff-F
STP-T-eff-F
STP-T-eff-F
STP-T-eff-F
STP-T-eff-F
STP-T-eff-F
STP-T-eff-F
STP-T-eff-F
STP-T-eff-F
STP-T-eff-F
STP-T-eff-F
STP-T-eff-F
STP-T-eff-F
STP-T-eff-F
STP-T-eff-F
STP-T-eff-F
STP-T-eff-F
STP-T-eff-F
STP-T-eff-F
STP-T-eff-F
STP-T-eff-F
STP-T-eff-F
STP-T-eff-F
STP-T-eff-F
STP-T-eff-F
STP-T-eff-F
STP-T-eff-F
STP-T-eff-F

STP-T-eff-F
STP-T-eff-F
STP-T-eff-F
STP-T-eff-F
STP-T-eff-F
STP-T-eff-F
STP-T-eff-F
STP-T-eff-F
STP-T-eff-F
STP-T-eff-F
STP-T-eff-F
STP-T-eff-F
STP-T-eff-F
STP-T-eff-F
STP-T-eff-F
STP-T-eff-F
STP-T-eff-F
STP-T-eff-F
STP-T-eff-F
STP-T-eff-F
STP-T-eff-F
STP-T-eff-F
STP-T-eff-F
STP-T-eff-F
STP-T-eff-F
STP-T-eff-F
STP-T-eff-F
STP-T-eff-F
STP-T-eff-F
STP-T-eff-F
STP-T-eff-F
STP-T-eff-F
STP-T-eff-F
STP-T-eff-F
STP-T-eff-F
STP-T-eff-F
STP-T-eff-F
STP-T-eff-F
STP-T-eff-F
STP-T-eff-F
STP-T-eff-F
STP-T-eff-F
STP-T-eff-F
STP-T-eff-F
STP-T-eff-F
STP-T-eff-F

STP-T-eff-F
STP-T-eff-F
STP-T-eff-F
STP-T-eff-F
STP-T-eff-F
STP-T-eff-F
STP-T-eff-F
STP-T-eff-A
STP-T-eff-A
STP-T-eff-A
STP-T-eff-A
STP-T-eff-A
STP-T-eff-A
STP-T-eff-A
STP-T-eff-A
STP-T-eff-A
STP-T-eff-A
STP-T-eff-A
STP-T-eff-A
STP-T-eff-A
STP-T-eff-A
STP-T-eff-A
STP-T-eff-A
STP-T-eff-A
STP-T-eff-A
STP-T-eff-A
STP-T-eff-A
STP-T-eff-A
STP-T-eff-A
STP-T-eff-A
STP-T-eff-A
STP-T-eff-A
STP-T-eff-A
STP-T-eff-A
STP-T-eff-A
STP-T-eff-A
STP-T-eff-A
STP-T-eff-A
STP-T-eff-A
STP-T-eff-A
STP-T-eff-A
STP-T-eff-A
STP-T-eff-A
STP-T-eff-A
STP-T-eff-A
STP-T-eff-A

STP-T-eff-A
STP-T-eff-A
STP-T-eff-A
STP-T-eff-A
STP-T-eff-A
STP-T-eff-A
STP-T-eff-A
STP-T-eff-A
STP-T-eff-A
STP-T-eff-A
STP-T-eff-A
STP-T-eff-A
STP-T-eff-A
STP-T-eff-A
STP-T-eff-A
STP-T-eff-A
STP-T-eff-A
STP-T-eff-A
STP-T-eff-A
STP-T-eff-A
STP-T-eff-A
STP-T-eff-A
STP-T-eff-A
STP-T-eff-A
STP-T-eff-A
STP-T-eff-A
STP-T-eff-A
STP-T-eff-A
STP-T-eff-A
STP-T-eff-A
STP-T-eff-A
STP-T-eff-A
STP-T-eff-A
STP-T-eff-A
STP-T-eff-A
STP-T-eff-A
STP-T-eff-A
STP-T-eff-A
STP-T-eff-A
STP-T-eff-A
STP-T-eff-A
STP-T-eff-A
STP-T-eff-A
STP-T-eff-A
STP-T-eff-A
STP-T-eff-A

STP-T-eff-A
STP-T-eff-A
STP-T-eff-A
STP-T-eff-A
STP-T-eff-A
STP-T-eff-A
STP-T-eff-A
STP-T-eff-A
STP-T-eff-A
STP-T-eff-A
STP-T-eff-A
STP-T-eff-A
STP-T-eff-A
STP-T-eff-A
STP-T-eff-A
STP-T-eff-A
STP-T-eff-A
STP-T-eff-A
STP-T-eff-A
STP-T-eff-A
STP-T-eff-A
STP-T-eff-A
STP-T-eff-A
STP-T-eff-A
STP-T-eff-A
STP-T-eff-A
STP-T-eff-A
STP-T-eff-A
STP-T-eff-A
STP-T-eff-A
STP-T-eff-A
STP-T-eff-A
STP-T-eff-A
STP-T-eff-A
STP-T-eff-A
STP-T-eff-A
STP-T-eff-A
STP-T-eff-A
STP-T-eff-A
STP-T-eff-A
STP-T-eff-A
STP-T-eff-A
STP-T-eff-A
STP-T-eff-A
STP-T-eff-A
STP-T-eff-A

STP-T-eff-A
STP-T-eff-A
STP-T-eff-A
STP-T-eff-A
STP-T-eff-A
STP-T-eff-A
STP-T-eff-A
STP-T-eff-A
STP-T-eff-A
STP-T-eff-A
STP-T-eff-A
STP-T-eff-A
STP-T-eff-A
STP-T-eff-A
STP-T-eff-A
STP-T-eff-A
STP-T-eff-A
STP-T-eff-A
STP-T-eff-A
STP-T-eff-A
STP-T-eff-A
STP-T-eff-A
STP-T-eff-A
STP-T-eff-A
STP-T-eff-A
STP-T-eff-A
STP-T-eff-A
STP-T-eff-A
STP-T-eff-A
STP-T-eff-A
STP-T-eff-A
STP-T-eff-A
STP-T-eff-A
STP-T-eff-A
STP-T-eff-A
STP-T-eff-A
STP-T-eff-A
STP-T-eff-A
STP-T-eff-A
STP-T-eff-A
STP-T-eff-A
STP-T-eff-A
STP-T-eff-A
STP-T-eff-A
STP-T-eff-A
STP-T-eff-A

| |
|---|
| STP-T-eff-A |
| STP-T-eff-A |
| STP-T-eff-A |
| STP-T-eff-A |
| STP-T-eff-A |
| STP-T-eff-A |
| STP-T-eff-A |
| STP-T-eff-A |
| STP-T-eff-A |
| STP-T-eff-A |
| STP-T-eff-A |
| STP-T-eff-A |
| STP-T-eff-A |
| STP-T-eff-A |
| STP-T-eff-A |
| STP-T-eff-A |
| STP-T-eff-A |
| STP-T-eff-A |
| STP-T-eff-A |
| STP-T-eff-A |
| STP-T-eff-A |
| STP-T-eff-A |
| STP-T-eff-A |
| STP-T-eff-A |
| STP-T-eff-A |
| STP-T-eff-A |
| STP-T-eff-A |
| STP-T-eff-A |
| STP-T-eff-A |
| STP-T-eff-A |
| STP-T-eff-A |
| STP-T-eff-A |
| STP-T-eff-A |
| STP-T-eff-A |
| STP-T-eff-A |
| STP-T-eff-A |
| STP-T-eff-A |
| STP-T-eff-A |
| STP-T-eff-A |
| STP-T-eff-A |

**Legend:** eff (effluent); F (Fe[illegible]

Detected biocide/metal resistance genes (BMRGs) across all samples.

| |
|---|
| BAC0688\|merR2\|tr\|Q79B70\|Q79B70_PSEST Organomercurial resistance regulatory protein OS=Pseudomonas stutzeri GN |
| BAC0648\|merA\|sp\|P08662\|MERA_SERMA Mercuric reductase (Fragments) OS=Serratia marcescens GN=merA PE=3 SV |
| BAC0324\|qacF\|sp\|Q9X2N9\|QACF_ENTAE Quaternary ammonium compound-resistance protein QacF OS=Enterobacter a |
| BAC0322\|qacE\|sp\|P0AGC9\|QACE_ECOLX Quaternary ammonium compound-resistance protein QacE OS=Escherichia co |
| BAC0018\|adeJ\|tr\|Q24LT7\|Q24LT7_ACIBA AdeJ OS=Acinetobacter baumannii GN=adeJ PE=4 SV=1 |
| BAC0323\|qacEdelta1\|tr\|Q7BQY4\|Q7BQY4_PSEAI Disinfectant resistance protein OS=Pseudomonas aeruginosa GN=qacE |
| BAC0019\|adeK\|tr\|Q24LT6\|Q24LT6_ACIBA AdeK OS=Acinetobacter baumannii GN=adeK PE=4 SV=1 |
| BAC0335\|rpoS\|sp\|P35540\|RPOS_SHIFL RNA polymerase sigma factor RpoS OS=Shigella flexneri GN=rpoS PE=3 SV=3 |
| BAC0533\|cpxR\|tr\|C4WZK6\|C4WZK6_KLEPN Response regulator of stress-related two-component regulatory system OS= |
| BAC0536\|oxyRkp\|tr\|C4WZN6\|C4WZN6_KLEPN Activator of hydrogen peroxide-inducible genes OS=Klebsiella pneumon |
| BAC0532\|cpxA\|tr\|C4WZK5\|C4WZK5_KLEPN Sensor protein of stress-related two-component regulatory system OS=Kle |
| BAC0194\|ibpA\|sp\|P0C054\|IBPA_ECOLI Small heat shock protein IbpA OS=Escherichia coli (strain K12) GN=ibpA PE=1 |
| BAC0564\|actP/yjcG\|sp\|P32705\|ACTP_ECOLI Cation/acetate symporter ActP OS=Escherichia coli (strain K12) GN=actP |
| BAC0017\|adeI\|tr\|Q2FD95\|Q2FD95_ACIBA AdeI OS=Acinetobacter baumannii GN=adeI PE=4 SV=1 |
| BAC0368\|sodA\|sp\|P00448\|SODM_ECOLI Superoxide dismutase [Mn] OS=Escherichia coli (strain K12) GN=sodA PE=1 |
| BAC0001\|abeM\|tr\|Q5FAM9\|Q5FAM9_ACIBA Multidrug efflux pump AbeM OS=Acinetobacter baumannii GN=abeM PE |
| BAC0211\|mdtB/yegN\|sp\|P76398\|MDTB_ECOLI Multidrug resistance protein MdtB OS=Escherichia coli (strain K12) GN |
| BAC0559\|emrR\|sp\|P0ACR9\|MPRA_ECOLI Transcriptional repressor MprA OS=Escherichia coli (strain K12) GN=mprA |
| BAC0047\|bexA\|tr\|Q93HR0\|Q93HR0_BACT4 BexA OS=Bacteroides thetaiotaomicron GN=bexA PE=4 SV=1 |
| BAC0061\|cepA\|sp\|Q8RR17\|FIEF_KLEPN Cation-efflux pump FieF OS=Klebsiella pneumoniae GN=fieF PE=3 SV=1 |
| BAC0006\|acrB\|sp\|P31224\|ACRB_ECOLI Multidrug efflux pump subunit AcrB OS=Escherichia coli (strain K12) GN=acrB |
| BAC0196\|iclR\|sp\|P16528\|ICLR_ECOLI Acetate operon repressor OS=Escherichia coli (strain K12) GN=iclR PE=1 SV=1 |
| BAC0357\|recG\|tr\|B5L350\|B5L350_9PSED ATP-dependent DNA helicase (Fragment) OS=Pseudomonas corrugata GN=rec |
| BAC0364\|smrA\|tr\|C7SLZ1\|C7SLZ1_STEMA ABC-type multidrug efflux pump (Fragment) OS=Stenotrophomonas maltop |
| BAC0472\|adeB\|tr\|Q93E19\|Q93E19_ACIBA AdeB RND protein OS=Acinetobacter baumannii GN=adeB PE=4 SV=1 |
| BAC0370\|soxR\|sp\|P0ACS2\|SOXR_ECOLI Redox-sensitive transcriptional activator SoxR OS=Escherichia coli (strain K12 |
| BAC0530\|phoB\|tr\|C4X6T6\|C4X6T6_KLEPN Response regulator in two-component regulatory system with PhoQ OS=Kle |
| BAC0156\|fabI\|sp\|P0AEK4\|FABI_ECOLI Enoyl-[acyl-carrier-protein] reductase [NADH] FabI OS=Escherichia coli (strain |
| BAC0148\|emrB\|sp\|P0AEJ0\|EMRB_ECOLI Multidrug resistance protein B OS=Escherichia coli (strain K12) GN=emrB PE |
| BAC0295\|oqxB\|tr\|Q69HW2\|Q69HW2_ECOLX OqxB integral membrane protein OS=Escherichia coli GN=oqxB PE=4 SV |
| BAC0039\|baeR\|sp\|P69228\|BAER_ECOLI Transcriptional regulatory protein BaeR OS=Escherichia coli (strain K12) GN=b |
| BAC0008\|acrD/yffA\|sp\|P24177\|ACRD_ECOLI Probable aminoglycoside efflux pump OS=Escherichia coli (strain K12) G |
| BAC0149\|emrD\|sp\|P31442\|EMRD_ECOLI Multidrug resistance protein D OS=Escherichia coli (strain K12) GN=emrD PE |
| BAC0494\|eefA\|tr\|A8CY69\|A8CY69_KLEPN EefA OS=Klebsiella pneumoniae GN=eefA PE=4 SV=1 |
| BAC0541\|yieF\|sp\|P0AGE6\|YIEF_ECOLI Uncharacterized protein YieF OS=Escherichia coli (strain K12) GN=yieF PE=1 |
| BAC0242\|mexK\|tr\|Q9HXW4\|Q9HXW4_PSEAE Probable Resistance-Nodulation-Cell Division (RND) efflux transporter |
| BAC0181\|glpF\|sp\|P0AER0\|GLPF_ECOLI Glycerol uptake facilitator protein OS=Escherichia coli (strain K12) GN=glpF P |
| BAC0212\|mdtC/yegO\|sp\|P76399\|MDTC_ECOLI Multidrug resistance protein MdtC OS=Escherichia coli (strain K12) GN |
| BAC0393\|tolC\|sp\|P02930\|TOLC_ECOLI Outer membrane protein TolC OS=Escherichia coli (strain K12) GN=tolC PE=1 |
| BAC0015\|adeG\|tr\|Q2FD81\|Q2FD81_ACIBA Cation/multidrug efflux pump OS=Acinetobacter baumannii GN=29_167 PE |
| BAC0371\|soxS\|sp\|P0A9E2\|SOXS_ECOLI Regulatory protein SoxS OS=Escherichia coli (strain K12) GN=soxS PE=1 SV |
| BAC0294\|oqxA\|tr\|Q69HW3\|Q69HW3_ECOLX OqxA membrane-fusion protein OS=Escherichia coli GN=oqxA PE=4 SV |
| BAC0560\|marA\|sp\|P0ACH5\|MARA_ECOLI Multiple antibiotic resistance protein MarA OS=Escherichia coli (strain K12) |

BAC0334|robA|sp|P0ACI0|ROB_ECOLI Right origin-binding protein OS=Escherichia coli (strain K12) GN=rob PE=1 SV
BAC0491|kdeA|tr|A6T6T9|A6T6T9_KLEP7 Multidrug/chloramphenicol efflux transport protein (MFS family) OS=Klebsi
BAC0529|kpnO|tr|C4XBC3|C4XBC3_KLEPN Outer membrane porin protein C OS=Klebsiella pneumoniae subsp. pneumo
BAC0707|sodB|sp|P0AGD3|SODF_ECOLI Superoxide dismutase [Fe] OS=Escherichia coli (strain K12) GN=sodB PE=1 S
BAC0351|sitC|tr|Q9XCS0|Q9XCS0_SALTM SitC OS=Salmonella typhimurium GN=sitC PE=3 SV=1
BAC0010|acrF/envD|sp|P24181|ACRF_ECOLI Acriflavine resistance protein F OS=Escherichia coli (strain K12) GN=acrF
BAC0172|gadA|sp|P69908|DCEA_ECOLI Glutamate decarboxylase alpha OS=Escherichia coli (strain K12) GN=gadA PE
BAC0218|mdtK/ydhE|sp|P37340|MDTK_ECOLI Multidrug resistance protein MdtK OS=Escherichia coli (strain K12) GN
BAC0195|ibpB|sp|P0C058|IBPB_ECOLI Small heat shock protein IbpB OS=Escherichia coli (strain K12) GN=ibpB PE=1
BAC0041|bcr|sp|P28246|BCR_ECOLI Bicyclomycin resistance protein OS=Escherichia coli (strain K12) GN=bcr PE=1 S
BAC0531|phoR|tr|C4X6T5|C4X6T5_KLEPN Sensor kinase in two-component regulatory system with PhoP OS=Klebsiella
BAC0378|sugE|sp|P69937|SUGE_ECOLI Quaternary ammonium compound-resistance protein SugE OS=Escherichia coli
BAC0210|mdtA/yegM|sp|P76397|MDTA_ECOLI Multidrug resistance protein MdtA OS=Escherichia coli (strain K12) GN
BAC0215|mdtG/yceE|sp|P25744|MDTG_ECOLI Multidrug resistance protein MdtG OS=Escherichia coli (strain K12) GN
BAC0296|ostA/lptD|sp|P31554|LPTD_ECOLI LPS-assembly protein LptD OS=Escherichia coli (strain K12) GN=lptD PE
BAC0005|acrA|sp|P0AE06|ACRA_ECOLI Multidrug efflux pump subunit AcrA OS=Escherichia coli (strain K12) GN=ac
BAC0185|hdeA/yhiB|sp|P0AES9|HDEA_ECOLI Acid stress chaperone HdeA OS=Escherichia coli (strain K12) GN=hdeA
BAC0237|mexD|tr|Q51396|Q51396_PSEAI RND family exporter MexD OS=Pseudomonas aeruginosa GN=mexD PE=4 S
BAC0434|ychH|sp|P0AB49|YCHH_ECOLI Uncharacterized protein YchH OS=Escherichia coli (strain K12) GN=ychH PE
BAC0493|kmrA|tr|C4X8X9|C4X8X9_KLEPN Energy-dependent efflux protein for methyl viologen resistance OS=Klebsie
BAC0496|adeN|tr|B7H1T7|B7H1T7_ACIB3 Bacterial regulatory protein, tetR family protein OS=Acinetobacter baumanni
BAC0011|acrR/ybaH|sp|P0ACS9|ACRR_ECOLI HTH-type transcriptional regulator AcrR OS=Escherichia coli (strain K1
BAC0445|ygiW|sp|P0ADU5|YGIW_ECOLI Protein YgiW OS=Escherichia coli (strain K12) GN=ygiW PE=1 SV=1
BAC0166|fetB/ybbM|sp|P77307|YBBM_ECOLI UPF0014 inner membrane protein YbbM OS=Escherichia coli (strain K1
BAC0186|hdeB/yhiC|sp|P0AET2|HDEB_ECOLI Acid stress chaperone HdeB OS=Escherichia coli (strain K12) GN=hdeB
BAC0565|actR|sp|A6UEL7|ACTR_SINMW Acid tolerance regulatory protein ActR OS=Sinorhizobium medicae (strain W
BAC0220|mdtN/yjcR|sp|P32716|MDTN_ECOLI Multidrug resistance protein MdtN OS=Escherichia coli (strain K12) GN
BAC0214|mdtF/yhiV|sp|P37637|MDTF_ECOLI Multidrug resistance protein MdtF OS=Escherichia coli (strain K12) GN
BAC0477|kpnF|tr|C4X7Z4|C4X7Z4_KLEPN Spermidine export protein MdtI OS=Klebsiella pneumoniae subsp. pneumoni
BAC0134|dpr/dps|sp|P0CB53|DPS_STRSU DNA protection during starvation protein OS=Streptococcus suis GN=dps PE
BAC0177|gadX|sp|P37639|GADX_ECOLI HTH-type transcriptional regulator GadX OS=Escherichia coli (strain K12) GN
BAC0471|adeA|tr|Q93E20|Q93E20_ACIBA AdeA membrane fusion protein OS=Acinetobacter baumannii GN=adeA PE
BAC0384|tehA|sp|P25396|TEHA_ECOLI Tellurite resistance protein TehA OS=Escherichia coli (strain K12) GN=tehA PE
BAC0040|baeS|sp|P30847|BAES_ECOLI Signal transduction histidine-protein kinase BaeS OS=Escherichia coli (strain K1
BAC0213|mdtE/yhiU|sp|P37636|MDTE_ECOLI Multidrug resistance protein MdtE OS=Escherichia coli (strain K12) GN
BAC0417|vcaM|tr|Q9KKV4|Q9KKV4_VIBCH ABC transporter, ATP-binding protein OS=Vibrio cholerae serotype O1 (s
BAC0476|kpnE|tr|C4X7Z3|C4X7Z3_KLEPN Multidrug transport protein OS=Klebsiella pneumoniae subsp. pneumoniae N
BAC0176|gadW/yhiW|sp|P63201|GADW_ECOLI HTH-type transcriptional regulator GadW OS=Escherichia coli (strain K
BAC0009|acrE/envC|sp|P24180|ACRE_ECOLI Acriflavine resistance protein E OS=Escherichia coli (strain K12) GN=acrE
BAC0147|emrA|sp|P27303|EMRA_ECOLI Multidrug resistance protein A OS=Escherichia coli (strain K12) GN=emrA PE
BAC0175|gadE/yhiE|sp|P63204|GADE_ECOLI Transcriptional regulator GadE OS=Escherichia coli (strain K12) GN=gad
BAC0385|tehB|sp|P25397|TEHB_ECOLI Tellurite methyltransferase OS=Escherichia coli (strain K12) GN=tehB PE=1 SV
BAC0495|eefX|tr|A8CY68|A8CY68_KLEPN EefX OS=Klebsiella pneumoniae GN=eefX PE=4 SV=1
BAC0492|kexD|tr|A6TA71|A6TA71_KLEP7 Acridine efflux pump OS=Klebsiella pneumoniae subsp. pneumoniae (strain
BAC0451|yodD|sp|P64519|YODD_ECOLI Uncharacterized protein YodD OS=Escherichia coli (strain K12) GN=yodD PE
BAC0293|ruvB|tr|B5L348|B5L348_9PSED Malic enzyme family protein (Fragment) OS=Pseudomonas corrugata PE=3 SV

BAC0446|yhcN|sp|P64614|YHCN_ECOLI Uncharacterized protein YhcN OS=Escherichia coli (strain K12) GN=yhcN PE=
BAC0219|mdtM/yjiO|sp|P39386|MDTM_ECOLI Multidrug resistance protein MdtM OS=Escherichia coli (strain K12) GN
BAC0208|mdfA/cmr|sp|P0AEY8|MDFA_ECOLI Multidrug transporter MdfA OS=Escherichia coli (strain K12) GN=mdfA
BAC0146|emmdR|tr|D5CJ69|D5CJ69_ENTCC MATE efflux family protein OS=Enterobacter cloacae subsp. cloacae (strai
BAC0561|marR|sp|P27245|MARR_ECOLI Multiple antibiotic resistance protein MarR OS=Escherichia coli (strain K12) G
BAC0706|sodB|sp|P53641|SODF_PSEAE Superoxide dismutase [Fe] OS=Pseudomonas aeruginosa (strain ATCC 15692 /
BAC0252|mntP/yebN|sp|P76264|MNTP_ECOLI Probable manganese efflux pump MntP OS=Escherichia coli (strain K12)
BAC0151|emrK|sp|P52599|EMRK_ECOLI Multidrug resistance protein K OS=Escherichia coli (strain K12) GN=emrK PE
BAC0153|emrY|sp|P52600|EMRY_ECOLI Multidrug resistance protein Y OS=Escherichia coli (strain K12) GN=emrY PE
BAC0450|ymgB/ariR|sp|P75993|ARIR_ECOLI Probable two-component-system connector protein AriR OS=Escherichia c
BAC0154|evgA|sp|P0ACZ4|EVGA_ECOLI Positive transcription regulator EvgA OS=Escherichia coli (strain K12) GN=ev
BAC0155|evgS|sp|P58402|EVGS_ECO57 Sensor protein EvgS OS=Escherichia coli O157:H7 GN=evgS PE=3 SV=1
BAC0217|mdtJ/ebrB/ydgF|sp|Q3Z1V3|MDTJ_SHISS Spermidine export protein MdtJ OS=Shigella sonnei (strain Ss046) G
BAC0165|fetA/ybbL|sp|P77279|YBBL_ECOLI Uncharacterized ABC transporter ATP-binding protein YbbL OS=Escheric
BAC0350|sitB|tr|Q9XCS1|Q9XCS1_SALTM SitB OS=Salmonella typhimurium GN=sitB PE=3 SV=1
BAC0659|merB|sp|P08664|MERB_SERMA Alkylmercury lyase OS=Serratia marcescens GN=merB PE=3 SV=1
BAC0216|mdtI/ydgE|sp|P69210|MDTI_ECOLI Spermidine export protein MdtI OS=Escherichia coli (strain K12) GN=mdt
BAC0353|smdA|tr|A7VN01|A7VN01_SERMA Multidrug efflux pump SmdA OS=Serratia marcescens GN=smdA PE=3 S
BAC0359|smeE|tr|I0KSX8|I0KSX8_STEMA RND efflux system, inner membrane transporter OS=Stenotrophomonas malt
BAC0436|ydeI|sp|P31130|YDEI_ECOLI Uncharacterized protein YdeI OS=Escherichia coli (strain K12) GN=ydeI PE=4 S
BAC0447|yjaA|sp|P09162|YJAA_ECOLI Uncharacterized protein YjaA OS=Escherichia coli (strain K12) GN=yjaA PE=4
BAC0174|gadC/xasA|sp|P63235|GADC_ECOLI Probable glutamate/gamma-aminobutyrate antiporter OS=Escherichia coli
BAC0150|emrE/mvrC|sp|P23895|EMRE_ECOLI Multidrug transporter EmrE OS=Escherichia coli (strain K12) GN=emrE
BAC0438|ydeP|sp|P77561|YDEP_ECOLI Protein YdeP OS=Escherichia coli (strain K12) GN=ydeP PE=2 SV=1
BAC0106|cuiD|sp|Q8ZRS2|CUEO_SALTY Blue copper oxidase CueO OS=Salmonella typhimurium (strain LT2 / SGSC1
BAC0437|ydeO|sp|P76135|YDEO_ECOLI HTH-type transcriptional regulator YdeO OS=Escherichia coli (strain K12) GN
BAC0349|sitA|tr|Q9XCS2|Q9XCS2_SALTI Iron transport protein, periplasmic-binding protein OS=Salmonella typhi GN=
BAC0135|dpsA|tr|Q8KR86|Q8KR86_BURPE DpsA OS=Burkholderia pseudomallei GN=dpsA PE=3 SV=1
BAC0352|sitD|tr|Q9XCR9|Q9XCR9_SALTM SitD OS=Salmonella typhimurium GN=sitD PE=3 SV=1
BAC0013|adeE|tr|Q8GKU1|Q8GKU1_ACIG3 AdeE OS=Acinetobacter sp. 4365 GN=adeE PE=4 SV=2
BAC0235|mexB|sp|P52002|MEXB_PSEAE Multidrug resistance protein MexB OS=Pseudomonas aeruginosa (strain ATC
BAC0596|baeR|tr|D0ZNE3|D0ZNE3_SALT1 DNA-binding transcriptional regulator BaeR OS=Salmonella typhimurium (s
BAC0337|sdeB|tr|Q84GI9|Q84GI9_SERMA Putative resistance-nodulation cell division protein SdeB OS=Serratia marcesc
BAC0339|sdeY|tr|Q7WSD5|Q7WSD5_SERMA Multidrug efflux pump SdeY OS=Serratia marcescens GN=sdeY PE=4 SV
BAC0511|vmeD|tr|Q87TN1|Q87TN1_VIBPA Putative multidrug resistance protein OS=Vibrio parahaemolyticus serotype
BAC0595|arsH|tr|P74312|P74312_SYNY3 Slr0945 protein OS=Synechocystis sp. (strain PCC 6803 / Kazusa) GN=slr0945
BAC0239|mexF|tr|Q4KBN7|Q4KBN7_PSEF5 Multidrug efflux RND transporter, permease protein MexF OS=Pseudomona
BAC0179|gesB|tr|Q8ZRG9|Q8ZRG9_SALTY Putative cation efflux system protein OS=Salmonella typhimurium (strain L
BAC0038|asr|sp|P36560|ASR_ECOLI Acid shock protein OS=Escherichia coli (strain K12) GN=asr PE=1 SV=3
BAC0424|vexB|tr|Q9KVI2|Q9KVI2_VIBCH Multidrug resistance protein, putative OS=Vibrio cholerae serotype O1 (strai
BAC0012|actP|sp|Q9X5X3|ATCU_SINMW Copper-transporting P-type ATPase OS=Sinorhizobium medicae (strain WSM
BAC0157|fabK|tr|Q9FBC5|Q9FBC5_STREE Trans-2-enoyl-ACP reductase II OS=Streptococcus pneumoniae GN=fabK P
BAC0029|chrF|tr|A4UQR2|A4UQR2_9RHIZ ChrF OS=Ochrobactrum tritici GN=chrF PE=4 SV=1
BAC0143|emhB|tr|C1KA85|C1KA85_PSEFL EmhB OS=Pseudomonas fluorescens GN=emhB PE=4 SV=1
BAC0290|opmD/nmpC|sp|P37592|OMPD_SALTY Outer membrane porin protein OmpD OS=Salmonella typhimurium (st
BAC0650|merA|tr|O08449|O08449_9PSED Mercuric reductase OS=Pseudomonas sp. K-62 GN=merA PE=4 SV=1

BAC0681|merR2|tr|Q9WWL1|Q9WWL1_BACSR Mercury resistance operon negative regulator MerR2 OS=Bacillus sp. (s
BAC0654|merB1|sp|P16172|MERB_BACCE Alkylmercury lyase OS=Bacillus cereus GN=merB1 PE=3 SV=2
BAC0222|mepB|sp|P0C070|MEPB_PSEPU Multidrug/solvent efflux pump membrane transporter MepB OS=Pseudomonas
BAC0435|yddg/emrE|sp|D0ZXP9|YDDG_SALT1 Methyl viologen resistance protein YddG OS=Salmonella typhimurium
BAC0244|mexT|tr|O87785|O87785_PSEAI MexT protein OS=Pseudomonas aeruginosa GN=mexT PE=4 SV=1
BAC0705|sodA|sp|P53652|SODM_PSEAE Superoxide dismutase [Mn] OS=Pseudomonas aeruginosa (strain ATCC 15692
BAC0246|mexW|tr|Q9HW27|Q9HW27_PSEAE Probable Resistance-Nodulation-Cell Division (RND) efflux transporter O
BAC0014|adeF|tr|Q2FD82|Q2FD82_ACIBA Putative RND family drug transporter OS=Acinetobacter baumannii GN=29_
BAC0508|adeL|tr|A3M732|A3M732_ACIBT Transcriptional regulator LysR family OS=Acinetobacter baumannii (strain A
BAC0656|merB3|tr|Q7DHE7|Q7DHE7_BACCE Organomercurial lyase enzyme OS=Bacillus cereus GN=merB3 PE=4 SV
BAC0478|adeT1|tr|C7F8K6|C7F8K6_ACIBA AdeT1 OS=Acinetobacter baumannii PE=4 SV=1
BAC0500|emrBsm|tr|B2FID0|B2FID0_STRMK Putative multidrug resistance protein B OS=Stenotrophomonas maltophili
BAC0240|mexI|tr|Q9HWH4|Q9HWH4_PSEAE Probable Resistance-Nodulation-Cell Division (RND) efflux transporter O
BAC0354|smdB|tr|A7VN02|A7VN02_SERMA Multidrug efflux pump SmdB OS=Serratia marcescens GN=smdB PE=3 S
BAC0661|merB2|tr|Q7DJN2|Q7DJN2_BACME MerB2 OS=Bacillus megaterium GN=merB2 PE=4 SV=1
BAC0238|mexE|tr|Q1IB41|Q1IB41_PSEE4 Multidrug efflux RND membrane fusion protein MexE OS=Pseudomonas ento
BAC0236|mexC|tr|Q51395|Q51395_PSEAI Membrane fusion protein MexC OS=Pseudomonas aeruginosa GN=mexC PE=
BAC0367|smvA/emrB|sp|D0ZXQ3|SMVA_SALT1 Methyl viologen resistance protein SmvA OS=Salmonella typhimuriu
BAC0507|tolCsm|tr|R4ITT0|R4ITT0_STEMA Outer membrane protein OS=Stenotrophomonas maltophilia GN=tolCsm P
BAC0016|adeH|tr|Q2FD80|Q2FD80_ACIBA Putative RND family drug transporter OS=Acinetobacter baumannii GN=29_
BAC0338|sdeX|tr|Q7WSD6|Q7WSD6_SERMA Multidrug efflux pump SdeX OS=Serratia marcescens GN=sdeX PE=4 S
BAC0499|emrAsm|tr|B2FIC9|B2FIC9_STRMK Putative multidrug resistance protein A OS=Stenotrophomonas maltophili
BAC0419|vceB|tr|O51919|O51919_VIBCL VceB OS=Vibrio cholerae GN=vceB PE=4 SV=1
BAC0184|hasF|tr|Q6GW09|Q6GW09_SERMA TolC-like protein OS=Serratia marcescens PE=4 SV=1
BAC0657|merB|tr|O07303|O07303_9PSED Alkylmercury lyase OS=Pseudomonas sp. K-62 GN=merB PE=3 SV=2
BAC0566|actS|tr|Q52912|Q52912_9RHIZ Histidine protein kinase OS=Sinorhizobium medicae GN=actS PE=4 SV=1
BAC0141|emeA|tr|Q8GR72|Q8GR72_ENTFL Multidrug efflux pump OS=Enterococcus faecalis GN=emeA PE=4 SV=1
BAC0044|bepE|sp|Q8G2M6|BEPE_BRUSU Efflux pump membrane transporter BepE OS=Brucella suis biovar 1 (strain 1
BAC0502|emrRsm|tr|B2FIC7|B2FIC7_STRMK Putative MarR-family transcriptional regulator OS=Stenotrophomonas ma
BAC0506|pcm|tr|R4IUI7|R4IUI7_STEMA Protein-L-isoaspartate O-methyltransferase OS=Stenotrophomonas maltophilia
BAC0358|oscA|tr|B6CM35|B6CM35_9PSED Putative uncharacterized protein oscA OS=Pseudomonas corrugata GN=osc
BAC0043|bepD|sp|Q8G2M7|BEPD_BRUSU Efflux pump periplasmic linker BepD OS=Brucella suis biovar 1 (strain 1330
BAC0207|mdeA|sp|P13254|MEGL_PSEPU Methionine gamma-lyase OS=Pseudomonas putida GN=mdeA PE=1 SV=2
BAC0159|fabV|sp|Q9KRA3|Y1738_VIBCH Putative reductase VC_1738/VC_1739 OS=Vibrio cholerae serotype O1 (strai
BAC0145|emhR|tr|Q4KH21|Q4KH21_PSEF5 Transcriptional regulator EmhR OS=Pseudomonas fluorescens (strain Pf-5 /
BAC0404|ttgA|sp|Q9WWZ9|TTGA_PSEPT Toluene efflux pump periplasmic linker protein TtgA OS=Pseudomonas putid
BAC0142|emhA|tr|Q4KH22|Q4KH22_PSEF5 Efflux transporter, membrane fusion protein subunit EmhA OS=Pseudomona
BAC0597|baeS|tr|D0ZNE2|D0ZNE2_SALT1 Signal transduction histidine-protein kinase BaeS OS=Salmonella typhimuri
BAC0027|chrB|tr|A4UQR5|A4UQR5_9RHIZ ChrB OS=Ochrobactrum tritici GN=chrB PE=4 SV=1
BAC0358|smeD|tr|I0KSX9|I0KSX9_STEMA Membrane fusion protein of RND family multidrug efflux pump OS=Stenot
BAC0662|merB3|tr|Q9RHR0|Q9RHR0_BACME MerB3 OS=Bacillus megaterium GN=merB3 PE=4 SV=2
BAC0144|emhC|tr|Q4KH24|Q4KH24_PSEF5 Efflux transporter, outer membrane factor lipoprotein EmhC OS=Pseudomo
BAC0538|chrR|tr|Q7BD45|Q7BD45_PSEPU Chromate reductase OS=Pseudomonas putida GN=chrR PE=4 SV=1
BAC0248|mexY|tr|Q9ZNG8|Q9ZNG8_PSEAI MexY OS=Pseudomonas aeruginosa GN=mexY PE=4 SV=1
BAC0362|smeT|tr|Q5MXJ5|Q5MXJ5_STEMA Repressor SmeT OS=Stenotrophomonas maltophilia GN=smeT PE=4 SV=
BAC0498|ideR|sp|P0A672|IDER_MYCTU Iron-dependent repressor IdeR OS=Mycobacterium tuberculosis GN=ideR PE=

BAC0473|adeC|tr|Q93E18|Q93E18_ACIBA AdeC outer membrane protein OS=Acinetobacter baumannii GN=adeC PE=4
BAC0173|gadB|sp|P69910|DCEB_ECOLI Glutamate decarboxylase beta OS=Escherichia coli (strain K12) GN=gadB PE=
BAC0360|smeF|tr|Q9F239|Q9F239_STEMA Outer membrane protein OS=Stenotrophomonas maltophilia GN=smeF PE=4
BAC0223|mepC|sp|P0C071|MEPC_PSEPU Multidrug/solvent efflux pump outer membrane protein MepC OS=Pseudomon
BAC0292|oprM/oprK|sp|Q51487|OPRM_PSEAE Outer membrane protein OprM OS=Pseudomonas aeruginosa (strain AT
BAC0501|emrCsm|tr|B2FIC8|B2FIC8_STRMK Putative outer membrane multidrug efflux protein OS=Stenotrophomonas
BAC0418|vceA|tr|O51918|O51918_VIBCL VceA OS=Vibrio cholerae GN=vceA PE=4 SV=1
BAC0505|farR|tr|Q7DD70|Q7DD70_NEIMB Transcriptional regulator, MarR family OS=Neisseria meningitidis serogroup
BAC0190|hmrR|sp|Q9X5X4|HMRR_SINMW HTH-type transcriptional regulator HmrR OS=Sinorhizobium medicae (strai
BAC0313|pmpM|sp|Q9I3Y3|PMPM_PSEAE Multidrug resistance protein PmpM OS=Pseudomonas aeruginosa (strain AT
BAC0258|mtrD|tr|Q5F725|Q5F725_NEIG1 Antibiotic resistance efflux pump component OS=Neisseria gonorrhoeae (strai
BAC0028|chrC|tr|A4UQR3|A4UQR3_9RHIZ Superoxide dismutase OS=Ochrobactrum tritici GN=chrC PE=3 SV=1
BAC0411|ttgH|sp|Q93PU4|TTGH_PSEPT Toluene efflux pump membrane transporter TtgH OS=Pseudomonas putida (stra
BAC0375|srpR|sp|Q9R9T9|SRPR_PSEPU HTH-type transcriptional regulator SrpR OS=Pseudomonas putida GN=srpR PE
BAC0046|bepG|sp|Q8FWV9|BEPG_BRUSU Efflux pump membrane transporter BepG OS=Brucella suis biovar 1 (strain 1
BAC0229|merG|tr|O07302|O07302_9PSED Mercuric resistance protein OS=Pseudomonas sp. K-62 GN=merG PE=4 SV=
BAC0481|pdrM|sp|Q8DPQ6|NORM_STRR6 Probable multidrug resistance protein NorM OS=Streptococcus pneumoniae (
BAC0562|oprN|tr|P95423|P95423_PSEAI Uncharacterized protein OS=Pseudomonas aeruginosa GN=oprN PE=4 SV=1
BAC0479|adeT2|tr|C7F8K7|C7F8K7_ACIBA AdeT2 OS=Acinetobacter baumannii PE=4 SV=1
BAC0363|smfY|tr|Q2AAU2|Q2AAU2_SERMA Multidrug efflux pump SmfY OS=Serratia marcescens GN=smfY PE=4 S
BAC0430|vmeB|tr|Q2AAU3|Q2AAU3_VIBPH Inner membrane protein VmeB OS=Vibrio parahaemolyticus GN=vmeB P
BAC0241|mexJ|tr|Q9HXW3|Q9HXW3_PSEAE Probable Resistance-Nodulation-Cell Division (RND) efflux membrane fu
BAC0510|vmeC|tr|Q87TN0|Q87TN0_VIBPA Uncharacterized protein OS=Vibrio parahaemolyticus serotype O3:K6 (strai
BAC0245|mexV|tr|Q9HW28|Q9HW28_PSEAE Probable Resistance-Nodulation-Cell Division (RND) efflux membrane fu
BAC0234|mexA|sp|P52477|MEXA_PSEAE Multidrug resistance protein MexA OS=Pseudomonas aeruginosa (strain ATC
BAC0503|farA|tr|Q9RQ30|Q9RQ30_NEIGO Efflux pump protein FarA OS=Neisseria gonorrhoeae PE=4 SV=1
BAC0202|lde|tr|G2JVL2|G2JVL2_LISMN Efflux pump Lde OS=Listeria monocytogenes J0161 GN=LMOG_01756 PE=4
BAC0042|bepC|sp|Q8G0Y6|BEPC_BRUSU Outer membrane efflux protein BepC OS=Brucella suis biovar 1 (strain 1330)
BAC0412|ttgI|sp|Q93PU3|TTGI_PSEPT Toluene efflux pump outer membrane protein TtgI OS=Pseudomonas putida (strai
BAC0291|oprJ|sp|Q51397|OPRJ_PSEAE Outer membrane protein OprJ OS=Pseudomonas aeruginosa (strain ATCC 15692
BAC0504|farB|tr|Q9RQ29|Q9RQ29_NEIGO Efflux pump protein FarB OS=Neisseria gonorrhoeae PE=4 SV=1
BAC0260|mtrF|tr|B4RN92|B4RN92_NEIG2 Antibiotic resistance efflux pump component OS=Neisseria gonorrhoeae (stra
BAC0613|cmeB|tr|Q8RTE4|Q8RTE4_CAMJU CmeB OS=Campylobacter jejuni GN=cmeB PE=4 SV=1
BAC0380|tbtB|tr|Q71UZ6|Q71UZ6_PSEST Resistance nodulation cell division family member TbtB OS=Pseudomonas stu
BAC0405|ttgB|sp|O52248|TTGB_PSEPT Toluene efflux pump membrane transporter TtgB OS=Pseudomonas putida (strai
BAC0688|merR2|tr|Q79B70|Q79B70_PSEST Organomercurial resistance regulatory protein OS=Pseudomonas stutzeri GN
BAC0018|adeJ|tr|Q24LT7|Q24LT7_ACIBA AdeJ OS=Acinetobacter baumannii GN=adeJ PE=4 SV=1
BAC0648|merA|sp|P08662|MERA_SERMA Mercuric reductase (Fragments) OS=Serratia marcescens GN=merA PE=3 SV
BAC0322|qacE|sp|P0AGC9|QACE_ECOLX Quaternary ammonium compound-resistance protein QacE OS=Escherichia co
BAC0324|qacF|sp|Q9X2N9|QACF_ENTAE Quaternary ammonium compound-resistance protein QacF OS=Enterobacter a
BAC0019|adeK|tr|Q24LT6|Q24LT6_ACIBA AdeK OS=Acinetobacter baumannii GN=adeK PE=4 SV=1
BAC0323|qacEdelta1|tr|Q7BQY4|Q7BQY4_PSEAI Disinfectant resistance protein OS=Pseudomonas aeruginosa GN=qacE
BAC0017|adeI|tr|Q2FD95|Q2FD95_ACIBA AdeI OS=Acinetobacter baumannii GN=adeI PE=4 SV=1
BAC0001|abeM|tr|Q5FAM9|Q5FAM9_ACIBA Multidrug efflux pump AbeM OS=Acinetobacter baumannii GN=abeM PE
BAC0472|adeB|tr|Q93E19|Q93E19_ACIBA AdeB RND protein OS=Acinetobacter baumannii GN=adeB PE=4 SV=1
BAC0335|rpoS|sp|P35540|RPOS_SHIFL RNA polymerase sigma factor RpoS OS=Shigella flexneri GN=rpoS PE=3 SV=3

BAC0536|oxyRkp|tr|C4WZN6|C4WZN6_KLEPN Activator of hydrogen peroxide-inducible genes OS=Klebsiella pneumo
BAC0357|recG|tr|B5L350|B5L350_9PSED ATP-dependent DNA helicase (Fragment) OS=Pseudomonas corrugata GN=re
BAC0194|ibpA|sp|P0C054|IBPA_ECOLI Small heat shock protein IbpA OS=Escherichia coli (strain K12) GN=ibpA PE=1
BAC0185|hdeA/yhiB|sp|P0AES9|HDEA_ECOLI Acid stress chaperone HdeA OS=Escherichia coli (strain K12) GN=hdeA
BAC0564|actP/yjcG|sp|P32705|ACTP_ECOLI Cation/acetate symporter ActP OS=Escherichia coli (strain K12) GN=actP
BAC0368|sodA|sp|P00448|SODM_ECOLI Superoxide dismutase [Mn] OS=Escherichia coli (strain K12) GN=sodA PE=1
BAC0533|cpxR|tr|C4WZK6|C4WZK6_KLEPN Response regulator of stress-related two-component regulatory system OS
BAC0471|adeA|tr|Q93E20|Q93E20_ACIBA AdeA membrane fusion protein OS=Acinetobacter baumannii GN=adeA PE=
BAC0175|gadE/yhiE|sp|P63204|GADE_ECOLI Transcriptional regulator GadE OS=Escherichia coli (strain K12) GN=gad
BAC0559|emrR|sp|P0ACR9|MPRA_ECOLI Transcriptional repressor MprA OS=Escherichia coli (strain K12) GN=mprA
BAC0532|cpxA|tr|C4WZK5|C4WZK5_KLEPN Sensor protein of stress-related two-component regulatory system OS=Kle
BAC0181|glpF|sp|P0AER0|GLPF_ECOLI Glycerol uptake facilitator protein OS=Escherichia coli (strain K12) GN=glpF P
BAC0047|bexA|tr|Q93HR0|Q93HR0_BACT4 BexA OS=Bacteroides thetaiotaomicron GN=bexA PE=4 SV=1
BAC0177|gadX|sp|P37639|GADX_ECOLI HTH-type transcriptional regulator GadX OS=Escherichia coli (strain K12) GN
BAC0293|ruvB|tr|B5L348|B5L348_9PSED Malic enzyme family protein (Fragment) OS=Pseudomonas corrugata PE=3 S
BAC0196|iclR|sp|P16528|ICLR_ECOLI Acetate operon repressor OS=Escherichia coli (strain K12) GN=iclR PE=1 SV=1
BAC0172|gadA|sp|P69908|DCEA_ECOLI Glutamate decarboxylase alpha OS=Escherichia coli (strain K12) GN=gadA PE
BAC0148|emrB|sp|P0AEJ0|EMRB_ECOLI Multidrug resistance protein B OS=Escherichia coli (strain K12) GN=emrB PE
BAC0370|soxR|sp|P0ACS2|SOXR_ECOLI Redox-sensitive transcriptional activator SoxR OS=Escherichia coli (strain K1
BAC0149|emrD|sp|P31442|EMRD_ECOLI Multidrug resistance protein D OS=Escherichia coli (strain K12) GN=emrD PE
BAC0530|phoB|tr|C4X6T6|C4X6T6_KLEPN Response regulator in two-component regulatory system with PhoQ OS=Kle
BAC0006|acrB|sp|P31224|ACRB_ECOLI Multidrug efflux pump subunit AcrB OS=Escherichia coli (strain K12) GN=acr
BAC0541|yieF|sp|P0AGE6|YIEF_ECOLI Uncharacterized protein YieF OS=Escherichia coli (strain K12) GN=yieF PE=1
BAC0176|gadW/yhiW|sp|P63201|GADW_ECOLI HTH-type transcriptional regulator GadW OS=Escherichia coli (strain K
BAC0195|ibpB|sp|P0C058|IBPB_ECOLI Small heat shock protein IbpB OS=Escherichia coli (strain K12) GN=ibpB PE=1
BAC0242|mexK|tr|Q9HXW4|Q9HXW4_PSEAE Probable Resistance-Nodulation-Cell Division (RND) efflux transporter
BAC0371|soxS|sp|P0A9E2|SOXS_ECOLI Regulatory protein SoxS OS=Escherichia coli (strain K12) GN=soxS PE=1 SV
BAC0214|mdtF/yhiV|sp|P37637|MDTF_ECOLI Multidrug resistance protein MdtF OS=Escherichia coli (strain K12) GN
BAC0061|cepA|sp|Q8RR17|FIEF_KLEPN Cation-efflux pump FieF OS=Klebsiella pneumoniae GN=fieF PE=3 SV=1
BAC0334|robA|sp|P0ACI0|ROB_ECOLI Right origin-binding protein OS=Escherichia coli (strain K12) GN=rob PE=1 SV
BAC0005|acrA|sp|P0AE06|ACRA_ECOLI Multidrug efflux pump subunit AcrA OS=Escherichia coli (strain K12) GN=ac
BAC0445|ygiW|sp|P0ADU5|YGIW_ECOLI Protein YgiW OS=Escherichia coli (strain K12) GN=ygiW PE=1 SV=1
BAC0156|fabI|sp|P0AEK4|FABI_ECOLI Enoyl-[acyl-carrier-protein] reductase [NADH] FabI OS=Escherichia coli (strain
BAC0186|hdeB/yhiC|sp|P0AET2|HDEB_ECOLI Acid stress chaperone HdeB OS=Escherichia coli (strain K12) GN=hdeB
BAC0211|mdtB/yegN|sp|P76398|MDTB_ECOLI Multidrug resistance protein MdtB OS=Escherichia coli (strain K12) GN
BAC0010|acrF/envD|sp|P24181|ACRF_ECOLI Acriflavine resistance protein F OS=Escherichia coli (strain K12) GN=acr
BAC0008|acrD/yffA|sp|P24177|ACRD_ECOLI Probable aminoglycoside efflux pump OS=Escherichia coli (strain K12) G
BAC0213|mdtE/yhiU|sp|P37636|MDTE_ECOLI Multidrug resistance protein MdtE OS=Escherichia coli (strain K12) GN
BAC0212|mdtC/yegO|sp|P76399|MDTC_ECOLI Multidrug resistance protein MdtC OS=Escherichia coli (strain K12) GN
BAC0039|baeR|sp|P69228|BAER_ECOLI Transcriptional regulatory protein BaeR OS=Escherichia coli (strain K12) GN=
BAC0560|marA|sp|P0ACH5|MARA_ECOLI Multiple antibiotic resistance protein MarA OS=Escherichia coli (strain K12)
BAC0296|ostA/lptD|sp|P31554|LPTD_ECOLI LPS-assembly protein LptD OS=Escherichia coli (strain K12) GN=lptD PE
BAC0295|oqxB|tr|Q69HW2|Q69HW2_ECOLX OqxB integral membrane protein OS=Escherichia coli GN=oqxB PE=4 S
BAC0220|mdtN/yjcR|sp|P32716|MDTN_ECOLI Multidrug resistance protein MdtN OS=Escherichia coli (strain K12) GN
BAC0218|mdtK/ydhE|sp|P37340|MDTK_ECOLI Multidrug resistance protein MdtK OS=Escherichia coli (strain K12) GN
BAC0434|ychH|sp|P0AB49|YCHH_ECOLI Uncharacterized protein YchH OS=Escherichia coli (strain K12) GN=ychH PE

BAC0215|mdtG/yceE|sp|P25744|MDTG_ECOLI Multidrug resistance protein MdtG OS=Escherichia coli (strain K12) GN=
BAC0147|emrA|sp|P27303|EMRA_ECOLI Multidrug resistance protein A OS=Escherichia coli (strain K12) GN=emrA PE
BAC0364|smrA|tr|C7SLZ1|C7SLZ1_STEMA ABC-type multidrug efflux pump (Fragment) OS=Stenotrophomonas maltop
BAC0015|adeG|tr|Q2FD81|Q2FD81_ACIBA Cation/multidrug efflux pump OS=Acinetobacter baumannii GN=29_167 PE=
BAC0496|adeN|tr|B7H1T7|B7H1T7_ACIB3 Bacterial regulatory protein, tetR family protein OS=Acinetobacter baumannii
BAC0393|tolC|sp|P02930|TOLC_ECOLI Outer membrane protein TolC OS=Escherichia coli (strain K12) GN=tolC PE=1 S
BAC0378|sugE|sp|P69937|SUGE_ECOLI Quaternary ammonium compound-resistance protein SugE OS=Escherichia coli
BAC0495|eefX|tr|A8CY68|A8CY68_KLEPN EefX OS=Klebsiella pneumoniae GN=eefX PE=4 SV=1
BAC0450|ymgB/ariR|sp|P75993|ARIR_ECOLI Probable two-component-system connector protein AriR OS=Escherichia c
BAC0009|acrE/envC|sp|P24180|ACRE_ECOLI Acriflavine resistance protein E OS=Escherichia coli (strain K12) GN=acrE
BAC0561|marR|sp|P27245|MARR_ECOLI Multiple antibiotic resistance protein MarR OS=Escherichia coli (strain K12) G
BAC0706|sodB|sp|P53641|SODF_PSEAE Superoxide dismutase [Fe] OS=Pseudomonas aeruginosa (strain ATCC 15692 /
BAC0040|baeS|sp|P30847|BAES_ECOLI Signal transduction histidine-protein kinase BaeS OS=Escherichia coli (strain K1
BAC0446|yhcN|sp|P64614|YHCN_ECOLI Uncharacterized protein YhcN OS=Escherichia coli (strain K12) GN=yhcN PE=
BAC0165|fetA/ybbL|sp|P77279|YBBL_ECOLI Uncharacterized ABC transporter ATP-binding protein YbbL OS=Escheric
BAC0207|mdeA|sp|P13254|MEGL_PSEPU Methionine gamma-lyase OS=Pseudomonas putida GN=mdeA PE=1 SV=2
BAC0041|bcr|sp|P28246|BCR_ECOLI Bicyclomycin resistance protein OS=Escherichia coli (strain K12) GN=bcr PE=1 SV
BAC0219|mdtM/yjiO|sp|P39386|MDTM_ECOLI Multidrug resistance protein MdtM OS=Escherichia coli (strain K12) GN
BAC0166|fetB/ybbM|sp|P77307|YBBM_ECOLI UPF0014 inner membrane protein YbbM OS=Escherichia coli (strain K12
BAC0222|mepB|sp|P0C070|MEPB_PSEPU Multidrug/solvent efflux pump membrane transporter MepB OS=Pseudomonas
BAC0384|tehA|sp|P25396|TEHA_ECOLI Tellurite resistance protein TehA OS=Escherichia coli (strain K12) GN=tehA PE
BAC0210|mdtA/yegM|sp|P76397|MDTA_ECOLI Multidrug resistance protein MdtA OS=Escherichia coli (strain K12) GN
BAC0174|gadC/xasA|sp|P63235|GADC_ECOLI Probable glutamate/gamma-aminobutyrate antiporter OS=Escherichia coli
BAC0150|emrE/mvrC|sp|P23895|EMRE_ECOLI Multidrug transporter EmrE OS=Escherichia coli (strain K12) GN=emrE
BAC0707|sodB|sp|P0AGD3|SODF_ECOLI Superoxide dismutase [Fe] OS=Escherichia coli (strain K12) GN=sodB PE=1 S
BAC0151|emrK|sp|P52599|EMRK_ECOLI Multidrug resistance protein K OS=Escherichia coli (strain K12) GN=emrK PE
BAC0494|eefA|tr|A8CY69|A8CY69_KLEPN EefA OS=Klebsiella pneumoniae GN=eefA PE=4 SV=1
BAC0155|evgS|sp|P58402|EVGS_ECO57 Sensor protein EvgS OS=Escherichia coli O157:H7 GN=evgS PE=3 SV=1
BAC0294|oqxA|tr|Q69HW3|Q69HW3_ECOLX OqxA membrane-fusion protein OS=Escherichia coli GN=oqxA PE=4 SV
BAC0239|mexF|tr|Q4KBN7|Q4KBN7_PSEF5 Multidrug efflux RND transporter, permease protein MexF OS=Pseudomona
BAC0153|emrY|sp|P52600|EMRY_ECOLI Multidrug resistance protein Y OS=Escherichia coli (strain K12) GN=emrY PE
BAC0154|evgA|sp|P0ACZ4|EVGA_ECOLI Positive transcription regulator EvgA OS=Escherichia coli (strain K12) GN=ev
BAC0437|ydeO|sp|P76135|YDEO_ECOLI HTH-type transcriptional regulator YdeO OS=Escherichia coli (strain K12) GN
BAC0134|dpr/dps|sp|P0CB53|DPS_STRSU DNA protection during starvation protein OS=Streptococcus suis GN=dps PE=
BAC0491|kdeA|tr|A6T6T9|A6T6T9_KLEP7 Multidrug/chloramphenicol efflux transport protein (MFS family) OS=Klebsi
BAC0208|mdfA/cmr|sp|P0AEY8|MDFA_ECOLI Multidrug transporter MdfA OS=Escherichia coli (strain K12) GN=mdfA
BAC0216|mdtI/ydgE|sp|P69210|MDTI_ECOLI Spermidine export protein MdtI OS=Escherichia coli (strain K12) GN=mdt
BAC0038|asr|sp|P36560|ASR_ECOLI Acid shock protein OS=Escherichia coli (strain K12) GN=asr PE=1 SV=3
BAC0011|acrR/ybaH|sp|P0ACS9|ACRR_ECOLI HTH-type transcriptional regulator AcrR OS=Escherichia coli (strain K1
BAC0529|kpnO|tr|C4XBC3|C4XBC3_KLEPN Outer membrane porin protein C OS=Klebsiella pneumoniae subsp. pneumo
BAC0013|adeE|tr|Q8GKU1|Q8GKU1_ACIG3 AdeE OS=Acinetobacter sp. 4365 GN=adeE PE=4 SV=2
BAC0531|phoR|tr|C4X6T5|C4X6T5_KLEPN Sensor kinase in two-component regulatory system with PhoP OS=Klebsiella
BAC0385|tehB|sp|P25397|TEHB_ECOLI Tellurite methyltransferase OS=Escherichia coli (strain K12) GN=tehB PE=1 SV
BAC0451|yodD|sp|P64519|YODD_ECOLI Uncharacterized protein YodD OS=Escherichia coli (strain K12) GN=yodD PE
BAC0436|ydeI|sp|P31130|YDEI_ECOLI Uncharacterized protein YdeI OS=Escherichia coli (strain K12) GN=ydeI PE=4 S
BAC0217|mdtJ/ebrB/ydgF|sp|Q3Z1V3|MDTJ_SHISS Spermidine export protein MdtJ OS=Shigella sonnei (strain Ss046) G

BAC0659|merB|sp|P08664|MERB_SERMA Alkylmercury lyase OS=Serratia marcescens GN=merB PE=3 SV=1
BAC0351|sitC|tr|Q9XCS0|Q9XCS0_SALTM SitC OS=Salmonella typhimurium GN=sitC PE=3 SV=1
BAC0252|mntP/yebN|sp|P76264|MNTP_ECOLI Probable manganese efflux pump MntP OS=Escherichia coli (strain K12)
BAC0438|ydeP|sp|P77561|YDEP_ECOLI Protein YdeP OS=Escherichia coli (strain K12) GN=ydeP PE=2 SV=1
BAC0223|mepC|sp|P0C071|MEPC_PSEPU Multidrug/solvent efflux pump outer membrane protein MepC OS=Pseudomor
BAC0238|mexE|tr|Q1IB41|Q1IB41_PSEE4 Multidrug efflux RND membrane fusion protein MexE OS=Pseudomonas ento
BAC0476|kpnE|tr|C4X7Z3|C4X7Z3_KLEPN Multidrug transport protein OS=Klebsiella pneumoniae subsp. pneumoniae N
BAC0157|fabK|tr|Q9FBC5|Q9FBC5_STREE Trans-2-enoyl-ACP reductase II OS=Streptococcus pneumoniae GN=fabK Pl
BAC0447|yjaA|sp|P09162|YJAA_ECOLI Uncharacterized protein YjaA OS=Escherichia coli (strain K12) GN=yjaA PE=4
BAC0246|mexW|tr|Q9HW27|Q9HW27_PSEAE Probable Resistance-Nodulation-Cell Division (RND) efflux transporter O
BAC0417|vcaM|tr|Q9KKV4|Q9KKV4_VIBCH ABC transporter, ATP-binding protein OS=Vibrio cholerae serotype O1 (st
BAC0508|adeL|tr|A3M732|A3M732_ACIBT Transcriptional regulator LysR family OS=Acinetobacter baumannii (strain A
BAC0349|sitA|tr|Q9XCS2|Q9XCS2_SALTI Iron transport protein, periplasmic-binding protein OS=Salmonella typhi GN=
BAC0477|kpnF|tr|C4X7Z4|C4X7Z4_KLEPN Spermidine export protein MdtI OS=Klebsiella pneumoniae subsp. pneumoni
BAC0237|mexD|tr|Q51396|Q51396_PSEAI RND family exporter MexD OS=Pseudomonas aeruginosa GN=mexD PE=4 S
BAC0353|smdA|tr|A7VN01|A7VN01_SERMA Multidrug efflux pump SmdA OS=Serratia marcescens GN=smdA PE=3 S
BAC0493|kmrA|tr|C4X8X9|C4X8X9_KLEPN Energy-dependent efflux protein for methyl viologen resistance OS=Klebsie
BAC0106|cuiD|sp|Q8ZRS2|CUEO_SALTY Blue copper oxidase CueO OS=Salmonella typhimurium (strain LT2 / SGSC1
BAC0143|emhB|tr|C1KA85|C1KA85_PSEFL EmhB OS=Pseudomonas fluorescens GN=emhB PE=4 SV=1
BAC0012|actP|sp|Q9X5X3|ATCU_SINMW Copper-transporting P-type ATPase OS=Sinorhizobium medicae (strain WSM
BAC0014|adeF|tr|Q2FD82|Q2FD82_ACIBA Putative RND family drug transporter OS=Acinetobacter baumannii GN=29_
BAC0565|actR|sp|A6UEL7|ACTR_SINMW Acid tolerance regulatory protein ActR OS=Sinorhizobium medicae (strain W
BAC0478|adeT1|tr|C7F8K6|C7F8K6_ACIBA AdeT1 OS=Acinetobacter baumannii PE=4 SV=1
BAC0244|mexT|tr|O87785|O87785_PSEAI MexT protein OS=Pseudomonas aeruginosa GN=mexT PE=4 SV=1
BAC0146|emmdR|tr|D5CJ69|D5CJ69_ENTCC MATE efflux family protein OS=Enterobacter cloacae subsp. cloacae (strai
BAC0350|sitB|tr|Q9XCS1|Q9XCS1_SALTM SitB OS=Salmonella typhimurium GN=sitB PE=3 SV=1
BAC0240|mexI|tr|Q9HWH4|Q9HWH4_PSEAE Probable Resistance-Nodulation-Cell Division (RND) efflux transporter O
BAC0492|kexD|tr|A6TA71|A6TA71_KLEP7 Acridine efflux pump OS=Klebsiella pneumoniae subsp. pneumoniae (strain
BAC0359|smeE|tr|I0KSX8|I0KSX8_STEMA RND efflux system, inner membrane transporter OS=Stenotrophomonas malt
BAC0404|ttgA|sp|Q9WWZ9|TTGA_PSEPT Toluene efflux pump periplasmic linker protein TtgA OS=Pseudomonas putid
BAC0290|opmD/nmpC|sp|P37592|OMPD_SALTY Outer membrane porin protein OmpD OS=Salmonella typhimurium (st
BAC0654|merB1|sp|P16172|MERB_BACCE Alkylmercury lyase OS=Bacillus cereus GN=merB1 PE=3 SV=2
BAC0235|mexB|sp|P52002|MEXB_PSEAE Multidrug resistance protein MexB OS=Pseudomonas aeruginosa (strain ATC
BAC0681|merR2|tr|Q9WWL1|Q9WWL1_BACSR Mercury resistance operon negative regulator MerR2 OS=Bacillus sp. (s
BAC0135|dpsA|tr|Q8KR86|Q8KR86_BURPE DpsA OS=Burkholderia pseudomallei GN=dpsA PE=3 SV=1
BAC0144|emhC|tr|Q4KH24|Q4KH24_PSEF5 Efflux transporter, outer membrane factor lipoprotein EmhC OS=Pseudomor
BAC0435|yddg/emrE|sp|D0ZXP9|YDDG_SALT1 Methyl viologen resistance protein YddG OS=Salmonella typhimurium
BAC0595|arsH|tr|P74312|P74312_SYNY3 Slr0945 protein OS=Synechocystis sp. (strain PCC 6803 / Kazusa) GN=slr0945
BAC0511|vmeD|tr|Q87TN1|Q87TN1_VIBPA Putative multidrug resistance protein OS=Vibrio parahaemolyticus serotype
BAC0184|hasF|tr|Q6GW09|Q6GW09_SERMA TolC-like protein OS=Serratia marcescens PE=4 SV=1
BAC0656|merB3|tr|Q7DHE7|Q7DHE7_BACCE Organomercurial lyase enzyme OS=Bacillus cereus GN=merB3 PE=4 SV
BAC0424|vexB|tr|Q9KVI2|Q9KVI2_VIBCH Multidrug resistance protein, putative OS=Vibrio cholerae serotype O1 (strai
BAC0179|gesB|tr|Q8ZRG9|Q8ZRG9_SALTY Putative cation efflux system protein OS=Salmonella typhimurium (strain L
BAC0339|sdeY|tr|Q7WSD5|Q7WSD5_SERMA Multidrug efflux pump SdeY OS=Serratia marcescens GN=sdeY PE=4 SV
BAC0705|sodA|sp|P53652|SODM_PSEAE Superoxide dismutase [Mn] OS=Pseudomonas aeruginosa (strain ATCC 15692
BAC0661|merB2|tr|Q7DJN2|Q7DJN2_BACME MerB2 OS=Bacillus megaterium GN=merB2 PE=4 SV=1

BAC0145|emhR|tr|Q4KH21|Q4KH21_PSEF5 Transcriptional regulator EmhR OS=Pseudomonas fluorescens (strain Pf-5 /
BAC0352|sitD|tr|Q9XCR9|Q9XCR9_SALTM SitD OS=Salmonella typhimurium GN=sitD PE=3 SV=1
BAC0016|adeH|tr|Q2FD80|Q2FD80_ACIBA Putative RND family drug transporter OS=Acinetobacter baumannii GN=29_
BAC0367|smvA/emrB|sp|D0ZXQ3|SMVA_SALT1 Methyl viologen resistance protein SmvA OS=Salmonella typhimuriu
BAC0142|emhA|tr|Q4KH22|Q4KH22_PSEF5 Efflux transporter, membrane fusion protein subunit EmhA OS=Pseudomona
BAC0419|vceB|tr|O51919|O51919_VIBCL VceB OS=Vibrio cholerae GN=vceB PE=4 SV=1
BAC0413|ttgR|sp|Q9AIU0|TTGR_PSEPT HTH-type transcriptional regulator TtgR OS=Pseudomonas putida (strain DOT-
BAC0329|qacZ|tr|Q82YU7|Q82YU7_ENTFA Multidrug resistance protein OS=Enterococcus faecalis (strain ATCC 70080
BAC0321|qacC/qacD/smr|sp|P14319|QACC_STAAU Quaternary ammonium compound-resistance protein QacC OS=Stap
BAC0002|abeS|tr|Q2FD83|Q2FD83_ACIBA QacEdelta1 SMR family efflux pump OS=Acinetobacter baumannii GN=qac
BAC0507|tolCsm|tr|R4ITT0|R4ITT0_STEMA Outer membrane protein OS=Stenotrophomonas maltophilia GN=tolCsm P
BAC0337|sdeB|tr|Q84GI9|Q84GI9_SERMA Putative resistance-nodulation cell division protein SdeB OS=Serratia marces
BAC0173|gadB|sp|P69910|DCEB_ECOLI Glutamate decarboxylase beta OS=Escherichia coli (strain K12) GN=gadB PE=
BAC0597|baeS|tr|D0ZNE2|D0ZNE2_SALT1 Signal transduction histidine-protein kinase BaeS OS=Salmonella typhimuri
BAC0499|emrAsm|tr|B2FIC9|B2FIC9_STRMK Putative multidrug resistance protein A OS=Stenotrophomonas maltophili
BAC0405|ttgB|sp|O52248|TTGB_PSEPT Toluene efflux pump membrane transporter TtgB OS=Pseudomonas putida (strai
BAC0505|farR|tr|Q7DD70|Q7DD70_NEIMB Transcriptional regulator, MarR family OS=Neisseria meningitidis serogroup
BAC0502|emrRsm|tr|B2FIC7|B2FIC7_STRMK Putative MarR-family transcriptional regulator OS=Stenotrophomonas ma
BAC0029|chrF|tr|A4UQR2|A4UQR2_9RHIZ ChrF OS=Ochrobactrum tritici GN=chrF PE=4 SV=1
BAC0538|chrR|tr|Q7BD45|Q7BD45_PSEPU Chromate reductase OS=Pseudomonas putida GN=chrR PE=4 SV=1
BAC0328|qacR|sp|P0A0N4|QACR_STAAU HTH-type transcriptional regulator QacR OS=Staphylococcus aureus GN=qac
BAC0650|merA|tr|O08449|O08449_9PSED Mercuric reductase OS=Pseudomonas sp. K-62 GN=merA PE=4 SV=1
BAC0141|emeA|tr|Q8GR72|Q8GR72_ENTFL Multidrug efflux pump OS=Enterococcus faecalis GN=emeA PE=4 SV=1
BAC0159|fabV|sp|Q9KRA3|Y1738_VIBCH Putative reductase VC_1738/VC_1739 OS=Vibrio cholerae serotype O1 (strai
BAC0657|merB|tr|O07303|O07303_9PSED Alkylmercury lyase OS=Pseudomonas sp. K-62 GN=merB PE=3 SV=2
BAC0506|pcm|tr|R4IUI7|R4IUI7_STEMA Protein-L-isoaspartate O-methyltransferase OS=Stenotrophomonas maltophilia
BAC0229|merG|tr|O07302|O07302_9PSED Mercuric resistance protein OS=Pseudomonas sp. K-62 GN=merG PE=4 SV=
BAC0498|ideR|sp|P0A672|IDER_MYCTU Iron-dependent repressor IdeR OS=Mycobacterium tuberculosis GN=ideR PE=
BAC0313|pmpM|sp|Q9I3Y3|PMPM_PSEAE Multidrug resistance protein PmpM OS=Pseudomonas aeruginosa (strain AT
BAC0596|baeR|tr|D0ZNE3|D0ZNE3_SALT1 DNA-binding transcriptional regulator BaeR OS=Salmonella typhimurium (s
BAC0415|ttgV|sp|Q93PU6|TTGV_PSEPT HTH-type transcriptional regulator TtgV OS=Pseudomonas putida (strain DOT-
BAC0044|bepE|sp|Q8G2M6|BEPE_BRUSU Efflux pump membrane transporter BepE OS=Brucella suis biovar 1 (strain 1
BAC0500|emrBsm|tr|B2FID0|B2FID0_STRMK Putative multidrug resistance protein B OS=Stenotrophomonas maltophili
BAC0027|chrB|tr|A4UQR5|A4UQR5_9RHIZ ChrB OS=Ochrobactrum tritici GN=chrB PE=4 SV=1
BAC0248|mexY|tr|Q9ZNG8|Q9ZNG8_PSEAI MexY OS=Pseudomonas aeruginosa GN=mexY PE=4 SV=1
BAC0411|ttgH|sp|Q93PU4|TTGH_PSEPT Toluene efflux pump membrane transporter TtgH OS=Pseudomonas putida (stra
BAC0258|mtrD|tr|Q5F725|Q5F725_NEIG1 Antibiotic resistance efflux pump component OS=Neisseria gonorrhoeae (strai
BAC0282|norA|sp|P0A0J7|NORA_STAAU Quinolone resistance protein NorA OS=Staphylococcus aureus GN=norA PE=
BAC0358|smeD|tr|I0KSX9|I0KSX9_STEMA Membrane fusion protein of RND family multidrug efflux pump OS=Stenot
BAC0418|vceA|tr|O51918|O51918_VIBCL VceA OS=Vibrio cholerae GN=vceA PE=4 SV=1
BAC0043|bepD|sp|Q8G2M7|BEPD_BRUSU Efflux pump periplasmic linker BepD OS=Brucella suis biovar 1 (strain 1330
BAC0221|mepA|sp|Q7A7N0|MEPA_STAAN Multidrug export protein MepA OS=Staphylococcus aureus (strain N315) G
BAC0473|adeC|tr|Q93E18|Q93E18_ACIBA AdeC outer membrane protein OS=Acinetobacter baumannii GN=adeC PE=4
BAC0292|oprM/oprK|sp|Q51487|OPRM_PSEAE Outer membrane protein OprM OS=Pseudomonas aeruginosa (strain AT
BAC0025|amvA|tr|C4PAW9|C4PAW9_ACIBA Major facilitator superfamily efflux pump OS=Acinetobacter baumannii P
BAC0501|emrCsm|tr|B2FIC8|B2FIC8_STRMK Putative outer membrane multidrug efflux protein OS=Stenotrophomonas

BAC0567|actA|sp|Q52910|LNT_SINMW Apolipoprotein N-acyltransferase OS=Sinorhizobium medicae (strain WSM419)
BAC0398|triC|tr|Q663E8|Q663E8_YERPS TriC protein OS=Yersinia pseudotuberculosis serotype I (strain IP32953) GN=t
BAC0430|vmeB|tr|Q2AAU3|Q2AAU3_VIBPH Inner membrane protein VmeB OS=Vibrio parahaemolyticus GN=vmeB P
BAC0688|merR2|tr|Q79B70|Q79B70_PSEST Organomercurial resistance regulatory protein OS=Pseudomonas stutzeri GN
BAC0648|merA|sp|P08662|MERA_SERMA Mercuric reductase (Fragments) OS=Serratia marcescens GN=merA PE=3 SV
BAC0324|qacF|sp|Q9X2N9|QACF_ENTAE Quaternary ammonium compound-resistance protein QacF OS=Enterobacter a
BAC0357|recG|tr|B5L350|B5L350_9PSED ATP-dependent DNA helicase (Fragment) OS=Pseudomonas corrugata GN=rec
BAC0047|bexA|tr|Q93HR0|Q93HR0_BACT4 BexA OS=Bacteroides thetaiotaomicron GN=bexA PE=4 SV=1
BAC0018|adeJ|tr|Q24LT7|Q24LT7_ACIBA AdeJ OS=Acinetobacter baumannii GN=adeJ PE=4 SV=1
BAC0293|ruvB|tr|B5L348|B5L348_9PSED Malic enzyme family protein (Fragment) OS=Pseudomonas corrugata PE=3 SV
BAC0335|rpoS|sp|P35540|RPOS_SHIFL RNA polymerase sigma factor RpoS OS=Shigella flexneri GN=rpoS PE=3 SV=3
BAC0706|sodB|sp|P53641|SODF_PSEAE Superoxide dismutase [Fe] OS=Pseudomonas aeruginosa (strain ATCC 15692 /
BAC0564|actP/yjcG|sp|P32705|ACTP_ECOLI Cation/acetate symporter ActP OS=Escherichia coli (strain K12) GN=actP P
BAC0322|qacE|sp|P0AGC9|QACE_ECOLX Quaternary ammonium compound-resistance protein QacE OS=Escherichia co
BAC0194|ibpA|sp|P0C054|IBPA_ECOLI Small heat shock protein IbpA OS=Escherichia coli (strain K12) GN=ibpA PE=1
BAC0368|sodA|sp|P00448|SODM_ECOLI Superoxide dismutase [Mn] OS=Escherichia coli (strain K12) GN=sodA PE=1 S
BAC0019|adeK|tr|Q24LT6|Q24LT6_ACIBA AdeK OS=Acinetobacter baumannii GN=adeK PE=4 SV=1
BAC0532|cpxA|tr|C4WZK5|C4WZK5_KLEPN Sensor protein of stress-related two-component regulatory system OS=Kle
BAC0242|mexK|tr|Q9HXW4|Q9HXW4_PSEAE Probable Resistance-Nodulation-Cell Division (RND) efflux transporter O
BAC0536|oxyRkp|tr|C4WZN6|C4WZN6_KLEPN Activator of hydrogen peroxide-inducible genes OS=Klebsiella pneumon
BAC0364|smrA|tr|C7SLZ1|C7SLZ1_STEMA ABC-type multidrug efflux pump (Fragment) OS=Stenotrophomonas maltop
BAC0533|cpxR|tr|C4WZK6|C4WZK6_KLEPN Response regulator of stress-related two-component regulatory system OS=
BAC0565|actR|sp|A6UEL7|ACTR_SINMW Acid tolerance regulatory protein ActR OS=Sinorhizobium medicae (strain W
BAC0239|mexF|tr|Q4KBN7|Q4KBN7_PSEF5 Multidrug efflux RND transporter, permease protein MexF OS=Pseudomona
BAC0211|mdtB/yegN|sp|P76398|MDTB_ECOLI Multidrug resistance protein MdtB OS=Escherichia coli (strain K12) GN=
BAC0196|iclR|sp|P16528|ICLR_ECOLI Acetate operon repressor OS=Escherichia coli (strain K12) GN=iclR PE=1 SV=1
BAC0006|acrB|sp|P31224|ACRB_ECOLI Multidrug efflux pump subunit AcrB OS=Escherichia coli (strain K12) GN=acrB
BAC0181|glpF|sp|P0AER0|GLPF_ECOLI Glycerol uptake facilitator protein OS=Escherichia coli (strain K12) GN=glpF P
BAC0295|oqxB|tr|Q69HW2|Q69HW2_ECOLX OqxB integral membrane protein OS=Escherichia coli GN=oqxB PE=4 SV
BAC0370|soxR|sp|P0ACS2|SOXR_ECOLI Redox-sensitive transcriptional activator SoxR OS=Escherichia coli (strain K12
BAC0559|emrR|sp|P0ACR9|MPRA_ECOLI Transcriptional repressor MprA OS=Escherichia coli (strain K12) GN=mprA P
BAC0039|baeR|sp|P69228|BAER_ECOLI Transcriptional regulatory protein BaeR OS=Escherichia coli (strain K12) GN=b
BAC0001|abeM|tr|Q5FAM9|Q5FAM9_ACIBA Multidrug efflux pump AbeM OS=Acinetobacter baumannii GN=abeM PE
BAC0017|adeI|tr|Q2FD95|Q2FD95_ACIBA AdeI OS=Acinetobacter baumannii GN=adeI PE=4 SV=1
BAC0323|qacEdelta1|tr|Q7BQY4|Q7BQY4_PSEAI Disinfectant resistance protein OS=Pseudomonas aeruginosa GN=qacE
BAC0149|emrD|sp|P31442|EMRD_ECOLI Multidrug resistance protein D OS=Escherichia coli (strain K12) GN=emrD PE
BAC0148|emrB|sp|P0AEJ0|EMRB_ECOLI Multidrug resistance protein B OS=Escherichia coli (strain K12) GN=emrB PE
BAC0143|emhB|tr|C1KA85|C1KA85_PSEFL EmhB OS=Pseudomonas fluorescens GN=emhB PE=4 SV=1
BAC0156|fabI|sp|P0AEK4|FABI_ECOLI Enoyl-[acyl-carrier-protein] reductase [NADH] FabI OS=Escherichia coli (strain
BAC0061|cepA|sp|Q8RR17|FIEF_KLEPN Cation-efflux pump FieF OS=Klebsiella pneumoniae GN=fieF PE=3 SV=1
BAC0212|mdtC/yegO|sp|P76399|MDTC_ECOLI Multidrug resistance protein MdtC OS=Escherichia coli (strain K12) GN=
BAC0530|phoB|tr|C4X6T6|C4X6T6_KLEPN Response regulator in two-component regulatory system with PhoQ OS=Kleb
BAC0015|adeG|tr|Q2FD81|Q2FD81_ACIBA Cation/multidrug efflux pump OS=Acinetobacter baumannii GN=29_167 PE=
BAC0494|eefA|tr|A8CY69|A8CY69_KLEPN EefA OS=Klebsiella pneumoniae GN=eefA PE=4 SV=1
BAC0541|yieF|sp|P0AGE6|YIEF_ECOLI Uncharacterized protein YieF OS=Escherichia coli (strain K12) GN=yieF PE=1 S
BAC0334|robA|sp|P0ACI0|ROB_ECOLI Right origin-binding protein OS=Escherichia coli (strain K12) GN=rob PE=1 SV

BAC0008|acrD/yffA|sp|P24177|ACRD_ECOLI Probable aminoglycoside efflux pump OS=Escherichia coli (strain K12) G
BAC0246|mexW|tr|Q9HW27|Q9HW27_PSEAE Probable Resistance-Nodulation-Cell Division (RND) efflux transporter O
BAC0010|acrF/envD|sp|P24181|ACRF_ECOLI Acriflavine resistance protein F OS=Escherichia coli (strain K12) GN=acrF
BAC0215|mdtG/yceE|sp|P25744|MDTG_ECOLI Multidrug resistance protein MdtG OS=Escherichia coli (strain K12) GN=
BAC0172|gadA|sp|P69908|DCEA_ECOLI Glutamate decarboxylase alpha OS=Escherichia coli (strain K12) GN=gadA PE
BAC0210|mdtA/yegM|sp|P76397|MDTA_ECOLI Multidrug resistance protein MdtA OS=Escherichia coli (strain K12) GN
BAC0005|acrA|sp|P0AE06|ACRA_ECOLI Multidrug efflux pump subunit AcrA OS=Escherichia coli (strain K12) GN=acr
BAC0451|yodD|sp|P64519|YODD_ECOLI Uncharacterized protein YodD OS=Escherichia coli (strain K12) GN=yodD PE
BAC0393|tolC|sp|P02930|TOLC_ECOLI Outer membrane protein TolC OS=Escherichia coli (strain K12) GN=tolC PE=1 S
BAC0218|mdtK/ydhE|sp|P37340|MDTK_ECOLI Multidrug resistance protein MdtK OS=Escherichia coli (strain K12) GN
BAC0294|oqxA|tr|Q69HW3|Q69HW3_ECOLX OqxA membrane-fusion protein OS=Escherichia coli GN=oqxA PE=4 SV=
BAC0166|fetB/ybbM|sp|P77307|YBBM_ECOLI UPF0014 inner membrane protein YbbM OS=Escherichia coli (strain K12
BAC0220|mdtN/yjcR|sp|P32716|MDTN_ECOLI Multidrug resistance protein MdtN OS=Escherichia coli (strain K12) GN=
BAC0011|acrR/ybaH|sp|P0ACS9|ACRR_ECOLI HTH-type transcriptional regulator AcrR OS=Escherichia coli (strain K12
BAC0222|mepB|sp|P0C070|MEPB_PSEPU Multidrug/solvent efflux pump membrane transporter MepB OS=Pseudomonas
BAC0529|kpnO|tr|C4XBC3|C4XBC3_KLEPN Outer membrane porin protein C OS=Klebsiella pneumoniae subsp. pneumo
BAC0491|kdeA|tr|A6T6T9|A6T6T9_KLEP7 Multidrug/chloramphenicol efflux transport protein (MFS family) OS=Klebsi
BAC0195|ibpB|sp|P0C058|IBPB_ECOLI Small heat shock protein IbpB OS=Escherichia coli (strain K12) GN=ibpB PE=1
BAC0296|ostA/lptD|sp|P31554|LPTD_ECOLI LPS-assembly protein LptD OS=Escherichia coli (strain K12) GN=lptD PE=
BAC0040|baeS|sp|P30847|BAES_ECOLI Signal transduction histidine-protein kinase BaeS OS=Escherichia coli (strain K1
BAC0185|hdeA/yhiB|sp|P0AES9|HDEA_ECOLI Acid stress chaperone HdeA OS=Escherichia coli (strain K12) GN=hdeA
BAC0707|sodB|sp|P0AGD3|SODF_ECOLI Superoxide dismutase [Fe] OS=Escherichia coli (strain K12) GN=sodB PE=1 S
BAC0147|emrA|sp|P27303|EMRA_ECOLI Multidrug resistance protein A OS=Escherichia coli (strain K12) GN=emrA PE
BAC0208|mdfA/cmr|sp|P0AEY8|MDFA_ECOLI Multidrug transporter MdfA OS=Escherichia coli (strain K12) GN=mdfA
BAC0495|eefX|tr|A8CY68|A8CY68_KLEPN EefX OS=Klebsiella pneumoniae GN=eefX PE=4 SV=1
BAC0371|soxS|sp|P0A9E2|SOXS_ECOLI Regulatory protein SoxS OS=Escherichia coli (strain K12) GN=soxS PE=1 SV=
BAC0531|phoR|tr|C4X6T5|C4X6T5_KLEPN Sensor kinase in two-component regulatory system with PhoP OS=Klebsiella
BAC0560|marA|sp|P0ACH5|MARA_ECOLI Multiple antibiotic resistance protein MarA OS=Escherichia coli (strain K12)
BAC0213|mdtE/yhiU|sp|P37636|MDTE_ECOLI Multidrug resistance protein MdtE OS=Escherichia coli (strain K12) GN=
BAC0493|kmrA|tr|C4X8X9|C4X8X9_KLEPN Energy-dependent efflux protein for methyl viologen resistance OS=Klebsie
BAC0434|ychH|sp|P0AB49|YCHH_ECOLI Uncharacterized protein YchH OS=Escherichia coli (strain K12) GN=ychH PE
BAC0041|bcr|sp|P28246|BCR_ECOLI Bicyclomycin resistance protein OS=Escherichia coli (strain K12) GN=bcr PE=1 SV
BAC0595|arsH|tr|P74312|P74312_SYNY3 Slr0945 protein OS=Synechocystis sp. (strain PCC 6803 / Kazusa) GN=slr0945
BAC0561|marR|sp|P27245|MARR_ECOLI Multiple antibiotic resistance protein MarR OS=Escherichia coli (strain K12) G
BAC0384|tehA|sp|P25396|TEHA_ECOLI Tellurite resistance protein TehA OS=Escherichia coli (strain K12) GN=tehA PE
BAC0498|ideR|sp|P0A672|IDER_MYCTU Iron-dependent repressor IdeR OS=Mycobacterium tuberculosis GN=ideR PE=
BAC0445|ygiW|sp|P0ADU5|YGIW_ECOLI Protein YgiW OS=Escherichia coli (strain K12) GN=ygiW PE=1 SV=1
BAC0417|vcaM|tr|Q9KKV4|Q9KKV4_VIBCH ABC transporter, ATP-binding protein OS=Vibrio cholerae serotype O1 (st
BAC0177|gadX|sp|P37639|GADX_ECOLI HTH-type transcriptional regulator GadX OS=Escherichia coli (strain K12) GN
BAC0351|sitC|tr|Q9XCS0|Q9XCS0_SALTM SitC OS=Salmonella typhimurium GN=sitC PE=3 SV=1
BAC0009|acrE/envC|sp|P24180|ACRE_ECOLI Acriflavine resistance protein E OS=Escherichia coli (strain K12) GN=acrE
BAC0214|mdtF/yhiV|sp|P37637|MDTF_ECOLI Multidrug resistance protein MdtF OS=Escherichia coli (strain K12) GN=
BAC0219|mdtM/yjiO|sp|P39386|MDTM_ECOLI Multidrug resistance protein MdtM OS=Escherichia coli (strain K12) GN
BAC0146|emmdR|tr|D5CJ69|D5CJ69_ENTCC MATE efflux family protein OS=Enterobacter cloacae subsp. cloacae (strai
BAC0235|mexB|sp|P52002|MEXB_PSEAE Multidrug resistance protein MexB OS=Pseudomonas aeruginosa (strain ATC
BAC0186|hdeB/yhiC|sp|P0AET2|HDEB_ECOLI Acid stress chaperone HdeB OS=Escherichia coli (strain K12) GN=hdeB

BAC0216|mdtI/ydgE|sp|P69210|MDTI_ECOLI Spermidine export protein MdtI OS=Escherichia coli (strain K12) GN=mdt
BAC0175|gadE/yhiE|sp|P63204|GADE_ECOLI Transcriptional regulator GadE OS=Escherichia coli (strain K12) GN=gad
BAC0165|fetA/ybbL|sp|P77279|YBBL_ECOLI Uncharacterized ABC transporter ATP-binding protein YbbL OS=Escheric
BAC0385|tehB|sp|P25397|TEHB_ECOLI Tellurite methyltransferase OS=Escherichia coli (strain K12) GN=tehB PE=1 SV
BAC0153|emrY|sp|P52600|EMRY_ECOLI Multidrug resistance protein Y OS=Escherichia coli (strain K12) GN=emrY PE
BAC0378|sugE|sp|P69937|SUGE_ECOLI Quaternary ammonium compound-resistance protein SugE OS=Escherichia coli
BAC0353|smdA|tr|A7VN01|A7VN01_SERMA Multidrug efflux pump SmdA OS=Serratia marcescens GN=smdA PE=3 S
BAC0144|emhC|tr|Q4KH24|Q4KH24_PSEF5 Efflux transporter, outer membrane factor lipoprotein EmhC OS=Pseudomo
BAC0477|kpnF|tr|C4X7Z4|C4X7Z4_KLEPN Spermidine export protein MdtI OS=Klebsiella pneumoniae subsp. pneumon
BAC0155|evgS|sp|P58402|EVGS_ECO57 Sensor protein EvgS OS=Escherichia coli O157:H7 GN=evgS PE=3 SV=1
BAC0446|yhcN|sp|P64614|YHCN_ECOLI Uncharacterized protein YhcN OS=Escherichia coli (strain K12) GN=yhcN PE=
BAC0450|ymgB/ariR|sp|P75993|ARIR_ECOLI Probable two-component-system connector protein AriR OS=Escherichia c
BAC0244|mexT|tr|O87785|O87785_PSEAI MexT protein OS=Pseudomonas aeruginosa GN=mexT PE=4 SV=1
BAC0176|gadW/yhiW|sp|P63201|GADW_ECOLI HTH-type transcriptional regulator GadW OS=Escherichia coli (strain K
BAC0252|mntP/yebN|sp|P76264|MNTP_ECOLI Probable manganese efflux pump MntP OS=Escherichia coli (strain K12)
BAC0142|emhA|tr|Q4KH22|Q4KH22_PSEF5 Efflux transporter, membrane fusion protein subunit EmhA OS=Pseudomon
BAC0154|evgA|sp|P0ACZ4|EVGA_ECOLI Positive transcription regulator EvgA OS=Escherichia coli (strain K12) GN=e
BAC0359|smeE|tr|I0KSX8|I0KSX8_STEMA RND efflux system, inner membrane transporter OS=Stenotrophomonas mal
BAC0151|emrK|sp|P52599|EMRK_ECOLI Multidrug resistance protein K OS=Escherichia coli (strain K12) GN=emrK PE
BAC0145|emhR|tr|Q4KH21|Q4KH21_PSEF5 Transcriptional regulator EmhR OS=Pseudomonas fluorescens (strain Pf-5 /
BAC0174|gadC/xasA|sp|P63235|GADC_ECOLI Probable glutamate/gamma-aminobutyrate antiporter OS=Escherichia col
BAC0496|adeN|tr|B7H1T7|B7H1T7_ACIB3 Bacterial regulatory protein, tetR family protein OS=Acinetobacter bauman
BAC0012|actP|sp|Q9X5X3|ATCU_SINMW Copper-transporting P-type ATPase OS=Sinorhizobium medicae (strain WSM
BAC0150|emrE/mvrC|sp|P23895|EMRE_ECOLI Multidrug transporter EmrE OS=Escherichia coli (strain K12) GN=emrE
BAC0472|adeB|tr|Q93E19|Q93E19_ACIBA AdeB RND protein OS=Acinetobacter baumannii GN=adeB PE=4 SV=1
BAC0029|chrF|tr|A4UQR2|A4UQR2_9RHIZ ChrF OS=Ochrobactrum tritici GN=chrF PE=4 SV=1
BAC0438|ydeP|sp|P77561|YDEP_ECOLI Protein YdeP OS=Escherichia coli (strain K12) GN=ydeP PE=2 SV=1
BAC0352|sitD|tr|Q9XCR9|Q9XCR9_SALTM SitD OS=Salmonella typhimurium GN=sitD PE=3 SV=1
BAC0135|dpsA|tr|Q8KR86|Q8KR86_BURPE DpsA OS=Burkholderia pseudomallei GN=dpsA PE=3 SV=1
BAC0350|sitB|tr|Q9XCS1|Q9XCS1_SALTM SitB OS=Salmonella typhimurium GN=sitB PE=3 SV=1
BAC0436|ydeI|sp|P31130|YDEI_ECOLI Uncharacterized protein YdeI OS=Escherichia coli (strain K12) GN=ydeI PE=4 S
BAC0476|kpnE|tr|C4X7Z3|C4X7Z3_KLEPN Multidrug transport protein OS=Klebsiella pneumoniae subsp. pneumoniae N
BAC0358|oscA|tr|B6CM35|B6CM35_9PSED Putative uncharacterized protein oscA OS=Pseudomonas corrugata GN=osc
BAC0217|mdtJ/ebrB/ydgF|sp|Q3Z1V3|MDTJ_SHISS Spermidine export protein MdtJ OS=Shigella sonnei (strain Ss046)
BAC0013|adeE|tr|Q8GKU1|Q8GKU1_ACIG3 AdeE OS=Acinetobacter sp. 4365 GN=adeE PE=4 SV=2
BAC0492|kexD|tr|A6TA71|A6TA71_KLEP7 Acridine efflux pump OS=Klebsiella pneumoniae subsp. pneumoniae (strain
BAC0435|yddg/emrE|sp|D0ZXP9|YDDG_SALT1 Methyl viologen resistance protein YddG OS=Salmonella typhimurium
BAC0659|merB|sp|P08664|MERB_SERMA Alkylmercury lyase OS=Serratia marcescens GN=merB PE=3 SV=1
BAC0437|ydeO|sp|P76135|YDEO_ECOLI HTH-type transcriptional regulator YdeO OS=Escherichia coli (strain K12) GN
BAC0157|fabK|tr|Q9FBC5|Q9FBC5_STREE Trans-2-enoyl-ACP reductase II OS=Streptococcus pneumoniae GN=fabK P
BAC0650|merA|tr|O08449|O08449_9PSED Mercuric reductase OS=Pseudomonas sp. K-62 GN=merA PE=4 SV=1
BAC0349|sitA|tr|Q9XCS2|Q9XCS2_SALTI Iron transport protein, periplasmic-binding protein OS=Salmonella typhi GN=
BAC0511|vmeD|tr|Q87TN1|Q87TN1_VIBPA Putative multidrug resistance protein OS=Vibrio parahaemolyticus serotype
BAC0424|vexB|tr|Q9KVI2|Q9KVI2_VIBCH Multidrug resistance protein, putative OS=Vibrio cholerae serotype O1 (strai
BAC0337|sdeB|tr|Q84GI9|Q84GI9_SERMA Putative resistance-nodulation cell division protein SdeB OS=Serratia marces
BAC0339|sdeY|tr|Q7WSD5|Q7WSD5_SERMA Multidrug efflux pump SdeY OS=Serratia marcescens GN=sdeY PE=4 S

BAC0106|cuiD|sp|Q8ZRS2|CUEO_SALTY Blue copper oxidase CueO OS=Salmonella typhimurium (strain LT2 / SGSC1
BAC0447|yjaA|sp|P09162|YJAA_ECOLI Uncharacterized protein YjaA OS=Escherichia coli (strain K12) GN=yjaA PE=4
BAC0038|asr|sp|P36560|ASR_ECOLI Acid shock protein OS=Escherichia coli (strain K12) GN=asr PE=1 SV=3
BAC0179|gesB|tr|Q8ZRG9|Q8ZRG9_SALTY Putative cation efflux system protein OS=Salmonella typhimurium (strain L
BAC0500|emrBsm|tr|B2FID0|B2FID0_STRMK Putative multidrug resistance protein B OS=Stenotrophomonas maltophili
BAC0238|mexE|tr|Q1IB41|Q1IB41_PSEE4 Multidrug efflux RND membrane fusion protein MexE OS=Pseudomonas ento
BAC0223|mepC|sp|P0C071|MEPC_PSEPU Multidrug/solvent efflux pump outer membrane protein MepC OS=Pseudomon
BAC0229|merG|tr|O07302|O07302_9PSED Mercuric resistance protein OS=Pseudomonas sp. K-62 GN=merG PE=4 SV=
BAC0367|smvA/emrB|sp|D0ZXQ3|SMVA_SALT1 Methyl viologen resistance protein SmvA OS=Salmonella typhimuriu
BAC0499|emrAsm|tr|B2FIC9|B2FIC9_STRMK Putative multidrug resistance protein A OS=Stenotrophomonas maltophili
BAC0134|dpr/dps|sp|P0CB53|DPS_STRSU DNA protection during starvation protein OS=Streptococcus suis GN=dps PE=
BAC0237|mexD|tr|Q51396|Q51396_PSEAI RND family exporter MexD OS=Pseudomonas aeruginosa GN=mexD PE=4 S
BAC0596|baeR|tr|D0ZNE3|D0ZNE3_SALT1 DNA-binding transcriptional regulator BaeR OS=Salmonella typhimurium (
BAC0506|pcm|tr|R4IUI7|R4IUI7_STEMA Protein-L-isoaspartate O-methyltransferase OS=Stenotrophomonas maltophilia
BAC0526|vmeV|tr|Q87HZ7|Q87HZ7_VIBPA Transporter, AcrB/D/F family OS=Vibrio parahaemolyticus serotype O3:K6
BAC0419|vceB|tr|O51919|O51919_VIBCL VceB OS=Vibrio cholerae GN=vceB PE=4 SV=1
BAC0505|farR|tr|Q7DD70|Q7DD70_NEIMB Transcriptional regulator, MarR family OS=Neisseria meningitidis serogroup
BAC0404|ttgA|sp|Q9WWZ9|TTGA_PSEPT Toluene efflux pump periplasmic linker protein TtgA OS=Pseudomonas putid
BAC0381|tbtM|tr|Q71UZ5|Q71UZ5_PSEST Outer membrane protein TbtM OS=Pseudomonas stutzeri GN=tbtM PE=4 SV
BAC0354|smdB|tr|A7VN02|A7VN02_SERMA Multidrug efflux pump SmdB OS=Serratia marcescens GN=smdB PE=3 S
BAC0566|actS|tr|Q52912|Q52912_9RHIZ Histidine protein kinase OS=Sinorhizobium medicae GN=actS PE=4 SV=1
BAC0538|chrR|tr|Q7BD45|Q7BD45_PSEPU Chromate reductase OS=Pseudomonas putida GN=chrR PE=4 SV=1
BAC0705|sodA|sp|P53652|SODM_PSEAE Superoxide dismutase [Mn] OS=Pseudomonas aeruginosa (strain ATCC 15692
BAC0027|chrB|tr|A4UQR5|A4UQR5_9RHIZ ChrB OS=Ochrobactrum tritici GN=chrB PE=4 SV=1
BAC0657|merB|tr|O07303|O07303_9PSED Alkylmercury lyase OS=Pseudomonas sp. K-62 GN=merB PE=3 SV=2
BAC0377|ssmE|tr|A7VN75|A7VN75_SERMA Multidrug efflux pump SsmE OS=Serratia marcescens GN=ssmE PE=3 SV
BAC0508|adeL|tr|A3M732|A3M732_ACIBT Transcriptional regulator LysR family OS=Acinetobacter baumannii (strain A
BAC0173|gadB|sp|P69910|DCEB_ECOLI Glutamate decarboxylase beta OS=Escherichia coli (strain K12) GN=gadB PE=
BAC0597|baeS|tr|D0ZNE2|D0ZNE2_SALT1 Signal transduction histidine-protein kinase BaeS OS=Salmonella typhimuriu
BAC0290|opmD/nmpC|sp|P37592|OMPD_SALTY Outer membrane porin protein OmpD OS=Salmonella typhimurium (st
BAC0184|hasF|tr|Q6GW09|Q6GW09_SERMA TolC-like protein OS=Serratia marcescens PE=4 SV=1
BAC0681|merR2|tr|Q9WWL1|Q9WWL1_BACSR Mercury resistance operon negative regulator MerR2 OS=Bacillus sp. (
BAC0509|vexH|tr|Q9KTI8|Q9KTI8_VIBCH Multidrug resistance protein, putative OS=Vibrio cholerae serotype O1 (strai
BAC0141|emeA|tr|Q8GR72|Q8GR72_ENTFL Multidrug efflux pump OS=Enterococcus faecalis GN=emeA PE=4 SV=1
BAC0358|smeD|tr|I0KSX9|I0KSX9_STEMA Membrane fusion protein of RND family multidrug efflux pump OS=Stenotr
BAC0044|bepE|sp|Q8G2M6|BEPE_BRUSU Efflux pump membrane transporter BepE OS=Brucella suis biovar 1 (strain 1
BAC0207|mdeA|sp|P13254|MEGL_PSEPU Methionine gamma-lyase OS=Pseudomonas putida GN=mdeA PE=1 SV=2
BAC0478|adeT1|tr|C7F8K6|C7F8K6_ACIBA AdeT1 OS=Acinetobacter baumannii PE=4 SV=1
BAC0379|tbtA|tr|Q71UZ7|Q71UZ7_PSEST Membrane fusion protein TbtA OS=Pseudomonas stutzeri GN=tbtA PE=4 SV
BAC0236|mexC|tr|Q51395|Q51395_PSEAI Membrane fusion protein MexC OS=Pseudomonas aeruginosa GN=mexC PE=
BAC0471|adeA|tr|Q93E20|Q93E20_ACIBA AdeA membrane fusion protein OS=Acinetobacter baumannii GN=adeA PE=
BAC0159|fabV|sp|Q9KRA3|Y1738_VIBCH Putative reductase VC_1738/VC_1739 OS=Vibrio cholerae serotype O1 (strai
BAC0028|chrC|tr|A4UQR3|A4UQR3_9RHIZ Superoxide dismutase OS=Ochrobactrum tritici GN=chrC PE=3 SV=1
BAC0240|mexI|tr|Q9HWH4|Q9HWH4_PSEAE Probable Resistance-Nodulation-Cell Division (RND) efflux transporter O
BAC0513|vmeF|tr|Q87R57|Q87R57_VIBPA Putative multidrug resistance protein OS=Vibrio parahaemolyticus serotype O
BAC0408|ttgE|sp|Q9KWV4|TTGE_PSEPT Toluene efflux pump membrane transporter TtgE OS=Pseudomonas putida (str

BAC0258|mtrD|tr|Q5F725|Q5F725_NEIG1 Antibiotic resistance efflux pump component OS=Neisseria gonorrhoeae (strai
BAC0507|tolCsm|tr|R4ITT0|R4ITT0_STEMA Outer membrane protein OS=Stenotrophomonas maltophilia GN=tolCsm PE
BAC0313|pmpM|sp|Q9I3Y3|PMPM_PSEAE Multidrug resistance protein PmpM OS=Pseudomonas aeruginosa (strain AT
BAC0474|adeD|tr|Q67GM1|Q67GM1_ACIG3 AdeD OS=Acinetobacter sp. 4365 GN=adeD PE=4 SV=1
BAC0372|srpA|sp|O31099|SRPA_PSEPU Solvent efflux pump periplasmic linker SrpA OS=Pseudomonas putida GN=srp
BAC0338|sdeX|tr|Q7WSD6|Q7WSD6_SERMA Multidrug efflux pump SdeX OS=Serratia marcescens GN=sdeX PE=4 S
BAC0418|vceA|tr|O51918|O51918_VIBCL VceA OS=Vibrio cholerae GN=vceA PE=4 SV=1
BAC0314|pmrA|sp|P0A4K4|PMRA_STRPN Multi-drug resistance efflux pump PmrA OS=Streptococcus pneumoniae sero
BAC0221|mepA|sp|Q7A7N0|MEPA_STAAN Multidrug export protein MepA OS=Staphylococcus aureus (strain N315) G
BAC0473|adeC|tr|Q93E18|Q93E18_ACIBA AdeC outer membrane protein OS=Acinetobacter baumannii GN=adeC PE=4
BAC0291|oprJ|sp|Q51397|OPRJ_PSEAE Outer membrane protein OprJ OS=Pseudomonas aeruginosa (strain ATCC 15692
BAC0501|emrCsm|tr|B2FIC8|B2FIC8_STRMK Putative outer membrane multidrug efflux protein OS=Stenotrophomonas
BAC0521|vmeK|tr|Q87LY6|Q87LY6_VIBPA Putative multidrug resistance protein OS=Vibrio parahaemolyticus serotype
BAC0430|vmeB|tr|Q2AAU3|Q2AAU3_VIBPH Inner membrane protein VmeB OS=Vibrio parahaemolyticus GN=vmeB P
BAC0046|bepG|sp|Q8FWV9|BEPG_BRUSU Efflux pump membrane transporter BepG OS=Brucella suis biovar 1 (strain
BAC0380|tbtB|tr|Q71UZ6|Q71UZ6_PSEST Resistance nodulation cell division family member TbtB OS=Pseudomonas st
BAC0248|mexY|tr|Q9ZNG8|Q9ZNG8_PSEAI MexY OS=Pseudomonas aeruginosa GN=mexY PE=4 SV=1
BAC0405|ttgB|sp|O52248|TTGB_PSEPT Toluene efflux pump membrane transporter TtgB OS=Pseudomonas putida (strai
BAC0688|merR2|tr|Q79B70|Q79B70_PSEST Organomercurial resistance regulatory protein OS=Pseudomonas stutzeri G
BAC0648|merA|sp|P08662|MERA_SERMA Mercuric reductase (Fragments) OS=Serratia marcescens GN=merA PE=3 S
BAC0018|adeJ|tr|Q24LT7|Q24LT7_ACIBA AdeJ OS=Acinetobacter baumannii GN=adeJ PE=4 SV=1
BAC0324|qacF|sp|Q9X2N9|QACF_ENTAE Quaternary ammonium compound-resistance protein QacF OS=Enterobacter a
BAC0019|adeK|tr|Q24LT6|Q24LT6_ACIBA AdeK OS=Acinetobacter baumannii GN=adeK PE=4 SV=1
BAC0357|recG|tr|B5L350|B5L350_9PSED ATP-dependent DNA helicase (Fragment) OS=Pseudomonas corrugata GN=re
BAC0293|ruvB|tr|B5L348|B5L348_9PSED Malic enzyme family protein (Fragment) OS=Pseudomonas corrugata PE=3 S
BAC0017|adeI|tr|Q2FD95|Q2FD95_ACIBA AdeI OS=Acinetobacter baumannii GN=adeI PE=4 SV=1
BAC0706|sodB|sp|P53641|SODF_PSEAE Superoxide dismutase [Fe] OS=Pseudomonas aeruginosa (strain ATCC 15692 /
BAC0335|rpoS|sp|P35540|RPOS_SHIFL RNA polymerase sigma factor RpoS OS=Shigella flexneri GN=rpoS PE=3 SV=3
BAC0001|abeM|tr|Q5FAM9|Q5FAM9_ACIBA Multidrug efflux pump AbeM OS=Acinetobacter baumannii GN=abeM PE
BAC0364|smrA|tr|C7SLZ1|C7SLZ1_STEMA ABC-type multidrug efflux pump (Fragment) OS=Stenotrophomonas malto
BAC0322|qacE|sp|P0AGC9|QACE_ECOLX Quaternary ammonium compound-resistance protein QacE OS=Escherichia c
BAC0242|mexK|tr|Q9HXW4|Q9HXW4_PSEAE Probable Resistance-Nodulation-Cell Division (RND) efflux transporter
BAC0564|actP/yjcG|sp|P32705|ACTP_ECOLI Cation/acetate symporter ActP OS=Escherichia coli (strain K12) GN=actP
BAC0047|bexA|tr|Q93HR0|Q93HR0_BACT4 BexA OS=Bacteroides thetaiotaomicron GN=bexA PE=4 SV=1
BAC0533|cpxR|tr|C4WZK6|C4WZK6_KLEPN Response regulator of stress-related two-component regulatory system OS
BAC0239|mexF|tr|Q4KBN7|Q4KBN7_PSEF5 Multidrug efflux RND transporter, permease protein MexF OS=Pseudomona
BAC0532|cpxA|tr|C4WZK5|C4WZK5_KLEPN Sensor protein of stress-related two-component regulatory system OS=Kle
BAC0536|oxyRkp|tr|C4WZN6|C4WZN6_KLEPN Activator of hydrogen peroxide-inducible genes OS=Klebsiella pneumo
BAC0194|ibpA|sp|P0C054|IBPA_ECOLI Small heat shock protein IbpA OS=Escherichia coli (strain K12) GN=ibpA PE=1
BAC0565|actR|sp|A6UEL7|ACTR_SINMW Acid tolerance regulatory protein ActR OS=Sinorhizobium medicae (strain W
BAC0015|adeG|tr|Q2FD81|Q2FD81_ACIBA Cation/multidrug efflux pump OS=Acinetobacter baumannii GN=29_167 PE
BAC0143|emhB|tr|C1KA85|C1KA85_PSEFL EmhB OS=Pseudomonas fluorescens GN=emhB PE=4 SV=1
BAC0006|acrB|sp|P31224|ACRB_ECOLI Multidrug efflux pump subunit AcrB OS=Escherichia coli (strain K12) GN=acr
BAC0211|mdtB/yegN|sp|P76398|MDTB_ECOLI Multidrug resistance protein MdtB OS=Escherichia coli (strain K12) GN
BAC0156|fabI|sp|P0AEK4|FABI_ECOLI Enoyl-[acyl-carrier-protein] reductase [NADH] FabI OS=Escherichia coli (strain
BAC0148|emrB|sp|P0AEJ0|EMRB_ECOLI Multidrug resistance protein B OS=Escherichia coli (strain K12) GN=emrB PE

BAC0181|glpF|sp|P0AER0|GLPF_ECOLI Glycerol uptake facilitator protein OS=Escherichia coli (strain K12) GN=glpF P
BAC0295|oqxB|tr|Q69HW2|Q69HW2_ECOLX OqxB integral membrane protein OS=Escherichia coli GN=oqxB PE=4 SV
BAC0246|mexW|tr|Q9HW27|Q9HW27_PSEAE Probable Resistance-Nodulation-Cell Division (RND) efflux transporter O
BAC0323|qacEdelta1|tr|Q7BQY4|Q7BQY4_PSEAI Disinfectant resistance protein OS=Pseudomonas aeruginosa GN=qacE
BAC0370|soxR|sp|P0ACS2|SOXR_ECOLI Redox-sensitive transcriptional activator SoxR OS=Escherichia coli (strain K12
BAC0196|iclR|sp|P16528|ICLR_ECOLI Acetate operon repressor OS=Escherichia coli (strain K12) GN=iclR PE=1 SV=1
BAC0559|emrR|sp|P0ACR9|MPRA_ECOLI Transcriptional repressor MprA OS=Escherichia coli (strain K12) GN=mprA
BAC0472|adeB|tr|Q93E19|Q93E19_ACIBA AdeB RND protein OS=Acinetobacter baumannii GN=adeB PE=4 SV=1
BAC0496|adeN|tr|B7H1T7|B7H1T7_ACIB3 Bacterial regulatory protein, tetR family protein OS=Acinetobacter baumanni
BAC0061|cepA|sp|Q8RR17|FIEF_KLEPN Cation-efflux pump FieF OS=Klebsiella pneumoniae GN=fieF PE=3 SV=1
BAC0368|sodA|sp|P00448|SODM_ECOLI Superoxide dismutase [Mn] OS=Escherichia coli (strain K12) GN=sodA PE=1
BAC0530|phoB|tr|C4X6T6|C4X6T6_KLEPN Response regulator in two-component regulatory system with PhoQ OS=Kle
BAC0039|baeR|sp|P69228|BAER_ECOLI Transcriptional regulatory protein BaeR OS=Escherichia coli (strain K12) GN=b
BAC0212|mdtC/yegO|sp|P76399|MDTC_ECOLI Multidrug resistance protein MdtC OS=Escherichia coli (strain K12) GN
BAC0010|acrF/envD|sp|P24181|ACRF_ECOLI Acriflavine resistance protein F OS=Escherichia coli (strain K12) GN=acrF
BAC0008|acrD/yffA|sp|P24177|ACRD_ECOLI Probable aminoglycoside efflux pump OS=Escherichia coli (strain K12) G
BAC0707|sodB|sp|P0AGD3|SODF_ECOLI Superoxide dismutase [Fe] OS=Escherichia coli (strain K12) GN=sodB PE=1
BAC0417|vcaM|tr|Q9KKV4|Q9KKV4_VIBCH ABC transporter, ATP-binding protein OS=Vibrio cholerae serotype O1 (st
BAC0172|gadA|sp|P69908|DCEA_ECOLI Glutamate decarboxylase alpha OS=Escherichia coli (strain K12) GN=gadA PE
BAC0144|emhC|tr|Q4KH24|Q4KH24_PSEF5 Efflux transporter, outer membrane factor lipoprotein EmhC OS=Pseudomo
BAC0498|ideR|sp|P0A672|IDER_MYCTU Iron-dependent repressor IdeR OS=Mycobacterium tuberculosis GN=ideR PE=
BAC0541|yieF|sp|P0AGE6|YIEF_ECOLI Uncharacterized protein YieF OS=Escherichia coli (strain K12) GN=yieF PE=1
BAC0294|oqxA|tr|Q69HW3|Q69HW3_ECOLX OqxA membrane-fusion protein OS=Escherichia coli GN=oqxA PE=4 SV
BAC0494|eefA|tr|A8CY69|A8CY69_KLEPN EefA OS=Klebsiella pneumoniae GN=eefA PE=4 SV=1
BAC0222|mepB|sp|P0C070|MEPB_PSEPU Multidrug/solvent efflux pump membrane transporter MepB OS=Pseudomona
BAC0371|soxS|sp|P0A9E2|SOXS_ECOLI Regulatory protein SoxS OS=Escherichia coli (strain K12) GN=soxS PE=1 SV=
BAC0041|bcr|sp|P28246|BCR_ECOLI Bicyclomycin resistance protein OS=Escherichia coli (strain K12) GN=bcr PE=1 S
BAC0235|mexB|sp|P52002|MEXB_PSEAE Multidrug resistance protein MexB OS=Pseudomonas aeruginosa (strain ATC
BAC0296|ostA/lptD|sp|P31554|LPTD_ECOLI LPS-assembly protein LptD OS=Escherichia coli (strain K12) GN=lptD PE
BAC0351|sitC|tr|Q9XCS0|Q9XCS0_SALTM SitC OS=Salmonella typhimurium GN=sitC PE=3 SV=1
BAC0166|fetB/ybbM|sp|P77307|YBBM_ECOLI UPF0014 inner membrane protein YbbM OS=Escherichia coli (strain K12
BAC0334|robA|sp|P0ACI0|ROB_ECOLI Right origin-binding protein OS=Escherichia coli (strain K12) GN=rob PE=1 SV
BAC0393|tolC|sp|P02930|TOLC_ECOLI Outer membrane protein TolC OS=Escherichia coli (strain K12) GN=tolC PE=1
BAC0358|oscA|tr|B6CM35|B6CM35_9PSED Putative uncharacterized protein oscA OS=Pseudomonas corrugata GN=osc
BAC0149|emrD|sp|P31442|EMRD_ECOLI Multidrug resistance protein D OS=Escherichia coli (strain K12) GN=emrD PE
BAC0218|mdtK/ydhE|sp|P37340|MDTK_ECOLI Multidrug resistance protein MdtK OS=Escherichia coli (strain K12) GN
BAC0216|mdtI/ydgE|sp|P69210|MDTI_ECOLI Spermidine export protein MdtI OS=Escherichia coli (strain K12) GN=md
BAC0244|mexT|tr|O87785|O87785_PSEAI MexT protein OS=Pseudomonas aeruginosa GN=mexT PE=4 SV=1
BAC0147|emrA|sp|P27303|EMRA_ECOLI Multidrug resistance protein A OS=Escherichia coli (strain K12) GN=emrA PE
BAC0220|mdtN/yjcR|sp|P32716|MDTN_ECOLI Multidrug resistance protein MdtN OS=Escherichia coli (strain K12) GN
BAC0154|evgA|sp|P0ACZ4|EVGA_ECOLI Positive transcription regulator EvgA OS=Escherichia coli (strain K12) GN=e
BAC0215|mdtG/yceE|sp|P25744|MDTG_ECOLI Multidrug resistance protein MdtG OS=Escherichia coli (strain K12) GN
BAC0491|kdeA|tr|A6T6T9|A6T6T9_KLEP7 Multidrug/chloramphenicol efflux transport protein (MFS family) OS=Klebsi
BAC0595|arsH|tr|P74312|P74312_SYNY3 Slr0945 protein OS=Synechocystis sp. (strain PCC 6803 / Kazusa) GN=slr0945
BAC0005|acrA|sp|P0AE06|ACRA_ECOLI Multidrug efflux pump subunit AcrA OS=Escherichia coli (strain K12) GN=ac
BAC0040|baeS|sp|P30847|BAES_ECOLI Signal transduction histidine-protein kinase BaeS OS=Escherichia coli (strain K1

BAC0531|phoR|tr|C4X6T5|C4X6T5_KLEPN Sensor kinase in two-component regulatory system with PhoP OS=Klebsiella
BAC0378|sugE|sp|P69937|SUGE_ECOLI Quaternary ammonium compound-resistance protein SugE OS=Escherichia coli
BAC0145|emhR|tr|Q4KH21|Q4KH21_PSEF5 Transcriptional regulator EmhR OS=Pseudomonas fluorescens (strain Pf-5 /
BAC0185|hdeA/yhiB|sp|P0AES9|HDEA_ECOLI Acid stress chaperone HdeA OS=Escherichia coli (strain K12) GN=hdeA
BAC0142|emhA|tr|Q4KH22|Q4KH22_PSEF5 Efflux transporter, membrane fusion protein subunit EmhA OS=Pseudomona
BAC0445|ygiW|sp|P0ADU5|YGIW_ECOLI Protein YgiW OS=Escherichia coli (strain K12) GN=ygiW PE=1 SV=1
BAC0213|mdtE/yhiU|sp|P37636|MDTE_ECOLI Multidrug resistance protein MdtE OS=Escherichia coli (strain K12) GN=
BAC0359|smeE|tr|I0KSX8|I0KSX8_STEMA RND efflux system, inner membrane transporter OS=Stenotrophomonas malt
BAC0012|actP|sp|Q9X5X3|ATCU_SINMW Copper-transporting P-type ATPase OS=Sinorhizobium medicae (strain WSM
BAC0210|mdtA/yegM|sp|P76397|MDTA_ECOLI Multidrug resistance protein MdtA OS=Escherichia coli (strain K12) GN
BAC0214|mdtF/yhiV|sp|P37637|MDTF_ECOLI Multidrug resistance protein MdtF OS=Escherichia coli (strain K12) GN=
BAC0208|mdfA/cmr|sp|P0AEY8|MDFA_ECOLI Multidrug transporter MdfA OS=Escherichia coli (strain K12) GN=mdfA
BAC0177|gadX|sp|P37639|GADX_ECOLI HTH-type transcriptional regulator GadX OS=Escherichia coli (strain K12) GN
BAC0493|kmrA|tr|C4X8X9|C4X8X9_KLEPN Energy-dependent efflux protein for methyl viologen resistance OS=Klebsie
BAC0476|kpnE|tr|C4X7Z3|C4X7Z3_KLEPN Multidrug transport protein OS=Klebsiella pneumoniae subsp. pneumoniae N
BAC0217|mdtJ/ebrB/ydgF|sp|Q3Z1V3|MDTJ_SHISS Spermidine export protein MdtJ OS=Shigella sonnei (strain Ss046) G
BAC0174|gadC/xasA|sp|P63235|GADC_ECOLI Probable glutamate/gamma-aminobutyrate antiporter OS=Escherichia coli
BAC0529|kpnO|tr|C4XBC3|C4XBC3_KLEPN Outer membrane porin protein C OS=Klebsiella pneumoniae subsp. pneumo
BAC0134|dpr/dps|sp|P0CB53|DPS_STRSU DNA protection during starvation protein OS=Streptococcus suis GN=dps PE=
BAC0252|mntP/yebN|sp|P76264|MNTP_ECOLI Probable manganese efflux pump MntP OS=Escherichia coli (strain K12)
BAC0011|acrR/ybaH|sp|P0ACS9|ACRR_ECOLI HTH-type transcriptional regulator AcrR OS=Escherichia coli (strain K12
BAC0176|gadW/yhiW|sp|P63201|GADW_ECOLI HTH-type transcriptional regulator GadW OS=Escherichia coli (strain K
BAC0135|dpsA|tr|Q8KR86|Q8KR86_BURPE DpsA OS=Burkholderia pseudomallei GN=dpsA PE=3 SV=1
BAC0146|emmdR|tr|D5CJ69|D5CJ69_ENTCC MATE efflux family protein OS=Enterobacter cloacae subsp. cloacae (strai
BAC0155|evgS|sp|P58402|EVGS_ECO57 Sensor protein EvgS OS=Escherichia coli O157:H7 GN=evgS PE=3 SV=1
BAC0477|kpnF|tr|C4X7Z4|C4X7Z4_KLEPN Spermidine export protein MdtI OS=Klebsiella pneumoniae subsp. pneumoni
BAC0219|mdtM/yjiO|sp|P39386|MDTM_ECOLI Multidrug resistance protein MdtM OS=Escherichia coli (strain K12) GN
BAC0009|acrE/envC|sp|P24180|ACRE_ECOLI Acriflavine resistance protein E OS=Escherichia coli (strain K12) GN=acrE
BAC0013|adeE|tr|Q8GKU1|Q8GKU1_ACIG3 AdeE OS=Acinetobacter sp. 4365 GN=adeE PE=4 SV=2
BAC0471|adeA|tr|Q93E20|Q93E20_ACIBA AdeA membrane fusion protein OS=Acinetobacter baumannii GN=adeA PE=4
BAC0424|vexB|tr|Q9KVI2|Q9KVI2_VIBCH Multidrug resistance protein, putative OS=Vibrio cholerae serotype O1 (strain
BAC0511|vmeD|tr|Q87TN1|Q87TN1_VIBPA Putative multidrug resistance protein OS=Vibrio parahaemolyticus serotype
BAC0384|tehA|sp|P25396|TEHA_ECOLI Tellurite resistance protein TehA OS=Escherichia coli (strain K12) GN=tehA PE
BAC0029|chrF|tr|A4UQR2|A4UQR2_9RHIZ ChrF OS=Ochrobactrum tritici GN=chrF PE=4 SV=1
BAC0153|emrY|sp|P52600|EMRY_ECOLI Multidrug resistance protein Y OS=Escherichia coli (strain K12) GN=emrY PE
BAC0495|eefX|tr|A8CY68|A8CY68_KLEPN EefX OS=Klebsiella pneumoniae GN=eefX PE=4 SV=1
BAC0434|ychH|sp|P0AB49|YCHH_ECOLI Uncharacterized protein YchH OS=Escherichia coli (strain K12) GN=ychH PE
BAC0337|sdeB|tr|Q84GI9|Q84GI9_SERMA Putative resistance-nodulation cell division protein SdeB OS=Serratia marcesc
BAC0165|fetA/ybbL|sp|P77279|YBBL_ECOLI Uncharacterized ABC transporter ATP-binding protein YbbL OS=Escheric
BAC0436|ydeI|sp|P31130|YDEI_ECOLI Uncharacterized protein YdeI OS=Escherichia coli (strain K12) GN=ydeI PE=4 S
BAC0141|emeA|tr|Q8GR72|Q8GR72_ENTFL Multidrug efflux pump OS=Enterococcus faecalis GN=emeA PE=4 SV=1
BAC0385|tehB|sp|P25397|TEHB_ECOLI Tellurite methyltransferase OS=Escherichia coli (strain K12) GN=tehB PE=1 SV
BAC0349|sitA|tr|Q9XCS2|Q9XCS2_SALTI Iron transport protein, periplasmic-binding protein OS=Salmonella typhi GN=
BAC0438|ydeP|sp|P77561|YDEP_ECOLI Protein YdeP OS=Escherichia coli (strain K12) GN=ydeP PE=2 SV=1
BAC0339|sdeY|tr|Q7WSD5|Q7WSD5_SERMA Multidrug efflux pump SdeY OS=Serratia marcescens GN=sdeY PE=4 SV
BAC0195|ibpB|sp|P0C058|IBPB_ECOLI Small heat shock protein IbpB OS=Escherichia coli (strain K12) GN=ibpB PE=1

BAC0561|marR|sp|P27245|MARR_ECOLI Multiple antibiotic resistance protein MarR OS=Escherichia coli (strain K12) G
BAC0186|hdeB/yhiC|sp|P0AET2|HDEB_ECOLI Acid stress chaperone HdeB OS=Escherichia coli (strain K12) GN=hdeB
BAC0353|smdA|tr|A7VN01|A7VN01_SERMA Multidrug efflux pump SmdA OS=Serratia marcescens GN=smdA PE=3 S
BAC0451|yodD|sp|P64519|YODD_ECOLI Uncharacterized protein YodD OS=Escherichia coli (strain K12) GN=yodD PE
BAC0151|emrK|sp|P52599|EMRK_ECOLI Multidrug resistance protein K OS=Escherichia coli (strain K12) GN=emrK PE
BAC0596|baeR|tr|D0ZNE3|D0ZNE3_SALT1 DNA-binding transcriptional regulator BaeR OS=Salmonella typhimurium (s
BAC0157|fabK|tr|Q9FBC5|Q9FBC5_STREE Trans-2-enoyl-ACP reductase II OS=Streptococcus pneumoniae GN=fabK P
BAC0705|sodA|sp|P53652|SODM_PSEAE Superoxide dismutase [Mn] OS=Pseudomonas aeruginosa (strain ATCC 15692
BAC0437|ydeO|sp|P76135|YDEO_ECOLI HTH-type transcriptional regulator YdeO OS=Escherichia coli (strain K12) GN
BAC0560|marA|sp|P0ACH5|MARA_ECOLI Multiple antibiotic resistance protein MarA OS=Escherichia coli (strain K12)
BAC0447|yjaA|sp|P09162|YJAA_ECOLI Uncharacterized protein YjaA OS=Escherichia coli (strain K12) GN=yjaA PE=4
BAC0175|gadE/yhiE|sp|P63204|GADE_ECOLI Transcriptional regulator GadE OS=Escherichia coli (strain K12) GN=gad
BAC0238|mexE|tr|Q1IB41|Q1IB41_PSEE4 Multidrug efflux RND membrane fusion protein MexE OS=Pseudomonas ento
BAC0499|emrAsm|tr|B2FIC9|B2FIC9_STRMK Putative multidrug resistance protein A OS=Stenotrophomonas maltophili
BAC0229|merG|tr|O07302|O07302_9PSED Mercuric resistance protein OS=Pseudomonas sp. K-62 GN=merG PE=4 SV=
BAC0150|emrE/mvrC|sp|P23895|EMRE_ECOLI Multidrug transporter EmrE OS=Escherichia coli (strain K12) GN=emrE
BAC0179|gesB|tr|Q8ZRG9|Q8ZRG9_SALTY Putative cation efflux system protein OS=Salmonella typhimurium (strain L
BAC0526|vmeV|tr|Q87HZ7|Q87HZ7_VIBPA Transporter, AcrB/D/F family OS=Vibrio parahaemolyticus serotype O3:K6
BAC0159|fabV|sp|Q9KRA3|Y1738_VIBCH Putative reductase VC_1738/VC_1739 OS=Vibrio cholerae serotype O1 (strai
BAC0290|opmD/nmpC|sp|P37592|OMPD_SALTY Outer membrane porin protein OmpD OS=Salmonella typhimurium (st
BAC0509|vexH|tr|Q9KTI8|Q9KTI8_VIBCH Multidrug resistance protein, putative OS=Vibrio cholerae serotype O1 (strain
BAC0044|bepE|sp|Q8G2M6|BEPE_BRUSU Efflux pump membrane transporter BepE OS=Brucella suis biovar 1 (strain 1
BAC0681|merR2|tr|Q9WWL1|Q9WWL1_BACSR Mercury resistance operon negative regulator MerR2 OS=Bacillus sp. (
BAC0106|cuiD|sp|Q8ZRS2|CUEO_SALTY Blue copper oxidase CueO OS=Salmonella typhimurium (strain LT2 / SGSC1
BAC0508|adeL|tr|A3M732|A3M732_ACIBT Transcriptional regulator LysR family OS=Acinetobacter baumannii (strain A
BAC0659|merB|sp|P08664|MERB_SERMA Alkylmercury lyase OS=Serratia marcescens GN=merB PE=3 SV=1
BAC0656|merB3|tr|Q7DHE7|Q7DHE7_BACCE Organomercurial lyase enzyme OS=Bacillus cereus GN=merB3 PE=4 SV
BAC0505|farR|tr|Q7DD70|Q7DD70_NEIMB Transcriptional regulator, MarR family OS=Neisseria meningitidis serogroup
BAC0354|smdB|tr|A7VN02|A7VN02_SERMA Multidrug efflux pump SmdB OS=Serratia marcescens GN=smdB PE=3 S
BAC0492|kexD|tr|A6TA71|A6TA71_KLEP7 Acridine efflux pump OS=Klebsiella pneumoniae subsp. pneumoniae (strain
BAC0478|adeT1|tr|C7F8K6|C7F8K6_ACIBA AdeT1 OS=Acinetobacter baumannii PE=4 SV=1
BAC0313|pmpM|sp|Q9I3Y3|PMPM_PSEAE Multidrug resistance protein PmpM OS=Pseudomonas aeruginosa (strain AT
BAC0237|mexD|tr|Q51396|Q51396_PSEAI RND family exporter MexD OS=Pseudomonas aeruginosa GN=mexD PE=4 S
BAC0446|yhcN|sp|P64614|YHCN_ECOLI Uncharacterized protein YhcN OS=Escherichia coli (strain K12) GN=yhcN PE=
BAC0450|ymgB/ariR|sp|P75993|ARIR_ECOLI Probable two-component-system connector protein AriR OS=Escherichia c
BAC0350|sitB|tr|Q9XCS1|Q9XCS1_SALTM SitB OS=Salmonella typhimurium GN=sitB PE=3 SV=1
BAC0538|chrR|tr|Q7BD45|Q7BD45_PSEPU Chromate reductase OS=Pseudomonas putida GN=chrR PE=4 SV=1
BAC0404|ttgA|sp|Q9WWZ9|TTGA_PSEPT Toluene efflux pump periplasmic linker protein TtgA OS=Pseudomonas putid
BAC0223|mepC|sp|P0C071|MEPC_PSEPU Multidrug/solvent efflux pump outer membrane protein MepC OS=Pseudomon
BAC0038|asr|sp|P36560|ASR_ECOLI Acid shock protein OS=Escherichia coli (strain K12) GN=asr PE=1 SV=3
BAC0027|chrB|tr|A4UQR5|A4UQR5_9RHIZ ChrB OS=Ochrobactrum tritici GN=chrB PE=4 SV=1
BAC0002|abeS|tr|Q2FD83|Q2FD83_ACIBA QacEdelta1 SMR family efflux pump OS=Acinetobacter baumannii GN=qac
BAC0507|tolCsm|tr|R4ITT0|R4ITT0_STEMA Outer membrane protein OS=Stenotrophomonas maltophilia GN=tolCsm P
BAC0650|merA|tr|O08449|O08449_9PSED Mercuric reductase OS=Pseudomonas sp. K-62 GN=merA PE=4 SV=1
BAC0240|mexI|tr|Q9HWH4|Q9HWH4_PSEAE Probable Resistance-Nodulation-Cell Division (RND) efflux transporter O
BAC0501|emrCsm|tr|B2FIC8|B2FIC8_STRMK Putative outer membrane multidrug efflux protein OS=Stenotrophomonas

BAC0376|srpS|sp|Q9R9U0|SRPS_PSEPU HTH-type transcriptional regulator SrpS OS=Pseudomonas putida GN=srpS PE=
BAC0500|emrBsm|tr|B2FID0|B2FID0_STRMK Putative multidrug resistance protein B OS=Stenotrophomonas maltophili
BAC0207|mdeA|sp|P13254|MEGL_PSEPU Methionine gamma-lyase OS=Pseudomonas putida GN=mdeA PE=1 SV=2
BAC0502|emrRsm|tr|B2FIC7|B2FIC7_STRMK Putative MarR-family transcriptional regulator OS=Stenotrophomonas ma
BAC0190|hmrR|sp|Q9X5X4|HMRR_SINMW HTH-type transcriptional regulator HmrR OS=Sinorhizobium medicae (stra
BAC0310|perR|sp|Q2G282|PERR_STAA8 Peroxide-responsive repressor PerR OS=Staphylococcus aureus (strain NCTC 8
BAC0258|mtrD|tr|Q5F725|Q5F725_NEIG1 Antibiotic resistance efflux pump component OS=Neisseria gonorrhoeae (strai
BAC0473|adeC|tr|Q93E18|Q93E18_ACIBA AdeC outer membrane protein OS=Acinetobacter baumannii GN=adeC PE=4
BAC0363|smfY|tr|Q2AAU2|Q2AAU2_SERMA Multidrug efflux pump SmfY OS=Serratia marcescens GN=smfY PE=4 S
BAC0184|hasF|tr|Q6GW09|Q6GW09_SERMA TolC-like protein OS=Serratia marcescens PE=4 SV=1
BAC0407|ttgD|sp|Q9KWV5|TTGD_PSEPT Toluene efflux pump periplasmic linker protein TtgD OS=Pseudomonas putid
BAC0236|mexC|tr|Q51395|Q51395_PSEAI Membrane fusion protein MexC OS=Pseudomonas aeruginosa GN=mexC PE=
BAC0338|sdeX|tr|Q7WSD6|Q7WSD6_SERMA Multidrug efflux pump SdeX OS=Serratia marcescens GN=sdeX PE=4 S
BAC0358|smeD|tr|I0KSX9|I0KSX9_STEMA Membrane fusion protein of RND family multidrug efflux pump OS=Stenot
BAC0014|adeF|tr|Q2FD82|Q2FD82_ACIBA Putative RND family drug transporter OS=Acinetobacter baumannii GN=29_
BAC0513|vmeF|tr|Q87R57|Q87R57_VIBPA Putative multidrug resistance protein OS=Vibrio parahaemolyticus serotype
BAC0661|merB2|tr|Q7DJN2|Q7DJN2_BACME MerB2 OS=Bacillus megaterium GN=merB2 PE=4 SV=1
BAC0413|ttgR|sp|Q9AIU0|TTGR_PSEPT HTH-type transcriptional regulator TtgR OS=Pseudomonas putida (strain DOT-
BAC0657|merB|tr|O07303|O07303_9PSED Alkylmercury lyase OS=Pseudomonas sp. K-62 GN=merB PE=3 SV=2
BAC0506|pcm|tr|R4IUI7|R4IUI7_STEMA Protein-L-isoaspartate O-methyltransferase OS=Stenotrophomonas maltophilia
BAC0566|actS|tr|Q52912|Q52912_9RHIZ Histidine protein kinase OS=Sinorhizobium medicae GN=actS PE=4 SV=1
BAC0654|merB1|sp|P16172|MERB_BACCE Alkylmercury lyase OS=Bacillus cereus GN=merB1 PE=3 SV=2
BAC0362|smeT|tr|Q5MXJ5|Q5MXJ5_STEMA Repressor SmeT OS=Stenotrophomonas maltophilia GN=smeT PE=4 SV=
BAC0481|pdrM|sp|Q8DPQ6|NORM_STRR6 Probable multidrug resistance protein NorM OS=Streptococcus pneumoniae
BAC0597|baeS|tr|D0ZNE2|D0ZNE2_SALT1 Signal transduction histidine-protein kinase BaeS OS=Salmonella typhimuri
BAC0367|smvA/emrB|sp|D0ZXQ3|SMVA_SALT1 Methyl viologen resistance protein SmvA OS=Salmonella typhimuriu
BAC0248|mexY|tr|Q9ZNG8|Q9ZNG8_PSEAI MexY OS=Pseudomonas aeruginosa GN=mexY PE=4 SV=1
BAC0260|mtrF|tr|B4RN92|B4RN92_NEIG2 Antibiotic resistance efflux pump component OS=Neisseria gonorrhoeae (stra
BAC0521|vmeK|tr|Q87LY6|Q87LY6_VIBPA Putative multidrug resistance protein OS=Vibrio parahaemolyticus serotype
BAC0408|ttgE|sp|Q9KWV4|TTGE_PSEPT Toluene efflux pump membrane transporter TtgE OS=Pseudomonas putida (str
BAC0380|tbtB|tr|Q71UZ6|Q71UZ6_PSEST Resistance nodulation cell division family member TbtB OS=Pseudomonas st
BAC0474|adeD|tr|Q67GM1|Q67GM1_ACIG3 AdeD OS=Acinetobacter sp. 4365 GN=adeD PE=4 SV=1
BAC0042|bepC|sp|Q8G0Y6|BEPC_BRUSU Outer membrane efflux protein BepC OS=Brucella suis biovar 1 (strain 1330)
BAC0173|gadB|sp|P69910|DCEB_ECOLI Glutamate decarboxylase beta OS=Escherichia coli (strain K12) GN=gadB PE=
BAC0360|smeF|tr|Q9F239|Q9F239_STEMA Outer membrane protein OS=Stenotrophomonas maltophilia GN=smeF PE=4
BAC0562|oprN|tr|P95423|P95423_PSEAI Uncharacterized protein OS=Pseudomonas aeruginosa GN=oprN PE=4 SV=1
BAC0016|adeH|tr|Q2FD80|Q2FD80_ACIBA Putative RND family drug transporter OS=Acinetobacter baumannii GN=29_
BAC0406|ttgC|sp|Q9WWZ8|TTGC_PSEPT Toluene efflux pump outer membrane protein TtgC OS=Pseudomonas putida (
BAC0504|farB|tr|Q9RQ29|Q9RQ29_NEIGO Efflux pump protein FarB OS=Neisseria gonorrhoeae PE=4 SV=1
BAC0419|vceB|tr|O51919|O51919_VIBCL VceB OS=Vibrio cholerae GN=vceB PE=4 SV=1
BAC0046|bepG|sp|Q8FWV9|BEPG_BRUSU Efflux pump membrane transporter BepG OS=Brucella suis biovar 1 (strain 1
BAC0425|vexD|tr|Q9KR85|Q9KR85_VIBCH Transporter, AcrB/D/F family OS=Vibrio cholerae serotype O1 (strain ATC
BAC0411|ttgH|sp|Q93PU4|TTGH_PSEPT Toluene efflux pump membrane transporter TtgH OS=Pseudomonas putida (stra
BAC0324|qacF|sp|Q9X2N9|QACF_ENTAE Quaternary ammonium compound-resistance protein QacF OS=Enterobacter a
BAC0688|merR2|tr|Q79B70|Q79B70_PSEST Organomercurial resistance regulatory protein OS=Pseudomonas stutzeri GN
BAC0648|merA|sp|P08662|MERA_SERMA Mercuric reductase (Fragments) OS=Serratia marcescens GN=merA PE=3 SV

BAC0357|recG|tr|B5L350|B5L350_9PSED ATP-dependent DNA helicase (Fragment) OS=Pseudomonas corrugata GN=rec
BAC0293|ruvB|tr|B5L348|B5L348_9PSED Malic enzyme family protein (Fragment) OS=Pseudomonas corrugata PE=3 SV
BAC0706|sodB|sp|P53641|SODF_PSEAE Superoxide dismutase [Fe] OS=Pseudomonas aeruginosa (strain ATCC 15692 /
BAC0565|actR|sp|A6UEL7|ACTR_SINMW Acid tolerance regulatory protein ActR OS=Sinorhizobium medicae (strain W
BAC0322|qacE|sp|P0AGC9|QACE_ECOLX Quaternary ammonium compound-resistance protein QacE OS=Escherichia co
BAC0018|adeJ|tr|Q24LT7|Q24LT7_ACIBA AdeJ OS=Acinetobacter baumannii GN=adeJ PE=4 SV=1
BAC0047|bexA|tr|Q93HR0|Q93HR0_BACT4 BexA OS=Bacteroides thetaiotaomicron GN=bexA PE=4 SV=1
BAC0335|rpoS|sp|P35540|RPOS_SHIFL RNA polymerase sigma factor RpoS OS=Shigella flexneri GN=rpoS PE=3 SV=3
BAC0564|actP/yjcG|sp|P32705|ACTP_ECOLI Cation/acetate symporter ActP OS=Escherichia coli (strain K12) GN=actP
BAC0364|smrA|tr|C7SLZ1|C7SLZ1_STEMA ABC-type multidrug efflux pump (Fragment) OS=Stenotrophomonas maltop
BAC0242|mexK|tr|Q9HXW4|Q9HXW4_PSEAE Probable Resistance-Nodulation-Cell Division (RND) efflux transporter
BAC0019|adeK|tr|Q24LT6|Q24LT6_ACIBA AdeK OS=Acinetobacter baumannii GN=adeK PE=4 SV=1
BAC0323|qacEdelta1|tr|Q7BQY4|Q7BQY4_PSEAI Disinfectant resistance protein OS=Pseudomonas aeruginosa GN=qac
BAC0017|adeI|tr|Q2FD95|Q2FD95_ACIBA AdeI OS=Acinetobacter baumannii GN=adeI PE=4 SV=1
BAC0239|mexF|tr|Q4KBN7|Q4KBN7_PSEF5 Multidrug efflux RND transporter, permease protein MexF OS=Pseudomona
BAC0532|cpxA|tr|C4WZK5|C4WZK5_KLEPN Sensor protein of stress-related two-component regulatory system OS=Kle
BAC0536|oxyRkp|tr|C4WZN6|C4WZN6_KLEPN Activator of hydrogen peroxide-inducible genes OS=Klebsiella pneumo
BAC0533|cpxR|tr|C4WZK6|C4WZK6_KLEPN Response regulator of stress-related two-component regulatory system OS
BAC0194|ibpA|sp|P0C054|IBPA_ECOLI Small heat shock protein IbpA OS=Escherichia coli (strain K12) GN=ibpA PE=1
BAC0143|emhB|tr|C1KA85|C1KA85_PSEFL EmhB OS=Pseudomonas fluorescens GN=emhB PE=4 SV=1
BAC0001|abeM|tr|Q5FAM9|Q5FAM9_ACIBA Multidrug efflux pump AbeM OS=Acinetobacter baumannii GN=abeM PE
BAC0368|sodA|sp|P00448|SODM_ECOLI Superoxide dismutase [Mn] OS=Escherichia coli (strain K12) GN=sodA PE=1
BAC0211|mdtB/yegN|sp|P76398|MDTB_ECOLI Multidrug resistance protein MdtB OS=Escherichia coli (strain K12) GN
BAC0659|merB|sp|P08664|MERB_SERMA Alkylmercury lyase OS=Serratia marcescens GN=merB PE=3 SV=1
BAC0559|emrR|sp|P0ACR9|MPRA_ECOLI Transcriptional repressor MprA OS=Escherichia coli (strain K12) GN=mprA
BAC0295|oqxB|tr|Q69HW2|Q69HW2_ECOLX OqxB integral membrane protein OS=Escherichia coli GN=oqxB PE=4 SV
BAC0006|acrB|sp|P31224|ACRB_ECOLI Multidrug efflux pump subunit AcrB OS=Escherichia coli (strain K12) GN=acr
BAC0015|adeG|tr|Q2FD81|Q2FD81_ACIBA Cation/multidrug efflux pump OS=Acinetobacter baumannii GN=29_167 PE
BAC0417|vcaM|tr|Q9KKV4|Q9KKV4_VIBCH ABC transporter, ATP-binding protein OS=Vibrio cholerae serotype O1 (s
BAC0595|arsH|tr|P74312|P74312_SYNY3 Slr0945 protein OS=Synechocystis sp. (strain PCC 6803 / Kazusa) GN=slr0945
BAC0156|fabI|sp|P0AEK4|FABI_ECOLI Enoyl-[acyl-carrier-protein] reductase [NADH] FabI OS=Escherichia coli (strain
BAC0148|emrB|sp|P0AEJ0|EMRB_ECOLI Multidrug resistance protein B OS=Escherichia coli (strain K12) GN=emrB PE
BAC0246|mexW|tr|Q9HW27|Q9HW27_PSEAE Probable Resistance-Nodulation-Cell Division (RND) efflux transporter O
BAC0370|soxR|sp|P0ACS2|SOXR_ECOLI Redox-sensitive transcriptional activator SoxR OS=Escherichia coli (strain K1
BAC0530|phoB|tr|C4X6T6|C4X6T6_KLEPN Response regulator in two-component regulatory system with PhoQ OS=Kle
BAC0181|glpF|sp|P0AER0|GLPF_ECOLI Glycerol uptake facilitator protein OS=Escherichia coli (strain K12) GN=glpF P
BAC0039|baeR|sp|P69228|BAER_ECOLI Transcriptional regulatory protein BaeR OS=Escherichia coli (strain K12) GN=b
BAC0371|soxS|sp|P0A9E2|SOXS_ECOLI Regulatory protein SoxS OS=Escherichia coli (strain K12) GN=soxS PE=1 SV=
BAC0196|iclR|sp|P16528|ICLR_ECOLI Acetate operon repressor OS=Escherichia coli (strain K12) GN=iclR PE=1 SV=1
BAC0005|acrA|sp|P0AE06|ACRA_ECOLI Multidrug efflux pump subunit AcrA OS=Escherichia coli (strain K12) GN=ac
BAC0149|emrD|sp|P31442|EMRD_ECOLI Multidrug resistance protein D OS=Escherichia coli (strain K12) GN=emrD PE
BAC0222|mepB|sp|P0C070|MEPB_PSEPU Multidrug/solvent efflux pump membrane transporter MepB OS=Pseudomona
BAC0351|sitC|tr|Q9XCS0|Q9XCS0_SALTM SitC OS=Salmonella typhimurium GN=sitC PE=3 SV=1
BAC0212|mdtC/yegO|sp|P76399|MDTC_ECOLI Multidrug resistance protein MdtC OS=Escherichia coli (strain K12) GN
BAC0008|acrD/yffA|sp|P24177|ACRD_ECOLI Probable aminoglycoside efflux pump OS=Escherichia coli (strain K12) G
BAC0446|yhcN|sp|P64614|YHCN_ECOLI Uncharacterized protein YhcN OS=Escherichia coli (strain K12) GN=yhcN PE

BAC0334|robA|sp|P0ACI0|ROB_ECOLI Right origin-binding protein OS=Escherichia coli (strain K12) GN=rob PE=1 SV
BAC0010|acrF/envD|sp|P24181|ACRF_ECOLI Acriflavine resistance protein F OS=Escherichia coli (strain K12) GN=acrF
BAC0145|emhR|tr|Q4KH21|Q4KH21_PSEF5 Transcriptional regulator EmhR OS=Pseudomonas fluorescens (strain Pf-5 /
BAC0434|ychH|sp|P0AB49|YCHH_ECOLI Uncharacterized protein YchH OS=Escherichia coli (strain K12) GN=ychH PE
BAC0041|bcr|sp|P28246|BCR_ECOLI Bicyclomycin resistance protein OS=Escherichia coli (strain K12) GN=bcr PE=1 S
BAC0541|yieF|sp|P0AGE6|YIEF_ECOLI Uncharacterized protein YieF OS=Escherichia coli (strain K12) GN=yieF PE=1
BAC0185|hdeA/yhiB|sp|P0AES9|HDEA_ECOLI Acid stress chaperone HdeA OS=Escherichia coli (strain K12) GN=hdeA
BAC0244|mexT|tr|O87785|O87785_PSEAI MexT protein OS=Pseudomonas aeruginosa GN=mexT PE=4 SV=1
BAC0393|tolC|sp|P02930|TOLC_ECOLI Outer membrane protein TolC OS=Escherichia coli (strain K12) GN=tolC PE=1
BAC0061|cepA|sp|Q8RR17|FIEF_KLEPN Cation-efflux pump FieF OS=Klebsiella pneumoniae GN=fieF PE=3 SV=1
BAC0235|mexB|sp|P52002|MEXB_PSEAE Multidrug resistance protein MexB OS=Pseudomonas aeruginosa (strain ATC
BAC0215|mdtG/yceE|sp|P25744|MDTG_ECOLI Multidrug resistance protein MdtG OS=Escherichia coli (strain K12) GN
BAC0218|mdtK/ydhE|sp|P37340|MDTK_ECOLI Multidrug resistance protein MdtK OS=Escherichia coli (strain K12) GN
BAC0296|ostA/lptD|sp|P31554|LPTD_ECOLI LPS-assembly protein LptD OS=Escherichia coli (strain K12) GN=lptD PE
BAC0491|kdeA|tr|A6T6T9|A6T6T9_KLEP7 Multidrug/chloramphenicol efflux transport protein (MFS family) OS=Klebsi
BAC0560|marA|sp|P0ACH5|MARA_ECOLI Multiple antibiotic resistance protein MarA OS=Escherichia coli (strain K12)
BAC0176|gadW/yhiW|sp|P63201|GADW_ECOLI HTH-type transcriptional regulator GadW OS=Escherichia coli (strain K
BAC0210|mdtA/yegM|sp|P76397|MDTA_ECOLI Multidrug resistance protein MdtA OS=Escherichia coli (strain K12) GN
BAC0144|emhC|tr|Q4KH24|Q4KH24_PSEF5 Efflux transporter, outer membrane factor lipoprotein EmhC OS=Pseudomon
BAC0531|phoR|tr|C4X6T5|C4X6T5_KLEPN Sensor kinase in two-component regulatory system with PhoP OS=Klebsiella
BAC0009|acrE/envC|sp|P24180|ACRE_ECOLI Acriflavine resistance protein E OS=Escherichia coli (strain K12) GN=acr
BAC0195|ibpB|sp|P0C058|IBPB_ECOLI Small heat shock protein IbpB OS=Escherichia coli (strain K12) GN=ibpB PE=1
BAC0494|eefA|tr|A8CY69|A8CY69_KLEPN EefA OS=Klebsiella pneumoniae GN=eefA PE=4 SV=1
BAC0208|mdfA/cmr|sp|P0AEY8|MDFA_ECOLI Multidrug transporter MdfA OS=Escherichia coli (strain K12) GN=mdfA
BAC0012|actP|sp|Q9X5X3|ATCU_SINMW Copper-transporting P-type ATPase OS=Sinorhizobium medicae (strain WSM
BAC0166|fetB/ybbM|sp|P77307|YBBM_ECOLI UPF0014 inner membrane protein YbbM OS=Escherichia coli (strain K1
BAC0214|mdtF/yhiV|sp|P37637|MDTF_ECOLI Multidrug resistance protein MdtF OS=Escherichia coli (strain K12) GN=
BAC0172|gadA|sp|P69908|DCEA_ECOLI Glutamate decarboxylase alpha OS=Escherichia coli (strain K12) GN=gadA PE
BAC0529|kpnO|tr|C4XBC3|C4XBC3_KLEPN Outer membrane porin protein C OS=Klebsiella pneumoniae subsp. pneum
BAC0165|fetA/ybbL|sp|P77279|YBBL_ECOLI Uncharacterized ABC transporter ATP-binding protein YbbL OS=Escheric
BAC0147|emrA|sp|P27303|EMRA_ECOLI Multidrug resistance protein A OS=Escherichia coli (strain K12) GN=emrA PE
BAC0011|acrR/ybaH|sp|P0ACS9|ACRR_ECOLI HTH-type transcriptional regulator AcrR OS=Escherichia coli (strain K1
BAC0477|kpnF|tr|C4X7Z4|C4X7Z4_KLEPN Spermidine export protein MdtI OS=Klebsiella pneumoniae subsp. pneumon
BAC0450|ymgB/ariR|sp|P75993|ARIR_ECOLI Probable two-component-system connector protein AriR OS=Escherichia c
BAC0437|ydeO|sp|P76135|YDEO_ECOLI HTH-type transcriptional regulator YdeO OS=Escherichia coli (strain K12) GN
BAC0294|oqxA|tr|Q69HW3|Q69HW3_ECOLX OqxA membrane-fusion protein OS=Escherichia coli GN=oqxA PE=4 SV
BAC0142|emhA|tr|Q4KH22|Q4KH22_PSEF5 Efflux transporter, membrane fusion protein subunit EmhA OS=Pseudomona
BAC0359|smeE|tr|I0KSX8|I0KSX8_STEMA RND efflux system, inner membrane transporter OS=Stenotrophomonas malt
BAC0040|baeS|sp|P30847|BAES_ECOLI Signal transduction histidine-protein kinase BaeS OS=Escherichia coli (strain K1
BAC0029|chrF|tr|A4UQR2|A4UQR2_9RHIZ ChrF OS=Ochrobactrum tritici GN=chrF PE=4 SV=1
BAC0038|asr|sp|P36560|ASR_ECOLI Acid shock protein OS=Escherichia coli (strain K12) GN=asr PE=1 SV=3
BAC0493|kmrA|tr|C4X8X9|C4X8X9_KLEPN Energy-dependent efflux protein for methyl viologen resistance OS=Klebsie
BAC0186|hdeB/yhiC|sp|P0AET2|HDEB_ECOLI Acid stress chaperone HdeB OS=Escherichia coli (strain K12) GN=hdeB
BAC0350|sitB|tr|Q9XCS1|Q9XCS1_SALTM SitB OS=Salmonella typhimurium GN=sitB PE=3 SV=1
BAC0219|mdtM/yjiO|sp|P39386|MDTM_ECOLI Multidrug resistance protein MdtM OS=Escherichia coli (strain K12) GN
BAC0177|gadX|sp|P37639|GADX_ECOLI HTH-type transcriptional regulator GadX OS=Escherichia coli (strain K12) GN

BAC0707|sodB|sp|P0AGD3|SODF_ECOLI Superoxide dismutase [Fe] OS=Escherichia coli (strain K12) GN=sodB PE=1 S
BAC0175|gadE/yhiE|sp|P63204|GADE_ECOLI Transcriptional regulator GadE OS=Escherichia coli (strain K12) GN=gad
BAC0358|oscA|tr|B6CM35|B6CM35_9PSED Putative uncharacterized protein oscA OS=Pseudomonas corrugata GN=osc
BAC0438|ydeP|sp|P77561|YDEP_ECOLI Protein YdeP OS=Escherichia coli (strain K12) GN=ydeP PE=2 SV=1
BAC0496|adeN|tr|B7H1T7|B7H1T7_ACIB3 Bacterial regulatory protein, tetR family protein OS=Acinetobacter baumanni
BAC0220|mdtN/yjcR|sp|P32716|MDTN_ECOLI Multidrug resistance protein MdtN OS=Escherichia coli (strain K12) GN=
BAC0155|evgS|sp|P58402|EVGS_ECO57 Sensor protein EvgS OS=Escherichia coli O157:H7 GN=evgS PE=3 SV=1
BAC0106|cuiD|sp|Q8ZRS2|CUEO_SALTY Blue copper oxidase CueO OS=Salmonella typhimurium (strain LT2 / SGSC1
BAC0151|emrK|sp|P52599|EMRK_ECOLI Multidrug resistance protein K OS=Escherichia coli (strain K12) GN=emrK PE
BAC0013|adeE|tr|Q8GKU1|Q8GKU1_ACIG3 AdeE OS=Acinetobacter sp. 4365 GN=adeE PE=4 SV=2
BAC0135|dpsA|tr|Q8KR86|Q8KR86_BURPE DpsA OS=Burkholderia pseudomallei GN=dpsA PE=3 SV=1
BAC0498|ideR|sp|P0A672|IDER_MYCTU Iron-dependent repressor IdeR OS=Mycobacterium tuberculosis GN=ideR PE=
BAC0384|tehA|sp|P25396|TEHA_ECOLI Tellurite resistance protein TehA OS=Escherichia coli (strain K12) GN=tehA PE
BAC0650|merA|tr|O08449|O08449_9PSED Mercuric reductase OS=Pseudomonas sp. K-62 GN=merA PE=4 SV=1
BAC0157|fabK|tr|Q9FBC5|Q9FBC5_STREE Trans-2-enoyl-ACP reductase II OS=Streptococcus pneumoniae GN=fabK P
BAC0339|sdeY|tr|Q7WSD5|Q7WSD5_SERMA Multidrug efflux pump SdeY OS=Serratia marcescens GN=sdeY PE=4 S
BAC0216|mdtI/ydgE|sp|P69210|MDTI_ECOLI Spermidine export protein MdtI OS=Escherichia coli (strain K12) GN=md
BAC0451|yodD|sp|P64519|YODD_ECOLI Uncharacterized protein YodD OS=Escherichia coli (strain K12) GN=yodD PE
BAC0238|mexE|tr|Q1IB41|Q1IB41_PSEE4 Multidrug efflux RND membrane fusion protein MexE OS=Pseudomonas ento
BAC0213|mdtE/yhiU|sp|P37636|MDTE_ECOLI Multidrug resistance protein MdtE OS=Escherichia coli (strain K12) GN=
BAC0153|emrY|sp|P52600|EMRY_ECOLI Multidrug resistance protein Y OS=Escherichia coli (strain K12) GN=emrY PE
BAC0385|tehB|sp|P25397|TEHB_ECOLI Tellurite methyltransferase OS=Escherichia coli (strain K12) GN=tehB PE=1 SV
BAC0596|baeR|tr|D0ZNE3|D0ZNE3_SALT1 DNA-binding transcriptional regulator BaeR OS=Salmonella typhimurium (s
BAC0353|smdA|tr|A7VN01|A7VN01_SERMA Multidrug efflux pump SmdA OS=Serratia marcescens GN=smdA PE=3 S
BAC0447|yjaA|sp|P09162|YJAA_ECOLI Uncharacterized protein YjaA OS=Escherichia coli (strain K12) GN=yjaA PE=4
BAC0445|ygiW|sp|P0ADU5|YGIW_ECOLI Protein YgiW OS=Escherichia coli (strain K12) GN=ygiW PE=1 SV=1
BAC0027|chrB|tr|A4UQR5|A4UQR5_9RHIZ ChrB OS=Ochrobactrum tritici GN=chrB PE=4 SV=1
BAC0146|emmdR|tr|D5CJ69|D5CJ69_ENTCC MATE efflux family protein OS=Enterobacter cloacae subsp. cloacae (strai
BAC0190|hmrR|sp|Q9X5X4|HMRR_SINMW HTH-type transcriptional regulator HmrR OS=Sinorhizobium medicae (strai
BAC0705|sodA|sp|P53652|SODM_PSEAE Superoxide dismutase [Mn] OS=Pseudomonas aeruginosa (strain ATCC 15692
BAC0349|sitA|tr|Q9XCS2|Q9XCS2_SALTI Iron transport protein, periplasmic-binding protein OS=Salmonella typhi GN=
BAC0472|adeB|tr|Q93E19|Q93E19_ACIBA AdeB RND protein OS=Acinetobacter baumannii GN=adeB PE=4 SV=1
BAC0337|sdeB|tr|Q84GI9|Q84GI9_SERMA Putative resistance-nodulation cell division protein SdeB OS=Serratia marcesc
BAC0378|sugE|sp|P69937|SUGE_ECOLI Quaternary ammonium compound-resistance protein SugE OS=Escherichia coli
BAC0352|sitD|tr|Q9XCR9|Q9XCR9_SALTM SitD OS=Salmonella typhimurium GN=sitD PE=3 SV=1
BAC0134|dpr/dps|sp|P0CB53|DPS_STRSU DNA protection during starvation protein OS=Streptococcus suis GN=dps PE=
BAC0217|mdtJ/ebrB/ydgF|sp|Q3Z1V3|MDTJ_SHISS Spermidine export protein MdtJ OS=Shigella sonnei (strain Ss046) G
BAC0174|gadC/xasA|sp|P63235|GADC_ECOLI Probable glutamate/gamma-aminobutyrate antiporter OS=Escherichia coli
BAC0154|evgA|sp|P0ACZ4|EVGA_ECOLI Positive transcription regulator EvgA OS=Escherichia coli (strain K12) GN=e
BAC0561|marR|sp|P27245|MARR_ECOLI Multiple antibiotic resistance protein MarR OS=Escherichia coli (strain K12) G
BAC0229|merG|tr|O07302|O07302_9PSED Mercuric resistance protein OS=Pseudomonas sp. K-62 GN=merG PE=4 SV=
BAC0492|kexD|tr|A6TA71|A6TA71_KLEP7 Acridine efflux pump OS=Klebsiella pneumoniae subsp. pneumoniae (strain
BAC0511|vmeD|tr|Q87TN1|Q87TN1_VIBPA Putative multidrug resistance protein OS=Vibrio parahaemolyticus serotype
BAC0404|ttgA|sp|Q9WWZ9|TTGA_PSEPT Toluene efflux pump periplasmic linker protein TtgA OS=Pseudomonas putid
BAC0499|emrAsm|tr|B2FIC9|B2FIC9_STRMK Putative multidrug resistance protein A OS=Stenotrophomonas maltophili
BAC0141|emeA|tr|Q8GR72|Q8GR72_ENTFL Multidrug efflux pump OS=Enterococcus faecalis GN=emeA PE=4 SV=1

BAC0471|adeA|tr|Q93E20|Q93E20_ACIBA AdeA membrane fusion protein OS=Acinetobacter baumannii GN=adeA PE=4
BAC0237|mexD|tr|Q51396|Q51396_PSEAI RND family exporter MexD OS=Pseudomonas aeruginosa GN=mexD PE=4 S
BAC0179|gesB|tr|Q8ZRG9|Q8ZRG9_SALTY Putative cation efflux system protein OS=Salmonella typhimurium (strain L
BAC0252|mntP/yebN|sp|P76264|MNTP_ECOLI Probable manganese efflux pump MntP OS=Escherichia coli (strain K12)
BAC0358|smeD|tr|I0KSX9|I0KSX9_STEMA Membrane fusion protein of RND family multidrug efflux pump OS=Stenotr
BAC0028|chrC|tr|A4UQR3|A4UQR3_9RHIZ Superoxide dismutase OS=Ochrobactrum tritici GN=chrC PE=3 SV=1
BAC0566|actS|tr|Q52912|Q52912_9RHIZ Histidine protein kinase OS=Sinorhizobium medicae GN=actS PE=4 SV=1
BAC0150|emrE/mvrC|sp|P23895|EMRE_ECOLI Multidrug transporter EmrE OS=Escherichia coli (strain K12) GN=emrE
BAC0236|mexC|tr|Q51395|Q51395_PSEAI Membrane fusion protein MexC OS=Pseudomonas aeruginosa GN=mexC PE=
BAC0436|ydeI|sp|P31130|YDEI_ECOLI Uncharacterized protein YdeI OS=Escherichia coli (strain K12) GN=ydeI PE=4 S
BAC0258|mtrD|tr|Q5F725|Q5F725_NEIG1 Antibiotic resistance efflux pump component OS=Neisseria gonorrhoeae (strain
BAC0505|farR|tr|Q7DD70|Q7DD70_NEIMB Transcriptional regulator, MarR family OS=Neisseria meningitidis serogroup
BAC0435|yddg/emrE|sp|D0ZXP9|YDDG_SALT1 Methyl viologen resistance protein YddG OS=Salmonella typhimurium
BAC0313|pmpM|sp|Q9I3Y3|PMPM_PSEAE Multidrug resistance protein PmpM OS=Pseudomonas aeruginosa (strain AT
BAC0184|hasF|tr|Q6GW09|Q6GW09_SERMA TolC-like protein OS=Serratia marcescens PE=4 SV=1
BAC0424|vexB|tr|Q9KVI2|Q9KVI2_VIBCH Multidrug resistance protein, putative OS=Vibrio cholerae serotype O1 (strai
BAC0248|mexY|tr|Q9ZNG8|Q9ZNG8_PSEAI MexY OS=Pseudomonas aeruginosa GN=mexY PE=4 SV=1
BAC0290|opmD/nmpC|sp|P37592|OMPD_SALTY Outer membrane porin protein OmpD OS=Salmonella typhimurium (st
BAC0354|smdB|tr|A7VN02|A7VN02_SERMA Multidrug efflux pump SmdB OS=Serratia marcescens GN=smdB PE=3 S
BAC0207|mdeA|sp|P13254|MEGL_PSEPU Methionine gamma-lyase OS=Pseudomonas putida GN=mdeA PE=1 SV=2
BAC0014|adeF|tr|Q2FD82|Q2FD82_ACIBA Putative RND family drug transporter OS=Acinetobacter baumannii GN=29_
BAC0661|merB2|tr|Q7DJN2|Q7DJN2_BACME MerB2 OS=Bacillus megaterium GN=merB2 PE=4 SV=1
BAC0380|tbtB|tr|Q71UZ6|Q71UZ6_PSEST Resistance nodulation cell division family member TbtB OS=Pseudomonas stu
BAC0506|pcm|tr|R4IUI7|R4IUI7_STEMA Protein-L-isoaspartate O-methyltransferase OS=Stenotrophomonas maltophilia
BAC0481|pdrM|sp|Q8DPQ6|NORM_STRR6 Probable multidrug resistance protein NorM OS=Streptococcus pneumoniae (
BAC0173|gadB|sp|P69910|DCEB_ECOLI Glutamate decarboxylase beta OS=Escherichia coli (strain K12) GN=gadB PE=
BAC0597|baeS|tr|D0ZNE2|D0ZNE2_SALT1 Signal transduction histidine-protein kinase BaeS OS=Salmonella typhimuriu
BAC0479|adeT2|tr|C7F8K7|C7F8K7_ACIBA AdeT2 OS=Acinetobacter baumannii PE=4 SV=1
BAC0662|merB3|tr|Q9RHR0|Q9RHR0_BACME MerB3 OS=Bacillus megaterium GN=merB3 PE=4 SV=2
BAC0223|mepC|sp|P0C071|MEPC_PSEPU Multidrug/solvent efflux pump outer membrane protein MepC OS=Pseudomor
BAC0367|smvA/emrB|sp|D0ZXQ3|SMVA_SALT1 Methyl viologen resistance protein SmvA OS=Salmonella typhimuriu
BAC0501|emrCsm|tr|B2FIC8|B2FIC8_STRMK Putative outer membrane multidrug efflux protein OS=Stenotrophomonas
BAC0240|mexI|tr|Q9HWH4|Q9HWH4_PSEAE Probable Resistance-Nodulation-Cell Division (RND) efflux transporter O
BAC0509|vexH|tr|Q9KTI8|Q9KTI8_VIBCH Multidrug resistance protein, putative OS=Vibrio cholerae serotype O1 (strai
BAC0408|ttgE|sp|Q9KWV4|TTGE_PSEPT Toluene efflux pump membrane transporter TtgE OS=Pseudomonas putida (str
BAC0508|adeL|tr|A3M732|A3M732_ACIBT Transcriptional regulator LysR family OS=Acinetobacter baumannii (strain A
BAC0044|bepE|sp|Q8G2M6|BEPE_BRUSU Efflux pump membrane transporter BepE OS=Brucella suis biovar 1 (strain 13
BAC0430|vmeB|tr|Q2AAU3|Q2AAU3_VIBPH Inner membrane protein VmeB OS=Vibrio parahaemolyticus GN=vmeB P
BAC0043|bepD|sp|Q8G2M7|BEPD_BRUSU Efflux pump periplasmic linker BepD OS=Brucella suis biovar 1 (strain 1330
BAC0159|fabV|sp|Q9KRA3|Y1738_VIBCH Putative reductase VC_1738/VC_1739 OS=Vibrio cholerae serotype O1 (strai
BAC0360|smeF|tr|Q9F239|Q9F239_STEMA Outer membrane protein OS=Stenotrophomonas maltophilia GN=smeF PE=4
BAC0412|ttgI|sp|Q93PU3|TTGI_PSEPT Toluene efflux pump outer membrane protein TtgI OS=Pseudomonas putida (strai
BAC0016|adeH|tr|Q2FD80|Q2FD80_ACIBA Putative RND family drug transporter OS=Acinetobacter baumannii GN=29_
BAC0614|cmeC|tr|Q8RTE3|Q8RTE3_CAMJU CmeC OS=Campylobacter jejuni GN=cmeC PE=4 SV=1
BAC0526|vmeV|tr|Q87HZ7|Q87HZ7_VIBPA Transporter, AcrB/D/F family OS=Vibrio parahaemolyticus serotype O3:K6
BAC0419|vceB|tr|O51919|O51919_VIBCL VceB OS=Vibrio cholerae GN=vceB PE=4 SV=1

BAC0319|qacA|sp|P0A0J9|QACA_STAAU Antiseptic resistance protein OS=Staphylococcus aureus GN=qacA PE=1 SV=
BAC0411|ttgH|sp|Q93PU4|TTGH_PSEPT Toluene efflux pump membrane transporter TtgH OS=Pseudomonas putida (stra
BAC0500|emrBsm|tr|B2FID0|B2FID0_STRMK Putative multidrug resistance protein B OS=Stenotrophomonas maltophili
BAC0188|hefC|tr|B6JLJ0|B6JLJ0_HELP2 Cytoplasmic pump protein of the HefABC efflux system HefC OS=Helicobacte
BAC0427|vexF|tr|A6P7H3|A6P7H3_VIBCL Multidrug efflux transporter VexF OS=Vibrio cholerae GN=vexF PE=4 SV=1
BAC0521|vmeK|tr|Q87LY6|Q87LY6_VIBPA Putative multidrug resistance protein OS=Vibrio parahaemolyticus serotype
BAC0046|bepG|sp|Q8FWV9|BEPG_BRUSU Efflux pump membrane transporter BepG OS=Brucella suis biovar 1 (strain 1
BAC0688|merR2|tr|Q79B70|Q79B70_PSEST Organomercurial resistance regulatory protein OS=Pseudomonas stutzeri GN
BAC0018|adeJ|tr|Q24LT7|Q24LT7_ACIBA AdeJ OS=Acinetobacter baumannii GN=adeJ PE=4 SV=1
BAC0648|merA|sp|P08662|MERA_SERMA Mercuric reductase (Fragments) OS=Serratia marcescens GN=merA PE=3 SV
BAC0324|qacF|sp|Q9X2N9|QACF_ENTAE Quaternary ammonium compound-resistance protein QacF OS=Enterobacter a
BAC0019|adeK|tr|Q24LT6|Q24LT6_ACIBA AdeK OS=Acinetobacter baumannii GN=adeK PE=4 SV=1
BAC0357|recG|tr|B5L350|B5L350_9PSED ATP-dependent DNA helicase (Fragment) OS=Pseudomonas corrugata GN=rec
BAC0565|actR|sp|A6UEL7|ACTR_SINMW Acid tolerance regulatory protein ActR OS=Sinorhizobium medicae (strain W
BAC0017|adeI|tr|Q2FD95|Q2FD95_ACIBA AdeI OS=Acinetobacter baumannii GN=adeI PE=4 SV=1
BAC0001|abeM|tr|Q5FAM9|Q5FAM9_ACIBA Multidrug efflux pump AbeM OS=Acinetobacter baumannii GN=abeM PE
BAC0706|sodB|sp|P53641|SODF_PSEAE Superoxide dismutase [Fe] OS=Pseudomonas aeruginosa (strain ATCC 15692 /
BAC0322|qacE|sp|P0AGC9|QACE_ECOLX Quaternary ammonium compound-resistance protein QacE OS=Escherichia c
BAC0293|ruvB|tr|B5L348|B5L348_9PSED Malic enzyme family protein (Fragment) OS=Pseudomonas corrugata PE=3 S
BAC0335|rpoS|sp|P35540|RPOS_SHIFL RNA polymerase sigma factor RpoS OS=Shigella flexneri GN=rpoS PE=3 SV=3
BAC0564|actP/yjcG|sp|P32705|ACTP_ECOLI Cation/acetate symporter ActP OS=Escherichia coli (strain K12) GN=actP
BAC0364|smrA|tr|C7SLZ1|C7SLZ1_STEMA ABC-type multidrug efflux pump (Fragment) OS=Stenotrophomonas malto
BAC0242|mexK|tr|Q9HXW4|Q9HXW4_PSEAE Probable Resistance-Nodulation-Cell Division (RND) efflux transporter
BAC0047|bexA|tr|Q93HR0|Q93HR0_BACT4 BexA OS=Bacteroides thetaiotaomicron GN=bexA PE=4 SV=1
BAC0239|mexF|tr|Q4KBN7|Q4KBN7_PSEF5 Multidrug efflux RND transporter, permease protein MexF OS=Pseudomon
BAC0496|adeN|tr|B7H1T7|B7H1T7_ACIB3 Bacterial regulatory protein, tetR family protein OS=Acinetobacter baumanni
BAC0536|oxyRkp|tr|C4WZN6|C4WZN6_KLEPN Activator of hydrogen peroxide-inducible genes OS=Klebsiella pneumo
BAC0194|ibpA|sp|P0C054|IBPA_ECOLI Small heat shock protein IbpA OS=Escherichia coli (strain K12) GN=ibpA PE=1
BAC0143|emhB|tr|C1KA85|C1KA85_PSEFL EmhB OS=Pseudomonas fluorescens GN=emhB PE=4 SV=1
BAC0533|cpxR|tr|C4WZK6|C4WZK6_KLEPN Response regulator of stress-related two-component regulatory system OS
BAC0659|merB|sp|P08664|MERB_SERMA Alkylmercury lyase OS=Serratia marcescens GN=merB PE=3 SV=1
BAC0595|arsH|tr|P74312|P74312_SYNY3 Slr0945 protein OS=Synechocystis sp. (strain PCC 6803 / Kazusa) GN=slr0945
BAC0323|qacEdelta1|tr|Q7BQY4|Q7BQY4_PSEAI Disinfectant resistance protein OS=Pseudomonas aeruginosa GN=qac
BAC0211|mdtB/yegN|sp|P76398|MDTB_ECOLI Multidrug resistance protein MdtB OS=Escherichia coli (strain K12) GN
BAC0246|mexW|tr|Q9HW27|Q9HW27_PSEAE Probable Resistance-Nodulation-Cell Division (RND) efflux transporter O
BAC0532|cpxA|tr|C4WZK5|C4WZK5_KLEPN Sensor protein of stress-related two-component regulatory system OS=Kle
BAC0559|emrR|sp|P0ACR9|MPRA_ECOLI Transcriptional repressor MprA OS=Escherichia coli (strain K12) GN=mprA
BAC0006|acrB|sp|P31224|ACRB_ECOLI Multidrug efflux pump subunit AcrB OS=Escherichia coli (strain K12) GN=acr
BAC0417|vcaM|tr|Q9KKV4|Q9KKV4_VIBCH ABC transporter, ATP-binding protein OS=Vibrio cholerae serotype O1 (s
BAC0368|sodA|sp|P00448|SODM_ECOLI Superoxide dismutase [Mn] OS=Escherichia coli (strain K12) GN=sodA PE=1
BAC0371|soxS|sp|P0A9E2|SOXS_ECOLI Regulatory protein SoxS OS=Escherichia coli (strain K12) GN=soxS PE=1 SV=
BAC0196|iclR|sp|P16528|ICLR_ECOLI Acetate operon repressor OS=Escherichia coli (strain K12) GN=iclR PE=1 SV=1
BAC0015|adeG|tr|Q2FD81|Q2FD81_ACIBA Cation/multidrug efflux pump OS=Acinetobacter baumannii GN=29_167 PE
BAC0148|emrB|sp|P0AEJ0|EMRB_ECOLI Multidrug resistance protein B OS=Escherichia coli (strain K12) GN=emrB PE
BAC0156|fabI|sp|P0AEK4|FABI_ECOLI Enoyl-[acyl-carrier-protein] reductase [NADH] FabI OS=Escherichia coli (strain
BAC0295|oqxB|tr|Q69HW2|Q69HW2_ECOLX OqxB integral membrane protein OS=Escherichia coli GN=oqxB PE=4 S

BAC0541|yieF|sp|P0AGE6|YIEF_ECOLI Uncharacterized protein YieF OS=Escherichia coli (strain K12) GN=yieF PE=1 S
BAC0472|adeB|tr|Q93E19|Q93E19_ACIBA AdeB RND protein OS=Acinetobacter baumannii GN=adeB PE=4 SV=1
BAC0222|mepB|sp|P0C070|MEPB_PSEPU Multidrug/solvent efflux pump membrane transporter MepB OS=Pseudomonas
BAC0010|acrF/envD|sp|P24181|ACRF_ECOLI Acriflavine resistance protein F OS=Escherichia coli (strain K12) GN=acrF
BAC0370|soxR|sp|P0ACS2|SOXR_ECOLI Redox-sensitive transcriptional activator SoxR OS=Escherichia coli (strain K12
BAC0212|mdtC/yegO|sp|P76399|MDTC_ECOLI Multidrug resistance protein MdtC OS=Escherichia coli (strain K12) GN=
BAC0530|phoB|tr|C4X6T6|C4X6T6_KLEPN Response regulator in two-component regulatory system with PhoQ OS=Kleb
BAC0061|cepA|sp|Q8RR17|FIEF_KLEPN Cation-efflux pump FieF OS=Klebsiella pneumoniae GN=fieF PE=3 SV=1
BAC0008|acrD/yffA|sp|P24177|ACRD_ECOLI Probable aminoglycoside efflux pump OS=Escherichia coli (strain K12) GN
BAC0351|sitC|tr|Q9XCS0|Q9XCS0_SALTM SitC OS=Salmonella typhimurium GN=sitC PE=3 SV=1
BAC0181|glpF|sp|P0AER0|GLPF_ECOLI Glycerol uptake facilitator protein OS=Escherichia coli (strain K12) GN=glpF P
BAC0172|gadA|sp|P69908|DCEA_ECOLI Glutamate decarboxylase alpha OS=Escherichia coli (strain K12) GN=gadA PE
BAC0039|baeR|sp|P69228|BAER_ECOLI Transcriptional regulatory protein BaeR OS=Escherichia coli (strain K12) GN=b
BAC0215|mdtG/yceE|sp|P25744|MDTG_ECOLI Multidrug resistance protein MdtG OS=Escherichia coli (strain K12) GN=
BAC0244|mexT|tr|O87785|O87785_PSEAI MexT protein OS=Pseudomonas aeruginosa GN=mexT PE=4 SV=1
BAC0393|tolC|sp|P02930|TOLC_ECOLI Outer membrane protein TolC OS=Escherichia coli (strain K12) GN=tolC PE=1 S
BAC0494|eefA|tr|A8CY69|A8CY69_KLEPN EefA OS=Klebsiella pneumoniae GN=eefA PE=4 SV=1
BAC0707|sodB|sp|P0AGD3|SODF_ECOLI Superoxide dismutase [Fe] OS=Escherichia coli (strain K12) GN=sodB PE=1 S
BAC0029|chrF|tr|A4UQR2|A4UQR2_9RHIZ ChrF OS=Ochrobactrum tritici GN=chrF PE=4 SV=1
BAC0149|emrD|sp|P31442|EMRD_ECOLI Multidrug resistance protein D OS=Escherichia coli (strain K12) GN=emrD PE
BAC0012|actP|sp|Q9X5X3|ATCU_SINMW Copper-transporting P-type ATPase OS=Sinorhizobium medicae (strain WSM
BAC0135|dpsA|tr|Q8KR86|Q8KR86_BURPE DpsA OS=Burkholderia pseudomallei GN=dpsA PE=3 SV=1
BAC0294|oqxA|tr|Q69HW3|Q69HW3_ECOLX OqxA membrane-fusion protein OS=Escherichia coli GN=oqxA PE=4 SV
BAC0476|kpnE|tr|C4X7Z3|C4X7Z3_KLEPN Multidrug transport protein OS=Klebsiella pneumoniae subsp. pneumoniae N
BAC0217|mdtJ/ebrB/ydgF|sp|Q3Z1V3|MDTJ_SHISS Spermidine export protein MdtJ OS=Shigella sonnei (strain Ss046) G
BAC0296|ostA/lptD|sp|P31554|LPTD_ECOLI LPS-assembly protein LptD OS=Escherichia coli (strain K12) GN=lptD PE
BAC0235|mexB|sp|P52002|MEXB_PSEAE Multidrug resistance protein MexB OS=Pseudomonas aeruginosa (strain ATC
BAC0334|robA|sp|P0ACI0|ROB_ECOLI Right origin-binding protein OS=Escherichia coli (strain K12) GN=rob PE=1 SV
BAC0147|emrA|sp|P27303|EMRA_ECOLI Multidrug resistance protein A OS=Escherichia coli (strain K12) GN=emrA PE
BAC0142|emhA|tr|Q4KH22|Q4KH22_PSEF5 Efflux transporter, membrane fusion protein subunit EmhA OS=Pseudomona
BAC0040|baeS|sp|P30847|BAES_ECOLI Signal transduction histidine-protein kinase BaeS OS=Escherichia coli (strain K1
BAC0195|ibpB|sp|P0C058|IBPB_ECOLI Small heat shock protein IbpB OS=Escherichia coli (strain K12) GN=ibpB PE=1
BAC0531|phoR|tr|C4X6T5|C4X6T5_KLEPN Sensor kinase in two-component regulatory system with PhoP OS=Klebsiella
BAC0529|kpnO|tr|C4XBC3|C4XBC3_KLEPN Outer membrane porin protein C OS=Klebsiella pneumoniae subsp. pneumo
BAC0220|mdtN/yjcR|sp|P32716|MDTN_ECOLI Multidrug resistance protein MdtN OS=Escherichia coli (strain K12) GN=
BAC0213|mdtE/yhiU|sp|P37636|MDTE_ECOLI Multidrug resistance protein MdtE OS=Escherichia coli (strain K12) GN=
BAC0166|fetB/ybbM|sp|P77307|YBBM_ECOLI UPF0014 inner membrane protein YbbM OS=Escherichia coli (strain K12
BAC0359|smeE|tr|I0KSX8|I0KSX8_STEMA RND efflux system, inner membrane transporter OS=Stenotrophomonas malt
BAC0378|sugE|sp|P69937|SUGE_ECOLI Quaternary ammonium compound-resistance protein SugE OS=Escherichia coli
BAC0214|mdtF/yhiV|sp|P37637|MDTF_ECOLI Multidrug resistance protein MdtF OS=Escherichia coli (strain K12) GN=
BAC0218|mdtK/ydhE|sp|P37340|MDTK_ECOLI Multidrug resistance protein MdtK OS=Escherichia coli (strain K12) GN
BAC0009|acrE/envC|sp|P24180|ACRE_ECOLI Acriflavine resistance protein E OS=Escherichia coli (strain K12) GN=acrE
BAC0491|kdeA|tr|A6T6T9|A6T6T9_KLEP7 Multidrug/chloramphenicol efflux transport protein (MFS family) OS=Klebsi
BAC0011|acrR/ybaH|sp|P0ACS9|ACRR_ECOLI HTH-type transcriptional regulator AcrR OS=Escherichia coli (strain K12
BAC0041|bcr|sp|P28246|BCR_ECOLI Bicyclomycin resistance protein OS=Escherichia coli (strain K12) GN=bcr PE=1 SV
BAC0471|adeA|tr|Q93E20|Q93E20_ACIBA AdeA membrane fusion protein OS=Acinetobacter baumannii GN=adeA PE=

BAC0208|mdfA/cmr|sp|P0AEY8|MDFA_ECOLI Multidrug transporter MdfA OS=Escherichia coli (strain K12) GN=mdfA
BAC0176|gadW/yhiW|sp|P63201|GADW_ECOLI HTH-type transcriptional regulator GadW OS=Escherichia coli (strain K
BAC0477|kpnF|tr|C4X7Z4|C4X7Z4_KLEPN Spermidine export protein MdtI OS=Klebsiella pneumoniae subsp. pneumoni
BAC0493|kmrA|tr|C4X8X9|C4X8X9_KLEPN Energy-dependent efflux protein for methyl viologen resistance OS=Klebsie
BAC0005|acrA|sp|P0AE06|ACRA_ECOLI Multidrug efflux pump subunit AcrA OS=Escherichia coli (strain K12) GN=acr
BAC0175|gadE/yhiE|sp|P63204|GADE_ECOLI Transcriptional regulator GadE OS=Escherichia coli (strain K12) GN=gad
BAC0450|ymgB/ariR|sp|P75993|ARIR_ECOLI Probable two-component-system connector protein AriR OS=Escherichia c
BAC0210|mdtA/yegM|sp|P76397|MDTA_ECOLI Multidrug resistance protein MdtA OS=Escherichia coli (strain K12) GN
BAC0038|asr|sp|P36560|ASR_ECOLI Acid shock protein OS=Escherichia coli (strain K12) GN=asr PE=1 SV=3
BAC0174|gadC/xasA|sp|P63235|GADC_ECOLI Probable glutamate/gamma-aminobutyrate antiporter OS=Escherichia coli
BAC0434|ychH|sp|P0AB49|YCHH_ECOLI Uncharacterized protein YchH OS=Escherichia coli (strain K12) GN=ychH PE
BAC0353|smdA|tr|A7VN01|A7VN01_SERMA Multidrug efflux pump SmdA OS=Serratia marcescens GN=smdA PE=3 S
BAC0154|evgA|sp|P0ACZ4|EVGA_ECOLI Positive transcription regulator EvgA OS=Escherichia coli (strain K12) GN=ev
BAC0146|emmdR|tr|D5CJ69|D5CJ69_ENTCC MATE efflux family protein OS=Enterobacter cloacae subsp. cloacae (strai
BAC0445|ygiW|sp|P0ADU5|YGIW_ECOLI Protein YgiW OS=Escherichia coli (strain K12) GN=ygiW PE=1 SV=1
BAC0384|tehA|sp|P25396|TEHA_ECOLI Tellurite resistance protein TehA OS=Escherichia coli (strain K12) GN=tehA PE
BAC0153|emrY|sp|P52600|EMRY_ECOLI Multidrug resistance protein Y OS=Escherichia coli (strain K12) GN=emrY PE
BAC0144|emhC|tr|Q4KH24|Q4KH24_PSEF5 Efflux transporter, outer membrane factor lipoprotein EmhC OS=Pseudomon
BAC0498|ideR|sp|P0A672|IDER_MYCTU Iron-dependent repressor IdeR OS=Mycobacterium tuberculosis GN=ideR PE=
BAC0155|evgS|sp|P58402|EVGS_ECO57 Sensor protein EvgS OS=Escherichia coli O157:H7 GN=evgS PE=3 SV=1
BAC0186|hdeB/yhiC|sp|P0AET2|HDEB_ECOLI Acid stress chaperone HdeB OS=Escherichia coli (strain K12) GN=hdeB
BAC0252|mntP/yebN|sp|P76264|MNTP_ECOLI Probable manganese efflux pump MntP OS=Escherichia coli (strain K12)
BAC0185|hdeA/yhiB|sp|P0AES9|HDEA_ECOLI Acid stress chaperone HdeA OS=Escherichia coli (strain K12) GN=hdeA
BAC0219|mdtM/yjiO|sp|P39386|MDTM_ECOLI Multidrug resistance protein MdtM OS=Escherichia coli (strain K12) GN
BAC0560|marA|sp|P0ACH5|MARA_ECOLI Multiple antibiotic resistance protein MarA OS=Escherichia coli (strain K12)
BAC0177|gadX|sp|P37639|GADX_ECOLI HTH-type transcriptional regulator GadX OS=Escherichia coli (strain K12) GN
BAC0151|emrK|sp|P52599|EMRK_ECOLI Multidrug resistance protein K OS=Escherichia coli (strain K12) GN=emrK PE
BAC0650|merA|tr|O08449|O08449_9PSED Mercuric reductase OS=Pseudomonas sp. K-62 GN=merA PE=4 SV=1
BAC0511|vmeD|tr|Q87TN1|Q87TN1_VIBPA Putative multidrug resistance protein OS=Vibrio parahaemolyticus serotype
BAC0013|adeE|tr|Q8GKU1|Q8GKU1_ACIG3 AdeE OS=Acinetobacter sp. 4365 GN=adeE PE=4 SV=2
BAC0157|fabK|tr|Q9FBC5|Q9FBC5_STREE Trans-2-enoyl-ACP reductase II OS=Streptococcus pneumoniae GN=fabK P
BAC0705|sodA|sp|P53652|SODM_PSEAE Superoxide dismutase [Mn] OS=Pseudomonas aeruginosa (strain ATCC 15692
BAC0436|ydeI|sp|P31130|YDEI_ECOLI Uncharacterized protein YdeI OS=Escherichia coli (strain K12) GN=ydeI PE=4 S
BAC0451|yodD|sp|P64519|YODD_ECOLI Uncharacterized protein YodD OS=Escherichia coli (strain K12) GN=yodD PE
BAC0437|ydeO|sp|P76135|YDEO_ECOLI HTH-type transcriptional regulator YdeO OS=Escherichia coli (strain K12) GN
BAC0337|sdeB|tr|Q84GI9|Q84GI9_SERMA Putative resistance-nodulation cell division protein SdeB OS=Serratia marcesc
BAC0495|eefX|tr|A8CY68|A8CY68_KLEPN EefX OS=Klebsiella pneumoniae GN=eefX PE=4 SV=1
BAC0237|mexD|tr|Q51396|Q51396_PSEAI RND family exporter MexD OS=Pseudomonas aeruginosa GN=mexD PE=4 S
BAC0561|marR|sp|P27245|MARR_ECOLI Multiple antibiotic resistance protein MarR OS=Escherichia coli (strain K12) G
BAC0350|sitB|tr|Q9XCS1|Q9XCS1_SALTM SitB OS=Salmonella typhimurium GN=sitB PE=3 SV=1
BAC0145|emhR|tr|Q4KH21|Q4KH21_PSEF5 Transcriptional regulator EmhR OS=Pseudomonas fluorescens (strain Pf-5 /
BAC0424|vexB|tr|Q9KVI2|Q9KVI2_VIBCH Multidrug resistance protein, putative OS=Vibrio cholerae serotype O1 (strai
BAC0446|yhcN|sp|P64614|YHCN_ECOLI Uncharacterized protein YhcN OS=Escherichia coli (strain K12) GN=yhcN PE
BAC0179|gesB|tr|Q8ZRG9|Q8ZRG9_SALTY Putative cation efflux system protein OS=Salmonella typhimurium (strain L
BAC0165|fetA/ybbL|sp|P77279|YBBL_ECOLI Uncharacterized ABC transporter ATP-binding protein YbbL OS=Escheric
BAC0339|sdeY|tr|Q7WSD5|Q7WSD5_SERMA Multidrug efflux pump SdeY OS=Serratia marcescens GN=sdeY PE=4 S

BAC0385|tehB|sp|P25397|TEHB_ECOLI Tellurite methyltransferase OS=Escherichia coli (strain K12) GN=tehB PE=1 SV
BAC0141|emeA|tr|Q8GR72|Q8GR72_ENTFL Multidrug efflux pump OS=Enterococcus faecalis GN=emeA PE=4 SV=1
BAC0657|merB|tr|O07303|O07303_9PSED Alkylmercury lyase OS=Pseudomonas sp. K-62 GN=merB PE=3 SV=2
BAC0216|mdtI/ydgE|sp|P69210|MDTI_ECOLI Spermidine export protein MdtI OS=Escherichia coli (strain K12) GN=mdt
BAC0438|ydeP|sp|P77561|YDEP_ECOLI Protein YdeP OS=Escherichia coli (strain K12) GN=ydeP PE=2 SV=1
BAC0106|cuiD|sp|Q8ZRS2|CUEO_SALTY Blue copper oxidase CueO OS=Salmonella typhimurium (strain LT2 / SGSC1
BAC0358|oscA|tr|B6CM35|B6CM35_9PSED Putative uncharacterized protein oscA OS=Pseudomonas corrugata GN=osc
BAC0352|sitD|tr|Q9XCR9|Q9XCR9_SALTM SitD OS=Salmonella typhimurium GN=sitD PE=3 SV=1
BAC0492|kexD|tr|A6TA71|A6TA71_KLEP7 Acridine efflux pump OS=Klebsiella pneumoniae subsp. pneumoniae (strain
BAC0238|mexE|tr|Q1IB41|Q1IB41_PSEE4 Multidrug efflux RND membrane fusion protein MexE OS=Pseudomonas ento
BAC0681|merR2|tr|Q9WWL1|Q9WWL1_BACSR Mercury resistance operon negative regulator MerR2 OS=Bacillus sp. (
BAC0526|vmeV|tr|Q87HZ7|Q87HZ7_VIBPA Transporter, AcrB/D/F family OS=Vibrio parahaemolyticus serotype O3:K6
BAC0134|dpr/dps|sp|P0CB53|DPS_STRSU DNA protection during starvation protein OS=Streptococcus suis GN=dps PE=
BAC0044|bepE|sp|Q8G2M6|BEPE_BRUSU Efflux pump membrane transporter BepE OS=Brucella suis biovar 1 (strain 1
BAC0150|emrE/mvrC|sp|P23895|EMRE_ECOLI Multidrug transporter EmrE OS=Escherichia coli (strain K12) GN=emrE
BAC0027|chrB|tr|A4UQR5|A4UQR5_9RHIZ ChrB OS=Ochrobactrum tritici GN=chrB PE=4 SV=1
BAC0508|adeL|tr|A3M732|A3M732_ACIBT Transcriptional regulator LysR family OS=Acinetobacter baumannii (strain A
BAC0349|sitA|tr|Q9XCS2|Q9XCS2_SALTI Iron transport protein, periplasmic-binding protein OS=Salmonella typhi GN=
BAC0505|farR|tr|Q7DD70|Q7DD70_NEIMB Transcriptional regulator, MarR family OS=Neisseria meningitidis serogroup
BAC0290|opmD/nmpC|sp|P37592|OMPD_SALTY Outer membrane porin protein OmpD OS=Salmonella typhimurium (st
BAC0500|emrBsm|tr|B2FID0|B2FID0_STRMK Putative multidrug resistance protein B OS=Stenotrophomonas maltophilia
BAC0229|merG|tr|O07302|O07302_9PSED Mercuric resistance protein OS=Pseudomonas sp. K-62 GN=merG PE=4 SV=
BAC0404|ttgA|sp|Q9WWZ9|TTGA_PSEPT Toluene efflux pump periplasmic linker protein TtgA OS=Pseudomonas putid
BAC0207|mdeA|sp|P13254|MEGL_PSEPU Methionine gamma-lyase OS=Pseudomonas putida GN=mdeA PE=1 SV=2
BAC0435|yddg/emrE|sp|D0ZXP9|YDDG_SALT1 Methyl viologen resistance protein YddG OS=Salmonella typhimurium
BAC0354|smdB|tr|A7VN02|A7VN02_SERMA Multidrug efflux pump SmdB OS=Serratia marcescens GN=smdB PE=3 S
BAC0596|baeR|tr|D0ZNE3|D0ZNE3_SALT1 DNA-binding transcriptional regulator BaeR OS=Salmonella typhimurium (
BAC0509|vexH|tr|Q9KTI8|Q9KTI8_VIBCH Multidrug resistance protein, putative OS=Vibrio cholerae serotype O1 (strai
BAC0566|actS|tr|Q52912|Q52912_9RHIZ Histidine protein kinase OS=Sinorhizobium medicae GN=actS PE=4 SV=1
BAC0499|emrAsm|tr|B2FIC9|B2FIC9_STRMK Putative multidrug resistance protein A OS=Stenotrophomonas maltophili
BAC0014|adeF|tr|Q2FD82|Q2FD82_ACIBA Putative RND family drug transporter OS=Acinetobacter baumannii GN=29_
BAC0184|hasF|tr|Q6GW09|Q6GW09_SERMA TolC-like protein OS=Serratia marcescens PE=4 SV=1
BAC0513|vmeF|tr|Q87R57|Q87R57_VIBPA Putative multidrug resistance protein OS=Vibrio parahaemolyticus serotype
BAC0223|mepC|sp|P0C071|MEPC_PSEPU Multidrug/solvent efflux pump outer membrane protein MepC OS=Pseudomo
BAC0338|sdeX|tr|Q7WSD6|Q7WSD6_SERMA Multidrug efflux pump SdeX OS=Serratia marcescens GN=sdeX PE=4 S
BAC0028|chrC|tr|A4UQR3|A4UQR3_9RHIZ Superoxide dismutase OS=Ochrobactrum tritici GN=chrC PE=3 SV=1
BAC0413|ttgR|sp|Q9AIU0|TTGR_PSEPT HTH-type transcriptional regulator TtgR OS=Pseudomonas putida (strain DOT-
BAC0258|mtrD|tr|Q5F725|Q5F725_NEIG1 Antibiotic resistance efflux pump component OS=Neisseria gonorrhoeae (strai
BAC0326|qacH/qacI|sp|O87868|QACH_STASA Quaternary ammonium compound-resistance protein QacH OS=Staphyloc
BAC0321|qacC/qacD/smr|sp|P14319|QACC_STAAU Quaternary ammonium compound-resistance protein QacC OS=Stap
BAC0506|pcm|tr|R4IUI7|R4IUI7_STEMA Protein-L-isoaspartate O-methyltransferase OS=Stenotrophomonas maltophilia
BAC0656|merB3|tr|Q7DHE7|Q7DHE7_BACCE Organomercurial lyase enzyme OS=Bacillus cereus GN=merB3 PE=4 SV
BAC0002|abeS|tr|Q2FD83|Q2FD83_ACIBA QacEdelta1 SMR family efflux pump OS=Acinetobacter baumannii GN=qac
BAC0654|merB1|sp|P16172|MERB_BACCE Alkylmercury lyase OS=Bacillus cereus GN=merB1 PE=3 SV=2
BAC0240|mexI|tr|Q9HWH4|Q9HWH4_PSEAE Probable Resistance-Nodulation-Cell Division (RND) efflux transporter O
BAC0479|adeT2|tr|C7F8K7|C7F8K7_ACIBA AdeT2 OS=Acinetobacter baumannii PE=4 SV=1

BAC0313|pmpM|sp|Q9I3Y3|PMPM_PSEAE Multidrug resistance protein PmpM OS=Pseudomonas aeruginosa (strain AT
BAC0016|adeH|tr|Q2FD80|Q2FD80_ACIBA Putative RND family drug transporter OS=Acinetobacter baumannii GN=29_
BAC0367|smvA/emrB|sp|D0ZXQ3|SMVA_SALT1 Methyl viologen resistance protein SmvA OS=Salmonella typhimuriu
BAC0447|yjaA|sp|P09162|YJAA_ECOLI Uncharacterized protein YjaA OS=Escherichia coli (strain K12) GN=yjaA PE=4
BAC0358|smeD|tr|I0KSX9|I0KSX9_STEMA Membrane fusion protein of RND family multidrug efflux pump OS=Stenotr
BAC0507|tolCsm|tr|R4ITT0|R4ITT0_STEMA Outer membrane protein OS=Stenotrophomonas maltophilia GN=tolCsm P
BAC0478|adeT1|tr|C7F8K6|C7F8K6_ACIBA AdeT1 OS=Acinetobacter baumannii PE=4 SV=1
BAC0473|adeC|tr|Q93E18|Q93E18_ACIBA AdeC outer membrane protein OS=Acinetobacter baumannii GN=adeC PE=4
BAC0597|baeS|tr|D0ZNE2|D0ZNE2_SALT1 Signal transduction histidine-protein kinase BaeS OS=Salmonella typhimuri
BAC0419|vceB|tr|O51919|O51919_VIBCL VceB OS=Vibrio cholerae GN=vceB PE=4 SV=1
BAC0260|mtrF|tr|B4RN92|B4RN92_NEIG2 Antibiotic resistance efflux pump component OS=Neisseria gonorrhoeae (stra
BAC0538|chrR|tr|Q7BD45|Q7BD45_PSEPU Chromate reductase OS=Pseudomonas putida GN=chrR PE=4 SV=1
BAC0236|mexC|tr|Q51395|Q51395_PSEAI Membrane fusion protein MexC OS=Pseudomonas aeruginosa GN=mexC PE=
BAC0043|bepD|sp|Q8G2M7|BEPD_BRUSU Efflux pump periplasmic linker BepD OS=Brucella suis biovar 1 (strain 1330
BAC0159|fabV|sp|Q9KRA3|Y1738_VIBCH Putative reductase VC_1738/VC_1739 OS=Vibrio cholerae serotype O1 (strai
BAC0661|merB2|tr|Q7DJN2|Q7DJN2_BACME MerB2 OS=Bacillus megaterium GN=merB2 PE=4 SV=1
BAC0248|mexY|tr|Q9ZNG8|Q9ZNG8_PSEAI MexY OS=Pseudomonas aeruginosa GN=mexY PE=4 SV=1
BAC0173|gadB|sp|P69910|DCEB_ECOLI Glutamate decarboxylase beta OS=Escherichia coli (strain K12) GN=gadB PE=
BAC0360|smeF|tr|Q9F239|Q9F239_STEMA Outer membrane protein OS=Stenotrophomonas maltophilia GN=smeF PE=4
BAC0291|oprJ|sp|Q51397|OPRJ_PSEAE Outer membrane protein OprJ OS=Pseudomonas aeruginosa (strain ATCC 15692
BAC0501|emrCsm|tr|B2FIC8|B2FIC8_STRMK Putative outer membrane multidrug efflux protein OS=Stenotrophomonas
BAC0170|frnE|tr|Q9RWK7|Q9RWK7_DEIRA FrnE protein OS=Deinococcus radiodurans (strain ATCC 13939 / DSM 20
BAC0521|vmeK|tr|Q87LY6|Q87LY6_VIBPA Putative multidrug resistance protein OS=Vibrio parahaemolyticus serotype
BAC0380|tbtB|tr|Q71UZ6|Q71UZ6_PSEST Resistance nodulation cell division family member TbtB OS=Pseudomonas st
BAC0411|ttgH|sp|Q93PU4|TTGH_PSEPT Toluene efflux pump membrane transporter TtgH OS=Pseudomonas putida (stra
BAC0046|bepG|sp|Q8FWV9|BEPG_BRUSU Efflux pump membrane transporter BepG OS=Brucella suis biovar 1 (strain 1
BAC0241|mexJ|tr|Q9HXW3|Q9HXW3_PSEAE Probable Resistance-Nodulation-Cell Division (RND) efflux membrane fu
BAC0474|adeD|tr|Q67GM1|Q67GM1_ACIG3 AdeD OS=Acinetobacter sp. 4365 GN=adeD PE=4 SV=1
BAC0234|mexA|sp|P52477|MEXA_PSEAE Multidrug resistance protein MexA OS=Pseudomonas aeruginosa (strain ATC
BAC0503|farA|tr|Q9RQ30|Q9RQ30_NEIGO Efflux pump protein FarA OS=Neisseria gonorrhoeae PE=4 SV=1
BAC0418|vceA|tr|O51918|O51918_VIBCL VceA OS=Vibrio cholerae GN=vceA PE=4 SV=1
BAC0042|bepC|sp|Q8G0Y6|BEPC_BRUSU Outer membrane efflux protein BepC OS=Brucella suis biovar 1 (strain 1330)
BAC0412|ttgI|sp|Q93PU3|TTGI_PSEPT Toluene efflux pump outer membrane protein TtgI OS=Pseudomonas putida (strai
BAC0292|oprM/oprK|sp|Q51487|OPRM_PSEAE Outer membrane protein OprM OS=Pseudomonas aeruginosa (strain AT
BAC0025|amvA|tr|C4PAW9|C4PAW9_ACIBA Major facilitator superfamily efflux pump OS=Acinetobacter baumannii P
BAC0319|qacA|sp|P0A0J9|QACA_STAAU Antiseptic resistance protein OS=Staphylococcus aureus GN=qacA PE=1 SV=
BAC0408|ttgE|sp|Q9KWV4|TTGE_PSEPT Toluene efflux pump membrane transporter TtgE OS=Pseudomonas putida (str
BAC0430|vmeB|tr|Q2AAU3|Q2AAU3_VIBPH Inner membrane protein VmeB OS=Vibrio parahaemolyticus GN=vmeB P
BAC0427|vexF|tr|A6P7H3|A6P7H3_VIBCL Multidrug efflux transporter VexF OS=Vibrio cholerae GN=vexF PE=4 SV=1
BAC0515|vmeZ|tr|Q87GX5|Q87GX5_VIBPA Putative acriflavin resistance protein OS=Vibrio parahaemolyticus serotype

[illegible]ruary); A (April); STP (sewage treatment plant); inf (influent); T (treated).

| Relativ abundance |
| ---: |
| 0.0464085353258943 |
| 0.0367843207742229 |
| 0.0323810361849341 |
| 0.0280635646936096 |
| 0.0192187527213741 |
| 0.0177579479599696 |
| 0.00985448850130247 |
| 0.00900501196761978 |
| 0.00845150361509899 |
| 0.00814151766935487 |
| 0.00773473870483933 |
| 0.00772552771544623 |
| 0.00710590871262758 |
| 0.00653996110859636 |
| 0.00642231369548624 |
| 0.00614854198987738 |
| 0.00549291496602308 |
| 0.00547393706935791 |
| 0.00540623727855957 |
| 0.00540506311937726 |
| 0.00521295141523675 |
| 0.00517511311707776 |
| 0.00508844854334679 |
| 0.00502727593790765 |
| 0.0048508237321438 |
| 0.0047139739752217 |
| 0.00469482734801054 |
| 0.00450580176357426 |
| 0.00449214597967334 |
| 0.00443260406442654 |
| 0.00438171957899307 |
| 0.00428535364306287 |
| 0.00416718967001666 |
| 0.00406350258007017 |
| 0.00404188820464426 |
| 0.00391851907338055 |
| 0.00391140528598899 |
| 0.00387218523298585 |
| 0.00364689347934253 |
| 0.00363254683547703 |
| 0.00348741021662397 |
| 0.00343567881187609 |
| 0.00336558801292229 |

0.00323970426289207
0.00306134302607856
0.00295548244161512
0.0029528820562427
0.00294157398310024
0.00287393998966589
0.00283904854349821
0.00277619220308052
0.00277117385459262
0.00274125174052352
0.00272496151501631
0.00264922400352023
0.00264844550689857
0.00264399777252333
0.00263941633625981
0.00254636048012988
0.00252880473063295
0.00243933295462454
0.0023598601940159
0.00235217527855443
0.00234490716283262
0.00233516398268318
0.00224413628065551
0.00217421482161021
0.00213589198115791
0.00213330145116257
0.00211647811132403
0.00206089526345771
0.00205405262300237
0.00201171221481152
0.00200566584920238
0.00191887621836646
0.00191202308901516
0.00190317418751871
0.00188558959008868
0.00185233775729802
0.00180922614874552
0.00176623833736004
0.00174461092914747
0.00173964052763993
0.00170584179738864
0.00158422086628732
0.00155712086572361
0.00154254830601136
0.00153784222643369
0.00152098042101717

0.00148169382871401
0.00143965861226397
0.00140656301198204
0.00138041605504934
0.00136634266441719
0.00133582759687169
0.00133526663903426
0.00133237584597538
0.00132511680816322
0.00131066098843781
0.00129705551104918
0.00125829638164632
0.00123356328323558
0.00117599699668459
0.00116804435427252
0.00115210155698418
0.00112039233981947
0.00111354655094109
0.00110902083637045
0.00109597353241315
0.00106844063902294
0.00104889089347526
0.00104852879075024
0.00104584713143886
0.000999968743593027
0.000992213945211233
0.000978761687026269
0.000879484933417964
0.000842060042634221
0.000772763099247003
0.000765375306711181
0.000706728964353721
0.000693363889383135
0.000639082024473622
0.000619746937971725
0.000592828956506423
0.000557374911263672
0.000546626383803447
0.000532125337866331
0.000510809506281918
0.000484028156481194
0.000481622701633647
0.000452306537186382
0.000450590003838426
0.000449807606041705
0.00042925400717161

| |
|---|
| 0.000420750267150122 |
| 0.000404586122712589 |
| 0.000394152839548132 |
| 0.000393645621100776 |
| 0.000379401865074103 |
| 0.000367638318402724 |
| 0.000353225635621978 |
| 0.000317505820438716 |
| 0.000281852738306054 |
| 0.000280098084954869 |
| 0.000267813081228778 |
| 0.00025699235067408 |
| 0.000230768641421623 |
| 0.00022920939384445 |
| 0.00022723534164866 |
| 0.000213558776637638 |
| 0.000210375133575061 |
| 0.000205593880539264 |
| 0.000195132245025098 |
| 0.000182608229298849 |
| 0.000172636082895565 |
| 0.000172636082895565 |
| 0.00017260229892631 |
| 0.000161218171670393 |
| 0.000160014105136691 |
| 0.000156688176854404 |
| 0.000155372474606009 |
| 0.000142018227660805 |
| 0.000138461184852974 |
| 0.000126225080145036 |
| 0.000113076634296595 |
| 0.00010253800591127 |
| 0.000102280372730588 |
| 0.000101515182909661 |
| 0.0000969228293970818 |
| 0.0000883411205442152 |
| 0.0000881116630882562 |
| 0.0000871682833978038 |
| 0.0000869820263819965 |
| 0.0000860989601243112 |
| 0.0000841065874933355 |
| 0.00008376046984933 |
| 0.0000729526672881261 |
| 0.0000648623141280662 |
| 0.0000619597996145728 |
| 0.0000589965048503976 |

| |
|---|
| 0.0000583621338305009 |
| 0.000058236893199633 |
| 0.000058236893199633 |
| 0.0000560710583288903 |
| 0.0000559554478993462 |
| 0.0000546044109279334 |
| 0.0000515285928440182 |
| 0.0000464698497109296 |
| 0.0000461537282843246 |
| 0.0000426704280364511 |
| 0.0000381514417495542 |
| 0.0000335871190979986 |
| 0.0000323384082831064 |
| 0.0000318525730412945 |
| 0.0000315856520381551 |
| 0.0000312654288377683 |
| 0.0000299540753103564 |
| 0.0000287482968550666 |
| 0.0000285067145285534 |
| 0.0000276922369705948 |
| 0.0000257724522613323 |
| 0.0000184866432092526 |
| 0.0000184364077657492 |
| 0.0000180441437707333 |
| 0.0000177143552422865 |
| 0.0000172197920248622 |
| 0.0000168771095965067 |
| 0.0000148785045127099 |
| 0.0000144353150165866 |
| 0.0000141640878033313 |
| 0.0000133818502126148 |
| 0.0000127290770315116 |
| 6.52365197864974E-06 |
| 6.48623141280662E-06 |
| 6.46152195980545E-06 |
| 0.0272050507455579 |
| 0.0267527540849148 |
| 0.0189737789815173 |
| 0.0178576743355457 |
| 0.0172488899831975 |
| 0.0138360080079124 |
| 0.0115492567713583 |
| 0.00909517968712438 |
| 0.00749882809012172 |
| 0.00734364506190013 |
| 0.00673044922873786 |

0.00651365990108019
0.00651061043483437
0.00529538700643462
0.00507320293623457
0.00489947576829065
0.0048220055092948
0.00481079588780864
0.00479135832866598
0.00452819599222766
0.0045024676059081
0.0044937254892511
0.00444853097326761
0.00443418550092556
0.00439924458996107
0.00427324700724645
0.0042363096051477
0.00404767564740775
0.00377121296392749
0.0035512420553642
0.00351261360661114
0.00348488370472147
0.00346854256040747
0.00338393429895646
0.00336676194859203
0.00330115458667658
0.00326664774430714
0.00323357420795512
0.0030674042825642
0.00305068603232239
0.00301232395798565
0.00298002751821134
0.00283318871669715
0.0028115613219132
0.00279026161492901
0.00277952983948698
0.00275248244412726
0.00272299397715348
0.00266705525790617
0.00265687349870314
0.00265074853418256
0.00254858541206114
0.00251977707062467
0.00251920762947876
0.00250554104197707
0.00244224211372342
0.00242631444776436

| |
|---|
| 0.00235257351846956 |
| 0.00234668156332491 |
| 0.00234199007679287 |
| 0.00232917022436 |
| 0.00231450272206093 |
| 0.00226390394720407 |
| 0.00223220929194321 |
| 0.00219561569699332 |
| 0.00215611124789969 |
| 0.00211625036768642 |
| 0.00209269621119676 |
| 0.00208185322564651 |
| 0.00207925276658521 |
| 0.00205260624546502 |
| 0.00203379068821492 |
| 0.00199104095135638 |
| 0.00191654333146639 |
| 0.0018783224529766 |
| 0.0018098994258999 |
| 0.00176450829744082 |
| 0.00175871035122798 |
| 0.00174811571055793 |
| 0.00168180152132707 |
| 0.00152196088087037 |
| 0.00150356066296693 |
| 0.00147083557996258 |
| 0.00146227614044408 |
| 0.00145457247094043 |
| 0.00142724379280256 |
| 0.00140171782732977 |
| 0.00137333188859787 |
| 0.00136777530143579 |
| 0.00136755905237627 |
| 0.00136268590496533 |
| 0.00133388116225874 |
| 0.00133388116225874 |
| 0.00133113398143402 |
| 0.00131306428937836 |
| 0.00129779609996698 |
| 0.00122312837914696 |
| 0.00119582640639814 |
| 0.00118929183587138 |
| 0.0010764461052518 |
| 0.00104169766957349 |
| 0.00103025044243532 |
| 0.00101464058724691 |

| |
|---|
| 0.00100028246572926 |
| 0.000975615949275879 |
| 0.00094987629444392 |
| 0.000941115907011631 |
| 0.000922400533860831 |
| 0.000864778418186232 |
| 0.000837078484478704 |
| 0.000792296507942806 |
| 0.000790940300294839 |
| 0.000734567890767167 |
| 0.000667138479821477 |
| 0.000629257812268858 |
| 0.000622091114148108 |
| 0.000614369529892627 |
| 0.000588549909189245 |
| 0.000585435601101857 |
| 0.000581537490887009 |
| 0.000562216892560324 |
| 0.000550639863287699 |
| 0.000539832960566677 |
| 0.000522314982104938 |
| 0.000517780505863116 |
| 0.000513995560644818 |
| 0.000513995560644818 |
| 0.000507320293623457 |
| 0.000490595448778728 |
| 0.000477245912757538 |
| 0.000462355489178989 |
| 0.000440002793123421 |
| 0.000435978377332658 |
| 0.000431642680762499 |
| 0.000409579686595084 |
| 0.00037345757752396 |
| 0.000346078959991195 |
| 0.000344476742583829 |
| 0.000344476742583829 |
| 0.00034283077862609 |
| 0.000325079023098526 |
| 0.000311221487819005 |
| 0.000287314067279819 |
| 0.000255987304121928 |
| 0.000247783849974391 |
| 0.000243321392012767 |
| 0.000223008540108503 |
| 0.000219922097728395 |
| 0.000213608544683561 |

| |
|---|
| 0.000212591361137448 |
| 0.000158312715740653 |
| 0.000138646539872249 |
| 0.000135285411632921 |
| 0.00011595892425679 |
| 0.000109207890995264 |
| 0.000106295680568724 |
| 0.000104308845417907 |
| 0.000104308845417907 |
| 0.000102394921648771 |
| 0.0000987703226523545 |
| 0.000095940227447416 |
| 0.0000958029739031421 |
| 0.0000955978283487457 |
| 0.0000851988279367638 |
| 0.0000850365444549795 |
| 0.0000764455236966853 |
| 0.0000759254861205174 |
| 0.000074406976398107 |
| 0.0000600056261275057 |
| 0.000059367268402745 |
| 0.0000588455876610688 |
| 0.0000567992186245092 |
| 0.0000556660671307534 |
| 0.0000526464455646984 |
| 0.0000519118439986793 |
| 0.0000514333938235763 |
| 0.0000485262889552872 |
| 0.0000467968405019541 |
| 0.0000465043602488169 |
| 0.0000430928434738072 |
| 0.0000424778171635245 |
| 0.0000422766911352881 |
| 0.0000357725848067822 |
| 0.0000320106495020537 |
| 0.0000319191033166331 |
| 0.0000313806367189767 |
| 0.000028765583659062 |
| 0.0000283275290855737 |
| 0.0000282558138220659 |
| 0.0000281134671529371 |
| 0.0000247473313962662 |
| 0.0000240022504510022 |
| 0.0000230124669272496 |
| 0.0000226850537799106 |
| 0.0000224568339229699 |

| |
|---|
| 0.0000210189198864709 |
| 0.0000121978649832962 |
| 0.0000105992843871947 |
| 0.0174169741179377 |
| 0.0166452177135879 |
| 0.0138984821172863 |
| 0.00975971500660258 |
| 0.00608932060751207 |
| 0.00568454845147645 |
| 0.00531496797249099 |
| 0.00457539562814247 |
| 0.00412437293164879 |
| 0.00365931223157563 |
| 0.00292901895860166 |
| 0.00281289784059212 |
| 0.00276006926424486 |
| 0.00266274450781969 |
| 0.00261270451578189 |
| 0.00247768949496634 |
| 0.00242343130808122 |
| 0.00236129204377145 |
| 0.00215121490451728 |
| 0.00198642785650062 |
| 0.00195669247766757 |
| 0.00193776914135242 |
| 0.00191369279318972 |
| 0.00189103188527078 |
| 0.00184353855324729 |
| 0.00179296435971919 |
| 0.00176397500307943 |
| 0.00172295232858921 |
| 0.0017109873818629 |
| 0.00170628686707756 |
| 0.0017008631961714 |
| 0.00164804135778098 |
| 0.00163549281952377 |
| 0.00160405067049647 |
| 0.00157038222991591 |
| 0.00149497899249089 |
| 0.00143196482420525 |
| 0.00142374694860006 |
| 0.00138611635999019 |
| 0.00137803647055246 |
| 0.00135133515967781 |
| 0.0013441472067008 |
| 0.00131159000792258 |

| |
|---|
| 0.00127933779461301 |
| 0.00125977490168294 |
| 0.00117307392584797 |
| 0.00105291531191563 |
| 0.00104387741224253 |
| 0.0010199324226267 |
| 0.00101843697676474 |
| 0.001010798699439 |
| 0.00098670765538544 |
| 0.000953844505761644 |
| 0.000937121556385268 |
| 0.000926890699292525 |
| 0.000920917182433497 |
| 0.000910894176820035 |
| 0.000908515497710059 |
| 0.000900026239226512 |
| 0.000893693972064975 |
| 0.000889787587534336 |
| 0.000886382788092241 |
| 0.000865780470611568 |
| 0.000861476164294607 |
| 0.000851061081133878 |
| 0.000842332249532505 |
| 0.000832059905026011 |
| 0.000828523524130333 |
| 0.00082658772150386 |
| 0.00080263216400126 |
| 0.000795904487747249 |
| 0.000771226280416125 |
| 0.000759618261101457 |
| 0.000755352288982953 |
| 0.000733850065880592 |
| 0.000705351034195908 |
| 0.000701943541277087 |
| 0.000689180931435686 |
| 0.000686683899075412 |
| 0.000680345278468562 |
| 0.000673593640579305 |
| 0.000668639650632554 |
| 0.000640584840116503 |
| 0.000623544652251335 |
| 0.00062139264309775 |
| 0.000616340670389638 |
| 0.000615340117353291 |
| 0.00059792705093488 |
| 0.000584952951064239 |

| |
|---|
| 0.000579586410228788 |
| 0.000577599256822289 |
| 0.00056155483302167 |
| 0.000545164273174591 |
| 0.000542909457706497 |
| 0.000541499303270896 |
| 0.000534474777622148 |
| 0.000532957133191863 |
| 0.000521627769205909 |
| 0.000517221556730485 |
| 0.000508303943683408 |
| 0.000502527762505187 |
| 0.000498749358275825 |
| 0.000496001427927198 |
| 0.000470451522345282 |
| 0.000443044884494369 |
| 0.000433553363729966 |
| 0.000425215799042851 |
| 0.000424430978446611 |
| 0.000421166124766252 |
| 0.000420341924913481 |
| 0.000407580120741534 |
| 0.000404869491159819 |
| 0.00040202221000415 |
| 0.000396750697243571 |
| 0.000379049512289627 |
| 0.000374554854041133 |
| 0.000358439255120215 |
| 0.000350971770638543 |
| 0.000347114937994164 |
| 0.000340172639234281 |
| 0.000315874593574689 |
| 0.000315874593574689 |
| 0.000313264059743493 |
| 0.000310996221473149 |
| 0.000310396999466457 |
| 0.000301859679866597 |
| 0.000297994899598763 |
| 0.000274673559630164 |
| 0.000272978043829978 |
| 0.000266467143964588 |
| 0.0002485570572391 |
| 0.000236905945181017 |
| 0.000213428779442357 |
| 0.000211186452246688 |
| 0.000204371763682958 |

| |
|---|
| 0.000200368212342153 |
| 0.000198976121936812 |
| 0.000185808584455699 |
| 0.000185632462574699 |
| 0.000143579360715767 |
| 0.000122372723903027 |
| 0.00011747402240381 |
| 0.000116451463069009 |
| 0.000114863488572614 |
| 0.000112525300510067 |
| 0.000110188811712101 |
| 0.000109026705356556 |
| 0.000105291531191563 |
| 0.0000881510493696808 |
| 0.0000867957666348509 |
| 0.0000865409845410108 |
| 0.0000865409845410108 |
| 0.0000822590087434087 |
| 0.0000796322504810141 |
| 0.0000747000728048252 |
| 0.000072950252557665 |
| 0.0000679300201235891 |
| 0.0000622412992265398 |
| 0.0000607451141489787 |
| 0.0000595989799197527 |
| 0.0000574317442863071 |
| 0.0000562388000429714 |
| 0.0000542273980385733 |
| 0.000054111279413223 |
| 0.0000523549050123794 |
| 0.0000500395395761884 |
| 0.0000489728052053782 |
| 0.000048783721015396 |
| 0.0000482251287900289 |
| 0.0000481027299859933 |
| 0.0000480874738077548 |
| 0.000047619285463521 |
| 0.0000415624465229854 |
| 0.0000330758736727423 |
| 0.0000326485368035854 |
| 0.000031906524603504 |
| 0.000031508687638373 |
| 0.0000312747122351177 |
| 0.0000306972394144499 |
| 0.0000304604236812622 |
| 0.000030198335905802 |

0.0000296039919001583
0.0000279535038561672
0.0000264884355198901
0.0000168916894959727
0.0000165379368363711
0.0000160750429300096
0.0000159936503075792
0.0000158333129611373
0.0000140077425088554
0.0000135860040247178
0.0000131889183120955
0.0000127112512504905
0.0000121140783729507
0.0000119990348936254
0.000011764416892912
6.03966718116041E-06
6.03966718116041E-06
6.01665892523218E-06
0.020453502188662
0.0180052889999286
0.0164508217956181
0.013297896236875
0.00806613792009467
0.0077294597842543
0.00624880718735313
0.00522688878853262
0.00506981130162335
0.00407322947796173
0.00405932402018507
0.00390482088213954
0.00323462340896961
0.00289275264216794
0.00279044369170875
0.00240953888252815
0.00230048647620683
0.00221190410953304
0.00190318226348321
0.00184709369528592
0.00180357144660714
0.00178311857453221
0.00167059424306504
0.00160350164249205
0.00157345704964441
0.00140650825582332
0.00139577155158039
0.00138987724604162

0.00133656839160666
0.00131780953698761
0.00130098092796911
0.00128916150357484
0.00128358071784508
0.001262500012625
0.00117928978452017
0.00109817461415634
0.00109311390164871
0.00108719286801478
0.00105552705632503
0.00104257083622438
0.00102953870077157
0.001022105933566
0.000994092300628958
0.000962623649246018
0.000921783873022428
0.00091011886439775
0.000890794000324249
0.000874472377939315
0.00085944100238323
0.000841155023609118
0.00078360797276118
0.000766373193301623
0.000762446946399979
0.000738958618871562
0.000723796904784866
0.000718118007727478
0.000705969394814796
0.000691158848070429
0.000686889140900882
0.000683984188747899
0.000661577810150985
0.000658904768493809
0.000652215379981182
0.000648812135278368
0.000634724776989449
0.000617723220462946
0.000608219786301978
0.000605116624126968
0.000581386560435714
0.000581386560435714
0.000578550528433589
0.000575742030722746
0.00057259986178933
0.000571427047623111

0.000567090169605328
0.000564775515851836
0.000564775515851836
0.000539103901494935
0.000539103901494935
0.000532192313014231
0.000513432287138033
0.000494178576370357
0.000466093457761642
0.000452500865110206
0.000447953579352107
0.000433912896325191
0.000432857147185714
0.000415941887926954
0.000411815480308631
0.000408412046587072
0.000406174172359197
0.000406174172359197
0.000402238376115407
0.000368005322828989
0.0003677608010198
0.000367570841928364
0.000366058204718783
0.00036368120338944
0.00036330588739007
0.000362699872565399
0.000361594080270993
0.000359402600996623
0.000352984697407398
0.000349419195413383
0.000339044443493484
0.000332620195633894
0.000329452384246904
0.000329452384246904
0.00032816546087094
0.000324051525488758
0.000322290375893711
0.000320956477489821
0.000307488891963777
0.000304109893150989
0.000301788443584951
0.000301022483068237
0.000291646372939883
0.000286480334127743
0.000282119073094399
0.000278410465560764

| |
|---|
| 0.000274543653539087 |
| 0.000274543653539087 |
| 0.0002675755405051 |
| 0.000263561907397523 |
| 0.000255389445152639 |
| 0.000247089288185178 |
| 0.000244038803145855 |
| 0.000243437722349929 |
| 0.000234393000649971 |
| 0.000233470193560798 |
| 0.000233470193560798 |
| 0.000225910206340734 |
| 0.000215380493333327 |
| 0.0002011922957233 |
| 0.000182185650274786 |
| 0.000179701300498311 |
| 0.000177997970635768 |
| 0.000174587131985602 |
| 0.000172531173795137 |
| 0.000163816102664206 |
| 0.000162182158268263 |
| 0.000159867474753493 |
| 0.000153233667091583 |
| 0.000147515992946375 |
| 0.000146640527112865 |
| 0.000139861861236893 |
| 0.000136012452212024 |
| 0.000135391390786399 |
| 0.000133561777397393 |
| 0.000133304432932273 |
| 0.000130046993781672 |
| 0.000124321654432794 |
| 0.000113713191111108 |
| 0.000113604270429967 |
| 0.000112313312811444 |
| 0.000108610676125353 |
| 0.000106274962660291 |
| 0.000102953870077157 |
| 0.000102103011646768 |
| 0.000096897760072619 |
| 0.000095034341609684 |
| 0.0000906749681413499 |
| 0.0000874652347558154 |
| 0.0000868503649157042 |
| 0.0000864452320181383 |
| 0.0000795458473030756 |

0.0000763210156556536
0.0000748755418742965
0.0000744992828699031
0.000067235180458552
0.000067235180458552
0.0000667808886986969
0.0000648406754375351
0.0000637649775961751
0.0000605116624126968
0.0000587142863014285
0.0000517464477874719
0.0000510778890305278
0.0000502980739308251
0.0000501704138447063
0.0000486875444699859
0.000047654636101288
0.0000472898159206083
0.0000470646263209864
0.0000466206204122978
0.000045970100127475
0.0000456516005884856
0.0000453374840706749
0.0000451304635954664
0.000043636077383696
0.0000423279294535637
0.0000399336223329581
0.0000377956846172357
0.0000370865723354864
0.000028428297777777
0.0000283467634629268
0.0000283467634629268
0.000026426661837987
0.0000216744989636121
0.0000212093809601011
0.0000212093809601011
0.0000209397701851846
0.0000204628810091245
0.0000204206023293536
0.0000194942239199352
0.0000193416272551998
0.000018405161131112
9.72792473169994E-06
9.42189850086477E-06
0.018216226609859
0.0173359864930053
0.0133404159547744

0.00813502427662508
0.00513143914996448
0.00449899897272856
0.00430366178497528
0.00387094815459504
0.00376419900054861
0.00322279178453205
0.00301073745357392
0.00287428165770316
0.00261422760295859
0.00252509482681379
0.00213902660949102
0.00188762348203611
0.00168587674153743
0.00165562966118879
0.00131539491931148
0.00131395732923573
0.00129555059940807
0.00127978964321089
0.00125157344433645
0.00123000190836659
0.00117535928166363
0.00113997379665864
0.00106332926555191
0.0010436379828565
0.00104165010098439
0.00103468398681674
0.00100126174748084
0.000944163603230442
0.000932181499250472
0.000924137450468357
0.000880569548035177
0.000869356590827424
0.000867439362374239
0.000810250585846402
0.000802227061555179
0.000800122453523322
0.000780290080640378
0.000761779549530297
0.000757097579502957
0.00074167166294371
0.000739492056424039
0.000729816771228326
0.000724946579311056
0.000716557612144486
0.000671766977470855

0.000664461553098952
0.000637890914086375
0.000636122199074442
0.000635257902608308
0.000632507868397882
0.000621741777020897
0.000607207553661967
0.00060421146375903
0.000592735568356636
0.000584437270399643
0.000574699309871269
0.000572977716078082
0.000566350590259111
0.000553767092944269
0.000529455366912569
0.000525927802384381
0.000517506437780086
0.000502958064027232
0.000498197555602283
0.000479046942950527
0.000477091649305831
0.000470372048611383
0.00046879994951843
0.000468364363038042
0.000464399973024419
0.000451302911505516
0.000418662874511385
0.000412080233574243
0.000411736628852978
0.000408178411072767
0.000406751213831253
0.000388330412225677
0.000382986415727158
0.000379504721038729
0.000363004515776176
0.000363004515776176
0.000346975744949695
0.000345203332790998
0.000339685167867642
0.000333964154514082
0.000327415837758904
0.000317901750288956
0.00030922606899452
0.000305827980324251
0.000305455019372636
0.000304711819812118

0.000302817238549038
0.000286254989583499
0.000278303462095068
0.000275003421042557
0.000269325931059743
0.000267755517467558
0.000265050916281017
0.000264803667291949
0.000258886941483784
0.000257887378003152
0.000257688390828767
0.000254103161043323
0.000253003147359153
0.000234772692101288
0.00023191955174589
0.000230374892504956
0.000229791849436295
0.000222642769676054
0.000222373226371362
0.000216859840593559
0.000211988965267727
0.000211906189412488
0.000208727596571301
0.000197779110118322
0.000197222925894143
0.000192671627604278
0.000187319637948603
0.000180716533827966
0.000170389874752082
0.000164514361829597
0.000164244666154466
0.000161335340344967
0.000159486224696314
0.00015903054976861
0.000148033756433547
0.000145623904584628
0.000138001716741356
0.000130710040905707
0.000122780939159589
0.000115959775872945
0.000115425399025604
0.0001126083372639
0.000112391782769162
0.000108712289880886
0.000106222695456133
0.000106222695456133

0.000105417978066313
0.000104063614781473
0.0000949661102883646
0.0000888202538601282
0.0000847624757649954
0.0000826643946817035
0.0000771279802572938
0.0000759009442077459
0.0000647217353709461
0.0000642238758680927
0.0000625987168723678
0.0000571856428962469
0.0000569904700535976
0.0000525100871877487
0.0000495986368090221
0.000048353883375591
0.0000478916091559391
0.0000461276456511163
0.0000423096479536421
0.0000419552957932264
0.0000411285904573992
0.0000399478653724978
0.0000399096742966159
0.0000388330412225676
0.000036861385707956
0.0000358330637890646
0.0000357563334597518
0.0000350802683313111
0.000034500429185339
0.000034500429185339
0.0000337337529812204
0.0000335980034722416
0.0000324552142384919
0.000032235922250394
0.0000319277394372927
0.0000247747889105402
0.0000238318854315472
0.0000237866206918862
0.0000210304883195265
0.0000208207078874116
0.0000179165318945323
0.0000177640507720256
0.000017285929322675
0.0000169697232984797
0.0000163868574344495
0.0000163387551132134

| |
|---|
| 0.0000162433927292841 |
| 0.0000159182151817961 |
| 0.000015812696709947 |
| 8.12169636464207E-06 |
| 8.03571112882777E-06 |
| 8.00489344472871E-06 |
| 7.77383972332593E-06 |
| 0.0189471970864551 |
| 0.0145816086482583 |
| 0.0143903602499636 |
| 0.0139832395337255 |
| 0.00859764930746518 |
| 0.00739498244572024 |
| 0.00637122935308739 |
| 0.00605244098980079 |
| 0.00487418007503308 |
| 0.00486247920499492 |
| 0.00466107984457518 |
| 0.00460090847378564 |
| 0.00421716747842516 |
| 0.00380236411989154 |
| 0.00375983767907696 |
| 0.00375160085051173 |
| 0.00229409000734694 |
| 0.00194093765617469 |
| 0.00177206952616337 |
| 0.00162100786163797 |
| 0.00157050200706815 |
| 0.00155924745071728 |
| 0.001460175105251 |
| 0.00144677217210744 |
| 0.00140002094846159 |
| 0.00135345020331608 |
| 0.00134254145591944 |
| 0.00134007680768641 |
| 0.00133228348949153 |
| 0.00130052451020512 |
| 0.00130047491918472 |
| 0.00129422148349977 |
| 0.00126668562003668 |
| 0.00115515747149435 |
| 0.00115281530306066 |
| 0.00112392826415461 |
| 0.000983521660842629 |
| 0.000960998264029439 |
| 0.000941728376761108 |

| |
|---|
| 0.000925309353643819 |
| 0.000919992295064528 |
| 0.000863251012031016 |
| 0.000841190321494381 |
| 0.000832335686531282 |
| 0.000830711616899026 |
| 0.000830579704070246 |
| 0.000808752842079563 |
| 0.000803440715735597 |
| 0.000800322775510848 |
| 0.000749398322748806 |
| 0.00071713042519809 |
| 0.000686676941388308 |
| 0.000684432899096189 |
| 0.000662583013620297 |
| 0.000659284149270722 |
| 0.000648732226267029 |
| 0.000640424090413966 |
| 0.000610379503456274 |
| 0.000604182757228291 |
| 0.000597832403627064 |
| 0.000593424517249155 |
| 0.000585402337074431 |
| 0.000572230784490256 |
| 0.000567501604453147 |
| 0.000566391694852601 |
| 0.000555818811703729 |
| 0.000554410448641079 |
| 0.000539951099211319 |
| 0.000511291921726359 |
| 0.000499936102937954 |
| 0.000483575310836836 |
| 0.000478422459163985 |
| 0.000476597694479556 |
| 0.000467127171012454 |
| 0.000463729882496 |
| 0.00044187705366043 |
| 0.000440177526530966 |
| 0.000435985359611624 |
| 0.000423792905003391 |
| 0.000420721101944696 |
| 0.000416167843265641 |
| 0.000413122712705161 |
| 0.000383261548681846 |
| 0.000381487189660171 |
| 0.000381487189660171 |

0.000379626276539877
0.000378334402968764
0.000377987307186224
0.000376135665957523
0.000368995165817394
0.000366227702073764
0.000364146862857436
0.000364021511097416
0.000359046766738984
0.000358344131187636
0.000348314390559286
0.000340821042539036
0.000336606343817798
0.000317080261535726
0.000316927819102296
0.000305189751728137
0.000303997604260449
0.000301421977015444
0.00029855519190796
0.000298305772365848
0.000296712258624577
0.000292202953782258
0.000291317490285948
0.000290302446765788
0.00028836826934942
0.000284026958725091
0.000283897443468034
0.000273544417190421
0.000272910066449199
0.000256288691123049
0.00025432479310678
0.000248060635641096
0.000246499414857341
0.000244151801382509
0.000235225302715362
0.000231734338704745
0.000225139980783051
0.00022384483228382
0.000222534193968433
0.000217992679805812
0.000217992679805812
0.000212100985757006
0.000210475690846991
0.000208281157577496
0.000203459834485424
0.000196006738740715

0.000185902386839474
0.000174726957096261
0.00017274891607253
0.000167994358749433
0.000156816868476908
0.000153733643594397
0.000152594875864068
0.000146101476891129
0.000145538465419487
0.000144096856142825
0.000141948721734017
0.000134774669556095
0.000133076926625641
0.000126315453855128
0.000124850352979692
0.000117380673741591
0.000108672908627229
0.000105065324365424
0.0000940653344367545
0.0000885218893134099
0.0000867016340136752
0.0000843842631506369
0.0000834503227381624
0.0000805148842498853
0.0000781202436164173
0.0000773284843905752
0.0000762974379320342
0.0000751190991222731
0.0000634343594815989
0.0000582423190320872
0.0000563774171911583
0.0000543902527832323
0.0000529741131254239
0.0000472918003710956
0.0000465938552256697
0.0000453252106526936
0.0000435985359611624
0.0000429039013675918
0.0000427836100553463
0.0000427836100553463
0.0000425846165202051
0.0000419985896873583
0.0000419985896873583
0.0000419985896873583
0.0000400394718010675
0.0000384692964363197

| |
|---|
| 0.0000383886480161178 |
| 0.0000379117704010108 |
| 0.0000369926971791681 |
| 0.0000360460336686775 |
| 0.0000348566975324014 |
| 0.0000303839354596596 |
| 0.0000301174097100135 |
| 0.0000295344921027229 |
| 0.0000294080060551738 |
| 0.0000268758098390727 |
| 0.0000257664893579102 |
| 0.0000246120767522691 |
| 0.0000236581202890028 |
| 0.0000230621978635871 |
| 0.000022832151002105 |
| 0.0000219035706981916 |
| 0.0000218826303820365 |
| 0.0000196474089095367 |
| 0.0000196474089095367 |
| 0.0000191141806927851 |
| 0.000018421916603308 |
| 0.0000181660566504843 |
| 0.0000175564574340251 |
| 0.0000175061043056292 |
| 0.0000130920293877656 |
| 0.0000127872801003409 |
| 0.0000124736955747194 |
| 0.0000122402306842835 |
| 0.0000119526012426163 |
| 0.0000116188991774671 |
| 0.0000115894842428406 |
| 0.0000100391365700045 |
| 9.74009845940862E-06 |
| 9.43885830087021E-06 |
| 9.30456560146759E-06 |
| 8.90631571191061E-06 |
| 8.75305215281463E-06 |
| 8.69486472159934E-06 |
| 4.40601181513191E-06 |
| 4.35985359611624E-06 |

**Supplementary Table S6:** [illegible]

| Sample |
|---|
| Hospital-eff-F |
| Hospital-eff-F |
| Hospital-eff-F |
| Hospital-eff-F |
| Hospital-eff-F |
| Hospital-eff-F |
| Hospital-eff-F |
| Hospital-eff-F |
| Hospital-eff-F |
| Hospital-eff-F |
| Hospital-eff-F |
| Hospital-eff-F |
| Hospital-eff-F |
| Hospital-eff-F |
| Hospital-eff-F |
| Hospital-eff-F |
| Hospital-eff-F |
| Hospital-eff-F |
| Hospital-eff-F |
| Hospital-eff-F |
| Hospital-eff-F |
| Hospital-eff-F |
| Hospital-eff-F |
| Hospital-eff-F |
| Hospital-eff-F |
| Hospital-eff-F |
| Hospital-eff-F |
| Hospital-eff-F |
| Hospital-eff-F |
| Hospital-eff-F |
| Hospital-eff-F |
| Hospital-eff-F |
| Hospital-eff-F |
| Hospital-eff-F |
| Hospital-eff-F |
| Hospital-eff-F |
| Hospital-eff-F |
| Hospital-eff-F |
| Hospital-eff-F |
| Hospital-eff-F |
| Hospital-eff-F |
| Hospital-eff-F |
| Hospital-eff-F |

Hospital-eff-F
Hospital-eff-F
Hospital-eff-F
Hospital-eff-F
Hospital-eff-F
Hospital-eff-F
Hospital-eff-F
Hospital-eff-F
Hospital-eff-F
Hospital-eff-F
Hospital-eff-F
Hospital-eff-F
Hospital-eff-F
Hospital-eff-F
Hospital-eff-F
Hospital-eff-F
Hospital-eff-F
Hospital-eff-F
Hospital-eff-F
Hospital-eff-F
Hospital-eff-F
Hospital-eff-F
Hospital-eff-F
Hospital-eff-F
Hospital-eff-F
Hospital-eff-F
Hospital-eff-F
Hospital-eff-F
Hospital-eff-F
Hospital-eff-F
Hospital-eff-F
Hospital-eff-F
Hospital-eff-F
Hospital-eff-F
Hospital-eff-F
Hospital-eff-F
Hospital-eff-F
Hospital-eff-F
Hospital-eff-F
Hospital-eff-F
Hospital-eff-F
Hospital-eff-F
Hospital-eff-F
Hospital-eff-F
Hospital-eff-F
Hospital-eff-F

Hospital-eff-F
Hospital-eff-F
Hospital-eff-F
Hospital-eff-F
Hospital-eff-F
Hospital-eff-F
Hospital-eff-F
Hospital-eff-F
Hospital-eff-F
Hospital-eff-F
Hospital-eff-F
Hospital-eff-F
Hospital-eff-F
Hospital-eff-F
Hospital-eff-F
Hospital-eff-F
Hospital-eff-F
Hospital-eff-F
Hospital-eff-F
Hospital-eff-F
Hospital-eff-F
Hospital-eff-F
Hospital-eff-F
Hospital-eff-F
Hospital-eff-F
Hospital-eff-F
Hospital-eff-F
Hospital-eff-F
Hospital-eff-F
Hospital-eff-F
Hospital-eff-F
Hospital-eff-F
Hospital-eff-F
Hospital-eff-F
Hospital-eff-F
Hospital-eff-F
Hospital-eff-F
Hospital-eff-F
Hospital-eff-F
Hospital-eff-F
Hospital-eff-F
Hospital-eff-F
Hospital-eff-F
Hospital-eff-F
Hospital-eff-F
Hospital-eff-F

Hospital-eff-F
Hospital-eff-F
Hospital-eff-F
Hospital-eff-F
Hospital-eff-F
Hospital-eff-F
Hospital-eff-F
Hospital-eff-F
Hospital-eff-F
Hospital-eff-F
Hospital-eff-F
Hospital-eff-F
Hospital-eff-F
Hospital-eff-F
Hospital-eff-F
Hospital-eff-F
Hospital-eff-F
Hospital-eff-F
Hospital-eff-F
Hospital-eff-F
Hospital-eff-F
Hospital-eff-F
Hospital-eff-F
Hospital-eff-F
Hospital-eff-F
Hospital-eff-F
Hospital-eff-F
Hospital-eff-F
Hospital-eff-F
Hospital-eff-F
Hospital-eff-F
Hospital-eff-F
Hospital-eff-F
Hospital-eff-F
Hospital-eff-F
Hospital-eff-F
Hospital-eff-F
Hospital-eff-F
Hospital-eff-F
Hospital-eff-F
Hospital-eff-F
Hospital-eff-F
Hospital-eff-F
Hospital-eff-F
Hospital-eff-F
Hospital-eff-F

Hospital-eff-F
Hospital-eff-F
Hospital-eff-F
Hospital-eff-F
Hospital-eff-F
Hospital-eff-F
Hospital-eff-F
Hospital-eff-F
Hospital-eff-F
Hospital-eff-F
Hospital-eff-F
Hospital-eff-F
Hospital-eff-F
Hospital-eff-F
Hospital-eff-F
Hospital-eff-F
Hospital-eff-F
Hospital-eff-F
Hospital-eff-F
Hospital-eff-F
Hospital-eff-F
Hospital-eff-F
Hospital-eff-F
Hospital-eff-F
Hospital-eff-F
Hospital-eff-F
Hospital-eff-F
Hospital-eff-F
Hospital-eff-F
Hospital-eff-F
Hospital-eff-F
Hospital-eff-F
Hospital-eff-F
Hospital-eff-F
Hospital-eff-F
Hospital-eff-F
Hospital-eff-F
Hospital-eff-F
Hospital-eff-F
Hospital-eff-F
Hospital-eff-F
Hospital-eff-F
Hospital-eff-F
Hospital-eff-F
Hospital-eff-F
Hospital-eff-F

Hospital-eff-F
Hospital-eff-F
Hospital-eff-F
Hospital-eff-F
Hospital-eff-F
Hospital-eff-F
Hospital-eff-F
Hospital-eff-F
Hospital-eff-F
Hospital-eff-F
Hospital-eff-F
Hospital-eff-F
Hospital-eff-F
Hospital-eff-F
Hospital-eff-F
Hospital-eff-F
Hospital-eff-F
Hospital-eff-F
Hospital-eff-F
Hospital-eff-F
Hospital-eff-F
Hospital-eff-F
Hospital-eff-F
Hospital-eff-F
Hospital-eff-F
Hospital-eff-F
Hospital-eff-F
Hospital-eff-F
Hospital-eff-F
Hospital-eff-F
Hospital-eff-F
Hospital-eff-F
Hospital-eff-F
Hospital-eff-F
Hospital-eff-F
Hospital-eff-F
Hospital-eff-F
Hospital-eff-F
Hospital-eff-F
Hospital-eff-F
Hospital-eff-F
Hospital-eff-F
Hospital-eff-F
Hospital-eff-F
Hospital-eff-F
Hospital-eff-F

Hospital-eff-F
Hospital-eff-F
Hospital-eff-F
Hospital-eff-F
Hospital-eff-F
Hospital-eff-F
Hospital-eff-F
Hospital-eff-F
Hospital-eff-F
Hospital-eff-F
Hospital-eff-F
Hospital-eff-F
Hospital-eff-F
Hospital-eff-F
Hospital-eff-F
Hospital-eff-F
Hospital-eff-F
Hospital-eff-F
Hospital-eff-F
Hospital-eff-A
Hospital-eff-A
Hospital-eff-A
Hospital-eff-A
Hospital-eff-A
Hospital-eff-A
Hospital-eff-A
Hospital-eff-A
Hospital-eff-A
Hospital-eff-A
Hospital-eff-A
Hospital-eff-A
Hospital-eff-A
Hospital-eff-A
Hospital-eff-A
Hospital-eff-A
Hospital-eff-A
Hospital-eff-A
Hospital-eff-A
Hospital-eff-A
Hospital-eff-A
Hospital-eff-A
Hospital-eff-A
Hospital-eff-A
Hospital-eff-A
Hospital-eff-A
Hospital-eff-A

Hospital-eff-A
Hospital-eff-A
Hospital-eff-A
Hospital-eff-A
Hospital-eff-A
Hospital-eff-A
Hospital-eff-A
Hospital-eff-A
Hospital-eff-A
Hospital-eff-A
Hospital-eff-A
Hospital-eff-A
Hospital-eff-A
Hospital-eff-A
Hospital-eff-A
Hospital-eff-A
Hospital-eff-A
Hospital-eff-A
Hospital-eff-A
Hospital-eff-A
Hospital-eff-A
Hospital-eff-A
Hospital-eff-A
Hospital-eff-A
Hospital-eff-A
Hospital-eff-A
Hospital-eff-A
Hospital-eff-A
Hospital-eff-A
Hospital-eff-A
Hospital-eff-A
Hospital-eff-A
Hospital-eff-A
Hospital-eff-A
Hospital-eff-A
Hospital-eff-A
Hospital-eff-A
Hospital-eff-A
Hospital-eff-A
Hospital-eff-A
Hospital-eff-A
Hospital-eff-A
Hospital-eff-A
Hospital-eff-A
Hospital-eff-A
Hospital-eff-A

Hospital-eff-A
Hospital-eff-A
Hospital-eff-A
Hospital-eff-A
Hospital-eff-A
Hospital-eff-A
Hospital-eff-A
Hospital-eff-A
Hospital-eff-A
Hospital-eff-A
Hospital-eff-A
Hospital-eff-A
Hospital-eff-A
Hospital-eff-A
Hospital-eff-A
Hospital-eff-A
Hospital-eff-A
Hospital-eff-A
Hospital-eff-A
Hospital-eff-A
Hospital-eff-A
Hospital-eff-A
Hospital-eff-A
Hospital-eff-A
Hospital-eff-A
Hospital-eff-A
Hospital-eff-A
Hospital-eff-A
Hospital-eff-A
Hospital-eff-A
Hospital-eff-A
Hospital-eff-A
Hospital-eff-A
Hospital-eff-A
Hospital-eff-A
Hospital-eff-A
Hospital-eff-A
Hospital-eff-A
Hospital-eff-A
Hospital-eff-A
Hospital-eff-A
Hospital-eff-A
Hospital-eff-A
Hospital-eff-A
Hospital-eff-A
Hospital-eff-A

Hospital-eff-A
Hospital-eff-A
Hospital-eff-A
Hospital-eff-A
Hospital-eff-A
Hospital-eff-A
Hospital-eff-A
Hospital-eff-A
Hospital-eff-A
Hospital-eff-A
Hospital-eff-A
Hospital-eff-A
Hospital-eff-A
Hospital-eff-A
Hospital-eff-A
Hospital-eff-A
Hospital-eff-A
Hospital-eff-A
Hospital-eff-A
Hospital-eff-A
Hospital-eff-A
Hospital-eff-A
Hospital-eff-A
Hospital-eff-A
Hospital-eff-A
Hospital-eff-A
Hospital-eff-A
Hospital-eff-A
Hospital-eff-A
Hospital-eff-A
Hospital-eff-A
Hospital-eff-A
Hospital-eff-A
Hospital-eff-A
Hospital-eff-A
Hospital-eff-A
Hospital-eff-A
Hospital-eff-A
Hospital-eff-A
Hospital-eff-A
Hospital-eff-A
Hospital-eff-A
Hospital-eff-A
Hospital-eff-A
Hospital-eff-A
Hospital-eff-A

Hospital-eff-A
Hospital-eff-A
Hospital-eff-A
Hospital-eff-A
Hospital-eff-A
Hospital-eff-A
Hospital-eff-A
Hospital-eff-A
Hospital-eff-A
Hospital-eff-A
Hospital-eff-A
Hospital-eff-A
Hospital-eff-A
Hospital-eff-A
Hospital-eff-A
Hospital-eff-A
Hospital-eff-A
Hospital-eff-A
Hospital-eff-A
Hospital-eff-A
Hospital-eff-A
Hospital-eff-A
Hospital-eff-A
Hospital-eff-A
Hospital-eff-A
Hospital-eff-A
Hospital-eff-A
Hospital-eff-A
Hospital-eff-A
Hospital-eff-A
Hospital-eff-A
Hospital-eff-A
Hospital-eff-A
Hospital-eff-A
Hospital-eff-A
Hospital-eff-A
Hospital-eff-A
Hospital-eff-A
Hospital-eff-A
Hospital-eff-A
Hospital-eff-A
Hospital-eff-A
Hospital-eff-A
Hospital-eff-A
Hospital-eff-A
Hospital-eff-A

Hospital-eff-A
Hospital-eff-A
Hospital-eff-A
Hospital-eff-A
Hospital-eff-A
Hospital-eff-A
Hospital-eff-A
Hospital-eff-A
Hospital-eff-A
Hospital-eff-A
Hospital-eff-A
Hospital-eff-A
Hospital-eff-A
Hospital-eff-A
Hospital-eff-A
Hospital-eff-A
Hospital-eff-A
Hospital-eff-A
Hospital-eff-A
Hospital-eff-A
Hospital-eff-A
Hospital-eff-A
Hospital-eff-A
Hospital-eff-A
Hospital-eff-A
Hospital-eff-A
Hospital-eff-A
Hospital-eff-A
Hospital-eff-A
Hospital-eff-A
Hospital-eff-A
Hospital-eff-A
Hospital-eff-A
Hospital-eff-A
Hospital-eff-A
Hospital-eff-A
Hospital-eff-A
Hospital-eff-A
Hospital-eff-A
Hospital-eff-A
Hospital-eff-A
Hospital-eff-A
Hospital-eff-A
Hospital-eff-A
Hospital-eff-A
Hospital-eff-A

Hospital-eff-A
Hospital-eff-A
Hospital-eff-A
Hospital-eff-A
Hospital-eff-A
Hospital-eff-A
Hospital-eff-A
Hospital-eff-A
STP-inf-F
STP-inf-F
STP-inf-F
STP-inf-F
STP-inf-F
STP-inf-F
STP-inf-F
STP-inf-F
STP-inf-F
STP-inf-F
STP-inf-F
STP-inf-F
STP-inf-F
STP-inf-F
STP-inf-F
STP-inf-F
STP-inf-F
STP-inf-F
STP-inf-F
STP-inf-F
STP-inf-F
STP-inf-F
STP-inf-F
STP-inf-F
STP-inf-F
STP-inf-F
STP-inf-F
STP-inf-F
STP-inf-F
STP-inf-F
STP-inf-F
STP-inf-F
STP-inf-F
STP-inf-F
STP-inf-F
STP-inf-F
STP-inf-F
STP-inf-F

STP-inf-F
STP-inf-F
STP-inf-F
STP-inf-F
STP-inf-F
STP-inf-F
STP-inf-F
STP-inf-F
STP-inf-F
STP-inf-F
STP-inf-F
STP-inf-F
STP-inf-F
STP-inf-F
STP-inf-F
STP-inf-F
STP-inf-F
STP-inf-F
STP-inf-F
STP-inf-F
STP-inf-F
STP-inf-F
STP-inf-F
STP-inf-F
STP-inf-F
STP-inf-F
STP-inf-F
STP-inf-F
STP-inf-F
STP-inf-F
STP-inf-F
STP-inf-F
STP-inf-F
STP-inf-F
STP-inf-F
STP-inf-F
STP-inf-F
STP-inf-F
STP-inf-F
STP-inf-F
STP-inf-F
STP-inf-F
STP-inf-F
STP-inf-F
STP-inf-F
STP-inf-F

STP-inf-F
STP-inf-F
STP-inf-F
STP-inf-F
STP-inf-F
STP-inf-F
STP-inf-F
STP-inf-F
STP-inf-F
STP-inf-F
STP-inf-F
STP-inf-F
STP-inf-F
STP-inf-F
STP-inf-F
STP-inf-F
STP-inf-F
STP-inf-F
STP-inf-F
STP-inf-F
STP-inf-F
STP-inf-F
STP-inf-F
STP-inf-F
STP-inf-F
STP-inf-F
STP-inf-F
STP-inf-F
STP-inf-F
STP-inf-F
STP-inf-F
STP-inf-F
STP-inf-F
STP-inf-F
STP-inf-F
STP-inf-F
STP-inf-F
STP-inf-F
STP-inf-F
STP-inf-F
STP-inf-F
STP-inf-F
STP-inf-F
STP-inf-F
STP-inf-F
STP-inf-F

STP-inf-F
STP-inf-F
STP-inf-F
STP-inf-F
STP-inf-F
STP-inf-F
STP-inf-F
STP-inf-F
STP-inf-F
STP-inf-F
STP-inf-F
STP-inf-F
STP-inf-F
STP-inf-F
STP-inf-F
STP-inf-F
STP-inf-F
STP-inf-F
STP-inf-F
STP-inf-F
STP-inf-F
STP-inf-F
STP-inf-F
STP-inf-F
STP-inf-F
STP-inf-F
STP-inf-F
STP-inf-F
STP-inf-F
STP-inf-F
STP-inf-F
STP-inf-F
STP-inf-F
STP-inf-F
STP-inf-F
STP-inf-F
STP-inf-F
STP-inf-F
STP-inf-F
STP-inf-F
STP-inf-F
STP-inf-F
STP-inf-F
STP-inf-F
STP-inf-F
STP-inf-F

STP-inf-F
STP-inf-F
STP-inf-F
STP-inf-F
STP-inf-F
STP-inf-F
STP-inf-F
STP-inf-F
STP-inf-F
STP-inf-F
STP-inf-F
STP-inf-F
STP-inf-F
STP-inf-F
STP-inf-F
STP-inf-F
STP-inf-F
STP-inf-F
STP-inf-F
STP-inf-F
STP-inf-F
STP-inf-F
STP-inf-F
STP-inf-F
STP-inf-F
STP-inf-F
STP-inf-F
STP-inf-F
STP-inf-F
STP-inf-F
STP-inf-F
STP-inf-F
STP-inf-F
STP-inf-F
STP-inf-F
STP-inf-F
STP-inf-F
STP-inf-F
STP-inf-F
STP-inf-F
STP-inf-F
STP-inf-F
STP-inf-F
STP-inf-F
STP-inf-F
STP-inf-F

STP-inf-F
STP-inf-F
STP-inf-F
STP-inf-F
STP-inf-F
STP-inf-F
STP-inf-F
STP-inf-F
STP-inf-F
STP-inf-F
STP-inf-F
STP-inf-F
STP-inf-F
STP-inf-F
STP-inf-F
STP-inf-F
STP-inf-F
STP-inf-F
STP-inf-F
STP-inf-F
STP-inf-F
STP-inf-F
STP-inf-F
STP-inf-F
STP-inf-F
STP-inf-F
STP-inf-F
STP-inf-F
STP-inf-F
STP-inf-F
STP-inf-F
STP-inf-F
STP-inf-F
STP-inf-F
STP-inf-F
STP-inf-F
STP-inf-F
STP-inf-F
STP-inf-F
STP-inf-F
STP-inf-F
STP-inf-F
STP-inf-F
STP-inf-F
STP-inf-F
STP-inf-F

STP-inf-F
STP-inf-F
STP-inf-F
STP-inf-F
STP-inf-F
STP-inf-F
STP-inf-F
STP-inf-F
STP-inf-F
STP-inf-F
STP-inf-A
STP-inf-A
STP-inf-A
STP-inf-A
STP-inf-A
STP-inf-A
STP-inf-A
STP-inf-A
STP-inf-A
STP-inf-A
STP-inf-A
STP-inf-A
STP-inf-A
STP-inf-A
STP-inf-A
STP-inf-A
STP-inf-A
STP-inf-A
STP-inf-A
STP-inf-A
STP-inf-A
STP-inf-A
STP-inf-A
STP-inf-A
STP-inf-A
STP-inf-A
STP-inf-A
STP-inf-A
STP-inf-A
STP-inf-A
STP-inf-A
STP-inf-A
STP-inf-A
STP-inf-A
STP-inf-A
STP-inf-A

STP-inf-A
STP-inf-A
STP-inf-A
STP-inf-A
STP-inf-A
STP-inf-A
STP-inf-A
STP-inf-A
STP-inf-A
STP-inf-A
STP-inf-A
STP-inf-A
STP-inf-A
STP-inf-A
STP-inf-A
STP-inf-A
STP-inf-A
STP-inf-A
STP-inf-A
STP-inf-A
STP-inf-A
STP-inf-A
STP-inf-A
STP-inf-A
STP-inf-A
STP-inf-A
STP-inf-A
STP-inf-A
STP-inf-A
STP-inf-A
STP-inf-A
STP-inf-A
STP-inf-A
STP-inf-A
STP-inf-A
STP-inf-A
STP-inf-A
STP-inf-A
STP-inf-A
STP-inf-A
STP-inf-A
STP-inf-A
STP-inf-A
STP-inf-A
STP-inf-A
STP-inf-A

STP-inf-A
STP-inf-A
STP-inf-A
STP-inf-A
STP-inf-A
STP-inf-A
STP-inf-A
STP-inf-A
STP-inf-A
STP-inf-A
STP-inf-A
STP-inf-A
STP-inf-A
STP-inf-A
STP-inf-A
STP-inf-A
STP-inf-A
STP-inf-A
STP-inf-A
STP-inf-A
STP-inf-A
STP-inf-A
STP-inf-A
STP-inf-A
STP-inf-A
STP-inf-A
STP-inf-A
STP-inf-A
STP-inf-A
STP-inf-A
STP-inf-A
STP-inf-A
STP-inf-A
STP-inf-A
STP-inf-A
STP-inf-A
STP-inf-A
STP-inf-A
STP-inf-A
STP-inf-A
STP-inf-A
STP-inf-A
STP-inf-A
STP-inf-A
STP-inf-A
STP-inf-A

STP-inf-A
STP-inf-A
STP-inf-A
STP-inf-A
STP-inf-A
STP-inf-A
STP-inf-A
STP-inf-A
STP-inf-A
STP-inf-A
STP-inf-A
STP-inf-A
STP-inf-A
STP-inf-A
STP-inf-A
STP-inf-A
STP-inf-A
STP-inf-A
STP-inf-A
STP-inf-A
STP-inf-A
STP-inf-A
STP-inf-A
STP-inf-A
STP-inf-A
STP-inf-A
STP-inf-A
STP-inf-A
STP-inf-A
STP-inf-A
STP-inf-A
STP-inf-A
STP-inf-A
STP-inf-A
STP-inf-A
STP-inf-A
STP-inf-A
STP-inf-A
STP-inf-A
STP-inf-A
STP-inf-A
STP-inf-A
STP-inf-A
STP-inf-A
STP-inf-A
STP-inf-A

STP-inf-A
STP-inf-A
STP-inf-A
STP-inf-A
STP-inf-A
STP-inf-A
STP-inf-A
STP-inf-A
STP-inf-A
STP-inf-A
STP-inf-A
STP-inf-A
STP-inf-A
STP-inf-A
STP-inf-A
STP-inf-A
STP-inf-A
STP-inf-A
STP-inf-A
STP-inf-A
STP-inf-A
STP-inf-A
STP-inf-A
STP-inf-A
STP-inf-A
STP-inf-A
STP-inf-A
STP-inf-A
STP-inf-A
STP-inf-A
STP-inf-A
STP-inf-A
STP-inf-A
STP-inf-A
STP-inf-A
STP-inf-A
STP-inf-A
STP-inf-A
STP-inf-A
STP-inf-A
STP-inf-A
STP-inf-A
STP-inf-A
STP-inf-A
STP-inf-A
STP-inf-A

STP-inf-A
STP-inf-A
STP-inf-A
STP-inf-A
STP-inf-A
STP-inf-A
STP-inf-A
STP-inf-A
STP-inf-A
STP-inf-A
STP-inf-A
STP-inf-A
STP-inf-A
STP-inf-A
STP-inf-A
STP-inf-A
STP-inf-A
STP-inf-A
STP-inf-A
STP-inf-A
STP-inf-A
STP-inf-A
STP-inf-A
STP-inf-A
STP-inf-A
STP-inf-A
STP-inf-A
STP-inf-A
STP-inf-A
STP-inf-A
STP-inf-A
STP-inf-A
STP-inf-A
STP-inf-A
STP-inf-A
STP-inf-A
STP-inf-A
STP-inf-A
STP-inf-A
STP-inf-A
STP-inf-A
STP-inf-A
STP-inf-A
STP-inf-A
STP-inf-A
STP-inf-A

STP-inf-A
STP-T-eff-F
STP-T-eff-F
STP-T-eff-F
STP-T-eff-F
STP-T-eff-F
STP-T-eff-F
STP-T-eff-F
STP-T-eff-F
STP-T-eff-F
STP-T-eff-F
STP-T-eff-F
STP-T-eff-F
STP-T-eff-F
STP-T-eff-F
STP-T-eff-F
STP-T-eff-F
STP-T-eff-F
STP-T-eff-F
STP-T-eff-F
STP-T-eff-F
STP-T-eff-F
STP-T-eff-F
STP-T-eff-F
STP-T-eff-F
STP-T-eff-F
STP-T-eff-F
STP-T-eff-F
STP-T-eff-F
STP-T-eff-F
STP-T-eff-F
STP-T-eff-F
STP-T-eff-F
STP-T-eff-F
STP-T-eff-F
STP-T-eff-F
STP-T-eff-F
STP-T-eff-F
STP-T-eff-F
STP-T-eff-F
STP-T-eff-F
STP-T-eff-F
STP-T-eff-F
STP-T-eff-F
STP-T-eff-F
STP-T-eff-F

STP-T-eff-F
STP-T-eff-F
STP-T-eff-F
STP-T-eff-F
STP-T-eff-F
STP-T-eff-F
STP-T-eff-F
STP-T-eff-F
STP-T-eff-F
STP-T-eff-F
STP-T-eff-F
STP-T-eff-F
STP-T-eff-F
STP-T-eff-F
STP-T-eff-F
STP-T-eff-F
STP-T-eff-F
STP-T-eff-F
STP-T-eff-F
STP-T-eff-F
STP-T-eff-F
STP-T-eff-F
STP-T-eff-F
STP-T-eff-F
STP-T-eff-F
STP-T-eff-F
STP-T-eff-F
STP-T-eff-F
STP-T-eff-F
STP-T-eff-F
STP-T-eff-F
STP-T-eff-F
STP-T-eff-F
STP-T-eff-F
STP-T-eff-F
STP-T-eff-F
STP-T-eff-F
STP-T-eff-F
STP-T-eff-F
STP-T-eff-F
STP-T-eff-F
STP-T-eff-F
STP-T-eff-F
STP-T-eff-F
STP-T-eff-F
STP-T-eff-F

STP-T-eff-F
STP-T-eff-F
STP-T-eff-F
STP-T-eff-F
STP-T-eff-F
STP-T-eff-F
STP-T-eff-F
STP-T-eff-F
STP-T-eff-F
STP-T-eff-F
STP-T-eff-F
STP-T-eff-F
STP-T-eff-F
STP-T-eff-F
STP-T-eff-F
STP-T-eff-F
STP-T-eff-F
STP-T-eff-F
STP-T-eff-F
STP-T-eff-F
STP-T-eff-F
STP-T-eff-F
STP-T-eff-F
STP-T-eff-F
STP-T-eff-F
STP-T-eff-F
STP-T-eff-F
STP-T-eff-F
STP-T-eff-F
STP-T-eff-F
STP-T-eff-F
STP-T-eff-F
STP-T-eff-F
STP-T-eff-F
STP-T-eff-F
STP-T-eff-F
STP-T-eff-F
STP-T-eff-F
STP-T-eff-F
STP-T-eff-F
STP-T-eff-F
STP-T-eff-F
STP-T-eff-F
STP-T-eff-F
STP-T-eff-F
STP-T-eff-F

STP-T-eff-F
STP-T-eff-F
STP-T-eff-F
STP-T-eff-F
STP-T-eff-F
STP-T-eff-F
STP-T-eff-F
STP-T-eff-F
STP-T-eff-F
STP-T-eff-F
STP-T-eff-F
STP-T-eff-F
STP-T-eff-F
STP-T-eff-F
STP-T-eff-F
STP-T-eff-F
STP-T-eff-F
STP-T-eff-F
STP-T-eff-F
STP-T-eff-F
STP-T-eff-F
STP-T-eff-F
STP-T-eff-F
STP-T-eff-F
STP-T-eff-F
STP-T-eff-F
STP-T-eff-F
STP-T-eff-F
STP-T-eff-F
STP-T-eff-F
STP-T-eff-F
STP-T-eff-F
STP-T-eff-F
STP-T-eff-F
STP-T-eff-F
STP-T-eff-F
STP-T-eff-F
STP-T-eff-F
STP-T-eff-F
STP-T-eff-F
STP-T-eff-F
STP-T-eff-F
STP-T-eff-F
STP-T-eff-F
STP-T-eff-F
STP-T-eff-F

STP-T-eff-F
STP-T-eff-F
STP-T-eff-F
STP-T-eff-F
STP-T-eff-F
STP-T-eff-F
STP-T-eff-F
STP-T-eff-F
STP-T-eff-F
STP-T-eff-F
STP-T-eff-F
STP-T-eff-F
STP-T-eff-F
STP-T-eff-F
STP-T-eff-F
STP-T-eff-F
STP-T-eff-F
STP-T-eff-F
STP-T-eff-F
STP-T-eff-F
STP-T-eff-F
STP-T-eff-F
STP-T-eff-F
STP-T-eff-F
STP-T-eff-F
STP-T-eff-F
STP-T-eff-F
STP-T-eff-F
STP-T-eff-F
STP-T-eff-F
STP-T-eff-F
STP-T-eff-F
STP-T-eff-F
STP-T-eff-F
STP-T-eff-F
STP-T-eff-F
STP-T-eff-F
STP-T-eff-F
STP-T-eff-F
STP-T-eff-F
STP-T-eff-F
STP-T-eff-F
STP-T-eff-F
STP-T-eff-F
STP-T-eff-F
STP-T-eff-F

STP-T-eff-F
STP-T-eff-F
STP-T-eff-F
STP-T-eff-F
STP-T-eff-F
STP-T-eff-F
STP-T-eff-F
STP-T-eff-F
STP-T-eff-F
STP-T-eff-F
STP-T-eff-F
STP-T-eff-F
STP-T-eff-F
STP-T-eff-F
STP-T-eff-F
STP-T-eff-F
STP-T-eff-F
STP-T-eff-F
STP-T-eff-F
STP-T-eff-F
STP-T-eff-F
STP-T-eff-F
STP-T-eff-F
STP-T-eff-F
STP-T-eff-F
STP-T-eff-F
STP-T-eff-F
STP-T-eff-F
STP-T-eff-A
STP-T-eff-A
STP-T-eff-A
STP-T-eff-A
STP-T-eff-A
STP-T-eff-A
STP-T-eff-A
STP-T-eff-A
STP-T-eff-A
STP-T-eff-A
STP-T-eff-A
STP-T-eff-A
STP-T-eff-A
STP-T-eff-A
STP-T-eff-A
STP-T-eff-A
STP-T-eff-A
STP-T-eff-A

STP-T-eff-A
STP-T-eff-A
STP-T-eff-A
STP-T-eff-A
STP-T-eff-A
STP-T-eff-A
STP-T-eff-A
STP-T-eff-A
STP-T-eff-A
STP-T-eff-A
STP-T-eff-A
STP-T-eff-A
STP-T-eff-A
STP-T-eff-A
STP-T-eff-A
STP-T-eff-A
STP-T-eff-A
STP-T-eff-A
STP-T-eff-A
STP-T-eff-A
STP-T-eff-A
STP-T-eff-A
STP-T-eff-A
STP-T-eff-A
STP-T-eff-A
STP-T-eff-A
STP-T-eff-A
STP-T-eff-A
STP-T-eff-A
STP-T-eff-A
STP-T-eff-A
STP-T-eff-A
STP-T-eff-A
STP-T-eff-A
STP-T-eff-A
STP-T-eff-A
STP-T-eff-A
STP-T-eff-A
STP-T-eff-A
STP-T-eff-A
STP-T-eff-A
STP-T-eff-A
STP-T-eff-A
STP-T-eff-A
STP-T-eff-A
STP-T-eff-A

STP-T-eff-A
STP-T-eff-A
STP-T-eff-A
STP-T-eff-A
STP-T-eff-A
STP-T-eff-A
STP-T-eff-A
STP-T-eff-A
STP-T-eff-A
STP-T-eff-A
STP-T-eff-A
STP-T-eff-A
STP-T-eff-A
STP-T-eff-A
STP-T-eff-A
STP-T-eff-A
STP-T-eff-A
STP-T-eff-A
STP-T-eff-A
STP-T-eff-A
STP-T-eff-A
STP-T-eff-A
STP-T-eff-A
STP-T-eff-A
STP-T-eff-A
STP-T-eff-A
STP-T-eff-A
STP-T-eff-A
STP-T-eff-A
STP-T-eff-A
STP-T-eff-A
STP-T-eff-A
STP-T-eff-A
STP-T-eff-A
STP-T-eff-A
STP-T-eff-A
STP-T-eff-A
STP-T-eff-A
STP-T-eff-A
STP-T-eff-A
STP-T-eff-A
STP-T-eff-A
STP-T-eff-A
STP-T-eff-A
STP-T-eff-A
STP-T-eff-A

STP-T-eff-A
STP-T-eff-A
STP-T-eff-A
STP-T-eff-A
STP-T-eff-A
STP-T-eff-A
STP-T-eff-A
STP-T-eff-A
STP-T-eff-A
STP-T-eff-A
STP-T-eff-A
STP-T-eff-A
STP-T-eff-A
STP-T-eff-A
STP-T-eff-A
STP-T-eff-A
STP-T-eff-A
STP-T-eff-A
STP-T-eff-A
STP-T-eff-A
STP-T-eff-A
STP-T-eff-A
STP-T-eff-A
STP-T-eff-A
STP-T-eff-A
STP-T-eff-A
STP-T-eff-A
STP-T-eff-A
STP-T-eff-A
STP-T-eff-A
STP-T-eff-A
STP-T-eff-A
STP-T-eff-A
STP-T-eff-A
STP-T-eff-A
STP-T-eff-A
STP-T-eff-A
STP-T-eff-A
STP-T-eff-A
STP-T-eff-A
STP-T-eff-A
STP-T-eff-A
STP-T-eff-A
STP-T-eff-A
STP-T-eff-A
STP-T-eff-A

STP-T-eff-A
STP-T-eff-A
STP-T-eff-A
STP-T-eff-A
STP-T-eff-A
STP-T-eff-A
STP-T-eff-A
STP-T-eff-A
STP-T-eff-A
STP-T-eff-A
STP-T-eff-A
STP-T-eff-A
STP-T-eff-A
STP-T-eff-A
STP-T-eff-A
STP-T-eff-A
STP-T-eff-A
STP-T-eff-A
STP-T-eff-A
STP-T-eff-A
STP-T-eff-A
STP-T-eff-A
STP-T-eff-A
STP-T-eff-A
STP-T-eff-A
STP-T-eff-A
STP-T-eff-A
STP-T-eff-A
STP-T-eff-A
STP-T-eff-A
STP-T-eff-A
STP-T-eff-A
STP-T-eff-A
STP-T-eff-A
STP-T-eff-A
STP-T-eff-A
STP-T-eff-A
STP-T-eff-A
STP-T-eff-A
STP-T-eff-A
STP-T-eff-A
STP-T-eff-A
STP-T-eff-A
STP-T-eff-A
STP-T-eff-A
STP-T-eff-A

STP-T-eff-A
STP-T-eff-A
STP-T-eff-A
STP-T-eff-A
STP-T-eff-A
STP-T-eff-A
STP-T-eff-A
STP-T-eff-A
STP-T-eff-A
STP-T-eff-A
STP-T-eff-A
STP-T-eff-A
STP-T-eff-A
STP-T-eff-A
STP-T-eff-A
STP-T-eff-A
STP-T-eff-A
STP-T-eff-A
STP-T-eff-A
STP-T-eff-A
STP-T-eff-A
STP-T-eff-A
STP-T-eff-A
STP-T-eff-A
STP-T-eff-A
STP-T-eff-A
STP-T-eff-A
STP-T-eff-A
STP-T-eff-A
STP-T-eff-A
STP-T-eff-A
STP-T-eff-A
STP-T-eff-A
STP-T-eff-A
STP-T-eff-A
STP-T-eff-A
STP-T-eff-A
STP-T-eff-A
STP-T-eff-A
STP-T-eff-A
STP-T-eff-A
STP-T-eff-A
STP-T-eff-A
STP-T-eff-A
STP-T-eff-A
STP-T-eff-A

| |
|---|
| STP-T-eff-A |
| STP-T-eff-A |
| STP-T-eff-A |
| STP-T-eff-A |
| STP-T-eff-A |
| STP-T-eff-A |
| STP-T-eff-A |
| STP-T-eff-A |
| STP-T-eff-A |
| STP-T-eff-A |
| STP-T-eff-A |
| STP-T-eff-A |
| STP-T-eff-A |
| STP-T-eff-A |
| STP-T-eff-A |
| STP-T-eff-A |
| STP-T-eff-A |
| STP-T-eff-A |
| STP-T-eff-A |
| STP-T-eff-A |
| STP-T-eff-A |
| STP-T-eff-A |
| STP-T-eff-A |
| STP-T-eff-A |
| STP-T-eff-A |
| STP-T-eff-A |
| STP-T-eff-A |
| STP-T-eff-A |
| STP-T-eff-A |
| STP-T-eff-A |
| STP-T-eff-A |
| STP-T-eff-A |
| STP-T-eff-A |
| STP-T-eff-A |
| STP-T-eff-A |
| STP-T-eff-A |
| STP-T-eff-A |
| STP-T-eff-A |

**Legend:** eff (effluent); F (F[illegible]

Detected biocide/metal resistance genes (BMRGs) across all samples.

| |
|---|
| BAC0116\|cutF/nlpE\|sp\|P40710\|NLPE_ECOLI Lipoprotein NlpE OS=Escherichia coli (strain K12) GN=nlpE PE= |
| BAC0307\|pcoE\|sp\|Q47459\|PCOE_ECOLX Probable copper-binding protein PcoE OS=Escherichia coli GN=pco |
| BAC0646\|mdtB\|tr\|D0ZND9\|D0ZND9_SALT1 Multidrug resistance protein MdtB OS=Salmonella typhimurium ( |
| BAC0571\|arsA\|sp\|O50593\|ARSA_ACIMA Arsenical pump-driving ATPase OS=Acidiphilium multivorum (strai |
| BAC0648\|merA\|sp\|P08662\|MERA_SERMA Mercuric reductase (Fragments) OS=Serratia marcescens GN=merA |
| BAC0667\|merD\|sp\|P08654\|MERD_SERMA HTH-type transcriptional regulator MerD OS=Serratia marcescens G |
| BAC0694\|merT-P\|tr\|H6WCN3\|H6WCN3_9FLAO MerT-P OS=Tenacibaculum discolor GN=merT-P PE=4 SV= |
| BAC0594\|arsR\|sp\|P37309\|ARSR_ECOLI Arsenical resistance operon repressor OS=Escherichia coli (strain K12) |
| BAC0063\|chrA\|sp\|P14285\|CHRA_PSEAI Chromate transport protein OS=Pseudomonas aeruginosa GN=chrA P |
| BAC0612\|perO\|tr\|D5AQ60\|D5AQ60_RHOCB Divalent ion symporter OS=Rhodobacter capsulatus (strain ATCC |
| BAC0309\|pcoS\|sp\|Q47457\|PCOS_ECOLX Probable sensor protein PcoS OS=Escherichia coli GN=pcoS PE=3 S |
| BAC0278\|nirC\|tr\|Q6RUG1\|Q6RUG1_KLEOX NirC OS=Klebsiella oxytoca GN=nirC PE=4 SV=1 |
| BAC0583\|arsC\|sp\|P52147\|ARSC2_ECOLX Arsenate reductase OS=Escherichia coli GN=arsC PE=3 SV=1 |
| BAC0182\|golS\|tr\|Q8ZRG6\|Q8ZRG6_SALTY Putative transcriptional regulator OS=Salmonella typhimurium (st |
| BAC0613\|cmeB\|tr\|Q8RTE4\|Q8RTE4_CAMJU CmeB OS=Campylobacter jejuni GN=cmeB PE=4 SV=1 |
| BAC0350\|sitB\|tr\|Q9XCS1\|Q9XCS1_SALTM SitB OS=Salmonella typhimurium GN=sitB PE=3 SV=1 |
| BAC0275\|nikR\|sp\|P0A6Z6\|NIKR_ECOLI Nickel-responsive regulator OS=Escherichia coli (strain K12) GN=nik |
| BAC0102\|cueA\|tr\|Q8KWW2\|Q8KWW2_PSEPU Copper transporter OS=Pseudomonas putida GN=cueA PE=3 S |
| BAC0168\|fptA\|sp\|P42512\|FPTA_PSEAE Fe(3+)-pyochelin receptor OS=Pseudomonas aeruginosa (strain ATCC |
| BAC0098\|ctpC\|sp\|P0A502\|CTPC_MYCTU Probable manganese/zinc-exporting P-type ATPase OS=Mycobacter |
| BAC0672\|merE\|tr\|Q79BE4\|Q79BE4_PSEST Urf1 OS=Pseudomonas stutzeri PE=4 SV=1 |
| BAC0637\|copS\|tr\|C6FFR5\|C6FFR5_PSEFL CopS OS=Pseudomonas fluorescens GN=copS PE=4 SV=1 |
| BAC0596\|baeR\|tr\|D0ZNE3\|D0ZNE3_SALT1 DNA-binding transcriptional regulator BaeR OS=Salmonella typhi |
| BAC0103\|cueO\|sp\|P36649\|CUEO_ECOLI Blue copper oxidase CueO OS=Escherichia coli (strain K12) GN=cue |
| BAC0255\|mreA\|tr\|Q88IN0\|Q88IN0_PSEPK Putative uncharacterized protein OS=Pseudomonas putida (strain K |
| BAC0445\|ygiW\|sp\|P0ADU5\|YGIW_ECOLI Protein YgiW OS=Escherichia coli (strain K12) GN=ygiW PE=1 S |
| BAC0652\|merA\|tr\|O66017\|O66017_PSEST MerA OS=Pseudomonas stutzeri GN=merA PE=4 SV=1 |
| BAC0274\|nikE\|sp\|P33594\|NIKE_ECOLI Nickel import ATP-binding protein NikE OS=Escherichia coli (strain K |
| BAC0654\|merB1\|sp\|P16172\|MERB_BACCE Alkylmercury lyase OS=Bacillus cereus GN=merB1 PE=3 SV=2 |
| BAC0640\|copD\|tr\|C6FFR7\|C6FFR7_PSEFL CopD OS=Pseudomonas fluorescens GN=copD PE=4 SV=1 |
| BAC0389\|terD\|sp\|P18781\|TERD_ALCSP Tellurium resistance protein TerD OS=Alcaligenes sp. GN=terD PE=3 |
| BAC0034\|arsH\|tr\|E8PS81\|E8PS81_YERPE Arsenic resistance protein ArsH OS=Yersinia pestis Java 9 GN=arsH |
| BAC0131\|czrB\|tr\|Q9RLI9\|Q9RLI9_PSEAI CzrB protein OS=Pseudomonas aeruginosa GN=czrB PE=4 SV=1 |
| BAC0650\|merA\|tr\|O08449\|O08449_9PSED Mercuric reductase OS=Pseudomonas sp. K-62 GN=merA PE=4 SV |
| BAC0661\|merB2\|tr\|Q7DJN2\|Q7DJN2_BACME MerB2 OS=Bacillus megaterium GN=merB2 PE=4 SV=1 |
| BAC0112\|cusS\|sp\|P77485\|CUSS_ECOLI Sensor kinase CusS OS=Escherichia coli (strain K12) GN=cusS PE=1 |
| BAC0639\|copC\|tr\|C6FFR6\|C6FFR6_PSEFL CopC OS=Pseudomonas fluorescens GN=copC PE=4 SV=1 |
| BAC0293\|ruvB\|tr\|B5L348\|B5L348_9PSED Malic enzyme family protein (Fragment) OS=Pseudomonas corrugat |
| BAC0621\|copA\|tr\|F4ZBX3\|F4ZBX3_XANCI CopA OS=Xanthomonas citri subsp. citri GN=copA PE=4 SV=1 |
| BAC0122\|czcD\|sp\|P13512\|CZCD_RALME Cobalt-zinc-cadmium resistance protein CzcD OS=Ralstonia metalli |
| BAC0641\|corA\|sp\|P0A2R8\|CORA_SALTY Magnesium transport protein CorA OS=Salmonella typhimurium (st |
| BAC0110\|cusF/cusX\|sp\|P77214\|CUSF_ECOLI Cation efflux system protein CusF OS=Escherichia coli (strain K |
| BAC0169\|fpvA\|sp\|P48632\|FPVA_PSEAE Ferripyoverdine receptor OS=Pseudomonas aeruginosa (strain ATCC |

BAC0272|nikC|sp|P0AFA9|NIKC_ECOLI Nickel transport system permease protein NikC OS=Escherichia coli (
BAC0228|merF|tr|Q2QCN0|Q2QCN0_9PSED MerF OS=Pseudomonas sp. CT14 GN=merF PE=4 SV=1
BAC0312|pitA|sp|P0AFJ7|PITA_ECOLI Low-affinity inorganic phosphate transporter 1 OS=Escherichia coli (st
BAC0291|cnrR/cnrX|sp|P37975|CNRR_RALME Nickel and cobalt resistance protein CnrR OS=Ralstonia metall
BAC0549|nccA|sp|Q44586|NCCA_ALCXX Nickel-cobalt-cadmium resistance protein NccA OS=Alcaligenes xy
BAC0229|merG|tr|O07302|O07302_9PSED Mercuric resistance protein OS=Pseudomonas sp. K-62 GN=merG P
BAC0101|ctpV|sp|P77894|CTPV_MYCTU Probable copper-exporting P-type ATPase V OS=Mycobacterium tub
BAC0124|czcP|tr|Q1LAJ7|Q1LAJ7_RALME CzcP cation efflux P1-ATPase OS=Ralstonia metallidurans (strain
BAC0383|tcrB|tr|Q8VPE6|Q8VPE6_ENTFC TcrB OS=Enterococcus faecium GN=tcrB PE=3 SV=1
BAC0078|copA|sp|O32220|COPA_BACSU Copper-exporting P-type ATPase A OS=Bacillus subtilis (strain 168)
BAC0083|copR|sp|Q02540|COPR_PSEUB Transcriptional activator protein CopR OS=Pseudomonas syringae pv
BAC0468|zraS/hydG|sp|P14377|ZRAS_ECOLI Sensor protein ZraS OS=Escherichia coli (strain K12) GN=zraS
BAC0570|actP|tr|D5AU53|D5AU53_RHOCB Cation/acetate symporter ActP-1 OS=Rhodobacter capsulatus (stra
BAC0254|mrdH|tr|Q88IN1|Q88IN1_PSEPK Membrane protein, putative OS=Pseudomonas putida (strain KT244
BAC0252|mntP/yebN|sp|P76264|MNTP_ECOLI Probable manganese efflux pump MntP OS=Escherichia coli (st
BAC0134|dpr/dps|sp|P0CB53|DPS_STRSU DNA protection during starvation protein OS=Streptococcus suis GN
BAC0685|merR|tr|H6WCN2|H6WCN2_9FLAO MerR OS=Tenacibaculum discolor GN=merR PE=4 SV=1
BAC0012|actP|sp|Q9X5X3|ATCU_SINMW Copper-transporting P-type ATPase OS=Sinorhizobium medicae (st
BAC0345|silF|tr|Q9ZHD1|Q9ZHD1_SALTM Uncharacterized protein OS=Salmonella typhimurium GN=ORF96
BAC0113|cutA|sp|P69488|CUTA_ECOLI Divalent-cation tolerance protein CutA OS=Escherichia coli (strain K1
BAC0233|merT|sp|P94185|MERT_ALCSP Mercuric transport protein OS=Alcaligenes sp. GN=merT PE=3 SV=
BAC0088|corC|sp|P0A2L3|CORC_SALTY Magnesium and cobalt efflux protein CorC OS=Salmonella typhimur
BAC0591|arsR|sp|P52144|ARSR2_ECOLX Arsenical resistance operon repressor OS=Escherichia coli GN=arsR
BAC0540|nfsA|sp|P17117|NFSA_ECOLI Oxygen-insensitive NADPH nitroreductase OS=Escherichia coli (strai
BAC0611|modC|sp|P09833|MODC_ECOLI Molybdenum import ATP-binding protein ModC OS=Escherichia co
BAC0464|znuB/yebI|sp|P39832|ZNUB_ECOLI High-affinity zinc uptake system membrane protein ZnuB OS=E
BAC0040|baeS|sp|P30847|BAES_ECOLI Signal transduction histidine-protein kinase BaeS OS=Escherichia coli
BAC0270|nikA|sp|P33590|NIKA_ECOLI Nickel-binding periplasmic protein OS=Escherichia coli (strain K12) G
BAC0086|corA|sp|P0ABI4|CORA_ECOLI Magnesium transport protein CorA OS=Escherichia coli (strain K12)
BAC0356|recG|tr|Q9HTL3|Q9HTL3_PSEAE ATP-dependent DNA helicase RecG OS=Pseudomonas aeruginosa
BAC0181|glpF|sp|P0AER0|GLPF_ECOLI Glycerol uptake facilitator protein OS=Escherichia coli (strain K12) G
BAC0092|corT/coaT|tr|H0P0Y3|H0P0Y3_9SYNC Cation-transporting ATPase E1-E2 ATPase OS=Synechocysti
BAC0125|czcR|sp|Q44006|CZCR_RALME Transcriptional activator protein CzcR OS=Ralstonia metallidurans (
BAC0114|cutC|sp|P67826|CUTC_ECOLI Copper homeostasis protein CutC OS=Escherichia coli (strain K12) GN
BAC0447|yjaA|sp|P09162|YJAA_ECOLI Uncharacterized protein YjaA OS=Escherichia coli (strain K12) GN=y
BAC0629|copB|sp|O30085|COPB_ARCFU Copper-exporting P-type ATPase B OS=Archaeoglobus fulgidus (str
BAC0190|hmrR|sp|Q9X5X4|HMRR_SINMW HTH-type transcriptional regulator HmrR OS=Sinorhizobium me
BAC0433|ybtQ|tr|Q9Z375|Q9Z375_YERPE Inner membrane ABC-transporter YbtQ OS=Yersinia pestis GN=yb
BAC0123|czcE|tr|Q1LAJ1|Q1LAJ1_RALME CzcE, involved in Cd(II), Zn(II), Co(II) resistance OS=Ralstonia m
BAC0066|chrF|tr|Q5NUZ7|Q5NUZ7_RALME ChrF, regulatory protein, involved in Chromate resistance OS=Ra
BAC0082|copL|tr|Q5YKV8|Q5YKV8_9XANT CopL OS=Xanthomonas perforans GN=copL PE=4 SV=1
BAC0023|aioS/aoxS|tr|Q2VGB2|Q2VGB2_RHIRD Putative sensor histidine kinase OS=Rhizobium radiobacter
BAC0673|merE|sp|P06690|MERE_PSEAI Uncharacterized mercuric resistance protein MerE OS=Pseudomonas
BAC0688|merR2|tr|Q79B70|Q79B70_PSEST Organomercurial resistance regulatory protein OS=Pseudomonas s
BAC0687|merR|tr|Q79BG7|Q79BG7_PSEST MerR OS=Pseudomonas stutzeri GN=merR PE=4 SV=1
BAC0565|actR|sp|A6UEL7|ACTR_SINMW Acid tolerance regulatory protein ActR OS=Sinorhizobium medicae

BAC0659|merB|sp|P08664|MERB_SERMA Alkylmercury lyase OS=Serratia marcescens GN=merB PE=3 SV=1
BAC0100|ctpG|sp|P63689|CTPG_MYCTU Probable cation-transporting ATPase G OS=Mycobacterium tubercul
BAC0120|czcB|sp|P13510|CZCB_RALME Cobalt-zinc-cadmium resistance protein CzcB OS=Ralstonia metallid
BAC0704|ncrC|tr|Q8VTR4|Q8VTR4_HAFAL NcrC OS=Hafnia alvei GN=ncrC PE=4 SV=1
BAC0035|arsM|tr|Q6N3Y0|Q6N3Y0_RHOPA UbiE/COQ5 methyltransferase OS=Rhodopseudomonas palustris
BAC0231|merP|sp|P13113|MERP_SERMA Mercuric transport protein periplasmic component OS=Serratia marc
BAC0265|ncrC|tr|D5CKG5|D5CKG5_ENTCC Nickel-resistant membrane protein-like protein NcrC OS=Enterob
BAC0631|copC|tr|F4ZBX9|F4ZBX9_XANCI CopC OS=Xanthomonas citri subsp. citri GN=copC PE=4 SV=1
BAC0346|silP|sp|Q9ZHC7|SILP_SALTM Silver exporting P-type ATPase OS=Salmonella typhimurium GN=silI
BAC0111|cusR/ylcA|sp|P0ACZ8|CUSR_ECOLI Transcriptional regulatory protein CusR OS=Escherichia coli (st
BAC0568|actP|sp|Q9X5V3|ATCU_RHILV Copper-transporting P-type ATPase OS=Rhizobium leguminosarum b
BAC0455|ziaA|sp|Q59998|ATZN_SYNY3 Zinc-transporting ATPase OS=Synechocystis sp. (strain PCC 6803 / K
BAC0485|pmrC|tr|Q70FH1|Q70FH1_PECCC Putative cytoplasmic membrane protein pmrC OS=Pectobacterium
BAC0644|corD|sp|Q56017|APAG_SALTY Protein ApaG OS=Salmonella typhimurium (strain LT2 / SGSC1412
BAC0627|copB|tr|F4ZD00|F4ZD00_9XANT Copper resistance protein B OS=Xanthomonas alfalfae subsp. citrun
BAC0003|acn|tr|O53166|O53166_MYCTU Aconitate hydratase OS=Mycobacterium tuberculosis H37Rv GN=R
BAC0318|pstS|sp|P0AG82|PSTS_ECOLI Phosphate-binding protein PstS OS=Escherichia coli (strain K12) GN=
BAC0461|zntA/yhhO|sp|P37617|ATZN_ECOLI Lead, cadmium, zinc and mercury-transporting ATPase OS=Esc
BAC0108|cusB|sp|P77239|CUSB_ECOLI Cation efflux system protein CusB OS=Escherichia coli (strain K12) G
BAC0029|chrF|tr|A4UQR2|A4UQR2_9RHIZ ChrF OS=Ochrobactrum tritici GN=chrF PE=4 SV=1
BAC0022|aioR/aoxR|tr|Q2VGB1|Q2VGB1_RHIRD Putative transcriptional regulator OS=Rhizobium radiobacte
BAC0351|sitC|tr|Q9XCS0|Q9XCS0_SALTM SitC OS=Salmonella typhimurium GN=sitC PE=3 SV=1
BAC0020|aioA/aoxB|sp|Q8GGJ6|AIOA_HERAR Arsenite oxidase subunit AioA OS=Herminiimonas arsenicoxy
BAC0638|copR|tr|C6FFR4|C6FFR4_PSEFL CopR OS=Pseudomonas fluorescens GN=copR PE=4 SV=1
BAC0386|terA|sp|P18778|TERA_ALCSP Tellurium resistance protein TerA OS=Alcaligenes sp. GN=terA PE=4
BAC0578|arsB|tr|O50594|O50594_ACIMU ArsB OS=Acidiphilium multivorum GN=arsB PE=4 SV=1
BAC0388|terC|sp|P18780|TERC_ALCSP Tellurium resistance protein TerC OS=Alcaligenes sp. GN=terC PE=3
BAC0434|ychH|sp|P0AB49|YCHH_ECOLI Uncharacterized protein YchH OS=Escherichia coli (strain K12) GN
BAC0263|ncrA|tr|Q06VT3|Q06VT3_9BACT NcrA OS=Leptospirillum ferriphilum GN=ncrA PE=4 SV=1
BAC0109|cusC/ylcB|sp|P77211|CUSC_ECOLI Cation efflux system protein CusC OS=Escherichia coli (strain K
BAC0633|copD|tr|Q7WYG8|Q7WYG8_PSEPU CopD OS=Pseudomonas putida GN=copD PE=4 SV=1
BAC0563|acrD|tr|Q8ZN77|Q8ZN77_SALTY RND family aminoglycoside/multidrug efflux pump OS=Salmonell
BAC0681|merR2|tr|Q9WWL1|Q9WWL1_BACSR Mercury resistance operon negative regulator MerR2 OS=Bac
BAC0601|tunR|tr|Q72FN4|Q72FN4_DESVH Molybdenum-pterin binding domain protein/site-specific recombina
BAC0582|arsC|sp|P08692|ARSC1_ECOLX Arsenate reductase OS=Escherichia coli GN=arsC PE=1 SV=1
BAC0161|fbpB|sp|P71338|FBPB2_HAEIN Fe(3+)-transport system permease protein FbpB 2 OS=Haemophilus i
BAC0253|mntR|sp|P0A9F1|MNTR_ECOLI Transcriptional regulator MntR OS=Escherichia coli (strain K12) GN
BAC0348|silS|sp|Q9ZHD4|SILS_SALTM Probable sensor kinase SilS OS=Salmonella typhimurium GN=silS PE
BAC0470|zur/yjbK|sp|P0AC51|ZUR_ECOLI Zinc uptake regulation protein OS=Escherichia coli (strain K12) GN
BAC0651|merA|sp|P0A0E5|MERA_STAAU Mercuric reductase OS=Staphylococcus aureus GN=merA PE=3 SV
BAC0179|gesB|tr|Q8ZRG9|Q8ZRG9_SALTY Putative cation efflux system protein OS=Salmonella typhimurium
BAC0285|nreB|tr|F0KND8|F0KND8_ACICP NrsD, nreB nickel permease involved in nickel and cobalt tolerance
BAC0031|arsB|sp|P08691|ARSB1_ECOLX Arsenical pump membrane protein OS=Escherichia coli GN=arsB PE
BAC0352|sitD|tr|Q9XCR9|Q9XCR9_SALTM SitD OS=Salmonella typhimurium GN=sitD PE=3 SV=1
BAC0138|dsbC|sp|P0AEG6|DSBC_ECOLI Thiol:disulfide interchange protein DsbC OS=Escherichia coli (strain
BAC0597|baeS|tr|D0ZNE2|D0ZNE2_SALT1 Signal transduction histidine-protein kinase BaeS OS=Salmonella t

BAC0299|pbrB/pbrC|tr|Q58AJ7|Q58AJ7_RALME Lipoprotein signal peptidase OS=Ralstonia metallidurans (stra
BAC0137|dsbB|sp|P0A6M2|DSBB_ECOLI Disulfide bond formation protein B OS=Escherichia coli (strain K12)
BAC0115|cutE/lnt|sp|P23930|LNT_ECOLI Apolipoprotein N-acyltransferase OS=Escherichia coli (strain K12) G
BAC0030|arsA|sp|P52145|ARSA2_ECOLX Arsenical pump-driving ATPase OS=Escherichia coli GN=arsA PE=
BAC0266|ncrY|tr|Q06VT0|Q06VT0_9BACT NcrY OS=Leptospirillum ferriphilum GN=ncrY PE=4 SV=1
BAC0308|pcoR|sp|Q47456|PCOR_ECOLX Transcriptional regulatory protein PcoR OS=Escherichia coli GN=pc
BAC0121|czcC|sp|P13509|CZCC_RALME Cobalt-zinc-cadmium resistance protein CzcC OS=Ralstonia metallid
BAC0498|ideR|sp|P0A672|IDER_MYCTU Iron-dependent repressor IdeR OS=Mycobacterium tuberculosis GN=
BAC0347|silR|sp|Q9ZHD3|SILR_SALTM Probable transcriptional regulatory protein SilR OS=Salmonella typhi
BAC0105|cueR/ybbI|sp|P0A9G4|CUER_ECOLI HTH-type transcriptional regulator CueR OS=Escherichia coli (
BAC0315|pstA|sp|P07654|PSTA_ECOLI Phosphate transport system permease protein PstA OS=Escherichia col
BAC0357|recG|tr|B5L350|B5L350_9PSED ATP-dependent DNA helicase (Fragment) OS=Pseudomonas corruga
BAC0084|copY/tcrY|sp|Q47839|COPY_ENTHA Transcriptional repressor CopY OS=Enterococcus hirae (strain
BAC0584|arsC|sp|O50595|ARSC_ACIMA Arsenate reductase OS=Acidiphilium multivorum (strain DSM 11245
BAC0224|merA|sp|P16171|MERA_BACCE Mercuric reductase OS=Bacillus cereus GN=merA PE=1 SV=1
BAC0269|nia|tr|Q92Z60|Q92Z60_RHIME Cation transport P-type ATPase OS=Rhizobium meliloti (strain 1021)
BAC0657|merB|tr|O07303|O07303_9PSED Alkylmercury lyase OS=Pseudomonas sp. K-62 GN=merB PE=3 SV
BAC0643|corB|tr|Q9X621|Q9X621_SALTM CorB OS=Salmonella typhimurium GN=corB PE=4 SV=1
BAC0024|aioX/aoxX|tr|G8XNW6|G8XNW6_RHIRD AioX (Fragment) OS=Rhizobium radiobacter GN=aioX P
BAC0341|silA|sp|Q9ZHC9|SILA_SALTM Putative cation efflux system protein SilA OS=Salmonella typhimuri
BAC0334|robA|sp|P0ACI0|ROB_ECOLI Right origin-binding protein OS=Escherichia coli (strain K12) GN=rob
BAC0076|comR/ycfQ|sp|P75952|COMR_ECOLI HTH-type transcriptional repressor ComR OS=Escherichia coli
BAC0387|terB|sp|P18779|TERB_ALCSP Tellurium resistance protein TerB OS=Alcaligenes sp. GN=terB PE=4
BAC0371|soxS|sp|P0A9E2|SOXS_ECOLI Regulatory protein SoxS OS=Escherichia coli (strain K12) GN=soxS
BAC0619|copA|tr|Q7WYH1|Q7WYH1_PSEPU CopA OS=Pseudomonas putida GN=copA PE=4 SV=1
BAC0057|cadD|tr|Q7A320|Q7A320_STAAN CadD OS=Staphylococcus aureus (strain N315) GN=cadD PE=4 S
BAC0048|bfrA|sp|P63697|BFR_MYCTU Bacterioferritin OS=Mycobacterium tuberculosis GN=bfr PE=1 SV=1
BAC0698|ncrA|tr|Q1KLR2|Q1KLR2_SERMA NcrA OS=Serratia marcescens GN=ncrA PE=4 SV=1
BAC0467|zraR/hydH|sp|P14375|ZRAR_ECOLI Transcriptional regulatory protein ZraR OS=Escherichia coli (str
BAC0107|cusA/ybdE|sp|P38054|CUSA_ECOLI Cation efflux system protein CusA OS=Escherichia coli (strain
BAC0026|chrA|tr|A4UQR4|A4UQR4_9RHIZ Chromate transporter OS=Ochrobactrum tritici GN=chrA PE=4 S
BAC0689|merR|tr|Q934S8|Q934S8_THIFE Mer operon regulatory protein OS=Thiobacillus ferrooxidans GN=m
BAC0649|merA|tr|E3VST6|E3VST6_9FLAO MerA OS=Tenacibaculum discolor GN=merA PE=3 SV=1
BAC0385|tehB|sp|P25397|TEHB_ECOLI Tellurite methyltransferase OS=Escherichia coli (strain K12) GN=tehB
BAC0081|copK|sp|Q58AD3|COPK_RALME Copper resistance protein K OS=Ralstonia metallidurans (strain CH
BAC0692|merT|tr|Q79BG6|Q79BG6_PSEST MerT OS=Pseudomonas stutzeri GN=merT PE=4 SV=1
BAC0572|arsA|sp|P08690|ARSA1_ECOLX Arsenical pump-driving ATPase OS=Escherichia coli GN=arsA PE=
BAC0028|chrC|tr|A4UQR3|A4UQR3_9RHIZ Superoxide dismutase OS=Ochrobactrum tritici GN=chrC PE=3 S
BAC0126|czcS|sp|Q44007|CZCS_RALME Sensor protein CzcS OS=Ralstonia metallidurans (strain CH34 / ATC
BAC0488|pmrB|sp|Q70FG9|PMRB_PECSS Sensor histidine kinase PmrB OS=Pectobacterium sp. (strain SCC31
BAC0656|merB3|tr|Q7DHE7|Q7DHE7_BACCE Organomercurial lyase enzyme OS=Bacillus cereus GN=merB3
BAC0645|mdtA|tr|D0ZND8|D0ZND8_SALT1 Multidrug resistance protein MdtA OS=Salmonella typhimurium
BAC0441|yfeC|sp|Q56954|YFEC_YERPE Chelated iron transport system membrane protein YfeC OS=Yersinia
BAC0690|merT|tr|Q934S7|Q934S7_THIFE Mercuric ion transport protein OS=Thiobacillus ferrooxidans GN=m
BAC0463|znuA/yebL|sp|P39172|ZNUA_ECOLI High-affinity zinc uptake system protein ZnuA OS=Escherichia
BAC0183|golT|tr|Q8ZRG7|Q8ZRG7_SALTY Putative cation transport ATPase OS=Salmonella typhimurium (st

BAC0585|arsC|sp|P74984|ARSC_YEREN Arsenate reductase OS=Yersinia enterocolitica GN=arsC PE=3 SV=1
BAC0301|pbrR|tr|D8IQ72|D8IQ72_HERSS Pb-specific transcription regulator protein OS=Herbaspirillum serope
BAC0251|mntH/yfeP|sp|P0A769|MNTH_ECOLI Divalent metal cation transporter MntH OS=Escherichia coli (s
BAC0119|czcA|sp|P13511|CZCA_RALME Cobalt-zinc-cadmium resistance protein CzcA OS=Ralstonia metallic
BAC0691|merT|tr|Q52397|Q52397_PSEST Mercury transport protein OS=Pseudomonas stutzeri GN=merT PE=
BAC0264|ncrB|tr|Q06VT2|Q06VT2_9BACT NcrB OS=Leptospirillum ferriphilum GN=ncrB PE=4 SV=1
BAC0064|chrB1|sp|P17552|CHRB1_RALME Protein ChrB OS=Ralstonia metallidurans (strain CH34 / ATCC 4
BAC0587|arsD|sp|P52148|ARSD2_ECOLX Arsenical resistance operon trans-acting repressor ArsD OS=Escheri
BAC0642|mgtA|sp|P36640|ATMA_SALTY Magnesium-transporting ATPase, P-type 1 OS=Salmonella typhimu
BAC0302|pbrT|tr|Q5GR69|Q5GR69_ALCXX Lead uptake protein PbrT OS=Alcaligenes xylosoxydans xylosoxy
BAC0675|merP|tr|O07301|O07301_9PSED Mercuric transport protein periplasmic component OS=Pseudomonas
BAC0106|cuiD|sp|Q8ZRS2|CUEO_SALTY Blue copper oxidase CueO OS=Salmonella typhimurium (strain LT2
BAC0647|mdtC|tr|D0ZNE0|D0ZNE0_SALT1 Multidrug resistance protein MdtC OS=Salmonella typhimurium (
BAC0072|cnrC|sp|P37974|CNRC_RALME Nickel and cobalt resistance protein CnrC OS=Ralstonia metallidurar
BAC0451|yodD|sp|P64519|YODD_ECOLI Uncharacterized protein YodD OS=Escherichia coli (strain K12) GN=
BAC0541|yieF|sp|P0AGE6|YIEF_ECOLI Uncharacterized protein YieF OS=Escherichia coli (strain K12) GN=y
BAC0039|baeR|sp|P69228|BAER_ECOLI Transcriptional regulatory protein BaeR OS=Escherichia coli (strain K
BAC0462|zntR/yhdM|sp|P0ACS5|ZNTR_ECOLI HTH-type transcriptional regulator ZntR OS=Escherichia coli (
BAC0610|modB|sp|P0AF01|MODB_ECOLI Molybdenum transport system permease protein ModB OS=Escheri
BAC0490|G2alt|tr|B0FSM1|B0FSM1_9BACI 7-cyano-7-deazaguanine synthase OS=Anoxybacillus gonensis GN
BAC0678|merP|tr|O66016|O66016_PSEST MerP OS=Pseudomonas stutzeri GN=merP PE=4 SV=1
BAC0342|silB|sp|Q9ZHD0|SILB_SALTM Putative membrane fusion protein SilB OS=Salmonella typhimurium
BAC0163|fecD|sp|P15029|FECD_ECOLI Fe(3+) dicitrate transport system permease protein FecD OS=Escherich
BAC0390|terE|sp|P18782|TERE_ALCSP Tellurium resistance protein TerE OS=Alcaligenes sp. GN=terE PE=3 S
BAC0682|merR1|sp|P22853|MERR_BACCE Mercuric resistance operon regulatory protein OS=Bacillus cereus
BAC0609|modA|sp|P37329|MODA_ECOLI Molybdate-binding periplasmic protein OS=Escherichia coli (strain
BAC0577|arsB|sp|P74985|ARSB_YEREN Arsenical pump membrane protein OS=Yersinia enterocolitica GN=ar
BAC0599|modB|tr|Q72FN2|Q72FN2_DESVH Molybdenum ABC transporter, permease protein OS=Desulfovib
BAC0162|fbpC|sp|P44513|FBPC2_HAEIN Fe(3+) ions import ATP-binding protein FbpC 2 OS=Haemophilus in
BAC0579|arsB|sp|P74311|Y944_SYNY3 Uncharacterized transporter slr0944 OS=Synechocystis sp. (strain PCC
BAC0548|chrA1|sp|P17551|CHRA1_RALME Chromate transport protein OS=Ralstonia metallidurans (strain CH
BAC0459|zitB/ybgR|sp|P75757|ZITB_ECOLI Zinc transporter ZitB OS=Escherichia coli (strain K12) GN=zitB
BAC0676|merP|tr|Q7DHE4|Q7DHE4_BACCE Mercury-binding protein OS=Bacillus cereus GN=merP PE=4 S
BAC0027|chrB|tr|A4UQR5|A4UQR5_9RHIZ ChrB OS=Ochrobactrum tritici GN=chrB PE=4 SV=1
BAC0628|copB|tr|F4ZBX4|F4ZBX4_XANCI CopB OS=Xanthomonas citri subsp. citri GN=copB PE=4 SV=1
BAC0679|merP|tr|O66047|O66047_PSEST Mercury transport protein OS=Pseudomonas stutzeri GN=merP PE=
BAC0164|fecE|sp|P15031|FECE_ECOLI Fe(3+) dicitrate transport ATP-binding protein FecE OS=Escherichia c
BAC0695|merT|sp|P13112|MERT_SERMA Mercuric transport protein OS=Serratia marcescens GN=merT PE=4
BAC0355|ruvB|sp|Q51426|RUVB_PSEAE Holliday junction ATP-dependent DNA helicase RuvB OS=Pseudom
BAC0332|rcnR/yohL|sp|P64530|RCNR_ECOLI Transcriptional repressor RcnR OS=Escherichia coli (strain K12
BAC0165|fetA/ybbL|sp|P77279|YBBL_ECOLI Uncharacterized ABC transporter ATP-binding protein YbbL OS
BAC0073|cnrH|sp|P37978|CNRH_RALME RNA polymerase sigma factor CnrH OS=Ralstonia metallidurans (st
BAC0666|merD|tr|O66018|O66018_PSEST MerD OS=Pseudomonas stutzeri GN=merD PE=4 SV=1
BAC0033|arsD|sp|P46003|ARSD1_ECOLX Arsenical resistance operon trans-acting repressor ArsD OS=Escheri
BAC0358|oscA|tr|B6CM35|B6CM35_9PSED Putative uncharacterized protein oscA OS=Pseudomonas corrugata
BAC0686|merR|sp|P13111|MERR_SERMA Mercuric resistance operon regulatory protein OS=Serratia marcesce

BAC0305|pcoC|sp|Q47454|PCOC_ECOLX Copper resistance protein C OS=Escherichia coli GN=pcoC PE=1 S
BAC0049|bhsA/ycfR/comC|sp|P0AB40|BHSA_ECOLI Multiple stress resistance protein BhsA OS=Escherichia c
BAC0276|nirA|tr|Q6RUG3|Q6RUG3_KLEOX NirA OS=Klebsiella oxytoca GN=nirA PE=4 SV=1
BAC0130|czrA|tr|Q9RLI8|Q9RLI8_PSEAI CzrA protein OS=Pseudomonas aeruginosa GN=czrA PE=4 SV=1
BAC0306|pcoD|sp|Q47455|PCOD_ECOLX Copper resistance protein D OS=Escherichia coli GN=pcoD PE=3 S
BAC0625|copA|tr|F4ZCZ9|F4ZCZ9_9XANT Copper resistance protein A OS=Xanthomonas alfalfae subsp. citru
BAC0566|actS|tr|Q52912|Q52912_9RHIZ Histidine protein kinase OS=Sinorhizobium medicae GN=actS PE=4 S
BAC0298|pbrA|tr|Q58AJ6|Q58AJ6_RALME P-type ATPase involved in Pb(II) resistance PbrA OS=Ralstonia m
BAC0166|fetB/ybbM|sp|P77307|YBBM_ECOLI UPF0014 inner membrane protein YbbM OS=Escherichia coli (
BAC0683|merR1|tr|O07300|O07300_9PSED Mercuric resistance operon regulatory protein OS=Pseudomonas sp
BAC0330|rcnA/yohM|sp|P76425|RCNA_ECOLI Nickel/cobalt efflux system RcnA OS=Escherichia coli (strain
BAC0707|sodB|sp|P0AGD3|SODF_ECOLI Superoxide dismutase [Fe] OS=Escherichia coli (strain K12) GN=so
BAC0059|cadX|tr|A7LHQ4|A7LHQ4_STRSL CadX OS=Streptococcus salivarius GN=cadX PE=4 SV=1
BAC0058|cadR|tr|Q93TP7|Q93TP7_PSEPU CadR OS=Pseudomonas putida GN=cadR PE=4 SV=1
BAC0317|pstC|sp|P0AGH8|PSTC_ECOLI Phosphate transport system permease protein PstC OS=Escherichia co
BAC0466|zraP|sp|Q9L9I0|ZRAP_SALTY Zinc resistance-associated protein OS=Salmonella typhimurium (strai
BAC0331|rcnB/yohN|sp|P64534|RCNB_ECOLI Nickel/cobalt homeostasis protein RcnB OS=Escherichia coli (s
BAC0273|nikD|sp|P33593|NIKD_ECOLI Nickel import ATP-binding protein NikD OS=Escherichia coli (strain
BAC0079|copB|sp|P05425|COPB_ENTHA Copper-exporting P-type ATPase B OS=Enterococcus hirae (strain A
BAC0344|silE|sp|Q9Z4N3|SILE_SALTM Silver-binding protein SilE OS=Salmonella typhimurium GN=silE PE
BAC0668|merD|tr|Q5NUV1|Q5NUV1_RALME MerD from Tn4378, regulatory protein involved in Hg(II) resist
BAC0432|ybtP|tr|Q9R7V3|Q9R7V3_YERPE Lipoprotein inner membrane ABC-transporter OS=Yersinia pestis
BAC0469|zupT/ygiE|sp|P0A8H3|ZUPT_ECOLI Zinc transporter ZupT OS=Escherichia coli (strain K12) GN=zu
BAC0665|merD|tr|O66022|O66022_PSEST Mercury operon coregulator protein OS=Pseudomonas stutzeri GN=
BAC0457|zinT/yodA|sp|P76344|ZINT_ECOLI Metal-binding protein ZinT OS=Escherichia coli (strain K12) GN
BAC0702|ncrA|tr|Q8VTR6|Q8VTR6_HAFAL NcrA OS=Hafnia alvei GN=ncrA PE=4 SV=1
BAC0576|arsB|sp|P52146|ARSB2_ECOLX Arsenical pump membrane protein OS=Escherichia coli GN=arsB P
BAC0446|yhcN|sp|P64614|YHCN_ECOLI Uncharacterized protein YhcN OS=Escherichia coli (strain K12) GN=
BAC0304|pcoB|sp|Q47453|PCOB_ECOLX Copper resistance protein B OS=Escherichia coli GN=pcoB PE=4 S
BAC0608|modE|sp|P0A9G8|MODE_ECOLI Transcriptional regulator ModE OS=Escherichia coli (strain K12) G
BAC0203|cnrA|sp|P37972|CNRA_RALME Nickel and cobalt resistance protein CnrA OS=Ralstonia metallidura
BAC0316|pstB|sp|P0AAH0|PSTB_ECOLI Phosphate import ATP-binding protein PstB OS=Escherichia coli (str
BAC0167|fieF/yiip|sp|P69380|FIEF_ECOLI Ferrous-iron efflux pump FieF OS=Escherichia coli (strain K12) GN
BAC0384|tehA|sp|P25396|TEHA_ECOLI Tellurite resistance protein TehA OS=Escherichia coli (strain K12) G
BAC0703|ncrB|tr|Q8VTR5|Q8VTR5_HAFAL NcrB OS=Hafnia alvei GN=ncrB PE=4 SV=1
BAC0653|merA|tr|Q934S5|Q934S5_THIFE Mercuric ion reductase OS=Thiobacillus ferrooxidans GN=merA PE
BAC0452|yqjH|sp|Q46871|YQJH_ECOLI NADPH-dependent ferric-chelate reductase OS=Escherichia coli (strai
BAC0135|dpsA|tr|Q8KR86|Q8KR86_BURPE DpsA OS=Burkholderia pseudomallei GN=dpsA PE=3 SV=1
BAC0279|nirD|tr|Q6RUG0|Q6RUG0_KLEOX NirD OS=Klebsiella oxytoca GN=nirD PE=4 SV=1
BAC0620|copA|sp|P32113|COPA_ENTHA Probable copper-importing P-type ATPase A OS=Enterococcus hira
BAC0662|merB3|tr|Q9RHR0|Q9RHR0_BACME MerB3 OS=Bacillus megaterium GN=merB3 PE=4 SV=2
BAC0133|dnaK|sp|P0A5B9|DNAK_MYCTU Chaperone protein DnaK OS=Mycobacterium tuberculosis GN=dn
BAC0588|arsR|tr|P74986|P74986_YEREN Arsenite inducible repressor OS=Yersinia enterocolitica GN=arsR PE
BAC0693|merT|tr|Q79F00|Q79F00_9PSED Mercuric transport protein OS=Pseudomonas sp. K-62 GN=merT PE
BAC0573|arsB|sp|P45946|ARSB_BACSU Arsenite resistance protein ArsB OS=Bacillus subtilis (strain 168) GN
BAC0077|copA|sp|P12374|COPA_PSEUB Copper resistance protein A OS=Pseudomonas syringae pv. tomato G

BAC0240|mexI|tr|Q9HWH4|Q9HWH4_PSEAE Probable Resistance-Nodulation-Cell Division (RND) efflux tran
BAC0670|merE|tr|Q52104|Q52104_9ZZZZ Uncharacterized protein OS=Plasmid pDU1358 PE=4 SV=1
BAC0589|arsR|sp|P15905|ARSR1_ECOLX Arsenical resistance operon repressor OS=Escherichia coli GN=arsR
BAC0368|sodA|sp|P00448|SODM_ECOLI Superoxide dismutase [Mn] OS=Escherichia coli (strain K12) GN=so
BAC0700|ncrC|tr|Q1KLR0|Q1KLR0_SERMA NcrC OS=Serratia marcescens GN=ncrC PE=4 SV=1
BAC0267|nczA|tr|B8GZE9|B8GZE9_CAUCN Cobalt-zinc-cadmium resistance protein czcA OS=Caulobacter cr
BAC0136|dsbA|sp|P0AEG4|DSBA_ECOLI Thiol:disulfide interchange protein DsbA OS=Escherichia coli (strai
BAC0271|nikB|sp|P33591|NIKB_ECOLI Nickel transport system permease protein NikB OS=Escherichia coli (s
BAC0538|chrR|tr|Q7BD45|Q7BD45_PSEPU Chromate reductase OS=Pseudomonas putida GN=chrR PE=4 SV=
BAC0465|znuC/yebM|sp|P0A9X1|ZNUC_ECOLI Zinc import ATP-binding protein ZnuC OS=Escherichia coli (
BAC0087|mgtA|sp|P0ABB8|ATMA_ECOLI Magnesium-transporting ATPase, P-type 1 OS=Escherichia coli (st
BAC0392|terZ|sp|Q52353|TERZ_SERMA Tellurium resistance protein TerZ OS=Serratia marcescens GN=terZ P
BAC0349|sitA|tr|Q9XCS2|Q9XCS2_SALTI Iron transport protein, periplasmic-binding protein OS=Salmonella t
BAC0204|cnrB|sp|P37973|CNRB_RALME Nickel and cobalt resistance protein CnrB OS=Ralstonia metallidura
BAC0343|silC|sp|Q9ZHD2|SILC_SALTM Probable outer membrane lipoprotein SilC OS=Salmonella typhimuri
BAC0630|copC|sp|P12376|COPC_PSEUB Copper resistance protein C OS=Pseudomonas syringae pv. tomato G
BAC0391|terW|sp|P75010|TERW_SERMA Tellurium resistance protein TerW OS=Serratia marcescens GN=ter
BAC0303|pcoA|sp|Q47452|PCOA_ECOLX Copper resistance protein A OS=Escherichia coli GN=pcoA PE=3 S
BAC0684|merR2|tr|Q7DKL2|Q7DKL2_9PSED MerR2 OS=Pseudomonas sp. K-62 GN=merR2 PE=4 SV=1
BAC0024|aioX/aoxX|tr|G8XNW6|G8XNW6_RHIRD AioX (Fragment) OS=Rhizobium radiobacter GN=aioX P
BAC0203|cnrA|sp|P37972|CNRA_RALME Nickel and cobalt resistance protein CnrA OS=Ralstonia metallidura
BAC0685|merR|tr|H6WCN2|H6WCN2_9FLAO MerR OS=Tenacibaculum discolor GN=merR PE=4 SV=1
BAC0700|ncrC|tr|Q1KLR0|Q1KLR0_SERMA NcrC OS=Serratia marcescens GN=ncrC PE=4 SV=1
BAC0539|chrR|sp|P96977|CHRR_PSEUG CR(VI) reductase OS=Pseudomonas sp. (strain G-1) GN=chrR PE=3
BAC0315|pstA|sp|P07654|PSTA_ECOLI Phosphate transport system permease protein PstA OS=Escherichia col
BAC0316|pstB|sp|P0AAH0|PSTB_ECOLI Phosphate import ATP-binding protein PstB OS=Escherichia coli (str
BAC0076|comR/ycfQ|sp|P75952|COMR_ECOLI HTH-type transcriptional repressor ComR OS=Escherichia coli
BAC0134|dpr/dps|sp|P0CB53|DPS_STRSU DNA protection during starvation protein OS=Streptococcus suis G
BAC0368|sodA|sp|P00448|SODM_ECOLI Superoxide dismutase [Mn] OS=Escherichia coli (strain K12) GN=so
BAC0390|terE|sp|P18782|TERE_ALCSP Tellurium resistance protein TerE OS=Alcaligenes sp. GN=terE PE=3 S
BAC0252|mntP/yebN|sp|P76264|MNTP_ECOLI Probable manganese efflux pump MntP OS=Escherichia coli (st
BAC0699|ncrB|tr|Q1KLR1|Q1KLR1_SERMA NcrB OS=Serratia marcescens GN=ncrB PE=4 SV=1
BAC0490|G2alt|tr|B0FSM1|B0FSM1_9BACI 7-cyano-7-deazaguanine synthase OS=Anoxybacillus gonensis G
BAC0459|zitB/ybgR|sp|P75757|ZITB_ECOLI Zinc transporter ZitB OS=Escherichia coli (strain K12) GN=zitB
BAC0466|zraP|sp|Q9L9I0|ZRAP_SALTY Zinc resistance-associated protein OS=Salmonella typhimurium (strai
BAC0654|merB1|sp|P16172|MERB_BACCE Alkylmercury lyase OS=Bacillus cereus GN=merB1 PE=3 SV=2
BAC0577|arsB|sp|P74985|ARSB_YEREN Arsenical pump membrane protein OS=Yersinia enterocolitica GN=a
BAC0278|nirC|tr|Q6RUG1|Q6RUG1_KLEOX NirC OS=Klebsiella oxytoca GN=nirC PE=4 SV=1
BAC0641|corA|sp|P0A2R8|CORA_SALTY Magnesium transport protein CorA OS=Salmonella typhimurium (st
BAC0645|mdtA|tr|D0ZND8|D0ZND8_SALT1 Multidrug resistance protein MdtA OS=Salmonella typhimurium
BAC0630|copC|sp|P12376|COPC_PSEUB Copper resistance protein C OS=Pseudomonas syringae pv. tomato G
BAC0627|copB|tr|F4ZD00|F4ZD00_9XANT Copper resistance protein B OS=Xanthomonas alfalfae subsp. citru
BAC0122|czcD|sp|P13512|CZCD_RALME Cobalt-zinc-cadmium resistance protein CzcD OS=Ralstonia metalli
BAC0161|fbpB|sp|P71338|FBPB2_HAEIN Fe(3+)-transport system permease protein FbpB 2 OS=Haemophilus
BAC0167|fieF/yiip|sp|P69380|FIEF_ECOLI Ferrous-iron efflux pump FieF OS=Escherichia coli (strain K12) G
BAC0679|merP|tr|O66047|O66047_PSEST Mercury transport protein OS=Pseudomonas stutzeri GN=merP PE=

BAC0342|silB|sp|Q9ZHD0|SILB_SALTM Putative membrane fusion protein SilB OS=Salmonella typhimurium
BAC0121|czcC|sp|P13509|CZCC_RALME Cobalt-zinc-cadmium resistance protein CzcC OS=Ralstonia metallid
BAC0626|copB|sp|P12375|COPB_PSEUB Copper resistance protein B OS=Pseudomonas syringae pv. tomato G
BAC0681|merR2|tr|Q9WWL1|Q9WWL1_BACSR Mercury resistance operon negative regulator MerR2 OS=Bac
BAC0665|merD|tr|O66022|O66022_PSEST Mercury operon coregulator protein OS=Pseudomonas stutzeri GN=
BAC0349|sitA|tr|Q9XCS2|Q9XCS2_SALTI Iron transport protein, periplasmic-binding protein OS=Salmonella t
BAC0584|arsC|sp|O50595|ARSC_ACIMA Arsenate reductase OS=Acidiphilium multivorum (strain DSM 11245
BAC0690|merT|tr|Q934S7|Q934S7_THIFE Mercuric ion transport protein OS=Thiobacillus ferrooxidans GN=m
BAC0130|czrA|tr|Q9RLI8|Q9RLI8_PSEAI CzrA protein OS=Pseudomonas aeruginosa GN=czrA PE=4 SV=1
BAC0023|aioS/aoxS|tr|Q2VGB2|Q2VGB2_RHIRD Putative sensor histidine kinase OS=Rhizobium radiobacter
BAC0357|recG|tr|B5L350|B5L350_9PSED ATP-dependent DNA helicase (Fragment) OS=Pseudomonas corruga
BAC0224|merA|sp|P16171|MERA_BACCE Mercuric reductase OS=Bacillus cereus GN=merA PE=1 SV=1
BAC0355|ruvB|sp|Q51426|RUVB_PSEAE Holliday junction ATP-dependent DNA helicase RuvB OS=Pseudom
BAC0350|sitB|tr|Q9XCS1|Q9XCS1_SALTM SitB OS=Salmonella typhimurium GN=sitB PE=3 SV=1
BAC0388|terC|sp|P18780|TERC_ALCSP Tellurium resistance protein TerC OS=Alcaligenes sp. GN=terC PE=3
BAC0110|cusF/cusX|sp|P77214|CUSF_ECOLI Cation efflux system protein CusF OS=Escherichia coli (strain K
BAC0276|nirA|tr|Q6RUG3|Q6RUG3_KLEOX NirA OS=Klebsiella oxytoca GN=nirA PE=4 SV=1
BAC0498|ideR|sp|P0A672|IDER_MYCTU Iron-dependent repressor IdeR OS=Mycobacterium tuberculosis GN=
BAC0056|cadC|sp|P20047|CADC_STAAU Cadmium resistance transcriptional regulatory protein CadC OS=Sta
BAC0625|copA|tr|F4ZCZ9|F4ZCZ9_9XANT Copper resistance protein A OS=Xanthomonas alfalfae subsp. citru
BAC0668|merD|tr|Q5NUV1|Q5NUV1_RALME MerD from Tn4378, regulatory protein involved in Hg(II) resist
BAC0675|merP|tr|O07301|O07301_9PSED Mercuric transport protein periplasmic component OS=Pseudomonas
BAC0100|ctpG|sp|P63689|CTPG_MYCTU Probable cation-transporting ATPase G OS=Mycobacterium tubercul
BAC0548|chrA1|sp|P17551|CHRA1_RALME Chromate transport protein OS=Ralstonia metallidurans (strain CH
BAC0101|ctpV|sp|P77894|CTPV_MYCTU Probable copper-exporting P-type ATPase V OS=Mycobacterium tub
BAC0608|modE|sp|P0A9G8|MODE_ECOLI Transcriptional regulator ModE OS=Escherichia coli (strain K12) G
BAC0079|copB|sp|P05425|COPB_ENTHA Copper-exporting P-type ATPase B OS=Enterococcus hirae (strain A
BAC0114|cutC|sp|P67826|CUTC_ECOLI Copper homeostasis protein CutC OS=Escherichia coli (strain K12) G
BAC0341|silA|sp|Q9ZHC9|SILA_SALTM Putative cation efflux system protein SilA OS=Salmonella typhimuriu
BAC0666|merD|tr|O66018|O66018_PSEST MerD OS=Pseudomonas stutzeri GN=merD PE=4 SV=1
BAC0649|merA|tr|E3VST6|E3VST6_9FLAO MerA OS=Tenacibaculum discolor GN=merA PE=3 SV=1
BAC0642|mgtA|sp|P36640|ATMA_SALTY Magnesium-transporting ATPase, P-type 1 OS=Salmonella typhimu
BAC0285|nreB|tr|F0KND8|F0KND8_ACICP NrsD, nreB nickel permease involved in nickel and cobalt toleranc
BAC0087|mgtA|sp|P0ABB8|ATMA_ECOLI Magnesium-transporting ATPase, P-type 1 OS=Escherichia coli (st
BAC0464|znuB/yebI|sp|P39832|ZNUB_ECOLI High-affinity zinc uptake system membrane protein ZnuB OS=E
BAC0240|mexI|tr|Q9HWH4|Q9HWH4_PSEAE Probable Resistance-Nodulation-Cell Division (RND) efflux tra
BAC0589|arsR|sp|P15905|ARSR1_ECOLX Arsenical resistance operon repressor OS=Escherichia coli GN=arsR
BAC0264|ncrB|tr|Q06VT2|Q06VT2_9BACT NcrB OS=Leptospirillum ferriphilum GN=ncrB PE=4 SV=1
BAC0352|sitD|tr|Q9XCR9|Q9XCR9_SALTM SitD OS=Salmonella typhimurium GN=sitD PE=3 SV=1
BAC0568|actP|sp|Q9X5V3|ATCU_RHILV Copper-transporting P-type ATPase OS=Rhizobium leguminosarum
BAC0656|merB3|tr|Q7DHE7|Q7DHE7_BACCE Organomercurial lyase enzyme OS=Bacillus cereus GN=merB3
BAC0389|terD|sp|P18781|TERD_ALCSP Tellurium resistance protein TerD OS=Alcaligenes sp. GN=terD PE=3
BAC0455|ziaA|sp|Q59998|ATZN_SYNY3 Zinc-transporting ATPase OS=Synechocystis sp. (strain PCC 6803 /
BAC0135|dpsA|tr|Q8KR86|Q8KR86_BURPE DpsA OS=Burkholderia pseudomallei GN=dpsA PE=3 SV=1
BAC0030|arsA|sp|P52145|ARSA2_ECOLX Arsenical pump-driving ATPase OS=Escherichia coli GN=arsA PE=
BAC0273|nikD|sp|P33593|NIKD_ECOLI Nickel import ATP-binding protein NikD OS=Escherichia coli (strain

BAC0585|arsC|sp|P74984|ARSC_YEREN Arsenate reductase OS=Yersinia enterocolitica GN=arsC PE=3 SV=1
BAC0231|merP|sp|P13113|MERP_SERMA Mercuric transport protein periplasmic component OS=Serratia marc
BAC0702|ncrA|tr|Q8VTR6|Q8VTR6_HAFAL NcrA OS=Hafnia alvei GN=ncrA PE=4 SV=1
BAC0266|ncrY|tr|Q06VT0|Q06VT0_9BACT NcrY OS=Leptospirillum ferriphilum GN=ncrY PE=4 SV=1
BAC0181|glpF|sp|P0AER0|GLPF_ECOLI Glycerol uptake facilitator protein OS=Escherichia coli (strain K12) G
BAC0077|copA|sp|P12374|COPA_PSEUB Copper resistance protein A OS=Pseudomonas syringae pv. tomato G
BAC0638|copR|tr|C6FFR4|C6FFR4_PSEFL CopR OS=Pseudomonas fluorescens GN=copR PE=4 SV=1
BAC0063|chrA|sp|P14285|CHRA_PSEAI Chromate transport protein OS=Pseudomonas aeruginosa GN=chrA P
BAC0330|rcnA/yohM|sp|P76425|RCNA_ECOLI Nickel/cobalt efflux system RcnA OS=Escherichia coli (strain K
BAC0164|fecE|sp|P15031|FECE_ECOLI Fe(3+) dicitrate transport ATP-binding protein FecE OS=Escherichia co
BAC0345|silF|tr|Q9ZHD1|Q9ZHD1_SALTM Uncharacterized protein OS=Salmonella typhimurium GN=ORF96
BAC0179|gesB|tr|Q8ZRG9|Q8ZRG9_SALTY Putative cation efflux system protein OS=Salmonella typhimurium
BAC0470|zur/yjbK|sp|P0AC51|ZUR_ECOLI Zinc uptake regulation protein OS=Escherichia coli (strain K12) GN
BAC0703|ncrB|tr|Q8VTR5|Q8VTR5_HAFAL NcrB OS=Hafnia alvei GN=ncrB PE=4 SV=1
BAC0432|ybtP|tr|Q9R7V3|Q9R7V3_YERPE Lipoprotein inner membrane ABC-transporter OS=Yersinia pestis G
BAC0434|ychH|sp|P0AB49|YCHH_ECOLI Uncharacterized protein YchH OS=Escherichia coli (strain K12) GN
BAC0098|ctpC|sp|P0A502|CTPC_MYCTU Probable manganese/zinc-exporting P-type ATPase OS=Mycobacteri
BAC0540|nfsA|sp|P17117|NFSA_ECOLI Oxygen-insensitive NADPH nitroreductase OS=Escherichia coli (strain
BAC0302|pbrT|tr|Q5GR69|Q5GR69_ALCXX Lead uptake protein PbrT OS=Alcaligenes xylosoxydans xylosoxy
BAC0451|yodD|sp|P64519|YODD_ECOLI Uncharacterized protein YodD OS=Escherichia coli (strain K12) GN=
BAC0640|copD|tr|C6FFR7|C6FFR7_PSEFL CopD OS=Pseudomonas fluorescens GN=copD PE=4 SV=1
BAC0263|ncrA|tr|Q06VT3|Q06VT3_9BACT NcrA OS=Leptospirillum ferriphilum GN=ncrA PE=4 SV=1
BAC0620|copA|sp|P32113|COPA_ENTHA Probable copper-importing P-type ATPase A OS=Enterococcus hirae
BAC0113|cutA|sp|P69488|CUTA_ECOLI Divalent-cation tolerance protein CutA OS=Escherichia coli (strain K1
BAC0452|yqjH|sp|Q46871|YQJH_ECOLI NADPH-dependent ferric-chelate reductase OS=Escherichia coli (strai
BAC0346|silP|sp|Q9ZHC7|SILP_SALTM Silver exporting P-type ATPase OS=Salmonella typhimurium GN=silP
BAC0270|nikA|sp|P33590|NIKA_ECOLI Nickel-binding periplasmic protein OS=Escherichia coli (strain K12) G
BAC0034|arsH|tr|E8PS81|E8PS81_YERPE Arsenic resistance protein ArsH OS=Yersinia pestis Java 9 GN=arsH
BAC0136|dsbA|sp|P0AEG4|DSBA_ECOLI Thiol:disulfide interchange protein DsbA OS=Escherichia coli (strain
BAC0384|tehA|sp|P25396|TEHA_ECOLI Tellurite resistance protein TehA OS=Escherichia coli (strain K12) GN
BAC0088|corC|sp|P0A2L3|CORC_SALTY Magnesium and cobalt efflux protein CorC OS=Salmonella typhimur
BAC0670|merE|tr|Q52104|Q52104_9ZZZZ Uncharacterized protein OS=Plasmid pDU1358 PE=4 SV=1
BAC0684|merR2|tr|Q7DKL2|Q7DKL2_9PSED MerR2 OS=Pseudomonas sp. K-62 GN=merR2 PE=4 SV=1
BAC0573|arsB|sp|P45946|ARSB_BACSU Arsenite resistance protein ArsB OS=Bacillus subtilis (strain 168) GN
BAC0107|cusA/ybdE|sp|P38054|CUSA_ECOLI Cation efflux system protein CusA OS=Escherichia coli (strain K
BAC0582|arsC|sp|P08692|ARSC1_ECOLX Arsenate reductase OS=Escherichia coli GN=arsC PE=1 SV=1
BAC0563|acrD|tr|Q8ZN77|Q8ZN77_SALTY RND family aminoglycoside/multidrug efflux pump OS=Salmonell
BAC0108|cusB|sp|P77239|CUSB_ECOLI Cation efflux system protein CusB OS=Escherichia coli (strain K12) G
BAC0691|merT|tr|Q52397|Q52397_PSEST Mercury transport protein OS=Pseudomonas stutzeri GN=merT PE=4
BAC0571|arsA|sp|O50593|ARSA_ACIMA Arsenical pump-driving ATPase OS=Acidiphilium multivorum (strain
BAC0347|silR|sp|Q9ZHD3|SILR_SALTM Probable transcriptional regulatory protein SilR OS=Salmonella typhi
BAC0657|merB|tr|O07303|O07303_9PSED Alkylmercury lyase OS=Pseudomonas sp. K-62 GN=merB PE=3 SV
BAC0137|dsbB|sp|P0A6M2|DSBB_ECOLI Disulfide bond formation protein B OS=Escherichia coli (strain K12)
BAC0565|actR|sp|A6UEL7|ACTR_SINMW Acid tolerance regulatory protein ActR OS=Sinorhizobium medicae
BAC0387|terB|sp|P18779|TERB_ALCSP Tellurium resistance protein TerB OS=Alcaligenes sp. GN=terB PE=4
BAC0611|modC|sp|P09833|MODC_ECOLI Molybdenum import ATP-binding protein ModC OS=Escherichia co

BAC0331|rcnB/yohN|sp|P64534|RCNB_ECOLI Nickel/cobalt homeostasis protein RcnB OS=Escherichia coli (st
BAC0698|ncrA|tr|Q1KLR2|Q1KLR2_SERMA NcrA OS=Serratia marcescens GN=ncrA PE=4 SV=1
BAC0083|copR|sp|Q02540|COPR_PSEUB Transcriptional activator protein CopR OS=Pseudomonas syringae pv
BAC0461|zntA/yhhO|sp|P37617|ATZN_ECOLI Lead, cadmium, zinc and mercury-transporting ATPase OS=Esc
BAC0653|merA|tr|Q934S5|Q934S5_THIFE Mercuric ion reductase OS=Thiobacillus ferrooxidans GN=merA PE
BAC0462|zntR/yhdM|sp|P0ACS5|ZNTR_ECOLI HTH-type transcriptional regulator ZntR OS=Escherichia coli (
BAC0695|merT|sp|P13112|MERT_SERMA Mercuric transport protein OS=Serratia marcescens GN=merT PE=4
BAC0029|chrF|tr|A4UQR2|A4UQR2_9RHIZ ChrF OS=Ochrobactrum tritici GN=chrF PE=4 SV=1
BAC0385|tehB|sp|P25397|TEHB_ECOLI Tellurite methyltransferase OS=Escherichia coli (strain K12) GN=tehB
BAC0457|zinT/yodA|sp|P76344|ZINT_ECOLI Metal-binding protein ZinT OS=Escherichia coli (strain K12) GN
BAC0467|zraR/hydH|sp|P14375|ZRAR_ECOLI Transcriptional regulatory protein ZraR OS=Escherichia coli (str
BAC0661|merB2|tr|Q7DJN2|Q7DJN2_BACME MerB2 OS=Bacillus megaterium GN=merB2 PE=4 SV=1
BAC0538|chrR|tr|Q7BD45|Q7BD45_PSEPU Chromate reductase OS=Pseudomonas putida GN=chrR PE=4 SV=
BAC0468|zraS/hydG|sp|P14377|ZRAS_ECOLI Sensor protein ZraS OS=Escherichia coli (strain K12) GN=zraS
BAC0572|arsA|sp|P08690|ARSA1_ECOLX Arsenical pump-driving ATPase OS=Escherichia coli GN=arsA PE=
BAC0646|mdtB|tr|D0ZND9|D0ZND9_SALT1 Multidrug resistance protein MdtB OS=Salmonella typhimurium (
BAC0303|pcoA|sp|Q47452|PCOA_ECOLX Copper resistance protein A OS=Escherichia coli GN=pcoA PE=3 S
BAC0463|znuA/yebL|sp|P39172|ZNUA_ECOLI High-affinity zinc uptake system protein ZnuA OS=Escherichia
BAC0253|mntR|sp|P0A9F1|MNTR_ECOLI Transcriptional regulator MntR OS=Escherichia coli (strain K12) GN
BAC0138|dsbC|sp|P0AEG6|DSBC_ECOLI Thiol:disulfide interchange protein DsbC OS=Escherichia coli (strain
BAC0026|chrA|tr|A4UQR4|A4UQR4_9RHIZ Chromate transporter OS=Ochrobactrum tritici GN=chrA PE=4 S
BAC0704|ncrC|tr|Q8VTR4|Q8VTR4_HAFAL NcrC OS=Hafnia alvei GN=ncrC PE=4 SV=1
BAC0265|ncrC|tr|D5CKG5|D5CKG5_ENTCC Nickel-resistant membrane protein-like protein NcrC OS=Enterob
BAC0650|merA|tr|O08449|O08449_9PSED Mercuric reductase OS=Pseudomonas sp. K-62 GN=merA PE=4 SV
BAC0707|sodB|sp|P0AGD3|SODF_ECOLI Superoxide dismutase [Fe] OS=Escherichia coli (strain K12) GN=so
BAC0183|golT|tr|Q8ZRG7|Q8ZRG7_SALTY Putative cation transport ATPase OS=Salmonella typhimurium (st
BAC0549|nccA|sp|Q44586|NCCA_ALCXX Nickel-cobalt-cadmium resistance protein NccA OS=Alcaligenes xy
BAC0298|pbrA|tr|Q58AJ6|Q58AJ6_RALME P-type ATPase involved in Pb(II) resistance PbrA OS=Ralstonia m
BAC0039|baeR|sp|P69228|BAER_ECOLI Transcriptional regulatory protein BaeR OS=Escherichia coli (strain K
BAC0233|merT|sp|P94185|MERT_ALCSP Mercuric transport protein OS=Alcaligenes sp. GN=merT PE=3 SV=
BAC0567|actA|sp|Q52910|LNT_SINMW Apolipoprotein N-acyltransferase OS=Sinorhizobium medicae (strain
BAC0116|cutF/nlpE|sp|P40710|NLPE_ECOLI Lipoprotein NlpE OS=Escherichia coli (strain K12) GN=nlpE PE=
BAC0597|baeS|tr|D0ZNE2|D0ZNE2_SALT1 Signal transduction histidine-protein kinase BaeS OS=Salmonella t
BAC0255|mreA|tr|Q88IN0|Q88IN0_PSEPK Putative uncharacterized protein OS=Pseudomonas putida (strain K
BAC0343|silC|sp|Q9ZHD2|SILC_SALTM Probable outer membrane lipoprotein SilC OS=Salmonella typhimuri
BAC0594|arsR|sp|P37309|ARSR_ECOLI Arsenical resistance operon repressor OS=Escherichia coli (strain K12)
BAC0066|chrF|tr|Q5NUZ7|Q5NUZ7_RALME ChrF, regulatory protein, involved in Chromate resistance OS=Ra
BAC0469|zupT/ygiE|sp|P0A8H3|ZUPT_ECOLI Zinc transporter ZupT OS=Escherichia coli (strain K12) GN=zu
BAC0356|recG|tr|Q9HTL3|Q9HTL3_PSEAE ATP-dependent DNA helicase RecG OS=Pseudomonas aeruginosa
BAC0012|actP|sp|Q9X5X3|ATCU_SINMW Copper-transporting P-type ATPase OS=Sinorhizobium medicae (st
BAC0678|merP|tr|O66016|O66016_PSEST MerP OS=Pseudomonas stutzeri GN=merP PE=4 SV=1
BAC0694|merT-P|tr|H6WCN3|H6WCN3_9FLAO MerT-P OS=Tenacibaculum discolor GN=merT-P PE=4 SV=
BAC0596|baeR|tr|D0ZNE3|D0ZNE3_SALT1 DNA-binding transcriptional regulator BaeR OS=Salmonella typhi
BAC0648|merA|sp|P08662|MERA_SERMA Mercuric reductase (Fragments) OS=Serratia marcescens GN=merA
BAC0639|copC|tr|C6FFR6|C6FFR6_PSEFL CopC OS=Pseudomonas fluorescens GN=copC PE=4 SV=1
BAC0591|arsR|sp|P52144|ARSR2_ECOLX Arsenical resistance operon repressor OS=Escherichia coli GN=arsR

BAC0446|yhcN|sp|P64614|YHCN_ECOLI Uncharacterized protein YhcN OS=Escherichia coli (strain K12) GN=
BAC0583|arsC|sp|P52147|ARSC2_ECOLX Arsenate reductase OS=Escherichia coli GN=arsC PE=3 SV=1
BAC0652|merA|tr|O66017|O66017_PSEST MerA OS=Pseudomonas stutzeri GN=merA PE=4 SV=1
BAC0667|merD|sp|P08654|MERD_SERMA HTH-type transcriptional regulator MerD OS=Serratia marcescens G
BAC0541|yieF|sp|P0AGE6|YIEF_ECOLI Uncharacterized protein YieF OS=Escherichia coli (strain K12) GN=y
BAC0629|copB|sp|O30085|COPB_ARCFU Copper-exporting P-type ATPase B OS=Archaeoglobus fulgidus (str
BAC0392|terZ|sp|Q52353|TERZ_SERMA Tellurium resistance protein TerZ OS=Serratia marcescens GN=terZ P
BAC0163|fecD|sp|P15029|FECD_ECOLI Fe(3+) dicitrate transport system permease protein FecD OS=Escherich
BAC0386|terA|sp|P18778|TERA_ALCSP Tellurium resistance protein TerA OS=Alcaligenes sp. GN=terA PE=4
BAC0119|czcA|sp|P13511|CZCA_RALME Cobalt-zinc-cadmium resistance protein CzcA OS=Ralstonia metallic
BAC0306|pcoD|sp|Q47455|PCOD_ECOLX Copper resistance protein D OS=Escherichia coli GN=pcoD PE=3 S
BAC0162|fbpC|sp|P44513|FBPC2_HAEIN Fe(3+) ions import ATP-binding protein FbpC 2 OS=Haemophilus in
BAC0383|tcrB|tr|Q8VPE6|Q8VPE6_ENTFC TcrB OS=Enterococcus faecium GN=tcrB PE=3 SV=1
BAC0309|pcoS|sp|Q47457|PCOS_ECOLX Probable sensor protein PcoS OS=Escherichia coli GN=pcoS PE=3 S
BAC0687|merR|tr|Q79BG7|Q79BG7_PSEST MerR OS=Pseudomonas stutzeri GN=merR PE=4 SV=1
BAC0692|merT|tr|Q79BG6|Q79BG6_PSEST MerT OS=Pseudomonas stutzeri GN=merT PE=4 SV=1
BAC0348|silS|sp|Q9ZHD4|SILS_SALTM Probable sensor kinase SilS OS=Salmonella typhimurium GN=silS PE
BAC0587|arsD|sp|P52148|ARSD2_ECOLX Arsenical resistance operon trans-acting repressor ArsD OS=Escheri
BAC0111|cusR/ylcA|sp|P0ACZ8|CUSR_ECOLI Transcriptional regulatory protein CusR OS=Escherichia coli (s
BAC0619|copA|tr|Q7WYH1|Q7WYH1_PSEPU CopA OS=Pseudomonas putida GN=copA PE=4 SV=1
BAC0040|baeS|sp|P30847|BAES_ECOLI Signal transduction histidine-protein kinase BaeS OS=Escherichia coli
BAC0271|nikB|sp|P33591|NIKB_ECOLI Nickel transport system permease protein NikB OS=Escherichia coli (s
BAC0254|mrdH|tr|Q88IN1|Q88IN1_PSEPK Membrane protein, putative OS=Pseudomonas putida (strain KT244
BAC0686|merR|sp|P13111|MERR_SERMA Mercuric resistance operon regulatory protein OS=Serratia marcesce
BAC0033|arsD|sp|P46003|ARSD1_ECOLX Arsenical resistance operon trans-acting repressor ArsD OS=Escheri
BAC0106|cuiD|sp|Q8ZRS2|CUEO_SALTY Blue copper oxidase CueO OS=Salmonella typhimurium (strain LT2
BAC0609|modA|sp|P37329|MODA_ECOLI Molybdate-binding periplasmic protein OS=Escherichia coli (strain 
BAC0102|cueA|tr|Q8KWW2|Q8KWW2_PSEPU Copper transporter OS=Pseudomonas putida GN=cueA PE=3 S
BAC0251|mntH/yfeP|sp|P0A769|MNTH_ECOLI Divalent metal cation transporter MntH OS=Escherichia coli (s
BAC0693|merT|tr|Q79F00|Q79F00_9PSED Mercuric transport protein OS=Pseudomonas sp. K-62 GN=merT PE
BAC0049|bhsA/ycfR/comC|sp|P0AB40|BHSA_ECOLI Multiple stress resistance protein BhsA OS=Escherichia c
BAC0643|corB|tr|Q9X621|Q9X621_SALTM CorB OS=Salmonella typhimurium GN=corB PE=4 SV=1
BAC0576|arsB|sp|P52146|ARSB2_ECOLX Arsenical pump membrane protein OS=Escherichia coli GN=arsB PE
BAC0082|copL|tr|Q5YKV8|Q5YKV8_9XANT CopL OS=Xanthomonas perforans GN=copL PE=4 SV=1
BAC0628|copB|tr|F4ZBX4|F4ZBX4_XANCI CopB OS=Xanthomonas citri subsp. citri GN=copB PE=4 SV=1
BAC0027|chrB|tr|A4UQR5|A4UQR5_9RHIZ ChrB OS=Ochrobactrum tritici GN=chrB PE=4 SV=1
BAC0133|dnaK|sp|P0A5B9|DNAK_MYCTU Chaperone protein DnaK OS=Mycobacterium tuberculosis GN=dn
BAC0305|pcoC|sp|Q47454|PCOC_ECOLX Copper resistance protein C OS=Escherichia coli GN=pcoC PE=1 SV
BAC0308|pcoR|sp|Q47456|PCOR_ECOLX Transcriptional regulatory protein PcoR OS=Escherichia coli GN=pc
BAC0659|merB|sp|P08664|MERB_SERMA Alkylmercury lyase OS=Serratia marcescens GN=merB PE=3 SV=1
BAC0612|perO|tr|D5AQ60|D5AQ60_RHOCB Divalent ion symporter OS=Rhodobacter capsulatus (strain ATCC
BAC0182|golS|tr|Q8ZRG6|Q8ZRG6_SALTY Putative transcriptional regulator OS=Salmonella typhimurium (str
BAC0307|pcoE|sp|Q47459|PCOE_ECOLX Probable copper-binding protein PcoE OS=Escherichia coli GN=pcoE
BAC0199|klaB/telA/kilB|sp|Q52328|KLAB_ECOLX Protein KlaB OS=Escherichia coli GN=klaB PE=3 SV=1
BAC0003|acn|tr|O53166|O53166_MYCTU Aconitate hydratase OS=Mycobacterium tuberculosis H37Rv GN=RV
BAC0293|ruvB|tr|B5L348|B5L348_9PSED Malic enzyme family protein (Fragment) OS=Pseudomonas corrugata

BAC0351|sitC|tr|Q9XCS0|Q9XCS0_SALTM SitC OS=Salmonella typhimurium GN=sitC PE=3 SV=1
BAC0676|merP|tr|Q7DHE4|Q7DHE4_BACCE Mercury-binding protein OS=Bacillus cereus GN=merP PE=4 SV
BAC0058|cadR|tr|Q93TP7|Q93TP7_PSEPU CadR OS=Pseudomonas putida GN=cadR PE=4 SV=1
BAC0579|arsB|sp|P74311|Y944_SYNY3 Uncharacterized transporter slr0944 OS=Synechocystis sp. (strain PCC
BAC0267|nczA|tr|B8GZE9|B8GZE9_CAUCN Cobalt-zinc-cadmium resistance protein czcA OS=Caulobacter cre
BAC0672|merE|tr|Q79BE4|Q79BE4_PSEST Urf1 OS=Pseudomonas stutzeri PE=4 SV=1
BAC0115|cutE/lnt|sp|P23930|LNT_ECOLI Apolipoprotein N-acyltransferase OS=Escherichia coli (strain K12) G
BAC0304|pcoB|sp|Q47453|PCOB_ECOLX Copper resistance protein B OS=Escherichia coli GN=pcoB PE=4 SV
BAC0334|robA|sp|P0ACI0|ROB_ECOLI Right origin-binding protein OS=Escherichia coli (strain K12) GN=rob
BAC0644|corD|sp|Q56017|APAG_SALTY Protein ApaG OS=Salmonella typhimurium (strain LT2 / SGSC1412
BAC0447|yjaA|sp|P09162|YJAA_ECOLI Uncharacterized protein YjaA OS=Escherichia coli (strain K12) GN=y
BAC0124|czcP|tr|Q1LAJ7|Q1LAJ7_RALME CzcP cation efflux P1-ATPase OS=Ralstonia metallidurans (strain
BAC0433|ybtQ|tr|Q9Z375|Q9Z375_YERPE Inner membrane ABC-transporter YbtQ OS=Yersinia pestis GN=yb
BAC0166|fetB/ybbM|sp|P77307|YBBM_ECOLI UPF0014 inner membrane protein YbbM OS=Escherichia coli (
BAC0570|actP|tr|D5AU53|D5AU53_RHOCB Cation/acetate symporter ActP-1 OS=Rhodobacter capsulatus (stra
BAC0578|arsB|tr|O50594|O50594_ACIMU ArsB OS=Acidiphilium multivorum GN=arsB PE=4 SV=1
BAC0169|fpvA|sp|P48632|FPVA_PSEAE Ferripyoverdine receptor OS=Pseudomonas aeruginosa (strain ATCC
BAC0610|modB|sp|P0AF01|MODB_ECOLI Molybdenum transport system permease protein ModB OS=Escheri
BAC0333|ricR|tr|O07434|O07434_MYCTU Regulated in copper repressor OS=Mycobacterium tuberculosis H37
BAC0689|merR|tr|Q934S8|Q934S8_THIFE Mer operon regulatory protein OS=Thiobacillus ferrooxidans GN=m
BAC0631|copC|tr|F4ZBX9|F4ZBX9_XANCI CopC OS=Xanthomonas citri subsp. citri GN=copC PE=4 SV=1
BAC0465|znuC/yebM|sp|P0A9X1|ZNUC_ECOLI Zinc import ATP-binding protein ZnuC OS=Escherichia coli (
BAC0312|pitA|sp|P0AFJ7|PITA_ECOLI Low-affinity inorganic phosphate transporter 1 OS=Escherichia coli (str
BAC0673|merE|sp|P06690|MERE_PSEAI Uncharacterized mercuric resistance protein MerE OS=Pseudomonas a
BAC0317|pstC|sp|P0AGH8|PSTC_ECOLI Phosphate transport system permease protein PstC OS=Escherichia co
BAC0059|cadX|tr|A7LHQ4|A7LHQ4_STRSL CadX OS=Streptococcus salivarius GN=cadX PE=4 SV=1
BAC0344|silE|sp|Q9Z4N3|SILE_SALTM Silver-binding protein SilE OS=Salmonella typhimurium GN=silE PE=
BAC0651|merA|sp|P0A0E5|MERA_STAAU Mercuric reductase OS=Staphylococcus aureus GN=merA PE=3 SV
BAC0371|soxS|sp|P0A9E2|SOXS_ECOLI Regulatory protein SoxS OS=Escherichia coli (strain K12) GN=soxS
BAC0682|merR1|sp|P22853|MERR_BACCE Mercuric resistance operon regulatory protein OS=Bacillus cereus
BAC0621|copA|tr|F4ZBX3|F4ZBX3_XANCI CopA OS=Xanthomonas citri subsp. citri GN=copA PE=4 SV=1
BAC0229|merG|tr|O07302|O07302_9PSED Mercuric resistance protein OS=Pseudomonas sp. K-62 GN=merG P
BAC0683|merR1|tr|O07300|O07300_9PSED Mercuric resistance operon regulatory protein OS=Pseudomonas sp
BAC0637|copS|tr|C6FFR5|C6FFR5_PSEFL CopS OS=Pseudomonas fluorescens GN=copS PE=4 SV=1
BAC0120|czcB|sp|P13510|CZCB_RALME Cobalt-zinc-cadmium resistance protein CzcB OS=Ralstonia metallid
BAC0647|mdtC|tr|D0ZNE0|D0ZNE0_SALT1 Multidrug resistance protein MdtC OS=Salmonella typhimurium (
BAC0165|fetA/ybbL|sp|P77279|YBBL_ECOLI Uncharacterized ABC transporter ATP-binding protein YbbL OS
BAC0109|cusC/ylcB|sp|P77211|CUSC_ECOLI Cation efflux system protein CusC OS=Escherichia coli (strain K
BAC0445|ygiW|sp|P0ADU5|YGIW_ECOLI Protein YgiW OS=Escherichia coli (strain K12) GN=ygiW PE=1 SV
BAC0274|nikE|sp|P33594|NIKE_ECOLI Nickel import ATP-binding protein NikE OS=Escherichia coli (strain K
BAC0086|corA|sp|P0ABI4|CORA_ECOLI Magnesium transport protein CorA OS=Escherichia coli (strain K12)
BAC0031|arsB|sp|P08691|ARSB1_ECOLX Arsenical pump membrane protein OS=Escherichia coli GN=arsB PE
BAC0112|cusS|sp|P77485|CUSS_ECOLI Sensor kinase CusS OS=Escherichia coli (strain K12) GN=cusS PE=1
BAC0035|arsM|tr|Q6N3Y0|Q6N3Y0_RHOPA UbiE/COQ5 methyltransferase OS=Rhodopseudomonas palustris
BAC0131|czrB|tr|Q9RLI9|Q9RLI9_PSEAI CzrB protein OS=Pseudomonas aeruginosa GN=czrB PE=4 SV=1
BAC0318|pstS|sp|P0AG82|PSTS_ECOLI Phosphate-binding protein PstS OS=Escherichia coli (strain K12) GN=

BAC0272|nikC|sp|P0AFA9|NIKC_ECOLI Nickel transport system permease protein NikC OS=Escherichia coli (
BAC0688|merR2|tr|Q79B70|Q79B70_PSEST Organomercurial resistance regulatory protein OS=Pseudomonas st
BAC0105|cueR/ybbI|sp|P0A9G4|CUER_ECOLI HTH-type transcriptional regulator CueR OS=Escherichia coli (
BAC0332|rcnR/yohL|sp|P64530|RCNR_ECOLI Transcriptional repressor RcnR OS=Escherichia coli (strain K12
BAC0228|merF|tr|Q2QCN0|Q2QCN0_9PSED MerF OS=Pseudomonas sp. CT14 GN=merF PE=4 SV=1
BAC0275|nikR|sp|P0A6Z6|NIKR_ECOLI Nickel-responsive regulator OS=Escherichia coli (strain K12) GN=nik
BAC0103|cueO|sp|P36649|CUEO_ECOLI Blue copper oxidase CueO OS=Escherichia coli (strain K12) GN=cue
BAC0022|aioR/aoxR|tr|Q2VGB1|Q2VGB1_RHIRD Putative transcriptional regulator OS=Rhizobium radiobacte
BAC0451|yodD|sp|P64519|YODD_ECOLI Uncharacterized protein YodD OS=Escherichia coli (strain K12) GN=
BAC0386|terA|sp|P18778|TERA_ALCSP Tellurium resistance protein TerA OS=Alcaligenes sp. GN=terA PE=4
BAC0124|czcP|tr|Q1LAJ7|Q1LAJ7_RALME CzcP cation efflux P1-ATPase OS=Ralstonia metallidurans (strain
BAC0392|terZ|sp|Q52353|TERZ_SERMA Tellurium resistance protein TerZ OS=Serratia marcescens GN=terZ P
BAC0631|copC|tr|F4ZBX9|F4ZBX9_XANCI CopC OS=Xanthomonas citri subsp. citri GN=copC PE=4 SV=1
BAC0266|ncrY|tr|Q06VT0|Q06VT0_9BACT NcrY OS=Leptospirillum ferriphilum GN=ncrY PE=4 SV=1
BAC0584|arsC|sp|O50595|ARSC_ACIMA Arsenate reductase OS=Acidiphilium multivorum (strain DSM 11245
BAC0112|cusS|sp|P77485|CUSS_ECOLI Sensor kinase CusS OS=Escherichia coli (strain K12) GN=cusS PE=1
BAC0265|ncrC|tr|D5CKG5|D5CKG5_ENTCC Nickel-resistant membrane protein-like protein NcrC OS=Enterob
BAC0447|yjaA|sp|P09162|YJAA_ECOLI Uncharacterized protein YjaA OS=Escherichia coli (strain K12) GN=y
BAC0683|merR1|tr|O07300|O07300_9PSED Mercuric resistance operon regulatory protein OS=Pseudomonas sp
BAC0102|cueA|tr|Q8KWW2|Q8KWW2_PSEPU Copper transporter OS=Pseudomonas putida GN=cueA PE=3 S
BAC0071|cmtR|sp|P67731|CMTR_MYCTU HTH-type transcriptional regulator CmtR OS=Mycobacterium tuber
BAC0630|copC|sp|P12376|COPC_PSEUB Copper resistance protein C OS=Pseudomonas syringae pv. tomato G
BAC0113|cutA|sp|P69488|CUTA_ECOLI Divalent-cation tolerance protein CutA OS=Escherichia coli (strain K1
BAC0169|fpvA|sp|P48632|FPVA_PSEAE Ferripyoverdine receptor OS=Pseudomonas aeruginosa (strain ATCC
BAC0138|dsbC|sp|P0AEG6|DSBC_ECOLI Thiol:disulfide interchange protein DsbC OS=Escherichia coli (strain
BAC0670|merE|tr|Q52104|Q52104_9ZZZZ Uncharacterized protein OS=Plasmid pDU1358 PE=4 SV=1
BAC0077|copA|sp|P12374|COPA_PSEUB Copper resistance protein A OS=Pseudomonas syringae pv. tomato G
BAC0490|G2alt|tr|B0FSM1|B0FSM1_9BACI 7-cyano-7-deazaguanine synthase OS=Anoxybacillus gonensis G
BAC0087|mgtA|sp|P0ABB8|ATMA_ECOLI Magnesium-transporting ATPase, P-type 1 OS=Escherichia coli (st
BAC0684|merR2|tr|Q7DKL2|Q7DKL2_9PSED MerR2 OS=Pseudomonas sp. K-62 GN=merR2 PE=4 SV=1
BAC0694|merT-P|tr|H6WCN3|H6WCN3_9FLAO MerT-P OS=Tenacibaculum discolor GN=merT-P PE=4 SV=
BAC0652|merA|tr|O66017|O66017_PSEST MerA OS=Pseudomonas stutzeri GN=merA PE=4 SV=1
BAC0082|copL|tr|Q5YKV8|Q5YKV8_9XANT CopL OS=Xanthomonas perforans GN=copL PE=4 SV=1
BAC0391|terW|sp|P75010|TERW_SERMA Tellurium resistance protein TerW OS=Serratia marcescens GN=ter
BAC0467|zraR/hydH|sp|P14375|ZRAR_ECOLI Transcriptional regulatory protein ZraR OS=Escherichia coli (st
BAC0224|merA|sp|P16171|MERA_BACCE Mercuric reductase OS=Bacillus cereus GN=merA PE=1 SV=1
BAC0056|cadC|sp|P20047|CADC_STAAU Cadmium resistance transcriptional regulatory protein CadC OS=Sta
BAC0027|chrB|tr|A4UQR5|A4UQR5_9RHIZ ChrB OS=Ochrobactrum tritici GN=chrB PE=4 SV=1
BAC0568|actP|sp|Q9X5V3|ATCU_RHILV Copper-transporting P-type ATPase OS=Rhizobium leguminosarum
BAC0276|nirA|tr|Q6RUG3|Q6RUG3_KLEOX NirA OS=Klebsiella oxytoca GN=nirA PE=4 SV=1
BAC0079|copB|sp|P05425|COPB_ENTHA Copper-exporting P-type ATPase B OS=Enterococcus hirae (strain A
BAC0576|arsB|sp|P52146|ARSB2_ECOLX Arsenical pump membrane protein OS=Escherichia coli GN=arsB P
BAC0646|mdtB|tr|D0ZND9|D0ZND9_SALT1 Multidrug resistance protein MdtB OS=Salmonella typhimurium (
BAC0103|cueO|sp|P36649|CUEO_ECOLI Blue copper oxidase CueO OS=Escherichia coli (strain K12) GN=cue
BAC0040|baeS|sp|P30847|BAES_ECOLI Signal transduction histidine-protein kinase BaeS OS=Escherichia coli
BAC0026|chrA|tr|A4UQR4|A4UQR4_9RHIZ Chromate transporter OS=Ochrobactrum tritici GN=chrA PE=4 S

BAC0571|arsA|sp|O50593|ARSA_ACIMA Arsenical pump-driving ATPase OS=Acidiphilium multivorum (strai
BAC0137|dsbB|sp|P0A6M2|DSBB_ECOLI Disulfide bond formation protein B OS=Escherichia coli (strain K12)
BAC0668|merD|tr|Q5NUV1|Q5NUV1_RALME MerD from Tn4378, regulatory protein involved in Hg(II) resist
BAC0691|merT|tr|Q52397|Q52397_PSEST Mercury transport protein OS=Pseudomonas stutzeri GN=merT PE=4
BAC0341|silA|sp|Q9ZHC9|SILA_SALTM Putative cation efflux system protein SilA OS=Salmonella typhimuriu
BAC0368|sodA|sp|P00448|SODM_ECOLI Superoxide dismutase [Mn] OS=Escherichia coli (strain K12) GN=so
BAC0383|tcrB|tr|Q8VPE6|Q8VPE6_ENTFC TcrB OS=Enterococcus faecium GN=tcrB PE=3 SV=1
BAC0031|arsB|sp|P08691|ARSB1_ECOLX Arsenical pump membrane protein OS=Escherichia coli GN=arsB P
BAC0252|mntP/yebN|sp|P76264|MNTP_ECOLI Probable manganese efflux pump MntP OS=Escherichia coli (st
BAC0305|pcoC|sp|Q47454|PCOC_ECOLX Copper resistance protein C OS=Escherichia coli GN=pcoC PE=1 S
BAC0678|merP|tr|O66016|O66016_PSEST MerP OS=Pseudomonas stutzeri GN=merP PE=4 SV=1
BAC0092|corT/coaT|tr|H0P0Y3|H0P0Y3_9SYNC Cation-transporting ATPase E1-E2 ATPase OS=Synechocysti
BAC0538|chrR|tr|Q7BD45|Q7BD45_PSEPU Chromate reductase OS=Pseudomonas putida GN=chrR PE=4 SV=
BAC0035|arsM|tr|Q6N3Y0|Q6N3Y0_RHOPA UbiE/COQ5 methyltransferase OS=Rhodopseudomonas palustris
BAC0285|nreB|tr|F0KND8|F0KND8_ACICP NrsD, nreB nickel permease involved in nickel and cobalt toleranc
BAC0673|merE|sp|P06690|MERE_PSEAI Uncharacterized mercuric resistance protein MerE OS=Pseudomonas a
BAC0666|merD|tr|O66018|O66018_PSEST MerD OS=Pseudomonas stutzeri GN=merD PE=4 SV=1
BAC0651|merA|sp|P0A0E5|MERA_STAAU Mercuric reductase OS=Staphylococcus aureus GN=merA PE=3 S
BAC0101|ctpV|sp|P77894|CTPV_MYCTU Probable copper-exporting P-type ATPase V OS=Mycobacterium tub
BAC0390|terE|sp|P18782|TERE_ALCSP Tellurium resistance protein TerE OS=Alcaligenes sp. GN=terE PE=3 S
BAC0355|ruvB|sp|Q51426|RUVB_PSEAE Holliday junction ATP-dependent DNA helicase RuvB OS=Pseudom
BAC0316|pstB|sp|P0AAH0|PSTB_ECOLI Phosphate import ATP-binding protein PstB OS=Escherichia coli (stra
BAC0587|arsD|sp|P52148|ARSD2_ECOLX Arsenical resistance operon trans-acting repressor ArsD OS=Escheri
BAC0434|ychH|sp|P0AB49|YCHH_ECOLI Uncharacterized protein YchH OS=Escherichia coli (strain K12) GN
BAC0549|nccA|sp|Q44586|NCCA_ALCXX Nickel-cobalt-cadmium resistance protein NccA OS=Alcaligenes xy
BAC0032|arsC|sp|P0A006|ARSC_STAAU Protein ArsC OS=Staphylococcus aureus GN=arsC PE=1 SV=1
BAC0099|ctpD|sp|A0R3A7|CTPD_MYCS2 Probable cobalt/nickel-exporting P-type ATPase OS=Mycobacteriun
BAC0498|ideR|sp|P0A672|IDER_MYCTU Iron-dependent repressor IdeR OS=Mycobacterium tuberculosis GN=
BAC0330|rcnA/yohM|sp|P76425|RCNA_ECOLI Nickel/cobalt efflux system RcnA OS=Escherichia coli (strain K
BAC0302|pbrT|tr|Q5GR69|Q5GR69_ALCXX Lead uptake protein PbrT OS=Alcaligenes xylosoxydans xylosoxy
BAC0608|modE|sp|P0A9G8|MODE_ECOLI Transcriptional regulator ModE OS=Escherichia coli (strain K12) G
BAC0679|merP|tr|O66047|O66047_PSEST Mercury transport protein OS=Pseudomonas stutzeri GN=merP PE=4
BAC0470|zur/yjbK|sp|P0AC51|ZUR_ECOLI Zinc uptake regulation protein OS=Escherichia coli (strain K12) GN
BAC0573|arsB|sp|P45946|ARSB_BACSU Arsenite resistance protein ArsB OS=Bacillus subtilis (strain 168) GN
BAC0255|mreA|tr|Q88IN0|Q88IN0_PSEPK Putative uncharacterized protein OS=Pseudomonas putida (strain KT
BAC0022|aioR/aoxR|tr|Q2VGB1|Q2VGB1_RHIRD Putative transcriptional regulator OS=Rhizobium radiobacte
BAC0299|pbrB/pbrC|tr|Q58AJ7|Q58AJ7_RALME Lipoprotein signal peptidase OS=Ralstonia metallidurans (stra
BAC0621|copA|tr|F4ZBX3|F4ZBX3_XANCI CopA OS=Xanthomonas citri subsp. citri GN=copA PE=4 SV=1
BAC0273|nikD|sp|P33593|NIKD_ECOLI Nickel import ATP-binding protein NikD OS=Escherichia coli (strain K
BAC0076|comR/ycfQ|sp|P75952|COMR_ECOLI HTH-type transcriptional repressor ComR OS=Escherichia coli
BAC0204|cnrB|sp|P37973|CNRB_RALME Nickel and cobalt resistance protein CnrB OS=Ralstonia metalliduran
BAC0626|copB|sp|P12375|COPB_PSEUB Copper resistance protein B OS=Pseudomonas syringae pv. tomato G
BAC0012|actP|sp|Q9X5X3|ATCU_SINMW Copper-transporting P-type ATPase OS=Sinorhizobium medicae (st
BAC0541|yieF|sp|P0AGE6|YIEF_ECOLI Uncharacterized protein YieF OS=Escherichia coli (strain K12) GN=y
BAC0385|tehB|sp|P25397|TEHB_ECOLI Tellurite methyltransferase OS=Escherichia coli (strain K12) GN=tehB
BAC0645|mdtA|tr|D0ZND8|D0ZND8_SALT1 Multidrug resistance protein MdtA OS=Salmonella typhimurium (

BAC0439|yfeA|sp|Q56952|YFEA_YERPE Periplasmic chelated iron-binding protein YfeA OS=Yersinia pestis G
BAC0228|merF|tr|Q2QCN0|Q2QCN0_9PSED MerF OS=Pseudomonas sp. CT14 GN=merF PE=4 SV=1
BAC0489|ALU1-P|tr|O52119|O52119_ARTVI Aluminum resistance protein (Fragment) OS=Arthrobacter viscos
BAC0572|arsA|sp|P08690|ARSA1_ECOLX Arsenical pump-driving ATPase OS=Escherichia coli GN=arsA PE=
BAC0084|copY/tcrY|sp|Q47839|COPY_ENTHA Transcriptional repressor CopY OS=Enterococcus hirae (strain
BAC0433|ybtQ|tr|Q9Z375|Q9Z375_YERPE Inner membrane ABC-transporter YbtQ OS=Yersinia pestis GN=yb
BAC0334|robA|sp|P0ACI0|ROB_ECOLI Right origin-binding protein OS=Escherichia coli (strain K12) GN=rob
BAC0627|copB|tr|F4ZD00|F4ZD00_9XANT Copper resistance protein B OS=Xanthomonas alfalfae subsp. citru
BAC0263|ncrA|tr|Q06VT3|Q06VT3_9BACT NcrA OS=Leptospirillum ferriphilum GN=ncrA PE=4 SV=1
BAC0594|arsR|sp|P37309|ARSR_ECOLI Arsenical resistance operon repressor OS=Escherichia coli (strain K12)
BAC0136|dsbA|sp|P0AEG4|DSBA_ECOLI Thiol:disulfide interchange protein DsbA OS=Escherichia coli (strai
BAC0589|arsR|sp|P15905|ARSR1_ECOLX Arsenical resistance operon repressor OS=Escherichia coli GN=arsR
BAC0171|furA|sp|P0A582|FURA_MYCTU Transcriptional regulator FurA OS=Mycobacterium tuberculosis GN
BAC0267|nczA|tr|B8GZE9|B8GZE9_CAUCN Cobalt-zinc-cadmium resistance protein czcA OS=Caulobacter cr
BAC0270|nikA|sp|P33590|NIKA_ECOLI Nickel-binding periplasmic protein OS=Escherichia coli (strain K12) G
BAC0638|copR|tr|C6FFR4|C6FFR4_PSEFL CopR OS=Pseudomonas fluorescens GN=copR PE=4 SV=1
BAC0315|pstA|sp|P07654|PSTA_ECOLI Phosphate transport system permease protein PstA OS=Escherichia col
BAC0303|pcoA|sp|Q47452|PCOA_ECOLX Copper resistance protein A OS=Escherichia coli GN=pcoA PE=3 S
BAC0445|ygiW|sp|P0ADU5|YGIW_ECOLI Protein YgiW OS=Escherichia coli (strain K12) GN=ygiW PE=1 S
BAC0628|copB|tr|F4ZBX4|F4ZBX4_XANCI CopB OS=Xanthomonas citri subsp. citri GN=copB PE=4 SV=1
BAC0629|copB|sp|O30085|COPB_ARCFU Copper-exporting P-type ATPase B OS=Archaeoglobus fulgidus (str
BAC0695|merT|sp|P13112|MERT_SERMA Mercuric transport protein OS=Serratia marcescens GN=merT PE=4
BAC0588|arsR|tr|P74986|P74986_YEREN Arsenite inducible repressor OS=Yersinia enterocolitica GN=arsR PE
BAC0452|yqjH|sp|Q46871|YQJH_ECOLI NADPH-dependent ferric-chelate reductase OS=Escherichia coli (strai
BAC0612|perO|tr|D5AQ60|D5AQ60_RHOCB Divalent ion symporter OS=Rhodobacter capsulatus (strain ATCC
BAC0682|merR1|sp|P22853|MERR_BACCE Mercuric resistance operon regulatory protein OS=Bacillus cereus
BAC0181|glpF|sp|P0AER0|GLPF_ECOLI Glycerol uptake facilitator protein OS=Escherichia coli (strain K12) G
BAC0570|actP|tr|D5AU53|D5AU53_RHOCB Cation/acetate symporter ActP-1 OS=Rhodobacter capsulatus (stra
BAC0441|yfeC|sp|Q56954|YFEC_YERPE Chelated iron transport system membrane protein YfeC OS=Yersinia
BAC0384|tehA|sp|P25396|TEHA_ECOLI Tellurite resistance protein TehA OS=Escherichia coli (strain K12) GN
BAC0306|pcoD|sp|Q47455|PCOD_ECOLX Copper resistance protein D OS=Escherichia coli GN=pcoD PE=3 S
BAC0089|corR|tr|Q1D6V8|Q1D6V8_MYXXD Sigma-54 dependent DNA-binding response regulator OS=Myxo
BAC0133|dnaK|sp|P0A5B9|DNAK_MYCTU Chaperone protein DnaK OS=Mycobacterium tuberculosis GN=dn
BAC0692|merT|tr|Q79BG6|Q79BG6_PSEST MerT OS=Pseudomonas stutzeri GN=merT PE=4 SV=1
BAC0350|sitB|tr|Q9XCS1|Q9XCS1_SALTM SitB OS=Salmonella typhimurium GN=sitB PE=3 SV=1
BAC0254|mrdH|tr|Q88IN1|Q88IN1_PSEPK Membrane protein, putative OS=Pseudomonas putida (strain KT244
BAC0110|cusF/cusX|sp|P77214|CUSF_ECOLI Cation efflux system protein CusF OS=Escherichia coli (strain K
BAC0637|copS|tr|C6FFR5|C6FFR5_PSEFL CopS OS=Pseudomonas fluorescens GN=copS PE=4 SV=1
BAC0597|baeS|tr|D0ZNE2|D0ZNE2_SALT1 Signal transduction histidine-protein kinase BaeS OS=Salmonella t
BAC0685|merR|tr|H6WCN2|H6WCN2_9FLAO MerR OS=Tenacibaculum discolor GN=merR PE=4 SV=1
BAC0063|chrA|sp|P14285|CHRA_PSEAI Chromate transport protein OS=Pseudomonas aeruginosa GN=chrA P
BAC0272|nikC|sp|P0AFA9|NIKC_ECOLI Nickel transport system permease protein NikC OS=Escherichia coli (
BAC0231|merP|sp|P13113|MERP_SERMA Mercuric transport protein periplasmic component OS=Serratia marc
BAC0676|merP|tr|Q7DHE4|Q7DHE4_BACCE Mercury-binding protein OS=Bacillus cereus GN=merP PE=4 S
BAC0122|czcD|sp|P13512|CZCD_RALME Cobalt-zinc-cadmium resistance protein CzcD OS=Ralstonia metalli
BAC0033|arsD|sp|P46003|ARSD1_ECOLX Arsenical resistance operon trans-acting repressor ArsD OS=Escheri

BAC0111|cusR/ylcA|sp|P0ACZ8|CUSR_ECOLI Transcriptional regulatory protein CusR OS=Escherichia coli (s
BAC0347|silR|sp|Q9ZHD3|SILR_SALTM Probable transcriptional regulatory protein SilR OS=Salmonella typhi
BAC0116|cutF/nlpE|sp|P40710|NLPE_ECOLI Lipoprotein NlpE OS=Escherichia coli (strain K12) GN=nlpE PE=
BAC0563|acrD|tr|Q8ZN77|Q8ZN77_SALTY RND family aminoglycoside/multidrug efflux pump OS=Salmonel
BAC0199|klaB/telA/kilB|sp|Q52328|KLAB_ECOLX Protein KlaB OS=Escherichia coli GN=klaB PE=3 SV=1
BAC0086|corA|sp|P0ABI4|CORA_ECOLI Magnesium transport protein CorA OS=Escherichia coli (strain K12)
BAC0688|merR2|tr|Q79B70|Q79B70_PSEST Organomercurial resistance regulatory protein OS=Pseudomonas s
BAC0100|ctpG|sp|P63689|CTPG_MYCTU Probable cation-transporting ATPase G OS=Mycobacterium tubercul
BAC0649|merA|tr|E3VST6|E3VST6_9FLAO MerA OS=Tenacibaculum discolor GN=merA PE=3 SV=1
BAC0700|ncrC|tr|Q1KLR0|Q1KLR0_SERMA NcrC OS=Serratia marcescens GN=ncrC PE=4 SV=1
BAC0331|rcnB/yohN|sp|P64534|RCNB_ECOLI Nickel/cobalt homeostasis protein RcnB OS=Escherichia coli (s
BAC0274|nikE|sp|P33594|NIKE_ECOLI Nickel import ATP-binding protein NikE OS=Escherichia coli (strain K
BAC0049|bhsA/ycfR/comC|sp|P0AB40|BHSA_ECOLI Multiple stress resistance protein BhsA OS=Escherichia
BAC0387|terB|sp|P18779|TERB_ALCSP Tellurium resistance protein TerB OS=Alcaligenes sp. GN=terB PE=4
BAC0298|pbrA|tr|Q58AJ6|Q58AJ6_RALME P-type ATPase involved in Pb(II) resistance PbrA OS=Ralstonia m
BAC0388|terC|sp|P18780|TERC_ALCSP Tellurium resistance protein TerC OS=Alcaligenes sp. GN=terC PE=3
BAC0088|corC|sp|P0A2L3|CORC_SALTY Magnesium and cobalt efflux protein CorC OS=Salmonella typhimu
BAC0152|srpC|sp|Q55027|SRPC_SYNE7 Probable chromate transport protein OS=Synechococcus elongatus (st
BAC0591|arsR|sp|P52144|ARSR2_ECOLX Arsenical resistance operon repressor OS=Escherichia coli GN=arsR
BAC0066|chrF|tr|Q5NUZ7|Q5NUZ7_RALME ChrF, regulatory protein, involved in Chromate resistance OS=Ra
BAC0163|fecD|sp|P15029|FECD_ECOLI Fe(3+) dicitrate transport system permease protein FecD OS=Escheric
BAC0693|merT|tr|Q79F00|Q79F00_9PSED Mercuric transport protein OS=Pseudomonas sp. K-62 GN=merT PE
BAC0057|cadD|tr|Q7A320|Q7A320_STAAN CadD OS=Staphylococcus aureus (strain N315) GN=cadD PE=4 S
BAC0578|arsB|tr|O50594|O50594_ACIMU ArsB OS=Acidiphilium multivorum GN=arsB PE=4 SV=1
BAC0566|actS|tr|Q52912|Q52912_9RHIZ Histidine protein kinase OS=Sinorhizobium medicae GN=actS PE=4 S
BAC0342|silB|sp|Q9ZHD0|SILB_SALTM Putative membrane fusion protein SilB OS=Salmonella typhimurium
BAC0464|znuB/yebI|sp|P39832|ZNUB_ECOLI High-affinity zinc uptake system membrane protein ZnuB OS=E
BAC0346|silP|sp|Q9ZHC7|SILP_SALTM Silver exporting P-type ATPase OS=Salmonella typhimurium GN=sil
BAC0240|mexI|tr|Q9HWH4|Q9HWH4_PSEAE Probable Resistance-Nodulation-Cell Division (RND) efflux tra
BAC0548|chrA1|sp|P17551|CHRA1_RALME Chromate transport protein OS=Ralstonia metallidurans (strain C
BAC0609|modA|sp|P37329|MODA_ECOLI Molybdate-binding periplasmic protein OS=Escherichia coli (strain
BAC0698|ncrA|tr|Q1KLR2|Q1KLR2_SERMA NcrA OS=Serratia marcescens GN=ncrA PE=4 SV=1
BAC0643|corB|tr|Q9X621|Q9X621_SALTM CorB OS=Salmonella typhimurium GN=corB PE=4 SV=1
BAC0639|copC|tr|C6FFR6|C6FFR6_PSEFL CopC OS=Pseudomonas fluorescens GN=copC PE=4 SV=1
BAC0611|modC|sp|P09833|MODC_ECOLI Molybdenum import ATP-binding protein ModC OS=Escherichia c
BAC0565|actR|sp|A6UEL7|ACTR_SINMW Acid tolerance regulatory protein ActR OS=Sinorhizobium medicae
BAC0229|merG|tr|O07302|O07302_9PSED Mercuric resistance protein OS=Pseudomonas sp. K-62 GN=merG P
BAC0028|chrC|tr|A4UQR3|A4UQR3_9RHIZ Superoxide dismutase OS=Ochrobactrum tritici GN=chrC PE=3 S
BAC0468|zraS/hydG|sp|P14377|ZRAS_ECOLI Sensor protein ZraS OS=Escherichia coli (strain K12) GN=zraS
BAC0345|silF|tr|Q9ZHD1|Q9ZHD1_SALTM Uncharacterized protein OS=Salmonella typhimurium GN=ORF96
BAC0665|merD|tr|O66022|O66022_PSEST Mercury operon coregulator protein OS=Pseudomonas stutzeri GN=
BAC0114|cutC|sp|P67826|CUTC_ECOLI Copper homeostasis protein CutC OS=Escherichia coli (strain K12) G
BAC0233|merT|sp|P94185|MERT_ALCSP Mercuric transport protein OS=Alcaligenes sp. GN=merT PE=3 SV=
BAC0059|cadX|tr|A7LHQ4|A7LHQ4_STRSL CadX OS=Streptococcus salivarius GN=cadX PE=4 SV=1
BAC0461|zntA/yhhO|sp|P37617|ATZN_ECOLI Lead, cadmium, zinc and mercury-transporting ATPase OS=Esc
BAC0048|bfrA|sp|P63697|BFR_MYCTU Bacterioferritin OS=Mycobacterium tuberculosis GN=bfr PE=1 SV=1

BAC0312|pitA|sp|P0AFJ7|PITA_ECOLI Low-affinity inorganic phosphate transporter 1 OS=Escherichia coli (str
BAC0458|zipB|tr|Q7WJT8|Q7WJT8_BORBR Putative membrane protein OS=Bordetella bronchiseptica (strain A
BAC0553|nccX|sp|Q44582|NCCX_ALCXX Nickel-cobalt-cadmium resistance protein NccX OS=Alcaligenes xy
BAC0699|ncrB|tr|Q1KLR1|Q1KLR1_SERMA NcrB OS=Serratia marcescens GN=ncrB PE=4 SV=1
BAC0120|czcB|sp|P13510|CZCB_RALME Cobalt-zinc-cadmium resistance protein CzcB OS=Ralstonia metallid
BAC0275|nikR|sp|P0A6Z6|NIKR_ECOLI Nickel-responsive regulator OS=Escherichia coli (strain K12) GN=nik
BAC0690|merT|tr|Q934S7|Q934S7_THIFE Mercuric ion transport protein OS=Thiobacillus ferrooxidans GN=m
BAC0457|zinT/yodA|sp|P76344|ZINT_ECOLI Metal-binding protein ZinT OS=Escherichia coli (strain K12) GN
BAC0599|modB|tr|Q72FN2|Q72FN2_DESVH Molybdenum ABC transporter, permease protein OS=Desulfovibr
BAC0083|copR|sp|Q02540|COPR_PSEUB Transcriptional activator protein CopR OS=Pseudomonas syringae pv
BAC0642|mgtA|sp|P36640|ATMA_SALTY Magnesium-transporting ATPase, P-type 1 OS=Salmonella typhimu
BAC0648|merA|sp|P08662|MERA_SERMA Mercuric reductase (Fragments) OS=Serratia marcescens GN=merA
BAC0664|merC|tr|O66048|O66048_PSEST MerC OS=Pseudomonas stutzeri GN=merC PE=4 SV=1
BAC0389|terD|sp|P18781|TERD_ALCSP Tellurium resistance protein TerD OS=Alcaligenes sp. GN=terD PE=3
BAC0115|cutE/lnt|sp|P23930|LNT_ECOLI Apolipoprotein N-acyltransferase OS=Escherichia coli (strain K12) G
BAC0308|pcoR|sp|Q47456|PCOR_ECOLX Transcriptional regulatory protein PcoR OS=Escherichia coli GN=pc
BAC0106|cuiD|sp|Q8ZRS2|CUEO_SALTY Blue copper oxidase CueO OS=Salmonella typhimurium (strain LT2
BAC0583|arsC|sp|P52147|ARSC2_ECOLX Arsenate reductase OS=Escherichia coli GN=arsC PE=3 SV=1
BAC0446|yhcN|sp|P64614|YHCN_ECOLI Uncharacterized protein YhcN OS=Escherichia coli (strain K12) GN=
BAC0469|zupT/ygiE|sp|P0A8H3|ZUPT_ECOLI Zinc transporter ZupT OS=Escherichia coli (strain K12) GN=zu
BAC0253|mntR|sp|P0A9F1|MNTR_ECOLI Transcriptional regulator MntR OS=Escherichia coli (strain K12) GN
BAC0459|zitB/ybgR|sp|P75757|ZITB_ECOLI Zinc transporter ZitB OS=Escherichia coli (strain K12) GN=zitB P
BAC0098|ctpC|sp|P0A502|CTPC_MYCTU Probable manganese/zinc-exporting P-type ATPase OS=Mycobacteri
BAC0640|copD|tr|C6FFR7|C6FFR7_PSEFL CopD OS=Pseudomonas fluorescens GN=copD PE=4 SV=1
BAC0582|arsC|sp|P08692|ARSC1_ECOLX Arsenate reductase OS=Escherichia coli GN=arsC PE=1 SV=1
BAC0659|merB|sp|P08664|MERB_SERMA Alkylmercury lyase OS=Serratia marcescens GN=merB PE=3 SV=1
BAC0164|fecE|sp|P15031|FECE_ECOLI Fe(3+) dicitrate transport ATP-binding protein FecE OS=Escherichia co
BAC0264|ncrB|tr|Q06VT2|Q06VT2_9BACT NcrB OS=Leptospirillum ferriphilum GN=ncrB PE=4 SV=1
BAC0182|golS|tr|Q8ZRG6|Q8ZRG6_SALTY Putative transcriptional regulator OS=Salmonella typhimurium (str
BAC0707|sodB|sp|P0AGD3|SODF_ECOLI Superoxide dismutase [Fe] OS=Escherichia coli (strain K12) GN=so
BAC0003|acn|tr|O53166|O53166_MYCTU Aconitate hydratase OS=Mycobacterium tuberculosis H37Rv GN=R
BAC0161|fbpB|sp|P71338|FBPB2_HAEIN Fe(3+)-transport system permease protein FbpB 2 OS=Haemophilus i
BAC0317|pstC|sp|P0AGH8|PSTC_ECOLI Phosphate transport system permease protein PstC OS=Escherichia co
BAC0352|sitD|tr|Q9XCR9|Q9XCR9_SALTM SitD OS=Salmonella typhimurium GN=sitD PE=3 SV=1
BAC0585|arsC|sp|P74984|ARSC_YEREN Arsenate reductase OS=Yersinia enterocolitica GN=arsC PE=3 SV=1
BAC0203|cnrA|sp|P37972|CNRA_RALME Nickel and cobalt resistance protein CnrA OS=Ralstonia metallidura
BAC0034|arsH|tr|E8PS81|E8PS81_YERPE Arsenic resistance protein ArsH OS=Yersinia pestis Java 9 GN=arsH
BAC0432|ybtP|tr|Q9R7V3|Q9R7V3_YERPE Lipoprotein inner membrane ABC-transporter OS=Yersinia pestis C
BAC0540|nfsA|sp|P17117|NFSA_ECOLI Oxygen-insensitive NADPH nitroreductase OS=Escherichia coli (strain
BAC0030|arsA|sp|P52145|ARSA2_ECOLX Arsenical pump-driving ATPase OS=Escherichia coli GN=arsA PE=
BAC0166|fetB/ybbM|sp|P77307|YBBM_ECOLI UPF0014 inner membrane protein YbbM OS=Escherichia coli (
BAC0358|oscA|tr|B6CM35|B6CM35_9PSED Putative uncharacterized protein oscA OS=Pseudomonas corrugata
BAC0250|mntA/ytgA|sp|O34385|MNTA_BACSU Manganese-binding lipoprotein MntA OS=Bacillus subtilis (st
BAC0332|rcnR/yohL|sp|P64530|RCNR_ECOLI Transcriptional repressor RcnR OS=Escherichia coli (strain K12
BAC0293|ruvB|tr|B5L348|B5L348_9PSED Malic enzyme family protein (Fragment) OS=Pseudomonas corrugata
BAC0105|cueR/ybbI|sp|P0A9G4|CUER_ECOLI HTH-type transcriptional regulator CueR OS=Escherichia coli (

BAC0183|golT|tr|Q8ZRG7|Q8ZRG7_SALTY Putative cation transport ATPase OS=Salmonella typhimurium (st
BAC0455|ziaA|sp|Q59998|ATZN_SYNY3 Zinc-transporting ATPase OS=Synechocystis sp. (strain PCC 6803 / K
BAC0058|cadR|tr|Q93TP7|Q93TP7_PSEPU CadR OS=Pseudomonas putida GN=cadR PE=4 SV=1
BAC0357|recG|tr|B5L350|B5L350_9PSED ATP-dependent DNA helicase (Fragment) OS=Pseudomonas corruga
BAC0271|nikB|sp|P33591|NIKB_ECOLI Nickel transport system permease protein NikB OS=Escherichia coli (s
BAC0167|fieF/yiip|sp|P69380|FIEF_ECOLI Ferrous-iron efflux pump FieF OS=Escherichia coli (strain K12) GN
BAC0130|czrA|tr|Q9RLI8|Q9RLI8_PSEAI CzrA protein OS=Pseudomonas aeruginosa GN=czrA PE=4 SV=1
BAC0650|merA|tr|O08449|O08449_9PSED Mercuric reductase OS=Pseudomonas sp. K-62 GN=merA PE=4 SV
BAC0307|pcoE|sp|Q47459|PCOE_ECOLX Probable copper-binding protein PcoE OS=Escherichia coli GN=pcoE
BAC0356|recG|tr|Q9HTL3|Q9HTL3_PSEAE ATP-dependent DNA helicase RecG OS=Pseudomonas aeruginosa
BAC0109|cusC/ylcB|sp|P77211|CUSC_ECOLI Cation efflux system protein CusC OS=Escherichia coli (strain K
BAC0251|mntH/yfeP|sp|P0A769|MNTH_ECOLI Divalent metal cation transporter MntH OS=Escherichia coli (s
BAC0463|znuA/yebL|sp|P39172|ZNUA_ECOLI High-affinity zinc uptake system protein ZnuA OS=Escherichia
BAC0371|soxS|sp|P0A9E2|SOXS_ECOLI Regulatory protein SoxS OS=Escherichia coli (strain K12) GN=soxS
BAC0675|merP|tr|O07301|O07301_9PSED Mercuric transport protein periplasmic component OS=Pseudomonas
BAC0107|cusA/ybdE|sp|P38054|CUSA_ECOLI Cation efflux system protein CusA OS=Escherichia coli (strain K
BAC0344|silE|sp|Q9Z4N3|SILE_SALTM Silver-binding protein SilE OS=Salmonella typhimurium GN=silE PE=
BAC0687|merR|tr|Q79BG7|Q79BG7_PSEST MerR OS=Pseudomonas stutzeri GN=merR PE=4 SV=1
BAC0574|arsB|sp|P30329|ARSB_STAAU Arsenical pump membrane protein OS=Staphylococcus aureus GN=ar
BAC0348|silS|sp|Q9ZHD4|SILS_SALTM Probable sensor kinase SilS OS=Salmonella typhimurium GN=silS PE
BAC0686|merR|sp|P13111|MERR_SERMA Mercuric resistance operon regulatory protein OS=Serratia marcesce
BAC0689|merR|tr|Q934S8|Q934S8_THIFE Mer operon regulatory protein OS=Thiobacillus ferrooxidans GN=m
BAC0667|merD|sp|P08654|MERD_SERMA HTH-type transcriptional regulator MerD OS=Serratia marcescens G
BAC0134|dpr/dps|sp|P0CB53|DPS_STRSU DNA protection during starvation protein OS=Streptococcus suis GN
BAC0681|merR2|tr|Q9WWL1|Q9WWL1_BACSR Mercury resistance operon negative regulator MerR2 OS=Bac
BAC0644|corD|sp|Q56017|APAG_SALTY Protein ApaG OS=Salmonella typhimurium (strain LT2 / SGSC1412
BAC0596|baeR|tr|D0ZNE3|D0ZNE3_SALT1 DNA-binding transcriptional regulator BaeR OS=Salmonella typhi
BAC0351|sitC|tr|Q9XCS0|Q9XCS0_SALTM SitC OS=Salmonella typhimurium GN=sitC PE=3 SV=1
BAC0465|znuC/yebM|sp|P0A9X1|ZNUC_ECOLI Zinc import ATP-binding protein ZnuC OS=Escherichia coli (
BAC0442|yfeD|sp|Q56955|YFED_YERPE Chelated iron transport system membrane protein YfeD OS=Yersinia
BAC0309|pcoS|sp|Q47457|PCOS_ECOLX Probable sensor protein PcoS OS=Escherichia coli GN=pcoS PE=3 S
BAC0462|zntR/yhdM|sp|P0ACS5|ZNTR_ECOLI HTH-type transcriptional regulator ZntR OS=Escherichia coli (
BAC0029|chrF|tr|A4UQR2|A4UQR2_9RHIZ ChrF OS=Ochrobactrum tritici GN=chrF PE=4 SV=1
BAC0318|pstS|sp|P0AG82|PSTS_ECOLI Phosphate-binding protein PstS OS=Escherichia coli (strain K12) GN=
BAC0165|fetA/ybbL|sp|P77279|YBBL_ECOLI Uncharacterized ABC transporter ATP-binding protein YbbL OS
BAC0672|merE|tr|Q79BE4|Q79BE4_PSEST Urf1 OS=Pseudomonas stutzeri PE=4 SV=1
BAC0653|merA|tr|Q934S5|Q934S5_THIFE Mercuric ion reductase OS=Thiobacillus ferrooxidans GN=merA PE
BAC0619|copA|tr|Q7WYH1|Q7WYH1_PSEPU CopA OS=Pseudomonas putida GN=copA PE=4 SV=1
BAC0201|kmtR|sp|O53838|KMTR_MYCTU HTH-type transcriptional regulator KmtR OS=Mycobacterium tube
BAC0577|arsB|sp|P74985|ARSB_YEREN Arsenical pump membrane protein OS=Yersinia enterocolitica GN=ar
BAC0625|copA|tr|F4ZCZ9|F4ZCZ9_9XANT Copper resistance protein A OS=Xanthomonas alfalfae subsp. citru
BAC0610|modB|sp|P0AF01|MODB_ECOLI Molybdenum transport system permease protein ModB OS=Escheri
BAC0657|merB|tr|O07303|O07303_9PSED Alkylmercury lyase OS=Pseudomonas sp. K-62 GN=merB PE=3 SV
BAC0343|silC|sp|Q9ZHD2|SILC_SALTM Probable outer membrane lipoprotein SilC OS=Salmonella typhimuriu
BAC0349|sitA|tr|Q9XCS2|Q9XCS2_SALTI Iron transport protein, periplasmic-binding protein OS=Salmonella t
BAC0440|yfeB|sp|Q56953|YFEB_YERPE Chelated iron transport system membrane protein YfeB OS=Yersinia

BAC0135|dpsA|tr|Q8KR86|Q8KR86_BURPE DpsA OS=Burkholderia pseudomallei GN=dpsA PE=3 SV=1
BAC0119|czcA|sp|P13511|CZCA_RALME Cobalt-zinc-cadmium resistance protein CzcA OS=Ralstonia metallic
BAC0304|pcoB|sp|Q47453|PCOB_ECOLX Copper resistance protein B OS=Escherichia coli GN=pcoB PE=4 SV
BAC0108|cusB|sp|P77239|CUSB_ECOLI Cation efflux system protein CusB OS=Escherichia coli (strain K12) G
BAC0620|copA|sp|P32113|COPA_ENTHA Probable copper-importing P-type ATPase A OS=Enterococcus hirae
BAC0647|mdtC|tr|D0ZNE0|D0ZNE0_SALT1 Multidrug resistance protein MdtC OS=Salmonella typhimurium (s
BAC0641|corA|sp|P0A2R8|CORA_SALTY Magnesium transport protein CorA OS=Salmonella typhimurium (st
BAC0279|nirD|tr|Q6RUG0|Q6RUG0_KLEOX NirD OS=Klebsiella oxytoca GN=nirD PE=4 SV=1
BAC0039|baeR|sp|P69228|BAER_ECOLI Transcriptional regulatory protein BaeR OS=Escherichia coli (strain K
BAC0179|gesB|tr|Q8ZRG9|Q8ZRG9_SALTY Putative cation efflux system protein OS=Salmonella typhimurium
BAC0469|zupT/ygiE|sp|P0A8H3|ZUPT_ECOLI Zinc transporter ZupT OS=Escherichia coli (strain K12) GN=zu
BAC0583|arsC|sp|P52147|ARSC2_ECOLX Arsenate reductase OS=Escherichia coli GN=arsC PE=3 SV=1
BAC0057|cadD|tr|Q7A320|Q7A320_STAAN CadD OS=Staphylococcus aureus (strain N315) GN=cadD PE=4 S
BAC0434|ychH|sp|P0AB49|YCHH_ECOLI Uncharacterized protein YchH OS=Escherichia coli (strain K12) GN
BAC0358|oscA|tr|B6CM35|B6CM35_9PSED Putative uncharacterized protein oscA OS=Pseudomonas corrugata
BAC0023|aioS/aoxS|tr|Q2VGB2|Q2VGB2_RHIRD Putative sensor histidine kinase OS=Rhizobium radiobacter C
BAC0467|zraR/hydH|sp|P14375|ZRAR_ECOLI Transcriptional regulatory protein ZraR OS=Escherichia coli (str
BAC0565|actR|sp|A6UEL7|ACTR_SINMW Acid tolerance regulatory protein ActR OS=Sinorhizobium medicae
BAC0388|terC|sp|P18780|TERC_ALCSP Tellurium resistance protein TerC OS=Alcaligenes sp. GN=terC PE=3
BAC0105|cueR/ybbI|sp|P0A9G4|CUER_ECOLI HTH-type transcriptional regulator CueR OS=Escherichia coli (
BAC0392|terZ|sp|Q52353|TERZ_SERMA Tellurium resistance protein TerZ OS=Serratia marcescens GN=terZ P
BAC0110|cusF/cusX|sp|P77214|CUSF_ECOLI Cation efflux system protein CusF OS=Escherichia coli (strain K
BAC0082|copL|tr|Q5YKV8|Q5YKV8_9XANT CopL OS=Xanthomonas perforans GN=copL PE=4 SV=1
BAC0251|mntH/yfeP|sp|P0A769|MNTH_ECOLI Divalent metal cation transporter MntH OS=Escherichia coli (s
BAC0199|klaB/telA/kilB|sp|Q52328|KLAB_ECOLX Protein KlaB OS=Escherichia coli GN=klaB PE=3 SV=1
BAC0455|ziaA|sp|Q59998|ATZN_SYNY3 Zinc-transporting ATPase OS=Synechocystis sp. (strain PCC 6803 / K
BAC0678|merP|tr|O66016|O66016_PSEST MerP OS=Pseudomonas stutzeri GN=merP PE=4 SV=1
BAC0308|pcoR|sp|Q47456|PCOR_ECOLX Transcriptional regulatory protein PcoR OS=Escherichia coli GN=pc
BAC0344|silE|sp|Q9Z4N3|SILE_SALTM Silver-binding protein SilE OS=Salmonella typhimurium GN=silE PE=
BAC0384|tehA|sp|P25396|TEHA_ECOLI Tellurite resistance protein TehA OS=Escherichia coli (strain K12) GN
BAC0357|recG|tr|B5L350|B5L350_9PSED ATP-dependent DNA helicase (Fragment) OS=Pseudomonas corruga
BAC0224|merA|sp|P16171|MERA_BACCE Mercuric reductase OS=Bacillus cereus GN=merA PE=1 SV=1
BAC0119|czcA|sp|P13511|CZCA_RALME Cobalt-zinc-cadmium resistance protein CzcA OS=Ralstonia metallic
BAC0640|copD|tr|C6FFR7|C6FFR7_PSEFL CopD OS=Pseudomonas fluorescens GN=copD PE=4 SV=1
BAC0356|recG|tr|Q9HTL3|Q9HTL3_PSEAE ATP-dependent DNA helicase RecG OS=Pseudomonas aeruginosa
BAC0168|fptA|sp|P42512|FPTA_PSEAE Fe(3+)-pyochelin receptor OS=Pseudomonas aeruginosa (strain ATCC
BAC0621|copA|tr|F4ZBX3|F4ZBX3_XANCI CopA OS=Xanthomonas citri subsp. citri GN=copA PE=4 SV=1
BAC0026|chrA|tr|A4UQR4|A4UQR4_9RHIZ Chromate transporter OS=Ochrobactrum tritici GN=chrA PE=4 SV
BAC0341|silA|sp|Q9ZHC9|SILA_SALTM Putative cation efflux system protein SilA OS=Salmonella typhimuriu
BAC0343|silC|sp|Q9ZHD2|SILC_SALTM Probable outer membrane lipoprotein SilC OS=Salmonella typhimuriu
BAC0568|actP|sp|Q9X5V3|ATCU_RHILV Copper-transporting P-type ATPase OS=Rhizobium leguminosarum b
BAC0652|merA|tr|O66017|O66017_PSEST MerA OS=Pseudomonas stutzeri GN=merA PE=4 SV=1
BAC0566|actS|tr|Q52912|Q52912_9RHIZ Histidine protein kinase OS=Sinorhizobium medicae GN=actS PE=4 S
BAC0179|gesB|tr|Q8ZRG9|Q8ZRG9_SALTY Putative cation efflux system protein OS=Salmonella typhimurium
BAC0066|chrF|tr|Q5NUZ7|Q5NUZ7_RALME ChrF, regulatory protein, involved in Chromate resistance OS=Ra
BAC0573|arsB|sp|P45946|ARSB_BACSU Arsenite resistance protein ArsB OS=Bacillus subtilis (strain 168) GN

BAC0459|zitB/ybgR|sp|P75757|ZITB_ECOLI Zinc transporter ZitB OS=Escherichia coli (strain K12) GN=zitB P
BAC0031|arsB|sp|P08691|ARSB1_ECOLX Arsenical pump membrane protein OS=Escherichia coli GN=arsB PE
BAC0596|baeR|tr|D0ZNE3|D0ZNE3_SALT1 DNA-binding transcriptional regulator BaeR OS=Salmonella typhi
BAC0681|merR2|tr|Q9WWL1|Q9WWL1_BACSR Mercury resistance operon negative regulator MerR2 OS=Bac
BAC0563|acrD|tr|Q8ZN77|Q8ZN77_SALTY RND family aminoglycoside/multidrug efflux pump OS=Salmonell
BAC0694|merT-P|tr|H6WCN3|H6WCN3_9FLAO MerT-P OS=Tenacibaculum discolor GN=merT-P PE=4 SV=
BAC0666|merD|tr|O66018|O66018_PSEST MerD OS=Pseudomonas stutzeri GN=merD PE=4 SV=1
BAC0098|ctpC|sp|P0A502|CTPC_MYCTU Probable manganese/zinc-exporting P-type ATPase OS=Mycobacteri
BAC0650|merA|tr|O08449|O08449_9PSED Mercuric reductase OS=Pseudomonas sp. K-62 GN=merA PE=4 SV
BAC0351|sitC|tr|Q9XCS0|Q9XCS0_SALTM SitC OS=Salmonella typhimurium GN=sitC PE=3 SV=1
BAC0498|ideR|sp|P0A672|IDER_MYCTU Iron-dependent repressor IdeR OS=Mycobacterium tuberculosis GN=
BAC0386|terA|sp|P18778|TERA_ALCSP Tellurium resistance protein TerA OS=Alcaligenes sp. GN=terA PE=4
BAC0331|rcnB/yohN|sp|P64534|RCNB_ECOLI Nickel/cobalt homeostasis protein RcnB OS=Escherichia coli (st
BAC0058|cadR|tr|Q93TP7|Q93TP7_PSEPU CadR OS=Pseudomonas putida GN=cadR PE=4 SV=1
BAC0012|actP|sp|Q9X5X3|ATCU_SINMW Copper-transporting P-type ATPase OS=Sinorhizobium medicae (st
BAC0034|arsH|tr|E8PS81|E8PS81_YERPE Arsenic resistance protein ArsH OS=Yersinia pestis Java 9 GN=arsH
BAC0441|yfeC|sp|Q56954|YFEC_YERPE Chelated iron transport system membrane protein YfeC OS=Yersinia p
BAC0133|dnaK|sp|P0A5B9|DNAK_MYCTU Chaperone protein DnaK OS=Mycobacterium tuberculosis GN=dn
BAC0667|merD|sp|P08654|MERD_SERMA HTH-type transcriptional regulator MerD OS=Serratia marcescens G
BAC0647|mdtC|tr|D0ZNE0|D0ZNE0_SALT1 Multidrug resistance protein MdtC OS=Salmonella typhimurium (s
BAC0584|arsC|sp|O50595|ARSC_ACIMA Arsenate reductase OS=Acidiphilium multivorum (strain DSM 11245
BAC0685|merR|tr|H6WCN2|H6WCN2_9FLAO MerR OS=Tenacibaculum discolor GN=merR PE=4 SV=1
BAC0572|arsA|sp|P08690|ARSA1_ECOLX Arsenical pump-driving ATPase OS=Escherichia coli GN=arsA PE=
BAC0656|merB3|tr|Q7DHE7|Q7DHE7_BACCE Organomercurial lyase enzyme OS=Bacillus cereus GN=merB3
BAC0609|modA|sp|P37329|MODA_ECOLI Molybdate-binding periplasmic protein OS=Escherichia coli (strain K
BAC0124|czcP|tr|Q1LAJ7|Q1LAJ7_RALME CzcP cation efflux P1-ATPase OS=Ralstonia metallidurans (strain C
BAC0345|silF|tr|Q9ZHD1|Q9ZHD1_SALTM Uncharacterized protein OS=Salmonella typhimurium GN=ORF96
BAC0240|mexI|tr|Q9HWH4|Q9HWH4_PSEAE Probable Resistance-Nodulation-Cell Division (RND) efflux tran
BAC0673|merE|sp|P06690|MERE_PSEAI Uncharacterized mercuric resistance protein MerE OS=Pseudomonas a
BAC0165|fetA/ybbL|sp|P77279|YBBL_ECOLI Uncharacterized ABC transporter ATP-binding protein YbbL OS
BAC0231|merP|sp|P13113|MERP_SERMA Mercuric transport protein periplasmic component OS=Serratia marc
BAC0389|terD|sp|P18781|TERD_ALCSP Tellurium resistance protein TerD OS=Alcaligenes sp. GN=terD PE=3
BAC0686|merR|sp|P13111|MERR_SERMA Mercuric resistance operon regulatory protein OS=Serratia marcesce
BAC0610|modB|sp|P0AF01|MODB_ECOLI Molybdenum transport system permease protein ModB OS=Escheri
BAC0114|cutC|sp|P67826|CUTC_ECOLI Copper homeostasis protein CutC OS=Escherichia coli (strain K12) GN
BAC0468|zraS/hydG|sp|P14377|ZRAS_ECOLI Sensor protein ZraS OS=Escherichia coli (strain K12) GN=zraS P
BAC0439|yfeA|sp|Q56952|YFEA_YERPE Periplasmic chelated iron-binding protein YfeA OS=Yersinia pestis G
BAC0485|pmrC|tr|Q70FH1|Q70FH1_PECCC Putative cytoplasmic membrane protein pmrC OS=Pectobacterium
BAC0169|fpvA|sp|P48632|FPVA_PSEAE Ferripyoverdine receptor OS=Pseudomonas aeruginosa (strain ATCC
BAC0121|czcC|sp|P13509|CZCC_RALME Cobalt-zinc-cadmium resistance protein CzcC OS=Ralstonia metallid
BAC0348|silS|sp|Q9ZHD4|SILS_SALTM Probable sensor kinase SilS OS=Salmonella typhimurium GN=silS PE
BAC0682|merR1|sp|P22853|MERR_BACCE Mercuric resistance operon regulatory protein OS=Bacillus cereus G
BAC0136|dsbA|sp|P0AEG4|DSBA_ECOLI Thiol:disulfide interchange protein DsbA OS=Escherichia coli (strain
BAC0107|cusA/ybdE|sp|P38054|CUSA_ECOLI Cation efflux system protein CusA OS=Escherichia coli (strain K
BAC0228|merF|tr|Q2QCN0|Q2QCN0_9PSED MerF OS=Pseudomonas sp. CT14 GN=merF PE=4 SV=1
BAC0035|arsM|tr|Q6N3Y0|Q6N3Y0_RHOPA UbiE/COQ5 methyltransferase OS=Rhodopseudomonas palustris

BAC0657|merB|tr|O07303|O07303_9PSED Alkylmercury lyase OS=Pseudomonas sp. K-62 GN=merB PE=3 SV
BAC0303|pcoA|sp|Q47452|PCOA_ECOLX Copper resistance protein A OS=Escherichia coli GN=pcoA PE=3 S
BAC0167|fieF/yiip|sp|P69380|FIEF_ECOLI Ferrous-iron efflux pump FieF OS=Escherichia coli (strain K12) GN
BAC0585|arsC|sp|P74984|ARSC_YEREN Arsenate reductase OS=Yersinia enterocolitica GN=arsC PE=3 SV=1
BAC0688|merR2|tr|Q79B70|Q79B70_PSEST Organomercurial resistance regulatory protein OS=Pseudomonas s
BAC0271|nikB|sp|P33591|NIKB_ECOLI Nickel transport system permease protein NikB OS=Escherichia coli (s
BAC0670|merE|tr|Q52104|Q52104_9ZZZZ Uncharacterized protein OS=Plasmid pDU1358 PE=4 SV=1
BAC0687|merR|tr|Q79BG7|Q79BG7_PSEST MerR OS=Pseudomonas stutzeri GN=merR PE=4 SV=1
BAC0649|merA|tr|E3VST6|E3VST6_9FLAO MerA OS=Tenacibaculum discolor GN=merA PE=3 SV=1
BAC0332|rcnR/yohL|sp|P64530|RCNR_ECOLI Transcriptional repressor RcnR OS=Escherichia coli (strain K12
BAC0470|zur/yjbK|sp|P0AC51|ZUR_ECOLI Zinc uptake regulation protein OS=Escherichia coli (strain K12) G
BAC0571|arsA|sp|O50593|ARSA_ACIMA Arsenical pump-driving ATPase OS=Acidiphilium multivorum (strai
BAC0317|pstC|sp|P0AGH8|PSTC_ECOLI Phosphate transport system permease protein PstC OS=Escherichia co
BAC0086|corA|sp|P0ABI4|CORA_ECOLI Magnesium transport protein CorA OS=Escherichia coli (strain K12)
BAC0106|cuiD|sp|Q8ZRS2|CUEO_SALTY Blue copper oxidase CueO OS=Salmonella typhimurium (strain LT2
BAC0548|chrA1|sp|P17551|CHRA1_RALME Chromate transport protein OS=Ralstonia metallidurans (strain C
BAC0266|ncrY|tr|Q06VT0|Q06VT0_9BACT NcrY OS=Leptospirillum ferriphilum GN=ncrY PE=4 SV=1
BAC0276|nirA|tr|Q6RUG3|Q6RUG3_KLEOX NirA OS=Klebsiella oxytoca GN=nirA PE=4 SV=1
BAC0077|copA|sp|P12374|COPA_PSEUB Copper resistance protein A OS=Pseudomonas syringae pv. tomato G
BAC0631|copC|tr|F4ZBX9|F4ZBX9_XANCI CopC OS=Xanthomonas citri subsp. citri GN=copC PE=4 SV=1
BAC0115|cutE/lnt|sp|P23930|LNT_ECOLI Apolipoprotein N-acyltransferase OS=Escherichia coli (strain K12) G
BAC0576|arsB|sp|P52146|ARSB2_ECOLX Arsenical pump membrane protein OS=Escherichia coli GN=arsB P
BAC0597|baeS|tr|D0ZNE2|D0ZNE2_SALT1 Signal transduction histidine-protein kinase BaeS OS=Salmonella t
BAC0639|copC|tr|C6FFR6|C6FFR6_PSEFL CopC OS=Pseudomonas fluorescens GN=copC PE=4 SV=1
BAC0116|cutF/nlpE|sp|P40710|NLPE_ECOLI Lipoprotein NlpE OS=Escherichia coli (strain K12) GN=nlpE PE
BAC0020|aioA/aoxB|sp|Q8GGJ6|AIOA_HERAR Arsenite oxidase subunit AioA OS=Herminiimonas arsenicox
BAC0371|soxS|sp|P0A9E2|SOXS_ECOLI Regulatory protein SoxS OS=Escherichia coli (strain K12) GN=soxS
BAC0368|sodA|sp|P00448|SODM_ECOLI Superoxide dismutase [Mn] OS=Escherichia coli (strain K12) GN=so
BAC0627|copB|tr|F4ZD00|F4ZD00_9XANT Copper resistance protein B OS=Xanthomonas alfalfae subsp. citru
BAC0334|robA|sp|P0ACI0|ROB_ECOLI Right origin-binding protein OS=Escherichia coli (strain K12) GN=rob
BAC0113|cutA|sp|P69488|CUTA_ECOLI Divalent-cation tolerance protein CutA OS=Escherichia coli (strain K1
BAC0067|cinA|tr|Q9I036|Q9I036_PSEAE Uncharacterized protein OS=Pseudomonas aeruginosa (strain ATCC 1
BAC0632|copD|sp|P12377|COPD_PSEUB Copper resistance protein D OS=Pseudomonas syringae pv. tomato G
BAC0130|czrA|tr|Q9RLI8|Q9RLI8_PSEAI CzrA protein OS=Pseudomonas aeruginosa GN=czrA PE=4 SV=1
BAC0582|arsC|sp|P08692|ARSC1_ECOLX Arsenate reductase OS=Escherichia coli GN=arsC PE=1 SV=1
BAC0108|cusB|sp|P77239|CUSB_ECOLI Cation efflux system protein CusB OS=Escherichia coli (strain K12) G
BAC0642|mgtA|sp|P36640|ATMA_SALTY Magnesium-transporting ATPase, P-type 1 OS=Salmonella typhimu
BAC0452|yqjH|sp|Q46871|YQJH_ECOLI NADPH-dependent ferric-chelate reductase OS=Escherichia coli (strai
BAC0087|mgtA|sp|P0ABB8|ATMA_ECOLI Magnesium-transporting ATPase, P-type 1 OS=Escherichia coli (str
BAC0027|chrB|tr|A4UQR5|A4UQR5_9RHIZ ChrB OS=Ochrobactrum tritici GN=chrB PE=4 SV=1
BAC0299|pbrB/pbrC|tr|Q58AJ7|Q58AJ7_RALME Lipoprotein signal peptidase OS=Ralstonia metallidurans (str
BAC0040|baeS|sp|P30847|BAES_ECOLI Signal transduction histidine-protein kinase BaeS OS=Escherichia coli
BAC0611|modC|sp|P09833|MODC_ECOLI Molybdenum import ATP-binding protein ModC OS=Escherichia co
BAC0385|tehB|sp|P25397|TEHB_ECOLI Tellurite methyltransferase OS=Escherichia coli (strain K12) GN=tehB
BAC0003|acn|tr|O53166|O53166_MYCTU Aconitate hydratase OS=Mycobacterium tuberculosis H37Rv GN=R
BAC0608|modE|sp|P0A9G8|MODE_ECOLI Transcriptional regulator ModE OS=Escherichia coli (strain K12) G

BAC0048|bfrA|sp|P63697|BFR_MYCTU Bacterioferritin OS=Mycobacterium tuberculosis GN=bfr PE=1 SV=1
BAC0100|ctpG|sp|P63689|CTPG_MYCTU Probable cation-transporting ATPase G OS=Mycobacterium tubercul
BAC0024|aioX/aoxX|tr|G8XNW6|G8XNW6_RHIRD AioX (Fragment) OS=Rhizobium radiobacter GN=aioX P
BAC0252|mntP/yebN|sp|P76264|MNTP_ECOLI Probable manganese efflux pump MntP OS=Escherichia coli (s
BAC0063|chrA|sp|P14285|CHRA_PSEAI Chromate transport protein OS=Pseudomonas aeruginosa GN=chrA P
BAC0577|arsB|sp|P74985|ARSB_YEREN Arsenical pump membrane protein OS=Yersinia enterocolitica GN=a
BAC0665|merD|tr|O66022|O66022_PSEST Mercury operon coregulator protein OS=Pseudomonas stutzeri GN=
BAC0112|cusS|sp|P77485|CUSS_ECOLI Sensor kinase CusS OS=Escherichia coli (strain K12) GN=cusS PE=1
BAC0350|sitB|tr|Q9XCS1|Q9XCS1_SALTM SitB OS=Salmonella typhimurium GN=sitB PE=3 SV=1
BAC0540|nfsA|sp|P17117|NFSA_ECOLI Oxygen-insensitive NADPH nitroreductase OS=Escherichia coli (strai
BAC0695|merT|sp|P13112|MERT_SERMA Mercuric transport protein OS=Serratia marcescens GN=merT PE=4
BAC0690|merT|tr|Q934S7|Q934S7_THIFE Mercuric ion transport protein OS=Thiobacillus ferrooxidans GN=m
BAC0315|pstA|sp|P07654|PSTA_ECOLI Phosphate transport system permease protein PstA OS=Escherichia co
BAC0272|nikC|sp|P0AFA9|NIKC_ECOLI Nickel transport system permease protein NikC OS=Escherichia coli (
BAC0346|silP|sp|Q9ZHC7|SILP_SALTM Silver exporting P-type ATPase OS=Salmonella typhimurium GN=sil
BAC0265|ncrC|tr|D5CKG5|D5CKG5_ENTCC Nickel-resistant membrane protein-like protein NcrC OS=Entero
BAC0570|actP|tr|D5AU53|D5AU53_RHOCB Cation/acetate symporter ActP-1 OS=Rhodobacter capsulatus (stra
BAC0432|ybtP|tr|Q9R7V3|Q9R7V3_YERPE Lipoprotein inner membrane ABC-transporter OS=Yersinia pestis
BAC0395|trgB|tr|O07841|O07841_RHOSH Tellurite resistance protein OS=Rhodobacter sphaeroides GN=trgB
BAC0629|copB|sp|O30085|COPB_ARCFU Copper-exporting P-type ATPase B OS=Archaeoglobus fulgidus (str
BAC0541|yieF|sp|P0AGE6|YIEF_ECOLI Uncharacterized protein YieF OS=Escherichia coli (strain K12) GN=y
BAC0316|pstB|sp|P0AAH0|PSTB_ECOLI Phosphate import ATP-binding protein PstB OS=Escherichia coli (str
BAC0039|baeR|sp|P69228|BAER_ECOLI Transcriptional regulatory protein BaeR OS=Escherichia coli (strain K
BAC0683|merR1|tr|O07300|O07300_9PSED Mercuric resistance operon regulatory protein OS=Pseudomonas sp
BAC0181|glpF|sp|P0AER0|GLPF_ECOLI Glycerol uptake facilitator protein OS=Escherichia coli (strain K12) G
BAC0307|pcoE|sp|Q47459|PCOE_ECOLX Probable copper-binding protein PcoE OS=Escherichia coli GN=pco
BAC0463|znuA/yebL|sp|P39172|ZNUA_ECOLI High-affinity zinc uptake system protein ZnuA OS=Escherichia
BAC0451|yodD|sp|P64519|YODD_ECOLI Uncharacterized protein YodD OS=Escherichia coli (strain K12) GN
BAC0461|zntA/yhhO|sp|P37617|ATZN_ECOLI Lead, cadmium, zinc and mercury-transporting ATPase OS=Esc
BAC0229|merG|tr|O07302|O07302_9PSED Mercuric resistance protein OS=Pseudomonas sp. K-62 GN=merG P
BAC0648|merA|sp|P08662|MERA_SERMA Mercuric reductase (Fragments) OS=Serratia marcescens GN=merA
BAC0103|cueO|sp|P36649|CUEO_ECOLI Blue copper oxidase CueO OS=Escherichia coli (strain K12) GN=cue
BAC0030|arsA|sp|P52145|ARSA2_ECOLX Arsenical pump-driving ATPase OS=Escherichia coli GN=arsA PE=
BAC0183|golT|tr|Q8ZRG7|Q8ZRG7_SALTY Putative cation transport ATPase OS=Salmonella typhimurium (st
BAC0464|znuB/yebI|sp|P39832|ZNUB_ECOLI High-affinity zinc uptake system membrane protein ZnuB OS=E
BAC0440|yfeB|sp|Q56953|YFEB_YERPE Chelated iron transport system membrane protein YfeB OS=Yersinia
BAC0691|merT|tr|Q52397|Q52397_PSEST Mercury transport protein OS=Pseudomonas stutzeri GN=merT PE=
BAC0083|copR|sp|Q02540|COPR_PSEUB Transcriptional activator protein CopR OS=Pseudomonas syringae p
BAC0646|mdtB|tr|D0ZND9|D0ZND9_SALT1 Multidrug resistance protein MdtB OS=Salmonella typhimurium
BAC0071|cmtR|sp|P67731|CMTR_MYCTU HTH-type transcriptional regulator CmtR OS=Mycobacterium tube
BAC0355|ruvB|sp|Q51426|RUVB_PSEAE Holliday junction ATP-dependent DNA helicase RuvB OS=Pseudom
BAC0620|copA|sp|P32113|COPA_ENTHA Probable copper-importing P-type ATPase A OS=Enterococcus hira
BAC0347|silR|sp|Q9ZHD3|SILR_SALTM Probable transcriptional regulatory protein SilR OS=Salmonella typh
BAC0458|zipB|tr|Q7WJT8|Q7WJT8_BORBR Putative membrane protein OS=Bordetella bronchiseptica (strain
BAC0302|pbrT|tr|Q5GR69|Q5GR69_ALCXX Lead uptake protein PbrT OS=Alcaligenes xylosoxydans xylosox
BAC0588|arsR|tr|P74986|P74986_YEREN Arsenite inducible repressor OS=Yersinia enterocolitica GN=arsR P

BAC0659|merB|sp|P08664|MERB_SERMA Alkylmercury lyase OS=Serratia marcescens GN=merB PE=3 SV=1
BAC0270|nikA|sp|P33590|NIKA_ECOLI Nickel-binding periplasmic protein OS=Escherichia coli (strain K12) G
BAC0638|copR|tr|C6FFR4|C6FFR4_PSEFL CopR OS=Pseudomonas fluorescens GN=copR PE=4 SV=1
BAC0049|bhsA/ycfR/comC|sp|P0AB40|BHSA_ECOLI Multiple stress resistance protein BhsA OS=Escherichia c
BAC0625|copA|tr|F4ZCZ9|F4ZCZ9_9XANT Copper resistance protein A OS=Xanthomonas alfalfae subsp. citru
BAC0383|tcrB|tr|Q8VPE6|Q8VPE6_ENTFC TcrB OS=Enterococcus faecium GN=tcrB PE=3 SV=1
BAC0263|ncrA|tr|Q06VT3|Q06VT3_9BACT NcrA OS=Leptospirillum ferriphilum GN=ncrA PE=4 SV=1
BAC0643|corB|tr|Q9X621|Q9X621_SALTM CorB OS=Salmonella typhimurium GN=corB PE=4 SV=1
BAC0255|mreA|tr|Q88IN0|Q88IN0_PSEPK Putative uncharacterized protein OS=Pseudomonas putida (strain KT
BAC0059|cadX|tr|A7LHQ4|A7LHQ4_STRSL CadX OS=Streptococcus salivarius GN=cadX PE=4 SV=1
BAC0465|znuC/yebM|sp|P0A9X1|ZNUC_ECOLI Zinc import ATP-binding protein ZnuC OS=Escherichia coli (
BAC0079|copB|sp|P05425|COPB_ENTHA Copper-exporting P-type ATPase B OS=Enterococcus hirae (strain A
BAC0628|copB|tr|F4ZBX4|F4ZBX4_XANCI CopB OS=Xanthomonas citri subsp. citri GN=copB PE=4 SV=1
BAC0111|cusR/ylcA|sp|P0ACZ8|CUSR_ECOLI Transcriptional regulatory protein CusR OS=Escherichia coli (st
BAC0318|pstS|sp|P0AG82|PSTS_ECOLI Phosphate-binding protein PstS OS=Escherichia coli (strain K12) GN=
BAC0490|G2alt|tr|B0FSM1|B0FSM1_9BACI 7-cyano-7-deazaguanine synthase OS=Anoxybacillus gonensis GN
BAC0135|dpsA|tr|Q8KR86|Q8KR86_BURPE DpsA OS=Burkholderia pseudomallei GN=dpsA PE=3 SV=1
BAC0190|hmrR|sp|Q9X5X4|HMRR_SINMW HTH-type transcriptional regulator HmrR OS=Sinorhizobium med
BAC0391|terW|sp|P75010|TERW_SERMA Tellurium resistance protein TerW OS=Serratia marcescens GN=terW
BAC0612|perO|tr|D5AQ60|D5AQ60_RHOCB Divalent ion symporter OS=Rhodobacter capsulatus (strain ATCC
BAC0267|nczA|tr|B8GZE9|B8GZE9_CAUCN Cobalt-zinc-cadmium resistance protein czcA OS=Caulobacter cre
BAC0697|merT|tr|Q7DHE5|Q7DHE5_BACCE Mercury transport protein OS=Bacillus cereus GN=merT PE=4 S
BAC0390|terE|sp|P18782|TERE_ALCSP Tellurium resistance protein TerE OS=Alcaligenes sp. GN=terE PE=3 S
BAC0162|fbpC|sp|P44513|FBPC2_HAEIN Fe(3+) ions import ATP-binding protein FbpC 2 OS=Haemophilus in
BAC0549|nccA|sp|Q44586|NCCA_ALCXX Nickel-cobalt-cadmium resistance protein NccA OS=Alcaligenes xy
BAC0698|ncrA|tr|Q1KLR2|Q1KLR2_SERMA NcrA OS=Serratia marcescens GN=ncrA PE=4 SV=1
BAC0088|corC|sp|P0A2L3|CORC_SALTY Magnesium and cobalt efflux protein CorC OS=Salmonella typhimur
BAC0589|arsR|sp|P15905|ARSR1_ECOLX Arsenical resistance operon repressor OS=Escherichia coli GN=arsR
BAC0641|corA|sp|P0A2R8|CORA_SALTY Magnesium transport protein CorA OS=Salmonella typhimurium (st
BAC0137|dsbB|sp|P0A6M2|DSBB_ECOLI Disulfide bond formation protein B OS=Escherichia coli (strain K12)
BAC0056|cadC|sp|P20047|CADC_STAAU Cadmium resistance transcriptional regulatory protein CadC OS=Stap
BAC0707|sodB|sp|P0AGD3|SODF_ECOLI Superoxide dismutase [Fe] OS=Escherichia coli (strain K12) GN=so
BAC0166|fetB/ybbM|sp|P77307|YBBM_ECOLI UPF0014 inner membrane protein YbbM OS=Escherichia coli (
BAC0264|ncrB|tr|Q06VT2|Q06VT2_9BACT NcrB OS=Leptospirillum ferriphilum GN=ncrB PE=4 SV=1
BAC0442|yfeD|sp|Q56955|YFED_YERPE Chelated iron transport system membrane protein YfeD OS=Yersinia
BAC0101|ctpV|sp|P77894|CTPV_MYCTU Probable copper-exporting P-type ATPase V OS=Mycobacterium tub
BAC0672|merE|tr|Q79BE4|Q79BE4_PSEST Urf1 OS=Pseudomonas stutzeri PE=4 SV=1
BAC0342|silB|sp|Q9ZHD0|SILB_SALTM Putative membrane fusion protein SilB OS=Salmonella typhimurium
BAC0693|merT|tr|Q79F00|Q79F00_9PSED Mercuric transport protein OS=Pseudomonas sp. K-62 GN=merT PE
BAC0684|merR2|tr|Q7DKL2|Q7DKL2_9PSED MerR2 OS=Pseudomonas sp. K-62 GN=merR2 PE=4 SV=1
BAC0300|pbrD|tr|Q1LAL8|Q1LAL8_RALME PbrD, Pb(II) binding protein involved in Pb(II) resistance OS=Ra
BAC0163|fecD|sp|P15029|FECD_ECOLI Fe(3+) dicitrate transport system permease protein FecD OS=Escherich
BAC0462|zntR/yhdM|sp|P0ACS5|ZNTR_ECOLI HTH-type transcriptional regulator ZntR OS=Escherichia coli (
BAC0619|copA|tr|Q7WYH1|Q7WYH1_PSEPU CopA OS=Pseudomonas putida GN=copA PE=4 SV=1
BAC0689|merR|tr|Q934S8|Q934S8_THIFE Mer operon regulatory protein OS=Thiobacillus ferrooxidans GN=m
BAC0482|dmeF|tr|Q1MJL2|Q1MJL2_RHIL3 Putative cation efflux system protein OS=Rhizobium leguminosaru

BAC0445|ygiW|sp|P0ADU5|YGIW_ECOLI Protein YgiW OS=Escherichia coli (strain K12) GN=ygiW PE=1 S
BAC0102|cueA|tr|Q8KWW2|Q8KWW2_PSEPU Copper transporter OS=Pseudomonas putida GN=cueA PE=3 S
BAC0305|pcoC|sp|Q47454|PCOC_ECOLX Copper resistance protein C OS=Escherichia coli GN=pcoC PE=1 S
BAC0312|pitA|sp|P0AFJ7|PITA_ECOLI Low-affinity inorganic phosphate transporter 1 OS=Escherichia coli (st
BAC0022|aioR/aoxR|tr|Q2VGB1|Q2VGB1_RHIRD Putative transcriptional regulator OS=Rhizobium radiobacte
BAC0587|arsD|sp|P52148|ARSD2_ECOLX Arsenical resistance operon trans-acting repressor ArsD OS=Escheri
BAC0349|sitA|tr|Q9XCS2|Q9XCS2_SALTI Iron transport protein, periplasmic-binding protein OS=Salmonella t
BAC0675|merP|tr|O07301|O07301_9PSED Mercuric transport protein periplasmic component OS=Pseudomonas
BAC0076|comR/ycfQ|sp|P75952|COMR_ECOLI HTH-type transcriptional repressor ComR OS=Escherichia coli
BAC0594|arsR|sp|P37309|ARSR_ECOLI Arsenical resistance operon repressor OS=Escherichia coli (strain K12)
BAC0457|zinT/yodA|sp|P76344|ZINT_ECOLI Metal-binding protein ZinT OS=Escherichia coli (strain K12) GN
BAC0164|fecE|sp|P15031|FECE_ECOLI Fe(3+) dicitrate transport ATP-binding protein FecE OS=Escherichia co
BAC0651|merA|sp|P0A0E5|MERA_STAAU Mercuric reductase OS=Staphylococcus aureus GN=merA PE=3 S
BAC0254|mrdH|tr|Q88IN1|Q88IN1_PSEPK Membrane protein, putative OS=Pseudomonas putida (strain KT244
BAC0134|dpr/dps|sp|P0CB53|DPS_STRSU DNA protection during starvation protein OS=Streptococcus suis G
BAC0306|pcoD|sp|Q47455|PCOD_ECOLX Copper resistance protein D OS=Escherichia coli GN=pcoD PE=3 S
BAC0273|nikD|sp|P33593|NIKD_ECOLI Nickel import ATP-binding protein NikD OS=Escherichia coli (strain
BAC0253|mntR|sp|P0A9F1|MNTR_ECOLI Transcriptional regulator MntR OS=Escherichia coli (strain K12) G
BAC0591|arsR|sp|P52144|ARSR2_ECOLX Arsenical resistance operon repressor OS=Escherichia coli GN=arsR
BAC0653|merA|tr|Q934S5|Q934S5_THIFE Mercuric ion reductase OS=Thiobacillus ferrooxidans GN=merA PE
BAC0538|chrR|tr|Q7BD45|Q7BD45_PSEPU Chromate reductase OS=Pseudomonas putida GN=chrR PE=4 SV=
BAC0309|pcoS|sp|Q47457|PCOS_ECOLX Probable sensor protein PcoS OS=Escherichia coli GN=pcoS PE=3 S
BAC0203|cnrA|sp|P37972|CNRA_RALME Nickel and cobalt resistance protein CnrA OS=Ralstonia metallidura
BAC0692|merT|tr|Q79BG6|Q79BG6_PSEST MerT OS=Pseudomonas stutzeri GN=merT PE=4 SV=1
BAC0298|pbrA|tr|Q58AJ6|Q58AJ6_RALME P-type ATPase involved in Pb(II) resistance PbrA OS=Ralstonia m
BAC0387|terB|sp|P18779|TERB_ALCSP Tellurium resistance protein TerB OS=Alcaligenes sp. GN=terB PE=4
BAC0637|copS|tr|C6FFR5|C6FFR5_PSEFL CopS OS=Pseudomonas fluorescens GN=copS PE=4 SV=1
BAC0029|chrF|tr|A4UQR2|A4UQR2_9RHIZ ChrF OS=Ochrobactrum tritici GN=chrF PE=4 SV=1
BAC0109|cusC/ylcB|sp|P77211|CUSC_ECOLI Cation efflux system protein CusC OS=Escherichia coli (strain K
BAC0679|merP|tr|O66047|O66047_PSEST Mercury transport protein OS=Pseudomonas stutzeri GN=merP PE=
BAC0285|nreB|tr|F0KND8|F0KND8_ACICP NrsD, nreB nickel permease involved in nickel and cobalt toleranc
BAC0233|merT|sp|P94185|MERT_ALCSP Mercuric transport protein OS=Alcaligenes sp. GN=merT PE=3 SV=
BAC0645|mdtA|tr|D0ZND8|D0ZND8_SALT1 Multidrug resistance protein MdtA OS=Salmonella typhimurium
BAC0578|arsB|tr|O50594|O50594_ACIMU ArsB OS=Acidiphilium multivorum GN=arsB PE=4 SV=1
BAC0304|pcoB|sp|Q47453|PCOB_ECOLX Copper resistance protein B OS=Escherichia coli GN=pcoB PE=4 S
BAC0138|dsbC|sp|P0AEG6|DSBC_ECOLI Thiol:disulfide interchange protein DsbC OS=Escherichia coli (strain
BAC0275|nikR|sp|P0A6Z6|NIKR_ECOLI Nickel-responsive regulator OS=Escherichia coli (strain K12) GN=nik
BAC0274|nikE|sp|P33594|NIKE_ECOLI Nickel import ATP-binding protein NikE OS=Escherichia coli (strain K
BAC0654|merB1|sp|P16172|MERB_BACCE Alkylmercury lyase OS=Bacillus cereus GN=merB1 PE=3 SV=2
BAC0330|rcnA/yohM|sp|P76425|RCNA_ECOLI Nickel/cobalt efflux system RcnA OS=Escherichia coli (strain
BAC0433|ybtQ|tr|Q9Z375|Q9Z375_YERPE Inner membrane ABC-transporter YbtQ OS=Yersinia pestis GN=yb
BAC0630|copC|sp|P12376|COPC_PSEUB Copper resistance protein C OS=Pseudomonas syringae pv. tomato G
BAC0661|merB2|tr|Q7DJN2|Q7DJN2_BACME MerB2 OS=Bacillus megaterium GN=merB2 PE=4 SV=1
BAC0447|yjaA|sp|P09162|YJAA_ECOLI Uncharacterized protein YjaA OS=Escherichia coli (strain K12) GN=y
BAC0293|ruvB|tr|B5L348|B5L348_9PSED Malic enzyme family protein (Fragment) OS=Pseudomonas corrugat
BAC0644|corD|sp|Q56017|APAG_SALTY Protein ApaG OS=Salmonella typhimurium (strain LT2 / SGSC1412

BAC0446|yhcN|sp|P64614|YHCN_ECOLI Uncharacterized protein YhcN OS=Escherichia coli (strain K12) GN=
BAC0119|czcA|sp|P13511|CZCA_RALME Cobalt-zinc-cadmium resistance protein CzcA OS=Ralstonia metallic
BAC0269|nia|tr|Q92Z60|Q92Z60_RHIME Cation transport P-type ATPase OS=Rhizobium meliloti (strain 1021)
BAC0028|chrC|tr|A4UQR3|A4UQR3_9RHIZ Superoxide dismutase OS=Ochrobactrum tritici GN=chrC PE=3 S
BAC0565|actR|sp|A6UEL7|ACTR_SINMW Acid tolerance regulatory protein ActR OS=Sinorhizobium medicae
BAC0023|aioS/aoxS|tr|Q2VGB2|Q2VGB2_RHIRD Putative sensor histidine kinase OS=Rhizobium radiobacter
BAC0254|mrdH|tr|Q88IN1|Q88IN1_PSEPK Membrane protein, putative OS=Pseudomonas putida (strain KT244
BAC0049|bhsA/ycfR/comC|sp|P0AB40|BHSA_ECOLI Multiple stress resistance protein BhsA OS=Escherichia
BAC0549|nccA|sp|Q44586|NCCA_ALCXX Nickel-cobalt-cadmium resistance protein NccA OS=Alcaligenes xy
BAC0120|czcB|sp|P13510|CZCB_RALME Cobalt-zinc-cadmium resistance protein CzcB OS=Ralstonia metallid
BAC0389|terD|sp|P18781|TERD_ALCSP Tellurium resistance protein TerD OS=Alcaligenes sp. GN=terD PE=3
BAC0087|mgtA|sp|P0ABB8|ATMA_ECOLI Magnesium-transporting ATPase, P-type 1 OS=Escherichia coli (st
BAC0063|chrA|sp|P14285|CHRA_PSEAI Chromate transport protein OS=Pseudomonas aeruginosa GN=chrA P
BAC0434|ychH|sp|P0AB49|YCHH_ECOLI Uncharacterized protein YchH OS=Escherichia coli (strain K12) GN
BAC0263|ncrA|tr|Q06VT3|Q06VT3_9BACT NcrA OS=Leptospirillum ferriphilum GN=ncrA PE=4 SV=1
BAC0305|pcoC|sp|Q47454|PCOC_ECOLX Copper resistance protein C OS=Escherichia coli GN=pcoC PE=1 S
BAC0304|pcoB|sp|Q47453|PCOB_ECOLX Copper resistance protein B OS=Escherichia coli GN=pcoB PE=4 S
BAC0583|arsC|sp|P52147|ARSC2_ECOLX Arsenate reductase OS=Escherichia coli GN=arsC PE=3 SV=1
BAC0666|merD|tr|O66018|O66018_PSEST MerD OS=Pseudomonas stutzeri GN=merD PE=4 SV=1
BAC0348|silS|sp|Q9ZHD4|SILS_SALTM Probable sensor kinase SilS OS=Salmonella typhimurium GN=silS PE
BAC0027|chrB|tr|A4UQR5|A4UQR5_9RHIZ ChrB OS=Ochrobactrum tritici GN=chrB PE=4 SV=1
BAC0113|cutA|sp|P69488|CUTA_ECOLI Divalent-cation tolerance protein CutA OS=Escherichia coli (strain K1
BAC0022|aioR/aoxR|tr|Q2VGB1|Q2VGB1_RHIRD Putative transcriptional regulator OS=Rhizobium radiobacte
BAC0568|actP|sp|Q9X5V3|ATCU_RHILV Copper-transporting P-type ATPase OS=Rhizobium leguminosarum
BAC0182|golS|tr|Q8ZRG6|Q8ZRG6_SALTY Putative transcriptional regulator OS=Salmonella typhimurium (st
BAC0673|merE|sp|P06690|MERE_PSEAI Uncharacterized mercuric resistance protein MerE OS=Pseudomonas
BAC0620|copA|sp|P32113|COPA_ENTHA Probable copper-importing P-type ATPase A OS=Enterococcus hira
BAC0309|pcoS|sp|Q47457|PCOS_ECOLX Probable sensor protein PcoS OS=Escherichia coli GN=pcoS PE=3 S
BAC0343|silC|sp|Q9ZHD2|SILC_SALTM Probable outer membrane lipoprotein SilC OS=Salmonella typhimuri
BAC0659|merB|sp|P08664|MERB_SERMA Alkylmercury lyase OS=Serratia marcescens GN=merB PE=3 SV=1
BAC0083|copR|sp|Q02540|COPR_PSEUB Transcriptional activator protein CopR OS=Pseudomonas syringae p
BAC0355|ruvB|sp|Q51426|RUVB_PSEAE Holliday junction ATP-dependent DNA helicase RuvB OS=Pseudom
BAC0625|copA|tr|F4ZCZ9|F4ZCZ9_9XANT Copper resistance protein A OS=Xanthomonas alfalfae subsp. citru
BAC0614|cmeC|tr|Q8RTE3|Q8RTE3_CAMJU CmeC OS=Campylobacter jejuni GN=cmeC PE=4 SV=1
BAC0264|ncrB|tr|Q06VT2|Q06VT2_9BACT NcrB OS=Leptospirillum ferriphilum GN=ncrB PE=4 SV=1
BAC0597|baeS|tr|D0ZNE2|D0ZNE2_SALT1 Signal transduction histidine-protein kinase BaeS OS=Salmonella
BAC0352|sitD|tr|Q9XCR9|Q9XCR9_SALTM SitD OS=Salmonella typhimurium GN=sitD PE=3 SV=1
BAC0670|merE|tr|Q52104|Q52104_9ZZZZ Uncharacterized protein OS=Plasmid pDU1358 PE=4 SV=1
BAC0387|terB|sp|P18779|TERB_ALCSP Tellurium resistance protein TerB OS=Alcaligenes sp. GN=terB PE=4
BAC0302|pbrT|tr|Q5GR69|Q5GR69_ALCXX Lead uptake protein PbrT OS=Alcaligenes xylosoxydans xylosox
BAC0357|recG|tr|B5L350|B5L350_9PSED ATP-dependent DNA helicase (Fragment) OS=Pseudomonas corruga
BAC0356|recG|tr|Q9HTL3|Q9HTL3_PSEAE ATP-dependent DNA helicase RecG OS=Pseudomonas aeruginosa
BAC0293|ruvB|tr|B5L348|B5L348_9PSED Malic enzyme family protein (Fragment) OS=Pseudomonas corrugat
BAC0469|zupT/ygiE|sp|P0A8H3|ZUPT_ECOLI Zinc transporter ZupT OS=Escherichia coli (strain K12) GN=zu
BAC0350|sitB|tr|Q9XCS1|Q9XCS1_SALTM SitB OS=Salmonella typhimurium GN=sitB PE=3 SV=1
BAC0358|oscA|tr|B6CM35|B6CM35_9PSED Putative uncharacterized protein oscA OS=Pseudomonas corrugat

BAC0388|terC|sp|P18780|TERC_ALCSP Tellurium resistance protein TerC OS=Alcaligenes sp. GN=terC PE=3
BAC0181|glpF|sp|P0AER0|GLPF_ECOLI Glycerol uptake facilitator protein OS=Escherichia coli (strain K12) G
BAC0386|terA|sp|P18778|TERA_ALCSP Tellurium resistance protein TerA OS=Alcaligenes sp. GN=terA PE=4
BAC0548|chrA1|sp|P17551|CHRA1_RALME Chromate transport protein OS=Ralstonia metallidurans (strain CH
BAC0035|arsM|tr|Q6N3Y0|Q6N3Y0_RHOPA UbiE/COQ5 methyltransferase OS=Rhodopseudomonas palustris
BAC0441|yfeC|sp|Q56954|YFEC_YERPE Chelated iron transport system membrane protein YfeC OS=Yersinia
BAC0105|cueR/ybbI|sp|P0A9G4|CUER_ECOLI HTH-type transcriptional regulator CueR OS=Escherichia coli (
BAC0224|merA|sp|P16171|MERA_BACCE Mercuric reductase OS=Bacillus cereus GN=merA PE=1 SV=1
BAC0133|dnaK|sp|P0A5B9|DNAK_MYCTU Chaperone protein DnaK OS=Mycobacterium tuberculosis GN=dn
BAC0124|czcP|tr|Q1LAJ7|Q1LAJ7_RALME CzcP cation efflux P1-ATPase OS=Ralstonia metallidurans (strain
BAC0644|corD|sp|Q56017|APAG_SALTY Protein ApaG OS=Salmonella typhimurium (strain LT2 / SGSC1412
BAC0059|cadX|tr|A7LHQ4|A7LHQ4_STRSL CadX OS=Streptococcus salivarius GN=cadX PE=4 SV=1
BAC0695|merT|sp|P13112|MERT_SERMA Mercuric transport protein OS=Serratia marcescens GN=merT PE=4
BAC0030|arsA|sp|P52145|ARSA2_ECOLX Arsenical pump-driving ATPase OS=Escherichia coli GN=arsA PE=
BAC0251|mntH/yfeP|sp|P0A769|MNTH_ECOLI Divalent metal cation transporter MntH OS=Escherichia coli (s
BAC0351|sitC|tr|Q9XCS0|Q9XCS0_SALTM SitC OS=Salmonella typhimurium GN=sitC PE=3 SV=1
BAC0183|golT|tr|Q8ZRG7|Q8ZRG7_SALTY Putative cation transport ATPase OS=Salmonella typhimurium (st
BAC0318|pstS|sp|P0AG82|PSTS_ECOLI Phosphate-binding protein PstS OS=Escherichia coli (strain K12) GN=
BAC0649|merA|tr|E3VST6|E3VST6_9FLAO MerA OS=Tenacibaculum discolor GN=merA PE=3 SV=1
BAC0108|cusB|sp|P77239|CUSB_ECOLI Cation efflux system protein CusB OS=Escherichia coli (strain K12) G
BAC0003|acn|tr|O53166|O53166_MYCTU Aconitate hydratase OS=Mycobacterium tuberculosis H37Rv GN=R
BAC0135|dpsA|tr|Q8KR86|Q8KR86_BURPE DpsA OS=Burkholderia pseudomallei GN=dpsA PE=3 SV=1
BAC0034|arsH|tr|E8PS81|E8PS81_YERPE Arsenic resistance protein ArsH OS=Yersinia pestis Java 9 GN=arsH
BAC0306|pcoD|sp|Q47455|PCOD_ECOLX Copper resistance protein D OS=Escherichia coli GN=pcoD PE=3 S
BAC0106|cuiD|sp|Q8ZRS2|CUEO_SALTY Blue copper oxidase CueO OS=Salmonella typhimurium (strain LT2
BAC0648|merA|sp|P08662|MERA_SERMA Mercuric reductase (Fragments) OS=Serratia marcescens GN=merA
BAC0161|fbpB|sp|P71338|FBPB2_HAEIN Fe(3+)-transport system permease protein FbpB 2 OS=Haemophilus
BAC0312|pitA|sp|P0AFJ7|PITA_ECOLI Low-affinity inorganic phosphate transporter 1 OS=Escherichia coli (st
BAC0458|zipB|tr|Q7WJT8|Q7WJT8_BORBR Putative membrane protein OS=Bordetella bronchiseptica (strain A
BAC0383|tcrB|tr|Q8VPE6|Q8VPE6_ENTFC TcrB OS=Enterococcus faecium GN=tcrB PE=3 SV=1
BAC0587|arsD|sp|P52148|ARSD2_ECOLX Arsenical resistance operon trans-acting repressor ArsD OS=Escheri
BAC0252|mntP/yebN|sp|P76264|MNTP_ECOLI Probable manganese efflux pump MntP OS=Escherichia coli (st
BAC0088|corC|sp|P0A2L3|CORC_SALTY Magnesium and cobalt efflux protein CorC OS=Salmonella typhimur
BAC0653|merA|tr|Q934S5|Q934S5_THIFE Mercuric ion reductase OS=Thiobacillus ferrooxidans GN=merA PE
BAC0303|pcoA|sp|Q47452|PCOA_ECOLX Copper resistance protein A OS=Escherichia coli GN=pcoA PE=3 S
BAC0056|cadC|sp|P20047|CADC_STAAU Cadmium resistance transcriptional regulatory protein CadC OS=Sta
BAC0640|copD|tr|C6FFR7|C6FFR7_PSEFL CopD OS=Pseudomonas fluorescens GN=copD PE=4 SV=1
BAC0461|zntA/yhhO|sp|P37617|ATZN_ECOLI Lead, cadmium, zinc and mercury-transporting ATPase OS=Esc
BAC0272|nikC|sp|P0AFA9|NIKC_ECOLI Nickel transport system permease protein NikC OS=Escherichia coli (
BAC0662|merB3|tr|Q9RHR0|Q9RHR0_BACME MerB3 OS=Bacillus megaterium GN=merB3 PE=4 SV=2
BAC0275|nikR|sp|P0A6Z6|NIKR_ECOLI Nickel-responsive regulator OS=Escherichia coli (strain K12) GN=nik
BAC0315|pstA|sp|P07654|PSTA_ECOLI Phosphate transport system permease protein PstA OS=Escherichia col
BAC0631|copC|tr|F4ZBX9|F4ZBX9_XANCI CopC OS=Xanthomonas citri subsp. citri GN=copC PE=4 SV=1
BAC0167|fieF/yiip|sp|P69380|FIEF_ECOLI Ferrous-iron efflux pump FieF OS=Escherichia coli (strain K12) GN
BAC0639|copC|tr|C6FFR6|C6FFR6_PSEFL CopC OS=Pseudomonas fluorescens GN=copC PE=4 SV=1
BAC0463|znuA/yebL|sp|P39172|ZNUA_ECOLI High-affinity zinc uptake system protein ZnuA OS=Escherichia

BAC0102|cueA|tr|Q8KWW2|Q8KWW2_PSEPU Copper transporter OS=Pseudomonas putida GN=cueA PE=3 S
BAC0619|copA|tr|Q7WYH1|Q7WYH1_PSEPU CopA OS=Pseudomonas putida GN=copA PE=4 SV=1
BAC0115|cutE/lnt|sp|P23930|LNT_ECOLI Apolipoprotein N-acyltransferase OS=Escherichia coli (strain K12) G
BAC0082|copL|tr|Q5YKV8|Q5YKV8_9XANT CopL OS=Xanthomonas perforans GN=copL PE=4 SV=1
BAC0470|zur/yjbK|sp|P0AC51|ZUR_ECOLI Zinc uptake regulation protein OS=Escherichia coli (strain K12) GN
BAC0265|ncrC|tr|D5CKG5|D5CKG5_ENTCC Nickel-resistant membrane protein-like protein NcrC OS=Enterob
BAC0668|merD|tr|Q5NUV1|Q5NUV1_RALME MerD from Tn4378, regulatory protein involved in Hg(II) resist
BAC0316|pstB|sp|P0AAH0|PSTB_ECOLI Phosphate import ATP-binding protein PstB OS=Escherichia coli (stra
BAC0040|baeS|sp|P30847|BAES_ECOLI Signal transduction histidine-protein kinase BaeS OS=Escherichia coli
BAC0457|zinT/yodA|sp|P76344|ZINT_ECOLI Metal-binding protein ZinT OS=Escherichia coli (strain K12) GN
BAC0092|corT/coaT|tr|H0P0Y3|H0P0Y3_9SYNC Cation-transporting ATPase E1-E2 ATPase OS=Synechocysti
BAC0231|merP|sp|P13113|MERP_SERMA Mercuric transport protein periplasmic component OS=Serratia marc
BAC0459|zitB/ybgR|sp|P75757|ZITB_ECOLI Zinc transporter ZitB OS=Escherichia coli (strain K12) GN=zitB P
BAC0330|rcnA/yohM|sp|P76425|RCNA_ECOLI Nickel/cobalt efflux system RcnA OS=Escherichia coli (strain K
BAC0138|dsbC|sp|P0AEG6|DSBC_ECOLI Thiol:disulfide interchange protein DsbC OS=Escherichia coli (strain
BAC0573|arsB|sp|P45946|ARSB_BACSU Arsenite resistance protein ArsB OS=Bacillus subtilis (strain 168) GN
BAC0540|nfsA|sp|P17117|NFSA_ECOLI Oxygen-insensitive NADPH nitroreductase OS=Escherichia coli (strain
BAC0125|czcR|sp|Q44006|CZCR_RALME Transcriptional activator protein CzcR OS=Ralstonia metallidurans (
BAC0169|fpvA|sp|P48632|FPVA_PSEAE Ferripyoverdine receptor OS=Pseudomonas aeruginosa (strain ATCC
BAC0610|modB|sp|P0AF01|MODB_ECOLI Molybdenum transport system permease protein ModB OS=Escheri
BAC0162|fbpC|sp|P44513|FBPC2_HAEIN Fe(3+) ions import ATP-binding protein FbpC 2 OS=Haemophilus in
BAC0201|kmtR|sp|O53838|KMTR_MYCTU HTH-type transcriptional regulator KmtR OS=Mycobacterium tube
BAC0652|merA|tr|O66017|O66017_PSEST MerA OS=Pseudomonas stutzeri GN=merA PE=4 SV=1
BAC0109|cusC/ylcB|sp|P77211|CUSC_ECOLI Cation efflux system protein CusC OS=Escherichia coli (strain K
BAC0464|znuB/yebI|sp|P39832|ZNUB_ECOLI High-affinity zinc uptake system membrane protein ZnuB OS=Es
BAC0344|silE|sp|Q9Z4N3|SILE_SALTM Silver-binding protein SilE OS=Salmonella typhimurium GN=silE PE=
BAC0168|fptA|sp|P42512|FPTA_PSEAE Fe(3+)-pyochelin receptor OS=Pseudomonas aeruginosa (strain ATCC
BAC0190|hmrR|sp|Q9X5X4|HMRR_SINMW HTH-type transcriptional regulator HmrR OS=Sinorhizobium med
BAC0570|actP|tr|D5AU53|D5AU53_RHOCB Cation/acetate symporter ActP-1 OS=Rhodobacter capsulatus (stra
BAC0345|silF|tr|Q9ZHD1|Q9ZHD1_SALTM Uncharacterized protein OS=Salmonella typhimurium GN=ORF96
BAC0588|arsR|tr|P74986|P74986_YEREN Arsenite inducible repressor OS=Yersinia enterocolitica GN=arsR PE
BAC0112|cusS|sp|P77485|CUSS_ECOLI Sensor kinase CusS OS=Escherichia coli (strain K12) GN=cusS PE=1 S
BAC0307|pcoE|sp|Q47459|PCOE_ECOLX Probable copper-binding protein PcoE OS=Escherichia coli GN=pcoE
BAC0646|mdtB|tr|D0ZND9|D0ZND9_SALT1 Multidrug resistance protein MdtB OS=Salmonella typhimurium (
BAC0643|corB|tr|Q9X621|Q9X621_SALTM CorB OS=Salmonella typhimurium GN=corB PE=4 SV=1
BAC0368|sodA|sp|P00448|SODM_ECOLI Superoxide dismutase [Mn] OS=Escherichia coli (strain K12) GN=so
BAC0579|arsB|sp|P74311|Y944_SYNY3 Uncharacterized transporter slr0944 OS=Synechocystis sp. (strain PCC
BAC0270|nikA|sp|P33590|NIKA_ECOLI Nickel-binding periplasmic protein OS=Escherichia coli (strain K12) G
BAC0442|yfeD|sp|Q56955|YFED_YERPE Chelated iron transport system membrane protein YfeD OS=Yersinia
BAC0584|arsC|sp|O50595|ARSC_ACIMA Arsenate reductase OS=Acidiphilium multivorum (strain DSM 11245
BAC0077|copA|sp|P12374|COPA_PSEUB Copper resistance protein A OS=Pseudomonas syringae pv. tomato G
BAC0273|nikD|sp|P33593|NIKD_ECOLI Nickel import ATP-binding protein NikD OS=Escherichia coli (strain K
BAC0390|terE|sp|P18782|TERE_ALCSP Tellurium resistance protein TerE OS=Alcaligenes sp. GN=terE PE=3 S
BAC0078|copA|sp|O32220|COPA_BACSU Copper-exporting P-type ATPase A OS=Bacillus subtilis (strain 168)
BAC0678|merP|tr|O66016|O66016_PSEST MerP OS=Pseudomonas stutzeri GN=merP PE=4 SV=1
BAC0029|chrF|tr|A4UQR2|A4UQR2_9RHIZ ChrF OS=Ochrobactrum tritici GN=chrF PE=4 SV=1

BAC0116|cutF/nlpE|sp|P40710|NLPE_ECOLI Lipoprotein NlpE OS=Escherichia coli (strain K12) GN=nlpE PE=
BAC0455|ziaA|sp|Q59998|ATZN_SYNY3 Zinc-transporting ATPase OS=Synechocystis sp. (strain PCC 6803 / K
BAC0031|arsB|sp|P08691|ARSB1_ECOLX Arsenical pump membrane protein OS=Escherichia coli GN=arsB PE
BAC0611|modC|sp|P09833|MODC_ECOLI Molybdenum import ATP-binding protein ModC OS=Escherichia co
BAC0541|yieF|sp|P0AGE6|YIEF_ECOLI Uncharacterized protein YieF OS=Escherichia coli (strain K12) GN=yi
BAC0661|merB2|tr|Q7DJN2|Q7DJN2_BACME MerB2 OS=Bacillus megaterium GN=merB2 PE=4 SV=1
BAC0683|merR1|tr|O07300|O07300_9PSED Mercuric resistance operon regulatory protein OS=Pseudomonas sp
BAC0650|merA|tr|O08449|O08449_9PSED Mercuric reductase OS=Pseudomonas sp. K-62 GN=merA PE=4 SV
BAC0203|cnrA|sp|P37972|CNRA_RALME Nickel and cobalt resistance protein CnrA OS=Ralstonia metallidura
BAC0638|copR|tr|C6FFR4|C6FFR4_PSEFL CopR OS=Pseudomonas fluorescens GN=copR PE=4 SV=1
BAC0686|merR|sp|P13111|MERR_SERMA Mercuric resistance operon regulatory protein OS=Serratia marcesce
BAC0334|robA|sp|P0ACI0|ROB_ECOLI Right origin-binding protein OS=Escherichia coli (strain K12) GN=rob
BAC0440|yfeB|sp|Q56953|YFEB_YERPE Chelated iron transport system membrane protein YfeB OS=Yersinia
BAC0228|merF|tr|Q2QCN0|Q2QCN0_9PSED MerF OS=Pseudomonas sp. CT14 GN=merF PE=4 SV=1
BAC0266|ncrY|tr|Q06VT0|Q06VT0_9BACT NcrY OS=Leptospirillum ferriphilum GN=ncrY PE=4 SV=1
BAC0612|perO|tr|D5AQ60|D5AQ60_RHOCB Divalent ion symporter OS=Rhodobacter capsulatus (strain ATCC
BAC0591|arsR|sp|P52144|ARSR2_ECOLX Arsenical resistance operon repressor OS=Escherichia coli GN=arsR
BAC0679|merP|tr|O66047|O66047_PSEST Mercury transport protein OS=Pseudomonas stutzeri GN=merP PE=4
BAC0165|fetA/ybbL|sp|P77279|YBBL_ECOLI Uncharacterized ABC transporter ATP-binding protein YbbL OS
BAC0645|mdtA|tr|D0ZND8|D0ZND8_SALT1 Multidrug resistance protein MdtA OS=Salmonella typhimurium
BAC0277|nirB|tr|Q6RUG2|Q6RUG2_KLEOX NirB OS=Klebsiella oxytoca GN=nirB PE=4 SV=1
BAC0298|pbrA|tr|Q58AJ6|Q58AJ6_RALME P-type ATPase involved in Pb(II) resistance PbrA OS=Ralstonia me
BAC0240|mexI|tr|Q9HWH4|Q9HWH4_PSEAE Probable Resistance-Nodulation-Cell Division (RND) efflux tran
BAC0166|fetB/ybbM|sp|P77307|YBBM_ECOLI UPF0014 inner membrane protein YbbM OS=Escherichia coli (
BAC0274|nikE|sp|P33594|NIKE_ECOLI Nickel import ATP-binding protein NikE OS=Escherichia coli (strain K
BAC0698|ncrA|tr|Q1KLR2|Q1KLR2_SERMA NcrA OS=Serratia marcescens GN=ncrA PE=4 SV=1
BAC0229|merG|tr|O07302|O07302_9PSED Mercuric resistance protein OS=Pseudomonas sp. K-62 GN=merG P
BAC0341|silA|sp|Q9ZHC9|SILA_SALTM Putative cation efflux system protein SilA OS=Salmonella typhimuriu
BAC0349|sitA|tr|Q9XCS2|Q9XCS2_SALTI Iron transport protein, periplasmic-binding protein OS=Salmonella t
BAC0107|cusA/ybdE|sp|P38054|CUSA_ECOLI Cation efflux system protein CusA OS=Escherichia coli (strain K
BAC0114|cutC|sp|P67826|CUTC_ECOLI Copper homeostasis protein CutC OS=Escherichia coli (strain K12) GN
BAC0699|ncrB|tr|Q1KLR1|Q1KLR1_SERMA NcrB OS=Serratia marcescens GN=ncrB PE=4 SV=1
BAC0432|ybtP|tr|Q9R7V3|Q9R7V3_YERPE Lipoprotein inner membrane ABC-transporter OS=Yersinia pestis G
BAC0439|yfeA|sp|Q56952|YFEA_YERPE Periplasmic chelated iron-binding protein YfeA OS=Yersinia pestis G
BAC0384|tehA|sp|P25396|TEHA_ECOLI Tellurite resistance protein TehA OS=Escherichia coli (strain K12) GN
BAC0707|sodB|sp|P0AGD3|SODF_ECOLI Superoxide dismutase [Fe] OS=Escherichia coli (strain K12) GN=sod
BAC0271|nikB|sp|P33591|NIKB_ECOLI Nickel transport system permease protein NikB OS=Escherichia coli (s
BAC0317|pstC|sp|P0AGH8|PSTC_ECOLI Phosphate transport system permease protein PstC OS=Escherichia co
BAC0137|dsbB|sp|P0A6M2|DSBB_ECOLI Disulfide bond formation protein B OS=Escherichia coli (strain K12)
BAC0308|pcoR|sp|Q47456|PCOR_ECOLX Transcriptional regulatory protein PcoR OS=Escherichia coli GN=pc
BAC0447|yjaA|sp|P09162|YJAA_ECOLI Uncharacterized protein YjaA OS=Escherichia coli (strain K12) GN=y
BAC0285|nreB|tr|F0KND8|F0KND8_ACICP NrsD, nreB nickel permease involved in nickel and cobalt toleranc
BAC0688|merR2|tr|Q79B70|Q79B70_PSEST Organomercurial resistance regulatory protein OS=Pseudomonas st
BAC0685|merR|tr|H6WCN2|H6WCN2_9FLAO MerR OS=Tenacibaculum discolor GN=merR PE=4 SV=1
BAC0572|arsA|sp|P08690|ARSA1_ECOLX Arsenical pump-driving ATPase OS=Escherichia coli GN=arsA PE=
BAC0687|merR|tr|Q79BG7|Q79BG7_PSEST MerR OS=Pseudomonas stutzeri GN=merR PE=4 SV=1

BAC0641|corA|sp|P0A2R8|CORA_SALTY Magnesium transport protein CorA OS=Salmonella typhimurium (st
BAC0255|mreA|tr|Q88IN0|Q88IN0_PSEPK Putative uncharacterized protein OS=Pseudomonas putida (strain KT
BAC0039|baeR|sp|P69228|BAER_ECOLI Transcriptional regulatory protein BaeR OS=Escherichia coli (strain K
BAC0577|arsB|sp|P74985|ARSB_YEREN Arsenical pump membrane protein OS=Yersinia enterocolitica GN=ar
BAC0691|merT|tr|Q52397|Q52397_PSEST Mercury transport protein OS=Pseudomonas stutzeri GN=merT PE=4
BAC0276|nirA|tr|Q6RUG3|Q6RUG3_KLEOX NirA OS=Klebsiella oxytoca GN=nirA PE=4 SV=1
BAC0267|nczA|tr|B8GZE9|B8GZE9_CAUCN Cobalt-zinc-cadmium resistance protein czcA OS=Caulobacter cre
BAC0391|terW|sp|P75010|TERW_SERMA Tellurium resistance protein TerW OS=Serratia marcescens GN=terW
BAC0596|baeR|tr|D0ZNE3|D0ZNE3_SALT1 DNA-binding transcriptional regulator BaeR OS=Salmonella typhi
BAC0446|yhcN|sp|P64614|YHCN_ECOLI Uncharacterized protein YhcN OS=Escherichia coli (strain K12) GN=
BAC0563|acrD|tr|Q8ZN77|Q8ZN77_SALTY RND family aminoglycoside/multidrug efflux pump OS=Salmonel
BAC0098|ctpC|sp|P0A502|CTPC_MYCTU Probable manganese/zinc-exporting P-type ATPase OS=Mycobacteri
BAC0179|gesB|tr|Q8ZRG9|Q8ZRG9_SALTY Putative cation efflux system protein OS=Salmonella typhimurium
BAC0451|yodD|sp|P64519|YODD_ECOLI Uncharacterized protein YodD OS=Escherichia coli (strain K12) GN=
BAC0667|merD|sp|P08654|MERD_SERMA HTH-type transcriptional regulator MerD OS=Serratia marcescens G
BAC0609|modA|sp|P37329|MODA_ECOLI Molybdate-binding periplasmic protein OS=Escherichia coli (strain K
BAC0621|copA|tr|F4ZBX3|F4ZBX3_XANCI CopA OS=Xanthomonas citri subsp. citri GN=copA PE=4 SV=1
BAC0627|copB|tr|F4ZD00|F4ZD00_9XANT Copper resistance protein B OS=Xanthomonas alfalfae subsp. citrum
BAC0342|silB|sp|Q9ZHD0|SILB_SALTM Putative membrane fusion protein SilB OS=Salmonella typhimurium G
BAC0026|chrA|tr|A4UQR4|A4UQR4_9RHIZ Chromate transporter OS=Ochrobactrum tritici GN=chrA PE=4 SV
BAC0058|cadR|tr|Q93TP7|Q93TP7_PSEPU CadR OS=Pseudomonas putida GN=cadR PE=4 SV=1
BAC0130|czrA|tr|Q9RLI8|Q9RLI8_PSEAI CzrA protein OS=Pseudomonas aeruginosa GN=czrA PE=4 SV=1
BAC0689|merR|tr|Q934S8|Q934S8_THIFE Mer operon regulatory protein OS=Thiobacillus ferrooxidans GN=m
BAC0462|zntR/yhdM|sp|P0ACS5|ZNTR_ECOLI HTH-type transcriptional regulator ZntR OS=Escherichia coli (
BAC0647|mdtC|tr|D0ZNE0|D0ZNE0_SALT1 Multidrug resistance protein MdtC OS=Salmonella typhimurium (s
BAC0576|arsB|sp|P52146|ARSB2_ECOLX Arsenical pump membrane protein OS=Escherichia coli GN=arsB PE
BAC0684|merR2|tr|Q7DKL2|Q7DKL2_9PSED MerR2 OS=Pseudomonas sp. K-62 GN=merR2 PE=4 SV=1
BAC0171|furA|sp|P0A582|FURA_MYCTU Transcriptional regulator FurA OS=Mycobacterium tuberculosis GN
BAC0163|fecD|sp|P15029|FECD_ECOLI Fe(3+) dicitrate transport system permease protein FecD OS=Escherich
BAC0608|modE|sp|P0A9G8|MODE_ECOLI Transcriptional regulator ModE OS=Escherichia coli (strain K12) G
BAC0642|mgtA|sp|P36640|ATMA_SALTY Magnesium-transporting ATPase, P-type 1 OS=Salmonella typhimu
BAC0066|chrF|tr|Q5NUZ7|Q5NUZ7_RALME ChrF, regulatory protein, involved in Chromate resistance OS=Ra
BAC0672|merE|tr|Q79BE4|Q79BE4_PSEST Urf1 OS=Pseudomonas stutzeri PE=4 SV=1
BAC0467|zraR/hydH|sp|P14375|ZRAR_ECOLI Transcriptional regulatory protein ZraR OS=Escherichia coli (str
BAC0498|ideR|sp|P0A672|IDER_MYCTU Iron-dependent repressor IdeR OS=Mycobacterium tuberculosis GN=
BAC0103|cueO|sp|P36649|CUEO_ECOLI Blue copper oxidase CueO OS=Escherichia coli (strain K12) GN=cue
BAC0371|soxS|sp|P0A9E2|SOXS_ECOLI Regulatory protein SoxS OS=Escherichia coli (strain K12) GN=soxS P
BAC0594|arsR|sp|P37309|ARSR_ECOLI Arsenical resistance operon repressor OS=Escherichia coli (strain K12)
BAC0164|fecE|sp|P15031|FECE_ECOLI Fe(3+) dicitrate transport ATP-binding protein FecE OS=Escherichia co
BAC0136|dsbA|sp|P0AEG4|DSBA_ECOLI Thiol:disulfide interchange protein DsbA OS=Escherichia coli (strain
BAC0331|rcnB/yohN|sp|P64534|RCNB_ECOLI Nickel/cobalt homeostasis protein RcnB OS=Escherichia coli (st
BAC0086|corA|sp|P0ABI4|CORA_ECOLI Magnesium transport protein CorA OS=Escherichia coli (strain K12)
BAC0452|yqjH|sp|Q46871|YQJH_ECOLI NADPH-dependent ferric-chelate reductase OS=Escherichia coli (strai
BAC0692|merT|tr|Q79BG6|Q79BG6_PSEST MerT OS=Pseudomonas stutzeri GN=merT PE=4 SV=1
BAC0076|comR/ycfQ|sp|P75952|COMR_ECOLI HTH-type transcriptional repressor ComR OS=Escherichia coli
BAC0347|silR|sp|Q9ZHD3|SILR_SALTM Probable transcriptional regulatory protein SilR OS=Salmonella typhi

BAC0578|arsB|tr|O50594|O50594_ACIMU ArsB OS=Acidiphilium multivorum GN=arsB PE=4 SV=1
BAC0566|actS|tr|Q52912|Q52912_9RHIZ Histidine protein kinase OS=Sinorhizobium medicae GN=actS PE=4 S
BAC0693|merT|tr|Q79F00|Q79F00_9PSED Mercuric transport protein OS=Pseudomonas sp. K-62 GN=merT PE
BAC0346|silP|sp|Q9ZHC7|SILP_SALTM Silver exporting P-type ATPase OS=Salmonella typhimurium GN=sil
BAC0665|merD|tr|O66022|O66022_PSEST Mercury operon coregulator protein OS=Pseudomonas stutzeri GN=
BAC0233|merT|sp|P94185|MERT_ALCSP Mercuric transport protein OS=Alcaligenes sp. GN=merT PE=3 SV=
BAC0582|arsC|sp|P08692|ARSC1_ECOLX Arsenate reductase OS=Escherichia coli GN=arsC PE=1 SV=1
BAC0445|ygiW|sp|P0ADU5|YGIW_ECOLI Protein YgiW OS=Escherichia coli (strain K12) GN=ygiW PE=1 S
BAC0694|merT-P|tr|H6WCN3|H6WCN3_9FLAO MerT-P OS=Tenacibaculum discolor GN=merT-P PE=4 SV=
BAC0690|merT|tr|Q934S7|Q934S7_THIFE Mercuric ion transport protein OS=Thiobacillus ferrooxidans GN=m
BAC0465|znuC/yebM|sp|P0A9X1|ZNUC_ECOLI Zinc import ATP-binding protein ZnuC OS=Escherichia coli (
BAC0637|copS|tr|C6FFR5|C6FFR5_PSEFL CopS OS=Pseudomonas fluorescens GN=copS PE=4 SV=1
BAC0675|merP|tr|O07301|O07301_9PSED Mercuric transport protein periplasmic component OS=Pseudomonas
BAC0134|dpr/dps|sp|P0CB53|DPS_STRSU DNA protection during starvation protein OS=Streptococcus suis G
BAC0100|ctpG|sp|P63689|CTPG_MYCTU Probable cation-transporting ATPase G OS=Mycobacterium tubercul
BAC0385|tehB|sp|P25397|TEHB_ECOLI Tellurite methyltransferase OS=Escherichia coli (strain K12) GN=tehB
BAC0468|zraS/hydG|sp|P14377|ZRAS_ECOLI Sensor protein ZraS OS=Escherichia coli (strain K12) GN=zraS
BAC0012|actP|sp|Q9X5X3|ATCU_SINMW Copper-transporting P-type ATPase OS=Sinorhizobium medicae (st
BAC0651|merA|sp|P0A0E5|MERA_STAAU Mercuric reductase OS=Staphylococcus aureus GN=merA PE=3 S
BAC0253|mntR|sp|P0A9F1|MNTR_ECOLI Transcriptional regulator MntR OS=Escherichia coli (strain K12) G
BAC0101|ctpV|sp|P77894|CTPV_MYCTU Probable copper-exporting P-type ATPase V OS=Mycobacterium tu
BAC0332|rcnR/yohL|sp|P64530|RCNR_ECOLI Transcriptional repressor RcnR OS=Escherichia coli (strain K12
BAC0111|cusR/ylcA|sp|P0ACZ8|CUSR_ECOLI Transcriptional regulatory protein CusR OS=Escherichia coli (s
BAC0628|copB|tr|F4ZBX4|F4ZBX4_XANCI CopB OS=Xanthomonas citri subsp. citri GN=copB PE=4 SV=1
BAC0110|cusF/cusX|sp|P77214|CUSF_ECOLI Cation efflux system protein CusF OS=Escherichia coli (strain K
BAC0433|ybtQ|tr|Q9Z375|Q9Z375_YERPE Inner membrane ABC-transporter YbtQ OS=Yersinia pestis GN=yb
BAC0571|arsA|sp|O50593|ARSA_ACIMA Arsenical pump-driving ATPase OS=Acidiphilium multivorum (strai
BAC0079|copB|sp|P05425|COPB_ENTHA Copper-exporting P-type ATPase B OS=Enterococcus hirae (strain A
BAC0588|arsR|tr|P74986|P74986_YEREN Arsenite inducible repressor OS=Yersinia enterocolitica GN=arsR PE
BAC0293|ruvB|tr|B5L348|B5L348_9PSED Malic enzyme family protein (Fragment) OS=Pseudomonas corrugat
BAC0639|copC|tr|C6FFR6|C6FFR6_PSEFL CopC OS=Pseudomonas fluorescens GN=copC PE=4 SV=1
BAC0331|rcnB/yohN|sp|P64534|RCNB_ECOLI Nickel/cobalt homeostasis protein RcnB OS=Escherichia coli (s
BAC0611|modC|sp|P09833|MODC_ECOLI Molybdenum import ATP-binding protein ModC OS=Escherichia c
BAC0030|arsA|sp|P52145|ARSA2_ECOLX Arsenical pump-driving ATPase OS=Escherichia coli GN=arsA PE=
BAC0388|terC|sp|P18780|TERC_ALCSP Tellurium resistance protein TerC OS=Alcaligenes sp. GN=terC PE=3
BAC0441|yfeC|sp|Q56954|YFEC_YERPE Chelated iron transport system membrane protein YfeC OS=Yersinia
BAC0389|terD|sp|P18781|TERD_ALCSP Tellurium resistance protein TerD OS=Alcaligenes sp. GN=terD PE=3
BAC0285|nreB|tr|F0KND8|F0KND8_ACICP NrsD, nreB nickel permease involved in nickel and cobalt toleranc
BAC0251|mntH/yfeP|sp|P0A769|MNTH_ECOLI Divalent metal cation transporter MntH OS=Escherichia coli (s
BAC0119|czcA|sp|P13511|CZCA_RALME Cobalt-zinc-cadmium resistance protein CzcA OS=Ralstonia metalli
BAC0652|merA|tr|O66017|O66017_PSEST MerA OS=Pseudomonas stutzeri GN=merA PE=4 SV=1
BAC0040|baeS|sp|P30847|BAES_ECOLI Signal transduction histidine-protein kinase BaeS OS=Escherichia coli
BAC0346|silP|sp|Q9ZHC7|SILP_SALTM Silver exporting P-type ATPase OS=Salmonella typhimurium GN=sil
BAC0109|cusC/ylcB|sp|P77211|CUSC_ECOLI Cation efflux system protein CusC OS=Escherichia coli (strain K
BAC0463|znuA/yebL|sp|P39172|ZNUA_ECOLI High-affinity zinc uptake system protein ZnuA OS=Escherichia
BAC0181|glpF|sp|P0AER0|GLPF_ECOLI Glycerol uptake facilitator protein OS=Escherichia coli (strain K12) G

BAC0467|zraR/hydH|sp|P14375|ZRAR_ECOLI Transcriptional regulatory protein ZraR OS=Escherichia coli (str
BAC0585|arsC|sp|P74984|ARSC_YEREN Arsenate reductase OS=Yersinia enterocolitica GN=arsC PE=3 SV=1
BAC0459|zitB/ybgR|sp|P75757|ZITB_ECOLI Zinc transporter ZitB OS=Escherichia coli (strain K12) GN=zitB P
BAC0059|cadX|tr|A7LHQ4|A7LHQ4_STRSL CadX OS=Streptococcus salivarius GN=cadX PE=4 SV=1
BAC0273|nikD|sp|P33593|NIKD_ECOLI Nickel import ATP-binding protein NikD OS=Escherichia coli (strain K
BAC0666|merD|tr|O66018|O66018_PSEST MerD OS=Pseudomonas stutzeri GN=merD PE=4 SV=1
BAC0549|nccA|sp|Q44586|NCCA_ALCXX Nickel-cobalt-cadmium resistance protein NccA OS=Alcaligenes xy
BAC0022|aioR/aoxR|tr|Q2VGB1|Q2VGB1_RHIRD Putative transcriptional regulator OS=Rhizobium radiobacter
BAC0563|acrD|tr|Q8ZN77|Q8ZN77_SALTY RND family aminoglycoside/multidrug efflux pump OS=Salmonell
BAC0027|chrB|tr|A4UQR5|A4UQR5_9RHIZ ChrB OS=Ochrobactrum tritici GN=chrB PE=4 SV=1
BAC0352|sitD|tr|Q9XCR9|Q9XCR9_SALTM SitD OS=Salmonella typhimurium GN=sitD PE=3 SV=1
BAC0168|fptA|sp|P42512|FPTA_PSEAE Fe(3+)-pyochelin receptor OS=Pseudomonas aeruginosa (strain ATCC
BAC0026|chrA|tr|A4UQR4|A4UQR4_9RHIZ Chromate transporter OS=Ochrobactrum tritici GN=chrA PE=4 SV
BAC0355|ruvB|sp|Q51426|RUVB_PSEAE Holliday junction ATP-dependent DNA helicase RuvB OS=Pseudom
BAC0687|merR|tr|Q79BG7|Q79BG7_PSEST MerR OS=Pseudomonas stutzeri GN=merR PE=4 SV=1
BAC0368|sodA|sp|P00448|SODM_ECOLI Superoxide dismutase [Mn] OS=Escherichia coli (strain K12) GN=so
BAC0279|nirD|tr|Q6RUG0|Q6RUG0_KLEOX NirD OS=Klebsiella oxytoca GN=nirD PE=4 SV=1
BAC0670|merE|tr|Q52104|Q52104_9ZZZZ Uncharacterized protein OS=Plasmid pDU1358 PE=4 SV=1
BAC0344|silE|sp|Q9Z4N3|SILE_SALTM Silver-binding protein SilE OS=Salmonella typhimurium GN=silE PE=
BAC0371|soxS|sp|P0A9E2|SOXS_ECOLI Regulatory protein SoxS OS=Escherichia coli (strain K12) GN=soxS P
BAC0049|bhsA/ycfR/comC|sp|P0AB40|BHSA_ECOLI Multiple stress resistance protein BhsA OS=Escherichia c
BAC0665|merD|tr|O66022|O66022_PSEST Mercury operon coregulator protein OS=Pseudomonas stutzeri GN=r
BAC0101|ctpV|sp|P77894|CTPV_MYCTU Probable copper-exporting P-type ATPase V OS=Mycobacterium tub
BAC0107|cusA/ybdE|sp|P38054|CUSA_ECOLI Cation efflux system protein CusA OS=Escherichia coli (strain K
BAC0298|pbrA|tr|Q58AJ6|Q58AJ6_RALME P-type ATPase involved in Pb(II) resistance PbrA OS=Ralstonia me
BAC0114|cutC|sp|P67826|CUTC_ECOLI Copper homeostasis protein CutC OS=Escherichia coli (strain K12) GN
BAC0086|corA|sp|P0ABI4|CORA_ECOLI Magnesium transport protein CorA OS=Escherichia coli (strain K12)
BAC0102|cueA|tr|Q8KWW2|Q8KWW2_PSEPU Copper transporter OS=Pseudomonas putida GN=cueA PE=3 S
BAC0137|dsbB|sp|P0A6M2|DSBB_ECOLI Disulfide bond formation protein B OS=Escherichia coli (strain K12)
BAC0110|cusF/cusX|sp|P77214|CUSF_ECOLI Cation efflux system protein CusF OS=Escherichia coli (strain K
BAC0686|merR|sp|P13111|MERR_SERMA Mercuric resistance operon regulatory protein OS=Serratia marcesce
BAC0452|yqjH|sp|Q46871|YQJH_ECOLI NADPH-dependent ferric-chelate reductase OS=Escherichia coli (strai
BAC0573|arsB|sp|P45946|ARSB_BACSU Arsenite resistance protein ArsB OS=Bacillus subtilis (strain 168) GN
BAC0672|merE|tr|Q79BE4|Q79BE4_PSEST Urf1 OS=Pseudomonas stutzeri PE=4 SV=1
BAC0651|merA|sp|P0A0E5|MERA_STAAU Mercuric reductase OS=Staphylococcus aureus GN=merA PE=3 SV
BAC0240|mexI|tr|Q9HWH4|Q9HWH4_PSEAE Probable Resistance-Nodulation-Cell Division (RND) efflux tran
BAC0387|terB|sp|P18779|TERB_ALCSP Tellurium resistance protein TerB OS=Alcaligenes sp. GN=terB PE=4
BAC0565|actR|sp|A6UEL7|ACTR_SINMW Acid tolerance regulatory protein ActR OS=Sinorhizobium medicae
BAC0657|merB|tr|O07303|O07303_9PSED Alkylmercury lyase OS=Pseudomonas sp. K-62 GN=merB PE=3 SV
BAC0447|yjaA|sp|P09162|YJAA_ECOLI Uncharacterized protein YjaA OS=Escherichia coli (strain K12) GN=y
BAC0108|cusB|sp|P77239|CUSB_ECOLI Cation efflux system protein CusB OS=Escherichia coli (strain K12) G
BAC0637|copS|tr|C6FFR5|C6FFR5_PSEFL CopS OS=Pseudomonas fluorescens GN=copS PE=4 SV=1
BAC0384|tehA|sp|P25396|TEHA_ECOLI Tellurite resistance protein TehA OS=Escherichia coli (strain K12) GN
BAC0299|pbrB/pbrC|tr|Q58AJ7|Q58AJ7_RALME Lipoprotein signal peptidase OS=Ralstonia metallidurans (stra
BAC0668|merD|tr|Q5NUV1|Q5NUV1_RALME MerD from Tn4378, regulatory protein involved in Hg(II) resist
BAC0629|copB|sp|O30085|COPB_ARCFU Copper-exporting P-type ATPase B OS=Archaeoglobus fulgidus (str

BAC0577|arsB|sp|P74985|ARSB_YEREN Arsenical pump membrane protein OS=Yersinia enterocolitica GN=ar
BAC0644|corD|sp|Q56017|APAG_SALTY Protein ApaG OS=Salmonella typhimurium (strain LT2 / SGSC1412
BAC0169|fpvA|sp|P48632|FPVA_PSEAE Ferripyoverdine receptor OS=Pseudomonas aeruginosa (strain ATCC
BAC0540|nfsA|sp|P17117|NFSA_ECOLI Oxygen-insensitive NADPH nitroreductase OS=Escherichia coli (strain
BAC0383|tcrB|tr|Q8VPE6|Q8VPE6_ENTFC TcrB OS=Enterococcus faecium GN=tcrB PE=3 SV=1
BAC0035|arsM|tr|Q6N3Y0|Q6N3Y0_RHOPA UbiE/COQ5 methyltransferase OS=Rhodopseudomonas palustris
BAC0445|ygiW|sp|P0ADU5|YGIW_ECOLI Protein YgiW OS=Escherichia coli (strain K12) GN=ygiW PE=1 SV
BAC0334|robA|sp|P0ACI0|ROB_ECOLI Right origin-binding protein OS=Escherichia coli (strain K12) GN=rob
BAC0077|copA|sp|P12374|COPA_PSEUB Copper resistance protein A OS=Pseudomonas syringae pv. tomato G
BAC0163|fecD|sp|P15029|FECD_ECOLI Fe(3+) dicitrate transport system permease protein FecD OS=Escherich
BAC0130|czrA|tr|Q9RLI8|Q9RLI8_PSEAI CzrA protein OS=Pseudomonas aeruginosa GN=czrA PE=4 SV=1
BAC0270|nikA|sp|P33590|NIKA_ECOLI Nickel-binding periplasmic protein OS=Escherichia coli (strain K12) G
BAC0078|copA|sp|O32220|COPA_BACSU Copper-exporting P-type ATPase A OS=Bacillus subtilis (strain 168)
BAC0699|ncrB|tr|Q1KLR1|Q1KLR1_SERMA NcrB OS=Serratia marcescens GN=ncrB PE=4 SV=1
BAC0012|actP|sp|Q9X5X3|ATCU_SINMW Copper-transporting P-type ATPase OS=Sinorhizobium medicae (st
BAC0570|actP|tr|D5AU53|D5AU53_RHOCB Cation/acetate symporter ActP-1 OS=Rhodobacter capsulatus (stra
BAC0134|dpr/dps|sp|P0CB53|DPS_STRSU DNA protection during starvation protein OS=Streptococcus suis GN
BAC0457|zinT/yodA|sp|P76344|ZINT_ECOLI Metal-binding protein ZinT OS=Escherichia coli (strain K12) GN
BAC0349|sitA|tr|Q9XCS2|Q9XCS2_SALTI Iron transport protein, periplasmic-binding protein OS=Salmonella t
BAC0684|merR2|tr|Q7DKL2|Q7DKL2_9PSED MerR2 OS=Pseudomonas sp. K-62 GN=merR2 PE=4 SV=1
BAC0692|merT|tr|Q79BG6|Q79BG6_PSEST MerT OS=Pseudomonas stutzeri GN=merT PE=4 SV=1
BAC0631|copC|tr|F4ZBX9|F4ZBX9_XANCI CopC OS=Xanthomonas citri subsp. citri GN=copC PE=4 SV=1
BAC0099|ctpD|sp|A0R3A7|CTPD_MYCS2 Probable cobalt/nickel-exporting P-type ATPase OS=Mycobacterium
BAC0575|arsB|sp|Q01255|ARSB_STAXY Arsenical pump membrane protein OS=Staphylococcus xylosus GN=
BAC0170|frnE|tr|Q9RWK7|Q9RWK7_DEIRA FrnE protein OS=Deinococcus radiodurans (strain ATCC 13939 /
BAC0307|pcoE|sp|Q47459|PCOE_ECOLX Probable copper-binding protein PcoE OS=Escherichia coli GN=pcoE
BAC0392|terZ|sp|Q52353|TERZ_SERMA Tellurium resistance protein TerZ OS=Serratia marcescens GN=terZ P
BAC0253|mntR|sp|P0A9F1|MNTR_ECOLI Transcriptional regulator MntR OS=Escherichia coli (strain K12) GN
BAC0266|ncrY|tr|Q06VT0|Q06VT0_9BACT NcrY OS=Leptospirillum ferriphilum GN=ncrY PE=4 SV=1
BAC0433|ybtQ|tr|Q9Z375|Q9Z375_YERPE Inner membrane ABC-transporter YbtQ OS=Yersinia pestis GN=yb
BAC0272|nikC|sp|P0AFA9|NIKC_ECOLI Nickel transport system permease protein NikC OS=Escherichia coli (
BAC0439|yfeA|sp|Q56952|YFEA_YERPE Periplasmic chelated iron-binding protein YfeA OS=Yersinia pestis G
BAC0076|comR/ycfQ|sp|P75952|COMR_ECOLI HTH-type transcriptional repressor ComR OS=Escherichia coli
BAC0120|czcB|sp|P13510|CZCB_RALME Cobalt-zinc-cadmium resistance protein CzcB OS=Ralstonia metallid
BAC0596|baeR|tr|D0ZNE3|D0ZNE3_SALT1 DNA-binding transcriptional regulator BaeR OS=Salmonella typhi
BAC0278|nirC|tr|Q6RUG1|Q6RUG1_KLEOX NirC OS=Klebsiella oxytoca GN=nirC PE=4 SV=1
BAC0673|merE|sp|P06690|MERE_PSEAI Uncharacterized mercuric resistance protein MerE OS=Pseudomonas a
BAC0582|arsC|sp|P08692|ARSC1_ECOLX Arsenate reductase OS=Escherichia coli GN=arsC PE=1 SV=1
BAC0451|yodD|sp|P64519|YODD_ECOLI Uncharacterized protein YodD OS=Escherichia coli (strain K12) GN=
BAC0656|merB3|tr|Q7DHE7|Q7DHE7_BACCE Organomercurial lyase enzyme OS=Bacillus cereus GN=merB3
BAC0263|ncrA|tr|Q06VT3|Q06VT3_9BACT NcrA OS=Leptospirillum ferriphilum GN=ncrA PE=4 SV=1
BAC0594|arsR|sp|P37309|ARSR_ECOLI Arsenical resistance operon repressor OS=Escherichia coli (strain K12)
BAC0164|fecE|sp|P15031|FECE_ECOLI Fe(3+) dicitrate transport ATP-binding protein FecE OS=Escherichia co
BAC0591|arsR|sp|P52144|ARSR2_ECOLX Arsenical resistance operon repressor OS=Escherichia coli GN=arsR
BAC0356|recG|tr|Q9HTL3|Q9HTL3_PSEAE ATP-dependent DNA helicase RecG OS=Pseudomonas aeruginosa
BAC0228|merF|tr|Q2QCN0|Q2QCN0_9PSED MerF OS=Pseudomonas sp. CT14 GN=merF PE=4 SV=1

BAC0432|ybtP|tr|Q9R7V3|Q9R7V3_YERPE Lipoprotein inner membrane ABC-transporter OS=Yersinia pestis C
BAC0689|merR|tr|Q934S8|Q934S8_THIFE Mer operon regulatory protein OS=Thiobacillus ferrooxidans GN=m
BAC0391|terW|sp|P75010|TERW_SERMA Tellurium resistance protein TerW OS=Serratia marcescens GN=ter
BAC0683|merR1|tr|O07300|O07300_9PSED Mercuric resistance operon regulatory protein OS=Pseudomonas sp
BAC0031|arsB|sp|P08691|ARSB1_ECOLX Arsenical pump membrane protein OS=Escherichia coli GN=arsB P
BAC0645|mdtA|tr|D0ZND8|D0ZND8_SALT1 Multidrug resistance protein MdtA OS=Salmonella typhimurium
BAC0084|copY/tcrY|sp|Q47839|COPY_ENTHA Transcriptional repressor CopY OS=Enterococcus hirae (strain
BAC0308|pcoR|sp|Q47456|PCOR_ECOLX Transcriptional regulatory protein PcoR OS=Escherichia coli GN=pc
BAC0695|merT|sp|P13112|MERT_SERMA Mercuric transport protein OS=Serratia marcescens GN=merT PE=4
BAC0707|sodB|sp|P0AGD3|SODF_ECOLI Superoxide dismutase [Fe] OS=Escherichia coli (strain K12) GN=so
BAC0020|aioA/aoxB|sp|Q8GGJ6|AIOA_HERAR Arsenite oxidase subunit AioA OS=Herminiimonas arsenicoxy
BAC0171|furA|sp|P0A582|FURA_MYCTU Transcriptional regulator FurA OS=Mycobacterium tuberculosis GN
BAC0648|merA|sp|P08662|MERA_SERMA Mercuric reductase (Fragments) OS=Serratia marcescens GN=merA
BAC0182|golS|tr|Q8ZRG6|Q8ZRG6_SALTY Putative transcriptional regulator OS=Salmonella typhimurium (st
BAC0058|cadR|tr|Q93TP7|Q93TP7_PSEPU CadR OS=Pseudomonas putida GN=cadR PE=4 SV=1
BAC0649|merA|tr|E3VST6|E3VST6_9FLAO MerA OS=Tenacibaculum discolor GN=merA PE=3 SV=1
BAC0317|pstC|sp|P0AGH8|PSTC_ECOLI Phosphate transport system permease protein PstC OS=Escherichia co
BAC0597|baeS|tr|D0ZNE2|D0ZNE2_SALT1 Signal transduction histidine-protein kinase BaeS OS=Salmonella t
BAC0698|ncrA|tr|Q1KLR2|Q1KLR2_SERMA NcrA OS=Serratia marcescens GN=ncrA PE=4 SV=1
BAC0032|arsC|sp|P0A006|ARSC_STAAU Protein ArsC OS=Staphylococcus aureus GN=arsC PE=1 SV=1
BAC0676|merP|tr|Q7DHE4|Q7DHE4_BACCE Mercury-binding protein OS=Bacillus cereus GN=merP PE=4 S
BAC0440|yfeB|sp|Q56953|YFEB_YERPE Chelated iron transport system membrane protein YfeB OS=Yersinia
BAC0566|actS|tr|Q52912|Q52912_9RHIZ Histidine protein kinase OS=Sinorhizobium medicae GN=actS PE=4
BAC0079|copB|sp|P05425|COPB_ENTHA Copper-exporting P-type ATPase B OS=Enterococcus hirae (strain A
BAC0231|merP|sp|P13113|MERP_SERMA Mercuric transport protein periplasmic component OS=Serratia marc
BAC0610|modB|sp|P0AF01|MODB_ECOLI Molybdenum transport system permease protein ModB OS=Escheri
BAC0541|yieF|sp|P0AGE6|YIEF_ECOLI Uncharacterized protein YieF OS=Escherichia coli (strain K12) GN=y
BAC0113|cutA|sp|P69488|CUTA_ECOLI Divalent-cation tolerance protein CutA OS=Escherichia coli (strain K1
BAC0229|merG|tr|O07302|O07302_9PSED Mercuric resistance protein OS=Pseudomonas sp. K-62 GN=merG P
BAC0489|ALU1-P|tr|O52119|O52119_ARTVI Aluminum resistance protein (Fragment) OS=Arthrobacter viscos
BAC0233|merT|sp|P94185|MERT_ALCSP Mercuric transport protein OS=Alcaligenes sp. GN=merT PE=3 SV=
BAC0303|pcoA|sp|Q47452|PCOA_ECOLX Copper resistance protein A OS=Escherichia coli GN=pcoA PE=3 S
BAC0166|fetB/ybbM|sp|P77307|YBBM_ECOLI UPF0014 inner membrane protein YbbM OS=Escherichia coli (
BAC0203|cnrA|sp|P37972|CNRA_RALME Nickel and cobalt resistance protein CnrA OS=Ralstonia metallidura
BAC0442|yfeD|sp|Q56955|YFED_YERPE Chelated iron transport system membrane protein YfeD OS=Yersinia
BAC0117|cutO|tr|D5AV58|D5AV58_RHOCB Multicopper oxidase family protein OS=Rhodobacter capsulatus (
BAC0661|merB2|tr|Q7DJN2|Q7DJN2_BACME MerB2 OS=Bacillus megaterium GN=merB2 PE=4 SV=1
BAC0088|corC|sp|P0A2L3|CORC_SALTY Magnesium and cobalt efflux protein CorC OS=Salmonella typhimu
BAC0469|zupT/ygiE|sp|P0A8H3|ZUPT_ECOLI Zinc transporter ZupT OS=Escherichia coli (strain K12) GN=zu
BAC0138|dsbC|sp|P0AEG6|DSBC_ECOLI Thiol:disulfide interchange protein DsbC OS=Escherichia coli (strain
BAC0275|nikR|sp|P0A6Z6|NIKR_ECOLI Nickel-responsive regulator OS=Escherichia coli (strain K12) GN=nik
BAC0116|cutF/nlpE|sp|P40710|NLPE_ECOLI Lipoprotein NlpE OS=Escherichia coli (strain K12) GN=nlpE PE=
BAC0482|dmeF|tr|Q1MJL2|Q1MJL2_RHIL3 Putative cation efflux system protein OS=Rhizobium leguminosaru
BAC0199|klaB/telA/kilB|sp|Q52328|KLAB_ECOLX Protein KlaB OS=Escherichia coli GN=klaB PE=3 SV=1
BAC0446|yhcN|sp|P64614|YHCN_ECOLI Uncharacterized protein YhcN OS=Escherichia coli (strain K12) GN=
BAC0316|pstB|sp|P0AAH0|PSTB_ECOLI Phosphate import ATP-binding protein PstB OS=Escherichia coli (str

BAC0638|copR|tr|C6FFR4|C6FFR4_PSEFL CopR OS=Pseudomonas fluorescens GN=copR PE=4 SV=1
BAC0548|chrA1|sp|P17551|CHRA1_RALME Chromate transport protein OS=Ralstonia metallidurans (strain CH
BAC0048|bfrA|sp|P63697|BFR_MYCTU Bacterioferritin OS=Mycobacterium tuberculosis GN=bfr PE=1 SV=1
BAC0627|copB|tr|F4ZD00|F4ZD00_9XANT Copper resistance protein B OS=Xanthomonas alfalfae subsp. citru
BAC0267|nczA|tr|B8GZE9|B8GZE9_CAUCN Cobalt-zinc-cadmium resistance protein czcA OS=Caulobacter cr
BAC0642|mgtA|sp|P36640|ATMA_SALTY Magnesium-transporting ATPase, P-type 1 OS=Salmonella typhimu
BAC0572|arsA|sp|P08690|ARSA1_ECOLX Arsenical pump-driving ATPase OS=Escherichia coli GN=arsA PE=
BAC0063|chrA|sp|P14285|CHRA_PSEAI Chromate transport protein OS=Pseudomonas aeruginosa GN=chrA P
BAC0678|merP|tr|O66016|O66016_PSEST MerP OS=Pseudomonas stutzeri GN=merP PE=4 SV=1
BAC0133|dnaK|sp|P0A5B9|DNAK_MYCTU Chaperone protein DnaK OS=Mycobacterium tuberculosis GN=dn
BAC0694|merT-P|tr|H6WCN3|H6WCN3_9FLAO MerT-P OS=Tenacibaculum discolor GN=merT-P PE=4 SV=
BAC0098|ctpC|sp|P0A502|CTPC_MYCTU Probable manganese/zinc-exporting P-type ATPase OS=Mycobacteri
BAC0269|nia|tr|Q92Z60|Q92Z60_RHIME Cation transport P-type ATPase OS=Rhizobium meliloti (strain 1021)
BAC0125|czcR|sp|Q44006|CZCR_RALME Transcriptional activator protein CzcR OS=Ralstonia metallidurans (
BAC0608|modE|sp|P0A9G8|MODE_ECOLI Transcriptional regulator ModE OS=Escherichia coli (strain K12) G
BAC0028|chrC|tr|A4UQR3|A4UQR3_9RHIZ Superoxide dismutase OS=Ochrobactrum tritici GN=chrC PE=3 S
BAC0254|mrdH|tr|Q88IN1|Q88IN1_PSEPK Membrane protein, putative OS=Pseudomonas putida (strain KT244
BAC0112|cusS|sp|P77485|CUSS_ECOLI Sensor kinase CusS OS=Escherichia coli (strain K12) GN=cusS PE=1
BAC0619|copA|tr|Q7WYH1|Q7WYH1_PSEPU CopA OS=Pseudomonas putida GN=copA PE=4 SV=1
BAC0003|acn|tr|O53166|O53166_MYCTU Aconitate hydratase OS=Mycobacterium tuberculosis H37Rv GN=R
BAC0039|baeR|sp|P69228|BAER_ECOLI Transcriptional regulatory protein BaeR OS=Escherichia coli (strain K
BAC0357|recG|tr|B5L350|B5L350_9PSED ATP-dependent DNA helicase (Fragment) OS=Pseudomonas corruga
BAC0115|cutE/lnt|sp|P23930|LNT_ECOLI Apolipoprotein N-acyltransferase OS=Escherichia coli (strain K12) G
BAC0315|pstA|sp|P07654|PSTA_ECOLI Phosphate transport system permease protein PstA OS=Escherichia col
BAC0697|merT|tr|Q7DHE5|Q7DHE5_BACCE Mercury transport protein OS=Bacillus cereus GN=merT PE=4 S
BAC0688|merR2|tr|Q79B70|Q79B70_PSEST Organomercurial resistance regulatory protein OS=Pseudomonas s
BAC0621|copA|tr|F4ZBX3|F4ZBX3_XANCI CopA OS=Xanthomonas citri subsp. citri GN=copA PE=4 SV=1
BAC0465|znuC/yebM|sp|P0A9X1|ZNUC_ECOLI Zinc import ATP-binding protein ZnuC OS=Escherichia coli (
BAC0650|merA|tr|O08449|O08449_9PSED Mercuric reductase OS=Pseudomonas sp. K-62 GN=merA PE=4 SV
BAC0333|ricR|tr|O07434|O07434_MYCTU Regulated in copper repressor OS=Mycobacterium tuberculosis H37
BAC0641|corA|sp|P0A2R8|CORA_SALTY Magnesium transport protein CorA OS=Salmonella typhimurium (st
BAC0274|nikE|sp|P33594|NIKE_ECOLI Nickel import ATP-binding protein NikE OS=Escherichia coli (strain K
BAC0302|pbrT|tr|Q5GR69|Q5GR69_ALCXX Lead uptake protein PbrT OS=Alcaligenes xylosoxydans xylosoxy
BAC0679|merP|tr|O66047|O66047_PSEST Mercury transport protein OS=Pseudomonas stutzeri GN=merP PE=
BAC0647|mdtC|tr|D0ZNE0|D0ZNE0_SALT1 Multidrug resistance protein MdtC OS=Salmonella typhimurium (
BAC0341|silA|sp|Q9ZHC9|SILA_SALTM Putative cation efflux system protein SilA OS=Salmonella typhimuri
BAC0464|znuB/yebI|sp|P39832|ZNUB_ECOLI High-affinity zinc uptake system membrane protein ZnuB OS=E
BAC0034|arsH|tr|E8PS81|E8PS81_YERPE Arsenic resistance protein ArsH OS=Yersinia pestis Java 9 GN=arsH
BAC0583|arsC|sp|P52147|ARSC2_ECOLX Arsenate reductase OS=Escherichia coli GN=arsC PE=3 SV=1
BAC0103|cueO|sp|P36649|CUEO_ECOLI Blue copper oxidase CueO OS=Escherichia coli (strain K12) GN=cue
BAC0167|fieF/yiip|sp|P69380|FIEF_ECOLI Ferrous-iron efflux pump FieF OS=Escherichia coli (strain K12) GN
BAC0612|perO|tr|D5AQ60|D5AQ60_RHOCB Divalent ion symporter OS=Rhodobacter capsulatus (strain ATCC
BAC0309|pcoS|sp|Q47457|PCOS_ECOLX Probable sensor protein PcoS OS=Escherichia coli GN=pcoS PE=3 S
BAC0574|arsB|sp|P30329|ARSB_STAAU Arsenical pump membrane protein OS=Staphylococcus aureus GN=a
BAC0358|oscA|tr|B6CM35|B6CM35_9PSED Putative uncharacterized protein oscA OS=Pseudomonas corrugat
BAC0271|nikB|sp|P33591|NIKB_ECOLI Nickel transport system permease protein NikB OS=Escherichia coli (s

BAC0386|terA|sp|P18778|TERA_ALCSP Tellurium resistance protein TerA OS=Alcaligenes sp. GN=terA PE=4
BAC0276|nirA|tr|Q6RUG3|Q6RUG3_KLEOX NirA OS=Klebsiella oxytoca GN=nirA PE=4 SV=1
BAC0461|zntA/yhhO|sp|P37617|ATZN_ECOLI Lead, cadmium, zinc and mercury-transporting ATPase OS=Esc
BAC0087|mgtA|sp|P0ABB8|ATMA_ECOLI Magnesium-transporting ATPase, P-type 1 OS=Escherichia coli (st
BAC0350|sitB|tr|Q9XCS1|Q9XCS1_SALTM SitB OS=Salmonella typhimurium GN=sitB PE=3 SV=1
BAC0136|dsbA|sp|P0AEG4|DSBA_ECOLI Thiol:disulfide interchange protein DsbA OS=Escherichia coli (strai
BAC0342|silB|sp|Q9ZHD0|SILB_SALTM Putative membrane fusion protein SilB OS=Salmonella typhimurium
BAC0122|czcD|sp|P13512|CZCD_RALME Cobalt-zinc-cadmium resistance protein CzcD OS=Ralstonia metalli
BAC0131|czrB|tr|Q9RLI9|Q9RLI9_PSEAI CzrB protein OS=Pseudomonas aeruginosa GN=czrB PE=4 SV=1
BAC0568|actP|sp|Q9X5V3|ATCU_RHILV Copper-transporting P-type ATPase OS=Rhizobium leguminosarum
BAC0264|ncrB|tr|Q06VT2|Q06VT2_9BACT NcrB OS=Leptospirillum ferriphilum GN=ncrB PE=4 SV=1
BAC0082|copL|tr|Q5YKV8|Q5YKV8_9XANT CopL OS=Xanthomonas perforans GN=copL PE=4 SV=1
BAC0306|pcoD|sp|Q47455|PCOD_ECOLX Copper resistance protein D OS=Escherichia coli GN=pcoD PE=3 S
BAC0224|merA|sp|P16171|MERA_BACCE Mercuric reductase OS=Bacillus cereus GN=merA PE=1 SV=1
BAC0675|merP|tr|O07301|O07301_9PSED Mercuric transport protein periplasmic component OS=Pseudomonas
BAC0654|merB1|sp|P16172|MERB_BACCE Alkylmercury lyase OS=Bacillus cereus GN=merB1 PE=3 SV=2
BAC0183|golT|tr|Q8ZRG7|Q8ZRG7_SALTY Putative cation transport ATPase OS=Salmonella typhimurium (st
BAC0385|tehB|sp|P25397|TEHB_ECOLI Tellurite methyltransferase OS=Escherichia coli (strain K12) GN=tehB
BAC0330|rcnA/yohM|sp|P76425|RCNA_ECOLI Nickel/cobalt efflux system RcnA OS=Escherichia coli (strain K
BAC0100|ctpG|sp|P63689|CTPG_MYCTU Probable cation-transporting ATPase G OS=Mycobacterium tubercul
BAC0312|pitA|sp|P0AFJ7|PITA_ECOLI Low-affinity inorganic phosphate transporter 1 OS=Escherichia coli (st
BAC0576|arsB|sp|P52146|ARSB2_ECOLX Arsenical pump membrane protein OS=Escherichia coli GN=arsB P
BAC0305|pcoC|sp|Q47454|PCOC_ECOLX Copper resistance protein C OS=Escherichia coli GN=pcoC PE=1 S
BAC0111|cusR/ylcA|sp|P0ACZ8|CUSR_ECOLI Transcriptional regulatory protein CusR OS=Escherichia coli (s
BAC0343|silC|sp|Q9ZHD2|SILC_SALTM Probable outer membrane lipoprotein SilC OS=Salmonella typhimuri
BAC0265|ncrC|tr|D5CKG5|D5CKG5_ENTCC Nickel-resistant membrane protein-like protein NcrC OS=Entero
BAC0083|copR|sp|Q02540|COPR_PSEUB Transcriptional activator protein CopR OS=Pseudomonas syringae p
BAC0458|zipB|tr|Q7WJT8|Q7WJT8_BORBR Putative membrane protein OS=Bordetella bronchiseptica (strain A
BAC0056|cadC|sp|P20047|CADC_STAAU Cadmium resistance transcriptional regulatory protein CadC OS=Sta
BAC0033|arsD|sp|P46003|ARSD1_ECOLX Arsenical resistance operon trans-acting repressor ArsD OS=Escheri
BAC0348|silS|sp|Q9ZHD4|SILS_SALTM Probable sensor kinase SilS OS=Salmonella typhimurium GN=silS P
BAC0682|merR1|sp|P22853|MERR_BACCE Mercuric resistance operon regulatory protein OS=Bacillus cereus
BAC0434|ychH|sp|P0AB49|YCHH_ECOLI Uncharacterized protein YchH OS=Escherichia coli (strain K12) GN
BAC0640|copD|tr|C6FFR7|C6FFR7_PSEFL CopD OS=Pseudomonas fluorescens GN=copD PE=4 SV=1
BAC0455|ziaA|sp|Q59998|ATZN_SYNY3 Zinc-transporting ATPase OS=Synechocystis sp. (strain PCC 6803 / 
BAC0646|mdtB|tr|D0ZND9|D0ZND9_SALT1 Multidrug resistance protein MdtB OS=Salmonella typhimurium (
BAC0667|merD|sp|P08654|MERD_SERMA HTH-type transcriptional regulator MerD OS=Serratia marcescens 
BAC0165|fetA/ybbL|sp|P77279|YBBL_ECOLI Uncharacterized ABC transporter ATP-binding protein YbbL OS
BAC0625|copA|tr|F4ZCZ9|F4ZCZ9_9XANT Copper resistance protein A OS=Xanthomonas alfalfae subsp. citru
BAC0462|zntR/yhdM|sp|P0ACS5|ZNTR_ECOLI HTH-type transcriptional regulator ZntR OS=Escherichia coli (
BAC0345|silF|tr|Q9ZHD1|Q9ZHD1_SALTM Uncharacterized protein OS=Salmonella typhimurium GN=ORF96
BAC0106|cuiD|sp|Q8ZRS2|CUEO_SALTY Blue copper oxidase CueO OS=Salmonella typhimurium (strain LT2
BAC0390|terE|sp|P18782|TERE_ALCSP Tellurium resistance protein TerE OS=Alcaligenes sp. GN=terE PE=3 S
BAC0609|modA|sp|P37329|MODA_ECOLI Molybdate-binding periplasmic protein OS=Escherichia coli (strain 
BAC0628|copB|tr|F4ZBX4|F4ZBX4_XANCI CopB OS=Xanthomonas citri subsp. citri GN=copB PE=4 SV=1
BAC0626|copB|sp|P12375|COPB_PSEUB Copper resistance protein B OS=Pseudomonas syringae pv. tomato G

| |
|---|
| BAC0681\|merR2\|tr\|Q9WWL1\|Q9WWL1_BACSR Mercury resistance operon negative regulator MerR2 OS=Bac |
| BAC0351\|sitC\|tr\|Q9XCS0\|Q9XCS0_SALTM SitC OS=Salmonella typhimurium GN=sitC PE=3 SV=1 |
| BAC0304\|pcoB\|sp\|Q47453\|PCOB_ECOLX Copper resistance protein B OS=Escherichia coli GN=pcoB PE=4 S |
| BAC0490\|G2alt\|tr\|B0FSM1\|B0FSM1_9BACI 7-cyano-7-deazaguanine synthase OS=Anoxybacillus gonensis G |
| BAC0071\|cmtR\|sp\|P67731\|CMTR_MYCTU HTH-type transcriptional regulator CmtR OS=Mycobacterium tube |
| BAC0685\|merR\|tr\|H6WCN2\|H6WCN2_9FLAO MerR OS=Tenacibaculum discolor GN=merR PE=4 SV=1 |
| BAC0538\|chrR\|tr\|Q7BD45\|Q7BD45_PSEPU Chromate reductase OS=Pseudomonas putida GN=chrR PE=4 SV= |
| BAC0255\|mreA\|tr\|Q88IN0\|Q88IN0_PSEPK Putative uncharacterized protein OS=Pseudomonas putida (strain K |
| BAC0587\|arsD\|sp\|P52148\|ARSD2_ECOLX Arsenical resistance operon trans-acting repressor ArsD OS=Escheri |
| BAC0643\|corB\|tr\|Q9X621\|Q9X621_SALTM CorB OS=Salmonella typhimurium GN=corB PE=4 SV=1 |
| BAC0466\|zraP\|sp\|Q9L9I0\|ZRAP_SALTY Zinc resistance-associated protein OS=Salmonella typhimurium (strai |
| BAC0690\|merT\|tr\|Q934S7\|Q934S7_THIFE Mercuric ion transport protein OS=Thiobacillus ferrooxidans GN=m |
| BAC0653\|merA\|tr\|Q934S5\|Q934S5_THIFE Mercuric ion reductase OS=Thiobacillus ferrooxidans GN=merA PE |
| BAC0066\|chrF\|tr\|Q5NUZ7\|Q5NUZ7_RALME ChrF, regulatory protein, involved in Chromate resistance OS=Ra |
| BAC0468\|zraS/hydG\|sp\|P14377\|ZRAS_ECOLI Sensor protein ZraS OS=Escherichia coli (strain K12) GN=zraS |
| BAC0498\|ideR\|sp\|P0A672\|IDER_MYCTU Iron-dependent repressor IdeR OS=Mycobacterium tuberculosis GN= |
| BAC0135\|dpsA\|tr\|Q8KR86\|Q8KR86_BURPE DpsA OS=Burkholderia pseudomallei GN=dpsA PE=3 SV=1 |
| BAC0659\|merB\|sp\|P08664\|MERB_SERMA Alkylmercury lyase OS=Serratia marcescens GN=merB PE=3 SV=1 |
| BAC0620\|copA\|sp\|P32113\|COPA_ENTHA Probable copper-importing P-type ATPase A OS=Enterococcus hira |
| BAC0162\|fbpC\|sp\|P44513\|FBPC2_HAEIN Fe(3+) ions import ATP-binding protein FbpC 2 OS=Haemophilus in |
| BAC0693\|merT\|tr\|Q79F00\|Q79F00_9PSED Mercuric transport protein OS=Pseudomonas sp. K-62 GN=merT PE |
| BAC0584\|arsC\|sp\|O50595\|ARSC_ACIMA Arsenate reductase OS=Acidiphilium multivorum (strain DSM 11245 |
| BAC0179\|gesB\|tr\|Q8ZRG9\|Q8ZRG9_SALTY Putative cation efflux system protein OS=Salmonella typhimurium |
| BAC0470\|zur/yjbK\|sp\|P0AC51\|ZUR_ECOLI Zinc uptake regulation protein OS=Escherichia coli (strain K12) G |
| BAC0124\|czcP\|tr\|Q1LAJ7\|Q1LAJ7_RALME CzcP cation efflux P1-ATPase OS=Ralstonia metallidurans (strain |
| BAC0578\|arsB\|tr\|O50594\|O50594_ACIMU ArsB OS=Acidiphilium multivorum GN=arsB PE=4 SV=1 |
| BAC0579\|arsB\|sp\|P74311\|Y944_SYNY3 Uncharacterized transporter slr0944 OS=Synechocystis sp. (strain PCC |
| BAC0691\|merT\|tr\|Q52397\|Q52397_PSEST Mercury transport protein OS=Pseudomonas stutzeri GN=merT PE= |
| BAC0252\|mntP/yebN\|sp\|P76264\|MNTP_ECOLI Probable manganese efflux pump MntP OS=Escherichia coli (st |
| BAC0029\|chrF\|tr\|A4UQR2\|A4UQR2_9RHIZ ChrF OS=Ochrobactrum tritici GN=chrF PE=4 SV=1 |
| BAC0589\|arsR\|sp\|P15905\|ARSR1_ECOLX Arsenical resistance operon repressor OS=Escherichia coli GN=arsR |
| BAC0347\|silR\|sp\|Q9ZHD3\|SILR_SALTM Probable transcriptional regulatory protein SilR OS=Salmonella typhi |
| BAC0204\|cnrB\|sp\|P37973\|CNRB_RALME Nickel and cobalt resistance protein CnrB OS=Ralstonia metallidura |
| BAC0092\|corT/coaT\|tr\|H0P0Y3\|H0P0Y3_9SYNC Cation-transporting ATPase E1-E2 ATPase OS=Synechocysti |
| BAC0332\|rcnR/yohL\|sp\|P64530\|RCNR_ECOLI Transcriptional repressor RcnR OS=Escherichia coli (strain K12 |
| BAC0105\|cueR/ybbI\|sp\|P0A9G4\|CUER_ECOLI HTH-type transcriptional regulator CueR OS=Escherichia coli ( |
| BAC0571\|arsA\|sp\|O50593\|ARSA_ACIMA Arsenical pump-driving ATPase OS=Acidiphilium multivorum (strai |
| BAC0318\|pstS\|sp\|P0AG82\|PSTS_ECOLI Phosphate-binding protein PstS OS=Escherichia coli (strain K12) GN= |

ebruary); A (April); STP (sewage treatment plant); inf (influent); T (treated).

| Relativ abundance |
|---:|
| 0.0015524080301735994 |
| 0.002544224271673399 |
| 0.00401204596686959 |
| 0.0018503449248533808 |
| 0.016710759998875134 |
| 0.005607105832889033 |
| 0.0020619856842320357 |
| 0.0010437843165839586 |
| 0.0007828382374379688 |
| 1.1402685811421398e-05 |
| 0.004149378640497388 |
| 0.00011049833970351353 |
| 0.0019247086688782213 |
| 8.811166308825624e-05 |
| 6.523651978649741e-06 |
| 0.001168044354272525 |
| 0.0028566728664403073 |
| 0.0006895262768901558 |
| 1.884610571609925e-05 |
| 9.449300916150041e-05 |
| 0.0176573513555453 |
| 0.00013216749463238435 |
| 0.0007067289643537219 |
| 0.0018539308646302285 |
| 0.00014283364332201538 |
| 0.002244136280655511 |
| 0.04189277659929485 |
| 0.002329041124317937 |
| 0.00040458612271258946 |
| 0.00041316462531448356 |
| 0.000742065412571408 |
| 0.0015499297287205763 |
| 1.6752093969866e-05 |
| 0.00014308466905720344 |
| 0.0002272353416486608 |
| 0.0014134579287074438 |
| 0.0003739542236580323 |
| 0.001520980421017179 |
| 0.0010345695272722221 |
| 8.588098807336367e-05 |
| 0.0015887982793572279 |
| 0.0016653104323680428 |
| 1.6649320387228787e-05 |

0.0034780249971371607
0.00971621450252228
0.006036796668659927
0.00022920939384445035
1.2610777059099869e-05
3.126542883776834e-05
8.811166308825624e-06
0.000597437464679238
0.005456345761973749
8.45959857580515e-06
0.000597761943418126
0.0013861006784743966
0.00039402384364693895
0.00026994421980619613
0.001335266639034266
0.0020117122148115246
0.0001641435013982838
0.00019689280941607922
0.0028269158574148875
0.0032105687237783363
0.006667622229213045
0.004995508843924938
0.003189340967339873
0.0026007625888216966
0.003083908208088968
0.0032753232003151804
0.0019031741875187168
0.0021493192320497924
0.005324621260548547
0.0007167520379436879
0.003911405285988998
2.113581949469075e-05
9.046130743727642e-05
0.0013405052614193175
0.0010684406390229497
3.9331003233598434e-05
4.6153728284324696e-05
0.0015265345630040394
0.00010202403094429668
9.692282939708185e-05
0.0005561145949012893
6.951432436266116e-05
0.01104671735051356
0.015500921951491633
0.02690281590973168
0.002133301451162575

0.0011521015569841806
0.00012319633308578498
6.523651978649741e-05
0.0003124485947669086
0.00021576460254474052
0.016327768952277635
0.0029051071470880653
0.0005342203195114749
0.004289776199164533
0.005200528907737696
0.00010487488079826931
0.00010350981780270877
2.4761306780276383e-05
0.0024967320852688287
0.0004823173974736301
0.0013310187069906787
0.004588427588220234
0.003040093118793715
0.0018003356025600463
0.00036184522974910566
9.209861616917281e-05
0.0029415739831002465
6.571039281158093e-05
0.002290903240294662
0.0001400949451462245
0.001549861560988302
0.0006078686121146463
0.002359860194015906
0.002795505681221389
0.0012915974420748982
2.331476995806093e-05
0.0048349257133182685
0.00042075026715012283
1.6835230912644492e-05
0.002598356702985599
1.3408296556908558e-05
0.0035892712305758056
0.003986121997739141
0.004919825492202752
0.0005953577820369196
0.0005787808769683562
0.024643095349349393
0.0017554554415275666
0.0008420600426342218
0.0018973875924343988
8.716828339780382e-05

9.475695611446551e-05
0.0008480747572244663
0.00237195908661217\9
0.0022576535561104148
0.0026169163937212105
0.004142807663609782
1.62310958320472e-05
5.899650485039765e-05
0.003064971929618246
0.002110763840203116
0.007999407845171315
0.004566556385054818
0.00018716132573229601
0.0032238870203710208
0.000967692274487505
5.094817064677644e-05
0.00016001410513669174
0.003257802591628787
8.86875563110553e-05
0.0054833535829704036
0.0032397042628920787
0.0020030718075396917
0.0007785604328618051
0.003487410216623973
8.420600426342218e-05
6.619120056386078e-05
8.534085607290227e-05
0.0001884610571609925
0.0033846067408504774
0.003738980973589433
0.0007851172424960124
0.00013479333889660392
0.0006330698891957283
0.001584220866287328
0.0001443531501658666
0.019768915030473764
0.0002094730103607601
3.3587119097998664e-05
1.4253357264276743e-05
1.8639005653284975e-05
0.0002800980849548696
0.0009856558921737138
4.6153728284324696e-05
0.0035092748574805503
0.002035379417338719
0.0036505055166617447

0.0012991783514927995
4.523065371863821e-05
0.003408766499911932
0.013135186079909326
0.015791736858662475
0.0030492575540654963
2.09401174623325e-05
0.002544224271673399
0.0021587357456622778
0.00020461486206050614
0.002087568633167917
0.000683522938658525
0.0030021515772312487
1.62310958320472e-05
0.0015378422264336988
0.004041888204644265
0.004381719578993076
0.002213414969209955
0.003407112561775148
9.209861616917281e-05
0.032357313814102714
0.005001668800747085
0.0018988340476220754
0.0004262574695997317
0.0004111877610785291
0.0029039135655935027
0.0007591158666065153
8.735533979994502e-05
7.623143885163741e-05
3.5428710484573e-05
0.0001522727743644927
0.0011921817673442978
0.0002924395714567125
8.698202638199654e-05
0.00014469521924208905
0.0044733613567883935
0.001303707077772513
0.0073109892864178135
0.014359447593914259
0.0027892236459826887
0.0011759969966845933
3.5521455799977643e-05
0.022764849681529475
0.0003392299028897865
0.0001130766342965955
0.014181694551364686

0.004792295453522381
0.0023147452197185433
0.00013340501799036548
0.0035956433094121994
0.0038643665313011282
0.000294508275820715
0.00015668817685440483
0.026289256053736648
0.0021742148216102146
0.008904784950856896
0.0010647361915518845
0.0029528820562427006
0.003755759639136922
0.00013846118485297406
0.007805478016335526
8.986222593106928e-05
0.0006057676837317616
0.0018697711182901618
0.0001730300175813676
0.0029890187247631535
0.000168213174986671
0.001809226148745528
0.0032207041363855214
0.02186771274826723
0.001036535814385459
7.34794013479682e-05
0.003716504763594397
0.0014816938287140102
0.003919480634740101
0.0019421559325751134
5.0443108236399476e-05
0.013173207902101436
0.00526937115822135
0.0019120230890151604
4.508038576608459e-05
0.0029939151571767636
0.0014691058786565556
0.0008794849334179649
8.273900070482598e-05
0.00011198786340240544
8.41065874933355e-05
0.003202330283279584
0.00017396405276399308
0.011405143286811789
1.9608664906924074e-05
0.00040105998371206287

0.00023076864142162348
0.0075674362952337
0.0008698202638199653
0.006422313695486249
0.0002643349892647687
0.00024068085152295494
0.002054950373274668
0.004278186036444441
7.295266728812614e-05
0.0036220563336439356
0.0029994269810076886
0.0004218402937489573
0.0009787616870262693
1.7176197614672733e-05
0.0047094823828517
5.384601633171214e-05
0.00017508640149150272
0.0036446187913778713
0.0032038379717368726
3.647400803828779e-05
4.1490879032401716e-05
9.000843919125858e-05
9.663243688065854e-05
4.593023234451055e-05
0.00467557351690808
0.008685639268261528
0.0020196179308057637
0.0013626859049653333
0.004822005509294803
0.00040904358752886095
0.0009498762944439203
0.00012540501640130407
0.00015150741800519545
0.0016046232929304244
0.00014782842993001408
0.0004095796865950849
0.0006504106328505864
0.00018177600097257432
0.0012008720874378044
0.0004053648835247965
8.857973380727034e-05
0.0004231676382830735
3.531976727758248e-05
2.2057404070585103e-05
0.003422720914312926
0.003188870417061732

| |
|---|
| 0.0035040494699108567 |
| 5.340213617089025e-05 |
| 3.402758066986605e-05 |
| 0.0003460789599911958 |
| 0.008209364751361402 |
| 0.0006220911141481085 |
| 0.0023746907361098003 |
| 0.0015394546840987672 |
| 0.002134510312467106 |
| 2.287099684368046e-05 |
| 0.005795158738698725 |
| 0.0007252027018199026 |
| 0.009195180321925166 |
| 0.0004905954487787281 |
| 0.0008064339927540509 |
| 0.0014204968221456806 |
| 0.00012540501640130407 |
| 4.852628895528723e-05 |
| 0.00018296797474944368 |
| 0.00026915706896421375 |
| 0.00018448010677216633 |
| 0.0012264886219468202 |
| 2.8952130894205092e-05 |
| 5.566606713075343e-05 |
| 5.797946212839514e-05 |
| 0.002513365424134533 |
| 0.0002696628674830726 |
| 0.0016651561250382834 |
| 0.003567700919851986 |
| 0.008578324964905733 |
| 0.0004912432420649675 |
| 0.0014600925523797067 |
| 0.030907513273059864 |
| 0.0028213335705518333 |
| 0.0029506214778559706 |
| 0.0004772459127575382 |
| 0.0003815742379390107 |
| 0.0008778351148091286 |
| 0.00015831271574065335 |
| 3.981348320945088e-05 |
| 0.00025598730412192805 |
| 0.001104478555909402 |
| 7.739976740441098e-05 |
| 0.0003444767425838291 |
| 0.0010912172353410214 |
| 0.0018455273673546246 |

0.0003957817893516335
0.012510183943857564
0.0001208777594915097
0.0005580523229858032
0.004448530973267612
0.000531478402843622
0.0017877000822921832
0.0008585420353627741
0.0016700835943370754
0.0010942202411486335
0.0019764353105747195
0.0002644797739269209
0.004046695207616351
3.7079888570485266e-05
0.0011905116223697135
0.0024263144477643613
1.554463295225078e-05
0.0020926962111967617
8.857973380727034e-05
0.0010416976695734991
0.0003934984328746048
0.001265952028995572
1.5352195955592933e-05
0.0021923484117299407
0.0018455273673546246
0.002939256167182507
0.0027476622009606337
0.0005772955065370377
0.0028439204921391886
0.0017587103512279856
0.003363603042654156
0.002432535766861193
0.0030227834161731
9.677207913048609e-05
0.003091407328861183
0.002453847093980128
0.00273375679919757
0.0015630949587317334
0.011930773801765444
0.0010529289112939683
0.001958078326265976
5.264644556469841e-05
0.001078055623949847
0.0005177805058631163
0.0007318718989977747
0.0024097713947114226

0.0005979132031990747
0.00010334302277514873
0.0005900112665929196
0.003095208239511422
0.0018270752211634836
0.0016622835152768605
0.0033675571214660536
7.440697639810709e-05
0.001076446105251803
0.0016018168530148053
0.002910476967953168
0.000213608544683561
6.000562612750572e-05
0.002184204791041208
5.743248607058008e-05
0.0018351336005879295
0.0025458254734558954
0.0019081789108546818
0.002016189037884192
0.001797117650293264
0.00026806006780977443
0.00029371174893989637
0.0010982944654507828
5.884558766106887e-05
0.0015035606629669308
0.0030172907751988306
1.0372719758100429e-05
0.011999172601872462
0.0026507485341825646
0.004907011805564821
2.1018919886470925e-05
0.0021281656385051815
9.559782834874572e-05
0.00023496939915191713
0.0029536825771916695
0.00143090339227129
7.97217604265433e-05
0.0033005429219393804
0.001130641464804811
0.00021593318422667108
0.020482359986511895
0.0009847982170337703
4.650436024881693e-05
0.010093468102699744
0.0007030580447065235
0.0015262969517560428

| |
|---|
| 0.002052606245465023 |
| 0.0007124072208329401 |
| 0.0215660862073658 |
| 0.002767201601582495 |
| 0.0033839342989564663 |
| 4.852628895528723e-05 |
| 0.00011565851253591776 |
| 0.0010529289112939683 |
| 0.0003292344088411818 |
| 0.005900760216707834 |
| 0.002564512293332817 |
| 6.270250820065204e-05 |
| 0.010862370568540563 |
| 0.0030656951649005496 |
| 0.015036409813784141 |
| 0.010391319117666678 |
| 0.003076586247446882 |
| 0.0009300872049763384 |
| 0.002655050699668138 |
| 0.00019789089467581674 |
| 0.002079252766585219 |
| 0.0028080339819030855 |
| 0.0003256538754824315 |
| 0.010851017391390615 |
| 9.300872049763386e-05 |
| 0.00033316556596167354 |
| 0.0026056917804784586 |
| 0.0007562064100685915 |
| 0.0032507902309852604 |
| 0.007985921173762355 |
| 0.0011817578604405242 |
| 0.0025567154885693185 |
| 0.0020813140251218767 |
| 0.0002744519621241655 |
| 5.2895954785384186e-05 |
| 3.577258480678225e-05 |
| 0.001964344176910027 |
| 0.003188870417061732 |
| 0.002568028388961218 |
| 0.0010002824657292697 |
| 1.875806127683372e-05 |
| 7.247432766049392e-05 |
| 0.0018601744099526769 |
| 2.9526577935756775e-05 |
| 0.0009468544186397509 |
| 0.004273247007246452 |

| |
|---|
| 0.0009756159492758796 |
| 9.621591775617295e-05 |
| 0.0006074038889641395 |
| 2.9141113471843508e-05 |
| 9.630816695824024e-05 |
| 0.009300872049763386 |
| 0.0025286745885294204 |
| 0.002677142900810272 |
| 0.0030123239579856504 |
| 0.001250037203488199 |
| 0.0007909403002948391 |
| 0.00022887550038018465 |
| 0.000818476740379178 |
| 0.0018098994258999022 |
| 0.00024307179222612113 |
| 0.0011187062885030087 |
| 0.00020541803300090915 |
| 0.0033141972020117568 |
| 0.0001162609006220423 |
| 0.00022174264489502113 |
| 0.00035152902235326177 |
| 0.0020009844250486967 |
| 0.004898335019394425 |
| 0.009443962388990515 |
| 0.006157818736395069 |
| 0.014150612475711435 |
| 0.0016390347947834777 |
| 0.001326266946766991 |
| 0.0032335742079551205 |
| 0.00016910676454115248 |
| 0.0005911950773204336 |
| 5.1433393823576323e-05 |
| 0.006355595900671647 |
| 0.0004348459659629635 |
| 2.1463550884069353e-05 |
| 0.0016643665773260795 |
| 0.002033790688214927 |
| 0.0015630349527829936 |
| 0.0028331887166971543 |
| 0.0022072218744960868 |
| 0.003955813935089238 |
| 0.0006764270581646099 |
| 0.0015346438882109588 |
| 0.00011831498013833282 |
| 2.755813940670633e-05 |
| 0.005032148114785277 |

| |
|---|
| 0.002095214497852835 |
| 0.011858611863448316 |
| 0.0028936046377041644 |
| 0.0014881395279621418 |
| 0.0055116278813412654 |
| 0.002349693991519171 |
| 0.0017736546699548782 |
| 7.575370900259773e-05 |
| 0.0010107986994390065 |
| 0.0001490853538995585 |
| 0.0008001648329395031 |
| 3.2733118505149175e-05 |
| 0.0009451365791998586 |
| 0.0005414993032708964 |
| 0.00026882944134016135 |
| 0.0005922648629525429 |
| 0.0008568938442717642 |
| 0.0001989761219368123 |
| 0.0010967867832454498 |
| 0.0013475202235306706 |
| 0.00016061420012272352 |
| 5.0138824376934846e-05 |
| 0.0009589050162088789 |
| 3.875761884352019e-05 |
| 0.0009101471340287666 |
| 0.0021058306238312635 |
| 0.0006846542914919379 |
| 0.00011434374428043061 |
| 0.0010341551281843958 |
| 0.0007019435412770879 |
| 0.0009290429222784987 |
| 0.0143354138189155 |
| 0.0006731753633558958 |
| 8.151602414830698e-05 |
| 0.0011746810282596166 |
| 0.0001702018412288343 |
| 0.0004142617620651667 |
| 6.074511414897877e-05 |
| 0.0001427257378815482 |
| 3.5491527367942646e-05 |
| 2.5439564583196476e-05 |
| 0.0016198697106394335 |
| 0.001719086730416099 |
| 0.0005509440585605051 |
| 0.0008657804706115688 |
| 0.0008000317217175542 |

0.00021672356334455546
0.0005025277625051879
5.221067662391563e-05
0.007679020292074349
0.002097793102366259
0.00276006926424486
0.006984832562144544
0.0006774000608128541
0.0004704515223452823
0.0007520823656540227
0.009302680338243606
1.9680660035806204e-05
6.793002012358915e-05
0.0001116164641606677
0.0061504428075841
0.0031587459357468955
0.0058998064585024665
0.00021943756043579899
1.640906979608777e-05
0.00013230349469096946
0.0131195640854033
0.005530878097611296
0.0010002695463198503
0.0007553522889829533
3.5227649841043444e-05
4.822512879002894e-05
4.882142095435696e-05
0.000686683899075412
0.0005994700316016005
0.00020055529750773938
0.0006028141098753618
0.0027074965163544815
0.001256109494916894
3.65172940548774e-05
0.0013964982031723118
1.4292968035053826e-05
1.7646625339368134e-05
0.0010241840754749163
0.0011192406858945693
0.0007520823656540227
1.5993650307579217e-05
3.8521291899352386e-05
0.00032083997412664963
0.0013441472067008066
0.000545164273174591
0.00045889771463635286

| |
|---|
| 4.062695737295042e-05 |
| 0.004913604788939615 |
| 3.378337899194541e-05 |
| 4.334471266891109e-05 |
| 8.713781891715574e-05 |
| 0.00045275358412372174 |
| 0.0013115900079225865 |
| 0.0003892293570114658 |
| 0.0005849529510642399 |
| 0.0002699782851065723 |
| 0.0008808041551601921 |
| 0.00016198697106394337 |
| 4.2976135180229874e-05 |
| 0.0002604526850184401 |
| 0.0009283337292080572 |
| 0.003008329462616091 |
| 0.0026465168650852368 |
| 0.001326151186247457 |
| 0.0006803452784685623 |
| 8.982215931033827e-05 |
| 2.7467355963016482e-05 |
| 0.00697102551337246 |
| 0.00021598262808525783 |
| 0.0005969283658104369 |
| 6.370580038481134e-05 |
| 0.000191439147621024 |
| 0.0018435385532472987 |
| 0.000286637562227486 |
| 4.2976135180229874e-05 |
| 0.0006891809314356863 |
| 0.001533371813469367 |
| 1.3556849509643329e-05 |
| 0.004932697653262352 |
| 0.0056094970927919015 |
| 0.00034711493799416437 |
| 0.0009384072806462925 |
| 0.0004020222100041503 |
| 0.0005059463187127061 |
| 5.411127941322305e-05 |
| 0.00010189503018538373 |
| 0.0005467060273408089 |
| 0.0009578868541615134 |
| 0.009996910214231932 |
| 0.00010892227364644468 |
| 5.997618865342207e-05 |
| 5.264576559578159e-05 |

0.0018924645253813998
0.0012745816933715543
0.0005889187337833195
0.00169359955667818
3.342588291795656e-05
0.0019392300997939803
0.00710717835543051
0.00012290840216913992
0.0006673406906507526
2.734844966014628e-05
0.0004512494193924136
0.0005893182715945701
0.00074323433782279
0.0002589136012907291
0.00691840474037179
0.00018258647027438702
0.0016659139524144585
3.21500858600193e-05
0.0018898479957460059
0.0002707496516354482
0.0005959897991975275
0.00304982366210045
3.081703351948191e-05
0.0002208913241781046
7.295025255766502e-05
0.00167486993802393
0.0012102474849604965
0.00170204271295099
4.9115583063119846e-05
0.00012603475055349207
0.0010324305782207753
5.849529510642399e-05
0.0008070804593538323
0.00064667239629464
0.0007358442236683108
0.00198642785650062
0.00011645146306900998
3.127471223511778e-05
0.00043475212879097056
0.0009871081049209049
0.0060564384883742125
0.0005604226660196105
0.00403012412491845
0.0033843706454431024
0.0011305893923028506
0.00023839591967901

| |
|---|
| 0.0015572174352579888 |
| 4.088991502584978e-05 |
| 4.268575588847157e-05 |
| 7.098305473588529e-05 |
| 1.2149022829795752e-05 |
| 0.001139998533201887 |
| 0.0020150620624592263 |
| 0.0003217241230853319 |
| 5.4227398038573314e-05 |
| 0.0009462322626906999 |
| 0.0013657548946134025 |
| 0.00976464504485236 |
| 4.480490689002688e-05 |
| 0.000329036034973635 |
| 0.0008267030378712578 |
| 0.0017610707429385347 |
| 0.00018858184691026244 |
| 0.0015233668342609142 |
| 0.0005083039436834084 |
| 0.001106175619522259 |
| 0.0014672884346695256 |
| 0.0007871635239241465 |
| 0.00014957849835013154 |
| 0.0005264576559578159 |
| 0.0012993422998107797 |
| 0.00029799489959876375 |
| 0.0003220682130565462 |
| 0.0004968813831511971 |
| 0.00012306802347065827 |
| 0.0008510610811338787 |
| 0.0021236955707990795 |
| 1.248516180137113e-05 |
| 0.003089431761608249 |
| 0.00033603680167520165 |
| 0.00022402453445013445 |
| 8.806912460260861e-05 |
| 0.0017972175151663374 |
| 0.00046328273724287805 |
| 0.0006843949527451607 |
| 0.001332849914569016 |
| 0.0009268906992925255 |
| 0.00031587459357468957 |
| 2.0645398272855528e-05 |
| 0.0010529153119156317 |
| 0.005314967972490998 |
| 0.000982720957787923 |

| |
|---|
| 0.0011772753881261395 |
| 9.63833711323602e-05 |
| 0.0003867852166220688 |
| 0.008990276894048856 |
| 0.0015089550648472432 |
| 0.0013687899054903214 |
| 0.0016169413067857563 |
| 0.00015543916731267676 |
| 0.00065807206994727 |
| 0.0024227718465207734 |
| 0.0005114818364229109 |
| 0.0012726986051795743 |
| 0.0007336442173347628 |
| 0.0008265877215038604 |
| 0.003054611454348646 |
| 0.00163518653025293 |
| 0.0005743174428630719 |
| 0.007809121896707603 |
| 4.4178264835620925e-05 |
| 0.0011821463662956189 |
| 0.009432366335910868 |
| 8.367538902640784e-05 |
| 0.005012224955895901 |
| 0.000110188811712101 |
| 4.897280520537823e-05 |
| 0.0005559392846914535 |
| 0.00010529153119156318 |
| 0.0006405848401165032 |
| 0.0010822794839610876 |
| 6.3813049207008e-05 |
| 0.0014641397470414796 |
| 0.0007616834171304571 |
| 0.00025269967485975163 |
| 0.0015154677032774124 |
| 0.0005615548330216703 |
| 0.007856368096601254 |
| 0.0019671905293797394 |
| 0.0003136343482301882 |
| 0.00043736482187264707 |
| 0.00032397394212788674 |
| 0.0004367397917592171 |
| 0.0017104126464306335 |
| 5.959897991975275e-05 |
| 0.0012607575969141623 |
| 0.0002485570572391 |
| 2.1342877944235784e-05 |

0.00035097177063854397
0.005776671777132611
0.0008964008736579027
0.0006829720942155451
1.7379620004109465e-05
0.0010344431134609716
0.0006597380751876428
3.8521291899352386e-05
0.0017109873818629015
0.0002455139021149246
0.0005768621514050863
0.0011215400314788248
9.642508807226483e-05
0.0003222903758937112
0.0006589047684938096
2.025322034304743e-05
0.0005827048973074508
0.001783118574532217
0.00045704377005350953
0.0002928465637750265
0.00010242043033582534
0.0006289545517440909
0.0006481030509775178
0.0010315378050449204
2.614701462277022e-05
4.1124430765910445e-05
0.009449128822905732
0.0008309197301802467
0.00041469530884225784
0.0003294523842469048
0.0074760348732951475
0.0004229103506180553
0.005429920763881254
0.0006652403912677886
0.0025602884275048895
1.3727182676954368e-05
0.000951868846941298
0.0007337149168817531
0.0015278039956488144
0.0007289401994183143
0.00010576949315893496
0.015873614877350867
4.565160058848565e-05
0.00021547122761171974
0.00021179081844443883
2.8565235628344346e-05

0.0004736535875754222
0.00025342491095915753
0.00024708928818517866
0.00015323366709158365
0.0013629225924968388
0.0007751820805809525
0.0062078631081234955
0.00017895045940987867
3.47401459662817e-05
0.0006911588480704297
0.00085944100238323
5.831015650387696e-05
8.82461743518495e-05
0.000201705541375656
0.0003346312004442564
0.0018744704620944584
3.3617590229276004e-05
0.0066417600664176015
0.005636086242901594
0.000857346847893992
0.0001401925039348531
7.97062219952189e-05
3.390590575439844e-05
0.00013601245221202496
0.0007306920584464426
0.000596114084885835
0.0009265848306944198
8.64452320181383e-05
0.0026609615650711542
0.0003074888919637778
0.009992182203532497
0.0004632924153472099
0.0097462997006376
0.0007768746178747974
0.0003188248879808756
0.0003825898655770508
9.533991827080848e-05
1.8035714466071428e-05
3.638124488616127e-05
2.3644907960304173e-05
0.0008948907821596006
0.00022462662562288966
0.00042765453724357837
0.0011422274640078935
0.0058569312755005305
3.492428101557295e-05

4.6620620412297854e-05
0.0010291983573994217
0.0009224666758913335
0.00021028875590227967
0.00885403282663556 7
0.0006610031913234077
0.00291438647603031 2
0.00789313003924876 2
0.0005046189688288859
0.0005490873070781747
0.00115597327805931 5
0.00020343543452639064
0.00250962161040745 7
0.0012823621285559903
0.00011063699470978146
4.9294621084324906e-05
0.00021179081844443883
2.776284136912119e-05
0.0005193338076798499
0.0006225871828287964
0.0005791155191840124
0.0009906610155676159
4.2327929453563786e-05
0.0006225871828287964
0.0001256386211111078
3.589674888888794e-05
0.0007389586188715621
0.0010555270563250348
0.0002576286417096649
0.0006839841887478994
0.00052947704611109 7
4.821254403613241e-05
3.188248879808757e-05
0.0022193367083426126
0.00042057751180455934
0.0002428395952679888
0.0011724414117877653
0.0003112935914143982
0.0006713784667837816
9.503434160968409e-05
2.7607741696668003e-05
0.0005714270476231112
0.0007581148614772525
0.0003010224830682379
0.0030918914110128395
0.0006413004426180208

| |
|---|
| 0.00024864330886558854 |
| 0.00023074486056203449 |
| 6.459850671507938e-05 |
| 0.0003680053228289894 |
| 0.0007127575620726306 |
| 0.00016127039788310025 |
| 0.008821700206280758 |
| 0.00043240625432406255 |
| 0.00010861067612535325 |
| 0.0006177232204629465 |
| 0.008009101065312685 |
| 0.0013632512451596062 |
| 0.0016695222174674229 |
| 0.0009276998545580714 |
| 0.001127494810165378 |
| 0.0001840026614144947 |
| 0.00034081281128990154 |
| 0.000411815480308631 |
| 2.73783144803522e-05 |
| 2.8648033412774332e-05 |
| 0.0008411550236091187 |
| 0.00419186496687696 |
| 0.0010295387007715777 |
| 0.001715897834619296 |
| 0.00133656839160666 |
| 0.000411815480308631 |
| 0.00047823733197131346 |
| 0.0002635619073975239 |
| 0.0007021116385589775 |
| 0.00018218565027478608 |
| 0.010571124329313729 |
| 0.00032562154256961527 |
| 0.0011188948898951485 |
| 0.002697877792258118 |
| 0.0009845703437263821 |
| 0.00013356177739739386 |
| 0.00792389786249021 |
| 0.00043539962675802406 |
| 0.001045377757706525 |
| 8.375908074073851e-05 |
| 0.013000265957924738 |
| 6.79750448927589e-05 |
| 0.00047683897719946744 |
| 0.0001919140102409154 |
| 0.0001411938789629592 |
| 0.00025342491095915753 |

0.00013986186123689356
0.00045268266537742643
0.00333730986639721８
0.0003488319362614286
0.0003495797003262977
0.016941276829372527
0.00038893684251370707
0.0007837193281644432
0.0012484511403040605
0.006441970727685014
0.0008662891378603872
7.959923377777567e-05
9.368314243987816e-05
0.0014368187683014792
0.0010283484826203967
4.472204311043957e-05
0.00036605820471878316
6.723518045855201e-05
6.376497759617513e-05
4.983313375163267e-05
0.00027480687851851125
0.000100852770687828
0.00015523934336241586
2.776284136912119e-05
3.674190158887414e-05
9.151455117969579e-05
0.001049283278594594
8.447497031971919e-05
0.0005004340013877036
0.0003930965948400569
0.0003240515254887589
0.000921783873022428
0.0006868891409008826
0.00022210273095296952
9.983405583239541e-05
8.985065024915585e-05
0.0083630220616522
0.0014020880538879901
0.002896908895964163
0.0013727182676954368
6.839841887478993e-05
0.00046620620412297854
0.0003504812598371328
0.0003329571968452761
0.000130908232151088
3.0599292654511284e-05

0.000532192313014231
0.0013641064843347376
0.0007844104386831067
0.0013072459334847123
2.2361021555219786e-05
0.0009883571527407146
0.00029164637293983
0.00271526690313383
0.0002823877579259184
0.00025342491095915753
9.151455117969579e-05
0.0001550364161161905
0.0004878545360877384
0.0012583857647627135
0.00040223837611540707
0.0005757420307227464
0.00035020529034119805
0.0004463548431732259
0.0005068498219183151
0.001688296915713676
0.00010627496266029189
0.0009120033812843502
3.674190158887414e-05
0.005282598574993473
0.006469471725699545
0.00040506440686094853
0.0008557204785633891
0.0002635619073975239
0.00045416849469485785
0.002823877579259184
0.018769282467912606
0.004345363343946244
0.0002153804933333276
0.0003916566805732434
0.0010017133304804538
0.0002931567825925848
0.0006688131108771752
0.00047942697707571973
4.533748407067498e-05
0.0006853571497107145
0.0003129797650345596
7.844104386831067e-05
4.728981592060835e-05
0.00023347019356079868
0.006248807187353132
0.00023720571665777147

0.00011360427042996718
0.004783258939300942
1.0449441630603325e-05
8.266439468170352e-05
0.004303661784975286
5.132645817327083e-05
0.0008415542355129392
0.00019644950265534247
4.655634124266945e-05
6.422387586809273e-05
0.00021742457976177228
0.0005671440263184582
0.0007024486423072643
0.0006352579026083086
0.00030922606899452056
0.0003975763744215264
0.0003384771836291374
0.0008882025386012824
0.004899060944318149
0.001024739105903371
0.0001873196379486038
0.0005218189914282535
1.8889375255321393e-05
0.00012905868040080464
0.0001626448804451699
0.001926716276042782
1.1484324433083982e-05
0.0007883274033594215
0.000905542718313672
0.0010633292655519127
0.0021332512072485426
0.015891760193496807
0.0002013449484610624
1.6969723298479788e-05
0.00018762031152476526
3.5756333459751844e-05
0.0001480337564335471
0.0014985571035888304
6.843527756436112e-05
0.00011927291232645792
0.007492785517944153
0.0019211396732177666
0.005131439149964488
0.0006822224946299345
0.0003058279803242511
0.0002783034620950685

0.00033782501179170166
0.0008022270615551797
4.925724992833071e-05
0.00024984849464893926
0.00011800853516398665
2.839831245868046e-05
0.0006802973517879452
0.00010585234691413064
0.0024179004786819554
0.0009164878787931688
0.0007347211399309808
0.0014163658338766881
0.004318501998026926
0.0009595025022488639
0.0006687389016362083
0.0007298167712283264
0.0009532441418216914
0.0008445625294792542
0.0007496554524743923
0.00043078914280072025
0.0014343105257497695
0.00025768839082876714
0.0023391885822645845
0.0005944345792321852
0.00018692023573549374
0.007677545508666128
1.650020526255347e-05
0.0009369735797990282
5.403950720292593e-05
0.003774735690106352
0.0006261827897139041
8.882025386012825e-05
0.001086527215028692
0.0012033405923327696
0.0009108113304929515
6.843527756436112e-05
0.00024083953450534774
0.00046764106335646756
0.0008138115678592256
3.4500429185339074e-05
0.00025110086805570087
0.0011564637107328859
0.0003287048764902384
0.0007792496938661918
0.00046018682708633374
0.00026932593105974373

| |
|---|
| 0.0013304092730015195 |
| 0.00013323038079019237 |
| 0.000407671087053323 |
| 0.00020530583269308332 |
| 0.0010741537133493872 |
| 0.0005773316500908336 |
| 6.900085837067815e-05 |
| 0.008089209579183508 |
| 0.00033968516786764253 |
| 0.00019326629312157534 |
| 1.300483467733965e-05 |
| 0.008165607074657504 |
| 0.00010669781294379624 |
| 0.000243769455849695 |
| 0.0004245307048907825 |
| 4.826071597024309e-05 |
| 0.00038266726038071916 |
| 3.7107128279342466e-05 |
| 5.12214960911169e-05 |
| 0.0005833435013346414 |
| 2.3452538940595658e-05 |
| 0.00025689550347237093 |
| 0.012620392291797757 |
| 0.0004201955992245017 |
| 0.001055633821739915 |
| 0.000467082733586129 |
| 1.1595977587294522e-05 |
| 0.00017038987475208274 |
| 0.000666716097940999 |
| 0.0003478793276188356 |
| 0.0001427197241513172 |
| 0.0002783034620950685 |
| 0.001159597758729452 |
| 0.0009553301535378794 |
| 0.0008827246374821556 |
| 0.0011753592816636388 |
| 2.179922679595837e-05 |
| 0.0003664682993236589 |
| 2.8111460817683687e-05 |
| 0.0002368540102936753 |
| 0.0003290287236591943 |
| 0.0002629639011921907 |
| 8.742517133876498e-05 |
| 1.0410353943705803e-05 |
| 0.007706865104171128 |
| 0.0002783034620950685 |

0.000530663381113478
0.0001736984160094047 8
0.00019461780566088705
0.0004506618562334916
0.0006217417770208977
3.994786537249787e-05
0.002029296077776541
0.000117386346050644
8.535329227822733e-05
0.0026023180871227185
0.008117184311106165
0.0006644615530989525
5.641286393818957e-05
0.005256843172906849
0.00011927291232645792
0.0002946742539830137
0.0010703979311348787
0.0022937098524318834
0.00040817841107276717
0.0001617259828155362
9.381015576238263e-05
0.01125404863615978
3.245521423849195e-05
0.00045130291150551657
0.0006853741976968105
7.730651724863014e-05
0.00011542539902560446
0.001330439260588066
0.00016424466615446668
0.001267915486335699
0.00026932593105974373
0.00018762031152476526
0.00019481242346654795
0.00010738397251256662
0.0002530031473591532
0.00030281723854903825
0.0007445060769422214
0.003952083646679186
0.00023719045064920613
0.0007019158114787127
0.00019722292589414303
0.00569986898329323
0.007943244647296748
0.0004039888965896156
1.432093286938603e-05
0.006551727336821404

0.00021136971804688745
0.0014940501649314203
0.000800122453523322
0.00017515602509479835
0.00568602763073545
2.3452538940595658e-05
0.00016009786889457442
5.386518621194875e-05
0.0002087275965713014
0.000671766977470855
0.0010144523497775883
3.4884835081554546e-05
0.00011079379533642538
0.00022264276967605482
0.0034500429185339073
0.0007147092800885027
0.000335036270579938
0.00013849224417053174
0.0010096590717867601
0.001549506175420141
5.679662491736092e-05
0.0016444000947767607
0.00011058415712386828
0.0006513485283076071
0.0005614894410689978
0.0008563183449079031
0.0013915173104753424
5.679662491736092e-05
0.00018378530515712069
0.0005417357468262783
0.001277357353740115
5.963645616322896e-05
0.007171666138603689
0.0005301018325620352
0.0002541031610433234
0.0002912478091692577
0.0007802900806403789
7.13598620756586e-05
0.00013096633510356165
0.0005619589138458114
0.00022363671061210858
0.0010568485902344375
0.00036157536413926225
0.006189852863838593
0.0006361221990744424
0.0005492831488718458

7.784712226435482e-05
7.71279802572938e-05
0.003022951398618848
0.000962578722052118
0.007935098712627986
0.0024471511322152572
0.0006513485283076071
0.0001926716276042782
0.0019235680468335616
0.0019433258991121162
0.00043242370604413036
0.0003072181075075432
0.0018349678819455067
0.00014562390458462885
3.248678545856831e-05
0.0002119061894124887
0.00034114617934234205
0.00031296519921210846
0.00018316132788706517
0.0005925170483314362
1.0842992029677994e-05
0.0003710712827934247
0.0013976473426800795
1.978460631007596e-05
0.00022770283262323786
0.00018089725036179454
0.0001002465300857022
3.362055246786063e-05
0.00019563445623598527
0.004600908473785642
0.0005767365386988415
0.0002043681373179489
0.00045518357857179525
0.0008087790161577559
0.0003175384700061541
0.0001868508684049818
0.0003338012909526499
0.022658138178181516
0.0011444615689805138
0.004866384093877631
0.013194968677657688
0.0004999361029379547
0.0012555743426679422
0.0003906695946629325
0.0002362759368217835
0.0007493983227488062

0.000404843548210794
0.00016233497432347713
0.00026326272513289774
0.0042917308836769264
0.0003965063703554536
0.00548584884304709
1.701801589562102e-05
4.142847308526747e-05
0.0012581351867288193
0.00011738067374159115
0.0001461014768911294
1.9074359483008564e-05
0.0011794451103903985
0.014396806327970782
0.007375419000096644
0.001266685620036685
2.7913696804402778e-05
0.00252368448544421
0.00038415493224520746
0.0011551574714943503
0.00032314209006508625
0.007150520216109656
3.567152942276926e-05
0.0010930865033242732
0.009287470354530227
0.00047993549666924774
0.0010140798712485566
0.0013038533307833207
0.0004161678432656414
0.00016646713730625654
0.008519880569077157
0.00032441430301809843
2.6461539167179507e-05
0.007453672782591039
0.0003431292455444319
4.893713220130477e-05
7.50466602610173e-05
0.006371229353087396
0.00017274891607253038
3.6046033668677594e-05
0.00039367228417020133
0.0004954379086495731
0.000305189751728137
2.5574560200681873e-05
3.783344029687649e-05
6.634559820176891e-06

0.0001493935847620251
0.000329604931866388
1.1233978591219767e-05
0.0004768589870752141З
0.009013843793998638
1.617613525060797e-05
0.0003169278191022961З
0.0005544104486410793
0.0004660533154469087
0.0003742893181571491
0.0019992687351553027
0.0004979718277243457
5.708037750526253e-06
5.143647501036017e-05
0.0004373033322827598
0.0004901868063147029
0.0001330769266256411
0.0002119373275889840
0.0001050653243654242
0.0013987863620872948
0.004735703044057299
0.0003965063703554536
7.0754965624761285e-06
1.0670970340144652e-05
1.8166056650484344e-05
0.0005404401853519093
4.743882151214565e-05
0.0008269657788762422
0.0002615912157669746
0.0001831138510368822
0.0005288486672545334
1.4719762945086993e-05
0.0004141860916310430
2.641065159185801e-05
7.629743793203426e-05
1.4911551387368257e-05
0.0025823748223150057
0.0003896039383763451
0.0002441518013825096
4.199858968735831e-05
0.0003920840560396205
0.0001565075649887882
0.0001795233833694923
0.0006651571512023499
0.0020934868642422133
0.003673580344875723

0.00023652205758930618
6.063372550890139e-05
2.9534492102722936e-05
0.003083688116419718
0.00028811619918390563
0.00015518122969227305
3.157135362704866e-05
0.0006684465801125124
0.004222668547617758
0.0006404240904139663
3.325312064834423e-05
3.1141811400830306e-05
0.008250075136389964
5.945254903794877e-05
9.34254342024909e-05
0.0006367074926018352
0.0025257082901638927
2.9408006055173802e-05
7.41780646561444e-05
6.989078283850466e-05
0.0001578567681352433
7.732848439057526e-05
6.343435948159892e-05
3.6868560611452795e-05
0.008401102506362453
0.0008595955889285956
0.0009253093536438197
0.00044960990209948757
8.438426315063695e-05
4.896092273713428e-05
0.0024467799060962706
0.0007945022462344062
0.00044187705366043
3.403603179124204e-05
0.00016954986207118723
1.8647031673816926e-05
2.190357069819165e-05
0.0010347186788043
0.0006946926255290277
0.00032976011309608026
0.0006539780394174364
0.0003879530742306826
2.8345797374130374e-05
1.2110704433656229e-05
0.00021047569084699103
0.005789105212741898

0.002655547190361712
0.00011416075501052507
0.000143957430060442
0.00026035144697187046
0.00026334686151037706
0.0011216230897769116
2.355667037352687e-05
0.001298523703266352
0.0081495724912019
0.004738986464834511
0.0012117828377440733
0.00010201328748572824
1.7188409046015225e-05
2.0345983448542467e-05
0.00038439930561177564
4.5325210652693616e-05
0.001104997376946703
0.0002956525719866328
0.00031655319993078044
0.0020923136213387124
0.0006866769413883082
0.0066320080663999
0.0005543485724749364
0.0012991185377616641
4.671271710124545e-05
0.007884068586310206
0.000558453157255339
0.0008754447061524249
0.000160908480700248
4.76858987075214e-05
0.0004201188038029734
0.00032454880314372776
0.00011626276256309982
0.0024649941485734143
0.0006291192250536157
0.0013366612217864018
0.0008243631224840482
0.00240731571406246
0.0006493398972939085
0.00041697437009367556
0.0008545313048387836
0.0008001613658754517
0.0008350148786553104
1.0670970340144652e-05
0.0001525948758640685
0.0005977442589579753

1.3503971315404292e-05
5.143647501036017e-05
0.00041901051979067994
0.0006372280450893729
0.00021799267980581212
0.0007262929187760952
0.0011072000295253342
2.8973710607101613e-05
1.1303324138079148e-05
0.0002667235047802386
0.00015430942503108052
0.0003377099711745778
0.0006963067151078854
0.0002539217427215086
0.002112852127348641
4.199858968735831e-05
0.002445122617191964
0.00018590238683947432
0.0003675642995265884
0.00010093824473498696
0.0012109733635705636
0.0008109937458509935
0.0007629743793203424
0.001593171170915605
0.0010228159792190275
0.0005478805383417354
0.001250336868313513
1.4815036491657137e-05
0.0004127566314355951
0.00011444615689805139
0.0007000328309257066
0.0002080839216328207
0.0003483143905592868
0.0004988678634017624
0.00015238323248561626
0.0010652296142049397
0.003972511231172032
0.00020345983448542468
0.00016927727386850043
0.0005194719178351268
0.00042917308836769264
6.832606381973217e-05
4.7935563098660264e-05
0.0004631284170193519
8.678381565729015e-05
1.3956848402201389e-05

| |
|---|
| 0.00014194872173401723 |
| 0.0008003227755108487 |
| 0.0006804906626370623 |
| 8.285694617053494e-05 |
| 7.759061484613652e-05 |
| 0.0001476724605136147 |
| 2.4612076752269116e-05 |
| 0.0014938235216166707 |
| 0.0005722307844902569 |
| 0.0007663354999340885 |
| 3.0316862754450695e-05 |
| 0.001065533184912892 |
| 0.0014417772078971595 |
| 0.00022889231379610278 |
| 0.0002559655982235988 |
| 0.0002985551919079601 |
| 0.0005934245172491552 |
| 0.0014467721721074418 |
| 6.926589963568447e-05 |
| 1.2859118752590042e-05 |
| 0.0029203502105020008 |
| 0.0005519389126998223 |
| 0.0002299771114918189 |
| 0.0008299019564537059 |
| 0.0008393638527625481 |
| 0.00022409037714303767 |
| 2.3905202485232665e-05 |
| 0.006708912645747839 |
| 0.0002922029537822588 |
| 0.00045778462759220556 |
| 0.00015650756498878821 |
| 0.0007027395599003155 |
| 1.1589484242840645e-05 |
| 2.1391805027673154e-05 |
| 0.00040691966897084936 |
| 0.0002712797793138996 |
| 0.0001491922456990035 |
| 0.001018769257936411 |
