## Supplementary material for "Novel antibiotic resistance genes from the hospital effluent are disseminated into the marine environment in Norway": supplemantary table S7

**Supplementary Table S7:**

| Sample |
|---|
| Hospital-eff-F |
| Hospital-eff-F |
| Hospital-eff-F |
| Hospital-eff-F |
| Hospital-eff-F |
| Hospital-eff-F |
| Hospital-eff-F |
| Hospital-eff-F |
| Hospital-eff-F |
| Hospital-eff-F |
| Hospital-eff-F |
| Hospital-eff-F |
| Hospital-eff-F |
| Hospital-eff-F |
| Hospital-eff-F |
| Hospital-eff-F |
| Hospital-eff-F |
| Hospital-eff-F |
| Hospital-eff-F |
| Hospital-eff-F |
| Hospital-eff-F |
| Hospital-eff-F |
| Hospital-eff-F |
| Hospital-eff-F |
| Hospital-eff-F |
| Hospital-eff-F |
| Hospital-eff-F |
| Hospital-eff-F |
| Hospital-eff-F |
| Hospital-eff-F |
| Hospital-eff-F |
| Hospital-eff-F |
| Hospital-eff-F |
| Hospital-eff-F |
| Hospital-eff-F |
| Hospital-eff-F |
| Hospital-eff-F |
| Hospital-eff-F |
| Hospital-eff-F |
| Hospital-eff-F |
| Hospital-eff-F |
| Hospital-eff-F |
| Hospital-eff-F |

Hospital-eff-F
Hospital-eff-F
Hospital-eff-F
Hospital-eff-F
Hospital-eff-F
Hospital-eff-F
Hospital-eff-F
Hospital-eff-F
Hospital-eff-F
Hospital-eff-F
Hospital-eff-F
Hospital-eff-F
Hospital-eff-F
Hospital-eff-F
Hospital-eff-F
Hospital-eff-F
Hospital-eff-F
Hospital-eff-F
Hospital-eff-F
Hospital-eff-F
Hospital-eff-F
Hospital-eff-F
Hospital-eff-F
Hospital-eff-F
Hospital-eff-F
Hospital-eff-F
Hospital-eff-F
Hospital-eff-F
Hospital-eff-F
Hospital-eff-F
Hospital-eff-F
Hospital-eff-F
Hospital-eff-F
Hospital-eff-F
Hospital-eff-F
Hospital-eff-F
Hospital-eff-F
Hospital-eff-F
Hospital-eff-F
Hospital-eff-F
Hospital-eff-F
Hospital-eff-F
Hospital-eff-F
Hospital-eff-F
Hospital-eff-F
Hospital-eff-F

Hospital-eff-F
Hospital-eff-F
Hospital-eff-F
Hospital-eff-F
Hospital-eff-F
Hospital-eff-F
Hospital-eff-F
Hospital-eff-F
Hospital-eff-F
Hospital-eff-F
Hospital-eff-F
Hospital-eff-F
Hospital-eff-F
Hospital-eff-F
Hospital-eff-F
Hospital-eff-F
Hospital-eff-F
Hospital-eff-F
Hospital-eff-F
Hospital-eff-F
Hospital-eff-F
Hospital-eff-F
Hospital-eff-F
Hospital-eff-F
Hospital-eff-F
Hospital-eff-F
Hospital-eff-F
Hospital-eff-F
Hospital-eff-F
Hospital-eff-F
Hospital-eff-F
Hospital-eff-F
Hospital-eff-F
Hospital-eff-F
Hospital-eff-F
Hospital-eff-F
Hospital-eff-F
Hospital-eff-F
Hospital-eff-F
Hospital-eff-F
Hospital-eff-F
Hospital-eff-F
Hospital-eff-F
Hospital-eff-F
Hospital-eff-F
Hospital-eff-F

Hospital-eff-F
Hospital-eff-F
Hospital-eff-F
Hospital-eff-F
Hospital-eff-F
Hospital-eff-F
Hospital-eff-F
Hospital-eff-F
Hospital-eff-F
Hospital-eff-F
Hospital-eff-F
Hospital-eff-F
Hospital-eff-F
Hospital-eff-F
Hospital-eff-F
Hospital-eff-F
Hospital-eff-F
Hospital-eff-F
Hospital-eff-F
Hospital-eff-F
Hospital-eff-F
Hospital-eff-F
Hospital-eff-F
Hospital-eff-F
Hospital-eff-F
Hospital-eff-F
Hospital-eff-F
Hospital-eff-F
Hospital-eff-F
Hospital-eff-F
Hospital-eff-F
Hospital-eff-F
Hospital-eff-F
Hospital-eff-F
Hospital-eff-F
Hospital-eff-F
Hospital-eff-F
Hospital-eff-F
Hospital-eff-F
Hospital-eff-F
Hospital-eff-F
Hospital-eff-F
Hospital-eff-F
Hospital-eff-F
Hospital-eff-F
Hospital-eff-F

Hospital-eff-F
Hospital-eff-F
Hospital-eff-F
Hospital-eff-F
Hospital-eff-F
Hospital-eff-F
Hospital-eff-F
Hospital-eff-F
Hospital-eff-F
Hospital-eff-F
Hospital-eff-F
Hospital-eff-F
Hospital-eff-F
Hospital-eff-F
Hospital-eff-F
Hospital-eff-F
Hospital-eff-F
Hospital-eff-F
Hospital-eff-F
Hospital-eff-F
Hospital-eff-F
Hospital-eff-F
Hospital-eff-F
Hospital-eff-F
Hospital-eff-F
Hospital-eff-F
Hospital-eff-F
Hospital-eff-F
Hospital-eff-F
Hospital-eff-F
Hospital-eff-F
Hospital-eff-F
Hospital-eff-F
Hospital-eff-F
Hospital-eff-F
Hospital-eff-F
Hospital-eff-F
Hospital-eff-F
Hospital-eff-F
Hospital-eff-F
Hospital-eff-F
Hospital-eff-F
Hospital-eff-F
Hospital-eff-F
Hospital-eff-F
Hospital-eff-F

Hospital-eff-F
Hospital-eff-F
Hospital-eff-F
Hospital-eff-F
Hospital-eff-F
Hospital-eff-F
Hospital-eff-F
Hospital-eff-F
Hospital-eff-F
Hospital-eff-F
Hospital-eff-F
Hospital-eff-F
Hospital-eff-F
Hospital-eff-F
Hospital-eff-F
Hospital-eff-F
Hospital-eff-F
Hospital-eff-F
Hospital-eff-F
Hospital-eff-F
Hospital-eff-F
Hospital-eff-F
Hospital-eff-F
Hospital-eff-F
Hospital-eff-F
Hospital-eff-F
Hospital-eff-F
Hospital-eff-F
Hospital-eff-F
Hospital-eff-F
Hospital-eff-F
Hospital-eff-F
Hospital-eff-F
Hospital-eff-F
Hospital-eff-F
Hospital-eff-F
Hospital-eff-F
Hospital-eff-F
Hospital-eff-F
Hospital-eff-F
Hospital-eff-F
Hospital-eff-F
Hospital-eff-F
Hospital-eff-F
Hospital-eff-F
Hospital-eff-F

Hospital-eff-F
Hospital-eff-F
Hospital-eff-F
Hospital-eff-F
Hospital-eff-F
Hospital-eff-F
Hospital-eff-F
Hospital-eff-F
Hospital-eff-F
Hospital-eff-F
Hospital-eff-F
Hospital-eff-F
Hospital-eff-F
Hospital-eff-F
Hospital-eff-F
Hospital-eff-F
Hospital-eff-F
Hospital-eff-F
Hospital-eff-F
Hospital-eff-F
Hospital-eff-F
Hospital-eff-F
Hospital-eff-F
Hospital-eff-F
Hospital-eff-F
Hospital-eff-F
Hospital-eff-F
Hospital-eff-F
Hospital-eff-F
Hospital-eff-F
Hospital-eff-F
Hospital-eff-F
Hospital-eff-F
Hospital-eff-F
Hospital-eff-F
Hospital-eff-F
Hospital-eff-F
Hospital-eff-F
Hospital-eff-F
Hospital-eff-F
Hospital-eff-F
Hospital-eff-F
Hospital-eff-F
Hospital-eff-F
Hospital-eff-F
Hospital-eff-F

Hospital-eff-F
Hospital-eff-F
Hospital-eff-F
Hospital-eff-F
Hospital-eff-F
Hospital-eff-F
Hospital-eff-F
Hospital-eff-F
Hospital-eff-F
Hospital-eff-F
Hospital-eff-F
Hospital-eff-F
Hospital-eff-F
Hospital-eff-F
Hospital-eff-F
Hospital-eff-F
Hospital-eff-F
Hospital-eff-F
Hospital-eff-F
Hospital-eff-F
Hospital-eff-F
Hospital-eff-F
Hospital-eff-F
Hospital-eff-F
Hospital-eff-F
Hospital-eff-F
Hospital-eff-F
Hospital-eff-F
Hospital-eff-F
Hospital-eff-F
Hospital-eff-F
Hospital-eff-F
Hospital-eff-F
Hospital-eff-F
Hospital-eff-F
Hospital-eff-F
Hospital-eff-F
Hospital-eff-F
Hospital-eff-F
Hospital-eff-F
Hospital-eff-F
Hospital-eff-F
Hospital-eff-F
Hospital-eff-F
Hospital-eff-F
Hospital-eff-F

Hospital-eff-F
Hospital-eff-F
Hospital-eff-F
Hospital-eff-F
Hospital-eff-F
Hospital-eff-F
Hospital-eff-F
Hospital-eff-F
Hospital-eff-F
Hospital-eff-F
Hospital-eff-F
Hospital-eff-F
Hospital-eff-F
Hospital-eff-F
Hospital-eff-F
Hospital-eff-F
Hospital-eff-F
Hospital-eff-F
Hospital-eff-F
Hospital-eff-F
Hospital-eff-F
Hospital-eff-F
Hospital-eff-F
Hospital-eff-F
Hospital-eff-F
Hospital-eff-F
Hospital-eff-F
Hospital-eff-F
Hospital-eff-F
Hospital-eff-F
Hospital-eff-F
Hospital-eff-F
Hospital-eff-F
Hospital-eff-F
Hospital-eff-F
Hospital-eff-F
Hospital-eff-F
Hospital-eff-F
Hospital-eff-F
Hospital-eff-F
Hospital-eff-F
Hospital-eff-F
Hospital-eff-F
Hospital-eff-F
Hospital-eff-F
Hospital-eff-F

Hospital-eff-F
Hospital-eff-F
Hospital-eff-F
Hospital-eff-F
Hospital-eff-F
Hospital-eff-F
Hospital-eff-F
Hospital-eff-F
Hospital-eff-F
Hospital-eff-F
Hospital-eff-F
Hospital-eff-F
Hospital-eff-F
Hospital-eff-F
Hospital-eff-F
Hospital-eff-F
Hospital-eff-F
Hospital-eff-F
Hospital-eff-F
Hospital-eff-F
Hospital-eff-F
Hospital-eff-F
Hospital-eff-F
Hospital-eff-F
Hospital-eff-F
Hospital-eff-F
Hospital-eff-F
Hospital-eff-F
Hospital-eff-F
Hospital-eff-F
Hospital-eff-F
Hospital-eff-F
Hospital-eff-F
Hospital-eff-F
Hospital-eff-F
Hospital-eff-F
Hospital-eff-F
Hospital-eff-F
Hospital-eff-F
Hospital-eff-F
Hospital-eff-F
Hospital-eff-F
Hospital-eff-F
Hospital-eff-F
Hospital-eff-F
Hospital-eff-F

Hospital-eff-F
Hospital-eff-F
Hospital-eff-F
Hospital-eff-F
Hospital-eff-F
Hospital-eff-F
Hospital-eff-F
Hospital-eff-F
Hospital-eff-F
Hospital-eff-F
Hospital-eff-F
Hospital-eff-F
Hospital-eff-F
Hospital-eff-F
Hospital-eff-F
Hospital-eff-F
Hospital-eff-F
Hospital-eff-F
Hospital-eff-F
Hospital-eff-F
Hospital-eff-F
Hospital-eff-F
Hospital-eff-F
Hospital-eff-F
Hospital-eff-F
Hospital-eff-F
Hospital-eff-F
Hospital-eff-F
Hospital-eff-F
Hospital-eff-F
Hospital-eff-F
Hospital-eff-F
Hospital-eff-F
Hospital-eff-F
Hospital-eff-F
Hospital-eff-F
Hospital-eff-F
Hospital-eff-F
Hospital-eff-F
Hospital-eff-F
Hospital-eff-F
Hospital-eff-F
Hospital-eff-F
Hospital-eff-F
Hospital-eff-F
Hospital-eff-F

Hospital-eff-F
Hospital-eff-F
Hospital-eff-F
Hospital-eff-F
Hospital-eff-F
Hospital-eff-F
Hospital-eff-F
Hospital-eff-F
Hospital-eff-F
Hospital-eff-F
Hospital-eff-F
Hospital-eff-F
Hospital-eff-F
Hospital-eff-F
Hospital-eff-F
Hospital-eff-F
Hospital-eff-F
Hospital-eff-F
Hospital-eff-F
Hospital-eff-F
Hospital-eff-F
Hospital-eff-F
Hospital-eff-F
Hospital-eff-F
Hospital-eff-F
Hospital-eff-F
Hospital-eff-F
Hospital-eff-F
Hospital-eff-F
Hospital-eff-F
Hospital-eff-F
Hospital-eff-F
Hospital-eff-F
Hospital-eff-F
Hospital-eff-F
Hospital-eff-F
Hospital-eff-F
Hospital-eff-F
Hospital-eff-F
Hospital-eff-F
Hospital-eff-F
Hospital-eff-F
Hospital-eff-F
Hospital-eff-F
Hospital-eff-F
Hospital-eff-F

Hospital-eff-F
Hospital-eff-F
Hospital-eff-F
Hospital-eff-F
Hospital-eff-F
Hospital-eff-F
Hospital-eff-F
Hospital-eff-F
Hospital-eff-F
Hospital-eff-F
Hospital-eff-F
Hospital-eff-F
Hospital-eff-F
Hospital-eff-F
Hospital-eff-F
Hospital-eff-F
Hospital-eff-F
Hospital-eff-F
Hospital-eff-F
Hospital-eff-F
Hospital-eff-F
Hospital-eff-F
Hospital-eff-F
Hospital-eff-F
Hospital-eff-F
Hospital-eff-F
Hospital-eff-F
Hospital-eff-F
Hospital-eff-F
Hospital-eff-F
Hospital-eff-F
Hospital-eff-F
Hospital-eff-F
Hospital-eff-F
Hospital-eff-F
Hospital-eff-F
Hospital-eff-F
Hospital-eff-F
Hospital-eff-F
Hospital-eff-F
Hospital-eff-F
Hospital-eff-F
Hospital-eff-F
Hospital-eff-F
Hospital-eff-F
Hospital-eff-F

Hospital-eff-F
Hospital-eff-F
Hospital-eff-F
Hospital-eff-F
Hospital-eff-F
Hospital-eff-F
Hospital-eff-F
Hospital-eff-F
Hospital-eff-F
Hospital-eff-F
Hospital-eff-F
Hospital-eff-F
Hospital-eff-F
Hospital-eff-F
Hospital-eff-F
Hospital-eff-F
Hospital-eff-F
Hospital-eff-F
Hospital-eff-F
Hospital-eff-F
Hospital-eff-F
Hospital-eff-F
Hospital-eff-F
Hospital-eff-F
Hospital-eff-F
Hospital-eff-F
Hospital-eff-F
Hospital-eff-F
Hospital-eff-F
Hospital-eff-F
Hospital-eff-F
Hospital-eff-F
Hospital-eff-F
Hospital-eff-F
Hospital-eff-F
Hospital-eff-F
Hospital-eff-F
Hospital-eff-F
Hospital-eff-F
Hospital-eff-F
Hospital-eff-F
Hospital-eff-F
Hospital-eff-F
Hospital-eff-F
Hospital-eff-F
Hospital-eff-F

Hospital-eff-F
Hospital-eff-F
Hospital-eff-F
Hospital-eff-F
Hospital-eff-F
Hospital-eff-F
Hospital-eff-F
Hospital-eff-F
Hospital-eff-F
Hospital-eff-F
Hospital-eff-F
Hospital-eff-F
Hospital-eff-F
Hospital-eff-F
Hospital-eff-F
Hospital-eff-F
Hospital-eff-F
Hospital-eff-F
Hospital-eff-F
Hospital-eff-F
Hospital-eff-F
Hospital-eff-F
Hospital-eff-F
Hospital-eff-F
Hospital-eff-F
Hospital-eff-F
Hospital-eff-F
Hospital-eff-F
Hospital-eff-F
Hospital-eff-F
Hospital-eff-F
Hospital-eff-F
Hospital-eff-F
Hospital-eff-F
Hospital-eff-F
Hospital-eff-F
Hospital-eff-F
Hospital-eff-F
Hospital-eff-F
Hospital-eff-F
Hospital-eff-F
Hospital-eff-F
Hospital-eff-F
Hospital-eff-F
Hospital-eff-F
Hospital-eff-F

Hospital-eff-F
Hospital-eff-F
Hospital-eff-F
Hospital-eff-F
Hospital-eff-F
Hospital-eff-F
Hospital-eff-F
Hospital-eff-F
Hospital-eff-F
Hospital-eff-F
Hospital-eff-F
Hospital-eff-F
Hospital-eff-F
Hospital-eff-F
Hospital-eff-F
Hospital-eff-F
Hospital-eff-F
Hospital-eff-F
Hospital-eff-F
Hospital-eff-F
Hospital-eff-F
Hospital-eff-F
Hospital-eff-F
Hospital-eff-F
Hospital-eff-F
Hospital-eff-F
Hospital-eff-F
Hospital-eff-F
Hospital-eff-F
Hospital-eff-F
Hospital-eff-F
Hospital-eff-F
Hospital-eff-F
Hospital-eff-F
Hospital-eff-F
Hospital-eff-F
Hospital-eff-F
Hospital-eff-F
Hospital-eff-F
Hospital-eff-F
Hospital-eff-F
Hospital-eff-F
Hospital-eff-F
Hospital-eff-F
Hospital-eff-F
Hospital-eff-F

Hospital-eff-F
Hospital-eff-F
Hospital-eff-F
Hospital-eff-F
Hospital-eff-F
Hospital-eff-F
Hospital-eff-F
Hospital-eff-F
Hospital-eff-F
Hospital-eff-F
Hospital-eff-F
Hospital-eff-F
Hospital-eff-F
Hospital-eff-F
Hospital-eff-F
Hospital-eff-F
Hospital-eff-F
Hospital-eff-F
Hospital-eff-F
Hospital-eff-F
Hospital-eff-F
Hospital-eff-F
Hospital-eff-F
Hospital-eff-F
Hospital-eff-F
Hospital-eff-F
Hospital-eff-F
Hospital-eff-F
Hospital-eff-F
Hospital-eff-F
Hospital-eff-F
Hospital-eff-F
Hospital-eff-F
Hospital-eff-F
Hospital-eff-F
Hospital-eff-F
Hospital-eff-F
Hospital-eff-F
Hospital-eff-F
Hospital-eff-F
Hospital-eff-F
Hospital-eff-F
Hospital-eff-F
Hospital-eff-F
Hospital-eff-F
Hospital-eff-F

Hospital-eff-F
Hospital-eff-F
Hospital-eff-F
Hospital-eff-F
Hospital-eff-F
Hospital-eff-F
Hospital-eff-F
Hospital-eff-F
Hospital-eff-F
Hospital-eff-F
Hospital-eff-F
Hospital-eff-F
Hospital-eff-F
Hospital-eff-F
Hospital-eff-F
Hospital-eff-F
Hospital-eff-F
Hospital-eff-F
Hospital-eff-F
Hospital-eff-F
Hospital-eff-F
Hospital-eff-F
Hospital-eff-F
Hospital-eff-F
Hospital-eff-F
Hospital-eff-F
Hospital-eff-F
Hospital-eff-F
Hospital-eff-F
Hospital-eff-F
Hospital-eff-F
Hospital-eff-F
Hospital-eff-F
Hospital-eff-F
Hospital-eff-F
Hospital-eff-F
Hospital-eff-F
Hospital-eff-F
Hospital-eff-F
Hospital-eff-F
Hospital-eff-F
Hospital-eff-F
Hospital-eff-F
Hospital-eff-F
Hospital-eff-F
Hospital-eff-F

Hospital-eff-F
Hospital-eff-F
Hospital-eff-F
Hospital-eff-F
Hospital-eff-F
Hospital-eff-F
Hospital-eff-F
Hospital-eff-F
Hospital-eff-F
Hospital-eff-F
Hospital-eff-F
Hospital-eff-F
Hospital-eff-F
Hospital-eff-F
Hospital-eff-F
Hospital-eff-F
Hospital-eff-F
Hospital-eff-F
Hospital-eff-F
Hospital-eff-F
Hospital-eff-F
Hospital-eff-F
Hospital-eff-F
Hospital-eff-F
Hospital-eff-F
Hospital-eff-F
Hospital-eff-F
Hospital-eff-F
Hospital-eff-F
Hospital-eff-F
Hospital-eff-F
Hospital-eff-F
Hospital-eff-F
Hospital-eff-F
Hospital-eff-F
Hospital-eff-F
Hospital-eff-F
Hospital-eff-F
Hospital-eff-F
Hospital-eff-F
Hospital-eff-A
Hospital-eff-A
Hospital-eff-A
Hospital-eff-A
Hospital-eff-A
Hospital-eff-A

Hospital-eff-A
Hospital-eff-A
Hospital-eff-A
Hospital-eff-A
Hospital-eff-A
Hospital-eff-A
Hospital-eff-A
Hospital-eff-A
Hospital-eff-A
Hospital-eff-A
Hospital-eff-A
Hospital-eff-A
Hospital-eff-A
Hospital-eff-A
Hospital-eff-A
Hospital-eff-A
Hospital-eff-A
Hospital-eff-A
Hospital-eff-A
Hospital-eff-A
Hospital-eff-A
Hospital-eff-A
Hospital-eff-A
Hospital-eff-A
Hospital-eff-A
Hospital-eff-A
Hospital-eff-A
Hospital-eff-A
Hospital-eff-A
Hospital-eff-A
Hospital-eff-A
Hospital-eff-A
Hospital-eff-A
Hospital-eff-A
Hospital-eff-A
Hospital-eff-A
Hospital-eff-A
Hospital-eff-A
Hospital-eff-A
Hospital-eff-A
Hospital-eff-A
Hospital-eff-A
Hospital-eff-A
Hospital-eff-A
Hospital-eff-A
Hospital-eff-A

Hospital-eff-A
Hospital-eff-A
Hospital-eff-A
Hospital-eff-A
Hospital-eff-A
Hospital-eff-A
Hospital-eff-A
Hospital-eff-A
Hospital-eff-A
Hospital-eff-A
Hospital-eff-A
Hospital-eff-A
Hospital-eff-A
Hospital-eff-A
Hospital-eff-A
Hospital-eff-A
Hospital-eff-A
Hospital-eff-A
Hospital-eff-A
Hospital-eff-A
Hospital-eff-A
Hospital-eff-A
Hospital-eff-A
Hospital-eff-A
Hospital-eff-A
Hospital-eff-A
Hospital-eff-A
Hospital-eff-A
Hospital-eff-A
Hospital-eff-A
Hospital-eff-A
Hospital-eff-A
Hospital-eff-A
Hospital-eff-A
Hospital-eff-A
Hospital-eff-A
Hospital-eff-A
Hospital-eff-A
Hospital-eff-A
Hospital-eff-A
Hospital-eff-A
Hospital-eff-A
Hospital-eff-A
Hospital-eff-A
Hospital-eff-A
Hospital-eff-A

Hospital-eff-A
Hospital-eff-A
Hospital-eff-A
Hospital-eff-A
Hospital-eff-A
Hospital-eff-A
Hospital-eff-A
Hospital-eff-A
Hospital-eff-A
Hospital-eff-A
Hospital-eff-A
Hospital-eff-A
Hospital-eff-A
Hospital-eff-A
Hospital-eff-A
Hospital-eff-A
Hospital-eff-A
Hospital-eff-A
Hospital-eff-A
Hospital-eff-A
Hospital-eff-A
Hospital-eff-A
Hospital-eff-A
Hospital-eff-A
Hospital-eff-A
Hospital-eff-A
Hospital-eff-A
Hospital-eff-A
Hospital-eff-A
Hospital-eff-A
Hospital-eff-A
Hospital-eff-A
Hospital-eff-A
Hospital-eff-A
Hospital-eff-A
Hospital-eff-A
Hospital-eff-A
Hospital-eff-A
Hospital-eff-A
Hospital-eff-A
Hospital-eff-A
Hospital-eff-A
Hospital-eff-A
Hospital-eff-A
Hospital-eff-A
Hospital-eff-A

Hospital-eff-A
Hospital-eff-A
Hospital-eff-A
Hospital-eff-A
Hospital-eff-A
Hospital-eff-A
Hospital-eff-A
Hospital-eff-A
Hospital-eff-A
Hospital-eff-A
Hospital-eff-A
Hospital-eff-A
Hospital-eff-A
Hospital-eff-A
Hospital-eff-A
Hospital-eff-A
Hospital-eff-A
Hospital-eff-A
Hospital-eff-A
Hospital-eff-A
Hospital-eff-A
Hospital-eff-A
Hospital-eff-A
Hospital-eff-A
Hospital-eff-A
Hospital-eff-A
Hospital-eff-A
Hospital-eff-A
Hospital-eff-A
Hospital-eff-A
Hospital-eff-A
Hospital-eff-A
Hospital-eff-A
Hospital-eff-A
Hospital-eff-A
Hospital-eff-A
Hospital-eff-A
Hospital-eff-A
Hospital-eff-A
Hospital-eff-A
Hospital-eff-A
Hospital-eff-A
Hospital-eff-A
Hospital-eff-A
Hospital-eff-A
Hospital-eff-A

Hospital-eff-A
Hospital-eff-A
Hospital-eff-A
Hospital-eff-A
Hospital-eff-A
Hospital-eff-A
Hospital-eff-A
Hospital-eff-A
Hospital-eff-A
Hospital-eff-A
Hospital-eff-A
Hospital-eff-A
Hospital-eff-A
Hospital-eff-A
Hospital-eff-A
Hospital-eff-A
Hospital-eff-A
Hospital-eff-A
Hospital-eff-A
Hospital-eff-A
Hospital-eff-A
Hospital-eff-A
Hospital-eff-A
Hospital-eff-A
Hospital-eff-A
Hospital-eff-A
Hospital-eff-A
Hospital-eff-A
Hospital-eff-A
Hospital-eff-A
Hospital-eff-A
Hospital-eff-A
Hospital-eff-A
Hospital-eff-A
Hospital-eff-A
Hospital-eff-A
Hospital-eff-A
Hospital-eff-A
Hospital-eff-A
Hospital-eff-A
Hospital-eff-A
Hospital-eff-A
Hospital-eff-A
Hospital-eff-A
Hospital-eff-A
Hospital-eff-A

Hospital-eff-A
Hospital-eff-A
Hospital-eff-A
Hospital-eff-A
Hospital-eff-A
Hospital-eff-A
Hospital-eff-A
Hospital-eff-A
Hospital-eff-A
Hospital-eff-A
Hospital-eff-A
Hospital-eff-A
Hospital-eff-A
Hospital-eff-A
Hospital-eff-A
Hospital-eff-A
Hospital-eff-A
Hospital-eff-A
Hospital-eff-A
Hospital-eff-A
Hospital-eff-A
Hospital-eff-A
Hospital-eff-A
Hospital-eff-A
Hospital-eff-A
Hospital-eff-A
Hospital-eff-A
Hospital-eff-A
Hospital-eff-A
Hospital-eff-A
Hospital-eff-A
Hospital-eff-A
Hospital-eff-A
Hospital-eff-A
Hospital-eff-A
Hospital-eff-A
Hospital-eff-A
Hospital-eff-A
Hospital-eff-A
Hospital-eff-A
Hospital-eff-A
Hospital-eff-A
Hospital-eff-A
Hospital-eff-A
Hospital-eff-A
Hospital-eff-A

Hospital-eff-A
Hospital-eff-A
Hospital-eff-A
Hospital-eff-A
Hospital-eff-A
Hospital-eff-A
Hospital-eff-A
Hospital-eff-A
Hospital-eff-A
Hospital-eff-A
Hospital-eff-A
Hospital-eff-A
Hospital-eff-A
Hospital-eff-A
Hospital-eff-A
Hospital-eff-A
Hospital-eff-A
Hospital-eff-A
Hospital-eff-A
Hospital-eff-A
Hospital-eff-A
Hospital-eff-A
Hospital-eff-A
Hospital-eff-A
Hospital-eff-A
Hospital-eff-A
Hospital-eff-A
Hospital-eff-A
Hospital-eff-A
Hospital-eff-A
Hospital-eff-A
Hospital-eff-A
Hospital-eff-A
Hospital-eff-A
Hospital-eff-A
Hospital-eff-A
Hospital-eff-A
Hospital-eff-A
Hospital-eff-A
Hospital-eff-A
Hospital-eff-A
Hospital-eff-A
Hospital-eff-A
Hospital-eff-A
Hospital-eff-A
Hospital-eff-A

Hospital-eff-A
Hospital-eff-A
Hospital-eff-A
Hospital-eff-A
Hospital-eff-A
Hospital-eff-A
Hospital-eff-A
Hospital-eff-A
Hospital-eff-A
Hospital-eff-A
Hospital-eff-A
Hospital-eff-A
Hospital-eff-A
Hospital-eff-A
Hospital-eff-A
Hospital-eff-A
Hospital-eff-A
Hospital-eff-A
Hospital-eff-A
Hospital-eff-A
Hospital-eff-A
Hospital-eff-A
Hospital-eff-A
Hospital-eff-A
Hospital-eff-A
Hospital-eff-A
Hospital-eff-A
Hospital-eff-A
Hospital-eff-A
Hospital-eff-A
Hospital-eff-A
Hospital-eff-A
Hospital-eff-A
Hospital-eff-A
Hospital-eff-A
Hospital-eff-A
Hospital-eff-A
Hospital-eff-A
Hospital-eff-A
Hospital-eff-A
Hospital-eff-A
Hospital-eff-A
Hospital-eff-A
Hospital-eff-A
Hospital-eff-A
Hospital-eff-A

Hospital-eff-A
Hospital-eff-A
Hospital-eff-A
Hospital-eff-A
Hospital-eff-A
Hospital-eff-A
Hospital-eff-A
Hospital-eff-A
Hospital-eff-A
Hospital-eff-A
Hospital-eff-A
Hospital-eff-A
Hospital-eff-A
Hospital-eff-A
Hospital-eff-A
Hospital-eff-A
Hospital-eff-A
Hospital-eff-A
Hospital-eff-A
Hospital-eff-A
Hospital-eff-A
Hospital-eff-A
Hospital-eff-A
Hospital-eff-A
Hospital-eff-A
Hospital-eff-A
Hospital-eff-A
Hospital-eff-A
Hospital-eff-A
Hospital-eff-A
Hospital-eff-A
Hospital-eff-A
Hospital-eff-A
Hospital-eff-A
Hospital-eff-A
Hospital-eff-A
Hospital-eff-A
Hospital-eff-A
Hospital-eff-A
Hospital-eff-A
Hospital-eff-A
Hospital-eff-A
Hospital-eff-A
Hospital-eff-A
Hospital-eff-A
Hospital-eff-A

Hospital-eff-A
Hospital-eff-A
Hospital-eff-A
Hospital-eff-A
Hospital-eff-A
Hospital-eff-A
Hospital-eff-A
Hospital-eff-A
Hospital-eff-A
Hospital-eff-A
Hospital-eff-A
Hospital-eff-A
Hospital-eff-A
Hospital-eff-A
Hospital-eff-A
Hospital-eff-A
Hospital-eff-A
Hospital-eff-A
Hospital-eff-A
Hospital-eff-A
Hospital-eff-A
Hospital-eff-A
Hospital-eff-A
Hospital-eff-A
Hospital-eff-A
Hospital-eff-A
Hospital-eff-A
Hospital-eff-A
Hospital-eff-A
Hospital-eff-A
Hospital-eff-A
Hospital-eff-A
Hospital-eff-A
Hospital-eff-A
Hospital-eff-A
Hospital-eff-A
Hospital-eff-A
Hospital-eff-A
Hospital-eff-A
Hospital-eff-A
Hospital-eff-A
Hospital-eff-A
Hospital-eff-A
Hospital-eff-A
Hospital-eff-A
Hospital-eff-A

Hospital-eff-A
Hospital-eff-A
Hospital-eff-A
Hospital-eff-A
Hospital-eff-A
Hospital-eff-A
Hospital-eff-A
Hospital-eff-A
Hospital-eff-A
Hospital-eff-A
Hospital-eff-A
Hospital-eff-A
Hospital-eff-A
Hospital-eff-A
Hospital-eff-A
Hospital-eff-A
Hospital-eff-A
Hospital-eff-A
Hospital-eff-A
Hospital-eff-A
Hospital-eff-A
Hospital-eff-A
Hospital-eff-A
Hospital-eff-A
Hospital-eff-A
Hospital-eff-A
Hospital-eff-A
Hospital-eff-A
Hospital-eff-A
Hospital-eff-A
Hospital-eff-A
Hospital-eff-A
Hospital-eff-A
Hospital-eff-A
Hospital-eff-A
Hospital-eff-A
Hospital-eff-A
Hospital-eff-A
Hospital-eff-A
Hospital-eff-A
Hospital-eff-A
Hospital-eff-A
Hospital-eff-A
Hospital-eff-A
Hospital-eff-A
Hospital-eff-A

Hospital-eff-A
Hospital-eff-A
Hospital-eff-A
Hospital-eff-A
Hospital-eff-A
Hospital-eff-A
Hospital-eff-A
Hospital-eff-A
Hospital-eff-A
Hospital-eff-A
Hospital-eff-A
Hospital-eff-A
Hospital-eff-A
Hospital-eff-A
Hospital-eff-A
Hospital-eff-A
Hospital-eff-A
Hospital-eff-A
Hospital-eff-A
Hospital-eff-A
Hospital-eff-A
Hospital-eff-A
Hospital-eff-A
Hospital-eff-A
Hospital-eff-A
Hospital-eff-A
Hospital-eff-A
Hospital-eff-A
Hospital-eff-A
Hospital-eff-A
Hospital-eff-A
Hospital-eff-A
Hospital-eff-A
Hospital-eff-A
Hospital-eff-A
Hospital-eff-A
Hospital-eff-A
Hospital-eff-A
Hospital-eff-A
Hospital-eff-A
Hospital-eff-A
Hospital-eff-A
Hospital-eff-A
Hospital-eff-A
Hospital-eff-A
Hospital-eff-A

Hospital-eff-A
Hospital-eff-A
Hospital-eff-A
Hospital-eff-A
Hospital-eff-A
Hospital-eff-A
Hospital-eff-A
Hospital-eff-A
Hospital-eff-A
Hospital-eff-A
Hospital-eff-A
Hospital-eff-A
Hospital-eff-A
Hospital-eff-A
Hospital-eff-A
Hospital-eff-A
Hospital-eff-A
Hospital-eff-A
Hospital-eff-A
Hospital-eff-A
Hospital-eff-A
Hospital-eff-A
Hospital-eff-A
Hospital-eff-A
Hospital-eff-A
Hospital-eff-A
Hospital-eff-A
Hospital-eff-A
Hospital-eff-A
Hospital-eff-A
Hospital-eff-A
Hospital-eff-A
Hospital-eff-A
Hospital-eff-A
Hospital-eff-A
Hospital-eff-A
Hospital-eff-A
Hospital-eff-A
Hospital-eff-A
Hospital-eff-A
Hospital-eff-A
Hospital-eff-A
Hospital-eff-A
Hospital-eff-A
Hospital-eff-A
Hospital-eff-A

Hospital-eff-A
Hospital-eff-A
Hospital-eff-A
Hospital-eff-A
Hospital-eff-A
Hospital-eff-A
Hospital-eff-A
Hospital-eff-A
Hospital-eff-A
Hospital-eff-A
Hospital-eff-A
Hospital-eff-A
Hospital-eff-A
Hospital-eff-A
Hospital-eff-A
Hospital-eff-A
Hospital-eff-A
Hospital-eff-A
Hospital-eff-A
Hospital-eff-A
Hospital-eff-A
Hospital-eff-A
Hospital-eff-A
Hospital-eff-A
Hospital-eff-A
Hospital-eff-A
Hospital-eff-A
Hospital-eff-A
Hospital-eff-A
Hospital-eff-A
Hospital-eff-A
Hospital-eff-A
Hospital-eff-A
Hospital-eff-A
Hospital-eff-A
Hospital-eff-A
Hospital-eff-A
Hospital-eff-A
Hospital-eff-A
Hospital-eff-A
Hospital-eff-A
Hospital-eff-A
Hospital-eff-A
Hospital-eff-A
Hospital-eff-A
Hospital-eff-A

Hospital-eff-A
Hospital-eff-A
Hospital-eff-A
Hospital-eff-A
Hospital-eff-A
Hospital-eff-A
Hospital-eff-A
Hospital-eff-A
Hospital-eff-A
Hospital-eff-A
Hospital-eff-A
Hospital-eff-A
Hospital-eff-A
Hospital-eff-A
Hospital-eff-A
Hospital-eff-A
Hospital-eff-A
Hospital-eff-A
Hospital-eff-A
Hospital-eff-A
Hospital-eff-A
Hospital-eff-A
Hospital-eff-A
Hospital-eff-A
Hospital-eff-A
Hospital-eff-A
Hospital-eff-A
Hospital-eff-A
Hospital-eff-A
Hospital-eff-A
Hospital-eff-A
Hospital-eff-A
Hospital-eff-A
Hospital-eff-A
Hospital-eff-A
Hospital-eff-A
Hospital-eff-A
Hospital-eff-A
Hospital-eff-A
Hospital-eff-A
Hospital-eff-A
Hospital-eff-A
Hospital-eff-A
Hospital-eff-A
Hospital-eff-A
Hospital-eff-A

Hospital-eff-A
Hospital-eff-A
Hospital-eff-A
Hospital-eff-A
Hospital-eff-A
Hospital-eff-A
Hospital-eff-A
Hospital-eff-A
Hospital-eff-A
Hospital-eff-A
Hospital-eff-A
Hospital-eff-A
Hospital-eff-A
Hospital-eff-A
Hospital-eff-A
Hospital-eff-A
Hospital-eff-A
Hospital-eff-A
Hospital-eff-A
Hospital-eff-A
Hospital-eff-A
Hospital-eff-A
Hospital-eff-A
Hospital-eff-A
Hospital-eff-A
Hospital-eff-A
Hospital-eff-A
Hospital-eff-A
Hospital-eff-A
Hospital-eff-A
Hospital-eff-A
Hospital-eff-A
Hospital-eff-A
Hospital-eff-A
Hospital-eff-A
Hospital-eff-A
Hospital-eff-A
Hospital-eff-A
Hospital-eff-A
Hospital-eff-A
Hospital-eff-A
Hospital-eff-A
Hospital-eff-A
Hospital-eff-A
Hospital-eff-A
Hospital-eff-A

Hospital-eff-A
Hospital-eff-A
Hospital-eff-A
Hospital-eff-A
Hospital-eff-A
Hospital-eff-A
Hospital-eff-A
Hospital-eff-A
Hospital-eff-A
Hospital-eff-A
Hospital-eff-A
Hospital-eff-A
Hospital-eff-A
Hospital-eff-A
Hospital-eff-A
Hospital-eff-A
Hospital-eff-A
Hospital-eff-A
Hospital-eff-A
Hospital-eff-A
Hospital-eff-A
Hospital-eff-A
Hospital-eff-A
Hospital-eff-A
Hospital-eff-A
Hospital-eff-A
Hospital-eff-A
Hospital-eff-A
Hospital-eff-A
Hospital-eff-A
Hospital-eff-A
Hospital-eff-A
Hospital-eff-A
Hospital-eff-A
Hospital-eff-A
Hospital-eff-A
Hospital-eff-A
Hospital-eff-A
Hospital-eff-A
Hospital-eff-A
Hospital-eff-A
Hospital-eff-A
Hospital-eff-A
Hospital-eff-A
Hospital-eff-A
Hospital-eff-A

Hospital-eff-A
Hospital-eff-A
Hospital-eff-A
Hospital-eff-A
Hospital-eff-A
Hospital-eff-A
Hospital-eff-A
Hospital-eff-A
Hospital-eff-A
Hospital-eff-A
Hospital-eff-A
Hospital-eff-A
Hospital-eff-A
Hospital-eff-A
Hospital-eff-A
Hospital-eff-A
Hospital-eff-A
Hospital-eff-A
Hospital-eff-A
Hospital-eff-A
Hospital-eff-A
Hospital-eff-A
Hospital-eff-A
Hospital-eff-A
Hospital-eff-A
Hospital-eff-A
Hospital-eff-A
Hospital-eff-A
Hospital-eff-A
Hospital-eff-A
Hospital-eff-A
Hospital-eff-A
Hospital-eff-A
Hospital-eff-A
Hospital-eff-A
Hospital-eff-A
STP-inf-F
STP-inf-F
STP-inf-F
STP-inf-F
STP-inf-F
STP-inf-F
STP-inf-F
STP-inf-F
STP-inf-F
STP-inf-F

STP-inf-F
STP-inf-F
STP-inf-F
STP-inf-F
STP-inf-F
STP-inf-F
STP-inf-F
STP-inf-F
STP-inf-F
STP-inf-F
STP-inf-F
STP-inf-F
STP-inf-F
STP-inf-F
STP-inf-F
STP-inf-F
STP-inf-F
STP-inf-F
STP-inf-F
STP-inf-F
STP-inf-F
STP-inf-F
STP-inf-F
STP-inf-F
STP-inf-F
STP-inf-F
STP-inf-F
STP-inf-F
STP-inf-F
STP-inf-F
STP-inf-F
STP-inf-F
STP-inf-F
STP-inf-F
STP-inf-F
STP-inf-F
STP-inf-F
STP-inf-F
STP-inf-F
STP-inf-F
STP-inf-F
STP-inf-F
STP-inf-F
STP-inf-F
STP-inf-F
STP-inf-F

STP-inf-F
STP-inf-F
STP-inf-F
STP-inf-F
STP-inf-F
STP-inf-F
STP-inf-F
STP-inf-F
STP-inf-F
STP-inf-F
STP-inf-F
STP-inf-F
STP-inf-F
STP-inf-F
STP-inf-F
STP-inf-F
STP-inf-F
STP-inf-F
STP-inf-F
STP-inf-F
STP-inf-F
STP-inf-F
STP-inf-F
STP-inf-F
STP-inf-F
STP-inf-F
STP-inf-F
STP-inf-F
STP-inf-F
STP-inf-F
STP-inf-F
STP-inf-F
STP-inf-F
STP-inf-F
STP-inf-F
STP-inf-F
STP-inf-F
STP-inf-F
STP-inf-F
STP-inf-F
STP-inf-F
STP-inf-F
STP-inf-F
STP-inf-F
STP-inf-F
STP-inf-F

STP-inf-F
STP-inf-F
STP-inf-F
STP-inf-F
STP-inf-F
STP-inf-F
STP-inf-F
STP-inf-F
STP-inf-F
STP-inf-F
STP-inf-F
STP-inf-F
STP-inf-F
STP-inf-F
STP-inf-F
STP-inf-F
STP-inf-F
STP-inf-F
STP-inf-F
STP-inf-F
STP-inf-F
STP-inf-F
STP-inf-F
STP-inf-F
STP-inf-F
STP-inf-F
STP-inf-F
STP-inf-F
STP-inf-F
STP-inf-F
STP-inf-F
STP-inf-F
STP-inf-F
STP-inf-F
STP-inf-F
STP-inf-F
STP-inf-F
STP-inf-F
STP-inf-F
STP-inf-F
STP-inf-F
STP-inf-F
STP-inf-F
STP-inf-F
STP-inf-F
STP-inf-F

STP-inf-F
STP-inf-F
STP-inf-F
STP-inf-F
STP-inf-F
STP-inf-F
STP-inf-F
STP-inf-F
STP-inf-F
STP-inf-F
STP-inf-F
STP-inf-F
STP-inf-F
STP-inf-F
STP-inf-F
STP-inf-F
STP-inf-F
STP-inf-F
STP-inf-F
STP-inf-F
STP-inf-F
STP-inf-F
STP-inf-F
STP-inf-F
STP-inf-F
STP-inf-F
STP-inf-F
STP-inf-F
STP-inf-F
STP-inf-F
STP-inf-F
STP-inf-F
STP-inf-F
STP-inf-F
STP-inf-F
STP-inf-F
STP-inf-F
STP-inf-F
STP-inf-F
STP-inf-F
STP-inf-F
STP-inf-F
STP-inf-F
STP-inf-F
STP-inf-F
STP-inf-F

STP-inf-F
STP-inf-F
STP-inf-F
STP-inf-F
STP-inf-F
STP-inf-F
STP-inf-F
STP-inf-F
STP-inf-F
STP-inf-F
STP-inf-F
STP-inf-F
STP-inf-F
STP-inf-F
STP-inf-F
STP-inf-F
STP-inf-F
STP-inf-F
STP-inf-F
STP-inf-F
STP-inf-F
STP-inf-F
STP-inf-F
STP-inf-F
STP-inf-F
STP-inf-F
STP-inf-F
STP-inf-F
STP-inf-F
STP-inf-F
STP-inf-F
STP-inf-F
STP-inf-F
STP-inf-F
STP-inf-F
STP-inf-F
STP-inf-F
STP-inf-F
STP-inf-F
STP-inf-F
STP-inf-F
STP-inf-F
STP-inf-F
STP-inf-F
STP-inf-F
STP-inf-F

STP-inf-F
STP-inf-F
STP-inf-F
STP-inf-F
STP-inf-F
STP-inf-F
STP-inf-F
STP-inf-F
STP-inf-F
STP-inf-F
STP-inf-F
STP-inf-F
STP-inf-F
STP-inf-F
STP-inf-F
STP-inf-F
STP-inf-F
STP-inf-F
STP-inf-F
STP-inf-F
STP-inf-F
STP-inf-F
STP-inf-F
STP-inf-F
STP-inf-F
STP-inf-F
STP-inf-F
STP-inf-F
STP-inf-F
STP-inf-F
STP-inf-F
STP-inf-F
STP-inf-F
STP-inf-F
STP-inf-F
STP-inf-F
STP-inf-F
STP-inf-F
STP-inf-F
STP-inf-F
STP-inf-F
STP-inf-F
STP-inf-F
STP-inf-F
STP-inf-F
STP-inf-F

STP-inf-F
STP-inf-F
STP-inf-F
STP-inf-F
STP-inf-F
STP-inf-F
STP-inf-F
STP-inf-F
STP-inf-F
STP-inf-F
STP-inf-F
STP-inf-F
STP-inf-F
STP-inf-F
STP-inf-F
STP-inf-F
STP-inf-F
STP-inf-F
STP-inf-F
STP-inf-F
STP-inf-F
STP-inf-F
STP-inf-F
STP-inf-F
STP-inf-F
STP-inf-F
STP-inf-F
STP-inf-F
STP-inf-F
STP-inf-F
STP-inf-F
STP-inf-F
STP-inf-F
STP-inf-F
STP-inf-F
STP-inf-F
STP-inf-F
STP-inf-F
STP-inf-F
STP-inf-F
STP-inf-F
STP-inf-F
STP-inf-F
STP-inf-F
STP-inf-F
STP-inf-F

STP-inf-F
STP-inf-F
STP-inf-F
STP-inf-F
STP-inf-F
STP-inf-F
STP-inf-F
STP-inf-F
STP-inf-F
STP-inf-F
STP-inf-F
STP-inf-F
STP-inf-F
STP-inf-F
STP-inf-F
STP-inf-F
STP-inf-F
STP-inf-F
STP-inf-F
STP-inf-F
STP-inf-F
STP-inf-F
STP-inf-F
STP-inf-F
STP-inf-F
STP-inf-F
STP-inf-F
STP-inf-F
STP-inf-F
STP-inf-F
STP-inf-F
STP-inf-F
STP-inf-F
STP-inf-F
STP-inf-F
STP-inf-F
STP-inf-F
STP-inf-F
STP-inf-F
STP-inf-F
STP-inf-F
STP-inf-F
STP-inf-F
STP-inf-F
STP-inf-F
STP-inf-F

STP-inf-F
STP-inf-F
STP-inf-F
STP-inf-F
STP-inf-F
STP-inf-F
STP-inf-F
STP-inf-F
STP-inf-F
STP-inf-F
STP-inf-F
STP-inf-F
STP-inf-F
STP-inf-F
STP-inf-F
STP-inf-F
STP-inf-F
STP-inf-F
STP-inf-F
STP-inf-F
STP-inf-F
STP-inf-F
STP-inf-F
STP-inf-F
STP-inf-F
STP-inf-F
STP-inf-F
STP-inf-F
STP-inf-F
STP-inf-F
STP-inf-F
STP-inf-F
STP-inf-F
STP-inf-F
STP-inf-F
STP-inf-F
STP-inf-F
STP-inf-F
STP-inf-F
STP-inf-F
STP-inf-F
STP-inf-F
STP-inf-F
STP-inf-F
STP-inf-F
STP-inf-F

STP-inf-F
STP-inf-F
STP-inf-F
STP-inf-F
STP-inf-F
STP-inf-F
STP-inf-F
STP-inf-F
STP-inf-F
STP-inf-F
STP-inf-F
STP-inf-F
STP-inf-F
STP-inf-F
STP-inf-F
STP-inf-F
STP-inf-F
STP-inf-F
STP-inf-F
STP-inf-F
STP-inf-F
STP-inf-F
STP-inf-F
STP-inf-F
STP-inf-F
STP-inf-F
STP-inf-F
STP-inf-F
STP-inf-F
STP-inf-F
STP-inf-F
STP-inf-F
STP-inf-F
STP-inf-F
STP-inf-F
STP-inf-F
STP-inf-F
STP-inf-F
STP-inf-F
STP-inf-F
STP-inf-F
STP-inf-F
STP-inf-F
STP-inf-F
STP-inf-F
STP-inf-F

STP-inf-F
STP-inf-F
STP-inf-F
STP-inf-F
STP-inf-F
STP-inf-F
STP-inf-F
STP-inf-F
STP-inf-F
STP-inf-F
STP-inf-F
STP-inf-F
STP-inf-F
STP-inf-F
STP-inf-F
STP-inf-F
STP-inf-F
STP-inf-F
STP-inf-F
STP-inf-F
STP-inf-F
STP-inf-F
STP-inf-F
STP-inf-F
STP-inf-F
STP-inf-F
STP-inf-F
STP-inf-F
STP-inf-F
STP-inf-F
STP-inf-F
STP-inf-F
STP-inf-F
STP-inf-F
STP-inf-F
STP-inf-F
STP-inf-F
STP-inf-F
STP-inf-F
STP-inf-F
STP-inf-F
STP-inf-F
STP-inf-F
STP-inf-F
STP-inf-F
STP-inf-F

STP-inf-F
STP-inf-F
STP-inf-F
STP-inf-F
STP-inf-F
STP-inf-F
STP-inf-F
STP-inf-F
STP-inf-F
STP-inf-F
STP-inf-F
STP-inf-F
STP-inf-F
STP-inf-F
STP-inf-F
STP-inf-F
STP-inf-F
STP-inf-F
STP-inf-F
STP-inf-F
STP-inf-F
STP-inf-F
STP-inf-F
STP-inf-F
STP-inf-F
STP-inf-F
STP-inf-F
STP-inf-F
STP-inf-F
STP-inf-F
STP-inf-F
STP-inf-F
STP-inf-F
STP-inf-F
STP-inf-F
STP-inf-F
STP-inf-F
STP-inf-F
STP-inf-F
STP-inf-F
STP-inf-F
STP-inf-F
STP-inf-F
STP-inf-F
STP-inf-F
STP-inf-F

STP-inf-F
STP-inf-F
STP-inf-F
STP-inf-F
STP-inf-F
STP-inf-F
STP-inf-F
STP-inf-F
STP-inf-F
STP-inf-F
STP-inf-F
STP-inf-F
STP-inf-F
STP-inf-F
STP-inf-F
STP-inf-F
STP-inf-F
STP-inf-F
STP-inf-F
STP-inf-F
STP-inf-F
STP-inf-F
STP-inf-F
STP-inf-F
STP-inf-F
STP-inf-F
STP-inf-F
STP-inf-F
STP-inf-F
STP-inf-F
STP-inf-F
STP-inf-F
STP-inf-F
STP-inf-F
STP-inf-F
STP-inf-F
STP-inf-F
STP-inf-F
STP-inf-F
STP-inf-F
STP-inf-F
STP-inf-F
STP-inf-F
STP-inf-F
STP-inf-F
STP-inf-F

STP-inf-F
STP-inf-F
STP-inf-F
STP-inf-F
STP-inf-F
STP-inf-F
STP-inf-F
STP-inf-F
STP-inf-F
STP-inf-F
STP-inf-F
STP-inf-F
STP-inf-F
STP-inf-F
STP-inf-F
STP-inf-F
STP-inf-F
STP-inf-F
STP-inf-F
STP-inf-F
STP-inf-F
STP-inf-F
STP-inf-F
STP-inf-F
STP-inf-F
STP-inf-F
STP-inf-F
STP-inf-F
STP-inf-F
STP-inf-F
STP-inf-F
STP-inf-F
STP-inf-F
STP-inf-F
STP-inf-F
STP-inf-F
STP-inf-F
STP-inf-F
STP-inf-F
STP-inf-F
STP-inf-F
STP-inf-F
STP-inf-F
STP-inf-F
STP-inf-F
STP-inf-F

STP-inf-F
STP-inf-F
STP-inf-F
STP-inf-F
STP-inf-F
STP-inf-F
STP-inf-F
STP-inf-F
STP-inf-F
STP-inf-F
STP-inf-F
STP-inf-F
STP-inf-F
STP-inf-F
STP-inf-F
STP-inf-F
STP-inf-F
STP-inf-F
STP-inf-F
STP-inf-F
STP-inf-F
STP-inf-F
STP-inf-F
STP-inf-F
STP-inf-F
STP-inf-F
STP-inf-F
STP-inf-F
STP-inf-F
STP-inf-F
STP-inf-F
STP-inf-F
STP-inf-F
STP-inf-F
STP-inf-F
STP-inf-F
STP-inf-F
STP-inf-F
STP-inf-F
STP-inf-F
STP-inf-F
STP-inf-F
STP-inf-F
STP-inf-F
STP-inf-F
STP-inf-F

STP-inf-F
STP-inf-F
STP-inf-F
STP-inf-F
STP-inf-F
STP-inf-F
STP-inf-F
STP-inf-F
STP-inf-F
STP-inf-F
STP-inf-F
STP-inf-F
STP-inf-F
STP-inf-F
STP-inf-F
STP-inf-F
STP-inf-F
STP-inf-F
STP-inf-F
STP-inf-F
STP-inf-F
STP-inf-F
STP-inf-F
STP-inf-F
STP-inf-F
STP-inf-F
STP-inf-F
STP-inf-F
STP-inf-F
STP-inf-F
STP-inf-F
STP-inf-F
STP-inf-F
STP-inf-F
STP-inf-F
STP-inf-F
STP-inf-F
STP-inf-F
STP-inf-F
STP-inf-F
STP-inf-F
STP-inf-F
STP-inf-F
STP-inf-F
STP-inf-F
STP-inf-F

STP-inf-F
STP-inf-F
STP-inf-F
STP-inf-F
STP-inf-F
STP-inf-F
STP-inf-F
STP-inf-F
STP-inf-F
STP-inf-F
STP-inf-F
STP-inf-F
STP-inf-F
STP-inf-F
STP-inf-F
STP-inf-F
STP-inf-F
STP-inf-F
STP-inf-F
STP-inf-F
STP-inf-F
STP-inf-A
STP-inf-A
STP-inf-A
STP-inf-A
STP-inf-A
STP-inf-A
STP-inf-A
STP-inf-A
STP-inf-A
STP-inf-A
STP-inf-A
STP-inf-A
STP-inf-A
STP-inf-A
STP-inf-A
STP-inf-A
STP-inf-A
STP-inf-A
STP-inf-A
STP-inf-A
STP-inf-A
STP-inf-A
STP-inf-A
STP-inf-A
STP-inf-A

STP-inf-A
STP-inf-A
STP-inf-A
STP-inf-A
STP-inf-A
STP-inf-A
STP-inf-A
STP-inf-A
STP-inf-A
STP-inf-A
STP-inf-A
STP-inf-A
STP-inf-A
STP-inf-A
STP-inf-A
STP-inf-A
STP-inf-A
STP-inf-A
STP-inf-A
STP-inf-A
STP-inf-A
STP-inf-A
STP-inf-A
STP-inf-A
STP-inf-A
STP-inf-A
STP-inf-A
STP-inf-A
STP-inf-A
STP-inf-A
STP-inf-A
STP-inf-A
STP-inf-A
STP-inf-A
STP-inf-A
STP-inf-A
STP-inf-A
STP-inf-A
STP-inf-A
STP-inf-A
STP-inf-A
STP-inf-A
STP-inf-A
STP-inf-A
STP-inf-A
STP-inf-A

STP-inf-A
STP-inf-A
STP-inf-A
STP-inf-A
STP-inf-A
STP-inf-A
STP-inf-A
STP-inf-A
STP-inf-A
STP-inf-A
STP-inf-A
STP-inf-A
STP-inf-A
STP-inf-A
STP-inf-A
STP-inf-A
STP-inf-A
STP-inf-A
STP-inf-A
STP-inf-A
STP-inf-A
STP-inf-A
STP-inf-A
STP-inf-A
STP-inf-A
STP-inf-A
STP-inf-A
STP-inf-A
STP-inf-A
STP-inf-A
STP-inf-A
STP-inf-A
STP-inf-A
STP-inf-A
STP-inf-A
STP-inf-A
STP-inf-A
STP-inf-A
STP-inf-A
STP-inf-A
STP-inf-A
STP-inf-A
STP-inf-A
STP-inf-A
STP-inf-A
STP-inf-A

STP-inf-A
STP-inf-A
STP-inf-A
STP-inf-A
STP-inf-A
STP-inf-A
STP-inf-A
STP-inf-A
STP-inf-A
STP-inf-A
STP-inf-A
STP-inf-A
STP-inf-A
STP-inf-A
STP-inf-A
STP-inf-A
STP-inf-A
STP-inf-A
STP-inf-A
STP-inf-A
STP-inf-A
STP-inf-A
STP-inf-A
STP-inf-A
STP-inf-A
STP-inf-A
STP-inf-A
STP-inf-A
STP-inf-A
STP-inf-A
STP-inf-A
STP-inf-A
STP-inf-A
STP-inf-A
STP-inf-A
STP-inf-A
STP-inf-A
STP-inf-A
STP-inf-A
STP-inf-A
STP-inf-A
STP-inf-A
STP-inf-A
STP-inf-A
STP-inf-A
STP-inf-A

STP-inf-A
STP-inf-A
STP-inf-A
STP-inf-A
STP-inf-A
STP-inf-A
STP-inf-A
STP-inf-A
STP-inf-A
STP-inf-A
STP-inf-A
STP-inf-A
STP-inf-A
STP-inf-A
STP-inf-A
STP-inf-A
STP-inf-A
STP-inf-A
STP-inf-A
STP-inf-A
STP-inf-A
STP-inf-A
STP-inf-A
STP-inf-A
STP-inf-A
STP-inf-A
STP-inf-A
STP-inf-A
STP-inf-A
STP-inf-A
STP-inf-A
STP-inf-A
STP-inf-A
STP-inf-A
STP-inf-A
STP-inf-A
STP-inf-A
STP-inf-A
STP-inf-A
STP-inf-A
STP-inf-A
STP-inf-A
STP-inf-A
STP-inf-A
STP-inf-A
STP-inf-A

STP-inf-A
STP-inf-A
STP-inf-A
STP-inf-A
STP-inf-A
STP-inf-A
STP-inf-A
STP-inf-A
STP-inf-A
STP-inf-A
STP-inf-A
STP-inf-A
STP-inf-A
STP-inf-A
STP-inf-A
STP-inf-A
STP-inf-A
STP-inf-A
STP-inf-A
STP-inf-A
STP-inf-A
STP-inf-A
STP-inf-A
STP-inf-A
STP-inf-A
STP-inf-A
STP-inf-A
STP-inf-A
STP-inf-A
STP-inf-A
STP-inf-A
STP-inf-A
STP-inf-A
STP-inf-A
STP-inf-A
STP-inf-A
STP-inf-A
STP-inf-A
STP-inf-A
STP-inf-A
STP-inf-A
STP-inf-A
STP-inf-A
STP-inf-A
STP-inf-A
STP-inf-A

STP-inf-A
STP-inf-A
STP-inf-A
STP-inf-A
STP-inf-A
STP-inf-A
STP-inf-A
STP-inf-A
STP-inf-A
STP-inf-A
STP-inf-A
STP-inf-A
STP-inf-A
STP-inf-A
STP-inf-A
STP-inf-A
STP-inf-A
STP-inf-A
STP-inf-A
STP-inf-A
STP-inf-A
STP-inf-A
STP-inf-A
STP-inf-A
STP-inf-A
STP-inf-A
STP-inf-A
STP-inf-A
STP-inf-A
STP-inf-A
STP-inf-A
STP-inf-A
STP-inf-A
STP-inf-A
STP-inf-A
STP-inf-A
STP-inf-A
STP-inf-A
STP-inf-A
STP-inf-A
STP-inf-A
STP-inf-A
STP-inf-A
STP-inf-A
STP-inf-A
STP-inf-A

STP-inf-A
STP-inf-A
STP-inf-A
STP-inf-A
STP-inf-A
STP-inf-A
STP-inf-A
STP-inf-A
STP-inf-A
STP-inf-A
STP-inf-A
STP-inf-A
STP-inf-A
STP-inf-A
STP-inf-A
STP-inf-A
STP-inf-A
STP-inf-A
STP-inf-A
STP-inf-A
STP-inf-A
STP-inf-A
STP-inf-A
STP-inf-A
STP-inf-A
STP-inf-A
STP-inf-A
STP-inf-A
STP-inf-A
STP-inf-A
STP-inf-A
STP-inf-A
STP-inf-A
STP-inf-A
STP-inf-A
STP-inf-A
STP-inf-A
STP-inf-A
STP-inf-A
STP-inf-A
STP-inf-A
STP-inf-A
STP-inf-A
STP-inf-A
STP-inf-A
STP-inf-A

STP-inf-A
STP-inf-A
STP-inf-A
STP-inf-A
STP-inf-A
STP-inf-A
STP-inf-A
STP-inf-A
STP-inf-A
STP-inf-A
STP-inf-A
STP-inf-A
STP-inf-A
STP-inf-A
STP-inf-A
STP-inf-A
STP-inf-A
STP-inf-A
STP-inf-A
STP-inf-A
STP-inf-A
STP-inf-A
STP-inf-A
STP-inf-A
STP-inf-A
STP-inf-A
STP-inf-A
STP-inf-A
STP-inf-A
STP-inf-A
STP-inf-A
STP-inf-A
STP-inf-A
STP-inf-A
STP-inf-A
STP-inf-A
STP-inf-A
STP-inf-A
STP-inf-A
STP-inf-A
STP-inf-A
STP-inf-A
STP-inf-A
STP-inf-A
STP-inf-A
STP-inf-A

STP-inf-A
STP-inf-A
STP-inf-A
STP-inf-A
STP-inf-A
STP-inf-A
STP-inf-A
STP-inf-A
STP-inf-A
STP-inf-A
STP-inf-A
STP-inf-A
STP-inf-A
STP-inf-A
STP-inf-A
STP-inf-A
STP-inf-A
STP-inf-A
STP-inf-A
STP-inf-A
STP-inf-A
STP-inf-A
STP-inf-A
STP-inf-A
STP-inf-A
STP-inf-A
STP-inf-A
STP-inf-A
STP-inf-A
STP-inf-A
STP-inf-A
STP-inf-A
STP-inf-A
STP-inf-A
STP-inf-A
STP-inf-A
STP-inf-A
STP-inf-A
STP-inf-A
STP-inf-A
STP-inf-A
STP-inf-A
STP-inf-A
STP-inf-A
STP-inf-A
STP-inf-A

STP-inf-A
STP-inf-A
STP-inf-A
STP-inf-A
STP-inf-A
STP-inf-A
STP-inf-A
STP-inf-A
STP-inf-A
STP-inf-A
STP-inf-A
STP-inf-A
STP-inf-A
STP-inf-A
STP-inf-A
STP-inf-A
STP-inf-A
STP-inf-A
STP-inf-A
STP-inf-A
STP-inf-A
STP-inf-A
STP-inf-A
STP-inf-A
STP-inf-A
STP-inf-A
STP-inf-A
STP-inf-A
STP-inf-A
STP-inf-A
STP-inf-A
STP-inf-A
STP-inf-A
STP-inf-A
STP-inf-A
STP-inf-A
STP-inf-A
STP-inf-A
STP-inf-A
STP-inf-A
STP-inf-A
STP-inf-A
STP-inf-A
STP-inf-A
STP-inf-A
STP-inf-A

STP-inf-A
STP-inf-A
STP-inf-A
STP-inf-A
STP-inf-A
STP-inf-A
STP-inf-A
STP-inf-A
STP-inf-A
STP-inf-A
STP-inf-A
STP-inf-A
STP-inf-A
STP-inf-A
STP-inf-A
STP-inf-A
STP-inf-A
STP-inf-A
STP-inf-A
STP-inf-A
STP-inf-A
STP-inf-A
STP-inf-A
STP-inf-A
STP-inf-A
STP-inf-A
STP-inf-A
STP-inf-A
STP-inf-A
STP-inf-A
STP-inf-A
STP-inf-A
STP-inf-A
STP-inf-A
STP-inf-A
STP-inf-A
STP-inf-A
STP-inf-A
STP-inf-A
STP-inf-A
STP-inf-A
STP-inf-A
STP-inf-A
STP-inf-A
STP-inf-A
STP-inf-A

STP-inf-A
STP-inf-A
STP-inf-A
STP-inf-A
STP-inf-A
STP-inf-A
STP-inf-A
STP-inf-A
STP-inf-A
STP-inf-A
STP-inf-A
STP-inf-A
STP-inf-A
STP-inf-A
STP-inf-A
STP-inf-A
STP-inf-A
STP-inf-A
STP-inf-A
STP-inf-A
STP-inf-A
STP-inf-A
STP-inf-A
STP-inf-A
STP-inf-A
STP-inf-A
STP-inf-A
STP-inf-A
STP-inf-A
STP-inf-A
STP-inf-A
STP-inf-A
STP-inf-A
STP-inf-A
STP-inf-A
STP-inf-A
STP-inf-A
STP-inf-A
STP-inf-A
STP-inf-A
STP-inf-A
STP-inf-A
STP-inf-A
STP-inf-A
STP-inf-A
STP-inf-A

STP-inf-A
STP-inf-A
STP-inf-A
STP-inf-A
STP-inf-A
STP-inf-A
STP-inf-A
STP-inf-A
STP-inf-A
STP-inf-A
STP-inf-A
STP-inf-A
STP-inf-A
STP-inf-A
STP-inf-A
STP-inf-A
STP-inf-A
STP-inf-A
STP-inf-A
STP-inf-A
STP-inf-A
STP-inf-A
STP-inf-A
STP-inf-A
STP-inf-A
STP-inf-A
STP-inf-A
STP-inf-A
STP-inf-A
STP-inf-A
STP-inf-A
STP-inf-A
STP-inf-A
STP-inf-A
STP-inf-A
STP-inf-A
STP-inf-A
STP-inf-A
STP-inf-A
STP-inf-A
STP-inf-A
STP-inf-A
STP-inf-A
STP-inf-A
STP-inf-A
STP-inf-A

STP-inf-A
STP-inf-A
STP-inf-A
STP-inf-A
STP-inf-A
STP-inf-A
STP-inf-A
STP-inf-A
STP-inf-A
STP-inf-A
STP-inf-A
STP-inf-A
STP-inf-A
STP-inf-A
STP-inf-A
STP-inf-A
STP-inf-A
STP-inf-A
STP-inf-A
STP-inf-A
STP-inf-A
STP-inf-A
STP-inf-A
STP-inf-A
STP-inf-A
STP-inf-A
STP-inf-A
STP-inf-A
STP-inf-A
STP-inf-A
STP-inf-A
STP-inf-A
STP-inf-A
STP-inf-A
STP-inf-A
STP-inf-A
STP-inf-A
STP-inf-A
STP-inf-A
STP-inf-A
STP-inf-A
STP-inf-A
STP-inf-A
STP-inf-A
STP-inf-A
STP-inf-A

STP-inf-A
STP-inf-A
STP-inf-A
STP-inf-A
STP-inf-A
STP-inf-A
STP-inf-A
STP-inf-A
STP-inf-A
STP-inf-A
STP-inf-A
STP-inf-A
STP-inf-A
STP-inf-A
STP-inf-A
STP-inf-A
STP-inf-A
STP-inf-A
STP-inf-A
STP-inf-A
STP-inf-A
STP-inf-A
STP-inf-A
STP-inf-A
STP-inf-A
STP-inf-A
STP-inf-A
STP-inf-A
STP-inf-A
STP-inf-A
STP-inf-A
STP-inf-A
STP-inf-A
STP-inf-A
STP-inf-A
STP-T-eff-F
STP-T-eff-F
STP-T-eff-F
STP-T-eff-F
STP-T-eff-F
STP-T-eff-F
STP-T-eff-F
STP-T-eff-F
STP-T-eff-F
STP-T-eff-F
STP-T-eff-F

STP-T-eff-F
STP-T-eff-F
STP-T-eff-F
STP-T-eff-F
STP-T-eff-F
STP-T-eff-F
STP-T-eff-F
STP-T-eff-F
STP-T-eff-F
STP-T-eff-F
STP-T-eff-F
STP-T-eff-F
STP-T-eff-F
STP-T-eff-F
STP-T-eff-F
STP-T-eff-F
STP-T-eff-F
STP-T-eff-F
STP-T-eff-F
STP-T-eff-F
STP-T-eff-F
STP-T-eff-F
STP-T-eff-F
STP-T-eff-F
STP-T-eff-F
STP-T-eff-F
STP-T-eff-F
STP-T-eff-F
STP-T-eff-F
STP-T-eff-F
STP-T-eff-F
STP-T-eff-F
STP-T-eff-F
STP-T-eff-F
STP-T-eff-F
STP-T-eff-F
STP-T-eff-F
STP-T-eff-F
STP-T-eff-F
STP-T-eff-F
STP-T-eff-F
STP-T-eff-F
STP-T-eff-F
STP-T-eff-F
STP-T-eff-F
STP-T-eff-F

STP-T-eff-F
STP-T-eff-F
STP-T-eff-F
STP-T-eff-F
STP-T-eff-F
STP-T-eff-F
STP-T-eff-F
STP-T-eff-F
STP-T-eff-F
STP-T-eff-F
STP-T-eff-F
STP-T-eff-F
STP-T-eff-F
STP-T-eff-F
STP-T-eff-F
STP-T-eff-F
STP-T-eff-F
STP-T-eff-F
STP-T-eff-F
STP-T-eff-F
STP-T-eff-F
STP-T-eff-F
STP-T-eff-F
STP-T-eff-F
STP-T-eff-F
STP-T-eff-F
STP-T-eff-F
STP-T-eff-F
STP-T-eff-F
STP-T-eff-F
STP-T-eff-F
STP-T-eff-F
STP-T-eff-F
STP-T-eff-F
STP-T-eff-F
STP-T-eff-F
STP-T-eff-F
STP-T-eff-F
STP-T-eff-F
STP-T-eff-F
STP-T-eff-F
STP-T-eff-F
STP-T-eff-F
STP-T-eff-F
STP-T-eff-F
STP-T-eff-F

STP-T-eff-F
STP-T-eff-F
STP-T-eff-F
STP-T-eff-F
STP-T-eff-F
STP-T-eff-F
STP-T-eff-F
STP-T-eff-F
STP-T-eff-F
STP-T-eff-F
STP-T-eff-F
STP-T-eff-F
STP-T-eff-F
STP-T-eff-F
STP-T-eff-F
STP-T-eff-F
STP-T-eff-F
STP-T-eff-F
STP-T-eff-F
STP-T-eff-F
STP-T-eff-F
STP-T-eff-F
STP-T-eff-F
STP-T-eff-F
STP-T-eff-F
STP-T-eff-F
STP-T-eff-F
STP-T-eff-F
STP-T-eff-F
STP-T-eff-F
STP-T-eff-F
STP-T-eff-F
STP-T-eff-F
STP-T-eff-F
STP-T-eff-F
STP-T-eff-F
STP-T-eff-F
STP-T-eff-F
STP-T-eff-F
STP-T-eff-F
STP-T-eff-F
STP-T-eff-F
STP-T-eff-F
STP-T-eff-F
STP-T-eff-F
STP-T-eff-F

STP-T-eff-F
STP-T-eff-F
STP-T-eff-F
STP-T-eff-F
STP-T-eff-F
STP-T-eff-F
STP-T-eff-F
STP-T-eff-F
STP-T-eff-F
STP-T-eff-F
STP-T-eff-F
STP-T-eff-F
STP-T-eff-F
STP-T-eff-F
STP-T-eff-F
STP-T-eff-F
STP-T-eff-F
STP-T-eff-F
STP-T-eff-F
STP-T-eff-F
STP-T-eff-F
STP-T-eff-F
STP-T-eff-F
STP-T-eff-F
STP-T-eff-F
STP-T-eff-F
STP-T-eff-F
STP-T-eff-F
STP-T-eff-F
STP-T-eff-F
STP-T-eff-F
STP-T-eff-F
STP-T-eff-F
STP-T-eff-F
STP-T-eff-F
STP-T-eff-F
STP-T-eff-F
STP-T-eff-F
STP-T-eff-F
STP-T-eff-F
STP-T-eff-F
STP-T-eff-F
STP-T-eff-F
STP-T-eff-F
STP-T-eff-F
STP-T-eff-F

STP-T-eff-F
STP-T-eff-F
STP-T-eff-F
STP-T-eff-F
STP-T-eff-F
STP-T-eff-F
STP-T-eff-F
STP-T-eff-F
STP-T-eff-F
STP-T-eff-F
STP-T-eff-F
STP-T-eff-F
STP-T-eff-F
STP-T-eff-F
STP-T-eff-F
STP-T-eff-F
STP-T-eff-F
STP-T-eff-F
STP-T-eff-F
STP-T-eff-F
STP-T-eff-F
STP-T-eff-F
STP-T-eff-F
STP-T-eff-F
STP-T-eff-F
STP-T-eff-F
STP-T-eff-F
STP-T-eff-F
STP-T-eff-F
STP-T-eff-F
STP-T-eff-F
STP-T-eff-F
STP-T-eff-F
STP-T-eff-F
STP-T-eff-F
STP-T-eff-F
STP-T-eff-F
STP-T-eff-F
STP-T-eff-F
STP-T-eff-F
STP-T-eff-F
STP-T-eff-F
STP-T-eff-F
STP-T-eff-F
STP-T-eff-F
STP-T-eff-F

STP-T-eff-F
STP-T-eff-F
STP-T-eff-F
STP-T-eff-F
STP-T-eff-F
STP-T-eff-F
STP-T-eff-F
STP-T-eff-F
STP-T-eff-F
STP-T-eff-F
STP-T-eff-F
STP-T-eff-F
STP-T-eff-F
STP-T-eff-F
STP-T-eff-F
STP-T-eff-F
STP-T-eff-F
STP-T-eff-F
STP-T-eff-F
STP-T-eff-F
STP-T-eff-F
STP-T-eff-F
STP-T-eff-F
STP-T-eff-F
STP-T-eff-F
STP-T-eff-F
STP-T-eff-F
STP-T-eff-F
STP-T-eff-F
STP-T-eff-F
STP-T-eff-F
STP-T-eff-F
STP-T-eff-F
STP-T-eff-F
STP-T-eff-F
STP-T-eff-F
STP-T-eff-F
STP-T-eff-F
STP-T-eff-F
STP-T-eff-F
STP-T-eff-F
STP-T-eff-F
STP-T-eff-F
STP-T-eff-F
STP-T-eff-F
STP-T-eff-F

STP-T-eff-F
STP-T-eff-F
STP-T-eff-F
STP-T-eff-F
STP-T-eff-F
STP-T-eff-F
STP-T-eff-F
STP-T-eff-F
STP-T-eff-F
STP-T-eff-F
STP-T-eff-F
STP-T-eff-F
STP-T-eff-F
STP-T-eff-F
STP-T-eff-F
STP-T-eff-F
STP-T-eff-F
STP-T-eff-F
STP-T-eff-F
STP-T-eff-F
STP-T-eff-F
STP-T-eff-F
STP-T-eff-F
STP-T-eff-F
STP-T-eff-F
STP-T-eff-F
STP-T-eff-F
STP-T-eff-F
STP-T-eff-F
STP-T-eff-F
STP-T-eff-F
STP-T-eff-F
STP-T-eff-F
STP-T-eff-F
STP-T-eff-F
STP-T-eff-F
STP-T-eff-F
STP-T-eff-F
STP-T-eff-F
STP-T-eff-F
STP-T-eff-F
STP-T-eff-F
STP-T-eff-F
STP-T-eff-F
STP-T-eff-F
STP-T-eff-F

STP-T-eff-F
STP-T-eff-F
STP-T-eff-F
STP-T-eff-F
STP-T-eff-F
STP-T-eff-F
STP-T-eff-F
STP-T-eff-F
STP-T-eff-F
STP-T-eff-F
STP-T-eff-F
STP-T-eff-F
STP-T-eff-F
STP-T-eff-F
STP-T-eff-F
STP-T-eff-F
STP-T-eff-F
STP-T-eff-F
STP-T-eff-F
STP-T-eff-F
STP-T-eff-F
STP-T-eff-F
STP-T-eff-F
STP-T-eff-F
STP-T-eff-F
STP-T-eff-F
STP-T-eff-F
STP-T-eff-F
STP-T-eff-F
STP-T-eff-F
STP-T-eff-F
STP-T-eff-F
STP-T-eff-F
STP-T-eff-F
STP-T-eff-F
STP-T-eff-F
STP-T-eff-F
STP-T-eff-F
STP-T-eff-F
STP-T-eff-F
STP-T-eff-F
STP-T-eff-F
STP-T-eff-F
STP-T-eff-F
STP-T-eff-F
STP-T-eff-F

STP-T-eff-F
STP-T-eff-F
STP-T-eff-F
STP-T-eff-F
STP-T-eff-F
STP-T-eff-F
STP-T-eff-F
STP-T-eff-F
STP-T-eff-F
STP-T-eff-F
STP-T-eff-F
STP-T-eff-F
STP-T-eff-F
STP-T-eff-F
STP-T-eff-F
STP-T-eff-F
STP-T-eff-F
STP-T-eff-F
STP-T-eff-F
STP-T-eff-F
STP-T-eff-F
STP-T-eff-F
STP-T-eff-F
STP-T-eff-F
STP-T-eff-F
STP-T-eff-F
STP-T-eff-F
STP-T-eff-F
STP-T-eff-F
STP-T-eff-F
STP-T-eff-F
STP-T-eff-F
STP-T-eff-F
STP-T-eff-F
STP-T-eff-F
STP-T-eff-F
STP-T-eff-F
STP-T-eff-F
STP-T-eff-F
STP-T-eff-F
STP-T-eff-F
STP-T-eff-F
STP-T-eff-F
STP-T-eff-F
STP-T-eff-F
STP-T-eff-F

STP-T-eff-F
STP-T-eff-F
STP-T-eff-F
STP-T-eff-F
STP-T-eff-F
STP-T-eff-F
STP-T-eff-F
STP-T-eff-F
STP-T-eff-F
STP-T-eff-F
STP-T-eff-F
STP-T-eff-F
STP-T-eff-F
STP-T-eff-F
STP-T-eff-F
STP-T-eff-F
STP-T-eff-F
STP-T-eff-F
STP-T-eff-F
STP-T-eff-F
STP-T-eff-F
STP-T-eff-F
STP-T-eff-F
STP-T-eff-F
STP-T-eff-F
STP-T-eff-F
STP-T-eff-F
STP-T-eff-F
STP-T-eff-F
STP-T-eff-F
STP-T-eff-F
STP-T-eff-F
STP-T-eff-F
STP-T-eff-F
STP-T-eff-F
STP-T-eff-F
STP-T-eff-F
STP-T-eff-F
STP-T-eff-F
STP-T-eff-F
STP-T-eff-F
STP-T-eff-F
STP-T-eff-F
STP-T-eff-F
STP-T-eff-F
STP-T-eff-F

STP-T-eff-F
STP-T-eff-F
STP-T-eff-F
STP-T-eff-F
STP-T-eff-F
STP-T-eff-F
STP-T-eff-F
STP-T-eff-F
STP-T-eff-F
STP-T-eff-F
STP-T-eff-F
STP-T-eff-F
STP-T-eff-F
STP-T-eff-F
STP-T-eff-F
STP-T-eff-F
STP-T-eff-F
STP-T-eff-F
STP-T-eff-F
STP-T-eff-F
STP-T-eff-F
STP-T-eff-F
STP-T-eff-F
STP-T-eff-F
STP-T-eff-F
STP-T-eff-F
STP-T-eff-F
STP-T-eff-F
STP-T-eff-F
STP-T-eff-F
STP-T-eff-F
STP-T-eff-F
STP-T-eff-F
STP-T-eff-F
STP-T-eff-F
STP-T-eff-F
STP-T-eff-F
STP-T-eff-F
STP-T-eff-F
STP-T-eff-F
STP-T-eff-F
STP-T-eff-F
STP-T-eff-F
STP-T-eff-F
STP-T-eff-F
STP-T-eff-F

STP-T-eff-F
STP-T-eff-F
STP-T-eff-F
STP-T-eff-F
STP-T-eff-F
STP-T-eff-F
STP-T-eff-F
STP-T-eff-F
STP-T-eff-F
STP-T-eff-F
STP-T-eff-F
STP-T-eff-F
STP-T-eff-F
STP-T-eff-F
STP-T-eff-F
STP-T-eff-F
STP-T-eff-F
STP-T-eff-F
STP-T-eff-F
STP-T-eff-F
STP-T-eff-F
STP-T-eff-F
STP-T-eff-F
STP-T-eff-F
STP-T-eff-F
STP-T-eff-F
STP-T-eff-F
STP-T-eff-F
STP-T-eff-F
STP-T-eff-F
STP-T-eff-F
STP-T-eff-F
STP-T-eff-F
STP-T-eff-F
STP-T-eff-F
STP-T-eff-F
STP-T-eff-F
STP-T-eff-F
STP-T-eff-F
STP-T-eff-F
STP-T-eff-F
STP-T-eff-F
STP-T-eff-F
STP-T-eff-F
STP-T-eff-F
STP-T-eff-F

STP-T-eff-F
STP-T-eff-F
STP-T-eff-F
STP-T-eff-F
STP-T-eff-F
STP-T-eff-F
STP-T-eff-F
STP-T-eff-F
STP-T-eff-F
STP-T-eff-F
STP-T-eff-F
STP-T-eff-F
STP-T-eff-F
STP-T-eff-F
STP-T-eff-F
STP-T-eff-F
STP-T-eff-F
STP-T-eff-F
STP-T-eff-F
STP-T-eff-F
STP-T-eff-F
STP-T-eff-F
STP-T-eff-F
STP-T-eff-F
STP-T-eff-F
STP-T-eff-F
STP-T-eff-F
STP-T-eff-F
STP-T-eff-F
STP-T-eff-F
STP-T-eff-F
STP-T-eff-F
STP-T-eff-F
STP-T-eff-F
STP-T-eff-F
STP-T-eff-F
STP-T-eff-F
STP-T-eff-F
STP-T-eff-F
STP-T-eff-F
STP-T-eff-F
STP-T-eff-F
STP-T-eff-F
STP-T-eff-F
STP-T-eff-F
STP-T-eff-F

STP-T-eff-F
STP-T-eff-F
STP-T-eff-F
STP-T-eff-F
STP-T-eff-F
STP-T-eff-F
STP-T-eff-F
STP-T-eff-F
STP-T-eff-F
STP-T-eff-F
STP-T-eff-F
STP-T-eff-F
STP-T-eff-F
STP-T-eff-F
STP-T-eff-F
STP-T-eff-F
STP-T-eff-F
STP-T-eff-F
STP-T-eff-F
STP-T-eff-F
STP-T-eff-F
STP-T-eff-F
STP-T-eff-F
STP-T-eff-F
STP-T-eff-F
STP-T-eff-F
STP-T-eff-F
STP-T-eff-F
STP-T-eff-F
STP-T-eff-F
STP-T-eff-F
STP-T-eff-F
STP-T-eff-F
STP-T-eff-F
STP-T-eff-F
STP-T-eff-F
STP-T-eff-F
STP-T-eff-A
STP-T-eff-A
STP-T-eff-A
STP-T-eff-A
STP-T-eff-A
STP-T-eff-A
STP-T-eff-A
STP-T-eff-A
STP-T-eff-A

STP-T-eff-A
STP-T-eff-A
STP-T-eff-A
STP-T-eff-A
STP-T-eff-A
STP-T-eff-A
STP-T-eff-A
STP-T-eff-A
STP-T-eff-A
STP-T-eff-A
STP-T-eff-A
STP-T-eff-A
STP-T-eff-A
STP-T-eff-A
STP-T-eff-A
STP-T-eff-A
STP-T-eff-A
STP-T-eff-A
STP-T-eff-A
STP-T-eff-A
STP-T-eff-A
STP-T-eff-A
STP-T-eff-A
STP-T-eff-A
STP-T-eff-A
STP-T-eff-A
STP-T-eff-A
STP-T-eff-A
STP-T-eff-A
STP-T-eff-A
STP-T-eff-A
STP-T-eff-A
STP-T-eff-A
STP-T-eff-A
STP-T-eff-A
STP-T-eff-A
STP-T-eff-A
STP-T-eff-A
STP-T-eff-A
STP-T-eff-A
STP-T-eff-A
STP-T-eff-A
STP-T-eff-A
STP-T-eff-A
STP-T-eff-A
STP-T-eff-A

STP-T-eff-A
STP-T-eff-A
STP-T-eff-A
STP-T-eff-A
STP-T-eff-A
STP-T-eff-A
STP-T-eff-A
STP-T-eff-A
STP-T-eff-A
STP-T-eff-A
STP-T-eff-A
STP-T-eff-A
STP-T-eff-A
STP-T-eff-A
STP-T-eff-A
STP-T-eff-A
STP-T-eff-A
STP-T-eff-A
STP-T-eff-A
STP-T-eff-A
STP-T-eff-A
STP-T-eff-A
STP-T-eff-A
STP-T-eff-A
STP-T-eff-A
STP-T-eff-A
STP-T-eff-A
STP-T-eff-A
STP-T-eff-A
STP-T-eff-A
STP-T-eff-A
STP-T-eff-A
STP-T-eff-A
STP-T-eff-A
STP-T-eff-A
STP-T-eff-A
STP-T-eff-A
STP-T-eff-A
STP-T-eff-A
STP-T-eff-A
STP-T-eff-A
STP-T-eff-A
STP-T-eff-A
STP-T-eff-A
STP-T-eff-A
STP-T-eff-A

STP-T-eff-A
STP-T-eff-A
STP-T-eff-A
STP-T-eff-A
STP-T-eff-A
STP-T-eff-A
STP-T-eff-A
STP-T-eff-A
STP-T-eff-A
STP-T-eff-A
STP-T-eff-A
STP-T-eff-A
STP-T-eff-A
STP-T-eff-A
STP-T-eff-A
STP-T-eff-A
STP-T-eff-A
STP-T-eff-A
STP-T-eff-A
STP-T-eff-A
STP-T-eff-A
STP-T-eff-A
STP-T-eff-A
STP-T-eff-A
STP-T-eff-A
STP-T-eff-A
STP-T-eff-A
STP-T-eff-A
STP-T-eff-A
STP-T-eff-A
STP-T-eff-A
STP-T-eff-A
STP-T-eff-A
STP-T-eff-A
STP-T-eff-A
STP-T-eff-A
STP-T-eff-A
STP-T-eff-A
STP-T-eff-A
STP-T-eff-A
STP-T-eff-A
STP-T-eff-A
STP-T-eff-A
STP-T-eff-A
STP-T-eff-A
STP-T-eff-A

STP-T-eff-A
STP-T-eff-A
STP-T-eff-A
STP-T-eff-A
STP-T-eff-A
STP-T-eff-A
STP-T-eff-A
STP-T-eff-A
STP-T-eff-A
STP-T-eff-A
STP-T-eff-A
STP-T-eff-A
STP-T-eff-A
STP-T-eff-A
STP-T-eff-A
STP-T-eff-A
STP-T-eff-A
STP-T-eff-A
STP-T-eff-A
STP-T-eff-A
STP-T-eff-A
STP-T-eff-A
STP-T-eff-A
STP-T-eff-A
STP-T-eff-A
STP-T-eff-A
STP-T-eff-A
STP-T-eff-A
STP-T-eff-A
STP-T-eff-A
STP-T-eff-A
STP-T-eff-A
STP-T-eff-A
STP-T-eff-A
STP-T-eff-A
STP-T-eff-A
STP-T-eff-A
STP-T-eff-A
STP-T-eff-A
STP-T-eff-A
STP-T-eff-A
STP-T-eff-A
STP-T-eff-A
STP-T-eff-A
STP-T-eff-A
STP-T-eff-A

STP-T-eff-A
STP-T-eff-A
STP-T-eff-A
STP-T-eff-A
STP-T-eff-A
STP-T-eff-A
STP-T-eff-A
STP-T-eff-A
STP-T-eff-A
STP-T-eff-A
STP-T-eff-A
STP-T-eff-A
STP-T-eff-A
STP-T-eff-A
STP-T-eff-A
STP-T-eff-A
STP-T-eff-A
STP-T-eff-A
STP-T-eff-A
STP-T-eff-A
STP-T-eff-A
STP-T-eff-A
STP-T-eff-A
STP-T-eff-A
STP-T-eff-A
STP-T-eff-A
STP-T-eff-A
STP-T-eff-A
STP-T-eff-A
STP-T-eff-A
STP-T-eff-A
STP-T-eff-A
STP-T-eff-A
STP-T-eff-A
STP-T-eff-A
STP-T-eff-A
STP-T-eff-A
STP-T-eff-A
STP-T-eff-A
STP-T-eff-A
STP-T-eff-A
STP-T-eff-A
STP-T-eff-A
STP-T-eff-A
STP-T-eff-A
STP-T-eff-A

STP-T-eff-A
STP-T-eff-A
STP-T-eff-A
STP-T-eff-A
STP-T-eff-A
STP-T-eff-A
STP-T-eff-A
STP-T-eff-A
STP-T-eff-A
STP-T-eff-A
STP-T-eff-A
STP-T-eff-A
STP-T-eff-A
STP-T-eff-A
STP-T-eff-A
STP-T-eff-A
STP-T-eff-A
STP-T-eff-A
STP-T-eff-A
STP-T-eff-A
STP-T-eff-A
STP-T-eff-A
STP-T-eff-A
STP-T-eff-A
STP-T-eff-A
STP-T-eff-A
STP-T-eff-A
STP-T-eff-A
STP-T-eff-A
STP-T-eff-A
STP-T-eff-A
STP-T-eff-A
STP-T-eff-A
STP-T-eff-A
STP-T-eff-A
STP-T-eff-A
STP-T-eff-A
STP-T-eff-A
STP-T-eff-A
STP-T-eff-A
STP-T-eff-A
STP-T-eff-A
STP-T-eff-A
STP-T-eff-A
STP-T-eff-A
STP-T-eff-A

STP-T-eff-A
STP-T-eff-A
STP-T-eff-A
STP-T-eff-A
STP-T-eff-A
STP-T-eff-A
STP-T-eff-A
STP-T-eff-A
STP-T-eff-A
STP-T-eff-A
STP-T-eff-A
STP-T-eff-A
STP-T-eff-A
STP-T-eff-A
STP-T-eff-A
STP-T-eff-A
STP-T-eff-A
STP-T-eff-A
STP-T-eff-A
STP-T-eff-A
STP-T-eff-A
STP-T-eff-A
STP-T-eff-A
STP-T-eff-A
STP-T-eff-A
STP-T-eff-A
STP-T-eff-A
STP-T-eff-A
STP-T-eff-A
STP-T-eff-A
STP-T-eff-A
STP-T-eff-A
STP-T-eff-A
STP-T-eff-A
STP-T-eff-A
STP-T-eff-A
STP-T-eff-A
STP-T-eff-A
STP-T-eff-A
STP-T-eff-A
STP-T-eff-A
STP-T-eff-A
STP-T-eff-A
STP-T-eff-A
STP-T-eff-A
STP-T-eff-A

STP-T-eff-A
STP-T-eff-A
STP-T-eff-A
STP-T-eff-A
STP-T-eff-A
STP-T-eff-A
STP-T-eff-A
STP-T-eff-A
STP-T-eff-A
STP-T-eff-A
STP-T-eff-A
STP-T-eff-A
STP-T-eff-A
STP-T-eff-A
STP-T-eff-A
STP-T-eff-A
STP-T-eff-A
STP-T-eff-A
STP-T-eff-A
STP-T-eff-A
STP-T-eff-A
STP-T-eff-A
STP-T-eff-A
STP-T-eff-A
STP-T-eff-A
STP-T-eff-A
STP-T-eff-A
STP-T-eff-A
STP-T-eff-A
STP-T-eff-A
STP-T-eff-A
STP-T-eff-A
STP-T-eff-A
STP-T-eff-A
STP-T-eff-A
STP-T-eff-A
STP-T-eff-A
STP-T-eff-A
STP-T-eff-A
STP-T-eff-A
STP-T-eff-A
STP-T-eff-A
STP-T-eff-A
STP-T-eff-A
STP-T-eff-A
STP-T-eff-A

STP-T-eff-A
STP-T-eff-A
STP-T-eff-A
STP-T-eff-A
STP-T-eff-A
STP-T-eff-A
STP-T-eff-A
STP-T-eff-A
STP-T-eff-A
STP-T-eff-A
STP-T-eff-A
STP-T-eff-A
STP-T-eff-A
STP-T-eff-A
STP-T-eff-A
STP-T-eff-A
STP-T-eff-A
STP-T-eff-A
STP-T-eff-A
STP-T-eff-A
STP-T-eff-A
STP-T-eff-A
STP-T-eff-A
STP-T-eff-A
STP-T-eff-A
STP-T-eff-A
STP-T-eff-A
STP-T-eff-A
STP-T-eff-A
STP-T-eff-A
STP-T-eff-A
STP-T-eff-A
STP-T-eff-A
STP-T-eff-A
STP-T-eff-A
STP-T-eff-A
STP-T-eff-A
STP-T-eff-A
STP-T-eff-A
STP-T-eff-A
STP-T-eff-A
STP-T-eff-A
STP-T-eff-A
STP-T-eff-A
STP-T-eff-A
STP-T-eff-A

STP-T-eff-A
STP-T-eff-A
STP-T-eff-A
STP-T-eff-A
STP-T-eff-A
STP-T-eff-A
STP-T-eff-A
STP-T-eff-A
STP-T-eff-A
STP-T-eff-A
STP-T-eff-A
STP-T-eff-A
STP-T-eff-A
STP-T-eff-A
STP-T-eff-A
STP-T-eff-A
STP-T-eff-A
STP-T-eff-A
STP-T-eff-A
STP-T-eff-A
STP-T-eff-A
STP-T-eff-A
STP-T-eff-A
STP-T-eff-A
STP-T-eff-A
STP-T-eff-A
STP-T-eff-A
STP-T-eff-A
STP-T-eff-A
STP-T-eff-A
STP-T-eff-A
STP-T-eff-A
STP-T-eff-A
STP-T-eff-A
STP-T-eff-A
STP-T-eff-A
STP-T-eff-A
STP-T-eff-A
STP-T-eff-A
STP-T-eff-A
STP-T-eff-A
STP-T-eff-A
STP-T-eff-A
STP-T-eff-A
STP-T-eff-A
STP-T-eff-A

STP-T-eff-A
STP-T-eff-A
STP-T-eff-A
STP-T-eff-A
STP-T-eff-A
STP-T-eff-A
STP-T-eff-A
STP-T-eff-A
STP-T-eff-A
STP-T-eff-A
STP-T-eff-A
STP-T-eff-A
STP-T-eff-A
STP-T-eff-A
STP-T-eff-A
STP-T-eff-A
STP-T-eff-A
STP-T-eff-A
STP-T-eff-A
STP-T-eff-A
STP-T-eff-A
STP-T-eff-A
STP-T-eff-A
STP-T-eff-A
STP-T-eff-A
STP-T-eff-A
STP-T-eff-A
STP-T-eff-A
STP-T-eff-A
STP-T-eff-A
STP-T-eff-A
STP-T-eff-A
STP-T-eff-A
STP-T-eff-A
STP-T-eff-A
STP-T-eff-A
STP-T-eff-A
STP-T-eff-A
STP-T-eff-A
STP-T-eff-A
STP-T-eff-A
STP-T-eff-A
STP-T-eff-A
STP-T-eff-A
STP-T-eff-A
STP-T-eff-A

STP-T-eff-A
STP-T-eff-A
STP-T-eff-A
STP-T-eff-A
STP-T-eff-A
STP-T-eff-A
STP-T-eff-A
STP-T-eff-A
STP-T-eff-A
STP-T-eff-A
STP-T-eff-A
STP-T-eff-A
STP-T-eff-A
STP-T-eff-A
STP-T-eff-A
STP-T-eff-A
STP-T-eff-A
STP-T-eff-A
STP-T-eff-A
STP-T-eff-A
STP-T-eff-A
STP-T-eff-A
STP-T-eff-A
STP-T-eff-A
STP-T-eff-A
STP-T-eff-A
STP-T-eff-A
STP-T-eff-A
STP-T-eff-A
STP-T-eff-A
STP-T-eff-A
STP-T-eff-A
STP-T-eff-A
STP-T-eff-A
STP-T-eff-A
STP-T-eff-A
STP-T-eff-A
STP-T-eff-A
STP-T-eff-A
STP-T-eff-A
STP-T-eff-A
STP-T-eff-A
STP-T-eff-A
STP-T-eff-A
STP-T-eff-A
STP-T-eff-A

STP-T-eff-A
STP-T-eff-A
STP-T-eff-A
STP-T-eff-A
STP-T-eff-A
STP-T-eff-A
STP-T-eff-A
STP-T-eff-A
STP-T-eff-A
STP-T-eff-A
STP-T-eff-A
STP-T-eff-A
STP-T-eff-A
STP-T-eff-A
STP-T-eff-A
STP-T-eff-A
STP-T-eff-A
STP-T-eff-A
STP-T-eff-A
STP-T-eff-A
STP-T-eff-A
STP-T-eff-A
STP-T-eff-A
STP-T-eff-A
STP-T-eff-A
STP-T-eff-A
STP-T-eff-A
STP-T-eff-A
STP-T-eff-A
STP-T-eff-A
STP-T-eff-A
STP-T-eff-A
STP-T-eff-A
STP-T-eff-A
STP-T-eff-A
STP-T-eff-A
STP-T-eff-A
STP-T-eff-A
STP-T-eff-A
STP-T-eff-A
STP-T-eff-A
STP-T-eff-A
STP-T-eff-A
STP-T-eff-A
STP-T-eff-A
STP-T-eff-A

STP-T-eff-A
STP-T-eff-A
STP-T-eff-A
STP-T-eff-A
STP-T-eff-A
STP-T-eff-A
STP-T-eff-A
STP-T-eff-A
STP-T-eff-A
STP-T-eff-A
STP-T-eff-A
STP-T-eff-A
STP-T-eff-A
STP-T-eff-A
STP-T-eff-A
STP-T-eff-A
STP-T-eff-A
STP-T-eff-A
STP-T-eff-A
STP-T-eff-A
STP-T-eff-A
STP-T-eff-A
STP-T-eff-A
STP-T-eff-A
STP-T-eff-A
STP-T-eff-A
STP-T-eff-A
STP-T-eff-A
STP-T-eff-A
STP-T-eff-A
STP-T-eff-A
STP-T-eff-A
STP-T-eff-A
STP-T-eff-A
STP-T-eff-A
STP-T-eff-A
STP-T-eff-A
STP-T-eff-A
STP-T-eff-A
STP-T-eff-A
STP-T-eff-A
STP-T-eff-A
STP-T-eff-A
STP-T-eff-A
STP-T-eff-A
STP-T-eff-A

STP-T-eff-A
STP-T-eff-A
STP-T-eff-A
STP-T-eff-A
STP-T-eff-A
STP-T-eff-A
STP-T-eff-A
STP-T-eff-A
STP-T-eff-A
STP-T-eff-A
STP-T-eff-A
STP-T-eff-A
STP-T-eff-A
STP-T-eff-A
STP-T-eff-A
STP-T-eff-A
STP-T-eff-A
STP-T-eff-A
STP-T-eff-A
STP-T-eff-A
STP-T-eff-A
STP-T-eff-A
STP-T-eff-A
STP-T-eff-A
STP-T-eff-A
STP-T-eff-A
STP-T-eff-A
STP-T-eff-A
STP-T-eff-A
STP-T-eff-A
STP-T-eff-A
STP-T-eff-A
STP-T-eff-A
STP-T-eff-A
STP-T-eff-A
STP-T-eff-A
STP-T-eff-A
STP-T-eff-A
STP-T-eff-A
STP-T-eff-A
STP-T-eff-A
STP-T-eff-A
STP-T-eff-A
STP-T-eff-A
STP-T-eff-A
STP-T-eff-A

STP-T-eff-A
STP-T-eff-A
STP-T-eff-A
STP-T-eff-A
STP-T-eff-A
STP-T-eff-A
STP-T-eff-A
STP-T-eff-A
STP-T-eff-A
STP-T-eff-A
STP-T-eff-A
STP-T-eff-A
STP-T-eff-A
STP-T-eff-A
STP-T-eff-A
STP-T-eff-A
STP-T-eff-A
STP-T-eff-A
STP-T-eff-A
STP-T-eff-A
STP-T-eff-A
STP-T-eff-A
STP-T-eff-A
STP-T-eff-A
STP-T-eff-A
STP-T-eff-A
STP-T-eff-A
STP-T-eff-A
STP-T-eff-A
STP-T-eff-A
STP-T-eff-A
STP-T-eff-A
STP-T-eff-A
STP-T-eff-A
STP-T-eff-A
STP-T-eff-A
STP-T-eff-A
STP-T-eff-A
STP-T-eff-A
STP-T-eff-A
STP-T-eff-A
STP-T-eff-A
STP-T-eff-A
STP-T-eff-A
STP-T-eff-A
STP-T-eff-A

STP-T-eff-A
STP-T-eff-A
STP-T-eff-A
STP-T-eff-A
STP-T-eff-A
STP-T-eff-A
STP-T-eff-A
STP-T-eff-A
STP-T-eff-A
STP-T-eff-A
STP-T-eff-A
STP-T-eff-A
STP-T-eff-A
STP-T-eff-A
STP-T-eff-A
STP-T-eff-A
STP-T-eff-A
STP-T-eff-A
STP-T-eff-A
STP-T-eff-A
STP-T-eff-A
STP-T-eff-A
STP-T-eff-A
STP-T-eff-A
STP-T-eff-A
STP-T-eff-A
STP-T-eff-A
STP-T-eff-A
STP-T-eff-A
STP-T-eff-A
STP-T-eff-A
STP-T-eff-A
STP-T-eff-A
STP-T-eff-A
STP-T-eff-A
STP-T-eff-A
STP-T-eff-A
STP-T-eff-A
STP-T-eff-A
STP-T-eff-A
STP-T-eff-A
STP-T-eff-A
STP-T-eff-A
STP-T-eff-A
STP-T-eff-A
STP-T-eff-A

STP-T-eff-A
STP-T-eff-A
STP-T-eff-A
STP-T-eff-A
STP-T-eff-A
STP-T-eff-A
STP-T-eff-A
STP-T-eff-A
STP-T-eff-A
STP-T-eff-A
STP-T-eff-A
STP-T-eff-A
STP-T-eff-A
STP-T-eff-A
STP-T-eff-A
STP-T-eff-A
STP-T-eff-A
STP-T-eff-A
STP-T-eff-A
STP-T-eff-A
STP-T-eff-A
STP-T-eff-A
STP-T-eff-A
STP-T-eff-A
STP-T-eff-A
STP-T-eff-A
STP-T-eff-A
STP-T-eff-A
STP-T-eff-A
STP-T-eff-A
STP-T-eff-A
STP-T-eff-A
STP-T-eff-A
STP-T-eff-A
STP-T-eff-A
STP-T-eff-A
STP-T-eff-A
STP-T-eff-A
STP-T-eff-A
STP-T-eff-A
STP-T-eff-A
STP-T-eff-A
STP-T-eff-A
STP-T-eff-A
STP-T-eff-A
STP-T-eff-A

STP-T-eff-A
STP-T-eff-A
STP-T-eff-A
STP-T-eff-A
STP-T-eff-A
STP-T-eff-A
STP-T-eff-A
STP-T-eff-A
STP-T-eff-A
STP-T-eff-A
STP-T-eff-A
STP-T-eff-A
STP-T-eff-A
STP-T-eff-A
STP-T-eff-A
STP-T-eff-A
STP-T-eff-A
STP-T-eff-A
STP-T-eff-A
STP-T-eff-A
STP-T-eff-A
STP-T-eff-A
STP-T-eff-A
STP-T-eff-A
STP-T-eff-A
STP-T-eff-A
STP-T-eff-A
STP-T-eff-A
STP-T-eff-A
STP-T-eff-A
STP-T-eff-A
STP-T-eff-A
STP-T-eff-A
STP-T-eff-A
STP-T-eff-A
STP-T-eff-A
STP-T-eff-A
STP-T-eff-A
STP-T-eff-A
STP-T-eff-A
STP-T-eff-A
STP-T-eff-A
STP-T-eff-A
STP-T-eff-A
STP-T-eff-A
STP-T-eff-A

STP-T-eff-A
STP-T-eff-A
STP-T-eff-A
STP-T-eff-A
STP-T-eff-A
STP-T-eff-A
STP-T-eff-A
STP-T-eff-A
STP-T-eff-A
STP-T-eff-A
STP-T-eff-A
STP-T-eff-A
STP-T-eff-A
STP-T-eff-A
STP-T-eff-A
STP-T-eff-A
STP-T-eff-A
STP-T-eff-A
STP-T-eff-A
STP-T-eff-A
STP-T-eff-A
STP-T-eff-A
STP-T-eff-A
STP-T-eff-A
STP-T-eff-A
STP-T-eff-A
STP-T-eff-A
STP-T-eff-A
STP-T-eff-A
STP-T-eff-A
STP-T-eff-A
STP-T-eff-A
STP-T-eff-A
STP-T-eff-A
STP-T-eff-A
STP-T-eff-A
STP-T-eff-A
STP-T-eff-A
STP-T-eff-A
STP-T-eff-A
STP-T-eff-A
STP-T-eff-A
STP-T-eff-A
STP-T-eff-A
STP-T-eff-A
STP-T-eff-A

| |
|---|
| STP-T-eff-A |
| STP-T-eff-A |
| STP-T-eff-A |
| STP-T-eff-A |
| STP-T-eff-A |
| STP-T-eff-A |

**Legend:** eff (effluent); F (I

: Detected virulence genes across samples.

| |
|---|
| VFG000881(gb\|WP_000930062) (papB) regulatory protein PapB [P fimbriae (VF0220) - Adherence (VFC0001)] |
| VFG022146(gb\|NP_214805) (mycP3) Type VII secretion system membrane-anchored mycosin MycP3 [ESX-3 ( |
| VFG000008(gb\|WP_010930612) (fimC) outer membrane usher protein precursor [Fimbriae (VF0031) - Adheren |
| VFG012513(gb\|WP_001221122) (iroB) glucosyltransferase IroB [Salmochelin siderophore (VF0230) - Nutrition |
| VFG049133(gb\|WP_004177236) (acrA) acriflavine resistance protein A [AcrAB (VF0568) - Antimicrobial activi |
| VFG048787(gb\|WP_004176417) (sciN/tssJ) type VI secretion system lipoprotein TssJ [T6SS (VF0569) - Effecto |
| VFG000630(gb\|NP_708742) (sigA) serine protease, autotransporter SigA [SigA (VF0119) - Exoenzyme (VFC02 |
| VFG000209(gb\|NP_250405) (exsD) type III secretion system regulatory protein ExsD [TTSS (VF0083) - Effecto |
| VFG001447(gb\|CAA50007) (cnf1) cytotoxic necrotizing factor 1 [CNF-1 (VF0240) - Exotoxin (VFC0235)] [Esc |
| VFG040545(gb\|WP_011190491) (sca1) cell surface antigen Sca1 [Sca1 (VF0498) - Adherence (VFC0001)] [Ric |
| VFG000212(gb\|NP_250408) (pscD) type III secretion system basal body protein PscD [TTSS (VF0083) - Effecto |
| VFG002063(gb\|NP_248767) (icmF1/tssM1) type VI secretion system protein IcmF1 [HSI-I (VF0334) - Effector |
| VFG049095(gb\|WP_002912373) (rfbA) O-antigen export ABC transporter permease RfbA [LPS (VF0561) - Imm |
| VFG049843(gb\|WP_003155066) (exlB) hemolysin activator protein [Exolysin (VF0582) - Effector delivery syste |
| VFG016058(gb\|NP_251114) (pvdL) peptide synthase PvdL [Pyoverdine (VF0094) - Nutritional/Metabolic factor |
| VFG038764(gb\|WP_017409846) (flrA) FleQ protein [Polar flagella (VF0473) - Motility (VFC0204)] [Aeromona |
| VFG000165(gb\|NP_252915) (pchF) pyochelin synthetase PchF [Pyochelin (VF0095) - Nutritional/Metabolic fact |
| VFG048697(gb\|WP_015874721) (clpV/tssH) type VI secretion system ATPase TssH [T6SS (VF0569) - Effector |
| VFG038176(gb\|WP_000777882) (ompA) outer membrane protein OmpA [OmpA (VF0464) - Immune modulatio |
| VFG011515(gb\|WP_004683743) (wbdA) mannosyltransferase [LPS (VF0367) - Immune modulation (VFC0258) |
| VFG000366(gb\|WP_002212761) (ybtQ) yersiniabactin ABC transporter ATP-binding/permease protein YbtQ [Y |
| VFG038859(gb\|WP_005314301) (lfiR) lateral flagellar biosynthetic protein LfiR [Lateral flagella (VF0474) - Mo |
| VFG012597(gb\|WP_000089593) (chuA) Outer membrane heme/hemoglobin receptor ChuA [Chu (VF0234) - Nu |
| VFG048287(gb\|WP_014906962) (fimF) type 1 fimbrial minor component [Type I fimbriae (VF0566) - Adherenc |
| VFG002518(gb\|WP_004200262) (flgL) flagellar hook-associated protein 3 FlgL [Flagella (VF0430) - Motility (V |
| VFG014893(gb\|NP_254009) (algC) phosphomannomutase AlgC [Alginate (VF0091) - Biofilm (VFC0271)] [Pse |
| VFG045626(gb\|WP_014748741) (scm) collagen adhesin protein Scm [Scm (VF0418) - Adherence (VFC0001)] [ |
| VFG041110(gb\|NP_459284) (STM0286) hypothetical protein [SCI (Salmonella centrisome island) (VF0974) - E |
| VFG002417(gb\|WP_001301550) (yagV/ecpE) E. coli common pilus chaperone EcpE [ECP (VF0404) - Adherenc |
| VFG013476(gb\|WP_032828451) (kpsF) KpsF/GutQ family sugar isomerase [LOS (VF0044) - Immune modulati |
| VFG045467(gb\|WP_011204972) (cdpA) cyclic di-GMP phosphodiesterase [CdpA (VF0432) - Regulation (VFC |
| VFG038753(gb\|WP_024944879) (motY) sodium-type flagellar protein MotY [Polar flagella (VF0473) - Motility |
| VFG048307(gb\|WP_041937815) (fimH) type 1 fimbrial adhesin precursor [Type I fimbriae (VF0566) - Adherenc |
| VFG002499(gb\|WP_004197164) (fliI) flagellum-specific ATP synthase FliI [Flagella (VF0430) - Motility (VFC |
| VFG049144(gb\|WP_002892069) (acrB) acriflavine resistance protein B [AcrAB (VF0568) - Antimicrobial activi |
| VFG038886(gb\|WP_005314222) (lafE) LafE, flagellar hook control length protein [Lateral flagella (VF0474) - M |
| VFG038883(gb\|WP_005314227) (lafB) flagellar hook associated protein LafB [Lateral flagella (VF0474) - Motil |
| VFG038820(gb\|WP_161785096) (flgA) flagella basal body P-ring formation protein FlgA [Polar flagella (VF047 |
| VFG007832(gb\|NP_217448) (ppsB) phthiocerol synthesis polyketide synthase type I PpsB [PDIM (VF0309) - In |
| VFG002413(gb\|WP_000389022) (ykgK/ecpR) regulator protein EcpR [ECP (VF0404) - Adherence (VFC0001)] |
| VFG048247(gb\|WP_002916190) (fimA) type 1 major fimbrial subunit precursor [Type I fimbriae (VF0566) - Ad |
| VFG048779(gb\|WP_004190503) (tssG) type VI secretion system baseplate subunit TssG [T6SS (VF0569) - Effe |
| VFG038862(gb\|WP_011898282) (lafK) two-component system lateral flagellar response regulator [Lateral flagel |

VFG002420(gb|WP_000645781) (adsA) Adenosine synthase A [AdsA (VF0422) - Immune modulation (VFC02
VFG049905(gb|WP_000343289) (tssL) Type VI secretion system protein TssL [T6SS (VF0579) - Effector delive
VFG031564(gb|YP_177826) (PE_PGRS30) PE-PGRS family protein PE_PGRS30 [PE/PE-PGRS (VF0316) - Im
VFG038367(gb|WP_011705045) (vgrG1) Type VI secretion system effector VgrG1, ADP-ribosyltransferase [T
VFG034636(gb|WP_000358674) (cfaC) colonisation factor antigen c, usher [Adhesive fimbriae (VF0213) - Adhe
VFG038119(gb|WP_000686134) (ACICU_RS00485) nucleotide sugar dehydrogenase [Capsule (VF0465) - Imm
VFG013070(gb|WP_001626196) (shuV) ATP-binding component of hemin transport system [Shu (VF0256) - N
VFG004773(gb|WP_000842036) (lip) triacylglycerol lipase precursor [Lipase (VF0012) - Exoenzyme (VFC0251
VFG038533(gb|WP_016350152) (flpF) traffic ATPase [Flp type IV pili (VF0476) - Adherence (VFC0001)] [Aer
VFG033511(gb|WP_000803678) (sepQ/escQ) Type III secetion system C ring protein EscQ [TTSS (VF0172) - E
VFG000166(gb|NP_252916) (pchE) dihydroaeruginoic acid synthetase PchE [Pyochelin (VF0095) - Nutritional/
VFG019180(gb|WP_011987710) (bont) botulinum neurotoxin, BoNT [BoNT (VF0375) - Exotoxin (VFC0235)] [
VFG013418(gb|WP_005694260) (gmhA/lpcA) phosphoheptose isomerase [LOS (VF0044) - Immune modulatio
VFG014369(gb|NP_249789) (fleS) two-component sensor [Flagella (VF0273) - Motility (VFC0204)] [Pseudomo
VFG000027(gb|WP_010931506) (brkA) serum resistance protein [Brk (VF0034) - Immune modulation (VFC02
VFG041038(gb|NP_251056) (PA2366) uricase [HSI-3 (VF0944) - Effector delivery system (VFC0086)] [Pseudo
VFG000169(gb|NP_252919) (pchC) pyochelin biosynthetic protein PchC [Pyochelin (VF0095) - Nutritional/Met
VFG002171(gb|WP_011109430) (EF0149) aggregation substance [AS (VF0352) - Adherence (VFC0001)] [Ente
VFG048449(gb|WP_012737621) (fepB) iron-enterobactin transporter periplasmic binding protein [Ent (VF0562)
VFG038383(gb|WP_011705715) (atsH) Type VI secretion system protein [T6SS (VF0480) - Effector delivery sy
VFG038861(gb|WP_005314296) (lfhA) lateral flagellar biosynthesis protein [Lateral flagella (VF0474) - Motilit
VFG001249(gb|NP_249790) (fleR) two-component response regulator [Flagella (VF0273) - Motility (VFC0204)
VFG049916(gb|WP_000103365) (vgrG/tssI) type VI secretion system effector VgrG [T6SS (VF0579) - Effector
VFG038301(gb|WP_016349342) (exeK) general secretion pathway protein K [Exe T2SS (VF0478) - Effector de
VFG037747(gb|WP_001984639) (lpxM) lysophospholipid acyltransferase family protein [LPS (VF0466) - Immu
VFG041118(gb|WP_045606108) (tssA-5) Type VI secretion system protein TssA [T6SS-1 (VF0429) - Effector d
VFG007857(gb|NP_217450) (ppsD) phthiocerol synthesis polyketide synthase type I PpsD [PDIM (VF0309) - In
VFG002076(gb|NP_248780) (clpV1) type VI secretion system AAA+ family ATPase [HSI-I (VF0334) - Effecto
VFG002077(gb|NP_248781) (vgrG1a) type VI secretion system substrate VgrG1 [HSI-I (VF0334) - Effector deli
VFG044334(gb|WP_015874979) (iroN) salmochelin receptor IroN [Sal (VF0563) - Nutritional/Metabolic factor
VFG039366(gb|NP_819695) (cvpA) Coxiella Dot/Icm type IVB secretion system translocated effector, Coxiella
VFG038878(gb|WP_005314254) (flgI) flagellar basal body P-ring protein [Lateral flagella (VF0474) - Motility (
VFG044097(gb|NP_252909) (PA4219) hypothetical protein [Pyochelin (VF0095) - Nutritional/Metabolic factor
VFG048708(gb|WP_041168969) (vgrG/tssI) type VI secretion system tip protein VgrG [T6SS (VF0569) - Effect
VFG049148(gb|WP_041168941) (clbB) colibactin hybrid non-ribosomal peptide synthetase/type I polyketide sy
VFG038792(gb|WP_011706640) (flgF) flagellar basal-body rod protein FlgF [Polar flagella (VF0473) - Motility
VFG000883(gb|WP_001239363) (papH) P pilus termination subunit PapH [P fimbriae (VF0220) - Adherence (V
VFG046610(gb|WP_003020688) (rpe) ribulose-phosphate 3-epimerase [Capsule (VF0543) - Immune modulatio
VFG044322(gb|WP_004190148) (iroE) siderophore esterase IroE [Sal (VF0563) - Nutritional/Metabolic factor (
VFG000362(gb|WP_002212777) (irp1) yersiniabactin biosynthetic protein Irp1 [Yersiniabactin (VF0136) - Nutri
VFG038444(gb|WP_075384315) (tapM) type IV pilus assembly protein PilM [Tap type IV pili (VF0475) - Adhe
VFG000157(gb|NP_249535) (plcH) hemolytic phospholipase C precursor [PLC (VF0092) - Exotoxin (VFC0235
VFG014950(gb|NP_249457) (mucD) serine protease MucD precursor [Alginate (VF0091) - Biofilm (VFC0271)]
VFG001713(gb|WP_001298062) (papX) PapX protein regulates flagellum synthesis to repress motility [P fimbri
VFG001405(gb|NP_217219) (sigA/rpoV) RNA polymerase sigma factor SigA [SigA (VF0257) - Regulation (VF
VFG041043(gb|NP_251061) (PA2371) ClpA/B-type protease [HSI-3 (VF0944) - Effector delivery system (VFC

VFG002178(gb|AAM75254) (cylI) cytolysin immunity protein [Cytolysin (VF0356) - Exotoxin (VFC0235)] [En
VFG038191(gb|WP_002023245) (abaI) N-acyl-L-homoserine lactone synthetase [Quorom sensing (VF0471) - B
VFG000678(gb|AAD32426) (cya) calmodulin sensitive adenylate cyclase, edema factor [Anthrax toxin (VF0142)
VFG001261(gb|NP_250139) (fliR) flagellar biosynthetic protein FliR [Flagella (VF0273) - Motility (VFC0204)]
VFG038371(gb|WP_011705707) (vgrG2) VgrG protein [T6SS (VF0480) - Effector delivery system (VFC0086)]
VFG001222(gb|NP_253731) (pilM) type IV pilus inner membrane platform protein PilM [Type IV pili (VF0082)
VFG000924(gb|WP_001234311) (fepB) ferrienterobactin ABC transporter periplasmic binding protein [Enteroba
VFG000877(gb|WP_001244821) (fimF) FimF protein precursor [Type 1 fimbriae (VF0221) - Adherence (VFC00
VFG000878(gb|WP_000872015) (fimG) FimG protein precursor [Type 1 fimbriae (VF0221) - Adherence (VFC0
VFG050400(gb|WP_000347039.1) (pilU) PilT/PilU family type 4a pilus ATPase [TFP (VF1334) - Adherence (V
VFG048409(gb|WP_086074289) (entA) 2,3-dihydroxybenzoate-2,3-dehydrogenase [Ent (VF0562) - Nutritional/
VFG002126(gb|NP_219593) (cdsV) Type III secretion protein CdsV [TTSS (VF0344) - Effector delivery system
VFG001827(gb|WP_001020414) (senB) enterotoxin [ShET2 (VF0258) - Exotoxin (VFC0235)] [Shigella boydii
VFG002529(gb|WP_011205290) (tsr) methyl-accepting chemotaxis protein I [Flagella (VF0430) - Motility (VFC
VFG012493(gb|WP_000528123) (cnf1) cytotoxic necrotizing factor 1 [CNF-1 (VF0226) - Exotoxin (VFC0235)]
VFG000365(gb|WP_002214412) (ybtP) yersiniabactin ABC transporter ATP-binding/permease protein YbtP [Ye
VFG049159(gb|WP_014907260) (clbM) precolibactin export MATE transporter ClbM [Colibactin (VF0573) - E
VFG015509(gb|NP_252901) (phzC1) phenazine biosynthesis protein PhzC [Pyocyanin (VF0100) - Nutritional/M
VFG050164(gb|WP_000723250) (neuC) UDP-N-acetylglucosamine 2-epimerase [K1 capsule (VF0239) - Invasio
VFG050742(gb|WP_001196426.1) (gspD) general secretion pathway protein D [T2SS (VF1335) - Effector deliv
VFG050163(gb|WP_000197083) (neuE) polysialic acid biosynthesis protein [K1 capsule (VF0239) - Invasion (V
VFG002274(gb|WP_011009584) (plc) phospholipase C [Alpha-toxin (CpPLC) (VF0378) - Exotoxin (VFC0235)
VFG038699(gb|WP_010634617) (cheY) chemotaxis protein CheY [Polar flagella (VF0473) - Motility (VFC020
VFG038687(gb|WP_016350057) (flhF) flagellar biosynthesis protein FlhF [Polar flagella (VF0473) - Motility (V
VFG001254(gb|NP_249795) (fliI) flagellum-specific ATP synthase FliI [Flagella (VF0273) - Motility (VFC020
VFG002187(gb|WP_002370472) (cpsD) glycosyl transferase, group 2 family protein [Capsule (VF0361) - Immu
VFG038369(gb|WP_011705706) (hcp) hemolysin co-regulated protein [T6SS (VF0480) - Effector delivery syste
VFG000919(gb|WP_001095619) (chuW) Putative oxygen independent coproporphyrinogen III oxidase [Chu (VF
VFG049029(gb|WP_014907241) (KP1_RS17220) glycosyltransferase [LPS (VF0561) - Immune modulation (VF
VFG034648(gb|WP_001245988) (cfaD/cfaE) minor pilin and initiator [Adhesive fimbriae (VF0213) - Adherence
VFG002450(gb|WP_004533397) (bipB) Type III secretion system translocator protein BipB [Bsa T3SS (VF042
VFG038281(gb|WP_016349337) (exeF) general secretion pathway protein F [Exe T2SS (VF0478) - Effector deli
VFG002174(gb|WP_002369251) (gelE) coccolysin [Gelatinase (VF0357) - Exoenzyme (VFC0251)] [Enterococc
VFG050161(gb|WP_000579525) (kpsC) Capsule polysaccharide export protein [K1 capsule (VF0239) - Invasion
VFG038417(gb|WP_016349294) (fimD) fimbrial chaperone protein [Type I pili (VF0486) - Adherence (VFC000
VFG048478(gb|WP_004147521) (fepG) iron-enterobactin transporter permease [Ent (VF0562) - Nutritional/Met
VFG002507(gb|WP_004534277) (flgA) flagellar basal body P-ring biosynthesis protein FlgA [Flagella (VF0430
VFG000845(gb|WP_000581721) (toxB) cytotoxin [ToxB (VF0203) - Effector delivery system (VFC0086)] [Esc
VFG014707(gb|NP_250151) (motC) flagellar motor protein [Flagella (VF0273) - Motility (VFC0204)] [Pseudom
VFG038723(gb|WP_011705294) (AHML_RS07540) CobQ/CobB/MinD/ParA family protein [Polar flagella (VF
VFG038468(gb|WP_016351972) (tapU) twitching ATPase [Tap type IV pili (VF0475) - Adherence (VFC0001)]
VFG039216(gb|NP_820634) (icmX) Type IVB secretion system protein IcmX [Dot/Icm T4SS (VF0503) - Effect
VFG049051(gb|WP_014907239) (KP1_RS17230) glycosyltransferase [LPS (VF0561) - Immune modulation (VF
VFG038373(gb|WP_011705708) (atsA) Type VI secretion system protein [T6SS (VF0480) - Effector delivery sy
VFG038836(gb|WP_011707085) (pomA2) chemotaxis protein PomA [Polar flagella (VF0473) - Motility (VFC0
VFG002547(gb|WP_004527661) (wcbS) UDP-3-O-[3-hydroxymyristoyl] N-acetylglucosamine deacetylase [Cap

VFG049161(gb|WP_001029878) (clbO) colibactin polyketide synthase ClbO [Colibactin (VF0573) - Exotoxin (V
VFG048643(gb|WP_014907727) (vipB/tssC) type VI secretion system contractile sheath large subunit VipB [T6S
VFG000872(gb|WP_000044711) (fimE) Type 1 fimbriae Regulatory protein fimE [Type 1 fimbriae (VF0221) - A
VFG000928(gb|WP_000640938) (fepG) iron-enterobactin ABC transporter permease [Enterobactin (VF0228) - N
VFG038882(gb|WP_005314247) (maf-5) modification accessory factor [Lateral flagella (VF0474) - Motility (VF
VFG045313(gb|WP_010946503) (LPG_RS03795) hypothetical protein [LPS (VF0171) - Immune modulation (V
VFG000906(gb|WP_001142375) (hlyA) Hemolysin A [-Hemolysin (VF0225) - Exotoxin (VFC0235)] [Escherich
VFG038715(gb|WP_024943899) (motA) chemotaxis protein MotA [Polar flagella (VF0473) - Motility (VFC020
VFG038772(gb|WP_016351319) (flgK) flagellar hook-associated protein FlgK [Polar flagella (VF0473) - Motilit
VFG001855(gb|WP_197535493) (htpB) Hsp60, 60K heat shock protein HtpB [Hsp60 (VF0159) - Adherence (VF
VFG048229(gb|WP_015875142) (fimB) tyrosine recombinase [Type I fimbriae (VF0566) - Adherence (VFC000
VFG034576(gb|WP_049794100) (aafC) aggregative adherence fimbria II usher protein AafC [AAFs (VF0214) -
VFG041316(gb|WP_010947871) (lpg2160) Dot/Icm type IV secretion system effector [Dot/Icm T4SS secreted ef
VFG041041(gb|NP_251059) (PA2369) hypothetical protein [HSI-3 (VF0944) - Effector delivery system (VFC00
VFG038545(gb|WP_043118798) (flpC) Flp pilus secretin FlpC [Flp type IV pili (VF0476) - Adherence (VFC000
VFG041011(gb|NP_248785) (vgrG1b) type VI secretion system substrate VgrG1b [HSI-I (VF0334) - Effector de
VFG042738(gb|NP_253646) (fimX) protein FimX [Type IV pili (VF0082) - Adherence (VFC0001)] [Pseudomon
VFG049909(gb|WP_001113715) (fha) Type VI secretion system protein Fha [T6SS (VF0579) - Effector delivery
VFG038261(gb|WP_016352107) (exeA) general secretion pathway protein A [Exe T2SS (VF0478) - Effector del
VFG002067(gb|NP_248771) (fha1) type VI secretion system forkhead-associated protein Fha1 [HSI-I (VF0334)
VFG001445(gb|AAA92657) (AAA92657) unknown protein [TraJ (VF0241) - Invasion (VFC0083)] [Escherichia
VFG002053(gb|WP_001250462) (gspJ) general secretion pathway protein J [T2SS (VF0333) - Effector delivery
VFG042979(gb|WP_002379311) (srtC) sortase [Ebp pili (VF0538) - Adherence (VFC0001)] [Enterococcus faeca
VFG001968(gb|YP_002344823) (Cj1440c) sugar transferase [Capsule (VF0323) - Immune modulation (VFC025
VFG038851(gb|WP_016350359) (nueB) pseudaminic acid synthase, NeuB [Polar flagella (VF0473) - Motility (V
VFG000367(gb|WP_001286280) (ybtX) putative signal transducer [Yersiniabactin (VF0136) - Nutritional/Metab
VFG045349(gb|WP_000092542) (fdeC) adhesin FdeC [FdeC (VF0506) - Adherence (VFC0001)] [Escherichia co
VFG038277(gb|WP_016349336) (exeE) general secretory pathway protein E [Exe T2SS (VF0478) - Effector del
VFG007904(gb|NP_217452) (ddrA) daunorubicin ABC transporter ATP-binding protein DrrA [PDIM (VF0309)
VFG038647(gb|WP_016350053) (fliK) flagellar hook-length control protein FliK [Polar flagella (VF0473) - Mot
VFG002179(gb|AAD09858) (esp) Enterococcal surface protein; Esp [Esp (VF0353) - Adherence (VFC0001)] [E
VFG022082(gb|YP_177711) (PPE4) PPE family protein [ESX-3 (VF0521) - Effector delivery system (VFC0086
VFG049154(gb|WP_014907257) (clbH) colibactin non-ribosomal peptide synthetase ClbH [Colibactin (VF0573)
VFG000326(gb|WP_105163169) (licA) choline kinase [LOS (VF0044) - Immune modulation (VFC0258)] [Haem
VFG042977(gb|WP_002379309) (ebpB) endocarditis and biofilm-associated pilus minor subunit EbpB [Ebp pili
VFG038711(gb|WP_016350060) (cheB-2) chemotaxis-specific methylesterase [Polar flagella (VF0473) - Motility
VFG000186(gb|NP_250382) (pscT) type III secretion system protein PscT [TTSS (VF0083) - Effector delivery sy
VFG046955(gb|WP_003021532) (lpcC) glycosyl transferase family protein [LPS (VF0542) - Immune modulation
VFG043623(gb|WP_002916122) (mrkF) type 3 fimbrial minor pilin subunit MrkF [Type 3 fimbriae (VF0567) - F
VFG043018(gb|NP_252039) (PA3349) chemotaxis protein [Flagella (VF0273) - Motility (VFC0204)] [Pseudomo
VFG022035(gb|NP_214797) (eccB3) Type VII secretion system protein EccB3 [ESX-3 (VF0521) - Effector deliv
VFG038289(gb|WP_016349339) (exeH) general secretion pathway protein H [Exe T2SS (VF0478) - Effector del
VFG000927(gb|WP_000096764) (fepE) LPS O-antigen length regulator [Enterobactin (VF0228) - Nutritional/Mo
VFG000175(gb|NP_251786) (xcpY) general secretion pathway protein L [Xcp secretion system (VF0084) - Effec
VFG000160(gb|NP_251087) (pvdE) pyoverdine biosynthesis protein PvdE [Pyoverdine (VF0094) - Nutritional/N
VFG050457(gb|WP_001274990.1) (pilB) type IV-A pilus assembly ATPase PilB [TFP (VF1334) - Adherence (V

VFG049122(gb|WP_012737539) (allC) allantoate amidohydrolase [Allantion utilization (VF0572) - Nutritional/M
VFG002463(gb|WP_011205639) (bsaO) Type III secretion system protein BsaO [Bsa T3SS (VF0428) - Effector
VFG038784(gb|WP_011706638) (flgH) flagellar basal body L-ring protein [Polar flagella (VF0473) - Motility (V
VFG000163(gb|NP_251088) (fpvA) ferripyoverdine receptor FpvA [Pyoverdine (VF0094) - Nutritional/Metabol
VFG016038(gb|NP_251090) (pvdJ) pyoverdine biosynthesis protein PvdJ [Pyoverdine (VF0094) - Nutritional/M
VFG050610(gb|WP_000477147.1) (fimT) GspH/FimT family pseudopilin [TFP (VF1334) - Adherence (VFC000
VFG038873(gb|WP_005314270) (flgC) flagellar basal body rod protein FlgC [Lateral flagella (VF0474) - Motilit
VFG000909(gb|WP_000768217) (focA) F1C major fimbrial subunit precursor [F1C fimbriae (VF0224) - Adhere
VFG000921(gb|WP_000189356) (chuY) ChuY [Chu (VF0227) - Nutritional/Metabolic factor (VFC0272)] [Esch
VFG038525(gb|WP_043118801) (flpH) Flp pilus assembly protein FlpH [Flp type IV pili (VF0476) - Adherence
VFG010749(gb|WP_010945896) (sdhB) Dot/Icm type IV secretion system effector SdhB [Dot/Icm T4SS secrete
VFG002415(gb|WP_001131063) (yagX/ecpC) E. coli common pilus usher EcpC [ECP (VF0404) - Adherence (V
VFG000182(gb|NP_251793) (xcpR) general secretion pathway protein E [Xcp secretion system (VF0084) - Effec
VFG038780(gb|WP_011706637) (flgI) flagellar basal body P-ring protein [Polar flagella (VF0473) - Motility (V
VFG002382(gb|NP_218387) (eccCa1) ESX-1 type VII secretion system FtsK/SpoIIIE family protein EccCa1 [ES
VFG000917(gb|WP_000089583) (chuA) Outer membrane heme/hemoglobin receptor ChuA [Chu (VF0227) - Nu
VFG049904 (clpV/tssH) Type VI secretion system ATPase ClpV/TssH [T6SS (VF0579) - Effector delivery syste
VFG002536(gb|WP_004530401) (fliD) flagellar capping protein FliD [Flagella (VF0430) - Motility (VFC0204)]
VFG049018(gb|WP_002913007) (rcsB) transcriptional regulator RcsB [RcsAB (VF0571) - Regulation (VFC030
VFG049040(gb|WP_074160401) (KP1_RS17225) glycosyltransferase family 4 protein [LPS (VF0561) - Immune
VFG002064(gb|NP_248768) (dotU1) type VI secretion system protein DotU [HSI-I (VF0334) - Effector delivery
VFG033788(gb|WP_001025657) (nleA/espI) Type III secretion system effector NleA [TTSS secreted effectors (V
VFG034908(gb|WP_000978349) (espY3) Type III secretion system effector EspY3 [TTSS secreted effectors (VF
VFG002176(gb|AAM75252) (cylB) ABC-type transporter [Cytolysin (VF0356) - Exotoxin (VFC0235)] [Enteroc
VFG000617(gb|NP_709456) (iucC) aerobactin synthesis protein IucC [Aerobactin (VF0123) - Nutritional/Metab
VFG049116(gb|WP_012737528) (allA) ureidoglycolate hydrolase [Allantion utilization (VF0572) - Nutritional/M
VFG001240(gb|NP_249773) (flgG) flagellar basal-body rod protein FlgG [Flagella (VF0273) - Motility (VFC020
VFG049152(gb|WP_000337350) (clbF) colibactin biosynthesis dehydrogenase ClbF [Colibactin (VF0573) - Exo
VFG048348(gb|WP_004149658) (mrkJ) phosphodiesterase [Type 3 fimbriae (VF0567) - Biofilm (VFC0271)] [K
VFG038413(gb|WP_043118350) (fimC) outer membrane usher protein [Type I pili (VF0486) - Adherence (VFC
VFG049906 (tssK) Type VI secretion system protein TssK [T6SS (VF0579) - Effector delivery system (VFC008
VFG043346(gb|WP_010947489) (fleS/flrB) flagellar sensor histidine kinase FleS [Flagella (VF0157) - Motility (
VFG038760(gb|WP_016351312) (flrB) two-component sensor kinase [Polar flagella (VF0473) - Motility (VFC0
VFG049898(gb|WP_001284199) (hcp1/tssD1) type VI secretion system protein, Hcp family [T6SS (VF0579) - E
VFG001218(gb|NP_253727) (pilQ) type 4 fimbrial biogenesis protein PilQ [Type IV pili (VF0082) - Adherence
VFG001278(gb|WP_000069296) (ebp) cell surface elastin binding protein [EbpS (VF0008) - Adherence (VFC00
VFG002279(gb|WP_003467250) (nagJ) hyaluronidase [Mu-toxin (VF0389) - Exoenzyme (VFC0251)] [Clostridi
VFG046118(gb|WP_011205945) (mhp107) P97 paralog, surface adhesin [P97/P102 paralog family (VF0426) - A
VFG002095(gb|WP_010947666) (ralF) Dot/Icm type IV secretion system effector guanine nucleotide exchange p
VFG049158(gb|WP_001297937) (clbL) colibactin biosynthesis amidase ClbL [Colibactin (VF0573) - Exotoxin (
VFG038425(gb|WP_016349296) (fimF) fimbrial protein [Type I pili (VF0486) - Adherence (VFC0001)] [Aerom
VFG045298(gb|WP_010946487) (hisH) imidazole glycerol phosphate synthase subunit HisH [LPS (VF0171) - In
VFG001814(gb|NP_215678) (narH) nitrate reductase subunit beta [Nitrate reductase (VF0302) - Nutritional/Meta
VFG038832(gb|WP_016351724) (pomB2) flagellar motor protein MotB [Polar flagella (VF0473) - Motility (VF
VFG014984(gb|NP_253136) (algW) AlgW protein [Alginate (VF0091) - Biofilm (VFC0271)] [Pseudomonas ae
VFG043627(gb|WP_000756959) (mrkA) type 3 fimbrial major pilin subunit MrkA [Type 3 fimbriae (VF0567) -

VFG000862(gb|WP_014639426) (pet) Pet serine protease precursor, autotransporter [Pet (VF0217) - Effector del
VFG048297(gb|WP_014906961) (fimG) type 1 fimbrial minor component [Type I fimbriae (VF0566) - Adherenc
VFG041027(gb|NP_250360) (PA1669) hypothetical protein [HSI-2 (VF0943) - Effector delivery system (VFC00
VFG012509(gb|WP_001111200) (iroC) ATP binding cassette transporter [Salmochelin siderophore (VF0230) - N
VFG040665(gb|WP_011190939) (sca4) cell surface antigen Sca4 [Sca4 (VF0502) - Others (VFC0346)] [Ricketts
VFG033484(gb|WP_000953248) (escD) Type III secretion system outer MS ring protein EscD [TTSS (VF0172)
VFG038497(gb|WP_016351992) (tppF) type IV pilus pseudopilin [Tap type IV pili (VF0475) - Adherence (VFC
VFG006217 (opa) outer membrane beta-barrel protein [Opa (VF0076) - Invasion (VFC0083)] [Neisseria meningi
VFG038913 (rtxA) multifunctional autoprocessing repeats-in-toxins (MARTX) toxin [RtxA (VF0482) - Exotoxin
VFG000932(gb|WP_000026784) (entE) 2,3-dihydroxybenzoate-AMP ligase component of enterobactin synthase
VFG001917(gb|YP_002344286) (flgL) flagellar hook-associated protein FlgL [Flagella (VF0114) - Motility (VF
VFG002548(gb|WP_004550042) (wcbR) capsular polysaccharide biosynthesis fatty acid synthase [Capsule I (VF
VFG038381(gb|WP_011705714) (atsG) Type VI secretion system protein [T6SS (VF0480) - Effector delivery sy
VFG048317(gb|WP_014906959) (fimK) transcriptional regulator [Type I fimbriae (VF0566) - Adherence (VFC0
VFG002122(gb|WP_115801843) (nleC) Type III secretion system effector NleC, zinc metalloprotease [TTSS se
VFG001251(gb|NP_249792) (fliF) flagellar M-ring protein FliF [Flagella (VF0273) - Motility (VFC0204)] [Pseu
VFG002480(gb|WP_011205629) (tssH-5/clpV) Clp-type ATPase chaperone protein [T6SS-1 (VF0429) - Effector
VFG038397(gb|WP_011705722) (vasH) sigma-54 dependent transcriptional regulator [T6SS (VF0480) - Effector
VFG000199(gb|NP_250395) (pcrR) type III secretion system regulatory protein PcrR [TTSS (VF0083) - Effector
VFG001966(gb|YP_002344821) (Cj1438c) sugar transferase [Capsule (VF0323) - Immune modulation (VFC025
VFG000429(gb|WP_010989299) (tviD) Vi polysaccharide biosynthesis protein TviD [Vi antigen (VF0101) - Imm
VFG000064(gb|WP_010930831) (bcrD) Type III secretion system LcrD homolog protein BcrD [TTSS (VF0035)
VFG034866(gb|WP_000900547) (espX4) Type III secretion system effector EspX4 [TTSS secreted effectors (VF
VFG038874(gb|WP_005314263) (flgE) flagellar hook protein FlgE [Lateral flagella (VF0474) - Motility (VFC02
VFG000933(gb|WP_001007140) (entB) isochorismatase [Enterobactin (VF0228) - Nutritional/Metabolic factor (
VFG000861(gb|WP_001045649) (pic) Pic serine protease precursor, autotransporter [Pic (VF0218) - Effector del
VFG050159(gb|WP_001331697) (kpsD) Polysialic acid transport protein precursor [K1 capsule (VF0239) - Invas
VFG038607(gb|WP_016349154) (mshJ) MSHA (mannose-sensitive hemagglutinin) pili biogenesis protein MshJ
VFG043518(gb|AAN12397) (acm) collagen adhesin precursor Acm [Acm (VF0419) - Adherence (VFC0001)] [E
VFG049062(gb|WP_015875020) (rfbD) UDP-galactopyranose mutase [LPS (VF0561) - Immune modulation (VF
VFG000025(gb|WP_010929995) (cyaA) bifunctional hemolysin-adenylate cyclase precursor [Cya (VF0028) - Ex
VFG038505(gb|WP_011705643) (tapF) type IV pilus biogenesis/stability protein, pilW-like [Tap type IV pili (VF
VFG000203(gb|NP_250399) (popB) type III secretion system hydrophobic translocator, pore protein PopB [TTS
VFG001262(gb|NP_250140) (flhB) flagellar biosynthetic protein FlhB [Flagella (VF0273) - Motility (VFC0204)
VFG038856(gb|WP_005314315) (lfiN) lateral flagellar motor switch protein, lfiN [Lateral flagella (VF0474) - M
VFG044330(gb|WP_032447695) (ybtU) yersiniabactin biosynthesis oxidoreductase YbtU [Ybt (VF0564) - Nutri
VFG002493(gb|WP_045606430) (fliO) flagellar protein FliO [Flagella (VF0430) - Motility (VFC0204)] [Burkho
VFG012505(gb|WP_000933673) (iroD) esterase [Salmochelin siderophore (VF0230) - Nutritional/Metabolic fact
VFG009561(gb|YP_177877) (mbtI) Isochorismate synthase MbtI [Mycobactin (VF0299) - Nutritional/Metabolic
VFG044331(gb|WP_015874966) (ybtT) type II thioesterase YbtT [Ybt (VF0564) - Nutritional/Metabolic factor (
VFG012591(gb|WP_001017167) (chuS) heme oxygenase ChuS [Chu (VF0234) - Nutritional/Metabolic factor (V
VFG039506 (coxCC8) Coxiella Dot/Icm type IVB secretion system translocated effector [T4SS secreted effector
VFG038391(gb|WP_011705719) (atsL) Type VI secretion system protein [T6SS (VF0480) - Effector delivery sy
VFG000038(gb|WP_003807115) (bplA) probable oxidoreductase [LPS (VF0033) - Immune modulation (VFC02
VFG038561(gb|WP_016349168) (mshQ) MSHA (mannose-sensitive hemagglutinin) pili biogenesis protein Msh
VFG045389 (APH_RS05905*) P44/Msp2 family outer membrane protein [P44/Msp2 family (VF0520) - Immune

VFG038731(gb|WP_016350347) (flaA) flagellin [Polar flagella (VF0473) - Motility (VFC0204)] [Aeromonas hy
VFG038796(gb|WP_016351321) (flgE) flagellar hook protein FlgE [Polar flagella (VF0473) - Motility (VFC020
VFG034897(gb|WP_013009101) (espY1) Type III secretion system effector EspY1 [TTSS secreted effectors (VF
VFG001444(gb|AAG10151) (aslA) putative arylsulfatase [AslA (VF0238) - Invasion (VFC0083)] [Escherichia c
VFG038845(gb|WP_016351249) (nueA) NeuA protein [Polar flagella (VF0473) - Motility (VFC0204)] [Aeromo
VFG041292(gb|WP_010947444) (lpg1717) Dot/Icm type IV secretion system effector [Dot/Icm T4SS secreted ef
VFG000935(gb|WP_001222189) (iroN) salmochelin receptor IroN [Salmochelin siderophore (VF0230) - Nutritic
VFG050167(gb|WP_000038461) (neuD) NeuD protein involved in sialic acid synthesis [K1 capsule (VF0239) - I
VFG045474(gb|AAM75251) (cylM) CylM; cytolysin subunit modifier [Cytolysin (VF0356) - Exotoxin (VFC023
VFG000902(gb|WP_001034089) (sat) Aecreted auto transpoter toxin [Sat (VF0231) - Effector delivery system (V
VFG041013(gb|NP_250203) (hcpA) secreted protein Hcp [HSI-2 (VF0943) - Effector delivery system (VFC008
VFG042975 (srtG2) surface protein cell wall anchor, sortase_B family [PI-2 (VF0530) - Adherence (VFC0001)]
VFG042861(gb|WP_011213223) (pilY1) type IV pilus biogenesis PilY1-related protein [Type IV pili (VF0155) -
VFG000929(gb|WP_001375021) (entD) phosphopantetheinyl transferase component of enterobactin synthase mu
VFG038452(gb|WP_016351631) (tapO) type IV pilus biogenesis protein PilO [Tap type IV pili (VF0475) - Adhe
VFG001289(gb|WP_001056172) (clfA) Clumping factor A, fibrinogen-binding protein [Clumping factor (VF000
VFG038776(gb|WP_016351320) (flgJ) flagellar rod assembly protein/muramidase FlgJ [Polar flagella (VF0473)
VFG014117(gb|NP_251831) (wbpM) nucleotide sugar epimerase/dehydratase WbpM [LPS (VF0085) - Immune
VFG001391(gb|NP_215677) (narG) nitrate reductase subunit alpha [Nitrate reductase (VF0302) - Nutritional/Me
VFG000161(gb|NP_251089) (pvdD) pyoverdine synthetase D [Pyoverdine (VF0094) - Nutritional/Metabolic fac
VFG038619(gb|WP_016349413) (motX) sodium-type flagellar motor component [Polar flagella (VF0473) - Mot
VFG050294(gb|NP_248789) (tse7) type VI secretion exported 7, a DNase and PAAR protein [HSI-I T6SS secret
VFG045788(gb|WP_001189322) (cgsG) curli production assembly/transport protein CsgG [Curli fibers (VF1138
VFG038581(gb|WP_043118279) (mshB) MSHA (mannose-sensitive hemagglutinin) pili minor prepilin protein M
VFG000358(gb|WP_002212885) (fyuA/psn) pesticin/yersiniabactin receptor protein [Yersiniabactin (VF0136) - I
VFG042734(gb|NP_249343) (vfr) cAMP-regulatory protein [Type IV pili (VF0082) - Adherence (VFC0001)] [P
VFG012603(gb|WP_001302516) (chuT) periplasmic heme-binding protein ChuT [Chu (VF0234) - Nutritional/M
VFG001258(gb|NP_250136) (fliO) flagellar protein FliO [Flagella (VF0273) - Motility (VFC0204)] [Pseudomon
VFG000710(gb|AAC38364) (AAC38364) Orf1 [Ler (VF0189) - Regulation (VFC0301)] [Escherichia coli O127:
VFG000167(gb|NP_252917) (pchR) transcriptional regulator PchR [Pyochelin (VF0095) - Nutritional/Metabolic
VFG002164(gb|WP_002395058) (prgB/asc10) aggregation substance PrgB/Asc10 [AS (VF0352) - Adherence (V
VFG050168(gb|WP_000590258) (kpsT) Polysialic acid transport ATP-binding protein [K1 capsule (VF0239) - In
VFG010455(gb|WP_015444381) (pilE/pilA) type IV pilus assembly protein, major pilin [Type IV pili (VF0155)
VFG002060(gb|NP_248764) (ppkA) serine/threonine protein kinase PpkA [HSI-I (VF0334) - Effector delivery sy
VFG002414(gb|WP_000730972) (yagZ/ecpA) E. coli common pilus structural subunit EcpA [ECP (VF0404) - A
VFG048654(gb|WP_004224345) (vasE/tssK) type VI secretion system baseplate subunit TssK [T6SS (VF0569) -
VFG038576(gb|WP_016349164) (mshC) MSHA (mannose-sensitive hemagglutinin) pili minor prepilin protein M
VFG000231 (pilC) pilus assembly protein [Type IV pili (VF0075) - Adherence (VFC0001)] [Neisseria meningiti
VFG049915(gb|WP_001142968) (hcp2/tssD2) Type VI secretion system protein, Hcp family [T6SS (VF0579) - I
VFG041047(gb|NP_252174) (tse3) type VI secretion system effector Tse3, glycoside hydrolase [HSI-I T6SS sec
VFG038317(gb|WP_016352170) (tapD/pilD) prepilin peptidase [Tap type IV pili (VF0475) - Adherence (VFC00
VFG045571(gb|WP_010948124) (LPG_RS12190) Dot/Icm type IV secretion system effector [Dot/Icm T4SS sec
VFG038756(gb|WP_011706626) (flrC) transcriptional activator [Polar flagella (VF0473) - Motility (VFC0204)]
VFG038707(gb|WP_016350059) (cheA-2) chemotaxis protein CheA [Polar flagella (VF0473) - Motility (VFC02
VFG000887(gb|WP_000597713) (papK) P pilus minor subunit PapK [P fimbriae (VF0220) - Adherence (VFC00
VFG034915(gb|WP_000225947) (espY4) Type III secretion system effector EspY4 [TTSS secreted effectors (VF

VFG038639(gb|WP_011705275) (fliI) flagellum-specific ATP synthase [Polar flagella (VF0473) - Motility (VFC
VFG045790(gb|WP_000833288) (cgsE) curli production assembly/transport protein CsgE [Curli fibers (VF1138)
VFG038890(gb|WP_005314213) (lafU) LafU, chemotaxis protein [Lateral flagella (VF0474) - Motility (VFC020
VFG002275(gb|WP_003467630) (pfoA) perfringolysin O [Theta-toxin/PFO (VF0382) - Exotoxin (VFC0235)] [C
VFG016472(gb|WP_011206327) (p216) protein P216, cilium adhesin p97 paralog [P97/P102 paralog family (VF
VFG048620(gb|WP_014907843) (iutA) ferric aerobactin receptor IutA [Aerobactin (VF0565) - Nutritional/Metal
VFG000925(gb|WP_000140634) (fepC) ferrienterobactin ABC transporter ATPase [Enterobactin (VF0228) - Nu
VFG049084(gb|WP_004175264) (rfbB) O-antigen export ABC transporter ATP-binding protein RfbB [LPS (VF
VFG034859(gb|WP_010904544) (espX2) Type III secretion system effector EspX2 [TTSS secreted effectors (VF
VFG038324(gb|WP_005320804) (ascH) Type III secretion system protein AscH, regulator needle assembling [T
VFG040863(gb|WP_011190526) (sca2) cell surface antigen Sca2 [Sca2 (VF0495) - Motility (VFC0204)] [Ricket
VFG049163(gb|WP_000065646) (clbQ) colibactin biosynthesis thioesterase ClbQ [Colibactin (VF0573) - Exoto
VFG046732(gb|WP_003020707) (FTT_RS04140) glycosyltransferase family 2 protein [Capsule (VF0543) - Imm
VFG049917(gb|WP_000508696) (rhs/PAAR) Type VI secretion system protein, PAAR family [T6SS (VF0579) -
VFG000886(gb|WP_000261297) (papJ) P pilus assembly protein PapJ [P fimbriae (VF0220) - Adherence (VFC0
VFG001403(gb|NP_216899) (mbtB) Phenyloxazoline synthase MbtB (phenyloxazoline synthetase) [Mycobactin
VFG038309(gb|WP_016349344) (exeM) general secretion pathway protein M [Exe T2SS (VF0478) - Effector de
VFG041315(gb|WP_010947866) (sidJ) Dot/Icm type IV secretion system effector SidJ [Dot/Icm T4SS secreted e
VFG050169(gb|WP_000124301) (kpsM) Polysialic acid transport protein [K1 capsule (VF0239) - Invasion (VFC
VFG012343(gb|WP_001234182) (sfaH) S fimbriae adhesin SfaH [S fimbriae (VF0222) - Adherence (VFC0001)]
VFG000477(gb|NP_461845) (rpoS) RNA polymerase sigma factor RpoS [RpoS (VF0112) - Regulation (VFC030
VFG049903(gb|WP_001240533) (tssA) Type VI secretion system protein TssA [T6SS (VF0579) - Effector deliv
VFG038892(gb|WP_043118302) (aerA/act) Aerolysin A [Aerolysin (VF0481) - Exotoxin (VFC0235)] [Aeromon
VFG001266(gb|NP_252912) (pchI) ABC transporter ATP-binding protein [Pyochelin (VF0095) - Nutritional/Me
VFG000361(gb|WP_000982866) (ybtU) yersiniabactin biosynthetic protein YbtU [Yersiniabactin (VF0136) - Nu
VFG000446(gb|NP_459541) (fimD) usher protein FimD [Type 1 fimbriae (VF0102) - Adherence (VFC0001)] [S
VFG043016(gb|NP_250155) (PA1464) purine-binding chemotaxis protein [Flagella (VF0273) - Motility (VFC02
VFG000905(gb|WP_001546261) (hlyC) Hemolysin C [-Hemolysin (VF0225) - Exotoxin (VFC0235)] [Escherich
VFG000139(gb|NP_253697) (waaG) B-band O-antigen polymerase [LPS (VF0085) - Immune modulation (VFC
VFG000114(gb|NP_253218) (xcpA/pilD) type 4 prepilin peptidase PilD [Type IV pili (VF0082) - Adherence (VF
VFG012122(gb|WP_010968250) (cpb2) beta2-toxin [Beta2-toxin (VF0417) - Exotoxin (VFC0235)] [Clostridium
VFG050157(gb|WP_001331698) (kpsF) Polysialic acid capsule expression protein [K1 capsule (VF0239) - Invas
VFG038331(gb|WP_005320790) (exsD) Type III secretion system protein ExsD [T3SS (VF0479) - Effector deliv
VFG002056 (gspM) general secretion pathway protein M [T2SS (VF0333) - Effector delivery system (VFC0086)
VFG041042(gb|NP_251060) (PA2370) hypothetical protein [HSI-3 (VF0944) - Effector delivery system (VFC00
VFG012609(gb|WP_000993317) (chuW) Putative oxygen independent coproporphyrinogen III oxidase [Chu (VF
VFG038889(gb|WP_005314215) (lafT) LafT, flagella motor stator protein [Lateral flagella (VF0474) - Motility (
VFG000001(gb|WP_010930610) (fhaB) filamentous hemagglutinin/adhesin [FHA (VF0025) - Adherence (VFC0
VFG038488(gb|WP_016349432) (tapY1) type IV pilus biogenesis protein [Tap type IV pili (VF0475) - Adherenc
VFG049120(gb|WP_012737535) (allB) allantoinase [Allantion utilization (VF0572) - Nutritional/Metabolic facto
VFG041109(gb|NP_459283) (STM0285) hypothetical protein [SCI (Salmonella centrisome island) (VF0974) - E
VFG038032 (ACICU_RS00455*) EpsG family protein [Capsule (VF0465) - Immune modulation (VFC0258)] [A
VFG038297(gb|WP_016349341) (exeJ) general secretion pathway protein J [Exe T2SS (VF0478) - Effector deliv
VFG038800(gb|WP_016351322) (flgD) flagellar hook capping protein [Polar flagella (VF0473) - Motility (VFC
VFG002521(gb|WP_004525693) (flhF) flagellar biosynthesis regulator FlhF [Flagella (VF0430) - Motility (VFC
VFG001224(gb|NP_249087) (pilU) twitching motility protein PilU [Type IV pili (VF0082) - Adherence (VFC00

VFG038572(gb|WP_016349165) (mshD) MSHA (mannose-sensitive hemagglutinin) pili minor prepilin protein M
VFG038816(gb|WP_011706646) (cheV) CheV [Polar flagella (VF0473) - Motility (VFC0204)] [Aeromonas hyd
VFG050166(gb|WP_000066003) (neuB) N-acylneuraminate-9-phosphate synthase [K1 capsule (VF0239) - Invas
VFG013368(gb|WP_005693225) (rffG) dTDP-glucose 46-dehydratase [LOS (VF0044) - Immune modulation (V
VFG046206(gb|NP_820798) (CBU_1819) Coxiella Dot/Icm type IVB secretion system translocated effector [T4
VFG045791(gb|WP_000481500) (cgsD) transcriptional regulator CsgD [Curli fibers (VF1138) - Adherence (VF
VFG038365(gb|WP_011705044) (hcp1) HcpA-like protein [T6SS (VF0480) - Effector delivery system (VFC008
VFG049153(gb|WP_000159201) (clbG) colibactin biosynthesis acyltransferase ClbG [Colibactin (VF0573) - Ex
VFG013065(gb|WP_024259428) (shuT) periplasmic binding protein ShuT [Shu (VF0256) - Nutritional/Metaboli
VFG000127(gb|NP_252235) (algG) outer membrane protein AlgE [Alginate (VF0091) - Biofilm (VFC0271)] [P
VFG050213(gb|WP_011861889) (CD3246) Cys-Gln thioester bond-forming surface protein [CD3246 (VF0599)
VFG010850 (rtxA) structural toxin protein RtxA [RtxA (VF0259) - Exotoxin (VFC0235)] [Legionella pneumoph
VFG048468(gb|WP_004176917) (fepD) iron-enterobactin transporter membrane protein [Ent (VF0562) - Nutriti
VFG049155(gb|WP_000829570) (clbI) colibactin polyketide synthase ClbI [Colibactin (VF0573) - Exotoxin (VF
VFG001822(gb|NP_216897) (mbtD) Polyketide synthetase MbtD (polyketide synthase) [Mycobactin (VF0299) -
VFG042390(gb|WP_011478152) (sfaX) regulatory protein SfaX [S fimbriae (VF0222) - Adherence (VFC0001)]
VFG013064(gb|WP_000089574) (shuA) outer membrane heme/hemoglobin receptor ShuA [Shu (VF0256) - Nut
VFG038387(gb|WP_011705717) (atsJ) Type VI secretion system protein [T6SS (VF0480) - Effector delivery sys
VFG001355(gb|WP_000698569) (cbpD) choline binding protein D [Autolysin (VF0143) - Exoenzyme (VFC025
VFG015515(gb|NP_252903) (phzE1) phenazine biosynthesis protein PhzE [Pyocyanin (VF0100) - Nutritional/M
VFG038273(gb|WP_016349335) (exeD) general secretion pathway protein D [Exe T2SS (VF0478) - Effector del
VFG038432(gb|WP_016352168) (tapB) type IV pilus assembly ATPase, PilB-like [Tap type IV pili (VF0475) -
VFG000184(gb|NP_251795) (xcpQ) general secretion pathway protein D [Xcp secretion system (VF0084) - Effe
VFG045635(gb|WP_002296629) (sgrA) cell wall anchored protein SgrA [SgrA (VF0540) - Adherence (VFC000
VFG048338(gb|WP_002916079) (mrkI) LuxR family regulatory protein [Type 3 fimbriae (VF0567) - Biofilm (V
VFG034815(gb|WP_000671689) (espR1) Type III secretion system effector espR1 [TTSS secreted effectors (VF
VFG038144(gb|WP_001062908) (galE) UDP-glucose 4-epimerase GalE [Capsule (VF0465) - Immune modulatio
VFG048419(gb|WP_004210873) (entB) 2,3-dihydro-2,3-dihydroxybenzoate synthetase, isochroismatase [Ent (V
VFG048518(gb|WP_012737617) (fepA) outer membrane receptor FepA [Ent (VF0562) - Nutritional/Metabolic f
VFG049835(gb|WP_012076956) (exlA) hemolysin [Exolysin (VF0582) - Effector delivery system (VFC0086)] [
VFG000112(gb|NP_253216) (pilB) type 4 fimbrial biogenesis protein PilB [Type IV pili (VF0082) - Adherence (
VFG002196(gb|WP_011109578) (EF3023) polysaccharide lyase, family 8 [Hyaluronidase (VF0359) - Exoenzym
VFG038695(gb|WP_017409955) (fliA) flagellar biosynthesis sigma factor [Polar flagella (VF0473) - Motility (V
VFG038493(gb|WP_016349428) (tppE) type IV pilus pseudopilin [Tap type IV pili (VF0475) - Adherence (VFC
VFG000347(gb|WP_010869038) (hgpC) hemoglobin-binding protein [Hgp (VF0270) - Nutritional/Metabolic fac
VFG002543(gb|WP_011205386) (bspR4) LuxR family transcriptional regulator [Quorum-sensing (VF0433) - Bi
VFG047478(gb|WP_003020972) (bioC) biotin synthesis protein BioC [Biotin synthesis (VF0552) - Nutritional/M
VFG038590(gb|WP_043118275) (mshE) ATPase protein MshE, GspE-like [MSHA type IV pili (VF0477) - Adh
VFG001265(gb|NP_250145) (fleN) flagellar synthesis regulator FleN [Flagella (VF0273) - Motility (VFC0204)]
VFG001381(gb|YP_177728) (icl) Isocitrate lyase Icl (isocitrase) (isocitratase) [Isocitrate lyase (VF0253) - Others
VFG038675(gb|WP_011705283) (fliR) flagellar biosynthesis protein FliR [Polar flagella (VF0473) - Motility (V
VFG034922(gb|WP_001303732) (espY5) hypothetical protein [TTSS secreted effectors (VF1110) - Effector deli
VFG050300(gb|NP_250641) (fapF) outer membrane secretin FapF [Fap (VF1191) - Adherence (VFC0001)] [Pse
VFG047777(gb|WP_003022126) (pdpB) Type VI secretion system TssM homolog [T6SS (VF0544) - Effector de
VFG001817(gb|NP_216900) (mbtA) Bifunctional enzyme MbtA: salicyl-AMP ligase (SAL-AMP ligase) + salicy
VFG038401(gb|WP_011705724) (atsQ) Type VI secretion system protein [T6SS (VF0480) - Effector delivery sy

VFG002190(gb|WP_002362225) (cpsA/uppS) undecaprenyl diphosphate synthase [Capsule (VF0361) - Immune
VFG006938(gb|WP_000252812) (mshK) MSHA biogenesis protein MshK [MSHA pili (VF0515) - Adherence (V
VFG048819(gb|WP_014907234) (KP1_RS17280) mannose-1-phosphate guanylyltransferase/mannose-6-phospha
VFG003561(gb|WP_005173823) (yst1E) type II secretion system ATPase GspE [Yst1 T2SS (VF0635) - Effector
VFG046169 (CBU_0820*) Coxiella Dot/Icm type IVB secretion system translocated effector [T4SS secreted effe
VFG038305(gb|WP_016349343) (exeL) general secretion pathway protein L [Exe T2SS (VF0478) - Effector deli
VFG000934(gb|WP_000348393) (entA) 2,3-dihydro-2,3-dihydroxybenzoate dehydrogenase EntA [Enterobactin (
VFG007866(gb|NP_217451) (ppsE) phthiocerol synthesis polyketide synthase type I PpsE [PDIM (VF0309) - Im
VFG038332(gb|WP_192914500) (exsA) Type III secretion system transcriptional regulator [T3SS (VF0479) - Ef
VFG013066 (shuW) heme anaerobic degradation radical SAM methyltransferase ChuW/HutW [Shu (VF0256) - I
VFG000155(gb|NP_252168) (rhlB) rhamnosyltransferase chain B [Rhamnolipid (VF0089) - Immune modulation
VFG002182(gb|WP_002376666) (cpsI) UDP-galactopyranose mutase [Capsule (VF0361) - Immune modulation (
VFG001231(gb|NP_249104) (chpA) still frameshift probable component of chemotactic signal transduction syste
VFG038812(gb|WP_011706645) (cheR-3) chemotaxis protein CheR [Polar flagella (VF0473) - Motility (VFC02
VFG000937(gb|WP_000750139) (iucD) L-lysine 6-monooxygenase IucD [Aerobactin (VF0229) - Nutritional/Me
VFG001362(gb|WP_000088129) (zmpC) immunoglobulin A1 protease [IgA1 protease (VF0147) - Immune modu
VFG038269(gb|WP_075384415) (exeC) general secretion pathway protein C [Exe T2SS (VF0478) - Effector del
VFG002482(gb|WP_011205630) (tagAB-5) Type VI secretion system associated protein TagAB-5 [T6SS-1 (VF
VFG048267(gb|WP_015875145) (fimC) periplasmic chaperone [Type I fimbriae (VF0566) - Adherence (VFC00
VFG002229(gb|WP_002963676) (wbkB) perosamine synthetase WbkB [LPS (VF0367) - Immune modulation (V
VFG012033(gb|YP_002344722) (pseD/maf2) motility accessory factor PseD [Flagella (VF0114) - Motility (VFC
VFG001443(gb|AAF37887) (ompA) outer membrane protein A [OmpA (VF0236) - Invasion (VFC0083)] [Esche
VFG000894(gb|WP_001239363) (papH) P pilus termination subunit PapH [P fimbriae (VF0220) - Adherence (V
VFG046465(gb|WP_003028672) (tufA) elongation factor Tu [EF-Tu (VF0460) - Adherence (VFC0001)] [Franci
VFG002189(gb|WP_002359680) (cpsB/cdsA) phosphatidate cytidylyltransferase [Capsule (VF0361) - Immune m
VFG019178(gb|WP_012291519) (bont) botulinum neurotoxin, BoNT [BoNT (VF0375) - Exotoxin (VFC0235)] [
VFG000133(gb|NP_252241) (algA) phosphomannose isomerase / guanosine 5'-diphospho-D-mannose pyrophosp
VFG048980(gb|WP_041937790) (KP1_RS17345) capsule assembly Wzi family protein [Capsule (VF0560) - Im
VFG048508(gb|WP_012737618) (fes) enterobactin/ferric enterobactin esterase [Ent (VF0562) - Nutritional/Meta
VFG000131(gb|NP_252239) (algJ) alginate o-acetyltransferase AlgJ [Alginate (VF0091) - Biofilm (VFC0271)] [
VFG000922(gb|WP_000910402) (chuU) heme permease protein ChuU [Chu (VF0227) - Nutritional/Metabolic fa
VFG022019(gb|NP_214796) (eccA3) Type VII secretion system protein EccA3 [ESX-3 (VF0521) - Effector deli
VFG038663(gb|WP_005298795) (fliO) flagellar biosynthesis protein, FliO [Polar flagella (VF0473) - Motility (V
VFG000895(gb|WP_000654937) (papC) usher protein PapC [P fimbriae (VF0220) - Adherence (VFC0001)] [Es
VFG049156(gb|WP_077265119) (clbJ) colibactin non-ribosomal peptide synthetase ClbJ [Colibactin (VF0573) -
VFG000908(gb|WP_000860472) (hlyD) Hemolysin D [-Hemolysin (VF0225) - Exotoxin (VFC0235)] [Escherich
VFG049160(gb|WP_001327259) (clbN) colibactin non-ribosomal peptide synthetase ClbN [Colibactin (VF0573)
VFG038285(gb|WP_005308483) (exeG) general secretion pathway protein G [Exe T2SS (VF0478) - Effector del
VFG001815(gb|NP_215679) (narJ) nitrate reductase subunit delta [Nitrate reductase (VF0302) - Nutritional/Meta
VFG047401(gb|WP_011242229) (feoB) Fe(2+) transporter permease subunit FeoB [FupA (VF0550) - Nutritiona
VFG002070(gb|NP_248774) (hsiC1/vipB/tssC) type VI secretion system tubule-forming protein VipB [HSI-I (V
VFG044336(gb|WP_050767835) (iroC) ABC transporter [Sal (VF0563) - Nutritional/Metabolic factor (VFC027
VFG048528(gb|WP_004183335) (entD) enterochelin synthetase component D [Ent (VF0562) - Nutritional/Meta
VFG000920(gb|WP_000020034) (chuX) putative heme-binding protein ChuX [Chu (VF0227) - Nutritional/Meta
VFG000936(gb|WP_016265865) (iutA) ferric aerobactin receptor precusor IutA [Aerobactin (VF0229) - Nutritio
VFG013236(gb|WP_005693864) (kdsB) 3-deoxy-manno-octulosonate cytidylyltransferase [LOS (VF0044) - Imm

VFG000618(gb|NP_709457) (iucD) aerobactin synthesis protein IucC, lysine 6-monooxygenase [Aerobactin (VF
VFG007182(gb|WP_005464333) (vopQ) type III secretion system effector VopQ [T3SS1 secreted effectors (VF
VFG038703(gb|WP_011705289) (cheZ) chemotaxis protein CheZ [Polar flagella (VF0473) - Motility (VFC0204
VFG002059(gb|NP_248763) (tagT) type six secretion associated protein TagT, ATP-binding component of ABC
VFG049912 (tssC) Type VI secretion system protein TssC [T6SS (VF0579) - Effector delivery system (VFC008
VFG038564(gb|WP_016349167) (mshP) hypothetical protein [MSHA type IV pili (VF0477) - Adherence (VFC0
VFG001282(gb|WP_000841389) (fnbB) fibronectin-binding protein B [FnBPs (VF0010) - Adherence (VFC0001
VFG034679(gb|WP_000074207) (ibeB) Cu(+)/Ag(+) efflux RND transporter outer membrane channel CusC [Ib
VFG041010(gb|NP_248760) (tagQ) type VI secretiona ssociated protein TagQ, outer membrane lipoprotein [HSI
VFG002434(gb|WP_011205650) (pilN) type IV pilus inner membrane platform protein PilN [Type IV pili (VF04
VFG049149(gb|WP_041169023) (clbC) colibactin polyketide synthase ClbC [Colibactin (VF0573) - Exotoxin (V
VFG000940(gb|WP_001299620) (iucA) aerobactin siderophore biosynthesis protein IucD [Aerobactin (VF0229)
VFG000359(gb|WP_001088826) (ybtE) yersiniabactin siderophore biosynthetic protein [Yersiniabactin (VF0136
VFG038615(gb|WP_016349152) (mshI) MSHA (mannose-sensitive hemagglutinin) pili biogenesis protein MshI,
VFG050160(gb|WP_000030755) (kpsU) 3-deoxy-manno-octulosonate cytidylyltransferase [K1 capsule (VF0239
VFG038623(gb|WP_011705271) (fliE) flagellar hook-basal body complex protein FliE [Polar flagella (VF0473)
VFG041098(gb|NP_459271) (STM0274) EvpB family type VI secretion protein [SCI (Salmonella centrisome isl
VFG011429(gb|WP_002963985) (acpXL) acyl carrier protein [LPS (VF0367) - Immune modulation (VFC0258)]
VFG002285(gb|WP_011590492) (nanH) sialidase [Sialidase (VF0391) - Exoenzyme (VFC0251)] [Clostridium p
VFG038347(gb|WP_005320757) (aopN) AopN, secretion control of translocators and immune suppressor [T3SS
VFG038436(gb|WP_016352169) (tapC) type IV fimbrial assembly protein, PilC-like [Tap type IV pili (VF0475)
VFG002416(gb|WP_001265657) (yagW/ecpD) polymerized tip adhesin of ECP fibers [ECP (VF0404) - Adheren
VFG001267(gb|NP_252913) (pchH) ABC transporter ATP-binding protein [Pyochelin (VF0095) - Nutritional/M
VFG012095(gb|WP_003435012) (groEL) chaperonin GroEL [GroEL (VF0594) - Adherence (VFC0001)] [Clostr
VFG043648(gb|NP_252312) (rpoS) RNA polymerase sigma factor RpoS [Type IV pili (VF0082) - Adherence (V
VFG012615(gb|WP_000020049) (chuX) putative heme-binding protein ChuX [Chu (VF0234) - Nutritional/Meta
VFG043625(gb|WP_000813718) (mrkC) fimbrial biogenesis outer membrane usher protein mrkC precursor [Typ
VFG010491(gb|WP_010947066) (LPG_RS06695) L,D-transpeptidase [Enh loci (VF0163) - Others (VFC0346)]
VFG002051(gb|WP_001115126) (gspH) general secretion pathway protein H [T2SS (VF0333) - Effector deliver
VFG041032(gb|NP_251050) (PA2360) hypothetical protein [HSI-3 (VF0944) - Effector delivery system (VFC00
VFG001210(gb|NP_253244) (pilY1) type 4 fimbrial biogenesis protein PilY1 [Type IV pili (VF0082) - Adherenc
VFG038155(gb|WP_000209960) (ACICU_RS00500) phosphomannomutase/phosphoglucomutase [Capsule (VF
VFG000142(gb|NP_253698) (waaC) 3-deoxy-D-manno-octulosonic-acid (KDO) transferase [LPS (VF0085) - Im
VFG002481(gb|WP_009940775) (vgrG-5) Type VI secretion system protein VgrG5 [T6SS-1 (VF0429) - Effecto
VFG038472(gb|WP_011706550) (tapW) twitching ATPase [Tap type IV pili (VF0475) - Adherence (VFC0001)]
VFG000844(gb|WP_001034100) (espP) autotransporter, serine protease [EspP (VF0208) - Effector delivery syst
VFG000036(gb|WP_010929614) (bplC) lipopolysaccharide biosynthesis protein [LPS (VF0033) - Immune modu
VFG000164(gb|NP_252911) (fptA) Fe(III)-pyochelin receptor precursor [Pyochelin (VF0095) - Nutritional/Meta
VFG000190(gb|NP_250386) (pscP) type III secretion system protein PscP [TTSS (VF0083) - Effector delivery s
VFG001228(gb|NP_249102) (pilJ) twitching motility protein PilJ [Type IV pili (VF0082) - Adherence (VFC000
VFG038364(gb|WP_005320615) (aexT) Type III secretion system effector AexT, bifunctional toxin with GTPas
VFG002270(gb|WP_011180104) (badA/vomp/brp) Surface protein/Bartonella adhesin BadA/VompA/VompC/V
VFG000120(gb|NP_253949) (algZ) sigma factor AlgU [Alginate (VF0091) - Biofilm (VFC0271)] [Pseudomona
VFG001816(gb|NP_215680) (narI) nitrate reductase subunit gamma [Nitrate reductase (VF0302) - Nutritional/M
VFG038840(gb|WP_005300916) (flmH) short chain dehydrogenase/reductase family oxidoreductase [Polar flage
VFG006902(gb|WP_000957200) (pilB) type IV-A pilus assembly ATPase PilB [ChiRP (VF0609) - Adherence (

VFG038480(gb|WP_016349429) (tppB) type IV pilus pseudopilin [Tap type IV pili (VF0475) - Adherence (VFC
VFG049911(gb|WP_000393837) (tssF) Type VI secretion system protein TssF [T6SS (VF0579) - Effector delive
VFG002478(gb|WP_004551874) (tssF-5) Type VI secretion system protein TssF [T6SS-1 (VF0429) - Effector de
VFG000876(gb|WP_000120946) (fimD) Outer membrane usher protein fimD precursor [Type 1 fimbriae (VF022
VFG000799(gb|WP_000935759) (espD) Type III secretion system translocator protein EspD, pore protein [TTSS
VFG014304(gb|NP_249784) (fleI/flaG) flagellar protein FlaG [Flagella (VF0273) - Motility (VFC0204)] [Pseudo
VFG002277(gb|WP_011009663) (nagH) hyaluronidase [Mu-toxin (VF0389) - Exoenzyme (VFC0251)] [Clostrid
VFG048632(gb|WP_004176434) (vipA/tssB) type VI secretion system contractile sheath small subunit VipA [T6
VFG049908(gb|WP_001080148) (tssJ) Type VI secretion system protein TssJ [T6SS (VF0579) - Effector deliver
VFG002054(gb|WP_000633233) (gspK) general secretion pathway protein K [T2SS (VF0333) - Effector delivery
VFG038593(gb|WP_016349158) (mshN) Tetratricopeptide repeat protein, PilF-like [MSHA type IV pili (VF047
VFG038867(gb|WP_005314284) (lfiI) lateral flagellar FliI-like assembly ATPase [Lateral flagella (VF0474) - M
VFG001225(gb|NP_249099) (pilG) twitching motility protein PilG [Type IV pili (VF0082) - Adherence (VFC00
VFG000739(gb|AAC38392) (eae) intimin [Intimin (VF0177) - Effector delivery system (VFC0086)] [Escherichia
VFG048609(gb|WP_004213923) (iucD) lysine 6-monooxygenase IucD [Aerobactin (VF0565) - Nutritional/Meta
VFG002116(gb|WP_010947864) (sdeC/laiC) Dot/Icm type IV secretion system effector SdeC [Dot/Icm T4SS se
VFG034612(gb|WP_000225871) (cfaA) colonisation factor antigen a, chaperone [Adhesive fimbriae (VF0213) -
VFG001950(gb|YP_002344805) (Cj1422c) sugar transferase [Capsule (VF0323) - Immune modulation (VFC025
VFG042976(gb|WP_154081516) (ebpA) endocarditis and biofilm-associated pilus tip protein EbpA [Ebp pili (V
VFG044083(gb|NP_250948) (ptxR) transcriptional regulator PtxR [Pyoverdine (VF0094) - Nutritional/Metabolic
VFG038521(gb|WP_016350155) (flpI) Flp pilus assembly protein FlpI [Flp type IV pili (VF0476) - Adherence (
VFG002055(gb|WP_000097229) (gspL) general secretion pathway protein L [T2SS (VF0333) - Effector delivery
VFG048429(gb|WP_004183353) (entE) enterobactin synthase subunit E [Ent (VF0562) - Nutritional/Metabolic f
VFG038320(gb|WP_005320812) (ascL) Type III secretion system C ring protein AscL [T3SS (VF0479) - Effecto
VFG000912(gb|WP_001443207) (focD) F1C fimbrial usher [F1C fimbriae (VF0224) - Adherence (VFC0001)] [
VFG038378(gb|WP_011705712) (vipA) Type VI secretion system contractile sheath small subunit TssB/VipA [T
VFG000478(gb|NP_459678) (fur) ferric iron uptake transcriptional regulator [Fur (VF0113) - Regulation (VFC0
VFG041348(gb|WP_010948513) (ceg34) Dot/Icm type IV secretion system effector Ceg34 [Dot/Icm T4SS secre
VFG002412(gb|WP_000716386) (yagY/ecpB) E. coli common pilus chaperone EcpB [ECP (VF0404) - Adherenc
VFG038568(gb|WP_016349166) (mshO) MSHA (mannose-sensitive hemagglutinin) pili biogenesis protein Msh
VFG001393(gb|NP_215861) (mbtM) Probable fatty acyl-AMP ligase MbtM [Mycobactin (VF0299) - Nutritional
VFG049007(gb|WP_002911561) (rcsA) transcriptional activator for ctr capsule biosynthesis [RcsAB (VF0571) -
VFG041016(gb|NP_250349) (PA1658) hypothetical protein [HSI-2 (VF0943) - Effector delivery system (VFC00
VFG038863(gb|WP_005314292) (lfiE) lateral flagellar hook basal body protein, LfiE [Lateral flagella (VF0474)
VFG042736(gb|NP_253152) (rpoN) RNA polymerase factor sigma-54 [Type IV pili (VF0082) - Adherence (VF
VFG000177(gb|NP_251788) (xcpW) general secretion pathway protein J [Xcp secretion system (VF0084) - Effe
VFG002513(gb|WP_004197257) (flgG) flagellar basal body rod protein FlgG [Flagella (VF0430) - Motility (VF
VFG002181(gb|WP_002387127) (cpsJ) ABC transporter, ATP-binding protein [Capsule (VF0361) - Immune mo
VFG050162(gb|WP_001161829) (kpsS) Capsule polysaccharide export protein [K1 capsule (VF0239) - Invasion
VFG048762(gb|WP_015874732) (impA/tssA) type VI secretion system protein TssA [T6SS (VF0569) - Effector
VFG038804(gb|WP_011706643) (flgC) flagellar basal body rod protein FlgC [Polar flagella (VF0473) - Motility
VFG012162(gb|WP_011590241) (nanJ) exo-alpha-sialidase [Sialidase (VF0391) - Exoenzyme (VFC0251)] [Clo
VFG001269(gb|WP_010929996) (cyaB) cyclolysin secretion ATP-binding protein [Cya (VF0028) - Exotoxin (V
VFG049899(gb|WP_001087735) (tssA) Type VI secretion system protein TssA [T6SS (VF0579) - Effector deliv
VFG000047(gb|WP_003820063) (bscO) Bordetella Bsc type III secretion system protein BscO [TTSS (VF0035)
VFG050158(gb|WP_000905916) (kpsE) Capsule polysaccharide export inner-membrane protein [K1 capsule (V

VFG049157(gb|WP_014907259) (clbK) colibactin hybrid non-ribosomal peptide synthetase/type I polyketide syn
VFG015950(gb|NP_251075) (pvdQ) 3-oxo-C12-homoserine lactone acylase PvdQ [Pyoverdine (VF0094) - Nutri
VFG038659(gb|WP_011705280) (fliN) flagellar motor switch protein FliN [Polar flagella (VF0473) - Motility (V
VFG043014(gb|NP_250149) (PA1458) two-component sensor [Flagella (VF0273) - Motility (VFC0204)] [Pseud
VFG002559(gb|WP_004194348) (wcbH) glycosyl transferase [Capsule I (VF0436) - Immune modulation (VFC0
VFG002062(gb|NP_248766) (tagF/pppB) Pseudomonas protein phosphatase PppB [HSI-I (VF0334) - Effector de
VFG038848(gb|WP_016350360) (flmD) FlmD protein [Polar flagella (VF0473) - Motility (VFC0204)] [Aeromo
VFG014109(gb|NP_251839) (wbpH) glycosyltransferase WbpH [LPS (VF0085) - Immune modulation (VFC025
VFG038655(gb|WP_011705279) (fliM) flagellar motor switch protein FliM [Polar flagella (VF0473) - Motility (
VFG038587(gb|WP_016349160) (mshG) MSHA (mannose-sensitive hemagglutinin) pili biogenesis protein Msh
VFG050414(gb|WP_002027454.1) (pilF) type IV pilus biogenesis/stability protein PilW [TFP (VF1334) - Adher
VFG048962(gb|WP_015875029) (KP1_RS17335) protein-tyrosine-phosphatase [Capsule (VF0560) - Immune m
VFG038456(gb|WP_016351630) (tapP) type IV pilus biogenesis protein PilP [Tap type IV pili (VF0475) - Adher
VFG034849(gb|WP_000129338) (espX1) Type III secretion system effector EspX1 [TTSS secreted effectors (V
VFG012621(gb|WP_000189352) (chuY) ChuY [Chu (VF0234) - Nutritional/Metabolic factor (VFC0272)] [Esch
VFG034942(gb|WP_000950813) (nleB2) Type III secretion system effector NleB2 [TTSS secreted effectors (VF
VFG002457(gb|WP_011205638) (bsaU) Type III secretion system protein BsaU [Bsa T3SS (VF0428) - Effector
VFG038355(gb|WP_005320739) (ascU) Type III secretion system inner membrane export apparatus protein Asc
VFG050874(gb|WP_001989189.1) (gspL) general secretion pathway protein L [T2SS (VF1335) - Effector delive
VFG000839(gb|WP_001449026) (paa) outer membrane adhesin Paa [Paa (VF0194) - Adherence (VFC0001)] [E
VFG002522(gb|WP_004198639) (flhA) flagellar biosynthesis protein FlhA [Flagella (VF0430) - Motility (VFC0
VFG039311(gb|NP_819427) (cetCb2) Coxiella Dot/Icm type IVB secretion system translocated effector [T4SS s
VFG014997(gb|NP_252722) (mucE) small envelope protein MucE [Alginate (VF0091) - Biofilm (VFC0271)] [P
VFG002492(gb|WP_004184995) (fliN) flagellar motor switch protein FliN [Flagella (VF0430) - Motility (VFC0
VFG038529(gb|WP_016350153) (flpG) Flp pilus assembly protein FlpG [Flp type IV pili (VF0476) - Adherence
VFG001271(gb|WP_010929998) (cyaE) cyclolysin secretion protein [Cya (VF0028) - Exotoxin (VFC0235)] [Bo
VFG001263(gb|NP_250143) (flhA) flagellar biosynthesis protein FlhA [Flagella (VF0273) - Motility (VFC0204)
VFG038768(gb|WP_016351318) (flgL) flagellar hook-associated protein FlgL [Polar flagella (VF0473) - Motilit
VFG038352(gb|WP_005320745) (ascR) Type III secretion system inner membrane export apparatus protein Asc
VFG042382(gb|WP_000260524) (etpB) two-partner secretion transporter EtpB [EtpA (VF0437) - Adherence (V
VFG001241(gb|NP_249774) (flgH) flagellar L-ring protein precursor FlgH [Flagella (VF0273) - Motility (VFC0
VFG012160(gb|WP_011590331) (nanI) exo-alpha-sialidase [Sialidase (VF0391) - Exoenzyme (VFC0251)] [Clos
VFG002180(gb|WP_002362627) (cpsK) ABC transporter, permease protein [Capsule (VF0361) - Immune modu
VFG034721(gb|WP_000556298) (ibeC) phosphoethanolamine transferase CptA [Ibes (VF0237) - Invasion (VFC
VFG002219(gb|WP_011005413) (cgs) cyclic beta 1-2 glucan synthetase [CG (VF0366) - Immune modulation (V
VFG000282(gb|WP_000488766) (cag3) type IV secretion system protein Cag3, component outer membrane sub
VFG012333(gb|WP_000703306) (sfaD) S fimbriae minor subunit SfaD [S fimbriae (VF0222) - Adherence (VFC
VFG048237(gb|WP_002916189) (fimE) tyrosine recombinase [Type I fimbriae (VF0566) - Adherence (VFC000
VFG022114(gb|NP_214803) (espG3) Type VII secretion system protein EspG3 [ESX-3 (VF0521) - Effector deli
VFG051042(gb|WP_000653195.1) (hcp/tssD) type VI secretion system tube protein Hcp [T6SS (VF1337) - Effe
VFG048771(gb|WP_004190505) (tssF) type VI secretion system baseplate subunit TssF [T6SS (VF0569) - Effec
VFG002165(gb|WP_002356954) (efaA) endocarditis specific antigen [EfaA (VF0354) - Adherence (VFC0001)]
VFG014105(gb|NP_251843) (wzx) O-antigen translocase [LPS (VF0085) - Immune modulation (VFC0258)] [Ps
VFG002287(gb|WP_011860904) (toxA) toxin A [TcdA (VF0376) - Exotoxin (VFC0235)] [Clostridium difficile
VFG002488(gb|WP_004551879) (tssL-5) Type VI secretion system protein TssL [T6SS-1 (VF0429) - Effector d
VFG034928(gb|WP_001239082) (lifA/efa1) lymphostatin Efa1/LifA [Lymphostatin/LifA (VF0175) - Effector de

VFG002050(gb|WP_001087302) (gspG) general secretion pathway protein G [T2SS (VF0333) - Effector delivery
VFG013390(gb|WP_005651701) (lpxA) UDP-N-acetylglucosamine acyltransferase [LOS (VF0044) - Immune m
VFG000871(gb|WP_000790574) (fimB) Type 1 fimbriae Regulatory protein fimB [Type 1 fimbriae (VF0221) - A
VFG001252(gb|NP_249793) (fliG) flagellar motor switch protein G [Flagella (VF0273) - Motility (VFC0204)] [
VFG001233(gb|NP_249106) (chpC) probable chemotaxis protein [Type IV pili (VF0082) - Adherence (VFC000
VFG016041(gb|NP_251092) (pvdI) peptide synthase [Pyoverdine (VF0094) - Nutritional/Metabolic factor (VFC
VFG038464(gb|WP_011707389) (tapT) twitching ATPase [Tap type IV pili (VF0475) - Adherence (VFC0001)]
VFG000938(gb|WP_001015709) (iucC) aerobactin siderophore biosynthesis protein IucC [Aerobactin (VF0229)
VFG038719(gb|WP_016350061) (pomB) ompA family protein [Polar flagella (VF0473) - Motility (VFC0204)] [
VFG049114(gb|WP_012737527) (allS) DNA-binding transcriptional activator AllS [Allantion utilization (VF057
VFG045632(gb|WP_002286924) (ecbA/fss3) Collagen binding MSCRAMM, EcbA/Fss3 [EcbA (VF0539) - Adh
VFG050165(gb|WP_001259305) (neuA) N-acylneuraminate cytidylyltransferase [K1 capsule (VF0239) - Invasio
VFG044332(gb|WP_015874967) (ybtE) yersiniabactin biosynthesis salycil-AMP ligase YbtE [Ybt (VF0564) - N
VFG037822(gb|WP_000240700) (lpxC) UDP-3-O-acyl-N-acetylglucosamine deacetylase [LPS (VF0466) - Immu
VFG047708(gb|WP_003022611) (carB) carbamoyl phosphate synthase large subunit [Pyrimidine biosynthesis (V
VFG045789(gb|WP_001264082) (cgsF) curli production assembly/transport protein CsgF [Curli fibers (VF1138)
VFG001232(gb|NP_249105) (chpB) probable methylesterase [Type IV pili (VF0082) - Adherence (VFC0001)] [
VFG000122(gb|NP_252230) (algD) GDP-mannose 6-dehydrogenase AlgD [Alginate (VF0091) - Biofilm (VFC0
VFG038691(gb|WP_010634619) (flhG) CobQ/CobB/MinD/ParA family protein [Polar flagella (VF0473) - Motil
VFG000918(gb|WP_001312167) (chuT) periplasmic heme-binding protein ChuT [Chu (VF0227) - Nutritional/M
VFG042739(gb|NP_254019) (crc) catabolite repression control protein [Type IV pili (VF0082) - Adherence (VF
VFG035916(gb|WP_032490785) (aatB) AatB [Dispersin (VF0215) - Others (VFC0346)] [Escherichia coli O44:H
VFG002458(gb|WP_004525314) (bsaT) Type III secretion system protein BsaT [Bsa T3SS (VF0428) - Effector d
VFG038860(gb|WP_005314298) (lfhB) lateral flagellar biosynthetic protein, LfhB [Lateral flagella (VF0474) - M
VFG002470(gb|WP_004533272) (orgB) Type III secretion system protein [Bsa T3SS (VF0428) - Effector delive
VFG038872(gb|WP_005314272) (lfgB) lateral flagellar basal body protein, LfgB [Lateral flagella (VF0474) - Mo
VFG038880(gb|WP_005314250) (lfgK) lateral flagellar hook-associated protein [Lateral flagella (VF0474) - Mot
VFG039501(gb|NP_820443) (coxCC7/cig44) Coxiella Dot/Icm type IVB secretion system translocated effector [
VFG002033(gb|WP_010874498) (p1/MgPa/gapA) ADP1_MYCPN adhesin P1 [Cytadherence organelle (VF033
VFG000147(gb|NP_252530) (exoS) type III secretion system effector ExoS, ADP ribosyltransferase activity and
VFG050699(gb|WP_000840548.1) (pilR) sigma-54 dependent transcriptional regulator [TFP (VF1334) - Adherer
VFG002047(gb|WP_000498818) (gspD) general secretion pathway protein D [T2SS (VF0333) - Effector delivery
VFG044324(gb|WP_032447693) (ybtX) yersiniabactin-associated zinc MFS transporter YbtX [Ybt (VF0564) - N
VFG038727(gb|WP_011705295) (cheW) chemotaxis protein CheW [Polar flagella (VF0473) - Motility (VFC020
VFG005197(gb|WP_000006706) (pavA) Fibronectin-binding protein-like protein A [PavA (VF0283) - Adherenc
VFG048621(gb|WP_004213924) (iutA) ferric aerobactin receptor IutA [Aerobactin (VF0565) - Nutritional/Metal
VFG038671(gb|WP_011705282) (fliQ) flagellar biosynthetic protein FliQ [Polar flagella (VF0473) - Motility (V
VFG044165(gb|WP_001041793) (entS) enterobactin exporter, iron-regulated [Enterobactin (VF0228) - Nutrition
VFG043773(gb|YP_398516) (bont) botulinum neurotoxin, BoNT [BoNT (VF0375) - Exotoxin (VFC0235)] [Clos
VFG049147(gb|WP_001217110) (clbA) colibactin biosynthesis phosphopantetheinyl transferase ClbA [Colibacti
VFG041031(gb|NP_251049) (PA2359) transcriptional regulator [HSI-3 (VF0944) - Effector delivery system (VF
VFG043775(gb|BAB86845) (bont) botulinum neurotoxin, BoNT [BoNT (VF0375) - Exotoxin (VFC0235)] [Clos
VFG038501(gb|WP_043119142) (tapV) type IV pilus assembly protein TapV [Tap type IV pili (VF0475) - Adhe
VFG038265(gb|WP_016352108) (exeB) general secretion pathway protein B [Exe T2SS (VF0478) - Effector del
VFG001213(gb|NP_253236) (pilS) two-component sensor PilS [Type IV pili (VF0082) - Adherence (VFC0001)]
VFG038868(gb|WP_005314282) (lfiJ) lateral flagellar protein, LfiJ [Lateral flagella (VF0474) - Motility (VFC02

VFG002502(gb|WP_004523066) (fliF) flagellar M-ring protein FliF [Flagella (VF0430) - Motility (VFC0204)] [
VFG038405(gb|WP_011705727) (AHA_RS09305) type VI secretion system PAAR protein [T6SS (VF0480) - E
VFG041033(gb|NP_251051) (PA2361) hypothetical protein [HSI-3 (VF0944) - Effector delivery system (VFC00
VFG049900 (tssM) Type VI secretion system protein TssM [T6SS (VF0579) - Effector delivery system (VFC008
VFG001227(gb|NP_249101) (pilI) twitching motility protein PilI [Type IV pili (VF0082) - Adherence (VFC0001
VFG045527(gb|WP_010947280) (ravY) Dot/Icm type IV secretion system effector RavY [Dot/Icm T4SS secrete
VFG001408(gb|NP_217456) (mas) multifunctional mycocerosic acid synthase [PDIM (VF0309) - Immune modu
VFG051244(gb|WP_004683766.1) (BPE043) type IV secretion system effector [T4SS secreted effectors (VF069
VFG015527(gb|NP_250591) (phzB2) phenazine biosynthesis protein PhzB [Pyocyanin (VF0100) - Nutritional/M
VFG038374(gb|WP_011705709) (atsB) Type VI secretion system protein [T6SS (VF0480) - Effector delivery sy
VFG000874(gb|WP_000824100) (fimI) Fimbrin-like protein fimI precursor [Type 1 fimbriae (VF0221) - Adhere
VFG038651(gb|WP_011705278) (fliL) flagellar basal body-associated protein FliL [Polar flagella (VF0473) - Mo
VFG000616(gb|NP_709455) (iucB) aerobactin synthesis protein IucB [Aerobactin (VF0123) - Nutritional/Metab
VFG038293(gb|WP_016349340) (exeI) general secretion pathway protein I [Exe T2SS (VF0478) - Effector deliv
VFG037028(gb|WP_002218620) (katA) catalase [KatA (VF0454) - Stress survival (VFC0282)] [Neisseria menin
VFG000364(gb|WP_000140406) (ybtA) transcriptional regulator YbtA [Yersiniabactin (VF0136) - Nutritional/M
VFG038407(gb|WP_011705728) (vgrG3) VgrG protein [T6SS (VF0480) - Effector delivery system (VFC0086)]
VFG000931(gb|WP_001336900) (entC) isochorismate synthase 1 [Enterobactin (VF0228) - Nutritional/Metaboli
VFG048808(gb|WP_004899416) (rfbK1) O9 family phosphomannomutase RfbK1 [Capsule (VF0560) - Immune
VFG000363(gb|WP_002212775) (irp2) yersiniabactin biosynthetic protein Irp2 [Yersiniabactin (VF0136) - Nutri
VFG038627(gb|WP_016350050) (fliF) flagellar MS-ring protein [Polar flagella (VF0473) - Motility (VFC0204)]
VFG013260(gb|WP_010868968) (lgtC) glycosyltransferase [LOS (VF0044) - Immune modulation (VFC0258)] [
VFG044326(gb|WP_001327262) (ybtP) yersiniabactin ABC transporter ATP-binding/permease protein YbtP [Y
VFG000181(gb|NP_251792) (xcpS) general secretion pathway protein F [Xcp secretion system (VF0084) - Effec
VFG002531(gb|WP_011205291) (cheA) chemotaxis two-component sensor kinase CheA [Flagella (VF0430) - M
VFG040864(gb|WP_001172770) (essC) type VII secretion system protein EssC [T7SS (VF0581) - Effector deliv
VFG046853(gb|WP_003029539) (waaY) hypothetical protein [Capsule (VF0543) - Immune modulation (VFC02
VFG048257(gb|WP_004174507) (fimI) type 1 pilus biosynthesis fimbrial protein [Type I fimbriae (VF0566) - A
VFG045818(gb|WP_001528154) (tsh) temperature-sensitive protein Tsh [Tsh (VF0233) - Effector delivery syste
VFG038380(gb|WP_043163044) (vipB) Type VI secretion system contractile sheath large subunit TssC/VipB [T
VFG048885(gb|WP_014907227) (gmd) GDP-mannose 4,6-dehydratase [Capsule (VF0560) - Immune modulatio
VFG002517(gb|WP_004203463) (flgK) flagellar hook-associated protein 1 FlgK [Flagella (VF0430) - Motility (
VFG045792(gb|WP_000791650) (csgB) curlin minor subunit CsgB [Curli fibers (VF1138) - Adherence (VFC000
VFG013292(gb|WP_105163174) (lic3A) alpha-2,3-sialyltransferase [LOS (VF0044) - Immune modulation (VFC
VFG002046(gb|WP_000135089) (gspC) general secretion pathway protein C [T2SS (VF0333) - Effector delivery
VFG012327(gb|WP_011076307) (sfaC) S fimbrial switch regulatory protein SfaC [S fimbriae (VF0222) - Adhere
VFG038635(gb|WP_016350051) (fliH) flagellar assembly protein FliH [Polar flagella (VF0473) - Motility (VFC
VFG015859(gb|NP_252177) (pldA) phospholipase D [HSI-2 T6SS secreted effectors (VF0915) - Effector delive
VFG038596(gb|WP_016349157) (mshM) membrane protein with ATPase domain, GspA-like [MSHA type IV p
VFG002389(gb|NP_218396) (espK) ESX-1 type VII secretion system EspK [ESX-1 (VF0315) - Effector delivery
VFG048030(gb|WP_003022143) (pdpC) Type VI secretion system secreted protein PdpC [T6SS (VF0544) - Eff
VFG038876(gb|WP_005314259) (lfgG) lateral flagellar basal-body rod protein LfgG [Lateral flagella (VF0474)
VFG038600(gb|WP_016349156) (mshL) MSHA (mannose-sensitive hemagglutinin) pili biogenesis protein Msh
VFG038683(gb|WP_016350056) (flhA) flagellar biosynthesis protein FlhA [Polar flagella (VF0473) - Motility (
VFG049118(gb|WP_012737529) (allR) DNA-binding transcriptional repressor AllR [Allantion utilization (VF05
VFG049910(gb|WP_000348788) (tssG) Type VI secretion system protein TssG [T6SS (VF0579) - Effector deliv

VFG038448(gb|WP_016351632) (tapN) type IV pilus biogenesis protein PilN [Tap type IV pili (VF0475) - Adhe
VFG000923(gb|WP_001034892) (fepA) ferrienterobactin outer membrane transporter [Enterobactin (VF0228) - 
VFG038402(gb|WP_011705725) (vasK/icmF) Type VI secretion system protein [T6SS (VF0480) - Effector deliv
VFG000123(gb|NP_252231) (alg8) alginate-c5-mannuronan-epimerase AlgG [Alginate (VF0091) - Biofilm (VF
VFG049150(gb|WP_000982270) (clbD) colibactin biosynthesis dehydrogenase ClbD [Colibactin (VF0573) - Exo
VFG038476(gb|WP_016349433) (tppA) type IV pilus pseudopilin [Tap type IV pili (VF0475) - Adherence (VFC
VFG000368(gb|WP_000703040) (ybtS) salicylate synthase Irp9 [Yersiniabactin (VF0136) - Nutritional/Metaboli
VFG000930(gb|WP_000077767) (entF) enterobactin synthase multienzyme complex component, ATP-dependent
VFG034875(gb|WP_001270087) (espX5) Type III secretion system effector EspX5 [TTSS secreted effectors (VF
VFG045794(gb|WP_000992818) (csgC) curli assembly protein CsgC [Curli fibers (VF1138) - Adherence (VFC0
VFG002072(gb|NP_248776) (hsiE1) type VI secretion system hcp secretion island protein HsiE1, interacting wit
VFG000474(gb|NP_460200) (phoQ) sensor protein PhoQ [PhoPQ (VF0111) - Regulation (VFC0301)] [Salmone
VFG000879(gb|WP_000832235) (fimH) FimH protein precursor [Type 1 fimbriae (VF0221) - Adherence (VFC0
VFG050757(gb|WP_001166319.1) (gspE1) general secretion pathway protein E [T2SS (VF1335) - Effector deliv
VFG044084(gb|NP_251073) (PA2383) transcriptional regulator [Pyoverdine (VF0094) - Nutritional/Metabolic fa
VFG044323(gb|WP_015874960) (ybtS) yersiniabactin biosynthesis salicylate synthase YbtS [Ybt (VF0564) - Nu
VFG038393(gb|WP_010634228) (dotU) Type VI secretion system protein DotU [T6SS (VF0480) - Effector deliv
VFG003997(gb|NP_461706) (pipB2) type III secretion system effector PipB3 [TTSS-2 secreted effectors (VF09
VFG038603(gb|WP_016349155) (mshK) MSHA (mannose-sensitive hemagglutinin) pili biogenesis protein Msh
VFG001221(gb|NP_253730) (pilN) type IV pilus inner membrane platform protein PilN [Type IV pili (VF0082)
VFG034754(gb|WP_000081129) (espL1) Type III secretion system effector espL1 [TTSS secreted effectors (VF
VFG034895(gb|WP_001304001) (espY1) Type III secretion system effector EspY1 [TTSS secreted effectors (VF
VFG038824(gb|WP_011706648) (flgM) anti-sigma-D factor [Polar flagella (VF0473) - Motility (VFC0204)] [Ae
VFG006349(gb|WP_000542818) (sabB/hopO) outer membrane protein adhesin [SabA (VF0055) - Adherence (V
VFG013647(gb|WP_041175806) (invC/ilp) intimin-like invasin C protein [Invasin C/Ilp (VF0575) - Invasion (V
VFG000926(gb|WP_001443194) (fepD) ferrienterobactin ABC transporter permease [Enterobactin (VF0228) - N
VFG000944(gb|AAK16477) (draC) usher protein DraC [Dr adhesins (VF0223) - Adherence (VFC0001)] [Escher
VFG038828(gb|WP_011706649) (flgN) flagellar protein FlgN [Polar flagella (VF0473) - Motility (VFC0204)] [
VFG044172(gb|WP_000622316) (chuV) ATP-binding hydrophilic protein ChuV [Chu (VF0227) - Nutritional/M
VFG002194(gb|WP_011109506) (fsrA) response regulator [Fsr (VF0360) - Biofilm (VFC0271)] [Enterococcus f
VFG012865 (ipaH) invasion plasmid antigen IpaH [TTSS secreted effectors (VF0978) - Effector delivery system
VFG002136(gb|NP_220093) (copB) Type III secretion system translocator CopB [TTSS (VF0344) - Effector del
VFG000903(gb|WP_001045652) (pic) Pic serine protease precursor, autotransporter [Pic (VF0232) - Effector del
VFG002049(gb|WP_001173470) (gspF) general secretion pathway protein F [T2SS (VF0333) - Effector delivery
VFG038375(gb|WP_011705710) (atsC) Type VI secretion system protein [T6SS (VF0480) - Effector delivery sy
VFG045326(gb|WP_010947454) (LPG_RS08655) DUF1189 family protein [PmiA (VF0162) - Others (VFC034
VFG006962(gb|WP_001122154) (mshE) MSHA biogenesis protein MshE [MSHA pili (VF0515) - Adherence (V
VFG038631(gb|WP_011705273) (fliG) flagellar motor switch protein G [Polar flagella (VF0473) - Motility (VF
VFG002489(gb|WP_181937968) (tssM-5) Type VI secretion system protein TssM [T6SS-1 (VF0429) - Effector
VFG048488(gb|WP_002893737) (fepC) iron-enterobactin transporter ATP-binding protein [Ent (VF0562) - Nutr
VFG002084(gb|WP_000619136) (clpB/vasG) type VI secretion system AAA+ family ATPase [VAS T6SS (VF0
VFG044329(gb|WP_015874964) (irp1) yersiniabactin polyketide synthase HMWP1 [Ybt (VF0564) - Nutritional
VFG048687(gb|WP_002902160) (hcp/tssD) type VI secretion system protein, Hcp family [T6SS (VF0569) - Effe
VFG000885(gb|WP_000265729) (papD) chaperone protein PapD [P fimbriae (VF0220) - Adherence (VFC0001)
VFG000939(gb|WP_001287497) (iucB) aerobactin siderophore biosynthesis protein IucB [Aerobactin (VF0229)
VFG045387(gb|WP_011451394) (APH_RS05670) P44/Msp2 family outer membrane protein [P44/Msp2 family

VFG002306(gb|NP_461449) (ratB) putative outer membrane protein [RatB (VF0399) - Adherence (VFC0001)] [
VFG051131(gb|WP_006191504.1) (bmaC) autotransporter outer membrane beta-barrel domain-containing protei
VFG045793(gb|WP_000771429) (csgA) curlin major subunit CsgA [Curli fibers (VF1138) - Adherence (VFC000
VFG050625(gb|WP_000389061.1) (pilG) twitching motility response regulator PilG [TFP (VF1334) - Adherence
VFG048797(gb|WP_004175261) (ugd) UDP-glucose 6-dehydrogenase [Capsule (VF0560) - Immune modulation
VFG000916(gb|WP_001017192) (chuS) heme oxygenase ChuS [Chu (VF0227) - Nutritional/Metabolic factor (V
VFG041018(gb|NP_250351) (PA1660) hypothetical protein [HSI-2 (VF0943) - Effector delivery system (VFC00
VFG001235(gb|NP_249108) (chpE) probable chemotaxis protein [Type IV pili (VF0082) - Adherence (VFC0001
VFG001242(gb|NP_249775) (flgI) flagellar P-ring protein precursor FlgI [Flagella (VF0273) - Motility (VFC020
VFG000210(gb|NP_250406) (pscB) type III secretion system protein PscB [TTSS (VF0083) - Effector delivery s
VFG038105(gb|WP_000591436) (galU) UTP--glucose-1-phosphate uridylyltransferase GalU [Capsule (VF0465)
VFG000261(gb|WP_002225480) (iga) IgA-specific serine endopeptidase [IgA1 protease (VF0080) - Effector del
VFG013063(gb|WP_001017208) (shuS) heme/hemoglobin transport protein ShuS [Shu (VF0256) - Nutritional/M
VFG034756 (espL1) Type III secretion system effector espL1 [TTSS secreted effectors (VF1111) - Effector deliv
VFG002282(gb|WP_041707714) (cloSI) alpha-clostripain [Alpha-clostripain (VF0390) - Exoenzyme (VFC0251)
VFG013069(gb|WP_005019013) (shuU) permease of iron compound ABC transport system [Shu (VF0256) - Nu
VFG050372(gb|WP_001017033.1) (pilQ) type IV pilus secretin PilQ [TFP (VF1334) - Adherence (VFC0001)] [
VFG050386(gb|WP_000355489.1) (pilT) type IV pilus twitching motility protein PilT [TFP (VF1334) - Adheren
VFG002233(gb|WP_011181129) (virB4) type IV secretion system virB4 protein homolog [VirB/VirD4 type IV s
VFG015524(gb|NP_250590) (phzA2) phenazine biosynthesis protein PhzA [Pyocyanin (VF0100) - Nutritional/M
VFG000884(gb|WP_000654937) (papC) usher protein PapC [P fimbriae (VF0220) - Adherence (VFC0001)] [Es
VFG034873(gb|WP_001270084) (espX5) Type III secretion system effector EspX5 [TTSS secreted effectors (VF
VFG038389(gb|WP_011705718) (atsK) Type VI secretion system protein [T6SS (VF0480) - Effector delivery sy
VFG002436(gb|WP_080341181) (boaA) surface-exposed protein, autotransporter [BoaA (VF0434) - Adherence
VFG050670(gb|WP_000505931.1) (pilJ) methyl-accepting chemotaxis protein [TFP (VF1334) - Adherence (VF
VFG041020(gb|NP_250353) (PA1662) ClpA/B-type protease [HSI-2 (VF0943) - Effector delivery system (VFC
VFG000119(gb|NP_253948) (algR) alginate biosynthesis regulatory protein AlgR [Alginate (VF0091) - Biofilm
VFG001223(gb|NP_249086) (pilT) twitching motility protein PilT [Type IV pili (VF0082) - Adherence (VFC000
VFG014733(gb|NP_252216) (motY) probable outer membrane protein precursor [Flagella (VF0273) - Motility (V
VFG038403(gb|WP_011705726) (atsS) Type VI secretion system protein [T6SS (VF0480) - Effector delivery sys
VFG013265(gb|WP_005694045) (orfM) deoxyribonucleotide triphosphate pyrophosphatase [LOS (VF0044) - Im
VFG038517(gb|WP_016350156) (flpJ) Flp pilus pseudopilin [Flp type IV pili (VF0476) - Adherence (VFC0001)
VFG001900(gb|YP_002343773) (flhB) flagellar biosynthetic protein [Flagella (VF0114) - Motility (VFC0204)] [
VFG048459(gb|WP_004147525) (entS) enterobactin exporter EntS [Ent (VF0562) - Nutritional/Metabolic factor
VFG034772(gb|WP_000627910) (espL4) Type III secretion system effector EspL4 [TTSS secreted effectors (VF
VFG044325(gb|WP_002212761) (ybtQ) yersiniabactin ABC transporter ATP-binding/permease protein YbtQ [Y
VFG048830(gb|WP_014907233) (gndA) NADP-dependent phosphogluconate dehydrogenase [Capsule (VF0560
VFG045475(gb|WP_010945770) (LPG_RS00040) hypothetical protein [Dot/Icm T4SS secreted effectors (VF07
VFG041015(gb|NP_250348) (PA1657) hypothetical protein [HSI-2 (VF0943) - Effector delivery system (VFC00
VFG041267(gb|WP_010946879) (lem7) Dot/Icm type IV secretion system effector Lem7 [Dot/Icm T4SS secrete
VFG002278(gb|WP_011010095) (nagI) hyaluronidase [Mu-toxin (VF0389) - Exoenzyme (VFC0251)] [Clostridi
VFG037777(gb|WP_000867091) (lpsB) glycosyltransferase family 4 protein [LPS (VF0466) - Immune modulatio
VFG001811(gb|YP_177694) (fbpC) Secreted antigen 85-C FbpC (85C) (antigen 85 complex C) (AG58C) (myco
VFG044327(gb|WP_000140406) (ybtA) yersiniabactin transcriptional regulator YbtA [Ybt (VF0564) - Nutrition
VFG038788(gb|WP_011706639) (flgG) flagellar basal body rod protein FlgG [Polar flagella (VF0473) - Motility
VFG041305(gb|WP_010947679) (pieA/lirC) Dot/Icm type IV secretion system effector PieA/LirC [Dot/Icm T4S

VFG048755(gb|WP_049127998) (icmF/tssM) type VI secretion protein TssM [T6SS (VF0569) - Effector deliver
VFG000310(gb|WP_001201399) (pseFG) CMP-N-acetylneuraminic acid synthetase [Flagellar glycosylation/Pse
VFG048990(gb|WP_001741945) (galF) GalU regulator GalF [Capsule (VF0560) - Immune modulation (VFC025
VFG000002(gb|WP_010930159) (prn) pertactin precursor [Pertactin (VF0027) - Adherence (VFC0001)] [Bordet
VFG000889(gb|WP_000113351) (papF) P pilus minor subunit PapF [P fimbriae (VF0220) - Adherence (VFC00
VFG044159(gb|WP_000125846) (fes) enterobactin/ferric enterobactin esterase [Enterobactin (VF0228) - Nutritio
VFG000890(gb|WP_000758683) (papG) P pilus tip adhesin PapG [P fimbriae (VF0220) - Adherence (VFC0001
VFG000079(gb|NP_463763) (clpC) endopeptidase Clp ATP-binding chain C [ClpC (VF0072) - Stress survival (
VFG038399(gb|WP_011705723) (atsP) Type VI secretion system protein [T6SS (VF0480) - Effector delivery sy
VFG015973(gb|NP_251082) (pvdP) tyrosinase required for pyoverdine maturation [Pyoverdine (VF0094) - Nutr
VFG050212(gb|WP_011861707) (CD2831) SrtB-anchored collagen-binding adhesin [CD2831 (VF0598) - Adher
VFG015521(gb|NP_252906) (phzG1) phenazine biosynthesis protein PhzG, pyridoxamine 5'-phosphate oxidase [
VFG038376(gb|WP_164927604) (atsD) Type VI secretion system protein [T6SS (VF0480) - Effector delivery sy
VFG014642(gb|NP_252040) (flgA) flagellar basal body P-ring biosynthesis protein FlgA [Flagella (VF0273) - M
VFG011631(gb|WP_004684547) (bvrS) stimulus-sensing domain-containing protein [BvrR-BvrS (VF0368) - Re
VFG038091(gb|WP_000826956) (ACICU_RS00475) sugar transferase [Capsule (VF0465) - Immune modulation
VFG049073(gb|WP_015875021) (KP1_RS17240) DUF4422 domain-containing protein [LPS (VF0561) - Immun
VFG000360(gb|WP_002212799) (ybtT) type II thioesterase YbtT [Yersiniabactin (VF0136) - Nutritional/Metabo
VFG000211(gb|NP_250407) (pscC) type III secretion system secretin PscC [TTSS (VF0083) - Effector delivery
VFG002068(gb|NP_248772) (hsiA1) type VI secretion system hcp secretion island protein HsiA1 [HSI-I (VF033
VFG012501(gb|WP_000271272) (iroE) esterase [Salmochelin siderophore (VF0230) - Nutritional/Metabolic fact
VFG042978(gb|WP_011109477) (ebpC) endocarditis and biofilm-associated pilus major subunit EbpC [Ebp pili
VFG048498(gb|WP_012737619) (entF) enterobactin synthase subunit F [Ent (VF0562) - Nutritional/Metabolic f
VFG038667(gb|WP_016350054) (fliP) flagellar biosynthesis protein FliP [Polar flagella (VF0473) - Motility (VF
VFG002276(gb|WP_011009653) (colA) collagenase [Kappa-toxin (VF0388) - Exoenzyme (VFC0251)] [Clostrid
VFG000875(gb|WP_000066579) (fimC) Chaperone protein fimC precursor [Type 1 fimbriae (VF0221) - Adhere
VFG002101(gb|WP_010948213) (sidC) Dot/Icm type IV secretion system effector SidC, ER interactor [Dot/Icm
VFG000150(gb|NP_250881) (exoY) type III secretion system effector ExoY, adenylate cyclase [TTSS secreted e
VFG012627(gb|WP_000910392) (chuU) heme permease protein ChuU [Chu (VF0234) - Nutritional/Metabolic fa
VFG049162(gb|WP_002430641) (clbP) precolibactin peptidase ClbP [Colibactin (VF0573) - Exotoxin (VFC023
VFG049124(gb|WP_012737540) (allD) ureidoglycolate dehydrogenase [Allantion utilization (VF0572) - Nutritio
VFG002052(gb|WP_000820136) (gspI) general secretion pathway protein I [T2SS (VF0333) - Effector delivery
VFG001245(gb|NP_249778) (flgL) flagellar hook-associated protein 3 FlgL [Flagella (VF0273) - Motility (VFC
VFG048277(gb|WP_009308599) (fimD) outer membrane usher protein [Type I fimbriae (VF0566) - Adherence
VFG000333(gb|WP_005694147) (lic2A) glycosyltransferase [LOS (VF0044) - Immune modulation (VFC0258)]
VFG034831(gb|WP_000484243) (espR4) Type III secretion system effector espR4 [TTSS secreted effectors (VF
VFG000144(gb|NP_249940) (aprA) alkaline metalloproteinase precursor [Alkaline protease (VF0090) - Exoenzy
VFG000907(gb|WP_000376545) (hlyB) Hemolysin B [-Hemolysin (VF0225) - Exotoxin (VFC0235)] [Escherich
VFG000264(gb|WP_002247545) (lbpA) lactoferrin-binding protein A [Lbp (VF0047) - Nutritional/Metabolic fac
VFG044333(gb|WP_015874968) (fyuA) yersiniabactin receptor FyuA [Ybt (VF0564) - Nutritional/Metabolic fac
VFG048985(gb|WP_014907219) (KP1_RS17355) phosphatase PAP2 family protein [Capsule (VF0560) - Immun
VFG002166(gb|WP_010714416) (ace) collagen adhesin protein [Ace (VF0355) - Adherence (VFC0001)] [Enter
VFG002300(gb|CAA88565) (tcnA) alpha-toxin [Alpha-toxin (novyi) (VF0386) - Exotoxin (VFC0235)] [Clostrid
VFG002184(gb|WP_002359685) (cpsG) MurB family protein [Capsule (VF0361) - Immune modulation (VFC02
VFG044328(gb|WP_015874963) (irp2) yersiniabactin non-ribosomal peptide synthetase HMWP2 [Ybt (VF0564)
VFG043624(gb|WP_004149659) (mrkD) fimbrial adhesin protein precursor MrkD [Type 3 fimbriae (VF0567) -

VFG038395(gb|WP_011705721) (clpB) type VI secretion system ATPase ClpV1 [T6SS (VF0480) - Effector deli
VFG038679(gb|WP_016350055) (flhB) flagellar biosynthetic protein FlhB [Polar flagella (VF0473) - Motility (V
VFG001248(gb|NP_249788) (fleQ) transcriptional regulator FleQ [Flagella (VF0273) - Motility (VFC0204)] [Ps
VFG043626(gb|WP_000820818) (mrkB) fimbrial chaperone protein mrkB precursor [Type 3 fimbriae (VF0567)
VFG001256(gb|NP_250134) (fliM) flagellar motor switch protein FliM [Flagella (VF0273) - Motility (VFC0204
VFG002048(gb|WP_000249362) (gspE) general secretion pathway protein E [T2SS (VF0333) - Effector delivery
VFG038460(gb|WP_043119325) (tapQ) type IV pilus secretin PilQ [Tap type IV pili (VF0475) - Adherence (VF
VFG037912(gb|WP_000939040) (pseB) UDP-N-acetylglucosamine 4,6-dehydratase (inverting) [Capsule (VF04
VFG048665(gb|WP_004176431) (dotU/tssL) type VI secretion system protein, DotU/TssL family [T6SS (VF056
VFG048676(gb|WP_014907726) (ompA) OmpA family protein [T6SS (VF0569) - Effector delivery system (VF
VFG034901(gb|WP_000868321) (espY2) Type III secretion system effector EspY2 [TTSS secreted effectors (V
VFG051134(gb|WP_002969669.1) (btaE) Trimeric autotransporter adhesin, YadA-like family protein [BtaE (VF
VFG049902(gb|WP_011310140) (tssM) Type VI secretion system protein TssM [T6SS (VF0579) - Effector deli
VFG048439(gb|WP_014907941) (entC) isochorismate synthase [Ent (VF0562) - Nutritional/Metabolic factor (V
VFG044096(gb|NP_252908) (PA4218) transporter [Pyochelin (VF0095) - Nutritional/Metabolic factor (VFC027
VFG038509(gb|WP_016350158) (flpL) Flp pilus pseudopilin [Flp type IV pili (VF0476) - Adherence (VFC0001
VFG041026(gb|NP_250359) (PA1668) hypothetical protein [HSI-2 (VF0943) - Effector delivery system (VFC00
VFG001259(gb|NP_250137) (fliP) flagellar biosynthetic protein FliP [Flagella (VF0273) - Motility (VFC0204)]
VFG001270(gb|WP_010929997) (cyaD) cyclolysin secretion protein [Cya (VF0028) - Exotoxin (VFC0235)] [Bo
VFG002197(gb|WP_002387027) (bopD) sugar-binding transcriptional regulator, LacI family [BopD (VF0362) -
VFG000141(gb|NP_253675) (waaA) lipopolysaccharide core biosynthesis protein WaaP [LPS (VF0085) - Immu
VFG000130(gb|NP_252238) (algI) alginate o-acetyltransferase AlgI [Alginate (VF0091) - Biofilm (VFC0271)] [
VFG002500(gb|WP_004556525) (fliH) flagellar assembly protein H [Flagella (VF0430) - Motility (VFC0204)] [
VFG002220(gb|WP_004686741) (pgm) phosphoglucomutase [LPS (VF0367) - Immune modulation (VFC0258)]
VFG037837(gb|WP_000868104) (lpxD) UDP-3-O-(3-hydroxymyristoyl)glucosamine N-acyltransferase [LPS (V
VFG049914(gb|WP_000037389) (tssB) Type VI secretion system protein TssB [T6SS (VF0579) - Effector deliv
VFG038584(gb|WP_016349161) (mshF) hypothetical protein [MSHA type IV pili (VF0477) - Adherence (VFC0
VFG038611(gb|WP_016349153) (mshI1) MSHA (mannose-sensitive hemagglutinin) pili biogenesis protein Msh
VFG000126(gb|NP_252234) (algE) alginate biosynthetic protein AlgK precursor [Alginate (VF0091) - Biofilm (
VFG038409(gb|WP_011704492) (fimA) fimbrial protein [Type I pili (VF0486) - Adherence (VFC0001)] [Aerom
VFG006914(gb|WP_000418747) (pilD/vcpD) A24 family peptidase [ChiRP (VF0609) - Adherence (VFC0001)]
VFG000498(gb|NP_460362) (sseA) chaperone for sseB and sseD [TTSS (SPI-2 encode) (VF0321) - Effector del
VFG002045(gb|WP_010930608) (bvgS) virulence factors two-component system sensor histidine kinase BvgS [
VFG001229(gb|NP_249103) (pilK) methyltransferase PilK [Type IV pili (VF0082) - Adherence (VFC0001)] [Ps
VFG038313(gb|WP_043118380) (exeN) general secretion pathway protein N [Exe T2SS (VF0478) - Effector del
VFG038385(gb|WP_011705716) (atsI) Type VI secretion system protein [T6SS (VF0480) - Effector delivery sys
VFG000873(gb|WP_000695571) (fimA) Type-1 fimbrial protein, A chain precursor [Type 1 fimbriae (VF0221) -
VFG048327(gb|WP_004152886) (mrkH) transcriptional activator [Type 3 fimbriae (VF0567) - Biofilm (VFC027
VFG034624(gb|WP_000755097) (cfaB) colonization factor antigen 1 [Adhesive fimbriae (VF0213) - Adherence
VFG001214(gb|NP_253237) (pilR) two-component response regulator PilR [Type IV pili (VF0082) - Adherence
VFG043624(gb|WP_004149659) (mrkD) fimbrial adhesin protein precursor MrkD [Type 3 fimbriae (VF0567) -
VFG001288(gb|WP_000723836) (icaC) intercellular adhesion protein C, involved in polysaccharide intercellular
VFG048697(gb|WP_015874721) (clpV/tssH) type VI secretion system ATPase TssH [T6SS (VF0569) - Effector
VFG000109(gb|WP_000021616) (zot) zona occludens toxin [Zot (VF0129) - Exotoxin (VFC0235)] [Vibrio chol
VFG000115(gb|NP_249839) (toxA) exotoxin A precursor [ExoA (VF0086) - Exotoxin (VFC0235)] [Pseudomon
VFG001938(gb|YP_002344530) (Cj1137c) glycosyltransferase [LOS (VF0326) - Immune modulation (VFC025

VFG038364(gb|WP_005320615) (aexT) Type III secretion system effector AexT, bifunctional toxin with GTPase
VFG002076(gb|NP_248780) (clpV1) type VI secretion system AAA+ family ATPase [HSI-I (VF0334) - Effector
VFG038820(gb|WP_161785096) (flgA) flagella basal body P-ring formation protein FlgA [Polar flagella (VF047
VFG048990(gb|WP_001741945) (galF) GalU regulator GalF [Capsule (VF0560) - Immune modulation (VFC025
VFG049122(gb|WP_012737539) (allC) allantoate amidohydrolase [Allantion utilization (VF0572) - Nutritional/M
VFG038413(gb|WP_043118350) (fimC) outer membrane usher protein [Type I pili (VF0486) - Adherence (VFC
VFG044325(gb|WP_002212761) (ybtQ) yersiniabactin ABC transporter ATP-binding/permease protein YbtQ [Y
VFG038780(gb|WP_011706637) (flgI) flagellar basal body P-ring protein [Polar flagella (VF0473) - Motility (V
VFG049147(gb|WP_001217110) (clbA) colibactin biosynthesis phosphopantetheinyl transferase ClbA [Colibacti
VFG038346(gb|WP_005320760) (acr1) Type III secretion system gatekeeper subunit [T3SS (VF0479) - Effector
VFG014707(gb|NP_250151) (motC) flagellar motor protein [Flagella (VF0273) - Motility (VFC0204)] [Pseudom
VFG000923(gb|WP_001034892) (fepA) ferrienterobactin outer membrane transporter [Enterobactin (VF0228) -
VFG038655(gb|WP_011705279) (fliM) flagellar motor switch protein FliM [Polar flagella (VF0473) - Motility (
VFG038389(gb|WP_011705718) (atsK) Type VI secretion system protein [T6SS (VF0480) - Effector delivery sy
VFG000028(gb|WP_003808614) (brkB) serum resistance protein [Brk (VF0034) - Immune modulation (VFC025
VFG048600(gb|WP_004213920) (iucA) aerobactin Synthetase IucA [Aerobactin (VF0565) - Nutritional/Metabol
VFG002471(gb|WP_004533443) (bprP) regulatory protein [Bsa T3SS (VF0428) - Effector delivery system (VFC
VFG001444(gb|AAG10151) (aslA) putative arylsulfatase [AslA (VF0238) - Invasion (VFC0083)] [Escherichia c
VFG002054(gb|WP_000633233) (gspK) general secretion pathway protein K [T2SS (VF0333) - Effector delivery
VFG038859(gb|WP_005314301) (lfiR) lateral flagellar biosynthetic protein LfiR [Lateral flagella (VF0474) - Mo
VFG014984(gb|NP_253136) (algW) AlgW protein [Alginate (VF0091) - Biofilm (VFC0271)] [Pseudomonas ae
VFG038888(gb|WP_011898286) (lafS) LafS, RNA polymerase sigma factor [Lateral flagella (VF0474) - Motility
VFG038261(gb|WP_016352107) (exeA) general secretion pathway protein A [Exe T2SS (VF0478) - Effector de
VFG049903(gb|WP_001240533) (tssA) Type VI secretion system protein TssA [T6SS (VF0579) - Effector deliv
VFG038600(gb|WP_016349156) (mshL) MSHA (mannose-sensitive hemagglutinin) pili biogenesis protein Msh
VFG038393(gb|WP_010634228) (dotU) Type VI secretion system protein DotU [T6SS (VF0480) - Effector deli
VFG048819(gb|WP_014907234) (KP1_RS17280) mannose-1-phosphate guanylyltransferase/mannose-6-phospha
VFG044097(gb|NP_252909) (PA4219) hypothetical protein [Pyochelin (VF0095) - Nutritional/Metabolic factor
VFG045342(gb|WP_002221634) (yapK) autotransporter protein YapK [YapK (VF0508) - Adherence (VFC0001
VFG050212(gb|WP_011861707) (CD2831) SrtB-anchored collagen-binding adhesin [CD2831 (VF0598) - Adhe
VFG041016(gb|NP_250349) (PA1658) hypothetical protein [HSI-2 (VF0943) - Effector delivery system (VFC0
VFG002305(gb|NP_461448) (shdA) AIDA autotransporter-like protein [ShdA (VF0398) - Adherence (VFC0001
VFG044165(gb|WP_001041793) (entS) enterobactin exporter, iron-regulated [Enterobactin (VF0228) - Nutrition
VFG048621(gb|WP_004213924) (iutA) ferric aerobactin receptor IutA [Aerobactin (VF0565) - Nutritional/Meta
VFG002172(gb|WP_010710127) (EF0485) aggregation substance [AS (VF0352) - Adherence (VFC0001)] [Ente
VFG001827(gb|WP_001020414) (senB) enterotoxin [ShET2 (VF0258) - Exotoxin (VFC0235)] [Shigella boydii
VFG038359(gb|WP_005321607) (aopH) Type III secretion system effector AopH, putative tyrosine phosphatase
VFG049007(gb|WP_002911561) (rcsA) transcriptional activator for ctr capsule biosynthesis [RcsAB (VF0571) -
VFG002055(gb|WP_000097229) (gspL) general secretion pathway protein L [T2SS (VF0333) - Effector delivery
VFG001405(gb|NP_217219) (sigA/rpoV) RNA polymerase sigma factor SigA [SigA (VF0257) - Regulation (VF
VFG000616(gb|NP_709455) (iucB) aerobactin synthesis protein IucB [Aerobactin (VF0123) - Nutritional/Metab
VFG013648(gb|WP_012104486) (invD) invasin D protein [Invasin D (VF0576) - Invasion (VFC0083)] [Yersinia
VFG002165(gb|WP_002356954) (efaA) endocarditis specific antigen [EfaA (VF0354) - Adherence (VFC0001)]
VFG048247(gb|WP_002916190) (fimA) type 1 major fimbrial subunit precursor [Type I fimbriae (VF0566) - Ad
VFG038792(gb|WP_011706640) (flgF) flagellar basal-body rod protein FlgF [Polar flagella (VF0473) - Motility
VFG014105(gb|NP_251843) (wzx) O-antigen translocase [LPS (VF0085) - Immune modulation (VFC0258)] [Ps

VFG048654(gb|WP_004224345) (vasE/tssK) type VI secretion system baseplate subunit TssK [T6SS (VF0569) -
VFG000359(gb|WP_001088826) (ybtE) yersiniabactin siderophore biosynthetic protein [Yersiniabactin (VF0136
VFG038848(gb|WP_016350360) (flmD) FlmD protein [Polar flagella (VF0473) - Motility (VFC0204)] [Aeromo
VFG049900 (tssM) Type VI secretion system protein TssM [T6SS (VF0579) - Effector delivery system (VFC00
VFG048348(gb|WP_004149658) (mrkJ) phosphodiesterase [Type 3 fimbriae (VF0567) - Biofilm (VFC0271)] [K
VFG001242(gb|NP_249775) (flgI) flagellar P-ring protein precursor FlgI [Flagella (VF0273) - Motility (VFC020
VFG004906(gb|WP_000778477) (set25) superantigen-like protein SSL10 [SSLs (VF0990) - Exotoxin (VFC0235
VFG043518(gb|AAN12397) (acm) collagen adhesin precursor Acm [Acm (VF0419) - Adherence (VFC0001)] [E
VFG000122(gb|NP_252230) (algD) GDP-mannose 6-dehydrogenase AlgD [Alginate (VF0091) - Biofilm (VFC0
VFG049160(gb|WP_001327259) (clbN) colibactin non-ribosomal peptide synthetase ClbN [Colibactin (VF0573)
VFG045789(gb|WP_001264082) (cgsF) curli production assembly/transport protein CsgF [Curli fibers (VF1138)
VFG049902(gb|WP_011310140) (tssM) Type VI secretion system protein TssM [T6SS (VF0579) - Effector deli
VFG044328(gb|WP_015874963) (irp2) yersiniabactin non-ribosomal peptide synthetase HMWP2 [Ybt (VF0564)
VFG000514(gb|NP_460379) (ssaV) type III secretion system major export apparatus protein ssaV [TTSS (SPI-2
VFG050115(gb|WP_002081542) (cesA) Cereulide synthetase A, CesA [Cereulide (VF0583) - Exotoxin (VFC02
VFG001259(gb|NP_250137) (fliP) flagellar biosynthetic protein FliP [Flagella (VF0273) - Motility (VFC0204)]
VFG002525(gb|WP_004185006) (cheY) chemotaxis protein CheY [Flagella (VF0430) - Motility (VFC0204)] [B
VFG038365(gb|WP_011705044) (hcp1) HcpA-like protein [T6SS (VF0480) - Effector delivery system (VFC00
VFG038880(gb|WP_005314250) (lfgK) lateral flagellar hook-associated protein [Lateral flagella (VF0474) - Mo
VFG048609(gb|WP_004213923) (iucD) lysine 6-monooxygenase IucD [Aerobactin (VF0565) - Nutritional/Meta
VFG050301(gb|NP_218420) (cpnT) NAD+ glycohydrolase [CpnT (VF1197) - Effector delivery system (VFC00
VFG048518(gb|WP_012737617) (fepA) outer membrane receptor FepA [Ent (VF0562) - Nutritional/Metabolic f
VFG050699(gb|WP_000840548.1) (pilR) sigma-54 dependent transcriptional regulator [TFP (VF1334) - Adheren
VFG002060(gb|NP_248764) (ppkA) serine/threonine protein kinase PpkA [HSI-I (VF0334) - Effector delivery s
VFG048985(gb|WP_014907219) (KP1_RS17355) phosphatase PAP2 family protein [Capsule (VF0560) - Immun
VFG012865 (ipaH) invasion plasmid antigen IpaH [TTSS secreted effectors (VF0978) - Effector delivery system
VFG038679(gb|WP_016350055) (flhB) flagellar biosynthetic protein FlhB [Polar flagella (VF0473) - Motility (V
VFG038603(gb|WP_016349155) (mshK) MSHA (mannose-sensitive hemagglutinin) pili biogenesis protein Msh
VFG050400(gb|WP_000347039.1) (pilU) PilT/PilU family type 4a pilus ATPase [TFP (VF1334) - Adherence (V
VFG001228(gb|NP_249102) (pilJ) twitching motility protein PilJ [Type IV pili (VF0082) - Adherence (VFC000
VFG002196(gb|WP_011109578) (EF3023) polysaccharide lyase, family 8 [Hyaluronidase (VF0359) - Exoenzym
VFG049149(gb|WP_041169023) (clbC) colibactin polyketide synthase ClbC [Colibactin (VF0573) - Exotoxin (V
VFG045474(gb|AAM75251) (cylM) CylM; cytolysin subunit modifier [Cytolysin (VF0356) - Exotoxin (VFC023
VFG050162(gb|WP_001161829) (kpsS) Capsule polysaccharide export protein [K1 capsule (VF0239) - Invasion
VFG000198(gb|NP_250394) (pcrD) type III secretion system protein PcrD [TTSS (VF0083) - Effector delivery s
VFG001447(gb|CAA50007) (cnf1) cytotoxic necrotizing factor 1 [CNF-1 (VF0240) - Exotoxin (VFC0235)] [Esc
VFG038581(gb|WP_043118279) (mshB) MSHA (mannose-sensitive hemagglutinin) pili minor prepilin protein M
VFG050263(gb|NP_220137) (CT_620) hypothetical protein [TTSS secreted effectors (VF0711) - Effector deliver
VFG049158(gb|WP_001297937) (clbL) colibactin biosynthesis amidase ClbL [Colibactin (VF0573) - Exotoxin (
VFG002535(gb|WP_004198207) (fliC) flagellin [Flagella (VF0430) - Motility (VFC0204)] [Burkholderia pseudo
VFG038667(gb|WP_016350054) (fliP) flagellar biosynthesis protein FliP [Polar flagella (VF0473) - Motility (VF
VFG014720(gb|NP_250152) (motD) flagellar motor protein [Flagella (VF0273) - Motility (VFC0204)] [Pseudon
VFG037176(gb|WP_001081735) (plc1) phospholipase C [Phospholipase C (VF0470) - Exotoxin (VFC0235)] [A
VFG001224(gb|NP_249087) (pilU) twitching motility protein PilU [Type IV pili (VF0082) - Adherence (VFC00
VFG049915(gb|WP_001142968) (hcp2/tssD2) Type VI secretion system protein, Hcp family [T6SS (VF0579) -
VFG001949(gb|YP_002344804) (Cj1421c) sugar transferase [Capsule (VF0323) - Immune modulation (VFC025

VFG000478(gb|NP_459678) (fur) ferric iron uptake transcriptional regulator [Fur (VF0113) - Regulation (VFC0
VFG002049(gb|WP_001173470) (gspF) general secretion pathway protein F [T2SS (VF0333) - Effector delivery
VFG037330(gb|WP_001177743) (basD) acinetobactin biosynthesis protein BasD [Acinetobactin (VF0467) - Nut
VFG013069(gb|WP_005019013) (shuU) permease of iron compound ABC transport system [Shu (VF0256) - Nu
VFG000366(gb|WP_002212761) (ybtQ) yersiniabactin ABC transporter ATP-binding/permease protein YbtQ [Y
VFG042735(gb|NP_250513) (fimL) hypothetical protein [Type IV pili (VF0082) - Adherence (VFC0001)] [Pseu
VFG050277(gb|WP_000562056) (vpsE) exopolysaccharide biosynthesis flippase VpsE [VPS (VF0615) - Biofilm
VFG001713(gb|WP_001298062) (papX) PapX protein regulates flagellum synthesis to repress motility [P fimbri
VFG041012(gb|NP_250202) (PA1511) hypothetical protein [HSI-2 (VF0943) - Effector delivery system (VFC00
VFG000181(gb|NP_251792) (xcpS) general secretion pathway protein F [Xcp secretion system (VF0084) - Effec
VFG000159(gb|NP_251076) (pvdA) L-ornithine N5-oxygenase PvdA [Pyoverdine (VF0094) - Nutritional/Metab
VFG016338(gb|WP_001253006) (inhA) immune inhibitor A metalloprotease [InhA (VF0536) - Exoenzyme (VF
VFG038699(gb|WP_010634617) (cheY) chemotaxis protein CheY [Polar flagella (VF0473) - Motility (VFC020
VFG038788(gb|WP_011706639) (flgG) flagellar basal body rod protein FlgG [Polar flagella (VF0473) - Motility
VFG007586(gb|WP_001881782) (flhF) flagellar biosynthesis protein [Flagella (VF0519) - Motility (VFC0204)]
VFG002493(gb|WP_045606430) (fliO) flagellar protein FliO [Flagella (VF0430) - Motility (VFC0204)] [Burkho
VFG001214(gb|NP_253237) (pilR) two-component response regulator PilR [Type IV pili (VF0082) - Adherence
VFG016058(gb|NP_251114) (pvdL) peptide synthase PvdL [Pyoverdine (VF0094) - Nutritional/Metabolic factor
VFG038297(gb|WP_016349341) (exeJ) general secretion pathway protein J [Exe T2SS (VF0478) - Effector deli
VFG048779(gb|WP_004190503) (tssG) type VI secretion system baseplate subunit TssG [T6SS (VF0569) - Effe
VFG012022(gb|YP_002344707) (pseI) N-acetylneuraminic acid synthetase [Flagella (VF0114) - Motility (VFC0
VFG000315(gb|WP_000694809) (rfbM) mannose-6-phosphate isomerase [LPS (VF0056) - Immune modulation
VFG038882(gb|WP_005314247) (maf-5) modification accessory factor [Lateral flagella (VF0474) - Motility (VF
VFG000068(gb|NP_463735) (actA) actin-assembly inducing protein precursor [ActA (VF0066) - Motility (VFC
VFG044324(gb|WP_032447693) (ybtX) yersiniabactin-associated zinc MFS transporter YbtX [Ybt (VF0564) - N
VFG041020(gb|NP_250353) (PA1662) ClpA/B-type protease [HSI-2 (VF0943) - Effector delivery system (VFC
VFG012509(gb|WP_001111200) (iroC) ATP binding cassette transporter [Salmochelin siderophore (VF0230) - N
VFG001210(gb|NP_253244) (pilY1) type 4 fimbrial biogenesis protein PilY1 [Type IV pili (VF0082) - Adherenc
VFG001270(gb|WP_010929997) (cyaD) cyclolysin secretion protein [Cya (VF0028) - Exotoxin (VFC0235)] [Bo
VFG038836(gb|WP_011707085) (pomA2) chemotaxis protein PomA [Polar flagella (VF0473) - Motility (VFC0
VFG048762(gb|WP_015874732) (impA/tssA) type VI secretion system protein TssA [T6SS (VF0569) - Effector
VFG038590(gb|WP_043118275) (mshE) ATPase protein MshE, GspE-like [MSHA type IV pili (VF0477) - Adh
VFG002077(gb|NP_248781) (vgrG1a) type VI secretion system substrate VgrG1 [HSI-I (VF0334) - Effector deli
VFG013299(gb|WP_010868996) (waaQ) heptosyltransferase-like protein [LOS (VF0044) - Immune modulation
VFG042739(gb|NP_254019) (crc) catabolite repression control protein [Type IV pili (VF0082) - Adherence (VF
VFG000928(gb|WP_000640938) (fepG) iron-enterobactin ABC transporter permease [Enterobactin (VF0228) - N
VFG002063(gb|NP_248767) (icmF1/tssM1) type VI secretion system protein IcmF1 [HSI-I (VF0334) - Effector
VFG001953(gb|YP_002344808) (hddA) D-glycero-D-manno-heptose 7-phosphate kinase [Capsule (VF0323) - In
VFG000036(gb|WP_010929614) (bplC) lipopolysaccharide biosynthesis protein [LPS (VF0033) - Immune modu
VFG002192(gb|WP_002386893) (fsrC) histidine kinase, putative [Fsr (VF0360) - Biofilm (VFC0271)] [Enteroco
VFG000303(gb|WP_000495985) (virB4/cagE) type IV secretion system protein Cag23/CagE, VirB4 homolog [T
VFG038663(gb|WP_005298795) (fliO) flagellar biosynthesis protein, FliO [Polar flagella (VF0473) - Motility (V
VFG038587(gb|WP_016349160) (mshG) MSHA (mannose-sensitive hemagglutinin) pili biogenesis protein Msh
VFG048297(gb|WP_014906961) (fimG) type 1 fimbrial minor component [Type I fimbriae (VF0566) - Adherenc
VFG037720(gb|WP_001027056) (adeG) cation/multidrug efflux pump [AdeFGH efflux pump (VF0504) - Biofil
VFG002069(gb|NP_248773) (hsiB1/vipA/tssB) type VI secretion system tubule-forming protein VipA [HSI-I (V

VFG000617(gb|NP_709456) (iucC) aerobactin synthesis protein IucC [Aerobactin (VF0123) - Nutritional/Metab
VFG051070(gb|WP_001066523.1) (tssC) type VI secretion system contractile sheath large subunit [T6SS (VF13
VFG049114(gb|WP_012737527) (allS) DNA-binding transcriptional activator AllS [Allantion utilization (VF057
VFG002174(gb|WP_002369251) (gelE) coccolysin [Gelatinase (VF0357) - Exoenzyme (VFC0251)] [Enterococc
VFG000877(gb|WP_001244821) (fimF) FimF protein precursor [Type 1 fimbriae (VF0221) - Adherence (VFC0
VFG048257(gb|WP_004174507) (fimI) type 1 pilus biosynthesis fimbrial protein [Type I fimbriae (VF0566) - A
VFG042979(gb|WP_002379311) (srtC) sortase [Ebp pili (VF0538) - Adherence (VFC0001)] [Enterococcus faec
VFG038889(gb|WP_005314215) (lafT) LafT, flagella motor stator protein [Lateral flagella (VF0474) - Motility (
VFG001230(gb|NP_251805) (fimV) a polar peptidoglycan-binding protein involved in type IV pilus assembly [T
VFG001299(gb|WP_000565297) (cap8C) type 8 capsular polysaccharide synthesis protein Cap8C [Capsule (VF
VFG013658(gb|WP_011988414) (pilN) type IV pilus inner membrane platform protein PilN [Type IV pili (VF0
VFG000136(gb|NP_249456) (mucC) negative regulator for alginate biosynthesis MucB [Alginate (VF0091) - Bi
VFG001965(gb|YP_002344820) (Cj1437c) aminotransferase [Capsule (VF0323) - Immune modulation (VFC025
VFG002046(gb|WP_000135089) (gspC) general secretion pathway protein C [T2SS (VF0333) - Effector delivery
VFG050160(gb|WP_000030755) (kpsU) 3-deoxy-manno-octulosonate cytidylyltransferase [K1 capsule (VF0239
VFG043209(gb|WP_005168769) (cheD) methyl-accepting chemotaxis protein CheD [Flagella (VF0394) - Motili
VFG048830(gb|WP_014907233) (gndA) NADP-dependent phosphogluconate dehydrogenase [Capsule (VF0560
VFG048267(gb|WP_015875145) (fimC) periplasmic chaperone [Type I fimbriae (VF0566) - Adherence (VFC00
VFG045584(gb|WP_010948248) (LPG_RS12855) Dot/Icm type IV secretion system effector [Dot/Icm T4SS sec
VFG000924(gb|WP_001234311) (fepB) ferrienterobactin ABC transporter periplasmic binding protein [Enteroba
VFG000185(gb|NP_250381) (pscU) type III secretion system protein PscU [TTSS (VF0083) - Effector delivery s
VFG005361(gb|WP_002352221) (spaP/pac) cell surface antigen SpaP [SpaP (VF0526) - Adherence (VFC0001)
VFG039311(gb|NP_819427) (cetCb2) Coxiella Dot/Icm type IVB secretion system translocated effector [T4SS s
VFG000361(gb|WP_000982866) (ybtU) yersiniabactin biosynthetic protein YbtU [Yersiniabactin (VF0136) - Nu
VFG001302(gb|WP_001028293) (cap8F) type 8 capsular polysaccharide synthesis protein Cap8F [Capsule (VF0
VFG041031(gb|NP_251049) (PA2359) transcriptional regulator [HSI-3 (VF0944) - Effector delivery system (VF
VFG002274(gb|WP_011009584) (plc) phospholipase C [Alpha-toxin (CpPLC) (VF0378) - Exotoxin (VFC0235)
VFG038816(gb|WP_011706646) (cheV) CheV [Polar flagella (VF0473) - Motility (VFC0204)] [Aeromonas hyd
VFG042978(gb|WP_011109477) (ebpC) endocarditis and biofilm-associated pilus major subunit EbpC [Ebp pili
VFG049051(gb|WP_014907239) (KP1_RS17230) glycosyltransferase [LPS (VF0561) - Immune modulation (VF
VFG045337(gb|AHK25022) (llsD) streptolysin-associated protein LlsD [LLS (VF0410) - Exotoxin (VFC0235)]
VFG000862(gb|WP_014639426) (pet) Pet serine protease precursor, autotransporter [Pet (VF0217) - Effector del
VFG045626(gb|WP_014748741) (scm) collagen adhesin protein Scm [Scm (VF0418) - Adherence (VFC0001)] [
VFG000365(gb|WP_002214412) (ybtP) yersiniabactin ABC transporter ATP-binding/permease protein YbtP [Ye
VFG002171(gb|WP_011109430) (EF0149) aggregation substance [AS (VF0352) - Adherence (VFC0001)] [Ente
VFG001225(gb|NP_249099) (pilG) twitching motility protein PilG [Type IV pili (VF0082) - Adherence (VFC00
VFG002187(gb|WP_002370472) (cpsD) glycosyl transferase, group 2 family protein [Capsule (VF0361) - Immu
VFG050959(gb|WP_000556915.1) (tssM) type VI secretion system membrane subunit TssM [T6SS (VF1337) -
VFG034895(gb|WP_001304001) (espY1) Type III secretion system effector EspY1 [TTSS secreted effectors (V
VFG034636(gb|WP_000358674) (cfaC) colonisation factor antigen c, usher [Adhesive fimbriae (VF0213) - Adh
VFG044085(gb|NP_251074) (PA2384) hypothetical protein [Pyoverdine (VF0094) - Nutritional/Metabolic facto
VFG007849(gb|NP_217449) (ppsC) phthiocerol synthesis polyketide synthase type I PpsC [PDIM (VF0309) - In
VFG049911(gb|WP_000393837) (tssF) Type VI secretion system protein TssF [T6SS (VF0579) - Effector delive
VFG038407(gb|WP_011705728) (vgrG3) VgrG protein [T6SS (VF0480) - Effector delivery system (VFC0086)]
VFG001223(gb|NP_249086) (pilT) twitching motility protein PilT [Type IV pili (VF0082) - Adherence (VFC00
VFG039316(gb|NP_819448) (coxCC3) Coxiella Dot/Icm type IVB secretion system translocated effector [T4SS

VFG038593(gb|WP_016349158) (mshN) Tetratricopeptide repeat protein, PilF-like [MSHA type IV pili (VF047
VFG044329(gb|WP_015874964) (irp1) yersiniabactin polyketide synthase HMWP1 [Ybt (VF0564) - Nutritional/
VFG038865(gb|WP_005314289) (fliG) flagellar motor switch protein G [Lateral flagella (VF0474) - Motility (V
VFG038488(gb|WP_016349432) (tapY1) type IV pilus biogenesis protein [Tap type IV pili (VF0475) - Adherenc
VFG050372(gb|WP_001017033.1) (pilQ) type IV pilus secretin PilQ [TFP (VF1334) - Adherence (VFC0001)] [
VFG042382(gb|WP_000260524) (etpB) two-partner secretion transporter EtpB [EtpA (VF0437) - Adherence (V
VFG001271(gb|WP_010929998) (cyaE) cyclolysin secretion protein [Cya (VF0028) - Exotoxin (VFC0235)] [Bo
VFG000161(gb|NP_251089) (pvdD) pyoverdine synthetase D [Pyoverdine (VF0094) - Nutritional/Metabolic fac
VFG038845(gb|WP_016351249) (nueA) NeuA protein [Polar flagella (VF0473) - Motility (VFC0204)] [Aeromo
VFG050386(gb|WP_000355489.1) (pilT) type IV pilus twitching motility protein PilT [TFP (VF1334) - Adheren
VFG048478(gb|WP_004147521) (fepG) iron-enterobactin transporter permease [Ent (VF0562) - Nutritional/Met
VFG038596(gb|WP_016349157) (mshM) membrane protein with ATPase domain, GspA-like [MSHA type IV p
VFG038800(gb|WP_016351322) (flgD) flagellar hook capping protein [Polar flagella (VF0473) - Motility (VFC
VFG000932(gb|WP_000026784) (entE) 2,3-dihydroxybenzoate-AMP ligase component of enterobactin synthase
VFG044096(gb|NP_252908) (PA4218) transporter [Pyochelin (VF0095) - Nutritional/Metabolic factor (VFC027
VFG002067(gb|NP_248771) (fha1) type VI secretion system forkhead-associated protein Fha1 [HSI-I (VF0334)
VFG034721(gb|WP_000556298) (ibeC) phosphoethanolamine transferase CptA [Ibes (VF0237) - Invasion (VFC
VFG038432(gb|WP_016352168) (tapB) type IV pilus assembly ATPase, PilB-like [Tap type IV pili (VF0475) -
VFG041120(gb|WP_011205628) (BPS_RS26880) sensor histidine kinase VirA [T6SS-1 (VF0429) - Effector del
VFG002489(gb|WP_181937968) (tssM-5) Type VI secretion system protein TssM [T6SS-1 (VF0429) - Effector
VFG001248(gb|NP_249788) (fleQ) transcriptional regulator FleQ [Flagella (VF0273) - Motility (VFC0204)] [Ps
VFG038695(gb|WP_017409955) (fliA) flagellar biosynthesis sigma factor [Polar flagella (VF0473) - Motility (V
VFG001304(gb|WP_001216935) (cap8H) type 8 capsular polysaccharide synthesis protein Cap8H [Capsule (VF
VFG014117(gb|NP_251831) (wbpM) nucleotide sugar epimerase/dehydratase WbpM [LPS (VF0085) - Immune
VFG002491(gb|WP_004524547) (fliM) flagellar motor switch protein FliM [Flagella (VF0430) - Motility (VFC
VFG038832(gb|WP_016351724) (pomB2) flagellar motor protein MotB [Polar flagella (VF0473) - Motility (VF
VFG002184(gb|WP_002359685) (cpsG) MurB family protein [Capsule (VF0361) - Immune modulation (VFC02
VFG031840(gb|NP_218383) (espG1) ESX-1 secretion-associated protein EspG1 [ESX-1 (VF0315) - Effector de
VFG041013(gb|NP_250203) (hcpA) secreted protein Hcp [HSI-2 (VF0943) - Effector delivery system (VFC008
VFG002277(gb|WP_011009663) (nagH) hyaluronidase [Mu-toxin (VF0389) - Exoenzyme (VFC0251)] [Clostrid
VFG000446(gb|NP_459541) (fimD) usher protein FimD [Type 1 fimbriae (VF0102) - Adherence (VFC0001)] [S
VFG001240(gb|NP_249773) (flgG) flagellar basal-body rod protein FlgG [Flagella (VF0273) - Motility (VFC02
VFG038796(gb|WP_016351321) (flgE) flagellar hook protein FlgE [Polar flagella (VF0473) - Motility (VFC020
VFG048338(gb|WP_002916079) (mrkI) LuxR family regulatory protein [Type 3 fimbriae (VF0567) - Biofilm (V
VFG000153(gb|NP_252166) (rhlI) autoinducer synthesis protein RhlL [Quorum sensing (VF0093) - Biofilm (VF
VFG034612(gb|WP_000225871) (cfaA) colonisation factor antigen a, chaperone [Adhesive fimbriae (VF0213) -
VFG038764(gb|WP_017409846) (flrA) FleQ protein [Polar flagella (VF0473) - Motility (VFC0204)] [Aeromona
VFG049144(gb|WP_002892069) (acrB) acriflavine resistance protein B [AcrAB (VF0568) - Antimicrobial activi
VFG048237(gb|WP_002916189) (fimE) tyrosine recombinase [Type I fimbriae (VF0566) - Adherence (VFC000
VFG041015(gb|NP_250348) (PA1657) hypothetical protein [HSI-2 (VF0943) - Effector delivery system (VFC00
VFG034817(gb|WP_000671691) (espR1) Type III secretion system effector espR1 [TTSS secreted effectors (VF
VFG038320(gb|WP_005320812) (ascL) Type III secretion system C ring protein AscL [T3SS (VF0479) - Effecto
VFG015045(gb|NP_249821) (rhlC) rhamnosyltransferase 2 [Rhamnolipid (VF0089) - Immune modulation (VFC
VFG038277(gb|WP_016349336) (exeE) general secretory pathway protein E [Exe T2SS (VF0478) - Effector del
VFG037987(gb|WP_000037963) (pseI) pseudaminic acid synthase [Capsule (VF0465) - Immune modulation (V
VFG000678(gb|AAD32426) (cya) calmodulin sensitive adenylate cyclase, edema factor [Anthrax toxin (VF0142)

VFG048676(gb|WP_014907726) (ompA) OmpA family protein [T6SS (VF0569) - Effector delivery system (VF
VFG001256(gb|NP_250134) (fliM) flagellar motor switch protein FliM [Flagella (VF0273) - Motility (VFC0204
VFG000119(gb|NP_253948) (algR) alginate biosynthesis regulatory protein AlgR [Alginate (VF0091) - Biofilm
VFG041035(gb|NP_251053) (PA2363) hypothetical protein [HSI-3 (VF0944) - Effector delivery system (VFC00
VFG000930(gb|WP_000077767) (entF) enterobactin synthase multienzyme complex component, ATP-dependen
VFG038367(gb|WP_011705045) (vgrG1) Type VI secretion system effector VgrG1, ADP-ribosyltransferase [T
VFG024730(gb|NP_217462) (pks1) polyketide synthase [PDIM (VF0309) - Immune modulation (VFC0258)] [M
VFG002198(gb|WP_003850266) (tox) Diphtheria toxin precursor [DT (VF0363) - Exotoxin (VFC0235)] [Coryn
VFG002219(gb|WP_011005413) (cgs) cyclic beta 1-2 glucan synthetase [CG (VF0366) - Immune modulation (V
VFG045464(gb|WP_000562556) (tcpC) Tir domain containing protein TcpC [TcpC (VF0413) - Immune modula
VFG043623(gb|WP_002916122) (mrkF) type 3 fimbrial minor pilin subunit MrkF [Type 3 fimbriae (VF0567) -
VFG000878(gb|WP_000872015) (fimG) FimG protein precursor [Type 1 fimbriae (VF0221) - Adherence (VFC0
VFG038269(gb|WP_075384415) (exeC) general secretion pathway protein C [Exe T2SS (VF0478) - Effector del
VFG001254(gb|NP_249795) (fliI) flagellum-specific ATP synthase FliI [Flagella (VF0273) - Motility (VFC0204
VFG041257(gb|WP_010946432) (ankN/ankX/legA8) Dot/Icm Type IV secretion system effector AnkX ankyrin
VFG034922(gb|WP_001303732) (espY5) hypothetical protein [TTSS secreted effectors (VF1110) - Effector deli
VFG034901(gb|WP_000868321) (espY2) Type III secretion system effector EspY2 [TTSS secreted effectors (V
VFG038561(gb|WP_016349168) (mshQ) MSHA (mannose-sensitive hemagglutinin) pili biogenesis protein Msh
VFG000145(gb|NP_250562) (lasA) LasA protease precursor [LasA (VF0088) - Effector delivery system (VFC0
VFG037678(gb|WP_000866237) (pgaC) poly-beta-1,6 N-acetyl-D-glucosamine synthase [PNAG (VF0472) - Bi
VFG001814(gb|NP_215678) (narH) nitrate reductase subunit beta [Nitrate reductase (VF0302) - Nutritional/Met
VFG000091(gb|WP_001176374) (tcpA) toxin-coregulated pilus (TCP) major pilin TcpA [TCP (VF0126) - Adhe
VFG002190(gb|WP_002362225) (cpsA/uppS) undecaprenyl diphosphate synthase [Capsule (VF0361) - Immune
VFG038812(gb|WP_011706645) (cheR-3) chemotaxis protein CheR [Polar flagella (VF0473) - Motility (VFC02
VFG038374(gb|WP_011705709) (atsB) Type VI secretion system protein [T6SS (VF0480) - Effector delivery sy
VFG000922(gb|WP_000910402) (chuU) heme permease protein ChuU [Chu (VF0227) - Nutritional/Metabolic f
VFG001266(gb|NP_252912) (pchI) ABC transporter ATP-binding protein [Pyochelin (VF0095) - Nutritional/Me
VFG034772(gb|WP_000627910) (espL4) Type III secretion system effector EspL4 [TTSS secreted effectors (VF
VFG045545(gb|WP_010947529) (LPG_RS09040) Dot/Icm type IV secretion system effector [Dot/Icm T4SS sec
VFG038373(gb|WP_011705708) (atsA) Type VI secretion system protein [T6SS (VF0480) - Effector delivery sy
VFG016038(gb|NP_251090) (pvdJ) pyoverdine biosynthesis protein PvdJ [Pyoverdine (VF0094) - Nutritional/M
VFG000902(gb|WP_001034089) (sat) Aecreted auto transpoter toxin [Sat (VF0231) - Effector delivery system (
VFG048468(gb|WP_004176917) (fepD) iron-enterobactin transporter membrane protein [Ent (VF0562) - Nutriti
VFG000894(gb|WP_001239363) (papH) P pilus termination subunit PapH [P fimbriae (VF0220) - Adherence (V
VFG047137(gb|WP_003022294) (wzy) membrane protein [LPS (VF0542) - Immune modulation (VFC0258)] [F
VFG038501(gb|WP_043119142) (tapV) type IV pilus assembly protein TapV [Tap type IV pili (VF0475) - Adhe
VFG048632(gb|WP_004176434) (vipA/tssB) type VI secretion system contractile sheath small subunit VipA [T
VFG042390(gb|WP_011478152) (sfaX) regulatory protein SfaX [S fimbriae (VF0222) - Adherence (VFC0001)]
VFG048439(gb|WP_014907941) (entC) isochorismate synthase [Ent (VF0562) - Nutritional/Metabolic factor (V
VFG038647(gb|WP_016350053) (fliK) flagellar hook-length control protein FliK [Polar flagella (VF0473) - Mot
VFG002182(gb|WP_002376666) (cpsI) UDP-galactopyranose mutase [Capsule (VF0361) - Immune modulation
VFG002458(gb|WP_004525314) (bsaT) Type III secretion system protein BsaT [Bsa T3SS (VF0428) - Effector
VFG048317(gb|WP_014906959) (fimK) transcriptional regulator [Type I fimbriae (VF0566) - Adherence (VFC0
VFG002548(gb|WP_004550042) (wcbR) capsular polysaccharide biosynthesis fatty acid synthase [Capsule I (VF
VFG041429(gb|WP_005771199) (CBUK_RS06760) ankyrin repeat domain-containing protein [T4SS secreted e
VFG038375(gb|WP_011705710) (atsC) Type VI secretion system protein [T6SS (VF0480) - Effector delivery sy

VFG035909(gb|WP_001216023) (aatA) outer membrane protein AatA [Dispersin (VF0215) - Others (VFC0346)
VFG038376(gb|WP_164927604) (atsD) Type VI secretion system protein [T6SS (VF0480) - Effector delivery sy
VFG049909(gb|WP_001113715) (fha) Type VI secretion system protein Fha [T6SS (VF0579) - Effector delivery
VFG000144(gb|NP_249940) (aprA) alkaline metalloproteinase precursor [Alkaline protease (VF0090) - Exoenzy
VFG001296(gb|WP_000676533) (sspA) serine protease; V8 protease; glutamyl endopeptidase [V8 protease (VF0
VFG037897(gb|WP_001165095) (tviB) Vi polysaccharide biosynthesis UDP-N-acetylglucosamine C-6 dehydrog
VFG048687(gb|WP_002902160) (hcp/tssD) type VI secretion system protein, Hcp family [T6SS (VF0569) - Effe
VFG050158(gb|WP_000905916) (kpsE) Capsule polysaccharide export inner-membrane protein [K1 capsule (VF
VFG000873(gb|WP_000695571) (fimA) Type-1 fimbrial protein, A chain precursor [Type 1 fimbriae (VF0221) -
VFG005930(gb|WP_000591736) (GBS_RS06565) glycosyltransferase [Capsule (VF0274) - Immune modulation
VFG007157(gb|WP_005464652) (vopN) type III secretion system protein VopN [T3SS1 (VF0408) - Effector del
VFG001443(gb|AAF37887) (ompA) outer membrane protein A [OmpA (VF0236) - Invasion (VFC0083)] [Esche
VFG001295(gb|WP_001089097) (sspB) staphopain cysteine proteinase SspB [Staphopain (VF0006) - Exoenzym
VFG047529(gb|WP_003029409) (bioA) adenosylmethionine-8-amino-7-oxononanoate aminotransferase [Biotin
VFG034815(gb|WP_000671689) (espR1) Type III secretion system effector espR1 [TTSS secreted effectors (VF
VFG043774(gb|AAB24244) (bont) botulinum neurotoxin, BoNT [BoNT (VF0375) - Exotoxin (VFC0235)] [Clos
VFG048498(gb|WP_012737619) (entF) enterobactin synthase subunit F [Ent (VF0562) - Nutritional/Metabolic fa
VFG044331(gb|WP_015874966) (ybtT) type II thioesterase YbtT [Ybt (VF0564) - Nutritional/Metabolic factor (
VFG000208(gb|NP_250404) (exsA) type III secretion system regulatory protein ExsA [TTSS (VF0083) - Effecto
VFG038399(gb|WP_011705723) (atsP) Type VI secretion system protein [T6SS (VF0480) - Effector delivery sy
VFG016472(gb|WP_011206327) (p216) protein P216, cilium adhesin p97 paralog [P97/P102 paralog family (VF
VFG024724(gb|NP_217463) (pks15) polyketide synthase [PDIM (VF0309) - Immune modulation (VFC0258)] [
VFG001375(gb|WP_000594468) (cps4K) capsular polysaccharide biosynthesis protein Cps4K [Capsule (VF014
VFG001281(gb|WP_000610231) (sdrE) Ser-Asp rich fibrinogen-binding bone sialoprotein-binding protein [SDr
VFG048528(gb|WP_004183335) (entD) enterochelin synthetase component D [Ent (VF0562) - Nutritional/Metal
VFG051244(gb|WP_004683766.1) (BPE043) type IV secretion system effector [T4SS secreted effectors (VF069
VFG002531(gb|WP_011205291) (cheA) chemotaxis two-component sensor kinase CheA [Flagella (VF0430) - M
VFG049150(gb|WP_000982270) (clbD) colibactin biosynthesis dehydrogenase ClbD [Colibactin (VF0573) - Exo
VFG000871(gb|WP_000790574) (fimB) Type 1 fimbriae Regulatory protein fimB [Type 1 fimbriae (VF0221) -
VFG002288(gb|WP_009902069) (toxB) toxin B [TcdB (VF0377) - Exotoxin (VFC0235)] [Clostridium difficile 6
VFG001280(gb|WP_000934435) (sdrD) Ser-Asp rich fibrinogen-binding bone sialoprotein-binding protein [SDr
VFG002526(gb|WP_004198645) (cheB) chemotaxis-specific methylesterase [Flagella (VF0430) - Motility (VFC
VFG043625(gb|WP_000813718) (mrkC) fimbrial biogenesis outer membrane usher protein mrkC precursor [Typ
VFG041099(gb|NP_459273) (STM0275) putative cytoplasmic protein [SCI (Salmonella centrisome island) (VF0
VFG000939(gb|WP_001287497) (iucB) aerobactin siderophore biosynthesis protein IucB [Aerobactin (VF0229)
VFG039131(gb|NP_820609) (icmE/dotG) Type IVB secretion system protein IcmE/DotG, inner membrane prote
VFG001857(gb|WP_010946483) (iraB) Di/tripeptide permease [IraAB (VF0293) - Nutritional/Metabolic factor (
VFG050567(gb|WP_000768964.1) (pilY1) VWA domain-containing protein [TFP (VF1334) - Adherence (VFC0
VFG045794(gb|WP_000992818) (csgC) curli assembly protein CsgC [Curli fibers (VF1138) - Adherence (VFC0
VFG000917(gb|WP_000089583) (chuA) Outer membrane heme/hemoglobin receptor ChuA [Chu (VF0227) - Nu
VFG038756(gb|WP_011706626) (flrC) transcriptional activator [Polar flagella (VF0473) - Motility (VFC0204)]
VFG002179(gb|AAD09858) (esp) Enterococcal surface protein; Esp [Esp (VF0353) - Adherence (VFC0001)] [E
VFG049073(gb|WP_015875021) (KP1_RS17240) DUF4422 domain-containing protein [LPS (VF0561) - Immun
VFG009561(gb|YP_177877) (mbtI) Isochorismate synthase MbtI [Mycobactin (VF0299) - Nutritional/Metabolic
VFG002164(gb|WP_002395058) (prgB/asc10) aggregation substance PrgB/Asc10 [AS (VF0352) - Adherence (V
VFG001263(gb|NP_250143) (flhA) flagellar biosynthesis protein FlhA [Flagella (VF0273) - Motility (VFC0204)

VFG002166(gb|WP_010714416) (ace) collagen adhesin protein [Ace (VF0355) - Adherence (VFC0001)] [Enterc
VFG001237(gb|NP_249770) (flgD) flagellar basal-body rod modification protein FlgD [Flagella (VF0273) - Mot
VFG000002(gb|WP_010930159) (prn) pertactin precursor [Pertactin (VF0027) - Adherence (VFC0001)] [Bordet
VFG038753(gb|WP_024944879) (motY) sodium-type flagellar protein MotY [Polar flagella (VF0473) - Motility
VFG000983(gb|WP_000517827) (rtxA) RTX toxin RtxA [MARTX (VF0265) - Exotoxin (VFC0235)] [Vibrio ch
VFG048620(gb|WP_014907843) (iutA) ferric aerobactin receptor IutA [Aerobactin (VF0565) - Nutritional/Metal
VFG002279(gb|WP_003467250) (nagJ) hyaluronidase [Mu-toxin (VF0389) - Exoenzyme (VFC0251)] [Clostridi
VFG050161(gb|WP_000579525) (kpsC) Capsule polysaccharide export protein [K1 capsule (VF0239) - Invasion
VFG000907(gb|WP_000376545) (hlyB) Hemolysin B [-Hemolysin (VF0225) - Exotoxin (VFC0235)] [Escherich
VFG011757(gb|WP_011117594) (virB4) Type IV secretion system protein VirB4 [Type IV secretion system (VF
VFG001926(gb|YP_002344844) (flgI) flagellar P-ring protein precursor FlgI [Flagella (VF0114) - Motility (VFC
VFG049906 (tssK) Type VI secretion system protein TssK [T6SS (VF0579) - Effector delivery system (VFC008
VFG013647(gb|WP_041175806) (invC/ilp) intimin-like invasin C protein [Invasin C/Ilp (VF0575) - Invasion (V
VFG045349(gb|WP_000092542) (fdeC) adhesin FdeC [FdeC (VF0506) - Adherence (VFC0001)] [Escherichia co
VFG002276(gb|WP_011009653) (colA) collagenase [Kappa-toxin (VF0388) - Exoenzyme (VFC0251)] [Clostrid
VFG002360(gb|WP_042661486) (YE_RS15405) O-antigen chain length determinant [O-antigen (VF0392) - Imn
VFG038683(gb|WP_016350056) (flhA) flagellar biosynthesis protein FlhA [Polar flagella (VF0473) - Motility (V
VFG001243(gb|NP_249776) (flgJ) flagellar rod assembly protein/muramidase FlgJ [Flagella (VF0273) - Motility
VFG001269(gb|WP_010929996) (cyaB) cyclolysin secretion ATP-binding protein [Cya (VF0028) - Exotoxin (V
VFG045791(gb|WP_000481500) (cgsD) transcriptional regulator CsgD [Curli fibers (VF1138) - Adherence (VF
VFG012505(gb|WP_000933673) (iroD) esterase [Salmochelin siderophore (VF0230) - Nutritional/Metabolic fac
VFG002515(gb|WP_004550967) (flgI) flagellar P-ring protein precursor FlgI [Flagella (VF0430) - Motility (VFC
VFG044079(gb|NP_250944) (pvcA) paerucumarin biosynthesis protein PvcA [Pyoverdine (VF0094) - Nutritiona
VFG049095(gb|WP_002912373) (rfbA) O-antigen export ABC transporter permease RfbA [LPS (VF0561) - Imn
VFG002412(gb|WP_000716386) (yagY/ecpB) E. coli common pilus chaperone EcpB [ECP (VF0404) - Adheren
VFG000368(gb|WP_000703040) (ybtS) salicylate synthase Irp9 [Yersiniabactin (VF0136) - Nutritional/Metaboli
VFG001213(gb|NP_253236) (pilS) two-component sensor PilS [Type IV pili (VF0082) - Adherence (VFC0001)]
VFG000903(gb|WP_001045652) (pic) Pic serine protease precursor, autotransporter [Pic (VF0232) - Effector del
VFG002194(gb|WP_011109506) (fsrA) response regulator [Fsr (VF0360) - Biofilm (VFC0271)] [Enterococcus f
VFG038391(gb|WP_011705719) (atsL) Type VI secretion system protein [T6SS (VF0480) - Effector delivery sy
VFG000123(gb|NP_252231) (alg8) alginate-c5-mannuronan-epimerase AlgG [Alginate (VF0091) - Biofilm (VF
VFG038675(gb|WP_011705283) (fliR) flagellar biosynthesis protein FliR [Polar flagella (VF0473) - Motility (V
VFG005303(gb|WP_000836217) (rrgB) cell wall surface anchor family protein, major pilin RrgB [RlrA islet (VF
VFG048488(gb|WP_002893737) (fepC) iron-enterobactin transporter ATP-binding protein [Ent (VF0562) - Nutr
VFG038436(gb|WP_016352169) (tapC) type IV fimbrial assembly protein, PilC-like [Tap type IV pili (VF0475)
VFG000184(gb|NP_251795) (xcpQ) general secretion pathway protein D [Xcp secretion system (VF0084) - Effe
VFG000915(gb|WP_001234179) (focH) F1C Putative fimbrial adhesin precursor [F1C fimbriae (VF0224) - Adhe
VFG016046(gb|NP_251103) (pvdH) diaminobutyrate-2-oxoglutarate aminotransferase PvdH [Pyoverdine (VF00
VFG015551(gb|NP_248741) (phzH) phenazine-modifying enzyme [Pyocyanin (VF0100) - Nutritional/Metabolic
VFG041327(gb|WP_010948034) (lem22) Dot/Icm type IV secretion system effector Lem22 [Dot/Icm T4SS secre
VFG002186(gb|WP_002387129) (cpsE) glycosyl transferase, group 2 family protein [Capsule (VF0361) - Immur
VFG022130(gb|NP_214804) (eccD3) Type IIV secretion system transmembrane protein EccD3 [ESX-3 (VF0521
VFG000927(gb|WP_000096764) (fepE) LPS O-antigen length regulator [Enterobactin (VF0228) - Nutritional/M
VFG038875(gb|WP_005314261) (lfgF) lateral flagellar basal-body rod protein LfgF [Lateral flagella (VF0474) -
VFG048409(gb|WP_086074289) (entA) 2,3-dihydroxybenzoate-2,3-dehydrogenase [Ent (VF0562) - Nutritional/
VFG038659(gb|WP_011705280) (fliN) flagellar motor switch protein FliN [Polar flagella (VF0473) - Motility (V

VFG001960(gb|YP_002344815) (Cj1432c) sugar transferase [Capsule (VF0323) - Immune modulation (VFC025
VFG002105(gb|WP_010947915) (wipC) Dot/Icm type IV secretion system effector WipC [Dot/Icm (VF0156) -
VFG038768(gb|WP_016351318) (flgL) flagellar hook-associated protein FlgL [Polar flagella (VF0473) - Motilit
VFG042977(gb|WP_002379309) (ebpB) endocarditis and biofilm-associated pilus minor subunit EbpB [Ebp pili
VFG048287(gb|WP_014906962) (fimF) type 1 fimbrial minor component [Type I fimbriae (VF0566) - Adherenc
VFG002395(gb|YP_007410461) (eccC5) ESX-5 type VII secretion system FtsK/SpoIIIE family protein EccC5 [
VFG048606(gb|WP_004213922) (iucC) aerobactin siderophore biosynthesis protein IucC [Aerobactin (VF0565)
VFG000171(gb|NP_252921) (pchA) salicylate biosynthesis isochorismate synthase PchA [Pyochelin (VF0095) -
VFG000879(gb|WP_000832235) (fimH) FimH protein precursor [Type 1 fimbriae (VF0221) - Adherence (VFC0
VFG000364(gb|WP_000140406) (ybtA) transcriptional regulator YbtA [Yersiniabactin (VF0136) - Nutritional/M
VFG002413(gb|WP_000389022) (ykgK/ecpR) regulator protein EcpR [ECP (VF0404) - Adherence (VFC0001)]
VFG049040(gb|WP_074160401) (KP1_RS17225) glycosyltransferase family 4 protein [LPS (VF0561) - Immune
VFG048665(gb|WP_004176431) (dotU/tssL) type VI secretion system protein, DotU/TssL family [T6SS (VF056
VFG047095(gb|WP_003014989) (wbtI) DegT/DnrJ/EryC1/StrS family aminotransferase [LPS (VF0542) - Immu
VFG002417(gb|WP_001301550) (yagV/ecpE) E. coli common pilus chaperone EcpE [ECP (VF0404) - Adherenc
VFG048111(gb|WP_014367985) (iglC/hcp) Type VI secretion system tube protein, Hcp homolog [T6SS (VF054
VFG000889(gb|WP_000113351) (papF) P pilus minor subunit PapF [P fimbriae (VF0220) - Adherence (VFC00
VFG002173(gb|WP_010785625) (asa1) aggregation substance Asa1 [AS (VF0352) - Adherence (VFC0001)] [E
VFG000921(gb|WP_000189356) (chuY) ChuY [Chu (VF0227) - Nutritional/Metabolic factor (VFC0272)] [Esch
VFG000874(gb|WP_000824100) (fimI) Fimbrin-like protein fimI precursor [Type 1 fimbriae (VF0221) - Adhere
VFG038383(gb|WP_011705715) (atsH) Type VI secretion system protein [T6SS (VF0480) - Effector delivery sy
VFG044199(gb|WP_003022888) (fslA) IucA/IucC family siderophore biosynthesis protein [Rhizoferrin (VF055
VFG042071(gb|WP_010930810) (BP_RS11155) hypothetical protein [TTSS (VF0035) - Effector delivery system
VFG000231 (pilC) pilus assembly protein [Type IV pili (VF0075) - Adherence (VFC0001)] [Neisseria meningiti
VFG038448(gb|WP_016351632) (tapN) type IV pilus biogenesis protein PilN [Tap type IV pili (VF0475) - Adhe
VFG049162(gb|WP_002430641) (clbP) precolibactin peptidase ClbP [Colibactin (VF0573) - Exotoxin (VFC023
VFG000918(gb|WP_001312167) (chuT) periplasmic heme-binding protein ChuT [Chu (VF0227) - Nutritional/M
VFG007946(gb|NP_217473) (Rv2957) PGL/p-HBAD biosynthesis glycosyltransferase [PDIM (VF0309) - Immu
VFG002220(gb|WP_004686741) (pgm) phosphoglucomutase [LPS (VF0367) - Immune modulation (VFC0258)]
VFG044465(gb|WP_005478084) (vopZ) type III secretion system effector VopZ [T3SS2 (VF0409) - Effector del
VFG000011(gb|WP_010931648) (ptxA) pertussis toxin subunit 1 precursor [Ptx (VF0026) - Exotoxin (VFC0235
VFG044333(gb|WP_015874968) (fyuA) yersiniabactin receptor FyuA [Ybt (VF0564) - Nutritional/Metabolic fac
VFG012603(gb|WP_001302516) (chuT) periplasmic heme-binding protein ChuT [Chu (VF0234) - Nutritional/M
VFG000906(gb|WP_001142375) (hlyA) Hemolysin A [-Hemolysin (VF0225) - Exotoxin (VFC0235)] [Escherich
VFG038401(gb|WP_011705724) (atsQ) Type VI secretion system protein [T6SS (VF0480) - Effector delivery sy
VFG048508(gb|WP_012737618) (fes) enterobactin/ferric enterobactin esterase [Ent (VF0562) - Nutritional/Meta
VFG049159(gb|WP_014907260) (clbM) precolibactin export MATE transporter ClbM [Colibactin (VF0573) - E
VFG049898(gb|WP_001284199) (hcp1/tssD1) type VI secretion system protein, Hcp family [T6SS (VF0579) - E
VFG001380(gb|NP_214946) (sodC) superoxide dismutase [SodC (VF0305) - Stress survival (VFC0282)] [Myco
VFG038317(gb|WP_016352170) (tapD/pilD) prepilin peptidase [Tap type IV pili (VF0475) - Adherence (VFC00
VFG000139(gb|NP_253697) (waaG) B-band O-antigen polymerase [LPS (VF0085) - Immune modulation (VFC
VFG002056 (gspM) general secretion pathway protein M [T2SS (VF0333) - Effector delivery system (VFC0086
VFG044326(gb|WP_001327262) (ybtP) yersiniabactin ABC transporter ATP-binding/permease protein YbtP [Y
VFG000985(gb|NP_858259) (ipaD) type III secretion system hydrophilic translocator, needle tip protein IpaD [T
VFG000358(gb|WP_002212885) (fyuA/psn) pesticin/yersiniabactin receptor protein [Yersiniabactin (VF0136) -
VFG038707(gb|WP_016350059) (cheA-2) chemotaxis protein CheA [Polar flagella (VF0473) - Motility (VFC02

VFG002051(gb|WP_001115126) (gspH) general secretion pathway protein H [T2SS (VF0333) - Effector delivery
VFG038576(gb|WP_016349164) (mshC) MSHA (mannose-sensitive hemagglutinin) pili minor prepilin protein M
VFG012095(gb|WP_003435012) (groEL) chaperonin GroEL [GroEL (VF0594) - Adherence (VFC0001)] [Clostr
VFG034875(gb|WP_001270087) (espX5) Type III secretion system effector EspX5 [TTSS secreted effectors (VF
VFG045818(gb|WP_001528154) (tsh) temperature-sensitive protein Tsh [Tsh (VF0233) - Effector delivery syster
VFG002189(gb|WP_002359680) (cpsB/cdsA) phosphatidate cytidylyltransferase [Capsule (VF0361) - Immune m
VFG034754(gb|WP_000081129) (espL1) Type III secretion system effector espL1 [TTSS secreted effectors (VF
VFG045788(gb|WP_001189322) (cgsG) curli production assembly/transport protein CsgG [Curli fibers (VF1138
VFG050157(gb|WP_001331698) (kpsF) Polysialic acid capsule expression protein [K1 capsule (VF0239) - Invas
VFG012341(gb|WP_000767895) (sfaS) S fimbriae minor subunit SfaS [S fimbriae (VF0222) - Adherence (VFC0
VFG038464(gb|WP_011707389) (tapT) twitching ATPase [Tap type IV pili (VF0475) - Adherence (VFC0001)]
VFG002148(gb|NP_220192) (pkn5) serine/threonine protein kinase [TTSS secreted effectors (VF0711) - Effector
VFG038851(gb|WP_016350359) (nueB) pseudaminic acid synthase, NeuB [Polar flagella (VF0473) - Motility (V
VFG002052(gb|WP_000820136) (gspI) general secretion pathway protein I [T2SS (VF0333) - Effector delivery s
VFG000872(gb|WP_000044711) (fimE) Type 1 fimbriae Regulatory protein fimE [Type 1 fimbriae (VF0221) - A
VFG038568(gb|WP_016349166) (mshO) MSHA (mannose-sensitive hemagglutinin) pili biogenesis protein Msh
VFG017841(gb|WP_012105808) (pilQ) type IV pilus biogenesis protein PilQ [Type IV pili (VF0401) - Adherenc
VFG038913 (rtxA) multifunctional autoprocessing repeats-in-toxins (MARTX) toxin [RtxA (VF0482) - Exotoxin
VFG048755(gb|WP_049127998) (icmF/tssM) type VI secretion protein TssM [T6SS (VF0569) - Effector deliver
VFG000180(gb|NP_251791) (xcpT) general secretion pathway protein G [Xcp secretion system (VF0084) - Effec
VFG002278(gb|WP_011010095) (nagI) hyaluronidase [Mu-toxin (VF0389) - Exoenzyme (VFC0251)] [Clostridi
VFG002434(gb|WP_011205650) (pilN) type IV pilus inner membrane platform protein PilN [Type IV pili (VF04
VFG001007(gb|NP_858281) (spa15) type III secretion system chaparone [TTSS (VF0118) - Effector delivery sys
VFG041278(gb|WP_010947155) (vpdC) Dot/Icm type IV secretion system effector VpdC [Dot/Icm T4SS secrete
VFG050802(gb|WP_000865310.1) (gspG) general secretion pathway protein G [T2SS (VF1335) - Effector delive
VFG012615(gb|WP_000020049) (chuX) putative heme-binding protein ChuX [Chu (VF0234) - Nutritional/Meta
VFG038403(gb|WP_011705726) (atsS) Type VI secretion system protein [T6SS (VF0480) - Effector delivery sys
VFG002072(gb|NP_248776) (hsiE1) type VI secretion system hcp secretion island protein HsiE1, interacting wit
VFG000114(gb|NP_253218) (xcpA/pilD) type 4 prepilin peptidase PilD [Type IV pili (VF0082) - Adherence (VF
VFG002195(gb|WP_002399773) (EF0818) polysaccharide lyase, family 8 [Hyaluronidase (VF0359) - Exoenzym
VFG038635(gb|WP_016350051) (fliH) flagellar assembly protein FliH [Polar flagella (VF0473) - Motility (VFC
VFG038505(gb|WP_011705643) (tapF) type IV pilus biogenesis/stability protein, pilW-like [Tap type IV pili (VF
VFG044330(gb|WP_032447695) (ybtU) yersiniabactin biosynthesis oxidoreductase YbtU [Ybt (VF0564) - Nutri
VFG041021(gb|NP_250354) (PA1663) transcriptional regulator [HSI-2 (VF0943) - Effector delivery system (VF
VFG038687(gb|WP_016350057) (flhF) flagellar biosynthesis protein FlhF [Polar flagella (VF0473) - Motility (V
VFG044084(gb|NP_251073) (PA2383) transcriptional regulator [Pyoverdine (VF0094) - Nutritional/Metabolic fa
VFG050245(gb|NP_219732) (CT_227) inclusion membrane protein [TTSS secreted effectors (VF0711) - Effecto
VFG022051(gb|NP_214798) (eccC3) Type VII secretion system protein EccC3 [ESX-3 (VF0521) - Effector deli
VFG038342(gb|WP_005320770) (ascV) Type III secretion system inner membrane export apparatus protein Asc
VFG000615(gb|NP_709454) (iucA) aerobactin synthesis protein IucA [Aerobactin (VF0123) - Nutritional/Metab
VFG038387(gb|WP_011705717) (atsJ) Type VI secretion system protein [T6SS (VF0480) - Effector delivery sys
VFG000545(gb|NP_461805) (sipC/sspC) type III secretion system hydrophilic translocator, pore protein SipC [T
VFG049155(gb|WP_000829570) (clbI) colibactin polyketide synthase ClbI [Colibactin (VF0573) - Exotoxin (VF
VFG047860(gb|WP_003022130) (iglF/clpV) Type VI secretion system secreted protein IglF [T6SS (VF0544) - E
VFG000165(gb|NP_252915) (pchF) pyochelin synthetase PchF [Pyochelin (VF0095) - Nutritional/Metabolic fact
VFG038497(gb|WP_016351992) (tppF) type IV pilus pseudopilin [Tap type IV pili (VF0475) - Adherence (VFC

VFG002438(gb|WP_011205635) (bprC) AraC family transcriptional regulator, regulates basal T6SS1 expression
VFG034849(gb|WP_000129338) (espX1) Type III secretion system effector EspX1 [TTSS secreted effectors (VI
VFG039952(gb|WP_011190507) (rvhB6a) VirB6-like protein of the type IV secretion system [Rvh T4SS (VF04
VFG038651(gb|WP_011705278) (fliL) flagellar basal body-associated protein FliL [Polar flagella (VF0473) - Mo
VFG001943(gb|YP_002344535) (neuC1) UDP-N-acetylglucosamine 2-epimerase [LOS (VF0326) - Immune mod
VFG044172(gb|WP_000622316) (chuV) ATP-binding hydrophilic protein ChuV [Chu (VF0227) - Nutritional/M
VFG034624(gb|WP_000755097) (cfaB) colonization factor antigen 1 [Adhesive fimbriae (VF0213) - Adherence
VFG045296(gb|WP_164919819) (LPG_RS03710) N-acetyl sugar amidotransferase [LPS (VF0171) - Immune mo
VFG041118(gb|WP_045606108) (tssA-5) Type VI secretion system protein TssA [T6SS-1 (VF0429) - Effector d
VFG000938(gb|WP_001015709) (iucC) aerobactin siderophore biosynthesis protein IucC [Aerobactin (VF0229)
VFG000618(gb|NP_709457) (iucD) aerobactin synthesis protein IucC, lysine 6-monooxygenase [Aerobactin (VF
VFG038176(gb|WP_000777882) (ompA) outer membrane protein OmpA [OmpA (VF0464) - Immune modulatio
VFG012591(gb|WP_001017167) (chuS) heme oxygenase ChuS [Chu (VF0234) - Nutritional/Metabolic factor (V
VFG001354(gb|WP_000728269) (pce/cbpE) choline binding protein E [CBPs (VF0145) - Adherence (VFC0001)
VFG002424(gb|WP_011205626) (bimA) YadA family of bacterial autotransporter mediating actin tail formation
VFG044327(gb|WP_000140406) (ybtA) yersiniabactin transcriptional regulator YbtA [Ybt (VF0564) - Nutritiona
VFG044159(gb|WP_000125846) (fes) enterobactin/ferric enterobactin esterase [Enterobactin (VF0228) - Nutritio
VFG037807(gb|WP_000013434) (lpxB) lipid-A-disaccharide synthase [LPS (VF0466) - Immune modulation (VI
VFG012621(gb|WP_000189352) (chuY) ChuY [Chu (VF0234) - Nutritional/Metabolic factor (VFC0272)] [Esch
VFG001408(gb|NP_217456) (mas) multifunctional mycocerosic acid synthase [PDIM (VF0309) - Immune modu
VFG039121(gb|NP_820607) (icmC/dotE) Type IVB secretion system protein IcmC [Dot/Icm T4SS (VF0503) - E
VFG012597(gb|WP_000089593) (chuA) Outer membrane heme/hemoglobin receptor ChuA [Chu (VF0234) - Nu
VFG001267(gb|NP_252913) (pchH) ABC transporter ATP-binding protein [Pyochelin (VF0095) - Nutritional/M
VFG002269(gb|WP_011181342) (trwD) Trw type IV secretion sysmtem VirB11 homolog, trwD protein [Trw typ
VFG051134(gb|WP_002969669.1) (btaE) Trimeric autotransporter adhesin, YadA-like family protein [BtaE (VF
VFG038703(gb|WP_011705289) (cheZ) chemotaxis protein CheZ [Polar flagella (VF0473) - Motility (VFC0204
VFG002282(gb|WP_041707714) (cloSI) alpha-clostripain [Alpha-clostripain (VF0390) - Exoenzyme (VFC0251)
VFG046628(gb|WP_003020689) (FTT_RS04105) sugar transferase [Capsule (VF0543) - Immune modulation (V
VFG001409(gb|NP_217457) (fadD28) long-chain-fatty-acid--AMP ligase FadD28 [PDIM (VF0309) - Immune m
VFG034648(gb|WP_001245988) (cfaD/cfaE) minor pilin and initiator [Adhesive fimbriae (VF0213) - Adherence
VFG048277(gb|WP_009308599) (fimD) outer membrane usher protein [Type I fimbriae (VF0566) - Adherence
VFG038395(gb|WP_011705721) (clpB) type VI secretion system ATPase ClpV1 [T6SS (VF0480) - Effector deli
VFG000348(gb|WP_005694042) (hxuC) heme-hemopexin utilization protein C, haem receptor [HxuABC (VF02
VFG043627(gb|WP_000756959) (mrkA) type 3 fimbrial major pilin subunit MrkA [Type 3 fimbriae (VF0567) -
VFG042566(gb|WP_011076307) (sfaC) S/F1C fimbrial major subunit operon transcriptional regulator [F1C fimb
VFG049118(gb|WP_012737529) (allR) DNA-binding transcriptional repressor AllR [Allantion utilization (VF05
VFG038804(gb|WP_011706643) (flgC) flagellar basal body rod protein FlgC [Polar flagella (VF0473) - Motility
VFG013070(gb|WP_001626196) (shuV) ATP-binding component of hemin transport system [Shu (VF0256) - Nu
VFG011671(gb|YP_002344801) (Cj1418c) hypothetical protein [Capsule (VF0323) - Immune modulation (VFC
VFG050169(gb|WP_000124301) (kpsM) Polysialic acid transport protein [K1 capsule (VF0239) - Invasion (VFC
VFG038631(gb|WP_011705273) (fliG) flagellar motor switch protein G [Polar flagella (VF0473) - Motility (VFC
VFG041025(gb|NP_250358) (PA1667) hypothetical protein [HSI-2 (VF0943) - Effector delivery system (VFC00
VFG038047(gb|WP_000994240) (ACICU_RS00460) hypothetical protein [Capsule (VF0465) - Immune modula
VFG045459 (APH_RS05665*) P44/Msp2 family outer membrane protein [P44/Msp2 family (VF0520) - Immune
VFG051111(gb|YP_001084320.1) (tse3) T6SS effector Tse3, unknown function [T6SS secreted effectors (VF13
VFG045635(gb|WP_002296629) (sgrA) cell wall anchored protein SgrA [SgrA (VF0540) - Adherence (VFC000

VFG043014(gb|NP_250149) (PA1458) two-component sensor [Flagella (VF0273) - Motility (VFC0204)] [Pseud
VFG001231(gb|NP_249104) (chpA) still frameshift probable component of chemotactic signal transduction syste
VFG000940(gb|WP_001299620) (iucA) aerobactin siderophore biosynthesis protein IucD [Aerobactin (VF0229)
VFG002096(gb|WP_010946675) (lidA) Dot/Icm type IV secretion system effector LidA, interacting with Rab G
VFG002414(gb|WP_000730972) (yagZ/ecpA) E. coli common pilus structural subunit EcpA [ECP (VF0404) - A
VFG038397(gb|WP_011705722) (vasH) sigma-54 dependent transcriptional regulator [T6SS (VF0480) - Effecto
VFG000192(gb|NP_250388) (pscN) type III secretion system ATPase PscN [TTSS (VF0083) - Effector delivery
VFG037650 (pgaA) poly-beta-1,6 N-acetyl-D-glucosamine export porin PgaA [PNAG (VF0472) - Biofilm (VFC
VFG038369(gb|WP_011705706) (hcp) hemolysin co-regulated protein [T6SS (VF0480) - Effector delivery syste
VFG043392(gb|YP_002343698) (eptC) phosphoethanolamine transferase [Flagella (VF0114) - Motility (VFC02
VFG000926(gb|WP_001443194) (fepD) ferrienterobactin ABC transporter permease [Enterobactin (VF0228) - N
VFG001445(gb|AAA92657) (AAA92657) unknown protein [TraJ (VF0241) - Invasion (VFC0083)] [Escherichia
VFG050037(gb|WP_001133943) (dhbF) non-ribosomal peptide synthetase, DhbF [Bacillibactin (VF0586) - Nutr
VFG048771(gb|WP_004190505) (tssF) type VI secretion system baseplate subunit TssF [T6SS (VF0569) - Effec
VFG045632(gb|WP_002286924) (ecbA/fss3) Collagen binding MSCRAMM, EcbA/Fss3 [EcbA (VF0539) - Adh
VFG002045(gb|WP_010930608) (bvgS) virulence factors two-component system sensor histidine kinase BvgS [
VFG000876(gb|WP_000120946) (fimD) Outer membrane usher protein fimD precursor [Type 1 fimbriae (VF02
VFG000931(gb|WP_001336900) (entC) isochorismate synthase 1 [Enterobactin (VF0228) - Nutritional/Metaboli
VFG010749(gb|WP_010945896) (sdhB) Dot/Icm type IV secretion system effector SdhB [Dot/Icm T4SS secrete
VFG034679(gb|WP_000074207) (ibeB) Cu(+)/Ag(+) efflux RND transporter outer membrane channel CusC [Ib
VFG037664(gb|WP_001061322) (pgaB) poly-beta-1,6-N-acetyl-D-glucosamine N-deacetylase PgaB [PNAG (V
VFG000163(gb|NP_251088) (fpvA) ferripyoverdine receptor FpvA [Pyoverdine (VF0094) - Nutritional/Metabol
VFG048797(gb|WP_004175261) (ugd) UDP-glucose 6-dehydrogenase [Capsule (VF0560) - Immune modulation
VFG002507(gb|WP_004534277) (flgA) flagellar basal body P-ring biosynthesis protein FlgA [Flagella (VF0430
VFG002070(gb|NP_248774) (hsiC1/vipB/tssC) type VI secretion system tubule-forming protein VipB [HSI-I (V
VFG050625(gb|WP_000389061.1) (pilG) twitching motility response regulator PilG [TFP (VF1334) - Adherenc
VFG050670(gb|WP_000505931.1) (pilJ) methyl-accepting chemotaxis protein [TFP (VF1334) - Adherence (VF
VFG002159(gb|NP_464601) (aut) autolysin [Auto (VF0348) - Invasion (VFC0083)] [Listeria monocytogenes E
VFG038409(gb|WP_011704492) (fimA) fimbrial protein [Type I pili (VF0486) - Adherence (VFC0001)] [Aerom
VFG042568(gb|WP_000012016) (sfaY) transcriptional regulator SfaY [F1C fimbriae (VF0224) - Adherence (VF
VFG049904 (clpV/tssH) Type VI secretion system ATPase ClpV/TssH [T6SS (VF0579) - Effector delivery syste
VFG046582(gb|WP_003023398) (acpC) acid phosphatase [Acid phosphatase (VF0547) - Stress survival (VFC02
VFG040863(gb|WP_011190526) (sca2) cell surface antigen Sca2 [Sca2 (VF0495) - Motility (VFC0204)] [Ricke
VFG050159(gb|WP_001331697) (kpsD) Polysialic acid transport protein precursor [K1 capsule (VF0239) - Inva
VFG048327(gb|WP_004152886) (mrkH) transcriptional activator [Type 3 fimbriae (VF0567) - Biofilm (VFC02
VFG001880(gb|WP_010947251) (pilB) type IV pilus assembly protein PilB [Type IV pili (VF0155) - Adherence
VFG000167(gb|NP_252917) (pchR) transcriptional regulator PchR [Pyochelin (VF0095) - Nutritional/Metabolic
VFG045790(gb|WP_000833288) (cgsE) curli production assembly/transport protein CsgE [Curli fibers (VF1138
VFG000920(gb|WP_000020034) (chuX) putative heme-binding protein ChuX [Chu (VF0227) - Nutritional/Meta
VFG001283(gb|WP_000794648) (fnbA) fibronectin-binding protein A [FnBPs (VF0010) - Adherence (VFC000
VFG001241(gb|NP_249774) (flgH) flagellar L-ring protein precursor FlgH [Flagella (VF0273) - Motility (VFC0
VFG002446(gb|WP_011205636) (bapA) Type III secretion system secreted protein BapA [Bsa T3SS (VF0428) -
VFG000166(gb|NP_252916) (pchE) dihydroaeruginoic acid synthetase PchE [Pyochelin (VF0095) - Nutritional/
VFG002048(gb|WP_000249362) (gspE) general secretion pathway protein E [T2SS (VF0333) - Effector delivery
VFG000861(gb|WP_001045649) (pic) Pic serine protease precursor, autotransporter [Pic (VF0218) - Effector de
VFG041018(gb|NP_250351) (PA1660) hypothetical protein [HSI-2 (VF0943) - Effector delivery system (VFC0

VFG039088(gb|WP_011451002) (aipA) Anaplasma invasion protein A [AipA (VF0501) - Invasion (VFC0083)]
VFG000929(gb|WP_001375021) (entD) phosphopantetheinyl transferase component of enterobactin synthase mu
VFG002530(gb|WP_004199265) (cheW) chemotaxis protein CheW [Flagella (VF0430) - Motility (VFC0204)] [I
VFG049163(gb|WP_000065646) (clbQ) colibactin biosynthesis thioesterase ClbQ [Colibactin (VF0573) - Exoto
VFG048419(gb|WP_004210873) (entB) 2,3-dihydro-2,3-dihydroxybenzoate synthetase, isochroismatase [Ent (V
VFG038784(gb|WP_011706638) (flgH) flagellar basal body L-ring protein [Polar flagella (VF0473) - Motility (V
VFG051158(gb|WP_004683443.1) (bspB) type IV secretion system effector BspB [T4SS secreted effectors (VF0
VFG011826(gb|YP_002343512) (fliK) flagellar hook-length control protein FliK [Flagella (VF0114) - Motility (
VFG043775(gb|BAB86845) (bont) botulinum neurotoxin, BoNT [BoNT (VF0375) - Exotoxin (VFC0235)] [Clos
VFG038301(gb|WP_016349342) (exeK) general secretion pathway protein K [Exe T2SS (VF0478) - Effector del
VFG002053(gb|WP_001250462) (gspJ) general secretion pathway protein J [T2SS (VF0333) - Effector delivery
VFG041038(gb|NP_251056) (PA2366) uricase [HSI-3 (VF0944) - Effector delivery system (VFC0086)] [Pseudo
VFG014486(gb|NP_250133) (fliL) flagellar basal body protein FliL [Flagella (VF0273) - Motility (VFC0204)] [I
VFG002188(gb|WP_002362618) (cpsC) teichoic acid biosynthesis protein, putative [Capsule (VF0361) - Immune
VFG000001(gb|WP_010930610) (fhaB) filamentous hemagglutinin/adhesin [FHA (VF0025) - Adherence (VFC0
VFG038607(gb|WP_016349154) (mshJ) MSHA (mannose-sensitive hemagglutinin) pili biogenesis protein MshJ
VFG000919(gb|WP_001095619) (chuW) Putative oxygen independent coproporphyrinogen III oxidase [Chu (VF
VFG037456(gb|WP_000160878) (bauF) siderophore-interacting protein [Acinetobactin (VF0467) - Nutritional/M
VFG031564(gb|YP_177826) (PE_PGRS30) PE-PGRS family protein PE_PGRS30 [PE/PE-PGRS (VF0316) - Im
VFG038331(gb|WP_005320790) (exsD) Type III secretion system protein ExsD [T3SS (VF0479) - Effector deli
VFG041427(gb|WP_143343646) (CBUD_RS08635) ankyrin repeat domain-containing protein [T4SS secreted ef
VFG002064(gb|NP_248768) (dotU1) type VI secretion system protein DotU [HSI-I (VF0334) - Effector delivery
VFG049120(gb|WP_012737535) (allB) allantoinase [Allantion utilization (VF0572) - Nutritional/Metabolic facto
VFG002178(gb|AAM75254) (cylI) cytolysin immunity protein [Cytolysin (VF0356) - Exotoxin (VFC0235)] [En
VFG002176(gb|AAM75252) (cylB) ABC-type transporter [Cytolysin (VF0356) - Exotoxin (VFC0235)] [Enteroc
VFG038265(gb|WP_016352108) (exeB) general secretion pathway protein B [Exe T2SS (VF0478) - Effector del
VFG002223(gb|WP_004686778) (pmm) phosphomannomutase [LPS (VF0367) - Immune modulation (VFC0258
VFG002529(gb|WP_011205290) (tsr) methyl-accepting chemotaxis protein I [Flagella (VF0430) - Motility (VFC
VFG038452(gb|WP_016351631) (tapO) type IV pilus biogenesis protein PilO [Tap type IV pili (VF0475) - Adhe
VFG038273(gb|WP_016349335) (exeD) general secretion pathway protein D [Exe T2SS (VF0478) - Effector del
VFG041047(gb|NP_252174) (tse3) type VI secretion system effector Tse3, glycoside hydrolase [HSI-I T6SS sec
VFG049905(gb|WP_000343289) (tssL) Type VI secretion system protein TssL [T6SS (VF0579) - Effector delive
VFG001446(gb|WP_000768218) (sfaA) S-fimbrial major subunit SfaA [S fimbriae (VF0222) - Adherence (VFC
VFG001262(gb|NP_250140) (flhB) flagellar biosynthetic protein FlhB [Flagella (VF0273) - Motility (VFC0204)
VFG038828(gb|WP_011706649) (flgN) flagellar protein FlgN [Polar flagella (VF0473) - Motility (VFC0204)] [A
VFG048307(gb|WP_041937815) (fimH) type 1 fimbrial adhesin precursor [Type I fimbriae (VF0566) - Adherenc
VFG049910(gb|WP_000348788) (tssG) Type VI secretion system protein TssG [T6SS (VF0579) - Effector deliv
VFG050166(gb|WP_000066003) (neuB) N-acylneuraminate-9-phosphate synthase [K1 capsule (VF0239) - Invas
VFG000140(gb|NP_253696) (waaP) UDP-glucose:(heptosyl) LPS alpha 1,3-glucosyltransferase WaaG [LPS (VF
VFG034873(gb|WP_001270084) (espX5) Type III secretion system effector EspX5 [TTSS secreted effectors (VF
VFG043016(gb|NP_250155) (PA1464) purine-binding chemotaxis protein [Flagella (VF0273) - Motility (VFC02
VFG002416(gb|WP_001265657) (yagW/ecpD) polymerized tip adhesin of ECP fibers [ECP (VF0404) - Adheren
VFG045792(gb|WP_000791650) (csgB) curlin minor subunit CsgB [Curli fibers (VF1138) - Adherence (VFC000
VFG045793(gb|WP_000771429) (csgA) curlin major subunit CsgA [Curli fibers (VF1138) - Adherence (VFC000
VFG050757(gb|WP_001166319.1) (gspE1) general secretion pathway protein E [T2SS (VF1335) - Effector deliv
VFG001892(gb|YP_002343523) (fliY) flagellar motor switch protein FliY [Flagella (VF0114) - Motility (VFC02

VFG000477(gb|NP_461845) (rpoS) RNA polymerase sigma factor RpoS [RpoS (VF0112) - Regulation (VFC030
VFG048429(gb|WP_004183353) (entE) enterobactin synthase subunit E [Ent (VF0562) - Nutritional/Metabolic fa
VFG038864(gb|WP_073531776) (fliF) flagellar MS-ring protein [Lateral flagella (VF0474) - Motility (VFC0204
VFG038385(gb|WP_011705716) (atsI) Type VI secretion system protein [T6SS (VF0480) - Effector delivery sys
VFG048787(gb|WP_004176417) (sciN/tssJ) type VI secretion system lipoprotein TssJ [T6SS (VF0569) - Effecto
VFG038615(gb|WP_016349152) (mshI) MSHA (mannose-sensitive hemagglutinin) pili biogenesis protein MshI,
VFG013260(gb|WP_010868968) (lgtC) glycosyltransferase [LOS (VF0044) - Immune modulation (VFC0258)] [
VFG050163(gb|WP_000197083) (neuE) polysialic acid biosynthesis protein [K1 capsule (VF0239) - Invasion (V
VFG000936(gb|WP_016265865) (iutA) ferric aerobactin receptor precusor IutA [Aerobactin (VF0229) - Nutritio
VFG000925(gb|WP_000140634) (fepC) ferrienterobactin ABC transporter ATPase [Enterobactin (VF0228) - Nu
VFG012501(gb|WP_000271272) (iroE) esterase [Salmochelin siderophore (VF0230) - Nutritional/Metabolic fact
VFG013064(gb|WP_000089574) (shuA) outer membrane heme/hemoglobin receptor ShuA [Shu (VF0256) - Nut
VFG044322(gb|WP_004190148) (iroE) siderophore esterase IroE [Sal (VF0563) - Nutritional/Metabolic factor (V
VFG041096(gb|NP_459269) (STM0272) ClpV1 family T6SS ATPase [SCI (Salmonella centrisome island) (VF0
VFG050300(gb|NP_250641) (fapF) outer membrane secretin FapF [Fap (VF1191) - Adherence (VFC0001)] [Pse
VFG038293(gb|WP_016349340) (exeI) general secretion pathway protein I [Exe T2SS (VF0478) - Effector deliv
VFG001822(gb|NP_216897) (mbtD) Polyketide synthetase MbtD (polyketide synthase) [Mycobactin (VF0299) -
VFG002406(gb|WP_000728972) (esaA) type VII secretion system protein EsaA [Type VII secretion system (VF
VFG002074(gb|NP_248778) (hsiG1/tssF) type VI secretion system hcp secretion island protein HsiG1 [HSI-I (V
VFG048708(gb|WP_041168969) (vgrG/tssI) type VI secretion system tip protein VgrG [T6SS (VF0569) - Effect
VFG038380(gb|WP_043163044) (vipB) Type VI secretion system contractile sheath large subunit TssC/VipB [T
VFG013342(gb|WP_010869051) (lpsA) glycosyltransferase [LOS (VF0044) - Immune modulation (VFC0258)] [
VFG038533(gb|WP_016350152) (flpF) traffic ATPase [Flp type IV pili (VF0476) - Adherence (VFC0001)] [Aer
VFG038627(gb|WP_016350050) (fliF) flagellar MS-ring protein [Polar flagella (VF0473) - Motility (VFC0204)]
VFG038378(gb|WP_011705712) (vipA) Type VI secretion system contractile sheath small subunit TssB/VipA [T
VFG038402(gb|WP_011705725) (vasK/icmF) Type VI secretion system protein [T6SS (VF0480) - Effector deliv
VFG049148(gb|WP_041168941) (clbB) colibactin hybrid non-ribosomal peptide synthetase/type I polyketide syn
VFG034851(gb|WP_000129342) (espX1) Type III secretion system effector EspX1 [TTSS secreted effectors (VF
VFG000064(gb|WP_010930831) (bcrD) Type III secretion system LcrD homolog protein BcrD [TTSS (VF0035)
VFG038305(gb|WP_016349343) (exeL) general secretion pathway protein L [Exe T2SS (VF0478) - Effector deli
VFG001250(gb|NP_249791) (fliE) flagellar hook-basal body complex protein FliE [Flagella (VF0273) - Motility
VFG001308(gb|WP_001291010) (cap8L) type 8 capsular polysaccharide synthesis protein Cap8L [Capsule (VF0
VFG000195(gb|NP_250391) (pcr2) type III secretion system protein Pcr2 [TTSS (VF0083) - Effector delivery sy
VFG000363(gb|WP_002212775) (irp2) yersiniabactin biosynthetic protein Irp2 [Yersiniabactin (VF0136) - Nutri
VFG050457(gb|WP_001274990.1) (pilB) type IV-A pilus assembly ATPase PilB [TFP (VF1334) - Adherence (V
VFG038808(gb|WP_011706644) (flgB) flagellar basal-body rod protein FlgB [Polar flagella (VF0473) - Motility
VFG038525(gb|WP_043118801) (flpH) Flp pilus assembly protein FlpH [Flp type IV pili (VF0476) - Adherence
VFG000881(gb|WP_000930062) (papB) regulatory protein PapB [P fimbriae (VF0220) - Adherence (VFC0001)]
VFG016463(gb|WP_011206020) (p97) protein p97; cilium adhesin [P97/P102 paralog family (VF0426) - Adhere
VFG044098(gb|NP_252910) (PA4220) hypothetical protein [Pyochelin (VF0095) - Nutritional/Metabolic factor (
VFG038381(gb|WP_011705714) (atsG) Type VI secretion system protein [T6SS (VF0480) - Effector delivery sy
VFG038723(gb|WP_011705294) (AHML_RS07540) CobQ/CobB/MinD/ParA family protein [Polar flagella (VF
VFG038772(gb|WP_016351319) (flgK) flagellar hook-associated protein FlgK [Polar flagella (VF0473) - Motilit
VFG049084(gb|WP_004175264) (rfbB) O-antigen export ABC transporter ATP-binding protein RfbB [LPS (VF
VFG041091(gb|NP_459264) (STM0267) sciB domain protein [SCI (Salmonella centrisome island) (VF0974) - E
VFG000933(gb|WP_001007140) (entB) isochorismatase [Enterobactin (VF0228) - Nutritional/Metabolic factor (

VFG042965(gb|WP_004188396) (pilP) type IV pilus biogenesis protein PilP [Type IV pili (VF0431) - Adherenc
VFG000188(gb|NP_250384) (pscR) type III secretion system protein PscR [TTSS (VF0083) - Effector delivery s
VFG043626(gb|WP_000820818) (mrkB) fimbrial chaperone protein mrkB precursor [Type 3 fimbriae (VF0567)
VFG050165(gb|WP_001259305) (neuA) N-acylneuraminate cytidylyltransferase [K1 capsule (VF0239) - Invasio
VFG000937(gb|WP_000750139) (iucD) L-lysine 6-monooxygenase IucD [Aerobactin (VF0229) - Nutritional/M
VFG038155(gb|WP_000209960) (ACICU_RS00500) phosphomannomutase/phosphoglucomutase [Capsule (VF
VFG041040(gb|NP_251058) (PA2368) hypothetical protein [HSI-3 (VF0944) - Effector delivery system (VFC0
VFG038727(gb|WP_011705295) (cheW) chemotaxis protein CheW [Polar flagella (VF0473) - Motility (VFC02
VFG041237(gb|WP_010945887) (cegC2) Dot/Icm type IV secretion system effector CegC2 [Dot/Icm T4SS secr
VFG034866(gb|WP_000900547) (espX4) Type III secretion system effector EspX4 [TTSS secreted effectors (V
VFG002281(gb|WP_003468301) (nagL) hyaluronidase [Mu-toxin (VF0389) - Exoenzyme (VFC0251)] [Clostrid
VFG038691(gb|WP_010634619) (flhG) CobQ/CobB/MinD/ParA family protein [Polar flagella (VF0473) - Moti
VFG000912(gb|WP_001443207) (focD) F1C fimbrial usher [F1C fimbriae (VF0224) - Adherence (VFC0001)] [
VFG038715(gb|WP_024943899) (motA) chemotaxis protein MotA [Polar flagella (VF0473) - Motility (VFC020
VFG038824(gb|WP_011706648) (flgM) anti-sigma-D factor [Polar flagella (VF0473) - Motility (VFC0204)] [A
VFG000935(gb|WP_001222189) (iroN) salmochelin receptor IroN [Salmochelin siderophore (VF0230) - Nutritio
VFG043321(gb|WP_010946642) (flgM) negative regulator of flagellin synthesis [Flagella (VF0157) - Motility (
VFG034908(gb|WP_000978349) (espY3) Type III secretion system effector EspY3 [TTSS secreted effectors (V
VFG010463(gb|WP_010946666) (pilQ) type IV pilus secretin protein PilQ  [Type IV pili (VF0155) - Adherence
VFG014681(gb|NP_253640) (motB) flagellar motor protein [Flagella (VF0273) - Motility (VFC0204)] [Pseudom
VFG002113(gb|WP_015444908) (sidE/laiD) Dot/Icm type IV secretion system effector SidE [Dot/Icm T4SS sec
VFG006582(gb|WP_001862801) (flhF) flagellar biosynthesis protein [Flagella (VF0051) - Motility (VFC0204)]
VFG001931(gb|YP_002344858) (cadF) outer membrane fibronectin-binding protein [CadF (VF0322) - Adherenc
VFG013066 (shuW) heme anaerobic degradation radical SAM methyltransferase ChuW/HutW [Shu (VF0256) -
VFG002425(gb|WP_004526211) (pilA) type IV fimbrial pilin protein [Type IV pili (VF0431) - Adherence (VFC
VFG000916(gb|WP_001017192) (chuS) heme oxygenase ChuS [Chu (VF0227) - Nutritional/Metabolic factor (V
VFG048643(gb|WP_014907727) (vipB/tssC) type VI secretion system contractile sheath large subunit VipB [T6
VFG012513(gb|WP_001221122) (iroB) glucosyltransferase IroB [Salmochelin siderophore (VF0230) - Nutrition
VFG051149(gb|WP_002966733.1) (bspA) type IV secretion system effector, DUF2062 domain-containing prote
VFG002057(gb|NP_248761) (tagR) type IV secretion associated protein TagR, positively regulates PpkA [HSI-I
VFG038867(gb|WP_005314284) (lfiI) lateral flagellar FliI-like assembly ATPase [Lateral flagella (VF0474) - M
VFG002463(gb|WP_011205639) (bsaO) Type III secretion system protein BsaO [Bsa T3SS (VF0428) - Effector
VFG049161(gb|WP_001029878) (clbO) colibactin polyketide synthase ClbO [Colibactin (VF0573) - Exotoxin (V
VFG012160(gb|WP_011590331) (nanI) exo-alpha-sialidase [Sialidase (VF0391) - Exoenzyme (VFC0251)] [Clo
VFG038289(gb|WP_016349339) (exeH) general secretion pathway protein H [Exe T2SS (VF0478) - Effector de
VFG000127(gb|NP_252235) (algG) outer membrane protein AlgE [Alginate (VF0091) - Biofilm (VFC0271)] [P
VFG034756 (espL1) Type III secretion system effector espL1 [TTSS secreted effectors (VF1111) - Effector deli
VFG002050(gb|WP_001087302) (gspG) general secretion pathway protein G [T2SS (VF0333) - Effector delivery
VFG001957(gb|YP_002344812) (Cj1429c) hypothetical protein [Capsule (VF0323) - Immune modulation (VFC
VFG050787(gb|WP_001112089.1) (gspF) general secretion pathway protein F [T2SS (VF1335) - Effector delive
VFG038760(gb|WP_016351312) (flrB) two-component sensor kinase [Polar flagella (VF0473) - Motility (VFC0
VFG049152(gb|WP_000337350) (clbF) colibactin biosynthesis dehydrogenase ClbF [Colibactin (VF0573) - Exo
VFG049914(gb|WP_000037389) (tssB) Type VI secretion system protein TssB [T6SS (VF0579) - Effector deliv
VFG041043(gb|NP_251061) (PA2371) ClpA/B-type protease [HSI-3 (VF0944) - Effector delivery system (VFC
VFG044323(gb|WP_015874960) (ybtS) yersiniabactin biosynthesis salicylate synthase YbtS [Ybt (VF0564) - Nu
VFG000035(gb|WP_010929613) (bplD) UDP-N-acetylglucosamine 2-epimerase [LPS (VF0033) - Immune modu

VFG000266(gb|WP_002222141) (hmbR) hemoglobin receptor [HmbR (VF0048) - Nutritional/Metabolic factor (
VFG012609(gb|WP_000993317) (chuW) Putative oxygen independent coproporphyrinogen III oxidase [Chu (VF
VFG038281(gb|WP_016349337) (exeF) general secretion pathway protein F [Exe T2SS (VF0478) - Effector deli
VFG043015(gb|NP_250150) (PA1459) chemotaxis-specific methylesterase [Flagella (VF0273) - Motility (VFC0
VFG000905(gb|WP_001546261) (hlyC) Hemolysin C [-Hemolysin (VF0225) - Exotoxin (VFC0235)] [Escherich
VFG001252(gb|NP_249793) (fliG) flagellar motor switch protein G [Flagella (VF0273) - Motility (VFC0204)] [
VFG049018(gb|WP_002913007) (rcsB) transcriptional regulator RcsB [RcsAB (VF0571) - Regulation (VFC030
VFG038572(gb|WP_016349165) (mshD) MSHA (mannose-sensitive hemagglutinin) pili minor prepilin protein M
VFG002098(gb|WP_010948481) (lepA) Dot/Icm type IV secretion system effector LepA, interaptin [Dot/Icm T4
VFG038356 (aopX) hypothetical protein [T3SS (VF0479) - Effector delivery system (VFC0086)] [Aeromonas sa
VFG044198(gb|WP_003022885) (fslB) MFS transporter, inner membrane protein responsible for uptake of the fe
VFG049062(gb|WP_015875020) (rfbD) UDP-galactopyranose mutase [LPS (VF0561) - Immune modulation (VF
VFG000886(gb|WP_000261297) (papJ) P pilus assembly protein PapJ [P fimbriae (VF0220) - Adherence (VFC0
VFG002275(gb|WP_003467630) (pfoA) perfringolysin O [Theta-toxin/PFO (VF0382) - Exotoxin (VFC0235)] [C
VFG049916(gb|WP_000103365) (vgrG/tssI) type VI secretion system effector VgrG [T6SS (VF0579) - Effector
VFG000211(gb|NP_250407) (pscC) type III secretion system secretin PscC [TTSS (VF0083) - Effector delivery
VFG042972(gb|WP_000924780) (sipA) signal peptidase I [PI-2 (VF0530) - Adherence (VFC0001)] [Streptococ
VFG014893(gb|NP_254009) (algC) phosphomannomutase AlgC [Alginate (VF0091) - Biofilm (VFC0271)] [Pse
VFG001260(gb|NP_250138) (fliQ) flagellar biosynthetic protein FliQ [Flagella (VF0273) - Motility (VFC0204)]
VFG007286(gb|WP_000290380) (flaC) flagellin [Flagella (VF0519) - Motility (VFC0204)] [Vibrio cholerae O1
VFG000908(gb|WP_000860472) (hlyD) Hemolysin D [-Hemolysin (VF0225) - Exotoxin (VFC0235)] [Escherich
VFG041045(gb|NP_251063) (PA2373) hypothetical protein [HSI-3 (VF0944) - Effector delivery system (VFC00
VFG002115(gb|WP_015444209) (sdeB/laiB) Dot/Icm type IV secretion system effector SdeB [Dot/Icm T4SS sec
VFG049164(gb|WP_004175305) (clbS) colibactin self-protection protein ClbS [Colibactin (VF0573) - Exotoxin (
VFG049133(gb|WP_004177236) (acrA) acriflavine resistance protein A [AcrAB (VF0568) - Antimicrobial activi
VFG038328(gb|WP_005320795) (ascD) Type III secretion system OM ring protein AscD [T3SS (VF0479) - Effe
VFG049908(gb|WP_001080148) (tssJ) Type VI secretion system protein TssJ [T6SS (VF0579) - Effector deliver
VFG049835(gb|WP_012076956) (exlA) hemolysin [Exolysin (VF0582) - Effector delivery system (VFC0086)] [
VFG002280(gb|WP_011010361) (nagK) hyaluronidase [Mu-toxin (VF0389) - Exoenzyme (VFC0251)] [Clostrid
VFG045321(gb|WP_010946511) (LPG_RS03835) hypothetical protein [LPS (VF0171) - Immune modulation (V
VFG049157(gb|WP_014907259) (clbK) colibactin hybrid non-ribosomal peptide synthetase/type I polyketide syn
VFG019180(gb|WP_011987710) (bont) botulinum neurotoxin, BoNT [BoNT (VF0375) - Exotoxin (VFC0235)] [
VFG049154(gb|WP_014907257) (clbH) colibactin non-ribosomal peptide synthetase ClbH [Colibactin (VF0573)
VFG038643(gb|WP_011705276) (fliJ) flagellar export protein FliJ [Polar flagella (VF0473) - Motility (VFC0204
VFG000913(gb|WP_000237770) (focF) F1C minor fimbrial subunit F precursor [F1C fimbriae (VF0224) - Adhe
VFG000690(gb|AAP42183) (AAP42183) Bcs3' [Capsule (VF0043) - Immune modulation (VFC0258)] [Haemop
VFG043382(gb|WP_000996418) (HP_RS03480) RNA polymerase factor sigma-54 [Flagella (VF0051) - Motility
VFG002177(gb|AAM75253) (cylA) cytolysin activator [Cytolysin (VF0356) - Exotoxin (VFC0235)] [Enterococ
VFG000888(gb|WP_000723799) (papE) P pilus minor subunit PapE [P fimbriae (VF0220) - Adherence (VFC000
VFG038619(gb|WP_016349413) (motX) sodium-type flagellar motor component [Polar flagella (VF0473) - Mot
VFG038371(gb|WP_011705707) (vgrG2) VgrG protein [T6SS (VF0480) - Effector delivery system (VFC0086)]
VFG037852(gb|WP_001075965) (ACICU_RS00395) polysaccharide biosynthesis tyrosine autokinase [Capsule (
VFG049116(gb|WP_012737528) (allA) ureidoglycolate hydrolase [Allantion utilization (VF0572) - Nutritional/M
VFG000143(gb|NP_253699) (waaF) heptosyltransferase I [LPS (VF0085) - Immune modulation (VFC0258)] [Ps
VFG001935(gb|YP_002344312) (ciaB) invasion antigen CiaB [CiaB (VF0324) - Invasion (VFC0083)] [Campylo
VFG043778(gb|CAA52275) (bont) botulinum neurotoxin, BoNT [BoNT (VF0375) - Exotoxin (VFC0235)] [Clos

VFG000353(gb|WP_002208792) (psaC) outer membrane usher protein PsaC [Myf/pH6 antigen (VF0134) - Adhe
VFG038711(gb|WP_016350060) (cheB-2) chemotaxis-specific methylesterase [Polar flagella (VF0473) - Motilit
VFG038444(gb|WP_075384315) (tapM) type IV pilus assembly protein PilM [Tap type IV pili (VF0475) - Adhe
VFG038776(gb|WP_016351320) (flgJ) flagellar rod assembly protein/muramidase FlgJ [Polar flagella (VF0473)
VFG038468(gb|WP_016351972) (tapU) twitching ATPase [Tap type IV pili (VF0475) - Adherence (VFC0001)]
VFG048808(gb|WP_004899416) (rfbK1) O9 family phosphomannomutase RfbK1 [Capsule (VF0560) - Immune
VFG000133(gb|NP_252241) (algA) phosphomannose isomerase / guanosine 5'-diphospho-D-mannose pyrophosp
VFG038623(gb|WP_011705271) (fliE) flagellar hook-basal body complex protein FliE [Polar flagella (VF0473)
VFG000116(gb|NP_254170) (algB) two-component response regulator AlgB [Alginate (VF0091) - Biofilm (VFC
VFG001964(gb|YP_002344819) (Cj1436c) aminotransferase [Capsule (VF0323) - Immune modulation (VFC025
VFG043631(gb|WP_011081073) (VV1_RS15595) pilus assembly protein N-terminal domain-containing protein
VFG001226(gb|NP_249100) (pilH) twitching motility protein PilH [Type IV pili (VF0082) - Adherence (VFC00
VFG049029(gb|WP_014907241) (KP1_RS17220) glycosyltransferase [LPS (VF0561) - Immune modulation (VF
VFG016041(gb|NP_251092) (pvdI) peptide synthase [Pyoverdine (VF0094) - Nutritional/Metabolic factor (VFC
VFG038886(gb|WP_005314222) (lafE) LafE, flagellar hook control length protein [Lateral flagella (VF0474) - M
VFG041033(gb|NP_251051) (PA2361) hypothetical protein [HSI-3 (VF0944) - Effector delivery system (VFC00
VFG050514(gb|WP_001214067.1) (fimU) GspH/FimT family pseudopilin [TFP (VF1334) - Adherence (VFC000
VFG000186(gb|NP_250382) (pscT) type III secretion system protein PscT [TTSS (VF0083) - Effector delivery s
VFG000934(gb|WP_000348393) (entA) 2,3-dihydro-2,3-dihydroxybenzoate dehydrogenase EntA [Enterobactin
VFG041105(gb|NP_459279) (STM0281) type VI secretion system-associated protein [SCI (Salmonella centrison
VFG012191(gb|WP_011948511) (bont) botulinum neurotoxin, BoNT [BoNT (VF0375) - Exotoxin (VFC0235)] [
VFG037762(gb|WP_000078878) (lpxL) lauroyl acyltransferase [LPS (VF0466) - Immune modulation (VFC0258
VFG038480(gb|WP_016349429) (tppB) type IV pilus pseudopilin [Tap type IV pili (VF0475) - Adherence (VFC
VFG002415(gb|WP_001131063) (yagX/ecpC) E. coli common pilus usher EcpC [ECP (VF0404) - Adherence (V
VFG046465(gb|WP_003028672) (tufA) elongation factor Tu [EF-Tu (VF0460) - Adherence (VFC0001)] [Franci
VFG002419(gb|WP_000744066) (coa) staphylocoagulase precursor [Staphylocoagulase (VF0421) - Exoenzyme
VFG041333(gb|WP_010948115) (lem24) Dot/Icm type IV secretion system effector Lem24 [Dot/Icm T4SS secre
VFG001396(gb|NP_216424) (katG) catalase-peroxidase [KatG (VF0303) - Stress survival (VFC0282)] [Mycoba
VFG000360(gb|WP_002212799) (ybtT) type II thioesterase YbtT [Yersiniabactin (VF0136) - Nutritional/Metabo
VFG049153(gb|WP_000159201) (clbG) colibactin biosynthesis acyltransferase ClbG [Colibactin (VF0573) - Exo
VFG037912(gb|WP_000939040) (pseB) UDP-N-acetylglucosamine 4,6-dehydratase (inverting) [Capsule (VF046
VFG049917(gb|WP_000508696) (rhs/PAAR) Type VI secretion system protein, PAAR family [T6SS (VF0579) -
VFG050772(gb|WP_001020922.1) (gspE2) putative secretion pathway ATPase [T2SS (VF1335) - Effector delive
VFG049899(gb|WP_001087735) (tssA) Type VI secretion system protein TssA [T6SS (VF0579) - Effector deliv
VFG048459(gb|WP_004147525) (entS) enterobactin exporter EntS [Ent (VF0562) - Nutritional/Metabolic factor
VFG012162(gb|WP_011590241) (nanJ) exo-alpha-sialidase [Sialidase (VF0391) - Exoenzyme (VFC0251)] [Clo
VFG038611(gb|WP_016349153) (mshI1) MSHA (mannose-sensitive hemagglutinin) pili biogenesis protein Msh
VFG001300(gb|WP_000940793) (cap8D) type 8 capsular polysaccharide synthesis protein Cap8D [Capsule (VF0
VFG038313(gb|WP_043118380) (exeN) general secretion pathway protein N [Exe T2SS (VF0478) - Effector del
VFG044332(gb|WP_015874967) (ybtE) yersiniabactin biosynthesis salycil-AMP ligase YbtE [Ybt (VF0564) - N
VFG007162(gb|WP_005377232) (vscR) type III secretion system C-ring protein VscR [T3SS1 (VF0408) - Effec
VFG012327(gb|WP_011076307) (sfaC) S fimbrial switch regulatory protein SfaC [S fimbriae (VF0222) - Adhere
VFG045473(gb|AAM75250) (cylS) CylS; cytolysin subunit [Cytolysin (VF0356) - Exotoxin (VFC0235)] [Entero
VFG002492(gb|WP_004184995) (fliN) flagellar motor switch protein FliN [Flagella (VF0430) - Motility (VFC0
VFG038840(gb|WP_005300916) (flmH) short chain dehydrogenase/reductase family oxidoreductase [Polar flage
VFG000875(gb|WP_000066579) (fimC) Chaperone protein fimC precursor [Type 1 fimbriae (VF0221) - Adhere

VFG000130(gb|NP_252238) (algI) alginate o-acetyltransferase AlgI [Alginate (VF0091) - Biofilm (VFC0271)] [
VFG038639(gb|WP_011705275) (fliI) flagellum-specific ATP synthase [Polar flagella (VF0473) - Motility (VFC
VFG047162(gb|WP_003022298) (wbtE) nucleotide sugar dehydrogenase [LPS (VF0542) - Immune modulation (
VFG042736(gb|NP_253152) (rpoN) RNA polymerase factor sigma-54 [Type IV pili (VF0082) - Adherence (VF
VFG047512(gb|WP_003020978) (bioB) biotin synthase [Biotin synthesis (VF0552) - Nutritional/Metabolic facto
VFG038285(gb|WP_005308483) (exeG) general secretion pathway protein G [Exe T2SS (VF0478) - Effector del
VFG049156(gb|WP_077265119) (clbJ) colibactin non-ribosomal peptide synthetase ClbJ [Colibactin (VF0573) -
VFG038417(gb|WP_016349294) (fimD) fimbrial chaperone protein [Type I pili (VF0486) - Adherence (VFC000
VFG048449(gb|WP_012737621) (fepB) iron-enterobactin transporter periplasmic binding protein [Ent (VF0562)
VFG000025(gb|WP_010929995) (cyaA) bifunctional hemolysin-adenylate cyclase precursor [Cya (VF0028) - Ex
VFG049912 (tssC) Type VI secretion system protein TssC [T6SS (VF0579) - Effector delivery system (VFC008
VFG000182(gb|NP_251793) (xcpR) general secretion pathway protein E [Xcp secretion system (VF0084) - Effec
VFG002499(gb|WP_004197164) (fliI) flagellum-specific ATP synthase FliI [Flagella (VF0430) - Motility (VFC
VFG038719(gb|WP_016350061) (pomB) ompA family protein [Polar flagella (VF0473) - Motility (VFC0204)] [
VFG000173(gb|NP_252907) (phzS) flavin dependent hydroxylase PhzS [Pyocyanin (VF0100) - Nutritional/Meta
VFG002191(gb|WP_002369252) (sprE) serine proteinase, V8 family [SprE (VF0358) - Exoenzyme (VFC0251)]
VFG048229(gb|WP_015875142) (fimB) tyrosine recombinase [Type I fimbriae (VF0566) - Adherence (VFC000
VFG002517(gb|WP_004203463) (flgK) flagellar hook-associated protein 1 FlgK [Flagella (VF0430) - Motility (
VFG038472(gb|WP_011706550) (tapW) twitching ATPase [Tap type IV pili (VF0475) - Adherence (VFC0001)]
VFG002432(gb|WP_004537540) (pilQ) type IV pilus biosynthesis protein [Type IV pili (VF0431) - Adherence (
VFG045478(gb|WP_016356699) (LPG_RS00200) hypothetical protein [Dot/Icm T4SS secreted effectors (VF07
VFG000362(gb|WP_002212777) (irp1) yersiniabactin biosynthetic protein Irp1 [Yersiniabactin (VF0136) - Nutri
VFG042976(gb|WP_154081516) (ebpA) endocarditis and biofilm-associated pilus tip protein EbpA [Ebp pili (V
VFG002518(gb|WP_004200262) (flgL) flagellar hook-associated protein 3 FlgL [Flagella (VF0430) - Motility (
VFG038460(gb|WP_043119325) (tapQ) type IV pilus secretin PilQ [Tap type IV pili (VF0475) - Adherence (VF
VFG034915(gb|WP_000225947) (espY4) Type III secretion system effector EspY4 [TTSS secreted effectors (V
VFG006622(gb|WP_000796857) (flhB) flagellar biosynthesis protein [Flagella (VF0051) - Motility (VFC0204)]
VFG000968(gb|WP_010922714) (emm) M protein [M protein (VF0242) - Immune modulation (VFC0258)] [Stre
VFG038892(gb|WP_043118302) (aerA/act) Aerolysin A [Aerolysin (VF0481) - Exotoxin (VFC0235)] [Aeromo
VFG000884(gb|WP_000654937) (papC) usher protein PapC [P fimbriae (VF0220) - Adherence (VFC0001)] [Es
VFG000944(gb|AAK16477) (draC) usher protein DraC [Dr adhesins (VF0223) - Adherence (VFC0001)] [Esche
VFG001310(gb|WP_001230668) (capN) capsular polysaccharide type 5/8 biosynthesis epimerase CapN [Capsul
VFG001403(gb|NP_216899) (mbtB) Phenyloxazoline synthase MbtB (phenyloxazoline synthetase) [Mycobactin
VFG002047(gb|WP_000498818) (gspD) general secretion pathway protein D [T2SS (VF0333) - Effector deliver
VFG037372(gb|WP_001104139) (bauB) ferric siderophore ABC transporter, periplasmic siderophore-binding pr
VFG000047(gb|WP_003820063) (bscO) Bordetella Bsc type III secretion system protein BscO [TTSS (VF0035)
VFG001391(gb|NP_215677) (narG) nitrate reductase subunit alpha [Nitrate reductase (VF0302) - Nutritional/Me
VFG042865(gb|WP_010946369) (fimT) type IV pilus biogenesis protein FimT [Type IV pili (VF0155) - Adhere
VFG038861(gb|WP_005314296) (lfhA) lateral flagellar biosynthesis protein [Lateral flagella (VF0474) - Motilit
VFG001827(gb|WP_001020414) (senB) enterotoxin [ShET2 (VF0258) - Exotoxin (VFC0235)] [Shigella boydii
VFG007823(gb|NP_217447) (ppsA) phthiocerol synthesis polyketide synthase type I PpsA [PDIM (VF0309) - In
VFG049118(gb|WP_012737529) (allR) DNA-binding transcriptional repressor AllR [Allantion utilization (VF05
VFG006902(gb|WP_000957200) (pilB) type IV-A pilus assembly ATPase PilB [ChiRP (VF0609) - Adherence (
VFG012333(gb|WP_000703306) (sfaD) S fimbriae minor subunit SfaD [S fimbriae (VF0222) - Adherence (VFC
VFG000180(gb|NP_251791) (xcpT) general secretion pathway protein G [Xcp secretion system (VF0084) - Effe
VFG000282(gb|WP_000488766) (cag3) type IV secretion system protein Cag3, component outer membrane sub

VFG000129(gb|NP_252237) (algL) poly(beta-d-mannuronate) lyase precursor AlgL [Alginate (VF0091) - Biofilm
VFG041100(gb|NP_459274) (STM0276) Hcp1 family type VI secretion system effector [SCI (Salmonella centris
VFG002410(gb|WP_001010291) (esxC) type VII secretion system secreted protein EsxC [Type VII secretion sys
VFG048498(gb|WP_012737619) (entF) enterobactin synthase subunit F [Ent (VF0562) - Nutritional/Metabolic fa
VFG000616(gb|NP_709455) (iucB) aerobactin synthesis protein IucB [Aerobactin (VF0123) - Nutritional/Metabo
VFG000469(gb|NP_459778) (slrP) type III secretion system effector SlrP, E3 ubiquitin ligase [TTSS effectors se
VFG038867(gb|WP_005314284) (lfiI) lateral flagellar FliI-like assembly ATPase [Lateral flagella (VF0474) - Mo
VFG002503(gb|WP_004534368) (fliE) flagellar hook-basal body complex protein FliE [Flagella (VF0430) - Mot
VFG041113(gb|NP_459287) (STM0289) type VI secretion protein Vgr [SCI (Salmonella centrisome island) (VF
VFG038397(gb|WP_011705722) (vasH) sigma-54 dependent transcriptional regulator [T6SS (VF0480) - Effector
VFG002059(gb|NP_248763) (tagT) type six secretion associated protein TagT, ATP-binding component of ABC
VFG000114(gb|NP_253218) (xcpA/pilD) type 4 prepilin peptidase PilD [Type IV pili (VF0082) - Adherence (VI
VFG048980(gb|WP_041937790) (KP1_RS17345) capsule assembly Wzi family protein [Capsule (VF0560) - Im
VFG000133(gb|NP_252241) (algA) phosphomannose isomerase / guanosine 5'-diphospho-D-mannose pyrophosp
VFG038581(gb|WP_043118279) (mshB) MSHA (mannose-sensitive hemagglutinin) pili minor prepilin protein M
VFG001918(gb|YP_002344570) (fliR) flagellar biosynthetic protein FliR [Flagella (VF0114) - Motility (VFC020
VFG038820(gb|WP_161785096) (flgA) flagella basal body P-ring formation protein FlgA [Polar flagella (VF047
VFG045495(gb|WP_010946171) (legY) Dot/Icm type IV secretion system effector LegY [Dot/Icm T4SS secrete
VFG038800(gb|WP_016351322) (flgD) flagellar hook capping protein [Polar flagella (VF0473) - Motility (VFC
VFG042075(gb|WP_003814626) (btcA) chaperone for BteA [TTSS (VF0035) - Effector delivery system (VFC00
VFG043016(gb|NP_250155) (PA1464) purine-binding chemotaxis protein [Flagella (VF0273) - Motility (VFC02
VFG038856(gb|WP_005314315) (lfiN) lateral flagellar motor switch protein, lfiN [Lateral flagella (VF0474) - M
VFG000363(gb|WP_002212775) (irp2) yersiniabactin biosynthetic protein Irp2 [Yersiniabactin (VF0136) - Nutri
VFG038321(gb|WP_005320811) (ascK) Type III secretion system C ring protein AscK [T3SS (VF0479) - Effect
VFG050699(gb|WP_000840548.1) (pilR) sigma-54 dependent transcriptional regulator [TFP (VF1334) - Adheren
VFG002056 (gspM) general secretion pathway protein M [T2SS (VF0333) - Effector delivery system (VFC0086)
VFG043014(gb|NP_250149) (PA1458) two-component sensor [Flagella (VF0273) - Motility (VFC0204)] [Pseud
VFG002126(gb|NP_219593) (cdsV) Type III secretion protein CdsV [TTSS (VF0344) - Effector delivery system
VFG001210(gb|NP_253244) (pilY1) type 4 fimbrial biogenesis protein PilY1 [Type IV pili (VF0082) - Adherenc
VFG051098(gb|WP_002001416.1) (clpV/tssH) type VI secretion system ATPase TssH [T6SS (VF1337) - Effect
VFG038389(gb|WP_011705718) (atsK) Type VI secretion system protein [T6SS (VF0480) - Effector delivery sy
VFG050159(gb|WP_001331697) (kpsD) Polysialic acid transport protein precursor [K1 capsule (VF0239) - Invas
VFG038269(gb|WP_075384415) (exeC) general secretion pathway protein C [Exe T2SS (VF0478) - Effector del
VFG049159(gb|WP_014907260) (clbM) precolibactin export MATE transporter ClbM [Colibactin (VF0573) - E
VFG038593(gb|WP_016349158) (mshN) Tetratricopeptide repeat protein, PilF-like [MSHA type IV pili (VF047
VFG001845(gb|WP_010946207) (icmF) Dot/Icm type IV secretion system protein IcmF [Dot/Icm (VF0156) - Ef
VFG051070(gb|WP_001066523.1) (tssC) type VI secretion system contractile sheath large subunit [T6SS (VF13
VFG000169(gb|NP_252919) (pchC) pyochelin biosynthetic protein PchC [Pyochelin (VF0095) - Nutritional/Met
VFG002521(gb|WP_004525693) (flhF) flagellar biosynthesis regulator FlhF [Flagella (VF0430) - Motility (VFC
VFG002446(gb|WP_011205636) (bapA) Type III secretion system secreted protein BapA [Bsa T3SS (VF0428) -
VFG038525(gb|WP_043118801) (flpH) Flp pilus assembly protein FlpH [Flp type IV pili (VF0476) - Adherence
VFG001296(gb|WP_000676533) (sspA) serine protease; V8 protease; glutamyl endopeptidase [V8 protease (VF
VFG043500(gb|WP_011206108) (mhp271) p97 cilium adhesin paralog [P97/P102 paralog family (VF0426) - Ad
VFG002025(gb|YP_002344527) (htrB) lipid A biosynthesis lauroyl acyltransferase [LOS (VF0326) - Immune m
VFG002329(gb|WP_005160381) (fliG) flagellar motor switch protein G [Flagella (VF0394) - Motility (VFC020
VFG013069(gb|WP_005019013) (shuU) permease of iron compound ABC transport system [Shu (VF0256) - Nu

VFG050787(gb|WP_001112089.1) (gspF) general secretion pathway protein F [T2SS (VF1335) - Effector delive
VFG001713(gb|WP_001298062) (papX) PapX protein regulates flagellum synthesis to repress motility [P fimbri
VFG034908(gb|WP_000978349) (espY3) Type III secretion system effector EspY3 [TTSS secreted effectors (V
VFG037792(gb|WP_000064875) (lpxA) acyl-ACP--UDP-N-acetylglucosamine O-acyltransferase [LPS (VF046
VFG001231(gb|NP_249104) (chpA) still frameshift probable component of chemotactic signal transduction syste
VFG038788(gb|WP_011706639) (flgG) flagellar basal body rod protein FlgG [Polar flagella (VF0473) - Motility
VFG002365(gb|WP_005167740) (gmd) GDP-mannose 4,6-dehydratase [O-antigen (VF0392) - Immune modulat
VFG047162(gb|WP_003022298) (wbtE) nucleotide sugar dehydrogenase [LPS (VF0542) - Immune modulation (
VFG006342(gb|WP_000753173) (babB/hopT) outer membrane protein adhesin [BabA (VF0053) - Adherence (V
VFG038365(gb|WP_011705044) (hcp1) HcpA-like protein [T6SS (VF0480) - Effector delivery system (VFC008
VFG001251(gb|NP_249792) (fliF) flagellar M-ring protein FliF [Flagella (VF0273) - Motility (VFC0204)] [Pseu
VFG038513(gb|WP_016350157) (flpK) Flp pilus pseudopilin [Flp type IV pili (VF0476) - Adherence (VFC0001
VFG001826(gb|NP_217099) (relA) Probable GTP pyrophosphokinase RelA (ATP:GTP 3'-pyrophosphotransfera
VFG038619(gb|WP_016349413) (motX) sodium-type flagellar motor component [Polar flagella (VF0473) - Mot
VFG049029(gb|WP_014907241) (KP1_RS17220) glycosyltransferase [LPS (VF0561) - Immune modulation (V
VFG000908(gb|WP_000860472) (hlyD) Hemolysin D [-Hemolysin (VF0225) - Exotoxin (VFC0235)] [Escheric
VFG002067(gb|NP_248771) (fha1) type VI secretion system forkhead-associated protein Fha1 [HSI-I (VF0334)
VFG000262(gb|WP_010980797) (tbpA) transferrin-binding protein A [Tbp (VF0046) - Nutritional/Metabolic fa
VFG000937(gb|WP_000750139) (iucD) L-lysine 6-monooxygenase IucD [Aerobactin (VF0229) - Nutritional/M
VFG038373(gb|WP_011705708) (atsA) Type VI secretion system protein [T6SS (VF0480) - Effector delivery sy
VFG038576(gb|WP_016349164) (mshC) MSHA (mannose-sensitive hemagglutinin) pili minor prepilin protein M
VFG014117(gb|NP_251831) (wbpM) nucleotide sugar epimerase/dehydratase WbpM [LPS (VF0085) - Immune
VFG035902(gb|WP_000209220) (aatP) permease AatP [Dispersin (VF0215) - Others (VFC0346)] [Escherichia c
VFG012621(gb|WP_000189352) (chuY) ChuY [Chu (VF0234) - Nutritional/Metabolic factor (VFC0272)] [Esch
VFG002220(gb|WP_004686741) (pgm) phosphoglucomutase [LPS (VF0367) - Immune modulation (VFC0258)]
VFG050567(gb|WP_000768964.1) (pilY1) VWA domain-containing protein [TFP (VF1334) - Adherence (VFC
VFG001447(gb|CAA50007) (cnf1) cytotoxic necrotizing factor 1 [CNF-1 (VF0240) - Exotoxin (VFC0235)] [Esc
VFG002076(gb|NP_248780) (clpV1) type VI secretion system AAA+ family ATPase [HSI-I (VF0334) - Effecto
VFG012501(gb|WP_000271272) (iroE) esterase [Salmochelin siderophore (VF0230) - Nutritional/Metabolic fac
VFG000615(gb|NP_709454) (iucA) aerobactin synthesis protein IucA [Aerobactin (VF0123) - Nutritional/Meta
VFG038261(gb|WP_016352107) (exeA) general secretion pathway protein A [Exe T2SS (VF0478) - Effector de
VFG038356 (aopX) hypothetical protein [T3SS (VF0479) - Effector delivery system (VFC0086)] [Aeromonas sa
VFG044330(gb|WP_032447695) (ybtU) yersiniabactin biosynthesis oxidoreductase YbtU [Ybt (VF0564) - Nutri
VFG038399(gb|WP_011705723) (atsP) Type VI secretion system protein [T6SS (VF0480) - Effector delivery sy
VFG045520(gb|WP_010947043) (legC1) Dot/Icm type IV secretion system effector LegC1 [Dot/Icm T4SS secre
VFG041032(gb|NP_251050) (PA2360) hypothetical protein [HSI-3 (VF0944) - Effector delivery system (VFC0
VFG038313(gb|WP_043118380) (exeN) general secretion pathway protein N [Exe T2SS (VF0478) - Effector de
VFG000923(gb|WP_001034892) (fepA) ferrienterobactin outer membrane transporter [Enterobactin (VF0228) -
VFG014629(gb|NP_250146) (fliA) flagellar biosynthesis sigma factor FliA [Flagella (VF0273) - Motility (VFC0
VFG037497(gb|WP_085947929) (ACICU_RS04580) TonB-dependent receptor [HemO cluster (VF0636) - Nutri
VFG000931(gb|WP_001336900) (entC) isochorismate synthase 1 [Enterobactin (VF0228) - Nutritional/Metaboli
VFG048762(gb|WP_015874732) (impA/tssA) type VI secretion system protein TssA [T6SS (VF0569) - Effector
VFG014984(gb|NP_253136) (algW) AlgW protein [Alginate (VF0091) - Biofilm (VFC0271)] [Pseudomonas ae
VFG033475(gb|WP_001273446) (sepL) Type III secretion system secretion gatekeeper SepL [TTSS (VF0172) -
VFG042739(gb|NP_254019) (crc) catabolite repression control protein [Type IV pili (VF0082) - Adherence (VF
VFG037028(gb|WP_002218620) (katA) catalase [KatA (VF0454) - Stress survival (VFC0282)] [Neisseria menin

VFG014369(gb|NP_249789) (fleS) two-component sensor [Flagella (VF0273) - Motility (VFC0204)] [Pseudomo
VFG049915(gb|WP_001142968) (hcp2/tssD2) Type VI secretion system protein, Hcp family [T6SS (VF0579) -
VFG048449(gb|WP_012737621) (fepB) iron-enterobactin transporter periplasmic binding protein [Ent (VF0562)
VFG002529(gb|WP_011205290) (tsr) methyl-accepting chemotaxis protein I [Flagella (VF0430) - Motility (VFC
VFG016041(gb|NP_251092) (pvdI) peptide synthase [Pyoverdine (VF0094) - Nutritional/Metabolic factor (VFC
VFG041260(gb|WP_010946680) (legL1) Dot/Icm type IV secretion system effector LegL1 [Dot/Icm T4SS secre
VFG002306(gb|NP_461449) (ratB) putative outer membrane protein [RatB (VF0399) - Adherence (VFC0001)] [
VFG048632(gb|WP_004176434) (vipA/tssB) type VI secretion system contractile sheath small subunit VipA [T6
VFG001248(gb|NP_249788) (fleQ) transcriptional regulator FleQ [Flagella (VF0273) - Motility (VFC0204)] [Ps
VFG000919(gb|WP_001095619) (chuW) Putative oxygen independent coproporphyrinogen III oxidase [Chu (VF
VFG000181(gb|NP_251792) (xcpS) general secretion pathway protein F [Xcp secretion system (VF0084) - Effec
VFG011510(gb|WP_002964609) (lpsB/lpcC) Lipopolysaccharide core biosynthesis mannosyltransferase LpcC [
VFG038607(gb|WP_016349154) (mshJ) MSHA (mannose-sensitive hemagglutinin) pili biogenesis protein MshJ
VFG013594(gb|WP_164927859) (hgpB) TonB-dependent hemoglobin/transferrin/lactoferrin family receptor [Hg
VFG048297(gb|WP_014906961) (fimG) type 1 fimbrial minor component [Type I fimbriae (VF0566) - Adherenc
VFG000922(gb|WP_000910402) (chuU) heme permease protein ChuU [Chu (VF0227) - Nutritional/Metabolic fa
VFG000364(gb|WP_000140406) (ybtA) transcriptional regulator YbtA [Yersiniabactin (VF0136) - Nutritional/M
VFG048194(gb|WP_003022155) (pdpD) Type VI secretion system secreted protein PdpD [T6SS (VF0544) - Eff
VFG048528(gb|WP_004183335) (entD) enterochelin synthetase component D [Ent (VF0562) - Nutritional/Meta
VFG012329(gb|WP_000046976) (sfaB) S fimbrial switch regulatory protein SfaB [S fimbriae (VF0222) - Adhere
VFG012609(gb|WP_000993317) (chuW) Putative oxygen independent coproporphyrinogen III oxidase [Chu (VF
VFG044322(gb|WP_004190148) (iroE) siderophore esterase IroE [Sal (VF0563) - Nutritional/Metabolic factor (
VFG000938(gb|WP_001015709) (iucC) aerobactin siderophore biosynthesis protein IucC [Aerobactin (VF0229)
VFG007857(gb|NP_217450) (ppsD) phthiocerol synthesis polyketide synthase type I PpsD [PDIM (VF0309) - In
VFG038432(gb|WP_016352168) (tapB) type IV pilus assembly ATPase, PilB-like [Tap type IV pili (VF0475) -
VFG014893(gb|NP_254009) (algC) phosphomannomutase AlgC [Alginate (VF0091) - Biofilm (VFC0271)] [Pse
VFG013948(gb|NP_251650) (pilZ) type 4 fimbrial biogenesis protein PilZ [Type IV pili (VF0082) - Adherence (
VFG044194(gb|WP_003028588) (FTT_RS09560) hypothetical protein [Rhizoferrin (VF0551) - Nutritional/Meta
VFG038892(gb|WP_043118302) (aerA/act) Aerolysin A [Aerolysin (VF0481) - Exotoxin (VFC0235)] [Aeromo
VFG049908(gb|WP_001080148) (tssJ) Type VI secretion system protein TssJ [T6SS (VF0579) - Effector delive
VFG000135(gb|NP_249455) (mucB) anti-sigma factor MucA, inhibitor of alg gene expression [Alginate (VF009
VFG002381(gb|NP_218386) (eccB1) ESX-1 type VII secretion system protein EccB1 [ESX-1 (VF0315) - Effect
VFG038659(gb|WP_011705280) (fliN) flagellar motor switch protein FliN [Polar flagella (VF0473) - Motility (V
VFG049120(gb|WP_012737535) (allB) allantoinase [Allantion utilization (VF0572) - Nutritional/Metabolic fact
VFG045598(gb|WP_010948515) (LPG_RS14265) Dot/Icm type IV secretion system effector [Dot/Icm T4SS sec
VFG000907(gb|WP_000376545) (hlyB) Hemolysin B [-Hemolysin (VF0225) - Exotoxin (VFC0235)] [Escherich
VFG038407(gb|WP_011705728) (vgrG3) VgrG protein [T6SS (VF0480) - Effector delivery system (VFC0086)]
VFG038380(gb|WP_043163044) (vipB) Type VI secretion system contractile sheath large subunit TssC/VipB [T
VFG034895(gb|WP_001304001) (espY1) Type III secretion system effector EspY1 [TTSS secreted effectors (V
VFG041338(gb|WP_010948206) (ceg32/sidI) Dot/Icm type IV secretion system effector Ceg32/SidI [Dot/Icm T
VFG043774(gb|AAB24244) (bont) botulinum neurotoxin, BoNT [BoNT (VF0375) - Exotoxin (VFC0235)] [Clos
VFG000921(gb|WP_000189356) (chuY) ChuY [Chu (VF0227) - Nutritional/Metabolic factor (VFC0272)] [Esch
VFG034754(gb|WP_000081129) (espL1) Type III secretion system effector espL1 [TTSS secreted effectors (VF
VFG016346(gb|WP_001035927) (BAS_RS06430) immune inhibitor A [InhA (VF0536) - Exoenzyme (VFC0251
VFG012865 (ipaH) invasion plasmid antigen IpaH [TTSS secreted effectors (VF0978) - Effector delivery system
VFG001239(gb|NP_249772) (flgF) flagellar basal-body rod protein FlgF [Flagella (VF0273) - Motility (VFC020

VFG002071(gb|NP_248775) (hcp1) type VI secretion system substrate Hcp1 [HSI-I (VF0334) - Effector delivery
VFG038776(gb|WP_016351320) (flgJ) flagellar rod assembly protein/muramidase FlgJ [Polar flagella (VF0473)
VFG002513(gb|WP_004197257) (flgG) flagellar basal body rod protein FlgG [Flagella (VF0430) - Motility (VFC
VFG038464(gb|WP_011707389) (tapT) twitching ATPase [Tap type IV pili (VF0475) - Adherence (VFC0001)]
VFG037650 (pgaA) poly-beta-1,6 N-acetyl-D-glucosamine export porin PgaA [PNAG (VF0472) - Biofilm (VFC
VFG038521(gb|WP_016350155) (flpI) Flp pilus assembly protein FlpI [Flp type IV pili (VF0476) - Adherence (
VFG038832(gb|WP_016351724) (pomB2) flagellar motor protein MotB [Polar flagella (VF0473) - Motility (VFC
VFG002046(gb|WP_000135089) (gspC) general secretion pathway protein C [T2SS (VF0333) - Effector delivery
VFG002499(gb|WP_004197164) (fliI) flagellum-specific ATP synthase FliI [Flagella (VF0430) - Motility (VFC
VFG044159(gb|WP_000125846) (fes) enterobactin/ferric enterobactin esterase [Enterobactin (VF0228) - Nutritic
VFG001241(gb|NP_249774) (flgH) flagellar L-ring protein precursor FlgH [Flagella (VF0273) - Motility (VFC0
VFG000909(gb|WP_000768217) (focA) F1C major fimbrial subunit precursor [F1C fimbriae (VF0224) - Adhere
VFG002055(gb|WP_000097229) (gspL) general secretion pathway protein L [T2SS (VF0333) - Effector delivery
VFG012335(gb|WP_000975448) (sfaE) S fimbriae periplasmic chaperone SfaE [S fimbriae (VF0222) - Adherenc
VFG013292(gb|WP_105163174) (lic3A) alpha-2,3-sialyltransferase [LOS (VF0044) - Immune modulation (VFC
VFG000112(gb|NP_253216) (pilB) type 4 fimbrial biogenesis protein PilB [Type IV pili (VF0082) - Adherence (
VFG002348(gb|WP_005160443) (flgE) flagellar hook protein FlgE [Flagella (VF0394) - Motility (VFC0204)] [Y
VFG048687(gb|WP_002902160) (hcp/tssD) type VI secretion system protein, Hcp family [T6SS (VF0569) - Effe
VFG038448(gb|WP_016351632) (tapN) type IV pilus biogenesis protein PilN [Tap type IV pili (VF0475) - Adhe
VFG001386(gb|NP_215271) (phoP) Possible two component system response transcriptional positive regulator P
VFG000895(gb|WP_000654937) (papC) usher protein PapC [P fimbriae (VF0220) - Adherence (VFC0001)] [Es
VFG002489(gb|WP_181937968) (tssM-5) Type VI secretion system protein TssM [T6SS-1 (VF0429) - Effector
VFG050655(gb|WP_000729757.1) (pilI) chemotaxis protein CheW [TFP (VF1334) - Adherence (VFC0001)] [A
VFG000139(gb|NP_253697) (waaG) B-band O-antigen polymerase [LPS (VF0085) - Immune modulation (VFC
VFG000916(gb|WP_001017192) (chuS) heme oxygenase ChuS [Chu (VF0227) - Nutritional/Metabolic factor (V
VFG049158(gb|WP_001297937) (clbL) colibactin biosynthesis amidase ClbL [Colibactin (VF0573) - Exotoxin (
VFG045464(gb|WP_000562556) (tcpC) Tir domain containing protein TcpC [TcpC (VF0413) - Immune modula
VFG045792(gb|WP_000791650) (csgB) curlin minor subunit CsgB [Curli fibers (VF1138) - Adherence (VFC00
VFG044333(gb|WP_015874968) (fyuA) yersiniabactin receptor FyuA [Ybt (VF0564) - Nutritional/Metabolic fac
VFG002051(gb|WP_001115126) (gspH) general secretion pathway protein H [T2SS (VF0333) - Effector delivery
VFG038417(gb|WP_016349294) (fimD) fimbrial chaperone protein [Type I pili (VF0486) - Adherence (VFC000
VFG006637(gb|WP_001085308) (motB) flagellar motor protein [Flagella (VF0051) - Motility (VFC0204)] [Heli
VFG038381(gb|WP_011705714) (atsG) Type VI secretion system protein [T6SS (VF0480) - Effector delivery sy
VFG001831(gb|WP_010946191) (icmS) Dot/Icm type IV secretion system chaperone IcmS [Dot/Icm (VF0156) -
VFG038375(gb|WP_011705710) (atsC) Type VI secretion system protein [T6SS (VF0480) - Effector delivery sy
VFG006217 (opa) outer membrane beta-barrel protein [Opa (VF0076) - Invasion (VFC0083)] [Neisseria meningi
VFG038703(gb|WP_011705289) (cheZ) chemotaxis protein CheZ [Polar flagella (VF0473) - Motility (VFC0204
VFG049148(gb|WP_041168941) (clbB) colibactin hybrid non-ribosomal peptide synthetase/type I polyketide syn
VFG045601(gb|WP_010948560) (LPG_RS14500) Dot/Icm type IV secretion system effector [Dot/Icm T4SS sec
VFG050164(gb|WP_000723250) (neuC) UDP-N-acetylglucosamine 2-epimerase [K1 capsule (VF0239) - Invasio
VFG002225(gb|WP_002963680) (gmd) GDP-mannose 4,6-dehydratase [LPS (VF0367) - Immune modulation (V
VFG047401(gb|WP_011242229) (feoB) Fe(2+) transporter permease subunit FeoB [FupA (VF0550) - Nutritiona
VFG000872(gb|WP_000044711) (fimE) Type 1 fimbriae Regulatory protein fimE [Type 1 fimbriae (VF0221) - A
VFG000243(gb|WP_002218264) (rfaC) lipopolysaccharide heptosyltransferase I [LOS (VF0078) - Immune modu
VFG038501(gb|WP_043119142) (tapV) type IV pilus assembly protein TapV [Tap type IV pili (VF0475) - Adhe
VFG038600(gb|WP_016349156) (mshL) MSHA (mannose-sensitive hemagglutinin) pili biogenesis protein MshL

VFG000154(gb|NP_250123) (lasI) autoinducer synthesis protein LasI [Quorum sensing (VF0093) - Biofilm (VFC
VFG012597(gb|WP_000089593) (chuA) Outer membrane heme/hemoglobin receptor ChuA [Chu (VF0234) - Nu
VFG001814(gb|NP_215678) (narH) nitrate reductase subunit beta [Nitrate reductase (VF0302) - Nutritional/Meta
VFG047618(gb|WP_003019748) (ileP) major facilitator transporter [IleP (VF0556) - Nutritional/Metabolic facto
VFG037612(gb|WP_000603301) (csuD) Csu pilus usher protein CsuD [Csu fimbriae (VF0461) - Biofilm (VFC0
VFG000918(gb|WP_001312167) (chuT) periplasmic heme-binding protein ChuT [Chu (VF0227) - Nutritional/M
VFG037344(gb|WP_000717759) (basC) acinetobactin biosynthesis protein BasC [Acinetobactin (VF0467) - Nut
VFG000001(gb|WP_010930610) (fhaB) filamentous hemagglutinin/adhesin [FHA (VF0025) - Adherence (VFC0
VFG000877(gb|WP_001244821) (fimF) FimF protein precursor [Type 1 fimbriae (VF0221) - Adherence (VFC0
VFG038886(gb|WP_005314222) (lafE) LafE, flagellar hook control length protein [Lateral flagella (VF0474) - M
VFG038401(gb|WP_011705724) (atsQ) Type VI secretion system protein [T6SS (VF0480) - Effector delivery sy
VFG000940(gb|WP_001299620) (iucA) aerobactin siderophore biosynthesis protein IucD [Aerobactin (VF0229)
VFG002278(gb|WP_011010095) (nagI) hyaluronidase [Mu-toxin (VF0389) - Exoenzyme (VFC0251)] [Clostridi
VFG038635(gb|WP_016350051) (fliH) flagellar assembly protein FliH [Polar flagella (VF0473) - Motility (VFC
VFG002413(gb|WP_000389022) (ykgK/ecpR) regulator protein EcpR [ECP (VF0404) - Adherence (VFC0001)]
VFG038784(gb|WP_011706638) (flgH) flagellar basal body L-ring protein [Polar flagella (VF0473) - Motility (V
VFG000175(gb|NP_251786) (xcpY) general secretion pathway protein L [Xcp secretion system (VF0084) - Effe
VFG000974(gb|WP_010922711) (scpA/scpB) streptococcal C5a peptidase [C5a peptidase (VF0245) - Adherence
VFG034915(gb|WP_000225947) (espY4) Type III secretion system effector EspY4 [TTSS secreted effectors (V
VFG000165(gb|NP_252915) (pchF) pyochelin synthetase PchF [Pyochelin (VF0095) - Nutritional/Metabolic fac
VFG038344(gb|WP_005320766) (ascX) Unknown function, chaperoned by AscY [T3SS (VF0479) - Effector de
VFG042736(gb|NP_253152) (rpoN) RNA polymerase factor sigma-54 [Type IV pili (VF0082) - Adherence (VF
VFG000009(gb|WP_050849356) (fimD) fimbrial adhesin [Fimbriae (VF0031) - Adherence (VFC0001)] [Bordet
VFG038865(gb|WP_005314289) (fliG) flagellar motor switch protein G [Lateral flagella (VF0474) - Motility (V
VFG002358(gb|WP_005164496) (flhC) flagellar biosynthesis transcription activator FlhC [Flagella (VF0394) - M
VFG002069(gb|NP_248773) (hsiB1/vipA/tssB) type VI secretion system tubule-forming protein VipA [HSI-I (V
VFG010467(gb|WP_010946665) (pilP) type IV pilus biogenesis protein PilP [Type IV pili (VF0155) - Adherenc
VFG042734(gb|NP_249343) (vfr) cAMP-regulatory protein [Type IV pili (VF0082) - Adherence (VFC0001)] [P
VFG034795 (espN) Type III secretion system effector EspN [TTSS secreted effectors (VF1110) - Effector delive
VFG041120(gb|WP_011205628) (BPS_RS26880) sensor histidine kinase VirA [T6SS-1 (VF0429) - Effector de
VFG000332(gb|WP_005632797) (rfaD) ADP-L-glycero-D-mannoheptose-6-epimerase [LOS (VF0044) - Immun
VFG011826(gb|YP_002343512) (fliK) flagellar hook-length control protein FliK [Flagella (VF0114) - Motility (
VFG038699(gb|WP_010634617) (cheY) chemotaxis protein CheY [Polar flagella (VF0473) - Motility (VFC020
VFG000802(gb|WP_000953255) (escD) Type III secretion system outer MS ring protein EscD [TTSS (VF0191)
VFG049164(gb|WP_004175305) (clbS) colibactin self-protection protein ClbS [Colibactin (VF0573) - Exotoxin
VFG037852(gb|WP_001075965) (ACICU_RS00395) polysaccharide biosynthesis tyrosine autokinase [Capsule
VFG001444(gb|AAG10151) (aslA) putative arylsulfatase [AslA (VF0238) - Invasion (VFC0083)] [Escherichia c
VFG000618(gb|NP_709457) (iucD) aerobactin synthesis protein IucC, lysine 6-monooxygenase [Aerobactin (VF
VFG039400(gb|NP_819902) (cetCB4) Coxiella Dot/Icm type IVB secretion system translocated effector [T4SS
VFG000333(gb|WP_005694147) (lic2A) glycosyltransferase [LOS (VF0044) - Immune modulation (VFC0258)]
VFG048237(gb|WP_002916189) (fimE) tyrosine recombinase [Type I fimbriae (VF0566) - Adherence (VFC000
VFG038277(gb|WP_016349336) (exeE) general secretory pathway protein E [Exe T2SS (VF0478) - Effector de
VFG038391(gb|WP_011705719) (atsL) Type VI secretion system protein [T6SS (VF0480) - Effector delivery sy
VFG038493(gb|WP_016349428) (tppE) type IV pilus pseudopilin [Tap type IV pili (VF0475) - Adherence (VFC
VFG038472(gb|WP_011706550) (tapW) twitching ATPase [Tap type IV pili (VF0475) - Adherence (VFC0001)]
VFG000885(gb|WP_000265729) (papD) chaperone protein PapD [P fimbriae (VF0220) - Adherence (VFC0001)

VFG049917(gb|WP_000508696) (rhs/PAAR) Type VI secretion system protein, PAAR family [T6SS (VF0579) -
VFG038289(gb|WP_016349339) (exeH) general secretion pathway protein H [Exe T2SS (VF0478) - Effector del
VFG041033(gb|NP_251051) (PA2361) hypothetical protein [HSI-3 (VF0944) - Effector delivery system (VFC00
VFG038816(gb|WP_011706646) (cheV) CheV [Polar flagella (VF0473) - Motility (VFC0204)] [Aeromonas hyd
VFG049152(gb|WP_000337350) (clbF) colibactin biosynthesis dehydrogenase ClbF [Colibactin (VF0573) - Exo
VFG000007(gb|WP_010930611) (fimB) chaperone protein [Fimbriae (VF0031) - Adherence (VFC0001)] [Borde
VFG038533(gb|WP_016350152) (flpF) traffic ATPase [Flp type IV pili (VF0476) - Adherence (VFC0001)] [Aer
VFG000358(gb|WP_002212885) (fyuA/psn) pesticin/yersiniabactin receptor protein [Yersiniabactin (VF0136) -
VFG011414(gb|WP_002964262) (kdsA) 2-dehydro-3-deoxyphosphooctonate aldolase [LPS (VF0367) - Immune
VFG041330(gb|WP_010948110) (lem23) Dot/Icm type IV secretion system effector Lem23 [Dot/Icm T4SS secre
VFG000920(gb|WP_000020034) (chuX) putative heme-binding protein ChuX [Chu (VF0227) - Nutritional/Meta
VFG048654(gb|WP_004224345) (vasE/tssK) type VI secretion system baseplate subunit TssK [T6SS (VF0569) -
VFG034624(gb|WP_000755097) (cfaB) colonization factor antigen 1 [Adhesive fimbriae (VF0213) - Adherence
VFG001420(gb|NP_218327) (erp) exported repetitive protein precursor Erp [Erp (VF0312) - Others (VFC0346)]
VFG038687(gb|WP_016350057) (flhF) flagellar biosynthesis protein FlhF [Polar flagella (VF0473) - Motility (V
VFG043640(gb|WP_043921118) (VV1_RS15640) VWA domain-containing protein [Flp pili (VF0612) - Adhere
VFG000204(gb|NP_250400) (popD) type III secretion system hydrophobic translocator, pore protein PopD [TTS
VFG044335(gb|WP_015874980) (iroB) glucosyltransferase IroB [Sal (VF0563) - Nutritional/Metabolic factor (V
VFG038505(gb|WP_011705643) (tapF) type IV pilus biogenesis/stability protein, pilW-like [Tap type IV pili (V
VFG000028(gb|WP_003808614) (brkB) serum resistance protein [Brk (VF0034) - Immune modulation (VFC025
VFG038529(gb|WP_016350153) (flpG) Flp pilus assembly protein FlpG [Flp type IV pili (VF0476) - Adherence
VFG038869(gb|WP_005314279) (lfgN) lateral flagellar protein, LfgN [Lateral flagella (VF0474) - Motility (VFC
VFG000862(gb|WP_014639426) (pet) Pet serine protease precursor, autotransporter [Pet (VF0217) - Effector del
VFG000166(gb|NP_252916) (pchE) dihydroaeruginoic acid synthetase PchE [Pyochelin (VF0095) - Nutritional/
VFG037521(gb|WP_000837764) (ACICU_RS04595) TonB family protein [HemO cluster (VF0636) - Nutritiona
VFG038305(gb|WP_016349343) (exeL) general secretion pathway protein L [Exe T2SS (VF0478) - Effector del
VFG038155(gb|WP_000209960) (ACICU_RS00500) phosphomannomutase/phosphoglucomutase [Capsule (VF
VFG000361(gb|WP_000982866) (ybtU) yersiniabactin biosynthetic protein YbtU [Yersiniabactin (VF0136) - Nu
VFG002531(gb|WP_011205291) (cheA) chemotaxis two-component sensor kinase CheA [Flagella (VF0430) - M
VFG049133(gb|WP_004177236) (acrA) acriflavine resistance protein A [AcrAB (VF0568) - Antimicrobial activi
VFG001142(gb|WP_042960771) (hopZ) outer membrane protein adhesin [HopZ (VF0054) - Adherence (VFC00
VFG000368(gb|WP_000703040) (ybtS) salicylate synthase Irp9 [Yersiniabactin (VF0136) - Nutritional/Metaboli
VFG022019(gb|NP_214796) (eccA3) Type VII secretion system protein EccA3 [ESX-3 (VF0521) - Effector deli
VFG034825(gb|WP_000484279) (espR3) Type III secretion system effector espR3 [TTSS secreted effectors (VF
VFG048990(gb|WP_001741945) (galF) GalU regulator GalF [Capsule (VF0560) - Immune modulation (VFC025
VFG000477(gb|NP_461845) (rpoS) RNA polymerase sigma factor RpoS [RpoS (VF0112) - Regulation (VFC030
VFG038764(gb|WP_017409846) (flrA) FleQ protein [Polar flagella (VF0473) - Motility (VFC0204)] [Aeromona
VFG038873(gb|WP_005314270) (flgC) flagellar basal body rod protein FlgC [Lateral flagella (VF0474) - Motilit
VFG002054(gb|WP_000633233) (gspK) general secretion pathway protein K [T2SS (VF0333) - Effector delivery
VFG001269(gb|WP_010929996) (cyaB) cyclolysin secretion ATP-binding protein [Cya (VF0028) - Exotoxin (V
VFG000813(gb|WP_001037814) (escV) Type III secretion system major export apparatus protein [TTSS (VF019
VFG001263(gb|NP_250143) (flhA) flagellar biosynthesis protein FlhA [Flagella (VF0273) - Motility (VFC0204)
VFG039912(gb|WP_011191202) (rvhB4b) type IV secretion system protein VirB4 precursor [Rvh T4SS (VF048
VFG001400(gb|NP_216865) (plcC) phospholipase C [Phospholipase C (VF0320) - Exotoxin (VFC0235)] [Myco
VFG002288(gb|WP_009902069) (toxB) toxin B [TcdB (VF0377) - Exotoxin (VFC0235)] [Clostridium difficile 6
VFG002287(gb|WP_011860904) (toxA) toxin A [TcdA (VF0376) - Exotoxin (VFC0235)] [Clostridium difficile

VFG049909(gb|WP_001113715) (fha) Type VI secretion system protein Fha [T6SS (VF0579) - Effector delivery
VFG000184(gb|NP_251795) (xcpQ) general secretion pathway protein D [Xcp secretion system (VF0084) - Effe
VFG050161(gb|WP_000579525) (kpsC) Capsule polysaccharide export protein [K1 capsule (VF0239) - Invasion
VFG000915(gb|WP_001234179) (focH) F1C Putative fimbrial adhesin precursor [F1C fimbriae (VF0224) - Adhe
VFG038596(gb|WP_016349157) (mshM) membrane protein with ATPase domain, GspA-like [MSHA type IV pi
VFG002534(gb|WP_004198197) (motA) flagellar motor protein MotA [Flagella (VF0430) - Motility (VFC0204)
VFG000873(gb|WP_000695571) (fimA) Type-1 fimbrial protein, A chain precursor [Type 1 fimbriae (VF0221) -
VFG005653 (nanA) neuraminidase A [Neuraminidase (VF0148) - Exoenzyme (VFC0251)] [Streptococcus pneu
VFG000157(gb|NP_249535) (plcH) hemolytic phospholipase C precursor [PLC (VF0092) - Exotoxin (VFC0235
VFG045794(gb|WP_000992818) (csgC) curli assembly protein CsgC [Curli fibers (VF1138) - Adherence (VFC0
VFG022082(gb|YP_177711) (PPE4) PPE family protein [ESX-3 (VF0521) - Effector delivery system (VFC0086
VFG045481(gb|WP_010945891) (LPG_RS00665) hypothetical protein [Dot/Icm T4SS secreted effectors (VF07
VFG002048(gb|WP_000249362) (gspE) general secretion pathway protein E [T2SS (VF0333) - Effector delivery
VFG037720(gb|WP_001027056) (adeG) cation/multidrug efflux pump [AdeFGH efflux pump (VF0504) - Biofil
VFG034636(gb|WP_000358674) (cfaC) colonisation factor antigen c, usher [Adhesive fimbriae (VF0213) - Adhe
VFG045343(gb|WP_002210880) (yapJ) autotransporter protein YapJ [YapJ (VF0509) - Adherence (VFC0001)]
VFG038804(gb|WP_011706643) (flgC) flagellar basal body rod protein FlgC [Polar flagella (VF0473) - Motility
VFG000936(gb|WP_016265865) (iutA) ferric aerobactin receptor precusor IutA [Aerobactin (VF0229) - Nutritio
VFG048676(gb|WP_014907726) (ompA) OmpA family protein [T6SS (VF0569) - Effector delivery system (VF
VFG002426(gb|WP_004194352) (pilB) type IV pilus assembly protein [Type IV pili (VF0431) - Adherence (VF
VFG043626(gb|WP_000820818) (mrkB) fimbrial chaperone protein mrkB precursor [Type 3 fimbriae (VF0567)
VFG045345(gb|WP_005457104) (mam7) multivalent adhesion molecule MAM7 [MAM7 (VF0512) - Adherence
VFG038845(gb|WP_016351249) (nueA) NeuA protein [Polar flagella (VF0473) - Motility (VFC0204)] [Aeromo
VFG000008(gb|WP_010930612) (fimC) outer membrane usher protein precursor [Fimbriae (VF0031) - Adheren
VFG002089(gb|WP_000073077) (vasL) type VI secretion system protein VasL [VAS T6SS (VF0335) - Effector
VFG000935(gb|WP_001222189) (iroN) salmochelin receptor IroN [Salmochelin siderophore (VF0230) - Nutritio
VFG034873(gb|WP_001270084) (espX5) Type III secretion system effector EspX5 [TTSS secreted effectors (V
VFG038796(gb|WP_016351321) (flgE) flagellar hook protein FlgE [Polar flagella (VF0473) - Motility (VFC020
VFG048708(gb|WP_041168969) (vgrG/tssI) type VI secretion system tip protein VgrG [T6SS (VF0569) - Effect
VFG005361(gb|WP_002352221) (spaP/pac) cell surface antigen SpaP [SpaP (VF0526) - Adherence (VFC0001)
VFG048439(gb|WP_014907941) (entC) isochorismate synthase [Ent (VF0562) - Nutritional/Metabolic factor (V
VFG000886(gb|WP_000261297) (papJ) P pilus assembly protein PapJ [P fimbriae (VF0220) - Adherence (VFC0
VFG001360(gb|WP_001042179) (nanB) neuraminidase B [Neuraminidase (VF0148) - Exoenzyme (VFC0251)]
VFG044165(gb|WP_001041793) (entS) enterobactin exporter, iron-regulated [Enterobactin (VF0228) - Nutrition
VFG001408(gb|NP_217456) (mas) multifunctional mycocerosic acid synthase [PDIM (VF0309) - Immune modu
VFG000190(gb|NP_250386) (pscP) type III secretion system protein PscP [TTSS (VF0083) - Effector delivery s
VFG043648(gb|NP_252312) (rpoS) RNA polymerase sigma factor RpoS [Type IV pili (VF0082) - Adherence (V
VFG024776(gb|NP_217466) (fadD29) long-chain-fatty-acid--AMP ligase FadD29 [PDIM (VF0309) - Immune m
VFG000174(gb|NP_251785) (xcpZ) general secretion pathway protein M [Xcp secretion system (VF0084) - Effe
VFG041016(gb|NP_250349) (PA1658) hypothetical protein [HSI-2 (VF0943) - Effector delivery system (VFC00
VFG000478(gb|NP_459678) (fur) ferric iron uptake transcriptional regulator [Fur (VF0113) - Regulation (VFC0
VFG001257(gb|NP_250135) (fliN) flagellar motor switch protein FliN [Flagella (VF0273) - Motility (VFC0204)
VFG048307(gb|WP_041937815) (fimH) type 1 fimbrial adhesin precursor [Type I fimbriae (VF0566) - Adheren
VFG050342(gb|WP_000076098.1) (pilO) type 4a pilus biogenesis protein PilO [TFP (VF1334) - Adherence (VF
VFG041013(gb|NP_250203) (hcpA) secreted protein Hcp [HSI-2 (VF0943) - Effector delivery system (VFC008
VFG000811(gb|WP_001062956) (escO) positive regulator EscO [TTSS (VF0191) - Effector delivery system (V

VFG041029(gb|NP_250362) (stk1) serine-threonine kinase Stk1 [HSI-2 (VF0943) - Effector delivery system (VF
VFG010850 (rtxA) structural toxin protein RtxA [RtxA (VF0259) - Exotoxin (VFC0235)] [Legionella pneumoph
VFG001932(gb|YP_002343539) (cdtC) cytolethal distending toxin C [CDT (VF0115) - Exotoxin (VFC0235)] [C
VFG002164(gb|WP_002395058) (prgB/asc10) aggregation substance PrgB/Asc10 [AS (VF0352) - Adherence (V
VFG001396(gb|NP_216424) (katG) catalase-peroxidase [KatG (VF0303) - Stress survival (VFC0282)] [Mycoba
VFG042738(gb|NP_253646) (fimX) protein FimX [Type IV pili (VF0082) - Adherence (VFC0001)] [Pseudomo
VFG038132(gb|WP_000045495) (pgi) glucose-6-phosphate isomerase [Capsule (VF0465) - Immune modulation
VFG001224(gb|NP_249087) (pilU) twitching motility protein PilU [Type IV pili (VF0082) - Adherence (VFC00
VFG042068(gb|NP_459947) (sopD2) type III secretion system effector SopD2 [TTSS-2 secreted effectors (VF0
VFG038395(gb|WP_011705721) (clpB) type VI secretion system ATPase ClpV1 [T6SS (VF0480) - Effector deli
VFG013368(gb|WP_005693225) (rffG) dTDP-glucose 46-dehydratase [LOS (VF0044) - Immune modulation (V
VFG034679(gb|WP_000074207) (ibeB) Cu(+)/Ag(+) efflux RND transporter outer membrane channel CusC [Ib
VFG036992(gb|WP_002224969) (mtrD) multiple transferable resistance system protein MtrD [MtrCDE (VF045
VFG038663(gb|WP_005298795) (fliO) flagellar biosynthesis protein, FliO [Polar flagella (VF0473) - Motility (V
VFG048797(gb|WP_004175261) (ugd) UDP-glucose 6-dehydrogenase [Capsule (VF0560) - Immune modulation
VFG034866(gb|WP_000900547) (espX4) Type III secretion system effector EspX4 [TTSS secreted effectors (V
VFG002053(gb|WP_001250462) (gspJ) general secretion pathway protein J [T2SS (VF0333) - Effector delivery
VFG005289(gb|WP_001868236) (GBS_RS03585) tip adhesin protein [PI-1 (VF0527) - Adherence (VFC0001)]
VFG038176(gb|WP_000777882) (ompA) outer membrane protein OmpA [OmpA (VF0464) - Immune modulatio
VFG001012(gb|NP_858286) (spa24) type III secretion system minor export apparatus protein Spa24 [TTSS (VF
VFG001817(gb|NP_216900) (mbtA) Bifunctional enzyme MbtA: salicyl-AMP ligase (SAL-AMP ligase) + salic
VFG013260(gb|WP_010868968) (lgtC) glycosyltransferase [LOS (VF0044) - Immune modulation (VFC0258)] [
VFG000906(gb|WP_001142375) (hlyA) Hemolysin A [-Hemolysin (VF0225) - Exotoxin (VFC0235)] [Escheric
VFG000939(gb|WP_001287497) (iucB) aerobactin siderophore biosynthesis protein IucB [Aerobactin (VF0229)
VFG000163(gb|NP_251088) (fpvA) ferripyoverdine receptor FpvA [Pyoverdine (VF0094) - Nutritional/Metabol
VFG044081(gb|NP_250946) (pvcC) paerucumarin biosynthesis protein PvcC [Pyoverdine (VF0094) - Nutritiona
VFG048348(gb|WP_004149658) (mrkJ) phosphodiesterase [Type 3 fimbriae (VF0567) - Biofilm (VFC0271)] [K
VFG038723(gb|WP_011705294) (AHML_RS07540) CobQ/CobB/MinD/ParA family protein [Polar flagella (VF
VFG039311(gb|NP_819427) (cetCb2) Coxiella Dot/Icm type IVB secretion system translocated effector [T4SS s
VFG000845(gb|WP_000581721) (toxB) cytotoxin [ToxB (VF0203) - Effector delivery system (VFC0086)] [Esc
VFG038364(gb|WP_005320615) (aexT) Type III secretion system effector AexT, bifunctional toxin with GTPas
VFG049902(gb|WP_011310140) (tssM) Type VI secretion system protein TssM [T6SS (VF0579) - Effector deli
VFG001238(gb|NP_249771) (flgE) flagellar hook protein FlgE [Flagella (VF0273) - Motility (VFC0204)] [Pseu
VFG015983(gb|NP_251083) (pvdM) dipeptidase precursor [Pyoverdine (VF0094) - Nutritional/Metabolic factor
VFG049124(gb|WP_012737540) (allD) ureidoglycolate dehydrogenase [Allantion utilization (VF0572) - Nutriti
VFG038651(gb|WP_011705278) (fliL) flagellar basal body-associated protein FliL [Polar flagella (VF0473) - M
VFG000903(gb|WP_001045652) (pic) Pic serine protease precursor, autotransporter [Pic (VF0232) - Effector del
VFG038281(gb|WP_016349337) (exeF) general secretion pathway protein F [Exe T2SS (VF0478) - Effector deli
VFG038545(gb|WP_043118798) (flpC) Flp pilus secretin FlpC [Flp type IV pili (VF0476) - Adherence (VFC00
VFG001259(gb|NP_250137) (fliP) flagellar biosynthetic protein FliP [Flagella (VF0273) - Motility (VFC0204)]
VFG049154(gb|WP_014907257) (clbH) colibactin non-ribosomal peptide synthetase ClbH [Colibactin (VF0573)
VFG047068(gb|WP_003022285) (wzx) O-antigen flippase [LPS (VF0542) - Immune modulation (VFC0258)] [F
VFG048621(gb|WP_004213924) (iutA) ferric aerobactin receptor IutA [Aerobactin (VF0565) - Nutritional/Meta
VFG047055(gb|WP_003022283) (wbtK) glycosyltransferase [LPS (VF0542) - Immune modulation (VFC0258)]
VFG012603(gb|WP_001302516) (chuT) periplasmic heme-binding protein ChuT [Chu (VF0234) - Nutritional/M
VFG000126(gb|NP_252234) (algE) alginate biosynthetic protein AlgK precursor [Alginate (VF0091) - Biofilm (

VFG038497(gb|WP_016351992) (tppF) type IV pilus pseudopilin [Tap type IV pili (VF0475) - Adherence (VFC
VFG037274(gb|WP_001281538) (barA) siderophore efflux system of the ABC superfamily [Acinetobactin (VF0
VFG000065(gb|WP_010930832) (bscD) putative type III secretion protein [TTSS (VF0035) - Effector delivery s
VFG037625(gb|WP_001022730) (csuE) Csu pilus tip adhesin CsuE [Csu fimbriae (VF0461) - Biofilm (VFC027
VFG048478(gb|WP_004147521) (fepG) iron-enterobactin transporter permease [Ent (VF0562) - Nutritional/Met
VFG001218(gb|NP_253727) (pilQ) type 4 fimbrial biogenesis protein PilQ [Type IV pili (VF0082) - Adherence
VFG049914(gb|WP_000037389) (tssB) Type VI secretion system protein TssB [T6SS (VF0579) - Effector deliv
VFG000367(gb|WP_001286280) (ybtX) putative signal transducer [Yersiniabactin (VF0136) - Nutritional/Metab
VFG038348(gb|WP_005320755) (ascN) Type III secretion system ATPase [T3SS (VF0479) - Effector delivery s
VFG048665(gb|WP_004176431) (dotU/tssL) type VI secretion system protein, DotU/TssL family [T6SS (VF056
VFG043373(gb|WP_000342347) (cheA) histidine kinase CheA [Flagella (VF0051) - Motility (VFC0204)] [Helic
VFG048468(gb|WP_004176917) (fepD) iron-enterobactin transporter membrane protein [Ent (VF0562) - Nutriti
VFG034875(gb|WP_001270087) (espX5) Type III secretion system effector EspX5 [TTSS secreted effectors (V
VFG001919(gb|YP_002344702) (pseG) UDP-2,4-diacetamido-2,4,6-trideoxy-beta-L-altropyranose hydrolase [F
VFG011671(gb|YP_002344801) (Cj1418c) hypothetical protein [Capsule (VF0323) - Immune modulation (VFC
VFG000123(gb|NP_252231) (alg8) alginate-c5-mannuronan-epimerase AlgG [Alginate (VF0091) - Biofilm (VF
VFG048257(gb|WP_004174507) (fimI) type 1 pilus biosynthesis fimbrial protein [Type I fimbriae (VF0566) - A
VFG000319 (wbpB) lipopolysaccharide biosynthesis protein [LPS (VF0056) - Immune modulation (VFC0258)]
VFG000162(gb|NP_251116) (pvdS) extracytoplasmic-function sigma-70 factor [Pyoverdine (VF0094) - Nutritio
VFG002300(gb|CAA88565) (tcnA) alpha-toxin [Alpha-toxin (novyi) (VF0386) - Exotoxin (VFC0235)] [Clostrid
VFG038387(gb|WP_011705717) (atsJ) Type VI secretion system protein [T6SS (VF0480) - Effector delivery sys
VFG012509(gb|WP_001111200) (iroC) ATP binding cassette transporter [Salmochelin siderophore (VF0230) -
VFG048779(gb|WP_004190503) (tssG) type VI secretion system baseplate subunit TssG [T6SS (VF0569) - Effe
VFG000366(gb|WP_002212761) (ybtQ) yersiniabactin ABC transporter ATP-binding/permease protein YbtQ [Y
VFG002050(gb|WP_001087302) (gspG) general secretion pathway protein G [T2SS (VF0333) - Effector deliver
VFG045632(gb|WP_002286924) (ecbA/fss3) Collagen binding MSCRAMM, EcbA/Fss3 [EcbA (VF0539) - Adh
VFG034761(gb|WP_001121629) (espL2) Type III secretion system secreted effector EspL2, cysteine protease [T
VFG005197(gb|WP_000006706) (pavA) Fibronectin-binding protein-like protein A [PavA (VF0283) - Adherenc
VFG039517 (CBU_1525*) Coxiella Dot/Icm type IVB secretion system translocated effector [T4SS secreted eff
VFG001413(gb|NP_217803) (sigF) RNA polymerase sigma factor SigF [SigF (VF0294) - Regulation (VFC0301
VFG001445(gb|AAA92657) (AAA92657) unknown protein [TraJ (VF0241) - Invasion (VFC0083)] [Escherichia
VFG037177(gb|WP_000632986) (plc2) phospholipase C [Phospholipase C (VF0470) - Exotoxin (VFC0235)] [A
VFG000029(gb|WP_010929608) (bplL) lipopolysaccharide biosynthesis protein [LPS (VF0033) - Immune modu
VFG050166(gb|WP_000066003) (neuB) N-acylneuraminate-9-phosphate synthase [K1 capsule (VF0239) - Invas
VFG038460(gb|WP_043119325) (tapQ) type IV pilus secretin PilQ [Tap type IV pili (VF0475) - Adherence (VF
VFG045351 (APH_RS00640*) P44/Msp2 family outer membrane protein [P44/Msp2 family (VF0520) - Immune
VFG044329(gb|WP_015874964) (irp1) yersiniabactin polyketide synthase HMWP1 [Ybt (VF0564) - Nutritional/
VFG038405(gb|WP_011705727) (AHA_RS09305) type VI secretion system PAAR protein [T6SS (VF0480) - E
VFG038436(gb|WP_016352169) (tapC) type IV fimbrial assembly protein, PilC-like [Tap type IV pili (VF0475)
VFG001232(gb|NP_249105) (chpB) probable methylesterase [Type IV pili (VF0082) - Adherence (VFC0001)] [
VFG050912(gb|WP_000898330.1) (vgrG/tssI) type VI secretion system tip protein VgrG [T6SS (VF1337) - Effe
VFG038369(gb|WP_011705706) (hcp) hemolysin co-regulated protein [T6SS (VF0480) - Effector delivery syste
VFG049073(gb|WP_015875021) (KP1_RS17240) DUF4422 domain-containing protein [LPS (VF0561) - Immun
VFG038488(gb|WP_016349432) (tapY1) type IV pilus biogenesis protein [Tap type IV pili (VF0475) - Adheren
VFG038780(gb|WP_011706637) (flgI) flagellar basal body P-ring protein [Polar flagella (VF0473) - Motility (V
VFG000933(gb|WP_001007140) (entB) isochorismatase [Enterobactin (VF0228) - Nutritional/Metabolic factor (

VFG012162(gb|WP_011590241) (nanJ) exo-alpha-sialidase [Sialidase (VF0391) - Exoenzyme (VFC0251)] [Clos
VFG051042(gb|WP_000653195.1) (hcp/tssD) type VI secretion system tube protein Hcp [T6SS (VF1337) - Effec
VFG000347(gb|WP_010869038) (hgpC) hemoglobin-binding protein [Hgp (VF0270) - Nutritional/Metabolic fac
VFG000928(gb|WP_000640938) (fepG) iron-enterobactin ABC transporter permease [Enterobactin (VF0228) - N
VFG044325(gb|WP_002212761) (ybtQ) yersiniabactin ABC transporter ATP-binding/permease protein YbtQ [Y
VFG038627(gb|WP_016350050) (fliF) flagellar MS-ring protein [Polar flagella (VF0473) - Motility (VFC0204)]
VFG037777(gb|WP_000867091) (lpsB) glycosyltransferase family 4 protein [LPS (VF0466) - Immune modulatio
VFG048808(gb|WP_004899416) (rfbK1) O9 family phosphomannomutase RfbK1 [Capsule (VF0560) - Immune
VFG000912(gb|WP_001443207) (focD) F1C fimbrial usher [F1C fimbriae (VF0224) - Adherence (VFC0001)] [
VFG034575(gb|WP_000715633) (aafB) invasin homolog AafB [AAFs (VF0214) - Adherence (VFC0001)] [Esch
VFG001230(gb|NP_251805) (fimV) a polar peptidoglycan-binding protein involved in type IV pilus assembly [T
VFG037316(gb|WP_000744381) (entE) non-ribosomal peptide synthetase adenylate-forming enzyme of acinetob
VFG045525(gb|WP_010947213) (LPG_RS07435) Dot/Icm type IV secretion system effector [Dot/Icm T4SS sec
VFG048327(gb|WP_004152886) (mrkH) transcriptional activator [Type 3 fimbriae (VF0567) - Biofilm (VFC027
VFG041340(gb|WP_010948225) (lem26) Dot/Icm type IV secretion system effector Lem26 [Dot/Icm T4SS secre
VFG000925(gb|WP_000140634) (fepC) ferrienterobactin ABC transporter ATPase [Enterobactin (VF0228) - Nu
VFG038707(gb|WP_016350059) (cheA-2) chemotaxis protein CheA [Polar flagella (VF0473) - Motility (VFC02
VFG044331(gb|WP_015874966) (ybtT) type II thioesterase YbtT [Ybt (VF0564) - Nutritional/Metabolic factor (
VFG048459(gb|WP_004147525) (entS) enterobactin exporter EntS [Ent (VF0562) - Nutritional/Metabolic factor
VFG000429(gb|WP_010989299) (tviD) Vi polysaccharide biosynthesis protein TviD [Vi antigen (VF0101) - Imm
VFG002173(gb|WP_010785625) (asa1) aggregation substance Asa1 [AS (VF0352) - Adherence (VFC0001)] [En
VFG050162(gb|WP_001161829) (kpsS) Capsule polysaccharide export protein [K1 capsule (VF0239) - Invasion
VFG043623(gb|WP_002916122) (mrkF) type 3 fimbrial minor pilin subunit MrkF [Type 3 fimbriae (VF0567) - B
VFG034815(gb|WP_000671689) (espR1) Type III secretion system effector espR1 [TTSS secreted effectors (VF
VFG048547(gb|WP_004213617) (iroN) salmochelin receptor IroN [Sal (VF0563) - Nutritional/Metabolic factor (
VFG044172(gb|WP_000622316) (chuV) ATP-binding hydrophilic protein ChuV [Chu (VF0227) - Nutritional/M
VFG001271(gb|WP_010929998) (cyaE) cyclolysin secretion protein [Cya (VF0028) - Exotoxin (VFC0235)] [Bo
VFG000801(gb|WP_001273445) (sepL) Type III secretion system secretion gatekeeper SepL [TTSS (VF0191) -
VFG048429(gb|WP_004183353) (entE) enterobactin synthase subunit E [Ent (VF0562) - Nutritional/Metabolic f
VFG000942(gb|AAK16475) (draA) cytoplamic protein DraA [Dr adhesins (VF0223) - Adherence (VFC0001)] [I
VFG038611(gb|WP_016349153) (mshI1) MSHA (mannose-sensitive hemagglutinin) pili biogenesis protein Msh
VFG049155(gb|WP_000829570) (clbI) colibactin polyketide synthase ClbI [Colibactin (VF0573) - Exotoxin (VF
VFG041018(gb|NP_250351) (PA1660) hypothetical protein [HSI-2 (VF0943) - Effector delivery system (VFC00
VFG038836(gb|WP_011707085) (pomA2) chemotaxis protein PomA [Polar flagella (VF0473) - Motility (VFC02
VFG041010(gb|NP_248760) (tagQ) type VI secretiona ssociated protein TagQ, outer membrane lipoprotein [HSI
VFG038768(gb|WP_016351318) (flgL) flagellar hook-associated protein FlgL [Polar flagella (VF0473) - Motility
VFG012337(gb|WP_001370792) (sfaF) S fimbriae outer membrane usher SfaF [S fimbriae (VF0222) - Adherenc
VFG045788(gb|WP_001189322) (cgsG) curli production assembly/transport protein CsgG [Curli fibers (VF1138
VFG000905(gb|WP_001546261) (hlyC) Hemolysin C [-Hemolysin (VF0225) - Exotoxin (VFC0235)] [Escherich
VFG000218(gb|NP_250414) (pscJ) type III secretion system inner MS ring protein [TTSS (VF0083) - Effector de
VFG049904 (clpV/tssH) Type VI secretion system ATPase ClpV/TssH [T6SS (VF0579) - Effector delivery syste
VFG037586(gb|WP_000876480) (csuB) Csu pilus subunit CsuB [Csu fimbriae (VF0461) - Biofilm (VFC0271)]
VFG038760(gb|WP_016351312) (flrB) two-component sensor kinase [Polar flagella (VF0473) - Motility (VFC0
VFG002536(gb|WP_004530401) (fliD) flagellar capping protein FliD [Flagella (VF0430) - Motility (VFC0204)]
VFG034780(gb|WP_001132163) (espM1) Type III secretion system effector EspM1, Rho guanine exchange facto
VFG000930(gb|WP_000077767) (entF) enterobactin synthase multienzyme complex component, ATP-dependen

VFG000130(gb|NP_252238) (algI) alginate o-acetyltransferase AlgI [Alginate (VF0091) - Biofilm (VFC0271)] [
VFG038444(gb|WP_075384315) (tapM) type IV pilus assembly protein PilM [Tap type IV pili (VF0475) - Adhe
VFG000122(gb|NP_252230) (algD) GDP-mannose 6-dehydrogenase AlgD [Alginate (VF0091) - Biofilm (VFC0
VFG034721(gb|WP_000556298) (ibeC) phosphoethanolamine transferase CptA [Ibes (VF0237) - Invasion (VFC
VFG038517(gb|WP_016350156) (flpJ) Flp pilus pseudopilin [Flp type IV pili (VF0476) - Adherence (VFC0001)
VFG037428(gb|WP_000939834) (basB) non-ribosomal peptide synthetase with condensation and peptidyl carrie
VFG045349(gb|WP_000092542) (fdeC) adhesin FdeC [FdeC (VF0506) - Adherence (VFC0001)] [Escherichia co
VFG038590(gb|WP_043118275) (mshE) ATPase protein MshE, GspE-like [MSHA type IV pili (VF0477) - Adh
VFG048277(gb|WP_009308599) (fimD) outer membrane usher protein [Type I fimbriae (VF0566) - Adherence
VFG034756 (espL1) Type III secretion system effector espL1 [TTSS secreted effectors (VF1111) - Effector deli
VFG038711(gb|WP_016350060) (cheB-2) chemotaxis-specific methylesterase [Polar flagella (VF0473) - Motilit
VFG038639(gb|WP_011705275) (fliI) flagellum-specific ATP synthase [Polar flagella (VF0473) - Motility (VFC
VFG049906 (tssK) Type VI secretion system protein TssK [T6SS (VF0579) - Effector delivery system (VFC008
VFG000929(gb|WP_001375021) (entD) phosphopantetheinyl transferase component of enterobactin synthase mu
VFG000875(gb|WP_000066579) (fimC) Chaperone protein fimC precursor [Type 1 fimbriae (VF0221) - Adhere
VFG051000(gb|WP_000568832.1) (tssF) type VI secretion system baseplate subunit TssF [T6SS (VF1337) - Eff
VFG038293(gb|WP_016349340) (exeI) general secretion pathway protein I [Exe T2SS (VF0478) - Effector deliv
VFG000883(gb|WP_001239363) (papH) P pilus termination subunit PapH [P fimbriae (VF0220) - Adherence (V
VFG015515(gb|NP_252903) (phzE1) phenazine biosynthesis protein PhzE [Pyocyanin (VF0100) - Nutritional/M
VFG038285(gb|WP_005308483) (exeG) general secretion pathway protein G [Exe T2SS (VF0478) - Effector del
VFG000362(gb|WP_002212777) (irp1) yersiniabactin biosynthetic protein Irp1 [Yersiniabactin (VF0136) - Nutri
VFG044025(gb|WP_012669495) (C2II) C2 toxin (component-II) [C2 toxin (VF0384) - Exotoxin (VFC0235)] [C
VFG000926(gb|WP_001443194) (fepD) ferrienterobactin ABC transporter permease [Enterobactin (VF0228) - N
VFG038792(gb|WP_011706640) (flgF) flagellar basal-body rod protein FlgF [Polar flagella (VF0473) - Motility
VFG046118(gb|WP_011205945) (mhp107) P97 paralog, surface adhesin [P97/P102 paralog family (VF0426) - A
VFG049900 (tssM) Type VI secretion system protein TssM [T6SS (VF0579) - Effector delivery system (VFC00
VFG050163(gb|WP_000197083) (neuE) polysialic acid biosynthesis protein [K1 capsule (VF0239) - Invasion (V
VFG048317(gb|WP_014906959) (fimK) transcriptional regulator [Type I fimbriae (VF0566) - Adherence (VFC0
VFG043625(gb|WP_000813718) (mrkC) fimbrial biogenesis outer membrane usher protein mrkC precursor [Typ
VFG002420(gb|WP_000645781) (adsA) Adenosine synthase A [AdsA (VF0422) - Immune modulation (VFC02
VFG001265(gb|NP_250145) (fleN) flagellar synthesis regulator FleN [Flagella (VF0273) - Motility (VFC0204)]
VFG012513(gb|WP_001221122) (iroB) glucosyltransferase IroB [Salmochelin siderophore (VF0230) - Nutrition
VFG002437(gb|WP_011325557) (boaB) autotransporter protein [BoaB (VF0435) - Adherence (VFC0001)] [Bur
VFG048419(gb|WP_004210873) (entB) 2,3-dihydro-2,3-dihydroxybenzoate synthetase, isochroismatase [Ent (V
VFG034648(gb|WP_001245988) (cfaD/cfaE) minor pilin and initiator [Adhesive fimbriae (VF0213) - Adherence
VFG034849(gb|WP_000129338) (espX1) Type III secretion system effector EspX1 [TTSS secreted effectors (V
VFG001359(gb|WP_000733059) (psaA) manganese ABC transporter, manganese-binding adhesion liprotein [Ps
VFG049051(gb|WP_014907239) (KP1_RS17230) glycosyltransferase [LPS (VF0561) - Immune modulation (V
VFG001254(gb|NP_249795) (fliI) flagellum-specific ATP synthase FliI [Flagella (VF0273) - Motility (VFC0204
VFG000879(gb|WP_000832235) (fimH) FimH protein precursor [Type 1 fimbriae (VF0221) - Adherence (VFC0
VFG048985(gb|WP_014907219) (KP1_RS17355) phosphatase PAP2 family protein [Capsule (VF0560) - Immu
VFG044044(gb|AAB29385) (trhX) TDH-related hemolysin [TRH (VF0518) - Exotoxin (VFC0235)] [Vibrio par
VFG047149(gb|WP_003029668) (wbtF) NAD dependent epimerase [LPS (VF0542) - Immune modulation (VFC
VFG048755(gb|WP_049127998) (icmF/tssM) type VI secretion protein TssM [T6SS (VF0569) - Effector delive
VFG000434(gb|NP_490509) (pefC) plasmid-encoded fimbriae usher protein PefC [Pef (VF0104) - Adherence (V
VFG002370(gb|WP_011816870) (YE_RS15455) EpsG family protein [O-antigen (VF0392) - Immune modulatio

VFG012036(gb|YP_002344725) (pseE/maf5) motility accessory factor PseE [Flagella (VF0114) - Motility (VFC
VFG000934(gb|WP_000348393) (entA) 2,3-dihydro-2,3-dihydroxybenzoate dehydrogenase EntA [Enterobactin
VFG000874(gb|WP_000824100) (fimI) Fimbrin-like protein fimI precursor [Type 1 fimbriae (VF0221) - Adhere
VFG048267(gb|WP_015875145) (fimC) periplasmic chaperone [Type I fimbriae (VF0566) - Adherence (VFC00
VFG048247(gb|WP_002916190) (fimA) type 1 major fimbrial subunit precursor [Type I fimbriae (VF0566) - Ad
VFG010463(gb|WP_010946666) (pilQ) type IV pilus secretin protein PilQ [Type IV pili (VF0155) - Adherence
VFG049160(gb|WP_001327259) (clbN) colibactin non-ribosomal peptide synthetase ClbN [Colibactin (VF0573)
VFG045790(gb|WP_000833288) (cgsE) curli production assembly/transport protein CsgE [Curli fibers (VF1138)
VFG037505(gb|WP_002002089) (ACICU_RS04585) transferrin-binding protein-like solute binding protein [He
VFG038331(gb|WP_005320790) (exsD) Type III secretion system protein ExsD [T3SS (VF0479) - Effector deli
VFG001267(gb|NP_252913) (pchH) ABC transporter ATP-binding protein [Pyochelin (VF0095) - Nutritional/M
VFG038342(gb|WP_005320770) (ascV) Type III secretion system inner membrane export apparatus protein Asc
VFG002356(gb|WP_011816614) (flhA) flagellar biosynthesis protein FlhA [Flagella (VF0394) - Motility (VFC0
VFG045789(gb|WP_001264082) (cgsF) curli production assembly/transport protein CsgF [Curli fibers (VF1138)
VFG048229(gb|WP_015875142) (fimB) tyrosine recombinase [Type I fimbriae (VF0566) - Adherence (VFC000
VFG041235(gb|WP_010945851) (lem1) Dot/Icm type IV secretion system effector Lem1 [Dot/Icm T4SS secrete
VFG002431(gb|WP_004187516) (pilR) type IV pilus biosynthesis protein PilR [Type IV pili (VF0431) - Adhere
VFG002319(gb|WP_005160341) (fliA) flagellar biosynthesis sigma factor [Flagella (VF0394) - Motility (VFC02
VFG050160(gb|WP_000030755) (kpsU) 3-deoxy-manno-octulosonate cytidylyltransferase [K1 capsule (VF0239
VFG042568(gb|WP_000012016) (sfaY) transcriptional regulator SfaY [F1C fimbriae (VF0224) - Adherence (VF
VFG002380(gb|NP_218385) (eccA1) ESX-1 type VII secretion system AAA+ ATPase EccA1 [ESX-1 (VF0315)
VFG048338(gb|WP_002916079) (mrkI) LuxR family regulatory protein [Type 3 fimbriae (VF0567) - Biofilm (V
VFG038403(gb|WP_011705726) (atsS) Type VI secretion system protein [T6SS (VF0480) - Effector delivery sy
VFG000878(gb|WP_000872015) (fimG) FimG protein precursor [Type 1 fimbriae (VF0221) - Adherence (VFC0
VFG038144(gb|WP_001062908) (galE) UDP-glucose 4-epimerase GalE [Capsule (VF0465) - Immune modulati
VFG040422(gb|WP_011190749) (rvhD4) type IV secretion system component VirD4 [Rvh T4SS (VF0488) - Ef
VFG002070(gb|NP_248774) (hsiC1/vipB/tssC) type VI secretion system tubule-forming protein VipB [HSI-I (V
VFG000120(gb|NP_253949) (algZ) sigma factor AlgU [Alginate (VF0091) - Biofilm (VFC0271)] [Pseudomona
VFG038671(gb|WP_011705282) (fliQ) flagellar biosynthetic protein FliQ [Polar flagella (VF0473) - Motility (V
VFG041302(gb|WP_010947674) (legL5) Dot/Icm type IV secretion system effector LegL5 [Dot/Icm T4SS secre
VFG043624(gb|WP_004149659) (mrkD) fimbrial adhesin protein precursor MrkD [Type 3 fimbriae (VF0567) -
VFG002033(gb|WP_010874498) (p1/MgPa/gapA) ADP1_MYCPN adhesin P1 [Cytadherence organelle (VF033
VFG001950(gb|YP_002344805) (Cj1422c) sugar transferase [Capsule (VF0323) - Immune modulation (VFC025
VFG044079(gb|NP_250944) (pvcA) paerucumarin biosynthesis protein PvcA [Pyoverdine (VF0094) - Nutritiona
VFG015000(gb|NP_252339) (mucP) metalloprotease protease [Alginate (VF0091) - Biofilm (VFC0271)] [Pseud
VFG049150(gb|WP_000982270) (clbD) colibactin biosynthesis dehydrogenase ClbD [Colibactin (VF0573) - Ex
VFG022130(gb|NP_214804) (eccD3) Type IIV secretion system transmembrane protein EccD3 [ESX-3 (VF052
VFG038840(gb|WP_005300916) (flmH) short chain dehydrogenase/reductase family oxidoreductase [Polar flage
VFG044323(gb|WP_015874960) (ybtS) yersiniabactin biosynthesis salicylate synthase YbtS [Ybt (VF0564) - Nu
VFG001266(gb|NP_252912) (pchI) ABC transporter ATP-binding protein [Pyochelin (VF0095) - Nutritional/Me
VFG051131(gb|WP_006191504.1) (bmaC) autotransporter outer membrane beta-barrel domain-containing protei
VFG038675(gb|WP_011705283) (fliR) flagellar biosynthesis protein FliR [Polar flagella (VF0473) - Motility (V
VFG038352(gb|WP_005320745) (ascR) Type III secretion system inner membrane export apparatus protein Asc
VFG002219(gb|WP_011005413) (cgs) cyclic beta 1-2 glucan synthetase [CG (VF0366) - Immune modulation (V
VFG002355(gb|WP_011816613) (flhE) flagellar biosynthesis protein FlhE [Flagella (VF0394) - Motility (VFC0
VFG038691(gb|WP_010634619) (flhG) CobQ/CobB/MinD/ParA family protein [Polar flagella (VF0473) - Moti

VFG034612(gb|WP_000225871) (cfaA) colonisation factor antigen a, chaperone [Adhesive fimbriae (VF0213) -
VFG049912 (tssC) Type VI secretion system protein TssC [T6SS (VF0579) - Effector delivery system (VFC008
VFG000917(gb|WP_000089583) (chuA) Outer membrane heme/hemoglobin receptor ChuA [Chu (VF0227) - Nu
VFG038328(gb|WP_005320795) (ascD) Type III secretion system OM ring protein AscD [T3SS (VF0479) - Eff
VFG000343(gb|WP_164927854) (hitB) iron ABC transporter permease [HitABC (VF0268) - Nutritional/Metabo
VFG049084(gb|WP_004175264) (rfbB) O-antigen export ABC transporter ATP-binding protein RfbB [LPS (VF
VFG038301(gb|WP_016349342) (exeK) general secretion pathway protein K [Exe T2SS (VF0478) - Effector de
VFG038679(gb|WP_016350055) (flhB) flagellar biosynthetic protein FlhB [Polar flagella (VF0473) - Motility (
VFG038587(gb|WP_016349160) (mshG) MSHA (mannose-sensitive hemagglutinin) pili biogenesis protein Msh
VFG000932(gb|WP_000026784) (entE) 2,3-dihydroxybenzoate-AMP ligase component of enterobactin synthase
VFG000944(gb|AAK16477) (draC) usher protein DraC [Dr adhesins (VF0223) - Adherence (VFC0001)] [Esche
VFG001825(gb|NP_217648) (devS) two component sensor histidine kinase DevS [DevRS (VF0317) - Regulatio
VFG049018(gb|WP_002913007) (rcsB) transcriptional regulator RcsB [RcsAB (VF0571) - Regulation (VFC030
VFG038756(gb|WP_011706626) (flrC) transcriptional activator [Polar flagella (VF0473) - Motility (VFC0204)]
VFG001214(gb|NP_253237) (pilR) two-component response regulator PilR [Type IV pili (VF0082) - Adherence
VFG002052(gb|WP_000820136) (gspI) general secretion pathway protein I [T2SS (VF0333) - Effector delivery s
VFG038509(gb|WP_016350158) (flpL) Flp pilus pseudopilin [Flp type IV pili (VF0476) - Adherence (VFC0001
VFG049162(gb|WP_002430641) (clbP) precolibactin peptidase ClbP [Colibactin (VF0573) - Exotoxin (VFC023
VFG038561(gb|WP_016349168) (mshQ) MSHA (mannose-sensitive hemagglutinin) pili biogenesis protein Msh
VFG048600(gb|WP_004213920) (iucA) aerobactin Synthetase IucA [Aerobactin (VF0565) - Nutritional/Metabo
VFG038429(gb|WP_016352167) (tapA) type IV pilus pilin subunit, pilA-like [Tap type IV pili (VF0475) - Adhe
VFG037807(gb|WP_000013434) (lpxB) lipid-A-disaccharide synthase [LPS (VF0466) - Immune modulation (V
VFG049144(gb|WP_002892069) (acrB) acriflavine resistance protein B [AcrAB (VF0568) - Antimicrobial activi
VFG038402(gb|WP_011705725) (vasK/icmF) Type VI secretion system protein [T6SS (VF0480) - Effector deli
VFG049916(gb|WP_000103365) (vgrG/tssI) type VI secretion system effector VgrG [T6SS (VF0579) - Effector
VFG002492(gb|WP_004184995) (fliN) flagellar motor switch protein FliN [Flagella (VF0430) - Motility (VFC0
VFG038882(gb|WP_005314247) (maf-5) modification accessory factor [Lateral flagella (VF0474) - Motility (V
VFG003977(gb|NP_461229) (sseL) type III secretion system effector SseL, deubiquitinase [TTSS-2 secreted effe
VFG002409(gb|WP_000549274) (essC) type VII secretion system protein EssC, FtsK/SpoIIIE family ATPase [T
VFG038878(gb|WP_005314254) (flgI) flagellar basal body P-ring protein [Lateral flagella (VF0474) - Motility (
VFG045793(gb|WP_000771429) (csgA) curlin major subunit CsgA [Curli fibers (VF1138) - Adherence (VFC00
VFG001225(gb|NP_249099) (pilG) twitching motility protein PilG [Type IV pili (VF0082) - Adherence (VFC00
VFG022146(gb|NP_214805) (mycP3) Type VII secretion system membrane-anchored mycosin MycP3 [ESX-3 (
VFG038553(gb|WP_016350147) (flpA) prepilin peptidase [Flp type IV pili (VF0476) - Adherence (VFC0001)] [
VFG048771(gb|WP_004190505) (tssF) type VI secretion system baseplate subunit TssF [T6SS (VF0569) - Effec
VFG044024(gb|ACA60977) (ACA60977) alpha-toxin [Alpha-toxin (septicum) (VF0416) - Exotoxin (VFC0235)
VFG016776(gb|WP_005767781) (fliI) flagellar specific ATP synthase [Flagella (VF0373) - Motility (VFC0204)
VFG002074(gb|NP_248778) (hsiG1/tssF) type VI secretion system hcp secretion island protein HsiG1 [HSI-I (V
VFG038731(gb|WP_016350347) (flaA) flagellin [Polar flagella (VF0473) - Motility (VFC0204)] [Aeromonas hy
VFG038413(gb|WP_043118350) (fimC) outer membrane usher protein [Type I pili (VF0486) - Adherence (VFC
VFG038468(gb|WP_016351972) (tapU) twitching ATPase [Tap type IV pili (VF0475) - Adherence (VFC0001)]
VFG038385(gb|WP_011705716) (atsI) Type VI secretion system protein [T6SS (VF0480) - Effector delivery sys
VFG043504(gb|WP_000671444) (pavB/pfbB) cell wall surface anchor family protein, Plasminogen- and Fibrone
VFG000359(gb|WP_001088826) (ybtE) yersiniabactin siderophore biosynthetic protein [Yersiniabactin (VF0136
VFG046465(gb|WP_003028672) (tufA) elongation factor Tu [EF-Tu (VF0460) - Adherence (VFC0001)] [Franci
VFG049157(gb|WP_014907259) (clbK) colibactin hybrid non-ribosomal peptide synthetase/type I polyketide sy

VFG015551(gb|NP_248741) (phzH) phenazine-modifying enzyme [Pyocyanin (VF0100) - Nutritional/Metabolic
VFG048697(gb|WP_015874721) (clpV/tssH) type VI secretion system ATPase TssH [T6SS (VF0569) - Effector
VFG003561(gb|WP_005173823) (yst1E) type II secretion system ATPase GspE [Yst1 T2SS (VF0635) - Effector
VFG000884(gb|WP_000654937) (papC) usher protein PapC [P fimbriae (VF0220) - Adherence (VFC0001)] [Es
VFG047529(gb|WP_003029409) (bioA) adenosylmethionine-8-amino-7-oxononanoate aminotransferase [Biotin
VFG000876(gb|WP_000120946) (fimD) Outer membrane usher protein fimD precursor [Type 1 fimbriae (VF02
VFG049911(gb|WP_000393837) (tssF) Type VI secretion system protein TssF [T6SS (VF0579) - Effector delive
VFG044332(gb|WP_015874967) (ybtE) yersiniabactin biosynthesis salycil-AMP ligase YbtE [Ybt (VF0564) - N
VFG038409(gb|WP_011704492) (fimA) fimbrial protein [Type I pili (VF0486) - Adherence (VFC0001)] [Aeron
VFG002517(gb|WP_004203463) (flgK) flagellar hook-associated protein 1 FlgK [Flagella (VF0430) - Motility (
VFG002120(gb|WP_010948516) (sidH) Dot/Icm type IV secretion system effector SidH [Dot/Icm T4SS secreted
VFG002049(gb|WP_001173470) (gspF) general secretion pathway protein F [T2SS (VF0333) - Effector delivery
VFG001381(gb|YP_177728) (icl) Isocitrate lyase Icl (isocitrase) (isocitratase) [Isocitrate lyase (VF0253) - Others
VFG001252(gb|NP_249793) (fliG) flagellar motor switch protein G [Flagella (VF0273) - Motility (VFC0204)] [
VFG002047(gb|WP_000498818) (gspD) general secretion pathway protein D [T2SS (VF0333) - Effector delivery
VFG049161(gb|WP_001029878) (clbO) colibactin polyketide synthase ClbO [Colibactin (VF0573) - Exotoxin (V
VFG000159(gb|NP_251076) (pvdA) L-ornithine N5-oxygenase PvdA [Pyoverdine (VF0094) - Nutritional/Metab
VFG001374(gb|WP_000077424) (cps4J) capsular polysaccharide biosynthesis protein Cps4J [Capsule (VF0144)
VFG037822(gb|WP_000240700) (lpxC) UDP-3-O-acyl-N-acetylglucosamine deacetylase [LPS (VF0466) - Imm
VFG002166(gb|WP_010714416) (ace) collagen adhesin protein [Ace (VF0355) - Adherence (VFC0001)] [Enter
VFG050115(gb|WP_002081542) (cesA) Cereulide synthetase A, CesA [Cereulide (VF0583) - Exotoxin (VFC02
VFG012615(gb|WP_000020049) (chuX) putative heme-binding protein ChuX [Chu (VF0234) - Nutritional/Meta
VFG038695(gb|WP_017409955) (fliA) flagellar biosynthesis sigma factor [Polar flagella (VF0473) - Motility (V
VFG048819(gb|WP_014907234) (KP1_RS17280) mannose-1-phosphate guanylyltransferase/mannose-6-phosph
VFG043642(gb|WP_011205946) (MHP_RS00555) P110/LppT family adhesin N-terminal domain [P97/P102 par
VFG049122(gb|WP_012737539) (allC) allantoate amidohydrolase [Allantion utilization (VF0572) - Nutritional/
VFG038667(gb|WP_016350054) (fliP) flagellar biosynthesis protein FliP [Polar flagella (VF0473) - Motility (V
VFG000859(gb|CBG37421) (set1A) enterotoxin subunit Set1A [ShET1 (VF0219) - Exotoxin (VFC0235)] [Esch
VFG049905(gb|WP_000343289) (tssL) Type VI secretion system protein TssL [T6SS (VF0579) - Effector delive
VFG038647(gb|WP_016350053) (fliK) flagellar hook-length control protein FliK [Polar flagella (VF0473) - Mot
VFG038374(gb|WP_011705709) (atsB) Type VI secretion system protein [T6SS (VF0480) - Effector delivery sy
VFG000326(gb|WP_105163169) (licA) choline kinase [LOS (VF0044) - Immune modulation (VFC0258)] [Hae
VFG048508(gb|WP_012737618) (fes) enterobactin/ferric enterobactin esterase [Ent (VF0562) - Nutritional/Meta
VFG044327(gb|WP_000140406) (ybtA) yersiniabactin transcriptional regulator YbtA [Ybt (VF0564) - Nutrition
VFG048620(gb|WP_014907843) (iutA) ferric aerobactin receptor IutA [Aerobactin (VF0565) - Nutritional/Meta
VFG048409(gb|WP_086074289) (entA) 2,3-dihydroxybenzoate-2,3-dehydrogenase [Ent (VF0562) - Nutritional/
VFG048830(gb|WP_014907233) (gndA) NADP-dependent phosphogluconate dehydrogenase [Capsule (VF0560
VFG038119(gb|WP_000686134) (ACICU_RS00485) nucleotide sugar dehydrogenase [Capsule (VF0465) - Imm
VFG043017(gb|NP_252038) (PA3348) chemotaxis protein methyltransferase [Flagella (VF0273) - Motility (VF
VFG048488(gb|WP_002893737) (fepC) iron-enterobactin transporter ATP-binding protein [Ent (VF0562) - Nutr
VFG049007(gb|WP_002911561) (rcsA) transcriptional activator for ctr capsule biosynthesis [RcsAB (VF0571) -
VFG012591(gb|WP_001017167) (chuS) heme oxygenase ChuS [Chu (VF0234) - Nutritional/Metabolic factor (V
VFG012505(gb|WP_000933673) (iroD) esterase [Salmochelin siderophore (VF0230) - Nutritional/Metabolic fac
VFG049826(gb|WP_000142058) (esaG) TIGR01741 family protein [Type VII secretion system (VF0403) - Effe
VFG038615(gb|WP_016349152) (mshI) MSHA (mannose-sensitive hemagglutinin) pili biogenesis protein MshI
VFG000902(gb|WP_001034089) (sat) Aecreted auto transpoter toxin [Sat (VF0231) - Effector delivery system (

VFG001443(gb|AAF37887) (ompA) outer membrane protein A [OmpA (VF0236) - Invasion (VFC0083)] [Esche
VFG049903(gb|WP_001240533) (tssA) Type VI secretion system protein TssA [T6SS (VF0579) - Effector deliv
VFG002066(gb|NP_248770) (lip1) lipoprotein [HSI-I (VF0334) - Effector delivery system (VFC0086)] [Pseudor
VFG000927(gb|WP_000096764) (fepE) LPS O-antigen length regulator [Enterobactin (VF0228) - Nutritional/Me
VFG041043(gb|NP_251061) (PA2371) ClpA/B-type protease [HSI-3 (VF0944) - Effector delivery system (VFC
VFG038631(gb|WP_011705273) (fliG) flagellar motor switch protein G [Polar flagella (VF0473) - Motility (VF
VFG038889(gb|WP_005314215) (lafT) LafT, flagella motor stator protein [Lateral flagella (VF0474) - Motility (
VFG002328(gb|WP_011816597) (fliF) flagellar M-ring protein FliF [Flagella (VF0394) - Motility (VFC0204)] [
VFG048518(gb|WP_012737617) (fepA) outer membrane receptor FepA [Ent (VF0562) - Nutritional/Metabolic f
VFG050165(gb|WP_001259305) (neuA) N-acylneuraminate cytidylyltransferase [K1 capsule (VF0239) - Invasio
VFG038393(gb|WP_010634228) (dotU) Type VI secretion system protein DotU [T6SS (VF0480) - Effector deli
VFG038349(gb|WP_005320751) (ascO) Type III secretion system protein AscO [T3SS (VF0479) - Effector deli
VFG000140(gb|NP_253696) (waaP) UDP-glucose:(heptosyl) LPS alpha 1,3-glucosyltransferase WaaG [LPS (V
VFG037246(gb|WP_001032906) (basH) non-ribosomal peptide biosynthesis thioesterase BasH [Acinetobactin (
VFG045474(gb|AAM75251) (cylM) CylM; cytolysin subunit modifier [Cytolysin (VF0356) - Exotoxin (VFC023
VFG050400(gb|WP_000347039.1) (pilU) PilT/PilU family type 4a pilus ATPase [TFP (VF1334) - Adherence (V
VFG050685(gb|WP_001160337.1) (pilS) PAS domain-containing sensor histidine kinase [TFP (VF1334) - Adhe
VFG001223(gb|NP_249086) (pilT) twitching motility protein PilT [Type IV pili (VF0082) - Adherence (VFC000
VFG002497(gb|WP_011204776) (fliK) flagellar hook-length control protein FliK [Flagella (VF0430) - Motility (
VFG051244(gb|WP_004683766.1) (BPE043) type IV secretion system effector [T4SS secreted effectors (VF069
VFG038683(gb|WP_016350056) (flhA) flagellar biosynthesis protein FlhA [Polar flagella (VF0473) - Motility (
VFG049095(gb|WP_002912373) (rfbA) O-antigen export ABC transporter permease RfbA [LPS (VF0561) - Imm
VFG016472(gb|WP_011206327) (p216) protein P216, cilium adhesin p97 paralog [P97/P102 paralog family (VF
VFG012160(gb|WP_011590331) (nanI) exo-alpha-sialidase [Sialidase (VF0391) - Exoenzyme (VFC0251)] [Clos
VFG048643(gb|WP_014907727) (vipB/tssC) type VI secretion system contractile sheath large subunit VipB [T6
VFG002277(gb|WP_011009663) (nagH) hyaluronidase [Mu-toxin (VF0389) - Exoenzyme (VFC0251)] [Clostrid
VFG002274(gb|WP_011009584) (plc) phospholipase C [Alpha-toxin (CpPLC) (VF0378) - Exotoxin (VFC0235)
VFG043627(gb|WP_000756959) (mrkA) type 3 fimbrial major pilin subunit MrkA [Type 3 fimbriae (VF0567) -
VFG001270(gb|WP_010929997) (cyaD) cyclolysin secretion protein [Cya (VF0028) - Exotoxin (VFC0235)] [Bo
VFG044324(gb|WP_032447693) (ybtX) yersiniabactin-associated zinc MFS transporter YbtX [Ybt (VF0564) - N
VFG002417(gb|WP_001301550) (yagV/ecpE) E. coli common pilus chaperone EcpE [ECP (VF0404) - Adherenc
VFG050157(gb|WP_001331698) (kpsF) Polysialic acid capsule expression protein [K1 capsule (VF0239) - Invas
VFG048287(gb|WP_014906962) (fimF) type 1 fimbrial minor component [Type I fimbriae (VF0566) - Adherenc
VFG002060(gb|NP_248764) (ppkA) serine/threonine protein kinase PpkA [HSI-I (VF0334) - Effector delivery s
VFG002452(gb|WP_004525318) (bsaZ) YscU homolog, component of the inner membrane ring [Bsa T3SS (VF
VFG050670(gb|WP_000505931.1) (pilJ) methyl-accepting chemotaxis protein [TFP (VF1334) - Adherence (VF
VFG038772(gb|WP_016351319) (flgK) flagellar hook-associated protein FlgK [Polar flagella (VF0473) - Motilit
VFG000820(gb|WP_000087467) (cesD) chaperone for EspB and EspD [TTSS (VF0191) - Effector delivery syste
VFG038297(gb|WP_016349341) (exeJ) general secretion pathway protein J [Exe T2SS (VF0478) - Effector deliv
VFG038753(gb|WP_024944879) (motY) sodium-type flagellar protein MotY [Polar flagella (VF0473) - Motility
VFG038913 (rtxA) multifunctional autoprocessing repeats-in-toxins (MARTX) toxin [RtxA (VF0482) - Exotoxin
VFG034901(gb|WP_000868321) (espY2) Type III secretion system effector EspY2 [TTSS secreted effectors (V
VFG000871(gb|WP_000790574) (fimB) Type 1 fimbriae Regulatory protein fimB [Type 1 fimbriae (VF0221) -
VFG000182(gb|NP_251793) (xcpR) general secretion pathway protein E [Xcp secretion system (VF0084) - Effe
VFG049040(gb|WP_074160401) (KP1_RS17225) glycosyltransferase family 4 protein [LPS (VF0561) - Immune
VFG002415(gb|WP_001131063) (yagX/ecpC) E. coli common pilus usher EcpC [ECP (VF0404) - Adherence (V

VFG049163(gb|WP_000065646) (clbQ) colibactin biosynthesis thioesterase ClbQ  [Colibactin (VF0573) - Exoto
VFG002414(gb|WP_000730972) (yagZ/ecpA) E. coli common pilus structural subunit EcpA [ECP (VF0404) - A
VFG000687(gb|AAP42180) (bexD') BexD' [Capsule (VF0043) - Immune modulation (VFC0258)] [Haemophilus
VFG038655(gb|WP_011705279) (fliM) flagellar motor switch protein FliM [Polar flagella (VF0473) - Motility (
VFG002280(gb|WP_011010361) (nagK) hyaluronidase [Mu-toxin (VF0389) - Exoenzyme (VFC0251)] [Clostrid
VFG000669(gb|NP_706257) (gtrA) bactoprenol-linked glucose translocase/flippase [LPS (VF0124) - Immune m
VFG016058(gb|NP_251114) (pvdL) peptide synthase PvdL [Pyoverdine (VF0094) - Nutritional/Metabolic factor
VFG041042(gb|NP_251060) (PA2370) hypothetical protein [HSI-3 (VF0944) - Effector delivery system (VFC0
VFG001930(gb|YP_002345095) (flgE2) flagellar hook protein [Flagella (VF0114) - Motility (VFC0204)] [Camp
VFG002412(gb|WP_000716386) (yagY/ecpB) E. coli common pilus chaperone EcpB [ECP (VF0404) - Adheren
VFG045818(gb|WP_001528154) (tsh) temperature-sensitive protein Tsh [Tsh (VF0233) - Effector delivery syste
VFG034576(gb|WP_049794100) (aafC) aggregative adherence fimbria II usher protein AafC [AAFs (VF0214) -
VFG000423(gb|WP_000052242) (vexE) Vi polysaccharide export protein VexE [Vi antigen (VF0101) - Immune
VFG000088(gb|WP_000591844) (tcpI) negative regulator of the major pilin TcpA [TCP (VF0126) - Adherence (
VFG039410(gb|NP_819986) (cem3) Coxiella Dot/Icm type IVB secretion system translocated effector  [T4SS se
VFG045791(gb|WP_000481500) (cgsD) transcriptional regulator CsgD [Curli fibers (VF1138) - Adherence (VF
VFG002349(gb|WP_005168797) (flgD) flagellar basal-body rod modification protein FlgD [Flagella (VF0394) -
VFG002538(gb|WP_004184923) (pmlR/bspR1) N-acylhomoserine lactone dependent regulatory protein [Quorur
VFG051113(gb|WP_080515433.1) (bigA) adhesin with a bacterial immunoglobulin-like domain [BigA (VF1344
VFG041020(gb|NP_250353) (PA1662) ClpA/B-type protease [HSI-2 (VF0943) - Effector delivery system (VFC
VFG034772(gb|WP_000627910) (espL4) Type III secretion system effector EspL4 [TTSS secreted effectors (VF
VFG038874(gb|WP_005314263) (flgE) flagellar hook protein FlgE [Lateral flagella (VF0474) - Motility (VFC0
VFG050158(gb|WP_000905916) (kpsE) Capsule polysaccharide export inner-membrane protein [K1 capsule (V
VFG050386(gb|WP_000355489.1) (pilT) type IV pilus twitching motility protein PilT [TFP (VF1334) - Adheren
VFG041102(gb|NP_459276) (STM0278) putative periplasmic protein [SCI (Salmonella centrisome island) (VF0
VFG049156(gb|WP_077265119) (clbJ) colibactin non-ribosomal peptide synthetase ClbJ [Colibactin (VF0573) -
VFG037762(gb|WP_000078878) (lpxL) lauroyl acyltransferase [LPS (VF0466) - Immune modulation (VFC025
VFG000427(gb|WP_000720235) (vexA) Vi polysaccharide export protein VexA [Vi antigen (VF0101) - Immune
VFG002416(gb|WP_001265657) (yagW/ecpD) polymerized tip adhesin of ECP fibers [ECP (VF0404) - Adheren
VFG034817(gb|WP_000671691) (espR1) Type III secretion system effector espR1 [TTSS secreted effectors (VF
VFG038812(gb|WP_011706645) (cheR-3) chemotaxis protein CheR [Polar flagella (VF0473) - Motility (VFC02
VFG049899(gb|WP_001087735) (tssA) Type VI secretion system protein TssA [T6SS (VF0579) - Effector deliv
VFG000064(gb|WP_010930831) (bcrD) Type III secretion system LcrD homolog protein BcrD [TTSS (VF0035)
VFG041027(gb|NP_250360) (PA1669) hypothetical protein [HSI-2 (VF0943) - Effector delivery system (VFC0
VFG040868(gb|WP_000404139) (esxL) type VII secretion system protein EsxL [T7SS (VF0581) - Effector deliv
VFG049898(gb|WP_001284199) (hcp1/tssD1) type VI secretion system protein, Hcp family [T6SS (VF0579) - E
VFG038273(gb|WP_016349335) (exeD) general secretion pathway protein D [Exe T2SS (VF0478) - Effector del
VFG011626(gb|WP_002965156) (bvrR) response regulator transcription factor [BvrR-BvrS (VF0368) - Regulati
VFG031564(gb|YP_177826) (PE_PGRS30) PE-PGRS family protein PE_PGRS30 [PE/PE-PGRS (VF0316) - Im
VFG038329(gb|WP_011899438) (ascC) Type III secretion system OM ring protein AscC [T3SS (VF0479) - Effe
VFG034928(gb|WP_001239082) (lifA/efa1) lymphostatin Efa1/LifA [Lymphostatin/LifA (VF0175) - Effector de
VFG038383(gb|WP_011705715) (atsH) Type VI secretion system protein [T6SS (VF0480) - Effector delivery sy
VFG000924(gb|WP_001234311) (fepB) ferrienterobactin ABC transporter periplasmic binding protein [Enteroba
VFG048787(gb|WP_004176417) (sciN/tssJ) type VI secretion system lipoprotein TssJ [T6SS (VF0569) - Effecto
VFG000943(gb|AAK16476) (draB) chaperone protein DraB [Dr adhesins (VF0223) - Adherence (VFC0001)] [E
VFG038345(gb|WP_005320764) (acr2) chaperone protein [T3SS (VF0479) - Effector delivery system (VFC008

VFG008007(gb|NP_218342) (pks2) phthioceranic/hydroxyphthioceranic acid synthase [Sulfolipid-1 biosynthesis
VFG038371(gb|WP_011705707) (vgrG2) VgrG protein [T6SS (VF0480) - Effector delivery system (VFC0086)]
VFG000115(gb|NP_249839) (toxA) exotoxin A precursor [ExoA (VF0086) - Exotoxin (VFC0235)] [Pseudomon
VFG038875(gb|WP_005314261) (lfgF) lateral flagellar basal-body rod protein LfgF [Lateral flagella (VF0474) -
VFG041011(gb|NP_248785) (vgrG1b) type VI secretion system substrate VgrG1b [HSI-I (VF0334) - Effector de
VFG038537(gb|WP_016350151) (flpE) Flp pilus assembly protein, ATPase [Flp type IV pili (VF0476) - Adherer
VFG049153(gb|WP_000159201) (clbG) colibactin biosynthesis acyltransferase ClbG [Colibactin (VF0573) - Exo
VFG001264(gb|NP_250144) (flhF) flagellar biosynthesis protein FlhF [Flagella (VF0273) - Motility (VFC0204)]
VFG001364(gb|WP_001193686) (hysA) hyaluronidase [Hyaluronidase (VF0146) - Exoenzyme (VFC0251)] [Str
VFG001228(gb|NP_249102) (pilJ) twitching motility protein PilJ [Type IV pili (VF0082) - Adherence (VFC000
VFG050169(gb|WP_000124301) (kpsM) Polysialic acid transport protein [K1 capsule (VF0239) - Invasion (VFC
VFG049149(gb|WP_041169023) (clbC) colibactin polyketide synthase ClbC [Colibactin (VF0573) - Exotoxin (V
VFG043206(gb|WP_005164479) (cheY) chemotaxis regulatory protein CheY [Flagella (VF0394) - Motility (VF
VFG050262(gb|NP_220136) (CT_619) hypothetical protein [TTSS secreted effectors (VF0711) - Effector delive
VFG038367(gb|WP_011705045) (vgrG1) Type VI secretion system effector VgrG1, ADP-ribosyltransferase [T
VFG001256(gb|NP_250134) (fliM) flagellar motor switch protein FliM [Flagella (VF0273) - Motility (VFC0204
VFG041304(gb|WP_010947678) (lirB) Dot/Icm type IV secretion system effector LirB [Dot/Icm T4SS secreted
VFG001405(gb|NP_217219) (sigA/rpoV) RNA polymerase sigma factor SigA [SigA (VF0257) - Regulation (VF
VFG044326(gb|WP_001327262) (ybtP) yersiniabactin ABC transporter ATP-binding/permease protein YbtP [Y
VFG049062(gb|WP_015875020) (rfbD) UDP-galactopyranose mutase [LPS (VF0561) - Immune modulation (V
VFG044328(gb|WP_015874963) (irp2) yersiniabactin non-ribosomal peptide synthetase HMWP2 [Ybt (VF0564)
VFG038679(gb|WP_016350055) (flhB) flagellar biosynthetic protein FlhB [Polar flagella (VF0473) - Motility (V
VFG001248(gb|NP_249788) (fleQ) transcriptional regulator FleQ [Flagella (VF0273) - Motility (VFC0204)] [Ps
VFG001263(gb|NP_250143) (flhA) flagellar biosynthesis protein FlhA [Flagella (VF0273) - Motility (VFC0204)
VFG049903(gb|WP_001240533) (tssA) Type VI secretion system protein TssA [T6SS (VF0579) - Effector deliv
VFG037497(gb|WP_085947929) (ACICU_RS04580) TonB-dependent receptor [HemO cluster (VF0636) - Nutri
VFG035025(gb|WP_001443810) (nleG6-1) DUF1076 domain-containing protein [TTSS secreted effectors (VF1
VFG002417(gb|WP_001301550) (yagV/ecpE) E. coli common pilus chaperone EcpE [ECP (VF0404) - Adherenc
VFG010515(gb|WP_010946906) (lidL) Dot/Icm type IV secretion system effector LidL [Dot/Icm T4SS secreted
VFG050297(gb|NP_250644) (fapC) major amyloid subunit FapC [Fap (VF1191) - Adherence (VFC0001)] [Pseu
VFG038391(gb|WP_011705719) (atsL) Type VI secretion system protein [T6SS (VF0480) - Effector delivery sy
VFG000944(gb|AAK16477) (draC) usher protein DraC [Dr adhesins (VF0223) - Adherence (VFC0001)] [Esche
VFG050787(gb|WP_001112089.1) (gspF) general secretion pathway protein F [T2SS (VF1335) - Effector delive
VFG048797(gb|WP_004175261) (ugd) UDP-glucose 6-dehydrogenase [Capsule (VF0560) - Immune modulation
VFG007574(gb|WP_001030417) (fliA) flagellar biosynthesis sigma factor FliA [Flagella (VF0519) - Motility (V
VFG038432(gb|WP_016352168) (tapB) type IV pilus assembly ATPase, PilB-like [Tap type IV pili (VF0475) -
VFG050158(gb|WP_000905916) (kpsE) Capsule polysaccharide export inner-membrane protein [K1 capsule (V
VFG000931(gb|WP_001336900) (entC) isochorismate synthase 1 [Enterobactin (VF0228) - Nutritional/Metaboli
VFG013070(gb|WP_001626196) (shuV) ATP-binding component of hemin transport system [Shu (VF0256) - N
VFG000887(gb|WP_000597713) (papK) P pilus minor subunit PapK [P fimbriae (VF0220) - Adherence (VFC00
VFG002269(gb|WP_011181342) (trwD) Trw type IV secretion sysmtem VirB11 homolog, trwD protein [Trw ty
VFG038596(gb|WP_016349157) (mshM) membrane protein with ATPase domain, GspA-like [MSHA type IV p
VFG000919(gb|WP_001095619) (chuW) Putative oxygen independent coproporphyrinogen III oxidase [Chu (V
VFG002063(gb|NP_248767) (icmF1/tssM1) type VI secretion system protein IcmF1 [HSI-I (VF0334) - Effector
VFG049835(gb|WP_012076956) (exlA) hemolysin [Exolysin (VF0582) - Effector delivery system (VFC0086)] [
VFG037720(gb|WP_001027056) (adeG) cation/multidrug efflux pump [AdeFGH efflux pump (VF0504) - Biofil

VFG000363(gb|WP_002212775) (irp2) yersiniabactin biosynthetic protein Irp2 [Yersiniabactin (VF0136) - Nutri
VFG034901(gb|WP_000868321) (espY2) Type III secretion system effector EspY2 [TTSS secreted effectors (VF
VFG038707(gb|WP_016350059) (cheA-2) chemotaxis protein CheA [Polar flagella (VF0473) - Motility (VFC02
VFG007027(gb|WP_001092347) (rtxD) RTX toxin transporter RtxD [MARTX (VF0265) - Exotoxin (VFC0235)
VFG002280(gb|WP_011010361) (nagK) hyaluronidase [Mu-toxin (VF0389) - Exoenzyme (VFC0251)] [Clostrid
VFG034754(gb|WP_000081129) (espL1) Type III secretion system effector espL1 [TTSS secreted effectors (VF
VFG034648(gb|WP_001245988) (cfaD/cfaE) minor pilin and initiator [Adhesive fimbriae (VF0213) - Adherence
VFG038711(gb|WP_016350060) (cheB-2) chemotaxis-specific methylesterase [Polar flagella (VF0473) - Motility
VFG041014(gb|NP_250347) (PA1656) hypothetical protein [HSI-2 (VF0943) - Effector delivery system (VFC00
VFG044328(gb|WP_015874963) (irp2) yersiniabactin non-ribosomal peptide synthetase HMWP2 [Ybt (VF0564)
VFG038881(gb|WP_005314249) (lfgL) lateral flagellar hook-associated protein, LfgL [Lateral flagella (VF0474)
VFG002189(gb|WP_002359680) (cpsB/cdsA) phosphatidate cytidylyltransferase [Capsule (VF0361) - Immune m
VFG048606(gb|WP_004213922) (iucC) aerobactin siderophore biosynthesis protein IucC [Aerobactin (VF0565)
VFG038468(gb|WP_016351972) (tapU) twitching ATPase [Tap type IV pili (VF0475) - Adherence (VFC0001)]
VFG038373(gb|WP_011705708) (atsA) Type VI secretion system protein [T6SS (VF0480) - Effector delivery sy
VFG016069(gb|NP_251115) (pvdG) pyoverdine biosynthesis protein PvdG [Pyoverdine (VF0094) - Nutritional/M
VFG012636(gb|NP_858256) (virB) transcriptional activator VirB [TTSS (VF0118) - Effector delivery system (V
VFG013066 (shuW) heme anaerobic degradation radical SAM methyltransferase ChuW/HutW [Shu (VF0256) - N
VFG038691(gb|WP_010634619) (flhG) CobQ/CobB/MinD/ParA family protein [Polar flagella (VF0473) - Motil
VFG000337(gb|WP_005693332) (iga1) immunoglobin A1 protease [IgA1 protease (VF0045) - Exoenzyme (VFC
VFG038824(gb|WP_011706648) (flgM) anti-sigma-D factor [Polar flagella (VF0473) - Motility (VFC0204)] [Ae
VFG002529(gb|WP_011205290) (tsr) methyl-accepting chemotaxis protein I [Flagella (VF0430) - Motility (VFC
VFG042734(gb|NP_249343) (vfr) cAMP-regulatory protein [Type IV pili (VF0082) - Adherence (VFC0001)] [P
VFG001927(gb|YP_002344848) (flgK) flagellar hook-associated protein 1 FlgK [Flagella (VF0114) - Motility (V
VFG002195(gb|WP_002399773) (EF0818) polysaccharide lyase, family 8 [Hyaluronidase (VF0359) - Exoenzym
VFG048429(gb|WP_004183353) (entE) enterobactin synthase subunit E [Ent (VF0562) - Nutritional/Metabolic f
VFG012597(gb|WP_000089593) (chuA) Outer membrane heme/hemoglobin receptor ChuA [Chu (VF0234) - Nu
VFG040815(gb|WP_041471735) (rickA) WASP, N-WASP, MENA proteins [RickA (VF0490) - Motility (VFC0
VFG002462(gb|WP_004555023) (bsaP) Type III secretion system gate keeper protein [Bsa T3SS (VF0428) - Eff
VFG048327(gb|WP_004152886) (mrkH) transcriptional activator [Type 3 fimbriae (VF0567) - Biofilm (VFC027
VFG000116(gb|NP_254170) (algB) two-component response regulator AlgB [Alginate (VF0091) - Biofilm (VFC
VFG037442(gb|WP_000910253) (basA) acinetobactin biosynthesis protein [Acinetobactin (VF0467) - Nutritiona
VFG002047(gb|WP_000498818) (gspD) general secretion pathway protein D [T2SS (VF0333) - Effector delivery
VFG000902(gb|WP_001034089) (sat) Aecreted auto transpoter toxin [Sat (VF0231) - Effector delivery system (V
VFG038731(gb|WP_016350347) (flaA) flagellin [Polar flagella (VF0473) - Motility (VFC0204)] [Aeromonas hy
VFG041109(gb|NP_459283) (STM0285) hypothetical protein [SCI (Salmonella centrisome island) (VF0974) - E
VFG000211(gb|NP_250407) (pscC) type III secretion system secretin PscC [TTSS (VF0083) - Effector delivery s
VFG000263(gb|WP_010980796) (tbpB) transferrin-binding protein B [Tbp (VF0046) - Nutritional/Metabolic fac
VFG002070(gb|NP_248774) (hsiC1/vipB/tssC) type VI secretion system tubule-forming protein VipB [HSI-I (VF
VFG048985(gb|WP_014907219) (KP1_RS17355) phosphatase PAP2 family protein [Capsule (VF0560) - Immun
VFG042568(gb|WP_000012016) (sfaY) transcriptional regulator SfaY [F1C fimbriae (VF0224) - Adherence (VF
VFG045788(gb|WP_001189322) (cgsG) curli production assembly/transport protein CsgG [Curli fibers (VF1138
VFG000907(gb|WP_000376545) (hlyB) Hemolysin B [-Hemolysin (VF0225) - Exotoxin (VFC0235)] [Escherich
VFG043016(gb|NP_250155) (PA1464) purine-binding chemotaxis protein [Flagella (VF0273) - Motility (VFC02
VFG050959(gb|WP_000556915.1) (tssM) type VI secretion system membrane subunit TssM [T6SS (VF1337) - E
VFG048830(gb|WP_014907233) (gndA) NADP-dependent phosphogluconate dehydrogenase [Capsule (VF0560

VFG038405(gb|WP_011705727) (AHA_RS09305) type VI secretion system PAAR protein [T6SS (VF0480) - E
VFG038397(gb|WP_011705722) (vasH) sigma-54 dependent transcriptional regulator [T6SS (VF0480) - Effecto
VFG000130(gb|NP_252238) (algI) alginate o-acetyltransferase AlgI [Alginate (VF0091) - Biofilm (VFC0271)] [
VFG038695(gb|WP_017409955) (fliA) flagellar biosynthesis sigma factor [Polar flagella (VF0473) - Motility (V
VFG034772(gb|WP_000627910) (espL4) Type III secretion system effector EspL4 [TTSS secreted effectors (VF
VFG034895(gb|WP_001304001) (espY1) Type III secretion system effector EspY1 [TTSS secreted effectors (VI
VFG001410(gb|NP_217458) (mmpL7) transmembrane transport protein MmpL7 [PDIM (VF0309) - Immune mo
VFG001266(gb|NP_252912) (pchI) ABC transporter ATP-binding protein [Pyochelin (VF0095) - Nutritional/Me
VFG049900 (tssM) Type VI secretion system protein TssM [T6SS (VF0579) - Effector delivery system (VFC00
VFG037897(gb|WP_001165095) (tviB) Vi polysaccharide biosynthesis UDP-N-acetylglucosamine C-6 dehydrog
VFG016046(gb|NP_251103) (pvdH) diaminobutyrate-2-oxoglutarate aminotransferase PvdH [Pyoverdine (VF00
VFG033393(gb|WP_001234181) (sfaH) S fimbriae adhesin SfaH [S fimbriae (VF0235) - Adherence (VFC0001)
VFG048478(gb|WP_004147521) (fepG) iron-enterobactin transporter permease [Ent (VF0562) - Nutritional/Met
VFG049051(gb|WP_014907239) (KP1_RS17230) glycosyltransferase [LPS (VF0561) - Immune modulation (VI
VFG001959(gb|YP_002344814) (hddC) capsular polysaccharide heptosyltransferase [Capsule (VF0323) - Immu
VFG038402(gb|WP_011705725) (vasK/icmF) Type VI secretion system protein [T6SS (VF0480) - Effector deli
VFG001145(gb|CAA83901) (hifB) periplasmic chaperone [Haemagglutinating pili (VF0036) - Adherence (VFC
VFG002067(gb|NP_248771) (fha1) type VI secretion system forkhead-associated protein Fha1 [HSI-I (VF0334)
VFG002481(gb|WP_009940775) (vgrG-5) Type VI secretion system protein VgrG5 [T6SS-1 (VF0429) - Effecto
VFG000193(gb|NP_250389) (popN) type III secretion system outer membrane protein PopN [TTSS (VF0083) -
VFG001228(gb|NP_249102) (pilJ) twitching motility protein PilJ [Type IV pili (VF0082) - Adherence (VFC000
VFG038623(gb|WP_011705271) (fliE) flagellar hook-basal body complex protein FliE [Polar flagella (VF0473)
VFG000340(gb|WP_005694039) (hxuA) heme-hemopexin utilization protein A, two-partner secretion (TPS) pat
VFG006543(gb|WP_001266636) (flgL) flagellar hook-associated protein 3 FlgL [Flagella (VF0051) - Motility (V
VFG001214(gb|NP_253237) (pilR) two-component response regulator PilR [Type IV pili (VF0082) - Adherence
VFG037482(gb|WP_004748062) (ACICU_RS04570) sigma-70 family RNA polymerase sigma factor [HemO clu
VFG002054(gb|WP_000633233) (gspK) general secretion pathway protein K [T2SS (VF0333) - Effector delivery
VFG050457(gb|WP_001274990.1) (pilB) type IV-A pilus assembly ATPase PilB [TFP (VF1334) - Adherence (V
VFG001940(gb|YP_002344532) (wlaN) beta-1,3 galactosyltransferase [LOS (VF0326) - Immune modulation (V
VFG000878(gb|WP_000872015) (fimG) FimG protein precursor [Type 1 fimbriae (VF0221) - Adherence (VFC0
VFG038525(gb|WP_043118801) (flpH) Flp pilus assembly protein FlpH [Flp type IV pili (VF0476) - Adherence
VFG012115 (cdtB) CdtB [CDT (VF0385) - Exotoxin (VFC0235)] [Clostridium difficile 630]
VFG018682(gb|WP_005491021) (vopL) type III secretion system effector VopL, actin filament nucleation activi
VFG002400(gb|YP_177838) (esxN) ESX-5 type VII secretion system EsxA (ESAT-6) homolog [ESX-5 (VF040
VFG038509(gb|WP_016350158) (flpL) Flp pilus pseudopilin [Flp type IV pili (VF0476) - Adherence (VFC0001
VFG042861(gb|WP_011213223) (pilY1) type IV pilus biogenesis PilY1-related protein [Type IV pili (VF0155) -
VFG002290(gb|WP_004456321) (cpb) beta toxin [Beta-toxin (CPB) (VF0379) - Exotoxin (VFC0235)] [Clostridi
VFG000182(gb|NP_251793) (xcpR) general secretion pathway protein E [Xcp secretion system (VF0084) - Effec
VFG002276(gb|WP_011009653) (colA) collagenase [Kappa-toxin (VF0388) - Exoenzyme (VFC0251)] [Clostrid
VFG002433(gb|WP_004187526) (pilO) type IV pilus inner membrane platform protein PilO [Type IV pili (VF04
VFG038816(gb|WP_011706646) (cheV) CheV [Polar flagella (VF0473) - Motility (VFC0204)] [Aeromonas hyd
VFG002248(gb|WP_011181145) (bepG) Bartonella effector protein BepG [VirB/VirD4 T4SS secreted effectors
VFG045792(gb|WP_000791650) (csgB) curlin minor subunit CsgB [Curli fibers (VF1138) - Adherence (VFC00
VFG038913 (rtxA) multifunctional autoprocessing repeats-in-toxins (MARTX) toxin [RtxA (VF0482) - Exotoxi
VFG038380(gb|WP_043163044) (vipB) Type VI secretion system contractile sheath large subunit TssC/VipB [T
VFG000265(gb|WP_002225048) (lbpB) lactoferrin-binding protein B [Lbp (VF0047) - Nutritional/Metabolic fac

VFG001814(gb|NP_215678) (narH) nitrate reductase subunit beta [Nitrate reductase (VF0302) - Nutritional/Meta
VFG034866(gb|WP_000900547) (espX4) Type III secretion system effector EspX4 [TTSS secreted effectors (VF
VFG050400(gb|WP_000347039.1) (pilU) PilT/PilU family type 4a pilus ATPase [TFP (VF1334) - Adherence (V
VFG038389(gb|WP_011705718) (atsK) Type VI secretion system protein [T6SS (VF0480) - Effector delivery sy
VFG043211(gb|WP_032902679) (cheA) chemotaxis protein CheA [Flagella (VF0394) - Motility (VFC0204)] [Y
VFG002380(gb|NP_218385) (eccA1) ESX-1 type VII secretion system AAA+ ATPase EccA1 [ESX-1 (VF0315)
VFG038593(gb|WP_016349158) (mshN) Tetratricopeptide repeat protein, PilF-like [MSHA type IV pili (VF047
VFG038393(gb|WP_010634228) (dotU) Type VI secretion system protein DotU [T6SS (VF0480) - Effector deli
VFG001142(gb|WP_042960771) (hopZ) outer membrane protein adhesin [HopZ (VF0054) - Adherence (VFC00
VFG038768(gb|WP_016351318) (flgL) flagellar hook-associated protein FlgL [Polar flagella (VF0473) - Motilit
VFG038880(gb|WP_005314250) (lfgK) lateral flagellar hook-associated protein [Lateral flagella (VF0474) - Mot
VFG042736(gb|NP_253152) (rpoN) RNA polymerase factor sigma-54 [Type IV pili (VF0082) - Adherence (VF
VFG038890(gb|WP_005314213) (lafU) LafU, chemotaxis protein [Lateral flagella (VF0474) - Motility (VFC020
VFG038305(gb|WP_016349343) (exeL) general secretion pathway protein L [Exe T2SS (VF0478) - Effector del
VFG038600(gb|WP_016349156) (mshL) MSHA (mannose-sensitive hemagglutinin) pili biogenesis protein Msh
VFG048687(gb|WP_002902160) (hcp/tssD) type VI secretion system protein, Hcp family [T6SS (VF0569) - Eff
VFG000195(gb|NP_250391) (pcr2) type III secretion system protein Pcr2 [TTSS (VF0083) - Effector delivery sy
VFG038719(gb|WP_016350061) (pomB) ompA family protein [Polar flagella (VF0473) - Motility (VFC0204)] [
VFG001224(gb|NP_249087) (pilU) twitching motility protein PilU [Type IV pili (VF0082) - Adherence (VFC00
VFG000353(gb|WP_002208792) (psaC) outer membrane usher protein PsaC [Myf/pH6 antigen (VF0134) - Adhe
VFG049007(gb|WP_002911561) (rcsA) transcriptional activator for ctr capsule biosynthesis [RcsAB (VF0571) -
VFG038627(gb|WP_016350050) (fliF) flagellar MS-ring protein [Polar flagella (VF0473) - Motility (VFC0204)]
VFG041104(gb|NP_459278) (STM0280) type VI secretion system-associated lipoprotein [SCI (Salmonella centr
VFG038727(gb|WP_011705295) (cheW) chemotaxis protein CheW [Polar flagella (VF0473) - Motility (VFC020
VFG011671(gb|YP_002344801) (Cj1418c) hypothetical protein [Capsule (VF0323) - Immune modulation (VFC
VFG049912 (tssC) Type VI secretion system protein TssC [T6SS (VF0579) - Effector delivery system (VFC008
VFG038277(gb|WP_016349336) (exeE) general secretory pathway protein E [Exe T2SS (VF0478) - Effector del
VFG043778(gb|CAA52275) (bont) botulinum neurotoxin, BoNT [BoNT (VF0375) - Exotoxin (VFC0235)] [Clos
VFG038521(gb|WP_016350155) (flpI) Flp pilus assembly protein FlpI [Flp type IV pili (VF0476) - Adherence (
VFG016058(gb|NP_251114) (pvdL) peptide synthase PvdL [Pyoverdine (VF0094) - Nutritional/Metabolic factor
VFG038488(gb|WP_016349432) (tapY1) type IV pilus biogenesis protein [Tap type IV pili (VF0475) - Adheren
VFG038545(gb|WP_043118798) (flpC) Flp pilus secretin FlpC [Flp type IV pili (VF0476) - Adherence (VFC00
VFG012509(gb|WP_001111200) (iroC) ATP binding cassette transporter [Salmochelin siderophore (VF0230) - N
VFG000360(gb|WP_002212799) (ybtT) type II thioesterase YbtT [Yersiniabactin (VF0136) - Nutritional/Metab
VFG000923(gb|WP_001034892) (fepA) ferrienterobactin outer membrane transporter [Enterobactin (VF0228) -
VFG043625(gb|WP_000813718) (mrkC) fimbrial biogenesis outer membrane usher protein mrkC precursor [Ty
VFG002278(gb|WP_011010095) (nagI) hyaluronidase [Mu-toxin (VF0389) - Exoenzyme (VFC0251)] [Clostridi
VFG048676(gb|WP_014907726) (ompA) OmpA family protein [T6SS (VF0569) - Effector delivery system (VF
VFG000877(gb|WP_001244821) (fimF) FimF protein precursor [Type 1 fimbriae (VF0221) - Adherence (VFC0
VFG039226(gb|NP_819077) (cetCb1) Coxiella Dot/Icm type IVB secretion system translocated effector, Coxiell
VFG038723(gb|WP_011705294) (AHML_RS07540) CobQ/CobB/MinD/ParA family protein [Polar flagella (VF
VFG001218(gb|NP_253727) (pilQ) type 4 fimbrial biogenesis protein PilQ [Type IV pili (VF0082) - Adherence
VFG002440(gb|WP_011204593) (bprB) two-component response regulator [Bsa T3SS (VF0428) - Effector deliv
VFG049062(gb|WP_015875020) (rfbD) UDP-galactopyranose mutase [LPS (VF0561) - Immune modulation (V
VFG002416(gb|WP_001265657) (yagW/ecpD) polymerized tip adhesin of ECP fibers [ECP (VF0404) - Adheren
VFG044159(gb|WP_000125846) (fes) enterobactin/ferric enterobactin esterase [Enterobactin (VF0228) - Nutritio

VFG002415(gb|WP_001131063) (yagX/ecpC) E. coli common pilus usher EcpC [ECP (VF0404) - Adherence (V
VFG001826(gb|NP_217099) (relA) Probable GTP pyrophosphokinase RelA (ATP:GTP 3'-pyrophosphotransfera
VFG048885(gb|WP_014907227) (gmd) GDP-mannose 4,6-dehydratase [Capsule (VF0560) - Immune modulatio
VFG046732(gb|WP_003020707) (FTT_RS04140) glycosyltransferase family 2 protein [Capsule (VF0543) - Imm
VFG050266(gb|NP_220214) (CT_695) hypothetical protein [TTSS secreted effectors (VF0711) - Effector delive
VFG038472(gb|WP_011706550) (tapW) twitching ATPase [Tap type IV pili (VF0475) - Adherence (VFC0001)]
VFG038331(gb|WP_005320790) (exsD) Type III secretion system protein ExsD [T3SS (VF0479) - Effector deli
VFG044096(gb|NP_252908) (PA4218) transporter [Pyochelin (VF0095) - Nutritional/Metabolic factor (VFC027
VFG012513(gb|WP_001221122) (iroB) glucosyltransferase IroB [Salmochelin siderophore (VF0230) - Nutrition
VFG050166(gb|WP_000066003) (neuB) N-acylneuraminate-9-phosphate synthase [K1 capsule (VF0239) - Invas
VFG038369(gb|WP_011705706) (hcp) hemolysin co-regulated protein [T6SS (VF0480) - Effector delivery syste
VFG016041(gb|NP_251092) (pvdI) peptide synthase [Pyoverdine (VF0094) - Nutritional/Metabolic factor (VFC
VFG002348(gb|WP_005160443) (flgE) flagellar hook protein FlgE [Flagella (VF0394) - Motility (VFC0204)] [Y
VFG034624(gb|WP_000755097) (cfaB) colonization factor antigen 1 [Adhesive fimbriae (VF0213) - Adherence
VFG038864(gb|WP_073531776) (fliF) flagellar MS-ring protein [Lateral flagella (VF0474) - Motility (VFC0204
VFG048498(gb|WP_012737619) (entF) enterobactin synthase subunit F [Ent (VF0562) - Nutritional/Metabolic f
VFG037852(gb|WP_001075965) (ACICU_RS00395) polysaccharide biosynthesis tyrosine autokinase [Capsule (
VFG034815(gb|WP_000671689) (espR1) Type III secretion system effector espR1 [TTSS secreted effectors (VF
VFG000181(gb|NP_251792) (xcpS) general secretion pathway protein F [Xcp secretion system (VF0084) - Effec
VFG001223(gb|NP_249086) (pilT) twitching motility protein PilT [Type IV pili (VF0082) - Adherence (VFC000
VFG041291(gb|WP_010947429) (ppeB) Dot/Icm type IV secretion system effector PpeB [Dot/Icm T4SS secrete
VFG034922(gb|WP_001303732) (espY5) hypothetical protein [TTSS secreted effectors (VF1110) - Effector deli
VFG050270(gb|NP_220318) (glgA) glycogen synthase [TTSS secreted effectors (VF0711) - Effector delivery sy
VFG044172(gb|WP_000622316) (chuV) ATP-binding hydrophilic protein ChuV [Chu (VF0227) - Nutritional/M
VFG037274(gb|WP_001281538) (barA) siderophore efflux system of the ABC superfamily [Acinetobactin (VF0
VFG014893(gb|NP_254009) (algC) phosphomannomutase AlgC [Alginate (VF0091) - Biofilm (VFC0271)] [Pse
VFG041011(gb|NP_248785) (vgrG1b) type VI secretion system substrate VgrG1b [HSI-I (VF0334) - Effector de
VFG048643(gb|WP_014907727) (vipB/tssC) type VI secretion system contractile sheath large subunit VipB [T6
VFG001444(gb|AAG10151) (aslA) putative arylsulfatase [AslA (VF0238) - Invasion (VFC0083)] [Escherichia c
VFG000929(gb|WP_001375021) (entD) phosphopantetheinyl transferase component of enterobactin synthase mu
VFG000190(gb|NP_250386) (pscP) type III secretion system protein PscP [TTSS (VF0083) - Effector delivery s
VFG007460(gb|WP_000012402) (fliH) flagellar assembly protein [Flagella (VF0519) - Motility (VFC0204)] [Vi
VFG000229 (pilS) pilin [Type IV pili (VF0075) - Adherence (VFC0001)] [Neisseria meningitidis MC58]
VFG000368(gb|WP_000703040) (ybtS) salicylate synthase Irp9 [Yersiniabactin (VF0136) - Nutritional/Metaboli
VFG038529(gb|WP_016350153) (flpG) Flp pilus assembly protein FlpG [Flp type IV pili (VF0476) - Adherence
VFG046191 (CBU_1495*) Coxiella Dot/Icm type IVB secretion system translocated effector [T4SS secreted effe
VFG000903(gb|WP_001045652) (pic) Pic serine protease precursor, autotransporter [Pic (VF0232) - Effector del
VFG038832(gb|WP_016351724) (pomB2) flagellar motor protein MotB [Polar flagella (VF0473) - Motility (VF
VFG048277(gb|WP_009308599) (fimD) outer membrane usher protein [Type I fimbriae (VF0566) - Adherence
VFG016463(gb|WP_011206020) (p97) protein p97; cilium adhesin [P97/P102 paralog family (VF0426) - Adhere
VFG002277(gb|WP_011009663) (nagH) hyaluronidase [Mu-toxin (VF0389) - Exoenzyme (VFC0251)] [Clostrid
VFG002003(gb|WP_010947482) (fliJ) flagellar protein FliJ [Flagella (VF0157) - Motility (VFC0204)] [Legionel
VFG048459(gb|WP_004147525) (entS) enterobactin exporter EntS [Ent (VF0562) - Nutritional/Metabolic factor
VFG038651(gb|WP_011705278) (fliL) flagellar basal body-associated protein FliL [Polar flagella (VF0473) - M
VFG050912(gb|WP_000898330.1) (vgrG/tssI) type VI secretion system tip protein VgrG [T6SS (VF1337) - Effe
VFG000060(gb|WP_010930827) (bopN) Bordetella Bsc type III secretion system secreted protein BopN [TTSS

VFG038417(gb|WP_016349294) (fimD) fimbrial chaperone protein [Type I pili (VF0486) - Adherence (VFC000
VFG041013(gb|NP_250203) (hcpA) secreted protein Hcp [HSI-2 (VF0943) - Effector delivery system (VFC008
VFG048990(gb|WP_001741945) (galF) GalU regulator GalF [Capsule (VF0560) - Immune modulation (VFC025
VFG002369(gb|WP_011816869) (YE_RS15450) hypothetical protein [O-antigen (VF0392) - Immune modulatio
VFG014117(gb|NP_251831) (wbpM) nucleotide sugar epimerase/dehydratase WbpM [LPS (VF0085) - Immune
VFG041284(gb|WP_010947327) (lem11) Dot/Icm type IV secretion system effector Lem11 [Dot/Icm T4SS secr
VFG002548(gb|WP_004550042) (wcbR) capsular polysaccharide biosynthesis fatty acid synthase [Capsule I (V
VFG001225(gb|NP_249099) (pilG) twitching motility protein PilG [Type IV pili (VF0082) - Adherence (VFC00
VFG038273(gb|WP_016349335) (exeD) general secretion pathway protein D [Exe T2SS (VF0478) - Effector de
VFG050217(gb|WP_011860976) (CD0873) ABC transporter substrate-binding protein [CD0873 (VF0593) - Adh
VFG001231(gb|NP_249104) (chpA) still frameshift probable component of chemotactic signal transduction syste
VFG000028(gb|WP_003808614) (brkB) serum resistance protein [Brk (VF0034) - Immune modulation (VFC025
VFG007953(gb|NP_217474) (Rv2958c) PGL/p-HBAD biosynthesis glycosyltransferase [PDIM (VF0309) - Imm
VFG000123(gb|NP_252231) (alg8) alginate-c5-mannuronan-epimerase AlgG [Alginate (VF0091) - Biofilm (VF
VFG043208(gb|WP_005164483) (cheR) chemotaxis methyltransferase CheR [Flagella (VF0394) - Motility (VF
VFG038537(gb|WP_016350151) (flpE) Flp pilus assembly protein, ATPase [Flp type IV pili (VF0476) - Adhere
VFG038643(gb|WP_011705276) (fliJ) flagellar export protein FliJ [Polar flagella (VF0473) - Motility (VFC0204
VFG002412(gb|WP_000716386) (yagY/ecpB) E. coli common pilus chaperone EcpB [ECP (VF0404) - Adheren
VFG001254(gb|NP_249795) (fliI) flagellum-specific ATP synthase FliI [Flagella (VF0273) - Motility (VFC0204
VFG048755(gb|WP_049127998) (icmF/tssM) type VI secretion protein TssM [T6SS (VF0569) - Effector delive
VFG001447(gb|CAA50007) (cnf1) cytotoxic necrotizing factor 1 [CNF-1 (VF0240) - Exotoxin (VFC0235)] [Esc
VFG038671(gb|WP_011705282) (fliQ) flagellar biosynthetic protein FliQ [Polar flagella (VF0473) - Motility (V
VFG045632(gb|WP_002286924) (ecbA/fss3) Collagen binding MSCRAMM, EcbA/Fss3 [EcbA (VF0539) - Adh
VFG037400(gb|WP_001223274) (bauC) ferric siderophore ABC transporter, permease protein BauC [Acinetoba
VFG002107(gb|WP_010946358) (sidA) Dot/Icm type IV secretion system effector SidA [Dot/Icm T4SS secreted
VFG038857(gb|WP_005314307) (fliP) flagellar biosynthesis protein FliP [Lateral flagella (VF0474) - Motility (V
VFG050670(gb|WP_000505931.1) (pilJ) methyl-accepting chemotaxis protein [TFP (VF1334) - Adherence (VF
VFG038348(gb|WP_005320755) (ascN) Type III secretion system ATPase [T3SS (VF0479) - Effector delivery s
VFG038703(gb|WP_011705289) (cheZ) chemotaxis protein CheZ [Polar flagella (VF0473) - Motility (VFC0204
VFG001259(gb|NP_250137) (fliP) flagellar biosynthetic protein FliP [Flagella (VF0273) - Motility (VFC0204)]
VFG050699(gb|WP_000840548.1) (pilR) sigma-54 dependent transcriptional regulator [TFP (VF1334) - Adhere
VFG049909(gb|WP_001113715) (fha) Type VI secretion system protein Fha [T6SS (VF0579) - Effector delivery
VFG049906 (tssK) Type VI secretion system protein TssK [T6SS (VF0579) - Effector delivery system (VFC008
VFG038876(gb|WP_005314259) (lfgG) lateral flagellar basal-body rod protein LfgG [Lateral flagella (VF0474) -
VFG049029(gb|WP_014907241) (KP1_RS17220) glycosyltransferase [LPS (VF0561) - Immune modulation (VF
VFG038281(gb|WP_016349337) (exeF) general secretion pathway protein F [Exe T2SS (VF0478) - Effector deli
VFG038615(gb|WP_016349152) (mshI) MSHA (mannose-sensitive hemagglutinin) pili biogenesis protein MshI,
VFG000872(gb|WP_000044711) (fimE) Type 1 fimbriae Regulatory protein fimE [Type 1 fimbriae (VF0221) -
VFG040864(gb|WP_001172770) (essC) type VII secretion system protein EssC [T7SS (VF0581) - Effector deliv
VFG038371(gb|WP_011705707) (vgrG2) VgrG protein [T6SS (VF0480) - Effector delivery system (VFC0086)]
VFG002450(gb|WP_004533397) (bipB) Type III secretion system translocator protein BipB [Bsa T3SS (VF042
VFG002401(gb|NP_216314) (eccA5) ESX-5 type VII secretion system AAA+ ATPase EccA5 [ESX-5 (VF0402)
VFG038155(gb|WP_000209960) (ACICU_RS00500) phosphomannomutase/phosphoglucomutase [Capsule (VF
VFG002049(gb|WP_001173470) (gspF) general secretion pathway protein F [T2SS (VF0333) - Effector delivery
VFG002053(gb|WP_001250462) (gspJ) general secretion pathway protein J [T2SS (VF0333) - Effector delivery
VFG001845(gb|WP_010946207) (icmF) Dot/Icm type IV secretion system protein IcmF [Dot/Icm (VF0156) - Ef

VFG000030(gb|WP_003807102) (bplI) lipopolysaccharide biosynthesis protein [LPS (VF0033) - Immune modul
VFG040466(gb|WP_012151379) (ompA) surface cell antigen 0 (Sca0), autotransporter [ROmpA/Sca0 (VF0492)
VFG046465(gb|WP_003028672) (tufA) elongation factor Tu [EF-Tu (VF0460) - Adherence (VFC0001)] [Franci
VFG048468(gb|WP_004176917) (fepD) iron-enterobactin transporter membrane protein [Ent (VF0562) - Nutritic
VFG040031(gb|WP_011190505) (rvhB6c) VirB6-like protein of the type IV secretion system [Rvh T4SS (VF048
VFG009810(gb|NP_215272) (phoR) Possible two component system response sensor kinase membrane associate
VFG048287(gb|WP_014906962) (fimF) type 1 fimbrial minor component [Type I fimbriae (VF0566) - Adherenc
VFG038675(gb|WP_011705283) (fliR) flagellar biosynthesis protein FliR [Polar flagella (VF0473) - Motility (V
VFG038301(gb|WP_016349342) (exeK) general secretion pathway protein K [Exe T2SS (VF0478) - Effector del
VFG038590(gb|WP_043118275) (mshE) ATPase protein MshE, GspE-like [MSHA type IV pili (VF0477) - Adh
VFG038796(gb|WP_016351321) (flgE) flagellar hook protein FlgE [Polar flagella (VF0473) - Motility (VFC020
VFG002115(gb|WP_015444209) (sdeB/laiB) Dot/Icm type IV secretion system effector SdeB [Dot/Icm T4SS sec
VFG044333(gb|WP_015874968) (fyuA) yersiniabactin receptor FyuA [Ybt (VF0564) - Nutritional/Metabolic fac
VFG044165(gb|WP_001041793) (entS) enterobactin exporter, iron-regulated [Enterobactin (VF0228) - Nutrition
VFG038367(gb|WP_011705045) (vgrG1) Type VI secretion system effector VgrG1, ADP-ribosyltransferase [T6
VFG002055(gb|WP_000097229) (gspL) general secretion pathway protein L [T2SS (VF0333) - Effector delivery
VFG011850(gb|YP_002343527) (flhG) ATP-binding protein [Flagella (VF0114) - Motility (VFC0204)] [Campyl
VFG038581(gb|WP_043118279) (mshB) MSHA (mannose-sensitive hemagglutinin) pili minor prepilin protein M
VFG002172(gb|WP_010710127) (EF0485) aggregation substance [AS (VF0352) - Adherence (VFC0001)] [Ente
VFG038261(gb|WP_016352107) (exeA) general secretion pathway protein A [Exe T2SS (VF0478) - Effector del
VFG050299(gb|NP_250642) (fapE) chaprone FapE [Fap (VF1191) - Adherence (VFC0001)] [Pseudomonas aeru
VFG048267(gb|WP_015875145) (fimC) periplasmic chaperone [Type I fimbriae (VF0566) - Adherence (VFC00
VFG038764(gb|WP_017409846) (flrA) FleQ protein [Polar flagella (VF0473) - Motility (VFC0204)] [Aeromona
VFG012865 (ipaH) invasion plasmid antigen IpaH [TTSS secreted effectors (VF0978) - Effector delivery system
VFG037735(gb|WP_000633124) (adeH) outer membrane protein [AdeFGH efflux pump (VF0504) - Biofilm (VF
VFG037372(gb|WP_001104139) (bauB) ferric siderophore ABC transporter, periplasmic siderophore-binding pr
VFG038647(gb|WP_016350053) (fliK) flagellar hook-length control protein FliK [Polar flagella (VF0473) - Mot
VFG045314(gb|WP_011213311) (LPG_RS03800) phosphotransferase [LPS (VF0171) - Immune modulation (V
VFG000918(gb|WP_001312167) (chuT) periplasmic heme-binding protein ChuT [Chu (VF0227) - Nutritional/M
VFG049164(gb|WP_004175305) (clbS) colibactin self-protection protein ClbS [Colibactin (VF0573) - Exotoxin
VFG002050(gb|WP_001087302) (gspG) general secretion pathway protein G [T2SS (VF0333) - Effector delivery
VFG007280(gb|WP_000782181) (hap/vvp) hemagglutinin/proteinase HapA [HAP (VF0620) - Exoenzyme (VFC
VFG048951(gb|WP_015875028) (KP1_RS17330) polysaccharide biosynthesis tyrosine autokinase [Capsule (VF
VFG038364(gb|WP_005320615) (aexT) Type III secretion system effector AexT, bifunctional toxin with GTPase
VFG048609(gb|WP_004213923) (iucD) lysine 6-monooxygenase IucD [Aerobactin (VF0565) - Nutritional/Meta
VFG049156(gb|WP_077265119) (clbJ) colibactin non-ribosomal peptide synthetase ClbJ [Colibactin (VF0573) -
VFG002531(gb|WP_011205291) (cheA) chemotaxis two-component sensor kinase CheA [Flagella (VF0430) - M
VFG001872(gb|WP_025862367) (lspH) general secretion pathway protein H [Lsp T2SS (VF0154) - Effector deli
VFG038871(gb|WP_005314274) (lfgA) lateral flagellar basal body P-ring biosynthesis protein, LfgA [Lateral fla
VFG049154(gb|WP_014907257) (clbH) colibactin non-ribosomal peptide synthetase ClbH [Colibactin (VF0573)
VFG048620(gb|WP_014907843) (iutA) ferric aerobactin receptor IutA [Aerobactin (VF0565) - Nutritional/Metal
VFG000980(gb|WP_010921949) (speJ) streptococcal exotoxin J precursor [Spes (VF0248) - Exotoxin (VFC023
VFG000937(gb|WP_000750139) (iucD) L-lysine 6-monooxygenase IucD [Aerobactin (VF0229) - Nutritional/M
VFG000677(gb|AAD32414) (pagA) anthrax toxin moiety, protective antigen [Anthrax toxin (VF0142) - Exotoxin
VFG013418(gb|WP_005694260) (gmhA/lpcA) phosphoheptose isomerase [LOS (VF0044) - Immune modulation
VFG000616(gb|NP_709455) (iucB) aerobactin synthesis protein IucB [Aerobactin (VF0123) - Nutritional/Metab

VFG049904 (clpV/tssH) Type VI secretion system ATPase ClpV/TssH [T6SS (VF0579) - Effector delivery syste
VFG000364(gb|WP_000140406) (ybtA) transcriptional regulator YbtA [Yersiniabactin (VF0136) - Nutritional/M
VFG000114(gb|NP_253218) (xcpA/pilD) type 4 prepilin peptidase PilD [Type IV pili (VF0082) - Adherence (V
VFG000930(gb|WP_000077767) (entF) enterobactin synthase multienzyme complex component, ATP-dependent
VFG044330(gb|WP_032447695) (ybtU) yersiniabactin biosynthesis oxidoreductase YbtU [Ybt (VF0564) - Nutri
VFG000932(gb|WP_000026784) (entE) 2,3-dihydroxybenzoate-AMP ligase component of enterobactin synthase
VFG002499(gb|WP_004197164) (fliI) flagellum-specific ATP synthase FliI [Flagella (VF0430) - Motility (VFC
VFG022082(gb|YP_177711) (PPE4) PPE family protein [ESX-3 (VF0521) - Effector delivery system (VFC0086
VFG002306(gb|NP_461449) (ratB) putative outer membrane protein [RatB (VF0399) - Adherence (VFC0001)] [
VFG001256(gb|NP_250134) (fliM) flagellar motor switch protein FliM [Flagella (VF0273) - Motility (VFC0204
VFG038395(gb|WP_011705721) (clpB) type VI secretion system ATPase ClpV1 [T6SS (VF0480) - Effector deli
VFG011524(gb|WP_002966241) (manBcore) phosphomannomutase [LPS (VF0367) - Immune modulation (VFC
VFG038687(gb|WP_016350057) (flhF) flagellar biosynthesis protein FlhF [Polar flagella (VF0473) - Motility (V
VFG000264(gb|WP_002247545) (lbpA) lactoferrin-binding protein A [Lbp (VF0047) - Nutritional/Metabolic fac
VFG000361(gb|WP_000982866) (ybtU) yersiniabactin biosynthetic protein YbtU [Yersiniabactin (VF0136) - Nu
VFG012162(gb|WP_011590241) (nanJ) exo-alpha-sialidase [Sialidase (VF0391) - Exoenzyme (VFC0251)] [Clo
VFG038269(gb|WP_075384415) (exeC) general secretion pathway protein C [Exe T2SS (VF0478) - Effector del
VFG002259(gb|WP_011181332) (trwK) Trw type IV secretion sysmtem VirB4-like trwK protein [Trw type IV s
VFG049902(gb|WP_011310140) (tssM) Type VI secretion system protein TssM [T6SS (VF0579) - Effector deliv
VFG001243(gb|NP_249776) (flgJ) flagellar rod assembly protein/muramidase FlgJ [Flagella (VF0273) - Motility
VFG000429(gb|WP_010989299) (tviD) Vi polysaccharide biosynthesis protein TviD [Vi antigen (VF0101) - Imm
VFG040863(gb|WP_011190526) (sca2) cell surface antigen Sca2 [Sca2 (VF0495) - Motility (VFC0204)] [Ricket
VFG008136(gb|NP_218340) (mmpL8) integral membrane transport protein MmpL8 [Sulfolipid-1 biosynthesis an
VFG002048(gb|WP_000249362) (gspE) general secretion pathway protein E [T2SS (VF0333) - Effector delivery
VFG000143(gb|NP_253699) (waaF) heptosyltransferase I [LPS (VF0085) - Immune modulation (VFC0258)] [Ps
VFG038788(gb|WP_011706639) (flgG) flagellar basal body rod protein FlgG [Polar flagella (VF0473) - Motility
VFG022051(gb|NP_214798) (eccC3) Type VII secretion system protein EccC3 [ESX-3 (VF0521) - Effector deli
VFG049162(gb|WP_002430641) (clbP) precolibactin peptidase ClbP [Colibactin (VF0573) - Exotoxin (VFC023
VFG002343(gb|WP_042661457) (flgJ) <beta>-N-acetylglucosaminidase [Flagella (VF0394) - Motility (VFC020
VFG002414(gb|WP_000730972) (yagZ/ecpA) E. coli common pilus structural subunit EcpA [ECP (VF0404) - A
VFG042738(gb|NP_253646) (fimX) protein FimX [Type IV pili (VF0082) - Adherence (VFC0001)] [Pseudomon
VFG048551(gb|WP_011251260) (iroC) ABC transporter [Sal (VF0563) - Nutritional/Metabolic factor (VFC027
VFG043012(gb|NP_250147) (cheY) chemotaxis protein CheY [Flagella (VF0273) - Motility (VFC0204)] [Pseud
VFG000001(gb|WP_010930610) (fhaB) filamentous hemagglutinin/adhesin [FHA (VF0025) - Adherence (VFC
VFG000008(gb|WP_010930612) (fimC) outer membrane usher protein precursor [Fimbriae (VF0031) - Adheren
VFG000928(gb|WP_000640938) (fepG) iron-enterobactin ABC transporter permease [Enterobactin (VF0228) - N
VFG048874(gb|WP_041937789) (KP1_RS17305) GDP-L-fucose synthase [Capsule (VF0560) - Immune modula
VFG038576(gb|WP_016349164) (mshC) MSHA (mannose-sensitive hemagglutinin) pili minor prepilin protein M
VFG045349(gb|WP_000092542) (fdeC) adhesin FdeC [FdeC (VF0506) - Adherence (VFC0001)] [Escherichia c
VFG000126(gb|NP_252234) (algE) alginate biosynthetic protein AlgK precursor [Alginate (VF0091) - Biofilm (
VFG000926(gb|WP_001443194) (fepD) ferrienterobactin ABC transporter permease [Enterobactin (VF0228) - N
VFG002179(gb|AAD09858) (esp) Enterococcal surface protein; Esp [Esp (VF0353) - Adherence (VFC0001)] [E
VFG001221(gb|NP_253730) (pilN) type IV pilus inner membrane platform protein PilN [Type IV pili (VF0082)
VFG043014(gb|NP_250149) (PA1458) two-component sensor [Flagella (VF0273) - Motility (VFC0204)] [Pseud
VFG048307(gb|WP_041937815) (fimH) type 1 fimbrial adhesin precursor [Type I fimbriae (VF0566) - Adheren
VFG002335(gb|WP_005160396) (fliM) flagellar motor switch protein FliM [Flagella (VF0394) - Motility (VFC

VFG001354(gb|WP_000728269) (pce/cbpE) choline binding protein E [CBPs (VF0145) - Adherence (VFC0001)
VFG000876(gb|WP_000120946) (fimD) Outer membrane usher protein fimD precursor [Type 1 fimbriae (VF022
VFG048488(gb|WP_002893737) (fepC) iron-enterobactin transporter ATP-binding protein [Ent (VF0562) - Nutr
VFG000359(gb|WP_001088826) (ybtE) yersiniabactin siderophore biosynthetic protein [Yersiniabactin (VF0136
VFG003522(gb|WP_005173749) (YE_RS17760) hypothetical protein [Ysa TTSS (VF1025) - Effector delivery sy
VFG000875(gb|WP_000066579) (fimC) Chaperone protein fimC precursor [Type 1 fimbriae (VF0221) - Adheren
VFG038611(gb|WP_016349153) (mshI1) MSHA (mannose-sensitive hemagglutinin) pili biogenesis protein Msh
VFG000908(gb|WP_000860472) (hlyD) Hemolysin D [-Hemolysin (VF0225) - Exotoxin (VFC0235)] [Escherich
VFG001971(gb|YP_002344826) (kpsF) D-arabinose 5-phosphate isomerase [Capsule (VF0323) - Immune modul
VFG000384(gb|WP_002212971) (yscV/lcrD) type III secretion system major export apparatus protein YscV [TT
VFG006171(gb|WP_002227268) (pilW) type IV fimbrial biogenesis and twitching motility protein PilW [Type I
VFG037028(gb|WP_002218620) (katA) catalase [KatA (VF0454) - Stress survival (VFC0282)] [Neisseria menin
VFG037822(gb|WP_000240700) (lpxC) UDP-3-O-acyl-N-acetylglucosamine deacetylase [LPS (VF0466) - Immu
VFG038309(gb|WP_016349344) (exeM) general secretion pathway protein M [Exe T2SS (VF0478) - Effector de
VFG038812(gb|WP_011706645) (cheR-3) chemotaxis protein CheR [Polar flagella (VF0473) - Motility (VFC02
VFG006582(gb|WP_001862801) (flhF) flagellar biosynthesis protein [Flagella (VF0051) - Motility (VFC0204)]
VFG040665(gb|WP_011190939) (sca4) cell surface antigen Sca4 [Sca4 (VF0502) - Others (VFC0346)] [Ricketts
VFG038892(gb|WP_043118302) (aerA/act) Aerolysin A [Aerolysin (VF0481) - Exotoxin (VFC0235)] [Aeromon
VFG049120(gb|WP_012737535) (allB) allantoinase [Allantion utilization (VF0572) - Nutritional/Metabolic facto
VFG014950(gb|NP_249457) (mucD) serine protease MucD precursor [Alginate (VF0091) - Biofilm (VFC0271)]
VFG000204(gb|NP_250400) (popD) type III secretion system hydrophobic translocator, pore protein PopD [TTS
VFG034756 (espL1) Type III secretion system effector espL1 [TTSS secreted effectors (VF1111) - Effector deliv
VFG040507(gb|WP_011191135) (ompB) outer membrane protein OmpB [ROmpB/Sca5 (VF0493) - Invasion (V
VFG038541(gb|WP_016350150) (flpD) Flp pilus assembly protein FlpD [Flp type IV pili (VF0476) - Adherence
VFG014984(gb|NP_253136) (algW) AlgW protein [Alginate (VF0091) - Biofilm (VFC0271)] [Pseudomonas aer
VFG001827(gb|WP_001020414) (senB) enterotoxin [ShET2 (VF0258) - Exotoxin (VFC0235)] [Shigella boydii S
VFG001252(gb|NP_249793) (fliG) flagellar motor switch protein G [Flagella (VF0273) - Motility (VFC0204)] [
VFG048348(gb|WP_004149658) (mrkJ) phosphodiesterase [Type 3 fimbriae (VF0567) - Biofilm (VFC0271)] [K
VFG039501(gb|NP_820443) (coxCC7/cig44) Coxiella Dot/Icm type IVB secretion system translocated effector [
VFG038760(gb|WP_016351312) (flrB) two-component sensor kinase [Polar flagella (VF0473) - Motility (VFC0
VFG044327(gb|WP_000140406) (ybtA) yersiniabactin transcriptional regulator YbtA [Ybt (VF0564) - Nutritiona
VFG001381(gb|YP_177728) (icl) Isocitrate lyase Icl (isocitrase) (isocitratase) [Isocitrate lyase (VF0253) - Others
VFG038403(gb|WP_011705726) (atsS) Type VI secretion system protein [T6SS (VF0480) - Effector delivery sys
VFG044326(gb|WP_001327262) (ybtP) yersiniabactin ABC transporter ATP-binding/permease protein YbtP [Yb
VFG048409(gb|WP_086074289) (entA) 2,3-dihydroxybenzoate-2,3-dehydrogenase [Ent (VF0562) - Nutritional/
VFG001895(gb|YP_002343528) (flhF) flagellar biosynthesis regulator FlhF [Flagella (VF0114) - Motility (VFC0
VFG000924(gb|WP_001234311) (fepB) ferrienterobactin ABC transporter periplasmic binding protein [Enteroba
VFG038505(gb|WP_011705643) (tapF) type IV pilus biogenesis/stability protein, pilW-like [Tap type IV pili (VF
VFG044325(gb|WP_002212761) (ybtQ) yersiniabactin ABC transporter ATP-binding/permease protein YbtQ [Y
VFG000879(gb|WP_000832235) (fimH) FimH protein precursor [Type 1 fimbriae (VF0221) - Adherence (VFC0
VFG050037(gb|WP_001133943) (dhbF) non-ribosomal peptide synthetase, DhbF [Bacillibactin (VF0586) - Nutr
VFG024730(gb|NP_217462) (pks1) polyketide synthase [PDIM (VF0309) - Immune modulation (VFC0258)] [M
VFG038385(gb|WP_011705716) (atsI) Type VI secretion system protein [T6SS (VF0480) - Effector delivery sys
VFG002196(gb|WP_011109578) (EF3023) polysaccharide lyase, family 8 [Hyaluronidase (VF0359) - Exoenzym
VFG000935(gb|WP_001222189) (iroN) salmochelin receptor IroN [Salmochelin siderophore (VF0230) - Nutritio
VFG013374(gb|WP_005693262) (lpxD) UDP-3-O-(3-hydroxymyristoyl) glucosamine N-acyltransferase [LOS (V

VFG001229(gb|NP_249103) (pilK) methyltransferase PilK [Type IV pili (VF0082) - Adherence (VFC0001)] [Ps
VFG048508(gb|WP_012737618) (fes) enterobactin/ferric enterobactin esterase [Ent (VF0562) - Nutritional/Meta
VFG038683(gb|WP_016350056) (flhA) flagellar biosynthesis protein FlhA [Polar flagella (VF0473) - Motility (V
VFG000161(gb|NP_251089) (pvdD) pyoverdine synthetase D [Pyoverdine (VF0094) - Nutritional/Metabolic fact
VFG002164(gb|WP_002395058) (prgB/asc10) aggregation substance PrgB/Asc10 [AS (VF0352) - Adherence (V
VFG044324(gb|WP_032447693) (ybtX) yersiniabactin-associated zinc MFS transporter YbtX [Ybt (VF0564) - N
VFG050625(gb|WP_000389061.1) (pilG) twitching motility response regulator PilG [TFP (VF1334) - Adherence
VFG038655(gb|WP_011705279) (fliM) flagellar motor switch protein FliM [Polar flagella (VF0473) - Motility (
VFG038663(gb|WP_005298795) (fliO) flagellar biosynthesis protein, FliO [Polar flagella (VF0473) - Motility (V
VFG043623(gb|WP_002916122) (mrkF) type 3 fimbrial minor pilin subunit MrkF [Type 3 fimbriae (VF0567) - I
VFG005942(gb|WP_000660895) (GBS_RS06570) glycosyltransferase [Capsule (VF0274) - Immune modulation
VFG040422(gb|WP_011190749) (rvhD4) type IV secretion system component VirD4 [Rvh T4SS (VF0488) - Eff
VFG050802(gb|WP_000865310.1) (gspG) general secretion pathway protein G [T2SS (VF1335) - Effector delive
VFG045464(gb|WP_000562556) (tcpC) Tir domain containing protein TcpC [TcpC (VF0413) - Immune modula
VFG038497(gb|WP_016351992) (tppF) type IV pilus pseudopilin [Tap type IV pili (VF0475) - Adherence (VFC
VFG001396(gb|NP_216424) (katG) catalase-peroxidase [KatG (VF0303) - Stress survival (VFC0282)] [Mycoba
VFG000933(gb|WP_001007140) (entB) isochorismatase [Enterobactin (VF0228) - Nutritional/Metabolic factor (
VFG050163(gb|WP_000197083) (neuE) polysialic acid biosynthesis protein [K1 capsule (VF0239) - Invasion (V
VFG000347(gb|WP_010869038) (hgpC) hemoglobin-binding protein [Hgp (VF0270) - Nutritional/Metabolic fac
VFG002154(gb|NP_219969) (tarp) Type III secretion system translocated actin-recruiting protein [TTSS secreted
VFG000131(gb|NP_252239) (algJ) alginate o-acetyltransferase AlgJ [Alginate (VF0091) - Biofilm (VFC0271)] [
VFG000362(gb|WP_002212777) (irp1) yersiniabactin biosynthetic protein Irp1 [Yersiniabactin (VF0136) - Nutri
VFG000884(gb|WP_000654937) (papC) usher protein PapC [P fimbriae (VF0220) - Adherence (VFC0001)] [Es
VFG050386(gb|WP_000355489.1) (pilT) type IV pilus twitching motility protein PilT [TFP (VF1334) - Adheren
VFG034908(gb|WP_000978349) (espY3) Type III secretion system effector EspY3 [TTSS secreted effectors (V
VFG038448(gb|WP_016351632) (tapN) type IV pilus biogenesis protein PilN [Tap type IV pili (VF0475) - Adhe
VFG045793(gb|WP_000771429) (csgA) curlin major subunit CsgA [Curli fibers (VF1138) - Adherence (VFC00
VFG000165(gb|NP_252915) (pchF) pyochelin synthetase PchF [Pyochelin (VF0095) - Nutritional/Metabolic fact
VFG013368(gb|WP_005693225) (rffG) dTDP-glucose 46-dehydratase [LOS (VF0044) - Immune modulation (V
VFG049084(gb|WP_004175264) (rfbB) O-antigen export ABC transporter ATP-binding protein RfbB [LPS (VF
VFG000873(gb|WP_000695571) (fimA) Type-1 fimbrial protein, A chain precursor [Type 1 fimbriae (VF0221) -
VFG037762(gb|WP_000078878) (lpxL) lauroyl acyltransferase [LPS (VF0466) - Immune modulation (VFC0258
VFG034928(gb|WP_001239082) (lifA/efa1) lymphostatin Efa1/LifA [Lymphostatin/LifA (VF0175) - Effector de
VFG012603(gb|WP_001302516) (chuT) periplasmic heme-binding protein ChuT [Chu (VF0234) - Nutritional/M
VFG000227(gb|WP_002221783) (pilS) pilin [Type IV pili (VF0075) - Adherence (VFC0001)] [Neisseria mening
VFG043774(gb|AAB24244) (bont) botulinum neurotoxin, BoNT [BoNT (VF0375) - Exotoxin (VFC0235)] [Clos
VFG002413(gb|WP_000389022) (ykgK/ecpR) regulator protein EcpR [ECP (VF0404) - Adherence (VFC0001)]
VFG049155(gb|WP_000829570) (clbI) colibactin polyketide synthase ClbI [Colibactin (VF0573) - Exotoxin (VF
VFG013315(gb|WP_005694490) (lgtF) beta-1,4-glucosyltransferase [LOS (VF0044) - Immune modulation (VFC
VFG049149(gb|WP_041169023) (clbC) colibactin polyketide synthase ClbC [Colibactin (VF0573) - Exotoxin (V
VFG000690(gb|AAP42183) (AAP42183) Bcs3' [Capsule (VF0043) - Immune modulation (VFC0258)] [Haemop
VFG038413(gb|WP_043118350) (fimC) outer membrane usher protein [Type I pili (VF0486) - Adherence (VFC
VFG002148(gb|NP_220192) (pkn5) serine/threonine protein kinase [TTSS secreted effectors (VF0711) - Effector
VFG002129(gb|NP_220075) (CT_560) hypothetical protein [TTSS (VF0344) - Effector delivery system (VFC00
VFG033466(gb|WP_000381567) (espA) Type III secretion system translocator protein EspA [TTSS (VF0172) - I
VFG000133(gb|NP_252241) (algA) phosphomannose isomerase / guanosine 5'-diphospho-D-mannose pyrophosp

VFG038317(gb|WP_016352170) (tapD/pilD) prepilin peptidase [Tap type IV pili (VF0475) - Adherence (VFC00
VFG001242(gb|NP_249775) (flgI) flagellar P-ring protein precursor FlgI [Flagella (VF0273) - Motility (VFC020
VFG019178(gb|WP_012291519) (bont) botulinum neurotoxin, BoNT [BoNT (VF0375) - Exotoxin (VFC0235)] [
VFG000833(gb|WP_001301460) (espG) Type III secretion system effector EspG, GTPase activating protein [TT
VFG012609(gb|WP_000993317) (chuW) Putative oxygen independent coproporphyrinogen III oxidase [Chu (VF
VFG000139(gb|NP_253697) (waaG) B-band O-antigen polymerase [LPS (VF0085) - Immune modulation (VFC0
VFG050160(gb|WP_000030755) (kpsU) 3-deoxy-manno-octulosonate cytidylyltransferase [K1 capsule (VF0239
VFG048621(gb|WP_004213924) (iutA) ferric aerobactin receptor IutA [Aerobactin (VF0565) - Nutritional/Metal
VFG049916(gb|WP_000103365) (vgrG/tssI) type VI secretion system effector VgrG [T6SS (VF0579) - Effector
VFG000112(gb|NP_253216) (pilB) type 4 fimbrial biogenesis protein PilB [Type IV pili (VF0082) - Adherence (
VFG038867(gb|WP_005314284) (lfiI) lateral flagellar FliI-like assembly ATPase [Lateral flagella (VF0474) - Mo
VFG038460(gb|WP_043119325) (tapQ) type IV pilus secretin PilQ [Tap type IV pili (VF0475) - Adherence (VF
VFG002437(gb|WP_011325557) (boaB) autotransporter protein [BoaB (VF0435) - Adherence (VFC0001)] [Burl
VFG038421(gb|WP_043118352) (fimE) fimbrial protein [Type I pili (VF0486) - Adherence (VFC0001)] [Aerom
VFG002076(gb|NP_248780) (clpV1) type VI secretion system AAA+ family ATPase [HSI-I (VF0334) - Effector
VFG000157(gb|NP_249535) (plcH) hemolytic phospholipase C precursor [PLC (VF0092) - Exotoxin (VFC0235)
VFG013647(gb|WP_041175806) (invC/ilp) intimin-like invasin C protein [Invasin C/Ilp (VF0575) - Invasion (VF
VFG034679(gb|WP_000074207) (ibeB) Cu(+)/Ag(+) efflux RND transporter outer membrane channel CusC [Ibe
VFG037176(gb|WP_001081735) (plc1) phospholipase C [Phospholipase C (VF0470) - Exotoxin (VFC0235)] [A
VFG051042(gb|WP_000653195.1) (hcp/tssD) type VI secretion system tube protein Hcp [T6SS (VF1337) - Effec
VFG038635(gb|WP_016350051) (fliH) flagellar assembly protein FliH [Polar flagella (VF0473) - Motility (VFC
VFG001935(gb|YP_002344312) (ciaB) invasion antigen CiaB [CiaB (VF0324) - Invasion (VFC0083)] [Campylo
VFG038359(gb|WP_005321607) (aopH) Type III secretion system effector AopH, putative tyrosine phosphatase
VFG000478(gb|NP_459678) (fur) ferric iron uptake transcriptional regulator [Fur (VF0113) - Regulation (VFC0
VFG002176(gb|AAM75252) (cylB) ABC-type transporter [Cytolysin (VF0356) - Exotoxin (VFC0235)] [Enteroc
VFG048665(gb|WP_004176431) (dotU/tssL) type VI secretion system protein, DotU/TssL family [T6SS (VF056
VFG050159(gb|WP_001331697) (kpsD) Polysialic acid transport protein precursor [K1 capsule (VF0239) - Invas
VFG049157(gb|WP_014907259) (clbK) colibactin hybrid non-ribosomal peptide synthetase/type I polyketide syn
VFG001443(gb|AAF37887) (ompA) outer membrane protein A [OmpA (VF0236) - Invasion (VFC0083)] [Esche
VFG000936(gb|WP_016265865) (iutA) ferric aerobactin receptor precusor IutA [Aerobactin (VF0229) - Nutritio
VFG005486(gb|WP_000417180) (iga) G5 domain-containing protein [IgA1 protease (VF0147) - Immune modula
VFG050946(gb|WP_000083625.1) (tssL) type IVB secretion system protein TssL [T6SS (VF1337) - Effector del
VFG002321(gb|WP_011816589) (fliC) flagellin [Flagella (VF0394) - Motility (VFC0204)] [Yersinia enterocoliti
VFG044323(gb|WP_015874960) (ybtS) yersiniabactin biosynthesis salicylate synthase YbtS [Ybt (VF0564) - Nu
VFG038587(gb|WP_016349160) (mshG) MSHA (mannose-sensitive hemagglutinin) pili biogenesis protein Msh
VFG034612(gb|WP_000225871) (cfaA) colonisation factor antigen a, chaperone [Adhesive fimbriae (VF0213) -
VFG000916(gb|WP_001017192) (chuS) heme oxygenase ChuS [Chu (VF0227) - Nutritional/Metabolic factor (V
VFG045790(gb|WP_000833288) (cgsE) curli production assembly/transport protein CsgE [Curli fibers (VF1138)
VFG038699(gb|WP_010634617) (cheY) chemotaxis protein CheY [Polar flagella (VF0473) - Motility (VFC0204
VFG048317(gb|WP_014906959) (fimK) transcriptional regulator [Type I fimbriae (VF0566) - Adherence (VFC0
VFG038464(gb|WP_011707389) (tapT) twitching ATPase [Tap type IV pili (VF0475) - Adherence (VFC0001)]
VFG043624(gb|WP_004149659) (mrkD) fimbrial adhesin protein precursor MrkD [Type 3 fimbriae (VF0567) -
VFG038578(gb|WP_016349163) (mshA) prepilin-type N-terminal cleavage/methylation domain-containing prote
VFG000912(gb|WP_001443207) (focD) F1C fimbrial usher [F1C fimbriae (VF0224) - Adherence (VFC0001)] [
VFG037330(gb|WP_001177743) (basD) acinetobactin biosynthesis protein BasD [Acinetobactin (VF0467) - Nut
VFG038293(gb|WP_016349340) (exeI) general secretion pathway protein I [Exe T2SS (VF0478) - Effector deliv

VFG038176(gb|WP_000777882) (ompA) outer membrane protein OmpA [OmpA (VF0464) - Immune modulatio
VFG001892(gb|YP_002343523) (fliY) flagellar motor switch protein FliY [Flagella (VF0114) - Motility (VFC02
VFG001267(gb|NP_252913) (pchH) ABC transporter ATP-binding protein [Pyochelin (VF0095) - Nutritional/M
VFG048654(gb|WP_004224345) (vasE/tssK) type VI secretion system baseplate subunit TssK [T6SS (VF0569) -
VFG001405(gb|NP_217219) (sigA/rpoV) RNA polymerase sigma factor SigA [SigA (VF0257) - Regulation (VF
VFG038501(gb|WP_043119142) (tapV) type IV pilus assembly protein TapV [Tap type IV pili (VF0475) - Adhe
VFG002051(gb|WP_001115126) (gspH) general secretion pathway protein H [T2SS (VF0333) - Effector delivery
VFG000889(gb|WP_000113351) (papF) P pilus minor subunit PapF [P fimbriae (VF0220) - Adherence (VFC000
VFG041251(gb|WP_010946186) (ceg14/sidL) Dot/Icm type IV secretion system effector Ceg14/SidL [Dot/Icm T
VFG043015(gb|NP_250150) (PA1459) chemotaxis-specific methylesterase [Flagella (VF0273) - Motility (VFC0
VFG048980(gb|WP_041937790) (KP1_RS17345) capsule assembly Wzi family protein [Capsule (VF0560) - Im
VFG037777(gb|WP_000867091) (lpsB) glycosyltransferase family 4 protein [LPS (VF0466) - Immune modulatio
VFG001846(gb|WP_010946208) (icmH/dotU) Dot/Icm type IV secretion system protein IcmH/DotU [Dot/Icm (V
VFG000874(gb|WP_000824100) (fimI) Fimbrin-like protein fimI precursor [Type 1 fimbriae (VF0221) - Adhere
VFG000172(gb|NP_252898) (phzM) phenazine-specific methyltransferase PhzM (adenosylmethionine dependen
VFG050313(gb|WP_004644147.1) (pilM) pilus assembly protein PilM [TFP (VF1334) - Adherence (VFC0001)]
VFG038436(gb|WP_016352169) (tapC) type IV fimbrial assembly protein, PilC-like [Tap type IV pili (VF0475)
VFG048297(gb|WP_014906961) (fimG) type 1 fimbrial minor component [Type I fimbriae (VF0566) - Adherenc
VFG043627(gb|WP_000756959) (mrkA) type 3 fimbrial major pilin subunit MrkA [Type 3 fimbriae (VF0567) -
VFG038667(gb|WP_016350054) (fliP) flagellar biosynthesis protein FliP [Polar flagella (VF0473) - Motility (VF
VFG034721(gb|WP_000556298) (ibeC) phosphoethanolamine transferase CptA [Ibes (VF0237) - Invasion (VFC
VFG041020(gb|NP_250353) (PA1662) ClpA/B-type protease [HSI-2 (VF0943) - Effector delivery system (VFC
VFG034810(gb|NP_309848) (espO1-2) type III secreted effector [TTSS secreted effectors (VF1110) - Effector de
VFG002352(gb|WP_005168790) (flgA) periplasmic flagellar chaperone protein FlgA required for P-ring assembl
VFG051070(gb|WP_001066523.1) (tssC) type VI secretion system contractile sheath large subunit [T6SS (VF13
VFG043213(gb|WP_005164494) (motA) flagellar motor protein MotA [Flagella (VF0394) - Motility (VFC0204)
VFG048779(gb|WP_004190503) (tssG) type VI secretion system baseplate subunit TssG [T6SS (VF0569) - Effe
VFG013063(gb|WP_001017208) (shuS) heme/hemoglobin transport protein ShuS [Shu (VF0256) - Nutritional/M
VFG037792(gb|WP_000064875) (lpxA) acyl-ACP--UDP-N-acetylglucosamine O-acyltransferase [LPS (VF0466
VFG048787(gb|WP_004176417) (sciN/tssJ) type VI secretion system lipoprotein TssJ [T6SS (VF0569) - Effecto
VFG048518(gb|WP_012737617) (fepA) outer membrane receptor FepA [Ent (VF0562) - Nutritional/Metabolic f
VFG000036(gb|WP_010929614) (bplC) lipopolysaccharide biosynthesis protein [LPS (VF0033) - Immune modu
VFG043626(gb|WP_000820818) (mrkB) fimbrial chaperone protein mrkB precursor [Type 3 fimbriae (VF0567)
VFG001857(gb|WP_010946483) (iraB) Di/tripeptide permease [IraAB (VF0293) - Nutritional/Metabolic factor (
VFG038378(gb|WP_011705712) (vipA) Type VI secretion system contractile sheath small subunit TssB/VipA [T
VFG049118(gb|WP_012737529) (allR) DNA-binding transcriptional repressor AllR [Allantion utilization (VF05
VFG002074(gb|NP_248778) (hsiG1/tssF) type VI secretion system hcp secretion island protein HsiG1 [HSI-I (V
VFG000065(gb|WP_010930832) (bscD) putative type III secretion protein [TTSS (VF0035) - Effector delivery s
VFG049040(gb|WP_074160401) (KP1_RS17225) glycosyltransferase family 4 protein [LPS (VF0561) - Immune
VFG044336(gb|WP_050767835) (iroC) ABC transporter [Sal (VF0563) - Nutritional/Metabolic factor (VFC0272
VFG006587(gb|WP_000602430) (fliA) flagellar biosynthesis sigma factor FliA [Flagella (VF0051) - Motility (V
VFG042978(gb|WP_011109477) (ebpC) endocarditis and biofilm-associated pilus major subunit EbpC [Ebp pili
VFG038639(gb|WP_011705275) (fliI) flagellum-specific ATP synthase [Polar flagella (VF0473) - Motility (VFC
VFG050972(gb|WP_000972583.1) (tagX) type VI-associated gene X, peptidoglycan hydrolase [T6SS (VF1337)
VFG038659(gb|WP_011705280) (fliN) flagellar motor switch protein FliN [Polar flagella (VF0473) - Motility (V
VFG038285(gb|WP_005308483) (exeG) general secretion pathway protein G [Exe T2SS (VF0478) - Effector del

VFG002224(gb|WP_002963682) (wbkA) mannosyltransferase [LPS (VF0367) - Immune modulation (VFC0258)
VFG038346(gb|WP_005320760) (acr1) Type III secretion system gatekeeper subunit [T3SS (VF0479) - Effector
VFG001392(gb|NP_215737) (sigE) ECF RNA polymerase sigma factor SigE [SigE (VF0295) - Regulation (VFC
VFG034636(gb|WP_000358674) (cfaC) colonisation factor antigen c, usher [Adhesive fimbriae (VF0213) - Adhe
VFG002056 (gspM) general secretion pathway protein M [T2SS (VF0333) - Effector delivery system (VFC0086)
VFG022146(gb|NP_214805) (mycP3) Type VII secretion system membrane-anchored mycosin MycP3 [ESX-3 (
VFG000917(gb|WP_000089583) (chuA) Outer membrane heme/hemoglobin receptor ChuA [Chu (VF0227) - Nu
VFG002173(gb|WP_010785625) (asa1) aggregation substance Asa1 [AS (VF0352) - Adherence (VFC0001)] [En
VFG038549(gb|WP_016350148) (flpB) Flp pilus assembly protein [Flp type IV pili (VF0476) - Adherence (VFC
VFG000890(gb|WP_000758683) (papG) P pilus tip adhesin PapG [P fimbriae (VF0220) - Adherence (VFC0001)
VFG038772(gb|WP_016351319) (flgK) flagellar hook-associated protein FlgK [Polar flagella (VF0473) - Motilit
VFG001915(gb|YP_002344227) (fliP) flagellar biosynthesis protein FliP [Flagella (VF0114) - Motility (VFC020
VFG000333(gb|WP_005694147) (lic2A) glycosyltransferase [LOS (VF0044) - Immune modulation (VFC0258)]
VFG000426(gb|WP_001023498) (vexB) Vi polysaccharide export inner-membrane protein (ABC-2 type transpo
VFG000267(gb|WP_002225846) (hpuA) haemoglobin-haptoglobin-utilization protein [HpuAB (VF0049) - Nutri
VFG002046(gb|WP_000135089) (gspC) general secretion pathway protein C [T2SS (VF0333) - Effector delivery
VFG002034(gb|WP_010874808) (hmw3) cytadherence accessory protein HMW3 [Cytadherence organelle (VF0
VFG015045(gb|NP_249821) (rhlC) rhamnosyltransferase 2 [Rhamnolipid (VF0089) - Immune modulation (VFC
VFG006717(gb|NP_465159) (lap) Listeria adhesion protein Lap [Lap (VF0444) - Adherence (VFC0001)] [Lister
VFG038882(gb|WP_005314247) (maf-5) modification accessory factor [Lateral flagella (VF0474) - Motility (VF
VFG038297(gb|WP_016349341) (exeJ) general secretion pathway protein J [Exe T2SS (VF0478) - Effector deli
VFG000010(gb|WP_010930243) (tcfA) tracheal colonization factor precursor [TcfA (VF0032) - Adherence (VF
VFG000140(gb|NP_253696) (waaP) UDP-glucose:(heptosyl) LPS alpha 1,3-glucosyltransferase WaaG [LPS (V
VFG038756(gb|WP_011706626) (flrC) transcriptional activator [Polar flagella (VF0473) - Motility (VFC0204)]
VFG038840(gb|WP_005300916) (flmH) short chain dehydrogenase/reductase family oxidoreductase [Polar flage
VFG001226(gb|NP_249100) (pilH) twitching motility protein PilH [Type IV pili (VF0082) - Adherence (VFC00
VFG038874(gb|WP_005314263) (flgE) flagellar hook protein FlgE [Lateral flagella (VF0474) - Motility (VFC02
VFG038383(gb|WP_011705715) (atsH) Type VI secretion system protein [T6SS (VF0480) - Effector delivery sy
VFG038780(gb|WP_011706637) (flgI) flagellar basal body P-ring protein [Polar flagella (VF0473) - Motility (V
VFG050157(gb|WP_001331698) (kpsF) Polysialic acid capsule expression protein [K1 capsule (VF0239) - Invas
VFG001391(gb|NP_215677) (narG) nitrate reductase subunit alpha [Nitrate reductase (VF0302) - Nutritional/Me
VFG037513(gb|WP_085947930) (ACICU_RS04590) porin family protein [HemO cluster (VF0636) - Nutritional
VFG038804(gb|WP_011706643) (flgC) flagellar basal body rod protein FlgC [Polar flagella (VF0473) - Motility
VFG038265(gb|WP_016352108) (exeB) general secretion pathway protein B [Exe T2SS (VF0478) - Effector del
VFG049152(gb|WP_000337350) (clbF) colibactin biosynthesis dehydrogenase ClbF [Colibactin (VF0573) - Exo
VFG038561(gb|WP_016349168) (mshQ) MSHA (mannose-sensitive hemagglutinin) pili biogenesis protein Msh
VFG007832(gb|NP_217448) (ppsB) phthiocerol synthesis polyketide synthase type I PpsB [PDIM (VF0309) - In
VFG043500(gb|WP_011206108) (mhp271) p97 cilium adhesin paralog [P97/P102 paralog family (VF0426) - Ad
VFG049843(gb|WP_003155066) (exlB) hemolysin activator protein [Exolysin (VF0582) - Effector delivery syste
VFG000940(gb|WP_001299620) (iucA) aerobactin siderophore biosynthesis protein IucD [Aerobactin (VF0229)
VFG049144(gb|WP_002892069) (acrB) acriflavine resistance protein B [AcrAB (VF0568) - Antimicrobial activi
VFG000477(gb|NP_461845) (rpoS) RNA polymerase sigma factor RpoS [RpoS (VF0112) - Regulation (VFC03
VFG039436 (CBU_1107*) Coxiella Dot/Icm type IVB secretion system translocated effector [T4SS secreted effe
VFG041038(gb|NP_251056) (PA2366) uricase [HSI-3 (VF0944) - Effector delivery system (VFC0086)] [Pseudo
VFG048257(gb|WP_004174507) (fimI) type 1 pilus biosynthesis fimbrial protein [Type I fimbriae (VF0566) - A
VFG038800(gb|WP_016351322) (flgD) flagellar hook capping protein [Polar flagella (VF0473) - Motility (VFC

VFG000294(gb|WP_000938081) (cagQ) type IV secretion system protein Cag14/CagQ [Type IV secretion syste
VFG049148(gb|WP_041168941) (clbB) colibactin hybrid non-ribosomal peptide synthetase/type I polyketide syn
VFG019180(gb|WP_011987710) (bont) botulinum neurotoxin, BoNT [BoNT (VF0375) - Exotoxin (VFC0235)] [
VFG043209(gb|WP_005168769) (cheD) methyl-accepting chemotaxis protein CheD [Flagella (VF0394) - Motili
VFG042858(gb|WP_003018618) (pilN) Type IV pilus associated protein [Type IV pili (VF0458) - Adherence (V
VFG038776(gb|WP_016351320) (flgJ) flagellar rod assembly protein/muramidase FlgJ [Polar flagella (VF0473)
VFG001230(gb|NP_251805) (fimV) a polar peptidoglycan-binding protein involved in type IV pilus assembly [T
VFG038784(gb|WP_011706638) (flgH) flagellar basal body L-ring protein [Polar flagella (VF0473) - Motility (V
VFG041329(gb|WP_010948104) (legL7) Dot/Icm type IV secretion system effector LegL7 [Dot/Icm T4SS secre
VFG048338(gb|WP_002916079) (mrkI) LuxR family regulatory protein [Type 3 fimbriae (VF0567) - Biofilm (V
VFG049133(gb|WP_004177236) (acrA) acriflavine resistance protein A [AcrAB (VF0568) - Antimicrobial activ
VFG038407(gb|WP_011705728) (vgrG3) VgrG protein [T6SS (VF0480) - Effector delivery system (VFC0086)]
VFG050168(gb|WP_000590258) (kpsT) Polysialic acid transport ATP-binding protein [K1 capsule (VF0239) - I
VFG049095(gb|WP_002912373) (rfbA) O-antigen export ABC transporter permease RfbA [LPS (VF0561) - Imm
VFG044332(gb|WP_015874967) (ybtE) yersiniabactin biosynthesis salycil-AMP ligase YbtE [Ybt (VF0564) - N
VFG001810(gb|YP_178017) (fbpD) MPT51/MPB51 antigen [Antigen 85 (VF0311) - Adherence (VFC0001)] [M
VFG000064(gb|WP_010930831) (bcrD) Type III secretion system LcrD homolog protein BcrD [TTSS (VF0035)
VFG032992(gb|NP_464816) (oatA) peptidoglycan O-acetyltransferase [OatA (VF0441) - Immune modulation (V
VFG038845(gb|WP_016351249) (nueA) NeuA protein [Polar flagella (VF0473) - Motility (VFC0204)] [Aeromo
VFG038533(gb|WP_016350152) (flpF) traffic ATPase [Flp type IV pili (VF0476) - Adherence (VFC0001)] [Ae
VFG048708(gb|WP_041168969) (vgrG/tssI) type VI secretion system tip protein VgrG [T6SS (VF0569) - Effect
VFG038355(gb|WP_005320739) (ascU) Type III secretion system inner membrane export apparatus protein Asc
VFG038572(gb|WP_016349165) (mshD) MSHA (mannose-sensitive hemagglutinin) pili minor prepilin protein M
VFG039131(gb|NP_820609) (icmE/dotG) Type IVB secretion system protein IcmE/DotG, inner membrane prote
VFG000927(gb|WP_000096764) (fepE) LPS O-antigen length regulator [Enterobactin (VF0228) - Nutritional/M
VFG050772(gb|WP_001020922.1) (gspE2) putative secretion pathway ATPase [T2SS (VF1335) - Effector deliv
VFG034875(gb|WP_001270087) (espX5) Type III secretion system effector EspX5 [TTSS secreted effectors (V
VFG045791(gb|WP_000481500) (cgsD) transcriptional regulator CsgD [Curli fibers (VF1138) - Adherence (VF
VFG038352(gb|WP_005320745) (ascR) Type III secretion system inner membrane export apparatus protein Asc
VFG006217 (opa) outer membrane beta-barrel protein [Opa (VF0076) - Invasion (VFC0083)] [Neisseria mening
VFG038401(gb|WP_011705724) (atsQ) Type VI secretion system protein [T6SS (VF0480) - Effector delivery sy
VFG038836(gb|WP_011707085) (pomA2) chemotaxis protein PomA [Polar flagella (VF0473) - Motility (VFC0
VFG049898(gb|WP_001284199) (hcp1/tssD1) type VI secretion system protein, Hcp family [T6SS (VF0579) - E
VFG038631(gb|WP_011705273) (fliG) flagellar motor switch protein G [Polar flagella (VF0473) - Motility (VF
VFG049899(gb|WP_001087735) (tssA) Type VI secretion system protein TssA [T6SS (VF0579) - Effector deliv
VFG038313(gb|WP_043118380) (exeN) general secretion pathway protein N [Exe T2SS (VF0478) - Effector del
VFG001939(gb|YP_002344531) (Cj1138) glycosyltransferase [LOS (VF0326) - Immune modulation (VFC0258)
VFG000180(gb|NP_251791) (xcpT) general secretion pathway protein G [Xcp secretion system (VF0084) - Effe
VFG038792(gb|WP_011706640) (flgF) flagellar basal-body rod protein FlgF [Polar flagella (VF0473) - Motility
VFG044335(gb|WP_015874980) (iroB) glucosyltransferase IroB [Sal (VF0563) - Nutritional/Metabolic factor (V
VFG001862(gb|WP_015444101) (katB) catalase-peroxidase KatB [KatAB (VF0168) - Stress survival (VFC028
VFG013487(gb|WP_005694185) (lsgE) glycosyltransferase [LOS (VF0044) - Immune modulation (VFC0258)] [
VFG044322(gb|WP_004190148) (iroE) siderophore esterase IroE [Sal (VF0563) - Nutritional/Metabolic factor (
VFG012095(gb|WP_003435012) (groEL) chaperonin GroEL [GroEL (VF0594) - Adherence (VFC0001)] [Clost
VFG000038(gb|WP_003807115) (bplA) probable oxidoreductase [LPS (VF0033) - Immune modulation (VFC02
VFG000934(gb|WP_000348393) (entA) 2,3-dihydro-2,3-dihydroxybenzoate dehydrogenase EntA [Enterobactin

VFG038865(gb|WP_005314289) (fliG) flagellar motor switch protein G [Lateral flagella (VF0474) - Motility (VF
VFG002182(gb|WP_002376666) (cpsI) UDP-galactopyranose mutase [Capsule (VF0361) - Immune modulation
VFG048419(gb|WP_004210873) (entB) 2,3-dihydro-2,3-dihydroxybenzoate synthetase, isochroismatase [Ent (VF
VFG049018(gb|WP_002913007) (rcsB) transcriptional regulator RcsB [RcsAB (VF0571) - Regulation (VFC030
VFG002508(gb|WP_004525899) (flgB) flagellar basal-body rod protein FlgB [Flagella (VF0430) - Motility (VFC
VFG000198(gb|NP_250394) (pcrD) type III secretion system protein PcrD [TTSS (VF0083) - Effector delivery s
VFG038375(gb|WP_011705710) (atsC) Type VI secretion system protein [T6SS (VF0480) - Effector delivery sy
VFG002521(gb|WP_004525693) (flhF) flagellar biosynthesis regulator FlhF [Flagella (VF0430) - Motility (VFC
VFG000266(gb|WP_002222141) (hmbR) hemoglobin receptor [HmbR (VF0048) - Nutritional/Metabolic factor (
VFG000938(gb|WP_001015709) (iucC) aerobactin siderophore biosynthesis protein IucC [Aerobactin (VF0229)
VFG041019(gb|NP_250352) (PA1661) hypothetical protein [HSI-2 (VF0943) - Effector delivery system (VFC00
VFG038828(gb|WP_011706649) (flgN) flagellar protein FlgN [Polar flagella (VF0473) - Motility (VFC0204)] [
VFG037414(gb|WP_012391956) (bauD) ferric siderophore ABC transporter, permease protein BauD [Acinetoba
VFG048439(gb|WP_014907941) (entC) isochorismate synthase [Ent (VF0562) - Nutritional/Metabolic factor (V
VFG048808(gb|WP_004899416) (rfbK1) O9 family phosphomannomutase RfbK1 [Capsule (VF0560) - Immune
VFG000862(gb|WP_014639426) (pet) Pet serine protease precursor, autotransporter [Pet (VF0217) - Effector del
VFG034849(gb|WP_000129338) (espX1) Type III secretion system effector EspX1 [TTSS secreted effectors (VF
VFG045530(gb|WP_010947381) (LPG_RS08285) Dot/Icm type IV secretion system effector [Dot/Icm T4SS sec
VFG000906(gb|WP_001142375) (hlyA) Hemolysin A [-Hemolysin (VF0225) - Exotoxin (VFC0235)] [Escherich
VFG001239(gb|NP_249772) (flgF) flagellar basal-body rod protein FlgF [Flagella (VF0273) - Motility (VFC020
VFG050567(gb|WP_000768964.1) (pilY1) VWA domain-containing protein [TFP (VF1334) - Adherence (VFC0
VFG000834(gb|WP_000823897) (rorf1) SdiA-regulated domain-containing protein [TTSS (VF0191) - Effector d
VFG010745(gb|WP_010946125) (sdhA) Dot/Icm type IV secretion system effector SdhA [Dot/Icm T4SS secrete
VFG044469(gb|WP_011106485) (VP_RS21635) putative type III secretion system protein [T3SS2 (VF0409) - E
VFG049160(gb|WP_001327259) (clbN) colibactin non-ribosomal peptide synthetase ClbN [Colibactin (VF0573)
VFG039106(gb|NP_820604) (icmB/dotO) Type IVB secretion system protein IcmB [Dot/Icm T4SS (VF0503) - 
VFG050161(gb|WP_000579525) (kpsC) Capsule polysaccharide export protein [K1 capsule (VF0239) - Invasion
VFG038452(gb|WP_016351631) (tapO) type IV pilus biogenesis protein PilO [Tap type IV pili (VF0475) - Adhe
VFG041018(gb|NP_250351) (PA1660) hypothetical protein [HSI-2 (VF0943) - Effector delivery system (VFC00
VFG038365(gb|WP_011705044) (hcp1) HcpA-like protein [T6SS (VF0480) - Effector delivery system (VFC008
VFG045520(gb|WP_010947043) (legC1) Dot/Icm type IV secretion system effector LegC1 [Dot/Icm T4SS secre
VFG049159(gb|WP_014907260) (clbM) precolibactin export MATE transporter ClbM [Colibactin (VF0573) - E
VFG045626(gb|WP_014748741) (scm) collagen adhesin protein Scm [Scm (VF0418) - Adherence (VFC0001)] [
VFG041043(gb|NP_251061) (PA2371) ClpA/B-type protease [HSI-3 (VF0944) - Effector delivery system (VFC
VFG048697(gb|WP_015874721) (clpV/tssH) type VI secretion system ATPase TssH [T6SS (VF0569) - Effector
VFG044081(gb|NP_250946) (pvcC) paerucumarin biosynthesis protein PvcC [Pyoverdine (VF0094) - Nutritiona
VFG050470(gb|WP_000279216.1) (pilC) type II secretion system F family protein [TFP (VF1334) - Adherence (
VFG035909(gb|WP_001216023) (aatA) outer membrane protein AatA [Dispersin (VF0215) - Others (VFC0346)
VFG001916(gb|YP_002344282) (flhA) flagellar biosynthesis protein FlhA [Flagella (VF0114) - Motility (VFC0
VFG038144(gb|WP_001062908) (galE) UDP-glucose 4-epimerase GalE [Capsule (VF0465) - Immune modulatio
VFG049158(gb|WP_001297937) (clbL) colibactin biosynthesis amidase ClbL [Colibactin (VF0573) - Exotoxin (
VFG000922(gb|WP_000910402) (chuU) heme permease protein ChuU [Chu (VF0227) - Nutritional/Metabolic fa
VFG049917(gb|WP_000508696) (rhs/PAAR) Type VI secretion system protein, PAAR family [T6SS (VF0579) -
VFG001238(gb|NP_249771) (flgE) flagellar hook protein FlgE [Flagella (VF0273) - Motility (VFC0204)] [Pseu
VFG012501(gb|WP_000271272) (iroE) esterase [Salmochelin siderophore (VF0230) - Nutritional/Metabolic fact
VFG050162(gb|WP_001161829) (kpsS) Capsule polysaccharide export protein [K1 capsule (VF0239) - Invasion

VFG001257(gb|NP_250135) (fliN) flagellar motor switch protein FliN [Flagella (VF0273) - Motility (VFC0204)
VFG002443(gb|WP_004528812) (bopE) Type III secretion system secreted effector, G-nucleotide exchange facto
VFG038357(gb|WP_005320736) (sycX) hypothetical protein [T3SS (VF0479) - Effector delivery system (VFC0
VFG002069(gb|NP_248773) (hsiB1/vipA/tssB) type VI secretion system tubule-forming protein VipA [HSI-I (V
VFG000341(gb|WP_005694040) (hxuB) heme-hemopexin utilization protein B HxuB, two-partner secretion (TP
VFG002491(gb|WP_004524547) (fliM) flagellar motor switch protein FliM [Flagella (VF0430) - Motility (VFC
VFG002406(gb|WP_000728972) (esaA) type VII secretion system protein EsaA [Type VII secretion system (VF
VFG000816(gb|WP_001302733) (escI) Type III secretion system inner rod component EscI [TTSS (VF0191) - E
VFG044329(gb|WP_015874964) (irp1) yersiniabactin polyketide synthase HMWP1 [Ybt (VF0564) - Nutritional/
VFG038861(gb|WP_005314296) (lfhA) lateral flagellar biosynthesis protein [Lateral flagella (VF0474) - Motility
VFG000871(gb|WP_000790574) (fimB) Type 1 fimbriae Regulatory protein fimB [Type 1 fimbriae (VF0221) - A
VFG000615(gb|NP_709454) (iucA) aerobactin synthesis protein IucA [Aerobactin (VF0123) - Nutritional/Metab
VFG050757(gb|WP_001166319.1) (gspE1) general secretion pathway protein E [T2SS (VF1335) - Effector deliv
VFG001249(gb|NP_249790) (fleR) two-component response regulator [Flagella (VF0273) - Motility (VFC0204)
VFG049911(gb|WP_000393837) (tssF) Type VI secretion system protein TssF [T6SS (VF0579) - Effector delive
VFG038753(gb|WP_024944879) (motY) sodium-type flagellar protein MotY [Polar flagella (VF0473) - Motility
VFG050655(gb|WP_000729757.1) (pilI) chemotaxis protein CheW [TFP (VF1334) - Adherence (VFC0001)] [A
VFG000423(gb|WP_000052242) (vexE) Vi polysaccharide export protein VexE [Vi antigen (VF0101) - Immune
VFG049163(gb|WP_000065646) (clbQ) colibactin biosynthesis thioesterase ClbQ  [Colibactin (VF0573) - Exoto
VFG038872(gb|WP_005314272) (lfgB) lateral flagellar basal body protein, LfgB [Lateral flagella (VF0474) - Mo
VFG042739(gb|NP_254019) (crc) catabolite repression control protein [Type IV pili (VF0082) - Adherence (VF
VFG002191(gb|WP_002369252) (sprE) serine proteinase, V8 family [SprE (VF0358) - Exoenzyme (VFC0251)]
VFG002300(gb|CAA88565) (tcnA) alpha-toxin [Alpha-toxin (novyi) (VF0386) - Exotoxin (VFC0235)] [Clostrid
VFG000945(gb|AAK16478) (draD) DraD, periplasmic protein [Dr adhesins (VF0223) - Adherence (VFC0001)]
VFG038387(gb|WP_011705717) (atsJ) Type VI secretion system protein [T6SS (VF0480) - Effector delivery sys
VFG048819(gb|WP_014907234) (KP1_RS17280) mannose-1-phosphate guanylyltransferase/mannose-6-phospha
VFG034859(gb|WP_010904544) (espX2) Type III secretion system effector EspX2 [TTSS secreted effectors (V
VFG014148(gb|NP_249768) (flgB) flagellar basal body rod protein FlgB [Flagella (VF0273) - Motility (VFC020
VFG000925(gb|WP_000140634) (fepC) ferrienterobactin ABC transporter ATPase [Enterobactin (VF0228) - Nu
VFG038342(gb|WP_005320770) (ascV) Type III secretion system inner membrane export apparatus protein Asc
VFG002171(gb|WP_011109430) (EF0149) aggregation substance [AS (VF0352) - Adherence (VFC0001)] [Ente
VFG002534(gb|WP_004198197) (motA) flagellar motor protein MotA [Flagella (VF0430) - Motility (VFC0204)
VFG048771(gb|WP_004190505) (tssF) type VI secretion system baseplate subunit TssF [T6SS (VF0569) - Effec
VFG038820(gb|WP_161785096) (flgA) flagella basal body P-ring formation protein FlgA [Polar flagella (VF047
VFG007550(gb|WP_000985860) (cheB) chemotaxis-specific methylesterase [Flagella (VF0519) - Motility (VFC
VFG011515(gb|WP_004683743) (wbdA) mannosyltransferase [LPS (VF0367) - Immune modulation (VFC0258)
VFG007823(gb|NP_217447) (ppsA) phthiocerol synthesis polyketide synthase type I PpsA [PDIM (VF0309) - In
VFG048419(gb|WP_004210873) (entB) 2,3-dihydro-2,3-dihydroxybenzoate synthetase, isochroismatase [Ent (V
VFG000933(gb|WP_001007140) (entB) isochorismatase [Enterobactin (VF0228) - Nutritional/Metabolic factor (
VFG002416(gb|WP_001265657) (yagW/ecpD) polymerized tip adhesin of ECP fibers [ECP (VF0404) - Adheren
VFG000916(gb|WP_001017192) (chuS) heme oxygenase ChuS [Chu (VF0227) - Nutritional/Metabolic factor (V
VFG000932(gb|WP_000026784) (entE) 2,3-dihydroxybenzoate-AMP ligase component of enterobactin synthase
VFG050428(gb|WP_000128073.1) (fimV) hypothetical protein [TFP (VF1334) - Adherence (VFC0001)] [Acine
VFG038671(gb|WP_011705282) (fliQ) flagellar biosynthetic protein FliQ [Polar flagella (VF0473) - Motility (V
VFG034908(gb|WP_000978349) (espY3) Type III secretion system effector EspY3 [TTSS secreted effectors (V
VFG038501(gb|WP_043119142) (tapV) type IV pilus assembly protein TapV [Tap type IV pili (VF0475) - Adhe

VFG000212(gb|NP_250408) (pscD) type III secretion system basal body protein PscD [TTSS (VF0083) - Effecto
VFG000929(gb|WP_001375021) (entD) phosphopantetheinyl transferase component of enterobactin synthase mu
VFG046118(gb|WP_011205945) (mhp107) P97 paralog, surface adhesin [P97/P102 paralog family (VF0426) - A
VFG000873(gb|WP_000695571) (fimA) Type-1 fimbrial protein, A chain precursor [Type 1 fimbriae (VF0221) -
VFG048528(gb|WP_004183335) (entD) enterochelin synthetase component D [Ent (VF0562) - Nutritional/Metab
VFG051042(gb|WP_000653195.1) (hcp/tssD) type VI secretion system tube protein Hcp [T6SS (VF1337) - Effec
VFG002180(gb|WP_002362627) (cpsK) ABC transporter, permease protein [Capsule (VF0361) - Immune modul
VFG000930(gb|WP_000077767) (entF) enterobactin synthase multienzyme complex component, ATP-dependent
VFG002277(gb|WP_011009663) (nagH) hyaluronidase [Mu-toxin (VF0389) - Exoenzyme (VFC0251)] [Clostrid
VFG049157(gb|WP_014907259) (clbK) colibactin hybrid non-ribosomal peptide synthetase/type I polyketide syn
VFG002047(gb|WP_000498818) (gspD) general secretion pathway protein D [T2SS (VF0333) - Effector delivery
VFG044332(gb|WP_015874967) (ybtE) yersiniabactin biosynthesis salycil-AMP ligase YbtE [Ybt (VF0564) - N
VFG037260(gb|WP_001095752) (barB) siderophore efflux system of the ABC superfamily [Acinetobactin (VF0
VFG045818(gb|WP_001528154) (tsh) temperature-sensitive protein Tsh [Tsh (VF0233) - Effector delivery system
VFG004931(gb|WP_000669009) (set16) superantigen-like protein SSL1 [SSLs (VF0990) - Exotoxin (VFC0235)
VFG038659(gb|WP_011705280) (fliN) flagellar motor switch protein FliN [Polar flagella (VF0473) - Motility (V
VFG049148(gb|WP_041168941) (clbB) colibactin hybrid non-ribosomal peptide synthetase/type I polyketide syn
VFG038541(gb|WP_016350150) (flpD) Flp pilus assembly protein FlpD [Flp type IV pili (VF0476) - Adherence
VFG002051(gb|WP_001115126) (gspH) general secretion pathway protein H [T2SS (VF0333) - Effector delivery
VFG000363(gb|WP_002212775) (irp2) yersiniabactin biosynthetic protein Irp2 [Yersiniabactin (VF0136) - Nutri
VFG043209(gb|WP_005168769) (cheD) methyl-accepting chemotaxis protein CheD [Flagella (VF0394) - Motili
VFG040545(gb|WP_011190491) (sca1) cell surface antigen Sca1 [Sca1 (VF0498) - Adherence (VFC0001)] [Ric
VFG038683(gb|WP_016350056) (flhA) flagellar biosynthesis protein FlhA [Polar flagella (VF0473) - Motility (V
VFG010749(gb|WP_010945896) (sdhB) Dot/Icm type IV secretion system effector SdhB [Dot/Icm T4SS secrete
VFG000934(gb|WP_000348393) (entA) 2,3-dihydro-2,3-dihydroxybenzoate dehydrogenase EntA [Enterobactin
VFG038365(gb|WP_011705044) (hcp1) HcpA-like protein [T6SS (VF0480) - Effector delivery system (VFC008
VFG012505(gb|WP_000933673) (iroD) esterase [Salmochelin siderophore (VF0230) - Nutritional/Metabolic fac
VFG000925(gb|WP_000140634) (fepC) ferrienterobactin ABC transporter ATPase [Enterobactin (VF0228) - Nu
VFG000540(gb|NP_461799) (sptP) type III secretion system effector SptP, tyrosine phosphatase and GTPase-act
VFG038401(gb|WP_011705724) (atsQ) Type VI secretion system protein [T6SS (VF0480) - Effector delivery sy
VFG001224(gb|NP_249087) (pilU) twitching motility protein PilU [Type IV pili (VF0082) - Adherence (VFC00
VFG010523(gb|WP_011213649) (LPG_RS06765) saccharopine dehydrogenase NADP-binding domain-containi
VFG041041(gb|NP_251059) (PA2369) hypothetical protein [HSI-3 (VF0944) - Effector delivery system (VFC00
VFG049906 (tssK) Type VI secretion system protein TssK [T6SS (VF0579) - Effector delivery system (VFC008
VFG038590(gb|WP_043118275) (mshE) ATPase protein MshE, GspE-like [MSHA type IV pili (VF0477) - Adh
VFG000365(gb|WP_002214412) (ybtP) yersiniabactin ABC transporter ATP-binding/permease protein YbtP [Ye
VFG000165(gb|NP_252915) (pchF) pyochelin synthetase PchF [Pyochelin (VF0095) - Nutritional/Metabolic fact
VFG000130(gb|NP_252238) (algI) alginate o-acetyltransferase AlgI [Alginate (VF0091) - Biofilm (VFC0271)] [
VFG002045(gb|WP_010930608) (bvgS) virulence factors two-component system sensor histidine kinase BvgS [
VFG016770(gb|WP_005767766) (flgI) flagellar P-ring protein FlgI [Flagella (VF0373) - Motility (VFC0204)] [
VFG002054(gb|WP_000633233) (gspK) general secretion pathway protein K [T2SS (VF0333) - Effector delivery
VFG049163(gb|WP_000065646) (clbQ) colibactin biosynthesis thioesterase ClbQ [Colibactin (VF0573) - Exoto
VFG048808(gb|WP_004899416) (rfbK1) O9 family phosphomannomutase RfbK1 [Capsule (VF0560) - Immune
VFG005215(gb|WP_000743858) (fbsB) Fibrinogen-binding surface protein B [FbsB (VF0523) - Adherence (VF
VFG048654(gb|WP_004224345) (vasE/tssK) type VI secretion system baseplate subunit TssK [T6SS (VF0569) -
VFG034648(gb|WP_001245988) (cfaD/cfaE) minor pilin and initiator [Adhesive fimbriae (VF0213) - Adherence

VFG000914(gb|WP_000767894) (focG) F1C minor fimbrial subunit protein G presursor [F1C fimbriae (VF0224
VFG001386(gb|NP_215271) (phoP) Possible two component system response transcriptional positive regulator P
VFG013661(gb|WP_011988426) (pilO) type IV pilus inner membrane platform protein PilO [Type IV pili (VF04
VFG001257(gb|NP_250135) (fliN) flagellar motor switch protein FliN [Flagella (VF0273) - Motility (VFC0204)
VFG016362(gb|WP_000959117) (hal) heme-acquisition leucine-rich repeat protein [Hal (VF0587) - Nutritional/M
VFG050670(gb|WP_000505931.1) (pilJ) methyl-accepting chemotaxis protein [TFP (VF1334) - Adherence (VFC
VFG002415(gb|WP_001131063) (yagX/ecpC) E. coli common pilus usher EcpC [ECP (VF0404) - Adherence (V
VFG042736(gb|NP_253152) (rpoN) RNA polymerase factor sigma-54 [Type IV pili (VF0082) - Adherence (VFC
VFG038913 (rtxA) multifunctional autoprocessing repeats-in-toxins (MARTX) toxin [RtxA (VF0482) - Exotoxin
VFG000919(gb|WP_001095619) (chuW) Putative oxygen independent coproporphyrinogen III oxidase [Chu (VF
VFG018682(gb|WP_005491021) (vopL) type III secretion system effector VopL, actin filament nucleation activi
VFG034849(gb|WP_000129338) (espX1) Type III secretion system effector EspX1 [TTSS secreted effectors (VF
VFG049902(gb|WP_011310140) (tssM) Type VI secretion system protein TssM [T6SS (VF0579) - Effector deliv
VFG000164(gb|NP_252911) (fptA) Fe(III)-pyochelin receptor precursor [Pyochelin (VF0095) - Nutritional/Meta
VFG001229(gb|NP_249103) (pilK) methyltransferase PilK [Type IV pili (VF0082) - Adherence (VFC0001)] [Ps
VFG002434(gb|WP_011205650) (pilN) type IV pilus inner membrane platform protein PilN [Type IV pili (VF04
VFG000913(gb|WP_000237770) (focF) F1C minor fimbrial subunit F precursor [F1C fimbriae (VF0224) - Adhe
VFG049911(gb|WP_000393837) (tssF) Type VI secretion system protein TssF [T6SS (VF0579) - Effector delive
VFG044333(gb|WP_015874968) (fyuA) yersiniabactin receptor FyuA [Ybt (VF0564) - Nutritional/Metabolic fac
VFG050499(gb|WP_000755268.1) (tsaP) LysM peptidoglycan-binding domain-containing protein [TFP (VF1334
VFG041039(gb|NP_251057) (PA2367) hypothetical protein [HSI-3 (VF0944) - Effector delivery system (VFC00
VFG034612(gb|WP_000225871) (cfaA) colonisation factor antigen a, chaperone [Adhesive fimbriae (VF0213) -
VFG050164(gb|WP_000723250) (neuC) UDP-N-acetylglucosamine 2-epimerase [K1 capsule (VF0239) - Invasio
VFG040222(gb|WP_011190745) (rvhB8b) VirB8-like protein of the Type IV secretion system [Rvh T4SS (VF04
VFG048247(gb|WP_002916190) (fimA) type 1 major fimbrial subunit precursor [Type I fimbriae (VF0566) - Ad
VFG038472(gb|WP_011706550) (tapW) twitching ATPase [Tap type IV pili (VF0475) - Adherence (VFC0001)]
VFG000229 (pilS) pilin [Type IV pili (VF0075) - Adherence (VFC0001)] [Neisseria meningitidis MC58]
VFG007991(gb|YP_178020) (papA2) trehalose-2-sulfate acyltransferase [Sulfolipid-1 biosynthesis and transport
VFG007857(gb|NP_217450) (ppsD) phthiocerol synthesis polyketide synthase type I PpsD [PDIM (VF0309) - In
VFG038389(gb|WP_011705718) (atsK) Type VI secretion system protein [T6SS (VF0480) - Effector delivery sy
VFG039216(gb|NP_820634) (icmX) Type IVB secretion system protein IcmX [Dot/Icm T4SS (VF0503) - Effect
VFG038796(gb|WP_016351321) (flgE) flagellar hook protein FlgE [Polar flagella (VF0473) - Motility (VFC020
VFG038655(gb|WP_011705279) (fliM) flagellar motor switch protein FliM [Polar flagella (VF0473) - Motility (
VFG006218(gb|WP_079760670) (opa) outer membrane beta-barrel protein [Opa (VF0076) - Invasion (VFC0083
VFG000423(gb|WP_000052242) (vexE) Vi polysaccharide export protein VexE [Vi antigen (VF0101) - Immune
VFG050787(gb|WP_001112089.1) (gspF) general secretion pathway protein F [T2SS (VF1335) - Effector delive
VFG034795 (espN) Type III secretion system effector EspN [TTSS secreted effectors (VF1110) - Effector delive
VFG002389(gb|NP_218396) (espK) ESX-1 type VII secretion system EspK [ESX-1 (VF0315) - Effector delivery
VFG001256(gb|NP_250134) (fliM) flagellar motor switch protein FliM [Flagella (VF0273) - Motility (VFC0204
VFG034679(gb|WP_000074207) (ibeB) Cu(+)/Ag(+) efflux RND transporter outer membrane channel CusC [Ibe
VFG038611(gb|WP_016349153) (mshI1) MSHA (mannose-sensitive hemagglutinin) pili biogenesis protein Msh
VFG049144(gb|WP_002892069) (acrB) acriflavine resistance protein B [AcrAB (VF0568) - Antimicrobial activi
VFG001259(gb|NP_250137) (fliP) flagellar biosynthetic protein FliP [Flagella (VF0273) - Motility (VFC0204)] [
VFG013260(gb|WP_010868968) (lgtC) glycosyltransferase [LOS (VF0044) - Immune modulation (VFC0258)] [
VFG012865 (ipaH) invasion plasmid antigen IpaH [TTSS secreted effectors (VF0978) - Effector delivery system
VFG001391(gb|NP_215677) (narG) nitrate reductase subunit alpha [Nitrate reductase (VF0302) - Nutritional/Me

VFG001297 (capA) capsular polysaccharide type 5/8 biosynthesis protein CapA [Capsule (VF0003) - Immune m
VFG049164(gb|WP_004175305) (clbS) colibactin self-protection protein ClbS [Colibactin (VF0573) - Exotoxin
VFG000871(gb|WP_000790574) (fimB) Type 1 fimbriae Regulatory protein fimB [Type 1 fimbriae (VF0221) -
VFG046680(gb|WP_003020700) (FTT_RS04120) ABC transporter ATP-binding protein [Capsule (VF0543) - In
VFG001405(gb|NP_217219) (sigA/rpoV) RNA polymerase sigma factor SigA [SigA (VF0257) - Regulation (VF
VFG016058(gb|NP_251114) (pvdL) peptide synthase PvdL [Pyoverdine (VF0094) - Nutritional/Metabolic factor
VFG038373(gb|WP_011705708) (atsA) Type VI secretion system protein [T6SS (VF0480) - Effector delivery sy
VFG017846(gb|WP_011988432) (pilR) type IV pilus biosynthesis protein PilR [Type IV pili (VF0401) - Adherer
VFG038635(gb|WP_016350051) (fliH) flagellar assembly protein FliH [Polar flagella (VF0473) - Motility (VFC
VFG000312(gb|WP_000542820) (sabA/hopP) outer membrane protein adhesin [SabA (VF0055) - Adherence (V
VFG000879(gb|WP_000832235) (fimH) FimH protein precursor [Type 1 fimbriae (VF0221) - Adherence (VFC0
VFG001938(gb|YP_002344530) (Cj1137c) glycosyltransferase [LOS (VF0326) - Immune modulation (VFC025
VFG038889(gb|WP_005314215) (lafT) LafT, flagella motor stator protein [Lateral flagella (VF0474) - Motility (
VFG000111(gb|NP_253215) (pilA) type 4 fimbrial precursor PilA [Type IV pili (VF0082) - Adherence (VFC000
VFG001985(gb|YP_002344650) (porA) major outer membrane protein [MOMP (VF0328) - Adherence (VFC000
VFG001940(gb|YP_002344532) (wlaN) beta-1,3 galactosyltransferase [LOS (VF0326) - Immune modulation (V
VFG013503(gb|WP_005694190) (lsgA) lipopolysaccharide biosynthesis transporter [LOS (VF0044) - Immune m
VFG001248(gb|NP_249788) (fleQ) transcriptional regulator FleQ [Flagella (VF0273) - Motility (VFC0204)] [Ps
VFG000935(gb|WP_001222189) (iroN) salmochelin receptor IroN [Salmochelin siderophore (VF0230) - Nutritio
VFG043625(gb|WP_000813718) (mrkC) fimbrial biogenesis outer membrane usher protein mrkC precursor [Typ
VFG050372(gb|WP_001017033.1) (pilQ) type IV pilus secretin PilQ [TFP (VF1334) - Adherence (VFC0001)] [
VFG048002(gb|WP_003022141) (iglJ) Type VI secretion system protein IglJ [T6SS (VF0544) - Effector delivery
VFG048687(gb|WP_002902160) (hcp/tssD) type VI secretion system protein, Hcp family [T6SS (VF0569) - Eff
VFG037882(gb|WP_000872593) (ACICU_RS00405) polysaccharide biosynthesis/export family protein [Capsul
VFG048951(gb|WP_015875028) (KP1_RS17330) polysaccharide biosynthesis tyrosine autokinase [Capsule (VF
VFG000352(gb|WP_002208793) (psaB) chaperone protein PsaB [Myf/pH6 antigen (VF0134) - Adherence (VFC
VFG043623(gb|WP_002916122) (mrkF) type 3 fimbrial minor pilin subunit MrkF [Type 3 fimbriae (VF0567) -
VFG007370(gb|WP_000145786) (cheV) chemotaxis protein CheV [Flagella (VF0519) - Motility (VFC0204)] [V
VFG001375(gb|WP_000594468) (cps4K) capsular polysaccharide biosynthesis protein Cps4K [Capsule (VF014
VFG001960(gb|YP_002344815) (Cj1432c) sugar transferase [Capsule (VF0323) - Immune modulation (VFC025
VFG012597(gb|WP_000089593) (chuA) Outer membrane heme/hemoglobin receptor ChuA [Chu (VF0234) - Nu
VFG000885(gb|WP_000265729) (papD) chaperone protein PapD [P fimbriae (VF0220) - Adherence (VFC0001)
VFG038756(gb|WP_011706626) (flrC) transcriptional activator [Polar flagella (VF0473) - Motility (VFC0204)]
VFG006199(gb|WP_010980842) (pilT2) twitching motility protein PilT [Type IV pili (VF0075) - Adherence (V
VFG000917(gb|WP_000089583) (chuA) Outer membrane heme/hemoglobin receptor ChuA [Chu (VF0227) - Nu
VFG005957(gb|WP_001193768) (wzy) O-antigen polysaccharide polymerase Wzy [Capsule (VF0144) - Immune
VFG050157(gb|WP_001331698) (kpsF) Polysialic acid capsule expression protein [K1 capsule (VF0239) - Invas
VFG038413(gb|WP_043118350) (fimC) outer membrane usher protein [Type I pili (VF0486) - Adherence (VFC
VFG042857(gb|WP_003015347) (pilO) Type IV pilus glycosylation protein [Type IV pili (VF0458) - Adherence
VFG000001(gb|WP_010930610) (fhaB) filamentous hemagglutinin/adhesin [FHA (VF0025) - Adherence (VFC
VFG005781(gb|WP_000650746) (cylE) haemolysin CylE [-haemolysin/cytolysin (VF0279) - Exotoxin (VFC023
VFG000927(gb|WP_000096764) (fepE) LPS O-antigen length regulator [Enterobactin (VF0228) - Nutritional/M
VFG000361(gb|WP_000982866) (ybtU) yersiniabactin biosynthetic protein YbtU [Yersiniabactin (VF0136) - Nu
VFG002492(gb|WP_004184995) (fliN) flagellar motor switch protein FliN [Flagella (VF0430) - Motility (VFC0
VFG038105(gb|WP_000591436) (galU) UTP--glucose-1-phosphate uridylyltransferase GalU [Capsule (VF0465)
VFG038545(gb|WP_043118798) (flpC) Flp pilus secretin FlpC [Flp type IV pili (VF0476) - Adherence (VFC000

VFG000333(gb|WP_005694147) (lic2A) glycosyltransferase [LOS (VF0044) - Immune modulation (VFC0258)]
VFG038707(gb|WP_016350059) (cheA-2) chemotaxis protein CheA [Polar flagella (VF0473) - Motility (VFC02
VFG050214(gb|WP_011860844) (cwpV) hemagglutinin/adhesin [CwpV (VF0596) - Adherence (VFC0001)] [Cl
VFG000926(gb|WP_001443194) (fepD) ferrienterobactin ABC transporter permease [Enterobactin (VF0228) - N
VFG049161(gb|WP_001029878) (clbO) colibactin polyketide synthase ClbO [Colibactin (VF0573) - Exotoxin (V
VFG049007(gb|WP_002911561) (rcsA) transcriptional activator for ctr capsule biosynthesis [RcsAB (VF0571) -
VFG002300(gb|CAA88565) (tcnA) alpha-toxin [Alpha-toxin (novyi) (VF0386) - Exotoxin (VFC0235)] [Clostrid
VFG038549(gb|WP_016350148) (flpB) Flp pilus assembly protein [Flp type IV pili (VF0476) - Adherence (VFC
VFG000887(gb|WP_000597713) (papK) P pilus minor subunit PapK [P fimbriae (VF0220) - Adherence (VFC00
VFG003512(gb|WP_011817126) (yspD) IpaD/SipD/SspD family type III secretion system needle tip protein [Ys
VFG048478(gb|WP_004147521) (fepG) iron-enterobactin transporter permease [Ent (VF0562) - Nutritional/Met
VFG049900 (tssM) Type VI secretion system protein TssM [T6SS (VF0579) - Effector delivery system (VFC008
VFG042977(gb|WP_002379309) (ebpB) endocarditis and biofilm-associated pilus minor subunit EbpB [Ebp pili
VFG043773(gb|YP_398516) (bont) botulinum neurotoxin, BoNT [BoNT (VF0375) - Exotoxin (VFC0235)] [Clo
VFG001291(gb|WP_001039417) (cna) collagen adhesin precursor [CNA (VF0005) - Adherence (VFC0001)] [St
VFG038851(gb|WP_016350359) (nueB) pseudaminic acid synthase, NeuB [Polar flagella (VF0473) - Motility (V
VFG002536(gb|WP_004530401) (fliD) flagellar capping protein FliD [Flagella (VF0430) - Motility (VFC0204)]
VFG037176(gb|WP_001081735) (plc1) phospholipase C [Phospholipase C (VF0470) - Exotoxin (VFC0235)] [A
VFG041013(gb|NP_250203) (hcpA) secreted protein Hcp [HSI-2 (VF0943) - Effector delivery system (VFC008
VFG038784(gb|WP_011706638) (flgH) flagellar basal body L-ring protein [Polar flagella (VF0473) - Motility (V
VFG001241(gb|NP_249774) (flgH) flagellar L-ring protein precursor FlgH [Flagella (VF0273) - Motility (VFC0
VFG048863(gb|WP_014907229) (KP1_RS17300) GDP-mannose mannosyl hydrolase [Capsule (VF0560) - Imm
VFG050159(gb|WP_001331697) (kpsD) Polysialic acid transport protein precursor [K1 capsule (VF0239) - Inva
VFG000826(gb|WP_000379425) (escS) Type III secretion systemminor export apparatus protein EscS [TTSS (V
VFG000358(gb|WP_002212885) (fyuA/psn) pesticin/yersiniabactin receptor protein [Yersiniabactin (VF0136) -
VFG000939(gb|WP_001287497) (iucB) aerobactin siderophore biosynthesis protein IucB [Aerobactin (VF0229)
VFG000906(gb|WP_001142375) (hlyA) Hemolysin A [-Hemolysin (VF0225) - Exotoxin (VFC0235)] [Escherich
VFG000127(gb|NP_252235) (algG) outer membrane protein AlgE [Alginate (VF0091) - Biofilm (VFC0271)] [Ps
VFG043624(gb|WP_004149659) (mrkD) fimbrial adhesin protein precursor MrkD [Type 3 fimbriae (VF0567) -
VFG000862(gb|WP_014639426) (pet) Pet serine protease precursor, autotransporter [Pet (VF0217) - Effector del
VFG001408(gb|NP_217456) (mas) multifunctional mycocerosic acid synthase [PDIM (VF0309) - Immune modu
VFG000360(gb|WP_002212799) (ybtT) type II thioesterase YbtT [Yersiniabactin (VF0136) - Nutritional/Metabo
VFG038537(gb|WP_016350151) (flpE) Flp pilus assembly protein, ATPase [Flp type IV pili (VF0476) - Adheren
VFG000877(gb|WP_001244821) (fimF) FimF protein precursor [Type 1 fimbriae (VF0221) - Adherence (VFC0
VFG050386(gb|WP_000355489.1) (pilT) type IV pilus twitching motility protein PilT [TFP (VF1334) - Adheren
VFG038553(gb|WP_016350147) (flpA) prepilin peptidase [Flp type IV pili (VF0476) - Adherence (VFC0001)] [
VFG002069(gb|NP_248773) (hsiB1/vipA/tssB) type VI secretion system tubule-forming protein VipA [HSI-I (V
VFG048620(gb|WP_014907843) (iutA) ferric aerobactin receptor IutA [Aerobactin (VF0565) - Nutritional/Meta
VFG000739(gb|AAC38392) (eae) intimin [Intimin (VF0177) - Effector delivery system (VFC0086)] [Escherichi
VFG001279(gb|WP_001060526) (sdrC) Ser-Asp rich fibrinogen-binding bone sialoprotein-binding protein [SDr
VFG000878(gb|WP_000872015) (fimG) FimG protein precursor [Type 1 fimbriae (VF0221) - Adherence (VFC0
VFG039912(gb|WP_011191202) (rvhB4b) type IV secretion system protein VirB4 precursor [Rvh T4SS (VF048
VFG000874(gb|WP_000824100) (fimI) Fimbrin-like protein fimI precursor [Type 1 fimbriae (VF0221) - Adhere
VFG038687(gb|WP_016350057) (flhF) flagellar biosynthesis protein FlhF [Polar flagella (VF0473) - Motility (V
VFG002413(gb|WP_000389022) (ykgK/ecpR) regulator protein EcpR [ECP (VF0404) - Adherence (VFC0001)]
VFG044323(gb|WP_015874960) (ybtS) yersiniabactin biosynthesis salicylate synthase YbtS [Ybt (VF0564) - Nu

VFG001231(gb|NP_249104) (chpA) still frameshift probable component of chemotactic signal transduction syste
VFG038405(gb|WP_011705727) (AHA_RS09305) type VI secretion system PAAR protein [T6SS (VF0480) - E
VFG000465(gb|NP_460064) (sopB/sigD) type III secretion system effector SopB, phosphoinositide phosphatase
VFG000036(gb|WP_010929614) (bplC) lipopolysaccharide biosynthesis protein [LPS (VF0033) - Immune modu
VFG044328(gb|WP_015874963) (irp2) yersiniabactin non-ribosomal peptide synthetase HMWP2 [Ybt (VF0564)
VFG002412(gb|WP_000716386) (yagY/ecpB) E. coli common pilus chaperone EcpB [ECP (VF0404) - Adheren
VFG000168(gb|NP_252918) (pchD) pyochelin biosynthesis protein PchD [Pyochelin (VF0095) - Nutritional/Me
VFG038297(gb|WP_016349341) (exeJ) general secretion pathway protein J [Exe T2SS (VF0478) - Effector deli
VFG001282(gb|WP_000841389) (fnbB) fibronectin-binding protein B [FnBPs (VF0010) - Adherence (VFC0001
VFG000922(gb|WP_000910402) (chuU) heme permease protein ChuU [Chu (VF0227) - Nutritional/Metabolic f
VFG040665(gb|WP_011190939) (sca4) cell surface antigen Sca4 [Sca4 (VF0502) - Others (VFC0346)] [Ricketts
VFG034866(gb|WP_000900547) (espX4) Type III secretion system effector EspX4 [TTSS secreted effectors (V
VFG041011(gb|NP_248785) (vgrG1b) type VI secretion system substrate VgrG1b [HSI-I (VF0334) - Effector de
VFG000315(gb|WP_000694809) (rfbM) mannose-6-phosphate isomerase [LPS (VF0056) - Immune modulation
VFG038892(gb|WP_043118302) (aerA/act) Aerolysin A [Aerolysin (VF0481) - Exotoxin (VFC0235)] [Aeromo
VFG038679(gb|WP_016350055) (flhB) flagellar biosynthetic protein FlhB [Polar flagella (VF0473) - Motility (V
VFG002409(gb|WP_000549274) (essC) type VII secretion system protein EssC, FtsK/SpoIIIE family ATPase [T
VFG000940(gb|WP_001299620) (iucA) aerobactin siderophore biosynthesis protein IucD [Aerobactin (VF0229)
VFG038627(gb|WP_016350050) (fliF) flagellar MS-ring protein [Polar flagella (VF0473) - Motility (VFC0204)]
VFG008007(gb|NP_218342) (pks2) phthioceranic/hydroxyphthioceranic acid synthase [Sulfolipid-1 biosynthesis
VFG038305(gb|WP_016349343) (exeL) general secretion pathway protein L [Exe T2SS (VF0478) - Effector del
VFG002179(gb|AAD09858) (esp) Enterococcal surface protein; Esp [Esp (VF0353) - Adherence (VFC0001)] [E
VFG045789(gb|WP_001264082) (cgsF) curli production assembly/transport protein CsgF [Curli fibers (VF1138)
VFG015045(gb|NP_249821) (rhlC) rhamnosyltransferase 2 [Rhamnolipid (VF0089) - Immune modulation (VFC
VFG001270(gb|WP_010929997) (cyaD) cyclolysin secretion protein [Cya (VF0028) - Exotoxin (VFC0235)] [Bo
VFG038380(gb|WP_043163044) (vipB) Type VI secretion system contractile sheath large subunit TssC/VipB [T
VFG038323(gb|WP_005320806) (ascI) Type III secretion system inner rod protein AscI [T3SS (VF0479) - Effec
VFG038691(gb|WP_010634619) (flhG) CobQ/CobB/MinD/ParA family protein [Polar flagella (VF0473) - Moti
VFG000249(gb|WP_002214251) (fbpB) iron(III) ABC transporter, permease protein [FbpABC (VF0272) - Nutri
VFG044329(gb|WP_015874964) (irp1) yersiniabactin polyketide synthase HMWP1 [Ybt (VF0564) - Nutritional
VFG022051(gb|NP_214798) (eccC3) Type VII secretion system protein EccC3 [ESX-3 (VF0521) - Effector deli
VFG038409(gb|WP_011704492) (fimA) fimbrial protein [Type I pili (VF0486) - Adherence (VFC0001)] [Aerom
VFG012513(gb|WP_001221122) (iroB) glucosyltransferase IroB [Salmochelin siderophore (VF0230) - Nutrition
VFG001827(gb|WP_001020414) (senB) enterotoxin [ShET2 (VF0258) - Exotoxin (VFC0235)] [Shigella boydii
VFG043014(gb|NP_250149) (PA1458) two-component sensor [Flagella (VF0273) - Motility (VFC0204)] [Pseud
VFG044324(gb|WP_032447693) (ybtX) yersiniabactin-associated zinc MFS transporter YbtX [Ybt (VF0564) -
VFG002115(gb|WP_015444209) (sdeB/laiB) Dot/Icm type IV secretion system effector SdeB [Dot/Icm T4SS se
VFG046120(gb|WP_011206514) (MHP_RS03455) P102 paralog, surface adhesin [P97/P102 paralog family (VF
VFG010471(gb|WP_014841277) (pilO) type IV pilus inner membrane platform protein PilO [Type IV pili (VF01
VFG038840(gb|WP_005300916) (flmH) short chain dehydrogenase/reductase family oxidoreductase [Polar flage
VFG034754(gb|WP_000081129) (espL1) Type III secretion system effector espL1 [TTSS secreted effectors (VF
VFG038383(gb|WP_011705715) (atsH) Type VI secretion system protein [T6SS (VF0480) - Effector delivery sy
VFG038878(gb|WP_005314254) (flgI) flagellar basal body P-ring protein [Lateral flagella (VF0474) - Motility (
VFG048468(gb|WP_004176917) (fepD) iron-enterobactin transporter membrane protein [Ent (VF0562) - Nutritio
VFG049122(gb|WP_012737539) (allC) allantoate amidohydrolase [Allantion utilization (VF0572) - Nutritional/
VFG000232(gb|WP_002218487) (pilT) twitching motility protein PilT [Type IV pili (VF0075) - Adherence (VF

VFG000907(gb|WP_000376545) (hlyB) Hemolysin B [-Hemolysin (VF0225) - Exotoxin (VFC0235)] [Escherich
VFG016260(gb|WP_000714434) (hblD) Hemolysin BL lytic component L1 [HBL (VF0532) - Exotoxin (VFC02
VFG013315(gb|WP_005694490) (lgtF) beta-1,4-glucosyltransferase [LOS (VF0044) - Immune modulation (VFC
VFG041033(gb|NP_251051) (PA2361) hypothetical protein [HSI-3 (VF0944) - Effector delivery system (VFC00
VFG001142(gb|WP_042960771) (hopZ) outer membrane protein adhesin [HopZ (VF0054) - Adherence (VFC00
VFG001861(gb|WP_010945955) (katA) catalase/(hydro)peroxidase [KatAB (VF0168) - Stress survival (VFC028
VFG038568(gb|WP_016349166) (mshO) MSHA (mannose-sensitive hemagglutinin) pili biogenesis protein Msh
VFG001267(gb|NP_252913) (pchH) ABC transporter ATP-binding protein [Pyochelin (VF0095) - Nutritional/M
VFG002414(gb|WP_000730972) (yagZ/ecpA) E. coli common pilus structural subunit EcpA [ECP (VF0404) - A
VFG014707(gb|NP_250151) (motC) flagellar motor protein [Flagella (VF0273) - Motility (VFC0204)] [Pseudom
VFG002052(gb|WP_000820136) (gspI) general secretion pathway protein I [T2SS (VF0333) - Effector delivery s
VFG038596(gb|WP_016349157) (mshM) membrane protein with ATPase domain, GspA-like [MSHA type IV pi
VFG001381(gb|YP_177728) (icl) Isocitrate lyase Icl (isocitrase) (isocitratase) [Isocitrate lyase (VF0253) - Others
VFG037513(gb|WP_085947930) (ACICU_RS04590) porin family protein [HemO cluster (VF0636) - Nutritional
VFG002270(gb|WP_011180104) (badA/vomp/brp) Surface protein/Bartonella adhesin BadA/VompA/VompC/V
VFG000434(gb|NP_490509) (pefC) plasmid-encoded fimbriae usher protein PefC [Pef (VF0104) - Adherence (V
VFG006300(gb|WP_010980816) (frpA) iron-regulated protein FrpA [RTX protein (VF0897) - Adherence (VFC
VFG013476(gb|WP_032828451) (kpsF) KpsF/GutQ family sugar isomerase [LOS (VF0044) - Immune modulati
VFG045791(gb|WP_000481500) (cgsD) transcriptional regulator CsgD [Curli fibers (VF1138) - Adherence (VF
VFG038883(gb|WP_005314227) (lafB) flagellar hook associated protein LafB [Lateral flagella (VF0474) - Moti
VFG000872(gb|WP_000044711) (fimE) Type 1 fimbriae Regulatory protein fimE [Type 1 fimbriae (VF0221) - A
VFG001223(gb|NP_249086) (pilT) twitching motility protein PilT [Type IV pili (VF0082) - Adherence (VFC000
VFG016038(gb|NP_251090) (pvdJ) pyoverdine biosynthesis protein PvdJ [Pyoverdine (VF0094) - Nutritional/M
VFG038393(gb|WP_010634228) (dotU) Type VI secretion system protein DotU [T6SS (VF0480) - Effector deli
VFG001225(gb|NP_249099) (pilG) twitching motility protein PilG [Type IV pili (VF0082) - Adherence (VFC00
VFG034577(gb|WP_014639428) (aafD) chaperone protein AafD [AAFs (VF0214) - Adherence (VFC0001)] [Es
VFG038402(gb|WP_011705725) (vasK/icmF) Type VI secretion system protein [T6SS (VF0480) - Effector deli
VFG034772(gb|WP_000627910) (espL4) Type III secretion system effector EspL4 [TTSS secreted effectors (VF
VFG034831(gb|WP_000484243) (espR4) Type III secretion system effector espR4 [TTSS secreted effectors (VF
VFG034928(gb|WP_001239082) (lifA/efa1) lymphostatin Efa1/LifA [Lymphostatin/LifA (VF0175) - Effector de
VFG002436(gb|WP_080341181) (boaA) surface-exposed protein, autotransporter [BoaA (VF0434) - Adherence
VFG038369(gb|WP_011705706) (hcp) hemolysin co-regulated protein [T6SS (VF0480) - Effector delivery syste
VFG002342(gb|WP_011816603) (flgK) flagellar hook-associated protein 1 FlgK [Flagella (VF0394) - Motility (
VFG019178(gb|WP_012291519) (bont) botulinum neurotoxin, BoNT [BoNT (VF0375) - Exotoxin (VFC0235)] [
VFG002287(gb|WP_011860904) (toxA) toxin A [TcdA (VF0376) - Exotoxin (VFC0235)] [Clostridium difficile
VFG050291(gb|WP_014302131) (DIP_RS19245) C40 family peptidase [DIP1621 (VF0603) - Adherence (VFC
VFG038695(gb|WP_017409955) (fliA) flagellar biosynthesis sigma factor [Polar flagella (VF0473) - Motility (V
VFG016216(gb|WP_000576719) (alo) thiol-activated cytolysin [ALO (VF0534) - Exotoxin (VFC0235)] [Bacillu
VFG045632(gb|WP_002286924) (ecbA/fss3) Collagen binding MSCRAMM, EcbA/Fss3 [EcbA (VF0539) - Adh
VFG048307(gb|WP_041937815) (fimH) type 1 fimbrial adhesin precursor [Type I fimbriae (VF0566) - Adherenc
VFG050959(gb|WP_000556915.1) (tssM) type VI secretion system membrane subunit TssM [T6SS (VF1337) -
VFG037490(gb|WP_001240499) (ACICU_RS04575) FecR domain-containing protein [HemO cluster (VF0636)
VFG050296(gb|NP_250645) (fapB) minor amyloid subunit FapB [Fap (VF1191) - Adherence (VFC0001)] [Pseu
VFG038261(gb|WP_016352107) (exeA) general secretion pathway protein A [Exe T2SS (VF0478) - Effector del
VFG038639(gb|WP_011705275) (fliI) flagellum-specific ATP synthase [Polar flagella (VF0473) - Motility (VFC
VFG038593(gb|WP_016349158) (mshN) Tetratricopeptide repeat protein, PilF-like [MSHA type IV pili (VF047

VFG038381(gb|WP_011705714) (atsG) Type VI secretion system protein [T6SS (VF0480) - Effector delivery sy
VFG048621(gb|WP_004213924) (iutA) ferric aerobactin receptor IutA [Aerobactin (VF0565) - Nutritional/Metal
VFG038395(gb|WP_011705721) (clpB) type VI secretion system ATPase ClpV1 [T6SS (VF0480) - Effector deli
VFG005763(gb|WP_000859501) (cylD) malonyl-CoA-ACP transacylase CylD [-haemolysin/cytolysin (VF0279)
VFG044331(gb|WP_015874966) (ybtT) type II thioesterase YbtT [Ybt (VF0564) - Nutritional/Metabolic factor (
VFG048819(gb|WP_014907234) (KP1_RS17280) mannose-1-phosphate guanylyltransferase/mannose-6-phospha
VFG048429(gb|WP_004183353) (entE) enterobactin synthase subunit E [Ent (VF0562) - Nutritional/Metabolic fa
VFG000347(gb|WP_010869038) (hgpC) hemoglobin-binding protein [Hgp (VF0270) - Nutritional/Metabolic fac
VFG001254(gb|NP_249795) (fliI) flagellum-specific ATP synthase FliI [Flagella (VF0273) - Motility (VFC0204
VFG000044(gb|WP_004568087) (bscR) Bordetella Bsc type III secretion system protein BscR [TTSS (VF0035)
VFG002381(gb|NP_218386) (eccB1) ESX-1 type VII secretion system protein EccB1 [ESX-1 (VF0315) - Effecto
VFG044195(gb|WP_003022876) (fslE) outer membrane siderophore receptor FslE [Rhizoferrin (VF0551) - Nutri
VFG000844(gb|WP_001034100) (espP) autotransporter, serine protease [EspP (VF0208) - Effector delivery syste
VFG002173(gb|WP_010785625) (asa1) aggregation substance Asa1 [AS (VF0352) - Adherence (VFC0001)] [En
VFG013648(gb|WP_012104486) (invD) invasin D protein [Invasin D (VF0576) - Invasion (VFC0083)] [Yersinia
VFG000824(gb|WP_001291694) (escU) Type III secretion system export apparatus switch protein EscU [TTSS (
VFG017845(gb|WP_011988489) (pilR) type IV pilus biosynthesis protein PilR [Type IV pili (VF0401) - Adheren
VFG002529(gb|WP_011205290) (tsr) methyl-accepting chemotaxis protein I [Flagella (VF0430) - Motility (VFC
VFG048518(gb|WP_012737617) (fepA) outer membrane receptor FepA [Ent (VF0562) - Nutritional/Metabolic fa
VFG002099(gb|WP_010948192) (lepB) Dot/Icm type IV secretion system effector LepB, Rab1 GTPase activatin
VFG048348(gb|WP_004149658) (mrkJ) phosphodiesterase [Type 3 fimbriae (VF0567) - Biofilm (VFC0271)] [K
VFG000931(gb|WP_001336900) (entC) isochorismate synthase 1 [Enterobactin (VF0228) - Nutritional/Metaboli
VFG038488(gb|WP_016349432) (tapY1) type IV pilus biogenesis protein [Tap type IV pili (VF0475) - Adherenc
VFG015515(gb|NP_252903) (phzE1) phenazine biosynthesis protein PhzE [Pyocyanin (VF0100) - Nutritional/M
VFG000181(gb|NP_251792) (xcpS) general secretion pathway protein F [Xcp secretion system (VF0084) - Effec
VFG002055(gb|WP_000097229) (gspL) general secretion pathway protein L [T2SS (VF0333) - Effector delivery
VFG001936(gb|YP_002344528) (Cj1135) glucosyltransferase [LOS (VF0326) - Immune modulation (VFC0258)
VFG048267(gb|WP_015875145) (fimC) periplasmic chaperone [Type I fimbriae (VF0566) - Adherence (VFC00
VFG038743(gb|WP_016350352) (flaH) flagellar hook-associated protein 2 [Polar flagella (VF0473) - Motility (V
VFG042072(gb|WP_010930809) (BP_RS11150) SpoIIE family protein phosphatase [TTSS (VF0035) - Effector
VFG000231 (pilC) pilus assembly protein [Type IV pili (VF0075) - Adherence (VFC0001)] [Neisseria meningiti
VFG001443(gb|AAF37887) (ompA) outer membrane protein A [OmpA (VF0236) - Invasion (VFC0083)] [Esche
VFG038828(gb|WP_011706649) (flgN) flagellar protein FlgN [Polar flagella (VF0473) - Motility (VFC0204)] [A
VFG000178(gb|NP_251789) (xcpV) general secretion pathway protein I [Xcp secretion system (VF0084) - Effec
VFG000589(gb|NP_460061) (pipB) type III secretion system effector PipB [TTSS-2 secreted effectors (VF0947)
VFG048797(gb|WP_004175261) (ugd) UDP-glucose 6-dehydrogenase [Capsule (VF0560) - Immune modulation
VFG000974(gb|WP_010922711) (scpA/scpB) streptococcal C5a peptidase [C5a peptidase (VF0245) - Adherence
VFG041016(gb|NP_250349) (PA1658) hypothetical protein [HSI-2 (VF0943) - Effector delivery system (VFC00
VFG001863(gb|WP_010946430) (ligA) LigA, interaptin [LigA (VF0165) - Effector delivery system (VFC0086)]
VFG045790(gb|WP_000833288) (cgsE) curli production assembly/transport protein CsgE [Curli fibers (VF1138)
VFG002050(gb|WP_001087302) (gspG) general secretion pathway protein G [T2SS (VF0333) - Effector delivery
VFG002191(gb|WP_002369252) (sprE) serine proteinase, V8 family [SprE (VF0358) - Exoenzyme (VFC0251)]
VFG038753(gb|WP_024944879) (motY) sodium-type flagellar protein MotY [Polar flagella (VF0473) - Motility
VFG048830(gb|WP_014907233) (gndA) NADP-dependent phosphogluconate dehydrogenase [Capsule (VF0560
VFG049154(gb|WP_014907257) (clbH) colibactin non-ribosomal peptide synthetase ClbH [Colibactin (VF0573)
VFG038727(gb|WP_011705295) (cheW) chemotaxis protein CheW [Polar flagella (VF0473) - Motility (VFC020

VFG049903(gb|WP_001240533) (tssA) Type VI secretion system protein TssA [T6SS (VF0579) - Effector deliv
VFG000459(gb|NP_460116) (csgC) curli assembly protein CsgC [Agf (VF0103) - Adherence (VFC0001)] [Salm
VFG048885(gb|WP_014907227) (gmd) GDP-mannose 4,6-dehydratase [Capsule (VF0560) - Immune modulation
VFG005297(gb|WP_000723540) (pilB) major subunit PilB [PI-2a (VF0528) - Adherence (VFC0001)] [Streptoco
VFG038600(gb|WP_016349156) (mshL) MSHA (mannose-sensitive hemagglutinin) pili biogenesis protein Msh
VFG036992(gb|WP_002224969) (mtrD) multiple transferable resistance system protein MtrD [MtrCDE (VF045
VFG014893(gb|NP_254009) (algC) phosphomannomutase AlgC [Alginate (VF0091) - Biofilm (VFC0271)] [Pse
VFG044165(gb|WP_001041793) (entS) enterobactin exporter, iron-regulated [Enterobactin (VF0228) - Nutrition
VFG039676(gb|NP_821023) (coxCC15) Coxiella Dot/Icm type IVB secretion system translocated effector [T4S
VFG007959(gb|NP_217478) (Rv2962c) PGL/p-HBAD biosynthesis rhamnosyltransferase [PDIM (VF0309) - Im
VFG050910(gb|WP_001178477.1) (vgrG/tssI) type VI secretion system tip protein VgrG [T6SS (VF1337) - Effe
VFG021863(gb|YP_178022) (PPE68) PPE family protein [ESX-1 (VF0315) - Effector delivery system (VFC008
VFG000937(gb|WP_000750139) (iucD) L-lysine 6-monooxygenase IucD [Aerobactin (VF0229) - Nutritional/M
VFG038882(gb|WP_005314247) (maf-5) modification accessory factor [Lateral flagella (VF0474) - Motility (VF
VFG038397(gb|WP_011705722) (vasH) sigma-54 dependent transcriptional regulator [T6SS (VF0480) - Effecto
VFG000122(gb|NP_252230) (algD) GDP-mannose 6-dehydrogenase AlgD [Alginate (VF0091) - Biofilm (VFC0
VFG001218(gb|NP_253727) (pilQ) type 4 fimbrial biogenesis protein PilQ [Type IV pili (VF0082) - Adherence
VFG000936(gb|WP_016265865) (iutA) ferric aerobactin receptor precusor IutA [Aerobactin (VF0229) - Nutritio
VFG038836(gb|WP_011707085) (pomA2) chemotaxis protein PomA [Polar flagella (VF0473) - Motility (VFC0
VFG002481(gb|WP_009940775) (vgrG-5) Type VI secretion system protein VgrG5 [T6SS-1 (VF0429) - Effecto
VFG049916(gb|WP_000103365) (vgrG/tssI) type VI secretion system effector VgrG [T6SS (VF0579) - Effector
VFG001226(gb|NP_249100) (pilH) twitching motility protein PilH [Type IV pili (VF0082) - Adherence (VFC00
VFG039585(gb|NP_820667) (CBU_1685) Coxiella Dot/Icm type IVB secretion system translocated effector [T4
VFG049124(gb|WP_012737540) (allD) ureidoglycolate dehydrogenase [Allantion utilization (VF0572) - Nutritio
VFG034873(gb|WP_001270084) (espX5) Type III secretion system effector EspX5 [TTSS secreted effectors (V
VFG038832(gb|WP_016351724) (pomB2) flagellar motor protein MotB [Polar flagella (VF0473) - Motility (VF
VFG038342(gb|WP_005320770) (ascV) Type III secretion system inner membrane export apparatus protein Asc
VFG044322(gb|WP_004190148) (iroE) siderophore esterase IroE [Sal (VF0563) - Nutritional/Metabolic factor (
VFG038764(gb|WP_017409846) (flrA) FleQ protein [Polar flagella (VF0473) - Motility (VFC0204)] [Aeromona
VFG038432(gb|WP_016352168) (tapB) type IV pilus assembly ATPase, PilB-like [Tap type IV pili (VF0475) -
VFG000924(gb|WP_001234311) (fepB) ferrienterobactin ABC transporter periplasmic binding protein [Enteroba
VFG048277(gb|WP_009308599) (fimD) outer membrane usher protein [Type I fimbriae (VF0566) - Adherence
VFG000010(gb|WP_010930243) (tcfA) tracheal colonization factor precursor [TcfA (VF0032) - Adherence (VF
VFG038760(gb|WP_016351312) (flrB) two-component sensor kinase [Polar flagella (VF0473) - Motility (VFC0
VFG040863(gb|WP_011190526) (sca2) cell surface antigen Sca2 [Sca2 (VF0495) - Motility (VFC0204)] [Ricke
VFG034636(gb|WP_000358674) (cfaC) colonisation factor antigen c, usher [Adhesive fimbriae (VF0213) - Adhe
VFG002497(gb|WP_011204776) (fliK) flagellar hook-length control protein FliK [Flagella (VF0430) - Motility (
VFG048665(gb|WP_004176431) (dotU/tssL) type VI secretion system protein, DotU/TssL family [T6SS (VF056
VFG049156(gb|WP_077265119) (clbJ) colibactin non-ribosomal peptide synthetase ClbJ [Colibactin (VF0573) -
VFG049062(gb|WP_015875020) (rfbD) UDP-galactopyranose mutase [LPS (VF0561) - Immune modulation (V
VFG049162(gb|WP_002430641) (clbP) precolibactin peptidase ClbP [Colibactin (VF0573) - Exotoxin (VFC023
VFG012438(gb|WP_000022458) (espJ) Type III secretion system effector EspJ, adenosine diphosphate (ADP) ri
VFG002346(gb|WP_005160438) (flgG) flagellar basal-body rod protein FlgG [Flagella (VF0394) - Motility (VF
VFG038367(gb|WP_011705045) (vgrG1) Type VI secretion system effector VgrG1, ADP-ribosyltransferase [T
VFG049095(gb|WP_002912373) (rfbA) O-antigen export ABC transporter permease RfbA [LPS (VF0561) - Imm
VFG038780(gb|WP_011706637) (flgI) flagellar basal body P-ring protein [Polar flagella (VF0473) - Motility (V

VFG012636(gb|NP_858256) (virB) transcriptional activator VirB [TTSS (VF0118) - Effector delivery system (V
VFG000902(gb|WP_001034089) (sat) Aecreted auto transpoter toxin [Sat (VF0231) - Effector delivery system (V
VFG038354(gb|WP_005320741) (ascT) Type III secretion system inner membrane export apparatus protein AscT
VFG001238(gb|NP_249771) (flgE) flagellar hook protein FlgE [Flagella (VF0273) - Motility (VFC0204)] [Pseud
VFG002483(gb|WP_004556233) (tagB-5) Type VI secretion system associated protein TagB-5 [T6SS-1 (VF042
VFG012089(gb|WP_011861682) (cwp66) cell wall-binding protein Cwp66 [Cwp66 (VF0591) - Adherence (VFC
VFG006538(gb|WP_000832067) (flgI) flagellar P-ring protein precursor FlgI [Flagella (VF0051) - Motility (VFC
VFG000478(gb|NP_459678) (fur) ferric iron uptake transcriptional regulator [Fur (VF0113) - Regulation (VFC0
VFG049160(gb|WP_001327259) (clbN) colibactin non-ribosomal peptide synthetase ClbN [Colibactin (VF0573)
VFG002224(gb|WP_002963682) (wbkA) mannosyltransferase [LPS (VF0367) - Immune modulation (VFC0258)
VFG050167(gb|WP_000038461) (neuD) NeuD protein involved in sialic acid synthesis [K1 capsule (VF0239) -
VFG037573 (csuA) Csu pilus subunit CsuA [Csu fimbriae (VF0461) - Biofilm (VFC0271)] [Acinetobacter baum
VFG038804(gb|WP_011706643) (flgC) flagellar basal body rod protein FlgC [Polar flagella (VF0473) - Motility
VFG002046(gb|WP_000135089) (gspC) general secretion pathway protein C [T2SS (VF0333) - Effector delivery
VFG050166(gb|WP_000066003) (neuB) N-acylneuraminate-9-phosphate synthase [K1 capsule (VF0239) - Invas
VFG000123(gb|NP_252231) (alg8) alginate-c5-mannuronan-epimerase AlgG [Alginate (VF0091) - Biofilm (VF
VFG000908(gb|WP_000860472) (hlyD) Hemolysin D [-Hemolysin (VF0225) - Exotoxin (VFC0235)] [Escherich
VFG049051(gb|WP_014907239) (KP1_RS17230) glycosyltransferase [LPS (VF0561) - Immune modulation (VF
VFG048237(gb|WP_002916189) (fimE) tyrosine recombinase [Type I fimbriae (VF0566) - Adherence (VFC000
VFG038281(gb|WP_016349337) (exeF) general secretion pathway protein F [Exe T2SS (VF0478) - Effector deli
VFG002120(gb|WP_010948516) (sidH) Dot/Icm type IV secretion system effector SidH [Dot/Icm T4SS secreted
VFG034875(gb|WP_001270087) (espX5) Type III secretion system effector EspX5 [TTSS secreted effectors (V
VFG050168(gb|WP_000590258) (kpsT) Polysialic acid transport ATP-binding protein [K1 capsule (VF0239) - I
VFG038723(gb|WP_011705294) (AHML_RS07540) CobQ/CobB/MinD/ParA family protein [Polar flagella (VF
VFG038285(gb|WP_005308483) (exeG) general secretion pathway protein G [Exe T2SS (VF0478) - Effector del
VFG002195(gb|WP_002399773) (EF0818) polysaccharide lyase, family 8 [Hyaluronidase (VF0359) - Exoenzym
VFG001974(gb|YP_002344829) (kpsT) capsule polysaccharide ABC transporter ATP-binding protein [Capsule (
VFG002291(gb|WP_003458888) (ext) epsilon-toxin [Epsilon-toxin (ETX) (VF0380) - Exotoxin (VFC0235)] [Cl
VFG041018(gb|NP_250351) (PA1660) hypothetical protein [HSI-2 (VF0943) - Effector delivery system (VFC00
VFG016046(gb|NP_251103) (pvdH) diaminobutyrate-2-oxoglutarate aminotransferase PvdH [Pyoverdine (VF00
VFG000522(gb|NP_460387) (ssaU) type III secretion system export apparatus switch protein SsaU [TTSS (SPI-2
VFG000182(gb|NP_251793) (xcpR) general secretion pathway protein E [Xcp secretion system (VF0084) - Effec
VFG002307(gb|NP_461452) (sinH) intimin-like protein [SinH (VF0400) - Adherence (VFC0001)] [Salmonella e
VFG038675(gb|WP_011705283) (fliR) flagellar biosynthesis protein FliR [Polar flagella (VF0473) - Motility (V
VFG037316(gb|WP_000744381) (entE) non-ribosomal peptide synthetase adenylate-forming enzyme of acinetob
VFG001900(gb|YP_002343773) (flhB) flagellar biosynthetic protein [Flagella (VF0114) - Motility (VFC0204)] [
VFG038845(gb|WP_016351249) (nueA) NeuA protein [Polar flagella (VF0473) - Motility (VFC0204)] [Aeromo
VFG048449(gb|WP_012737621) (fepB) iron-enterobactin transporter periplasmic binding protein [Ent (VF0562)
VFG049133(gb|WP_004177236) (acrA) acriflavine resistance protein A [AcrAB (VF0568) - Antimicrobial activi
VFG038371(gb|WP_011705707) (vgrG2) VgrG protein [T6SS (VF0480) - Effector delivery system (VFC0086)]
VFG050162(gb|WP_001161829) (kpsS) Capsule polysaccharide export protein [K1 capsule (VF0239) - Invasion
VFG050273(gb|WP_000874844) (futC2) alpha-(1,2)-fucosyltransferase [Lewis antigen (VF0057) - Immune mod
VFG038269(gb|WP_075384415) (exeC) general secretion pathway protein C [Exe T2SS (VF0478) - Effector del
VFG049899(gb|WP_001087735) (tssA) Type VI secretion system protein TssA [T6SS (VF0579) - Effector deliv
VFG048508(gb|WP_012737618) (fes) enterobactin/ferric enterobactin esterase [Ent (VF0562) - Nutritional/Meta
VFG038460(gb|WP_043119325) (tapQ) type IV pilus secretin PilQ [Tap type IV pili (VF0475) - Adherence (VF

VFG048439(gb|WP_014907941) (entC) isochorismate synthase [Ent (VF0562) - Nutritional/Metabolic factor (V[illegible]
VFG039351(gb|NP_819656) (cetCb3) Coxiella Dot/Icm type IVB secretion system translocated effector [T4SS [illegible]
VFG049904 (clpV/tssH) Type VI secretion system ATPase ClpV/TssH [T6SS (VF0579) - Effector delivery syst[illegible]
VFG038403(gb|WP_011705726) (atsS) Type VI secretion system protein [T6SS (VF0480) - Effector delivery sy[illegible]
VFG038273(gb|WP_016349335) (exeD) general secretion pathway protein D [Exe T2SS (VF0478) - Effector de[illegible]
VFG042739(gb|NP_254019) (crc) catabolite repression control protein [Type IV pili (VF0082) - Adherence (VF[illegible]
VFG000912(gb|WP_001443207) (focD) F1C fimbrial usher [F1C fimbriae (VF0224) - Adherence (VFC0001)] [[illegible]
VFG000362(gb|WP_002212777) (irp1) yersiniabactin biosynthetic protein Irp1 [Yersiniabactin (VF0136) - Nutri[illegible]
VFG011757(gb|WP_011117594) (virB4) Type IV secretion system protein VirB4 [Type IV secretion system (V[illegible]
VFG048317(gb|WP_014906959) (fimK) transcriptional regulator [Type I fimbriae (VF0566) - Adherence (VFC[illegible]
VFG002070(gb|NP_248774) (hsiC1/vipB/tssC) type VI secretion system tubule-forming protein VipB [HSI-I (V[illegible]
VFG006217 (opa) outer membrane beta-barrel protein [Opa (VF0076) - Invasion (VFC0083)] [Neisseria mening[illegible]
VFG048459(gb|WP_004147525) (entS) enterobactin exporter EntS [Ent (VF0562) - Nutritional/Metabolic factor
VFG047137(gb|WP_003022294) (wzy) membrane protein [LPS (VF0542) - Immune modulation (VFC0258)] [F[illegible]
VFG003523(gb|WP_005173750) (ysaN) type III secretion system ATPase SctN [Ysa TTSS (VF1025) - Effector
VFG038464(gb|WP_011707389) (tapT) twitching ATPase [Tap type IV pili (VF0475) - Adherence (VFC0001)]
VFG000133(gb|NP_252241) (algA) phosphomannose isomerase / guanosine 5'-diphospho-D-mannose pyrophos[illegible]
VFG038525(gb|WP_043118801) (flpH) Flp pilus assembly protein FlpH [Flp type IV pili (VF0476) - Adherence
VFG000079(gb|NP_463763) (clpC) endopeptidase Clp ATP-binding chain C [ClpC (VF0072) - Stress survival ([illegible]
VFG050163(gb|WP_000197083) (neuE) polysialic acid biosynthesis protein [K1 capsule (VF0239) - Invasion (V[illegible]
VFG038788(gb|WP_011706639) (flgG) flagellar basal body rod protein FlgG [Polar flagella (VF0473) - Motility
VFG040507(gb|WP_011191135) (ompB) outer membrane protein OmpB [ROmpB/Sca5 (VF0493) - Invasion (V[illegible]
VFG000690(gb|AAP42183) (AAP42183) Bcs3' [Capsule (VF0043) - Immune modulation (VFC0258)] [Haemop[illegible]
VFG002095(gb|WP_010947666) (ralF) Dot/Icm type IV secretion system effector guanine nucleotide exchange [illegible]
VFG000876(gb|WP_000120946) (fimD) Outer membrane usher protein fimD precursor [Type 1 fimbriae (VF02[illegible]
VFG049084(gb|WP_004175264) (rfbB) O-antigen export ABC transporter ATP-binding protein RfbB [LPS (VF[illegible]
VFG031564(gb|YP_177826) (PE_PGRS30) PE-PGRS family protein PE_PGRS30 [PE/PE-PGRS (VF0316) - Im[illegible]
VFG047162(gb|WP_003022298) (wbtE) nucleotide sugar dehydrogenase [LPS (VF0542) - Immune modulation [illegible]
VFG019180(gb|WP_011987710) (bont) botulinum neurotoxin, BoNT [BoNT (VF0375) - Exotoxin (VFC0235)] [illegible]
VFG000297(gb|WP_000835501) (cagN) type IV secretion system protein Cag17/CagN, membrane-associated pr[illegible]
VFG049073(gb|WP_015875021) (KP1_RS17240) DUF4422 domain-containing protein [LPS (VF0561) - Immu[illegible]
VFG000038(gb|WP_003807115) (bplA) probable oxidoreductase [LPS (VF0033) - Immune modulation (VFC02[illegible]
VFG002158(gb|NP_464456) (lplA1) lipoate protein ligase [LplA1 (VF0347) - Nutritional/Metabolic factor (VFC[illegible]
VFG002172(gb|WP_010710127) (EF0485) aggregation substance [AS (VF0352) - Adherence (VFC0001)] [Ent[illegible]
VFG011429(gb|WP_002963985) (acpXL) acyl carrier protein [LPS (VF0367) - Immune modulation (VFC0258)[illegible]
VFG048755(gb|WP_049127998) (icmF/tssM) type VI secretion protein TssM [T6SS (VF0569) - Effector delive[illegible]
VFG002463(gb|WP_011205639) (bsaO) Type III secretion system protein BsaO [Bsa T3SS (VF0428) - Effector
VFG002359(gb|WP_005164498) (flhD) flagellar transcriptional activator FlhD [Flagella (VF0394) - Motility (V[illegible]
VFG002053(gb|WP_001250462) (gspJ) general secretion pathway protein J [T2SS (VF0333) - Effector delivery
VFG013594(gb|WP_164927859) (hgpB) TonB-dependent hemoglobin/transferrin/lactoferrin family receptor [Hg[illegible]
VFG049919(gb|AAD32394) (bslA) s-layer protein [BslA (VF0411) - Adherence (VFC0001)] [Bacillus anthracis
VFG043500(gb|WP_011206108) (mhp271) p97 cilium adhesin paralog [P97/P102 paralog family (VF0426) - Ad[illegible]
VFG001447(gb|CAA50007) (cnf1) cytotoxic necrotizing factor 1 [CNF-1 (VF0240) - Exotoxin (VFC0235)] [Es[illegible]
VFG043212(gb|WP_005168761) (motB) flagellar motor protein MotB [Flagella (VF0394) - Motility (VFC0204)[illegible]
VFG048771(gb|WP_004190505) (tssF) type VI secretion system baseplate subunit TssF [T6SS (VF0569) - Effec[illegible]
VFG012033(gb|YP_002344722) (pseD/maf2) motility accessory factor PseD [Flagella (VF0114) - Motility (VF[illegible]

VFG001963(gb|YP_002344818) (Cj1435c) phosphatase [Capsule (VF0323) - Immune modulation (VFC0258)] [
VFG001810(gb|YP_178017) (fbpD) MPT51/MPB51 antigen [Antigen 85 (VF0311) - Adherence (VFC0001)] [M
VFG048779(gb|WP_004190503) (tssG) type VI secretion system baseplate subunit TssG [T6SS (VF0569) - Effe
VFG045335(gb|AHK25020) (llsB) SagB family dehydrogenase LlsB [LLS (VF0410) - Exotoxin (VFC0235)] [Li
VFG001444(gb|AAG10151) (aslA) putative arylsulfatase [AslA (VF0238) - Invasion (VFC0083)] [Escherichia c
VFG038293(gb|WP_016349340) (exeI) general secretion pathway protein I [Exe T2SS (VF0478) - Effector deliv
VFG044326(gb|WP_001327262) (ybtP) yersiniabactin ABC transporter ATP-binding/permease protein YbtP [Yb
VFG038505(gb|WP_011705643) (tapF) type IV pilus biogenesis/stability protein, pilW-like [Tap type IV pili (V
VFG038533(gb|WP_016350152) (flpF) traffic ATPase [Flp type IV pili (VF0476) - Adherence (VFC0001)] [Ae
VFG038468(gb|WP_016351972) (tapU) twitching ATPase [Tap type IV pili (VF0475) - Adherence (VFC0001)]
VFG002049(gb|WP_001173470) (gspF) general secretion pathway protein F [T2SS (VF0333) - Effector delivery
VFG038772(gb|WP_016351319) (flgK) flagellar hook-associated protein FlgK [Polar flagella (VF0473) - Motilit
VFG002071(gb|NP_248775) (hcp1) type VI secretion system substrate Hcp1 [HSI-I (VF0334) - Effector delivery
VFG002465(gb|WP_009940818) (bsaM) Type III secretion system protein BsaM [Bsa T3SS (VF0428) - Effector
VFG012509(gb|WP_001111200) (iroC) ATP binding cassette transporter [Salmochelin siderophore (VF0230) - N
VFG038309(gb|WP_016349344) (exeM) general secretion pathway protein M [Exe T2SS (VF0478) - Effector de
VFG038587(gb|WP_016349160) (mshG) MSHA (mannose-sensitive hemagglutinin) pili biogenesis protein Msh
VFG049910(gb|WP_000348788) (tssG) Type VI secretion system protein TssG [T6SS (VF0579) - Effector deliv
VFG001400(gb|NP_216865) (plcC) phospholipase C [Phospholipase C (VF0320) - Exotoxin (VFC0235)] [Myco
VFG002276(gb|WP_011009653) (colA) collagenase [Kappa-toxin (VF0388) - Exoenzyme (VFC0251)] [Clostrid
VFG034815(gb|WP_000671689) (espR1) Type III secretion system effector espR1 [TTSS secreted effectors (VF
VFG037837(gb|WP_000868104) (lpxD) UDP-3-O-(3-hydroxymyristoyl)glucosamine N-acyltransferase [LPS (V
VFG043626(gb|WP_000820818) (mrkB) fimbrial chaperone protein mrkB precursor [Type 3 fimbriae (VF0567)
VFG042734(gb|NP_249343) (vfr) cAMP-regulatory protein [Type IV pili (VF0082) - Adherence (VFC0001)] [P
VFG037762(gb|WP_000078878) (lpxL) lauroyl acyltransferase [LPS (VF0466) - Immune modulation (VFC0258
VFG048676(gb|WP_014907726) (ompA) OmpA family protein [T6SS (VF0569) - Effector delivery system (VF
VFG051028(gb|WP_000471445.1) (tssK) type VI secretion system baseplate subunit TssK [T6SS (VF1337) - Ef
VFG000938(gb|WP_001015709) (iucC) aerobactin siderophore biosynthesis protein IucC [Aerobactin (VF0229)
VFG002076(gb|NP_248780) (clpV1) type VI secretion system AAA+ family ATPase [HSI-I (VF0334) - Effector
VFG001263(gb|NP_250143) (flhA) flagellar biosynthesis protein FlhA [Flagella (VF0273) - Motility (VFC0204)
VFG048257(gb|WP_004174507) (fimI) type 1 pilus biosynthesis fimbrial protein [Type I fimbriae (VF0566) - A
VFG049158(gb|WP_001297937) (clbL) colibactin biosynthesis amidase ClbL [Colibactin (VF0573) - Exotoxin (
VFG001214(gb|NP_253237) (pilR) two-component response regulator PilR [Type IV pili (VF0082) - Adherence
VFG050400(gb|WP_000347039.1) (pilU) PilT/PilU family type 4a pilus ATPase [TFP (VF1334) - Adherence (V
VFG012621(gb|WP_000189352) (chuY) ChuY [Chu (VF0234) - Nutritional/Metabolic factor (VFC0272)] [Esch
VFG022130(gb|NP_214804) (eccD3) Type IIV secretion system transmembrane protein EccD3 [ESX-3 (VF0521
VFG038407(gb|WP_011705728) (vgrG3) VgrG protein [T6SS (VF0480) - Effector delivery system (VFC0086)]
VFG045520(gb|WP_010947043) (legC1) Dot/Icm type IV secretion system effector LegC1 [Dot/Icm T4SS secre
VFG015000(gb|NP_252339) (mucP) metalloprotease protease [Alginate (VF0091) - Biofilm (VFC0271)] [Pseud
VFG048708(gb|WP_041168969) (vgrG/tssI) type VI secretion system tip protein VgrG [T6SS (VF0569) - Effect
VFG041276(gb|WP_010947021) (lem8) Dot/Icm type IV secretion system effector Lem8 [Dot/Icm T4SS secrete
VFG016346(gb|WP_001035927) (BAS_RS06430) immune inhibitor A [InhA (VF0536) - Exoenzyme (VFC0251
VFG002467(gb|WP_004525308) (bsaK) Type III secretion system protein BsaK [Bsa T3SS (VF0428) - Effector
VFG038364(gb|WP_005320615) (aexT) Type III secretion system effector AexT, bifunctional toxin with GTPas
VFG011626(gb|WP_002965156) (bvrR) response regulator transcription factor [BvrR-BvrS (VF0368) - Regulati
VFG001290(gb|WP_000745817) (clfB) Clumping factor B, adhesin [Clumping factor (VF0004) - Adherence (V

VFG050247(gb|NP_219793) (CT_288) hypothetical protein [TTSS secreted effectors (VF0711) - Effector deliver
VFG047708(gb|WP_003022611) (carB) carbamoyl phosphate synthase large subunit [Pyrimidine biosynthesis (V
VFG000131(gb|NP_252239) (algJ) alginate o-acetyltransferase AlgJ [Alginate (VF0091) - Biofilm (VFC0271)] [
VFG043656(gb|WP_003377818) (C2I) C2 toxin (component-I) [C2 toxin (VF0384) - Exotoxin (VFC0235)] [Clo
VFG000884(gb|WP_000654937) (papC) usher protein PapC [P fimbriae (VF0220) - Adherence (VFC0001)] [Esc
VFG000343(gb|WP_164927854) (hitB) iron ABC transporter permease [HitABC (VF0268) - Nutritional/Metabo
VFG034880(gb|WP_000535069) (espX6) Type III secretion system effector EspX6 [TTSS secreted effectors (VF
VFG000802(gb|WP_000953255) (escD) Type III secretion system outer MS ring protein EscD [TTSS (VF0191)
VFG050699(gb|WP_000840548.1) (pilR) sigma-54 dependent transcriptional regulator [TFP (VF1334) - Adheren
VFG002259(gb|WP_011181332) (trwK) Trw type IV secretion sysmtem VirB4-like trwK protein [Trw type IV s
VFG038329(gb|WP_011899438) (ascC) Type III secretion system OM ring protein AscC [T3SS (VF0479) - Effe
VFG000918(gb|WP_001312167) (chuT) periplasmic heme-binding protein ChuT [Chu (VF0227) - Nutritional/M
VFG037028(gb|WP_002218620) (katA) catalase [KatA (VF0454) - Stress survival (VFC0282)] [Neisseria menin
VFG002048(gb|WP_000249362) (gspE) general secretion pathway protein E [T2SS (VF0333) - Effector delivery
VFG000166(gb|NP_252916) (pchE) dihydroaeruginoic acid synthetase PchE [Pyochelin (VF0095) - Nutritional/N
VFG001353(gb|WP_001100143) (cbpG) choline binding protein G [CBPs (VF0145) - Adherence (VFC0001)] [S
VFG014950(gb|NP_249457) (mucD) serine protease MucD precursor [Alginate (VF0091) - Biofilm (VFC0271)]
VFG000364(gb|WP_000140406) (ybtA) transcriptional regulator YbtA [Yersiniabactin (VF0136) - Nutritional/M
VFG050772(gb|WP_001020922.1) (gspE2) putative secretion pathway ATPase [T2SS (VF1335) - Effector delive
VFG021876(gb|NP_218393) (espI) ESX-1 secretion-associated protein EspI [ESX-1 (VF0315) - Effector delivery
VFG044325(gb|WP_002212761) (ybtQ) yersiniabactin ABC transporter ATP-binding/permease protein YbtQ [Y
VFG035902(gb|WP_000209220) (aatP) permease AatP [Dispersin (VF0215) - Others (VFC0346)] [Escherichia c
VFG002382(gb|NP_218387) (eccCa1) ESX-1 type VII secretion system FtsK/SpoIIIE family protein EccCa1 [ES
VFG039436 (CBU_1107*) Coxiella Dot/Icm type IVB secretion system translocated effector [T4SS secreted effe
VFG014984(gb|NP_253136) (algW) AlgW protein [Alginate (VF0091) - Biofilm (VFC0271)] [Pseudomonas aer
VFG002333(gb|WP_005168827) (fliK) flagellar hook-length control protein FliK [Flagella (VF0394) - Motility (
VFG002067(gb|NP_248771) (fha1) type VI secretion system forkhead-associated protein Fha1 [HSI-I (VF0334)
VFG000064(gb|WP_010930831) (bcrD) Type III secretion system LcrD homolog protein BcrD [TTSS (VF0035)
VFG044096(gb|NP_252908) (PA4218) transporter [Pyochelin (VF0095) - Nutritional/Metabolic factor (VFC027
VFG001966(gb|YP_002344821) (Cj1438c) sugar transferase [Capsule (VF0323) - Immune modulation (VFC025
VFG045567(gb|WP_010948074) (LPG_RS11920) Dot/Icm type IV secretion system effector [Dot/Icm T4SS sec
VFG043211(gb|WP_032902679) (cheA) chemotaxis protein CheA [Flagella (VF0394) - Motility (VFC0204)] [Y
VFG038816(gb|WP_011706646) (cheV) CheV [Polar flagella (VF0473) - Motility (VFC0204)] [Aeromonas hyd
VFG050115(gb|WP_002081542) (cesA) Cereulide synthetase A, CesA [Cereulide (VF0583) - Exotoxin (VFC02
VFG048338(gb|WP_002916079) (mrkI) LuxR family regulatory protein [Type 3 fimbriae (VF0567) - Biofilm (V
VFG048488(gb|WP_002893737) (fepC) iron-enterobactin transporter ATP-binding protein [Ent (VF0562) - Nutr
VFG000025(gb|WP_010929995) (cyaA) bifunctional hemolysin-adenylate cyclase precursor [Cya (VF0028) - Ex
VFG049152(gb|WP_000337350) (clbF) colibactin biosynthesis dehydrogenase ClbF [Colibactin (VF0573) - Exo
VFG013604 (hgbA) TonB-dependent hemoglobin/transferrin/lactoferrin family receptor [Hgp (VF0270) - Nutriti
VFG002489(gb|WP_181937968) (tssM-5) Type VI secretion system protein TssM [T6SS-1 (VF0429) - Effector
VFG000477(gb|NP_461845) (rpoS) RNA polymerase sigma factor RpoS [RpoS (VF0112) - Regulation (VFC030
VFG002219(gb|WP_011005413) (cgs) cyclic beta 1-2 glucan synthetase [CG (VF0366) - Immune modulation (V
VFG049029(gb|WP_014907241) (KP1_RS17220) glycosyltransferase [LPS (VF0561) - Immune modulation (VF
VFG049120(gb|WP_012737535) (allB) allantoinase [Allantion utilization (VF0572) - Nutritional/Metabolic facto
VFG046177(gb|NP_820262) (CBU_1268) Coxiella Dot/Icm type IVB secretion system translocated effector [T4
VFG034915(gb|WP_000225947) (espY4) Type III secretion system effector EspY4 [TTSS secreted effectors (VF

VFG001228(gb|NP_249102) (pilJ) twitching motility protein PilJ [Type IV pili (VF0082) - Adherence (VFC000
VFG051435(gb|NP_220394.1) (pmpH) polymorphic membrane protein PmpH [Pmps (VF1346) - Adherence (VF
VFG048287(gb|WP_014906962) (fimF) type 1 fimbrial minor component [Type I fimbriae (VF0566) - Adherenc
VFG041038(gb|NP_251056) (PA2366) uricase [HSI-3 (VF0944) - Effector delivery system (VFC0086)] [Pseudo
VFG012095(gb|WP_003435012) (groEL) chaperonin GroEL [GroEL (VF0594) - Adherence (VFC0001)] [Clostr
VFG038667(gb|WP_016350054) (fliP) flagellar biosynthesis protein FliP [Polar flagella (VF0473) - Motility (VF
VFG048229(gb|WP_015875142) (fimB) tyrosine recombinase [Type I fimbriae (VF0566) - Adherence (VFC000
VFG038436(gb|WP_016352169) (tapC) type IV fimbrial assembly protein, PilC-like [Tap type IV pili (VF0475)
VFG012333(gb|WP_000703306) (sfaD) S fimbriae minor subunit SfaD [S fimbriae (VF0222) - Adherence (VFC
VFG034859(gb|WP_010904544) (espX2) Type III secretion system effector EspX2 [TTSS secreted effectors (VF
VFG002417(gb|WP_001301550) (yagV/ecpE) E. coli common pilus chaperone EcpE [ECP (VF0404) - Adherenc
VFG038301(gb|WP_016349342) (exeK) general secretion pathway protein K [Exe T2SS (VF0478) - Effector del
VFG038874(gb|WP_005314263) (flgE) flagellar hook protein FlgE [Lateral flagella (VF0474) - Motility (VFC02
VFG037807(gb|WP_000013434) (lpxB) lipid-A-disaccharide synthase [LPS (VF0466) - Immune modulation (VI
VFG000176(gb|NP_251787) (xcpX) general secretion pathway protein K [Xcp secretion system (VF0084) - Effe
VFG046465(gb|WP_003028672) (tufA) elongation factor Tu [EF-Tu (VF0460) - Adherence (VFC0001)] [Franci
VFG049018(gb|WP_002913007) (rcsB) transcriptional regulator RcsB [RcsAB (VF0571) - Regulation (VFC030
VFG038703(gb|WP_011705289) (cheZ) chemotaxis protein CheZ [Polar flagella (VF0473) - Motility (VFC0204
VFG000903(gb|WP_001045652) (pic) Pic serine protease precursor, autotransporter [Pic (VF0232) - Effector del
VFG041261(gb|WP_010946698) (lpg0963) Dot/Icm type IV secretion system effector [Dot/Icm T4SS secreted ef
VFG001280(gb|WP_000934435) (sdrD) Ser-Asp rich fibrinogen-binding bone sialoprotein-binding protein [SDr
VFG044159(gb|WP_000125846) (fes) enterobactin/ferric enterobactin esterase [Enterobactin (VF0228) - Nutritic
VFG038277(gb|WP_016349336) (exeE) general secretory pathway protein E [Exe T2SS (VF0478) - Effector del
VFG006652(gb|WP_001000225) (flgE) flagellar hook protein [Flagella (VF0051) - Motility (VFC0204)] [Helico
VFG038880(gb|WP_005314250) (lfgK) lateral flagellar hook-associated protein [Lateral flagella (VF0474) - Mot
VFG044330(gb|WP_032447695) (ybtU) yersiniabactin biosynthesis oxidoreductase YbtU [Ybt (VF0564) - Nutri
VFG000185(gb|NP_250381) (pscU) type III secretion system protein PscU [TTSS (VF0083) - Effector delivery s
VFG001909(gb|YP_002343979) (fliD) flagellar capping protein FliD [Flagella (VF0114) - Motility (VFC0204)]
VFG002166(gb|WP_010714416) (ace) collagen adhesin protein [Ace (VF0355) - Adherence (VFC0001)] [Enter
VFG041031(gb|NP_251049) (PA2359) transcriptional regulator [HSI-3 (VF0944) - Effector delivery system (VF
VFG005361(gb|WP_002352221) (spaP/pac) cell surface antigen SpaP [SpaP (VF0526) - Adherence (VFC0001)
VFG035006(gb|WP_001117988) (nleG-3) T3SS effector NleG family protein [TTSS secreted effectors (VF1111
VFG034721(gb|WP_000556298) (ibeC) phosphoethanolamine transferase CptA [Ibes (VF0237) - Invasion (VFC
VFG048632(gb|WP_004176434) (vipA/tssB) type VI secretion system contractile sheath small subunit VipA [T6
VFG002056 (gspM) general secretion pathway protein M [T2SS (VF0333) - Effector delivery system (VFC0086
VFG012162(gb|WP_011590241) (nanJ) exo-alpha-sialidase [Sialidase (VF0391) - Exoenzyme (VFC0251)] [Clos
VFG045474(gb|AAM75251) (cylM) CylM; cytolysin subunit modifier [Cytolysin (VF0356) - Exotoxin (VFC023
VFG048643(gb|WP_014907727) (vipB/tssC) type VI secretion system contractile sheath large subunit VipB [T6
VFG002178(gb|AAM75254) (cylI) cytolysin immunity protein [Cytolysin (VF0356) - Exotoxin (VFC0235)] [En
VFG001253(gb|NP_249794) (fliH) flagellar assembly protein H [Flagella (VF0273) - Motility (VFC0204)] [Pseu
VFG046166(gb|NP_819566) (CBU_0534) Coxiella Dot/Icm type IVB secretion system translocated effector [T4
VFG000320(gb|WP_001169234) (kdtB) lipopolysaccharide core biosynthesis protein [LPS (VF0056) - Immune
VFG038176(gb|WP_000777882) (ompA) outer membrane protein OmpA [OmpA (VF0464) - Immune modulatio
VFG049917(gb|WP_000508696) (rhs/PAAR) Type VI secretion system protein, PAAR family [T6SS (VF0579)
VFG000180(gb|NP_251791) (xcpT) general secretion pathway protein G [Xcp secretion system (VF0084) - Effe
VFG000065(gb|WP_010930832) (bscD) putative type III secretion protein [TTSS (VF0035) - Effector delivery s

VFG038631(gb|WP_011705273) (fliG) flagellar motor switch protein G [Polar flagella (VF0473) - Motility (VF
VFG000861(gb|WP_001045649) (pic) Pic serine protease precursor, autotransporter [Pic (VF0218) - Effector del
VFG048030(gb|WP_003022143) (pdpC) Type VI secretion system secreted protein PdpC [T6SS (VF0544) - Effe
VFG048498(gb|WP_012737619) (entF) enterobactin synthase subunit F [Ent (VF0562) - Nutritional/Metabolic f
VFG002499(gb|WP_004197164) (fliI) flagellum-specific ATP synthase FliI [Flagella (VF0430) - Motility (VFC
VFG000928(gb|WP_000640938) (fepG) iron-enterobactin ABC transporter permease [Enterobactin (VF0228) - N
VFG051113(gb|WP_080515433.1) (bigA) adhesin with a bacterial immunoglobulin-like domain [BigA (VF1344
VFG000875(gb|WP_000066579) (fimC) Chaperone protein fimC precursor [Type 1 fimbriae (VF0221) - Adhere
VFG005299(gb|WP_001233990) (pilA) tip adhesin PilA [PI-2a (VF0528) - Adherence (VFC0001)] [Streptococ
VFG038448(gb|WP_016351632) (tapN) type IV pilus biogenesis protein PilN [Tap type IV pili (VF0475) - Adhe
VFG049915(gb|WP_001142968) (hcp2/tssD2) Type VI secretion system protein, Hcp family [T6SS (VF0579) -
VFG038812(gb|WP_011706645) (cheR-3) chemotaxis protein CheR [Polar flagella (VF0473) - Motility (VFC02
VFG000112(gb|NP_253216) (pilB) type 4 fimbrial biogenesis protein PilB [Type IV pili (VF0082) - Adherence (
VFG041020(gb|NP_250353) (PA1662) ClpA/B-type protease [HSI-2 (VF0943) - Effector delivery system (VFC
VFG049912 (tssC) Type VI secretion system protein TssC [T6SS (VF0579) - Effector delivery system (VFC008
VFG000923(gb|WP_001034892) (fepA) ferrienterobactin outer membrane transporter [Enterobactin (VF0228) -
VFG050158(gb|WP_000905916) (kpsE) Capsule polysaccharide export inner-membrane protein [K1 capsule (V
VFG038348(gb|WP_005320755) (ascN) Type III secretion system ATPase [T3SS (VF0479) - Effector delivery s
VFG001252(gb|NP_249793) (fliG) flagellar motor switch protein G [Flagella (VF0273) - Motility (VFC0204)] [
VFG002480(gb|WP_011205629) (tssH-5/clpV) Clp-type ATPase chaperone protein [T6SS-1 (VF0429) - Effecto
VFG003561(gb|WP_005173823) (yst1E) type II secretion system ATPase GspE [Yst1 T2SS (VF0635) - Effector
VFG037537(gb|WP_000359621) (ACICU_RS04605) YbaN family protein [HemO cluster (VF0636) - Nutritiona
VFG043627(gb|WP_000756959) (mrkA) type 3 fimbrial major pilin subunit MrkA [Type 3 fimbriae (VF0567) -
VFG041045(gb|NP_251063) (PA2373) hypothetical protein [HSI-3 (VF0944) - Effector delivery system (VFC0
VFG000630(gb|NP_708742) (sigA) serine protease, autotransporter SigA [SigA (VF0119) - Exoenzyme (VFC02
VFG043213(gb|WP_005164494) (motA) flagellar motor protein MotA [Flagella (VF0394) - Motility (VFC0204)
VFG005757(gb|WP_001101136) (cfa/cfb) cAMP factor [CAMP factor (VF0280) - Exotoxin (VFC0235)] [Strept
VFG048409(gb|WP_086074289) (entA) 2,3-dihydroxybenzoate-2,3-dehydrogenase [Ent (VF0562) - Nutritional/
VFG001296(gb|WP_000676533) (sspA) serine protease; V8 protease; glutamyl endopeptidase [V8 protease (VF
VFG049040(gb|WP_074160401) (KP1_RS17225) glycosyltransferase family 4 protein [LPS (VF0561) - Immun
VFG048697(gb|WP_015874721) (clpV/tssH) type VI secretion system ATPase TssH [T6SS (VF0569) - Effector
VFG048990(gb|WP_001741945) (galF) GalU regulator GalF [Capsule (VF0560) - Immune modulation (VFC025
VFG049155(gb|WP_000829570) (clbI) colibactin polyketide synthase ClbI [Colibactin (VF0573) - Exotoxin (V
VFG045349(gb|WP_000092542) (fdeC) adhesin FdeC [FdeC (VF0506) - Adherence (VFC0001)] [Escherichia c
VFG045788(gb|WP_001189322) (cgsG) curli production assembly/transport protein CsgG [Curli fibers (VF1138
VFG000040(gb|WP_010930814) (bscW) Bordetella Bsc type III secretion system protein BscW [TTSS (VF0035
VFG050161(gb|WP_000579525) (kpsC) Capsule polysaccharide export protein [K1 capsule (VF0239) - Invasion
VFG049900 (tssM) Type VI secretion system protein TssM [T6SS (VF0579) - Effector delivery system (VFC00
VFG000905(gb|WP_001546261) (hlyC) Hemolysin C [-Hemolysin (VF0225) - Exotoxin (VFC0235)] [Escheric
VFG000310(gb|WP_001201399) (pseFG) CMP-N-acetylneuraminic acid synthetase [Flagellar glycosylation/Pse
VFG001259(gb|NP_250137) (fliP) flagellar biosynthetic protein FliP [Flagella (VF0273) - Motility (VFC0204)]
VFG002417(gb|WP_001301550) (yagV/ecpE) E. coli common pilus chaperone EcpE [ECP (VF0404) - Adheren
VFG049095(gb|WP_002912373) (rfbA) O-antigen export ABC transporter permease RfbA [LPS (VF0561) - Im
VFG038533(gb|WP_016350152) (flpF) traffic ATPase [Flp type IV pili (VF0476) - Adherence (VFC0001)] [Ae
VFG001969(gb|YP_002344824) (kfiD) UDP-glucose 6-dehydrogenase [Capsule (VF0323) - Immune modulatio
VFG007478(gb|WP_001234824) (fliE) flagellar hook-basal body protein [Flagella (VF0519) - Motility (VFC02

VFG047672(gb|WP_003021043) (argP) amino acid antiporter [ArgP (VF0557) - Nutritional/Metabolic factor (V
VFG002412(gb|WP_000716386) (yagY/ecpB) E. coli common pilus chaperone EcpB [ECP (VF0404) - Adherenc
VFG038707(gb|WP_016350059) (cheA-2) chemotaxis protein CheA [Polar flagella (VF0473) - Motility (VFC02
VFG048708(gb|WP_041168969) (vgrG/tssI) type VI secretion system tip protein VgrG [T6SS (VF0569) - Effect
VFG001301(gb|WP_000459062) (cap8E) type 8 capsular polysaccharide synthesis protein Cap8E [Capsule (VF0
VFG002343(gb|WP_042661457) (flgJ) <beta>-N-acetylglucosaminidase [Flagella (VF0394) - Motility (VFC020
VFG001928(gb|YP_002344934) (pflA) paralysed flagellum protein [Flagella (VF0114) - Motility (VFC0204)] [C
VFG041118(gb|WP_045606108) (tssA-5) Type VI secretion system protein TssA [T6SS-1 (VF0429) - Effector d
VFG048237(gb|WP_002916189) (fimE) tyrosine recombinase [Type I fimbriae (VF0566) - Adherence (VFC000
VFG045788(gb|WP_001189322) (cgsG) curli production assembly/transport protein CsgG [Curli fibers (VF1138
VFG021876(gb|NP_218393) (espI) ESX-1 secretion-associated protein EspI [ESX-1 (VF0315) - Effector deliver
VFG048755(gb|WP_049127998) (icmF/tssM) type VI secretion protein TssM [T6SS (VF0569) - Effector delive
VFG038768(gb|WP_016351318) (flgL) flagellar hook-associated protein FlgL [Polar flagella (VF0473) - Motilit
VFG013604 (hgbA) TonB-dependent hemoglobin/transferrin/lactoferrin family receptor [Hgp (VF0270) - Nutriti
VFG038349(gb|WP_005320751) (ascO) Type III secretion system protein AscO [T3SS (VF0479) - Effector deli
VFG000080(gb|NP_464522) (clpE) ATP-dependent protease [ClpE (VF0073) - Stress survival (VFC0282)] [List
VFG002280(gb|WP_011010361) (nagK) hyaluronidase [Mu-toxin (VF0389) - Exoenzyme (VFC0251)] [Clostrid
VFG000885(gb|WP_000265729) (papD) chaperone protein PapD [P fimbriae (VF0220) - Adherence (VFC0001)
VFG044165(gb|WP_001041793) (entS) enterobactin exporter, iron-regulated [Enterobactin (VF0228) - Nutrition
VFG010866(gb|WP_010946593) (ccmB) ABC transporter involved in cytochrome c biogenesis, CcmB subunit [
VFG000423(gb|WP_000052242) (vexE) Vi polysaccharide export protein VexE [Vi antigen (VF0101) - Immune
VFG001803(gb|WP_000608674) (seh) staphylococcal enterotoxin H precursor [SE (VF0020) - Exotoxin (VFC0
VFG011757(gb|WP_011117594) (virB4) Type IV secretion system protein VirB4 [Type IV secretion system (V
VFG041017(gb|NP_250350) (PA1659) hypothetical protein [HSI-2 (VF0943) - Effector delivery system (VFC0
VFG002499(gb|WP_004197164) (fliI) flagellum-specific ATP synthase FliI [Flagella (VF0430) - Motility (VFC
VFG038509(gb|WP_016350158) (flpL) Flp pilus pseudopilin [Flp type IV pili (VF0476) - Adherence (VFC0001
VFG038342(gb|WP_005320770) (ascV) Type III secretion system inner membrane export apparatus protein Asc
VFG049062(gb|WP_015875020) (rfbD) UDP-galactopyranose mutase [LPS (VF0561) - Immune modulation (V
VFG038380(gb|WP_043163044) (vipB) Type VI secretion system contractile sheath large subunit TssC/VipB [T
VFG038647(gb|WP_016350053) (fliK) flagellar hook-length control protein FliK [Polar flagella (VF0473) - Mot
VFG002369(gb|WP_011816869) (YE_RS15450) hypothetical protein [O-antigen (VF0392) - Immune modulatio
VFG001831(gb|WP_010946191) (icmS) Dot/Icm type IV secretion system chaperone IcmS [Dot/Icm (VF0156) -
VFG001364(gb|WP_001193686) (hysA) hyaluronidase [Hyaluronidase (VF0146) - Exoenzyme (VFC0251)] [Str
VFG001241(gb|NP_249774) (flgH) flagellar L-ring protein precursor FlgH [Flagella (VF0273) - Motility (VFC0
VFG043014(gb|NP_250149) (PA1458) two-component sensor [Flagella (VF0273) - Motility (VFC0204)] [Pseud
VFG001224(gb|NP_249087) (pilU) twitching motility protein PilU [Type IV pili (VF0082) - Adherence (VFC00
VFG000844(gb|WP_001034100) (espP) autotransporter, serine protease [EspP (VF0208) - Effector delivery syst
VFG043773(gb|YP_398516) (bont) botulinum neurotoxin, BoNT [BoNT (VF0375) - Exotoxin (VFC0235)] [Clo
VFG038389(gb|WP_011705718) (atsK) Type VI secretion system protein [T6SS (VF0480) - Effector delivery sy
VFG048841(gb|WP_014907232) (wcaJ) undecaprenyl-phosphate glucose phosphotransferase [Capsule (VF0560)
VFG038663(gb|WP_005298795) (fliO) flagellar biosynthesis protein, FliO [Polar flagella (VF0473) - Motility (V
VFG000364(gb|WP_000140406) (ybtA) transcriptional regulator YbtA [Yersiniabactin (VF0136) - Nutritional/M
VFG002322(gb|WP_011816590) (fliC) flagellin [Flagella (VF0394) - Motility (VFC0204)] [Yersinia enterocoliti
VFG048990(gb|WP_001741945) (galF) GalU regulator GalF [Capsule (VF0560) - Immune modulation (VFC025
VFG048297(gb|WP_014906961) (fimG) type 1 fimbrial minor component [Type I fimbriae (VF0566) - Adherenc
VFG001146(gb|CAA83902) (hifC) membrane anchor [Haemagglutinating pili (VF0036) - Adherence (VFC0001

VFG001355(gb|WP_000698569) (cbpD) choline binding protein D [Autolysin (VF0143) - Exoenzyme (VFC025
VFG002329(gb|WP_005160381) (fliG) flagellar motor switch protein G [Flagella (VF0394) - Motility (VFC0204
VFG049073(gb|WP_015875021) (KP1_RS17240) DUF4422 domain-containing protein [LPS (VF0561) - Immun
VFG001263(gb|NP_250143) (flhA) flagellar biosynthesis protein FlhA [Flagella (VF0273) - Motility (VFC0204)
VFG002335(gb|WP_005160396) (fliM) flagellar motor switch protein FliM [Flagella (VF0394) - Motility (VFC0
VFG013065(gb|WP_024259428) (shuT) periplasmic binding protein ShuT [Shu (VF0256) - Nutritional/Metaboli
VFG007174(gb|WP_011105906) (vscC) type III secretion system OM ring protein VscC [T3SS1 (VF0408) - Eff
VFG040545(gb|WP_011190491) (sca1) cell surface antigen Sca1 [Sca1 (VF0498) - Adherence (VFC0001)] [Ric
VFG049144(gb|WP_002892069) (acrB) acriflavine resistance protein B [AcrAB (VF0568) - Antimicrobial activi
VFG048409(gb|WP_086074289) (entA) 2,3-dihydroxybenzoate-2,3-dehydrogenase [Ent (VF0562) - Nutritional/
VFG048632(gb|WP_004176434) (vipA/tssB) type VI secretion system contractile sheath small subunit VipA [T6
VFG002385(gb|NP_218394) (eccD1) ESX-1 type VII secretion system EccD1 [ESX-1 (VF0315) - Effector deliv
VFG049152(gb|WP_000337350) (clbF) colibactin biosynthesis dehydrogenase ClbF [Colibactin (VF0573) - Exo
VFG041432(gb|WP_004683775) (vceC) type IV secretion system effector VceC [T4SS secreted effectors (VF06
VFG002056 (gspM) general secretion pathway protein M [T2SS (VF0333) - Effector delivery system (VFC0086)
VFG002177(gb|AAM75253) (cylA) cytolysin activator [Cytolysin (VF0356) - Exotoxin (VFC0235)] [Enterococ
VFG000935(gb|WP_001222189) (iroN) salmochelin receptor IroN [Salmochelin siderophore (VF0230) - Nutritic
VFG002521(gb|WP_004525693) (flhF) flagellar biosynthesis regulator FlhF [Flagella (VF0430) - Motility (VFC
VFG037987(gb|WP_000037963) (pseI) pseudaminic acid synthase [Capsule (VF0465) - Immune modulation (VF
VFG013260(gb|WP_010868968) (lgtC) glycosyltransferase [LOS (VF0044) - Immune modulation (VFC0258)] [
VFG002421(gb|WP_000792571) (sbi) IgG-binding protein SBI [Sbi (VF0423) - Immune modulation (VFC0258)
VFG000093(gb|WP_000742753) (tcpQ) toxin-coregulated pilus (TCP) outer membrane protein TcpQ [TCP (VF0
VFG001415(gb|NP_218004) (lipF) Probable esterase/lipase LipF [LipF (VF0307) - Nutritional/Metabolic factor
VFG049161(gb|WP_001029878) (clbO) colibactin polyketide synthase ClbO [Colibactin (VF0573) - Exotoxin (V
VFG049029(gb|WP_014907241) (KP1_RS17220) glycosyltransferase [LPS (VF0561) - Immune modulation (VF
VFG002174(gb|WP_002369251) (gelE) coccolysin [Gelatinase (VF0357) - Exoenzyme (VFC0251)] [Enterococc
VFG012191(gb|WP_011948511) (bont) botulinum neurotoxin, BoNT [BoNT (VF0375) - Exotoxin (VFC0235)] [
VFG043624(gb|WP_004149659) (mrkD) fimbrial adhesin protein precursor MrkD [Type 3 fimbriae (VF0567) -
VFG038587(gb|WP_016349160) (mshG) MSHA (mannose-sensitive hemagglutinin) pili biogenesis protein Msh
VFG038868(gb|WP_005314282) (lfiJ) lateral flagellar protein, LfiJ [Lateral flagella (VF0474) - Motility (VFC02
VFG002415(gb|WP_001131063) (yagX/ecpC) E. coli common pilus usher EcpC [ECP (VF0404) - Adherence (V
VFG042734(gb|NP_249343) (vfr) cAMP-regulatory protein [Type IV pili (VF0082) - Adherence (VFC0001)] [P
VFG002480(gb|WP_011205629) (tssH-5/clpV) Clp-type ATPase chaperone protein [T6SS-1 (VF0429) - Effecto
VFG011826(gb|YP_002343512) (fliK) flagellar hook-length control protein FliK [Flagella (VF0114) - Motility (
VFG002507(gb|WP_004534277) (flgA) flagellar basal body P-ring biosynthesis protein FlgA [Flagella (VF0430)
VFG038460(gb|WP_043119325) (tapQ) type IV pilus secretin PilQ [Tap type IV pili (VF0475) - Adherence (VF
VFG041237(gb|WP_010945887) (cegC2) Dot/Icm type IV secretion system effector CegC2 [Dot/Icm T4SS secre
VFG001447(gb|CAA50007) (cnf1) cytotoxic necrotizing factor 1 [CNF-1 (VF0240) - Exotoxin (VFC0235)] [Esc
VFG002244(gb|WP_011181141) (bepC) Bartonella effector protein BepC [VirB/VirD4 T4SS secreted effectors (
VFG000924(gb|WP_001234311) (fepB) ferrienterobactin ABC transporter periplasmic binding protein [Enteroba
VFG048985(gb|WP_014907219) (KP1_RS17355) phosphatase PAP2 family protein [Capsule (VF0560) - Immun
VFG002353(gb|WP_005168786) (flgM) negative regulator of flagellin synthesis [Flagella (VF0394) - Motility (V
VFG045467(gb|WP_011204972) (cdpA) cyclic di-GMP phosphodiesterase [CdpA (VF0432) - Regulation (VFC0
VFG001212(gb|NP_253246) (pilE) type 4 fimbrial biogenesis protein PilE [Type IV pili (VF0082) - Adherence (
VFG044443(gb|WP_001032773) (harA) haptoglobin-binding heme uptake protein HarA [Isd (VF0015) - Nutritic
VFG000998(gb|NP_858272) (mxiJ) type III secretion system inner MS ring protein MxiJ [TTSS (VF0118) - Effe

VFG002497(gb|WP_011204776) (fliK) flagellar hook-length control protein FliK [Flagella (VF0430) - Motility (
VFG011414(gb|WP_002964262) (kdsA) 2-dehydro-3-deoxyphosphooctonate aldolase [LPS (VF0367) - Immune
VFG038776(gb|WP_016351320) (flgJ) flagellar rod assembly protein/muramidase FlgJ [Polar flagella (VF0473)
VFG002495(gb|WP_004199882) (fliQ) flagellar biosynthesis protein FliQ [Flagella (VF0430) - Motility (VFC02
VFG050386(gb|WP_000355489.1) (pilT) type IV pilus twitching motility protein PilT [TFP (VF1334) - Adheren
VFG041304(gb|WP_010947678) (lirB) Dot/Icm type IV secretion system effector LirB [Dot/Icm T4SS secreted e
VFG038836(gb|WP_011707085) (pomA2) chemotaxis protein PomA [Polar flagella (VF0473) - Motility (VFC0
VFG050946(gb|WP_000083625.1) (tssL) type IVB secretion system protein TssL [T6SS (VF1337) - Effector del
VFG002380(gb|NP_218385) (eccA1) ESX-1 type VII secretion system AAA+ ATPase EccA1 [ESX-1 (VF0315)
VFG050757(gb|WP_001166319.1) (gspE1) general secretion pathway protein E [T2SS (VF1335) - Effector deliv
VFG042736(gb|NP_253152) (rpoN) RNA polymerase factor sigma-54 [Type IV pili (VF0082) - Adherence (VF
VFG041021(gb|NP_250354) (PA1663) transcriptional regulator [HSI-2 (VF0943) - Effector delivery system (VF
VFG037762(gb|WP_000078878) (lpxL) lauroyl acyltransferase [LPS (VF0466) - Immune modulation (VFC025
VFG038784(gb|WP_011706638) (flgH) flagellar basal body L-ring protein [Polar flagella (VF0473) - Motility (V
VFG048771(gb|WP_004190505) (tssF) type VI secretion system baseplate subunit TssF [T6SS (VF0569) - Effec
VFG000937(gb|WP_000750139) (iucD) L-lysine 6-monooxygenase IucD [Aerobactin (VF0229) - Nutritional/M
VFG001917(gb|YP_002344286) (flgL) flagellar hook-associated protein FlgL [Flagella (VF0114) - Motility (VF
VFG037807(gb|WP_000013434) (lpxB) lipid-A-disaccharide synthase [LPS (VF0466) - Immune modulation (V
VFG041012(gb|NP_250202) (PA1511) hypothetical protein [HSI-2 (VF0943) - Effector delivery system (VFC0
VFG000120(gb|NP_253949) (algZ) sigma factor AlgU [Alginate (VF0091) - Biofilm (VFC0271)] [Pseudomonas
VFG000939(gb|WP_001287497) (iucB) aerobactin siderophore biosynthesis protein IucB [Aerobactin (VF0229)
VFG048980(gb|WP_041937790) (KP1_RS17345) capsule assembly Wzi family protein [Capsule (VF0560) - Im
VFG049159(gb|WP_014907260) (clbM) precolibactin export MATE transporter ClbM [Colibactin (VF0573) - E
VFG000931(gb|WP_001336900) (entC) isochorismate synthase 1 [Enterobactin (VF0228) - Nutritional/Metaboli
VFG049904 (clpV/tssH) Type VI secretion system ATPase ClpV/TssH [T6SS (VF0579) - Effector delivery syste
VFG045523(gb|WP_010947178) (LPG_RS07260) Dot/Icm type IV secretion system effector [Dot/Icm T4SS sec
VFG045567(gb|WP_010948074) (LPG_RS11920) Dot/Icm type IV secretion system effector [Dot/Icm T4SS sec
VFG038885(gb|WP_005314224) (lafX) LafX [Lateral flagella (VF0474) - Motility (VFC0204)] [Aeromonas sal
VFG034859(gb|WP_010904544) (espX2) Type III secretion system effector EspX2 [TTSS secreted effectors (V
VFG041043(gb|NP_251061) (PA2371) ClpA/B-type protease [HSI-3 (VF0944) - Effector delivery system (VFC
VFG002409(gb|WP_000549274) (essC) type VII secretion system protein EssC, FtsK/SpoIIIE family ATPase [T
VFG001896(gb|YP_002343653) (fliI) flagellum-specific ATP synthase FliI [Flagella (VF0114) - Motility (VFC0
VFG045626(gb|WP_014748741) (scm) collagen adhesin protein Scm [Scm (VF0418) - Adherence (VFC0001)] [
VFG038845(gb|WP_016351249) (nueA) NeuA protein [Polar flagella (VF0473) - Motility (VFC0204)] [Aeromo
VFG001960(gb|YP_002344815) (Cj1432c) sugar transferase [Capsule (VF0323) - Immune modulation (VFC025
VFG038855(gb|WP_005314317) (lfiM) lateral flagellar motor switch protein, LfiM [Lateral flagella (VF0474) -
VFG038756(gb|WP_011706626) (flrC) transcriptional activator [Polar flagella (VF0473) - Motility (VFC0204)]
VFG045794(gb|WP_000992818) (csgC) curli assembly protein CsgC [Curli fibers (VF1138) - Adherence (VFC0
VFG002166(gb|WP_010714416) (ace) collagen adhesin protein [Ace (VF0355) - Adherence (VFC0001)] [Entero
VFG048307(gb|WP_041937815) (fimH) type 1 fimbrial adhesin precursor [Type I fimbriae (VF0566) - Adherenc
VFG044329(gb|WP_015874964) (irp1) yersiniabactin polyketide synthase HMWP1 [Ybt (VF0564) - Nutritional
VFG000165(gb|NP_252915) (pchF) pyochelin synthetase PchF [Pyochelin (VF0095) - Nutritional/Metabolic fac
VFG048620(gb|WP_014907843) (iutA) ferric aerobactin receptor IutA [Aerobactin (VF0565) - Nutritional/Meta
VFG038409(gb|WP_011704492) (fimA) fimbrial protein [Type I pili (VF0486) - Adherence (VFC0001)] [Aerom
VFG051070(gb|WP_001066523.1) (tssC) type VI secretion system contractile sheath large subunit [T6SS (VF13
VFG049122(gb|WP_012737539) (allC) allantoate amidohydrolase [Allantion utilization (VF0572) - Nutritional/

VFG002481(gb|WP_009940775) (vgrG-5) Type VI secretion system protein VgrG5 [T6SS-1 (VF0429) - Effecto
VFG000142(gb|NP_253698) (waaC) 3-deoxy-D-manno-octulosonic-acid (KDO) transferase [LPS (VF0085) - Im
VFG038261(gb|WP_016352107) (exeA) general secretion pathway protein A [Exe T2SS (VF0478) - Effector del
VFG038376(gb|WP_164927604) (atsD) Type VI secretion system protein [T6SS (VF0480) - Effector delivery sy
VFG001252(gb|NP_249793) (fliG) flagellar motor switch protein G [Flagella (VF0273) - Motility (VFC0204)] [
VFG000368(gb|WP_000703040) (ybtS) salicylate synthase Irp9 [Yersiniabactin (VF0136) - Nutritional/Metaboli
VFG049835(gb|WP_012076956) (exlA) hemolysin [Exolysin (VF0582) - Effector delivery system (VFC0086)] [
VFG000871(gb|WP_000790574) (fimB) Type 1 fimbriae Regulatory protein fimB [Type 1 fimbriae (VF0221) -
VFG050787(gb|WP_001112089.1) (gspF) general secretion pathway protein F [T2SS (VF1335) - Effector delive
VFG001405(gb|NP_217219) (sigA/rpoV) RNA polymerase sigma factor SigA [SigA (VF0257) - Regulation (VF
VFG000874(gb|WP_000824100) (fimI) Fimbrin-like protein fimI precursor [Type 1 fimbriae (VF0221) - Adhere
VFG005586(gb|WP_010922241) (hylA) hyaluronidase [Hyaluronidase (VF0246) - Exoenzyme (VFC0251)] [Str
VFG037288(gb|WP_000603876) (basG) acinetobactin biosynthesis protein BasF [Acinetobactin (VF0467) - Nutr
VFG035062(gb|WP_000951026) (nleH1) Type III secretion system effector NleH1 [TTSS secreted effectors (VF
VFG042568(gb|WP_000012016) (sfaY) transcriptional regulator SfaY [F1C fimbriae (VF0224) - Adherence (VF
VFG041329(gb|WP_010948104) (legL7) Dot/Icm type IV secretion system effector LegL7 [Dot/Icm T4SS secre
VFG050625(gb|WP_000389061.1) (pilG) twitching motility response regulator PilG [TFP (VF1334) - Adherence
VFG005009(gb|WP_001043551) (sec) staphylococcal enterotoxin C precursor [SE (VF0020) - Exotoxin (VFC02
VFG048348(gb|WP_004149658) (mrkJ) phosphodiesterase [Type 3 fimbriae (VF0567) - Biofilm (VFC0271)] [K
VFG038378(gb|WP_011705712) (vipA) Type VI secretion system contractile sheath small subunit TssB/VipA [T
VFG000889(gb|WP_000113351) (papF) P pilus minor subunit PapF [P fimbriae (VF0220) - Adherence (VFC00
VFG045563(gb|WP_010948017) (ceg28) Dot/Icm type IV secretion system effector Ceg28 [Dot/Icm T4SS secre
VFG038273(gb|WP_016349335) (exeD) general secretion pathway protein D [Exe T2SS (VF0478) - Effector del
VFG037482(gb|WP_004748062) (ACICU_RS04570) sigma-70 family RNA polymerase sigma factor [HemO clu
VFG038808(gb|WP_011706644) (flgB) flagellar basal-body rod protein FlgB [Polar flagella (VF0473) - Motility
VFG002427(gb|WP_004549962) (pilC) type IV pilus assembly protein PilC [Type IV pili (VF0431) - Adherence
VFG002198(gb|WP_003850266) (tox) Diphtheria toxin precursor [DT (VF0363) - Exotoxin (VFC0235)] [Coryn
VFG001261(gb|NP_250139) (fliR) flagellar biosynthetic protein FliR [Flagella (VF0273) - Motility (VFC0204)]
VFG044323(gb|WP_015874960) (ybtS) yersiniabactin biosynthesis salicylate synthase YbtS [Ybt (VF0564) - Nu
VFG038887(gb|WP_005314220) (lafF) LafF [Lateral flagella (VF0474) - Motility (VFC0204)] [Aeromonas salm
VFG000616(gb|NP_709455) (iucB) aerobactin synthesis protein IucB [Aerobactin (VF0123) - Nutritional/Metab
VFG000923(gb|WP_001034892) (fepA) ferrienterobactin outer membrane transporter [Enterobactin (VF0228) -
VFG000907(gb|WP_000376545) (hlyB) Hemolysin B [-Hemolysin (VF0225) - Exotoxin (VFC0235)] [Escherich
VFG038864(gb|WP_073531776) (fliF) flagellar MS-ring protein [Lateral flagella (VF0474) - Motility (VFC0204
VFG039991(gb|WP_011190506) (rvhB6b) VirB6-like protein of the type IV secretion system [Rvh T4SS (VF04
VFG048449(gb|WP_012737621) (fepB) iron-enterobactin transporter periplasmic binding protein [Ent (VF0562)
VFG001225(gb|NP_249099) (pilG) twitching motility protein PilG [Type IV pili (VF0082) - Adherence (VFC00
VFG038590(gb|WP_043118275) (mshE) ATPase protein MshE, GspE-like [MSHA type IV pili (VF0477) - Adh
VFG000918(gb|WP_001312167) (chuT) periplasmic heme-binding protein ChuT [Chu (VF0227) - Nutritional/M
VFG038679(gb|WP_016350055) (flhB) flagellar biosynthetic protein FlhB [Polar flagella (VF0473) - Motility (V
VFG049133(gb|WP_004177236) (acrA) acriflavine resistance protein A [AcrAB (VF0568) - Antimicrobial activi
VFG002416(gb|WP_001265657) (yagW/ecpD) polymerized tip adhesin of ECP fibers [ECP (VF0404) - Adheren
VFG049051(gb|WP_014907239) (KP1_RS17230) glycosyltransferase [LPS (VF0561) - Immune modulation (VF
VFG048528(gb|WP_004183335) (entD) enterochelin synthetase component D [Ent (VF0562) - Nutritional/Metab
VFG000944(gb|AAK16477) (draC) usher protein DraC [Dr adhesins (VF0223) - Adherence (VFC0001)] [Escher
VFG000919(gb|WP_001095619) (chuW) Putative oxygen independent coproporphyrinogen III oxidase [Chu (VF

VFG001269(gb|WP_010929996) (cyaB) cyclolysin secretion ATP-binding protein [Cya (VF0028) - Exotoxin (V
VFG000051(gb|WP_010930821) (bscJ) Bordetella Bsc type III secretion system protein BscJ [TTSS (VF0035) -
VFG048459(gb|WP_004147525) (entS) enterobactin exporter EntS [Ent (VF0562) - Nutritional/Metabolic factor
VFG002277(gb|WP_011009663) (nagH) hyaluronidase [Mu-toxin (VF0389) - Exoenzyme (VFC0251)] [Clostrid
VFG000114(gb|NP_253218) (xcpA/pilD) type 4 prepilin peptidase PilD [Type IV pili (VF0082) - Adherence (VI
VFG000908(gb|WP_000860472) (hlyD) Hemolysin D [-Hemolysin (VF0225) - Exotoxin (VFC0235)] [Escherich
VFG007953(gb|NP_217474) (Rv2958c) PGL/p-HBAD biosynthesis glycosyltransferase [PDIM (VF0309) - Imm
VFG043642(gb|WP_011205946) (MHP_RS00555) P110/LppT family adhesin N-terminal domain [P97/P102 par
VFG045791(gb|WP_000481500) (cgsD) transcriptional regulator CsgD [Curli fibers (VF1138) - Adherence (VF
VFG002278(gb|WP_011010095) (nagI) hyaluronidase [Mu-toxin (VF0389) - Exoenzyme (VFC0251)] [Clostridi
VFG002189(gb|WP_002359680) (cpsB/cdsA) phosphatidate cytidylyltransferase [Capsule (VF0361) - Immune m
VFG001862(gb|WP_015444101) (katB) catalase-peroxidase KatB [KatAB (VF0168) - Stress survival (VFC0282
VFG000881(gb|WP_000930062) (papB) regulatory protein PapB [P fimbriae (VF0220) - Adherence (VFC0001)
VFG034721(gb|WP_000556298) (ibeC) phosphoethanolamine transferase CptA [Ibes (VF0237) - Invasion (VFC
VFG002341(gb|WP_005168813) (flgL) flagellar hook-associated protein 3 FlgL [Flagella (VF0394) - Motility (V
VFG038860(gb|WP_005314298) (lfhB) lateral flagellar biosynthetic protein, LfhB [Lateral flagella (VF0474) - M
VFG043016(gb|NP_250155) (PA1464) purine-binding chemotaxis protein [Flagella (VF0273) - Motility (VFC02
VFG038269(gb|WP_075384415) (exeC) general secretion pathway protein C [Exe T2SS (VF0478) - Effector del
VFG000934(gb|WP_000348393) (entA) 2,3-dihydro-2,3-dihydroxybenzoate dehydrogenase EntA [Enterobactin
VFG038866(gb|WP_005314286) (lfiH) lateral flagellar assembly protein FliH [Lateral flagella (VF0474) - Motili
VFG013286(gb|WP_005694325) (galE) UDP-glucose 4-epimerase [LOS (VF0044) - Immune modulation (VFC0
VFG000121(gb|NP_249453) (algU) alginate biosynthesis protein AlgZ/FimS [Alginate (VF0091) - Biofilm (VFC
VFG034928(gb|WP_001239082) (lifA/efa1) lymphostatin Efa1/LifA [Lymphostatin/LifA (VF0175) - Effector de
VFG002338(gb|WP_042661456) (fliP) flagellar biosynthetic protein FliP [Flagella (VF0394) - Motility (VFC020
VFG041045(gb|NP_251063) (PA2373) hypothetical protein [HSI-3 (VF0944) - Effector delivery system (VFC00
VFG038391(gb|WP_011705719) (atsL) Type VI secretion system protein [T6SS (VF0480) - Effector delivery sy
VFG000911(gb|WP_000975445) (focC) F1C periplasmic chaperone [F1C fimbriae (VF0224) - Adherence (VFC
VFG000453(gb|NP_462538) (lpfD) long polar fimbrial protein LpfD [Lpf (VF0105) - Adherence (VFC0001)] [S
VFG047708(gb|WP_003022611) (carB) carbamoyl phosphate synthase large subunit [Pyrimidine biosynthesis (V
VFG038375(gb|WP_011705710) (atsC) Type VI secretion system protein [T6SS (VF0480) - Effector delivery sy
VFG002164(gb|WP_002395058) (prgB/asc10) aggregation substance PrgB/Asc10 [AS (VF0352) - Adherence (V
VFG038861(gb|WP_005314296) (lfhA) lateral flagellar biosynthesis protein [Lateral flagella (VF0474) - Motility
VFG007023(gb|WP_149591715) (rtxB) RTX toxin transporter RtxB [MARTX (VF0265) - Exotoxin (VFC0235)
VFG049007(gb|WP_002911561) (rcsA) transcriptional activator for ctr capsule biosynthesis [RcsAB (VF0571) -
VFG048654(gb|WP_004224345) (vasE/tssK) type VI secretion system baseplate subunit TssK [T6SS (VF0569) -
VFG000402(gb|WP_002212923) (yscC) type III secretion system secretin YscC [TTSS (VF0140) - Effector deliv
VFG038480(gb|WP_016349429) (tppB) type IV pilus pseudopilin [Tap type IV pili (VF0475) - Adherence (VFC
VFG000822(gb|WP_000605370) (glrR) type III secretion system LEE GrlA-binding negative regulator GrlR [TT
VFG002055(gb|WP_000097229) (gspL) general secretion pathway protein L [T2SS (VF0333) - Effector delivery
VFG038436(gb|WP_016352169) (tapC) type IV fimbrial assembly protein, PilC-like [Tap type IV pili (VF0475)
VFG037852(gb|WP_001075965) (ACICU_RS00395) polysaccharide biosynthesis tyrosine autokinase [Capsule (
VFG050328(gb|WP_000201227.1) (pilN) PilN domain-containing protein [TFP (VF1334) - Adherence (VFC000
VFG048439(gb|WP_014907941) (entC) isochorismate synthase [Ent (VF0562) - Nutritional/Metabolic factor (V
VFG038889(gb|WP_005314215) (lafT) LafT, flagella motor stator protein [Lateral flagella (VF0474) - Motility (
VFG050161(gb|WP_000579525) (kpsC) Capsule polysaccharide export protein [K1 capsule (VF0239) - Invasion
VFG038639(gb|WP_011705275) (fliI) flagellum-specific ATP synthase [Polar flagella (VF0473) - Motility (VFC

VFG001381(gb|YP_177728) (icl) Isocitrate lyase Icl (isocitrase) (isocitratase) [Isocitrate lyase (VF0253) - Others
VFG038549(gb|WP_016350148) (flpB) Flp pilus assembly protein [Flp type IV pili (VF0476) - Adherence (VFC
VFG012501(gb|WP_000271272) (iroE) esterase [Salmochelin siderophore (VF0230) - Nutritional/Metabolic fact
VFG001228(gb|NP_249102) (pilJ) twitching motility protein PilJ [Type IV pili (VF0082) - Adherence (VFC000
VFG045310(gb|WP_010946499) (LPG_RS03780) glycosyltransferase family 4 protein [LPS (VF0171) - Immun
VFG000075(gb|NP_463962) (inlA) Internalin A [InlA (VF0069) - Invasion (VFC0083)] [Listeria monocytogene
VFG038581(gb|WP_043118279) (mshB) MSHA (mannose-sensitive hemagglutinin) pili minor prepilin protein M
VFG000141(gb|NP_253675) (waaA) lipopolysaccharide core biosynthesis protein WaaP [LPS (VF0085) - Immu
VFG038521(gb|WP_016350155) (flpI) Flp pilus assembly protein FlpI [Flp type IV pili (VF0476) - Adherence (
VFG048874(gb|WP_041937789) (KP1_RS17305) GDP-L-fucose synthase [Capsule (VF0560) - Immune modula
VFG047111(gb|WP_003022291) (wbtH) asparagine synthase (glutamine-hydrolyzing) [LPS (VF0542) - Immune
VFG038353(gb|WP_005320743) (ascS) type III secretion system inner membrane export apparatus protein AscS
VFG006339(gb|WP_000716249) (babA/hopS) outer membrane protein adhesin [BabA (VF0053) - Adherence (V
VFG002348(gb|WP_005160443) (flgE) flagellar hook protein FlgE [Flagella (VF0394) - Motility (VFC0204)] [
VFG000636(gb|AAW31739) (set1B) ShET1B [ShET1 (VF0125) - Exotoxin (VFC0235)] [Shigella flexneri 2a str
VFG000112(gb|NP_253216) (pilB) type 4 fimbrial biogenesis protein PilB [Type IV pili (VF0082) - Adherence (
VFG038155(gb|WP_000209960) (ACICU_RS00500) phosphomannomutase/phosphoglucomutase [Capsule (VF
VFG038545(gb|WP_043118798) (flpC) Flp pilus secretin FlpC [Flp type IV pili (VF0476) - Adherence (VFC00
VFG050699(gb|WP_000840548.1) (pilR) sigma-54 dependent transcriptional regulator [TFP (VF1334) - Adhere
VFG038865(gb|WP_005314289) (fliG) flagellar motor switch protein G [Lateral flagella (VF0474) - Motility (V
VFG000053(gb|WP_010930823) (bcr4) Bordetella Bsc type III secretion system protein Bcr4 [TTSS (VF0035) -
VFG000115(gb|NP_249839) (toxA) exotoxin A precursor [ExoA (VF0086) - Exotoxin (VFC0235)] [Pseudomon
VFG001257(gb|NP_250135) (fliN) flagellar motor switch protein FliN [Flagella (VF0273) - Motility (VFC0204)
VFG038421(gb|WP_043118352) (fimE) fimbrial protein [Type I pili (VF0486) - Adherence (VFC0001)] [Aerom
VFG000360(gb|WP_002212799) (ybtT) type II thioesterase YbtT [Yersiniabactin (VF0136) - Nutritional/Metabo
VFG001245(gb|NP_249778) (flgL) flagellar hook-associated protein 3 FlgL [Flagella (VF0273) - Motility (VFC
VFG002281(gb|WP_003468301) (nagL) hyaluronidase [Mu-toxin (VF0389) - Exoenzyme (VFC0251)] [Clostrid
VFG038856(gb|WP_005314315) (lfiN) lateral flagellar motor switch protein, lfiN [Lateral flagella (VF0474) - M
VFG002193(gb|WP_002369249) (fsrB) agrBfs protein [Fsr (VF0360) - Biofilm (VFC0271)] [Enterococcus faeca
VFG038615(gb|WP_016349152) (mshI) MSHA (mannose-sensitive hemagglutinin) pili biogenesis protein MshI
VFG046118(gb|WP_011205945) (mhp107) P97 paralog, surface adhesin [P97/P102 paralog family (VF0426) - A
VFG001983(gb|YP_002344319) (pebA) bifunctional adhesin/ABC transporter aspartate/glutamate-binding protei
VFG000917(gb|WP_000089583) (chuA) Outer membrane heme/hemoglobin receptor ChuA [Chu (VF0227) - Nu
VFG038691(gb|WP_010634619) (flhG) CobQ/CobB/MinD/ParA family protein [Polar flagella (VF0473) - Moti
VFG012509(gb|WP_001111200) (iroC) ATP binding cassette transporter [Salmochelin siderophore (VF0230) - N
VFG000359(gb|WP_001088826) (ybtE) yersiniabactin siderophore biosynthetic protein [Yersiniabactin (VF013
VFG000347(gb|WP_010869038) (hgpC) hemoglobin-binding protein [Hgp (VF0270) - Nutritional/Metabolic fac
VFG038753(gb|WP_024944879) (motY) sodium-type flagellar protein MotY [Polar flagella (VF0473) - Motility
VFG038788(gb|WP_011706639) (flgG) flagellar basal body rod protein FlgG [Polar flagella (VF0473) - Motility
VFG000366(gb|WP_002212761) (ybtQ) yersiniabactin ABC transporter ATP-binding/permease protein YbtQ [Y
VFG038607(gb|WP_016349154) (mshJ) MSHA (mannose-sensitive hemagglutinin) pili biogenesis protein MshJ
VFG002450(gb|WP_004533397) (bipB) Type III secretion system translocator protein BipB [Bsa T3SS (VF042
VFG038867(gb|WP_005314284) (lfiI) lateral flagellar FliI-like assembly ATPase [Lateral flagella (VF0474) - M
VFG000261(gb|WP_002225480) (iga) IgA-specific serine endopeptidase [IgA1 protease (VF0080) - Effector del
VFG050414(gb|WP_002027454.1) (pilF) type IV pilus biogenesis/stability protein PilW [TFP (VF1334) - Adher
VFG050428(gb|WP_000128073.1) (fimV) hypothetical protein [TFP (VF1334) - Adherence (VFC0001)] [Acine

VFG037316(gb|WP_000744381) (entE) non-ribosomal peptide synthetase adenylate-forming enzyme of acinetob
VFG013063(gb|WP_001017208) (shuS) heme/hemoglobin transport protein ShuS [Shu (VF0256) - Nutritional/M
VFG002069(gb|NP_248773) (hsiB1/vipA/tssB) type VI secretion system tubule-forming protein VipA [HSI-I (V
VFG039686 (CBU_2059*) Coxiella Dot/Icm type IVB secretion system translocated effector [T4SS secreted effe
VFG048478(gb|WP_004147521) (fepG) iron-enterobactin transporter permease [Ent (VF0562) - Nutritional/Meta
VFG000920(gb|WP_000020034) (chuX) putative heme-binding protein ChuX [Chu (VF0227) - Nutritional/Meta
VFG006721(gb|NP_466343) (inlJ) internalin J [InlJ (VF0445) - Adherence (VFC0001)] [Listeria monocytogenes
VFG000878(gb|WP_000872015) (fimG) FimG protein precursor [Type 1 fimbriae (VF0221) - Adherence (VFC0
VFG038812(gb|WP_011706645) (cheR-3) chemotaxis protein CheR [Polar flagella (VF0473) - Motility (VFC02
VFG000065(gb|WP_010930832) (bscD) putative type III secretion protein [TTSS (VF0035) - Effector delivery s
VFG002531(gb|WP_011205291) (cheA) chemotaxis two-component sensor kinase CheA [Flagella (VF0430) - M
VFG038289(gb|WP_016349339) (exeH) general secretion pathway protein H [Exe T2SS (VF0478) - Effector del
VFG048554(gb|WP_004902400) (iroD) siderophore esterase IroD [Sal (VF0563) - Nutritional/Metabolic factor (
VFG000133(gb|NP_252241) (algA) phosphomannose isomerase / guanosine 5'-diphospho-D-mannose pyrophosp
VFG000895(gb|WP_000654937) (papC) usher protein PapC [P fimbriae (VF0220) - Adherence (VFC0001)] [Esc
VFG043627(gb|WP_000756959) (mrkA) type 3 fimbrial major pilin subunit MrkA [Type 3 fimbriae (VF0567) -
VFG002376(gb|WP_005167723) (rfbG) CDP-glucose 4,6-dehydratase [O-antigen (VF0392) - Immune modulatio
VFG048907(gb|WP_014907226) (KP1_RS17315) glycosyltransferase [Capsule (VF0560) - Immune modulation
VFG038875(gb|WP_005314261) (lfgF) lateral flagellar basal-body rod protein LfgF [Lateral flagella (VF0474) -
VFG038399(gb|WP_011705723) (atsP) Type VI secretion system protein [T6SS (VF0480) - Effector delivery sy
VFG014111(gb|NP_251837) (wbpJ) glycosyl transferase WbpJ [LPS (VF0085) - Immune modulation (VFC0258
VFG048277(gb|WP_009308599) (fimD) outer membrane usher protein [Type I fimbriae (VF0566) - Adherence
VFG037414(gb|WP_012391956) (bauD) ferric siderophore ABC transporter, permease protein BauD [Acinetoba
VFG041336(gb|WP_010948154) (ankF/legA14/ceg31) Dot/Icm type IV secretion system effector AnkF/LegA14
VFG038877(gb|WP_005314257) (lfgH) lateral flagellar L-ring protein, LfgH [Lateral flagella (VF0474) - Motilit
VFG000879(gb|WP_000832235) (fimH) FimH protein precursor [Type 1 fimbriae (VF0221) - Adherence (VFC0
VFG045531(gb|WP_010947388) (LPG_RS08320) Dot/Icm type IV secretion system effector [Dot/Icm T4SS sec
VFG038659(gb|WP_011705280) (fliN) flagellar motor switch protein FliN [Polar flagella (VF0473) - Motility (V
VFG001315(gb|WP_000222233) (hysA) hyaluronate lyase precursor [Hyaluronate lyase (VF0013) - Exoenzyme
VFG048643(gb|WP_014907727) (vipB/tssC) type VI secretion system contractile sheath large subunit VipB [T6
VFG002195(gb|WP_002399773) (EF0818) polysaccharide lyase, family 8 [Hyaluronidase (VF0359) - Exoenzym
VFG002432(gb|WP_004537540) (pilQ) type IV pilus biosynthesis protein [Type IV pili (VF0431) - Adherence (
VFG016472(gb|WP_011206327) (p216) protein P216, cilium adhesin p97 paralog [P97/P102 paralog family (VF
VFG001443(gb|AAF37887) (ompA) outer membrane protein A [OmpA (VF0236) - Invasion (VFC0083)] [Esche
VFG002046(gb|WP_000135089) (gspC) general secretion pathway protein C [T2SS (VF0333) - Effector delivery
VFG048687(gb|WP_002902160) (hcp/tssD) type VI secretion system protein, Hcp family [T6SS (VF0569) - Effe
VFG049906 (tssK) Type VI secretion system protein TssK [T6SS (VF0579) - Effector delivery system (VFC008
VFG001935(gb|YP_002344312) (ciaB) invasion antigen CiaB [CiaB (VF0324) - Invasion (VFC0083)] [Campylo
VFG010686(gb|WP_010948184) (sdbB) Dot/Icm type IV secretion system effector SdbB [Dot/Icm T4SS secrete
VFG000861(gb|WP_001045649) (pic) Pic serine protease precursor, autotransporter [Pic (VF0218) - Effector del
VFG045336(gb|AHK25021) (llsY) LlsY [LLS (VF0410) - Exotoxin (VFC0235)] [Listeria innocua SLCC6294]
VFG002171(gb|WP_011109430) (EF0149) aggregation substance [AS (VF0352) - Adherence (VFC0001)] [Ente
VFG012609(gb|WP_000993317) (chuW) Putative oxygen independent coproporphyrinogen III oxidase [Chu (VF
VFG014369(gb|NP_249789) (fleS) two-component sensor [Flagella (VF0273) - Motility (VFC0204)] [Pseudomo
VFG037573 (csuA) Csu pilus subunit CsuA [Csu fimbriae (VF0461) - Biofilm (VFC0271)] [Acinetobacter baum
VFG038322(gb|WP_180975055) (ascJ) type III secretion system OM ring protein AscJ [T3SS (VF0479) - Effect

VFG038667(gb|WP_016350054) (fliP) flagellar biosynthesis protein FliP [Polar flagella (VF0473) - Motility (VF
VFG001958(gb|YP_002344813) (rfbC) dTDP-4-dehydrorhamnose 3,5-epimerase [Capsule (VF0323) - Immune r
VFG038452(gb|WP_016351631) (tapO) type IV pilus biogenesis protein PilO [Tap type IV pili (VF0475) - Adhe
VFG051131(gb|WP_006191504.1) (bmaC) autotransporter outer membrane beta-barrel domain-containing protei
VFG000122(gb|NP_252230) (algD) GDP-mannose 6-dehydrogenase AlgD [Alginate (VF0091) - Biofilm (VFC0
VFG038715(gb|WP_024943899) (motA) chemotaxis protein MotA [Polar flagella (VF0473) - Motility (VFC020
VFG049905(gb|WP_000343289) (tssL) Type VI secretion system protein TssL [T6SS (VF0579) - Effector delive
VFG010759(gb|WP_010948114) (vpdA/vipD2) Dot/Icm type IV secretion system effector VpdA [Dot/Icm T4SS
VFG002172(gb|WP_010710127) (EF0485) aggregation substance [AS (VF0352) - Adherence (VFC0001)] [Ente
VFG043015(gb|NP_250150) (PA1459) chemotaxis-specific methylesterase [Flagella (VF0273) - Motility (VFC0
VFG039131(gb|NP_820609) (icmE/dotG) Type IVB secretion system protein IcmE/DotG, inner membrane prote
VFG012162(gb|WP_011590241) (nanJ) exo-alpha-sialidase [Sialidase (VF0391) - Exoenzyme (VFC0251)] [Clos
VFG037560(gb|WP_000790104) (csuA/B) Csu pilus major pilin subunit CsuA/B [Csu fimbriae (VF0461) - Biofi
VFG005486(gb|WP_000417180) (iga) G5 domain-containing protein [IgA1 protease (VF0147) - Immune modula
VFG050157(gb|WP_001331698) (kpsF) Polysialic acid capsule expression protein [K1 capsule (VF0239) - Invas
VFG002088(gb|WP_000225015) (icmF/vasK) type VI secretion system inner membrane protein VasK [VAS T6
VFG044327(gb|WP_000140406) (ybtA) yersiniabactin transcriptional regulator YbtA [Ybt (VF0564) - Nutrition
VFG034826(gb|WP_000484292) (espR3) Type III secretion system effector espR3 [TTSS secreted effectors (VF
VFG038772(gb|WP_016351319) (flgK) flagellar hook-associated protein FlgK [Polar flagella (VF0473) - Motilit
VFG001937(gb|YP_002344529) (Cj1136) glycosyltransferase [LOS (VF0326) - Immune modulation (VFC0258)
VFG001254(gb|NP_249795) (fliI) flagellum-specific ATP synthase FliI [Flagella (VF0273) - Motility (VFC0204
VFG000015(gb|WP_010931651) (ptxC) pertussis toxin subunit 3 precursor [Ptx (VF0026) - Exotoxin (VFC0235
VFG001231(gb|NP_249104) (chpA) still frameshift probable component of chemotactic signal transduction syste
VFG000922(gb|WP_000910402) (chuU) heme permease protein ChuU [Chu (VF0227) - Nutritional/Metabolic fa
VFG002049(gb|WP_001173470) (gspF) general secretion pathway protein F [T2SS (VF0333) - Effector delivery
VFG007959(gb|NP_217478) (Rv2962c) PGL/p-HBAD biosynthesis rhamnosyltransferase [PDIM (VF0309) - Im
VFG001966(gb|YP_002344821) (Cj1438c) sugar transferase [Capsule (VF0323) - Immune modulation (VFC025
VFG001142(gb|WP_042960771) (hopZ) outer membrane protein adhesin [HopZ (VF0054) - Adherence (VFC00
VFG038824(gb|WP_011706648) (flgM) anti-sigma-D factor [Polar flagella (VF0473) - Motility (VFC0204)] [Ae
VFG044197(gb|WP_003022882) (fslC) diaminopimelate decarboxylase [Rhizoferrin (VF0551) - Nutritional/Met
VFG047777(gb|WP_003022126) (pdpB) Type VI secretion system TssM homolog [T6SS (VF0544) - Effector de
VFG049164(gb|WP_004175305) (clbS) colibactin self-protection protein ClbS [Colibactin (VF0573) - Exotoxin
VFG048468(gb|WP_004176917) (fepD) iron-enterobactin transporter membrane protein [Ent (VF0562) - Nutritic
VFG035929(gb|WP_014639418) (aatD) AatD [Dispersin (VF0215) - Others (VFC0346)] [Escherichia coli O44:
VFG041091(gb|NP_459264) (STM0267) sciB domain protein [SCI (Salmonella centrisome island) (VF0974) - E
VFG038405(gb|WP_011705727) (AHA_RS09305) type VI secretion system PAAR protein [T6SS (VF0480) - E
VFG001916(gb|YP_002344282) (flhA) flagellar biosynthesis protein FlhA [Flagella (VF0114) - Motility (VFC0
VFG000903(gb|WP_001045652) (pic) Pic serine protease precursor, autotransporter [Pic (VF0232) - Effector del
VFG049154(gb|WP_014907257) (clbH) colibactin non-ribosomal peptide synthetase ClbH [Colibactin (VF0573)
VFG001015(gb|NP_858289) (spa40) type III secretion system export apparatus switch protein Spa40 [TTSS (VF
VFG038675(gb|WP_011705283) (fliR) flagellar biosynthesis protein FliR [Polar flagella (VF0473) - Motility (V
VFG001967(gb|YP_002344822) (glf) UDP-galactopyranose mutase [Capsule (VF0323) - Immune modulation (V
VFG037274(gb|WP_001281538) (barA) siderophore efflux system of the ABC superfamily [Acinetobactin (VF0
VFG049917(gb|WP_000508696) (rhs/PAAR) Type VI secretion system protein, PAAR family [T6SS (VF0579)
VFG044328(gb|WP_015874963) (irp2) yersiniabactin non-ribosomal peptide synthetase HMWP2 [Ybt (VF0564)
VFG000358(gb|WP_002212885) (fyuA/psn) pesticin/yersiniabactin receptor protein [Yersiniabactin (VF0136) -

VFG000630(gb|NP_708742) (sigA) serine protease, autotransporter SigA [SigA (VF0119) - Exoenzyme (VFC02
VFG050484(gb|WP_001152285.1) (gspO/pilD) A24 family peptidase [TFP (VF1334) - Adherence (VFC0001)] [
VFG038397(gb|WP_011705722) (vasH) sigma-54 dependent transcriptional regulator [T6SS (VF0480) - Effecto
VFG000139(gb|NP_253697) (waaG) B-band O-antigen polymerase [LPS (VF0085) - Immune modulation (VFC
VFG001408(gb|NP_217456) (mas) multifunctional mycocerosic acid synthase [PDIM (VF0309) - Immune modu
VFG002083(gb|WP_000083409) (vasF) type VI secretion system protein VasF [VAS T6SS (VF0335) - Effector
VFG038345(gb|WP_005320764) (acr2) chaperone protein [T3SS (VF0479) - Effector delivery system (VFC008
VFG038655(gb|WP_011705279) (fliM) flagellar motor switch protein FliM [Polar flagella (VF0473) - Motility (
VFG002414(gb|WP_000730972) (yagZ/ecpA) E. coli common pilus structural subunit EcpA [ECP (VF0404) - A
VFG037028(gb|WP_002218620) (katA) catalase [KatA (VF0454) - Stress survival (VFC0282)] [Neisseria menin
VFG000163(gb|NP_251088) (fpvA) ferripyoverdine receptor FpvA [Pyoverdine (VF0094) - Nutritional/Metabol
VFG041323(gb|WP_010947957) (lem21) Dot/Icm type IV secretion system effector Lem21 [Dot/Icm T4SS secr
VFG002065(gb|NP_248769) (hsiJ1) type VI secretion system hcp secretion island protein HsiJ1 [HSI-I (VF0334
VFG001988(gb|YP_002344798) (cysC) adenylyl-sulfate kinase [Capsule (VF0323) - Immune modulation (VFC
VFG002184(gb|WP_002359685) (cpsG) MurB family protein [Capsule (VF0361) - Immune modulation (VFC02
VFG000064(gb|WP_010930831) (bcrD) Type III secretion system LcrD homolog protein BcrD [TTSS (VF0035)
VFG007382(gb|WP_000907265) (flgM) negative regulator of flagellin synthesis FlgM, putative [Flagella (VF05
VFG005197(gb|WP_000006706) (pavA) Fibronectin-binding protein-like protein A [PavA (VF0283) - Adherenc
VFG000916(gb|WP_001017192) (chuS) heme oxygenase ChuS [Chu (VF0227) - Nutritional/Metabolic factor (V
VFG050912(gb|WP_000898330.1) (vgrG/tssI) type VI secretion system tip protein VgrG [T6SS (VF1337) - Effe
VFG049120(gb|WP_012737535) (allB) allantoinase [Allantion utilization (VF0572) - Nutritional/Metabolic fact
VFG049155(gb|WP_000829570) (clbI) colibactin polyketide synthase ClbI [Colibactin (VF0573) - Exotoxin (VF
VFG048229(gb|WP_015875142) (fimB) tyrosine recombinase [Type I fimbriae (VF0566) - Adherence (VFC000
VFG037118(gb|WP_002247552) (recN) DNA repair protein RecN [RecN (VF0457) - Stress survival (VFC0282)
VFG046214(gb|NP_819037) (CBUA0020) Coxiella Dot/Icm type IVB secretion system translocated effector [T
VFG038719(gb|WP_016350061) (pomB) ompA family protein [Polar flagella (VF0473) - Motility (VFC0204)] [
VFG000161(gb|NP_251089) (pvdD) pyoverdine synthetase D [Pyoverdine (VF0094) - Nutritional/Metabolic fac
VFG038627(gb|WP_016350050) (fliF) flagellar MS-ring protein [Polar flagella (VF0473) - Motility (VFC0204)]
VFG045481(gb|WP_010945891) (LPG_RS00665) hypothetical protein [Dot/Icm T4SS secreted effectors (VF07
VFG038723(gb|WP_011705294) (AHML_RS07540) CobQ/CobB/MinD/ParA family protein [Polar flagella (VF
VFG033511(gb|WP_000803678) (sepQ/escQ) Type III secetion system C ring protein EscQ [TTSS (VF0172) - E
VFG051042(gb|WP_000653195.1) (hcp/tssD) type VI secretion system tube protein Hcp [T6SS (VF1337) - Effe
VFG002179(gb|AAD09858) (esp) Enterococcal surface protein; Esp [Esp (VF0353) - Adherence (VFC0001)] [E
VFG038913 (rtxA) multifunctional autoprocessing repeats-in-toxins (MARTX) toxin [RtxA (VF0482) - Exotoxi
VFG000180(gb|NP_251791) (xcpT) general secretion pathway protein G [Xcp secretion system (VF0084) - Effe
VFG042739(gb|NP_254019) (crc) catabolite repression control protein [Type IV pili (VF0082) - Adherence (VF
VFG049040(gb|WP_074160401) (KP1_RS17225) glycosyltransferase family 4 protein [LPS (VF0561) - Immune
VFG048488(gb|WP_002893737) (fepC) iron-enterobactin transporter ATP-binding protein [Ent (VF0562) - Nutr
VFG038848(gb|WP_016350360) (flmD) FlmD protein [Polar flagella (VF0473) - Motility (VFC0204)] [Aeromo
VFG038576(gb|WP_016349164) (mshC) MSHA (mannose-sensitive hemagglutinin) pili minor prepilin protein M
VFG038568(gb|WP_016349166) (mshO) MSHA (mannose-sensitive hemagglutinin) pili biogenesis protein Msh
VFG044326(gb|WP_001327262) (ybtP) yersiniabactin ABC transporter ATP-binding/permease protein YbtP [Y
VFG038501(gb|WP_043119142) (tapV) type IV pilus assembly protein TapV [Tap type IV pili (VF0475) - Adhe
VFG002286(gb|WP_003470007) (cpe) enterotoxin Cpe [CPE (VF0383) - Exotoxin (VFC0235)] [Clostridium pe
VFG013948(gb|NP_251650) (pilZ) type 4 fimbrial biogenesis protein PilZ [Type IV pili (VF0082) - Adherence (
VFG049910(gb|WP_000348788) (tssG) Type VI secretion system protein TssG [T6SS (VF0579) - Effector deliv

VFG051123(gb|WP_032449063.1) (bmaA) autotransporter outer membrane beta-barrel domain-containing protei
VFG000615(gb|NP_709454) (iucA) aerobactin synthesis protein IucA [Aerobactin (VF0123) - Nutritional/Metab
VFG050118(gb|WP_000291846) (cesH) hydrolase, alpha/beta superfamily, CesH [Cereulide (VF0583) - Exotoxi
VFG000477(gb|NP_461845) (rpoS) RNA polymerase sigma factor RpoS [RpoS (VF0112) - Regulation (VFC030
VFG048267(gb|WP_015875145) (fimC) periplasmic chaperone [Type I fimbriae (VF0566) - Adherence (VFC00
VFG000365(gb|WP_002214412) (ybtP) yersiniabactin ABC transporter ATP-binding/permease protein YbtP [Ye
VFG001932(gb|YP_002343539) (cdtC) cytolethal distending toxin C [CDT (VF0115) - Exotoxin (VFC0235)] [C
VFG000876(gb|WP_000120946) (fimD) Outer membrane usher protein fimD precursor [Type 1 fimbriae (VF022
VFG045510(gb|WP_010946845) (ravN) Dot/Icm type IV secretion system effector RavN [Dot/Icm T4SS secrete
VFG000804(gb|WP_000098793) (cesT) multieffector chaperone [TTSS (VF0191) - Effector delivery system (VF
VFG012597(gb|WP_000089593) (chuA) Outer membrane heme/hemoglobin receptor ChuA [Chu (VF0234) - Nu
VFG012591(gb|WP_001017167) (chuS) heme oxygenase ChuS [Chu (VF0234) - Nutritional/Metabolic factor (V
VFG002366(gb|WP_005167739) (YE_RS15435) glycosyltransferase family 4 protein [O-antigen (VF0392) - Imm
VFG034636(gb|WP_000358674) (cfaC) colonisation factor antigen c, usher [Adhesive fimbriae (VF0213) - Adhe
VFG041020(gb|NP_250353) (PA1662) ClpA/B-type protease [HSI-2 (VF0943) - Effector delivery system (VFC
VFG001266(gb|NP_252912) (pchI) ABC transporter ATP-binding protein [Pyochelin (VF0095) - Nutritional/Me
VFG045325(gb|WP_010946516) (LPG_RS03855) glycosyltransferase family 2 protein [LPS (VF0171) - Immune
VFG000111(gb|NP_253215) (pilA) type 4 fimbrial precursor PilA [Type IV pili (VF0082) - Adherence (VFC000
VFG038611(gb|WP_016349153) (mshI1) MSHA (mannose-sensitive hemagglutinin) pili biogenesis protein Msh
VFG000361(gb|WP_000982866) (ybtU) yersiniabactin biosynthetic protein YbtU [Yersiniabactin (VF0136) - Nu
VFG014473(gb|NP_250132) (fliK) flagellar hook-length control protein FliK [Flagella (VF0273) - Motility (VFC
VFG001234(gb|NP_249107) (chpD) probable transcriptional regulator [Type IV pili (VF0082) - Adherence (VFC
VFG049914(gb|WP_000037389) (tssB) Type VI secretion system protein TssB [T6SS (VF0579) - Effector delive
VFG038313(gb|WP_043118380) (exeN) general secretion pathway protein N [Exe T2SS (VF0478) - Effector del
VFG045413(gb|WP_133121395) (APH_RS06575*) hypothetical protein [P44/Msp2 family (VF0520) - Immune
VFG041337(gb|WP_010948158) (ankD/legA15) Dot/Icm type IV secretion system effector AnkD/LegA15 [Dot/
VFG002306(gb|NP_461449) (ratB) putative outer membrane protein [RatB (VF0399) - Adherence (VFC0001)] [
VFG002288(gb|WP_009902069) (toxB) toxin B [TcdB (VF0377) - Exotoxin (VFC0235)] [Clostridium difficile 6
VFG015000(gb|NP_252339) (mucP) metalloprotease protease [Alginate (VF0091) - Biofilm (VFC0271)] [Pseud
VFG014099(gb|NP_251849) (wbpA) UDP-N-acetyl-d-glucosamine 6-dehydrogenase [LPS (VF0085) - Immune r
VFG049909(gb|WP_001113715) (fha) Type VI secretion system protein Fha [T6SS (VF0579) - Effector delivery
VFG044331(gb|WP_015874966) (ybtT) type II thioesterase YbtT [Ybt (VF0564) - Nutritional/Metabolic factor (
VFG048518(gb|WP_012737617) (fepA) outer membrane receptor FepA [Ent (VF0562) - Nutritional/Metabolic fa
VFG002522(gb|WP_004198639) (flhA) flagellar biosynthesis protein FlhA [Flagella (VF0430) - Motility (VFC0
VFG045319(gb|WP_010946509) (LPG_RS03825) ABC transporter permease [LPS (VF0171) - Immune modulat
VFG034679(gb|WP_000074207) (ibeB) Cu(+)/Ag(+) efflux RND transporter outer membrane channel CusC [Ibe
VFG050163(gb|WP_000197083) (neuE) polysialic acid biosynthesis protein [K1 capsule (VF0239) - Invasion (V
VFG000143(gb|NP_253699) (waaF) heptosyltransferase I [LPS (VF0085) - Immune modulation (VFC0258)] [Ps
VFG001242(gb|NP_249775) (flgI) flagellar P-ring protein precursor FlgI [Flagella (VF0273) - Motility (VFC020
VFG012513(gb|WP_001221122) (iroB) glucosyltransferase IroB [Salmochelin siderophore (VF0230) - Nutrition
VFG000884(gb|WP_000654937) (papC) usher protein PapC [P fimbriae (VF0220) - Adherence (VFC0001)] [Esc
VFG016041(gb|NP_251092) (pvdI) peptide synthase [Pyoverdine (VF0094) - Nutritional/Metabolic factor (VFC
VFG041096(gb|NP_459269) (STM0272) ClpV1 family T6SS ATPase [SCI (Salmonella centrisome island) (VF0
VFG049084(gb|WP_004175264) (rfbB) O-antigen export ABC transporter ATP-binding protein RfbB [LPS (VF
VFG038361(gb|WP_005320818) (ati1) Ati2 chaperone [T3SS (VF0479) - Effector delivery system (VFC0086)] [
VFG051098(gb|WP_002001416.1) (clpV/tssH) type VI secretion system ATPase TssH [T6SS (VF1337) - Effecto

VFG041039(gb|NP_251057) (PA2367) hypothetical protein [HSI-3 (VF0944) - Effector delivery system (VFC00
VFG006217 (opa) outer membrane beta-barrel protein [Opa (VF0076) - Invasion (VFC0083)] [Neisseria meningi
VFG041038(gb|NP_251056) (PA2366) uricase [HSI-3 (VF0944) - Effector delivery system (VFC0086)] [Pseudo
VFG043623(gb|WP_002916122) (mrkF) type 3 fimbrial minor pilin subunit MrkF [Type 3 fimbriae (VF0567) - I
VFG039464(gb|NP_820359) (CBU_1370) Coxiella Dot/Icm type IVB secretion system translocated effector [T4
VFG000927(gb|WP_000096764) (fepE) LPS O-antigen length regulator [Enterobactin (VF0228) - Nutritional/Me
VFG002437(gb|WP_011325557) (boaB) autotransporter protein [BoaB (VF0435) - Adherence (VFC0001)] [Burl
VFG038144(gb|WP_001062908) (galE) UDP-glucose 4-epimerase GalE [Capsule (VF0465) - Immune modulatio
VFG001814(gb|NP_215678) (narH) nitrate reductase subunit beta [Nitrate reductase (VF0302) - Nutritional/Meta
VFG005775(gb|WP_000403526) (cylA) ABC (ATP-binding cassette) transporter CylA [-haemolysin/cytolysin (V
VFG000831(gb|WP_000628731) (escE) chaperone for EscF [TTSS (VF0191) - Effector delivery system (VFC00
VFG012343(gb|WP_001234182) (sfaH) S fimbriae adhesin SfaH [S fimbriae (VF0222) - Adherence (VFC0001)]
VFG046519(gb|WP_003020038) (dipA) bacterial surface protein, sel1 repeat family protein [DipA (VF0545) - O
VFG000229 (pilS) pilin [Type IV pili (VF0075) - Adherence (VFC0001)] [Neisseria meningitidis MC58]
VFG002449(gb|WP_004203135) (bipC) Type III secretion system translocator protein BipC [Bsa T3SS (VF0428
VFG000938(gb|WP_001015709) (iucC) aerobactin siderophore biosynthesis protein IucC [Aerobactin (VF0229)
VFG038407(gb|WP_011705728) (vgrG3) VgrG protein [T6SS (VF0480) - Effector delivery system (VFC0086)]
VFG000009(gb|WP_050849356) (fimD) fimbrial adhesin [Fimbriae (VF0031) - Adherence (VFC0001)] [Bordete
VFG039425(gb|NP_820063) (cem4) Coxiella Dot/Icm type IVB secretion system translocated effector [T4SS se
VFG048338(gb|WP_002916079) (mrkI) LuxR family regulatory protein [Type 3 fimbriae (VF0567) - Biofilm (V
VFG038643(gb|WP_011705276) (fliJ) flagellar export protein FliJ [Polar flagella (VF0473) - Motility (VFC0204
VFG034915(gb|WP_000225947) (espY4) Type III secretion system effector EspY4 [TTSS secreted effectors (VF
VFG038285(gb|WP_005308483) (exeG) general secretion pathway protein G [Exe T2SS (VF0478) - Effector del
VFG048787(gb|WP_004176417) (sciN/tssJ) type VI secretion system lipoprotein TssJ [T6SS (VF0569) - Effecto
VFG016038(gb|NP_251090) (pvdJ) pyoverdine biosynthesis protein PvdJ [Pyoverdine (VF0094) - Nutritional/M
VFG048606(gb|WP_004213922) (iucC) aerobactin siderophore biosynthesis protein IucC [Aerobactin (VF0565)
VFG044330(gb|WP_032447695) (ybtU) yersiniabactin biosynthesis oxidoreductase YbtU [Ybt (VF0564) - Nutri
VFG049151(gb|WP_001297917) (clbE) colibactin biosynthesis aminomalonyl-acyl carrier protein ClbE [Colibac
VFG048830(gb|WP_014907233) (gndA) NADP-dependent phosphogluconate dehydrogenase [Capsule (VF0560
VFG000862(gb|WP_014639426) (pet) Pet serine protease precursor, autotransporter [Pet (VF0217) - Effector del
VFG038703(gb|WP_011705289) (cheZ) chemotaxis protein CheZ [Polar flagella (VF0473) - Motility (VFC0204
VFG048508(gb|WP_012737618) (fes) enterobactin/ferric enterobactin esterase [Ent (VF0562) - Nutritional/Meta
VFG043626(gb|WP_000820818) (mrkB) fimbrial chaperone protein mrkB precursor [Type 3 fimbriae (VF0567)
VFG000127(gb|NP_252235) (algG) outer membrane protein AlgE [Alginate (VF0091) - Biofilm (VFC0271)] [Ps
VFG000227(gb|WP_002221783) (pilS) pilin [Type IV pili (VF0075) - Adherence (VFC0001)] [Neisseria mening
VFG044322(gb|WP_004190148) (iroE) siderophore esterase IroE [Sal (VF0563) - Nutritional/Metabolic factor (V
VFG034873(gb|WP_001270084) (espX5) Type III secretion system effector EspX5 [TTSS secreted effectors (VF
VFG045307(gb|WP_025862603) (pgi) glucose-6-phosphate isomerase [LPS (VF0171) - Immune modulation (VF
VFG049162(gb|WP_002430641) (clbP) precolibactin peptidase ClbP [Colibactin (VF0573) - Exotoxin (VFC023
VFG000415(gb|WP_002213290) (yopO/ypkA) type III secretion system effector YopO, regulating actin polymer
VFG002382(gb|NP_218387) (eccCa1) ESX-1 type VII secretion system FtsK/SpoIIIE family protein EccCa1 [ES
VFG002152(gb|NP_220384) (scc3) chaperone [TTSS (VF0344) - Effector delivery system (VFC0086)] [Chlamy
VFG002342(gb|WP_011816603) (flgK) flagellar hook-associated protein 1 FlgK [Flagella (VF0394) - Motility (V
VFG000278(gb|WP_000946389) (flgE_1) flagellar hook protein [Flagella (VF0051) - Motility (VFC0204)] [Heli
VFG045789(gb|WP_001264082) (cgsF) curli production assembly/transport protein CsgF [Curli fibers (VF1138)
VFG042978(gb|WP_011109477) (ebpC) endocarditis and biofilm-associated pilus major subunit EbpC [Ebp pili

VFG038564(gb|WP_016349167) (mshP) hypothetical protein [MSHA type IV pili (VF0477) - Adherence (VFC0
VFG038760(gb|WP_016351312) (flrB) two-component sensor kinase [Polar flagella (VF0473) - Motility (VFC0
VFG001251(gb|NP_249792) (fliF) flagellar M-ring protein FliF [Flagella (VF0273) - Motility (VFC0204)] [Pseu
VFG013292(gb|WP_105163174) (lic3A) alpha-2,3-sialyltransferase [LOS (VF0044) - Immune modulation (VFC
VFG050267(gb|NP_220230) (CT_711) hypothetical protein [TTSS secreted effectors (VF0711) - Effector delive
VFG038561(gb|WP_016349168) (mshQ) MSHA (mannose-sensitive hemagglutinin) pili biogenesis protein Msh
VFG001218(gb|NP_253727) (pilQ) type 4 fimbrial biogenesis protein PilQ [Type IV pili (VF0082) - Adherence
VFG037735(gb|WP_000633124) (adeH) outer membrane protein [AdeFGH efflux pump (VF0504) - Biofilm (VF
VFG002406(gb|WP_000728972) (esaA) type VII secretion system protein EsaA [Type VII secretion system (VF
VFG000027(gb|WP_010931506) (brkA) serum resistance protein [Brk (VF0034) - Immune modulation (VFC025
VFG034772(gb|WP_000627910) (espL4) Type III secretion system effector EspL4 [TTSS secreted effectors (VF
VFG035902(gb|WP_000209220) (aatP) permease AatP [Dispersin (VF0215) - Others (VFC0346)] [Escherichia c
VFG000940(gb|WP_001299620) (iucA) aerobactin siderophore biosynthesis protein IucD [Aerobactin (VF0229)
VFG002196(gb|WP_011109578) (EF3023) polysaccharide lyase, family 8 [Hyaluronidase (VF0359) - Exoenzym
VFG049148(gb|WP_041168941) (clbB) colibactin hybrid non-ribosomal peptide synthetase/type I polyketide syn
VFG037442(gb|WP_000910253) (basA) acinetobactin biosynthesis protein [Acinetobactin (VF0467) - Nutritiona
VFG048951(gb|WP_015875028) (KP1_RS17330) polysaccharide biosynthesis tyrosine autokinase [Capsule (VF
VFG046119(gb|WP_011206220) (mhp385) P97 paralog, surface adhesin [P97/P102 paralog family (VF0426) - A
VFG048609(gb|WP_004213923) (iucD) lysine 6-monooxygenase IucD [Aerobactin (VF0565) - Nutritional/Meta
VFG050165(gb|WP_001259305) (neuA) N-acylneuraminate cytidylyltransferase [K1 capsule (VF0239) - Invasio
VFG045474(gb|AAM75251) (cylM) CylM; cytolysin subunit modifier [Cytolysin (VF0356) - Exotoxin (VFC023
VFG042218(gb|AAN07916) (cif) Type III secretion system effector Cif, cyclomodulin [TTSS secreted effectors (
VFG000902(gb|WP_001034089) (sat) Aecreted auto transpoter toxin [Sat (VF0231) - Effector delivery system (V
VFG034815(gb|WP_000671689) (espR1) Type III secretion system effector espR1 [TTSS secreted effectors (VF
VFG038695(gb|WP_017409955) (fliA) flagellar biosynthesis sigma factor [Polar flagella (VF0473) - Motility (V
VFG015509(gb|NP_252901) (phzC1) phenazine biosynthesis protein PhzC [Pyocyanin (VF0100) - Nutritional/M
VFG022130(gb|NP_214804) (eccD3) Type IIV secretion system transmembrane protein EccD3 [ESX-3 (VF0521
VFG000921(gb|WP_000189356) (chuY) ChuY [Chu (VF0227) - Nutritional/Metabolic factor (VFC0272)] [Esch
VFG001307(gb|WP_001238807) (cap8K) type 8 capsular polysaccharide synthesis protein Cap8K [Capsule (VF
VFG038281(gb|WP_016349337) (exeF) general secretion pathway protein F [Exe T2SS (VF0478) - Effector deli
VFG038878(gb|WP_005314254) (flgI) flagellar basal body P-ring protein [Lateral flagella (VF0474) - Motility (
VFG036956(gb|WP_002224867) (farB) fatty acid efflux system protein FarB [FarAB (VF0450) - Antimicrobial a
VFG038277(gb|WP_016349336) (exeE) general secretory pathway protein E [Exe T2SS (VF0478) - Effector del
VFG000886(gb|WP_000261297) (papJ) P pilus assembly protein PapJ [P fimbriae (VF0220) - Adherence (VFC0
VFG048429(gb|WP_004183353) (entE) enterobactin synthase subunit E [Ent (VF0562) - Nutritional/Metabolic f
VFG002533(gb|WP_009938926) (motB) flagellar motor protein MotB [Flagella (VF0430) - Motility (VFC0204)
VFG038309(gb|WP_016349344) (exeM) general secretion pathway protein M [Exe T2SS (VF0478) - Effector de
VFG037822(gb|WP_000240700) (lpxC) UDP-3-O-acyl-N-acetylglucosamine deacetylase [LPS (VF0466) - Immu
VFG001819(gb|NP_216894) (mbtG) Lysine-N-oxygenase MbtG (L-lysine 6-monooxygenase) (lysine N6-hydrox
VFG048665(gb|WP_004176431) (dotU/tssL) type VI secretion system protein, DotU/TssL family [T6SS (VF056
VFG034875(gb|WP_001270087) (espX5) Type III secretion system effector EspX5 [TTSS secreted effectors (VF
VFG002276(gb|WP_011009653) (colA) collagenase [Kappa-toxin (VF0388) - Exoenzyme (VFC0251)] [Clostrid
VFG034932(gb|WP_000953022) (nleB1) Type III secretion system effector nleB1 [TTSS secreted effectors (VF
VFG041040(gb|NP_251058) (PA2368) hypothetical protein [HSI-3 (VF0944) - Effector delivery system (VFC0
VFG038820(gb|WP_161785096) (flgA) flagella basal body P-ring formation protein FlgA [Polar flagella (VF047
VFG000190(gb|NP_250386) (pscP) type III secretion system protein PscP [TTSS (VF0083) - Effector delivery s

VFG048797(gb|WP_004175261) (ugd) UDP-glucose 6-dehydrogenase [Capsule (VF0560) - Immune modulation
VFG037513(gb|WP_085947930) (ACICU_RS04590) porin family protein [HemO cluster (VF0636) - Nutritional
VFG038464(gb|WP_011707389) (tapT) twitching ATPase [Tap type IV pili (VF0475) - Adherence (VFC0001)]
VFG049912 (tssC) Type VI secretion system protein TssC [T6SS (VF0579) - Effector delivery system (VFC008
VFG048779(gb|WP_004190503) (tssG) type VI secretion system baseplate subunit TssG [T6SS (VF0569) - Effe
VFG000925(gb|WP_000140634) (fepC) ferrienterobactin ABC transporter ATPase [Enterobactin (VF0228) - Nu
VFG034866(gb|WP_000900547) (espX4) Type III secretion system effector EspX4 [TTSS secreted effectors (V
VFG015950(gb|NP_251075) (pvdQ) 3-oxo-C12-homoserine lactone acylase PvdQ [Pyoverdine (VF0094) - Nutri
VFG038132(gb|WP_000045495) (pgi) glucose-6-phosphate isomerase [Capsule (VF0465) - Immune modulation
VFG000478(gb|NP_459678) (fur) ferric iron uptake transcriptional regulator [Fur (VF0113) - Regulation (VFC0
VFG048762(gb|WP_015874732) (impA/tssA) type VI secretion system protein TssA [T6SS (VF0569) - Effector
VFG048819(gb|WP_014907234) (KP1_RS17280) mannose-1-phosphate guanylyltransferase/mannose-6-phosph
VFG016058(gb|NP_251114) (pvdL) peptide synthase PvdL [Pyoverdine (VF0094) - Nutritional/Metabolic factor
VFG041278(gb|WP_010947155) (vpdC) Dot/Icm type IV secretion system effector VpdC [Dot/Icm T4SS secret
VFG038346(gb|WP_005320760) (acr1) Type III secretion system gatekeeper subunit [T3SS (VF0479) - Effector
VFG038631(gb|WP_011705273) (fliG) flagellar motor switch protein G [Polar flagella (VF0473) - Motility (VF
VFG045790(gb|WP_000833288) (cgsE) curli production assembly/transport protein CsgE [Curli fibers (VF1138
VFG000216(gb|NP_250412) (pscH) type III secretion system protein PscH [TTSS (VF0083) - Effector delivery
VFG001959(gb|YP_002344814) (hddC) capsular polysaccharide heptosyltransferase [Capsule (VF0323) - Immu
VFG048547(gb|WP_004213617) (iroN) salmochelin receptor IroN [Sal (VF0563) - Nutritional/Metabolic factor
VFG050457(gb|WP_001274990.1) (pilB) type IV-A pilus assembly ATPase PilB [TFP (VF1334) - Adherence (V
VFG050222(gb|NP_219545) (glgX) glycogen hydrolase [TTSS secreted effectors (VF0711) - Effector delivery s
VFG000270(gb|WP_000724295) (ureB) urease beta subunit UreB, urea amidohydrolase [Urease (VF0050) - Stre
VFG034849(gb|WP_000129338) (espX1) Type III secretion system effector EspX1 [TTSS secreted effectors (V
VFG048676(gb|WP_014907726) (ompA) OmpA family protein [T6SS (VF0569) - Effector delivery system (VF
VFG050400(gb|WP_000347039.1) (pilU) PilT/PilU family type 4a pilus ATPase [TFP (VF1334) - Adherence (V
VFG038876(gb|WP_005314259) (lfgG) lateral flagellar basal-body rod protein LfgG [Lateral flagella (VF0474)
VFG013374(gb|WP_005693262) (lpxD) UDP-3-O-(3-hydroxymyristoyl) glucosamine N-acyltransferase [LOS (
VFG034754(gb|WP_000081129) (espL1) Type III secretion system effector espL1 [TTSS secreted effectors (VF
VFG000192(gb|NP_250388) (pscN) type III secretion system ATPase PscN [TTSS (VF0083) - Effector delivery
VFG002413(gb|WP_000389022) (ykgK/ecpR) regulator protein EcpR [ECP (VF0404) - Adherence (VFC0001)]
VFG035909(gb|WP_001216023) (aatA) outer membrane protein AatA [Dispersin (VF0215) - Others (VFC0346)
VFG043465(gb|WP_010976622) (pfbA) cell wall surface anchor family protein, Plasminogen- and Fibronectin-b
VFG002501(gb|WP_004197167) (fliG) flagellar motor switch protein G [Flagella (VF0430) - Motility (VFC020
VFG037777(gb|WP_000867091) (lpsB) glycosyltransferase family 4 protein [LPS (VF0466) - Immune modulati
VFG049150(gb|WP_000982270) (clbD) colibactin biosynthesis dehydrogenase ClbD [Colibactin (VF0573) - Ex
VFG038367(gb|WP_011705045) (vgrG1) Type VI secretion system effector VgrG1, ADP-ribosyltransferase [T
VFG002188(gb|WP_002362618) (cpsC) teichoic acid biosynthesis protein, putative [Capsule (VF0361) - Immun
VFG050229(gb|NP_219638) (CT_135) hypothetical protein [TTSS secreted effectors (VF0711) - Effector delive
VFG000337(gb|WP_005693332) (iga1) immunoglobin A1 protease [IgA1 protease (VF0045) - Exoenzyme (VF
VFG049903(gb|WP_001240533) (tssA) Type VI secretion system protein TssA [T6SS (VF0579) - Effector deliv
VFG013066 (shuW) heme anaerobic degradation radical SAM methyltransferase ChuW/HutW [Shu (VF0256) -
VFG000123(gb|NP_252231) (alg8) alginate-c5-mannuronan-epimerase AlgG [Alginate (VF0091) - Biofilm (VF
VFG049118(gb|WP_012737529) (allR) DNA-binding transcriptional repressor AllR [Allantion utilization (VF05
VFG002063(gb|NP_248767) (icmF1/tssM1) type VI secretion system protein IcmF1 [HSI-I (VF0334) - Effector
VFG034612(gb|WP_000225871) (cfaA) colonisation factor antigen a, chaperone [Adhesive fimbriae (VF0213) -

VFG000137(gb|NP_251850) (wzz) O-antigen chain length regulator [LPS (VF0085) - Immune modulation (VFC
VFG014984(gb|NP_253136) (algW) AlgW protein [Alginate (VF0091) - Biofilm (VFC0271)] [Pseudomonas aer
VFG050910(gb|WP_001178477.1) (vgrG/tssI) type VI secretion system tip protein VgrG [T6SS (VF1337) - Effe
VFG037344(gb|WP_000717759) (basC) acinetobactin biosynthesis protein BasC [Acinetobactin (VF0467) - Nutr
VFG049149(gb|WP_041169023) (clbC) colibactin polyketide synthase ClbC [Colibactin (VF0573) - Exotoxin (V
VFG000877(gb|WP_001244821) (fimF) FimF protein precursor [Type 1 fimbriae (VF0221) - Adherence (VFC00
VFG034817(gb|WP_000671691) (espR1) Type III secretion system effector espR1 [TTSS secreted effectors (VF
VFG013368(gb|WP_005693225) (rffG) dTDP-glucose 46-dehydratase [LOS (VF0044) - Immune modulation (V
VFG000929(gb|WP_001375021) (entD) phosphopantetheinyl transferase component of enterobactin synthase mu
VFG000130(gb|NP_252238) (algI) alginate o-acetyltransferase AlgI [Alginate (VF0091) - Biofilm (VFC0271)] [
VFG000930(gb|WP_000077767) (entF) enterobactin synthase multienzyme complex component, ATP-dependent
VFG038351(gb|WP_005320747) (ascQ) type III secretion system protein AscQ [T3SS (VF0479) - Effector deliv
VFG046680(gb|WP_003020700) (FTT_RS04120) ABC transporter ATP-binding protein [Capsule (VF0543) - In
VFG050670(gb|WP_000505931.1) (pilJ) methyl-accepting chemotaxis protein [TFP (VF1334) - Adherence (VFC
VFG016046(gb|NP_251103) (pvdH) diaminobutyrate-2-oxoglutarate aminotransferase PvdH [Pyoverdine (VF00
VFG038557(gb|WP_011705350) (flp1) Flp pilus pilin subunit [Flp type IV pili (VF0476) - Adherence (VFC0001
VFG048998(gb|WP_011154608) (rmpA2) regulator of mucoid phenotype RmpA2 [RmpA (VF0570) - Regulation
VFG002489(gb|WP_181937968) (tssM-5) Type VI secretion system protein TssM [T6SS-1 (VF0429) - Effector
VFG051303(gb|NP_219923.1) (pmpB) polymorphic membrane protein PmpB [Pmps (VF1346) - Adherence (VF
VFG014893(gb|NP_254009) (algC) phosphomannomutase AlgC [Alginate (VF0091) - Biofilm (VFC0271)] [Pse
VFG001248(gb|NP_249788) (fleQ) transcriptional regulator FleQ [Flagella (VF0273) - Motility (VFC0204)] [Ps
VFG048697(gb|WP_015874721) (clpV/tssH) type VI secretion system ATPase TssH [T6SS (VF0569) - Effector
VFG014668(gb|NP_252042) (flgN) flagella synthesis protein FlgN [Flagella (VF0273) - Motility (VFC0204)] [P
VFG041906(gb|WP_005463180) (VP_RS21695) type III secretion system chaperone [T3SS2 (VF0409) - Effecto
VFG000429(gb|WP_010989299) (tviD) Vi polysaccharide biosynthesis protein TviD [Vi antigen (VF0101) - Imm
VFG037720(gb|WP_001027056) (adeG) cation/multidrug efflux pump [AdeFGH efflux pump (VF0504) - Biofil
VFG037177(gb|WP_000632986) (plc2) phospholipase C [Phospholipase C (VF0470) - Exotoxin (VFC0235)] [A
VFG001246(gb|NP_249783) (fliC) B-type flagellin [Flagella (VF0273) - Motility (VFC0204)] [Pseudomonas aer
VFG001278(gb|WP_000069296) (ebp) cell surface elastin binding protein [EbpS (VF0008) - Adherence (VFC00
VFG017842(gb|WP_011988521) (pilQ) type IV pilus biogenesis protein PilQ [Type IV pili (VF0401) - Adherenc
VFG050470(gb|WP_000279216.1) (pilC) type II secretion system F family protein [TFP (VF1334) - Adherence (
VFG002052(gb|WP_000820136) (gspI) general secretion pathway protein I [T2SS (VF0333) - Effector delivery s
VFG038293(gb|WP_016349340) (exeI) general secretion pathway protein I [Exe T2SS (VF0478) - Effector deliv
VFG024730(gb|NP_217462) (pks1) polyketide synthase [PDIM (VF0309) - Immune modulation (VFC0258)] [M
VFG045464(gb|WP_000562556) (tcpC) Tir domain containing protein TcpC [TcpC (VF0413) - Immune modula
VFG037218(gb|WP_000983821) (basJ) acinetobactin biosynthesis protein BasJ [Acinetobactin (VF0467) - Nutri
VFG045544(gb|WP_010947524) (mavB) Dot/Icm type IV secretion system effector mavB [Dot/Icm T4SS secret
VFG049163(gb|WP_000065646) (clbQ) colibactin biosynthesis thioesterase ClbQ  [Colibactin (VF0573) - Exoto
VFG001256(gb|NP_250134) (fliM) flagellar motor switch protein FliM [Flagella (VF0273) - Motility (VFC0204
VFG048551(gb|WP_011251260) (iroC) ABC transporter [Sal (VF0563) - Nutritional/Metabolic factor (VFC0272
VFG039753(gb|WP_011190903) (rvhB1) type IV secretion system protein [Rvh T4SS (VF0488) - Effector deliv
VFG050270(gb|NP_220318) (glgA) glycogen synthase [TTSS secreted effectors (VF0711) - Effector delivery sy
VFG011087(gb|WP_010929841) (bopC/bteA) Bordetella Bsc type III secretion system secreted protein BteA [T
VFG048808(gb|WP_004899416) (rfbK1) O9 family phosphomannomutase RfbK1 [Capsule (VF0560) - Immune
VFG002321(gb|WP_011816589) (fliC) flagellin [Flagella (VF0394) - Motility (VFC0204)] [Yersinia enterocoliti
VFG038727(gb|WP_011705295) (cheW) chemotaxis protein CheW [Polar flagella (VF0473) - Motility (VFC020

VFG045569(gb|WP_010948086) (LPG_RS11990) Dot/Icm type IV secretion system effector [Dot/Icm T4SS sec
VFG038880(gb|WP_005314250) (lfgK) lateral flagellar hook-associated protein [Lateral flagella (VF0474) - Mot
VFG038413(gb|WP_043118350) (fimC) outer membrane usher protein [Type I pili (VF0486) - Adherence (VFC
VFG041016(gb|NP_250349) (PA1658) hypothetical protein [HSI-2 (VF0943) - Effector delivery system (VFC00
VFG038619(gb|WP_016349413) (motX) sodium-type flagellar motor component [Polar flagella (VF0473) - Mot
VFG038374(gb|WP_011705709) (atsB) Type VI secretion system protein [T6SS (VF0480) - Effector delivery sy
VFG038890(gb|WP_005314213) (lafU) LafU, chemotaxis protein [Lateral flagella (VF0474) - Motility (VFC020
VFG038324(gb|WP_005320804) (ascH) Type III secretion system protein AscH, regulator needle assembling [T
VFG000056(gb|WP_010930825) (bopD) Bordetella Bsc type III secretion system secreted protein BopD, comple
VFG005969(gb|WP_000902274) (SP_RS01735) polysaccharide pyruvyl transferase family protein [Capsule (VF
VFG040815(gb|WP_041471735) (rickA) WASP, N-WASP, MENA proteins [RickA (VF0490) - Motility (VFC0
VFG002333(gb|WP_005168827) (fliK) flagellar hook-length control protein FliK [Flagella (VF0394) - Motility (
VFG049915(gb|WP_001142968) (hcp2/tssD2) Type VI secretion system protein, Hcp family [T6SS (VF0579) -
VFG041027(gb|NP_250360) (PA1669) hypothetical protein [HSI-2 (VF0943) - Effector delivery system (VFC00
VFG001145(gb|CAA83901) (hifB) periplasmic chaperone [Haemagglutinating pili (VF0036) - Adherence (VFC
VFG045793(gb|WP_000771429) (csgA) curlin major subunit CsgA [Curli fibers (VF1138) - Adherence (VFC00
VFG012505(gb|WP_000933673) (iroD) esterase [Salmochelin siderophore (VF0230) - Nutritional/Metabolic fact
VFG007832(gb|NP_217448) (ppsB) phthiocerol synthesis polyketide synthase type I PpsB [PDIM (VF0309) - Im
VFG038301(gb|WP_016349342) (exeK) general secretion pathway protein K [Exe T2SS (VF0478) - Effector del
VFG038832(gb|WP_016351724) (pomB2) flagellar motor protein MotB [Polar flagella (VF0473) - Motility (VF
VFG000928(gb|WP_000640938) (fepG) iron-enterobactin ABC transporter permease [Enterobactin (VF0228) - N
VFG034880(gb|WP_000535069) (espX6) Type III secretion system effector EspX6 [TTSS secreted effectors (VF
VFG034756 (espL1) Type III secretion system effector espL1 [TTSS secreted effectors (VF1111) - Effector deliv
VFG038448(gb|WP_016351632) (tapN) type IV pilus biogenesis protein PilN [Tap type IV pili (VF0475) - Adhe
VFG005584(gb|WP_000403400) (hylB) hyaluronidase [Hyaluronidase (VF0281) - Exoenzyme (VFC0251)] [Str
VFG038317(gb|WP_016352170) (tapD/pilD) prepilin peptidase [Tap type IV pili (VF0475) - Adherence (VFC00
VFG043018(gb|NP_252039) (PA3349) chemotaxis protein [Flagella (VF0273) - Motility (VFC0204)] [Pseudom
VFG040863(gb|WP_011190526) (sca2) cell surface antigen Sca2 [Sca2 (VF0495) - Motility (VFC0204)] [Ricke
VFG038176(gb|WP_000777882) (ompA) outer membrane protein OmpA [OmpA (VF0464) - Immune modulatio
VFG038635(gb|WP_016350051) (fliH) flagellar assembly protein FliH [Polar flagella (VF0473) - Motility (VFC
VFG000198(gb|NP_250394) (pcrD) type III secretion system protein PcrD [TTSS (VF0083) - Effector delivery s
VFG045349(gb|WP_000092542) (fdeC) adhesin FdeC [FdeC (VF0506) - Adherence (VFC0001)] [Escherichia c
VFG002054(gb|WP_000633233) (gspK) general secretion pathway protein K [T2SS (VF0333) - Effector delivery
VFG002051(gb|WP_001115126) (gspH) general secretion pathway protein H [T2SS (VF0333) - Effector delivery
VFG049158(gb|WP_001297937) (clbL) colibactin biosynthesis amidase ClbL [Colibactin (VF0573) - Exotoxin (
VFG038468(gb|WP_016351972) (tapU) twitching ATPase [Tap type IV pili (VF0475) - Adherence (VFC0001)]
VFG045491(gb|WP_010946113) (LPG_RS01820) Dot/Icm type IV secretion system effector [Dot/Icm T4SS sec
VFG000875(gb|WP_000066579) (fimC) Chaperone protein fimC precursor [Type 1 fimbriae (VF0221) - Adhere
VFG050282(gb|WP_000843487) (vpsj) exopolysaccharide biosynthesis protein VpsJ [VPS (VF0615) - Biofilm (
VFG002351(gb|WP_005160463) (flgB) flagellar basal-body rod protein FlgB [Flagella (VF0394) - Motility (VF
VFG003624(gb|WP_011815750) (yplA) DUF2974 domain-containing protein [YplA (VF0622) - Exotoxin (VFC
VFG000166(gb|NP_252916) (pchE) dihydroaeruginoic acid synthetase PchE [Pyochelin (VF0095) - Nutritional/
VFG001264(gb|NP_250144) (flhF) flagellar biosynthesis protein FlhF [Flagella (VF0273) - Motility (VFC0204)]
VFG001382(gb|YP_177730) (pcaA) cyclopropane mycolic acid synthase [PcaA (VF0285) - Others (VFC0346)]
VFG000926(gb|WP_001443194) (fepD) ferrienterobactin ABC transporter permease [Enterobactin (VF0228) - N
VFG001014(gb|NP_858288) (spa29) type III secretion system minor export apparatus protein Spa29 [TTSS (VF

VFG038529(gb|WP_016350153) (flpG) Flp pilus assembly protein FlpG [Flp type IV pili (VF0476) - Adherence
VFG001904(gb|YP_002343808) (Cj0371) hypothetical protein [Flagella (VF0114) - Motility (VFC0204)] [Camp
VFG037505(gb|WP_002002089) (ACICU_RS04585) transferrin-binding protein-like solute binding protein [He
VFG012333(gb|WP_000703306) (sfaD) S fimbriae minor subunit SfaD [S fimbriae (VF0222) - Adherence (VFC
VFG012095(gb|WP_003435012) (groEL) chaperonin GroEL [GroEL (VF0594) - Adherence (VFC0001)] [Clostr
VFG038472(gb|WP_011706550) (tapW) twitching ATPase [Tap type IV pili (VF0475) - Adherence (VFC0001)]
VFG000906(gb|WP_001142375) (hlyA) Hemolysin A [-Hemolysin (VF0225) - Exotoxin (VFC0235)] [Escherich
VFG049160(gb|WP_001327259) (clbN) colibactin non-ribosomal peptide synthetase ClbN [Colibactin (VF0573)
VFG050158(gb|WP_000905916) (kpsE) Capsule polysaccharide export inner-membrane protein [K1 capsule (V
VFG045609(gb|WP_010948684) (LPG_RS15170) Dot/Icm type IV secretion system effector [Dot/Icm T4SS sec
VFG038800(gb|WP_016351322) (flgD) flagellar hook capping protein [Polar flagella (VF0473) - Motility (VFC
VFG022051(gb|NP_214798) (eccC3) Type VII secretion system protein EccC3 [ESX-3 (VF0521) - Effector deli
VFG000430(gb|WP_000127915) (tviC) Vi polysaccharide biosynthesis protein, epimerase TviC [Vi antigen (VF
VFG045514(gb|WP_010946881) (LPG_RS05710) Dot/Icm type IV secretion system effector [Dot/Icm T4SS sec
VFG006167(gb|WP_002225356) (pilX) type IV pilus minor pilin PilX [Type IV pili (VF0075) - Adherence (VF
VFG001214(gb|NP_253237) (pilR) two-component response regulator PilR [Type IV pili (VF0082) - Adherence
VFG037400(gb|WP_001223274) (bauC) ferric siderophore ABC transporter, permease protein BauC [Acinetoba
VFG045608(gb|WP_010948659) (LPG_RS15050) Dot/Icm type IV secretion system effector [Dot/Icm T4SS sec
VFG000129(gb|NP_252237) (algL) poly(beta-d-mannuronate) lyase precursor AlgL [Alginate (VF0091) - Biofil
VFG043211(gb|WP_032902679) (cheA) chemotaxis protein CheA [Flagella (VF0394) - Motility (VFC0204)] [Y
VFG000363(gb|WP_002212775) (irp2) yersiniabactin biosynthetic protein Irp2 [Yersiniabactin (VF0136) - Nutri
VFG045792(gb|WP_000791650) (csgB) curlin minor subunit CsgB [Curli fibers (VF1138) - Adherence (VFC00
VFG000912(gb|WP_001443207) (focD) F1C fimbrial usher [F1C fimbriae (VF0224) - Adherence (VFC0001)] [
VFG044159(gb|WP_000125846) (fes) enterobactin/ferric enterobactin esterase [Enterobactin (VF0228) - Nutritic
VFG002492(gb|WP_004184995) (fliN) flagellar motor switch protein FliN [Flagella (VF0430) - Motility (VFC0
VFG034922(gb|WP_001303732) (espY5) hypothetical protein [TTSS secreted effectors (VF1110) - Effector deli
VFG038683(gb|WP_016350056) (flhA) flagellar biosynthesis protein FlhA [Polar flagella (VF0473) - Motility (
VFG000873(gb|WP_000695571) (fimA) Type-1 fimbrial protein, A chain precursor [Type 1 fimbriae (VF0221) -
VFG038840(gb|WP_005300916) (flmH) short chain dehydrogenase/reductase family oxidoreductase [Polar flage
VFG042979(gb|WP_002379311) (srtC) sortase [Ebp pili (VF0538) - Adherence (VFC0001)] [Enterococcus faeca
VFG038699(gb|WP_010634617) (cheY) chemotaxis protein CheY [Polar flagella (VF0473) - Motility (VFC020
VFG049156(gb|WP_077265119) (clbJ) colibactin non-ribosomal peptide synthetase ClbJ [Colibactin (VF0573) -
VFG001936(gb|YP_002344528) (Cj1135) glucosyltransferase [LOS (VF0326) - Immune modulation (VFC0258)
VFG050115(gb|WP_002081542) (cesA) Cereulide synthetase A, CesA [Cereulide (VF0583) - Exotoxin (VFC02
VFG038687(gb|WP_016350057) (flhF) flagellar biosynthesis protein FlhF [Polar flagella (VF0473) - Motility (V
VFG038804(gb|WP_011706643) (flgC) flagellar basal body rod protein FlgC [Polar flagella (VF0473) - Motility
VFG038513(gb|WP_016350157) (flpK) Flp pilus pseudopilin [Flp type IV pili (VF0476) - Adherence (VFC0001
VFG044333(gb|WP_015874968) (fyuA) yersiniabactin receptor FyuA [Ybt (VF0564) - Nutritional/Metabolic fa
VFG000983(gb|WP_000517827) (rtxA) RTX toxin RtxA [MARTX (VF0265) - Exotoxin (VFC0235)] [Vibrio ch
VFG048317(gb|WP_014906959) (fimK) transcriptional regulator [Type I fimbriae (VF0566) - Adherence (VFC
VFG038600(gb|WP_016349156) (mshL) MSHA (mannose-sensitive hemagglutinin) pili biogenesis protein Msh
VFG002070(gb|NP_248774) (hsiC1/vipB/tssC) type VI secretion system tubule-forming protein VipB [HSI-I (V
VFG044324(gb|WP_032447693) (ybtX) yersiniabactin-associated zinc MFS transporter YbtX [Ybt (VF0564) - N
VFG001820(gb|NP_216895) (mbtF) Peptide synthetase MbtF (peptide synthase) [Mycobactin (VF0299) - Nutriti
VFG000181(gb|NP_251792) (xcpS) general secretion pathway protein F [Xcp secretion system (VF0084) - Effec
VFG038884(gb|WP_005314225) (lafC) LafC, flagellar export chaperone [Lateral flagella (VF0474) - Motility (V

VFG038305(gb|WP_016349343) (exeL) general secretion pathway protein L [Exe T2SS (VF0478) - Effector del
VFG049911(gb|WP_000393837) (tssF) Type VI secretion system protein TssF [T6SS (VF0579) - Effector delive
VFG013660(gb|WP_041175640) (pilO) type IV pilus inner membrane platform protein PilO [Type IV pili (VF04
VFG000314(gb|WP_001186109) (gluE) UDP-glucose 4-epimerase [LPS (VF0056) - Immune modulation (VFC0
VFG048287(gb|WP_014906962) (fimF) type 1 fimbrial minor component [Type I fimbriae (VF0566) - Adherenc
VFG038792(gb|WP_011706640) (flgF) flagellar basal-body rod protein FlgF [Polar flagella (VF0473) - Motility
VFG038593(gb|WP_016349158) (mshN) Tetratricopeptide repeat protein, PilF-like [MSHA type IV pili (VF047
VFG019180(gb|WP_011987710) (bont) botulinum neurotoxin, BoNT [BoNT (VF0375) - Exotoxin (VFC0235)] [
VFG038105(gb|WP_000591436) (galU) UTP--glucose-1-phosphate uridylyltransferase GalU [Capsule (VF0465)
VFG045818(gb|WP_001528154) (tsh) temperature-sensitive protein Tsh [Tsh (VF0233) - Effector delivery syste
VFG035923(gb|WP_000621745) (aatC) ATP-binding protein AatC [Dispersin (VF0215) - Others (VFC0346)] [E
VFG050860(gb|WP_000301479.1) (gspK) general secretion pathway protein K [T2SS (VF1335) - Effector deliv
VFG015515(gb|NP_252903) (phzE1) phenazine biosynthesis protein PhzE [Pyocyanin (VF0100) - Nutritional/M
VFG038297(gb|WP_016349341) (exeJ) general secretion pathway protein J [Exe T2SS (VF0478) - Effector deli
VFG041309(gb|WP_010947685) (pieE) Dot/Icm type IV secretion system effector PieE [Dot/Icm T4SS secreted
VFG038488(gb|WP_016349432) (tapY1) type IV pilus biogenesis protein [Tap type IV pili (VF0475) - Adherenc
VFG049908(gb|WP_001080148) (tssJ) Type VI secretion system protein TssJ [T6SS (VF0579) - Effector delive
VFG037927(gb|WP_000470518) (pseC) UDP-4-amino-4, 6-dideoxy-N-acetyl-beta-L-altrosamine transaminase [
VFG000894(gb|WP_001239363) (papH) P pilus termination subunit PapH [P fimbriae (VF0220) - Adherence (V
VFG001391(gb|NP_215677) (narG) nitrate reductase subunit alpha [Nitrate reductase (VF0302) - Nutritional/Me
VFG049919(gb|AAD32394) (bslA) s-layer protein [BslA (VF0411) - Adherence (VFC0001)] [Bacillus anthracis
VFG000690(gb|AAP42183) (AAP42183) Bcs3' [Capsule (VF0043) - Immune modulation (VFC0258)] [Haemop
VFG038032 (ACICU_RS00455*) EpsG family protein [Capsule (VF0465) - Immune modulation (VFC0258)] [A
VFG036992(gb|WP_002224969) (mtrD) multiple transferable resistance system protein MtrD [MtrCDE (VF045
VFG002357(gb|WP_005164474) (flhB) flagellar biosynthetic protein FlhB [Flagella (VF0394) - Motility (VFC0
VFG001279(gb|WP_001060526) (sdrC) Ser-Asp rich fibrinogen-binding bone sialoprotein-binding protein [SDr
VFG037428(gb|WP_000939834) (basB) non-ribosomal peptide synthetase with condensation and peptidyl carrie
VFG002436(gb|WP_080341181) (boaA) surface-exposed protein, autotransporter [BoaA (VF0434) - Adherence
VFG002486(gb|WP_004533362) (tssJ-5) Type VI secretion system protein TssJ [T6SS-1 (VF0429) - Effector de
VFG050164(gb|WP_000723250) (neuC) UDP-N-acetylglucosamine 2-epimerase [K1 capsule (VF0239) - Invasio
VFG037521(gb|WP_000837764) (ACICU_RS04595) TonB family protein [HemO cluster (VF0636) - Nutritiona
VFG038373(gb|WP_011705708) (atsA) Type VI secretion system protein [T6SS (VF0480) - Effector delivery sy
VFG043774(gb|AAB24244) (bont) botulinum neurotoxin, BoNT [BoNT (VF0375) - Exotoxin (VFC0235)] [Clos
VFG000932(gb|WP_000026784) (entE) 2,3-dihydroxybenzoate-AMP ligase component of enterobactin synthase
VFG012865 (ipaH) invasion plasmid antigen IpaH [TTSS secreted effectors (VF0978) - Effector delivery system
VFG048498(gb|WP_012737619) (entF) enterobactin synthase subunit F [Ent (VF0562) - Nutritional/Metabolic fa
VFG012621(gb|WP_000189352) (chuY) ChuY [Chu (VF0234) - Nutritional/Metabolic factor (VFC0272)] [Esch
VFG002084(gb|WP_000619136) (clpB/vasG) type VI secretion system AAA+ family ATPase [VAS T6SS (VF0
VFG048621(gb|WP_004213924) (iutA) ferric aerobactin receptor IutA [Aerobactin (VF0565) - Nutritional/Meta
VFG000872(gb|WP_000044711) (fimE) Type 1 fimbriae Regulatory protein fimE [Type 1 fimbriae (VF0221) - A
VFG038780(gb|WP_011706637) (flgI) flagellar basal body P-ring protein [Polar flagella (VF0473) - Motility (V
VFG000002(gb|WP_010930159) (prn) pertactin precursor [Pertactin (VF0027) - Adherence (VFC0001)] [Bordet
VFG000617(gb|NP_709456) (iucC) aerobactin synthesis protein IucC [Aerobactin (VF0123) - Nutritional/Metab
VFG012327(gb|WP_011076307) (sfaC) S fimbrial switch regulatory protein SfaC [S fimbriae (VF0222) - Adher
VFG000182(gb|NP_251793) (xcpR) general secretion pathway protein E [Xcp secretion system (VF0084) - Effec
VFG050166(gb|WP_000066003) (neuB) N-acylneuraminate-9-phosphate synthase [K1 capsule (VF0239) - Invas

VFG001975(gb|YP_002344830) (kpsM) capsule polysaccharide ABC transporter permease [Capsule (VF0323) -
VFG038402(gb|WP_011705725) (vasK/icmF) Type VI secretion system protein [T6SS (VF0480) - Effector deliv
VFG002050(gb|WP_001087302) (gspG) general secretion pathway protein G [T2SS (VF0333) - Effector delivery
VFG050162(gb|WP_001161829) (kpsS) Capsule polysaccharide export protein [K1 capsule (VF0239) - Invasion
VFG042738(gb|NP_253646) (fimX) protein FimX [Type IV pili (VF0082) - Adherence (VFC0001)] [Pseudomon
VFG038879(gb|WP_005314251) (flgJ) peptidoglycan hydrolase [Lateral flagella (VF0474) - Motility (VFC0204)
VFG034795 (espN) Type III secretion system effector EspN [TTSS secreted effectors (VF1110) - Effector delive
VFG038395(gb|WP_011705721) (clpB) type VI secretion system ATPase ClpV1 [T6SS (VF0480) - Effector deli
VFG002274(gb|WP_011009584) (plc) phospholipase C [Alpha-toxin (CpPLC) (VF0378) - Exotoxin (VFC0235)
VFG034840(gb|WP_001301665) (espW) Type III secretion system effector EspW [TTSS secreted effectors (VF1
VFG002076(gb|NP_248780) (clpV1) type VI secretion system AAA+ family ATPase [HSI-I (VF0334) - Effector
VFG048002(gb|WP_003022141) (iglJ) Type VI secretion system protein IglJ [T6SS (VF0544) - Effector delivery
VFG001970(gb|YP_002344825) (Cj1442c) sugar transferase [Capsule (VF0323) - Immune modulation (VFC025
VFG038859(gb|WP_005314301) (lfiR) lateral flagellar biosynthetic protein LfiR [Lateral flagella (VF0474) - Mo
VFG038537(gb|WP_016350151) (flpE) Flp pilus assembly protein, ATPase [Flp type IV pili (VF0476) - Adherer
VFG037176(gb|WP_001081735) (plc1) phospholipase C [Phospholipase C (VF0470) - Exotoxin (VFC0235)] [A
VFG044325(gb|WP_002212761) (ybtQ) yersiniabactin ABC transporter ATP-binding/permease protein YbtQ [Y
VFG038329(gb|WP_011899438) (ascC) Type III secretion system OM ring protein AscC [T3SS (VF0479) - Effe
VFG000079(gb|NP_463763) (clpC) endopeptidase Clp ATP-binding chain C [ClpC (VF0072) - Stress survival (
VFG001265(gb|NP_250145) (fleN) flagellar synthesis regulator FleN [Flagella (VF0273) - Motility (VFC0204)]
VFG048030(gb|WP_003022143) (pdpC) Type VI secretion system secreted protein PdpC [T6SS (VF0544) - Effe
VFG043205(gb|WP_005164477) (cheZ) chemotaxis regulator CheZ [Flagella (VF0394) - Motility (VFC0204)] [
VFG002305(gb|NP_461448) (shdA) AIDA autotransporter-like protein [ShdA (VF0398) - Adherence (VFC0001
VFG034648(gb|WP_001245988) (cfaD/cfaE) minor pilin and initiator [Adhesive fimbriae (VF0213) - Adherence
VFG002395(gb|YP_007410461) (eccC5) ESX-5 type VII secretion system FtsK/SpoIIIE family protein EccC5 [
VFG037260(gb|WP_001095752) (barB) siderophore efflux system of the ABC superfamily [Acinetobactin (VF0
VFG034908(gb|WP_000978349) (espY3) Type III secretion system effector EspY3 [TTSS secreted effectors (V
VFG038385(gb|WP_011705716) (atsI) Type VI secretion system protein [T6SS (VF0480) - Effector delivery sys
VFG044332(gb|WP_015874967) (ybtE) yersiniabactin biosynthesis salycil-AMP ligase YbtE [Ybt (VF0564) - N
VFG001826(gb|NP_217099) (relA) Probable GTP pyrophosphokinase RelA (ATP:GTP 3'-pyrophosphotransfera
VFG051000(gb|WP_000568832.1) (tssF) type VI secretion system baseplate subunit TssF [T6SS (VF1337) - Eff
VFG046465(gb|WP_003028672) (tufA) elongation factor Tu [EF-Tu (VF0460) - Adherence (VFC0001)] [Franci
VFG016216(gb|WP_000576719) (alo) thiol-activated cytolysin [ALO (VF0534) - Exotoxin (VFC0235)] [Bacillu
VFG002099(gb|WP_010948192) (lepB) Dot/Icm type IV secretion system effector LepB, Rab1 GTPase activatin
VFG038525(gb|WP_043118801) (flpH) Flp pilus assembly protein FlpH [Flp type IV pili (VF0476) - Adherence
VFG000144(gb|NP_249940) (aprA) alkaline metalloproteinase precursor [Alkaline protease (VF0090) - Exoenzy
VFG043208(gb|WP_005164483) (cheR) chemotaxis methyltransferase CheR [Flagella (VF0394) - Motility (VFC
VFG041011(gb|NP_248785) (vgrG1b) type VI secretion system substrate VgrG1b [HSI-I (VF0334) - Effector de
VFG038596(gb|WP_016349157) (mshM) membrane protein with ATPase domain, GspA-like [MSHA type IV pi
VFG038651(gb|WP_011705278) (fliL) flagellar basal body-associated protein FliL [Polar flagella (VF0473) - Mo
VFG000164(gb|NP_252911) (fptA) Fe(III)-pyochelin receptor precursor [Pyochelin (VF0095) - Nutritional/Meta
VFG002548(gb|WP_004550042) (wcbR) capsular polysaccharide biosynthesis fatty acid synthase [Capsule I (VF
VFG016338(gb|WP_001253006) (inhA) immune inhibitor A metalloprotease [InhA (VF0536) - Exoenzyme (VF
VFG048419(gb|WP_004210873) (entB) 2,3-dihydro-2,3-dihydroxybenzoate synthetase, isochroismatase [Ent (V
VFG038882(gb|WP_005314247) (maf-5) modification accessory factor [Lateral flagella (VF0474) - Motility (VF
VFG007857(gb|NP_217450) (ppsD) phthiocerol synthesis polyketide synthase type I PpsD [PDIM (VF0309) - In

VFG002053(gb|WP_001250462) (gspJ) general secretion pathway protein J [T2SS (VF0333) - Effector delivery
VFG041018(gb|NP_250351) (PA1660) hypothetical protein [HSI-2 (VF0943) - Effector delivery system (VFC00
VFG038355(gb|WP_005320739) (ascU) Type III secretion system inner membrane export apparatus protein AscU
VFG007823(gb|NP_217447) (ppsA) phthiocerol synthesis polyketide synthase type I PpsA [PDIM (VF0309) - In
VFG001376(gb|WP_000695335) (cps4L) capsular polysaccharide biosynthesis protein Cps4L [Capsule (VF0144
VFG044097(gb|NP_252909) (PA4219) hypothetical protein [Pyochelin (VF0095) - Nutritional/Metabolic factor (
VFG043394(gb|YP_002343725) (cheA) chemotaxis histidine kinase [Flagella (VF0114) - Motility (VFC0204)] [
VFG019178(gb|WP_012291519) (bont) botulinum neurotoxin, BoNT [BoNT (VF0375) - Exotoxin (VFC0235)] [
VFG049018(gb|WP_002913007) (rcsB) transcriptional regulator RcsB [RcsAB (VF0571) - Regulation (VFC030
VFG031564(gb|YP_177826) (PE_PGRS30) PE-PGRS family protein PE_PGRS30 [PE/PE-PGRS (VF0316) - Im
VFG037358(gb|WP_001073030) (bauA) TonB-dependent siderophore receptor BauA [Acinetobactin (VF0467) -
VFG048257(gb|WP_004174507) (fimI) type 1 pilus biosynthesis fimbrial protein [Type I fimbriae (VF0566) - Ad
VFG045507(gb|WP_010946840) (LPG_RS05490) Dot/Icm type IV secretion system effector [Dot/Icm T4SS sec
VFG001940(gb|YP_002344532) (wlaN) beta-1,3 galactosyltransferase [LOS (VF0326) - Immune modulation (V
VFG050212(gb|WP_011861707) (CD2831) SrtB-anchored collagen-binding adhesin [CD2831 (VF0598) - Adher
VFG002159(gb|NP_464601) (aut) autolysin [Auto (VF0348) - Invasion (VFC0083)] [Listeria monocytogenes EG
VFG050216(gb|WP_011861847) (cbpA) collagen-binding adhesin CbpA [CbpA (VF0592) - Adherence (VFC000
VFG034882(gb|WP_000535079) (espX6) Type III secretion system effector EspX6 [TTSS secreted effectors (VF
VFG003645(gb|NP_461791) (orgA/sctK) type III secretion system accessory cytosolic protein OrgA [TTSS (SPI
VFG038432(gb|WP_016352168) (tapB) type IV pilus assembly ATPase, PilB-like [Tap type IV pili (VF0475) - A
VFG000362(gb|WP_002212777) (irp1) yersiniabactin biosynthetic protein Irp1 [Yersiniabactin (VF0136) - Nutri
VFG046120(gb|WP_011206514) (MHP_RS03455) P102 paralog, surface adhesin [P97/P102 paralog family (VF
VFG048327(gb|WP_004152886) (mrkH) transcriptional activator [Type 3 fimbriae (VF0567) - Biofilm (VFC027
VFG002487(gb|WP_004536933) (tssK-5) Type VI secretion system protein TssK [T6SS-1 (VF0429) - Effector d
VFG038892(gb|WP_043118302) (aerA/act) Aerolysin A [Aerolysin (VF0481) - Exotoxin (VFC0235)] [Aeromon
VFG038553(gb|WP_016350147) (flpA) prepilin peptidase [Flp type IV pili (VF0476) - Adherence (VFC0001)] [
VFG043209(gb|WP_005168769) (cheD) methyl-accepting chemotaxis protein CheD [Flagella (VF0394) - Motili
VFG051368(gb|NP_220393.1) (pmpG) polymorphic membrane protein PmpG [Pmps (VF1346) - Adherence (VF
VFG011946(gb|YP_002344419) (flgR) sigma-54 associated transcriptional activator [Flagella (VF0114) - Motilit
VFG038371(gb|WP_011705707) (vgrG2) VgrG protein [T6SS (VF0480) - Effector delivery system (VFC0086)]
VFG011429(gb|WP_002963985) (acpXL) acyl carrier protein [LPS (VF0367) - Immune modulation (VFC0258)]
VFG000845(gb|WP_000581721) (toxB) cytotoxin [ToxB (VF0203) - Effector delivery system (VFC0086)] [Esch
VFG002465(gb|WP_009940818) (bsaM) Type III secretion system protein BsaM [Bsa T3SS (VF0428) - Effector
VFG038365(gb|WP_011705044) (hcp1) HcpA-like protein [T6SS (VF0480) - Effector delivery system (VFC008
VFG050169(gb|WP_000124301) (kpsM) Polysialic acid transport protein [K1 capsule (VF0239) - Invasion (VFC
VFG038505(gb|WP_011705643) (tapF) type IV pilus biogenesis/stability protein, pilW-like [Tap type IV pili (VF
VFG014950(gb|NP_249457) (mucD) serine protease MucD precursor [Alginate (VF0091) - Biofilm (VFC0271)]
VFG049916(gb|WP_000103365) (vgrG/tssI) type VI secretion system effector VgrG [T6SS (VF0579) - Effector
VFG002048(gb|WP_000249362) (gspE) general secretion pathway protein E [T2SS (VF0333) - Effector delivery
VFG038862(gb|WP_011898282) (lafK) two-component system lateral flagellar response regulator [Lateral flagel
VFG045632(gb|WP_002286924) (ecbA/fss3) Collagen binding MSCRAMM, EcbA/Fss3 [EcbA (VF0539) - Adh
VFG038541(gb|WP_016350150) (flpD) Flp pilus assembly protein FlpD [Flp type IV pili (VF0476) - Adherence
VFG041109(gb|NP_459283) (STM0285) hypothetical protein [SCI (Salmonella centrisome island) (VF0974) - E
VFG002029(gb|WP_010874666) (hmw2) cytadherence protein [Cytadherence organelle (VF0330) - Adherence (
VFG001243(gb|NP_249776) (flgJ) flagellar rod assembly protein/muramidase FlgJ [Flagella (VF0273) - Motility
VFG040422(gb|WP_011190749) (rvhD4) type IV secretion system component VirD4 [Rvh T4SS (VF0488) - Eff

VFG002047(gb|WP_000498818) (gspD) general secretion pathway protein D [T2SS (VF0333) - Effector delivery
VFG050111(gb|WP_050412751) (cer) ADP-ribosyltransferase exoenzyme family protein [Certhrax (VF0588) - E
VFG000340(gb|WP_005694039) (hxuA) heme-hemopexin utilization protein A, two-partner secretion (TPS) path
VFG043635(gb|WP_011081077) (VV1_RS15615) type II secretion system F family protein [Flp pili (VF0612) -
VFG038517(gb|WP_016350156) (flpJ) Flp pilus pseudopilin [Flp type IV pili (VF0476) - Adherence (VFC0001)
VFG005361(gb|WP_002352221) (spaP/pac) cell surface antigen SpaP [SpaP (VF0526) - Adherence (VFC0001)
VFG002302(gb|NP_461708) (mig-14) antimicrobial peptide resistance protein Mig-14 [Mig-14 (VF0395) - Antim
VFG049153(gb|WP_000159201) (clbG) colibactin biosynthesis acyltransferase ClbG [Colibactin (VF0573) - Exo
VFG049902(gb|WP_011310140) (tssM) Type VI secretion system protein TssM [T6SS (VF0579) - Effector deli
VFG002511(gb|WP_004197255) (flgE) flagellar hook protein FlgE [Flagella (VF0430) - Motility (VFC0204)] [E
VFG038403(gb|WP_011705726) (atsS) Type VI secretion system protein [T6SS (VF0480) - Effector delivery sy
VFG001240(gb|NP_249773) (flgG) flagellar basal-body rod protein FlgG [Flagella (VF0273) - Motility (VFC020
VFG002346(gb|WP_005160438) (flgG) flagellar basal-body rod protein FlgG [Flagella (VF0394) - Motility (VF
VFG038369(gb|WP_011705706) (hcp) hemolysin co-regulated protein [T6SS (VF0480) - Effector delivery syste
VFG001308(gb|WP_001291010) (cap8L) type 8 capsular polysaccharide synthesis protein Cap8L [Capsule (VF0
VFG000417(gb|WP_002218234) (pla) plasminogen activator protease precursor [Pla (VF0139) - Exoenzyme (VF
VFG041338(gb|WP_010948206) (ceg32/sidI) Dot/Icm type IV secretion system effector Ceg32/SidI [Dot/Icm T4
VFG050159(gb|WP_001331697) (kpsD) Polysialic acid transport protein precursor [K1 capsule (VF0239) - Invas
VFG038364(gb|WP_005320615) (aexT) Type III secretion system effector AexT, bifunctional toxin with GTPase
VFG048600(gb|WP_004213920) (iucA) aerobactin Synthetase IucA [Aerobactin (VF0565) - Nutritional/Metabol
VFG001445(gb|AAA92657) (AAA92657) unknown protein [TraJ (VF0241) - Invasion (VFC0083)] [Escherichia
VFG000933(gb|WP_001007140) (entB) isochorismatase [Enterobactin (VF0228) - Nutritional/Metabolic factor (
VFG001884(gb|WP_010948340) (enhB) enhanced entry protein EnhB [Enh loci (VF0163) - Others (VFC0346)]
VFG005289(gb|WP_001868236) (GBS_RS03585) tip adhesin protein [PI-1 (VF0527) - Adherence (VFC0001)] [
VFG034895(gb|WP_001304001) (espY1) Type III secretion system effector EspY1 [TTSS secreted effectors (VF
VFG002219(gb|WP_011005413) (cgs) cyclic beta 1-2 glucan synthetase [CG (VF0366) - Immune modulation (V
VFG038383(gb|WP_011705715) (atsH) Type VI secretion system protein [T6SS (VF0480) - Effector delivery sy
VFG002457(gb|WP_011205638) (bsaU) Type III secretion system protein BsaU [Bsa T3SS (VF0428) - Effector
VFG050160(gb|WP_000030755) (kpsU) 3-deoxy-manno-octulosonate cytidylyltransferase [K1 capsule (VF0239
VFG001444(gb|AAG10151) (aslA) putative arylsulfatase [AslA (VF0238) - Invasion (VFC0083)] [Escherichia c
VFG038796(gb|WP_016351321) (flgE) flagellar hook protein FlgE [Polar flagella (VF0473) - Motility (VFC020
VFG050301(gb|NP_218420) (cpnT) NAD+ glycohydrolase [CpnT (VF1197) - Effector delivery system (VFC008
VFG037330(gb|WP_001177743) (basD) acinetobactin biosynthesis protein BasD [Acinetobactin (VF0467) - Nut
VFG040108(gb|WP_011190503) (rvhB6e) VirB6-like protein of the type IV secretion system [Rvh T4SS (VF048
VFG049157(gb|WP_014907259) (clbK) colibactin hybrid non-ribosomal peptide synthetase/type I polyketide syn
VFG038764(gb|WP_017409846) (flrA) FleQ protein [Polar flagella (VF0473) - Motility (VFC0204)] [Aeromona
VFG002287(gb|WP_011860904) (toxA) toxin A [TcdA (VF0376) - Exotoxin (VFC0235)] [Clostridium difficile
VFG000913(gb|WP_000237770) (focF) F1C minor fimbrial subunit F precursor [F1C fimbriae (VF0224) - Adhe
VFG037497(gb|WP_085947929) (ACICU_RS04580) TonB-dependent receptor [HemO cluster (VF0636) - Nutri
VFG001223(gb|NP_249086) (pilT) twitching motility protein PilT [Type IV pili (VF0082) - Adherence (VFC000
VFG000191(gb|NP_250387) (pscO) type III secretion system protein PscO [TTSS (VF0083) - Effector delivery s
VFG000974(gb|WP_010922711) (scpA/scpB) streptococcal C5a peptidase [C5a peptidase (VF0245) - Adherence
VFG002529(gb|WP_011205290) (tsr) methyl-accepting chemotaxis protein I [Flagella (VF0430) - Motility (VFC
VFG002059(gb|NP_248763) (tagT) type six secretion associated protein TagT, ATP-binding component of ABC
VFG038816(gb|WP_011706646) (cheV) CheV [Polar flagella (VF0473) - Motility (VFC0204)] [Aeromonas hyd
VFG034929(gb|WP_001239101) (lifA/efa1) lymphostatin Efa1/LifA [Efa-1/LifA (VF0204) - Effector delivery sy

| |
|---|
| VFG002033(gb\|WP_010874498) (p1/MgPa/gapA) ADP1_MYCPN adhesin P1 [Cytadherence organelle (VF033 |
| VFG043625(gb\|WP_000813718) (mrkC) fimbrial biogenesis outer membrane usher protein mrkC precursor [Typ |
| VFG001968(gb\|YP_002344823) (Cj1440c) sugar transferase [Capsule (VF0323) - Immune modulation (VFC025 |
| VFG049899(gb\|WP_001087735) (tssA) Type VI secretion system protein TssA [T6SS (VF0579) - Effector deliv |
| VFG012337(gb\|WP_001370792) (sfaF) S fimbriae outer membrane usher SfaF [S fimbriae (VF0222) - Adherenc |
| VFG000289(gb\|WP_000481884) (virB6/cagW) type IV secretion system protein Cag9/CagW, VirB6 homolog [T |

ebruary); A (April); STP (sewage treatment plant); inf (influent); T (treated).

| Realtive abundance |
| --- |
| 2.1538406532684858e-05 |
| 9.790184787584026e-06 |
| 5.175132004420847e-06 |
| 3.647633364406307e-05 |
| 0.0002954766323328124 |
| 0.0001499358686805686 |
| 3.5171581429734212e-06 |
| 8.164377927552023e-06 |
| 3.1193554288716e-05 |
| 1.999586813379231e-06 |
| 5.222939228480161e-06 |
| 8.208830075977895e-06 |
| 0.00011484345670747979 |
| 7.921305379796533e-06 |
| 6.2487663438137045e-06 |
| 0.0001431349801222728 |
| 8.746259006366502e-06 |
| 0.00020698773735647989 |
| 0.0014570098536816226 |
| 4.349101319099827e-06 |
| 3.762949560618819e-05 |
| 1.732975238262e-05 |
| 5.132071148105696e-05 |
| 0.00012849617533704035 |
| 5.502512617839197e-06 |
| 3.643435857657853e-05 |
| 0.00011232778241383658 |
| 8.598983596699277e-06 |
| 8.588098807336367e-05 |
| 7.248502198499712e-06 |
| 3.89248310831654e-06 |
| 0.00020136934874736182 |
| 0.00014977037655178213 |
| 1.7263608289556567e-05 |
| 0.00036219017277079206 |
| 6.230117592098925e-06 |
| 5.283954873672687e-06 |
| 2.9757009025419866e-05 |
| 1.469481927181228e-06 |
| 0.00012494655723380719 |
| 7.414861265350525e-05 |
| 0.00018117250798902043 |
| 4.9270864617252945e-06 |

2.925656773521229e-06
0.00010684406390229495
2.234716092818093e-06
6.823590000656625e-05
0.00010107408652209652
1.0743623211077957e-05
2.6399214232668212e-05
3.3208996856562556e-06
1.5489949903643218e-05
7.390629692587942e-06
1.5716001987018136e-06
1.7682038201187724e-06
1.1597603517599539e-05
1.1223487275096328e-05
2.236926494492493e-06
4.568752900872546e-06
1.7948672110570712e-05
1.0389889828171103e-05
0.0003109607443156376
7.679228135592224e-05
6.536221635641358e-06
4.7711660040757596e-06
0.00019321217624908475
9.367295148830395e-05
6.8949167254021656e-06
3.473936537529816e-06
1.2371623008380252e-06
5.2593783393765354e-05
7.023393434571149e-06
3.1193554288715997e-06
6.873959531707934e-06
6.128814867024147e-06
4.505045191099422e-06
9.54869356282362e-05
3.243857298187336e-05
7.265968468857542e-05
2.3076864142162348e-05
1.0141402179066861e-05
0.00020295806155799193
3.3594196029962e-05
5.701342905710697e-05
3.093751964339138e-06
1.4283364332201536e-05
0.00014749126212599412
0.00018810479807373164
1.0642506757326635e-05

6.8949167254021656e-06
1.1965781407047142e-05
5.646773248269439e-06
8.731786432169535e-06
9.727022305083484e-05
1.2741029216517801e-05
0.00029066721041758086
0.00010221616659579253
8.076902449756821e-05
6.06309031080941e-06
0.00019073167230751047
3.18974990963598e-06
4.6153728284324696e-05
2.684311793390991e-05
4.456222041245143e-06
3.168522151918613e-05
3.298068500317369e-05
5.570277551556429e-06
2.3076864142162348e-05
2.9796214570907905e-06
1.1809570161524335e-05
5.668001719127593e-06
0.0001943504651972735
0.00018607547415895462
3.5023736286556125e-05
4.624811218674663e-06
0.00018301420579795805
8.620191852206831e-05
3.3670461825288984e-05
3.115058796049463e-05
7.283519117333044e-06
0.00027227297693037736
8.851399944938982e-06
0.00011040026425407252
0.00015756580188869864
0.00022547002608988833
4.434377815552764e-06
7.134172510826213e-07
9.156002777052266e-06
5.120451364374136e-05
0.00019559201608059762
5.9987604401376925e-06
0.0001138355378824787
8.737739922918743e-05
0.0004380043541923462
1.4781259385175884e-05

5.791730049337819e-05
0.00017565302415005127
0.00018183177374326916
0.00015714577575961911
1.4497004396999424e-05
6.06309031080941e-06
2.8682853577673008e-05
6.435255610375354e-05
8.137448944882434e-05
4.111877610785291e-06
5.597853182999777e-05
2.6859058027694893e-06
4.6822622897140995e-06
1.1345481701999548e-05
9.706148866660557e-06
9.1436631506681e-06
3.268110817820679e-06
8.4741271604006e-05
0.0003130719200925777
4.541230293035964e-06
6.717423819599733e-05
0.0001455441827579942
1.5870404813557264e-05
1.1085944538881913e-05
2.5846087839221827e-05
2.11853179010015e-05
0.0001851360561525064
0.0004189692027722463
1.3623690879107892e-05
9.265763660115565e-05
4.947856996969494e-05
4.399869038778035e-06
2.1215128385852814e-05
1.3832004195302203e-05
4.741158670716792e-06
4.863511152541742e-05
8.598983596699277e-06
6.370514608258901e-06
0.00017068171214580454
7.271809279523827e-06
4.195793480393154e-06
4.916375404199804e-05
0.00026923008165856074
5.904785080762168e-06
4.111877610785291e-06
3.960652689898266e-06

8.233735507033653e-05
3.641759558666522e-06
0.00012115353674635231
8.314458404161434e-06
3.1439286644095136e-06
1.3148445848441339e-05
1.5926286520647255e-05
1.2494655723380719e-05
8.698202638199654e-05
7.66621249468444e-06
1.2055078283219138e-06
0.00021218655841878965
2.6976533032985007e-05
0.00011122291898025789
3.023439419695067e-06
8.895589990049873e-05
0.0001343242956007074
1.3381850212614853e-05
0.0003543415268280412
0.00015433031452553717
5.0256281909598e-06
5.1165897871762665e-06
6.905443315822627e-05
9.48894833258144e-06
3.89248310831654e-06
1.4046786869142299e-05
1.7263608289556567e-05
5.398884396123924e-05
0.00010408727843201259
9.367295148830395e-05
7.651802320822251e-05
6.574222924220669e-06
0.00019009984896239246
7.02339343457115e-05
6.3259655550542946e-06
4.643804283227742e-06
2.2570186486346407e-06
2.1892862400115294e-06
6.0307538291517596e-06
3.7074306326752624e-05
1.229093851049951e-05
1.0567909747345374e-05
4.045675645674257e-06
9.79997497237161e-05
1.7396405276399308e-05
0.00014482721634046715

1.7450097885277085e-06
0.000125640704773995
7.69228804738745e-06
4.263545227576552e-05
2.217188907776382e-06
5.556591365926069e-06
6.586988405626922e-05
8.174214527464735e-06
9.633493244457681e-05
0.0002105710135877011
3.01136176555514e-06
8.879201750812367e-07
1.5705088096749374e-05
0.00011043577871854337
1.3303133446658295e-05
3.775513665996511e-06
2.2325100552141265e-06
7.053513250469895e-05
1.5596777144358e-05
2.9105954773898458e-06
2.718188324437392e-06
0.00015507652703533096
0.00015019666794614204
5.682242929477161e-06
0.00020559388053926456
4.941440683026752e-06
7.68678372678109e-05
5.2350293655831245e-05
0.00010023413566457219
0.00012335632832355872
1.3248580468259578e-06
0.00010279694026963228
1.7351913191293427e-05
1.1934209424442799e-05
1.7948672110570712e-05
0.00010475764485243179
1.018708417086446e-05
5.5159333803217325e-05
5.014484891201574e-06
5.906988358777377e-05
6.593389754903527e-06
2.98880971709063e-06
9.14777266219649e-05
6.443113065333077e-06
6.195979961457287e-05
9.678456573175078e-06

3.01537691457588e-05
0.00013213449400950486
1.838644460107244e-05
0.00021380035896415115
0.00013355507987786871
4.01693194659309e-06
5.91861171249398e-05
1.0872753297749567e-05
2.2751837886638933e-06
2.9665166404971042e-05
2.6144886542565432e-05
2.5862508479526294e-05
1.932933919599923e-06
0.00020758029484399174
6.78459805779573e-05
2.38810209707699e-06
0.00011183403391970984
3.3956947236214867e-06
3.6683417452261318e-06
9.234514846598245e-07
4.523065371863821e-05
5.843753710418372e-06
0.00015456518357088593
6.057676837317615e-05
3.3553897417387386e-05
4.2075026715012275e-05
1.4829722530701051e-05
1.4977037655178211e-05
3.4792810552798615e-05
1.5229176336241818e-05
6.926593218780735e-06
2.0559388053926453e-05
1.638791801399935e-05
8.757144960046118e-06
0.00010384588863973056
0.00017668224108843047
9.479478324265491e-05
2.194952461273287e-06
0.0001568693192553926
5.52941977000467e-06
0.00013988861974836557
1.229093851049951e-05
0.0002221148173683126
0.00014560552909424627
1.2634260815262065e-05
5.632709055870262e-05

0.00038710919849284945
0.00015656764748759377
7.156749006113639e-06
2.2570186486346404e-05
1.2029429180488884e-06
0.00017038944894007542
0.0001496602512749058
6.409201943936587e-05
2.0222348905501432e-05
1.2224501005037351e-05
3.047887716889367e-06
3.753581221463751e-05
7.067289643537219e-06
8.54320971877343e-05
4.662953991612186e-05
1.598256315146226e-06
6.894916725402165e-05
2.5875660022104234e-06
6.112250502518677e-05
1.4003298364903465e-05
0.0002869618513871306
0.00011043577871854337
9.613802915904881e-05
7.866200646719687e-06
3.697328641850534e-05
2.596478399462583e-06
2.8269158574148875e-05
3.967601203389316e-05
6.046878839390134e-06
7.771589986020309e-06
8.50200257869139e-06
8.273900070482598e-05
8.314458404161434e-06
8.843982570683447e-05
6.67118786410593e-06
2.535350544766715e-05
1.5814913887635735e-05
1.8893339063758645e-06
0.00011934743154739059
5.479484481332822e-05
3.506252226251023e-06
6.923059242648704e-06
0.0001556100471971498
0.00012098734533792111
7.744974951821609e-06
4.723828064609734e-05

7.21765750829333e-05
0.0001339065406143894
1.3034770524103227e-05
6.67118786410593e-06
5.982890703523571e-06
0.00016674895380143116
0.00018301420579795805
3.2078477814637025e-05
5.9318890122804204e-05
8.314458404161434e-06
3.106500942214162e-06
1.4975384742954926e-06
0.0002423070734927046
3.5790823911879885e-05
4.5005625590684775e-06
2.458187702099902e-05
6.842761530807595e-06
0.0001518535136836774
1.0073642253594252e-05
1.4404666789375223e-05
0.00025646246953867015
0.0001669321139000707
1.029529295568396e-05
5.566849688447778e-05
4.736194106663686e-05
8.076902449756821e-05
1.334237572821186e-05
0.00043000973605747584
0.0003104661291588894
2.7379330338158714e-06
1.595437520939619e-05
6.588587577369002e-06
0.0001130766342965955
5.384601633171214e-05
4.168723845035779e-06
1.9247086688782215e-05
9.19322230053622e-06
0.00048007974561010714
8.04815902466872e-06
2.108655185018098e-05
0.00016276182209358443
5.68939040486015e-06
5.359082194151446e-06
2.067214520961527e-06
3.995640787865565e-06
8.498452681998827e-05

3.3257833616645734e-05
2.1335214018225568e-05
0.0002299863748405332
4.532129631126073e-06
5.09736893899003e-06
0.00010878258489292731
0.000127154448205007
3.0376530368460843e-06
8.66487619131e-06
4.0565608716267444e-05
1.059265895050075e-05
5.951401805083973e-06
1.0059385178023054e-05
0.00010690881788041757
3.549490762673401e-05
1.2178420495056058e-06
0.0002276217963113286
2.567006453952225e-06
8.41065874933355e-05
7.935202406778632e-06
3.4580010488255507e-06
0.00015641724628923874
3.4615296213243515e-05
1.1450798409781823e-05
8.470159872404157e-06
1.7504122956129332e-06
4.691976526829689e-06
8.989355864582906e-05
0.00021324625822683024
1.1538432071081174e-05
9.565395046237687e-05
1.0735123509170459e-05
0.00012367756876190133
2.701950640300968e-06
3.691867724235861e-05
4.249226340999413e-05
3.2618259893248705e-05
0.000376922114321985
1.1195706365999553e-05
3.023439419695067e-06
2.2660648155630363e-05
1.6752093969866e-05
7.5384422864397e-05
4.1118776107852906e-05
6.17905105445877e-06
1.7737511262211057e-05

3.042420653720058e-05
4.587287395399412e-06
0.00011902803610167947
9.423052858049625e-06
5.056272265294921e-05
1.4404666789375223e-05
4.7913828091777755e-06
0.00013245419201770407
1.4829722530701051e-05
3.83310624734222e-06
3.13014904627254e-05
3.9331003233598434e-05
3.8695426185146745e-05
0.000125640704773995
0.0001098720333246272
6.40056420546767e-05
4.496088838830835e-06
2.4058858360977764e-05
5.904785080762168e-06
7.718541590211297e-06
0.00015295390146399392
0.00021047110763234927
3.960652689898266e-06
4.164885241126906e-06
6.750843838602716e-06
4.1118776107852906e-05
0.00026189039547583033
9.082460586071927e-06
0.0001323237209853777
6.162214403084224e-06
3.89248310831654e-06
4.948649203352101e-06
1.9057859712909353e-05
2.243584013821339e-06
0.00028566728664403075
1.7383033712005458e-06
6.162214403084224e-06
3.1366611455366296e-06
1.2224501005037351e-05
4.6356453298750717e-05
9.502238176184495e-06
7.446600875640138e-07
1.2599067888200055e-05
9.156002777052266e-06
0.00011999969353924422
4.01693194659309e-06

9.832750808399608e-05
7.33073804191867e-05
7.78496621663308e-06
0.00023670194209981314
6.0307538291517596e-06
1.8238166822031534e-05
1.8047836044883258e-05
0.00016547800140965195
5.1692175678443654e-05
0.00017343042070029987
0.00014096274194155538
1.011871447844255e-05
0.00013303133446658294
2.405885836097777e-06
5.308762173549085e-06
1.4742716335931616e-06
4.7312399287278456e-05
3.6126720222554474e-06
1.174822174510083e-05
7.225344044510895e-06
8.868755631105529e-06
0.00027621773263290507
0.00031222652753537566
1.018708417086446e-05
1.547228747501877e-05
6.730752041464019e-05
0.00014977037655178213
7.82537261568135e-06
0.00016226243486506978
0.00011555276497462314
4.332438095655e-06
0.00018483680606174263
4.59661115026811e-06
2.0748006292953302e-05
2.2706151465179817e-05
9.502238176184495e-06
8.598983596699277e-06
4.895092393792013e-06
0.00011251406397671194
2.9701179740193943e-05
4.8461414698540924e-05
1.541078491265356e-05
6.34371019896749e-06
0.00010028969782403148
1.3303133446658295e-05
0.00012400048669600552

2.833474826921651e-05
8.892002696979986e-06
0.00017263608289556565
2.999380220068846e-05
3.775513665996511e-06
2.9888097170906303e-05
1.8128518524504292e-05
6.046878839390134e-06
0.0004117636543013281
0.000566772319324459
8.470159872404157e-06
1.5596777144358e-05
2.555404164894813e-05
7.633865606521215e-05
2.1745506595499134e-05
6.916002097651101e-06
5.308762173549085e-06
6.406608175444505e-06
5.951401805083973e-06
1.7877728742544745e-05
6.452304382116719e-06
3.246995959701235e-06
2.5128140954798998e-05
1.3623690879107892e-05
7.66621249468444e-06
9.522242888134357e-06
1.2777020824474066e-05
5.611743637548164e-05
2.0748006292953302e-05
3.744259413794553e-06
4.873992857611874e-05
6.50800772930046e-06
8.345139062479374e-06
0.00020737237431555576
3.942699940606538e-06
4.691976526829689e-06
1.2564070477399499e-05
0.00011140555103112858
7.640313128148346e-06
1.3461504082928035e-05
0.0001932933919599923
7.318876006252135e-06
5.489157004689102e-06
8.342060811257506e-07
1.018708417086446e-05
7.014679546935205e-07

0.00028269158574148874
1.7197967193398554e-05
0.0001687710959650679
1.334237572821186e-05
1.3381850212614853e-05
4.8304581641264095e-06
0.000452306537186382
6.61722128413812e-05
6.586988405626922e-05
7.342638590688019e-06
0.00012190417823796542
1.0794905422109356e-05
0.00010318780316039133
2.2540192883042296e-05
2.065326653819096e-06
0.0001627001932325115
6.574222924220669e-06
1.0350264008841694e-05
0.00039499372254290203
0.00010380805771490735
3.4792810552798615e-05
1.650753785351759e-05
1.4590533457625226e-05
5.967104712221399e-06
8.66487619131e-06
3.4792810552798615e-05
5.139847013481613e-06
8.631804144778283e-06
1.3891478414815172e-06
4.981349528484383e-06
4.7711660040757596e-06
0.00018238166822031533
0.00015359355478226087
4.662953991612186e-05
4.096979503499837e-06
7.702768003855278e-05
2.5128140954798998e-05
0.00024405028984876727
1.7504122956129332e-06
3.692298262745975e-05
1.2494655723380719e-05
1.8048944021802955e-06
0.00011415649301429831
2.9888097170906303e-05
4.259006941491356e-06
1.627001932325115e-05

3.775513665996511e-06
9.423052858049625e-05
8.889672507593986e-06
0.0001070125876623932
1.2634260815262065e-05
9.082460586071927e-06
2.1416029222840057e-06
1.4286371989462474e-06
1.3874433656023988e-05
7.5384422864397e-05
0.0001382047752513945
0.00011765198944154443
7.156749006113639e-06
0.0003957682200380843
8.956565092799643e-06
0.00010600934465305828
9.616487960707535e-05
0.00023076864142162348
0.0002870216537944218
3.776626292813602e-05
0.00013862284414643933
1.3664850066053837e-05
0.00017030806566562545
1.1140555103112858e-05
9.070318259085201e-06
1.6927639864759803e-06
6.230117592098925e-06
7.580556489157241e-05
2.1201868930611657e-05
0.00013761862186198234
6.0307538291517596e-06
6.771055945903921e-06
8.92710270762596e-05
2.037416834172892e-05
0.0001837495307319677
4.7611214440671785e-05
0.00015132932470919843
2.0559388053926456e-06
0.0004644898782553754
3.097989980728644e-06
1.7016799743656206e-06
4.315902072389141e-05
0.00023912263166663567
0.00017421221831715142
0.00010769203266342428
8.14402352274649e-05

8.202450156229726e-05
0.00015137434310119878
8.368838682880562e-05
1.3706258702617638e-05
2.3395165716537002e-05
0.00010126265757904075
7.2784960007004e-05
0.00021322023777719707
6.821328287033603e-05
0.0001018708417086446
8.019619453659256e-06
4.634288290844078e-06
0.00021788853120274215
9.100735154655573e-06
7.366555980234235e-06
0.00011437636572529199
0.0001130766342965955
6.443113065333077e-06
4.1496012585906605e-05
1.1364485858954323e-05
8.931803656919075e-05
4.5050451910994226e-05
6.340745848407224e-05
3.5897344221141435e-06
4.492516261287068e-07
0.00026328290970550596
2.629689169688267e-06
6.55516720559974e-05
3.519895231022428e-05
9.268576581688156e-06
0.00018428988688177537
4.634288290844078e-06
4.9450423161776455e-06
0.00017526878315972302
2.4852007537713297e-05
7.390629692587942e-06
3.9262720241873435e-06
0.00043441870803211183
3.41363424291609e-06
0.0002389543970041263
2.599462857393e-06
0.00014366879578770982
0.0002206373352128693
2.7692236970594818e-05
2.8626996024454557e-05
5.951401805083973e-06

9.283795919260714e-07
1.3221471417316048e-06
0.00014781259385175882
1.7668224108843046e-05
0.00018603867853424455
9.230745656864939e-05
1.2874000109669319e-05
1.1085944538881913e-05
1.2224501005037351e-05
1.6039238907318513e-05
7.744974951821609e-06
1.2453373821210957e-06
6.593389754903527e-06
3.2171434415258995e-05
1.297246282561325e-05
2.8357776626105456e-05
3.132316739517881e-06
0.00023530397888308887
2.8809333578750448e-06
1.3874433656023988e-05
8.646242048963098e-05
5.771893165951511e-05
2.6296891696882677e-05
2.640435126598844e-06
3.263394929194676e-05
2.0606220372955902e-05
2.7247381758215784e-05
9.177234087839635e-05
2.8093573738284597e-05
4.7411586707167926e-05
1.1538432071081174e-05
1.4313498012227279e-05
6.230117592098925e-06
0.00025128140954799
0.00013960078308221667
0.00016556978066728
0.000250745628290958
5.3846016331712146e-06
2.6763700425229705e-05
2.9370554362752077e-06
8.711605107595956e-06
6.162214403084224e-06
6.632060662556921e-06
9.894205500952107e-05
0.00010318780316039133
3.2307609799027284e-06

0.00013741952084655703
4.365893216084768e-06
0.0002817397622204736
2.4824727617254775e-06
1.3542111891807843e-05
0.0002537879572741545
2.684311793390991e-05
2.754607412828149e-06
1.1195706365999553e-05
4.149601258590661e-06
2.3242884747501644e-06
1.0518756678753069e-05
6.419197580156077e-05
9.706148866660557e-06
3.7566988138403823e-06
1.0925278675999565e-05
0.0001646052271406137
7.594699318428055e-05
1.1288848681856456e-05
6.555167205599738e-06
7.798388572179e-05
1.445068808902179e-05
0.00032157806353282184
0.0002870406870605886
1.0233179574352533e-05
0.0001869035277629678
2.4635432308626473e-06
1.1934209424442799e-05
1.3664850066053837e-05
2.2391412731999107e-05
1.2923043919610914e-05
7.295266728812614e-05
5.139847013481613e-06
0.00015319222556829242
2.2391412731999107e-05
5.951401805083973e-06
1.4134579287074437e-05
5.4302338504014786e-05
2.3956914045888877e-06
0.00017112487682867568
0.00021538406532684855
1.6752093969866e-05
1.0378764047415833e-06
2.764709885002335e-06
0.00013662501000472738
0.00017029613598884864

6.417516134880561e-05
0.0001499690110034423
2.7635837302630264e-05
0.00030926942713598775
0.00010470058731166249
0.000245226435823942
0.0001078387520539739
6.791389447242973e-06
0.00010816025889239569
0.0001947541328834158
3.5708410830503844e-05
3.390603727034348e-06
0.00013326889042098754
0.0002769223697059481
1.0898952703286313e-05
4.90571081547052e-06
7.798388572179e-06
4.417056027210762e-05
5.128192031591633e-06
2.0132338450432434e-05
5.308762173549085e-06
3.472602972640169e-05
9.962699056968765e-06
4.157229202080717e-06
6.334825450789664e-06
1.3542111891807843e-05
5.874110872550415e-05
0.00014848446927835773
4.605972883771711e-06
7.49679343402843e-05
7.744974951821609e-06
2.0748006292953302e-05
1.825288689210581e-05
7.744974951821609e-06
0.00017948672110570712
8.149667336691567e-05
8.65067147624228e-05
0.0001347296068214755
8.125267135084707e-05
2.0282804358133722e-05
0.0003025584883657969
1.0599284387194742e-05
0.00010509459943235463
9.300872049763387e-06
5.8221421281773925e-06
1.1205869939473958e-05

7.815858865347383e-06
4.119987618942806e-05
1.63173193855498e-05
8.768498902133832e-05
9.029972863847947e-05
5.283335602232456e-05
0.00012380528518819814
8.131909988864163e-05
3.0370194448206972e-05
3.957817893516334e-05
6.0248563885106946e-05
0.00022909778542924537
0.00027094977399870924
0.0001297796099966984
2.505285400609666e-05
6.470171860704964e-06
1.1482558086127637e-05
0.00030481849574854795
0.000239654371834394
2.8508420075903098e-05
0.0001240116273301785
1.5764189914853195e-05
0.00010164887486080203
0.00010268478696129429
0.00014001312763084668
2.8618067845425803e-05
0.00022069865880794477
1.4822106852212565e-05
2.929408519610515e-06
7.647171263937009e-06
0.00016635706105267844
3.6474008038287786e-06
0.00018735569596645666
5.068595122486859e-05
1.1394636508132784e-05
2.8472057295194035e-05
8.017993146347746e-06
8.943146201695562e-05
0.0002745295566851279
8.439354600919519e-05
3.531976727758248e-05
1.3305968597658634e-06
1.2039963818463929e-05
2.0329774972160407e-05
0.0001494115991929861
9.029972863847947e-06

6.643480035545275e-05
5.658325201376964e-05
7.455608857525761e-06
0.00014670145188901238
0.00023349469580995948
3.016499043166504e-05
3.26346387710996e-05
0.00011851526711609854
2.554015208173012e-05
4.343813869394988e-05
0.0002944160936615748
5.314784028436221e-05
0.00013704624827745655
5.455056920682338e-06
2.204651152536506e-06
5.813045031102115e-05
2.8184460756858746e-05
4.3009812946882704e-05
1.6910676454115246e-05
1.746642638453218e-05
4.392383494575388e-06
0.00015522316745230955
2.3546511518388317e-05
7.202998683263029e-06
7.086378704581627e-05
0.000276967618383296
0.00011841959108452055
3.41316405495904e-05
9.97412552253446e-06
9.804726026104887e-05
1.625789724649099e-05
5.1492717230523936e-05
1.4971222615313297e-05
0.00015732818392634582
5.262162404392297e-06
1.0996104886419766e-05
8.304350044431594e-05
4.434265577956322e-06
6.098932491648122e-05
9.563878714409651e-06
0.00018601744099526772
1.252642700304833e-05
5.145710677600767e-06
1.9427408981229006e-05
0.00010752453236720676
6.069084534919011e-06

0.00012319035827501173
0.00021392005714455787
3.7924045055100456e-06
3.4987606456790164e-05
2.4761057037639632e-05
6.608079609068125e-06
7.915635787032669e-06
6.065786119410904e-05
4.413225171892473e-06
9.16342073868314e-06
8.379164008795843e-06
4.650436024881693e-06
8.719567546653174e-05
7.072906501721205e-05
7.500703265938215e-06
1.6758328017591687e-05
2.5024767846897e-05
1.0279573069045418e-05
0.0001194607419235664
5.138603342410711e-05
1.0814967499724869e-05
7.898829766253406e-06
1.4309033922712902e-05
5.813045031102116e-06
0.00020910625685650704
4.237299339299947e-05
6.098932491648122e-05
3.2016771255639877e-06
8.436165124501937e-06
0.00014704936047056736
4.886011583365381e-05
0.00015011933834705812
5.776939161343718e-06
1.0721466339784882e-05
2.8618067845425803e-05
0.000191075317034414
1.0127991342755047e-05
2.1884404822972672e-05
1.0137190244973717e-05
3.3217400177726376e-05
7.561684593303566e-06
5.813045031102115e-05
0.0002559453733595821
6.889534851676582e-05
3.5097630376465605e-06
4.3009812946882704e-05

6.403354251127975e-06
1.5062140971276737e-05
3.623716383024696e-05
4.368315639810592e-05
0.0001471324392052965
4.1568143239165963e-05
2.610771101687968e-05
1.3008212657011729e-05
8.087714825881205e-06
1.4589603215315114e-05
6.538398629007653e-06
2.4475979078324696e-05
1.0109643532351506e-05
0.0001395130807464508
0.00010543498679893717
1.3334583583890158e-05
0.00018244780993139265
6.149336892405544e-05
7.915635787032669e-06
0.0001982631032551443
1.062956805687244e-05
7.140784683119682e-06
2.670745742932774e-06
3.041157073492115e-05
2.0109993621110022e-05
1.0277206684821422e-05
3.729673002411382e-05
7.342793723497409e-05
4.7544393864605173e-05
2.4968784026210432e-05
8.493947077409484e-06
5.741279043063819e-06
0.00017246650158501644
5.8639473518648315e-05
5.697318254066392e-06
2.7355506028715842e-05
3.0432301185319875e-05
2.9179206430630236e-06
0.00010375474796548798
9.144991512616512e-05
3.444767425838291e-05
1.6995654727754016e-06
3.617843260847994e-05
6.000562612750572e-05
2.156723953568321e-05
6.425472918662098e-06

2.016449224880951e-05
0.0001316937635364727
3.253949405165033e-05
3.6604505383684165e-05
5.152837700699936e-06
6.1595179137505865e-06
7.832313305063903e-06
3.0382595507597833e-06
0.0001171763407844206
0.00019354415826097217
2.247944906287223e-05
0.00011273784302743498
9.18604646890211e-05
0.00024940885943499583
8.964695951579167e-06
7.470579959649306e-06
0.0004119417378441915
7.846078354809182e-05
6.049347674642852e-06
5.615620860234497e-06
0.0001060791924209266
6.200581366508924e-05
1.0334302277514872e-05
5.586109339197229e-06
1.1172218678394458e-05
6.200581366508924e-05
1.8192414767263345e-05
1.3099819788399134e-05
0.0005376226618360339
4.5676474154761865e-06
4.271353409765045e-06
1.4199804656127308e-05
3.3441337707014426e-05
0.00011686959643681741
1.8417568415373042e-05
1.55663130539973e-05
7.848837172796106e-05
0.00014540924844243998
3.665368295473256e-05
2.2013898342635233e-05
5.314784028436221e-05
1.6758328017591687e-05
1.1412112944494952e-05
0.0001630431753743382
3.188870417061732e-05
4.644630237085337e-06

6.538398629007653e-05
0.00013779069703353164
4.482347975789583e-05
8.379164008795843e-06
0.0002443814912611709
5.612595202443422e-05
2.3007723066823462e-06
6.631637825143235e-06
2.5943855089995493e-06
3.623716383024696e-05
0.00017548815188232804
0.00017715946761454068
0.00010871149149074087
2.4692580663088636e-05
3.916156652531952e-06
9.359368100390827e-06
9.790391631329878e-05
1.4989318371899092e-05
8.879114128652398e-06
8.943146201695563e-06
1.3310729230430606e-05
1.6534883644023795e-05
5.4711012057431685e-05
6.764270581646098e-05
1.1273784302743499e-05
0.00015735614344010562
1.2940343721409928e-05
0.00020413436597560244
1.1924194935594084e-05
9.300872049763386e-05
1.7239799906883013e-06
3.1577034736851e-05
7.750726708136155e-05
5.694411459038807e-05
9.074021511964278e-06
6.598162982096809e-05
6.80551613397321e-05
2.0219287064703013e-05
0.0001328696007109055
3.951812100051211e-05
9.790391631329879e-06
2.4002250451002285e-05
6.319063813002725e-05
4.3820363014197345e-06
7.006306628823643e-06
2.0441477032447004e-05

9.008108522773254e-06
6.498425886297562e-05
5.227656421412676e-05
7.750726708136156e-06
1.1342526889955349e-05
8.753761929189069e-06
4.5370107559821395e-05
0.00013599186286860303
8.131909988864163e-05
1.1773255759194158e-05
1.2484392013105216e-05
0.000182264927776343
9.442509695191254e-06
8.672141771341151e-06
9.743770718799737e-05
2.9133506812101443e-06
0.00010062767287224682
0.00013881898581736396
1.3334583583890158e-05
1.8417568415373042e-05
1.978908946758167e-06
7.485611307656648e-06
9.074021511964278e-06
3.257748528813796e-06
0.00010629568056872442
2.3501887681019293e-06
9.947456737714852e-06
5.131515613662558e-05
0.0002406195754167642
3.1435140007649805e-06
5.5198053707794565e-06
1.019273649289138e-05
0.00041736120657563083
3.3516656035183374e-05
9.418604607355327e-05
3.5772584806782254e-06
7.396319721481817e-06
2.8952130894205093e-06
0.00020109993621110026
0.00010131055787941964
0.00016608700088863187
5.955734503583812e-05
4.709302303677663e-05
8.249110465422071e-06
2.5637932143298765e-05
3.6783109801324124e-05

5.511627881341265e-06
1.5631717730694766e-05
4.0838077057138905e-06
0.00012740920616114226
8.183785349549833e-07
7.644552369668536e-05
1.1138768921872316e-05
0.00017611118674108186
5.254729971617731e-06
4.520472442169325e-06
1.0660025271935113e-05
8.39176425542561e-05
7.390442629927204e-07
0.0002179173973550772
1.0100494533679696e-05
1.0248894820675907e-05
0.00012737285546038303
1.8555355710251145e-05
5.2178805328265845e-06
0.00018858911068644652
4.5370107559821395e-05
9.34761010026471e-06
1.1308051124332384e-05
4.359783773326587e-05
0.00026693085703356805
2.565757806831279e-05
7.006306628823643e-06
8.134873512912582e-06
1.5247331229120305e-05
2.5081003280260818e-05
0.00015031712403658
5.636892151371749e-05
5.586109339197229e-06
5.615620860234497e-05
8.087714825881205e-05
5.645445857216016e-06
1.1518107801564562e-05
3.1662543148130675e-05
6.0889506054097445e-06
2.9065225155510576e-05
1.3334583583890158e-05
7.865430908890813e-06
0.00017715946761454068
1.5185097224103486e-05
8.964695951579167e-05
0.00019879726518578228

7.209978333149911e-06
1.0137190244973717e-05
3.692653915538813e-05
3.899736708496179e-05
6.341503670293219e-05
2.672664382115915e-06
6.436589653815492e-06
7.799473416992356e-06
0.00011123966571145578
4.650436024881693e-05
0.0003699794406535711
7.811756052294707e-05
8.087714825881205e-05
1.0305675401399873e-05
0.00031395348691184424
1.7715946761454068e-05
4.4555075687489265e-05
2.00789836078161e-05
5.365887721017338e-05
0.0002686918592153867
6.948019867395399e-05
5.813045031102116e-06
9.39482025228625e-06
3.6108205951847502e-06
5.782925626795888e-05
3.6835136830746083e-05
0.00012197864983296244
1.3479524709802008e-05
6.8388765071789606e-06
1.4822106852212565e-05
1.3779069703353164e-05
9.383669130325077e-05
3.659359494988873e-05
1.4518434419142846e-05
2.330072329794585e-05
6.462144352192923e-05
5.425508695695309e-05
0.00020796980980837383
1.5437132032802302e-05
3.8354115454160024e-05
9.947456737714852e-06
0.00016627257295666385
0.00016288742644068977
1.1172218678394458e-05
2.7599026853897284e-05
6.625278720379397e-05

0.00025725816307856173
2.2277537843744632e-05
2.0554413369642844e-05
5.1791398884993326e-05
4.359783773326587e-05
1.3933890711256008e-05
0.0001528105360466654
0.00024088589481401575
0.0001361103226794642
2.2685053779910698e-05
0.00021567239535683213
7.5770851729233284e-06
1.062956805687244e-05
0.0002700253175737757
0.00011217132120317652
4.073374620334329e-05
7.1821405789678646e-06
5.692841453122338e-05
7.427779761963814e-05
2.4968784026210432e-05
2.8662163481551267e-06
1.2611351931882557e-05
2.7763797163472793e-05
4.203783977294186e-06
1.9177055772708012e-05
4.5095137210973995e-05
6.239578733593885e-05
1.3192726311721116e-05
5.113881539388803e-05
1.4807358487185489e-05
6.915146505400288e-05
9.86456126490056e-05
0.00015205785367460576
7.381644483939194e-06
9.418604607355327e-05
2.4236800129676578e-05
2.7763797163472793e-05
2.7951531329116105e-06
1.0539231784434431e-05
1.252642700304833e-05
5.353019884755905e-05
9.074021511964278e-06
3.679870247186305e-05
6.447744921846367e-06
1.0277206684821422e-05
3.611989145539179e-05

1.0509459943235462e-05
0.0002511627895294754
3.2749549470997836e-06
8.601962589376541e-05
1.0000937687917619e-05
0.000159236719918128
2.2277537843744632e-05
8.230860221029544e-06
5.714821535952925e-06
3.8420125506767856e-05
4.1707946411494996e-05
0.00220928277260486
7.592548612051743e-05
5.924122324690055e-06
1.4392065067332126e-05
8.138263043542962e-05
0.0002783303356537672
9.490685765064679e-06
7.15451696135645e-05
3.5230575946073433e-06
2.1884404822972672e-05
4.50269146130754e-05
2.6061988230510367e-05
1.0509459943235462e-05
2.7888671813383464e-06
4.5186422913830215e-05
1.4226955334246096e-05
8.000750150334096e-06
6.403354251127975e-06
3.074668446202772e-05
8.542706819530091e-05
5.91201855603575e-05
5.138603342410711e-06
0.00014661473181893023
3.9161566525319515e-05
4.088295406489401e-05
0.0001594435208530866
4.342819634130764e-05
4.7696779742376344e-06
4.30928434738072e-05
0.0003993594778429477
1.6758328017591687e-05
1.8237004019143896e-05
1.3933890711256008e-05
3.949414883126703e-06
8.013058996719225e-05

4.9341496285216896e-05
9.026321439317478e-06
7.11718904677546e-05
5.09636824644569e-06
0.00020879508683142292
2.1756425847399732e-05
8.436165124501937e-06
4.576074809231678e-06
8.601962589376541e-05
7.252141949133244e-06
0.00023321589617317147
0.00014734054732298433
1.5592409136233672e-06
7.631484758780215e-05
0.0002212846881728092
6.005405681848837e-06
0.00022855385241739377
0.00032268331601219914
1.9831283688194854e-06
0.00017754376147053753
6.098932491648122e-06
1.3677753014357921e-05
8.607490842968686e-05
1.4589603215315114e-05
5.9644870860206085e-05
5.813045031102115e-05
5.905315587151355e-05
6.492755357600967e-06
4.110882673928569e-05
1.5062140971276737e-05
0.00013660028064447306
1.500140653187643e-05
2.506973598318972e-06
9.983046922822953e-05
0.00011081890101845736
6.470171860704964e-06
1.252642700304833e-05
0.00022894454276340643
4.5095137210973995e-05
1.098528194853943e-05
3.2071972585390986e-05
4.2228703971683935e-06
5.170741931765606e-06
0.00028388203846667364
1.625789724649099e-05
2.823794170706151e-05

1.0450418033442008e-05
0.00026959049419604015
2.113834556764406e-05
6.174852813120921e-05
0.00017029765724918875
4.982610026658956e-05
1.978908946758167e-05
1.2421865842755774e-05
2.969153088511855e-06
4.4027796685270465e-05
0.00012892297890761129
3.0063424807316e-05
2.138131505692732e-05
9.539355948475269e-06
5.180101392238031e-06
5.1671511387574363e-05
8.341589282298999e-05
1.2962887874234684e-05
3.676234011764184e-06
1.3677753014357921e-05
6.7642705816460995e-06
8.267441822011898e-06
6.555680739921328e-05
2.2685053779910698e-05
5.2032850628046915e-06
4.916760554940997e-05
8.230860221029544e-06
1.6559416112338375e-05
5.580523229858031e-05
0.00018081224014267555
9.096207383631673e-06
2.929408519610515e-05
4.088295406489401e-05
9.816223799222571e-06
2.6959049419604016e-05
0.0001355093941025129
4.122270160559949e-05
3.2164399019354646e-05
1.3830293010800574e-05
0.00012947849721248333
4.650436024881693e-05
0.00019009081561560204
0.0002202838117049223
9.726402143543411e-05
1.4971222615313297e-05
1.3239675515677417e-05

0.000191075317034414
0.00015270088439910034
6.403354251127975e-06
5.586109339197228e-05
8.221765347857137e-05
3.875363354068077e-05
1.1239724531436116e-05
3.885481796245801e-05
5.082443743040102e-06
0.0002872328133015163
4.665014194238689e-05
5.628364326634425e-06
0.0001311661442915349
4.227669113528811e-06
8.815992464230698e-06
0.0004340406956556247
7.4036792435927445e-06
3.683513683074608e-06
6.000562612750571e-06
4.133720911005949e-05
3.7731732453401155e-05
1.3334583583890158e-05
8.493947077409484e-06
0.00016940292349831737
2.2144933451817585e-05
4.162180263233185e-05
4.8722996705963476e-05
1.5697674345592213e-05
1.594435208530866e-05
6.593023225148729e-05
3.382135290823049e-05
2.7763797163472793e-05
3.000281306375286e-05
3.289109958658958e-05
6.515497057627592e-06
5.594509503617074e-05
2.5222703863765114e-05
7.086378704581627e-05
3.3546878448199764e-06
3.957817893516334e-05
2.5835755693787182e-05
7.019526075293121e-05
7.80882810774737e-05
6.0248563885106946e-05
1.1205869939473958e-05
0.000286180678454258

2.2277537843744632e-05
1.70658202747952e-05
0.00033387745819663434
2.6637342385957194e-05
7.507430354069102e-05
2.4422421137234275e-05
2.619963957679827e-05
5.113881539388803e-05
3.3729363734409375e-06
0.00025414147536355363
3.298181577930279e-06
0.00026755933293839875
2.9694916464466905e-05
0.0001361103226794642
3.4769615139302375e-05
5.1244474103379534e-05
3.4769615139302375e-05
7.099902328063652e-05
5.322387439063454e-06
1.069065752846366e-05
2.486864184428713e-06
8.105335119619508e-06
1.1626090062204232e-05
5.0049535693794e-05
2.1504906473441352e-05
0.00010846498017216777
7.946376120186194e-05
3.000281306375286e-05
1.9376816770340386e-05
6.515497057627592e-06
8.32292800873681e-06
5.9908998710231144e-06
4.5370107559821395e-05
5.353019884755905e-06
8.087714825881205e-05
1.3677753014357921e-05
2.352169959898011e-05
9.790391631329878e-05
1.2039963818463929e-05
9.254599054490932e-06
1.0783619767841606e-05
3.947319702817352e-05
4.4555075687489265e-05
1.7507523858378138e-05
0.00013684041636433488
1.0248894820675907e-05

4.697410126143125e-06
2.5024767846897e-05
0.00010969087429696377
5.041123062202377e-05
2.1756425847399732e-05
4.389792117882424e-05
0.00024002250451002287
5.936726840274502e-05
3.2294694617233977e-06
1.149438358364167e-05
8.836933063908204e-06
5.797946212839514e-05
1.9177055772708012e-05
7.4258459479148795e-06
0.00013026431442245639
6.190264259409908e-06
2.0219287064703013e-05
1.712473564973696e-05
4.133720911005949e-05
9.842192645252259e-06
1.5533815531963902e-05
5.561059521532667e-06
1.9356653589517972e-06
2.1756425847399732e-05
0.00014956176160423537
8.55252602277093e-06
4.251827222748976e-05
4.504054261386627e-06
3.1961759621180017e-06
9.713704490614503e-06
4.143311910799467e-05
2.9087950116539126e-06
4.420677146854392e-05
2.5308495373505815e-05
2.113834556764406e-05
3.059497384790587e-06
8.964695951579167e-06
3.603243409109302e-05
4.276263011385464e-05
9.300872049763386e-05
7.500703265938215e-05
5.1033591493900604e-06
6.93232699361246e-05
1.0752453236720676e-05
6.0889506054097445e-06
2.8662163481551267e-06

1.3544959295771919e-05
5.000468843958809e-05
3.126343546138953e-05
6.132443109734101e-05
7.038497767388508e-05
8.954887750319235e-05
2.315569804920345e-05
3.5097630376465606e-05
4.960465093207139e-05
9.51495861868377e-06
8.417078778066413e-06
6.098932491648122e-05
5.538980873308219e-05
5.05678480375485e-06
1.0248894820675907e-05
5.849605062744268e-06
2.3546511518388317e-05
1.414581300344241e-05
0.0001643527591122847
8.304350044431594e-06
2.868426229688014e-06
2.384838987118817e-05
0.00013479524709802008
0.000256271058853338
3.767441842942131e-05
1.1736116151120991e-05
1.3479524709802008e-05
5.020713657092246e-06
5.552759432694559e-05
2.638545262344223e-05
1.1172218678394458e-05
0.00014584403784375944
1.2697436245410764e-05
0.00016498220930844142
0.00010783619767841606
2.2182659062468038e-05
3.7579281009145e-05
6.118994769581174e-06
8.857973380727034e-05
9.902069102409688e-05
1.7144464607858774e-05
3.9161566525319515e-05
5.813045031102115e-05
2.2411739878947916e-05
0.00012148077779282789
0.0002920935023892634

0.00012853412429615432
0.00012568746013193763
8.513384027243373e-06
3.735289979824652e-05
1.184824464938011e-05
0.0002583575569378718
3.991045842249214e-05
4.574199368736092e-05
0.0001162609006220423
1.0897331048346086e-05
5.293179215312496e-05
2.958527888592727e-05
7.099902328063654e-06
7.223978291078358e-05
9.231634788847033e-06
1.305385550843984e-05
1.8417568415373042e-05
5.569384460936158e-06
0.00016643665773260794
1.3856047746388656e-05
5.034301515433497e-06
3.409927805855097e-05
2.898973106419757e-05
9.051943600742954e-06
0.00010137190244973717
4.16975583800102e-05
1.0363088634833856e-05
7.670822309083205e-06
6.0248563885106946e-05
1.777944477852021e-05
1.297796099966984e-05
1.2568746013193766e-05
5.2584435617036805e-06
0.0002618427321927522
1.1518107801564562e-05
2.1884404822972672e-05
3.4157836558495767e-06
8.923011117929085e-06
3.0431078379064503e-06
2.686008448764367e-05
2.2438259177743883e-06
1.5427330577518416e-05
3.7403741098246245e-06
3.5097177063854396e-05
0.0001554640057862007
4.368943202969427e-06

5.722365825628434e-06
1.2998954468094221e-05
1.607504293000965e-05
0.00017250235403872794
1.9992062884474023e-05
2.749126140771885e-06
1.4133091435109152e-05
1.847219845466021e-05
2.884699484700361e-06
2.0524664949622452e-05
8.774294265963599e-06
7.23653135337204e-06
3.524402717709228e-05
8.737886405938854e-06
5.64061774240517e-05
8.225900874340873e-06
9.236099227330104e-06
4.679623608513919e-06
5.1995817872376886e-05
1.583331296113732e-05
3.9484324196836196e-05
1.671294145897828e-05
1.2411575386038882e-05
1.007574461163284e-05
4.442680640994228e-06
7.058650135747254e-05
2.7928788114472993e-06
2.9701419235983973e-06
1.8122466642265606e-06
2.3555152391848586e-06
4.897280520537823e-05
1.8835694309760857e-05
6.83711241503657e-05
8.774294265963599e-06
5.706858059163316e-06
2.162043761633741e-06
4.262815027998509e-06
8.35647072948914e-06
3.6058743558754516e-06
4.780546251603323e-06
1.4276817788686533e-05
6.420215316558732e-06
9.999195744687862e-06
7.114292648078594e-06
6.362026053870887e-06
1.320269983593269e-05

5.238384636396179e-06
2.2889463302513735e-05
8.037521465004825e-06
8.006960546886933e-06
5.109172560852237e-06
3.202784218754773e-05
1.1291317017862003e-05
4.818834379476576e-06
2.9701419235983973e-06
0.00015824160757113543
1.0546731004163257e-05
1.2460536235687951e-05
1.331119231246058e-05
1.052915311915632e-05
4.1803089306824095e-05
8.792612208063732e-06
4.228575549861975e-06
2.2989417290734325e-06
1.8886373307903712e-05
2.8715872143153593e-05
2.5219528429116928e-05
3.161907843590486e-06
5.585757622894599e-06
1.012418569149646e-05
3.871012176160411e-06
4.916336086765597e-06
2.074709974217994e-06
4.430872851915173e-05
1.320269983593269e-05
3.545169400389333e-06
0.00014192301107788117
6.506170825431298e-06
1.7213874309247386e-05
1.0424904078372593e-05
1.853724140696535e-06
5.737958103082462e-06
4.1782353647445706e-05
9.302866209696344e-05
8.491252515448644e-06
6.103856880670329e-06
9.132428725798848e-05
1.1852705199800733e-05
4.319652561705157e-05
5.9824733631 57e-06
8.099348553197169e-06
8.339923262698074e-06

5.225386163353012e-06
4.8689725406503204e-05
8.554936909314509e-05
1.5621888900825398e-05
1.6355966010339911e-06
7.090338800778666e-06
8.644624892574975e-07
7.704258379870477e-05
2.573316444600322e-05
4.721593326975927e-05
2.074709974217994e-05
5.931917250228911e-06
1.949843170214133e-05
2.1079385623936574e-06
3.899686340428266e-05
1.2724052107741774e-05
1.3161441398945398e-05
1.7607279463472103e-06
0.00010027764875386969
3.8287829524204793e-05
1.4164779980927782e-05
6.749457127664307e-05
2.537145329917185e-05
2.303972236139238e-06
3.330839299557359e-05
6.30052890904636e-05
3.539211132489519e-05
1.8311570642010988e-05
4.26281502799851e-05
1.2033317850464364e-05
6.6429988133478345e-06
4.378026244971442e-06
0.0001286003434400772
2.7830360667373533e-05
5.02584874422736e-06
5.948674078619388e-06
1.8526369124615224e-05
1.7085846846501127e-05
8.389763441558819e-06
2.2331183709769502e-06
1.6490451243784368e-06
9.111767122346814e-05
6.653493282247279e-05
2.6455158590845025e-06
2.965958625114456e-05
8.423322495325055e-06

1.2919206281173398e-05
3.471149379941644e-05
8.006960546886933e-06
0.00015870027889742857
2.590197569288147e-06
8.25815930914221e-06
2.105830623831264e-05
0.00010529153119156318
4.018760732502412e-06
9.97775108548479e-05
1.8153712274407445e-05
1.163442333608433e-05
4.2866781146692394e-05
9.076856137203722e-06
6.323815687180972e-06
1.1141960972652189e-05
5.0138824376934855e-06
5.1361722532469854e-05
1.0911039501716393e-05
8.491252515448644e-06
5.031853342488085e-06
1.5893061311934065e-06
1.1764416892912088e-05
1.126112633064847e-05
6.139447882889982e-05
8.630453376357638e-06
1.367422483007314e-05
1.3854148840995155e-05
7.186068894379683e-05
0.00010081104050256605
8.630453376357638e-06
8.162134200896371e-06
1.4623823776605998e-05
1.8311570642010988e-05
1.1570497933138812e-05
0.00020550877172329198
2.5576890167991057e-05
1.707252766436322e-05
7.1384088943432664e-06
1.0744033795057468e-05
5.801186291546181e-06
2.8152815826621175e-06
5.291031718169004e-05
6.5195994545859555e-06
1.7237358448823443e-05
1.886945003433032e-05

1.0424904078372593e-05
9.557476356268973e-06
3.767138861952171e-06
4.840989939841986e-06
2.528007951778228e-06
3.452181350543055e-05
4.818834379476576e-06
5.864189985606415e-07
8.328143710067146e-05
5.801186291546181e-06
4.835936714435053e-05
2.5636198898815386e-05
3.244731315610576e-06
1.5656733262685977e-05
4.653769334433732e-05
7.520823656540227e-05
1.649475684463131e-05
1.7815825920738274e-06
1.3112270385001643e-05
1.163442333608433e-06
1.7260906752715275e-05
2.960002884903383e-05
5.585757622894599e-06
1.8418343648669944e-05
1.0854797030058062e-05
2.4344862703251602e-05
1.0744033795057468e-05
1.9589122082151288e-05
1.8564478023784282e-06
3.4241148354979894e-06
6.814985837641629e-06
3.5155770013877524e-06
3.290360349736349e-05
5.174031016784432e-06
1.2314798969773472e-05
2.8886565484653822e-06
4.4240139156118984e-05
4.721593326975928e-06
5.413446333756462e-06
1.389987210449679e-05
5.1867749355449853e-05
0.00023491337636364696
2.366101824529 51e-05
1.2534706094233712e-05
0.00020504140284672827
8.595227036045974e-06

3.452181350543055e-05
1.1444731651256867e-05
3.311054439986264e-06
2.770829768199031e-05
2.2343030491578395e-05
8.595227036045974e-06
1.923132989800241e-05
9.37313334049524e-06
7.574930301551308e-06
5.753635584238426e-06
2.55252196828032e-05
4.700514785337642e-05
2.5219528429116928e-05
1.4777758763728168e-05
0.00015549382243479796
4.753567999619106e-06
7.114292648078594e-06
5.660835010299096e-06
4.7859786905256e-05
7.090338800778666e-06
1.4276817788686533e-05
1.4228585296157188e-05
3.2497386170235553e-06
1.4633986267069239e-06
1.3718766279031035e-05
3.1987300615158435e-05
9.215888944556953e-06
5.737958103082462e-06
2.8152815826621175e-06
0.00012169372951788709
3.1197490723426126e-06
2.9045939639051914e-05
6.664020961491341e-06
1.1083319072796124e-05
0.00011344542081245866
9.54303908080633e-05
5.331216769193073e-05
1.482979312557228e-05
5.167682512469359e-05
2.9534791358082244e-06
3.115134058921988e-06
1.4871685196548472e-05
2.596585232837563e-06
4.137191795346294e-06
8.896622829874371e-07
1.5535452776328025e-06

3.452181350543055e-05
3.1954941181051047e-06
4.049674276598584e-05
1.9558798363757867e-05
8.5084065609344e-05
7.3373889332099784e-06
1.1507271168476852e-05
2.1882548914076175e-06
2.8807532473751898e-06
0.00015177157649234332
4.09694673897133e-06
2.558724937826566e-06
0.00012262869094599725
7.946530655967032e-06
3.294036730015385e-05
1.6594409959269218e-06
3.0083294626160914e-05
5.753635584238426e-06
4.811212321582852e-05
1.0322699136427764e-05
0.00019798407574481968
2.384859143636765e-06
9.119738922103899e-05
2.409417189738288e-06
5.0019729782215285e-06
1.1602372583092362e-05
3.9087343365777516e-05
2.36610182452951e-05
3.977680067236832e-05
1.3473004631038156e-06
0.00011818437174563214
2.1709594060116124e-05
3.0169493178098333e-06
7.069934948114554e-05
9.970788938595e-07
5.691434118462875e-06
6.286061563675413e-06
3.3965010061794575e-06
1.2033317850464364e-05
4.280143544372487e-05
5.5783592684271884e-05
1.3328041922982682e-05
9.762128719747581e-05
8.525630055997018e-06
1.2172431351625801e-05
1.9680660035806204e-05

6.3813049207008e-06
6.972179529294548e-07
1.1083319072796124e-05
1.6124277364711056e-06
2.84187668533234e-06
3.0431078379064503e-06
3.7806653928748003e-06
7.697553194161277e-05
6.580720699472699e-06
3.1073550635421596e-05
6.211889745814937e-06
5.48155476919201e-05
1.9717515204412582e-06
9.871081049209048e-05
9.744203400761632e-05
6.393413658690744e-05
0.00010424904078372592
2.363446266926222e-06
0.00041290796545711053
9.704288589084164e-06
3.720548805355589e-06
0.00012087849502354686
8.217875605195174e-06
6.664020961491341e-06
2.580674784106941e-06
4.203254738186155e-06
0.00020265315626828064
7.946530655967032e-06
3.023446696096574e-06
6.642998813347835e-07
1.3272041746835695e-05
1.5041647313080457e-05
9.09646057810481e-06
9.380091865618101e-06
6.016658925232182e-06
4.8689725406503204e-05
7.674309853612477e-06
0.0001293507754196108
4.518949836547776e-05
4.1129504371704374e-05
2.502237764402377e-05
4.245626257724322e-06
5.737958103082462e-06
7.3373889332099784e-06
2.071308810325833e-05
4.288860741000537e-06

1.0222478756462446e-05
1.568588919054945e-05
4.931687643632936e-06
6.193619481856658e-06
6.998228659257977e-05
2.9452176557080615e-06
1.2609764214558464e-05
9.863375287265872e-06
9.550252262273306e-06
1.8311570642010988e-05
2.6191923181980894e-06
8.147558961251913e-05
1.9543671682888758e-05
7.657565904840959e-06
2.699782851065723e-06
1.7016813121868798e-05
8.235091825038462e-05
6.656998810846988e-06
2.240245344501344e-05
3.865682650447479e-06
5.049953534367539e-06
2.243917877852986e-05
3.490327000825299e-05
1.4015511639475967e-05
2.770829768199031e-05
7.828366631342988e-05
3.8287829524204795e-06
3.814910550418956e-06
2.0863579496346736e-06
8.037521465004825e-06
2.0849808156745185e-05
2.834226949974783e-06
3.3693289981300217e-06
1.8206028448108908e-05
2.868979051541231e-05
6.671057942443284e-06
5.990036540607261e-05
2.1935735664908998e-05
2.5432737002793036e-05
6.121600650672279e-06
5.774672643047341e-06
6.086215675812901e-05
3.3320104807456706e-05
3.767138861952171e-06
2.876817792119213e-05
5.154130897489106e-05

1.7937228482378737e-06
1.2534706094233712e-05
1.1645146306900998e-05
8.906836475419242e-05
5.956592446777285e-05
7.744736094650883e-05
5.737958103082462e-06
0.00011980655627924039
1.680822618793483e-05
1.4325378393409958e-05
2.2889463302513735e-06
3.87814111202811e-06
7.799372680856532e-06
7.168785102404302e-05
2.699782851065723e-06
6.96782191708874e-05
4.3270492270505416e-05
4.714546172756561e-05
0.00011699059021284799
5.06209284574823e-06
1.6236165179886381e-06
6.809900027315032e-05
0.00016886377643929946
1.5041647313080457e-05
8.713781891715574e-06
2.4581680433827982e-05
4.43332762911845e-06
5.982473363157e-06
0.0001414363851826968
2.0645398272855528e-05
1.0635508201168e-05
1.874626668099048e-05
3.995883536681715e-06
7.491097080822676e-05
1.3808725402172221e-05
5.225386163353012e-06
4.802350339409952e-06
7.574930301551308e-05
1.2314798969773472e-05
8.45715109972395e-06
3.8659957456945444e-05
1.2172431351625801e-05
6.103856880670033e-05
4.15351207856265e-06
1.0689495552442963e-05
8.136903492392828e-05

2.425140833586868e-05
1.179737044163173e-05
2.409417189738288e-05
0.00012022908405957042
2.6656083845965365e-05
6.221065358438002e-06
6.687535502639863e-05
0.0001588609067100778
0.00013055666324556628
1.3313997621693975e-05
1.1321670020598191e-05
0.00013280012943080042
2.849996333004718e-05
5.086547400558607e-05
5.221067662391563e-05
3.4864745427669928e-06
0.00024568023944698076
1.0744033795057468e-05
3.3532334774383177e-06
0.00027785265175551396
1.5973430774952697e-05
2.916662913893717e-06
9.429092345513122e-05
2.537145329917185e-05
2.0385582031280383e-06
1.9928996440043504e-05
1.649475684463131e-05
4.4709779699177565e-05
0.00018543502477645624
2.724231078695037e-06
2.248217747862559e-05
2.8304175051495477e-05
1.2724052107741775e-06
0.00011122344844179211
2.9005931457730904e-05
8.441093217889033e-05
2.0379006037076745e-05
4.2399274305327454e-05
1.8635669237444813e-05
5.596892023471797e-05
2.0055529750773942e-05
1.1083319072796124e-05
1.2998954468094221e-05
3.8387537413590744e-05
2.6224540770003287e-06
6.193619481856658e-06

3.3479024226252207e-06
0.0001014858131966874
4.6796236085139194e-05
5.221067662391563e-05
2.3014542336953704e-05
3.012633224365184e-06
1.4463122416423514e-05
4.859609131918301e-05
7.976631150876e-06
7.742024352320822e-06
3.6879695688813728e-06
1.193105169309498e-05
3.0387166289051424e-06
9.089916361861571e-05
2.0849808156745185e-05
3.178612262386813e-06
5.865823464711041e-06
8.737886405938854e-06
3.4102520223988073e-05
8.525630055997018e-06
3.6686944666049892e-06
6.615174734548473e-05
1.3244217759945056e-05
6.267353047116857e-05
2.484755898325975e-05
3.557146324039297e-06
3.798091305507289e-05
5.865823464711041e-06
7.019435412770879e-05
3.87814111202811e-06
0.00016491444644461703
1.293507754196108e-06
3.3639466834365237e-06
1.9202102952868666e-05
9.338495005903608e-06
1.4522969819525957e-05
4.45207320048893e-06
5.157136221627585e-05
4.356890945857787e-05
3.662314128402198e-06
6.155599601962186e-07
8.774294265963599e-05
1.9319547007626274e-05
2.2027516985682674e-06
1.4934968963342297e-05
0.00010817623067626353

8.81100679427307e-06
1.712057417748995e-05
4.460155632925521e-05
4.840989939841986e-06
4.15351207856265e-06
2.5576890167991057e-05
8.722375364981565e-05
5.0271818606051394e-05
0.00015004689948674852
6.666502905983516e-05
2.448640260268911e-06
7.2740263344775955e-06
8.733859730175749e-05
0.00011751286963344106
4.721593326975928e-06
3.3965010061794575e-05
2.740777384596005e-05
1.6679846525396148e-05
1.6968820498237419e-06
3.662314128402198e-06
1.4934968963342297e-05
1.0744033795057468e-05
0.00012446282047429776
2.3559206634945292e-05
3.539211132489519e-05
1.2685726649585926e-05
2.699782851065723e-06
6.622108879972528e-06
1.4228585296157188e-06
5.706858059163316e-06
8.258159309142211e-05
4.6452146113924936e-05
4.558074943357714e-06
1.5259642201675824e-05
7.919363029792786e-05
4.742861765385729e-06
3.914183315671494e-06
3.3965010061794575e-06
1.4038870825541758e-05
9.968428988550361e-06
5.1222907066165883e-05
1.2647631374361944e-05
1.8122466642265606e-06
2.0017401367217333e-05
5.331216769193073e-06
1.8566488098744322e-05

3.446531299232837e-06
4.7589392628955106e-05
4.220101450563655e-06
1.5095560027464256e-05
4.908696092846769e-06
6.70799288592439e-05
2.3891109184471386e-05
7.606612519542587e-05
1.163442333608433e-05
3.152441053639616e-06
9.460155542817898e-07
9.476237807240688e-05
7.363044139270153e-05
6.211889745814937e-06
4.940365099897393e-05
1.7976602886364447e-05
4.742861765385729e-06
5.982473363157e-06
1.3992230058679493e-05
3.1197490723426126e-06
1.247899628937045e-06
2.55252196828032e-05
3.5097177063854396e-05
0.00011599914453307809
2.082918520110053e-06
2.044495751292489e-05
0.0001295895768511547
1.183050912264755e-05
8.290671747367179e-06
6.390988236210209e-06
1.27626098414016e-05
1.2879698005084182e-05
0.00010450772326706024
3.9484324196836196e-05
8.365628505631048e-05
0.0002537145329917185
0.00011674114332540055
5.0019729782215285e-06
1.5315131809681918e-05
9.53583678716044e-05
6.074511414897877e-05
6.139447882889981e-06
5.136172253246985e-06
1.2919206281173398e-05
3.655955944151499e-05
1.2998954468094221e-05

8.496146609117491e-05
8.941955939835515e-06
1.358600402471783e-05
8.913568778121751e-05
1.2387238963713316e-05
0.00023794696314477555
7.363044139270153e-06
3.713986990884063e-06
0.0001332086666488148
5.02584874422736e-06
1.6198697106394338e-05
1.367422483007314e-05
7.828366631342988e-06
1.7190454072091948e-05
2.118541875081754e-06
1.6936975526793004e-05
4.026444787440274e-06
0.00015259642201675822
4.480490689002689e-06
1.330278347334974e-06
5.707672161596863e-05
4.935540524604524e-05
1.1201226722506722e-06
6.059944241241046e-06
6.951285554394463e-05
1.29271370400937e-06
5.277770987045773e-06
0.00022821809716397928
4.775126131136653e-06
4.438518879269643e-05
0.00010662433538386146
7.062236848214604e-05
9.5719573810512e-05
2.0385582031280383e-06
5.111239378231223e-06
3.9503316175766854e-05
3.15716735207086e-06
1.3854148840995155e-05
8.693796153431824e-05
2.8846994847003613e-05
6.967181551137349e-06
1.1083319072796124e-05
6.286061563675415e-05
2.5119251974130378e-05
7.185248847718224e-05
0.00010754241903176287

2.621365921781656e-05
8.595227036045975e-05
5.331216769193073e-06
0.00018875792706610767
1.80913283834301e-06
1.7403558874638543e-05
2.909275556755142e-06
6.211889745814937e-06
2.431675085255501e-06
7.554549323161485e-05
4.9355405246045245e-06
2.5009864891107642e-06
3.205221649667068e-06
3.3910316003724053e-06
1.3498914255328614e-05
6.793002012358915e-05
9.19576691629373e-06
8.774294265963599e-06
4.17823536474457e-06
7.195321038147826e-06
6.643910061470379e-05
1.058206343633801e-05
5.4982522815437694e-05
0.00012172431351625801
1.6451801748681746e-05
1.4732956789864252e-05
6.749457127664307e-06
5.915254561323775e-06
8.454064548227702e-05
5.0138824376934855e-06
1.5315131809681918e-05
2.334623751475902e-05
3.0083294626160914e-05
3.5813445983524895e-06
4.461505558964542e-06
1.3079693315722136e-05
0.00010234522914054743
3.5097177063854396e-05
2.0808603002285217e-06
1.0222478756462446e-05
6.531732704191265e-07
2.8602113736248065e-05
6.601349917966344e-05
0.00010470981002475895
8.491252515448644e-06
1.6982505030897288e-05

9.900473078661325e-07
2.2643340041196382e-05
3.295509583460506e-06
1.7190454072091948e-05
2.838046662845369e-06
2.2283921945304378e-05
1.4934968963342297e-05
4.897280520537822e-06
3.947198920021113e-06
4.933131768858012e-05
1.6261240338465356e-05
1.4573222310250962e-05
3.2397394212788676e-05
2.3984403460492755e-06
3.857663858311583e-05
8.449320404261244e-05
1.1141960972652189e-05
0.00016719260151401338
6.63834522398647e-05
6.016658925232183e-05
4.344051385113609e-05
0.00010486548039689278
7.380806979054895e-05
5.1186104897117984e-05
1.3989485530654133e-05
6.366229647283185e-06
1.5688208773662135e-05
2.780188896598353e-05
6.642185166268243e-06
9.661360241844716e-06
3.7017121825494925e-05
3.8308416772895905e-06
8.195332941465295e-06
4.2346064813226845e-05
1.34470360917104e-05
2.895012163856809e-05
3.440755971247048e-05
7.5639578015871e-05
1.2819158920113028e-05
3.6810322262224e-05
9.306564526748722e-06
6.655603722159693e-05
1.4773649517798423e-05
2.989586063946505e-06
1.9942638271604406e-06
6.216082721639713e-06

4.8544064476459455e-06
6.935839668355891e-05
5.41565563145597e-05
7.256660445967066e-06
2.8303469437019315e-06
2.0818476097750697e-05
1.8151646514980982e-05
2.6568740665072972e-05
6.347830139632078e-06
4.207152254626486e-05
1.0981746141563494e-05
1.2339040608498308e-05
5.699868239565828e-06
8.90411849315959e-05
1.497510837485931e-05
1.2919701343015876e-05
1.0627496266029186e-05
2.9547299035596846e-05
7.897831129206622e-05
1.9436718834625654e-06
6.157988490596352e-05
4.888017570428854e-06
4.597010012747509e-05
5.4097271633317705e-06
9.834399529758352e-06
5.531849735489073e-05
1.4952453142824728e-05
1.9080253913913407e-05
8.928248895580076e-06
5.6077001573941246e-05
7.321164094375662e-06
5.348252991021182e-06
6.279884157199066e-05
5.084141732205322e-06
1.0981746141563494e-05
2.553894451526394e-06
5.481736842710563e-06
4.620650550447473e-06
5.2818017514533845e-05
1.5688208773662135e-05
1.3338153208376712e-05
8.295563632116307e-05
1.3959846790123086e-05
6.177232204629467e-05
5.167880537206351e-06
9.834399529758353e-05

6.863591338477184e-05
3.853244260197717e-05
3.7940773617685774e-05
5.490873070781747e-05
1.8076948381174477e-05
1.3125592997884653e-05
3.577116007023939e-06
1.71888200476646e-05
1.0392819692331382e-05
7.751820805809525e-06
7.009625196742656e-06
1.0199764218170427e-05
4.9766221185333054e-05
4.42217965432087e-05
5.650984292399739e-06
1.984652917150029e-05
1.3900944482991764e-05
1.984652917150029e-05
3.2683768278462776e-06
1.1399736479131656e-05
6.753050335954125e-05
6.216082721639713e-05
3.636339781974667e-06
3.9740938992389e-06
2.2160474276697632e-05
1.8934045071661198e-05
0.00012127081628720422
5.7697440323450936e-06
2.1674498963612157e-05
5.8830782901232997e-05
5.5839387160492344e-05
6.307320693942019e-06
6.792832664884636e-06
3.4679198341779454e-05
4.287883525990085e-05
2.8158323439906392e-06
9.776035140857708e-06
1.96492475693979e-05
2.9814695406959716e-06
2.2825800294242825e-05
7.586074637264255e-05
3.1556741786101994e-06
2.1674498963612157e-05
2.5887490904430073e-05
2.0047812428817735e-05
4.464124447790038e-06

5.893602580445524e-06
3.125734195890937e-05
1.766500719822546e-05
1.9154208386447956e-05
4.823607382824375e-06
5.739588575730049e-06
1.7856497791160153e-05
2.5342491095915757e-05
4.8807760629171085e-06
1.634999425542952e-05
7.487554187429655e-06
7.938611668600115e-05
1.0425708362243824e-05
3.336226675918023e-05
5.904164592238438e-05
6.026568004516552e-05
2.6568740665072972e-05
2.132377891565727e-05
0.00012902834892176428
3.998208546685738e-06
9.503434160968409e-05
0.00010385539258221168
1.8405161131112e-05
1.1321387774807726e-05
4.22374851598596e-06
1.3392373343370115e-05
0.0002625118599576931
2.538153961840561e-06
3.875910402904762e-05
3.792912551771872e-06
5.893602580445524e-06
9.190731749377172e-05
1.3502146895364952e-05
1.2292999412197941e-05
3.969305834300058e-05
0.0001231969108764059
7.614461885521684e-06
2.316009731085447e-05
0.00014890503242797956
4.067313385764257e-06
1.2432165443279426e-05
9.21545130760573e-06
1.4707695725308249e-05
1.7114409571267783e-05
4.809523857619049e-05
4.1078850903604096e-05

7.43419869440759e-05
1.2495033536545063e-05
1.757079382650159e-05
1.0295387007715775e-05
8.256951986137965e-06
0.00017339599170889726
1.2112220009077383e-05
7.938611668600116e-06
1.7712493776715312e-05
1.8988610043049268e-05
7.617396167558493e-05
6.397133674697181e-07
7.844104386831068e-06
5.918306303836614e-05
1.1340873812285878e-05
6.364999695651174e-05
1.3557711285880856e-05
7.844104386831068e-06
8.114590744997655e-06
0.0002196349228312699
6.02289550725603e-06
4.144055147759809e-06
6.93583966835589e-06
3.845747676033909e-05
6.135053710370667e-06
6.444983351205273e-05
4.4400590868855095e-06
6.397133674697181e-05
5.537014861292518e-05
1.5915574118207964e-05
8.90411849315959e-06
1.2339040608498308e-05
3.013284002258276e-05
7.573618028664479e-06
4.467150972839387e-05
4.173805543668558e-06
4.8025128898965715e-06
3.294523842469049e-05
7.564922715198733e-05
2.970715818276869e-06
2.022421020545763e-06
2.1674498963612157e-05
1.5915574118207964e-05
3.808698083779246e-05
1.204579101451206e-05
1.8002862527153268e-05

1.3502146895364952e-05
1.904349041889623e-05
3.3278018610798465e-05
2.0399528436340854e-05
4.9467324961997724e-06
9.306564526748722e-06
1.2934918894656647e-06
7.26733200544643e-05
8.248439664502771e-05
9.661360241844716e-06
6.6609863373818206e-06
1.1092672870266156e-05
7.679542756338107e-06
1.996681116647908e-05
1.1479177151460098e-05
6.100970078646385e-05
2.2411726819517338e-05
4.4320948553395263e-05
7.28876956298462e-06
1.4299148621827466e-05
6.491672595998125e-06
0.00014642328188751324
0.00014696760635549657
1.0425708362243824e-05
6.93583966835589e-06
1.34470360917104e-05
7.131003988028243e-05
7.4705756065057785e-06
5.335261283350685e-05
5.147693503857887e-05
6.950472241495882e-06
3.0862050046548466e-05
7.431256787524169e-06
1.2574518482706293e-05
1.634999425542952e-05
6.475722540479702e-05
2.2878637794923947e-05
6.62215847732472e-05
2.465960959931922e-06
3.985311099760945e-05
5.305191372735987e-06
5.392019382109734e-06
7.209023725315204e-06
2.4708928818517860e-05
8.154761986309524e-05
3.3824680107485095e-06

1.4019250393485311e-05
1.4642328188751326e-06
8.340566689795058e-06
5.8830782901232997e-05
3.389427821470214e-06
6.778855642940428e-06
3.7437770937148275e-05
3.7437770937148275e-05
9.747111959967599e-05
0.00014449665975741437
5.18239705556929e-05
1.7141123009724495e-06
9.776035140857708e-06
7.110483113242551e-05
3.550133450936474e-05
5.8681086252629363e-05
1.1399736479131656e-05
1.4707695725308249e-05
2.522606311232043e-06
0.00010201622098720187
1.3125592997884653e-05
0.00010890987908988588
7.645519465645472e-05
6.297379578160153e-05
6.820960336374841e-06
1.0199764218170427e-05
9.998554908859024e-06
9.863843839727687e-06
4.320687006516785e-05
1.9266221300988587e-05
2.1674498963612157e-05
5.400858758145981e-06
9.176946636404033e-06
6.921268576615648e-06
7.733624043354574e-06
7.683124632623714e-06
1.3213330918993509e-05
2.0336566928821288e-05
1.2432165443279426e-05
8.241460518996995e-06
8.123483447183954e-05
1.4139587306734112e-05
7.386824758899211e-06
4.306567114338625e-06
1.689499406394384e-05
1.0425708362243824e-05

5.6924818012424154e-05
1.0295387007715775e-05
1.1321387774807726e-05
7.128799659129316e-05
1.7953808405825875e-05
4.908042968296533e-05
2.5149036965412586e-05
6.409579460056514e-06
1.3524317908329426e-06
0.00011185111810851708
1.869763815249176e-05
6.892309293868302e-06
0.00015986086155440532
3.4899616975307715e-06
8.976904202912937e-06
1.122495346667478e-05
2.139301196408473e-05
3.998208546685738e-06
5.883078290123301e-06
8.215770180720818e-06
7.919528467473673e-06
4.4400590868855095e-06
3.0224989380449982e-06
5.9539587514500876e-05
9.521745209448116e-06
1.2526706625357596e-05
7.425673574310403e-06
1.304761917809524e-05
2.052662830198784e-05
8.404397557319e-05
9.521745209448116e-06
2.7115422571761715e-06
2.6356190739752385e-05
1.8348782191417699e-06
7.5389561612564035e-06
8.95791981335995e-05
1.0199764218170427e-05
1.9727687679455375e-05
5.1149982028453816e-05
6.70982452641354e-06
7.867519623806681e-05
1.7580169917124056e-06
1.65553961933118e-05
3.0585765115760393e-05
4.3636077383696e-05
9.834399529758352e-06

5.246057073995299e-06
2.623625358052712e-05
2.4864330886558853e-05
2.505341325071519e-05
2.1674498963612157e-05
4.084120465870721e-05
1.6639009305399233e-05
1.3755840678367634e-05
1.0425708362243824e-05
4.673083464495103e-06
1.297056630893326e-05
1.304761917809524e-05
1.0945261935113117e-05
4.017712003011035e-05
4.792034679954979e-05
7.177611857231042e-06
3.229925335753969e-06
9.3367072458637e-05
7.256660445967066e-06
6.93583966835589e-06
2.2260296232898976e-05
5.2073611840922784e-06
2.0015333186324716e-06
3.6005725054306536e-05
7.602747328774727e-05
1.6808795114638002e-05
2.915507825193848e-05
6.892309293868301e-05
1.2574518482706293e-05
3.8197377883699104e-05
1.0295387007715775e-05
4.6077256538028646e-05
2.0720275738799047e-05
4.615795225876075e-05
3.969305834300058e-05
6.792832664884636e-06
4.131064379271534e-05
6.239628489524713e-05
4.3853894741684506e-05
3.283578580533936e-05
1.3807727755528283e-06
6.112289132595637e-06
1.978692998479909e-05
7.198522598257207e-06
9.075823257490491e-06
9.633110650494293e-06

1.1282615898866604e-05
2.452499138314428e-05
8.459547669678013e-05
1.345252691902429e-06
2.522606311232043e-06
4.629307506982269e-05
5.147693503857887e-05
0.00020302387824739233
2.5738467519289436e-05
9.324124082459571e-05
1.0199764218170427e-05
5.565074058224744e-06
1.698208166221159e-05
1.0696505982042364e-05
1.5992834186742952e-05
4.446051069458905e-06
3.4557942403521487e-05
8.60188992811762e-06
3.036427504579768e-06
3.2748745948996504e-06
8.404397557319001e-06
7.28876956298462e-06
2.361665836895375e-05
0.0002856523562834435
1.2574518482706293e-05
3.414014344527511e-05
4.3065671143386256e-05
3.640357836982374e-06
9.718359417312828e-06
1.3338153208376712e-05
3.6005725054306536e-05
8.447497031971919e-05
1.021874640964345e-06
1.0801717516291962e-05
1.65553961933118e-05
2.579893377031361e-06
5.4605367554735606e-05
2.281074866200132e-05
1.2919701343015876e-05
1.5199648638842207e-05
2.7093123704515196e-06
1.5595379135948157e-05
6.70982452641354e-06
1.1808329184476875e-05
1.7070071722637555e-05
6.83511170636732e-06

1.1321387774807726e-05
1.780823698631918e-05
7.649823163627821e-06
8.62440796457866e-06
7.386824758899211e-06
8.808887279329007e-06
2.6676306416753424e-05
1.3465356304369405e-05
4.6141790510770976e-05
5.810447693948939e-06
1.4740598847736234e-05
5.386142521747762e-05
1.9906488474133223e-06
9.605025779793143e-06
5.472630967556559e-05
4.506872561517166e-06
6.544544780431165e-07
8.57576705198017e-05
1.8226964550312852e-05
1.9610260967077667e-05
1.2247300529624715e-05
5.392019382109734e-06
1.4200533803745897e-05
2.1818038691848e-05
4.607725653802865e-06
1.4324016706387166e-05
2.3574410321782097e-05
1.07014695579041e-05
5.69658301291478e-05
4.500715631788317e-06
1.6431540361441637e-06
1.2247300529624715e-05
8.62440796457866e-06
2.2720854085993436e-05
0.0002428395952679888
1.3784618587736603e-05
1.9210051559586286e-05
2.5342491095915757e-05
0.00010295387007715774
2.0984228295981197e-05
0.00024828295624404424
4.961632292875072e-05
2.2260296232898976e-05
1.5026334515252215e-05
1.3433328613533328e-05
0.00010981746141563494

0.001172001478973583
1.1724284136900528e-05
5.7697440323450936e-06
4.412308717592475e-05
0.00016815150046628412
2.6967453007932183e-05
3.5048125983713276e-05
1.9727687679455375e-05
4.939316105650747e-06
8.928248895580076e-06
1.3784618587736603e-05
8.976904202912937e-06
1.2574518482706293e-05
9.151455117969579e-05
9.834399529758352e-06
9.332928732206937e-06
5.570450941372787e-05
6.100970078646385e-05
8.114590744997657e-05
2.5342491095915757e-05
6.269855063522409e-05
1.5009220239038945e-05
3.5424987553430624e-05
1.4642328188751325e-05
6.669076604188356e-06
1.1130148116449488e-05
4.5504472962279675e-05
9.605025779793143e-06
1.2526706625357596e-05
5.4605367554735606e-05
4.877491556818241e-05
8.976904202912937e-06
0.00012671245547957877
6.549749189799301e-06
5.8830782901232997e-05
1.2067852902817027e-05
2.1254992532058373e-05
7.715512511637116e-06
3.4588176823822025e-05
2.7115422571761715e-06
1.2869233759644718e-05
5.262817639726914e-06
0.0001409818761416935
1.0360137869399524e-05
0.0002514903696541258
0.00022878637794923945

8.83250359911273e-06
3.5048125983713276e-05
2.553894451526394e-05
2.20861933573344e-05
5.5215483393336006e-05
7.131003988028243e-06
2.4920755238041212e-05
5.080221807970776e-06
2.2185345740532312e-05
1.9552070281715416e-05
1.975726442260299e-05
1.34470360917104e-05
1.0873015981746035e-05
1.2432165443279426e-05
2.7607741696668003e-05
4.11815480308631e-05
4.89528059802236e-06
1.010590135726702e-05
3.7999121597105518e-06
1.2671245547957878e-05
0.0001662374415924749
5.084141732205322e-06
2.449460105924943e-05
0.00011030771793981189
8.068221655026242e-05
5.400858758145981e-05
8.2776980966559e-06
1.678025726215135e-05
2.7004293790729904e-05
1.0044280007527587e-05
2.6719576986772495e-06
6.737267571511346e-06
7.05969394814796e-05
1.4513320891934132e-05
8.738790033074398e-06
2.654733152674495e-06
2.1406912556653984e-06
6.2574052088680875e-06
5.7697440323450936e-06
1.71888200476646e-05
8.165645367416134e-05
8.957919813359995e-05
3.131676656339399e-06
6.6556037221596945e-06
3.6810322262224e-05
6.778855642940428e-05

2.594113261786652e-05
1.540937250920977e-05
5.15171828376708e-06
1.7712493776715312e-05
1.7524062991856638e-05
1.8101779354225542e-05
3.5810041765967915e-06
4.412308717592475e-05
9.280348852025488e-06
0.00010349289557494392
9.93323771598708e-05
4.830680120922358e-06
1.497510837485931e-05
2.5738467519289436e-05
4.384347318875159e-05
1.0981746141563494e-05
9.41292526419728e-06
5.2881602607849885e-06
0.00010376453047146607
6.769569539319962e-05
2.562407433031482e-05
9.332928732206937e-06
1.7524062991856638e-05
3.1678113869894696e-06
2.614701462277022e-05
5.622054338684382e-06
1.5287813654148717e-05
4.554641256869652e-05
3.0224989380449984e-05
1.1907917502900174e-05
2.751168135673527e-05
0.00015626200043331455
2.0462881009124525e-05
0.0002978100648559591
3.6524654572827586e-05
1.307350731138511e-05
8.44749703197192e-06
0.0001326653896296261
9.261713613366803e-05
1.7712493776715312e-05
4.563052413392034e-06
1.1167877432098468e-05
1.0559371289964899e-05
1.820178918491187e-05
9.38610781330213e-06
1.323101944766686e-05

9.605025779793143e-06
8.669799585444863e-06
6.960261639019116e-05
0.00013663923770608957
2.0088560015055174e-05
4.659863992176871e-06
3.6203558708451085e-05
1.1282615898866604e-05
4.159752326349808e-06
1.701131071842882e-05
9.805130483538833e-06
2.3873361177311945e-05
1.0661889457828635e-05
4.2021987786595e-05
5.767218980252163e-05
2.542070866102661e-06
6.950472241495882e-06
8.83250359911273e-06
1.285667840963531e-05
1.3178095369876193e-05
5.127663568045211e-06
1.2067852902817027e-05
2.3038628269014325e-06
9.776035140857708e-06
1.5839056934947346e-05
3.2813982494711633e-06
1.4620667939951398e-05
1.6472619212345245e-05
6.2514683917818745e-06
9.521745209448116e-05
2.9001090162579652e-06
1.3727182676954368e-05
5.454509672962e-06
3.875910402904762e-06
2.2335754864196936e-05
6.575895893151793e-06
1.611014103896845e-05
7.977055308641762e-06
4.544170817198687e-06
1.9436718834625655e-05
4.050644068609486e-05
2.9859732711199832e-05
1.878559568050776e-05
7.115602251553019e-06
4.131064379271534e-05
1.639066588293059e-05

6.255425017346294e-05
1.2574518482706293e-05
2.594113261786652e-05
1.904349041889623e-05
5.820713502595491e-06
9.893464992399545e-06
3.26190479452381e-06
2.614701462277022e-05
4.789762855675607e-05
1.4282617814172173e-05
4.9171997648791765e-05
5.546336435133078e-06
6.628820608589633e-06
6.950472241495882e-06
5.339584833823417e-06
3.384784769659981e-05
3.6810322262224e-05
5.014495955051824e-06
1.367022341273464e-05
2.5342491095915757e-05
1.2671245547957878e-05
1.1559732780593153e-05
4.535829062600801e-06
2.2260296232898976e-05
1.5801073584983443e-05
3.4899616975307716e-05
4.909871598314528e-06
2.4224440018154766e-05
4.844888003630953e-05
1.8665857464413873e-05
2.522606311232043e-06
1.1479177151460098e-05
2.2526658751925114e-05
1.4449665975741439e-05
8.715671540923408e-06
5.351989655674394e-06
1.4827035806876318e-06
2.939825303821146e-05
1.9461780566088706e-05
7.109942462282772e-05
8.113803559622988e-06
7.255583741770873e-05
6.1165596064850215e-06
3.092260689945205e-05
5.311134772806652e-06
7.593546032607599e-06

6.427331688107817e-06
2.6889223390827873e-05
2.6941283842697823e-06
1.5207839458746914e-05
1.3252545814050882e-05
4.96970468026908e-05
1.026950044631249e-05
2.7959389545872414e-05
1.7084313204117157e-06
6.457156893157042e-06
4.896835110177158e-05
3.174564206407626e-05
5.231268084493769e-06
6.522737392853168e-06
1.2260064409474381e-05
4.2489078182453214e-05
1.0413600078393583e-05
1.5207839458746914e-05
1.4803375643354706e-05
2.733825757318944e-06
9.97503448369421e-06
2.460684899160641e-06
1.9850460919762374e-05
2.9669878688173616e-06
3.353053760181548e-05
0.00012869524258731491
6.787889319391914e-06
4.092697971986302e-05
5.115872464982877e-06
1.1620186308771129e-05
5.8131271455888986e-05
7.66676204118646e-06
4.653904048412517e-06
6.277521701392523e-06
6.835523630405192e-05
9.747932122419213e-06
1.5375881883705441e-06
1.602515136823811e-05
2.246194205771336e-06
6.738582617314007e-06
6.82953281214892e-05
2.309572299544137e-05
3.653874775865232e-05
4.375840598979065e-06
1.24242617006727e-05
1.533352408237292e-05

1.65656822675636e-05
1.122191379415599e-05
6.254010384158843e-06
3.522828634114791e-05
2.5985383949119376e-06
3.21273837916385e-05
5.949480187305503e-05
3.353053760181548e-05
6.905793104095992e-06
1.8719963818053933e-05
5.738215713300382e-06
4.697104845486388e-05
1.490911404080724e-05
3.859964800208995e-06
9.530940482707825e-06
4.717007832119805e-06
1.5812696709947076e-05
1.8042363831122755e-05
4.129131485090037e-05
7.228661353118662e-06
1.676526880090774e-05
1.1644496321969393e-05
7.099578114670115e-06
1.1405879594060184e-05
4.5623518376240736e-05
0.00011718040509266041
2.545458494771968e-05
5.933975737634723e-06
1.52244782327718e-06
1.618043384273654e-05
7.382054697481923e-06
1.876203115247653e-05
3.897807592367906e-05
2.046348985993151e-05
4.235973547870145e-06
1.3845943387814355e-05
2.4534539708645475e-06
3.81237619308313e-06
8.589613027625572e-05
6.0369514554244784e-05
1.4055730408841843e-05
6.1019824863551716e-05
5.435614494044307e-05
9.248755537902579e-05
4.75971715856757e-05
9.02849836480352e-06

1.2498658477323436e-05
1.62750562628695e-05
1.3845943387814355e-05
4.960846026650063e-06
7.891402516873398e-05
8.971329655378676e-06
1.2650157367957658e-05
7.839534143523056e-06
5.1729268047410506e-05
4.355296746401698e-06
6.472173537094616e-05
8.38263440045387e-06
9.730890283044353e-06
1.8553564139671233e-05
6.548316755178082e-06
9.154719147864095e-06
6.9229716939071775e-06
3.400856970611835e-05
3.83338102059323e-06
0.00010742715062777071
3.854618588574356e-06
1.0786955895157694e-05
1.6969723298479788e-05
7.583200602045464e-06
3.876092786839394e-06
1.0157060660403959e-05
3.938256539081158e-05
9.006584533820986e-06
6.787889319391914e-06
2.6967389737894235e-06
8.420679639790273e-06
1.1359324983472183e-05
4.96970468026908e-05
7.501441026821254e-06
2.5262038919370818e-05
5.959388909958641e-06
5.939403154467926e-05
1.6467660478998137e-05
1.3985098597742137e-05
3.87501339592131e-06
4.166219492441145e-06
1.4725050904500977e-05
1.5166401204090927e-05
1.676526880090774e-05
1.906188096541565e-05
5.972177298177435e-06

9.184932742411502e-06
3.431138573774817e-05
1.7147471478439217e-06
3.3230264130754444e-05
1.0181833979087872e-05
4.013992241755796e-05
2.5544145212948006e-06
9.370486939227896e-06
1.554767944665187e-05
7.974311234815716e-06
5.044775747946861e-05
2.6337867075243076e-05
5.834454131972085e-06
2.154051564203316e-06
2.3505359974245653e-06
7.951527488430528e-06
5.489220159666046e-06
3.849287165906895e-06
1.6086905323414364e-05
8.69698319047089e-05
1.1995838883408125e-05
1.8189768764383564e-05
9.957190057068641e-06
3.092260689945205e-05
4.129131485090037e-06
8.807071585286978e-06
8.145467183270298e-06
5.115872464982877e-06
5.029580640272323e-05
2.147403256906393e-06
1.3177247258289229e-06
4.153783016344306e-05
7.362525452250489e-06
4.7170078321198055e-05
3.217381064682873e-05
2.0166917543120905e-05
3.217381064682873e-05
6.862277147549634e-05
2.9606751286709414e-06
2.911124080492348e-06
3.31313645351272e-05
6.863217314305018e-06
3.092260689945205e-05
5.2842429511721863e-05
1.5375881883705443e-05
1.2795561475635334e-05

2.2507356416908086e-06
2.8989943968236304e-05
4.952018898488763e-06
7.583200602045464e-06
3.5539734845146274e-05
1.2479975878702624e-05
5.0785303302019795e-06
2.55324277151439e-05
2.9481298950748785e-06
8.407959579911435e-06
2.7284653146575343e-06
5.808990669916042e-05
2.6255043593874386e-05
5.908778388430329e-06
1.690101996528756e-05
2.214616409244577e-05
1.880428797939652e-06
4.840060210349017e-06
3.899172848967685e-05
1.3084318857313987e-06
2.1136971804688745e-05
1.4850771723322757e-06
2.002183180540061e-05
8.53691601518615e-06
6.310736101928992e-06
1.1290201302031177e-05
2.4628624964165355e-05
6.671658337895478e-05
5.500068420851157e-06
2.111024996928459e-05
2.0909351021417614e-06
1.5375881883705443e-05
1.4962551725541317e-05
7.099578114670115e-06
1.1073082046222884e-05
4.5623518376240736e-05
1.4479888766652887e-06
2.3288992643938785e-06
1.3915173104753427e-05
3.4077974950416554e-05
3.5172633440135036e-05
9.450032668762937e-06
1.5084198487537589e-05
8.2828411337818e-05
1.3509876800731482e-05
1.5994451844544168e-05

7.861679720199674e-06
6.8379229015987356e-06
1.0913861258630137e-05
2.1879202994895324e-06
4.123014253260274e-06
3.7107128279342464e-06
1.0157060660403959e-05
4.873966061209607e-06
2.839831245868046e-05
1.1267346643525039e-05
2.244382758831198e-05
3.748194775691158e-05
2.594904075478494e-05
5.691277343457434e-06
9.163762334378285e-07
3.4657965391664824e-06
2.135866938565376e-06
1.783996551891465e-05
2.5650088672356543e-05
6.502417338669825e-06
2.7970197195484275e-05
0.00012100150525872543
1.2896360616082878e-06
1.0703979311348788e-05
8.185395943972604e-05
1.113213848380274e-05
9.580153600518707e-06
3.8176057900558095e-05
7.323775318291276e-06
1.726448276023998e-06
1.6246553537365354e-06
1.6086905323414364e-05
1.0028953589011477e-05
4.308103128406632e-06
2.0531424721141167e-06
7.997225922272084e-06
3.4787932761883565e-05
5.425018754289834e-06
4.9142804645039975e-05
5.529207856193414e-05
2.182772251726028e-06
8.209541654721784e-06
1.4647550636582551e-05
6.0831357834987655e-05
6.89490559244539e-05
1.5084198487537589e-05

1.9326629312157535e-05
7.583200602045464e-06
1.2635798506019002e-05
1.9668089193997774e-05
1.0384457540860765e-05
3.5377558740898537e-05
4.6730058933873435e-05
7.695026601706964e-06
6.157156241041339e-06
1.24242617006727e-05
5.785934762891237e-06
5.4569306293150686e-06
2.139150361991303e-06
2.1457475874716152e-06
9.953628830295724e-07
8.043452661707182e-06
7.730651724863013e-06
1.2387394455270112e-05
6.742210387229116e-05
2.1490614833596024e-06
4.657798528787757e-05
3.5497890573350575e-05
2.4892975142671602e-06
8.863167582645494e-06
1.3709530152466429e-05
5.665210424327094e-05
1.0786955895157694e-05
4.600057224711876e-05
5.972177298177435e-06
4.129131485090037e-06
2.701101217357507e-06
5.6141908779985e-05
2.0166917543120905e-05
2.1407958622697576e-05
9.530940482707825e-06
3.5771653225587215e-05
2.354513215694319e-06
2.2626297731306382e-05
1.950269531149744e-06
2.1407958622697576e-05
1.830943829572819e-05
9.765033757721703e-06
9.530940482707825e-06
7.71416845892514e-05
1.2183391086088051e-05
9.563692855500635e-06

2.3635113553721317e-05
2.55324277151439e-05
7.421425655868493e-06
4.123014253260274e-06
1.4962551725541317e-05
2.605837660066184e-06
6.405142971117802e-05
5.33915514810683e-05
9.245962195849452e-06
6.184521379890411e-06
2.932597071602408e-06
7.542099243768794e-06
3.1199939696756555e-05
3.56799310378293e-06
5.425018754289834e-06
6.368500276775023e-06
3.892356113217741e-06
3.8019598646867284e-06
2.2000273683404626e-05
2.7609470445939332e-06
3.118246073894325e-05
2.2811759188120368e-05
5.871381056857985e-06
2.3854582465291587e-05
1.2914313786314083e-05
3.7107128279342466e-05
1.97098769189142e-05
1.783996551891465e-05
3.522828634114791e-05
9.782195504220334e-06
5.234547876396273e-05
5.112348327808376e-05
4.294806513812786e-06
4.033383508624181e-05
3.750720513410627e-06
1.554767944665187e-05
5.921350257341883e-06
1.2100150525872543e-05
5.1922287704303825e-06
2.891464541247465e-05
1.1021919290893802e-05
1.276621385757195e-05
1.0662967896362778e-05
2.9989597208520308e-05
2.1742457976177225e-05
1.5207839458746914e-05

8.97753103532479e-06
2.147403256906393e-06
1.0581880688025417e-05
6.010873911340572e-06
7.839534143523056e-06
4.554884813339909e-06
8.113803559622988e-06
1.8430692853978046e-05
5.734274631079709e-06
7.461218822924088e-06
2.6759948278371973e-05
1.5235590990605938e-05
3.975763744215265e-05
5.218189914282535e-05
8.020272682855001e-06
2.248916865414695e-05
5.810093154385564e-06
9.339042352183507e-06
4.1128590457399286e-05
7.521715191758609e-05
1.2502401711368757e-06
1.9371470679471125e-05
3.795047210387297e-05
2.1004034875099506e-05
5.797988793647261e-05
2.7691886775628708e-06
1.2592916836880927e-05
8.45907179620269e-06
5.280900609014582e-06
1.1842700514683766e-05
7.88395076756568e-06
3.319723205905389e-05
3.8071609041733036e-06
1.0541797806631383e-05
5.125293961235148e-06
7.66676204118646e-06
4.3827316865365115e-05
2.6090949571412673e-05
2.7970197195484275e-05
1.8703189656926645e-05
6.9229716939071775e-06
1.783996551891465e-05
3.614330676559331e-05
3.702485083304681e-05
4.1434758624575956e-05
7.583200602045464e-06

5.679662491736092e-05
3.998612961136042e-06
2.7048932601032576e-05
5.834454131972085e-06
4.0987255095002724e-05
1.0703979311348788e-05
3.1733576065572237e-06
1.495309372824325e-05
3.381573050972886e-06
5.908778388430329e-06
0.00011154447378559 86
0.00014082825792762505
6.722305847706968e-05
6.787889319391914e-06
6.457156893157042e-06
8.066767017248362e-05
1.7321792246581027e-05
1.886803132847922e-05
3.389810744154306e-06
7.266408931986123e-06
2.1163761376050833e-05
3.381573050972886e-06
2.2886797869660237e-06
7.421425655868493e-06
3.799364670239843e-05
2.2534693287050078e-05
2.7500342104255783e-06
1.2737000553550046e-05
4.351891510478006e-06
9.065259351630897e-06
2.2017678963217445e-05
7.928873563962065e-06
8.38263440045387e-06
2.130960659227171e-06
5.9213502573418825e-05
2.8989943968236304e-05
4.481537231804646e-06
2.37866206918862e-05
2.7554798227234505e-05
2.785820441392077e-06
4.261921318454342e-06
2.642957854654022e-06
1.3709530152466429e-05
6.706107520363096e-06
2.37866206918862e-05
4.25540461919065e-06

1.3127521796937193e-05
9.276782069835617e-06
7.687940941852721e-06
9.530940482707825e-06
2.3386845554207438e-05
0.0001623436862221233
2.4369830306048027e-05
2.1083595613262766e-05
3.176980160902608e-05
3.7608575958793045e-05
4.1745519314260276e-05
8.344931397153478e-06
1.70738320303723e-05
6.487260188696235e-06
1.8249407350496295e-05
5.090916989543936e-05
9.573092062238229e-05
2.3127711531446136e-05
5.467651514637888e-06
2.5185833673761858e-06
3.31313645351272e-05
7.795615184735812e-06
7.13598620756586e-05
2.5888694148378463e-05
2.6759948278371973e-05
1.9564391008440667e-05
6.1165596064850215e-06
4.790076800259354e-06
6.163974797232967e-05
7.861679720199674e-06
3.109535889330374e-05
5.702939797030092e-06
2.4959951757405248e-05
2.2383656468772265e-05
1.3379974139185987e-05
5.88379412463147e-06
1.2242087775442895e-05
2.449854419850955e-06
1.2341616944348936e-05
2.164582482961644e-05
5.260935011248932e-06
3.4962746494355343e-06
2.7554798227234505e-05
1.1693422777103719e-05
1.1595977587294522e-05
3.0650161023685953e-06

4.934458547784903e-06
5.083168257444173e-06
1.419915622934023e-05
6.442209770719178e-06
6.650022989129474e-06
5.489220159666046e-06
3.8019598646867284e-06
6.8379229015987356e-06
1.174276211371597e-05
3.3774692001828705e-06
4.503292266910493e-06
2.7374111025744444e-05
5.510959645446901e-06
5.588422933635913e-05
3.868011981863356e-06
1.0157060660403959e-05
5.859020254633021e-06
8.696983190470891e-06
4.749205837799803e-06
8.344931397153478e-06
2.7784039477045107e-05
1.4764109394963846e-05
3.7206345199875468e-06
2.645470172006354e-06
5.708788966052688e-05
6.143564284659349e-06
5.588422933635913e-06
7.951527488430528e-06
6.706107520363096e-06
7.1635382778653415e-06
1.778296882396604e-05
4.074721260542731e-06
2.7464157443592286e-05
8.246028506520547e-07
1.4570861889794162e-05
5.251008718774877e-05
1.6303659173700558e-06
7.382054697481923e-06
2.841764418942156e-06
2.1004034875099506e-06
7.567163621920292e-05
9.703746934974494e-07
4.1434758624575956e-05
1.8390096614211577e-05
7.773839723325935e-06
6.931593078332965e-06

6.927026142922642e-05
2.7365138849072616e-06
1.5812696709947076e-05
5.622292163536738e-06
1.5375881883705443e-05
0.00010703979311348789
5.510959645446901e-05
1.3444611695413936e-05
1.5461303449726026e-05
2.4885555477949496e-05
2.348552422743194e-05
8.233830239499068e-06
1.3985098597742137e-05
7.099578114670115e-06
8.33243898488229e-06
1.4091314536459166e-05
5.1300177344713086e-05
1.1267346643525039e-05
2.028450889905747e-06
6.722305847706968e-06
2.0645657425450184e-06
2.7760943849882144e-05
0.00013859734168081102
4.592466371205751e-06
6.325078683978829e-06
2.2749601806136387e-05
7.951527488430528e-06
4.32820314300262e-06
4.123014253260274e-06
1.5375881883705443e-05
1.7805723742486788e-06
1.4570861889794162e-05
6.740914306800967e-05
3.3939446596959576e-05
1.554767944665187e-05
7.111672796296469e-06
2.7998336226868057e-06
4.863555648263333e-05
8.484861649239894e-06
1.03458536094821e-05
6.975024112658358e-06
1.7614143170573956e-05
0.00022607284035733858
1.7852681103746375e-05
0.00020545893174803715
6.517645482320106e-06

0.00011792519580299512
2.026973503969909e-06
2.094081731339868e-06
5.3768056818985414e-05
5.311134772806652e-06
6.726367663929148e-05
5.5218940891878665e-06
6.900085837067815e-05
3.085404236087234e-06
1.4419868502335156e-05
3.217381064682873e-05
3.0360377683098382e-05
4.908350301500327e-06
6.339486608088121e-06
1.1313148865653191e-05
5.588422933635913e-05
2.9065635727944493e-05
1.2621472203857984e-05
1.641908330944357e-05
2.747319467868396e-06
5.57722368927993e-06
1.8932208305786975e-05
9.5966711067265e-05
4.159991959567541e-06
2.16410157150131e-06
9.402143989698261e-06
1.0871228988088612e-05
6.706107520363096e-05
2.545458494771968e-05
2.9218211243576742e-05
3.4591390768878574e-05
9.370486939227896e-06
1.3763771616966792e-05
3.5352592220968476e-05
4.004366361080122e-05
2.0925072337975074e-05
4.1169151197495345e-05
1.9254873926065425e-05
2.6771030853345352e-05
2.945847024402867e-06
5.364663604596158e-05
5.150881885706954e-05
3.5764424030641056e-05
4.180681530522425e-05
3.872966392489048e-06
1.4672584217698894e-05

3.2193011785668464e-06
2.7371278181895697e-05
3.971647453996303e-05
1.8650484827830595e-05
4.448830200118615e-06
4.7537344505940345e-06
1.9340288449184857e-06
2.344007309739916e-06
3.0067955835284435e-05
5.489024311657141e-05
2.287779248336859e-06
1.85445161640361e-05
3.7864733465029404e-06
1.5581505363928027e-06
1.981751634598292e-05
4.2095138169398206e-06
2.62190508357506e-06
6.228362280166061e-06
3.65934954110476e-05
6.722241227492004e-06
4.6452017005804715e-06
6.305573382812748e-06
3.708259437765942e-06
1.1220211460593272e-05
5.824231903208722e-06
2.3170588525997386e-05
2.161400507989639e-06
1.1890509807589754e-05
1.547615373874934e-05
1.852441588638161e-05
4.7242995623550615e-06
1.3269119640353783e-05
1.4301300455863964e-06
6.57736533896847e-06
1.4166633037778243e-05
8.765240911251976e-05
1.1729045031826942e-06
1.1810748905887654e-06
2.6615385325128228e-05
3.2605742705997546e-06
0.00013113622144568385
2.38429493537607e-05
4.157898524906499e-06
4.624087147396015e-05
2.825831034519787e-05
1.8209412394280252e-06

3.3985495738099887e-06
1.8440468382364774e-05
1.4486855303550806e-05
1.5087063997859881e-05
1.3665212763946432e-05
5.00311068406782e-06
2.4106615460358372e-06
1.3492031464550707e-06
5.673212734698448e-05
3.0641541338166364e-05
9.304565601467593e-06
2.9803686692200876e-06
1.2142828318095636e-05
3.6418824788560504e-06
2.5574560200681873e-05
7.3895823662987165e-06
1.2611146765625496e-05
2.6129259565765158e-06
4.359853596116243e-06
2.766070257354716e-05
3.4839012754353536e-06
1.0105620918150232e-05
5.48902431165714e-06
7.443652481174073e-06
1.5145893386011762e-05
2.986201093230303e-06
1.1765217876952082e-06
7.353969921159928e-05
0.00011622705532644038
1.097804862331428e-05
3.8058104431656045e-05
2.838974434680344e-05
3.012732001264926e-06
2.547493753991127e-06
2.9920563894915394e-06
3.1184238936798744e-05
1.3834530903360697e-06
6.013591167056887e-06
2.862943261990028e-06
5.261892271174776e-05
1.4532845320387477e-05
1.5259487586406852e-05
2.6264178289856883e-06
2.1492236037192748e-05
1.7030678109829072e-06
6.305573382812748e-06

3.2466994864695428e-06
5.48902431165714e-06
1.6768667677370165e-05
1.6768667677370165e-05
0.0002557948786160686
8.07380295577082e-06
0.00023522530271536253
5.672671965207008e-06
2.6584473147050263e-06
9.210958301654035e-06
8.579631574686583e-05
3.0276761084140572e-06
2.934516843539779e-05
2.0436813731794887e-05
2.0867675331838427e-05
1.0264229318210885e-05
2.0318891593085022e-06
3.892726425103788e-06
5.430422628614538e-06
4.250553645238677e-06
1.9315807071401077e-05
1.2769445679001548e-05
1.2716239655339043e-05
5.449816995145303e-05
1.812682110696278e-05
1.7519503543521068e-06
4.875235650609218e-06
2.7007942630808584e-05
2.2741412200308273e-06
3.590467667389847e-06
2.062092917082007e-06
3.302919390997154e-06
7.579215688612674e-06
5.4069050503016394e-05
2.9572650361253586e-06
6.438602357133693e-06
8.855952617111119e-05
5.498914445552018e-05
2.2606648276158296e-06
3.536967321352581e-05
2.363194980195751e-05
2.5291968375259974e-06
4.180681530522425e-05
1.6861312250173316e-05
6.17793019692585e-06
1.4815036491657137e-05

1.5138380542070286e-06
4.286372917530015e-06
0.0001500933205220346
3.331765848560448e-06
9.002647543602861e-06
3.507928180783184e-06
1.8473955915746791e-06
3.0367139475436523e-05
7.591784868859131e-06
0.00016153710866517648
4.238746551779681e-05
3.7864733465029404e-06
3.973824892293451e-06
1.038060380027677e-05
1.23558603938517e-05
4.298447207438549e-06
1.192147467688035e-05
5.715163890040019e-06
3.830833703700464e-05
2.7249084975726514e-05
9.137417716411286e-06
1.3064629782882579e-06
0.00011910940236812417
2.630946135587388e-05
1.1473298937148007e-05
3.7584944794105544e-06
2.7200512631741266e-06
5.891694048805734e-06
1.4031712723132736e-05
9.973521298305132e-06
4.828951767850269e-06
3.472708018325521e-05
2.1552948568371255e-06
1.0505671315942753e-05
2.2606648276158296e-06
6.676025819052997e-05
0.000123422326066526
0.00014188646352273037
3.502177478847474e-05
9.309501710539988e-05
6.13446737141984e-05
5.5691560534331576e-05
2.048253367302933e-05
5.813138128154991e-05
1.774359021675215e-06
1.3685639090947849e-05

4.2803611743076725e-06
5.548904576875219e-06
6.265973163500398e-05
3.7469582778162925e-06
5.24381016715012e-06
9.557090346392599e-06
3.5569901133815502e-06
3.018691906311939e-06
2.1096065787659238e-05
4.702461505826456e-06
5.715163890040019e-06
2.1135024357904226e-06
1.4532845320387477e-05
5.016094535324052e-05
4.666509965262034e-06
8.052500045597284e-06
9.537179741504282e-06
6.936130721094023e-05
3.0641541338166364e-05
6.605838781994308e-06
4.501323771801431e-06
7.86571525072518e-06
9.466183366257351e-07
6.079477126058506e-06
2.280939848491308e-06
2.7432786672192094e-05
6.57736533896847e-06
4.238746551779681e-06
1.3935605101741417e-06
2.515300151605525e-05
1.1684140571521326e-06
1.1025641319658129e-05
2.1737161803998363e-06
3.668146054424723e-05
1.7030678109829073e-05
2.5097841425011267e-06
1.1057599700294819e-05
1.2306038376134558e-05
5.8242319032087216e-05
2.2115199400589638e-05
2.0932081737183606e-06
7.130601675891052e-06
4.281999067614167e-05
5.335485170072326e-06
2.257320648876753e-05
0.00010310464585410032

1.7784950566907754e-05
2.568937304108897e-05
2.870123057004423e-05
8.489905245731485e-05
4.555070921315477e-06
1.9050546300133399e-06
4.0873627463589773e-05
3.5820393395321246e-06
5.984112778983079e-06
9.448599124710123e-06
2.4259916671553023e-06
5.143647501036017e-05
2.0789492624532493e-06
3.6332113300968692e-06
2.4612076752269116e-05
8.07380295577082e-06
6.678112729280898e-06
2.6196545212715623e-05
1.2877204714267386e-05
4.448830200118615e-06
8.76982045195796e-06
4.776052452709499e-06
3.8631614142802155e-05
4.448830200118615e-06
1.1387677303288695e-05
3.4680653605470113e-06
2.7055829053912857e-06
1.2110704433656229e-05
6.279624521155082e-06
2.1193732758898404e-05
1.4772011216269943e-06
5.869033687079558e-06
2.7702549324793074e-05
8.883948252360153e-05
1.3758554381186505e-05
1.740625960426637e-05
7.032021929219747e-06
7.316192678414243e-05
1.740625960426637e-05
5.078032474677821e-06
7.0645775862994675e-06
4.914488755686586e-06
6.82751122434311e-06
8.402801534365005e-07
5.715163890040019e-06
3.353733535474033e-06

2.810218708362219e-06
4.448830200118615e-06
4.4102565278632516e-05
2.87553158035305e-06
5.071128805150313e-05
4.624087147396015e-05
1.7910196697660623e-06
9.083028325242172e-06
2.2195618307500875e-05
3.5736504886198716e-06
4.08007689476119e-06
8.293199775221114e-06
3.7218262405870366e-06
9.497606381581028e-06
1.823116796464379e-06
0.00013530580125877995
8.524853400227291e-06
4.157898524906499e-06
6.228362280166061e-06
7.554201775448936e-06
3.6858665667649396e-06
2.9783156023985818e-05
4.938345497219045e-06
1.655042037571242e-06
6.812271243931629e-06
5.06959720478633e-05
4.091015438714973e-06
0.00015143002948342675
1.870035243432212e-06
4.4445109474971405e-05
3.0367139475436515e-06
5.683235600151527e-06
1.6233497432347714e-06
3.957792169385062e-05
4.768589870752141e-06
1.8609131202935185e-05
6.883979362288805e-06
2.4974611434381096e-06
3.3985495738099887e-06
1.1113974935474763e-06
4.986760649152566e-06
1.1684140571521326e-06
1.3685639090947849e-05
3.7864733465029404e-06
8.353734080149734e-06
1.23558603938517e-05

0.00012324970742867074
8.384333838685082e-06
7.629743793203426e-06
8.921068451567875e-07
3.491873589566785e-06
6.203043734311728e-06
1.8023016834338797e-05
2.291214352313341e-06
3.5052421714563976e-06
4.135362489541152e-06
1.4672584217698896e-06
2.5995719908699917e-06
8.524853400227291e-06
2.283215100210501e-06
4.652282800733796e-06
1.30758248383949e-06
1.4305769612256424e-05
8.031309256003606e-06
9.151116993347436e-06
3.902682247162878e-06
6.751985657702146e-06
1.3385515426672676e-05
4.936348592448637e-06
4.610117095591193e-06
4.196359086261884e-05
3.390997241423744e-06
1.963898016268578e-06
2.2606648276158296e-06
5.7044813407128414e-05
3.6505951163652752e-06
4.184502994444292e-06
1.7847353902230235e-05
5.449816995145303e-05
3.7677747126930495e-06
4.596231200724955e-06
1.5895299569173805e-05
2.1047569084699103e-06
7.785452850207577e-06
9.543144206633429e-06
4.448830200118615e-06
6.936130721094023e-05
8.270724979082303e-06
5.683235600151527e-06
1.848975687590281e-05
2.3233993869283512e-05
6.792056789201863e-06

1.1865853488652293e-06
5.316894629410053e-06
2.379647186964031e-05
1.224023068428357e-05
7.225136167806274e-07
5.914530072250717e-06
1.2306038376134558e-05
0.00014532845320387475
6.228362280166061e-05
6.043361420359149e-06
1.870035243432212e-06
2.048253367302933e-06
1.3716393336096047e-05
8.923676951115117e-06
1.865463030123087e-06
6.5397803941743646e-06
1.4129155172598935e-05
2.7643999250737048e-06
8.89766040023723e-06
5.579337325925723e-06
6.722241227492004e-06
1.660280548471566e-05
3.0216807101795743e-05
2.734675194696568e-06
5.373059009298187e-06
4.938345497219045e-06
6.230905506903573e-07
6.413795132640358e-05
1.854129718882971e-06
1.1516594404835358e-05
9.973521298305132e-06
0.00011807936822814824
1.6285472344084151e-06
1.9563445623598527e-05
6.144760101908798e-05
5.869033687079558e-06
3.6046033668677594e-05
3.454978321450608e-05
6.116027088740222e-06
5.4824506298467725e-05
1.1138312106866314e-05
3.474139030180194e-05
2.2899640307022555e-05
4.768589870752141e-06
1.2823098812106596e-05
8.45400974316169e-06

2.8791486012088395e-06
5.137874608217795e-06
5.846546967971973e-06
4.1491053860320744e-05
4.4139013679689236e-05
2.672414638600149e-06
8.031309256003606e-06
4.5136140756152234e-05
7.164078679064249e-06
9.719418844845127e-06
2.3085457770660897e-06
4.448830200118615e-06
4.250553645238677e-06
2.898450156077279e-05
5.213947922462478e-06
2.6538239280707568e-06
4.488084584237309e-06
1.0172991724271233e-05
7.706811912326693e-06
1.6631594099625995e-05
3.565300837945526e-06
1.727489160725304e-05
9.137417716411286e-06
3.633211330096869e-05
7.6680842142748e-06
3.2329422852556887e-06
1.2528314931368516e-06
6.446762816394951e-07
3.3834784005336696e-06
3.491873589566785e-06
7.147300977239743e-06
2.277535460657739e-05
5.134417088293018e-05
6.5304511782055e-06
5.373059009298187e-06
5.627142927742223e-05
3.984200414205444e-06
4.410256527863251e-06
1.2372557502492043e-05
1.2306038376134558e-05
1.0938700778786272e-05
5.926014596662855e-07
5.202098040820517e-06
1.23558603938517e-05
1.0172991724271233e-05
1.7068778060857774e-06

9.19246240144991e-06
3.8608342086089626e-05
3.0827247649306772e-06
5.75829720241768e-05
4.6381421235279185e-06
2.825831034519787e-05
2.7660702573547163e-06
4.501323771801431e-06
5.459566220539124e-06
4.922415350453822e-06
2.0903407652612126e-05
4.7242995623550615e-06
4.310589713674251e-06
1.3956848402201389e-05
3.6332113300968692e-06
1.0505671315942753e-05
2.684953827520267e-05
4.047609439365213e-06
3.2536220866539128e-06
7.989260516443376e-06
3.1141811400830306e-05
9.50154893300551e-06
0.00022253419396843323
8.430656125086658e-06
7.071124924192238e-07
7.920149266301134e-06
1.6631594099625995e-05
3.676984960579964e-05
4.2297087126500864e-05
2.354859195433156e-06
3.088965098462925e-05
2.6505313425520585e-05
0.00012390182228279068
8.415158595444955e-06
2.30042526428244e-05
3.9126891247197054e-05
1.7702421793975463e-05
6.1654495298613544e-06
1.8130084261077446e-05
2.0817854824565963e-06
4.08007689476119e-06
7.6680842142748e-06
2.749457222776009e-06
2.122320943867434e-06
3.293414586994285e-05
7.312853475913826e-06

1.9438837689690254e-05
3.980735892106135e-05
1.0189975015964508e-05
4.582428704626682e-06
1.9869124461467256e-06
2.4592244297190735e-06
8.536776272115721e-06
3.1593142000842343e-06
1.5108403550897871e-06
1.5093459531559695e-06
4.186416347436722e-05
4.047609439365213e-06
2.6538239280707568e-06
3.334192480642429e-06
9.040544563197075e-06
2.477189543247865e-06
2.1252768226193384e-06
1.5429208884132306e-06
3.5820393395321246e-06
7.283764957712101e-06
4.605479150827018e-06
5.392045083535989e-06
2.354859195433156e-06
1.8166056650484344e-05
7.629743793203426e-05
3.7584944794105544e-06
3.226107312136755e-06
1.4672584217698894e-05
3.6947911831493582e-06
0.00011622705532644038
8.270724979082303e-06
2.9979346928107763e-06
0.00016414588240357968
7.86571525072518e-06
5.124454786479912e-05
4.474923045867112e-06
4.652282800733795e-05
1.0139194409572658e-05
3.5322887931497338e-06
3.980735892106135e-05
3.540484358795093e-06
1.3809491028422491e-06
4.624087147396015e-06
1.0746118018596374e-05
2.6771030853345352e-05
2.062092917082007e-05

5.884121177277706e-05
3.120549608672158e-06
0.00025211327316672187
2.481217493724691e-05
2.1076640312716646e-05
3.366063438177982e-05
2.895538441443425e-05
1.9999328422551573e-06
2.7395848449563464e-06
2.0211241836300463e-05
1.431471630995014e-05
4.2028249708323956e-05
3.864940443252944e-06
1.7242358854697007e-06
1.6233497432347712e-05
0.00017673417826064432
4.695226949663647e-05
1.0596866379449202e-05
2.6174078192807633e-06
4.2095138169398206e-06
8.017243915800448e-06
2.287779248336859e-06
2.6771030853345355e-06
3.541231296423531e-05
2.1454464093366396e-05
1.636406175485989e-05
1.1648463806417444e-05
4.461838475557559e-06
3.6159923190537556e-05
1.038060380027677e-05
3.372262450034663e-05
3.6947911831493582e-06
2.1193732758898404e-06
1.3788693602174867e-05
8.315797049812997e-06
5.261892271174776e-06
2.630946135587388e-05
7.825378249439412e-06
4.227004871580845e-06
9.002647543602862e-07
6.479612563230086e-06
3.421409772736962e-06
3.699269717916812e-05
5.589555892456722e-06
2.76941698482883e-06
6.384722839500774e-06

4.372346013297092e-06
5.4777647746075874e-05
1.6079544348163172e-06
6.98374717913357e-06
1.760033170289141e-06
3.448471770939401e-05
7.2664226601937385e-06
9.002647543602861e-06
6.634559820176891e-05
2.3431074988724533e-05
5.542472307272967e-05
4.938345497219045e-06
2.7200512631741266e-06
9.027979668725553e-05
1.9480196918817258e-05
4.488084584237309e-05
7.164078679064249e-06
1.1516594404835357e-06
1.7419506377176768e-06
6.848331598042199e-05
3.729406334763385e-05
2.586353828204551e-05
9.719418844845127e-06
1.573143050145036e-05
1.8340730272123618e-06
1.4395743006044197e-06
2.0537668353172073e-06
3.120549608672158e-06
3.133365007475739e-06
3.05189751728137e-06
3.730926060246174e-06
1.2306038376134558e-05
0.00012716239655339044
9.436912545706153e-07
4.95437908649573e-06
3.912689124719706e-06
3.824432979049336e-06
1.266347517544137e-05
0.00010832352298992517
1.2559249042310166e-06
5.8020865347554555e-06
3.212523702401442e-06
2.3226008502902358e-06
5.342490183424718e-05
7.989260516443376e-06
1.573143050145036e-05

3.1462861002900726e-06
6.936130721094023e-06
9.029282595507012e-06
2.481217493724691e-05
6.103795034562741e-05
4.624087147396015e-06
9.657903535700539e-06
8.248371668328028e-06
4.8597094224225635e-06
4.286372917530015e-06
2.945847024402867e-06
6.737080612100155e-06
1.7641026111453006e-05
1.2975754750345962e-06
6.438602357133693e-06
1.9947042596610264e-05
1.116547872176111e-05
1.9830393224700262e-06
4.0631771679781555e-05
1.0172991724271233e-05
2.766070257354716e-05
4.16925890338985e-06
1.2059658261122378e-05
2.3719410756072826e-05
1.422133046263453e-06
1.048762033430024e-05
4.906587648362332e-06
1.0282673575745857e-06
0.0010002017073443147
1.1345343930414016e-05
2.1583433644139818e-06
4.307547660241877e-05
5.14890685430906e-05
4.058374358086928e-05
6.253888355084775e-06
6.186278751246022e-05
1.4960281947457696e-05
6.30557338281275e-05
3.902682247162878e-06
1.1057599700294819e-05
9.390453899327292e-06
2.1208460856715564e-06
3.548718043335043e-06
5.298433189724601e-06
7.288113474104763e-05
5.937543807940409e-06

1.5518122969227306e-05
7.554201775448936e-06
5.780108934245019e-06
8.477493103559362e-06
3.372262450034663e-05
0.00020078273140009014
1.0421113473643704e-05
7.336292108849447e-06
2.3905202485232665e-05
2.489312819968491e-06
1.2559249042310164e-05
1.146467887784136e-06
4.372346013297092e-06
9.083028325242172e-06
9.657903535700539e-06
5.1321146591054425e-05
9.657903535700539e-06
1.7499412369732627e-06
4.146599887610557e-06
2.2341855909819693e-06
6.7453530588242465e-06
1.0039136570004507e-05
1.0439786262079944e-05
4.185894350385919e-05
2.7577387204349733e-05
1.5355459206447144e-05
4.353634118803666e-05
1.66770356135594e-05
4.9826898241328486e-05
1.0708412341338142e-05
4.76858987075214e-05
1.067594747183315e-05
2.9572650361253586e-06
4.521329655231659e-07
0.0001191141436069733
9.809670591261548e-05
1.8058565191014025e-05
1.584813250813768e-05
3.356684466873482e-07
1.619903140807521e-05
5.19588286992348e-05
4.892821670992177e-05
5.360475732929807e-05
2.0874812019708414e-06
7.516988958821109e-06
1.1648463806417444e-05

3.476845272852193e-05
7.419523948009814e-06
2.8469193258221737e-06
4.423039880117927e-06
1.7340326802735057e-05
3.0641541338166364e-05
4.135362489541152e-06
4.772318244380563e-06
1.0451703826306063e-05
5.960737338440175e-06
2.179926798058122e-05
4.768589870752141e-06
4.8597094224225635e-06
0.00018899365359311237
2.3992905010073664e-06
1.2284023996213028e-05
8.719707192232486e-06
3.943020048167145e-06
7.785452850207577e-06
6.187951170481286e-06
2.336828114304265e-06
2.5097841425011267e-06
4.671271710124547e-06
1.4287909725100047e-06
3.973824892293451e-06
3.192361419750387e-06
4.507972699086219e-06
1.781609759066766e-06
5.7366494685740035e-06
1.1678179275311363e-05
1.4911551387368257e-05
2.0808392163282067e-05
2.3898962547230777e-06
4.5465884801212216e-05
3.5308673489673795e-05
4.4811478229015485e-05
2.2008876326548345e-05
1.753964090391592e-06
1.2473695574719495e-05
7.6680842142748e-06
4.5861847937288344e-05
1.6750260797373052e-06
7.879253486957065e-06
1.6062618512007212e-05
2.1235867416470766e-05
1.3192640564616873e-05

5.846546967971973e-06
2.7577387204349733e-05
3.011740971001352e-05
1.8979462172147825e-05
2.2051282639316257e-06
2.2946597874296014e-05
2.690476800424364e-06
2.424890195342746e-05
3.824432979049336e-06
4.322801015979278e-06
2.534798602393165e-05
5.914530072250717e-06
2.79990597915722e-06
5.846546967971973e-06
1.614760591154164e-05
2.110579195906895e-06
3.300721108540583e-05
2.4691727486095226e-06
1.858646478246876e-06
2.7152113143072687e-05
1.1481931968703422e-06
7.09743608670086e-06
1.4960281947457697e-06
4.203715588541832e-06
1.096227556494745e-06
8.604974202861005e-06
2.912115951604361e-06
4.582428704626682e-06
5.8020865347554555e-06
5.787416278030412e-06
2.526405229537558e-06
7.72632282856043e-06
2.9745589837050387e-06
1.1783388097611468e-06
4.1381661251272816e-05
3.1790599138347606e-06
5.316894629410053e-06
8.226138860596685e-06
5.651662069039574e-05
6.174359139008552e-05
2.116433784522448e-06
2.396464481571551e-06
1.9074359483008565e-06
6.984976712087643e-05
3.912689124719706e-06
3.339056364640449e-06

3.777100887724468e-05
2.895538441443425e-06
1.2968403047937834e-05
8.129721676295604e-07
3.863161414280215e-06
3.039738563029253e-06
1.9817516345982923e-06
3.5432246717662968e-06
7.735224122141722e-05
1.5078545045856571e-06
2.005188907543607e-06
2.5574560200681873e-05
3.590467667389847e-06
5.019568285002253e-06
1.568292660473469e-06
2.663086838814459e-06
1.2812332146437322e-06
2.067681244770576e-06
7.629743793203426e-06
4.82725442100744e-05
8.681124417045618e-06
1.2769445679001548e-06
5.844059075645177e-05
6.549136303178906e-06
4.324551137848095e-05
3.3172799100884456e-05
2.734675194696568e-06
1.504880432584502e-06
3.5160109646098733e-06
3.076509594033639e-05
3.2466994864695424e-05
9.627436963032713e-07
3.5569901133815502e-06
5.2923078334359013e-05
1.1783388097611467e-05
6.358119827669522e-05
3.212523702401442e-06
4.2743662707021984e-05
2.7577387204349733e-05
6.649014198870088e-06
0.00010636259934763139
1.66770356135594e-05
1.1829060144501433e-06
8.38894314810712e-07
3.8053585003508354e-06
2.577616146352509e-06

2.9086999798136227e-05
3.088965098462925e-06
1.6842701530250387e-06
4.986760649152566e-06
1.9315807071401077e-05
1.952589582393711e-06
1.0207015107964448e-05
1.0822331621565143e-05
1.089963399029061e-05
3.6858665667649396e-06
9.597162004029466e-06
5.824231903208722e-06
1.1693093935943947e-05
7.938461750153852e-05
3.7958924344295654e-06
4.875235650609218e-06
1.618185321994364e-06
2.729783110269562e-05
9.617324109079947e-06
5.3076478561415136e-06
7.554201775448936e-06
4.2683881360578606e-05
8.07380295577082e-06
1.7126248694059317e-06
6.079477126058506e-06
2.128240946500258e-06
3.8861173819372286e-05
7.164078679064249e-06
2.47117207877034e-05
1.2823098812106596e-05
1.714549167012006e-05
1.8015923950893563e-06
6.222013287016046e-06
1.3211677563988615e-06
9.913356204626259e-06
3.8631614142802155e-05
2.2514920820961787e-06
8.670163401367529e-06
1.4743466267059759e-06
0.00018576767496495295
9.597162004029466e-06
1.2909887974963496e-06
4.528037859467908e-06
6.3581198276695521e-06
5.521525113502478e-05
4.7330916831286755e-07

| |
|---|
| 9.373149623100031e-07 |
| 8.467266935762547e-05 |
| 3.740070486864424e-06 |
| 1.3533913602134678e-05 |
| 1.739964377013324e-06 |
| 2.8469193258221737e-06 |
