## Supplementary material for "Novel antibiotic resistance genes from the hospital effluent are disseminated into the marine environment in Norway": supplemantary table S8

**Supplementary Table S8:** Assembled known and novel antibiotic resistance genes (ARGs) across all

| Gene name | Sample type | Resistance against |
|---|---|---|
| HiACLASS_a_sequ16ence | STP-inf-A | Betalactams |
| HiAAMG_B_sequ1ence | STP-inf-A | Aminoglycosides |
| HiAAMG_B_sequ3ence | STP-inf-A | Aminoglycosides |
| HiAAMG_D_sequ1ence | STP-inf-A | Aminoglycosides |
| HiAAMG_E_sequ1ence | STP-inf-A | Aminoglycosides |
| HiAAMG_E_sequ2ence | STP-inf-A | Aminoglycosides |
| HiAAMG_E_sequ7ence | STP-inf-A | Aminoglycosides |
| HiAAMG_G_sequ1ence | STP-inf-A | Aminoglycosides |
| HiAAMG_H_sequ2ence | STP-inf-A | Aminoglycosides |
| HiAAMG_H_sequ3ence | STP-inf-A | Aminoglycosides |
| HiAAMG_H_sequ6ence | STP-inf-A | Aminoglycosides |
| HiAAMG_H_sequ7ence | STP-inf-A | Aminoglycosides |
| HiAAMG_I_sequ3ence | STP-inf-A | Aminoglycosides |
| HiACLASS_a_sequ12ence | STP-inf-A | Betalactams |
| HiACLASS_a_sequ14ence | STP-inf-A | Betalactams |
| HiACLASS_a_sequ19ence | STP-inf-A | Betalactams |
| HiACLASS_a_sequ1ence | STP-inf-A | Betalactams |
| HiACLASS_a_sequ20ence | STP-inf-A | Betalactams |
| HiACLASS_a_sequ21ence | STP-inf-A | Betalactams |
| HiACLASS_a_sequ32ence | STP-inf-A | Betalactams |
| HiACLASS_a_sequ37ence | STP-inf-A | Betalactams |
| HiACLASS_a_sequ4ence | STP-inf-A | Betalactams |
| HiACLASS_a_sequ5ence | STP-inf-A | Betalactams |
| HiACLASS_b3_sequ1ence | STP-inf-A | Betalactams |
| HiACLASS_c_sequ11ence | STP-inf-A | Betalactams |
| HiACLASS_c_sequ12ence | STP-inf-A | Betalactams |
| HiACLASS_c_sequ1ence | STP-inf-A | Betalactams |
| HiACLASS_d2_sequ14ence | STP-inf-A | Betalactams |
| HiACLASS_d2_sequ16ence | STP-inf-A | Betalactams |
| HiACLASS_d2_sequ18ence | STP-inf-A | Betalactams |
| HiACLASS_d2_sequ21ence | STP-inf-A | Betalactams |
| HiACLASS_d2_sequ22ence | STP-inf-A | Betalactams |
| HiACLASS_d2_sequ23ence | STP-inf-A | Betalactams |
| HiACLASS_d2_sequ25ence | STP-inf-A | Betalactams |
| HiACLASS_d2_sequ30ence | STP-inf-A | Betalactams |
| HiACLASS_d2_sequ7ence | STP-inf-A | Betalactams |
| HiAERM_ermtype_sequ1ence | STP-inf-A | Erythromycin |
| HiAERM_ermtype_sequ1ence | STP-inf-A | Erythromycin |
| HiAERM_ermtype_sequ2ence | STP-inf-A | Erythromycin |
| HiAERM_ermtype_sequ2ence | STP-inf-A | Erythromycin |
| HiAERM_ermtype_sequ3ence | STP-inf-A | Erythromycin |
| HiAERM_ermtype_sequ8ence | STP-inf-A | Erythromycin |
| HiAMPH_sequ1ence | STP-inf-A | Macrolides |

| | | |
|---|---|---|
| HiAMPH_sequ2ence | STP-inf-A | Macrolides |
| HiAMPH_sequ3ence | STP-inf-A | Macrolides |
| HiAMPH_sequ4ence | STP-inf-A | Macrolides |
| HiApraminoglycosidemodelcSeq1 | STP-inf-A | Aminoglycosides |
| HiAQNR_sequ3ence | STP-inf-A | Quinolones |
| HiATET_eff_sequ1ence | STP-inf-A | Tetracyclines |
| HiATET_eff_sequ2ence | STP-inf-A | Tetracyclines |
| HiATET_enz_sequ1ence | STP-inf-A | Tetracyclines |
| HiATET_enz_sequ2ence | STP-inf-A | Tetracyclines |
| HiATET_rpg_sequ2ence | STP-inf-A | Tetracyclines |
| HiATET_rpg_sequ6ence | STP-inf-A | Tetracyclines |
| HiFAMG_B_sequ1ence | STP-inf-F | Aminoglycosides |
| HiFAMG_B_sequ5ence | STP-inf-F | Aminoglycosides |
| HiFAMG_C_sequ1ence | STP-inf-F | Aminoglycosides |
| HiFAMG_D_sequ1ence | STP-inf-F | Aminoglycosides |
| HiFAMG_D_sequ2ence | STP-inf-F | Aminoglycosides |
| HiFAMG_E_sequ1ence | STP-inf-F | Aminoglycosides |
| HiFAMG_E_sequ2ence | STP-inf-F | Aminoglycosides |
| HiFAMG_E_sequ3ence | STP-inf-F | Aminoglycosides |
| HiFAMG_E_sequ7ence | STP-inf-F | Aminoglycosides |
| HiFAMG_G_sequ1ence | STP-inf-F | Aminoglycosides |
| HiFAMG_G_sequ2ence | STP-inf-F | Aminoglycosides |
| HiFAMG_H_sequ2ence | STP-inf-F | Aminoglycosides |
| HiFAMG_H_sequ3ence | STP-inf-F | Aminoglycosides |
| HiFAMG_H_sequ4ence | STP-inf-F | Aminoglycosides |
| HiFAMG_I_sequ2ence | STP-inf-F | Aminoglycosides |
| HiFCLASS_a_sequ13ence | STP-inf-F | Betalactams |
| HiFCLASS_a_sequ1ence | STP-inf-F | Betalactams |
| HiFCLASS_a_sequ23ence | STP-inf-F | Betalactams |
| HiFCLASS_a_sequ25ence | STP-inf-F | Betalactams |
| HiFCLASS_a_sequ26ence | STP-inf-F | Betalactams |
| HiFCLASS_a_sequ2ence | STP-inf-F | Betalactams |
| HiFCLASS_a_sequ33ence | STP-inf-F | Betalactams |
| HiFCLASS_a_sequ34ence | STP-inf-F | Betalactams |
| HiFCLASS_a_sequ35ence | STP-inf-F | Betalactams |
| HiFCLASS_a_sequ41ence | STP-inf-F | Betalactams |
| HiFCLASS_a_sequ51ence | STP-inf-F | Betalactams |
| HiFCLASS_a_sequ5ence | STP-inf-F | Betalactams |
| HiFCLASS_a_sequ7ence | STP-inf-F | Betalactams |
| HiFCLASS_b3_sequ1ence | STP-inf-F | Betalactams |
| HiFCLASS_b3_sequ2ence | STP-inf-F | Betalactams |
| HiFCLASS_b3_sequ3ence | STP-inf-F | Betalactams |
| HiFCLASS_c_sequ5ence | STP-inf-F | Betalactams |
| HiFCLASS_c_sequ8ence | STP-inf-F | Betalactams |
| HiFCLASS_d2_sequ11ence | STP-inf-F | Betalactams |
| HiFCLASS_d2_sequ13ence | STP-inf-F | Betalactams |

| | | |
|---|---|---|
| HiFCLASS_d2_sequ17ence | STP-inf-F | Betalactams |
| HiFCLASS_d2_sequ20ence | STP-inf-F | Betalactams |
| HiFCLASS_d2_sequ22ence | STP-inf-F | Betalactams |
| HiFCLASS_d2_sequ24ence | STP-inf-F | Betalactams |
| HiFCLASS_d2_sequ25ence | STP-inf-F | Betalactams |
| HiFCLASS_d2_sequ26ence | STP-inf-F | Betalactams |
| HiFCLASS_d2_sequ29ence | STP-inf-F | Betalactams |
| HiFCLASS_d2_sequ30ence | STP-inf-F | Betalactams |
| HiFCLASS_d2_sequ31ence | STP-inf-F | Betalactams |
| HiFERM_ermtype_sequ10ence | STP-inf-F | Erythromycin |
| HiFERM_ermtype_sequ12ence | STP-inf-F | Erythromycin |
| HiFERM_ermtype_sequ1ence | STP-inf-F | Erythromycin |
| HiFERM_ermtype_sequ1ence | STP-inf-F | Erythromycin |
| HiFERM_ermtype_sequ2ence | STP-inf-F | Erythromycin |
| HiFERM_ermtype_sequ3ence | STP-inf-F | Erythromycin |
| HiFERM_ermtype_sequ4ence | STP-inf-F | Erythromycin |
| HiFERM_ermtype_sequ5ence | STP-inf-F | Erythromycin |
| HiFERM_ermtype_sequ6ence | STP-inf-F | Erythromycin |
| HiFERM_ermtype_sequ8ence | STP-inf-F | Erythromycin |
| HiFMPH_sequ1ence | STP-inf-F | Macrolides |
| HiFMPH_sequ2ence | STP-inf-F | Macrolides |
| HiFMPH_sequ4ence | STP-inf-F | Macrolides |
| HiFMPH_sequ6ence | STP-inf-F | Macrolides |
| HiFQNR_sequ1ence | STP-inf-F | Quinolones |
| HiFQNR_sequ4ence | STP-inf-F | Quinolones |
| HiFTET_eff_sequ1ence | STP-inf-F | Tetracyclines |
| HiFTET_eff_sequ2ence | STP-inf-F | Tetracyclines |
| HiFTET_enz_sequ1ence | STP-inf-F | Tetracyclines |
| HiFTET_enz_sequ2ence | STP-inf-F | Tetracyclines |
| HiFTET_enz_sequ3ence | STP-inf-F | Tetracyclines |
| HiFTET_rpg_sequ2ence | STP-inf-F | Tetracyclines |
| HiFTET_rpg_sequ3ence | STP-inf-F | Tetracyclines |
| HiFTET_rpg_sequ4ence | STP-inf-F | Tetracyclines |
| HiFTET_rpg_sequ7ence | STP-inf-F | Tetracyclines |
| HiFTET_rpg_sequ8ence | STP-inf-F | Tetracyclines |
| HiFTET_rpg_sequ9ence | STP-inf-F | Tetracyclines |
| HoAAMG_B_sequ1ence | STP-T-eff-A | Aminoglycosides |
| HoAAMG_B_sequ2ence | STP-T-eff-A | Aminoglycosides |
| HoAAMG_C_sequ1ence | STP-T-eff-A | Aminoglycosides |
| HoAAMG_D_sequ1ence | STP-T-eff-A | Aminoglycosides |
| HoAAMG_D_sequ2ence | STP-T-eff-A | Aminoglycosides |
| HoAAMG_E_sequ1ence | STP-T-eff-A | Aminoglycosides |
| HoAAMG_E_sequ4ence | STP-T-eff-A | Aminoglycosides |
| HoAAMG_E_sequ5ence | STP-T-eff-A | Aminoglycosides |
| HoAAMG_E_sequ8ence | STP-T-eff-A | Aminoglycosides |
| HoAAMG_E_sequ9ence | STP-T-eff-A | Aminoglycosides |

| | | |
|---|---|---|
| HoAAMG_G_sequ1ence | STP-T-eff-A | Aminoglycosides |
| HoAAMG_G_sequ2ence | STP-T-eff-A | Aminoglycosides |
| HoAAMG_H_sequ1ence | STP-T-eff-A | Aminoglycosides |
| HoAAMG_H_sequ5ence | STP-T-eff-A | Aminoglycosides |
| HoAAMG_H_sequ7ence | STP-T-eff-A | Aminoglycosides |
| HoAAMG_H_sequ8ence | STP-T-eff-A | Aminoglycosides |
| HoAAMG_I_sequ4ence | STP-T-eff-A | Aminoglycosides |
| HoACLASS_a_sequ14ence | STP-T-eff-A | Betalactams |
| HoACLASS_a_sequ19ence | STP-T-eff-A | Betalactams |
| HoACLASS_a_sequ1ence | STP-T-eff-A | Betalactams |
| HoACLASS_a_sequ23ence | STP-T-eff-A | Betalactams |
| HoACLASS_a_sequ30ence | STP-T-eff-A | Betalactams |
| HoACLASS_a_sequ41ence | STP-T-eff-A | Betalactams |
| HoACLASS_a_sequ43ence | STP-T-eff-A | Betalactams |
| HoACLASS_a_sequ44ence | STP-T-eff-A | Betalactams |
| HoACLASS_a_sequ48ence | STP-T-eff-A | Betalactams |
| HoACLASS_a_sequ4ence | STP-T-eff-A | Betalactams |
| HoACLASS_a_sequ52ence | STP-T-eff-A | Betalactams |
| HoACLASS_a_sequ55ence | STP-T-eff-A | Betalactams |
| HoACLASS_a_sequ57ence | STP-T-eff-A | Betalactams |
| HoACLASS_a_sequ60ence | STP-T-eff-A | Betalactams |
| HoACLASS_a_sequ61ence | STP-T-eff-A | Betalactams |
| HoACLASS_a_sequ65ence | STP-T-eff-A | Betalactams |
| HoACLASS_a_sequ67ence | STP-T-eff-A | Betalactams |
| HoACLASS_a_sequ73ence | STP-T-eff-A | Betalactams |
| HoACLASS_a_sequ76ence | STP-T-eff-A | Betalactams |
| HoACLASS_a_sequ77ence | STP-T-eff-A | Betalactams |
| HoACLASS_b3_sequ3ence | STP-T-eff-A | Betalactams |
| HoACLASS_b3_sequ5ence | STP-T-eff-A | Betalactams |
| HoACLASS_c_sequ3ence | STP-T-eff-A | Betalactams |
| HoACLASS_c_sequ4ence | STP-T-eff-A | Betalactams |
| HoACLASS_d1_sequ4ence | STP-T-eff-A | Betalactams |
| HoACLASS_d2_sequ13ence | STP-T-eff-A | Betalactams |
| HoACLASS_d2_sequ24ence | STP-T-eff-A | Betalactams |
| HoACLASS_d2_sequ27ence | STP-T-eff-A | Betalactams |
| HoACLASS_d2_sequ32ence | STP-T-eff-A | Betalactams |
| HoACLASS_d2_sequ37ence | STP-T-eff-A | Betalactams |
| HoACLASS_d2_sequ41ence | STP-T-eff-A | Betalactams |
| HoACLASS_d2_sequ49ence | STP-T-eff-A | Betalactams |
| HoACLASS_d2_sequ51ence | STP-T-eff-A | Betalactams |
| HoACLASS_d2_sequ52ence | STP-T-eff-A | Betalactams |
| HoACLASS_d2_sequ7ence | STP-T-eff-A | Betalactams |
| HoACLASS_d2_sequ9ence | STP-T-eff-A | Betalactams |
| HoAERM_ermtype_sequ12ence | STP-T-eff-A | Erythromycin |
| HoAERM_ermtype_sequ13ence | STP-T-eff-A | Erythromycin |
| HoAERM_ermtype_sequ1ence | STP-T-eff-A | Erythromycin |

| | | |
|---|---|---|
| HoAERM_ermtype_sequ1ence | STP-T-eff-A | Erythromycin |
| HoAERM_ermtype_sequ2ence | STP-T-eff-A | Erythromycin |
| HoAERM_ermtype_sequ3ence | STP-T-eff-A | Erythromycin |
| HoAERM_ermtype_sequ5ence | STP-T-eff-A | Erythromycin |
| HoAERM_ermtype_sequ5ence | STP-T-eff-A | Erythromycin |
| HoAERM_ermtype_sequ7ence | STP-T-eff-A | Erythromycin |
| HoAERM_ermtype_sequ8ence | STP-T-eff-A | Erythromycin |
| HoAERM_ermtype_sequ9ence | STP-T-eff-A | Erythromycin |
| HoAMPH_sequ1ence | STP-T-eff-A | Macrolides |
| HoAMPH_sequ2ence | STP-T-eff-A | Macrolides |
| HoAMPH_sequ5ence | STP-T-eff-A | Macrolides |
| HoAQNR_sequ2ence | STP-T-eff-A | Quinolones |
| HoATET_eff_sequ1ence | STP-T-eff-A | Tetracyclines |
| HoATET_eff_sequ2ence | STP-T-eff-A | Tetracyclines |
| HoATET_eff_sequ4ence | STP-T-eff-A | Tetracyclines |
| HoATET_enz_sequ1ence | STP-T-eff-A | Tetracyclines |
| HoATET_enz_sequ2ence | STP-T-eff-A | Tetracyclines |
| HoATET_rpg_sequ11ence | STP-T-eff-A | Tetracyclines |
| HoATET_rpg_sequ3ence | STP-T-eff-A | Tetracyclines |
| HoATET_rpg_sequ7ence | STP-T-eff-A | Tetracyclines |
| HoATET_rpg_sequ8ence | STP-T-eff-A | Tetracyclines |
| HoFAMG_B_sequ1ence | STP-T-eff-F | Aminoglycosides |
| HoFAMG_B_sequ2ence | STP-T-eff-F | Aminoglycosides |
| HoFAMG_C_sequ1ence | STP-T-eff-F | Aminoglycosides |
| HoFAMG_D_sequ4ence | STP-T-eff-F | Aminoglycosides |
| HoFAMG_E_sequ1ence | STP-T-eff-F | Aminoglycosides |
| HoFAMG_E_sequ2ence | STP-T-eff-F | Aminoglycosides |
| HoFAMG_E_sequ4ence | STP-T-eff-F | Aminoglycosides |
| HoFAMG_E_sequ5ence | STP-T-eff-F | Aminoglycosides |
| HoFAMG_G_sequ1ence | STP-T-eff-F | Aminoglycosides |
| HoFAMG_H_sequ1ence | STP-T-eff-F | Aminoglycosides |
| HoFAMG_H_sequ2ence | STP-T-eff-F | Aminoglycosides |
| HoFAMG_I_sequ5ence | STP-T-eff-F | Aminoglycosides |
| HoFCLASS_a_sequ12ence | STP-T-eff-F | Betalactams |
| HoFCLASS_a_sequ19ence | STP-T-eff-F | Betalactams |
| HoFCLASS_a_sequ25ence | STP-T-eff-F | Betalactams |
| HoFCLASS_a_sequ2ence | STP-T-eff-F | Betalactams |
| HoFCLASS_a_sequ46ence | STP-T-eff-F | Betalactams |
| HoFCLASS_a_sequ47ence | STP-T-eff-F | Betalactams |
| HoFCLASS_a_sequ49ence | STP-T-eff-F | Betalactams |
| HoFCLASS_a_sequ57ence | STP-T-eff-F | Betalactams |
| HoFCLASS_a_sequ5ence | STP-T-eff-F | Betalactams |
| HoFCLASS_a_sequ60ence | STP-T-eff-F | Betalactams |
| HoFCLASS_a_sequ7ence | STP-T-eff-F | Betalactams |
| HoFCLASS_b3_sequ2ence | STP-T-eff-F | Betalactams |
| HoFCLASS_c_sequ5ence | STP-T-eff-F | Betalactams |

| | | |
|---|---|---|
| HoFCLASS_d1_sequ2ence | STP-T-eff-F | Betalactams |
| HoFCLASS_d2_sequ13ence | STP-T-eff-F | Betalactams |
| HoFCLASS_d2_sequ14ence | STP-T-eff-F | Betalactams |
| HoFCLASS_d2_sequ17ence | STP-T-eff-F | Betalactams |
| HoFCLASS_d2_sequ19ence | STP-T-eff-F | Betalactams |
| HoFCLASS_d2_sequ21ence | STP-T-eff-F | Betalactams |
| HoFCLASS_d2_sequ29ence | STP-T-eff-F | Betalactams |
| HoFCLASS_d2_sequ34ence | STP-T-eff-F | Betalactams |
| HoFCLASS_d2_sequ4ence | STP-T-eff-F | Betalactams |
| HoFCLASS_d2_sequ8ence | STP-T-eff-F | Betalactams |
| HoFERM_ermtype_sequ1ence | STP-T-eff-F | Erythromycin |
| HoFERM_ermtype_sequ3ence | STP-T-eff-F | Erythromycin |
| HoFERM_ermtype_sequ4ence | STP-T-eff-F | Erythromycin |
| HoFERM_ermtype_sequ5ence | STP-T-eff-F | Erythromycin |
| HoFERM_ermtype_sequ7ence | STP-T-eff-F | Erythromycin |
| HoFERM_ermtype_sequ8ence | STP-T-eff-F | Erythromycin |
| HoFMPH_sequ1ence | STP-T-eff-F | Macrolides |
| HoFMPH_sequ2ence | STP-T-eff-F | Macrolides |
| HoFMPH_sequ4ence | STP-T-eff-F | Macrolides |
| HoFTET_eff_sequ2ence | STP-T-eff-F | Tetracyclines |
| HoFTET_eff_sequ3ence | STP-T-eff-F | Tetracyclines |
| HoFTET_enz_sequ1ence | STP-T-eff-F | Tetracyclines |
| HoFTET_enz_sequ2ence | STP-T-eff-F | Tetracyclines |
| HoFTET_rpg_sequ3ence | STP-T-eff-F | Tetracyclines |
| HoFTET_rpg_sequ5ence | STP-T-eff-F | Tetracyclines |
| SeAAMG_B_sequ2ence | Hospital-eff-A | Aminoglycosides |
| SeAAMG_B_sequ3ence | Hospital-eff-A | Aminoglycosides |
| SeAAMG_C_sequ1ence | Hospital-eff-A | Aminoglycosides |
| SeAAMG_D_sequ4ence | Hospital-eff-A | Aminoglycosides |
| SeAAMG_E_sequ11ence | Hospital-eff-A | Aminoglycosides |
| SeAAMG_E_sequ2ence | Hospital-eff-A | Aminoglycosides |
| SeAAMG_E_sequ4ence | Hospital-eff-A | Aminoglycosides |
| SeAAMG_E_sequ5ence | Hospital-eff-A | Aminoglycosides |
| SeAAMG_E_sequ6ence | Hospital-eff-A | Aminoglycosides |
| SeAAMG_F_sequ1ence | Hospital-eff-A | Aminoglycosides |
| SeAAMG_G_sequ1ence | Hospital-eff-A | Aminoglycosides |
| SeAAMG_H_sequ1ence | Hospital-eff-A | Aminoglycosides |
| SeAAMG_H_sequ4ence | Hospital-eff-A | Aminoglycosides |
| SeAAMG_H_sequ5ence | Hospital-eff-A | Aminoglycosides |
| SeAAMG_H_sequ7ence | Hospital-eff-A | Aminoglycosides |
| SeAAMG_H_sequ8ence | Hospital-eff-A | Aminoglycosides |
| SeACLASS_a_sequ11ence | Hospital-eff-A | Betalactams |
| SeACLASS_a_sequ13ence | Hospital-eff-A | Betalactams |
| SeACLASS_a_sequ14ence | Hospital-eff-A | Betalactams |
| SeACLASS_a_sequ16ence | Hospital-eff-A | Betalactams |
| SeACLASS_a_sequ17ence | Hospital-eff-A | Betalactams |

| | | |
|---|---|---|
| SeACLASS_a_sequ1ence | Hospital-eff-A | Betalactams |
| SeACLASS_a_sequ20ence | Hospital-eff-A | Betalactams |
| SeACLASS_a_sequ23ence | Hospital-eff-A | Betalactams |
| SeACLASS_a_sequ24ence | Hospital-eff-A | Betalactams |
| SeACLASS_a_sequ29ence | Hospital-eff-A | Betalactams |
| SeACLASS_a_sequ3ence | Hospital-eff-A | Betalactams |
| SeACLASS_a_sequ42ence | Hospital-eff-A | Betalactams |
| SeACLASS_a_sequ5ence | Hospital-eff-A | Betalactams |
| SeACLASS_a_sequ8ence | Hospital-eff-A | Betalactams |
| SeACLASS_b12_sequ2ence | Hospital-eff-A | Betalactams |
| SeACLASS_b12_sequ5ence | Hospital-eff-A | Betalactams |
| SeACLASS_b3_sequ2ence | Hospital-eff-A | Betalactams |
| SeACLASS_c_sequ11ence | Hospital-eff-A | Betalactams |
| SeACLASS_c_sequ12ence | Hospital-eff-A | Betalactams |
| SeACLASS_c_sequ7ence | Hospital-eff-A | Betalactams |
| SeACLASS_d1_sequ2ence | Hospital-eff-A | Betalactams |
| SeACLASS_d1_sequ3ence | Hospital-eff-A | Betalactams |
| SeACLASS_d1_sequ4ence | Hospital-eff-A | Betalactams |
| SeACLASS_d2_sequ13ence | Hospital-eff-A | Betalactams |
| SeACLASS_d2_sequ17ence | Hospital-eff-A | Betalactams |
| SeACLASS_d2_sequ18ence | Hospital-eff-A | Betalactams |
| SeACLASS_d2_sequ1ence | Hospital-eff-A | Betalactams |
| SeACLASS_d2_sequ2ence | Hospital-eff-A | Betalactams |
| SeACLASS_d2_sequ5ence | Hospital-eff-A | Betalactams |
| SeAERM_ermtype_sequ1ence | Hospital-eff-A | Erythromycin |
| SeAERM_ermtype_sequ1ence | Hospital-eff-A | Erythromycin |
| SeAERM_ermtype_sequ3ence | Hospital-eff-A | Erythromycin |
| SeAERM_ermtype_sequ4ence | Hospital-eff-A | Erythromycin |
| SeAERM_ermtype_sequ7ence | Hospital-eff-A | Erythromycin |
| SeAERM_ermtype_sequ8ence | Hospital-eff-A | Erythromycin |
| SeAMPH_sequ1ence | Hospital-eff-A | Macrolides |
| SeAMPH_sequ2ence | Hospital-eff-A | Macrolides |
| SeAMPH_sequ3ence | Hospital-eff-A | Macrolides |
| SeAMPH_sequ4ence | Hospital-eff-A | Macrolides |
| SeAMPH_sequ5ence | Hospital-eff-A | Macrolides |
| SeAQNR_sequ2ence | Hospital-eff-A | Quinolones |
| SeATET_eff_sequ4ence | Hospital-eff-A | Tetracyclines |
| SeATET_eff_sequ5ence | Hospital-eff-A | Tetracyclines |
| SeATET_eff_sequ6ence | Hospital-eff-A | Tetracyclines |
| SeATET_enz_sequ1ence | Hospital-eff-A | Tetracyclines |
| SeATET_rpg_sequ4ence | Hospital-eff-A | Tetracyclines |
| SeFAMG_B_sequ1ence | Hospital-eff-F | Aminoglycosides |
| SeFAMG_B_sequ5ence | Hospital-eff-F | Aminoglycosides |
| SeFAMG_C_sequ2ence | Hospital-eff-F | Aminoglycosides |
| SeFAMG_D_sequ1ence | Hospital-eff-F | Aminoglycosides |
| SeFAMG_D_sequ5ence | Hospital-eff-F | Aminoglycosides |

| | | |
|---|---|---|
| SeFAMG_E_sequ11ence | Hospital-eff-F | Aminoglycosides |
| SeFAMG_E_sequ12ence | Hospital-eff-F | Aminoglycosides |
| SeFAMG_E_sequ13ence | Hospital-eff-F | Aminoglycosides |
| SeFAMG_E_sequ14ence | Hospital-eff-F | Aminoglycosides |
| SeFAMG_E_sequ1ence | Hospital-eff-F | Aminoglycosides |
| SeFAMG_E_sequ2ence | Hospital-eff-F | Aminoglycosides |
| SeFAMG_F_sequ1ence | Hospital-eff-F | Aminoglycosides |
| SeFAMG_G_sequ1ence | Hospital-eff-F | Aminoglycosides |
| SeFAMG_H_sequ12ence | Hospital-eff-F | Aminoglycosides |
| SeFAMG_H_sequ3ence | Hospital-eff-F | Aminoglycosides |
| SeFAMG_H_sequ6ence | Hospital-eff-F | Aminoglycosides |
| SeFAMG_H_sequ7ence | Hospital-eff-F | Aminoglycosides |
| SeFAMG_H_sequ8ence | Hospital-eff-F | Aminoglycosides |
| SeFCLASS_a_sequ11ence | Hospital-eff-F | Betalactams |
| SeFCLASS_a_sequ12ence | Hospital-eff-F | Betalactams |
| SeFCLASS_a_sequ15ence | Hospital-eff-F | Betalactams |
| SeFCLASS_a_sequ16ence | Hospital-eff-F | Betalactams |
| SeFCLASS_a_sequ20ence | Hospital-eff-F | Betalactams |
| SeFCLASS_a_sequ22ence | Hospital-eff-F | Betalactams |
| SeFCLASS_a_sequ28ence | Hospital-eff-F | Betalactams |
| SeFCLASS_a_sequ36ence | Hospital-eff-F | Betalactams |
| SeFCLASS_a_sequ37ence | Hospital-eff-F | Betalactams |
| SeFCLASS_a_sequ38ence | Hospital-eff-F | Betalactams |
| SeFCLASS_a_sequ3ence | Hospital-eff-F | Betalactams |
| SeFCLASS_a_sequ43ence | Hospital-eff-F | Betalactams |
| SeFCLASS_a_sequ52ence | Hospital-eff-F | Betalactams |
| SeFCLASS_a_sequ53ence | Hospital-eff-F | Betalactams |
| SeFCLASS_a_sequ54ence | Hospital-eff-F | Betalactams |
| SeFCLASS_a_sequ56ence | Hospital-eff-F | Betalactams |
| SeFCLASS_a_sequ61ence | Hospital-eff-F | Betalactams |
| SeFCLASS_a_sequ62ence | Hospital-eff-F | Betalactams |
| SeFCLASS_a_sequ64ence | Hospital-eff-F | Betalactams |
| SeFCLASS_a_sequ65ence | Hospital-eff-F | Betalactams |
| SeFCLASS_a_sequ67ence | Hospital-eff-F | Betalactams |
| SeFCLASS_a_sequ8ence | Hospital-eff-F | Betalactams |
| SeFCLASS_b12_sequ1ence | Hospital-eff-F | Betalactams |
| SeFCLASS_b12_sequ2ence | Hospital-eff-F | Betalactams |
| SeFCLASS_b3_sequ3ence | Hospital-eff-F | Betalactams |
| SeFCLASS_b3_sequ5ence | Hospital-eff-F | Betalactams |
| SeFCLASS_b3_sequ7ence | Hospital-eff-F | Betalactams |
| SeFCLASS_c_sequ13ence | Hospital-eff-F | Betalactams |
| SeFCLASS_c_sequ4ence | Hospital-eff-F | Betalactams |
| SeFCLASS_d1_sequ1ence | Hospital-eff-F | Betalactams |
| SeFCLASS_d1_sequ2ence | Hospital-eff-F | Betalactams |
| SeFCLASS_d1_sequ5ence | Hospital-eff-F | Betalactams |
| SeFCLASS_d1_sequ6ence | Hospital-eff-F | Betalactams |

| | | |
|---|---|---|
| SeFCLASS_d2_sequ15ence | Hospital-eff-F | Betalactams |
| SeFCLASS_d2_sequ21ence | Hospital-eff-F | Betalactams |
| SeFCLASS_d2_sequ22ence | Hospital-eff-F | Betalactams |
| SeFCLASS_d2_sequ26ence | Hospital-eff-F | Betalactams |
| SeFCLASS_d2_sequ27ence | Hospital-eff-F | Betalactams |
| SeFCLASS_d2_sequ28ence | Hospital-eff-F | Betalactams |
| SeFCLASS_d2_sequ29ence | Hospital-eff-F | Betalactams |
| SeFCLASS_d2_sequ2ence | Hospital-eff-F | Betalactams |
| SeFCLASS_d2_sequ4ence | Hospital-eff-F | Betalactams |
| SeFCLASS_d2_sequ5ence | Hospital-eff-F | Betalactams |
| SeFERM_ermtype_sequ12ence | Hospital-eff-F | Erythromycin |
| SeFERM_ermtype_sequ13ence | Hospital-eff-F | Erythromycin |
| SeFERM_ermtype_sequ1ence | Hospital-eff-F | Erythromycin |
| SeFERM_ermtype_sequ1ence | Hospital-eff-F | Erythromycin |
| SeFERM_ermtype_sequ2ence | Hospital-eff-F | Erythromycin |
| SeFERM_ermtype_sequ3ence | Hospital-eff-F | Erythromycin |
| SeFERM_ermtype_sequ5ence | Hospital-eff-F | Erythromycin |
| SeFERM_ermtype_sequ7ence | Hospital-eff-F | Erythromycin |
| SeFERM_ermtype_sequ8ence | Hospital-eff-F | Erythromycin |
| SeFERM_ermtype_sequ9ence | Hospital-eff-F | Erythromycin |
| SeFMPH_sequ2ence | Hospital-eff-F | Macrolides |
| SeFMPH_sequ3ence | Hospital-eff-F | Macrolides |
| SeFMPH_sequ4ence | Hospital-eff-F | Macrolides |
| SeFQNR_sequ1ence | Hospital-eff-F | Quinolones |
| SeFQNR_sequ2ence | Hospital-eff-F | Quinolones |
| SeFQNR_sequ6ence | Hospital-eff-F | Quinolones |
| SeFTET_eff_sequ6ence | Hospital-eff-F | Tetracyclines |
| SeFTET_eff_sequ7ence | Hospital-eff-F | Tetracyclines |
| SeFTET_eff_sequ8ence | Hospital-eff-F | Tetracyclines |
| SeFTET_enz_sequ1ence | Hospital-eff-F | Tetracyclines |
| SeFTET_enz_sequ2ence | Hospital-eff-F | Tetracyclines |
| SeFTET_rpg_sequ1ence | Hospital-eff-F | Tetracyclines |
| SeFTET_rpg_sequ6ence | Hospital-eff-F | Tetracyclines |
| HiAAMG_G_sequ2ence | STP-inf-A | Aminoglycosides |
| SeFAMG_E_sequ6ence | Hospital-eff-F | Aminoglycosides |
| HiACLASS_a_sequ17ence | STP-inf-A | Betalactams |
| HiACLASS_a_sequ24ence | STP-inf-A | Betalactams |
| HiACLASS_a_sequ25ence | STP-inf-A | Betalactams |
| HiACLASS_a_sequ26ence | STP-inf-A | Betalactams |
| HiACLASS_a_sequ27ence | STP-inf-A | Betalactams |
| HiACLASS_a_sequ28ence | STP-inf-A | Betalactams |
| HiACLASS_a_sequ30ence | STP-inf-A | Betalactams |
| HoAAMG_E_sequ6ence | STP-T-eff-A | Aminoglycosides |
| HiACLASS_a_sequ31ence | STP-inf-A | Betalactams |
| HiACLASS_a_sequ35ence | STP-inf-A | Betalactams |
| HiACLASS_a_sequ41ence | STP-inf-A | Betalactams |

| | | |
|---|---|---|
| HiACLASS_a_sequ8ence | STP-inf-A | Betalactams |
| HiACLASS_b12_sequ3ence | STP-inf-A | Betalactams |
| HiACLASS_b12_sequ5ence | STP-inf-A | Betalactams |
| HiACLASS_c_sequ3ence | STP-inf-A | Betalactams |
| HiACLASS_c_sequ5ence | STP-inf-A | Betalactams |
| HiACLASS_d1_sequ2ence | STP-inf-A | Betalactams |
| HiACLASS_d2_sequ10ence | STP-inf-A | Betalactams |
| HiACLASS_d2_sequ13ence | STP-inf-A | Betalactams |
| HoFAMG_I_sequ2ence | STP-T-eff-F | Aminoglycosides |
| HiACLASS_d2_sequ15ence | STP-inf-A | Betalactams |
| HiACLASS_d2_sequ26ence | STP-inf-A | Betalactams |
| HiACLASS_d2_sequ27ence | STP-inf-A | Betalactams |
| SeAAMG_E_sequ3ence | Hospital-eff-A | Aminoglycosides |
| SeFAMG_E_sequ17ence | Hospital-eff-F | Aminoglycosides |
| HoAMPH_sequ6ence | STP-T-eff-A | Aminoglycosides |
| HiACLASS_d2_sequ28ence | STP-inf-A | Betalactams |
| HiACLASS_d2_sequ31ence | STP-inf-A | Betalactams |
| HiACLASS_d2_sequ3ence | STP-inf-A | Betalactams |
| SeFAMG_E_sequ16ence | Hospital-eff-F | Aminoglycosides |
| HiAAMG_H_sequ4ence | STP-inf-A | Aminoglycosides |
| HoAAMG_H_sequ3ence | STP-T-eff-A | Aminoglycosides |
| HiFAMG_I_sequ4ence | STP-inf-F | Aminoglycosides |
| HiACLASS_d2_sequ4ence | STP-inf-A | Betalactams |
| HiACLASS_d2_sequ5ence | STP-inf-A | Betalactams |
| HoAAMG_B_sequ6ence | STP-T-eff-A | Aminoglycosides |
| HiACLASS_d2_sequ6ence | STP-inf-A | Betalactams |
| SeFAMG_H_sequ9ence | Hospital-eff-F | Aminoglycosides |
| HiACLASS_d2_sequ9ence | STP-inf-A | Betalactams |
| HiFCLASS_a_sequ11ence | STP-inf-F | Betalactams |
| HiFCLASS_a_sequ14ence | STP-inf-F | Betalactams |
| HoAAMG_I_sequ5ence | STP-T-eff-A | Aminoglycosides |
| HiFCLASS_a_sequ18ence | STP-inf-F | Betalactams |
| HiFCLASS_a_sequ29ence | STP-inf-F | Betalactams |
| HiFCLASS_a_sequ30ence | STP-inf-F | Betalactams |
| HiFCLASS_a_sequ36ence | STP-inf-F | Betalactams |
| HiFCLASS_a_sequ38ence | STP-inf-F | Betalactams |
| HiFCLASS_a_sequ40ence | STP-inf-F | Betalactams |
| HiFCLASS_a_sequ42ence | STP-inf-F | Betalactams |
| HiFAMG_E_sequ8ence | STP-inf-F | Aminoglycosides |
| HoAAMG_I_sequ8ence | STP-T-eff-A | Aminoglycosides |
| HoFAMG_I_sequ7ence | STP-T-eff-F | Aminoglycosides |
| HiAAMG_I_sequ7ence | STP-inf-A | Aminoglycosides |
| HiFCLASS_a_sequ48ence | STP-inf-F | Betalactams |
| SeFAMG_I_sequ4ence | Hospital-eff-F | Aminoglycosides |
| HiFCLASS_a_sequ49ence | STP-inf-F | Betalactams |
| HiFCLASS_a_sequ56ence | STP-inf-F | Betalactams |

| | | |
|---|---|---|
| HiFCLASS_a_sequ9ence | STP-inf-F | Betalactams |
| HiFCLASS_b12_sequ1ence | STP-inf-F | Betalactams |
| HiFCLASS_b12_sequ4ence | STP-inf-F | Betalactams |
| HiFCLASS_b12_sequ5ence | STP-inf-F | Betalactams |
| HiFCLASS_b12_sequ6ence | STP-inf-F | Betalactams |
| HiFCLASS_c_sequ10ence | STP-inf-F | Betalactams |
| HiFCLASS_c_sequ11ence | STP-inf-F | Betalactams |
| HiAAMG_H_sequ9ence | STP-inf-A | Aminoglycosides |
| HiFCLASS_c_sequ1ence | STP-inf-F | Betalactams |
| HiFCLASS_d2_sequ14ence | STP-inf-F | Betalactams |
| HiFCLASS_d2_sequ15ence | STP-inf-F | Betalactams |
| HiFCLASS_d2_sequ16ence | STP-inf-F | Betalactams |
| HoAAMG_H_sequ6ence | STP-T-eff-A | Aminoglycosides |
| HoAAMG_H_sequ2ence | STP-T-eff-A | Aminoglycosides |
| HiFCLASS_d2_sequ21ence | STP-inf-F | Betalactams |
| HiFCLASS_d2_sequ23ence | STP-inf-F | Betalactams |
| HiFCLASS_d2_sequ27ence | STP-inf-F | Betalactams |
| SeFAMG_H_sequ13ence | Hospital-eff-F | Aminoglycosides |
| HiFCLASS_d2_sequ28ence | STP-inf-F | Betalactams |
| HiFCLASS_d2_sequ2ence | STP-inf-F | Betalactams |
| HiFCLASS_d2_sequ32ence | STP-inf-F | Betalactams |
| HiFCLASS_d2_sequ4ence | STP-inf-F | Betalactams |
| HoAAMG_I_sequ6ence | STP-T-eff-A | Aminoglycosides |
| HiFCLASS_d2_sequ5ence | STP-inf-F | Betalactams |
| HiFCLASS_d2_sequ9ence | STP-inf-F | Betalactams |
| HiFCLASS_d2_sequ9ence | STP-inf-F | Betalactams |
| HoACLASS_a_sequ10ence | STP-T-eff-A | Betalactams |
| HoACLASS_a_sequ11ence | STP-T-eff-A | Betalactams |
| HoACLASS_a_sequ13ence | STP-T-eff-A | Betalactams |
| HiAAMG_H_sequ1ence | STP-inf-A | Aminoglycosides |
| HiFAMG_A_sequ1ence | STP-inf-F | Aminoglycosides |
| HiFAMG_A_sequ3ence | STP-inf-F | Aminoglycosides |
| HoACLASS_a_sequ16ence | STP-T-eff-A | Betalactams |
| HoACLASS_a_sequ17ence | STP-T-eff-A | Betalactams |
| SeAAMG_H_sequ9ence | Hospital-eff-A | Aminoglycosides |
| SeAAMG_E_sequ8ence | Hospital-eff-A | Aminoglycosides |
| SeAAMG_I_sequ4ence | Hospital-eff-A | Aminoglycosides |
| HoACLASS_a_sequ20ence | STP-T-eff-A | Betalactams |
| SeFAMG_I_sequ5ence | Hospital-eff-F | Aminoglycosides |
| HoACLASS_a_sequ21ence | STP-T-eff-A | Betalactams |
| SeAERM_ermtype_sequ2ence | Hospital-eff-A | Erythromycin |
| SeFERM_ermtype_sequ2ence | Hospital-eff-F | Erythromycin |
| SeAAMG_D_sequ1ence | Hospital-eff-A | Aminoglycosides |
| SeFAMG_D_sequ2ence | Hospital-eff-F | Aminoglycosides |
| HiAAMG_I_sequ4ence | STP-inf-A | Aminoglycosides |
| HoACLASS_a_sequ22ence | STP-T-eff-A | Betalactams |

| | | |
|---|---|---|
| SeFERM_ermtype_sequ3ence | Hospital-eff-F | Erythromycin |
| HoACLASS_a_sequ24ence | STP-T-eff-A | Betalactams |
| HoACLASS_a_sequ33ence | STP-T-eff-A | Betalactams |
| HoACLASS_a_sequ35ence | STP-T-eff-A | Betalactams |
| HoACLASS_a_sequ36ence | STP-T-eff-A | Betalactams |
| HoACLASS_a_sequ37ence | STP-T-eff-A | Betalactams |
| HoACLASS_a_sequ39ence | STP-T-eff-A | Betalactams |
| HiAAMG_E_sequ6ence | STP-inf-A | Aminoglycosides |
| HiApraminoglycosidemodelcSeq3 | STP-inf-A | Aminoglycosides |
| HoACLASS_a_sequ45ence | STP-T-eff-A | Betalactams |
| SeFAMG_I_sequ6ence | Hospital-eff-F | Aminoglycosides |
| HoACLASS_a_sequ47ence | STP-T-eff-A | Betalactams |
| HoACLASS_a_sequ49ence | STP-T-eff-A | Betalactams |
| HiFERM_ermtype_sequ11ence | STP-inf-F | Erythromycin |
| HiAAMG_I_sequ2ence | STP-inf-A | Aminoglycosides |
| HiFAMG_I_sequ1ence | STP-inf-F | Aminoglycosides |
| SeAAMG_E_sequ1ence | Hospital-eff-A | Aminoglycosides |
| SeFAMG_E_sequ3ence | Hospital-eff-F | Aminoglycosides |
| HoACLASS_a_sequ53ence | STP-T-eff-A | Betalactams |
| HoACLASS_a_sequ54ence | STP-T-eff-A | Betalactams |
| HoACLASS_a_sequ56ence | STP-T-eff-A | Betalactams |
| HiFAMG_C_sequ2ence | STP-inf-F | Aminoglycosides |
| HiFAMG_H_sequ6ence | STP-inf-F | Aminoglycosides |
| HoACLASS_a_sequ62ence | STP-T-eff-A | Betalactams |
| HoACLASS_a_sequ68ence | STP-T-eff-A | Betalactams |
| HoACLASS_a_sequ69ence | STP-T-eff-A | Betalactams |
| SeFAMG_H_sequ10ence | Hospital-eff-F | Aminoglycosides |
| HoACLASS_a_sequ6ence | STP-T-eff-A | Betalactams |
| SeAAMG_H_sequ6ence | Hospital-eff-A | Aminoglycosides |
| HoACLASS_a_sequ71ence | STP-T-eff-A | Betalactams |
| HoACLASS_a_sequ72ence | STP-T-eff-A | Betalactams |
| HoACLASS_a_sequ74ence | STP-T-eff-A | Betalactams |
| HoACLASS_a_sequ79ence | STP-T-eff-A | Betalactams |
| HoACLASS_a_sequ7ence | STP-T-eff-A | Betalactams |
| HoACLASS_a_sequ80ence | STP-T-eff-A | Betalactams |
| HiFAMG_H_sequ5ence | STP-inf-F | Aminoglycosides |
| HoACLASS_a_sequ81ence | STP-T-eff-A | Betalactams |
| HoACLASS_a_sequ8ence | STP-T-eff-A | Betalactams |
| SeFAMG_I_sequ2ence | Hospital-eff-F | Aminoglycosides |
| HoACLASS_b12_sequ10ence | STP-T-eff-A | Betalactams |
| HiAAMG_E_sequ5ence | STP-inf-A | Aminoglycosides |
| SeAAMG_C_sequ4ence | Hospital-eff-A | Aminoglycosides |
| SeFAMG_H_sequ1ence | Hospital-eff-F | Aminoglycosides |
| HiFAMG_A_sequ2ence | STP-inf-F | Aminoglycosides |
| SeAAMG_C_sequ3ence | Hospital-eff-A | Aminoglycosides |
| SeFAMG_C_sequ1ence | Hospital-eff-F | Aminoglycosides |

| | | |
|---|---|---|
| HoACLASS_b12_sequ14ence | STP-T-eff-A | Betalactams |
| HoACLASS_b12_sequ15ence | STP-T-eff-A | Betalactams |
| HoACLASS_b12_sequ16ence | STP-T-eff-A | Betalactams |
| HiApraminoglycosidemodelcSeq2 | STP-inf-A | Aminoglycosides |
| HiFAMG_E_sequ4ence | STP-inf-F | Aminoglycosides |
| HoACLASS_b12_sequ2ence | STP-T-eff-A | Betalactams |
| HoACLASS_b12_sequ3ence | STP-T-eff-A | Betalactams |
| HoACLASS_b12_sequ4ence | STP-T-eff-A | Betalactams |
| HoACLASS_b12_sequ5ence | STP-T-eff-A | Betalactams |
| HoACLASS_b12_sequ6ence | STP-T-eff-A | Betalactams |
| HoACLASS_b12_sequ8ence | STP-T-eff-A | Betalactams |
| HoACLASS_b12_sequ9ence | STP-T-eff-A | Betalactams |
| HoACLASS_b3_sequ1ence | STP-T-eff-A | Betalactams |
| HoACLASS_b3_sequ7ence | STP-T-eff-A | Betalactams |
| HoACLASS_c_sequ1ence | STP-T-eff-A | Betalactams |
| SeAAMG_D_sequ3ence | Hospital-eff-A | Aminoglycosides |
| HoACLASS_c_sequ2ence | STP-T-eff-A | Betalactams |
| HoACLASS_c_sequ7ence | STP-T-eff-A | Betalactams |
| HiAAMG_I_sequ5ence | STP-inf-A | Aminoglycosides |
| HiAAMG_A_sequ1ence | STP-inf-A | Aminoglycosides |
| HoACLASS_c_sequ8ence | STP-T-eff-A | Betalactams |
| HoACLASS_d1_sequ10ence | STP-T-eff-A | Betalactams |
| HoACLASS_d1_sequ1ence | STP-T-eff-A | Betalactams |
| HoACLASS_d1_sequ2ence | STP-T-eff-A | Betalactams |
| HoACLASS_d1_sequ3ence | STP-T-eff-A | Betalactams |
| SeATET_eff_sequ7ence | Hospital-eff-A | Tetracyclines |
| HoACLASS_d1_sequ5ence | STP-T-eff-A | Betalactams |
| HoACLASS_d1_sequ6ence | STP-T-eff-A | Betalactams |
| SeAQNR_sequ1ence | Hospital-eff-A | Quinolones |
| SeFQNR_sequ5ence | Hospital-eff-F | Quinolones |
| HoACLASS_d1_sequ8ence | STP-T-eff-A | Betalactams |
| HoACLASS_d1_sequ9ence | STP-T-eff-A | Betalactams |
| HoACLASS_d2_sequ10ence | STP-T-eff-A | Betalactams |
| HoACLASS_d2_sequ11ence | STP-T-eff-A | Betalactams |
| HoACLASS_d2_sequ16ence | STP-T-eff-A | Betalactams |
| HoACLASS_d2_sequ19ence | STP-T-eff-A | Betalactams |
| HoACLASS_d2_sequ20ence | STP-T-eff-A | Betalactams |
| HoACLASS_d2_sequ21ence | STP-T-eff-A | Betalactams |
| HoACLASS_d2_sequ22ence | STP-T-eff-A | Betalactams |
| HoACLASS_d2_sequ23ence | STP-T-eff-A | Betalactams |
| HoACLASS_d2_sequ25ence | STP-T-eff-A | Betalactams |
| HoACLASS_d2_sequ26ence | STP-T-eff-A | Betalactams |
| HoACLASS_d2_sequ28ence | STP-T-eff-A | Betalactams |
| HoACLASS_d2_sequ29ence | STP-T-eff-A | Betalactams |
| HiATET_eff_sequ4ence | STP-inf-A | Tetracyclines |
| HoACLASS_d2_sequ31ence | STP-T-eff-A | Betalactams |

| | | |
|---|---|---|
| HoACLASS_d2_sequ38ence | STP-T-eff-A | Betalactams |
| SeAAMG_E_sequ10ence | Hospital-eff-A | Aminoglycosides |
| HoACLASS_d2_sequ39ence | STP-T-eff-A | Betalactams |
| SeFERM_ermtype_sequ6ence | Hospital-eff-F | Erythromycin |
| HoACLASS_d2_sequ3ence | STP-T-eff-A | Betalactams |
| HoACLASS_d2_sequ42ence | STP-T-eff-A | Betalactams |
| SeFAMG_I_sequ3ence | Hospital-eff-F | Aminoglycosides |
| HiAQNR_sequ1ence | STP-inf-A | Quinolones |
| HiFQNR_sequ2ence | STP-inf-F | Quinolones |
| HoACLASS_d2_sequ45ence | STP-T-eff-A | Betalactams |
| SeFAMG_D_sequ3ence | Hospital-eff-F | Aminoglycosides |
| HoACLASS_d2_sequ46ence | STP-T-eff-A | Betalactams |
| HiAERM_ermtype_sequ7ence | STP-inf-A | Erythromycin |
| SeFAMG_E_sequ9ence | Hospital-eff-F | Aminoglycosides |
| SeAAMG_E_sequ9ence | Hospital-eff-A | Aminoglycosides |
| SeFAMG_E_sequ8ence | Hospital-eff-F | Aminoglycosides |
| HiFERM_ermtype_sequ9ence | STP-inf-F | Erythromycin |
| HoACLASS_d2_sequ48ence | STP-T-eff-A | Betalactams |
| HiAAMG_I_sequ6ence | STP-inf-A | Aminoglycosides |
| HoACLASS_d2_sequ4ence | STP-T-eff-A | Betalactams |
| HoACLASS_d2_sequ53ence | STP-T-eff-A | Betalactams |
| HiACLASS_b3_sequ2ence | STP-inf-A | Betalactams |
| SeFCLASS_a_sequ59ence | Hospital-eff-F | Betalactams |
| HiACLASS_a_sequ38ence | STP-inf-A | Betalactams |
| SeACLASS_a_sequ4ence | Hospital-eff-A | Betalactams |
| SeFCLASS_a_sequ10ence | Hospital-eff-F | Betalactams |
| HiFCLASS_d2_sequ12ence | STP-inf-F | Betalactams |
| HiFCLASS_d2_sequ6ence | STP-inf-F | Betalactams |
| HiACLASS_c_sequ8ence | STP-inf-A | Betalactams |
| HiFCLASS_a_sequ54ence | STP-inf-F | Betalactams |
| SeFCLASS_b12_sequ4ence | Hospital-eff-F | Betalactams |
| SeFERM_ermtype_sequ10ence | Hospital-eff-F | Erythromycin |
| SeACLASS_c_sequ9ence | Hospital-eff-A | Betalactams |
| SeFCLASS_c_sequ11ence | Hospital-eff-F | Betalactams |
| HiACLASS_a_sequ29ence | STP-inf-A | Betalactams |
| HiFCLASS_a_sequ24ence | STP-inf-F | Betalactams |
| SeACLASS_a_sequ21ence | Hospital-eff-A | Betalactams |
| SeFCLASS_a_sequ6ence | Hospital-eff-F | Betalactams |
| SeACLASS_b3_sequ1ence | Hospital-eff-A | Betalactams |
| SeFCLASS_a_sequ40ence | Hospital-eff-F | Betalactams |
| SeFCLASS_b3_sequ1ence | Hospital-eff-F | Betalactams |
| HiFCLASS_a_sequ39ence | STP-inf-F | Betalactams |
| HiACLASS_b3_sequ3ence | STP-inf-A | Betalactams |
| HiFCLASS_d2_sequ19ence | STP-inf-F | Betalactams |
| HiACLASS_d2_sequ32ence | STP-inf-A | Betalactams |
| SeAERM_ermtype_sequ4ence | Hospital-eff-A | Erythromycin |

| | | |
|---|---|---|
| SeACLASS_d2_sequ4ence | Hospital-eff-A | Betalactams |
| SeFCLASS_d2_sequ6ence | Hospital-eff-F | Betalactams |
| SeFCLASS_d2_sequ25ence | Hospital-eff-F | Betalactams |
| SeFERM_ermtype_sequ4ence | Hospital-eff-F | Erythromycin |
| HiAAMG_H_sequ5ence | STP-inf-A | Aminoglycosides |
| SeACLASS_d2_sequ12ence | Hospital-eff-A | Betalactams |
| SeFCLASS_a_sequ29ence | Hospital-eff-F | Betalactams |
| HiACLASS_b12_sequ4ence | STP-inf-A | Betalactams |
| HiFCLASS_b12_sequ3ence | STP-inf-F | Betalactams |
| SeACLASS_a_sequ30ence | Hospital-eff-A | Betalactams |
| SeACLASS_b12_sequ1ence | Hospital-eff-A | Betalactams |
| SeAQNR_sequ6ence | Hospital-eff-A | Quinolones |
| HiACLASS_d2_sequ24ence | STP-inf-A | Betalactams |
| HiFCLASS_d2_sequ18ence | STP-inf-F | Betalactams |
| HiACLASS_a_sequ18ence | STP-inf-A | Betalactams |
| HiFCLASS_a_sequ4ence | STP-inf-F | Betalactams |
| SeACLASS_a_sequ43ence | Hospital-eff-A | Betalactams |
| SeFCLASS_a_sequ66ence | Hospital-eff-F | Betalactams |
| SeFCLASS_d2_sequ3ence | Hospital-eff-F | Betalactams |
| SeACLASS_a_sequ39ence | Hospital-eff-A | Betalactams |
| HiACLASS_a_sequ42ence | STP-inf-A | Betalactams |
| HiACLASS_c_sequ4ence | STP-inf-A | Betalactams |
| HiACLASS_d2_sequ12ence | STP-inf-A | Betalactams |
| HiFCLASS_c_sequ2ence | STP-inf-F | Betalactams |
| SeACLASS_a_sequ38ence | Hospital-eff-A | Betalactams |
| HiACLASS_d2_sequ8ence | STP-inf-A | Betalactams |
| HiFCLASS_a_sequ57ence | STP-inf-F | Betalactams |
| HiFCLASS_d2_sequ10ence | STP-inf-F | Betalactams |
| HiFCLASS_c_sequ4ence | STP-inf-F | Betalactams |
| SeACLASS_a_sequ32ence | Hospital-eff-A | Betalactams |
| HiFCLASS_a_sequ16ence | STP-inf-F | Betalactams |
| HiFCLASS_b3_sequ5ence | STP-inf-F | Betalactams |
| HiFCLASS_c_sequ7ence | STP-inf-F | Betalactams |
| SeAAMG_D_sequ2ence | Hospital-eff-A | Aminoglycosides |
| SeACLASS_a_sequ33ence | Hospital-eff-A | Betalactams |
| SeFAMG_D_sequ4ence | Hospital-eff-F | Aminoglycosides |
| SeFCLASS_d2_sequ14ence | Hospital-eff-F | Betalactams |
| SeAAMG_E_sequ12ence | Hospital-eff-A | Aminoglycosides |
| HiFCLASS_d2_sequ3ence | STP-inf-F | Betalactams |
| HiACLASS_a_sequ40ence | STP-inf-A | Betalactams |
| HiACLASS_a_sequ6ence | STP-inf-A | Betalactams |
| HiFTET_eff_sequ5ence | STP-inf-F | Tetracyclines |
| SeACLASS_a_sequ25ence | Hospital-eff-A | Betalactams |
| SeFCLASS_a_sequ35ence | Hospital-eff-F | Betalactams |
| HiFCLASS_a_sequ53ence | STP-inf-F | Betalactams |
| SeFAMG_B_sequ6ence | Hospital-eff-F | Aminoglycosides |

| | | |
|---|---|---|
| SeFCLASS_a_sequ44ence | Hospital-eff-F | Betalactams |
| SeFQNR_sequ4ence | Hospital-eff-F | Quinolones |
| HiFERM_ermtype_sequ7ence | STP-inf-F | Erythromycin |
| HiFCLASS_a_sequ47ence | STP-inf-F | Betalactams |
| HiFCLASS_a_sequ59ence | STP-inf-F | Betalactams |
| SeFCLASS_a_sequ13ence | Hospital-eff-F | Betalactams |
| SeFCLASS_d1_sequ4ence | Hospital-eff-F | Betalactams |
| HiACLASS_a_sequ43ence | STP-inf-A | Betalactams |
| SeFAMG_E_sequ5ence | Hospital-eff-F | Aminoglycosides |
| SeFCLASS_a_sequ51ence | Hospital-eff-F | Betalactams |
| SeACLASS_a_sequ49ence | Hospital-eff-A | Betalactams |
| SeFAMG_H_sequ11ence | Hospital-eff-F | Aminoglycosides |
| SeFTET_rpg_sequ3ence | Hospital-eff-F | Tetracyclines |
| HiFAMG_D_sequ5ence | STP-inf-F | Aminoglycosides |
| HiFCLASS_a_sequ27ence | STP-inf-F | Betalactams |
| SeFCLASS_d2_sequ8ence | Hospital-eff-F | Betalactams |
| HiFCLASS_a_sequ3ence | STP-inf-F | Betalactams |
| HiFTET_rpg_sequ5ence | STP-inf-F | Tetracyclines |
| HiACLASS_d2_sequ29ence | STP-inf-A | Betalactams |
| SeACLASS_a_sequ44ence | Hospital-eff-A | Betalactams |
| SeFAMG_C_sequ4ence | Hospital-eff-F | Aminoglycosides |
| SeFCLASS_b12_sequ7ence | Hospital-eff-F | Betalactams |
| HiAAMG_E_sequ3ence | STP-inf-A | Aminoglycosides |
| HiACLASS_c_sequ6ence | STP-inf-A | Betalactams |
| HiACLASS_c_sequ7ence | STP-inf-A | Betalactams |
| HiACLASS_d2_sequ2ence | STP-inf-A | Betalactams |
| SeACLASS_a_sequ45ence | Hospital-eff-A | Betalactams |
| SeACLASS_c_sequ2ence | Hospital-eff-A | Betalactams |
| SeFCLASS_a_sequ27ence | Hospital-eff-F | Betalactams |
| SeFCLASS_c_sequ3ence | Hospital-eff-F | Betalactams |
| HiAAMG_H_sequ8ence | STP-inf-A | Aminoglycosides |
| HiACLASS_a_sequ39ence | STP-inf-A | Betalactams |
| HiApraminoglycosidemodelcSeq4 | STP-inf-A | Aminoglycosides |
| HiFAMG_B_sequ4ence | STP-inf-F | Aminoglycosides |
| SeFAMG_E_sequ10ence | Hospital-eff-F | Aminoglycosides |
| SeFCLASS_a_sequ46ence | Hospital-eff-F | Betalactams |
| HiACLASS_c_sequ2ence | STP-inf-A | Betalactams |
| HiFCLASS_c_sequ9ence | STP-inf-F | Betalactams |
| HiFERM_ermtype_sequ2ence | STP-inf-F | Erythromycin |
| SeATET_rpg_sequ1ence | Hospital-eff-A | Tetracyclines |
| SeFAMG_H_sequ2ence | Hospital-eff-F | Aminoglycosides |
| SeFTET_eff_sequ5ence | Hospital-eff-F | Tetracyclines |
| SeFTET_rpg_sequ5ence | Hospital-eff-F | Tetracyclines |
| HiAAMG_D_sequ2ence | STP-inf-A | Aminoglycosides |
| HiAERM_ermtype_sequ6ence | STP-inf-A | Erythromycin |
| SeATET_eff_sequ1ence | Hospital-eff-A | Tetracyclines |

| | | |
|---|---|---|
| SeFTET_eff_sequ4ence | Hospital-eff-F | Tetracyclines |
| HiAAMG_E_sequ8ence | STP-inf-A | Aminoglycosides |
| HiACLASS_b12_sequ1ence | STP-inf-A | Betalactams |
| HiFCLASS_b12_sequ2ence | STP-inf-F | Betalactams |
| HiFERM_ermtype_sequ3ence | STP-inf-F | Erythromycin |
| SeFCLASS_a_sequ24ence | Hospital-eff-F | Betalactams |
| SeFCLASS_a_sequ47ence | Hospital-eff-F | Betalactams |
| SeFCLASS_c_sequ14ence | Hospital-eff-F | Betalactams |
| SeFTET_rpg_sequ4ence | Hospital-eff-F | Tetracyclines |
| HiATET_rpg_sequ3ence | STP-inf-A | Tetracyclines |
| HiFAMG_H_sequ1ence | STP-inf-F | Aminoglycosides |
| HiFCLASS_d1_sequ2ence | STP-inf-F | Betalactams |
| SeFCLASS_d2_sequ18ence | Hospital-eff-F | Betalactams |
| HiFCLASS_a_sequ46ence | STP-inf-F | Betalactams |
| HiFCLASS_a_sequ55ence | STP-inf-F | Betalactams |
| SeACLASS_d2_sequ6ence | Hospital-eff-A | Betalactams |
| SeFAMG_E_sequ18ence | Hospital-eff-F | Aminoglycosides |
| SeFAMG_E_sequ7ence | Hospital-eff-F | Aminoglycosides |
| SeFCLASS_a_sequ45ence | Hospital-eff-F | Betalactams |
| SeFCLASS_b3_sequ2ence | Hospital-eff-F | Betalactams |
| SeFERM_ermtype_sequ11ence | Hospital-eff-F | Erythromycin |
| HiFCLASS_a_sequ37ence | STP-inf-F | Betalactams |
| SeFAMG_B_sequ3ence | Hospital-eff-F | Aminoglycosides |
| SeFAMG_I_sequ7ence | Hospital-eff-F | Aminoglycosides |
| SeFCLASS_c_sequ12ence | Hospital-eff-F | Betalactams |
| SeFERM_ermtype_sequ4ence | Hospital-eff-F | Erythromycin |
| SeAAMG_H_sequ2ence | Hospital-eff-A | Aminoglycosides |
| SeACLASS_b12_sequ7ence | Hospital-eff-A | Betalactams |
| SeFAMG_H_sequ5ence | Hospital-eff-F | Aminoglycosides |
| SeFERM_ermtype_sequ5ence | Hospital-eff-F | Erythromycin |
| HiAAMG_D_sequ3ence | STP-inf-A | Aminoglycosides |
| HiFAMG_D_sequ4ence | STP-inf-F | Aminoglycosides |
| HiFCLASS_b3_sequ4ence | STP-inf-F | Betalactams |
| HiFTET_rpg_sequ6ence | STP-inf-F | Tetracyclines |
| SeAAMG_D_sequ5ence | Hospital-eff-A | Aminoglycosides |
| SeACLASS_b3_sequ3ence | Hospital-eff-A | Betalactams |
| SeAERM_ermtype_sequ5ence | Hospital-eff-A | Erythromycin |
| SeFAMG_D_sequ6ence | Hospital-eff-F | Aminoglycosides |
| HiACLASS_a_sequ36ence | STP-inf-A | Betalactams |
| HiACLASS_a_sequ9ence | STP-inf-A | Betalactams |
| SeACLASS_a_sequ10ence | Hospital-eff-A | Betalactams |
| SeACLASS_a_sequ18ence | Hospital-eff-A | Betalactams |
| SeACLASS_a_sequ31ence | Hospital-eff-A | Betalactams |
| SeACLASS_b3_sequ4ence | Hospital-eff-A | Betalactams |
| SeFCLASS_a_sequ1ence | Hospital-eff-F | Betalactams |
| SeFCLASS_c_sequ5ence | Hospital-eff-F | Betalactams |

| | | |
|---|---|---|
| HiATET_rpg_sequ4ence | STP-inf-A | Tetracyclines |
| HiATET_rpg_sequ5ence | STP-inf-A | Tetracyclines |
| HiFCLASS_a_sequ17ence | STP-inf-F | Betalactams |
| SeFCLASS_a_sequ9ence | Hospital-eff-F | Betalactams |
| SeACLASS_c_sequ4ence | Hospital-eff-A | Betalactams |
| SeACLASS_c_sequ8ence | Hospital-eff-A | Betalactams |
| SeACLASS_d1_sequ6ence | Hospital-eff-A | Betalactams |
| SeACLASS_d2_sequ10ence | Hospital-eff-A | Betalactams |
| SeFCLASS_a_sequ48ence | Hospital-eff-F | Betalactams |
| SeFCLASS_b12_sequ6ence | Hospital-eff-F | Betalactams |
| SeFCLASS_c_sequ2ence | Hospital-eff-F | Betalactams |
| SeFCLASS_d2_sequ9ence | Hospital-eff-F | Betalactams |
| HiAAMG_B_sequ4ence | STP-inf-A | Aminoglycosides |
| HiACLASS_a_sequ13ence | STP-inf-A | Betalactams |
| HiACLASS_a_sequ22ence | STP-inf-A | Betalactams |
| HiACLASS_a_sequ7ence | STP-inf-A | Betalactams |
| HiACLASS_d2_sequ11ence | STP-inf-A | Betalactams |
| HiACLASS_d2_sequ1ence | STP-inf-A | Betalactams |
| HiFAMG_E_sequ5ence | STP-inf-F | Aminoglycosides |
| HiFAMG_E_sequ6ence | STP-inf-F | Aminoglycosides |
| HiFCLASS_a_sequ10ence | STP-inf-F | Betalactams |
| HiFCLASS_a_sequ19ence | STP-inf-F | Betalactams |
| HiFCLASS_a_sequ21ence | STP-inf-F | Betalactams |
| HiFCLASS_a_sequ28ence | STP-inf-F | Betalactams |
| HiFCLASS_a_sequ32ence | STP-inf-F | Betalactams |
| HiFCLASS_a_sequ43ence | STP-inf-F | Betalactams |
| HiFCLASS_d2_sequ1ence | STP-inf-F | Betalactams |
| SeAAMG_B_sequ1ence | Hospital-eff-A | Aminoglycosides |
| SeAAMG_E_sequ7ence | Hospital-eff-A | Aminoglycosides |
| SeACLASS_a_sequ15ence | Hospital-eff-A | Betalactams |
| SeACLASS_a_sequ47ence | Hospital-eff-A | Betalactams |
| SeACLASS_a_sequ48ence | Hospital-eff-A | Betalactams |
| SeACLASS_a_sequ9ence | Hospital-eff-A | Betalactams |
| SeACLASS_c_sequ5ence | Hospital-eff-A | Betalactams |
| SeAERM_ermtype_sequ3ence | Hospital-eff-A | Erythromycin |
| SeAMPH_sequ6ence | Hospital-eff-A | Aminoglycosides |
| SeFAMG_B_sequ2ence | Hospital-eff-F | Aminoglycosides |
| SeFCLASS_a_sequ23ence | Hospital-eff-F | Betalactams |
| SeFCLASS_a_sequ26ence | Hospital-eff-F | Betalactams |
| SeFCLASS_a_sequ4ence | Hospital-eff-F | Betalactams |
| SeFCLASS_a_sequ63ence | Hospital-eff-F | Betalactams |
| SeFCLASS_a_sequ70ence | Hospital-eff-F | Betalactams |
| SeFCLASS_d2_sequ19ence | Hospital-eff-F | Betalactams |
| HiAAMG_D_sequ4ence | STP-inf-A | Aminoglycosides |
| HiAAMG_E_sequ4ence | STP-inf-A | Aminoglycosides |
| HiFAMG_B_sequ3ence | STP-inf-F | Aminoglycosides |

| | | |
|---|---|---|
| HiFAMG_D_sequ3ence | STP-inf-F | Aminoglycosides |
| SeFAMG_B_sequ4ence | Hospital-eff-F | Aminoglycosides |
| SeFAMG_E_sequ4ence | Hospital-eff-F | Aminoglycosides |
| HiATET_rpg_sequ1ence | STP-inf-A | Tetracyclines |
| HiFQNR_sequ5ence | STP-inf-F | Quinolones |
| HiFQNR_sequ6ence | STP-inf-F | Quinolones |
| SeACLASS_c_sequ6ence | Hospital-eff-A | Betalactams |
| SeAQNR_sequ5ence | Hospital-eff-A | Quinolones |
| SeFCLASS_c_sequ8ence | Hospital-eff-F | Betalactams |
| SeFCLASS_c_sequ9ence | Hospital-eff-F | Betalactams |
| SeFQNR_sequ3ence | Hospital-eff-F | Quinolones |
| SeFTET_rpg_sequ2ence | Hospital-eff-F | Tetracyclines |
| HiAAMG_I_sequ1ence | STP-inf-A | Aminoglycosides |
| HiACLASS_b12_sequ2ence | STP-inf-A | Betalactams |
| HiACLASS_d2_sequ19ence | STP-inf-A | Betalactams |
| HiACLASS_d2_sequ20ence | STP-inf-A | Betalactams |
| HiAERM_ermtype_sequ4ence | STP-inf-A | Erythromycin |
| HiFAMG_I_sequ3ence | STP-inf-F | Aminoglycosides |
| HiFCLASS_a_sequ58ence | STP-inf-F | Betalactams |
| HiFCLASS_d2_sequ8ence | STP-inf-F | Betalactams |
| HiFERM_ermtype_sequ13ence | STP-inf-F | Erythromycin |
| SeAAMG_C_sequ2ence | Hospital-eff-A | Aminoglycosides |
| SeAAMG_I_sequ1ence | Hospital-eff-A | Aminoglycosides |
| SeAAMG_I_sequ3ence | Hospital-eff-A | Aminoglycosides |
| SeACLASS_b12_sequ3ence | Hospital-eff-A | Betalactams |
| SeACLASS_b12_sequ4ence | Hospital-eff-A | Betalactams |
| SeACLASS_d2_sequ11ence | Hospital-eff-A | Betalactams |
| SeACLASS_d2_sequ15ence | Hospital-eff-A | Betalactams |
| SeACLASS_d2_sequ8ence | Hospital-eff-A | Betalactams |
| SeAERM_ermtype_sequ6ence | Hospital-eff-A | Erythromycin |
| SeAQNR_sequ4ence | Hospital-eff-A | Quinolones |
| SeFAMG_C_sequ3ence | Hospital-eff-F | Aminoglycosides |
| SeFAMG_I_sequ1ence | Hospital-eff-F | Aminoglycosides |
| SeFCLASS_a_sequ39ence | Hospital-eff-F | Betalactams |
| SeFCLASS_a_sequ42ence | Hospital-eff-F | Betalactams |
| SeFCLASS_a_sequ57ence | Hospital-eff-F | Betalactams |
| SeFCLASS_a_sequ58ence | Hospital-eff-F | Betalactams |
| SeFCLASS_b12_sequ3ence | Hospital-eff-F | Betalactams |
| SeFCLASS_d2_sequ16ence | Hospital-eff-F | Betalactams |
| SeFERM_ermtype_sequ6ence | Hospital-eff-F | Erythromycin |
| HiACLASS_a_sequ15ence | STP-inf-A | Betalactams |
| HiACLASS_a_sequ23ence | STP-inf-A | Betalactams |
| HiACLASS_a_sequ2ence | STP-inf-A | Betalactams |
| HiACLASS_a_sequ33ence | STP-inf-A | Betalactams |
| HiACLASS_a_sequ34ence | STP-inf-A | Betalactams |
| HiACLASS_a_sequ3ence | STP-inf-A | Betalactams |

| | | |
|---|---|---|
| HiACLASS_c_sequ10ence | STP-inf-A | Betalactams |
| HiACLASS_c_sequ9ence | STP-inf-A | Betalactams |
| HiACLASS_d1_sequ1ence | STP-inf-A | Betalactams |
| HiAERM_ermtype_sequ5ence | STP-inf-A | Erythromycin |
| HiFCLASS_a_sequ12ence | STP-inf-F | Betalactams |
| HiFCLASS_a_sequ15ence | STP-inf-F | Betalactams |
| HiFCLASS_a_sequ31ence | STP-inf-F | Betalactams |
| HiFCLASS_a_sequ44ence | STP-inf-F | Betalactams |
| HiFCLASS_a_sequ45ence | STP-inf-F | Betalactams |
| HiFCLASS_a_sequ50ence | STP-inf-F | Betalactams |
| HiFCLASS_a_sequ6ence | STP-inf-F | Betalactams |
| HiFCLASS_a_sequ8ence | STP-inf-F | Betalactams |
| HiFCLASS_c_sequ3ence | STP-inf-F | Betalactams |
| HiFCLASS_c_sequ6ence | STP-inf-F | Betalactams |
| HiFCLASS_d1_sequ1ence | STP-inf-F | Betalactams |
| HiFMPH_sequ3ence | STP-inf-F | Macrolides |
| HiFMPH_sequ5ence | STP-inf-F | Macrolides |
| HiFMPH_sequ7ence | STP-inf-F | Macrolides |
| HiFTET_eff_sequ4ence | STP-inf-F | Tetracyclines |
| HoACLASS_a_sequ25ence | STP-T-eff-A | Betalactams |
| HoACLASS_a_sequ28ence | STP-T-eff-A | Betalactams |
| HoACLASS_a_sequ2ence | STP-T-eff-A | Betalactams |
| HoACLASS_a_sequ31ence | STP-T-eff-A | Betalactams |
| HoACLASS_a_sequ34ence | STP-T-eff-A | Betalactams |
| HoACLASS_a_sequ38ence | STP-T-eff-A | Betalactams |
| HoACLASS_a_sequ3ence | STP-T-eff-A | Betalactams |
| HoACLASS_a_sequ40ence | STP-T-eff-A | Betalactams |
| HoACLASS_a_sequ46ence | STP-T-eff-A | Betalactams |
| HoACLASS_a_sequ50ence | STP-T-eff-A | Betalactams |
| HoACLASS_a_sequ58ence | STP-T-eff-A | Betalactams |
| HoACLASS_a_sequ78ence | STP-T-eff-A | Betalactams |
| HoACLASS_a_sequ9ence | STP-T-eff-A | Betalactams |
| HoACLASS_c_sequ12ence | STP-T-eff-A | Betalactams |
| HoACLASS_d1_sequ7ence | STP-T-eff-A | Betalactams |
| HoAMPH_sequ3ence | STP-T-eff-A | Aminoglycosides |
| HoAMPH_sequ4ence | STP-T-eff-A | Aminoglycosides |
| HoATET_rpg_sequ2ence | STP-T-eff-A | Tetracyclines |
| HoFCLASS_a_sequ17ence | STP-T-eff-F | Betalactams |
| HoFCLASS_a_sequ1ence | STP-T-eff-F | Betalactams |
| HoFCLASS_a_sequ22ence | STP-T-eff-F | Betalactams |
| HoFCLASS_a_sequ26ence | STP-T-eff-F | Betalactams |
| HoFCLASS_a_sequ37ence | STP-T-eff-F | Betalactams |
| HoFCLASS_a_sequ43ence | STP-T-eff-F | Betalactams |
| HoFCLASS_a_sequ54ence | STP-T-eff-F | Betalactams |
| HoFCLASS_a_sequ6ence | STP-T-eff-F | Betalactams |
| HoFCLASS_a_sequ9ence | STP-T-eff-F | Betalactams |

| | | |
|---|---|---|
| HoFCLASS_c_sequ9ence | STP-T-eff-F | Betalactams |
| HoFMPH_sequ3ence | STP-T-eff-F | Macrolides |
| HoFTET_rpg_sequ1ence | STP-T-eff-F | Tetracyclines |
| SeAAMG_G_sequ2ence | Hospital-eff-A | Aminoglycosides |
| SeACLASS_a_sequ12ence | Hospital-eff-A | Betalactams |
| SeACLASS_a_sequ19ence | Hospital-eff-A | Betalactams |
| SeACLASS_a_sequ26ence | Hospital-eff-A | Betalactams |
| SeACLASS_a_sequ35ence | Hospital-eff-A | Betalactams |
| SeACLASS_a_sequ40ence | Hospital-eff-A | Betalactams |
| SeACLASS_a_sequ6ence | Hospital-eff-A | Betalactams |
| SeACLASS_a_sequ7ence | Hospital-eff-A | Betalactams |
| SeACLASS_c_sequ10ence | Hospital-eff-A | Betalactams |
| SeACLASS_c_sequ3ence | Hospital-eff-A | Betalactams |
| SeATET_rpg_sequ3ence | Hospital-eff-A | Tetracyclines |
| SeFAMG_G_sequ3ence | Hospital-eff-F | Aminoglycosides |
| SeFCLASS_a_sequ14ence | Hospital-eff-F | Betalactams |
| SeFCLASS_a_sequ17ence | Hospital-eff-F | Betalactams |
| SeFCLASS_a_sequ21ence | Hospital-eff-F | Betalactams |
| SeFCLASS_a_sequ30ence | Hospital-eff-F | Betalactams |
| SeFCLASS_a_sequ33ence | Hospital-eff-F | Betalactams |
| SeFCLASS_a_sequ50ence | Hospital-eff-F | Betalactams |
| SeFCLASS_a_sequ5ence | Hospital-eff-F | Betalactams |
| SeFCLASS_a_sequ7ence | Hospital-eff-F | Betalactams |
| SeFCLASS_b3_sequ6ence | Hospital-eff-F | Betalactams |
| SeFCLASS_c_sequ10ence | Hospital-eff-F | Betalactams |
| SeFCLASS_d2_sequ1ence | Hospital-eff-F | Betalactams |
| SeFMPH_sequ1ence | Hospital-eff-F | Macrolides |
| SeFTET_rpg_sequ7ence | Hospital-eff-F | Tetracyclines |
| HiATET_eff_sequ3ence | STP-inf-A | Tetracyclines |
| HiFTET_eff_sequ3ence | STP-inf-F | Tetracyclines |
| HiFTET_rpg_sequ1ence | STP-inf-F | Tetracyclines |
| HoATET_eff_sequ3ence | STP-T-eff-A | Tetracyclines |
| HoATET_rpg_sequ4ence | STP-T-eff-A | Tetracyclines |
| HoFTET_eff_sequ1ence | STP-T-eff-F | Tetracyclines |
| HoFTET_rpg_sequ4ence | STP-T-eff-F | Tetracyclines |
| SeATET_eff_sequ2ence | Hospital-eff-A | Tetracyclines |
| SeATET_eff_sequ3ence | Hospital-eff-A | Tetracyclines |
| SeATET_rpg_sequ2ence | Hospital-eff-A | Tetracyclines |
| SeFTET_eff_sequ1ence | Hospital-eff-F | Tetracyclines |
| SeFTET_eff_sequ2ence | Hospital-eff-F | Tetracyclines |
| SeFTET_eff_sequ3ence | Hospital-eff-F | Tetracyclines |
| HoACLASS_d2_sequ55ence | STP-T-eff-A | Betalactams |
| HoACLASS_d2_sequ5ence | STP-T-eff-A | Betalactams |
| HoACLASS_d2_sequ8ence | STP-T-eff-A | Betalactams |
| HoFCLASS_a_sequ10ence | STP-T-eff-F | Betalactams |
| HoFCLASS_a_sequ14ence | STP-T-eff-F | Betalactams |

| | | |
|---|---|---|
| HoFCLASS_a_sequ18ence | STP-T-eff-F | Betalactams |
| HoFCLASS_a_sequ20ence | STP-T-eff-F | Betalactams |
| HoFCLASS_a_sequ21ence | STP-T-eff-F | Betalactams |
| HoFCLASS_a_sequ23ence | STP-T-eff-F | Betalactams |
| HoAERM_ermtype_sequ10ence | STP-T-eff-A | Erythromycin |
| HoFCLASS_a_sequ24ence | STP-T-eff-F | Betalactams |
| HoFCLASS_a_sequ27ence | STP-T-eff-F | Betalactams |
| HoFCLASS_a_sequ28ence | STP-T-eff-F | Betalactams |
| HoFCLASS_a_sequ31ence | STP-T-eff-F | Betalactams |
| HoFCLASS_a_sequ34ence | STP-T-eff-F | Betalactams |
| HoFCLASS_a_sequ35ence | STP-T-eff-F | Betalactams |
| HoFCLASS_a_sequ36ence | STP-T-eff-F | Betalactams |
| HoFCLASS_a_sequ38ence | STP-T-eff-F | Betalactams |
| HoFCLASS_a_sequ40ence | STP-T-eff-F | Betalactams |
| HoAAMG_I_sequ7ence | STP-T-eff-A | Aminoglycosides |
| HoAERM_ermtype_sequ11ence | STP-T-eff-A | Erythromycin |
| HoAERM_ermtype_sequ15ence | STP-T-eff-A | Erythromycin |
| HoFCLASS_a_sequ42ence | STP-T-eff-F | Betalactams |
| HoFCLASS_a_sequ44ence | STP-T-eff-F | Betalactams |
| HoFCLASS_a_sequ48ence | STP-T-eff-F | Betalactams |
| HoFCLASS_a_sequ4ence | STP-T-eff-F | Betalactams |
| HoFAMG_A_sequ1ence | STP-T-eff-F | Aminoglycosides |
| HoFCLASS_a_sequ52ence | STP-T-eff-F | Betalactams |
| HoFCLASS_a_sequ56ence | STP-T-eff-F | Betalactams |
| HoAAMG_C_sequ3ence | STP-T-eff-A | Aminoglycosides |
| HoFCLASS_a_sequ58ence | STP-T-eff-F | Betalactams |
| HoFCLASS_a_sequ8ence | STP-T-eff-F | Betalactams |
| HoAAMG_I_sequ1ence | STP-T-eff-A | Aminoglycosides |
| HoFCLASS_b12_sequ10ence | STP-T-eff-F | Betalactams |
| HoFCLASS_b12_sequ11ence | STP-T-eff-F | Betalactams |
| HoAAMG_I_sequ3ence | STP-T-eff-A | Aminoglycosides |
| HoFCLASS_b12_sequ13ence | STP-T-eff-F | Betalactams |
| HoAERM_ermtype_sequ14ence | STP-T-eff-A | Erythromycin |
| HoFCLASS_b12_sequ1ence | STP-T-eff-F | Betalactams |
| HoFAMG_I_sequ4ence | STP-T-eff-F | Aminoglycosides |
| HoFCLASS_b12_sequ2ence | STP-T-eff-F | Betalactams |
| HoFCLASS_b12_sequ3ence | STP-T-eff-F | Betalactams |
| HoFCLASS_b12_sequ5ence | STP-T-eff-F | Betalactams |
| HoFCLASS_b12_sequ6ence | STP-T-eff-F | Betalactams |
| HoFCLASS_b12_sequ7ence | STP-T-eff-F | Betalactams |
| HoFCLASS_b12_sequ8ence | STP-T-eff-F | Betalactams |
| HoFCLASS_b12_sequ9ence | STP-T-eff-F | Betalactams |
| HoFCLASS_b3_sequ1ence | STP-T-eff-F | Betalactams |
| HoFAMG_I_sequ6ence | STP-T-eff-F | Aminoglycosides |
| HoFCLASS_c_sequ11ence | STP-T-eff-F | Betalactams |
| HoFCLASS_c_sequ1ence | STP-T-eff-F | Betalactams |

| | | |
|---|---|---|
| HoFCLASS_c_sequ2ence | STP-T-eff-F | Betalactams |
| HoFCLASS_c_sequ8ence | STP-T-eff-F | Betalactams |
| HoFCLASS_d1_sequ1ence | STP-T-eff-F | Betalactams |
| HoAAMG_C_sequ4ence | STP-T-eff-A | Aminoglycosides |
| HoFCLASS_d1_sequ3ence | STP-T-eff-F | Betalactams |
| HoAAMG_H_sequ4ence | STP-T-eff-A | Aminoglycosides |
| HoFCLASS_d2_sequ15ence | STP-T-eff-F | Betalactams |
| HoAAMG_I_sequ10ence | STP-T-eff-A | Aminoglycosides |
| HoFCLASS_d2_sequ18ence | STP-T-eff-F | Betalactams |
| HoFCLASS_d2_sequ1ence | STP-T-eff-F | Betalactams |
| HoFCLASS_d2_sequ20ence | STP-T-eff-F | Betalactams |
| HoFCLASS_d2_sequ24ence | STP-T-eff-F | Betalactams |
| HoFCLASS_d2_sequ25ence | STP-T-eff-F | Betalactams |
| HoFCLASS_d2_sequ27ence | STP-T-eff-F | Betalactams |
| HoFCLASS_d2_sequ28ence | STP-T-eff-F | Betalactams |
| HoFCLASS_d2_sequ2ence | STP-T-eff-F | Betalactams |
| HoFCLASS_d2_sequ31ence | STP-T-eff-F | Betalactams |
| HoFCLASS_d2_sequ32ence | STP-T-eff-F | Betalactams |
| HoFCLASS_d2_sequ3ence | STP-T-eff-F | Betalactams |
| HoFCLASS_d2_sequ6ence | STP-T-eff-F | Betalactams |
| HoFCLASS_d2_sequ9ence | STP-T-eff-F | Betalactams |
| HoAAMG_E_sequ7ence | STP-T-eff-A | Aminoglycosides |
| HoFMPH_sequ5ence | STP-T-eff-F | Betalactams |
| SeACLASS_a_sequ22ence | Hospital-eff-A | Betalactams |
| SeACLASS_a_sequ27ence | Hospital-eff-A | Betalactams |
| HoAAMG_C_sequ2ence | STP-T-eff-A | Aminoglycosides |
| SeACLASS_a_sequ28ence | Hospital-eff-A | Betalactams |
| SeACLASS_a_sequ34ence | Hospital-eff-A | Betalactams |
| SeACLASS_a_sequ36ence | Hospital-eff-A | Betalactams |
| SeACLASS_a_sequ37ence | Hospital-eff-A | Betalactams |
| SeACLASS_a_sequ41ence | Hospital-eff-A | Betalactams |
| HoFAMG_E_sequ6ence | STP-T-eff-F | Aminoglycosides |
| SeACLASS_b12_sequ6ence | Hospital-eff-A | Betalactams |
| SeACLASS_c_sequ1ence | Hospital-eff-A | Betalactams |
| SeACLASS_d1_sequ5ence | Hospital-eff-A | Betalactams |
| SeACLASS_d2_sequ14ence | Hospital-eff-A | Betalactams |
| SeACLASS_d2_sequ3ence | Hospital-eff-A | Betalactams |
| SeACLASS_d2_sequ7ence | Hospital-eff-A | Betalactams |
| SeACLASS_d2_sequ9ence | Hospital-eff-A | Betalactams |
| SeFCLASS_a_sequ18ence | Hospital-eff-F | Betalactams |
| SeFCLASS_a_sequ19ence | Hospital-eff-F | Betalactams |
| SeFCLASS_a_sequ25ence | Hospital-eff-F | Betalactams |
| SeFCLASS_a_sequ31ence | Hospital-eff-F | Betalactams |
| SeFCLASS_a_sequ32ence | Hospital-eff-F | Betalactams |
| SeFCLASS_a_sequ34ence | Hospital-eff-F | Betalactams |
| SeFCLASS_a_sequ41ence | Hospital-eff-F | Betalactams |

| | | |
|---|---|---|
| SeFCLASS_a_sequ49ence | Hospital-eff-F | Betalactams |
| SeFCLASS_a_sequ55ence | Hospital-eff-F | Betalactams |
| HoATET_eff_sequ7ence | STP-T-eff-A | Tetracyclines |
| SeFCLASS_a_sequ60ence | Hospital-eff-F | Betalactams |
| HoAAMG_C_sequ6ence | STP-T-eff-A | Aminoglycosides |
| SeFCLASS_a_sequ68ence | Hospital-eff-F | Betalactams |
| SeFCLASS_a_sequ69ence | Hospital-eff-F | Betalactams |
| SeFCLASS_b12_sequ5ence | Hospital-eff-F | Betalactams |
| SeFCLASS_b12_sequ8ence | Hospital-eff-F | Betalactams |
| SeFCLASS_b3_sequ4ence | Hospital-eff-F | Betalactams |
| SeFCLASS_b3_sequ8ence | Hospital-eff-F | Betalactams |
| SeFCLASS_c_sequ1ence | Hospital-eff-F | Betalactams |
| SeFCLASS_c_sequ6ence | Hospital-eff-F | Betalactams |
| SeFCLASS_c_sequ7ence | Hospital-eff-F | Betalactams |
| SeFCLASS_d1_sequ3ence | Hospital-eff-F | Betalactams |
| SeFCLASS_d1_sequ7ence | Hospital-eff-F | Betalactams |
| SeFCLASS_d1_sequ8ence | Hospital-eff-F | Betalactams |
| SeFCLASS_d2_sequ10ence | Hospital-eff-F | Betalactams |
| HoAAMG_I_sequ9ence | STP-T-eff-A | Aminoglycosides |
| SeFCLASS_d2_sequ11ence | Hospital-eff-F | Betalactams |
| SeFCLASS_d2_sequ12ence | Hospital-eff-F | Betalactams |
| HoATET_eff_sequ9ence | STP-T-eff-A | Tetracyclines |
| HoAAMG_C_sequ5ence | STP-T-eff-A | Aminoglycosides |
| HoATET_eff_sequ8ence | STP-T-eff-A | Tetracyclines |
| HoAQNR_sequ1ence | STP-T-eff-A | Quinolones |
| HoFQNR_sequ1ence | STP-T-eff-F | Quinolones |
| HoAERM_ermtype_sequ2ence | STP-T-eff-A | Erythromycin |
| SeFCLASS_d2_sequ13ence | Hospital-eff-F | Betalactams |
| SeFCLASS_d2_sequ17ence | Hospital-eff-F | Betalactams |
| SeFCLASS_d2_sequ20ence | Hospital-eff-F | Betalactams |
| SeFCLASS_d2_sequ23ence | Hospital-eff-F | Betalactams |
| SeFCLASS_d2_sequ24ence | Hospital-eff-F | Betalactams |
| SeFCLASS_d2_sequ7ence | Hospital-eff-F | Betalactams |
| HoACLASS_d2_sequ18ence | STP-T-eff-A | Betalactams |
| HoACLASS_a_sequ51ence | STP-T-eff-A | Betalactams |
| HoACLASS_b12_sequ17ence | STP-T-eff-A | Betalactams |
| HoACLASS_a_sequ42ence | STP-T-eff-A | Betalactams |
| HoFCLASS_d2_sequ16ence | STP-T-eff-F | Betalactams |
| HoACLASS_d2_sequ14ence | STP-T-eff-A | Betalactams |
| HoACLASS_d2_sequ43ence | STP-T-eff-A | Betalactams |
| HoACLASS_a_sequ15ence | STP-T-eff-A | Betalactams |
| HoFCLASS_a_sequ11ence | STP-T-eff-F | Betalactams |
| HoACLASS_d2_sequ17ence | STP-T-eff-A | Betalactams |
| HoFCLASS_d2_sequ12ence | STP-T-eff-F | Betalactams |
| HoFCLASS_d2_sequ33ence | STP-T-eff-F | Betalactams |
| HoACLASS_d2_sequ33ence | STP-T-eff-A | Betalactams |

| | | |
|---|---|---|
| HoACLASS_a_sequ5ence | STP-T-eff-A | Betalactams |
| HoFCLASS_a_sequ13ence | STP-T-eff-F | Betalactams |
| HoACLASS_d2_sequ57ence | STP-T-eff-A | Betalactams |
| HoACLASS_d2_sequ54ence | STP-T-eff-A | Betalactams |
| HoATET_eff_sequ5ence | STP-T-eff-A | Tetracyclines |
| HoACLASS_c_sequ9ence | STP-T-eff-A | Betalactams |
| HoACLASS_b12_sequ11ence | STP-T-eff-A | Betalactams |
| HoACLASS_b12_sequ1ence | STP-T-eff-A | Betalactams |
| HoFCLASS_b12_sequ12ence | STP-T-eff-F | Betalactams |
| HoACLASS_b3_sequ4ence | STP-T-eff-A | Betalactams |
| HoACLASS_d2_sequ36ence | STP-T-eff-A | Betalactams |
| HoFCLASS_d2_sequ22ence | STP-T-eff-F | Betalactams |
| HoACLASS_a_sequ32ence | STP-T-eff-A | Betalactams |
| HoFCLASS_a_sequ55ence | STP-T-eff-F | Betalactams |
| HoACLASS_d2_sequ58ence | STP-T-eff-A | Betalactams |
| HoACLASS_c_sequ15ence | STP-T-eff-A | Betalactams |
| HoACLASS_d2_sequ15ence | STP-T-eff-A | Betalactams |
| HoFCLASS_c_sequ3ence | STP-T-eff-F | Betalactams |
| HoACLASS_d2_sequ50ence | STP-T-eff-A | Betalactams |
| HoACLASS_d2_sequ6ence | STP-T-eff-A | Betalactams |
| HoAAMG_H_sequ9ence | STP-T-eff-A | Aminoglycosides |
| HoACLASS_d2_sequ35ence | STP-T-eff-A | Betalactams |
| HoAAMG_D_sequ3ence | STP-T-eff-A | Aminoglycosides |
| HoFCLASS_c_sequ7ence | STP-T-eff-F | Betalactams |
| HoACLASS_a_sequ82ence | STP-T-eff-A | Betalactams |
| HoFCLASS_a_sequ33ence | STP-T-eff-F | Betalactams |
| HoFCLASS_d2_sequ10ence | STP-T-eff-F | Betalactams |
| HoACLASS_b12_sequ13ence | STP-T-eff-A | Betalactams |
| HoACLASS_d2_sequ12ence | STP-T-eff-A | Betalactams |
| HoACLASS_d2_sequ47ence | STP-T-eff-A | Betalactams |
| HoACLASS_c_sequ5ence | STP-T-eff-A | Betalactams |
| HoAAMG_E_sequ3ence | STP-T-eff-A | Aminoglycosides |
| HoACLASS_a_sequ64ence | STP-T-eff-A | Betalactams |
| HoFCLASS_a_sequ45ence | STP-T-eff-F | Betalactams |
| HoACLASS_d2_sequ34ence | STP-T-eff-A | Betalactams |
| HoACLASS_d2_sequ40ence | STP-T-eff-A | Betalactams |
| HoFCLASS_d2_sequ5ence | STP-T-eff-F | Betalactams |
| HoFAMG_I_sequ3ence | STP-T-eff-F | Aminoglycosides |
| HoAAMG_H_sequ10ence | STP-T-eff-A | Aminoglycosides |
| HoACLASS_b3_sequ2ence | STP-T-eff-A | Betalactams |
| HoACLASS_d2_sequ56ence | STP-T-eff-A | Betalactams |
| HoFAMG_D_sequ2ence | STP-T-eff-F | Aminoglycosides |
| HoFAMG_H_sequ3ence | STP-T-eff-F | Aminoglycosides |
| HoFCLASS_c_sequ6ence | STP-T-eff-F | Betalactams |
| HoAERM_ermtype_sequ3ence | STP-T-eff-A | Erythromycin |
| HoFQNR_sequ2ence | STP-T-eff-F | Quinolones |

| | | |
|---|---|---|
| HoFCLASS_a_sequ30ence | STP-T-eff-F | Betalactams |
| HoACLASS_b3_sequ6ence | STP-T-eff-A | Betalactams |
| HoFCLASS_a_sequ39ence | STP-T-eff-F | Betalactams |
| HoFCLASS_d2_sequ26ence | STP-T-eff-F | Betalactams |
| HoACLASS_a_sequ29ence | STP-T-eff-A | Betalactams |
| HoACLASS_c_sequ11ence | STP-T-eff-A | Betalactams |
| HoACLASS_c_sequ13ence | STP-T-eff-A | Betalactams |
| HoACLASS_c_sequ6ence | STP-T-eff-A | Betalactams |
| HoFCLASS_d2_sequ30ence | STP-T-eff-F | Betalactams |
| HoACLASS_a_sequ75ence | STP-T-eff-A | Betalactams |
| HoATET_eff_sequ6ence | STP-T-eff-A | Tetracyclines |
| HoAAMG_B_sequ4ence | STP-T-eff-A | Aminoglycosides |
| HoACLASS_c_sequ10ence | STP-T-eff-A | Betalactams |
| HoFCLASS_a_sequ3ence | STP-T-eff-F | Betalactams |
| HoFCLASS_b12_sequ4ence | STP-T-eff-F | Betalactams |
| HoAAMG_D_sequ4ence | STP-T-eff-A | Aminoglycosides |
| HoACLASS_d2_sequ30ence | STP-T-eff-A | Betalactams |
| HoFAMG_D_sequ1ence | STP-T-eff-F | Aminoglycosides |
| HoFCLASS_c_sequ10ence | STP-T-eff-F | Betalactams |
| HoFTET_rpg_sequ2ence | STP-T-eff-F | Tetracyclines |
| HoAERM_ermtype_sequ4ence | STP-T-eff-A | Erythromycin |
| HoATET_rpg_sequ10ence | STP-T-eff-A | Tetracyclines |
| HoFCLASS_a_sequ29ence | STP-T-eff-F | Betalactams |
| HoFCLASS_a_sequ50ence | STP-T-eff-F | Betalactams |
| HoFCLASS_a_sequ51ence | STP-T-eff-F | Betalactams |
| HoACLASS_c_sequ14ence | STP-T-eff-A | Betalactams |
| HoATET_rpg_sequ1ence | STP-T-eff-A | Tetracyclines |
| HoFAMG_D_sequ3ence | STP-T-eff-F | Aminoglycosides |
| HoACLASS_b12_sequ7ence | STP-T-eff-A | Betalactams |
| HoATET_rpg_sequ5ence | STP-T-eff-A | Tetracyclines |
| HoFCLASS_d2_sequ35ence | STP-T-eff-F | Betalactams |
| HoFTET_eff_sequ4ence | STP-T-eff-F | Tetracyclines |
| HoAAMG_B_sequ3ence | STP-T-eff-A | Aminoglycosides |
| HoAAMG_E_sequ2ence | STP-T-eff-A | Aminoglycosides |
| HoACLASS_a_sequ18ence | STP-T-eff-A | Betalactams |
| HoACLASS_a_sequ26ence | STP-T-eff-A | Betalactams |
| HoACLASS_a_sequ59ence | STP-T-eff-A | Betalactams |
| HoACLASS_a_sequ66ence | STP-T-eff-A | Betalactams |
| HoACLASS_d2_sequ44ence | STP-T-eff-A | Betalactams |
| HoFAMG_E_sequ3ence | STP-T-eff-F | Aminoglycosides |
| HoFCLASS_a_sequ41ence | STP-T-eff-F | Betalactams |
| HoFCLASS_a_sequ53ence | STP-T-eff-F | Betalactams |
| HoFCLASS_d2_sequ23ence | STP-T-eff-F | Betalactams |
| HoATET_rpg_sequ6ence | STP-T-eff-A | Tetracyclines |
| HoAQNR_sequ3ence | STP-T-eff-A | Quinolones |
| HoATET_rpg_sequ9ence | STP-T-eff-A | Tetracyclines |

| | | |
|---|---|---|
| HoFCLASS_c_sequ4ence | STP-T-eff-F | Betalactams |
| HoAAMG_I_sequ2ence | STP-T-eff-A | Aminoglycosides |
| HoACLASS_a_sequ12ence | STP-T-eff-A | Betalactams |
| HoACLASS_a_sequ63ence | STP-T-eff-A | Betalactams |
| HoACLASS_b12_sequ12ence | STP-T-eff-A | Betalactams |
| HoACLASS_d2_sequ1ence | STP-T-eff-A | Betalactams |
| HoAERM_ermtype_sequ4ence | STP-T-eff-A | Erythromycin |
| HoAERM_ermtype_sequ6ence | STP-T-eff-A | Erythromycin |
| HoFAMG_I_sequ1ence | STP-T-eff-F | Aminoglycosides |
| HoFCLASS_a_sequ15ence | STP-T-eff-F | Betalactams |
| HoFCLASS_d2_sequ7ence | STP-T-eff-F | Betalactams |
| HoFERM_ermtype_sequ2ence | STP-T-eff-F | Erythromycin |

**Legend:** eff (effluent); F (February); A (April); STP (sewage treatment plant); inf (influent); T (treated samples.

| Protein length (aa) |
|---|
| 302 |
| 154 |
| 154 |
| 254 |
| 152 |
| 145 |
| 154 |
| 361 |
| 264 |
| 260 |
| 272 |
| 267 |
| 293 |
| 293 |
| 295 |
| 295 |
| 330 |
| 308 |
| 298 |
| 317 |
| 286 |
| 321 |
| 322 |
| 291 |
| 374 |
| 384 |
| 404 |
| 271 |
| 264 |
| 266 |
| 282 |
| 274 |
| 266 |
| 253 |
| 283 |
| 275 |
| 264 |
| 245 |
| 290 |
| 298 |
| 266 |
| 244 |
| 294 |

294
295
294
294
218
424
396
388
392
646
614
154
138
294
213
173
152
154
209
177
321
312
267
264
272
293
290
321
298
345
301
319
284
295
296
293
274
322
275
294
292
293
404
392
274
266

264
266
277
271
279
266
253
283
282
247
247
264
244
266
245
290
304
266
266
294
295
294
294
218
226
396
424
392
388
396
639
680
646
644
662
639
154
154
294
255
242
152
145
177
154
148

361
292
264
260
272
267
293
308
300
330
293
301
332
298
289
300
321
298
322
289
294
290
272
286
295
284
291
292
291
391
407
275
266
260
253
266
267
274
271
283
282
275
264
266
245
264

244
269
290
258
266
257
263
304
294
295
301
218
424
396
405
388
392
599
646
657
660
154
139
294
232
152
154
177
145
305
267
264
293
319
321
296
295
293
269
290
281
267
295
322
273
404

276
266
274
264
257
266
282
259
253
271
245
244
290
266
266
252
294
295
302
396
424
392
388
646
599
154
154
294
184
144
145
152
154
157
182
361
267
272
260
271
264
301
331
286
295
346

321
296
319
293
301
308
266
322
317
246
267
319
407
404
391
291
266
279
270
283
282
275
281
274
245
269
244
266
266
268
294
301
294
294
294
218
396
405
412
388
632
154
154
294
173
184

157
144
145
144
152
155
182
361
264
271
272
266
267
308
296
286
300
304
298
295
301
294
288
321
297
311
287
293
290
284
273
291
302
274
322
267
246
319
291
292
404
391
291
264
275
279

243
270
267
274
282
271
244
275
281
266
266
245
269
244
266
309
264
263
290
266
301
294
280
218
235
218
405
396
374
388
396
639
680
335
210
301
318
296
302
365
314
314
206
336
356
276

293
271
238
406
389
322
310
274
292
274
251
246
164
145
291
246
258
283
179
309
309
312
283
258
144
259
307
261
314
296
312
293
282
314
301
318
299
302
169
309
283
254
308
283
317
312

302
251
242
251
265
354
377
261
406
258
274
314
277
267
261
277
295
256
251
283
256
283
267
268
310
310
293
291
296
260
228
172
301
333
239
155
273
318
277
293
290
290
217
217
277
308

264
368
365
325
308
314
298
146
318
298
306
302
301
239
331
331
145
145
296
294
292
271
270
299
297
272
256
302
257
303
270
298
291
314
266
258
291
346
302
225
149
264
258
180
266
266

255
235
241
264
157
254
243
252
242
241
251
241
321
300
405
218
406
389
290
181
385
252
263
265
281
390
283
268
218
218
285
245
259
260
301
282
265
274
261
277
264
265
264
283
376
269

279
148
270
269
269
258
304
218
218
264
171
271
301
146
151
151
243
272
296
268
246
273
298
315
315
315
273
266
396
288
252
284
396
396
293
293
293
293
316
302
316
319
271
254
244
283

253
268
265
283
270
268
298
252
252
313
252
231
279
279
304
304
304
304
301
291
294
387
303
387
298
277
281
277
442
295
295
319
396
184
291
184
271
139
301
296
295
413
292
292
303
155

309
225
289
303
265
301
268
288
163
299
268
254
650
200
324
287
287
656
271
269
268
225
153
394
402
284
282
402
290
402
248
312
265
152
147
296
394
388
286
656
274
385
656
239
299
401

401
133
258
258
260
312
320
382
640
656
274
264
289
296
280
277
146
149
290
283
289
300
155
301
387
304
260
265
260
283
184
184
293
640
184
270
266
184
301
303
290
305
303
294
290
420

662
640
320
317
396
386
247
260
283
261
396
260
137
305
300
304
270
274
144
160
301
319
305
304
294
301
274
157
144
304
267
278
304
442
269
295
157
297
304
304
304
267
276
178
177
155

178
155
177
657
228
216
392
214
392
409
228
606
281
244
267
277
263
281
273
267
236
263
281
266
244
252
271
264
276
266
226
263
281
276
282
280
270
252
266
266
320
317
318
301
290
331

375
396
291
304
300
317
294
286
297
306
304
313
407
391
291
294
302
301
378
317
340
318
290
301
301
331
295
320
293
299
294
322
396
291
302
294
644
317
331
317
294
293
345
294
318
304

396
294
657
326
311
293
317
294
294
312
298
381
396
639
296
311
320
311
338
300
313
317
301
318
396
313
294
605
410
410
606
410
662
410
639
410
404
614
424
404
410
277
258
283
294
296

367
333
308
293
255
330
301
346
291
298
293
299
320
304
315
244
242
308
307
302
302
204
333
302
318
309
314
270
237
252
331
244
238
242
296
249
252
251
238
235
262
265
321
277
382
406

405
385
315
268
268
288
265
268
266
283
274
269
266
293
274
260
245
254
310
283
264
157
283
344
290
264
305
309
299
337
294
148
251
388
256
269
257
267
288
290
299
317
345
337
301
299

347
342
397
293
257
282
276
251
227
362
295
388
386
445
296
256
256
257
252
287
269
371
266
367
218
218
301
271
305
265
262
267
288
268
315
225
325
266
266
252
293
293
279
254
259
250

291
291
244
266
428
442
244
252
252
300
279
279
304
304
250
387
303
387
253
277
239
273
184
420
287
295
301
245
274
283
388
163
301
301
275
267
267
282
248
297
262
171
260
394
286
225

339
365
296
258
269
394
404
402
268
288
371
150
396
327
252
184
270
184
367
640
299
601
301
296
309
351
657
212
258
640
255
391
157
153
304
305
269
297
287
144
300
274
268
651
219
639

| |
|---|
| 404 |
| 281 |
| 303 |
| 276 |
| 252 |
| 267 |
| 298 |
| 266 |
| 281 |
| 303 |
| 267 |
| 266 |

[illegible]). Novel genes <90% amino aci[illegible]

| Closest blasthit |
| --- |
| AMP48593.1 CARB-PSE [uncultured bacterium] |
| WP_034033273.1 AAC(3)-I family aminoglycoside N-acetyltransferase [Pseudomonas aeruginosa] |
| WP_010792467.1 MULTISPECIES: aminoglycoside N-acetyltransferase AAC(3)-Ib [Gammaproteobacteria] |
| EGQ3237364.1 aminoglycoside N-acetyltransferase AAC(6')-Ie [Staphylococcus pseudintermedius] |
| WP_117833108.1 MULTISPECIES: GNAT family N-acetyltransferase [Roseburia] |
| WP_153568177.1 AAC(6')-Ighjkrstuvwx family aminoglycoside N-acetyltransferase [Acinetobacter haemolyticu |
| HBA02228.1 GNAT family N-acetyltransferase [Ruminococcus sp.] |
| WP_240081656.1 aminoglycoside O-phosphotransferase APH(2")-Ia, partial [Enterococcus faecium] |
| WP_001096887.1 MULTISPECIES: aminoglycoside O-phosphotransferase APH(3')-IIIa [Bacteria] |
| WP_000422633.1 MULTISPECIES: aminoglycoside O-phosphotransferase APH(3')-VIb [Proteobacteria] |
| WP_055068576.1 MULTISPECIES: aminoglycoside 3'-phosphotransferase [Roseburia] |
| CAD2016259.1 Aminoglycoside 3'-phosphotransferase, partial [Enterobacter cloacae] |
| QIE40729.1 APH(6) family putative aminoglycoside O-phosphotransferase [Rhodobacteraceae bacterium SC52] |
| WP_028030786.1 MULTISPECIES: RCP family class A beta-lactamase [Rhodobacteraceae] |
| WP_254015947.1 class A beta-lactamase, subclass A2 [Bacteroides ovatus] |
| BBQ55396.1 class A beta-lactamase [Aeromonas veronii] |
| AOA60293.1 class A beta-lactamase [uncultured bacterium] |
| WP_001100753.1 MULTISPECIES: inhibitor-resistant extended-spectrum class A beta-lactamase PER-1 [Gamm |
| WP_015589625.1 MULTISPECIES: RTG family carbenicillin-hydrolyzing class A beta-lactamase CARB-16 [Pr |
| WP_227806170.1 class A beta-lactamase, subclass A2 [Parabacteroides distasonis] |
| WP_000027057.1 MULTISPECIES: broad-spectrum class A beta-lactamase TEM-1 [Bacteria] |
| WP_004339683.1 MULTISPECIES: CfxA family broad-spectrum class A beta-lactamase [Bacteroidota] |
| WP_004327564.1 class A beta-lactamase, subclass A2 [Alistipes putredinis] |
| MCH3945785.1 MBL fold metallo-hydrolase [Lachnospiraceae bacterium] |
| QLO40532.1 beta-lactamase [Klebsiella sp. RHBSTW-00484] |
| WP_042649345.1 MULTISPECIES: CMY-1/MOX family class C beta-lactamase MOX-9 [Gammaproteobacteri |
| WP_086374409.1 MULTISPECIES: MCA family class C beta-lactamase [Acinetobacter] |
| AMP48538.1 classD [uncultured bacterium] |
| MBP8205294.1 class D beta-lactamase [Giesbergeria sp.] |
| WP_000586782.1 MULTISPECIES: oxacillin-hydrolyzing class D beta-lactamase OXA-20 [Gammaproteobacter |
| AMP47933.1 classD, partial [uncultured bacterium] |
| WP_004295324.1 MULTISPECIES: class D beta-lactamase OXA-347 [Bacteria] |
| WP_000846390.1 MULTISPECIES: oxacillin-hydrolyzing class D beta-lactamase OXA-10 [Proteobacteria] |
| WP_120987132.1 class D beta-lactamase [Aliarcobacter cryaerophilus] |
| CDI28150.1 carbapenem-hydrolyzing oxacillinase, partial [Acinetobacter pittii 42F] |
| WP_012754353.1 MULTISPECIES: OXA-24 family carbapenem-hydrolyzing class D beta-lactamase OXA-24 [ |
| WP_219869572.1 rRNA adenine N(6)-methyltransferase family protein [Microbacterium sp. PAMC22086] |
| WP_001038790.1 MULTISPECIES: 23S rRNA (adenine(2058)-N(6))-methyltransferase Erm(B) [Bacteria] |
| WP_005423882.1 MULTISPECIES: 16S rRNA (adenine(1518)-N(6)/adenine(1519)-N(6))-dimethyltransferase R |
| WP_005390949.1 MULTISPECIES: 23S ribosomal RNA methyltransferase Erm [Actinomycetia] |
| WP_002682030.1 MULTISPECIES: 23S rRNA (adenine(2058)-N(6))-methyltransferase Erm(F) [Bacteria] |
| WP_118308821.1 23S ribosomal RNA methyltransferase Erm [Bacteroides eggerthii] |
| WP_000155092.1 MULTISPECIES: Mph(E) family macrolide 2'-phosphotransferase [Proteobacteria] |

MBP7262500.1 Mph(E)/Mph(G) family macrolide 2'-phosphotransferase [Bacteroidia bacterium]
WP_038557444.1 MULTISPECIES: Mph(E)/Mph(G) family macrolide 2'-phosphotransferase [Marinilabiliales]
WP_078213155.1 MULTISPECIES: Mph(E)/Mph(G) family macrolide 2'-phosphotransferase [Flavobacteriales]
AGP03376.1 AAC(3)-II, partial [Klebsiella pneumoniae]
WP_012537714.1 MULTISPECIES: quinolone resistance pentapeptide repeat protein QnrS2 [Gammaproteobacte
WP_000106218.1 tetracycline efflux MFS transporter Tet(A) [Klebsiella pneumoniae]
WP_000841446.1 MULTISPECIES: tetracycline efflux MFS transporter Tet(C) [Bacteria]
WP_005783159.1 MULTISPECIES: tetracycline-inactivating monooxygenase Tet(X) [Bacteria]
TXI98365.1 MAG: tetracycline destructase [Neisseriales bacterium]
ABB97394.1 tetracycline resistance protein [Enterococcus faecium]
WP_107208979.1 tetracycline resistance ribosomal protection protein Tet(W) [Streptococcus suis]
HBO8156621.1 AAC(3)-I family aminoglycoside N-acetyltransferase [Pseudomonas aeruginosa]
EJN7178204.1 GNAT family N-acetyltransferase [Citrobacter amalonaticus]
AGP03376.1 AAC(3)-II, partial [Klebsiella pneumoniae]
WP_110165650.1 MULTISPECIES: aminoglycoside N-acetyltransferase AAC(6')-Ie, partial [Bacilli]
WP_195287176.1 GNAT family N-acetyltransferase [Roseburia faecis]
WP_117833108.1 MULTISPECIES: GNAT family N-acetyltransferase [Roseburia]
HBA02228.1 GNAT family N-acetyltransferase [Ruminococcus sp.]
CUP37732.1 Acetyltransferase (GNAT) family [[Eubacterium] rectale]
WP_227248514.1 GNAT family N-acetyltransferase [Roseburia faecis]
WP_010714603.1 bifunctional aminoglycoside N-acetyltransferase AAC(6')-Ie/aminoglycoside O-phosphotransf
WP_002592615.1 MULTISPECIES: aminoglycoside O-phosphotransferase APH(2")-IIa [Lachnospiraceae]
CAD2016259.1 Aminoglycoside 3'-phosphotransferase, partial [Enterobacter cloacae]
WP_001096887.1 MULTISPECIES: aminoglycoside O-phosphotransferase APH(3')-IIIa [Bacteria]
WP_055068576.1 MULTISPECIES: aminoglycoside 3'-phosphotransferase [Roseburia]
QIE40729.1 APH(6) family putative aminoglycoside O-phosphotransferase [Rhodobacteraceae bacterium SC52]
MBP8038075.1 class A beta-lactamase [Prevotella sp.]
WP_004339683.1 MULTISPECIES: CfxA family broad-spectrum class A beta-lactamase [Bacteroidota]
WP_055170259.1 class A beta-lactamase, subclass A2 [Bacteroides caccae]
WP_086989205.1 serine hydrolase [Trichococcus flocculiformis]
WP_008861554.1 class A beta-lactamase [Barnesiella intestinihominis]
ACT97465.1 HGD-1 beta-lactamase [uncultured organism]
WP_006555379.1 MULTISPECIES: extended-spectrum class A beta-lactamase ACI-1 [Negativicutes]
BBQ55396.1 class A beta-lactamase [Aeromonas veronii]
WP_046451287.1 class A beta-lactamase, subclass A2 [Odoribacter splanchnicus]
WP_028030786.1 MULTISPECIES: RCP family class A beta-lactamase [Rhodobacteraceae]
MBP8721109.1 class A beta-lactamase, subclass A2 [Tidjanibacter sp.]
MBP8652476.1 class A beta-lactamase, subclass A2 [Alistipes sp.]
MCB1465501.1 class A beta-lactamase [Nitratireductor sp.]
MCH3945785.1 MBL fold metallo-hydrolase [Lachnospiraceae bacterium]
WP_227588532.1 MULTISPECIES: MBL fold metallo-hydrolase [Blautia]
HCJ59310.1 MBL fold metallo-hydrolase [Faecalibacterium sp.]
WP_086374409.1 MULTISPECIES: MCA family class C beta-lactamase [Acinetobacter]
ULG20519.1 Beta-lactamase [Acinetobacter nosocomialis]
WP_004295324.1 MULTISPECIES: class D beta-lactamase OXA-347 [Bacteria]
WP_000586782.1 MULTISPECIES: oxacillin-hydrolyzing class D beta-lactamase OXA-20 [Gammaproteobacter

MBP8205294.1 class D beta-lactamase [Giesbergeria sp.]
AMP47200.1 classD [uncultured bacterium]
QKE29104.1 putative class D beta-lactamase [Arcobacter acticola]
AMP48538.1 classD [uncultured bacterium]
OQA98182.1 Beta-lactamase OXA-2 precursor [Bacteroidetes bacterium ADurb.Bin217]
WP_000846390.1 MULTISPECIES: oxacillin-hydrolyzing class D beta-lactamase OXA-10 [Proteobacteria]
WP_120987132.1 class D beta-lactamase [Aliarcobacter cryaerophilus]
CDI28150.1 carbapenem-hydrolyzing oxacillinase, partial [Acinetobacter pittii 42F]
AMP47933.1 classD, partial [uncultured bacterium]
WP_227035317.1 23S ribosomal RNA methyltransferase Erm [Bacteroides faecis]
MCK6541170.1 23S ribosomal RNA methyltransferase Erm [bacterium]
WP_219869572.1 rRNA adenine N(6)-methyltransferase family protein [Microbacterium sp. PAMC22086]
WP_008021360.1 MULTISPECIES: 23S ribosomal RNA methyltransferase Erm [Bacteroidales]
WP_002682030.1 MULTISPECIES: 23S rRNA (adenine(2058)-N(6))-methyltransferase Erm(F) [Bacteria]
WP_001038790.1 MULTISPECIES: 23S rRNA (adenine(2058)-N(6))-methyltransferase Erm(B) [Bacteria]
WP_005423882.1 MULTISPECIES: 16S rRNA (adenine(1518)-N(6)/adenine(1519)-N(6))-dimethyltransferase R
MBO5349701.1 rRNA adenine N(6)-methyltransferase family protein [Clostridia bacterium]
WP_036880322.1 MULTISPECIES: 23S ribosomal RNA methyltransferase Erm [Bacteroidales]
WP_010539471.1 MULTISPECIES: 23S ribosomal RNA methyltransferase Erm [Bacteroidales]
WP_000155092.1 MULTISPECIES: Mph(E) family macrolide 2'-phosphotransferase [Proteobacteria]
WP_038557444.1 MULTISPECIES: Mph(E)/Mph(G) family macrolide 2'-phosphotransferase [Marinilabiliales]
MBP7262500.1 Mph(E)/Mph(G) family macrolide 2'-phosphotransferase [Bacteroidia bacterium]
WP_078213155.1 MULTISPECIES: Mph(E)/Mph(G) family macrolide 2'-phosphotransferase [Flavobacteriales]
WP_012537714.1 MULTISPECIES: quinolone resistance pentapeptide repeat protein QnrS2 [Gammaproteobacte
WP_012772744.1 QnrB family quinolone resistance pentapeptide repeat protein [Escherichia coli]
WP_000841446.1 MULTISPECIES: tetracycline efflux MFS transporter Tet(C) [Bacteria]
WP_000106218.1 tetracycline efflux MFS transporter Tet(A) [Klebsiella pneumoniae]
TXI98365.1 MAG: tetracycline destructase [Neisseriales bacterium]
WP_005783159.1 MULTISPECIES: tetracycline-inactivating monooxygenase Tet(X) [Bacteria]
WP_005790841.1 MULTISPECIES: NAD(P)/FAD-dependent oxidoreductase [Bacteroidales]
APO28173.1 ferrous iron transport protein B [uncultured bacterium]
ERI78723.1 putative translation elongation factor G [Clostridium sp. ATCC BAA-442]
ABB97394.1 tetracycline resistance protein [Enterococcus faecium]
APO27487.1 ferrous iron transport protein B [uncultured bacterium]
WP_113842634.1 MULTISPECIES: tetracycline resistance ribosomal protection protein [Firmicutes]
WP_088656287.1 MULTISPECIES: tetracycline resistance ribosomal protection protein Tet(36) [Bacteroidota]
WP_034033273.1 AAC(3)-I family aminoglycoside N-acetyltransferase [Pseudomonas aeruginosa]
HBO8156621.1 AAC(3)-I family aminoglycoside N-acetyltransferase [Pseudomonas aeruginosa]
AGP03376.1 AAC(3)-II, partial [Klebsiella pneumoniae]
EGO9274095.1 aminoglycoside N-acetyltransferase AAC(6')-Ie [Enterococcus faecalis]
CAB5652345.1 Bifunctional AAC/APH [Pseudomonas putida]
WP_117833108.1 MULTISPECIES: GNAT family N-acetyltransferase [Roseburia]
WP_153568177.1 AAC(6')-Ighjkrstuvwx family aminoglycoside N-acetyltransferase [Acinetobacter haemolyticu
WP_227248514.1 GNAT family N-acetyltransferase [Roseburia faecis]
HBA02228.1 GNAT family N-acetyltransferase [Ruminococcus sp.]
WP_128470137.1 MULTISPECIES: GNAT family N-acetyltransferase [Terrabacteria group]

WP_240081656.1 aminoglycoside O-phosphotransferase APH(2")-Ia, partial [Enterococcus faecium]
WP_010708503.1 MULTISPECIES: aminoglycoside O-phosphotransferase APH(2")-IIa [Bacteria]
WP_001096887.1 MULTISPECIES: aminoglycoside O-phosphotransferase APH(3')-IIIa [Bacteria]
WP_000422633.1 MULTISPECIES: aminoglycoside O-phosphotransferase APH(3')-VIb [Proteobacteria]
WP_055068576.1 MULTISPECIES: aminoglycoside 3'-phosphotransferase [Roseburia]
CAD2016259.1 Aminoglycoside 3'-phosphotransferase, partial [Enterobacter cloacae]
QIE40729.1 APH(6) family putative aminoglycoside O-phosphotransferase [Rhodobacteraceae bacterium SC52]
WP_001100753.1 MULTISPECIES: inhibitor-resistant extended-spectrum class A beta-lactamase PER-1 [Gamm
AMP51956.1 CblA [uncultured bacterium]
AOA60293.1 class A beta-lactamase [uncultured bacterium]
WP_028030786.1 MULTISPECIES: RCP family class A beta-lactamase [Rhodobacteraceae]
WP_149947951.1 class A beta-lactamase, subclass A2 [Bacteroides cellulosilyticus]
WP_086989205.1 serine hydrolase [Trichococcus flocculiformis]
WP_055256788.1 class A beta-lactamase, subclass A2 [Bacteroides caccae]
WP_063857830.1 MULTISPECIES: extended-spectrum class A beta-lactamase BEL-1 [Gammaproteobacteria]
WP_032567550.1 CepA family extended-spectrum class A beta-lactamase [Bacteroides fragilis]
WP_004339683.1 MULTISPECIES: CfxA family broad-spectrum class A beta-lactamase [Bacteroidota]
WP_015589625.1 MULTISPECIES: RTG family carbenicillin-hydrolyzing class A beta-lactamase CARB-16 [Pr
MCC6940111.1 serine hydrolase [Novosphingobium sp.]
WP_046403513.1 class A beta-lactamase, subclass A2 [Odoribacter splanchnicus]
WP_141418823.1 class A beta-lactamase [Alistipes communis]
MBP8038075.1 class A beta-lactamase [Prevotella sp.]
TAH91353.1 SHV family class A beta-lactamase [Klebsiella pneumoniae]
WP_000027057.1 MULTISPECIES: broad-spectrum class A beta-lactamase TEM-1 [Bacteria]
BBQ55396.1 class A beta-lactamase [Aeromonas veronii]
WP_006555379.1 MULTISPECIES: extended-spectrum class A beta-lactamase ACI-1 [Negativicutes]
MBS1939435.1 class A beta-lactamase, subclass A2 [Bacteroidetes bacterium]
WP_227588532.1 MULTISPECIES: MBL fold metallo-hydrolase [Blautia]
MCH3945785.1 MBL fold metallo-hydrolase [Lachnospiraceae bacterium]
QLO40532.1 beta-lactamase [Klebsiella sp. RHBSTW-00484]
WP_042649345.1 MULTISPECIES: CMY-1/MOX family class C beta-lactamase MOX-9 [Gammaproteobacteri
EIU1367781.1 class D beta-lactamase [Pseudomonas aeruginosa]
EIU5375759.1 OXA-10 family class D beta-lactamase [Pseudomonas aeruginosa]
WP_011013281.1 MULTISPECIES: oxacillin-hydrolyzing class D beta-lactamase NPS-1 [Proteobacteria]
WP_120987132.1 class D beta-lactamase [Aliarcobacter cryaerophilus]
WP_000586782.1 MULTISPECIES: oxacillin-hydrolyzing class D beta-lactamase OXA-20 [Gammaproteobacter
PKQ64175.1 class D beta-lactamase [Labilibaculum filiforme]
WP_004295324.1 MULTISPECIES: class D beta-lactamase OXA-347 [Bacteria]
AMP48538.1 classD [uncultured bacterium]
CDI28150.1 carbapenem-hydrolyzing oxacillinase, partial [Acinetobacter pittii 42F]
AMP47933.1 classD, partial [uncultured bacterium]
WP_012754353.1 MULTISPECIES: OXA-24 family carbapenem-hydrolyzing class D beta-lactamase OXA-24 [
MBP8205294.1 class D beta-lactamase [Giesbergeria sp.]
WP_002682030.1 MULTISPECIES: 23S rRNA (adenine(2058)-N(6))-methyltransferase Erm(F) [Bacteria]
WP_001038790.1 MULTISPECIES: 23S rRNA (adenine(2058)-N(6))-methyltransferase Erm(B) [Bacteria]
WP_219869572.1 rRNA adenine N(6)-methyltransferase family protein [Microbacterium sp. PAMC22086]

WP_118308821.1 23S ribosomal RNA methyltransferase Erm [Bacteroides eggerthii]
WP_182048909.1 MULTISPECIES: 23S ribosomal RNA methyltransferase Erm [Actinomycetia]
WP_005423882.1 MULTISPECIES: 16S rRNA (adenine(1518)-N(6)/adenine(1519)-N(6))-dimethyltransferase R
WP_130108831.1 MULTISPECIES: 23S ribosomal RNA methyltransferase Erm [Actinomycetia]
WP_036880322.1 MULTISPECIES: 23S ribosomal RNA methyltransferase Erm [Bacteroidales]
WP_007286427.1 MULTISPECIES: 23S rRNA (adenine(2058)-N(6))-methyltransferase Erm(Q) [Eubacteriales]
EJA2650198.1 23S ribosomal RNA methyltransferase Erm [Pseudomonas aeruginosa]
MBO5349701.1 rRNA adenine N(6)-methyltransferase family protein [Clostridia bacterium]
WP_000155092.1 MULTISPECIES: Mph(E) family macrolide 2'-phosphotransferase [Proteobacteria]
WP_038557444.1 MULTISPECIES: Mph(E)/Mph(G) family macrolide 2'-phosphotransferase [Marinilabiliales]
WP_000219391.1 MULTISPECIES: Mph(A) family macrolide 2'-phosphotransferase [Bacteria]
WP_012537714.1 MULTISPECIES: quinolone resistance pentapeptide repeat protein QnrS2 [Gammaproteobacte
WP_000106218.1 tetracycline efflux MFS transporter Tet(A) [Klebsiella pneumoniae]
WP_000841446.1 MULTISPECIES: tetracycline efflux MFS transporter Tet(C) [Bacteria]
WP_017411290.1 MULTISPECIES: tetracycline efflux MFS transporter Tet(E) [Gammaproteobacteria]
WP_005783159.1 MULTISPECIES: tetracycline-inactivating monooxygenase Tet(X) [Bacteria]
TXI98365.1 MAG: tetracycline destructase [Neisseriales bacterium]
MBQ9662135.1 tetracycline resistance ribosomal protection protein Tet(W) [Oscillospiraceae bacterium]
ABB97394.1 tetracycline resistance protein [Enterococcus faecium]
ERI78723.1 putative translation elongation factor G [Clostridium sp. ATCC BAA-442]
WP_006440995.1 MULTISPECIES: tetracycline resistance ribosomal protection protein TetB(P) [Firmicutes]
HBO8156621.1 AAC(3)-I family aminoglycoside N-acetyltransferase [Pseudomonas aeruginosa]
KJM97364.1 gentamicin 3'-acetyltransferase, partial [Enterobacter hormaechei subsp. xiangfangensis]
AGP03376.1 AAC(3)-II, partial [Klebsiella pneumoniae]
HCC0854438.1 aminoglycoside N-acetyltransferase AAC(6')-Ie [Enterococcus faecium]
WP_117833108.1 MULTISPECIES: GNAT family N-acetyltransferase [Roseburia]
HBA02228.1 GNAT family N-acetyltransferase [Ruminococcus sp.]
WP_227248514.1 GNAT family N-acetyltransferase [Roseburia faecis]
WP_153568177.1 AAC(6')-Ighjkrstuvwx family aminoglycoside N-acetyltransferase [Acinetobacter haemolyticu
4ORK_A Crystal Structure of the Phosphotransferase Domain of the Bifunctional Aminoglycoside Resistance En
CAD2016259.1 Aminoglycoside 3'-phosphotransferase, partial [Enterobacter cloacae]
WP_001096887.1 MULTISPECIES: aminoglycoside O-phosphotransferase APH(3')-IIIa [Bacteria]
QIE40729.1 APH(6) family putative aminoglycoside O-phosphotransferase [Rhodobacteraceae bacterium SC52]
ACT97465.1 HGD-1 beta-lactamase [uncultured organism]
WP_004339683.1 MULTISPECIES: CfxA family broad-spectrum class A beta-lactamase [Bacteroidota]
WP_166159251.1 class A beta-lactamase [Acidovorax sp. HDW3]
BBQ55396.1 class A beta-lactamase [Aeromonas veronii]
WP_028030786.1 MULTISPECIES: RCP family class A beta-lactamase [Rhodobacteraceae]
WP_004320670.1 MULTISPECIES: class A beta-lactamase, subclass A2 [Bacteroides]
MBP8038075.1 class A beta-lactamase [Prevotella sp.]
MCC6940111.1 serine hydrolase [Novosphingobium sp.]
MCB1465501.1 class A beta-lactamase [Nitratireductor sp.]
MBP8779047.1 class A beta-lactamase [Alicycliphilus sp.]
WP_004327564.1 class A beta-lactamase, subclass A2 [Alistipes putredinis]
MCH3945785.1 MBL fold metallo-hydrolase [Lachnospiraceae bacterium]
WP_086374409.1 MULTISPECIES: MCA family class C beta-lactamase [Acinetobacter]

WP_032491311.1 MULTISPECIES: OXA-1 family oxacillin-hydrolyzing class D beta-lactamase OXA-4 [Proteo
EIU5375759.1 OXA-10 family class D beta-lactamase [Pseudomonas aeruginosa]
WP_004295324.1 MULTISPECIES: class D beta-lactamase OXA-347 [Bacteria]
MBP8205294.1 class D beta-lactamase [Giesbergeria sp.]
WP_000085217.1 class D beta-lactamase [Acinetobacter baumannii]
WP_000586782.1 MULTISPECIES: oxacillin-hydrolyzing class D beta-lactamase OXA-20 [Gammaproteobacter
AMP47933.1 classD, partial [uncultured bacterium]
WP_032490445.1 OXA-46 family oxacillin-hydrolyzing class D beta-lactamase OXA-119 [Pseudomonas aerugi
WP_120987132.1 class D beta-lactamase [Aliarcobacter cryaerophilus]
AMP48538.1 classD [uncultured bacterium]
WP_001038790.1 MULTISPECIES: 23S rRNA (adenine(2058)-N(6))-methyltransferase Erm(B) [Bacteria]
WP_118308821.1 23S ribosomal RNA methyltransferase Erm [Bacteroides eggerthii]
WP_005423882.1 MULTISPECIES: 16S rRNA (adenine(1518)-N(6)/adenine(1519)-N(6))-dimethyltransferase R
WP_036880322.1 MULTISPECIES: 23S ribosomal RNA methyltransferase Erm [Bacteroidales]
WP_002682030.1 MULTISPECIES: 23S rRNA (adenine(2058)-N(6))-methyltransferase Erm(F) [Bacteria]
MBR2247486.1 rRNA adenine N(6)-methyltransferase family protein [Bacilli bacterium]
WP_000155092.1 MULTISPECIES: Mph(E) family macrolide 2'-phosphotransferase [Proteobacteria]
WP_038557444.1 MULTISPECIES: Mph(E)/Mph(G) family macrolide 2'-phosphotransferase [Marinilabiliales]
MBP6408896.1 Mph(B) family macrolide 2'-phosphotransferase [Fusobacteriaceae bacterium]
WP_000841446.1 MULTISPECIES: tetracycline efflux MFS transporter Tet(C) [Bacteria]
WP_000106218.1 tetracycline efflux MFS transporter Tet(A) [Klebsiella pneumoniae]
TXI98365.1 MAG: tetracycline destructase [Neisseriales bacterium]
WP_005783159.1 MULTISPECIES: tetracycline-inactivating monooxygenase Tet(X) [Bacteria]
ABB97394.1 tetracycline resistance protein [Enterococcus faecium]
WP_138721012.1 tetracycline resistance ribosomal protection protein Tet(W) [Eubacterium maltosivorans]
HBO8156621.1 AAC(3)-I family aminoglycoside N-acetyltransferase [Pseudomonas aeruginosa]
WP_010792467.1 MULTISPECIES: aminoglycoside N-acetyltransferase AAC(3)-Ib [Gammaproteobacteria]
AGP03376.1 AAC(3)-II, partial [Klebsiella pneumoniae]
KZM11363.1 aminoglycoside adenylyltransferase [Pseudomonas aeruginosa]
WP_065217783.1 GNAT family N-acetyltransferase [Clostridioides difficile]
WP_153568177.1 AAC(6')-Ighjkrstuvwx family aminoglycoside N-acetyltransferase [Acinetobacter haemolyticu
WP_117833108.1 MULTISPECIES: GNAT family N-acetyltransferase [Roseburia]
HBA02228.1 GNAT family N-acetyltransferase [Ruminococcus sp.]
WP_262313883.1 GNAT family N-acetyltransferase [Klebsiella quasipneumoniae]
WP_002289795.1 MULTISPECIES: aminoglycoside N-acetyltransferase AAC(6')-Ii [Enterococcus]
WP_240081656.1 aminoglycoside O-phosphotransferase APH(2")-Ia, partial [Enterococcus faecium]
CAD2016259.1 Aminoglycoside 3'-phosphotransferase, partial [Enterobacter cloacae]
WP_055068576.1 MULTISPECIES: aminoglycoside 3'-phosphotransferase [Roseburia]
WP_000422633.1 MULTISPECIES: aminoglycoside O-phosphotransferase APH(3')-VIb [Proteobacteria]
WP_000018326.1 MULTISPECIES: aminoglycoside O-phosphotransferase APH(3')-Ia [Bacteria]
WP_001096887.1 MULTISPECIES: aminoglycoside O-phosphotransferase APH(3')-IIIa [Bacteria]
WP_118435663.1 class A beta-lactamase, subclass A2 [Bacteroides cellulosilyticus]
AMP53257.1 CfxA [uncultured bacterium]
WP_000027057.1 MULTISPECIES: broad-spectrum class A beta-lactamase TEM-1 [Bacteria]
WP_120079028.1 class A beta-lactamase, subclass A2 [Bacteroides sp. OF03-11BH]
QCO89460.1 BlaCARB-2 [Escherichia coli]

WP_004339683.1 MULTISPECIES: CfxA family broad-spectrum class A beta-lactamase [Bacteroidota]
WP_046451287.1 class A beta-lactamase, subclass A2 [Odoribacter splanchnicus]
CDA71336.1 putative uncharacterized protein [Bacteroides coprocola CAG:162]
WP_151720985.1 RCP family class A beta-lactamase [Gemmobacter serpentinus]
WP_021892713.1 class A beta-lactamase [Barnesiella intestinihominis]
WP_001100753.1 MULTISPECIES: inhibitor-resistant extended-spectrum class A beta-lactamase PER-1 [Gamm
WP_143270028.1 class A beta-lactamase, subclass A2 [Bacteroides clarus]
MBP7019807.1 class A beta-lactamase, subclass A2 [Alistipes sp.]
WP_227806170.1 class A beta-lactamase, subclass A2 [Parabacteroides distasonis]
WP_063860581.1 MULTISPECIES: subclass B1 metallo-beta-lactamase IMP-22 [Gammaproteobacteria]
WP_003108247.1 MULTISPECIES: subclass B1 metallo-beta-lactamase VIM-2 [Proteobacteria]
WP_198260390.1 HARLDQ motif MBL-fold protein [Acinetobacter bereziniae]
WP_042649345.1 MULTISPECIES: CMY-1/MOX family class C beta-lactamase MOX-9 [Gammaproteobacteri
WP_086374409.1 MULTISPECIES: MCA family class C beta-lactamase [Acinetobacter]
QLO40532.1 beta-lactamase [Klebsiella sp. RHBSTW-00484]
CAE6180605.1 Beta-lactamase OXA-1, partial [Escherichia coli]
WP_161507793.1 MULTISPECIES: OXA-427 family carbapenem-hydrolyzing class D beta-lactamase OXA-91
AAD22145.1 oxacillinase [Klebsiella aerogenes]
OQA98182.1 Beta-lactamase OXA-2 precursor [Bacteroidetes bacterium ADurb.Bin217]
CDI28150.1 carbapenem-hydrolyzing oxacillinase, partial [Acinetobacter pittii 42F]
AMP47933.1 classD, partial [uncultured bacterium]
WP_012754353.1 MULTISPECIES: OXA-24 family carbapenem-hydrolyzing class D beta-lactamase OXA-24 [
WP_262087276.1 OXA-274 family carbapenem-hydrolyzing class D beta-lactamase [Acinetobacter sp. I-MWF]
WP_004295324.1 MULTISPECIES: class D beta-lactamase OXA-347 [Bacteria]
WP_001038790.1 MULTISPECIES: 23S rRNA (adenine(2058)-N(6))-methyltransferase Erm(B) [Bacteria]
WP_182048909.1 MULTISPECIES: 23S ribosomal RNA methyltransferase Erm [Actinomycetia]
WP_008021360.1 MULTISPECIES: 23S ribosomal RNA methyltransferase Erm [Bacteroidales]
MCK6541170.1 23S ribosomal RNA methyltransferase Erm [bacterium]
WP_010539471.1 MULTISPECIES: 23S ribosomal RNA methyltransferase Erm [Bacteroidales]
MBO5349701.1 rRNA adenine N(6)-methyltransferase family protein [Clostridia bacterium]
WP_000155092.1 MULTISPECIES: Mph(E) family macrolide 2'-phosphotransferase [Proteobacteria]
WP_000219391.1 MULTISPECIES: Mph(A) family macrolide 2'-phosphotransferase [Bacteria]
WP_014386803.1 MULTISPECIES: Mph(G) family macrolide 2'-phosphotransferase [Proteobacteria]
TXI65181.1 MAG: Mph(E)/Mph(G) family macrolide 2'-phosphotransferase [Flavobacterium sp.]
MBP7262500.1 Mph(E)/Mph(G) family macrolide 2'-phosphotransferase [Bacteroidia bacterium]
WP_012537714.1 MULTISPECIES: quinolone resistance pentapeptide repeat protein QnrS2 [Gammaproteobacte
WP_000841446.1 MULTISPECIES: tetracycline efflux MFS transporter Tet(C) [Bacteria]
WP_011899270.1 MULTISPECIES: tetracycline efflux MFS transporter Tet(E) [Gammaproteobacteria]
EMG61192.1 tetracycline efflux protein [Salmonella enterica subsp. enterica serovar Newport str. Henan_3]
WP_008651082.1 MULTISPECIES: tetracycline-inactivating monooxygenase Tet(X2) [Bacteria]
WP_107208979.1 tetracycline resistance ribosomal protection protein Tet(W) [Streptococcus suis]
HBO8156621.1 AAC(3)-I family aminoglycoside N-acetyltransferase [Pseudomonas aeruginosa]
WP_010792467.1 MULTISPECIES: aminoglycoside N-acetyltransferase AAC(3)-Ib [Gammaproteobacteria]
AGP03376.1 AAC(3)-II, partial [Klebsiella pneumoniae]
WP_195287176.1 GNAT family N-acetyltransferase [Roseburia faecis]
KZM11363.1 aminoglycoside adenylyltransferase [Pseudomonas aeruginosa]

WP_262313883.1 GNAT family N-acetyltransferase [Klebsiella quasipneumoniae]
EAC1778952.1 GNAT family N-acetyltransferase [Campylobacter coli]
WP_153568177.1 AAC(6')-Ighjkrstuvwx family aminoglycoside N-acetyltransferase [Acinetobacter haemolyticu
WP_065217783.1 GNAT family N-acetyltransferase [Clostridioides difficile]
WP_117833108.1 MULTISPECIES: GNAT family N-acetyltransferase [Roseburia]
WP_005202238.1 MULTISPECIES: AAC(6')-Ighjkrstuvwx family aminoglycoside N-acetyltransferase [Acineto
WP_002289795.1 MULTISPECIES: aminoglycoside N-acetyltransferase AAC(6')-Ii [Enterococcus]
WP_240081656.1 aminoglycoside O-phosphotransferase APH(2")-Ia, partial [Enterococcus faecium]
WP_001096887.1 MULTISPECIES: aminoglycoside O-phosphotransferase APH(3')-IIIa [Bacteria]
WP_000018326.1 MULTISPECIES: aminoglycoside O-phosphotransferase APH(3')-Ia [Bacteria]
WP_055068576.1 MULTISPECIES: aminoglycoside 3'-phosphotransferase [Roseburia]
WP_012091421.1 MULTISPECIES: aminoglycoside 3'-phosphotransferase [Brucella]
CAD2016259.1 Aminoglycoside 3'-phosphotransferase, partial [Enterobacter cloacae]
WP_001100753.1 MULTISPECIES: inhibitor-resistant extended-spectrum class A beta-lactamase PER-1 [Gamm
WP_046451287.1 class A beta-lactamase, subclass A2 [Odoribacter splanchnicus]
WP_000027057.1 MULTISPECIES: broad-spectrum class A beta-lactamase TEM-1 [Bacteria]
AMP50979.1 CblA [uncultured bacterium]
pir|A49789| beta-lactamase (EC 3.5.2.6) PSE-1 - Pseudomonas aeruginosa plasmid RPL11 transposon Tn1403 [P
WP_149921626.1 class A beta-lactamase, subclass A2 [Bacteroides caccae]
BBQ55396.1 class A beta-lactamase [Aeromonas veronii]
WP_008861554.1 class A beta-lactamase [Barnesiella intestinihominis]
WP_227043252.1 class A beta-lactamase [Alistipes communis]
AYN80769.1 SHV family beta-lactamase, partial [uncultured bacterium]
WP_004339683.1 MULTISPECIES: CfxA family broad-spectrum class A beta-lactamase [Bacteroidota]
WP_002567442.1 MULTISPECIES: class A beta-lactamase [Eubacteriales]
WP_237043943.1 PAU family class A beta-lactamase [Pseudomonas aeruginosa]
WP_012658785.1 MULTISPECIES: carbapenem-hydrolyzing class A beta-lactamase GES-5 [Proteobacteria]
WP_028030786.1 MULTISPECIES: RCP family class A beta-lactamase [Rhodobacteraceae]
WP_032748514.1 extended-spectrum class A beta-lactamase OXY-1-8 [Klebsiella michiganensis]
WP_006555379.1 MULTISPECIES: extended-spectrum class A beta-lactamase ACI-1 [Negativicutes]
CDA17792.1 beta-lactamase [Acetobacter sp. CAG:267]
MBS1939435.1 class A beta-lactamase, subclass A2 [Bacteroidetes bacterium]
WP_035471713.1 MULTISPECIES: class A beta-lactamase, subclass A2 [Rikenellaceae]
WP_004320670.1 MULTISPECIES: class A beta-lactamase, subclass A2 [Bacteroides]
MBP8652476.1 class A beta-lactamase, subclass A2 [Alistipes sp.]
WP_003108247.1 MULTISPECIES: subclass B1 metallo-beta-lactamase VIM-2 [Proteobacteria]
WP_063860581.1 MULTISPECIES: subclass B1 metallo-beta-lactamase IMP-22 [Gammaproteobacteria]
WP_198260390.1 HARLDQ motif MBL-fold protein [Acinetobacter bereziniae]
MCH3945785.1 MBL fold metallo-hydrolase [Lachnospiraceae bacterium]
WP_227588532.1 MULTISPECIES: MBL fold metallo-hydrolase [Blautia]
WP_086374409.1 MULTISPECIES: MCA family class C beta-lactamase [Acinetobacter]
QLO40532.1 beta-lactamase [Klebsiella sp. RHBSTW-00484]
CAE6180605.1 Beta-lactamase OXA-1, partial [Escherichia coli]
WP_139736287.1 class D beta-lactamase [Aeromonas caviae]
EIU1367781.1 class D beta-lactamase [Pseudomonas aeruginosa]
AAD22145.1 oxacillinase [Klebsiella aerogenes]

MBP8205294.1 class D beta-lactamase [Giesbergeria sp.]
OQA98182.1 Beta-lactamase OXA-2 precursor [Bacteroidetes bacterium ADurb.Bin217]
PKQ64175.1 class D beta-lactamase [Labilibaculum filiforme]
WP_004295324.1 MULTISPECIES: class D beta-lactamase OXA-347 [Bacteria]
AMP47933.1 classD, partial [uncultured bacterium]
AMP48538.1 classD [uncultured bacterium]
HAT7527105.1 OXA-10 family class D beta-lactamase [Citrobacter koseri]
WP_012754353.1 MULTISPECIES: OXA-24 family carbapenem-hydrolyzing class D beta-lactamase OXA-24 [[illegible]
WP_262087276.1 OXA-274 family carbapenem-hydrolyzing class D beta-lactamase [Acinetobacter sp. I-MWF]
AMP47200.1 classD [uncultured bacterium]
MCK6541170.1 23S ribosomal RNA methyltransferase Erm [bacterium]
WP_001038790.1 MULTISPECIES: 23S rRNA (adenine(2058)-N(6))-methyltransferase Erm(B) [Bacteria]
WP_182048909.1 MULTISPECIES: 23S ribosomal RNA methyltransferase Erm [Actinomycetia]
WP_008021360.1 MULTISPECIES: 23S ribosomal RNA methyltransferase Erm [Bacteroidales]
WP_015431539.1 MULTISPECIES: 23S rRNA (adenine(2058)-N(6))-methyltransferase Erm(F) [Bacteria]
WP_002571018.1 MULTISPECIES: 23S rRNA (adenine(2058)-N(6))-methyltransferase Erm(52) [Bacteria]
WP_219869572.1 rRNA adenine N(6)-methyltransferase family protein [Microbacterium sp. PAMC22086]
EJA2650198.1 23S ribosomal RNA methyltransferase Erm [Pseudomonas aeruginosa]
WP_005423882.1 MULTISPECIES: 16S rRNA (adenine(1518)-N(6)/adenine(1519)-N(6))-dimethyltransferase [illegible]
WP_010539471.1 MULTISPECIES: 23S ribosomal RNA methyltransferase Erm [Bacteroidales]
WP_000219391.1 MULTISPECIES: Mph(A) family macrolide 2'-phosphotransferase [Bacteria]
MBP7262500.1 Mph(E)/Mph(G) family macrolide 2'-phosphotransferase [Bacteroidia bacterium]
WP_038557444.1 MULTISPECIES: Mph(E)/Mph(G) family macrolide 2'-phosphotransferase [Marinilabiliales]
WP_012537714.1 MULTISPECIES: quinolone resistance pentapeptide repeat protein QnrS2 [Gammaproteobact[illegible]
WP_012772744.1 QnrB family quinolone resistance pentapeptide repeat protein [Escherichia coli]
WP_000415714.1 MULTISPECIES: quinolone resistance pentapeptide repeat protein QnrVC1 [Gammaproteoba[illegible]
WP_011899270.1 MULTISPECIES: tetracycline efflux MFS transporter Tet(E) [Gammaproteobacteria]
WP_000841446.1 MULTISPECIES: tetracycline efflux MFS transporter Tet(C) [Bacteria]
EIW8706509.1 tetracycline efflux MFS transporter Tet(D) [Klebsiella pneumoniae]
WP_005783159.1 MULTISPECIES: tetracycline-inactivating monooxygenase Tet(X) [Bacteria]
WP_005790841.1 MULTISPECIES: NAD(P)/FAD-dependent oxidoreductase [Bacteroidales]
WP_088656287.1 MULTISPECIES: tetracycline resistance ribosomal protection protein Tet(36) [Bacteroidota]
ERI78723.1 putative translation elongation factor G [Clostridium sp. ATCC BAA-442]
WP_225511234.1 aminoglycoside O-phosphotransferase APH(2")-Ia [Staphylococcus aureus]
MBB6732384.1 GNAT family N-acetyltransferase [Cohnella zeiphila]
NOT94262.1 class A beta-lactamase, subclass A2 [Ferruginibacter sp.]
WP_118155791.1 class A beta-lactamase [Tabrizicola alkalilacus]
WP_095951366.1 class A beta-lactamase, subclass A2 [Flavobacterium sp. ACN6]
SCY57217.1 beta-lactamase class A CARB-1 [Desulfoluna spongiiphila]
TMU75979.1 class A beta-lactamase [Hydrogenophaga intermedia]
WP_166066202.1 class A beta-lactamase [Diaphorobacter sp. HDW4B]
WP_149648923.1 class A beta-lactamase [Azospirillum argentinense]
MBR3555062.1 GNAT family N-acetyltransferase [Oscillospiraceae bacterium]
WP_027778134.1 class A beta-lactamase [Paraburkholderia caledonica]
WP_245961988.1 class A beta-lactamase [[Pseudomonas] urumqiensis]
WP_153045275.1 class A beta-lactamase [Sinorhizobium americanum]

WP_211681140.1 class A beta-lactamase [Moritella sp. 24]
WP_116797844.1 subclass B1 metallo-beta-lactamase [Flavobacterium sp. 103]
ALG03680.1 beta-lactamase [uncultured bacterium]
WP_058722424.1 class C beta-lactamase [Paucibacter sp. KCTC 42545]
WP_086206342.1 class C beta-lactamase [Acinetobacter sp. ANC 4648]
WP_057157847.1 class D beta-lactamase [Massilia sp. Root351]
ACI29753.2 GcuF1/OXA-28 fusion protein [Pseudomonas aeruginosa]
WP_177219092.1 class D beta-lactamase [Polaromonas sp. OV174]
WP_092305823.1 phosphotransferase [Pseudomonas sp. NFIX28]
MBX9912722.1 class D beta-lactamase [Pseudomonadaceae bacterium]
QKE29104.1 putative class D beta-lactamase [Arcobacter acticola]
WP_235831602.1 class D beta-lactamase [Acinetobacter rongchengensis]
WP_241326703.1 GNAT family N-acetyltransferase [Aeromonas sp. MR7]
QTE69679.1 GNAT family N-acetyltransferase [Clostridiales bacterium]
MCD8490272.1 macrolide 2'-phosphotransferase [Desertifilum sp.]
WP_235831602.1 class D beta-lactamase [Acinetobacter rongchengensis]
WP_256875622.1 OXA-274 family carbapenem-hydrolyzing class D beta-lactamase OXA-944 [Acinetobacter gu
WP_236496866.1 OXA-46 family oxacillin-hydrolyzing class D beta-lactamase [Thiothrix winogradskyi]
AIF25978.1 putative acetyltransferase, GNAT family [uncultured bacterium Ad_113_F04_contig1]
WP_205587823.1 aminoglycoside 3'-phosphotransferase [Streptomyces sp. 11-1-2]
WP_205587823.1 aminoglycoside 3'-phosphotransferase [Streptomyces sp. 11-1-2]
WP_207393872.1 APH(6) family putative aminoglycoside O-phosphotransferase [Legionella taurinensis]
WP_096735335.1 OXA-274 family carbapenem-hydrolyzing class D beta-lactamase OXA-667 [Acinetobacter gu
MBP7769491.1 class D beta-lactamase [Aliarcobacter sp.]
WP_146384574.1 AAC(3)-I family aminoglycoside N-acetyltransferase [Luteimonas marina]
MBP7769491.1 class D beta-lactamase [Aliarcobacter sp.]
ADI03324.1 aminoglycoside phosphotransferase [Streptomyces bingchenggensis BCW-1]
MBP7737654.1 class D beta-lactamase [Spirochaetes bacterium]
WP_149648923.1 class A beta-lactamase [Azospirillum argentinense]
WP_095951366.1 class A beta-lactamase, subclass A2 [Flavobacterium sp. ACN6]
WP_176950817.1 APH(6) family putative aminoglycoside O-phosphotransferase [Rhizobium rhizolycopersici]
WP_211681140.1 class A beta-lactamase [Moritella sp. 24]
WP_090023000.1 class A beta-lactamase, subclass A2 [Chryseobacterium oleae]
WP_166066202.1 class A beta-lactamase [Diaphorobacter sp. HDW4B]
NOT94262.1 class A beta-lactamase, subclass A2 [Ferruginibacter sp.]
WP_118155791.1 class A beta-lactamase [Tabrizicola alkalilacus]
WP_249093128.1 class A beta-lactamase [Mediterranea sp. ET5]
SCY57217.1 beta-lactamase class A CARB-1 [Desulfoluna spongiiphila]
AIF25978.1 putative acetyltransferase, GNAT family [uncultured bacterium Ad_113_F04_contig1]
WP_206662053.1 phosphotransferase [Propioniciclava tarda]
WP_235049612.1 phosphotransferase [Bordetella bronchiseptica]
WP_221080609.1 phosphotransferase [Alcaligenes faecalis]
MBK6595569.1 class A beta-lactamase [Burkholderiales bacterium]
WP_040159492.1 phosphotransferase [Nigerium massiliense]
MBO5026667.1 serine hydrolase [Bacteroidaceae bacterium]
AMP54275.1 classA [uncultured bacterium]

AMP48593.1 CARB-PSE [uncultured bacterium]
WP_116797844.1 subclass B1 metallo-beta-lactamase [Flavobacterium sp. 103]
WP_173852188.1 BlaB/IND/MUS family subclass B1 metallo-beta-lactamase [Flavobacterium sp. 28A]
WP_040474030.1 BlaB/IND/MUS family subclass B1 metallo-beta-lactamase [Flavobacterium frigoris]
WP_155076559.1 subclass B1 metallo-beta-lactamase [Flavobacterium sp. MC2016-06]
TXH04664.1 beta-lactamase [Rhodocyclaceae bacterium]
WP_086206342.1 class C beta-lactamase [Acinetobacter sp. ANC 4648]
WP_004861468.1 APH(3') family aminoglycoside O-phosphotransferase [Acinetobacter gerneri]
WP_058722424.1 class C beta-lactamase [Paucibacter sp. KCTC 42545]
MBP7769491.1 class D beta-lactamase [Aliarcobacter sp.]
MBX9912722.1 class D beta-lactamase [Pseudomonadaceae bacterium]
AQY21079.1 Beta-lactamase OXA-10 precursor [Riemerella anatipestifer]
WP_137932453.1 aminoglycoside 3'-phosphotransferase [Mesorhizobium comanense]
WP_004861468.1 APH(3') family aminoglycoside O-phosphotransferase [Acinetobacter gerneri]
MBP7737654.1 class D beta-lactamase [Spirochaetes bacterium]
RKG38860.1 class D beta-lactamase [Acinetobacter rongchengensis]
WP_143223192.1 class D beta-lactamase [Acinetobacter sp. ANC 4973]
WP_086191974.1 aminoglycoside 3'-phosphotransferase [Acinetobacter sp. ANC 3832]
QKE29104.1 putative class D beta-lactamase [Arcobacter acticola]
WP_096735335.1 OXA-274 family carbapenem-hydrolyzing class D beta-lactamase OXA-667 [Acinetobacter gu
WP_256875622.1 OXA-274 family carbapenem-hydrolyzing class D beta-lactamase OXA-944 [Acinetobacter gu
WP_236496866.1 OXA-46 family oxacillin-hydrolyzing class D beta-lactamase [Thiothrix winogradskyi]
CAI1772374.1 Aminoglycoside/hydroxyurea antibiotic resistance kinase [Serratia marcescens]
WP_177219092.1 class D beta-lactamase [Polaromonas sp. OV174]
ACI29753.2 GcuF1/OXA-28 fusion protein [Pseudomonas aeruginosa]
ACI29753.2 GcuF1/OXA-28 fusion protein [Pseudomonas aeruginosa]
WP_211681140.1 class A beta-lactamase [Moritella sp. 24]
WP_092836705.1 class A beta-lactamase [Roseovarius lutimaris]
WP_095951366.1 class A beta-lactamase, subclass A2 [Flavobacterium sp. ACN6]
WP_086191974.1 aminoglycoside 3'-phosphotransferase [Acinetobacter sp. ANC 3832]
TRW88068.1 GNAT family N-acetyltransferase [Mycolicibacterium sp. 018/SC-01/001]
OLP03714.1 aminoglycoside N-acetyltransferase AAC(2')-Ib [Mycobacterium porcinum]
NOT94262.1 class A beta-lactamase, subclass A2 [Ferruginibacter sp.]
UXD25037.1 Beta-lactamase [Yersinia enterocolitica]
MBP9019866.1 aminoglycoside 3'-phosphotransferase [Bacteroidales bacterium]
WP_183860772.1 MULTISPECIES: GNAT family N-acetyltransferase [unclassified Rhizobium]
WP_119762917.1 phosphotransferase [Deinococcus cavernae]
WP_118155791.1 class A beta-lactamase [Tabrizicola alkalilacus]
NLI49929.1 aminoglycoside resistance protein [Propionibacterium sp.]
WP_166159251.1 class A beta-lactamase [Acidovorax sp. HDW3]
WP_223303238.1 23S ribosomal RNA methyltransferase Erm [Aeromicrobium tamlense]
WP_223303238.1 23S ribosomal RNA methyltransferase Erm [Aeromicrobium tamlense]
CAF3175248.1 Aminoglycoside N(6')-acetyltransferase type 1 [Enterobacter cloacae]
CAF3175248.1 Aminoglycoside N(6')-acetyltransferase type 1 [Enterobacter cloacae]
NLI49929.1 aminoglycoside resistance protein [Propionibacterium sp.]
MBK6595569.1 class A beta-lactamase [Burkholderiales bacterium]

WP_201612657.1 23S ribosomal RNA methyltransferase Erm [Gulosibacter hominis]
WP_229822533.1 class A beta-lactamase [Novosphingobium arvoryzae]
TMU75979.1 class A beta-lactamase [Hydrogenophaga intermedia]
MBL9023700.1 class A beta-lactamase [Myxococcales bacterium]
MBK6595569.1 class A beta-lactamase [Burkholderiales bacterium]
WP_166066202.1 class A beta-lactamase [Diaphorobacter sp. HDW4B]
WP_090023000.1 class A beta-lactamase, subclass A2 [Chryseobacterium oleae]
MBK9234985.1 GNAT family N-acetyltransferase [Rhodoferax sp.]
WP_191090379.1 AAC(3) family N-acetyltransferase [Dechloromonas sp. CZR5]
WP_256118782.1 BKC/GPC family carbapenem-hydrolyzing class A beta-lactamase [Shinella lacus]
WP_203568402.1 aminoglycoside resistance protein [Aestuariimicrobium ganziense]
SCY57217.1 beta-lactamase class A CARB-1 [Desulfoluna spongiiphila]
WP_076387176.1 class A beta-lactamase, subclass A2 [Kaistella chaponensis]
QCT39786.1 methyltransferase [Candidatus Saccharibacteria bacterium oral taxon 955]
WP_044579084.1 aminoglycoside resistance protein [Arthrobacter alpinus]
WP_044579084.1 aminoglycoside resistance protein [Arthrobacter alpinus]
WP_068841060.1 GNAT family N-acetyltransferase [Flavobacterium chilense]
WP_068841060.1 GNAT family N-acetyltransferase [Flavobacterium chilense]
WP_188768813.1 serine hydrolase [Emticicia aquatilis]
WP_109740954.1 class A beta-lactamase, subclass A2 [Arcicella aurantiaca]
MBK6595569.1 class A beta-lactamase [Burkholderiales bacterium]
WP_191090379.1 AAC(3) family N-acetyltransferase [Dechloromonas sp. CZR5]
WP_209999764.1 aminoglycoside 3'-phosphotransferase [Paeniglutamicibacter kerguelensis]
WP_090078406.1 class A beta-lactamase, subclass A2 [Halpernia frigidisoli]
MBN8623456.1 class A beta-lactamase, subclass A2 [Flavobacteriales bacterium]
MBP6768229.1 class A beta-lactamase [Reyranella sp.]
WP_249393889.1 aminoglycoside 3'-phosphotransferase [Deinococcus sp. QL22]
AMP48593.1 CARB-PSE [uncultured bacterium]
WP_249393889.1 aminoglycoside 3'-phosphotransferase [Deinococcus sp. QL22]
MBU0747356.1 class A beta-lactamase [Gammaproteobacteria bacterium]
MBK6595569.1 class A beta-lactamase [Burkholderiales bacterium]
WP_143851291.1 class A beta-lactamase, subclass A2 [Chryseobacterium sp. SNU WT5]
MBV8248307.1 class A beta-lactamase [Comamonas sp.]
WP_149648923.1 class A beta-lactamase [Azospirillum argentinense]
WP_218381611.1 class A beta-lactamase [Acidovorax sp. sic0104]
HCJ31825.1 hypothetical protein [Firmicutes bacterium]
WP_063857835.1 PSE family carbenicillin-hydrolyzing class A beta-lactamase CARB-1 [Pseudomonas aerugino
TMU75979.1 class A beta-lactamase [Hydrogenophaga intermedia]
WP_232547141.1 aminoglycoside phosphotransferase family protein [Propioniciclava soli]
WP_116797844.1 subclass B1 metallo-beta-lactamase [Flavobacterium sp. 103]
MCP1583202.1 aminoglycoside 6'-N-acetyltransferase I [Pseudoxanthomonas mexicana]
WP_051618766.1 AAC(3) family N-acetyltransferase [Deinococcus radiodurans]
HCJ31825.1 hypothetical protein [Firmicutes bacterium]
MBN3510432.1 GNAT family N-acetyltransferase [Mycolicibacterium septicum]
WP_158034955.1 AAC(3) family N-acetyltransferase [Kocuria coralli]
WP_158034955.1 AAC(3) family N-acetyltransferase [Kocuria coralli]

WP_138950667.1 subclass B1 metallo-beta-lactamase [Aureibaculum algae]
OYU65816.1 subclass B1 metallo-beta-lactamase [Cytophagaceae bacterium BCCC1]
WP_255075702.1 subclass B1 metallo-beta-lactamase [Lacihabitans sp. CCS-44]
WP_263593348.1 AAC(3) family N-acetyltransferase [Brachybacterium huguangmaarense]
WP_245092121.1 MULTISPECIES: GNAT family N-acetyltransferase [Pseudoxanthomonas]
WP_188766146.1 subclass B1 metallo-beta-lactamase [Emticicia aquatilis]
WP_084059423.1 subclass B1 metallo-beta-lactamase [Cellulophaga tyrosinoxydans]
WP_185265411.1 subclass B1 metallo-beta-lactamase [Chryseobacterium indologenes]
WP_084059423.1 subclass B1 metallo-beta-lactamase [Cellulophaga tyrosinoxydans]
RZJ80005.1 MAG: subclass B1 metallo-beta-lactamase [Flavobacterium sp.]
WP_040474030.1 BlaB/IND/MUS family subclass B1 metallo-beta-lactamase [Flavobacterium frigoris]
MBI1185369.1 subclass B1 metallo-beta-lactamase [bacterium]
WP_246202904.1 subclass B3 metallo-beta-lactamase [Sphingomonas lacunae]
MBH1991830.1 subclass B3 metallo-beta-lactamase [Sphingomonadaceae bacterium]
WP_226749031.1 beta-lactamase [Loktanella sp. TSTF-M6]
CAF3175248.1 Aminoglycoside N(6')-acetyltransferase type 1 [Enterobacter cloacae]
WP_058722424.1 class C beta-lactamase [Paucibacter sp. KCTC 42545]
WP_086206342.1 class C beta-lactamase [Acinetobacter sp. ANC 4648]
NHF74677.1 APH(6) family putative aminoglycoside O-phosphotransferase [Paracoccus xiamenensis]
MBI3214810.1 GNAT family N-acetyltransferase [Mycobacterium sp.]
WP_163913602.1 class C beta-lactamase [Pseudomonas frederiksbergensis]
MBN9027240.1 class D beta-lactamase [Hyphomicrobiales bacterium]
WP_009450042.1 class D beta-lactamase [Nitratireductor indicus]
WP_009450042.1 class D beta-lactamase [Nitratireductor indicus]
WP_217694081.1 class D beta-lactamase [Vannielia litorea]
WP_103310573.1 TCR/Tet family MFS transporter [Deinococcus koreensis]
WP_061938040.1 class D beta-lactamase [Collimonas pratensis]
WP_063204786.1 penicillin-binding transpeptidase domain-containing protein [Bdellovibrio bacteriovorus]
WP_063866424.1 quinolone resistance pentapeptide repeat protein QnrS9 [Klebsiella pneumoniae]
WP_063866424.1 quinolone resistance pentapeptide repeat protein QnrS9 [Klebsiella pneumoniae]
WP_063244044.1 penicillin-binding transpeptidase domain-containing protein [Bdellovibrio bacteriovorus]
AOF91150.1 penicillin binding transpeptidase domain protein [Sinorhizobium sp. RAC02]
MBP7769491.1 class D beta-lactamase [Aliarcobacter sp.]
WP_240067735.1 OXA-198 family carbapenem-hydrolyzing class D beta-lactamase OXA-1057 [Pseudomonas c
WP_100886243.1 OXA-229 family carbapenem-hydrolyzing class D beta-lactamase OXA-895 [Acinetobacter be
PKO51440.1 hypothetical protein CVU27_06220 [Betaproteobacteria bacterium HGW-Betaproteobacteria-20]
NQU50985.1 class D beta-lactamase [Bacteroidetes bacterium]
MBX9912722.1 class D beta-lactamase [Pseudomonadaceae bacterium]
MBP7737654.1 class D beta-lactamase [Spirochaetes bacterium]
QKE29104.1 putative class D beta-lactamase [Arcobacter acticola]
AMP48561.1 classD [uncultured bacterium]
WP_157615101.1 class D beta-lactamase [Variovorax paradoxus]
QKE29104.1 putative class D beta-lactamase [Arcobacter acticola]
WP_096735335.1 OXA-274 family carbapenem-hydrolyzing class D beta-lactamase OXA-667 [Acinetobacter gu
WP_116131168.1 TCR/Tet family MFS transporter [Tropicimonas sp. IMCC34043]
MBK9061574.1 class D beta-lactamase [Flavobacteriales bacterium]

WP_117172494.1 class D beta-lactamase [Mariniflexile sp. TRM1-10]
WP_237986791.1 GNAT family N-acetyltransferase [Casaltella massiliensis]
MBP6022806.1 class D beta-lactamase [Ferruginibacter sp.]
WP_228163740.1 23S ribosomal RNA methyltransferase Erm [Microbacterium testaceum]
WP_188548411.1 class D beta-lactamase [Hymenobacter qilianensis]
MBP7769491.1 class D beta-lactamase [Aliarcobacter sp.]
WP_077687393.1 hypothetical protein [Tessaracoccus aquimaris]
WP_028868880.1 Qnr family pentapeptide repeat protein [Psychromonas arctica]
WP_028868880.1 Qnr family pentapeptide repeat protein [Psychromonas arctica]
NTV84765.1 class D beta-lactamase [Bacteroidales bacterium]
WP_134652716.1 GNAT family N-acetyltransferase [Shinella sumterensis]
WP_177219092.1 class D beta-lactamase [Polaromonas sp. OV174]
MCK6541170.1 23S ribosomal RNA methyltransferase Erm [bacterium]
WP_005259042.1 MULTISPECIES: AAC(6')-Ighjkrstuvwx family aminoglycoside N-acetyltransferase [Acineto
WP_239809455.1 GNAT family N-acetyltransferase [Comamonas sp. B21-038]
WP_239809455.1 GNAT family N-acetyltransferase [Comamonas sp. B21-038]
MCB5279398.1 23S rRNA (adenine(2058)-N(6))-methyltransferase Erm(A) [Candidatus Cloacimonetes bacteriu
WP_229988018.1 class D beta-lactamase [Flavobacterium sp. F-65]
WP_121900326.1 kinase [Tessaracoccus antarcticus]
WP_177219092.1 class D beta-lactamase [Polaromonas sp. OV174]
WP_118886044.1 class D beta-lactamase [Arcobacter suis]
WP_232703758.1 ESP-1 family subclass B3 metallo-beta-lactamase [Epilithonimonas sp. JDS]
WP_207822500.1 class A beta-lactamase [Brevundimonas pondensis]
WP_158387013.1 class A beta-lactamase [Comamonas testosteroni]
WP_158387013.1 class A beta-lactamase [Comamonas testosteroni]
WP_158387013.1 class A beta-lactamase [Comamonas testosteroni]
MCI4442682.1 class D beta-lactamase [Lentimicrobium sp.]
WP_177219092.1 class D beta-lactamase [Polaromonas sp. OV174]
AZM39680.1 class C beta-lactamase [Acinetobacter baumannii]
WP_158387013.1 class A beta-lactamase [Comamonas testosteroni]
MBG0505245.1 subclass B1 metallo-beta-lactamase [Elizabethkingia anophelis]
MCI6401910.1 23S ribosomal RNA methyltransferase Erm [Oscillospiraceae bacterium]
WP_046739866.1 class C beta-lactamase [Acinetobacter sp. AG1]
WP_046739866.1 class C beta-lactamase [Acinetobacter sp. AG1]
WP_011634137.1 class A beta-lactamase [Nitrosomonas eutropha]
WP_011634137.1 class A beta-lactamase [Nitrosomonas eutropha]
WP_011634137.1 class A beta-lactamase [Nitrosomonas eutropha]
WP_011634137.1 class A beta-lactamase [Nitrosomonas eutropha]
BAP35887.1 hypothetical protein AS4_09470 [Acinetobacter guillouiae]
MCP4561377.1 class A beta-lactamase [Bosea sp.]
BAP35887.1 hypothetical protein AS4_09470 [Acinetobacter guillouiae]
WP_092272362.1 MULTISPECIES: class A beta-lactamase [unclassified Duganella]
WP_121453593.1 MBL fold metallo-hydrolase [Acidovorax sp. 106]
WP_128990904.1 class D beta-lactamase [Arcobacter sp. CECT 8986]
WP_059015111.1 class D beta-lactamase [Flavobacterium psychrophilum]
WP_248138077.1 23S ribosomal RNA methyltransferase Erm [Micrococcus sp. EYE_162]

ALG03687.1 beta-lactamase [uncultured bacterium]
WP_011372581.1 class D beta-lactamase [Sulfurimonas denitrificans]
MBK8119780.1 class D beta-lactamase [Sulfuritalea sp.]
WP_248138077.1 23S ribosomal RNA methyltransferase Erm [Micrococcus sp. EYE_162]
WP_068733786.1 aminoglycoside 3'-phosphotransferase [Paeniglutamicibacter antarcticus]
WP_011372581.1 class D beta-lactamase [Sulfurimonas denitrificans]
WP_246093273.1 class A beta-lactamase [Zoogloea ramigera]
MBG0505245.1 subclass B1 metallo-beta-lactamase [Elizabethkingia anophelis]
MBG0505245.1 subclass B1 metallo-beta-lactamase [Elizabethkingia anophelis]
HBO3867382.1 class A beta-lactamase [Pseudomonas aeruginosa]
MBG0505245.1 subclass B1 metallo-beta-lactamase [Elizabethkingia anophelis]
AWT08553.1 QnrVC7 [Pseudomonas aeruginosa]
WP_258543728.1 class D beta-lactamase [Parvicella tangerina]
WP_258543728.1 class D beta-lactamase [Parvicella tangerina]
MBK7736138.1 class A beta-lactamase, subclass A2 [Saprospiraceae bacterium]
MBK7736138.1 class A beta-lactamase, subclass A2 [Saprospiraceae bacterium]
MBK7736138.1 class A beta-lactamase, subclass A2 [Saprospiraceae bacterium]
MBK7736138.1 class A beta-lactamase, subclass A2 [Saprospiraceae bacterium]
WP_004650739.1 class D beta-lactamase OXA-296 [Acinetobacter bohemicus]
WP_124447809.1 class A beta-lactamase [Paucibacter sp. KBW04]
HBO3867382.1 class A beta-lactamase [Pseudomonas aeruginosa]
WP_048709300.1 MULTISPECIES: class C beta-lactamase [Proteobacteria]
WP_059015111.1 class D beta-lactamase [Flavobacterium psychrophilum]
WP_048709300.1 MULTISPECIES: class C beta-lactamase [Proteobacteria]
WP_246093273.1 class A beta-lactamase [Zoogloea ramigera]
MBD3830034.1 class D beta-lactamase [Arcobacter sp.]
WP_241233047.1 class A beta-lactamase [Pseudomonas aeruginosa]
MBD3830034.1 class D beta-lactamase [Arcobacter sp.]
CQR83532.1 beta-lactamase/D-alanine carboxypeptidase [Escherichia coli K-12]
WP_076387176.1 class A beta-lactamase, subclass A2 [Kaistella chaponensis]
WP_076387176.1 class A beta-lactamase, subclass A2 [Kaistella chaponensis]
WP_121453593.1 MBL fold metallo-hydrolase [Acidovorax sp. 106]
AMP47568.1 classC-AmpC [uncultured bacterium]
WP_027814019.1 MULTISPECIES: aminoglycoside 6'-N-acetyltransferase AacA38 [Proteobacteria]
WP_069796862.1 class A beta-lactamase, subclass A2 [Cloacibacterium normanense]
WP_027814019.1 MULTISPECIES: aminoglycoside 6'-N-acetyltransferase AacA38 [Proteobacteria]
NLV82867.1 class D beta-lactamase [Synergistaceae bacterium]
CAD7546396.1 aminoglycoside N(6')-acetyltransferase type 1 [Aeromonas hydrophila]
WP_004650739.1 class D beta-lactamase OXA-296 [Acinetobacter bohemicus]
WP_068372448.1 class A beta-lactamase [Rhodococcus sp. EPR-157]
WP_076387176.1 class A beta-lactamase, subclass A2 [Kaistella chaponensis]
WP_227750903.1 MULTISPECIES: tetracycline efflux MFS transporter Tet(42) [Microbacterium]
WP_128321378.1 BES family class A beta-lactamase [Raoultella ornithinolytica]
WP_128321378.1 BES family class A beta-lactamase [Raoultella ornithinolytica]
WP_068372448.1 class A beta-lactamase [Rhodococcus sp. EPR-157]
MBP6216604.1 AAC(3)-I family aminoglycoside N-acetyltransferase [Luteimonas sp.]

WP_241233047.1 class A beta-lactamase [Pseudomonas aeruginosa]
WP_005328535.1 Qnr family pentapeptide repeat protein [Aeromonas media]
WP_073290566.1 rRNA adenine N(6)-methyltransferase family protein [Anaerosporobacter mobilis]
WP_237043943.1 PAU family class A beta-lactamase [Pseudomonas aeruginosa]
WP_099737147.1 class A beta-lactamase [Comamonas sp. 26]
WP_034369791.1 class A beta-lactamase [Comamonas testosteroni]
WP_242217332.1 class D beta-lactamase [Shinella zoogloeoides]
WP_099737147.1 class A beta-lactamase [Comamonas sp. 26]
MCP1583202.1 aminoglycoside 6'-N-acetyltransferase I [Pseudoxanthomonas mexicana]
WP_032494864.1 MULTISPECIES: extended-spectrum class A beta-lactamase VEB-9 [Proteobacteria]
WP_118308872.1 class A beta-lactamase, subclass A2 [Bacteroides eggerthii]
HCG1435102.1 APH(3')-II family aminoglycoside O-phosphotransferase [Pseudomonas aeruginosa]
APO27487.1 ferrous iron transport protein B [uncultured bacterium]
EAO3803148.1 AAC(6')-Ib family aminoglycoside 6'-N-acetyltransferase [Salmonella enterica]
EIY98793.1 hypothetical protein HMPREF1056_01267, partial [Bacteroides fragilis CL07T12C05]
WP_130145076.1 MULTISPECIES: class D beta-lactamase [Acinetobacter]
WP_185210947.1 class A beta-lactamase, subclass A2 [Elizabethkingia meningoseptica]
TVL95669.1 MAG: tetracycline resistance ribosomal protection protein [Candidatus Brocadia sp. WS118]
WP_171537671.1 class D beta-lactamase [Acinetobacter terrestris]
WP_008863796.1 class A beta-lactamase [Parasutterella excrementihominis]
WP_055186072.1 AAC(3) family N-acetyltransferase [Faecalibacterium prausnitzii]
WP_240040201.1 CphA family subclass B2 metallo-beta-lactamase [Aeromonas hydrophila]
MBP7086420.1 GNAT family N-acetyltransferase [Enterococcus sp.]
WP_092692487.1 class C beta-lactamase [Acinetobacter kyonggiensis]
WP_126037746.1 class C beta-lactamase [Acinetobacter johnsonii]
WP_004650739.1 class D beta-lactamase OXA-296 [Acinetobacter bohemicus]
WP_185209618.1 class A beta-lactamase [Chryseobacterium sp. C3]
WP_126037746.1 class C beta-lactamase [Acinetobacter johnsonii]
WP_054256025.1 class A beta-lactamase [Acidovorax caeni]
WP_126037746.1 class C beta-lactamase [Acinetobacter johnsonii]
HCE14906.1 hypothetical protein [Clostridiales bacterium]
MBP8061095.1 class A beta-lactamase, subclass A2 [Cloacibacterium sp.]
HAY63238.1 hypothetical protein [Oscillospiraceae bacterium]
WP_180551239.1 AAC(3)-I family aminoglycoside N-acetyltransferase [Ottowia beijingensis]
HBA02228.1 GNAT family N-acetyltransferase [Ruminococcus sp.]
MBS6989360.1 class A beta-lactamase [Azospirillum sp.]
WP_068886293.1 class C beta-lactamase [Acinetobacter celticus]
WP_126037746.1 class C beta-lactamase [Acinetobacter johnsonii]
WP_201612657.1 23S ribosomal RNA methyltransferase Erm [Gulosibacter hominis]
MCI1952108.1 tetracycline resistance ribosomal protection protein [Clostridiales bacterium]
MBD9173422.1 aminoglycoside 3'-phosphotransferase [Clostridiales bacterium]
WP_174835061.1 tetracycline efflux MFS transporter Tet(G) [Sphingomonas sp. CL5.1]
MCI1952108.1 tetracycline resistance ribosomal protection protein [Clostridiales bacterium]
SWV50123.1 putative aminoglycoside N(6')-acetyltransferase (Amikacin resistance protein) (AAC(6')) [Klebsiel
WP_073290566.1 rRNA adenine N(6)-methyltransferase family protein [Anaerosporobacter mobilis]
ATZ64700.1 tetracycline resistance MFS efflux pump [Acinetobacter bereziniae]

ATZ64700.1 tetracycline resistance MFS efflux pump [Acinetobacter bereziniae]
MBS6251134.1 GNAT family N-acetyltransferase [Clostridiales bacterium]
BDF64221.1 beta-lactamase [Alistipes finegoldii]
BDF64221.1 beta-lactamase [Alistipes finegoldii]
CAO72218.1 ErmX protein, partial [Bifidobacterium animalis]
UGK55404.1 Beta-lactamase [Raoultella ornithinolytica]
HBW02239.1 class A beta-lactamase [Alistipes sp.]
WP_113995806.1 FOX/MOX family class C beta-lactamase [Aeromonas hydrophila]
RHQ96343.1 TetM/TetW/TetO/TetS family tetracycline resistance ribosomal protection protein [Peptoclostridiu[illegible]
WP_050696799.1 tetracycline resistance ribosomal protection protein [Anaeromassilibacillus senegalensis]
MBD9280952.1 aminoglycoside 3'-phosphotransferase [Clostridiales bacterium]
WP_042016119.1 MULTISPECIES: class D beta-lactamase [Aeromonas]
MBJ7435079.1 OXA-211 family carbapenem-hydrolyzing class D beta-lactamase [Acinetobacter sp.]
MBS6995561.1 class A beta-lactamase [Azospirillum sp.]
WP_166159251.1 class A beta-lactamase [Acidovorax sp. HDW3]
WP_130145076.1 MULTISPECIES: class D beta-lactamase [Acinetobacter]
HIR78125.1 GNAT family N-acetyltransferase [Candidatus Egerieenecus merdigallinarum]
OLA09435.1 MAG: GNAT family N-acetyltransferase [Eubacterium sp. 45_250]
WP_185209618.1 class A beta-lactamase [Chryseobacterium sp. C3]
OJV32391.1 subclass B3 metallo-beta-lactamase [Sphingomonas sp. 67-36]
NCC07651.1 methyltransferase domain-containing protein [Clostridia bacterium]
CDC98626.1 hGB-1 beta-lactamase [Alistipes sp. CAG:268]
WP_187573687.1 AAC(3)-I family aminoglycoside N-acetyltransferase [Pseudoxanthomonas mexicana]
WP_164987546.1 aminoglycoside resistance protein [Propioniciclava flava]
WP_048709300.1 MULTISPECIES: class C beta-lactamase [Proteobacteria]
MBO5349701.1 rRNA adenine N(6)-methyltransferase family protein [Clostridia bacterium]
HBW3776649.1 APH(3')-VI family aminoglycoside O-phosphotransferase [Klebsiella pneumoniae]
MBC8672249.1 CphA family subclass B2 metallo-beta-lactamase [Aeromonas hydrophila]
WP_140967323.1 APH(3')-VI family aminoglycoside O-phosphotransferase [Acinetobacter baumannii]
OYX37921.1 hypothetical protein B7Z00_02650 [Candidatus Saccharibacteria bacterium 32-50-10]
HCF5415646.1 AacA4 family aminoglycoside N(6')-acetyltransferase [Pseudomonas aeruginosa]
HCF5415646.1 AacA4 family aminoglycoside N(6')-acetyltransferase [Pseudomonas aeruginosa]
WP_213603971.1 subclass B3 metallo-beta-lactamase PJM-1 [Pseudoxanthomonas japonensis]
WP_142534931.1 tetracycline resistance ribosomal protection protein Tet(44) [Peptacetobacter hominis]
HCF5415646.1 AacA4 family aminoglycoside N(6')-acetyltransferase [Pseudomonas aeruginosa]
WP_155165098.1 MBL fold metallo-hydrolase [Parasutterella excrementihominis]
WP_073342390.1 MULTISPECIES: 23S ribosomal RNA methyltransferase Erm [Bacteroidales]
HCF5415646.1 AacA4 family aminoglycoside N(6')-acetyltransferase [Pseudomonas aeruginosa]
WP_118435663.1 class A beta-lactamase, subclass A2 [Bacteroides cellulosilyticus]
WP_063857819.1 carbenicillin-hydrolyzing class A beta-lactamase AER-1 [Aeromonas hydrophila]
MBP8038075.1 class A beta-lactamase [Prevotella sp.]
WP_198846418.1 class A beta-lactamase [Acidovorax sp. IB03]
MBP8061095.1 class A beta-lactamase, subclass A2 [Cloacibacterium sp.]
WP_022675378.1 subclass B3 metallo-beta-lactamase [Novosphingobium sp. B-7]
MBP8038075.1 class A beta-lactamase [Prevotella sp.]
ACT97394.1 AmpC-EcolK12 beta-lactamase [mixed culture bacterium AX_gF3SD01_01]

WP_113842634.1 MULTISPECIES: tetracycline resistance ribosomal protection protein [Firmicutes]
WP_142534931.1 tetracycline resistance ribosomal protection protein Tet(44) [Peptacetobacter hominis]
WP_053397570.1 MULTISPECIES: class A beta-lactamase [Prevotella]
AMP49121.1 classA [uncultured bacterium]
WP_262087325.1 beta-lactamase [Acinetobacter sp. I-MWF]
WP_078481254.1 MULTISPECIES: class C beta-lactamase [Pseudomonas]
WP_042066448.1 OXA-12 family class D beta-lactamase [Aeromonas hydrophila]
WP_011013281.1 MULTISPECIES: oxacillin-hydrolyzing class D beta-lactamase NPS-1 [Proteobacteria]
MBP8061095.1 class A beta-lactamase, subclass A2 [Cloacibacterium sp.]
WP_009596813.1 MULTISPECIES: subclass B1 metallo-beta-lactamase [Alistipes]
WP_262087325.1 beta-lactamase [Acinetobacter sp. I-MWF]
WP_011013281.1 MULTISPECIES: oxacillin-hydrolyzing class D beta-lactamase NPS-1 [Proteobacteria]
MBP8928750.1 AAC(3)-I family aminoglycoside N-acetyltransferase [Ottowia sp.]
WP_198846418.1 class A beta-lactamase [Acidovorax sp. IB03]
AMP50979.1 CblA [uncultured bacterium]
AMP48925.1 CblA [uncultured bacterium]
OQA98182.1 Beta-lactamase OXA-2 precursor [Bacteroidetes bacterium ADurb.Bin217]
AMP47204.1 classD [uncultured bacterium]
MCI5676974.1 GNAT family N-acetyltransferase [Clostridia bacterium]
MBS6251134.1 GNAT family N-acetyltransferase [Clostridiales bacterium]
WP_007213314.1 class A beta-lactamase, subclass A2 [Bacteroides cellulosilyticus]
WP_204461399.1 class A beta-lactamase, subclass A2 [Phocaeicola coprocola]
WP_198846418.1 class A beta-lactamase [Acidovorax sp. IB03]
AHM79651.1 Beta-lactamase [Klebsiella pneumoniae 30684/NJST258_2]
WP_141418823.1 class A beta-lactamase [Alistipes communis]
MBP7261866.1 class A beta-lactamase, subclass A2 [Bacteroidia bacterium]
WP_140423313.1 OXA-211 family carbapenem-hydrolyzing class D beta-lactamase OXA-644 [Acinetobacter jo
MBP8928750.1 AAC(3)-I family aminoglycoside N-acetyltransferase [Ottowia sp.]
WP_227248514.1 GNAT family N-acetyltransferase [Roseburia faecis]
WP_063857819.1 carbenicillin-hydrolyzing class A beta-lactamase AER-1 [Aeromonas hydrophila]
MBN8623456.1 class A beta-lactamase, subclass A2 [Flavobacteriales bacterium]
MBS1939435.1 class A beta-lactamase, subclass A2 [Bacteroidetes bacterium]
AMP48925.1 CblA [uncultured bacterium]
CQR83532.1 beta-lactamase/D-alanine carboxypeptidase [Escherichia coli K-12]
WP_005390949.1 MULTISPECIES: 23S ribosomal RNA methyltransferase Erm [Actinomycetia]
WP_038557444.1 MULTISPECIES: Mph(E)/Mph(G) family macrolide 2'-phosphotransferase [Marinilabiliales]
MBP8928750.1 AAC(3)-I family aminoglycoside N-acetyltransferase [Ottowia sp.]
WP_117672447.1 class A beta-lactamase, subclass A2 [Phocaeicola plebeius]
WP_063857819.1 carbenicillin-hydrolyzing class A beta-lactamase AER-1 [Aeromonas hydrophila]
AMP48925.1 CblA [uncultured bacterium]
WP_064964607.1 class A beta-lactamase, subclass A2 [Riemerella anatipestifer]
WP_198846418.1 class A beta-lactamase [Acidovorax sp. IB03]
WP_042088348.1 OXA-229 family carbapenem-hydrolyzing class D beta-lactamase OXA-1001 [Acinetobacter b
WP_002592614.1 MULTISPECIES: aminoglycoside N-acetyltransferase AAC(6')-Im [Lachnospiraceae]
WP_227248514.1 GNAT family N-acetyltransferase [Roseburia faecis]
WP_187573687.1 AAC(3)-I family aminoglycoside N-acetyltransferase [Pseudoxanthomonas mexicana]

WP_002592614.1 MULTISPECIES: aminoglycoside N-acetyltransferase AAC(6')-Im [Lachnospiraceae]
WP_180551239.1 AAC(3)-I family aminoglycoside N-acetyltransferase [Ottowia beijingensis]
WP_227248514.1 GNAT family N-acetyltransferase [Roseburia faecis]
AMP53591.1 tet_ribosomoal_protect [uncultured bacterium]
WP_042877968.1 MULTISPECIES: Qnr family pentapeptide repeat protein [Aeromonas]
WP_111911606.1 Qnr family pentapeptide repeat protein [Aeromonas media]
MBI0396458.1 beta-lactamase [Acinetobacter bereziniae]
WP_012634451.1 MULTISPECIES: quinolone resistance pentapeptide repeat protein QnrD1 [Bacteria]
MBI0396458.1 beta-lactamase [Acinetobacter bereziniae]
ACT97517.1 AmpC-HG2 beta-lactamase, partial [mixed culture bacterium CF_gF3SD01_19]
WP_042877968.1 MULTISPECIES: Qnr family pentapeptide repeat protein [Aeromonas]
WP_243323146.1 tetracycline resistance ribosomal protection protein Tet(Q) [Parabacteroides sp. AGMB00274]
ABR80560.1 streptomycin resistance protein B [Klebsiella pneumoniae subsp. pneumoniae MGH 78578]
MBP6661912.1 subclass B1 metallo-beta-lactamase [Paludibacter sp.]
WP_032492122.1 MULTISPECIES: OXA-5 family class D beta-lactamase OXA-129 [Proteobacteria]
QKE29104.1 putative class D beta-lactamase [Arcobacter acticola]
EJA2650198.1 23S ribosomal RNA methyltransferase Erm [Pseudomonas aeruginosa]
ABR80560.1 streptomycin resistance protein B [Klebsiella pneumoniae subsp. pneumoniae MGH 78578]
CDA17792.1 beta-lactamase [Acetobacter sp. CAG:267]
WP_032492122.1 MULTISPECIES: OXA-5 family class D beta-lactamase OXA-129 [Proteobacteria]
MBS6249525.1 23S ribosomal RNA methyltransferase Erm [Clostridiales bacterium]
MBQ1574690.1 AAC(3) family N-acetyltransferase [Clostridiales bacterium]
ABR80560.1 streptomycin resistance protein B [Klebsiella pneumoniae subsp. pneumoniae MGH 78578]
WP_210015376.1 APH(6) family putative aminoglycoside O-phosphotransferase [Pseudomonas palmensis]
KAA3160692.1 subclass B1 metallo-beta-lactamase [Alistipes finegoldii]
WP_035589998.1 subclass B1 metallo-beta-lactamase [Elizabethkingia anophelis]
AMP48538.1 classD [uncultured bacterium]
MBP8205294.1 class D beta-lactamase [Giesbergeria sp.]
WP_042088348.1 OXA-229 family carbapenem-hydrolyzing class D beta-lactamase OXA-1001 [Acinetobacter b
WP_064964835.1 MULTISPECIES: 23S rRNA (adenine(2058)-N(6))-methyltransferase Erm(F) [Weeksellaceae
ABP68837.1 QnrB6 [Klebsiella pneumoniae]
MBQ1574690.1 AAC(3) family N-acetyltransferase [Clostridiales bacterium]
ABR80560.1 streptomycin resistance protein B [Klebsiella pneumoniae subsp. pneumoniae MGH 78578]
CUQ16647.1 alpha-glycosidase [Phocaeicola vulgatus]
MBN8623456.1 class A beta-lactamase, subclass A2 [Flavobacteriales bacterium]
WP_021980438.1 class A beta-lactamase, subclass A2 [Paraprevotella clara]
WP_004294668.1 class A beta-lactamase, subclass A2 [Bacteroides eggerthii]
WP_035589998.1 subclass B1 metallo-beta-lactamase [Elizabethkingia anophelis]
WP_063861588.1 OXA-46 family oxacillin-hydrolyzing class D beta-lactamase OXA-205 [Pseudomonas aerugi
WP_073342390.1 MULTISPECIES: 23S ribosomal RNA methyltransferase Erm [Bacteroidales]
WP_159375860.1 class A beta-lactamase [Moraxella osloensis]
AMP49121.1 classA [uncultured bacterium]
MCB1465502.1 class A beta-lactamase [Nitratireductor sp.]
MBP7261866.1 class A beta-lactamase, subclass A2 [Bacteroidia bacterium]
MCI1731167.1 class A beta-lactamase [Prevotella sp.]
QJA10405.1 class A extended-spectrum beta-lactamase, partial [uncultured bacterium]

EHU1405633.1 MCA family class C beta-lactamase [Acinetobacter baumannii]
AMP47568.1 classC-AmpC [uncultured bacterium]
AXQ85785.1 OXA-1-like protein [Pseudomonas aeruginosa]
MBO5349701.1 rRNA adenine N(6)-methyltransferase family protein [Clostridia bacterium]
AMP50979.1 CblA [uncultured bacterium]
AMP49121.1 classA [uncultured bacterium]
MBP7785603.1 class A beta-lactamase [Bifidobacterium sp.]
ANG12681.1 beta-lactamase TEM-1 variant [synthetic construct]
WP_032494864.1 MULTISPECIES: extended-spectrum class A beta-lactamase VEB-9 [Proteobacteria]
KAB3638900.1 class A beta-lactamase [Phocaeicola vulgatus]
AMP48925.1 CblA [uncultured bacterium]
MCB1465502.1 class A beta-lactamase [Nitratireductor sp.]
WP_042649345.1 MULTISPECIES: CMY-1/MOX family class C beta-lactamase MOX-9 [Gammaproteobacteri
QLO40532.1 beta-lactamase [Klebsiella sp. RHBSTW-00484]
AXQ85785.1 OXA-1-like protein [Pseudomonas aeruginosa]
TXG77876.1 macrolide 2'-phosphotransferase [Candidatus Dojkabacteria bacterium]
MBP6408896.1 Mph(B) family macrolide 2'-phosphotransferase [Fusobacteriaceae bacterium]
WP_000219391.1 MULTISPECIES: Mph(A) family macrolide 2'-phosphotransferase [Bacteria]
WP_017411290.1 MULTISPECIES: tetracycline efflux MFS transporter Tet(E) [Gammaproteobacteria]
AMP49121.1 classA [uncultured bacterium]
KAB3638900.1 class A beta-lactamase [Phocaeicola vulgatus]
MCB1465502.1 class A beta-lactamase [Nitratireductor sp.]
MCI1731167.1 class A beta-lactamase [Prevotella sp.]
WP_008861554.1 class A beta-lactamase [Barnesiella intestinihominis]
MBP7261866.1 class A beta-lactamase, subclass A2 [Bacteroidia bacterium]
QJA10405.1 class A extended-spectrum beta-lactamase, partial [uncultured bacterium]
WP_074557906.1 class A beta-lactamase, subclass A2 [Bacteroides ovatus]
WP_159375860.1 class A beta-lactamase [Moraxella osloensis]
ALG03677.1 beta-lactamase [uncultured bacterium]
WP_094009812.1 extended-spectrum class A beta-lactamase VEB-19 [Escherichia coli]
MBP7785603.1 class A beta-lactamase [Bifidobacterium sp.]
WP_004327564.1 class A beta-lactamase, subclass A2 [Alistipes putredinis]
AMP47568.1 classC-AmpC [uncultured bacterium]
AVA17849.1 beta-lactamase OXA-1 [uncultured bacterium]
MBP6408896.1 Mph(B) family macrolide 2'-phosphotransferase [Fusobacteriaceae bacterium]
TXG77876.1 macrolide 2'-phosphotransferase [Candidatus Dojkabacteria bacterium]
EFH13796.1 putative translation elongation factor G [Clostridioides difficile NAP07]
WP_181748465.1 class A beta-lactamase, subclass A2 [Bacteroides fragilis]
QJA10405.1 class A extended-spectrum beta-lactamase, partial [uncultured bacterium]
AMP49121.1 classA [uncultured bacterium]
MBP7785603.1 class A beta-lactamase [Bifidobacterium sp.]
ALG03677.1 beta-lactamase [uncultured bacterium]
WP_086989205.1 serine hydrolase [Trichococcus flocculiformis]
WP_141418823.1 class A beta-lactamase [Alistipes communis]
MCB1465502.1 class A beta-lactamase [Nitratireductor sp.]
AMP48925.1 CblA [uncultured bacterium]

AMP47568.1 classC-AmpC [uncultured bacterium]
TXG77876.1 macrolide 2'-phosphotransferase [Candidatus Dojkabacteria bacterium]
AMP53591.1 tet_ribosomoal_protect [uncultured bacterium]
EAM0355781.1 GNAT family N-acetyltransferase [Campylobacter coli]
ACQ42051.1 CTX-M-15 [Escherichia coli]
WP_011270171.1 MULTISPECIES: class A beta-lactamase [Proteobacteria]
AMP49121.1 classA [uncultured bacterium]
WP_117908041.1 class A beta-lactamase, subclass A2 [Bacteroides stercoris]
WP_179219264.1 class A beta-lactamase [Akkermansia muciniphila]
QHV83156.1 SHV-12 [Escherichia coli]
WP_122364884.1 class A beta-lactamase, subclass A2 [Bacteroides caccae]
WP_129324144.1 ACT family cephalosporin-hydrolyzing class C beta-lactamase [Enterobacter cloacae]
AMP47568.1 classC-AmpC [uncultured bacterium]
WP_146266213.1 tetracycline resistance ribosomal protection protein Tet(36) [Flavobacterium channae]
WP_231299036.1 MULTISPECIES: APH(2")-Ia/If/Ih family aminoglycoside O-phosphotransferase [Eubacterial
ACQ42051.1 CTX-M-15 [Escherichia coli]
WP_053397570.1 MULTISPECIES: class A beta-lactamase [Prevotella]
EIY98793.1 hypothetical protein HMPREF1056_01267, partial [Bacteroides fragilis CL07T12C05]
KAB5409037.1 class A beta-lactamase [Phocaeicola vulgatus]
CDA71336.1 putative uncharacterized protein [Bacteroides coprocola CAG:162]
KAB3638900.1 class A beta-lactamase [Phocaeicola vulgatus]
WP_181748465.1 class A beta-lactamase, subclass A2 [Bacteroides fragilis]
WP_195422944.1 class A beta-lactamase, subclass A2 [Bacteroides cellulosilyticus]
WP_063857820.1 subclass B3 metallo-beta-lactamase AIM-1 [Pseudomonas aeruginosa]
AMP47568.1 classC-AmpC [uncultured bacterium]
AGQ08547.1 Beta-lactamase class D [Acinetobacter baumannii BJAB0715]
TXI65181.1 MAG: Mph(E)/Mph(G) family macrolide 2'-phosphotransferase [Flavobacterium sp.]
APO30252.1 Ferrous iron transport protein B [uncultured bacterium]
EPR81956.1 Tetracycline resistance protein [Acinetobacter gerneri DSM 14967 = CIP 107464 = MTCC 9824]
EPR81956.1 Tetracycline resistance protein [Acinetobacter gerneri DSM 14967 = CIP 107464 = MTCC 9824]
WP_153828815.1 tetracycline resistance ribosomal protection protein Tet(Q) [Ornithobacterium rhinotracheale]
EPR81956.1 Tetracycline resistance protein [Acinetobacter gerneri DSM 14967 = CIP 107464 = MTCC 9824]
WP_113842634.1 MULTISPECIES: tetracycline resistance ribosomal protection protein [Firmicutes]
EPR81956.1 Tetracycline resistance protein [Acinetobacter gerneri DSM 14967 = CIP 107464 = MTCC 9824]
WP_118063230.1 tetracycline resistance ribosomal protection protein Tet(32) [Ruminococcus sp. OM08-9BH]
EPR81956.1 Tetracycline resistance protein [Acinetobacter gerneri DSM 14967 = CIP 107464 = MTCC 9824]
WP_262780998.1 TCR/Tet family MFS transporter [Acinetobacter guillouiae]
AMP53591.1 tet_ribosomoal_protect [uncultured bacterium]
BBI29161.1 Tetracycline resistance, MFS efflux pump [Klebsiella pneumoniae]
WP_262780998.1 TCR/Tet family MFS transporter [Acinetobacter guillouiae]
EPR81956.1 Tetracycline resistance protein [Acinetobacter gerneri DSM 14967 = CIP 107464 = MTCC 9824]
WP_169151336.1 class D beta-lactamase [Azoarcus sp. TTM-91]
MBP7769491.1 class D beta-lactamase [Aliarcobacter sp.]
WP_236496866.1 OXA-46 family oxacillin-hydrolyzing class D beta-lactamase [Thiothrix winogradskyi]
WP_109740954.1 class A beta-lactamase, subclass A2 [Arcicella aurantiaca]
WP_095951366.1 class A beta-lactamase, subclass A2 [Flavobacterium sp. ACN6]

WP_254063395.1 class A beta-lactamase [Rhodanobacter sp. L36]
UXD25037.1 Beta-lactamase [Yersinia enterocolitica]
MBK6595569.1 class A beta-lactamase [Burkholderiales bacterium]
WP_166159251.1 class A beta-lactamase [Acidovorax sp. HDW3]
MBP6925064.1 methyltransferase domain-containing protein [Candidatus Saccharibacteria bacterium]
MBK6595569.1 class A beta-lactamase [Burkholderiales bacterium]
NOT94262.1 class A beta-lactamase, subclass A2 [Ferruginibacter sp.]
TMU75979.1 class A beta-lactamase [Hydrogenophaga intermedia]
WP_092836705.1 class A beta-lactamase [Roseovarius lutimaris]
WP_090023000.1 class A beta-lactamase, subclass A2 [Chryseobacterium oleae]
WP_211681140.1 class A beta-lactamase [Moritella sp. 24]
WP_100410627.1 class A beta-lactamase [Acidovorax sp. 69]
WP_027990679.1 class A beta-lactamase [Sinorhizobium meliloti]
MBK9236258.1 class A beta-lactamase [Rhodoferax sp.]
WP_226344678.1 aminoglycoside phosphotransferase family protein [Agilicoccus flavus]
OJU87767.1 hypothetical protein BGO17_02145 [Candidatus Saccharibacteria bacterium 49-20]
MBC7459603.1 methyltransferase domain-containing protein [Candidatus Saccharibacteria bacterium]
MBK6595569.1 class A beta-lactamase [Burkholderiales bacterium]
MBQ0750512.1 class A beta-lactamase [Roseovarius sp.]
MBL0105859.1 class A beta-lactamase, subclass A2 [Ignavibacteria bacterium]
AMP48593.1 CARB-PSE [uncultured bacterium]
MCH9721032.1 GNAT family N-acetyltransferase [Actinomycetia bacterium]
WP_194659135.1 class A beta-lactamase, subclass A2 [Flavobacterium sp. LC2016-12]
WP_143851291.1 class A beta-lactamase, subclass A2 [Chryseobacterium sp. SNU WT5]
WP_191090379.1 AAC(3) family N-acetyltransferase [Dechloromonas sp. CZR5]
WP_224152065.1 class A beta-lactamase [Comamonas odontotermitis]
WP_149648923.1 class A beta-lactamase [Azospirillum argentinense]
WP_133599664.1 MULTISPECIES: APH(6) family putative aminoglycoside O-phosphotransferase [Raoultella]
WP_116797844.1 subclass B1 metallo-beta-lactamase [Flavobacterium sp. 103]
WP_249330969.1 subclass B1 metallo-beta-lactamase [Chryseobacterium arthrosphaerae]
WP_044579084.1 aminoglycoside resistance protein [Arthrobacter alpinus]
WP_142770653.1 BlaB/IND/MUS family subclass B1 metallo-beta-lactamase [Spirosoma sp. KCTC 42546]
OYX53972.1 hypothetical protein B7Y92_00935 [Candidatus Saccharibacteria bacterium 32-50-13]
WP_173852188.1 BlaB/IND/MUS family subclass B1 metallo-beta-lactamase [Flavobacterium sp. 28A]
WP_258068575.1 aminoglycoside phosphotransferase family protein [Arthrobacter sp. N199823]
WP_206567387.1 subclass B1 metallo-beta-lactamase [Algoriphagus aestuariicola]
WP_185265411.1 subclass B1 metallo-beta-lactamase [Chryseobacterium indologenes]
WP_040474030.1 BlaB/IND/MUS family subclass B1 metallo-beta-lactamase [Flavobacterium frigoris]
WP_162628159.1 subclass B1 metallo-beta-lactamase [Arcticibacterium luteifluviistationis]
WP_255075702.1 subclass B1 metallo-beta-lactamase [Lacihabitans sp. CCS-44]
WP_216784553.1 subclass B1 metallo-beta-lactamase [Cellulophaga sp. HaHaR_3_176]
WP_155076559.1 subclass B1 metallo-beta-lactamase [Flavobacterium sp. MC2016-06]
WP_246202904.1 subclass B3 metallo-beta-lactamase [Sphingomonas lacunae]
MBI3217072.1 aminoglycoside resistance protein [Mycobacterium sp.]
WP_258444838.1 beta-lactamase [Pseudomonas sp. LS1212]
WP_058722424.1 class C beta-lactamase [Paucibacter sp. KCTC 42545]

WP_226749031.1 beta-lactamase [Loktanella sp. TSTF-M6]
WP_163913602.1 class C beta-lactamase [Pseudomonas frederiksbergensis]
WP_230987056.1 class D beta-lactamase [Allorhizobium terrae]
MBK7822271.1 AAC(3) family N-acetyltransferase [Tessaracoccus sp.]
WP_063204786.1 penicillin-binding transpeptidase domain-containing protein [Bdellovibrio bacteriovorus]
WP_188057013.1 MULTISPECIES: aminoglycoside 3'-phosphotransferase [unclassified Leucobacter]
WP_157615101.1 class D beta-lactamase [Variovorax paradoxus]
HHU09552.1 kinase [Intrasporangiaceae bacterium]
WP_000586782.1 MULTISPECIES: oxacillin-hydrolyzing class D beta-lactamase OXA-20 [Gammaproteobacter
WP_236496866.1 OXA-46 family oxacillin-hydrolyzing class D beta-lactamase [Thiothrix winogradskyi]
MBX9912722.1 class D beta-lactamase [Pseudomonadaceae bacterium]
WP_116762922.1 class D beta-lactamase [Flavobacterium laiguense]
MBK9061574.1 class D beta-lactamase [Flavobacteriales bacterium]
PKO51440.1 hypothetical protein CVU27_06220 [Betaproteobacteria bacterium HGW-Betaproteobacteria-20]
MBU2019858.1 class D beta-lactamase [Bacteroidetes bacterium]
WP_240067735.1 OXA-198 family carbapenem-hydrolyzing class D beta-lactamase OXA-1057 [Pseudomonas c
MBK6366032.1 class D beta-lactamase [Polaromonas sp.]
WP_103457351.1 class D beta-lactamase [Stutzerimonas stutzeri]
ACI29753.2 GcuF1/OXA-28 fusion protein [Pseudomonas aeruginosa]
WP_096735335.1 OXA-274 family carbapenem-hydrolyzing class D beta-lactamase OXA-667 [Acinetobacter gu
QKE29104.1 putative class D beta-lactamase [Arcobacter acticola]
WP_245092121.1 MULTISPECIES: GNAT family N-acetyltransferase [Pseudoxanthomonas]
MCD8490272.1 macrolide 2'-phosphotransferase [Desertifilum sp.]
WP_082988147.1 class A beta-lactamase [Bordetella bronchialis]
WP_015060700.1 class A extended spectrum beta-lactamase BES-1 [Serratia marcescens]
WP_263593348.1 AAC(3) family N-acetyltransferase [Brachybacterium huguangmaarense]
MBL0105859.1 class A beta-lactamase, subclass A2 [Ignavibacteria bacterium]
MBD5252048.1 class A beta-lactamase [Bacteroides sp.]
WP_079667870.1 class A beta-lactamase, subclass A2 [Soonwooa buanensis]
WP_245968381.1 class A beta-lactamase [Salinisphaera halophila]
WP_109669559.1 class A beta-lactamase [Roseicyclus mahoneyensis]
MBD5462201.1 GNAT family N-acetyltransferase [Lachnospiraceae bacterium]
MBS1764015.1 subclass B1 metallo-beta-lactamase [Bacteroidetes bacterium]
WP_202979265.1 class C beta-lactamase [Citrobacter freundii]
MBK8803619.1 class D beta-lactamase [Fibrobacteres bacterium]
MBK9061574.1 class D beta-lactamase [Flavobacteriales bacterium]
WP_101260310.1 class D beta-lactamase [Labilibaculum filiforme]
MBK6366032.1 class D beta-lactamase [Polaromonas sp.]
MBN8702188.1 VOC family protein [Bacteroidetes bacterium]
WP_015060700.1 class A extended spectrum beta-lactamase BES-1 [Serratia marcescens]
WP_079667870.1 class A beta-lactamase, subclass A2 [Soonwooa buanensis]
MBO5026667.1 serine hydrolase [Bacteroidaceae bacterium]
WP_082988147.1 class A beta-lactamase [Bordetella bronchialis]
WP_245968381.1 class A beta-lactamase [Salinisphaera halophila]
NOT94262.1 class A beta-lactamase, subclass A2 [Ferruginibacter sp.]
WP_158387013.1 class A beta-lactamase [Comamonas testosteroni]

MTJ93634.1 class A beta-lactamase [Desulfovibrio sp.]
AZL61262.1 class A beta-lactamase [Tabrizicola piscis]
WP_116131168.1 TCR/Tet family MFS transporter [Tropicimonas sp. IMCC34043]
WP_158387013.1 class A beta-lactamase [Comamonas testosteroni]
MBS6844437.1 AAC(3) family N-acetyltransferase [Clostridiales bacterium]
WP_158387013.1 class A beta-lactamase [Comamonas testosteroni]
WP_184260379.1 class A beta-lactamase [Novispirillum itersonii]
MBS1764015.1 subclass B1 metallo-beta-lactamase [Bacteroidetes bacterium]
QDC28540.1 subclass B2 metallo-beta-lactamase [uncultured bacterium]
WP_021233414.1 subclass B3 metallo-beta-lactamase [Novosphingobium lindaniclasticum]
RZA34870.1 subclass B3 metallo-beta-lactamase [Xanthomonadaceae bacterium]
WP_202979265.1 class C beta-lactamase [Citrobacter freundii]
WP_233685854.1 beta-lactamase [Pseudomonas anguilliseptica]
RMT84180.1 Beta-lactamase/D-alanine carboxypeptidase [Pseudomonas viridiflava]
WP_213338545.1 class D beta-lactamase [Chelatococcus sp. HY11]
WP_220231395.1 class D beta-lactamase [Hoeflea sp. WL0058]
WP_063244044.1 penicillin-binding transpeptidase domain-containing protein [Bdellovibrio bacteriovorus]
WP_101260310.1 class D beta-lactamase [Labilibaculum filiforme]
WP_009778337.1 aminoglycoside phosphotransferase [Janibacter sp. HTCC2649]
WP_241150390.1 class D beta-lactamase [Delftia lacustris]
MBK9061574.1 class D beta-lactamase [Flavobacteriales bacterium]
WP_062172770.1 TCR/Tet family MFS transporter [Burkholderia sp. PAMC 26561]
NTW91342.1 AAC(3) family N-acetyltransferase [Erysipelotrichaceae bacterium]
WP_132957865.1 TCR/Tet family MFS transporter [Rhizobium sp. BK251]
WP_028868880.1 Qnr family pentapeptide repeat protein [Psychromonas arctica]
WP_028868880.1 Qnr family pentapeptide repeat protein [Psychromonas arctica]
MBW7942880.1 23S ribosomal RNA methyltransferase Erm [Candidatus Kuenenia stuttgartiensis]
ALG03679.1 beta-lactamase class D [uncultured bacterium]
WP_150552526.1 class D beta-lactamase [Pandoraea soli]
WP_088860312.1 class D beta-lactamase [Laribacter hongkongensis]
WP_091746303.1 class D beta-lactamase [Propionispora vibrioides]
MBK6366032.1 class D beta-lactamase [Polaromonas sp.]
MBN8702188.1 VOC family protein [Bacteroidetes bacterium]
PLW79849.1 class D beta-lactamase [Candidatus Woesearchaeota archaeon]
WP_158387013.1 class A beta-lactamase [Comamonas testosteroni]
WP_173504733.1 subclass B1 metallo-beta-lactamase [Flavobacterium sp. M31R6]
MBK6595569.1 class A beta-lactamase [Burkholderiales bacterium]
WP_177219092.1 class D beta-lactamase [Polaromonas sp. OV174]
WP_177219092.1 class D beta-lactamase [Polaromonas sp. OV174]
WP_122893269.1 class D beta-lactamase [Arcobacter peruensis]
WP_011634137.1 class A beta-lactamase [Nitrosomonas eutropha]
WP_011634137.1 class A beta-lactamase [Nitrosomonas eutropha]
WP_118886044.1 class D beta-lactamase [Arcobacter suis]
WP_128990904.1 class D beta-lactamase [Arcobacter sp. CECT 8986]
WP_101260308.1 class D beta-lactamase [Labilibaculum filiforme]
WP_128990904.1 class D beta-lactamase [Arcobacter sp. CECT 8986]

AMP48575.1 CARB-PSE [uncultured bacterium]
AMP48575.1 CARB-PSE [uncultured bacterium]
AMP47191.1 classD [uncultured bacterium]
WP_099736705.1 class D beta-lactamase [Comamonas sp. 26]
WP_244175872.1 tetracycline efflux MFS transporter Tet(42) [Microbacterium oxydans]
CQR83532.1 beta-lactamase/D-alanine carboxypeptidase [Escherichia coli K-12]
MBP6661912.1 subclass B1 metallo-beta-lactamase [Paludibacter sp.]
MBG0505245.1 subclass B1 metallo-beta-lactamase [Elizabethkingia anophelis]
MBG0505245.1 subclass B1 metallo-beta-lactamase [Elizabethkingia anophelis]
MBP6364397.1 subclass B3 metallo-beta-lactamase [Novosphingobium sp.]
WP_258543728.1 class D beta-lactamase [Parvicella tangerina]
WP_258543728.1 class D beta-lactamase [Parvicella tangerina]
MBK7736138.1 class A beta-lactamase, subclass A2 [Saprospiraceae bacterium]
MBK7736138.1 class A beta-lactamase, subclass A2 [Saprospiraceae bacterium]
WP_079458943.1 OXA-5 family class D beta-lactamase [Pseudomonas aeruginosa]
WP_048709300.1 MULTISPECIES: class C beta-lactamase [Proteobacteria]
WP_059015111.1 class D beta-lactamase [Flavobacterium psychrophilum]
WP_048709300.1 MULTISPECIES: class C beta-lactamase [Proteobacteria]
WP_141049904.1 class D beta-lactamase [Aliarcobacter cryaerophilus]
MBD3830034.1 class D beta-lactamase [Arcobacter sp.]
WP_068733786.1 aminoglycoside 3'-phosphotransferase [Paeniglutamicibacter antarcticus]
AMP47147.1 classD, partial [uncultured bacterium]
WP_027814019.1 MULTISPECIES: aminoglycoside 6'-N-acetyltransferase AacA38 [Proteobacteria]
WP_167655841.1 class C beta-lactamase [Pseudomonas sp. B14(2022)]
WP_241233047.1 class A beta-lactamase [Pseudomonas aeruginosa]
WP_076387176.1 class A beta-lactamase, subclass A2 [Kaistella chaponensis]
WP_004650739.1 class D beta-lactamase OXA-296 [Acinetobacter bohemicus]
WP_129025753.1 subclass B1 metallo-beta-lactamase [Flavobacterium sp. YO12]
WP_159374988.1 class D beta-lactamase [Moraxella osloensis]
MBP5992809.1 class D beta-lactamase [Acinetobacter sp.]
RUP42360.1 beta-lactamase [Acinetobacter sp.]
MCP1583202.1 aminoglycoside 6'-N-acetyltransferase I [Pseudoxanthomonas mexicana]
WP_185210947.1 class A beta-lactamase, subclass A2 [Elizabethkingia meningoseptica]
WP_185210947.1 class A beta-lactamase, subclass A2 [Elizabethkingia meningoseptica]
MCA0335223.1 class D beta-lactamase [Bacteroidetes bacterium]
HCB1475147.1 class D beta-lactamase [Citrobacter freundii]
HCB1475147.1 class D beta-lactamase [Citrobacter freundii]
MBP8161724.1 hypothetical protein [Ottowia sp.]
MBD9173422.1 aminoglycoside 3'-phosphotransferase [Clostridiales bacterium]
MCC6940552.1 subclass B3 metallo-beta-lactamase [Novosphingobium sp.]
WP_004650739.1 class D beta-lactamase OXA-296 [Acinetobacter bohemicus]
WP_195287176.1 GNAT family N-acetyltransferase [Roseburia faecis]
MCI6124210.1 aminoglycoside 3'-phosphotransferase [Christensenellaceae bacterium]
WP_068886293.1 class C beta-lactamase [Acinetobacter celticus]
WP_119932702.1 23S ribosomal RNA methyltransferase Erm [Kocuria sp. CPCC 104605]
WP_111911606.1 Qnr family pentapeptide repeat protein [Aeromonas media]

KAB3638900.1 class A beta-lactamase [Phocaeicola vulgatus]
WP_099657070.1 MBL fold metallo-hydrolase [Acidovorax sp. 56]
ACT97384.1 HGE-1 beta-lactamase [uncultured organism]
WP_240067734.1 OXA-198 family carbapenem-hydrolyzing class D beta-lactamase OXA-1056 [Pseudomonas a
WP_076387176.1 class A beta-lactamase, subclass A2 [Kaistella chaponensis]
VVN01565.1 Beta-lactamase [Pseudomonas fluorescens]
WP_086374409.1 MULTISPECIES: MCA family class C beta-lactamase [Acinetobacter]
WP_126037746.1 class C beta-lactamase [Acinetobacter johnsonii]
MBK7291810.1 class D beta-lactamase [Chitinophagaceae bacterium]
MBP8061095.1 class A beta-lactamase, subclass A2 [Cloacibacterium sp.]
AMP48647.1 TetA-G [uncultured bacterium]
WP_187573687.1 AAC(3)-I family aminoglycoside N-acetyltransferase [Pseudoxanthomonas mexicana]
EIU4863518.1 beta-lactamase [Pseudomonas aeruginosa]
MBH3313086.1 class A beta-lactamase [Pseudomonas mosselii]
WP_195289612.1 subclass B1 metallo-beta-lactamase [Alistipes finegoldii]
HCF5415646.1 AacA4 family aminoglycoside N(6')-acetyltransferase [Pseudomonas aeruginosa]
OQA98182.1 Beta-lactamase OXA-2 precursor [Bacteroidetes bacterium ADurb.Bin217]
HCF5415646.1 AacA4 family aminoglycoside N(6')-acetyltransferase [Pseudomonas aeruginosa]
WP_126037746.1 class C beta-lactamase [Acinetobacter johnsonii]
WP_009300547.1 MULTISPECIES: tetracycline resistance ribosomal protection protein Tet(44) [Firmicutes]
WP_073290566.1 rRNA adenine N(6)-methyltransferase family protein [Anaerosporobacter mobilis]
MCI1208181.1 tetracycline resistance ribosomal protection protein [Treponema sp.]
MBP7261866.1 class A beta-lactamase, subclass A2 [Bacteroidia bacterium]
WP_122364884.1 class A beta-lactamase, subclass A2 [Bacteroides caccae]
EIY98793.1 hypothetical protein HMPREF1056_01267, partial [Bacteroides fragilis CL07T12C05]
WP_265466831.1 FOX/MOX family class C beta-lactamase [Aeromonas salmonicida]
AMP53591.1 tet_ribosomoal_protect [uncultured bacterium]
GKN85319.1 hypothetical protein NUBL17189_52290 [Klebsiella pneumoniae]
WP_009596813.1 MULTISPECIES: subclass B1 metallo-beta-lactamase [Alistipes]
WP_059429510.1 tetracycline resistance ribosomal protection protein Tet(44) [Campylobacter hyointestinalis]
QKE29104.1 putative class D beta-lactamase [Arcobacter acticola]
WP_018426326.1 MULTISPECIES: tetracycline efflux MFS transporter Tet(G) [Proteobacteria]
MBP8928750.1 AAC(3)-I family aminoglycoside N-acetyltransferase [Ottowia sp.]
MBP7086420.1 GNAT family N-acetyltransferase [Enterococcus sp.]
AMP48925.1 CblA [uncultured bacterium]
WP_198846418.1 class A beta-lactamase [Acidovorax sp. IB03]
AMP50537.1 classA, partial [uncultured bacterium]
WP_117672447.1 class A beta-lactamase, subclass A2 [Phocaeicola plebeius]
BBE19185.1 beta-lactamase [Aquipluma nitroreducens]
MCI5676974.1 GNAT family N-acetyltransferase [Clostridia bacterium]
AMP50979.1 CblA [uncultured bacterium]
CDA71336.1 putative uncharacterized protein [Bacteroides coprocola CAG:162]
OQA98182.1 Beta-lactamase OXA-2 precursor [Bacteroidetes bacterium ADurb.Bin217]
WP_018579533.1 MULTISPECIES: tetracycline resistance ribosomal protection protein [Firmicutes]
ABO92047.1 quinolone resistance determinant QnrA [Aeromonas salmonicida subsp. salmonicida A449]
WP_088656287.1 MULTISPECIES: tetracycline resistance ribosomal protection protein Tet(36) [Bacteroidota]

ULG20519.1 Beta-lactamase [Acinetobacter nosocomialis]
ABR80560.1 streptomycin resistance protein B [Klebsiella pneumoniae subsp. pneumoniae MGH 78578]
WP_157057059.1 class A beta-lactamase [Herbaspirillum autotrophicum]
CUQ16647.1 alpha-glycosidase [Phocaeicola vulgatus]
WP_035589998.1 subclass B1 metallo-beta-lactamase [Elizabethkingia anophelis]
WP_032492122.1 MULTISPECIES: OXA-5 family class D beta-lactamase OXA-129 [Proteobacteria]
WP_005390949.1 MULTISPECIES: 23S ribosomal RNA methyltransferase Erm [Actinomycetia]
WP_118158801.1 MULTISPECIES: 23S ribosomal RNA methyltransferase Erm [Bacteria]
ABR80560.1 streptomycin resistance protein B [Klebsiella pneumoniae subsp. pneumoniae MGH 78578]
WP_157057059.1 class A beta-lactamase [Herbaspirillum autotrophicum]
WP_032492122.1 MULTISPECIES: OXA-5 family class D beta-lactamase OXA-129 [Proteobacteria]
MCK6541170.1 23S ribosomal RNA methyltransferase Erm [bacterium]

d (aa) identity.

| % identity | E-value |
|---|---|
| 66.8 | 1.12e-121 |
| 100 | 8.20e-105 |
| 100 | 7.88e-103 |
| 100 | 1.04e-188 |
| 100 | 4.25e-106 |
| 100 | 5.04e-97 |
| 100 | 1.79e-108 |
| 100 | 1.54e-263 |
| 100 | 1.04e-193 |
| 100 | 2.73e-180 |
| 100 | 3.27e-197 |
| 100 | 1.00e-197 |
| 100 | 4.57e-197 |
| 100 | 1.80e-204 |
| 100 | 1.56e-210 |
| 100 | 8.20e-202 |
| 100 | 2.18e-235 |
| 100 | 6.81e-212 |
| 100 | 6.07e-210 |
| 100 | 2.21e-217 |
| 100 | 4.97e-203 |
| 100 | 2.15e-226 |
| 100 | 6.49e-233 |
| 100 | 1.06e-220 |
| 100 | 1.73e-272 |
| 100 | 1.35e-272 |
| 100 | 2.15e-285 |
| 100 | 1.60e-196 |
| 100 | 8.55e-193 |
| 100 | 1.80e-195 |
| 100 | 3.20e-206 |
| 100 | 1.17e-195 |
| 100 | 2.17e-187 |
| 100 | 3.33e-180 |
| 100 | 1.40e-200 |
| 100 | 4.22e-194 |
| 100 | 4.12e-184 |
| 100 | 3.07e-172 |
| 100 | 1.43e-204 |
| 100 | 2.61e-203 |
| 100 | 1.31e-188 |
| 100 | 2.73e-170 |
| 100 | 1.81e-209 |

| | |
|---|---|
| 100 | 7.69e-211 |
| 100 | 1.56e-215 |
| 100 | 8.01e-213 |
| 100 | 6.04e-214 |
| 100 | 2.65e-107 |
| 100 | 6.76e-298 |
| 100 | 1.19e-270 |
| 100 | 2.02e-283 |
| 100 | 1.67e-284 |
| 100 | 0.0 |
| 100 | 0.0 |
| 100 | 5.77e-105 |
| 100 | 1.70e-90 |
| 100 | 6.04e-214 |
| 100 | 3.24e-158 |
| 100 | 5.37e-126 |
| 100 | 4.25e-106 |
| 100 | 1.79e-108 |
| 100 | 4.16e-133 |
| 100 | 6.82e-121 |
| 100 | 7.58e-234 |
| 100 | 1.79e-215 |
| 100 | 1.00e-197 |
| 100 | 1.04e-193 |
| 100 | 3.27e-197 |
| 100 | 4.57e-197 |
| 100 | 4.57e-201 |
| 100 | 2.15e-226 |
| 100 | 1.19e-211 |
| 100 | 4.40e-221 |
| 100 | 8.68e-216 |
| 100 | 1.79e-233 |
| 100 | 5.18e-204 |
| 100 | 8.20e-202 |
| 100 | 1.22e-208 |
| 100 | 1.80e-204 |
| 100 | 5.43e-194 |
| 100 | 1.31e-232 |
| 100 | 5.18e-191 |
| 100 | 3.17e-216 |
| 100 | 2.05e-214 |
| 100 | 3.86e-224 |
| 100 | 2.15e-285 |
| 100 | 2.26e-285 |
| 100 | 1.17e-195 |
| 100 | 1.80e-195 |

| | |
|---|---|
| 100 | 8.55e-193 |
| 100 | 2.35e-191 |
| 100 | 1.89e-188 |
| 100 | 1.60e-196 |
| 100 | 1.02e-203 |
| 100 | 2.17e-187 |
| 100 | 3.33e-180 |
| 100 | 1.40e-200 |
| 100 | 3.20e-206 |
| 100 | 3.60e-173 |
| 100 | 3.17e-175 |
| 100 | 4.12e-184 |
| 100 | 3.87e-170 |
| 100 | 1.31e-188 |
| 100 | 3.07e-172 |
| 100 | 1.43e-204 |
| 100 | 1.82e-214 |
| 100 | 2.98e-194 |
| 100 | 1.86e-188 |
| 100 | 1.81e-209 |
| 100 | 1.56e-215 |
| 100 | 7.69e-211 |
| 100 | 8.01e-213 |
| 100 | 2.65e-107 |
| 100 | 1.66e-114 |
| 100 | 1.19e-270 |
| 100 | 6.76e-298 |
| 100 | 1.67e-284 |
| 100 | 2.02e-283 |
| 100 | 1.06e-278 |
| 100 | 0.0 |
| 100 | 0.0 |
| 100 | 0.0 |
| 100 | 0.0 |
| 100 | 0.0 |
| 100 | 0.0 |
| 100 | 8.20e-105 |
| 100 | 5.77e-105 |
| 100 | 6.04e-214 |
| 100 | 7.91e-189 |
| 100 | 2.62e-176 |
| 100 | 4.25e-106 |
| 100 | 5.04e-97 |
| 100 | 6.82e-121 |
| 100 | 1.79e-108 |
| 100 | 2.58e-105 |

| | |
|---|---|
| 100 | 1.54e-263 |
| 100 | 9.96e-210 |
| 100 | 1.04e-193 |
| 100 | 2.73e-180 |
| 100 | 3.27e-197 |
| 100 | 1.00e-197 |
| 100 | 4.57e-197 |
| 100 | 6.81e-212 |
| 100 | 2.69e-214 |
| 100 | 2.18e-235 |
| 100 | 1.80e-204 |
| 100 | 9.72e-213 |
| 100 | 9.69e-210 |
| 100 | 5.87e-212 |
| 100 | 8.93e-197 |
| 100 | 1.45e-218 |
| 100 | 2.15e-226 |
| 100 | 6.07e-210 |
| 100 | 1.76e-221 |
| 100 | 9.89e-204 |
| 100 | 7.69e-211 |
| 100 | 4.57e-201 |
| 100 | 2.41e-188 |
| 100 | 4.97e-203 |
| 100 | 8.20e-202 |
| 100 | 5.18e-204 |
| 100 | 4.60e-206 |
| 100 | 2.05e-214 |
| 100 | 1.06e-220 |
| 100 | 2.19e-284 |
| 100 | 1.02e-273 |
| 100 | 3.83e-200 |
| 100 | 4.37e-187 |
| 100 | 9.19e-186 |
| 100 | 3.33e-180 |
| 100 | 1.80e-195 |
| 100 | 1.08e-192 |
| 100 | 1.17e-195 |
| 100 | 1.60e-196 |
| 100 | 1.40e-200 |
| 100 | 3.20e-206 |
| 100 | 4.22e-194 |
| 100 | 8.55e-193 |
| 100 | 1.31e-188 |
| 100 | 3.07e-172 |
| 100 | 4.12e-184 |

| | |
|---|---|
| 100 | 2.73e-170 |
| 100 | 2.37e-197 |
| 100 | 1.43e-204 |
| 100 | 1.53e-175 |
| 100 | 2.98e-194 |
| 100 | 1.21e-184 |
| 100 | 6.50e-192 |
| 100 | 1.82e-214 |
| 100 | 1.81e-209 |
| 100 | 1.56e-215 |
| 100 | 9.72e-213 |
| 100 | 2.65e-107 |
| 100 | 6.76e-298 |
| 100 | 1.19e-270 |
| 100 | 1.09e-282 |
| 100 | 2.02e-283 |
| 100 | 1.67e-284 |
| 100 | 0.0 |
| 100 | 0.0 |
| 100 | 0.0 |
| 100 | 0.0 |
| 100 | 5.77e-105 |
| 100 | 1.56e-89 |
| 100 | 6.04e-214 |
| 100 | 7.90e-173 |
| 100 | 4.25e-106 |
| 100 | 1.79e-108 |
| 100 | 6.82e-121 |
| 100 | 5.04e-97 |
| 100 | 3.85e-221 |
| 100 | 1.00e-197 |
| 100 | 1.04e-193 |
| 100 | 4.57e-197 |
| 100 | 1.79e-233 |
| 100 | 2.15e-226 |
| 100 | 3.93e-202 |
| 100 | 8.20e-202 |
| 100 | 1.80e-204 |
| 100 | 3.56e-191 |
| 100 | 4.57e-201 |
| 100 | 1.06e-193 |
| 100 | 1.15e-185 |
| 100 | 1.57e-203 |
| 100 | 6.49e-233 |
| 100 | 5.66e-209 |
| 100 | 2.15e-285 |

| | |
|---|---|
| 100 | 2.26e-194 |
| 100 | 4.37e-187 |
| 100 | 1.17e-195 |
| 100 | 8.55e-193 |
| 100 | 4.92e-184 |
| 100 | 1.80e-195 |
| 100 | 3.20e-206 |
| 100 | 3.84e-186 |
| 100 | 3.33e-180 |
| 100 | 1.60e-196 |
| 100 | 3.07e-172 |
| 100 | 2.73e-170 |
| 100 | 1.43e-204 |
| 100 | 2.98e-194 |
| 100 | 1.31e-188 |
| 100 | 1.74e-176 |
| 100 | 1.81e-209 |
| 100 | 1.56e-215 |
| 100 | 2.15e-221 |
| 100 | 1.19e-270 |
| 100 | 6.76e-298 |
| 100 | 1.67e-284 |
| 100 | 2.02e-283 |
| 100 | 0.0 |
| 100 | 0.0 |
| 100 | 5.77e-105 |
| 100 | 7.88e-103 |
| 100 | 6.04e-214 |
| 100 | 7.20e-132 |
| 100 | 3.56e-101 |
| 100 | 5.04e-97 |
| 100 | 4.25e-106 |
| 100 | 1.79e-108 |
| 100 | 1.25e-105 |
| 100 | 2.33e-129 |
| 100 | 1.54e-263 |
| 100 | 1.00e-197 |
| 100 | 3.27e-197 |
| 100 | 2.73e-180 |
| 100 | 4.62e-205 |
| 100 | 1.04e-193 |
| 100 | 1.38e-212 |
| 100 | 1.53e-231 |
| 100 | 4.97e-203 |
| 100 | 1.10e-210 |
| 100 | 3.98e-246 |

| | |
|---|---|
| 100 | 2.15e-226 |
| 100 | 1.22e-208 |
| 100 | 1.36e-212 |
| 100 | 1.80e-204 |
| 100 | 4.30e-216 |
| 100 | 6.81e-212 |
| 100 | 4.03e-190 |
| 100 | 2.64e-232 |
| 100 | 2.21e-217 |
| 100 | 7.88e-180 |
| 100 | 3.90e-188 |
| 100 | 9.91e-214 |
| 100 | 1.02e-273 |
| 100 | 2.15e-285 |
| 100 | 2.19e-284 |
| 100 | 8.91e-204 |
| 100 | 4.41e-191 |
| 100 | 1.17e-202 |
| 100 | 2.89e-203 |
| 100 | 1.40e-200 |
| 100 | 3.20e-206 |
| 100 | 4.22e-194 |
| 100 | 2.77e-202 |
| 100 | 1.17e-195 |
| 100 | 3.07e-172 |
| 100 | 2.37e-197 |
| 100 | 3.87e-170 |
| 100 | 9.58e-191 |
| 100 | 1.86e-188 |
| 100 | 6.97e-189 |
| 100 | 1.81e-209 |
| 100 | 9.72e-213 |
| 100 | 5.64e-213 |
| 100 | 2.80e-213 |
| 100 | 7.69e-211 |
| 100 | 2.65e-107 |
| 100 | 1.19e-270 |
| 100 | 2.19e-282 |
| 100 | 4.78e-288 |
| 100 | 2.02e-283 |
| 100 | 0.0 |
| 100 | 5.77e-105 |
| 100 | 7.88e-103 |
| 100 | 6.04e-214 |
| 100 | 5.37e-126 |
| 100 | 7.20e-132 |

| | |
|---|---|
| 100 | 1.25e-105 |
| 100 | 2.06e-100 |
| 100 | 5.04e-97 |
| 100 | 3.56e-101 |
| 100 | 4.25e-106 |
| 100 | 1.41e-99 |
| 100 | 2.33e-129 |
| 100 | 1.54e-263 |
| 100 | 1.04e-193 |
| 100 | 4.62e-205 |
| 100 | 3.27e-197 |
| 100 | 4.95e-193 |
| 100 | 1.00e-197 |
| 100 | 6.81e-212 |
| 100 | 1.22e-208 |
| 100 | 4.97e-203 |
| 100 | 7.71e-214 |
| 100 | 1.49e-213 |
| 100 | 8.34e-212 |
| 100 | 8.20e-202 |
| 100 | 8.68e-216 |
| 100 | 3.26e-212 |
| 100 | 1.12e-198 |
| 100 | 2.15e-226 |
| 100 | 1.61e-209 |
| 100 | 1.10e-212 |
| 100 | 8.90e-202 |
| 100 | 1.80e-204 |
| 100 | 3.76e-200 |
| 100 | 5.18e-204 |
| 100 | 8.12e-188 |
| 100 | 4.60e-206 |
| 100 | 3.01e-212 |
| 100 | 4.75e-195 |
| 100 | 1.31e-232 |
| 100 | 3.90e-188 |
| 100 | 7.88e-180 |
| 100 | 9.91e-214 |
| 100 | 1.06e-220 |
| 100 | 2.05e-214 |
| 100 | 2.15e-285 |
| 100 | 2.19e-284 |
| 100 | 8.91e-204 |
| 100 | 4.24e-193 |
| 100 | 3.83e-200 |
| 100 | 1.17e-202 |

| | |
|---|---|
| 100 | 9.12e-178 |
| 100 | 2.89e-203 |
| 100 | 1.08e-192 |
| 100 | 1.17e-195 |
| 100 | 3.20e-206 |
| 100 | 1.60e-196 |
| 100 | 1.03e-171 |
| 100 | 4.22e-194 |
| 100 | 2.77e-202 |
| 100 | 2.35e-191 |
| 100 | 9.58e-191 |
| 100 | 3.07e-172 |
| 100 | 2.37e-197 |
| 100 | 3.87e-170 |
| 100 | 1.31e-188 |
| 100 | 1.17e-208 |
| 100 | 4.12e-184 |
| 100 | 6.50e-192 |
| 100 | 1.43e-204 |
| 100 | 1.86e-188 |
| 100 | 9.72e-213 |
| 100 | 7.69e-211 |
| 100 | 3.28e-204 |
| 100 | 2.65e-107 |
| 100 | 3.36e-114 |
| 100 | 1.30e-74 |
| 100 | 2.19e-282 |
| 100 | 1.19e-270 |
| 100 | 9.40e-250 |
| 100 | 2.02e-283 |
| 100 | 1.06e-278 |
| 100 | 0.0 |
| 100 | 0.0 |
| 30.5 | 1.20e-31 |
| 31.3 | 6.82e-14 |
| 76.6 | 4.86e-160 |
| 81.3 | 5.08e-165 |
| 79.4 | 5.09e-169 |
| 66.6 | 1.42e-136 |
| 42.6 | 1.10e-67 |
| 73.9 | 1.92e-151 |
| 58.1 | 3.89e-97 |
| 39.8 | 1.98e-28 |
| 44.6 | 2.93e-74 |
| 50.8 | 4.36e-86 |
| 49.6 | 2.35e-70 |

| | |
|---|---|
| 81.0 | 3.01e-158 |
| 88.0 | 6.96e-161 |
| 79.8 | 1.01e-138 |
| 73.1 | 1.08e-196 |
| 82.8 | 1.08e-235 |
| 41.9 | 1.91e-66 |
| 34.9 | 1.52e-45 |
| 75.9 | 6.83e-145 |
| 45.2 | 2.18e-58 |
| 72.6 | 5.08e-130 |
| 82.7 | 1.88e-147 |
| 84.0 | 3.79e-145 |
| 46.7 | 4.05e-40 |
| 47.0 | 1.48e-30 |
| 47.2 | 3.57e-95 |
| 83.5 | 3.62e-143 |
| 78.5 | 3.58e-144 |
| 55.9 | 5.66e-99 |
| 47.9 | 1.44e-38 |
| 48.7 | 2.08e-90 |
| 48.7 | 2.95e-90 |
| 49.1 | 2.41e-83 |
| 73.2 | 2.24e-142 |
| 86.0 | 6.74e-162 |
| 51.4 | 9.05e-39 |
| 70.7 | 9.16e-128 |
| 51.4 | 3.87e-88 |
| 58.1 | 1.12e-100 |
| 58.1 | 3.89e-97 |
| 79.4 | 5.09e-169 |
| 52.9 | 2.60e-80 |
| 81.0 | 3.01e-158 |
| 83.0 | 6.08e-166 |
| 74.2 | 8.20e-153 |
| 76.6 | 4.86e-160 |
| 81.6 | 4.36e-166 |
| 58.5 | 2.76e-112 |
| 66.6 | 1.42e-136 |
| 54.0 | 4.45e-47 |
| 54.3 | 9.67e-107 |
| 54.5 | 4.19e-90 |
| 54.7 | 1.09e-83 |
| 68.0 | 9.99e-135 |
| 54.7 | 1.21e-85 |
| 53.5 | 8.21e-106 |
| 41.7 | 7.11e-61 |

| | |
|---|---|
| 66.8 | 1.12e-121 |
| 85.3 | 5.78e-157 |
| 81.1 | 6.25e-142 |
| 85.5 | 1.38e-153 |
| 72.8 | 7.35e-131 |
| 84.9 | 1.10e-214 |
| 84.1 | 1.68e-229 |
| 56.3 | 8.21e-98 |
| 73.1 | 1.08e-196 |
| 86.0 | 6.74e-162 |
| 71.5 | 4.83e-128 |
| 81.5 | 2.22e-173 |
| 57.6 | 4.03e-94 |
| 57.7 | 7.07e-103 |
| 57.9 | 3.69e-99 |
| 77.2 | 1.25e-148 |
| 71.7 | 1.46e-137 |
| 58.0 | 2.16e-103 |
| 83.5 | 1.97e-149 |
| 73.2 | 2.24e-142 |
| 78.2 | 3.68e-134 |
| 54.8 | 6.13e-96 |
| 58.1 | 1.53e-96 |
| 78.0 | 1.45e-149 |
| 36.1 | 2.36e-48 |
| 36.1 | 2.36e-48 |
| 81.0 | 3.01e-158 |
| 75.7 | 3.06e-143 |
| 79.4 | 5.09e-169 |
| 59.1 | 1.40e-107 |
| 60.0 | 6.77e-79 |
| 61.6 | 3.61e-57 |
| 76.6 | 4.86e-160 |
| 56.3 | 1.11e-102 |
| 63.6 | 1.68e-114 |
| 63.7 | 2.56e-56 |
| 63.9 | 3.58e-113 |
| 81.3 | 5.08e-165 |
| 64.2 | 2.26e-123 |
| 59.7 | 2.97e-122 |
| 64.7 | 1.97e-129 |
| 64.7 | 1.97e-129 |
| 64.8 | 3.34e-87 |
| 64.8 | 3.34e-87 |
| 65.6 | 9.32e-130 |
| 71.2 | 1.76e-142 |

| | |
|---|---|
| 66.5 | 3.49e-121 |
| 66.1 | 4.32e-145 |
| 42.6 | 1.55e-67 |
| 64.2 | 7.67e-123 |
| 68.0 | 2.01e-134 |
| 74.2 | 5.77e-153 |
| 79.9 | 4.20e-168 |
| 67.6 | 1.23e-67 |
| 68.0 | 1.22e-148 |
| 87.2 | 9.39e-177 |
| 68.6 | 1.16e-136 |
| 66.6 | 1.42e-136 |
| 81.9 | 1.86e-151 |
| 69.5 | 2.27e-109 |
| 69.7 | 9.08e-168 |
| 69.7 | 9.08e-168 |
| 69.7 | 1.52e-68 |
| 69.7 | 1.52e-68 |
| 66.6 | 3.15e-125 |
| 75.2 | 1.27e-155 |
| 73.9 | 2.54e-133 |
| 71.1 | 7.45e-133 |
| 71.1 | 4.07e-146 |
| 46.4 | 2.94e-84 |
| 73.2 | 2.16e-145 |
| 45.5 | 1.28e-59 |
| 71.9 | 1.46e-128 |
| 66.8 | 1.12e-121 |
| 72.4 | 3.22e-130 |
| 88.6 | 2.24e-185 |
| 76.3 | 5.13e-139 |
| 76.6 | 5.78e-152 |
| 80.4 | 1.07e-163 |
| 58.1 | 3.89e-97 |
| 52.7 | 6.60e-67 |
| 73.3 | 2.09e-138 |
| 64.0 | 1.33e-121 |
| 53.1 | 1.26e-102 |
| 74.4 | 7.04e-149 |
| 86.7 | 1.63e-147 |
| 74.8 | 6.80e-72 |
| 74.8 | 1.55e-133 |
| 75.0 | 1.54e-140 |
| 75.6 | 6.61e-94 |
| 75.7 | 6.67e-141 |
| 75.7 | 6.67e-141 |

| | |
|---|---|
| 63.5 | 8.18e-113 |
| 55.6 | 3.37e-87 |
| 55.3 | 1.57e-89 |
| 76.6 | 7.95e-137 |
| 76.6 | 2.57e-73 |
| 77.7 | 2.66e-145 |
| 68.8 | 1.32e-119 |
| 85.2 | 3.91e-158 |
| 69.0 | 5.43e-121 |
| 57.7 | 1.76e-83 |
| 85.1 | 9.33e-152 |
| 63.1 | 4.81e-112 |
| 59.7 | 1.85e-124 |
| 85.2 | 1.17e-174 |
| 88.3 | 5.25e-246 |
| 79.7 | 2.08e-116 |
| 73.1 | 1.08e-196 |
| 82.6 | 1.26e-234 |
| 80.3 | 4.61e-150 |
| 80.7 | 3.62e-103 |
| 54.7 | 1.24e-136 |
| 86.3 | 6.49e-156 |
| 64.1 | 3.30e-115 |
| 62.9 | 3.37e-113 |
| 65.3 | 1.52e-114 |
| 81.4 | 5.75e-213 |
| 71.9 | 3.17e-142 |
| 60.0 | 2.44e-105 |
| 81.7 | 1.08e-106 |
| 81.7 | 1.08e-106 |
| 58.3 | 2.65e-104 |
| 89.8 | 1.26e-165 |
| 71.4 | 7.88e-129 |
| 53.1 | 1.43e-91 |
| 70.3 | 4.39e-141 |
| 85.2 | 1.05e-170 |
| 66.9 | 4.85e-131 |
| 72.2 | 5.90e-129 |
| 57.9 | 3.69e-99 |
| 89.3 | 5.86e-167 |
| 59.1 | 2.77e-111 |
| 81.8 | 1.69e-158 |
| 83.1 | 3.25e-156 |
| 72.5 | 1.06e-140 |
| 84.8 | 6.87e-223 |
| 77.0 | 1.98e-150 |

| | |
|---|---|
| 78.1 | 1.11e-159 |
| 85.4 | 3.17e-87 |
| 72.2 | 4.73e-139 |
| 85.8 | 2.69e-150 |
| 52.3 | 5.08e-87 |
| 84.1 | 2.51e-157 |
| 87.2 | 1.80e-189 |
| 87.6 | 7.08e-119 |
| 87.6 | 7.08e-119 |
| 53.1 | 6.05e-90 |
| 87.8 | 2.07e-107 |
| 72.6 | 1.84e-139 |
| 88.4 | 2.84e-183 |
| 88.4 | 1.74e-84 |
| 88.5 | 9.66e-86 |
| 88.5 | 9.66e-86 |
| 88.9 | 7.02e-152 |
| 55.8 | 1.14e-92 |
| 89.5 | 7.87e-191 |
| 78.4 | 1.25e-150 |
| 89.4 | 6.45e-163 |
| 90.1 | 4.86e-166 |
| 90.6 | 5.10e-184 |
| 91.0 | 6.08e-188 |
| 91.0 | 6.08e-188 |
| 91.0 | 6.08e-188 |
| 91.2 | 2.47e-176 |
| 92.1 | 2.14e-178 |
| 92.2 | 2.30e-268 |
| 92.3 | 2.57e-182 |
| 92.5 | 1.02e-169 |
| 92.8 | 7.92e-176 |
| 93.0 | 1.94e-258 |
| 93.0 | 1.94e-258 |
| 93.2 | 1.64e-188 |
| 93.2 | 1.64e-188 |
| 93.2 | 1.64e-188 |
| 93.2 | 1.64e-188 |
| 93.4 | 4.52e-205 |
| 93.4 | 9.55e-190 |
| 93.4 | 4.52e-205 |
| 93.5 | 1.32e-190 |
| 93.7 | 2.98e-170 |
| 93.7 | 5.26e-173 |
| 93.9 | 2.93e-165 |
| 94.0 | 5.29e-192 |

| | |
|---|---|
| 94.1 | 1.56e-169 |
| 94.4 | 1.83e-180 |
| 94.7 | 4.53e-184 |
| 94.7 | 3.89e-194 |
| 94.8 | 1.71e-186 |
| 94.8 | 7.78e-182 |
| 95.0 | 3.17e-196 |
| 95.2 | 1.11e-173 |
| 95.2 | 1.11e-173 |
| 95.2 | 1.37e-209 |
| 95.2 | 1.11e-173 |
| 95.2 | 7.22e-83 |
| 95.3 | 8.17e-194 |
| 95.3 | 8.17e-194 |
| 95.4 | 1.79e-205 |
| 95.4 | 1.79e-205 |
| 95.4 | 1.79e-205 |
| 95.4 | 1.79e-205 |
| 95.4 | 2.90e-194 |
| 95.5 | 6.57e-191 |
| 95.6 | 1.99e-197 |
| 95.6 | 5.19e-265 |
| 95.6 | 2.15e-187 |
| 95.6 | 5.19e-265 |
| 95.6 | 3.31e-198 |
| 95.7 | 3.69e-177 |
| 95.7 | 2.97e-174 |
| 95.7 | 3.69e-177 |
| 95.9 | 0.0 |
| 95.9 | 2.83e-195 |
| 96.2 | 4.19e-197 |
| 96.2 | 5.78e-222 |
| 96.2 | 2.85e-275 |
| 96.2 | 1.64e-130 |
| 96.2 | 4.30e-191 |
| 96.2 | 1.64e-130 |
| 96.3 | 2.61e-181 |
| 96.4 | 9.59e-91 |
| 96.5 | 3.69e-197 |
| 96.6 | 4.67e-201 |
| 96.6 | 5.11e-198 |
| 96.6 | 1.13e-262 |
| 96.6 | 7.27e-199 |
| 96.6 | 7.27e-199 |
| 96.7 | 1.73e-207 |
| 96.8 | 2.38e-101 |

| | |
|---|---|
| 96.8 | 8.96e-213 |
| 96.8 | 1.70e-102 |
| 96.9 | 3.48e-200 |
| 97.0 | 1.17e-205 |
| 97.0 | 4.45e-179 |
| 97.0 | 2.86e-205 |
| 97.0 | 4.75e-192 |
| 97.2 | 6.50e-194 |
| 97.3 | 9.90e-96 |
| 97.3 | 7.34e-207 |
| 97.4 | 2.15e-187 |
| 97.4 | 7.01e-164 |
| 97.4 | 0.0 |
| 97.5 | 1.13e-138 |
| 97.5 | 3.47e-223 |
| 97.5 | 2.88e-191 |
| 97.6 | 6.80e-198 |
| 97.7 | 0.0 |
| 97.8 | 4.22e-186 |
| 97.8 | 1.32e-180 |
| 97.8 | 9.21e-195 |
| 97.8 | 1.72e-161 |
| 97.9 | 5.33e-101 |
| 97.9 | 1.53e-275 |
| 97.9 | 4.45e-277 |
| 97.9 | 3.35e-200 |
| 97.9 | 1.36e-187 |
| 97.9 | 1.89e-278 |
| 97.9 | 1.12e-196 |
| 97.9 | 1.89e-278 |
| 98.0 | 1.13e-177 |
| 98.0 | 3.16e-210 |
| 98.0 | 2.44e-169 |
| 98.0 | 3.85e-96 |
| 98.0 | 9.81e-102 |
| 98.0 | 5.12e-201 |
| 98.2 | 5.82e-280 |
| 98.2 | 8.84e-278 |
| 98.2 | 1.97e-195 |
| 98.2 | 0.0 |
| 98.2 | 1.80e-199 |
| 98.2 | 1.99e-258 |
| 98.2 | 0.0 |
| 98.3 | 3.89e-166 |
| 98.3 | 3.28e-203 |
| 98.3 | 3.87e-274 |

| | |
|---|---|
| 98.3 | 3.87e-274 |
| 98.4 | 5.27e-84 |
| 98.4 | 2.64e-184 |
| 98.4 | 4.56e-185 |
| 98.4 | 1.99e-172 |
| 98.4 | 6.54e-212 |
| 98.4 | 2.53e-229 |
| 98.4 | 9.02e-268 |
| 98.4 | 0.0 |
| 98.5 | 0.0 |
| 98.5 | 7.64e-201 |
| 98.5 | 2.02e-191 |
| 98.5 | 8.02e-191 |
| 98.6 | 1.86e-203 |
| 98.6 | 7.57e-189 |
| 98.6 | 2.01e-193 |
| 98.6 | 3.73e-103 |
| 98.6 | 2.08e-99 |
| 98.6 | 1.64e-197 |
| 98.6 | 1.64e-190 |
| 98.6 | 1.30e-204 |
| 98.7 | 2.21e-213 |
| 98.7 | 9.93e-102 |
| 98.7 | 2.21e-222 |
| 98.7 | 1.02e-271 |
| 98.7 | 2.48e-212 |
| 98.8 | 3.72e-178 |
| 98.8 | 5.23e-185 |
| 98.8 | 7.83e-180 |
| 98.8 | 1.29e-177 |
| 98.9 | 4.18e-133 |
| 98.9 | 4.18e-133 |
| 98.9 | 1.44e-196 |
| 98.9 | 0.0 |
| 98.9 | 4.18e-133 |
| 98.9 | 1.18e-190 |
| 98.9 | 3.34e-191 |
| 98.9 | 4.18e-133 |
| 99.0 | 1.32e-210 |
| 99.0 | 2.01e-204 |
| 99.0 | 1.26e-198 |
| 99.0 | 2.20e-211 |
| 99.0 | 3.25e-212 |
| 99.0 | 7.13e-209 |
| 99.0 | 1.26e-198 |
| 99.0 | 0.0 |

| | |
|---|---|
| 99.1 | 0.0 |
| 99.1 | 0.0 |
| 99.1 | 2.49e-220 |
| 99.1 | 3.72e-225 |
| 99.2 | 2.88e-279 |
| 99.2 | 9.60e-281 |
| 99.2 | 6.74e-181 |
| 99.2 | 4.37e-184 |
| 99.2 | 1.79e-181 |
| 99.2 | 7.47e-173 |
| 99.2 | 2.88e-279 |
| 99.2 | 8.82e-184 |
| 99.3 | 1.76e-89 |
| 99.3 | 7.67e-212 |
| 99.3 | 5.20e-212 |
| 99.3 | 4.47e-215 |
| 99.3 | 4.79e-202 |
| 99.3 | 9.20e-193 |
| 99.3 | 1.70e-99 |
| 99.3 | 8.67e-106 |
| 99.3 | 6.56e-211 |
| 99.3 | 7.88e-212 |
| 99.3 | 7.67e-212 |
| 99.3 | 3.23e-208 |
| 99.3 | 7.37e-209 |
| 99.3 | 4.82e-213 |
| 99.3 | 2.26e-193 |
| 99.3 | 4.87e-99 |
| 99.3 | 3.37e-101 |
| 99.3 | 3.23e-208 |
| 99.3 | 1.30e-182 |
| 99.3 | 6.05e-194 |
| 99.3 | 4.47e-215 |
| 99.3 | 0.0 |
| 99.3 | 5.38e-193 |
| 99.3 | 1.05e-213 |
| 99.3 | 4.87e-99 |
| 99.3 | 5.02e-213 |
| 99.3 | 3.23e-208 |
| 99.3 | 4.47e-215 |
| 99.3 | 2.48e-212 |
| 99.3 | 6.37e-185 |
| 99.3 | 1.42e-197 |
| 99.4 | 1.41e-128 |
| 99.4 | 3.95e-120 |
| 99.4 | 3.46e-102 |

| | |
|---|---|
| 99.4 | 9.95e-129 |
| 99.4 | 2.96e-103 |
| 99.4 | 2.29e-119 |
| 99.5 | 0.0 |
| 99.5 | 2.12e-80 |
| 99.5 | 1.14e-100 |
| 99.5 | 5.55e-278 |
| 99.5 | 1.23e-133 |
| 99.5 | 5.55e-278 |
| 99.5 | 4.54e-303 |
| 99.5 | 2.12e-80 |
| 99.5 | 0.0 |
| 99.6 | 1.18e-203 |
| 99.6 | 4.56e-178 |
| 99.6 | 3.32e-187 |
| 99.6 | 1.56e-187 |
| 99.6 | 3.76e-191 |
| 99.6 | 1.18e-203 |
| 99.6 | 4.69e-187 |
| 99.6 | 3.32e-187 |
| 99.6 | 5.69e-168 |
| 99.6 | 2.02e-195 |
| 99.6 | 1.18e-203 |
| 99.6 | 3.94e-199 |
| 99.6 | 3.81e-177 |
| 99.6 | 3.90e-183 |
| 99.6 | 3.23e-196 |
| 99.6 | 7.03e-192 |
| 99.6 | 2.46e-198 |
| 99.6 | 2.64e-188 |
| 99.6 | 1.01e-108 |
| 99.6 | 2.02e-195 |
| 99.6 | 1.18e-203 |
| 99.6 | 3.32e-192 |
| 99.6 | 1.79e-193 |
| 99.6 | 1.08e-201 |
| 99.6 | 6.26e-192 |
| 99.6 | 3.90e-183 |
| 99.6 | 1.80e-195 |
| 99.6 | 2.45e-193 |
| 99.7 | 8.84e-218 |
| 99.7 | 2.73e-227 |
| 99.7 | 3.58e-220 |
| 99.7 | 1.18e-213 |
| 99.7 | 7.58e-200 |
| 99.7 | 1.25e-230 |

| | |
|---|---|
| 99.7 | 1.29e-273 |
| 99.7 | 1.50e-286 |
| 99.7 | 1.04e-202 |
| 99.7 | 6.08e-213 |
| 99.7 | 2.21e-213 |
| 99.7 | 2.73e-227 |
| 99.7 | 5.34e-202 |
| 99.7 | 1.00e-202 |
| 99.7 | 1.17e-207 |
| 99.7 | 1.65e-217 |
| 99.7 | 5.43e-216 |
| 99.7 | 2.21e-215 |
| 99.7 | 8.34e-273 |
| 99.7 | 1.80e-283 |
| 99.7 | 1.04e-202 |
| 99.7 | 5.41e-211 |
| 99.7 | 1.76e-220 |
| 99.7 | 2.29e-211 |
| 99.7 | 2.68e-237 |
| 99.7 | 2.73e-227 |
| 99.7 | 2.05e-245 |
| 99.7 | 3.58e-220 |
| 99.7 | 9.23e-201 |
| 99.7 | 1.44e-214 |
| 99.7 | 1.18e-213 |
| 99.7 | 1.25e-230 |
| 99.7 | 1.28e-209 |
| 99.7 | 8.84e-218 |
| 99.7 | 4.94e-202 |
| 99.7 | 1.61e-210 |
| 99.7 | 5.34e-202 |
| 99.7 | 1.31e-232 |
| 99.7 | 1.50e-286 |
| 99.7 | 1.27e-203 |
| 99.7 | 1.76e-220 |
| 99.7 | 5.41e-211 |
| 99.7 | 0.0 |
| 99.7 | 6.33e-217 |
| 99.7 | 1.25e-230 |
| 99.7 | 2.73e-227 |
| 99.7 | 5.34e-202 |
| 99.7 | 4.94e-202 |
| 99.7 | 1.79e-220 |
| 99.7 | 2.57e-209 |
| 99.7 | 3.58e-220 |
| 99.7 | 5.43e-216 |

| | |
|---|---|
| 99.7 | 1.50e-286 |
| 99.7 | 5.41e-211 |
| 99.7 | 0.0 |
| 99.7 | 6.57e-234 |
| 99.7 | 1.89e-215 |
| 99.7 | 1.93e-199 |
| 99.7 | 2.73e-227 |
| 99.7 | 3.00e-208 |
| 99.7 | 8.98e-210 |
| 99.7 | 8.67e-217 |
| 99.7 | 1.68e-211 |
| 99.7 | 1.86e-280 |
| 99.7 | 1.50e-286 |
| 99.7 | 0.0 |
| 99.7 | 5.42e-212 |
| 99.7 | 1.89e-215 |
| 99.7 | 3.17e-223 |
| 99.7 | 4.83e-222 |
| 99.7 | 7.03e-236 |
| 99.7 | 1.82e-212 |
| 99.7 | 3.54e-223 |
| 99.7 | 6.33e-217 |
| 99.7 | 5.62e-212 |
| 99.7 | 1.86e-215 |
| 99.7 | 1.50e-286 |
| 99.7 | 4.15e-220 |
| 99.7 | 2.30e-212 |
| 99.7 | 0.0 |
| 99.8 | 1.32e-281 |
| 99.8 | 1.32e-281 |
| 99.8 | 0.0 |
| 99.8 | 1.32e-281 |
| 99.8 | 0.0 |
| 99.8 | 1.32e-281 |
| 99.8 | 0.0 |
| 99.8 | 1.32e-281 |
| 99.8 | 1.99e-273 |
| 99.8 | 0.0 |
| 99.8 | 9.61e-298 |
| 99.8 | 1.99e-273 |
| 99.8 | 1.32e-281 |
| 41.4 | 1.35e-50 |
| 86.0 | 6.74e-162 |
| 55.2 | 3.05e-96 |
| 75.2 | 1.27e-155 |
| 79.4 | 5.09e-169 |

| | |
|---|---|
| 44.0 | 5.11e-73 |
| 56.3 | 1.11e-102 |
| 68.0 | 1.42e-134 |
| 59.7 | 2.97e-122 |
| 62.2 | 5.16e-89 |
| 70.8 | 1.57e-139 |
| 76.6 | 4.86e-160 |
| 53.1 | 1.26e-102 |
| 75.7 | 3.06e-143 |
| 79.9 | 4.20e-168 |
| 81.0 | 3.01e-158 |
| 86.9 | 4.33e-184 |
| 37.5 | 2.67e-55 |
| 65.0 | 8.43e-137 |
| 65.4 | 1.76e-120 |
| 65.6 | 7.32e-113 |
| 65.7 | 1.02e-102 |
| 71.2 | 1.76e-142 |
| 71.8 | 6.23e-145 |
| 61.5 | 1.77e-125 |
| 66.8 | 1.12e-121 |
| 66.8 | 3.66e-86 |
| 43.5 | 3.70e-76 |
| 72.4 | 2.13e-141 |
| 68.0 | 3.48e-148 |
| 72.0 | 2.09e-134 |
| 58.1 | 3.89e-97 |
| 68.5 | 1.81e-140 |
| 86.1 | 2.12e-153 |
| 79.8 | 1.16e-143 |
| 69.7 | 9.08e-168 |
| 74.5 | 2.60e-134 |
| 70.5 | 2.28e-115 |
| 82.3 | 5.61e-145 |
| 71.0 | 5.05e-153 |
| 58.1 | 1.01e-95 |
| 85.2 | 3.91e-158 |
| 85.1 | 9.33e-152 |
| 56.7 | 1.00e-88 |
| 59.2 | 3.16e-97 |
| 63.0 | 3.34e-114 |
| 72.8 | 1.81e-131 |
| 60.1 | 3.22e-125 |
| 72.0 | 1.83e-143 |
| 71.5 | 7.74e-194 |
| 73.1 | 1.08e-196 |

| | |
|---|---|
| 88.3 | 5.25e-246 |
| 54.9 | 7.62e-138 |
| 40.1 | 8.89e-67 |
| 72.5 | 6.12e-130 |
| 60.0 | 2.11e-106 |
| 72.6 | 3.55e-144 |
| 81.8 | 1.69e-158 |
| 72.7 | 9.14e-124 |
| 83.7 | 1.93e-165 |
| 54.8 | 6.13e-96 |
| 72.2 | 4.16e-129 |
| 84.2 | 4.84e-160 |
| 77.0 | 3.55e-150 |
| 85.2 | 1.62e-170 |
| 51.4 | 1.25e-88 |
| 53.1 | 2.03e-91 |
| 71.4 | 9.81e-126 |
| 71.3 | 3.45e-129 |
| 36.1 | 2.36e-48 |
| 73.2 | 2.24e-142 |
| 83.1 | 6.56e-156 |
| 76.6 | 2.57e-73 |
| 46.6 | 8.09e-92 |
| 38.5 | 4.10e-53 |
| 63.2 | 1.50e-104 |
| 77.0 | 9.70e-138 |
| 61.5 | 5.66e-125 |
| 55.7 | 1.24e-101 |
| 66.8 | 1.92e-122 |
| 47.5 | 9.58e-78 |
| 79.9 | 2.83e-158 |
| 78.4 | 1.19e-81 |
| 89.6 | 4.12e-164 |
| 82.5 | 1.19e-227 |
| 64.5 | 1.97e-101 |
| 77.0 | 1.98e-150 |
| 82.4 | 3.25e-153 |
| 69.1 | 2.57e-129 |
| 83.3 | 2.82e-160 |
| 63.2 | 1.50e-104 |
| 66.7 | 2.35e-123 |
| 53.5 | 8.21e-106 |
| 39.0 | 7.60e-54 |
| 47.5 | 9.58e-78 |
| 76.6 | 4.86e-160 |
| 70.1 | 6.21e-142 |

| | |
|---|---|
| 82.9 | 7.05e-180 |
| 58.0 | 9.59e-116 |
| 83.7 | 1.05e-227 |
| 72.2 | 9.91e-142 |
| 83.9 | 1.01e-154 |
| 74.4 | 2.76e-143 |
| 56.5 | 7.37e-88 |
| 89.6 | 4.12e-164 |
| 80.8 | 1.03e-129 |
| 51.5 | 2.63e-101 |
| 70.7 | 1.98e-143 |
| 82.5 | 1.19e-227 |
| 87.6 | 3.08e-242 |
| 39.8 | 2.39e-100 |
| 47.3 | 2.43e-70 |
| 59.0 | 6.13e-97 |
| 64.1 | 3.22e-119 |
| 82.4 | 3.25e-153 |
| 86.9 | 9.74e-161 |
| 65.6 | 2.16e-121 |
| 77.0 | 1.98e-150 |
| 87.2 | 1.87e-212 |
| 87.4 | 6.29e-168 |
| 87.4 | 1.26e-221 |
| 87.6 | 7.08e-119 |
| 87.6 | 7.08e-119 |
| 88.0 | 9.49e-182 |
| 76.0 | 1.44e-148 |
| 43.0 | 9.24e-65 |
| 71.3 | 1.37e-124 |
| 89.0 | 1.67e-167 |
| 69.1 | 2.57e-129 |
| 83.3 | 2.82e-160 |
| 90.3 | 4.82e-176 |
| 91.0 | 6.08e-188 |
| 91.1 | 1.59e-154 |
| 91.5 | 4.79e-200 |
| 91.7 | 3.54e-177 |
| 92.5 | 1.06e-178 |
| 92.5 | 7.17e-170 |
| 93.2 | 1.64e-188 |
| 93.2 | 1.64e-188 |
| 93.7 | 2.03e-170 |
| 93.7 | 5.26e-173 |
| 93.8 | 8.41e-177 |
| 94.0 | 1.75e-170 |

| | |
|---|---|
| 94.2 | 6.61e-190 |
| 94.2 | 6.61e-190 |
| 94.3 | 1.75e-163 |
| 94.4 | 3.59e-186 |
| 94.6 | 2.63e-265 |
| 95.0 | 0.0 |
| 95.1 | 1.35e-170 |
| 95.2 | 1.11e-173 |
| 95.2 | 1.11e-173 |
| 95.3 | 1.96e-206 |
| 95.3 | 8.17e-194 |
| 95.3 | 8.17e-194 |
| 95.4 | 1.79e-205 |
| 95.4 | 1.79e-205 |
| 95.5 | 1.06e-151 |
| 95.6 | 5.19e-265 |
| 95.6 | 3.05e-187 |
| 95.6 | 5.19e-265 |
| 95.7 | 1.07e-176 |
| 95.7 | 3.69e-177 |
| 95.8 | 2.73e-168 |
| 96.0 | 4.46e-193 |
| 96.2 | 1.64e-130 |
| 96.4 | 2.69e-268 |
| 96.5 | 3.33e-200 |
| 96.6 | 5.11e-198 |
| 96.8 | 9.07e-198 |
| 97.1 | 1.71e-176 |
| 97.1 | 3.30e-186 |
| 97.1 | 1.26e-185 |
| 97.2 | 3.86e-272 |
| 97.3 | 3.45e-96 |
| 97.3 | 2.46e-206 |
| 97.3 | 2.46e-206 |
| 97.4 | 2.54e-190 |
| 97.4 | 1.84e-184 |
| 97.4 | 1.84e-184 |
| 97.5 | 7.58e-193 |
| 97.6 | 1.96e-178 |
| 97.6 | 2.67e-208 |
| 97.7 | 9.40e-183 |
| 97.7 | 1.86e-121 |
| 97.7 | 3.11e-193 |
| 97.7 | 1.95e-278 |
| 97.8 | 2.30e-194 |
| 98.1 | 6.24e-98 |

| | |
|---|---|
| 98.2 | 2.57e-241 |
| 98.3 | 4.80e-263 |
| 98.3 | 1.26e-201 |
| 98.4 | 1.31e-184 |
| 98.5 | 2.91e-188 |
| 98.5 | 5.14e-280 |
| 98.5 | 4.86e-282 |
| 98.5 | 4.65e-279 |
| 98.5 | 8.91e-197 |
| 98.6 | 1.75e-203 |
| 98.6 | 1.29e-248 |
| 98.7 | 2.60e-98 |
| 98.7 | 7.64e-283 |
| 98.8 | 5.28e-225 |
| 98.8 | 3.75e-181 |
| 98.9 | 4.18e-133 |
| 98.9 | 1.95e-201 |
| 98.9 | 4.18e-133 |
| 98.9 | 6.90e-265 |
| 98.9 | 0.0 |
| 99.0 | 5.67e-204 |
| 99.0 | 0.0 |
| 99.0 | 4.82e-213 |
| 99.0 | 3.51e-208 |
| 99.0 | 4.98e-219 |
| 99.1 | 8.32e-249 |
| 99.1 | 0.0 |
| 99.1 | 5.94e-151 |
| 99.2 | 7.37e-186 |
| 99.2 | 0.0 |
| 99.2 | 5.33e-171 |
| 99.2 | 2.58e-267 |
| 99.3 | 4.87e-99 |
| 99.3 | 3.21e-102 |
| 99.3 | 4.47e-215 |
| 99.3 | 7.67e-212 |
| 99.3 | 4.46e-188 |
| 99.3 | 2.49e-213 |
| 99.3 | 1.25e-202 |
| 99.3 | 1.70e-99 |
| 99.3 | 3.14e-213 |
| 99.3 | 2.14e-195 |
| 99.3 | 7.33e-201 |
| 99.4 | 0.0 |
| 99.5 | 7.37e-110 |
| 99.5 | 0.0 |

| | |
|---|---|
| 99.5 | 8.76e-293 |
| 99.6 | 1.18e-203 |
| 99.6 | 1.45e-194 |
| 99.6 | 3.32e-192 |
| 99.6 | 3.90e-183 |
| 99.6 | 3.32e-187 |
| 99.6 | 1.51e-202 |
| 99.6 | 1.53e-187 |
| 99.6 | 1.18e-203 |
| 99.6 | 1.45e-194 |
| 99.6 | 3.32e-187 |
| 99.6 | 1.12e-189 |
